# Emergence of Novel SARS-CoV-2 Variants in the Netherlands

**DOI:** 10.1101/2020.11.02.20224352

**Authors:** Aysun Urhan, Thomas Abeel

## Abstract

In this study, we analyzed SARS-CoV-2 genomes in the Netherlands, in the context of global viral population since the beginning of the pandemic. We have identified the most variant sites on the whole genome as well as the stable, conserved ones on the S and N proteins. We found four mutations, S:D614G, NSP12b:P314L, NSP3:F106F, to be the most frequent ones that dominate the SARS-CoV-2 population outside of China. We detected novel variants of SARS-CoV-2 almost unique to the Netherlands that form localized clusters, indicating community spread. We emphasize that while SARS-CoV-2 is evolving, and the number of mutations from the reference sequence is increasing, we observe only little diversity in the new variants as we enter the later stages of the pandemic. Our analyses suggest we have diverged away from the current SARS-CoV-2 reference enough that the reference should be re-evaluated to represent the current viral population more accurately. We assert our work provides valuable information on the genetic diversity of SARS-CoV-2 and its local dynamics in the Netherlands, especially for DNA-based diagnostic, therapeutic or vaccine development against COVID-19. We suggest sequence-based analyses should opt for a consensus representation to adequately cover the genomic variation observed.

## Background

In late December 2019, officials had reported the first case of coronavirus disease 2019 (COVID-19) in China, caused by a novel type of coronavirus named severe acute respiratory syndrome coronavirus 2, (SARS-CoV-2) [1]. COVID-19 has consequently led to the global pandemic we are going through at the moment; according a situation report released by the World Health Organization (September 9^th^) there are 27.4 million cases and almost 900,000 deaths in total [2]. SARS-CoV-2 has been placed under the betacoronavirus genus, closest relatives being bat and pangolin coronaviruses [3, 4].

Despite having major commonalities with recent outbreaks of betacoronaviruses, SARS in 2002 and Middle East respiratory syndrome (MERS) in 2012, it is unprecedented not only in its ease of spread but also in the collective effort of several international scientists to investigate and understand the biology of the disease and the virus causing it since the day the first complete SARS-CoV-2 genome sequence had been published [4–7]. Early studies on the SARS-CoV-2 genome has shown its closest relative, in terms of sequence identity, to be the bat coronavirus RaTF13 with over 93.1% match in the spike (S) protein and >96% sequence identity overall [8, 9]. Immediately a reference sequence had been established [10], paving the way for the exponential growth in both the number and the scale of studies on the SARS-CoV-2 genome [11–16].

At the moment, the GISAID database has established the SARS-CoV-2 population consists of six major clades: G, GH, GR, L, S and V [17]. There is a growing number of studies on the genetic variability of SARS-CoV-2 relative to the reference genome [18–21]. From previous viral outbreaks, it is known that as part of the natural evolution of a virus, subpopulations of clades that can affect the severity of a disease emerge and alter the trajectory of a pandemic [22]. It has been reported that while the two major structural proteins, S and nucleocapsid (N) protein are rich in sites of episodic selection, ORF3a and ORF8 had also been shown to carry a lot of mutations [23].

In this study, we investigate the genetic variability of SARS-CoV-2 genomes in the Netherlands, in the context of global viral population with a particular focus on the later stages of the first wave of the pandemic (from early April to the end of May). We have identified the most variant proteins in the SARS-CoV-2 genome, as well as the most frequent mutations in the Netherlands that also showed high dominance in the rest of the world. We found relatively conserved regions in the S and N proteins of SARS-CoV-2. Tracing the viral genome since its first introduction into the Netherlands, we detect novel mutations unique to the Netherlands, which indicates local clusters of viral sub-populations are emerging. Our work provides valuable insights into the SARS-CoV-2 population in the Netherlands that would prove beneficial for tracking routes of transmission through genetic variation, future efforts in primer/probe design in RT-qPCR tests, and development of potential vaccines against COVID-19.

## Methods

Our study of SARS-COV-2 genomes in the Netherlands consists of three main steps: data retrieval, preprocessing and multiple sequence alignment, phylogenetic tree construction and sequence variation analysis. We have also analyzed the global phylogenetic tree of SARS-COV-2 genomes using additional metadata on patients and travel history.

### Data retrieval and preprocessing, and multiple sequence alignment

Complete, high quality (number of undetermined bases less than 1% of the whole sequence) genome sequences of SARS-COV-2 that were isolated from human hosts only were obtained from GISAID, NCBI and China’s National Genomics Data Center (NGDC) on June 13^th^ [17, 24, 25]. The dataset contained 29503 sequences with unique identifiers in total, including the Wuhan-Hu-1 reference sequence (accession ID NC_045512.2). The acknowledgement table for GISAID sequences can be found in Supplementary File 2, and the full list of sequence identifiers for NCBI and NGDC records are provided in Supplementary File 3.

All sequences were aligned against the Wuhan-Hu-1 reference using MAFFT (v7.46) with the FFT-NS-fragment option, and the alignment was filtered to remove identical sequences to obtain 24365 non-redundant genomes [26].

### Sequence variation analysis

In order to determine mutations, the filtered multiple sequence alignment was trimmed to remove gaps from the Wuhan-Hu-1 reference (accession ID NC_045512.2) and used as input to the *coronapp* web application to obtain nucleotide variations [27]. Next, the trimmed alignment was used to cluster genomes according to the nomenclature on GISAID website. We assigned all 29503 sequences to one of the clades S, L, V, G, GH and GR.

### Phylogenetic tree construction

The maximum likelihood phylogenetic tree for the samples in the Netherlands was built using IQ-TREE (v2.05) with GTR model, allowing to collapse non-zero branches, and ultrafast bootstrap with 1000 replicates [28]. A time tree was also constructed for dating branches in IQ-TREE (v2.05) and the final tree was rooted at the ancestral node of S clades in the tree using ETE Toolkit (v3.1.1) [29]. ETE was also used for visualizing tree.

## Results

The global SARS-CoV-2 dataset was filtered considering only the sequence quality, hence we observe a large discrepancy in the distribution of genomes across different countries. Initially, most sequencing effort was concentrated in China and other countries where the outbreak had begun. However, at the time of data retrieval (June 13^th^) the dataset is dominated by samples from the UK, the USA and Australia (Table 1 and Figure 1).

**Table 1.** 20 countries with the largest number of genomes in the dataset.

| Country | Number of genomes |
| --- | --- |
| The UK | 9641 |
| The USA | 7294 |
| Australia | 1398 |
| The Netherlands | 1338 |
| Spain | 886 |
| India | 710 |
| China | 651 |
| Belgium | 645 |
| Denmark | 581 |
| Canada | 560 |
| Portugal | 500 |
| Iceland | 481 |
| France | 376 |
| Sweden | 353 |
| Switzerland | 314 |
| Singapore | 285 |
| Austria | 247 |
| Russia | 218 |
| Germany | 209 |
| Luxembourg | 192 |

**Figure 1.**
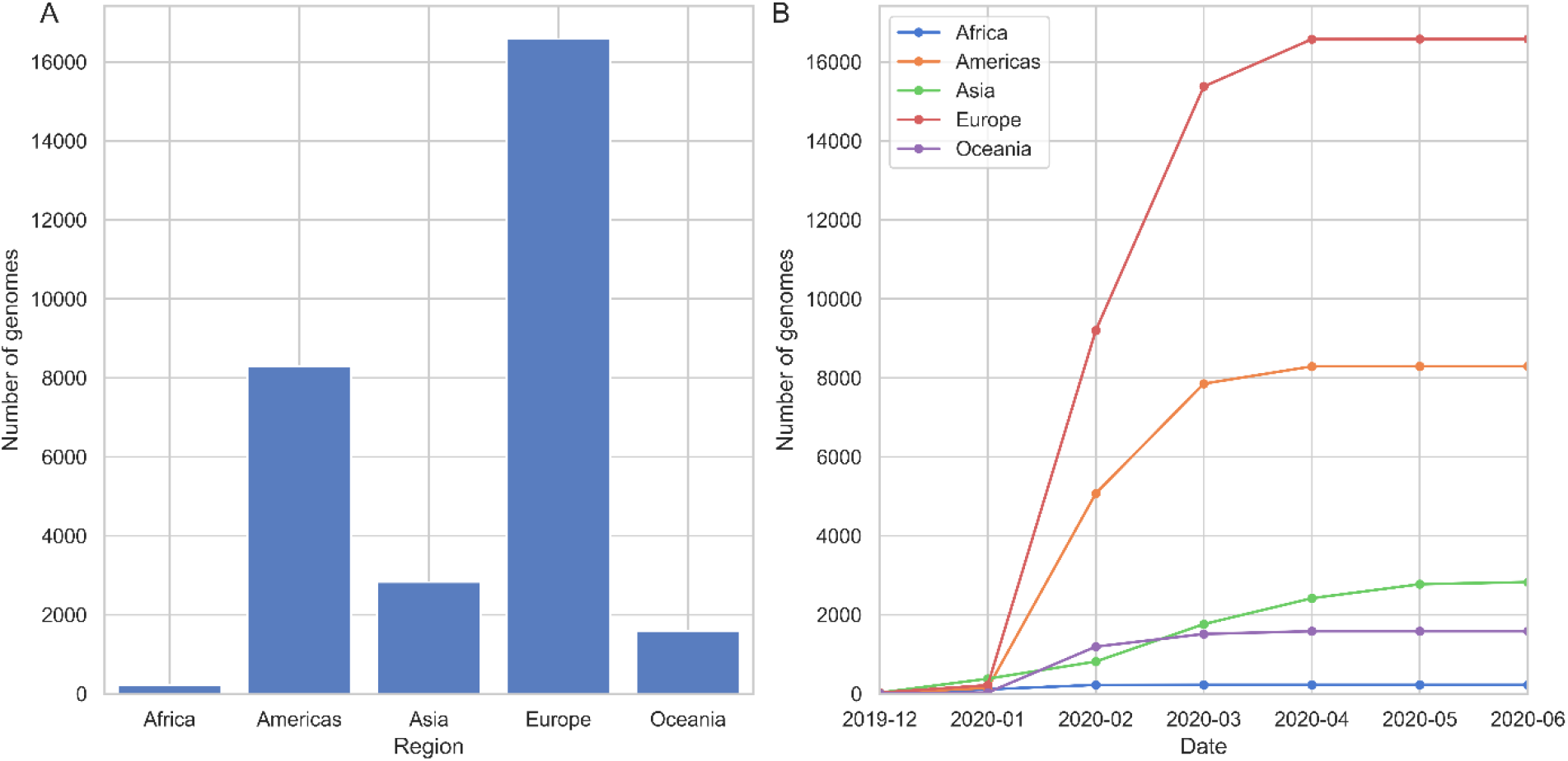
Distribution of SARS-CoV-2 genomes across five continents: total number of genomes is shown in subplot A and the change in number of genomes over the course of pandemic in subplot B. See legend of B for colors of each continent.

Since we have not corrected for sampling differences, in this section, we will provide a view of the current situation of pandemic mainly in Europe, focusing on the Netherlands, where most of the viral genomes are available today (Figure 1). While initially many genome sequences were generated, by April virtually no sequences were determined.

### Distinct genetic patterns in the SARS-CoV-2 population emerge across the globe

In order to get an overview of the viral diversity throughout the pandemic, we used the clade definitions proposed by the GISAID database [17]. We observe the distribution of different clades in the Netherlands resemble that of Australia where the first samples are genetically diverse and there is no dominating variant (subplots in Figure 2). A similar pattern is seen in other European counties such as the UK and Belgium, while the USA, Canada and Denmark have distinct trajectories with GH clade dominating the population (Additional file 1, Figure S3). Also note that clade S has gradually faded out despite its high prevalence before April in several countries, this is particularly noticeable in Australia, China (Figure 2), the USA, Spain and Canada (Additional file 1, Figure S3).

**Figure 2.**
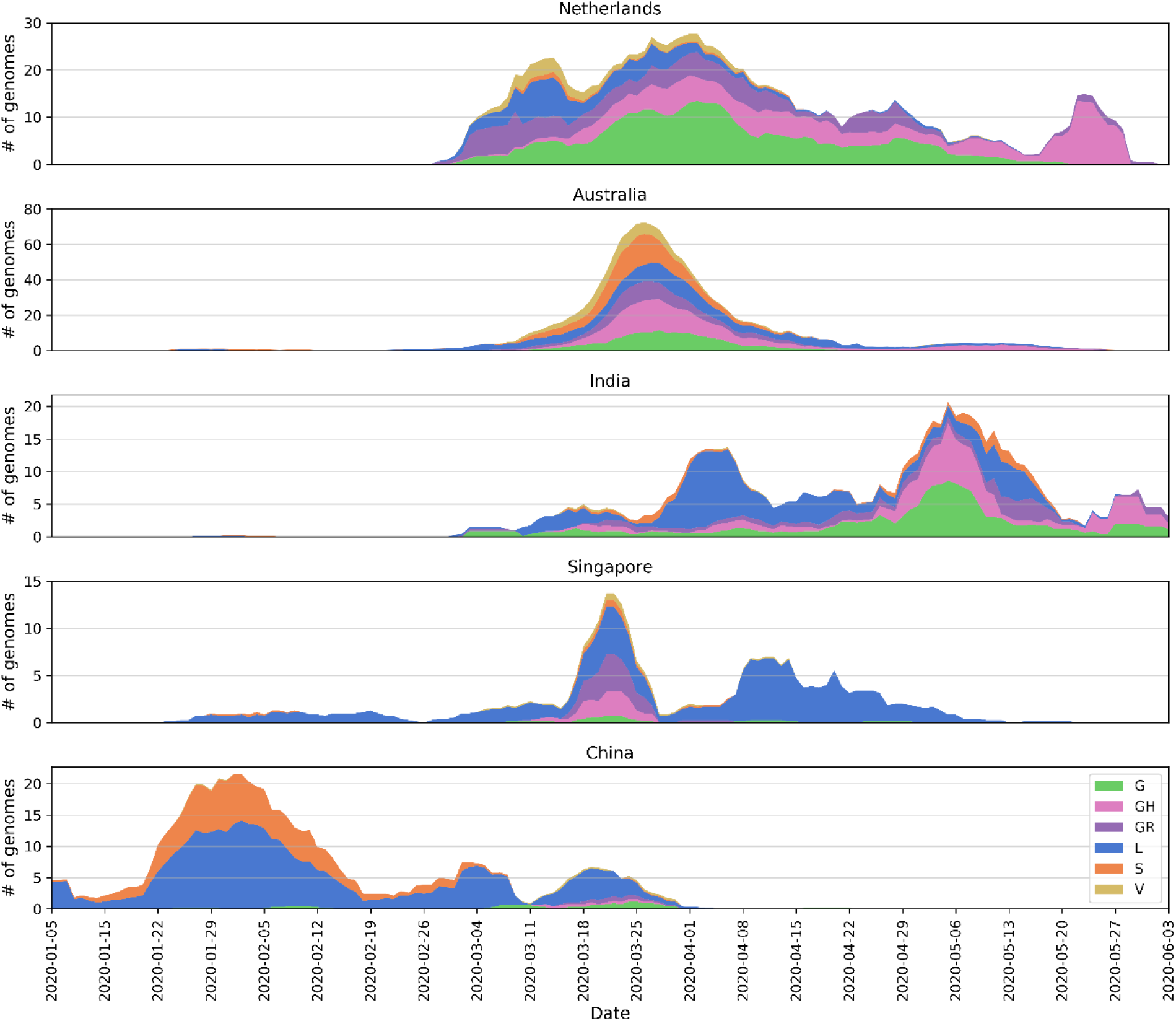
Distribution of SARS-CoV-2 clades in a selection of well-sampled countries in comparison to the Netherlands: moving average over seven days was calculated for six clades (see the legend for clade names and colors) discarding intervals of less than one genome per day.

Viral diversity can be observed more clearly when put into context with less diverse populations in other countries where the outbreak had begun the earliest. For instance, China, Singapore and Italy had experienced the outbreak the earliest in the world, and there are only few of the major clades circulating (Figure 2, Italy not shown due to small sample size, see Figure S3 for other countries). China had opted for possibly the most severe restrictions; similarly in Singapore, the initial cases of COVID-19 had been followed up with strict precautions, preventing both the spread and new introduction of the virus. While it is tricky to formulate any clear hypothesis since there has not been any submission from either country since April, it is certainly interesting to see the contrast between them and countries where COVID-19 arrived at relatively late stages of the pandemic, such as the Netherlands, Australia and India. The composition of viral genomes in these locations appears to be quite diverse, suggesting multiple introductions of virus.

### Evolution of the SARS-CoV-2 genome and increased mutation frequency in hotspot regions

As the pandemic progresses, we observe that across time different mutations dominate the viral population. It is essential to monitor these changes in the SARS-CoV-2 genome to identify conserved sites relevant for designing therapeutics and vaccines, as well as to study the viral evolution during a pandemic. Currently, each new sample has on average around 10 mutation sites in total compared to the Wuhan-Hu-1 reference (accession ID NC_045512.2) in the Netherlands where the trajectory has been in parallel with those in Europe and the world (Figure 3). Clearly showing a divergence away from the original reference.

**Figure 3.**
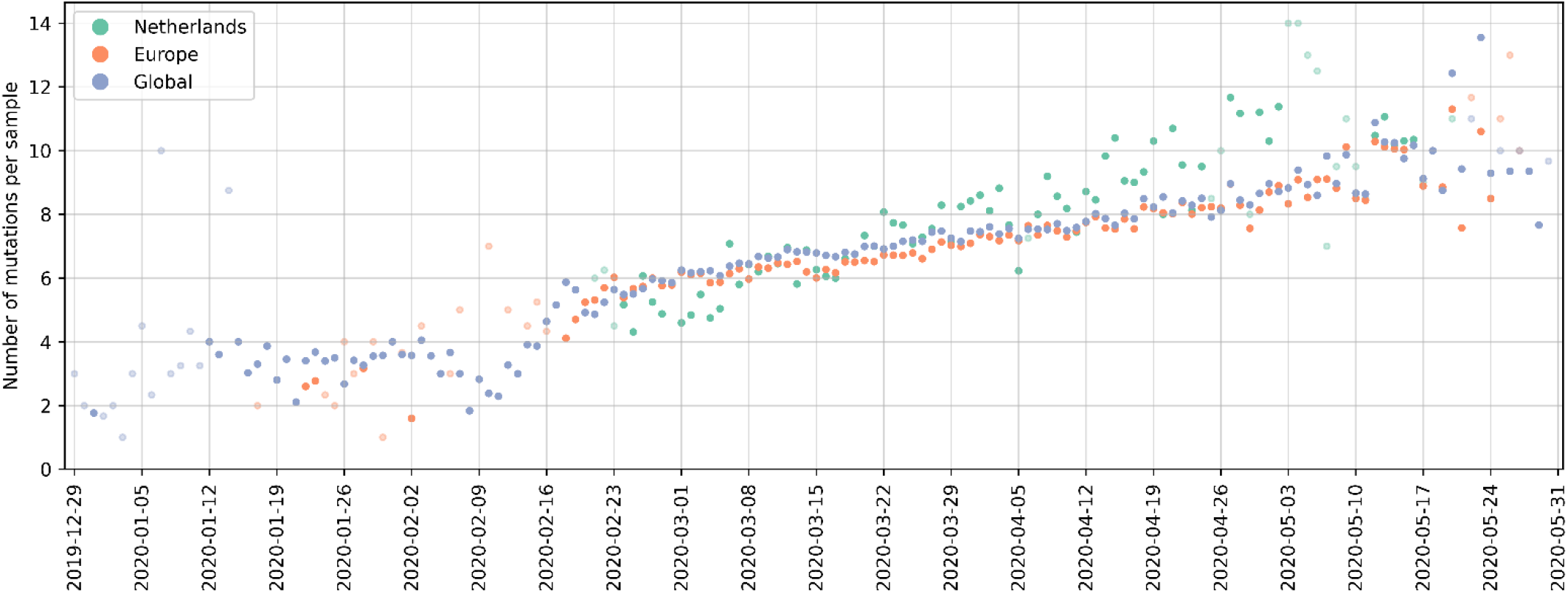
Number of mutations per sample over the course of pandemic in the Netherlands (green), Europe (orange) and globally (blue): data points corresponding to dates with fewer than five samples are faded on color to indicate uncertainty.

In particular, the S and N proteins have both been reported as the most variant proteins in the SARS-CoV-2 genome [23, 30]. S:D614G and N:RG203KR amino acid changes comprise a large fraction of the mutation in these regions, (Figure 4); former being one of the mutation that defines G, GR and GH clades. Apart from carrying the majority of mutations observed in the populations, both proteins play an important role in RT-qPCR based diagnostic tests as well as vaccine and drug development [31]. The S protein has been investigated in great detail for its significance in binding to the host cell and a potential target for COVID-19 treatment and vaccine design [32–34].

**Figure 4.**
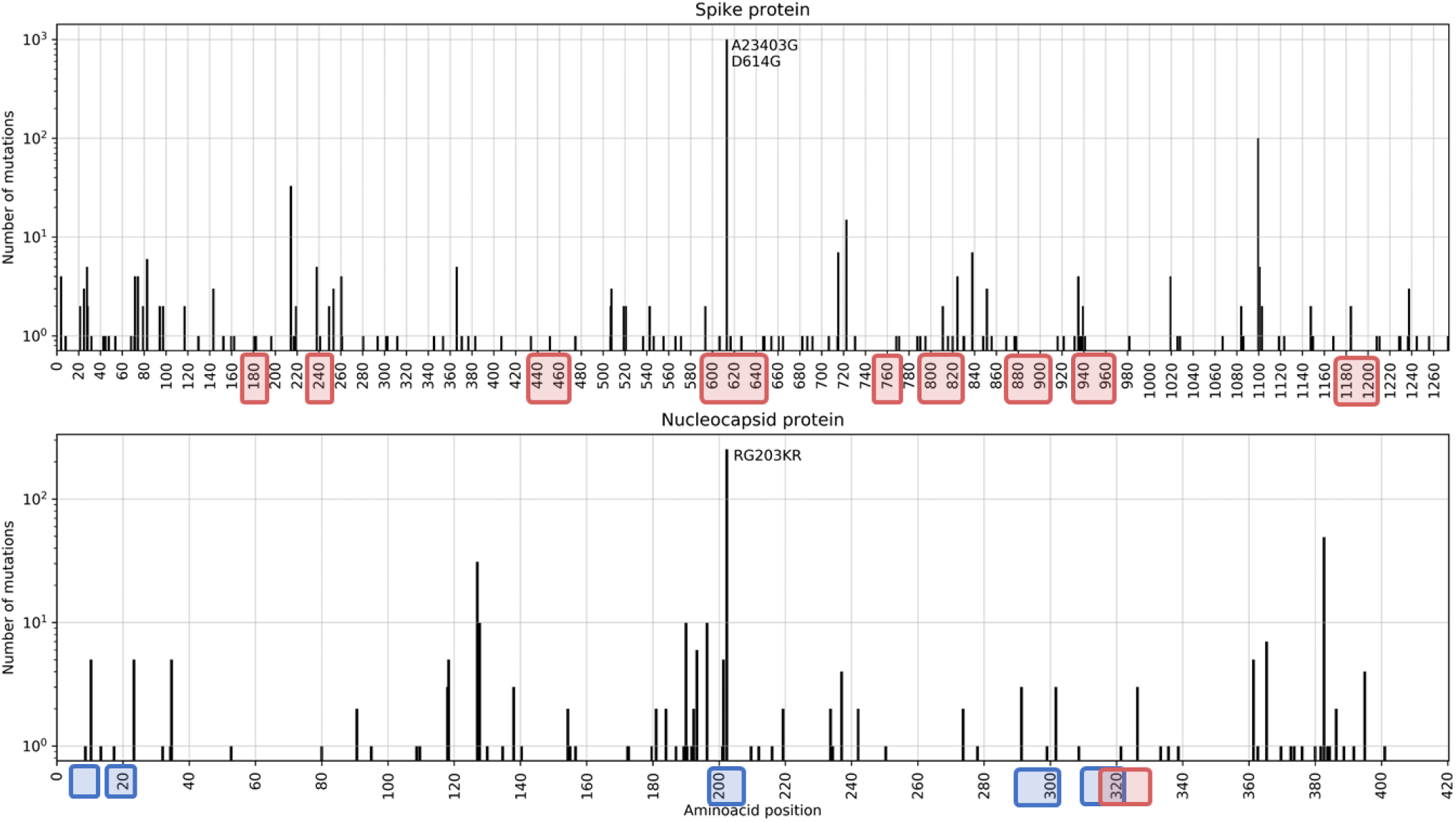
Total number of mutations in the S (top) and N proteins in samples from the Netherlands; predicted epitope regions from [37] and the recommended primer/probe sequences for RT-PCR diagnostic assays are shown in red and blue rectangles, respectively.

Prevalence of S:D614G variant has steadily increased over the course of the pandemic: it is observed in all the sequences sampled recently in the world (Additional file supplementary figure S4). The location of origin of this variant is still unclear, and its implications for humans infected with COVID-19 remain unknown; however, it appears to be very rare in China despite its ubiquity in rest of the world (discussed further in the following section). Note that there have been no sequences released publicly from China since April, making it more difficult to interpret (bottom subplot for China in Figure 2). Thus, it is also likely to be the result of a founder effect commonly observed in viral populations, when the virus spreads very rapidly once in introduced through only a few members of the whole population [35]. Moreover, a study on 453 COVID-19 patients in Sheffield, UK failed to detect any significant link between the trajectory of disease in humans and presence/absence of S:D614G mutation [36].

In order to determine the appropriate primers to use when diagnosing patients with RT-PCR tests or when designing novel primer/probe sequences, variations in the nucleotide sequence should be considered since it plays a crucial role in achieving accurate tests [10, 20]. In light of these, in Figure 4 we have highlighted the predicted epitope regions from [37] and the locations of RT-qPCR primer and probe sequences currently used in the Netherlands with mutations with red and blue rectangles, respectively. Without being too specific, amino-acid positions from 60 to 80, 160 to 170, 340 to 360 and 400 to 420 appear to be relatively stable sites, free of any mutations and could potentially be utilized as primer sequences. This finding is further supported by recent work that combined prior information on the SARS-CoV S and N proteins, and their known epitopes to identify regions in the SARS-CoV-2 genome that could potentially serve as epitopes for B-cells and T-cells [37]. The authors confirmed that the most abundant mutations in these regions, S:D614G in particular, should be taken into account for vaccine design and development of treatments. The N protein is also recommended as a screening assay by the WHO, and is utilized in many countries including the Netherlands [38].

### Population of SARS-CoV-2 is dominated by four mutations globally while emergence of locally distinct variants indicates local outbreaks

In addition to S:D614G and N:RG203KR, several other mutations, NSP12b:P314L, NSP3:F106F and 5’UTR:241 in particular, appear to dominate the most frequent mutations in the world; Figure 5 shows the 15 most dominant SNPs in some of the most-sampled countries in our dataset. Due to over-representation of few European countries, it is difficult to comment on the geographical dominance of any mutations. However, four mutations, S:D614G, NSP12b:P314L, NSP3:F106F and 5’UTR:241 (blue bars in Figure 5) are established within the global collection genomes, except for China where these mutations have very low frequencies.

**Figure 5.**
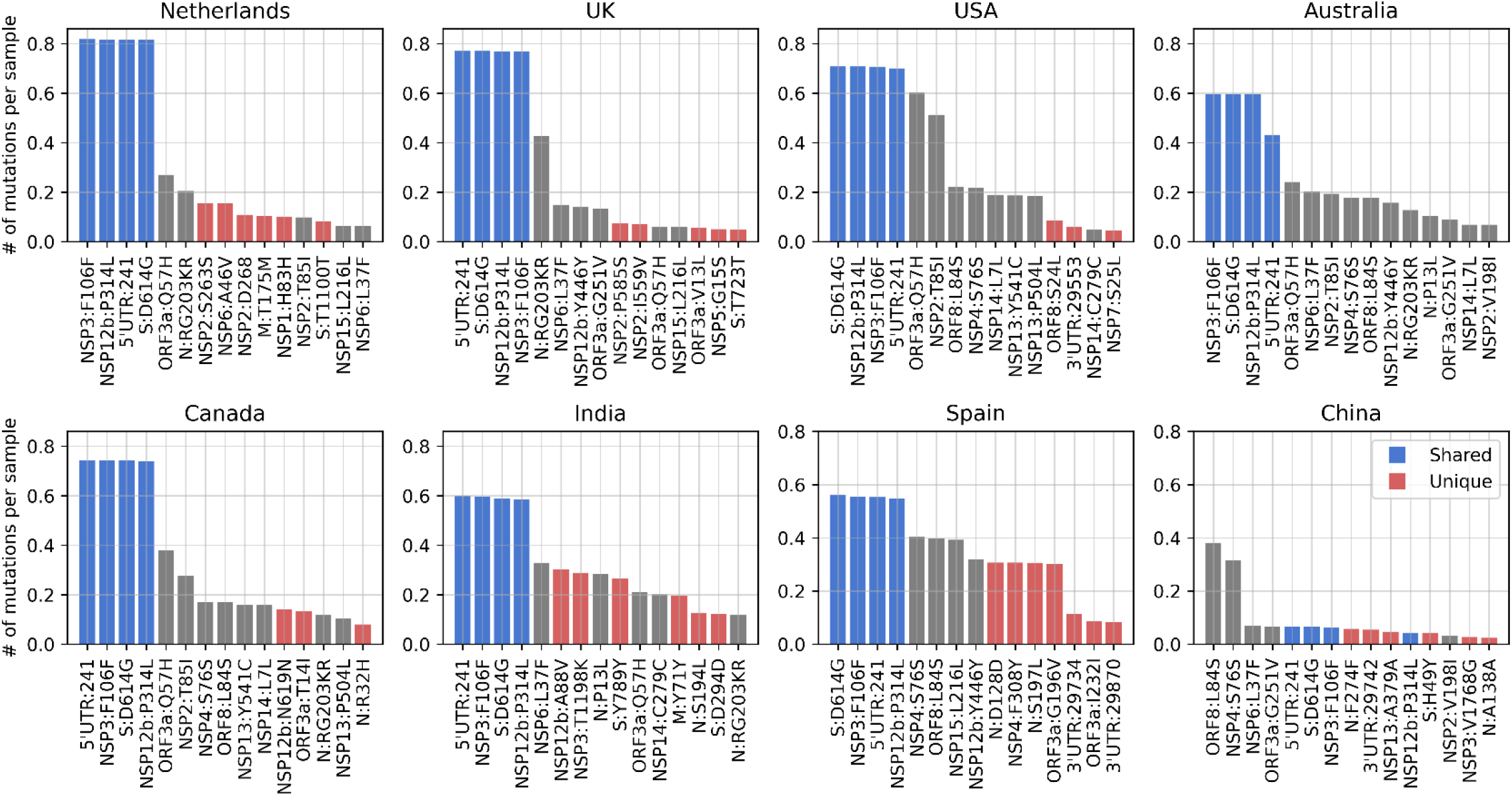
Total frequency (number of mutations per sample) of the top 15 mutations in the most-sampled countries in our dataset: blue bars are mutations shared across all these countries in the top 15, while the red bars are unique to that one country in the top 15.

While we observe a diverse mutational landscape in Australia, India and Spain, Viral population in China has remained relatively homogenous and with very few variants compared to the Wuhan-Hu-1 reference. The most frequent mutation is ORF8:L84S, which defines the S clade that appears to be fading out even though it had been circulating since the beginning of the pandemic along with the L clade. Recently, a possible link between two mutations, ORF8:L84S and NSP4:S76S, has been suggested as they co-occur several times outside of Europe; in China, the USA, Australia and Canada [42]. We also note a few region-specific mutations: first one being ORF8:L84S, which is more frequent in the USA and China and, second is NSP6:L37F which is frequent in in Australia and the USA.

Considering the fluctuations in rate of sequencing, and over-representation of samples from the USA, the UK and Europe in general, it is difficult to comment on the geographical spread. Nevertheless, when we look into the frequency of the top four mutations, S:D614G, NSP12b:P314L, NSP3:F106F and 5’UTR:241 over the course of pandemic (Figure 6), we see a steady increase of their abundance in the viral population, regardless of the date of introduction in each country.

**Figure 6.**
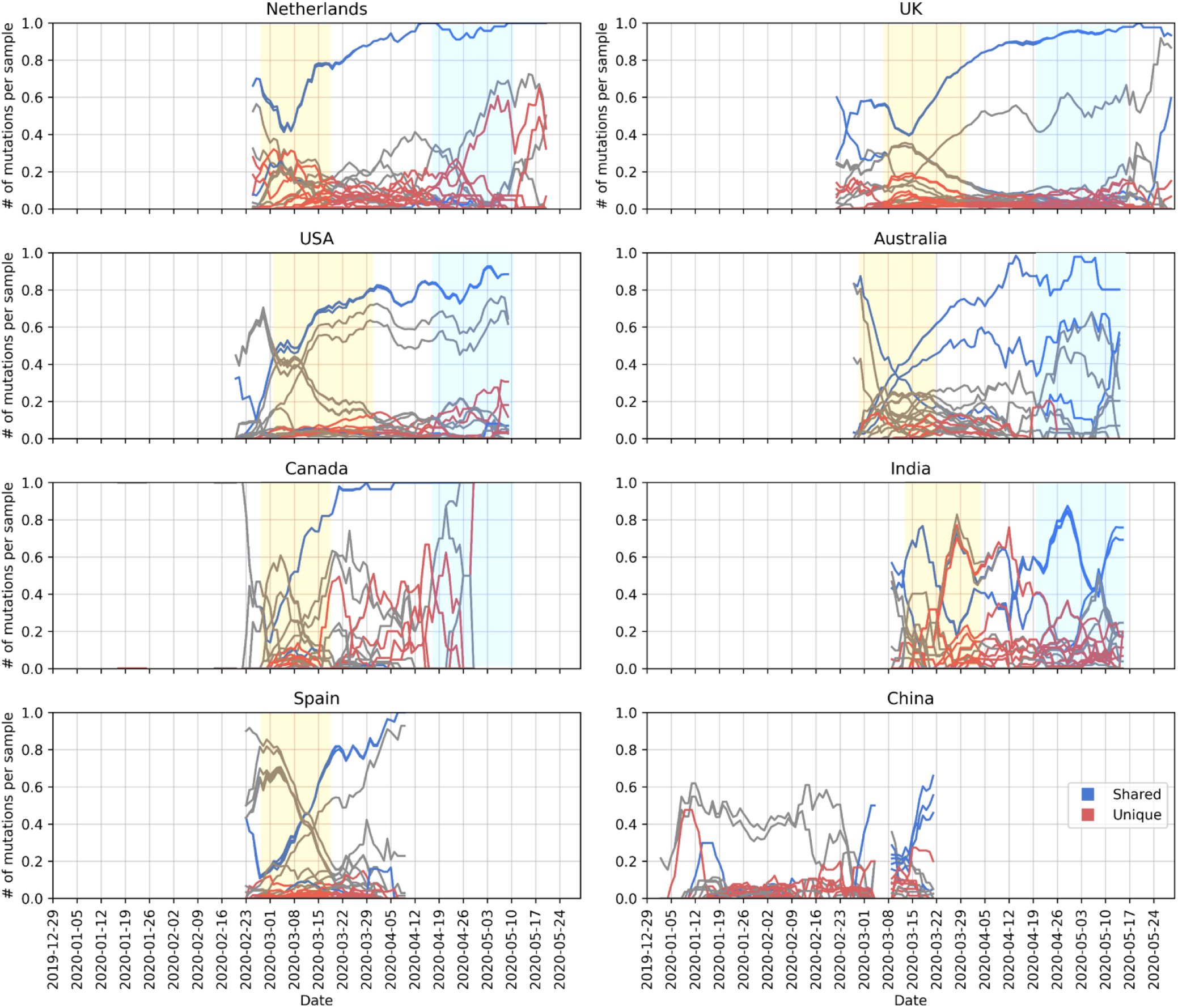
Change in frequency of the top 15 mutations in the most-sampled countries in our dataset, averaged over a period of 7 days and removed periods with less than one sample per day. Line colors were kept consistent with Figure 5: blue lines are mutations shared across all these countries in the top 15, while the red lines are unique to that one country in the top 15. Areas highlighted in yellow and blue as, mentioned in text, to indicate pre-lockdown and post-lockdown.

A common pattern emerges in the frequency change of shared and rarer mutations: before the introduction of lockdown the viral population is very diverse with unique mutations indicating local clusters (areas highlighted in yellow in Figure 6: first two weeks of March in the Netherlands, Australia, Canada and Spain, and end of March for the UK, the USA and India). In the absence of travel restrictions, we observe unique and rare mutations increase in frequency (late April in Figure 6, highlighted in blue). The UK and the USA have been among the latest to impose control measures, and we see rare mutations peak several times from February till mid-March. As the pandemic progresses, the four most abundant mutations shared across each country (blue lines in Figure 6) become well-established as part of the viral genome, while lifting the lockdown leads to again an increase in rare mutations (areas highlighted in blue in Figure 6) as they spread and form local clusters of variants. Abundance of small variants suggest community-driven spread, which can be elaborated by monitoring such variants to detect super-spreading events.

### Introduction of COV-19 in the Netherlands and local clusters with high genomic diversity

Next, we examined the Dutch phylogenetic tree to better understand the dynamics of COVID-19 in the Netherlands: from its introduction in the earliest samples to its further spread through localized infection clusters. We have identified multiple points of introduction in different provinces via highly diverse samples of virus. As the pandemic progresses, we see deeper branching in the tree with unique, localized mutations as well as similar patterns of evolution emerge in separate locations. While the virus population carries an increased number of mutations in general, these mutations are localized in their own clusters with little genomic diversity.

We observe two separate sections on the radial tree in Figure 7 below, representing the diversity of introduction to the Netherlands in terms of both the viral genome and location. First, at the top, starting from around 12 o’clock to 3 o’clock consists of some of the earliest samples from early March of V, L and S clades (denoted with a blue arch and text “Early March”). This is further broken down into four sections numbered from 1 to 4 where the second section is the early outbreak in Noord-Brabant in parallel with the first case reports. However, the remaining sections are mixed in location and date as we encounter samples isolated from Limburg, Zuid-Holland, Gelderland and Utrecht, also from later into the pandemic in late March and early April.

**Figure 7.**
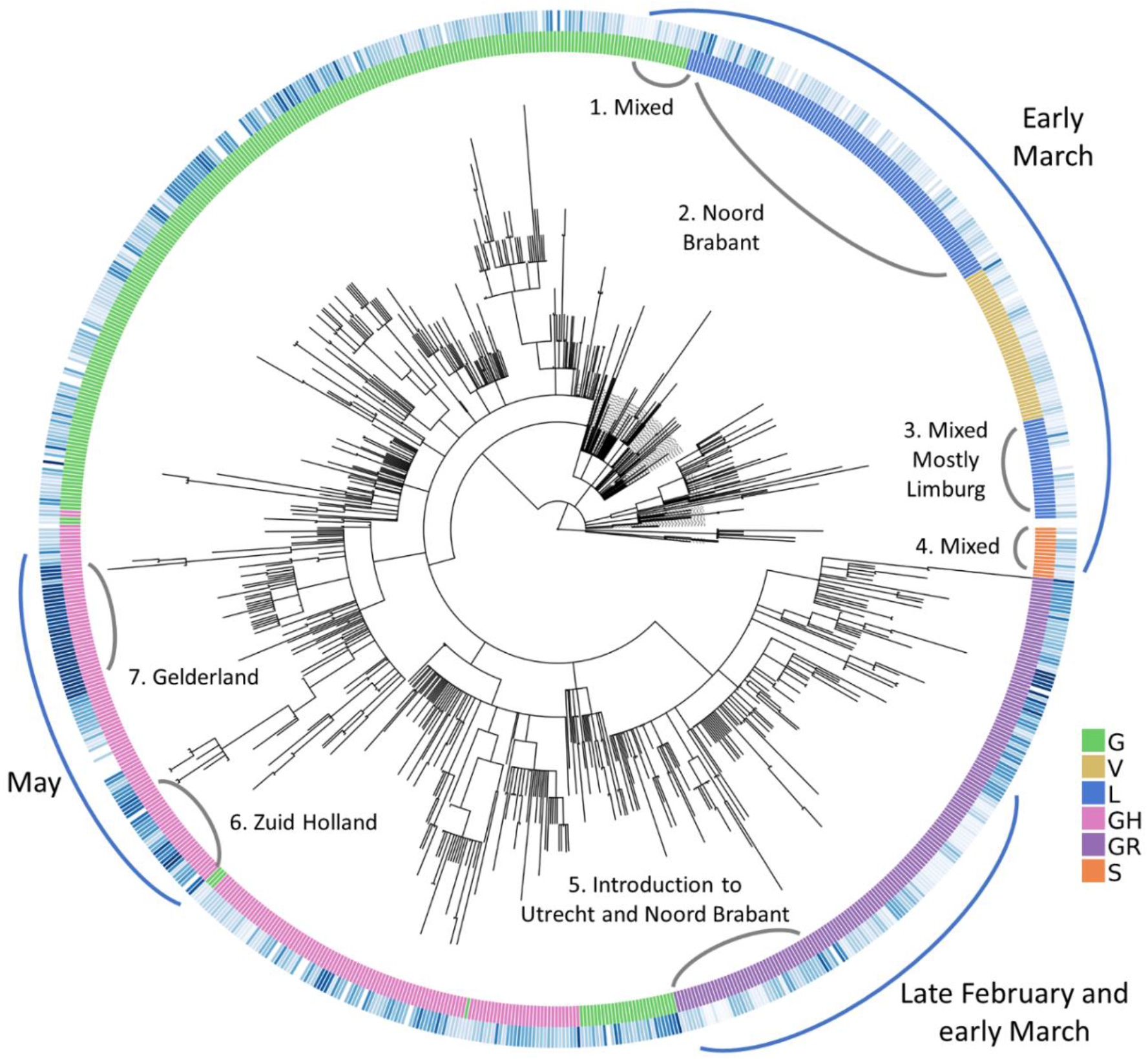
Radial representation of the Dutch phylogenetic tree: inner circle colored w.r.t. the assigned clades (see legend for clade names), outer circle is color-coded according to the sample collection date (if available), where the darker shade of blue represents more recent samples. Major points discussed in the text have been indicated with blue arches on the outer circle, along with more detailed information (numbered in clockwise direction) in gray arches on the inner circle.

The second point of introduction is from 4 o’clock to 6 o’clock on the tree, denoted as “late February and early March” with a blue arch. This section differs from the first one in that we observe only samples of G and GR clades, both of which are dominant in the Europe while absent in China. The earliest SARS-CoV-2 genome in our dataset with full sample collection date (accession ID EPI_IS_454750, collected on February 27) is also located in this section and it was first isolated in Utrecht.

Recall the clade distribution over time in the Netherlands (Figures 2 and 3) showed an initial phase of high diversity with L and GR dominating the dataset, also supported by the phylogenetic analysis. As part of the Dutch initiative to investigate transmission of COVID-19 in the Netherlands, Munnink et al. had conducted a detailed analysis on the earlier samples with patient data [11]. More recently, Sikkema et al. have published their findings on COVID-19 infection in health-care workers in early March [43]. Their studies suggest multiple introductions from Italy and Switzerland, as well as localized community transmissions in super-spreading events in late February and early March. In addition, the authors note the diversity of early strains even for patients with similar travel histories, also in parallel with our observations in our study. In addition to Noord-Brabant, Munnink et al. had detected local clusters in Zuid-Holland and Utrecht.

### Novel mutations appear in the later phase of pandemic

Munnink et al. have stated three phases of response to pandemic in the Netherlands in their study; (i) before the first case was reported, (ii) from the first reported case to the start of screening of healthcare workers and (iii) the period from the introduction of stricter measures along with events and large gatherings of people being banned until March 15^th^ when the most strict phase of lockdown had begun as retail and catering industries were closed, as well as schools and childcare centers [44]. Since March 15^th^, the spread of COVID-19 has been very limited due to more stringent measures on travel and widely adopted practice of social distancing. For this reason, it is particularly interesting to investigate the deeper branching in Figure 7 with later samples around 8-9 o’clock (denoted with a blue arch and the text “May”).

Below in Figure 8, we have zoomed into the two “May” regions from Figure 7 (numbered 3 and 4 in Figure 8) as well as the remaining deep branches (numbered 1 and 2 in Figure 8). To simplify, we have indicated the absence/presence of a mutation with a circle where the branch ends. Additional information about sample collection date and its location are also displayed aligned to the leaves, if available and in the case of duplicate sequences separated with a semicolon. Dates are expressed in format month-day. The large squares next to the leaf names are color-coded clade assignments, colors have been kept consistent throughout our study in Figures 2, 7 and 8.

**Figure 8.**
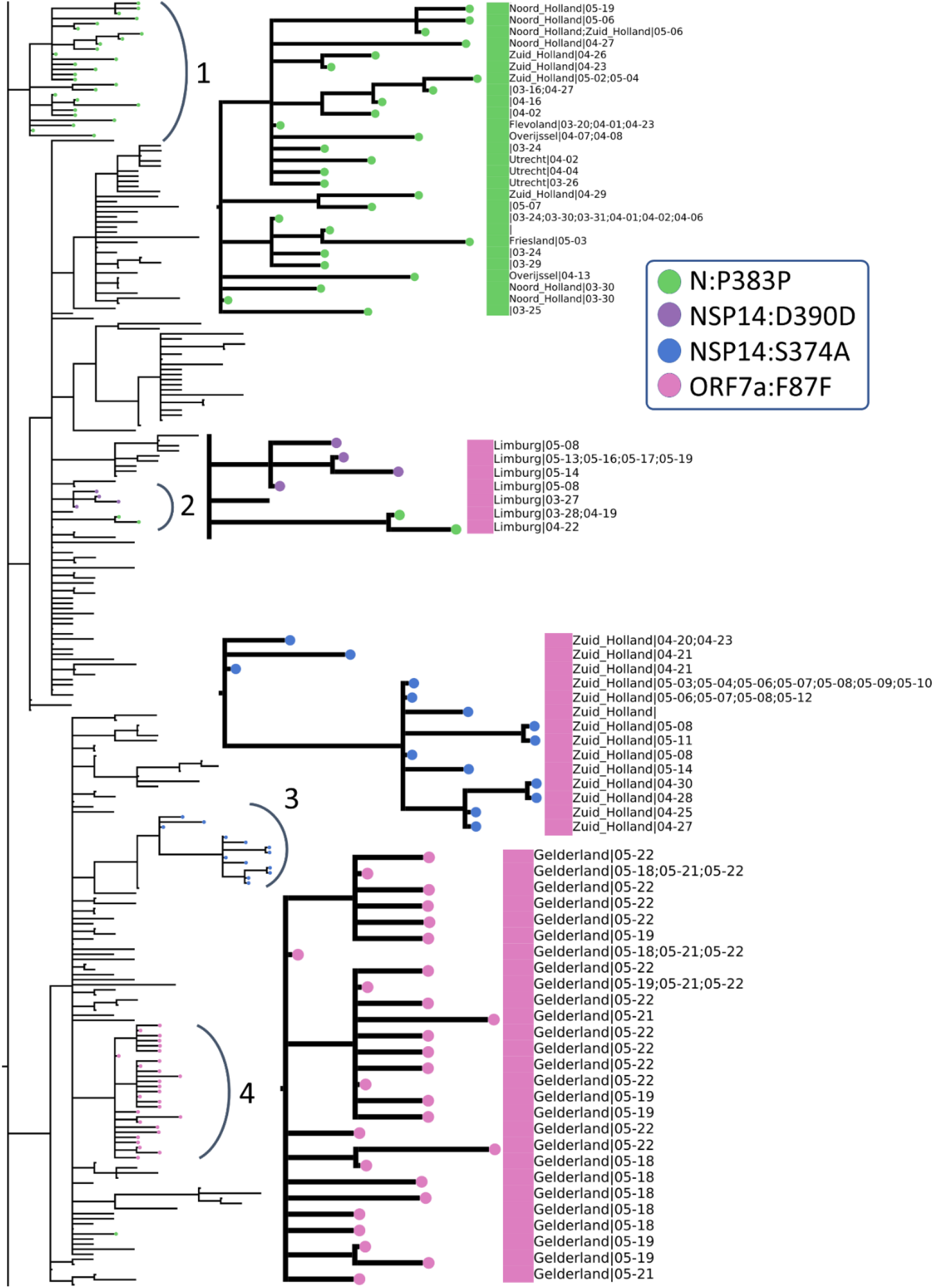
Zoomed-in view of rectangular representation of the Dutch phylogenetic tree: three regions of focus are numbered next to the corresponding arch. Newer, unique mutations that define deep branching in the tree are drawn in circles and the common mutations within Europe are rectangle (see legend for mutation annotations). Assigned clades are indicated with large rectangle aligned next to the leaves (pink: GH and green: G) and additional information about sequences (location and sequence collection date) are displayed next to the clade color, if available.

We have identified four mutations all of which have emerged after March 15^th^ and have led to deeper branching on the phylogenetic tree and are either unique to the Netherlands or very rarely observed in the rest of the world: N:P383P, NSP14:D390D, NSP14:S374A and ORF7a:F87F. These rare mutations could be further utilized to track local transmissions of disease within the Netherlands.

N:P383P (green circles), a silent mutation on N protein is fairly unique to the Netherlands; it is present in less than five sequences in many European countries, including the most well-sampled ones Denmark and the UK, as well as the USA and Canada. Considering the sample size, it is surely intriguing that this mutation has been observed only in the Netherlands in such abundance. This mutation is also one of the oldest circulating ones since its first occurrence was in a sequence from the Switzerland on February 27^th^. However, we observe it for the first time in the Netherlands two weeks later on March 16^th^ (province unknown). Later on, the same mutation has appeared in multiple provinces, Noord Holland, Zuid Holland, Flevoland, Utrecht and Limburg, in 50 sequences in total. Moreover, we observe it in two separate branching events in the phylogenetic tree in different provinces of the Netherlands; several provinces in arc number 1 and only in Limburg in arc number 3. In a recent study, this mutation had been detected as one of several homoplasies on the SARS-CoV-2 genome [45]. Since the Limburg branching contains only three sequences carrying the mutation, it is difficult to comment whether it is convergent or not. Given that branching with number 1 contains several provinces; it is also likely that this is a consequence of relaxations in domestic travel restrictions, rather than convergent evolution.

The second mutation, NSP14:D390D (purple circles), is tricky to interpret because it is present in only 9 genomes, 7 of which had been sequenced in the Netherlands and the remaining two in the UK. It has first appeared in the UK on March 24^th^, during strict lockdown conditions, and it has emerged in the Netherlands in May. We hypothesize this is a small cluster of variants genomes, localized in Limburg only and it has not found the chance to spread outside of the province yet.

NSP14:S374A (blue circles) is the only non-silent mutation in this list, and is very unique to Zuid Holland; it is present in 35 genomes in total, all collected in Zuid-Holland region within three weeks. Similar to NSP14:D390D, it is highly likely to be a small, contained cluster of individuals.

ORF7a:F87F (pink circles) is also incredibly rare since it was observed only in Gelderland in the Netherlands from late April to early May, and less than five times in any other country. It occurs in only one sequence from Canada in April 13^th^, twice in the USA in late March and four times in the UK in mid-April.

## Discussion

In this work, we retrieved 29,503 complete, high quality SARS-CoV-2 from publicly available databases to explore the viral population diversity In the Netherlands, within a global context. Considering the rapid increase in public data and research on this subject, our work is among the more comprehensive ones to lend insight into the genetic variation of SARS-CoV-2 in the later stages of the pandemic in April and early May.

As a consequence of the natural evolution of a virus, SARS-CoV-2 genome has been diverging from the initial reference sequence Wuhan-Hu-1 established based on viral samples from Wuhan, China. The six major clades designated by GISAID had varying distributions in different regions, at different points of time through the course of pandemic. We demonstrated that in most countries, viral population goes through an initial phase of high diversity followed by a decline in genetic variety in which the population is comprised of mostly G, GR or GH clades (Figure 2). With increased ease of travel, COVID-19 was able to spread rapidly across the world and several studies had reported multiple introductions of a diverse viral population into many countries outside of China that lends itself to a more homogeneous population diverged from the Wuhan-Hu-1 reference [11, 16, 46, 47]. Interesting, we have also observed that China and Singapore, both of which are countries that experienced the outbreak the earliest, harbor a markedly different viral population that remains mostly homogeneous with L being the dominant clade that also includes the Wuehan-Hu-1 reference (Figure 2). Note that this could also be the artifact of the dramatic decline in number of sequences from China.

The S and N proteins in SARS-CoV-2 genome has received much attention; both have been reported as the most variant proteins [23, 30] and are also significant in RT-qPCR based diagnostic tests as well as vaccine and drug development [31]. We have identified the most variant sites on the S and N proteins in sequences from the Netherlands (Figure 4). Koyama et al. had noted the effect of these variants on sequence-based vaccine and therapeutics against COVID-19 [37]. Following their discussion, we highlight their predicted epitope regions derived from SARS and the mutations we detected on the S and N proteins in Figure 4. In addition, Kim et al. discussed variations on SARS-CoV-2 genes targeted by diagnostic assays in [21]. Similarly, we analyzed primer/probe sequences currently in use in the Netherlands for diagnostics targeting S and N genes (Figure 4); while the specific implications of mutations on the accuracy of diagnostic tests is unknown, we reported the amino acid positions on N gene from 60 to 80, 160 to 170, 340 to 360 and 400 to 420 to be relatively stable, hence suitable for primer design in RT-qPCR tests.

When we observed the global landscape of variants, we found four mutations, S:D614G, N:RG203KR, NSP3:F106F and 5’UTR:241, are not only the most frequent ones, but also have been steadily increasing in the frequency outside of China since the beginning of pandemic. The implications of these mutations thriving in the population individually are currently unknown, speculative at best. However, some studies have suggested certain linked mutations which poses a different question on its own [36]. We also reported the increase in frequency of these shared mutations, regardless of the date of introduction (Figure 6). On one hand, the abundance of these mutations might suggest that viral genome has converged to a new variant, different than the Wuhan-Hu-1 reference. On the other hand, since most of the viral sequences are from diagnostic tests performed on hospitalized patients at the moment, we are looking at only a small portion of the whole virus population in humans and we do not know clearly whether milder, or even asymptomatic cases of COVID-19 also carry these mutations or not. To our knowledge, studies have not found any significant correlation between these specific mutations and the COVID-19 disease in patients [36]. Nevertheless, it is surely interesting consider that these four mutations, linked to one another, might also influence the infection in the human host.

We have analyzed the most frequent mutations around the world, and found that. Since the rate of sequencing has been fluctuating dramatically and the dataset has been over-represented by the UK, the USA and Europe in general, it is not possible to tell if any change in frequency of mutations over time is caused by the unbalanced sampling or evolutionary advantage/disadvantage.

With our phylogenetic study in the Netherlands, we confirmed multiple introductions in distinct provinces as well as the population diversity in the initial samples (Figure 7). We also detected emerging local clusters, defined by four mutations, N:P383P, NSP14:D390D, NSP14:S374A and ORF7a:F87F, all of which are either entirely unique to the Netherlands or very rarely observed elsewhere (Figure 8). N:P383P had occurred at two distinct sections in different regions, we presume this is likely a domestic travel event rather than a convergent mutation. We note the detection and monitoring of such unique mutations could be utilized for tracking the spread of virus and identifying possible routes of transmission during the outbreak. In addition, our findings are in line with previous studies in the Netherlands by Munnink et al and Sikkema et al.; they had also observed sequence diversity in the earliest days of the outbreak as well as community transmission [11, 43].

The single most prominent pattern that we encountered in our study was that despite the continual increase in number of mutations in the genome, diverging away from the Wuhan-Hu-1 reference, there is little diversity in the new variants as we enter the later stages of the pandemic. This suggests the current SARS-CoV-2 reference genome should be re-evaluated, perhaps replaced with a new one that represents the viral population more accurately. Further work is required to investigate implications of an inadequate reference in sequence-based analyses as well as develop alternative models. Having a good quality reference sequence is crucial in sequence-based analyses, and we assert this line of research will continue to supplement the global effort to fight COVID-19.

The major limitation of our study is the biased dataset of SARS-CoV-2 sequences. Despite our efforts to combine all genome sequences publicly available up to date, due to imbalanced sampling and dramatic changes in the frequency of genome sequencing, our dataset is over-represented by samples from the Europe and the USA and there are several gaps in time since the beginning of pandemic. In addition, most of the viral sequencing today is performed on hospitalized patients. These issues could be circumvented to some extend by stratified sampling or controlled sequencing efforts with random samples collected from individuals. Nevertheless, our findings are significant to understand the viral population diversity within the Netherlands from late March to early May, where our dataset has the most coverage.

## Availability of data and materials

Full list of sequence identifiers, and the corresponding acknowledgements for the sequences used in this work are provided in the Supplementary File 2 and Supplementary File 3.

## Conclusions

In this study, we have analyzed 29503 SARS-CoV-2 genomes retrieved from public databases to investigate genetic diversity in viral population as the pandemic progresses, with a focus on the Netherlands, in particular. Our dataset contained 1338 genomes from the Netherlands, most of them sequenced in the later stages of pandemic in April and early May. We assert our work provides valuable information on the genetic diversity of SARS-CoV-2 and its local dynamics in the Netherlands for tracking the transmission of COVID-19, as well as DNA-based therapeutic or vaccine development against COVID-19, and primer/probe design in RT-qPCR tests. We emphasize the little diversity observed globally in recent samples despite the increased number of mutations relative to the established reference sequence, suggesting the current reference may not be representative of the population; potential implications of an inadequate reference on downstream analyses should be investigated.

## Data Availability

All data is available in the supplementary files.

## Acknowledgements

We thank all the researchers, authors, originating and submitting laboratories of the sequences on GISAID EpiFlu™ database [17]. Full list of sequence identifiers, and the corresponding acknowledgements for the sequences used in this work are provided in the supplementary file.

## Author contributions

A.U. wrote the main manuscript text, prepared the figures and tables in the main manuscript and additional files.T.A. supervised the project. All authors reviewed the manuscript.

## Ethics declarations

### Ethics approval and consent to participate

Not applicable.

### Consent for publication

Not applicable.

## Additional Files

Supplementary File 1: supplementary-file-1.pdf

Document contains Supplementary Text: Annotation of mutations further elucidate conserved regions and show a general preference of non-silent changes in the genome, and Supplementary Figures: Figure S3 and Figure S4.

Supplementary File 2: supplementary-file-2.pdf

Document contains the acknowledgement table for the sequence records retrieved from the GISAID EpiFlu™ database.

Supplementary File 3: supplementary-file-3.pdf

Document contains the list of sequence records retrieved from the NCBI and NGDC databases.

