## Supplementary material for "Emergence of Novel SARS-CoV-2 Variants in the Netherlands": supplementary-file-1.pdf

### Supplementary Text: Annotation of mutations further elucidate conserved regions and show a general preference of non-silent changes in the genome

We have characterized and annotated point mutations in the SARS-CoV-2 genomes sampled within the Netherlands. A large portion of these mutations are found in the S and N proteins (subplots A and B in Figure S1, number of unique mutations and the total number of mutations at the top of each bar); overall NSP3, NSP12b, S and N proteins carry a majority of the mutations (protein names in orange color in Figure S1). To classify the mutations, we follow a similar nomenclature to *coronapp*'s: SNPs leading to a change in the amino acid sequence are non-silent, SNPs with no amino acid change are silent and SNP stop denotes SNPs where a stop codon is introduced.

Most variant sites, in terms of total number of unique mutations, also favor non-silent SNPs. More than half of unique mutations in NSP1, NSP5, NSP6, NSP7, NSP8 and NSP9 are non-silent (orange parts of bars in Figure S1A). According to a recent study analyzing SARS-CoV-2 proteins in terms of their codon usage, S, N, NSP7 and NSP8 proteins might play a critical role in adaptation to their host since they prefer a smaller, more "optimized" set of codons compared to the rest of the proteins [1].

Since there are only few dominant mutations in each protein, the percent breakdown becomes more skewed when the total number of mutations is considered in Figure S1B. More than 80% of total nucleotide substitutions in NSP3 are silent, whereas substitutions do cause a change in the amino acid sequence in more than 60% of the instances on the other proteins (compare blue and orange bars in Figure S1B). However, we have not observed a significant change in the relative frequency of silent and non-silent SNPs over time (Figure S2).

In terms of total number of mutations, NSP7, NSP10, ORF6, ORF7b and ORF10 appear to be the least variable proteins in the SARS-CoV-2 genome (protein names in blue color in Figure S1). ORF10, in particular is not only conserved, but it is also rather unique; there are currently no homologs of ORF10 on NCBI [2]. While studies have shown it is possible for ORF10 to be encoded in pangolin and bat viruses [3], ORF10 is mostly unique to SARS-CoV-2 and could be used as a specific marker.

Table S1. Percent breakdown of unique and total mutations observed in the Netherlands on different proteins of the SARS-CoV-2 genome, total number of unique and total mutations in each protein are also reported. Throughout this report, the term “nonsilent” refers to nucleotide changes accompanied with an amino acid change, whereas “silent” mutations are nucleotide changes with no change in the amino acid sequence.

| | Unique mutations | | | | | Total mutations | | | | |
|---|---|---|---|---|---|---|---|---|---|---|
| Protein | SNP nonsilent | SNP silent | SNP stop | Deletion | Total # | SNP nonsilent | SNP silent | SNP stop | Deletion | Total # |
| NSP1 | 29.2 | 58.3 | 0 | 12.5 | 24 | 6.4 | 90.3 | 0 | 3.4 | 236 |
| NSP2 | 62.0 | 36.6 | 0 | 1.4 | 71 | 34.5 | 44.3 | 0 | 21.2 | 707 |
| NSP3 | 67.9 | 30.9 | 0.6 | 0.6 | 165 | 19.0 | 80.9 | 0.1 | 0.1 | 1542 |
| NSP4 | 56.8 | 40.9 | 0 | 2.3 | 44 | 61.9 | 37.5 | 0 | 0.6 | 160 |
| NSP5 | 50.0 | 50.0 | 0 | 0 | 32 | 54.9 | 45.1 | 0 | 0 | 113 |
| NSP6 | 54.2 | 45.8 | 0 | 0 | 24 | 90.3 | 9.7 | 0 | 0 | 424 |
| NSP7 | 37.5 | 62.5 | 0 | 0 | 8 | 27.3 | 72.7 | 0 | 0 | 11 |
| NSP8 | 47.6 | 47.6 | 0 | 4.8 | 21 | 12.7 | 85.8 | 0 | 1.5 | 134 |
| NSP9 | 33.3 | 66.7 | 0 | 0 | 12 | 37.6 | 62.4 | 0 | 0 | 85 |
| NSP10 | 66.7 | 33.3 | 0 | 0 | 9 | 76.9 | 23.1 | 0 | 0 | 13 |
| NSP12a | 100.0 | 0.0 | 0 | 0 | 1 | 100.0 | 0.0 | 0 | 0 | 73 |
| NSP12b | 58.9 | 41.1 | 0 | 0 | 56 | 87.3 | 12.7 | 0 | 0 | 1312 |
| NSP13 | 62.5 | 37.5 | 0 | 0 | 64 | 67.5 | 32.5 | 0 | 0 | 326 |
| NSP14 | 58.0 | 42.0 | 0 | 0 | 50 | 47.2 | 52.8 | 0 | 0 | 218 |
| NSP15 | 65.8 | 31.6 | 0 | 2.6 | 38 | 37.6 | 62.0 | 0 | 0.4 | 271 |
| NSP16 | 42.9 | 57.1 | 0 | 0 | 21 | 85.0 | 15.0 | 0 | 0 | 107 |
| S | 61.4 | 35.7 | 0.7 | 2.1 | 140 | 85.7 | 13.9 | 0.1 | 0.3 | 1471 |
| ORF3a | 70.4 | 24.1 | 3.7 | 1.9 | 54 | 94.1 | 5.0 | 0.8 | 0.2 | 623 |
| E | 66.7 | 33.3 | 0 | 0 | 9 | 72.3 | 27.7 | 0 | 0 | 47 |
| M | 35.7 | 64.3 | 0 | 0 | 28 | 85.2 | 14.8 | 0 | 0 | 244 |
| ORF6 | 60.0 | 30.0 | 10 | 0 | 10 | 41.2 | 52.9 | 5.9 | 0 | 17 |
| ORF7a | 63.6 | 18.2 | 13.6 | 4.5 | 22 | 19.6 | 75.7 | 3.7 | 0.9 | 107 |
| ORF7b | 57.1 | 28.6 | 14.3 | 0 | 7 | 50.0 | 41.7 | 8.3 | 0 | 12 |
| ORF8 | 64.7 | 23.5 | 5.9 | 5.9 | 17 | 75.5 | 20.4 | 2.0 | 2.0 | 49 |
| N | 69.5 | 30.5 | 0 | 0 | 82 | 82.0 | 18.0 | 0 | 0 | 523 |
| ORF10 | 57.1 | 28.6 | 14.3 | 0 | 7 | 33.3 | 61.1 | 5.6 | 0 | 18 |

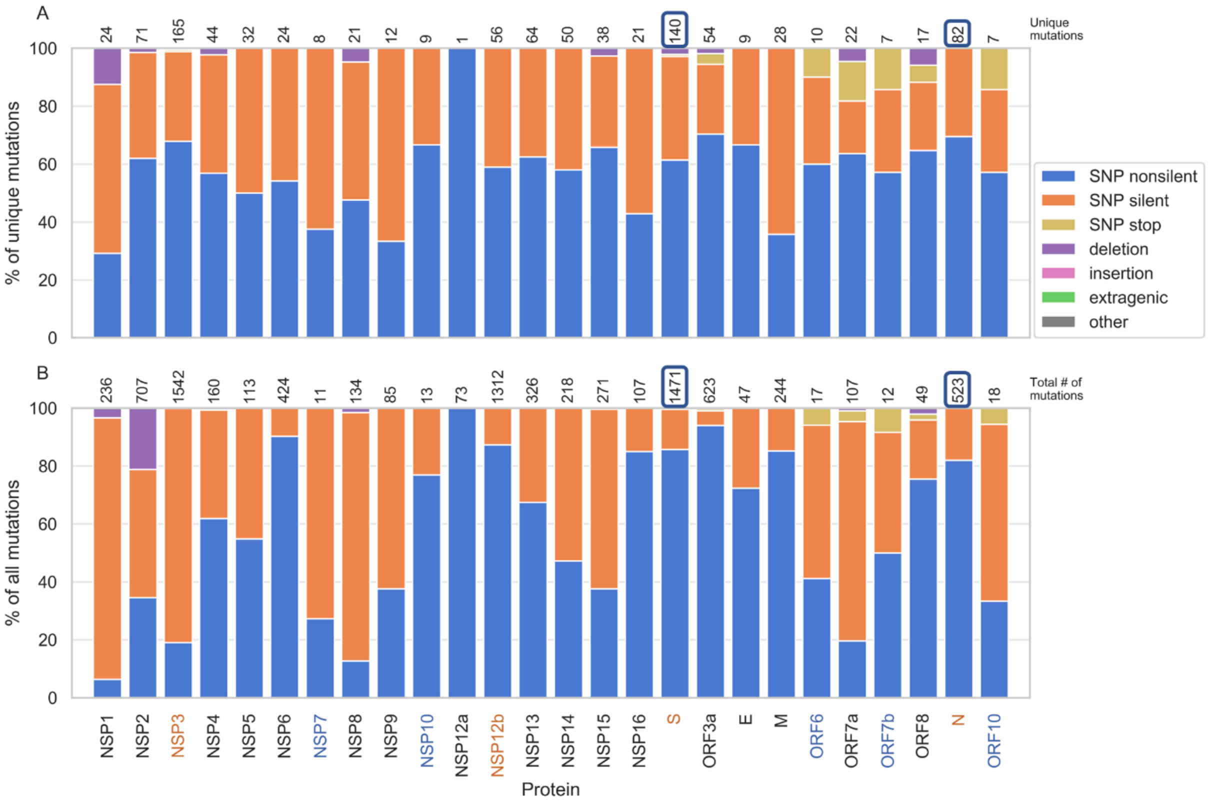


Figure S1. Percent breakdown of unique (A) and total (B) mutations observed in the Netherlands on different proteins of the SARS-CoV-2 genome, total number of unique and total mutations in each protein are placed at the top of the bars. Throughout this report, the term "nonsilent" refers to nucleotide changes accompanied with an amino acid change, whereas "silent" mutations are nucleotide changes with no change in the amino acid sequence.

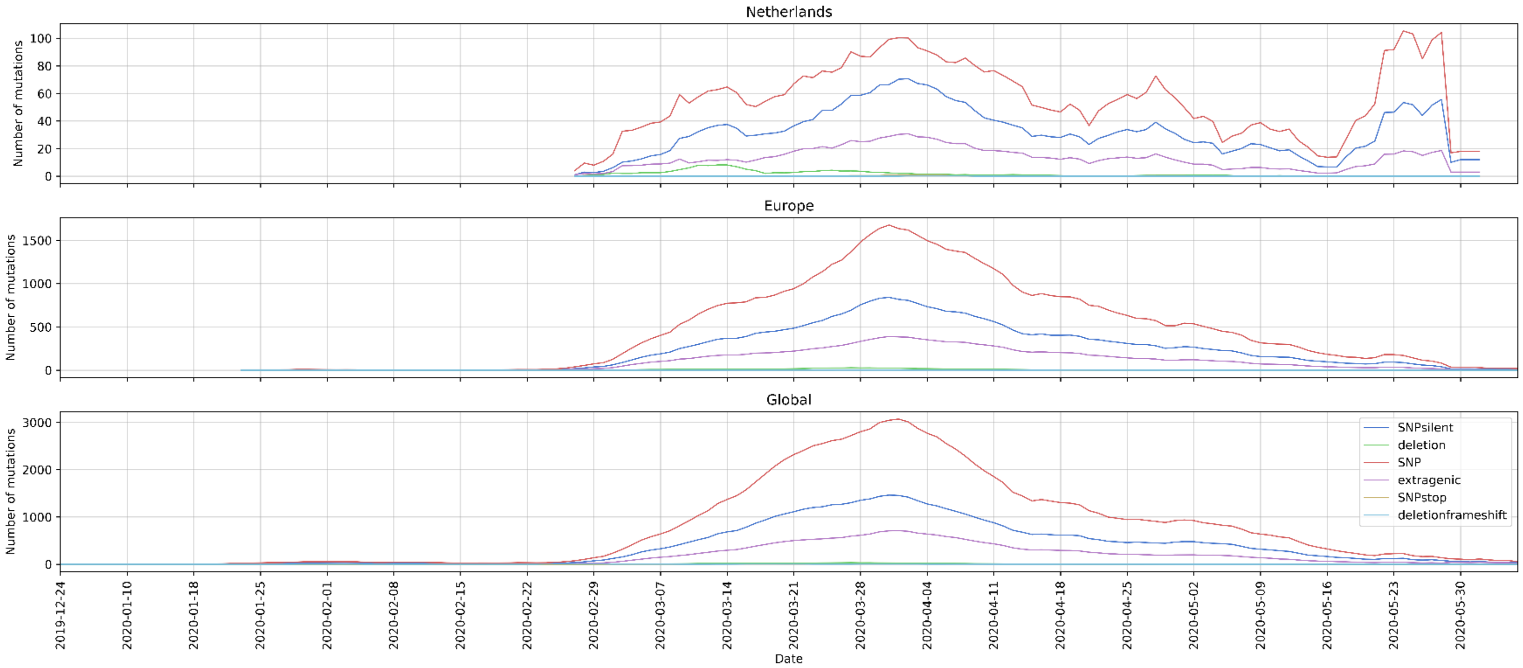


Figure S2. Changes in the percent breakdown of non-silent and silent SNPs observed on SARS-CoV-2 genome in the Netherlands over the course of pandemic.

### Supplementary Figures

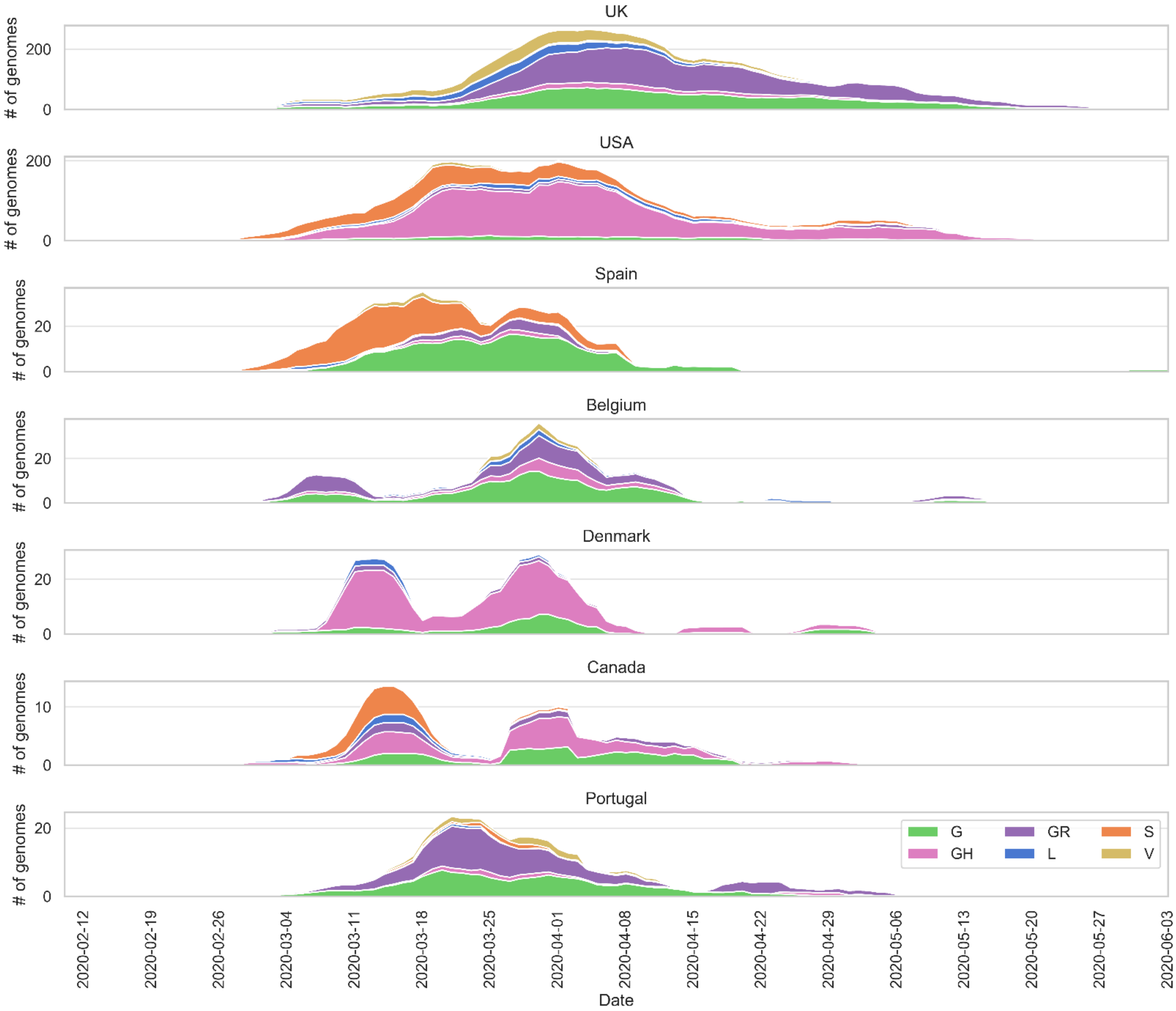


Figure S3. Distribution of SARS-CoV-2 clades in the UK, the USA, Spain, Belgium, Denmark, Canada and Portugal: moving average over seven days was calculated for six clades (see the legend for clade names and colors) discarding intervals of less than one genome per day.

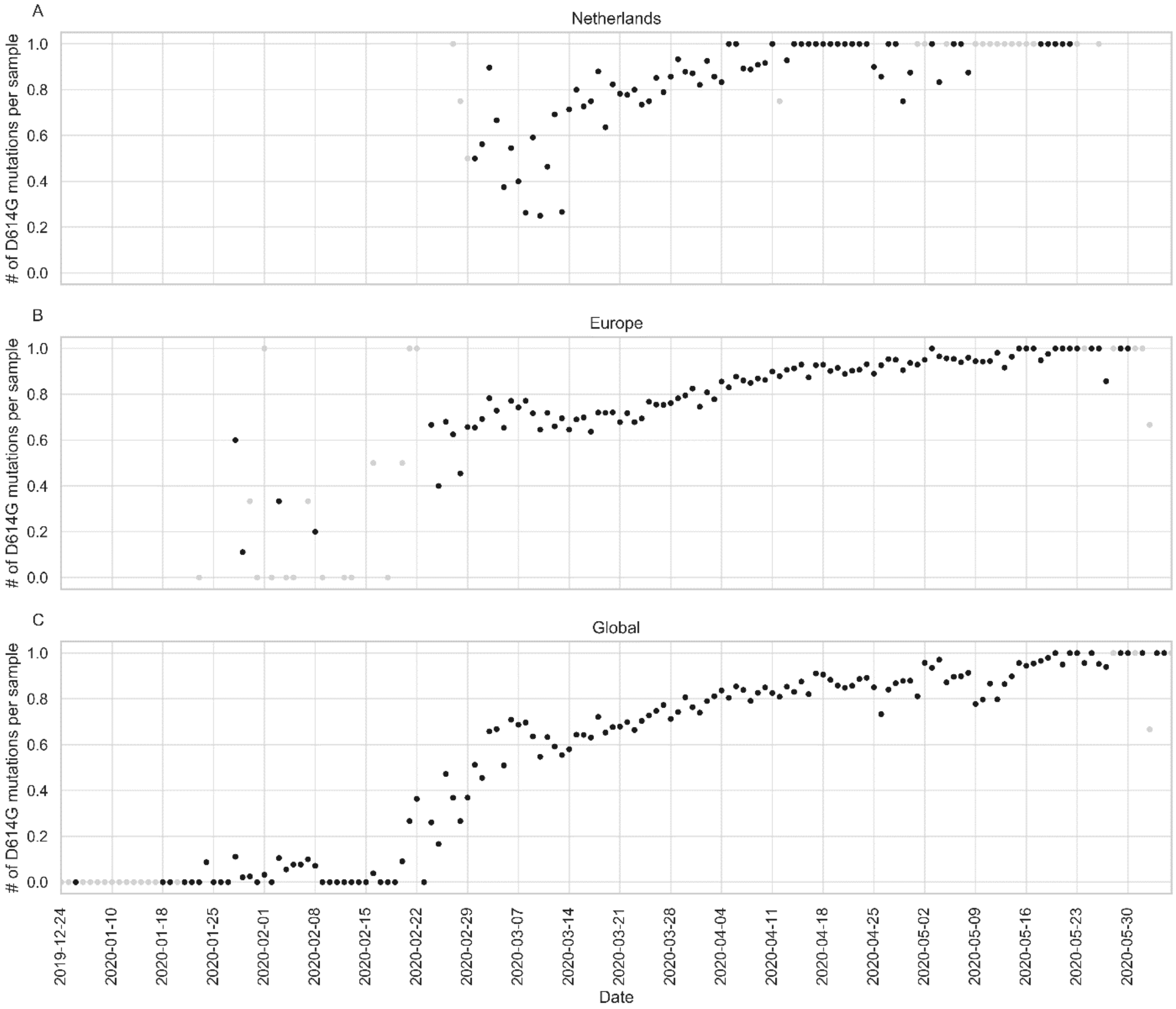


Figure S4. Number of S:D614G mutations observed per sample over the course of pandemic in the Netherlands (A), Europe (B) and globally (C): data points corresponding to dates with fewer than five samples are colored gray to indicate uncertainty.
