## Supplementary material for "Emergence of Novel SARS-CoV-2 Variants in the Netherlands": supplementary-file-2.pdf

Emergence of Novel SARS-CoV-2 Variants in the Netherlands - Supplementary File 2
Aysun Urhan1, Thomas Abeel1,2,*
1 Delft Bioinformatics Lab, Delft University of Technology Van Mourik Broekmanweg 6, 2628 XE Delft, The Netherlands
2 Infectious Disease and Microbiome Program, Broad Institute of MIT and Harvard, 415 Main Street, Cambridge, MA 02142, USA

We gratefully acknowledge the authors, originating and submitting laboratories of the sequences from GISAID's EpiFlu™ Database on which this research is based. The list is detailed below.
All submitters of data may be contacted directly via www.gisaid.org
Accession ID Virus name Location Collection date Originating lab Submitting lab Authors
EPI_ISL_406531 hCoV-19/Guangdong/20SF174/2020 Asia / China / Guangdong / Zhuhai 2020-01-22 Guangdong Provincial Center for Diseases Contorl and Prevention; Guangdong Provinical Public Health Guangdong Provincial Center for Disease Control and Prevention Min Kang, Jie Wu, Jing Lu, Tao Liu, Baisheng Li, Shujiang Mei, Feng Ruan, Lifeng Lin, Changwen Ke, Haojie Zhong, Yingtao Zhang, Lirong Zou, Xuguang Chen, Qi Zhu, Jianpeng Xiao, Jianxiang Geng, Zhe Liu, Jianxiong Hu, Weilin Zeng, Xing Li, Yuhuang Liao, Xiujuan Tang, Songjian Xiao, Ying Wang, Yingchao Song, Xue Zhuang, Lijun Liang, Guanhao He, Huihong Deng, Tie Song, Jianfeng He, Wenjun Ma
EPI_ISL_406533 hCoV-19/Guangzhou/20SF206/2020 Asia / China / Guangdong / Guangzhou 2020-01-22 Guangdong Provincial Center for Diseases Contorl and Prevention; Guangdong Provinical Public Health Guangdong Provincial Center for Diseases Control and Prevention Min Kang, Jie Wu, Jing Lu, Tao Liu, Baisheng Li, Shujiang Mei, Feng Ruan, Lifeng Lin, Changwen Ke, Haojie Zhong, Yingtao Zhang, Lirong Zou, Xuguang Chen, Qi Zhu, Jianpeng Xiao, Jianxiang Geng, Zhe Liu, Jianxiong Hu, Weilin Zeng, Xing Li, Yuhuang Liao, Xiujuan Tang, Songjian Xiao, Ying Wang, Yingchao Song, Xue Zhuang, Lijun Liang, Guanhao He, Huihong Deng, Tie Song, Jianfeng He, Wenjun Ma
EPI_ISL_406534 hCoV-19/Foshan/20SF207/2020 Asia / China / Guangdong / Foshan 2020-01-22 Guangdong Provincial Center for Diseases Control and Prevention; Guangdong Provincial Public Health Guangdong Provincial Center for Diseases Control and Prevention Min Kang, Jie Wu, Jing Lu, Tao Liu, Baisheng Li, Shujiang Mei, Feng Ruan, Lifeng Lin, Changwen Ke, Haojie Zhong, Yingtao Zhang, Lirong Zou, Xuguang Chen, Qi Zhu, Jianpeng Xiao, Jianxiang Geng, Zhe Liu, Jianxiong Hu, Weilin Zeng, Xing Li, Yuhuang Liao, Xiujuan Tang, Songjian Xiao, Ying Wang, Yingchao Song, Xue Zhuang, Lijun Liang, Guanhao He, Huihong Deng, Tie Song, Jianfeng He, Wenjun Ma
EPI_ISL_406535 hCoV-19/Foshan/20SF210/2020 Asia / China / Guangdong / Foshan 2020-01-22 Guangdong Provincial Center for Diseases Control and Prevention; Guangdong Provincial Public Health Guangdong Provincial Center for Diseases Control and Prevention Min Kang, Jie Wu, Jing Lu, Tao Liu, Baisheng Li, Shujiang Mei, Feng Ruan, Lifeng Lin, Changwen Ke, Haojie Zhong, Yingtao Zhang, Lirong Zou, Xuguang Chen, Qi Zhu, Jianpeng Xiao, Jianxiang Geng, Zhe Liu, Jianxiong Hu, Weilin Zeng, Xing Li, Yuhuang Liao, Xiujuan Tang, Songjian Xiao, Ying Wang, Yingchao Song, Xue Zhuang, Lijun Liang, Guanhao He, Huihong Deng, Tie Song, Jianfeng He, Wenjun Ma
EPI_ISL_406536 hCoV-19/Foshan/20SF211/2020 Asia / China / Guangdong / Foshan 2020-01-22 Guangdong Provincial Center for Diseases Control and Prevention; Guangdong Provincial Public Health Guangdong Provincial Center for Diseases Control and Prevention Min Kang, Jie Wu, Jing Lu, Tao Liu, Baisheng Li, Shujiang Mei, Feng Ruan, Lifeng Lin, Changwen Ke, Haojie Zhong, Yingtao Zhang, Lirong Zou, Xuguang Chen, Qi Zhu, Jianpeng Xiao, Jianxiang Geng, Zhe Liu, Jianxiong Hu, Weilin Zeng, Xing Li, Yuhuang Liao, Xiujuan Tang, Songjian Xiao, Ying Wang, Yingchao Song, Xue Zhuang, Lijun Liang, Guanhao He, Huihong Deng, Tie Song, Jianfeng He, Wenjun Ma
EPI_ISL_406538 hCoV-19/Guangdong/20SF201/2020 Asia / China / Guangdong 2020-01-23 Guangdong Provincial Center for Diseases Control and Prevention;Guangdong Provincial Institute of Public Health Guangdong Provincial Center for Diseases Control and Prevention Min Kang, Jie Wu, Jing Lu, Tao Liu, Baisheng Li, Shujiang Mei, Feng Ruan, Lifeng Lin, Changwen Ke, Haojie Zhong, Yingtao Zhang, Lirong Zou, Xuguang Chen, Qi Zhu, Jianpeng Xiao, Jianxiang Geng, Zhe Liu, Jianxiong Hu, Weilin Zeng, Xing Li, Yuhuang Liao, Xiujuan Tang, Songjian Xiao, Ying Wang, Yingchao Song, Xue Zhuang, Lijun Liang, Guanhao He, Huihong Deng, Tie Song, Jianfeng He, Wenjun Ma
EPI_ISL_413513 hCoV-19/South Korea/KUMC03/2020 Asia / South Korea 2020-02-27 Division of Infectious Diseases, Department of Internal Medicine, Korea University College of Medicine Department of Microbiology, Institute for Viral Diseases, College of Medicine, Korea University Changmin Kang, Joon-Yong Bae, Jungmin Lee, Jin Gu Yoon, Heedo Park, Juyoung Cho, Jeonghun Kim, Gee Eun Lee, Cui Chunguang, Kyeong-ryeol Shin, Ji Yun Noh, Joon Young Song, Hee Jin Cheong, Woo Joo Kim, Jin Il Kim, Man-Seong Park
EPI_ISL_413514 hCoV-19/South Korea/KUMC04/2020 Asia / South Korea 2020-02-27 Department of Microbiology, Institute for Viral Diseases, College of Medicine, Korea University Department of Microbiology, Institute for Viral Diseases, College of Medicine, Korea University Changmin Kang, Joon-Yong Bae, Jungmin Lee, Jin Gu Yoon, Heedo Park, Juyoung Cho, Jeonghun Kim, Gee Eun Lee, Cui Chunguang, Kyeong-ryeol Shin, Ji Yun Noh, Joon Young Song, Hee Jin Cheong, Woo Joo Kim, Jin Il Kim, Man-Seong Park
EPI_ISL_413515 hCoV-19/South Korea/KUMC05/2020 Asia / South Korea 2020-02-27 Division of Infectious Diseases, Department of Internal Medicine, Korea University College of Medicine Department of Microbiology, Institute for Viral Diseases, College of Medicine, Korea University Changmin Kang, Joon-Yong Bae, Jungmin Lee, Jin Gu Yoon, Heedo Park, Juyoung Cho, Jeonghun Kim, Gee Eun Lee, Cui Chunguang, Kyeong-ryeol Shin, Ji Yun Noh, Joon Young Song, Hee Jin Cheong, Woo Joo Kim, Jin Il Kim, Man-Seong Park

EPI_ISL_413516 hCoV-19/South Korea/KUMC06/2020 Asia / South Korea 2020-02-27 Department of Microbiology, Institute for Viral Diseases, College of Medicine, Korea University Department of Microbiology, Institute for Viral Diseases, College of Medicine, Korea University Changmin Kang, Joon-Yong Bae, Jungmin Lee, Jin Gu Yoon, Heedo Park, Juyoung Cho, Jeonghun Kim, Gee Eun Lee, Cui Chunguang, Kyeong-ryeol Shin, Ji Yun Noh, Joon Young Song, Hee Jin Cheong, Woo Joo Kim, Jin Il Kim, Man-Seong Park
EPI_ISL_413518 hCoV-19/Beijing/105/2020 Asia / China / Beijing 2020-01-26 unknown Infectious Disease Control Center Li,J., Li,L., Li,Z., Qiu,S., Song,H., Li,P. and Li,P.
EPI_ISL_413519 hCoV-19/Beijing/231/2020 Asia / China / Beijing 2020-01-28 unknown Infectious Disease Control Center Li,J., Li,L., Li,Z., Qiu,S., Song,H., Li,P. and Li,P.
EPI_ISL_413520 hCoV-19/Beijing/233/2020 Asia / China / Beijing 2020-01-28 unknown Infectious Disease Control Center Li,J., Li,L., Li,Z., Qiu,S., Song,H., Li,P. and Li,P.
EPI_ISL_413521 hCoV-19/Beijing/235/2020 Asia / China / Beijing 2020-01-28 unknown Infectious Disease Control Center Li,J., Li,L., Li,Z., Qiu,S., Song,H., Li,P. and Li,P.
EPI_ISL_413522 hCoV-19/India/1-27/2020 Asia / India / Kerala 2020-01-27 Indian Council of Medical Research - National Institute of Virology National Influenza Center, Indian Council of Medical Research - National Institute of Virology Potdar V, Yadav PD, Choudhary ML, Shete-Aich A
EPI_ISL_413523 hCoV-19/India/1-31/2020 Asia / India / Kerala 2020-01-31 Indian Council of Medical Research-National Institute of Virology National Influenza Center, Indian Council of Medical Research-National Institute of Virology Potdar V, Yadav PD, Choudhary ML, Shete-Aich A
EPI_ISL_413555 hCoV-19/Wales/PHW1/2020 Europe / United Kingdom / Wales 2020-02-27 Wales Specialist Virology Centre Public Health Wales Microbiology Cardiff Catherine Moore, Cen Sabu, Joanne Watkins, Sally Corden, Tom Connor
EPI_ISL_413556 hCoV-19/Wales/PHW2/2020 Europe / United Kingdom / Wales 2020-03-04 Wales Specialist Virology Centre Public Health Wales Microbiology Cardiff Catherine Moore, Tim Jones, Joanne Watkins, Sally Corden, Tom Connor
EPI_ISL_413557 hCoV-19/USA/CA-CDPH-UC1/2020 North America / USA / California / Sonoma County 2020-02-28 California Department of Public Health Chiu Laboratory, University of California, San Francisco Xianding Deng, Scot Federman, Chao-Yang Pan, Hugo Guevara,Wei Gu, Debra A. Wadford, and Charles Y. Chiu
EPI_ISL_413558 hCoV-19/USA/CA-CDPH-UC2/2020 North America / USA / California / Solano County 2020-02-27 California Department of Public Health Chiu Laboratory, University of California, San Francisco Xianding Deng, Scot Federman, Chao-Yang Pan, Hugo Guevara,Wei Gu, Debra A. Wadford, and Charles Y. Chiu
EPI_ISL_413559 hCoV-19/USA/CA-CDPH-UC3/2020 North America / USA / California / Solano County 2020-02-27 California Department of Public Health Chiu Laboratory, University of California, San Francisco Xianding Deng, Scot Federman, Chao-Yang Pan, Hugo Guevara,Wei Gu, Debra A. Wadford, and Charles Y. Chiu
EPI_ISL_413560 hCoV-19/USA/WA-S3/2020 North America / USA / Washington 2020-02-28 Seattle Flu Study Seattle Flu Study Chu et al
EPI_ISL_413561 hCoV-19/USA/CA-CDPH-UC4/2020 North America / USA / California / Solano County 2020-02-27 California Department of Public Health Chiu Laboratory, University of California, San Francisco Xianding Deng, Scot Federman, Chao-Yang Pan, Hugo Guevara,Wei Gu, Debra A. Wadford, and Charles Y. Chiu
EPI_ISL_413562 hCoV-19/USA/WA11-UW7/2020 North America / USA / Washington 2020-03-02 UW Virology Lab UW Virology Lab Pavitra Roychoudhury, Hong Xie, Keith Jerome, Alexander Greninger
EPI_ISL_413563 hCoV-19/USA/WA12-UW8/2020 North America / USA / Washington 2020-03-03 UW Virology Lab UW Virology Lab Pavitra Roychoudhury, Hong Xie, Keith Jerome, Alexander Greninger
EPI_ISL_413564 hCoV-19/Netherlands/Andel_1365066/2020 Europe / Netherlands / Andel 2020-03-01 MHC West-Brabant Erasmus Medical Center David Nieuwenhuijse, Bas Oude Munnink, Reina Sikkema, Claudia Schapendonk, Irina Chestakova, Anne van der Linden, Mark Pronk, Pascal Lexmond, Corien Swaan, Manon Haverkate, Madelief Mollers, Mart Stein, Sandra Kengne Kamga Mobou, Jeroen van Kampen, Jolanda Voermans, Aura Timen, Corine GeurtsvanKessel, Annemiek van der Eijk, Richard Molenkamp, Marion Koopmans, on behalf of the Dutch national COVID-19 response team.
EPI_ISL_413566 hCoV-19/Netherlands/Blaricum_1364780/2020 Europe / Netherlands / Blaricum 2020-03-02 MHC Gooi & Vechtstreek Erasmus Medical Center David Nieuwenhuijse, Bas Oude Munnink, Reina Sikkema, Claudia Schapendonk, Irina Chestakova, Anne van der Linden, Mark Pronk, Pascal Lexmond, Corien Swaan, Manon Haverkate, Madelief Mollers, Mart Stein, Sandra Kengne Kamga Mobou, Jeroen van Kampen, Jolanda Voermans, Aura Timen, Corine GeurtsvanKessel, Annemiek van der Eijk, Richard Molenkamp, Marion Koopmans, on behalf of the Dutch national COVID-19 response team.
EPI_ISL_413568 hCoV-19/Netherlands/Dalen_1363624/2020 Europe / Netherlands / Dalen 2020-03-01 MHC Drente Erasmus Medical Center David Nieuwenhuijse, Bas Oude Munnink, Reina Sikkema, Claudia Schapendonk, Irina Chestakova, Anne van der Linden, Mark Pronk, Pascal Lexmond, Corien Swaan, Manon Haverkate, Madelief Mollers, Mart Stein, Sandra Kengne Kamga Mobou, Jeroen van Kampen, Jolanda Voermans, Aura Timen, Corine GeurtsvanKessel, Annemiek van der Eijk, Richard Molenkamp, Marion Koopmans, on behalf of the Dutch national COVID-19 response team.
EPI_ISL_413571 hCoV-19/Netherlands/Eindhoven_1363782/2020 Europe / Netherlands / Eindhoven 2020-03-02 MHC Brabant Zuidoost Erasmus Medical Center David Nieuwenhuijse, Bas Oude Munnink, Reina Sikkema, Claudia Schapendonk, Irina Chestakova, Anne van der Linden, Mark Pronk, Pascal Lexmond, Corien Swaan, Manon Haverkate, Madelief Mollers, Mart Stein, Sandra Kengne Kamga Mobou, Jeroen van Kampen, Jolanda Voermans, Aura Timen, Corine GeurtsvanKessel, Annemiek van der Eijk, Richard Molenkamp, Marion Koopmans, on behalf of the Dutch national COVID-19 response team.
EPI_ISL_413572 hCoV-19/Netherlands/Haarlem_1363688/2020 Europe / Netherlands / Haarlem 2020-03-01 MHC Kennemerland Erasmus Medical Center David Nieuwenhuijse, Bas Oude Munnink, Reina Sikkema, Claudia Schapendonk, Irina Chestakova, Anne van der Linden, Mark Pronk, Pascal Lexmond, Corien Swaan, Manon Haverkate,

Madelief Mollers, Mart Stein, Sandra Kengne Kamga Mobou, Jeroen van Kampen, Jolanda Voermans, Aura Timen, Corine GeurtsvanKessel, Annemiek van der Eijk, Richard Molenkamp, Marion Koopmans, on behalf of the Dutch national COVID-19 response team.
EPI_ISL_413573 hCoV-19/Netherlands/Hardinxveld_Giessendam_1364806/2020 Europe / Netherlands / Hardinxveld Giessendam 2020-03-02 Dienst Gezondheid & Jeugd Zuid-Holland Zuid Erasmus Medical Center David Nieuwenhuijse, Bas Oude Munnink, Reina Sikkema, Claudia Schapendonk, Irina Chestakova, Anne van der Linden, Mark Pronk, Pascal Lexmond, Corien Swaan, Manon Haverkate, Madelief Mollers, Mart Stein, Sandra Kengne Kamga Mobou, Jeroen van Kampen, Jolanda Voermans, Aura Timen, Corine GeurtsvanKessel, Annemiek van der Eijk, Richard Molenkamp, Marion Koopmans, on behalf of the Dutch national COVID-19 response team.
EPI_ISL_413574 hCoV-19/Netherlands/Helmond_1363548/2020 Europe / Netherlands / Helmond 2020-02-29 MHC West-Brabant Erasmus Medical Center David Nieuwenhuijse, Bas Oude Munnink, Reina Sikkema, Claudia Schapendonk, Irina Chestakova, Anne van der Linden, Mark Pronk, Pascal Lexmond, Corien Swaan, Manon Haverkate, Madelief Mollers, Mart Stein, Sandra Kengne Kamga Mobou, Jeroen van Kampen, Jolanda Voermans, Aura Timen, Corine GeurtsvanKessel, Annemiek van der Eijk, Richard Molenkamp, Marion Koopmans, on behalf of the Dutch national COVID-19 response team.
EPI_ISL_413577 hCoV-19/Netherlands/Naarden_1364774/2020 Europe / Netherlands / Naarden 2020-03-02 MHC Gooi & Vechtstreek Erasmus Medical Center David Nieuwenhuijse, Bas Oude Munnink, Reina Sikkema, Claudia Schapendonk, Irina Chestakova, Anne van der Linden, Mark Pronk, Pascal Lexmond, Corien Swaan, Manon Haverkate, Madelief Mollers, Mart Stein, Sandra Kengne Kamga Mobou, Jeroen van Kampen, Jolanda Voermans, Aura Timen, Corine GeurtsvanKessel, Annemiek van der Eijk, Richard Molenkamp, Marion Koopmans, on behalf of the Dutch national COVID-19 response team.
EPI_ISL_413579 hCoV-19/Netherlands/Nootdorp_1364222/2020 Europe / Netherlands / Nootdorp 2020-03-03 MHC Haaglanden Erasmus Medical Center David Nieuwenhuijse, Bas Oude Munnink, Reina Sikkema, Claudia Schapendonk, Irina Chestakova, Anne van der Linden, Mark Pronk, Pascal Lexmond, Corien Swaan, Manon Haverkate, Madelief Mollers, Mart Stein, Sandra Kengne Kamga Mobou, Jeroen van Kampen, Jolanda Voermans, Aura Timen, Corine GeurtsvanKessel, Annemiek van der Eijk, Richard Molenkamp, Marion Koopmans, on behalf of the Dutch national COVID-19 response team.
EPI_ISL_413580 hCoV-19/Netherlands/Oisterwijk_1364072/2020 Europe / Netherlands / Oisterwijk 2020-03-02 MHC Hart voor Brabant Erasmus Medical Center David Nieuwenhuijse, Bas Oude Munnink, Reina Sikkema, Claudia Schapendonk, Irina Chestakova, Anne van der Linden, Mark Pronk, Pascal Lexmond, Corien Swaan, Manon Haverkate, Madelief Mollers, Mart Stein, Sandra Kengne Kamga Mobou, Jeroen van Kampen, Jolanda Voermans, Aura Timen, Corine GeurtsvanKessel, Annemiek van der Eijk, Richard Molenkamp, Marion Koopmans, on behalf of the Dutch national COVID-19 response team.
EPI_ISL_413582 hCoV-19/Netherlands/Rotterdam_1363790/2020 Europe / Netherlands / Rotterdam 2020-03-01 ErasmusMC Erasmus Medical Center David Nieuwenhuijse, Bas Oude Munnink, Reina Sikkema, Claudia Schapendonk, Irina Chestakova, Anne van der Linden, Mark Pronk, Pascal Lexmond, Corien Swaan, Manon Haverkate, Madelief Mollers, Mart Stein, Sandra Kengne Kamga Mobou, Jeroen van Kampen, Jolanda Voermans, Aura Timen, Corine GeurtsvanKessel, Annemiek van der Eijk, Richard Molenkamp, Marion Koopmans, on behalf of the Dutch national COVID-19 response team.
EPI_ISL_413583 hCoV-19/Netherlands/Rotterdam_1364040/2020 Europe / Netherlands / Rotterdam 2020-03-02 MHC Rotterdam-Rijnmond Erasmus Medical Center David Nieuwenhuijse, Bas Oude Munnink, Reina Sikkema, Claudia Schapendonk, Irina Chestakova, Anne van der Linden, Mark Pronk, Pascal Lexmond, Corien Swaan, Manon Haverkate, Madelief Mollers, Mart Stein, Sandra Kengne Kamga Mobou, Jeroen van Kampen, Jolanda Voermans, Aura Timen, Corine GeurtsvanKessel, Annemiek van der Eijk, Richard Molenkamp, Marion Koopmans, on behalf of the Dutch national COVID-19 response team.
EPI_ISL_413584 hCoV-19/Netherlands/Rotterdam_1364740/2020 Europe / Netherlands / Rotterdam 2020-03-03 unknown Erasmus Medical Center David Nieuwenhuijse, Bas Oude Munnink, Reina Sikkema, Claudia Schapendonk, Irina Chestakova, Anne van der Linden, Mark Pronk, Pascal Lexmond, Corien Swaan, Manon Haverkate, Madelief Mollers, Mart Stein, Sandra Kengne Kamga Mobou, Jeroen van Kampen, Jolanda Voermans, Aura Timen, Corine GeurtsvanKessel, Annemiek van der Eijk, Richard Molenkamp, Marion Koopmans, on behalf of the Dutch national COVID-19 response team.
EPI_ISL_413587 hCoV-19/Netherlands/Tilburg_1364286/2020 Europe / Netherlands / Tilburg 2020-03-03 Foundation Elisabeth-Tweesteden Ziekenhuis Erasmus Medical Center David Nieuwenhuijse, Bas Oude Munnink, Reina Sikkema, Claudia Schapendonk, Irina Chestakova, Anne van der Linden, Mark Pronk, Pascal Lexmond, Corien Swaan, Manon Haverkate, Madelief Mollers, Mart Stein, Sandra Kengne Kamga Mobou, Jeroen van Kampen, Jolanda Voermans, Aura Timen, Corine GeurtsvanKessel, Annemiek van der Eijk, Richard Molenkamp, Marion Koopmans, on behalf of the Dutch national COVID-19 response team.
EPI_ISL_413588 hCoV-19/Netherlands/Utrecht_1363564/2020 Europe / Netherlands / Utrecht 2020-03-01 MHC Utrecht Erasmus Medical Center David Nieuwenhuijse, Bas Oude Munnink, Reina Sikkema, Claudia Schapendonk, Irina Chestakova, Anne van der Linden, Mark Pronk, Pascal Lexmond, Corien Swaan, Manon Haverkate, Madelief Mollers, Mart Stein, Sandra Kengne Kamga Mobou, Jeroen van Kampen, Jolanda Voermans, Aura Timen, Corine GeurtsvanKessel, Annemiek van der Eijk, Richard Molenkamp, Marion Koopmans, on behalf of the Dutch national COVID-19 response team.
EPI_ISL_413589 hCoV-19/Netherlands/Utrecht_1363628/2020 Europe / Netherlands / Utrecht 2020-03-01 MHC Utrecht Erasmus Medical Center David Nieuwenhuijse, Bas Oude Munnink, Reina Sikkema, Claudia Schapendonk, Irina Chestakova, Anne van der Linden, Mark Pronk, Pascal Lexmond, Corien Swaan, Manon Haverkate, Madelief Mollers, Mart Stein, Sandra Kengne Kamga Mobou, Jeroen van Kampen, Jolanda Voermans, Aura Timen, Corine GeurtsvanKessel, Annemiek van der Eijk, Richard Molenkamp, Marion Koopmans, on behalf of the Dutch national COVID-19 response team.
EPI_ISL_413590 hCoV-19/Netherlands/Utrecht_1364066/2020 Europe / Netherlands / Utrecht 2020-03-02

MHC Utrecht	Erasmus Medical Center	David Nieuwenhuijse, Bas Oude Munnink, Reina Sikkema, Claudia Schapendonk, Irina Chestakova, Anne van der Linden, Mark Pronk, Pascal Lexmond, Corien Swaan, Manon Haverkate, Madelief Mollers, Mart Stein, Sandra Kengne Kamga Mobou, Jeroen van Kampen, Jolanda Voermans, Aura Timen, Corine GeurtsvanKessel, Annemiek van der Eijk, Richard Molenkamp, Marion Koopmans, on behalf of the Dutch national COVID-19 response team.
EPI_ISL_413591	hCoV-19/Netherlands/Zeewolde_1365080/2020	Europe / Netherlands / Zeewolde	2020-03-02	MHC Flevoland	Erasmus Medical Center	David Nieuwenhuijse, Bas Oude Munnink, Reina Sikkema, Claudia Schapendonk, Irina Chestakova, Anne van der Linden, Mark Pronk, Pascal Lexmond, Corien Swaan, Manon Haverkate, Madelief Mollers, Mart Stein, Sandra Kengne Kamga Mobou, Jeroen van Kampen, Jolanda Voermans, Aura Timen, Corine GeurtsvanKessel, Annemiek van der Eijk, Richard Molenkamp, Marion Koopmans, on behalf of the Dutch national COVID-19 response team.
EPI_ISL_413592	hCoV-19/Taiwan/NTU03/2020	Asia / Taiwan / Taipei	2020-03-02	Department of Laboratory Medicine, National Taiwan University Hospital	Microbial Genomics Core Lab, National Taiwan University Centers of Genomic and Precision Medicine	Shiou-Hwei Yeh, You-Yu Lin, Ya-Yun Lai, Chiao-Ling Li, Shan-Chwen Chang, Pei-Jer Chen, Sui-Yuan Chang
EPI_ISL_413593	hCoV-19/Luxembourg/Lux1/2020	Europe / Luxembourg	2020-02-29	Laboratoire National de Santé	Erasmus Medical Center	David Nieuwenhuijse, Bas Oude Munnink, Reina Sikkema, Claudia Schapendonk, Irina Chestakova, Anne van der Linden, Mark Pronk, Pascal Lexmond, T. Abdelrahman, G. Fournier, J. Mossong, T. Nguyen, Jeroen van Kampen, Jolanda Voermans, Corine GeurtsvanKessel, Annemiek van der Eijk, Richard Molenkamp, Marion Koopmans, on behalf of the Dutch national COVID-19 response team.
EPI_ISL_413594	hCoV-19/Australia/NSW08/2020	Oceania / Australia / NSW / Sydney	2020-02-28	Centre for Infectious Diseases and Microbiology Laboratory Services	NSW Health Pathology - Institute of Clinical Pathology and Medical Research; Westmead Hospital; University of Sydney	Rockett R, Eden J-S, Lam C, Gray K, Timms, V, Gall, M, Alicia, A, Carter I, Rahman H, Holmes EC, , O’Sullivan MV, Sintchenko V, Chen SC, Maddocks S, Kok J and Dwyer DE for the 2019-nCoV Study Group*
EPI_ISL_413595	hCoV-19/Australia/NSW09/2020	Oceania / Australia / NSW / Sydney	2020-02-28	Centre for Infectious Diseases and Microbiology Laboratory Services	NSW Health Pathology - Institute of Clinical Pathology and Medical Research; Westmead Hospital; University of Sydney	Rockett R, Eden J-S, Lam C, Gray K, Timms, V, Gall, M, Carter I, Rahman H, Holmes EC, O’Sullivan MV, Sintchenko V, Chen SC, Maddocks S, Kok J and Dwyer DE for the 2019-nCoV Study Group*
EPI_ISL_413596	hCoV-19/Australia/NSW10/2020	Oceania / Australia / NSW / Sydney	2020-02-28	Centre for Infectious Diseases and Microbiology - Public Health	NSW Health Pathology - Institute of Clinical Pathology and Medical Research; Westmead Hospital; University of Sydney	Rockett R, Eden J-S, Lam C, Gray K, Timms, V, Gall, M, Carter I, Rahman H, Holmes EC, O’Sullivan MV, Sintchenko V, Chen SC, Maddocks S, Kok J and Dwyer DE for the 2019-nCoV Study Group*
EPI_ISL_413597	hCoV-19/Australia/NSW11/2020	Oceania / Australia / NSW / Sydney	2020-03-02	Centre for Infectious Diseases and Microbiology- Public Health	NSW Health Pathology - Institute of Clinical Pathology and Medical Research; Westmead Hospital; University of Sydney	Lam C, Eden J-S, Rockett R, Gray K, Timms, V, Gall, M, Carter I, Rahman H, Holmes EC, O’Sullivan MV, Sintchenko V, Chen SC, Maddocks S, Kok J and Dwyer DE for the 2019-nCoV Study Group*
EPI_ISL_413598	hCoV-19/Australia/NSW12/2020	Oceania / Australia / NSW / Sydney	2020-03-04	Centre for Infectious Diseases and Microbiology - Public Health	NSW Health Pathology - Institute of Clinical Pathology and Medical Research; Westmead Hospital; University of Sydney	Gray K, Eden J-S, Lam C, Rockett R, Timms, V, Gall, M, Carter I, Rahman H, Holmes EC, O’Sullivan MV, Sintchenko V, Chen SC, Maddocks S, Kok J and Dwyer DE for the 2019-nCoV Study Group*
EPI_ISL_413599	hCoV-19/Australia/NSW13/2020	Oceania / Australia / NSW / Sydney	2020-03-04	Centre for Infectious Diseases and Microbiology - Public Health	NSW Health Pathology - Institute of Clinical Pathology and Medical Research; Westmead Hospital; University of Sydney	Timms, V, Eden J-S, Lam C, Gray K, Rockett R, Gall, M, Carter I, Rahman H, Holmes EC, O’Sullivan MV, Sintchenko V, Chen SC, Maddocks S, Kok J and Dwyer DE for the 2019-nCoV Study Group*
EPI_ISL_413600	hCoV-19/Australia/NSW14/2020	Oceania / Australia / NSW / Sydney	2020-03-03	Centre for Infectious Diseases and Microbiology - Public Health	NSW Health Pathology - Institute of Clinical Pathology and Medical Research; Westmead Hospital; University of Sydney	Gall, M, Eden J-S, Lam C, Gray K, Timms, V, Rockett R, Carter I, Rahman H, Holmes EC, O’Sullivan MV, Sintchenko V, Chen SC, Maddocks S, Kok J and Dwyer DE for the 2019-nCoV Study Group*
EPI_ISL_413601	hCoV-19/USA/WA13-UW9/2020	North America / USA / Washington	2020-03-02	UW Virology Lab	UW Virology Lab	Pavitra Roychoudhury, Hong Xie, Keith Jerome, Alexander Greninger
EPI_ISL_413711	hCoV-19/China/WF0014/2020	Asia / China	2020-02	Weifang Center for Disease Control and Prevention	Weifang Center for Disease Control and Prevention & BGI-Shenzhen	Qing Nie, Xingguang Li, Erik M Volz, Han Fu, Haowei Wang, Xiaoyue Xi, Wei Chen, Dehui Liu, Yingying Chen, Mengmeng Tian, Wei Tan, Junjie Zai, Wanying Sun, Jiandong Li, Junhua Li
EPI_ISL_417091	hCoV-19/USA/WA-S38/2020	North America / USA / Washington / Snohomish County	2020-03-04	Washington State Department of Health	Seattle Flu Study	Chu etl al
EPI_ISL_417092	hCoV-19/USA/WA-S39/2020	North America / USA / Washington / Snohomish County	2020-03-04	Washington State Department of Health	Seattle Flu Study	Chu etl al
EPI_ISL_417093	hCoV-19/USA/WA-S40/2020	North America / USA / Washington / Snohomish County	2020-02-28	Washington State Department of Health	Seattle Flu Study	Chu etl al
EPI_ISL_417095	hCoV-19/USA/WA-S42/2020	North America / USA / Washington / King County	2020-02-28	Washington State Department of Health	Seattle Flu Study	Chu etl al
EPI_ISL_417096	hCoV-19/USA/WA-S43/2020	North America / USA / Washington	2020-02-27	Washington

State Department of Health	Seattle Flu Study	Chu etl al
EPI_ISL_417097	hCoV-19/USA/WA-S44/2020 North America / USA / Washington	2020-02-28	Washington State Department of Health	Seattle Flu Study	Chu etl al
EPI_ISL_417098	hCoV-19/USA/WA-S45/2020 North America / USA / Washington	2020-02-29	Washington State Department of Health	Seattle Flu Study	Chu etl al
EPI_ISL_417099	hCoV-19/USA/WA-S46/2020 North America / USA / Washington	2020-02-29	Washington State Department of Health	Seattle Flu Study	Chu etl al
EPI_ISL_417100	hCoV-19/USA/WA-S47/2020 North America / USA / Washington / Snohomish County	2020-02-29	Washington State Department of Health	Seattle Flu Study	Chu etl al
EPI_ISL_417101	hCoV-19/USA/WA-S48/2020 North America / USA / Washington / Snohomish County	2020-02-29	Washington State Department of Health	Seattle Flu Study	Chu etl al
EPI_ISL_417102	hCoV-19/USA/WA-S49/2020 North America / USA / Washington / King County	2020-03-04	Washington State Department of Health	Seattle Flu Study	Chu etl al
EPI_ISL_417103	hCoV-19/USA/WA-S50/2020 North America / USA / Washington / King County	2020-03-05	Washington State Department of Health	Seattle Flu Study	Chu etl al
EPI_ISL_417104	hCoV-19/USA/WA-S51/2020 North America / USA / Washington	2020-03-03	Washington State Department of Health	Seattle Flu Study	Chu etl al
EPI_ISL_417105	hCoV-19/USA/WA-S52/2020 North America / USA / Washington / Snohomish County	2020-03-03	Washington State Department of Health	Seattle Flu Study	Chu etl al
EPI_ISL_417106	hCoV-19/USA/WA-S53/2020 North America / USA / Washington / Snohomish County	2020-03-03	Washington State Department of Health	Seattle Flu Study	Chu etl al
EPI_ISL_417107	hCoV-19/USA/WA-S54/2020 North America / USA / Washington / King County	2020-03-05	Washington State Department of Health	Seattle Flu Study	Chu etl al
EPI_ISL_417108	hCoV-19/USA/WA-S55/2020 North America / USA / Washington / King County	2020-02-29	Washington State Department of Health	Seattle Flu Study	Chu etl al
EPI_ISL_417109	hCoV-19/USA/WA-S56/2020 North America / USA / Washington / King County	2020-03-05	Washington State Department of Health	Seattle Flu Study	Chu etl al
EPI_ISL_417110	hCoV-19/USA/WA-S57/2020 North America / USA / Washington	2020-03-03	Washington State Department of Health	Seattle Flu Study	Chu etl al
EPI_ISL_417111	hCoV-19/USA/WA-S58/2020 North America / USA / Washington / Snohomish County	2020-03-05	Washington State Department of Health	Seattle Flu Study	Chu etl al
EPI_ISL_417112	hCoV-19/USA/WA-S59/2020 North America / USA / Washington / King County	2020-03-02	Washington State Department of Health	Seattle Flu Study	Chu etl al
EPI_ISL_417114	hCoV-19/USA/WA-S61/2020 North America / USA / Washington / King County	2020-03-05	Washington State Department of Health	Seattle Flu Study	Chu etl al
EPI_ISL_417115	hCoV-19/USA/WA-S62/2020 North America / USA / Washington / Snohomish County	2020-03-03	Washington State Department of Health	Seattle Flu Study	Chu etl al
EPI_ISL_417116	hCoV-19/USA/WA-S63/2020 North America / USA / Washington / King County	2020-03-04	Washington State Department of Health	Seattle Flu Study	Chu etl al
EPI_ISL_417117	hCoV-19/USA/WA-S64/2020 North America / USA / Washington / King County	2020-03-03	Washington State Department of Health	Seattle Flu Study	Chu etl al
EPI_ISL_417118	hCoV-19/USA/WA-S65/2020 North America / USA / Washington / King County	2020-03-03	Washington State Department of Health	Seattle Flu Study	Chu etl al
EPI_ISL_417119	hCoV-19/USA/WA-S66/2020 North America / USA / Washington / Snohomish County	2020-03-06	Washington State Department of Health	Seattle Flu Study	Chu etl al
EPI_ISL_417120	hCoV-19/USA/WA-S67/2020 North America / USA / Washington / King County	2020-03-04	Washington State Department of Health	Seattle Flu Study	Chu etl al
EPI_ISL_417121	hCoV-19/USA/WA-S68/2020 North America / USA / Washington / King County	2020-03-04	Washington State Department of Health	Seattle Flu Study	Chu etl al
EPI_ISL_417122	hCoV-19/USA/WA-S69/2020 North America / USA / Washington / King County	2020-03-05	Washington State Department of Health	Seattle Flu Study	Chu etl al
EPI_ISL_417123	hCoV-19/USA/WA-S70/2020 North America / USA / Washington / King County	2020-03-05	Washington State Department of Health	Seattle Flu Study	Chu etl al
EPI_ISL_417124	hCoV-19/USA/WA-S71/2020 North America / USA / Washington / King County	2020-03-05	Washington State Department of Health	Seattle Flu Study	Chu etl al
EPI_ISL_417125	hCoV-19/USA/WA-S72/2020 North America / USA / Washington / King County	2020-03-06	Washington State Department of Health	Seattle Flu Study	Chu etl al
EPI_ISL_417126	hCoV-19/USA/WA-S73/2020 North America / USA / Washington	2020-03-05	Washington State Department of Health	Seattle Flu Study	Chu etl al
EPI_ISL_417127	hCoV-19/USA/WA-S74/2020 North America / USA / Washington / King County	2020-03-05	Washington State Department of Health	Seattle Flu Study	Chu etl al
EPI_ISL_417128	hCoV-19/USA/WA-S75/2020 North America / USA / Washington / King County	2020-03-05	Washington State Department of Health	Seattle Flu Study	Chu etl al
EPI_ISL_417129	hCoV-19/USA/WA-S76/2020 North America / USA / Washington / King County	2020-03-05	Washington State Department of Health	Seattle Flu Study	Chu etl al
EPI_ISL_417130	hCoV-19/USA/WA-S77/2020 North America / USA / Washington / King County	2020-03-05	Washington State Department of Health	Seattle Flu Study	Chu etl al
EPI_ISL_417132	hCoV-19/USA/WA-S79/2020 North America / USA / Washington / King County	2020-03-05	Washington State Department of Health	Seattle Flu Study	Chu etl al
EPI_ISL_417133	hCoV-19/USA/WA-S80/2020 North America / USA / Washington / King County	2020-03-05

Washington State Department of Health	Seattle Flu Study	Chu etl al
EPI_ISL_417134	hCoV-19/USA/WA-S81/2020	North America / USA / Washington / King County	2020-02-26
Washington State Department of Health	Seattle Flu Study	Chu etl al
EPI_ISL_417135	hCoV-19/USA/WA-S82/2020	North America / USA / Washington / King County	2020-02-22
Washington State Department of Health	Seattle Flu Study	Chu etl al
EPI_ISL_417136	hCoV-19/USA/WA-S83/2020	North America / USA / Washington / King County	2020-03-05
Washington State Department of Health	Seattle Flu Study	Chu etl al
EPI_ISL_417137	hCoV-19/USA/WA-S84/2020	North America / USA / Washington / King County	2020-02-21
Washington State Department of Health	Seattle Flu Study	Chu etl al
EPI_ISL_417139	hCoV-19/USA/WA-S86/2020	North America / USA / Washington / King County	2020-03-01
Washington State Department of Health	Seattle Flu Study	Chu etl al
EPI_ISL_417140	hCoV-19/USA/WA-S87/2020	North America / USA / Washington / King County	2020-03-01
Washington State Department of Health	Seattle Flu Study	Chu etl al
EPI_ISL_417141	hCoV-19/USA/WA-S88/2020	North America / USA / Washington / King County	2020-03-01
Washington State Department of Health	Seattle Flu Study	Chu etl al
EPI_ISL_417142	hCoV-19/USA/WA-S89/2020	North America / USA / Washington / Umatilla County	2020-02-29
Washington State Department of Health	Seattle Flu Study	Chu etl al
EPI_ISL_417143	hCoV-19/USA/WA-S90/2020	North America / USA / Washington / Umatilla County	2020-02-29
Washington State Department of Health	Seattle Flu Study	Chu etl al
EPI_ISL_417144	hCoV-19/USA/WA-S91/2020	North America / USA / Washington / Snohomish County	2020-03-02
Washington State Department of Health	Seattle Flu Study	Chu etl al
EPI_ISL_417145	hCoV-19/USA/WA-S92/2020	North America / USA / Washington / King County	2020-02-29
Washington State Department of Health	Seattle Flu Study	Chu etl al
EPI_ISL_417146	hCoV-19/USA/WA-S93/2020	North America / USA / Washington / King County	2020-02-29
Washington State Department of Health	Seattle Flu Study	Chu etl al
EPI_ISL_417147	hCoV-19/USA/WA-S94/2020	North America / USA / Washington / Snohomish County	2020-02-28
Washington State Department of Health	Seattle Flu Study	Chu etl al
EPI_ISL_417148	hCoV-19/USA/WA-S95/2020	North America / USA / Washington / King County	2020-02-28
Washington State Department of Health	Seattle Flu Study	Chu etl al
EPI_ISL_417149	hCoV-19/USA/WA-S96/2020	North America / USA / Washington / King County	2020-02-28
Washington State Department of Health	Seattle Flu Study	Chu etl al
EPI_ISL_417150	hCoV-19/USA/WA-S97/2020	North America / USA / Washington / King County	2020-02-28
Washington State Department of Health	Seattle Flu Study	Chu etl al
EPI_ISL_417151	hCoV-19/USA/WA-S98/2020	North America / USA / Washington / King County	2020-02-29
Washington State Department of Health	Seattle Flu Study	Chu etl al
EPI_ISL_417152	hCoV-19/USA/WA-S99/2020	North America / USA / Washington / King County	2020-02-28
Washington State Department of Health	Seattle Flu Study	Chu etl al
EPI_ISL_417153	hCoV-19/USA/WA-S100/2020	North America / USA / Washington / King County	2020-02-29
Washington State Department of Health	Seattle Flu Study	Chu etl al
EPI_ISL_417154	hCoV-19/USA/WA-S101/2020	North America / USA / Washington / King County	2020-02-28
Washington State Department of Health	Seattle Flu Study	Chu etl al
EPI_ISL_417155	hCoV-19/USA/WA-S102/2020	North America / USA / Washington / King County	2020-02-28
Washington State Department of Health	Seattle Flu Study	Chu etl al
EPI_ISL_417156	hCoV-19/USA/WA-S103/2020	North America / USA / Washington / King County	2020-02-28
Washington State Department of Health	Seattle Flu Study	Chu etl al
EPI_ISL_417157	hCoV-19/USA/WA-S104/2020	North America / USA / Washington / Snohomish County	2020-
03-05	Washington State Department of Health	Seattle Flu Study	Chu etl al
EPI_ISL_417158	hCoV-19/USA/WA-S105/2020	North America / USA / Washington / King County	2020-02-28
Washington State Department of Health	Seattle Flu Study	Chu etl al
EPI_ISL_417159	hCoV-19/USA/WA-S106/2020	North America / USA / Washington / King County	2020-02-29
Washington State Department of Health	Seattle Flu Study	Chu etl al
EPI_ISL_417160	hCoV-19/USA/WA-S107/2020	North America / USA / Washington / King County	2020-02-29
Washington State Department of Health	Seattle Flu Study	Chu etl al
EPI_ISL_417161	hCoV-19/USA/WA-S108/2020	North America / USA / Washington / King County	2020-02-29
Washington State Department of Health	Seattle Flu Study	Chu etl al
EPI_ISL_417162	hCoV-19/USA/WA-S109/2020	North America / USA / Washington / Snohomish County	2020-
03-01	Washington State Department of Health	Seattle Flu Study	Chu etl al
EPI_ISL_417163	hCoV-19/USA/WA-S110/2020	North America / USA / Washington / King County	2020-03-05
Seattle Flu Study	Seattle Flu Study	Chu etl al
EPI_ISL_417164	hCoV-19/USA/WA-S111/2020	North America / USA / Washington / King County	2020-03-07
Seattle Flu Study	Seattle Flu Study	Chu etl al
EPI_ISL_417165	hCoV-19/USA/WA-S112/2020	North America / USA / Washington / King County	2020-03-08
Seattle Flu Study	Seattle Flu Study	Chu etl al
EPI_ISL_417167	hCoV-19/USA/WA-S114/2020	North America / USA / Washington / Snohomish County	2020-
03-05	Washington State Department of Health	Seattle Flu Study	Chu etl al
EPI_ISL_417168	hCoV-19/USA/WA-S115/2020	North America / USA / Washington / King County	2020-02-29
Washington State Department of Health	Seattle Flu Study	Chu etl al
EPI_ISL_417169	hCoV-19/USA/WA-S116/2020	North America / USA / Washington	2020-03-02
Washington State Department of Health	Seattle Flu Study	Chu etl al
EPI_ISL_417170	hCoV-19/USA/WA-S117/2020	North America / USA / Washington	2020-03-02

Washington State Department of Health   Seattle Flu Study       Chu etl al
EPI_ISL_417171  hCoV-19/USA/WA-S118/2020        North America / USA / Washington / King County  2020-03-01
Washington State Department of Health   Seattle Flu Study       Chu etl al
EPI_ISL_417172  hCoV-19/USA/WA-S119/2020        North America / USA / Washington / King County  2020-02-29
Washington State Department of Health   Seattle Flu Study       Chu etl al
EPI_ISL_417175  hCoV-19/USA/WA-S122/2020        North America / USA / Washington / Grant County 2020-03-02
Washington State Department of Health   Seattle Flu Study       Chu etl al
EPI_ISL_417176  hCoV-19/Hong Kong/HKPU1_2101/2020       Asia / Hong Kong        2020-01-21      Department of Pathology, Princess Margaret Hospital   Department of Health Technology and Informatics, Faculty of Health and Social Science, The Hong Kong Polytechnic University    Kenneth Siu-Sing LEUNG, Timothy Ting-Leung NG, Alan Ka-Lun WU, Miranda Chong-Yee YAU, Hiu-Yin LAO, Ming-Pan CHOI, Kingsley King-Gee TAM, Lam-Kwong LEE, Barry Kin-Chung WONG, Alex Yat-Man HO, Kam-Tong Yip, Kwok-Cheung LUNG, Raymond Wai-To LIU, Eugene Yuk-Keung TSO, Wai-Shing LEUNG, Man-Chun CHAN, Yuk-Yung NG, Kit-Man SIN, Kitty Sau-Chun FUNG, Sandy Ka-Yee CHAU, Wing-Kin TO, Tak-Lun Que, David Ho-Keung SHUM, Shea Ping YIP, Wing Cheong YAM, Gilman Kit-Hang SIU
EPI_ISL_417178  hCoV-19/Hong Kong/HKPU6_2101/2020       Asia / Hong Kong        2020-01-25      Department of Pathology, Princess Margaret Hospital   Department of Health Technology and Informatics, Faculty of Health and Social Science, The Hong Kong Polytechnic University    Kenneth Siu-Sing LEUNG, Timothy Ting-Leung NG, Alan Ka-Lun WU, Miranda Chong-Yee YAU, Hiu-Yin LAO, Ming-Pan CHOI, Kingsley King-Gee TAM, Lam-Kwong LEE, Barry Kin-Chung WONG, Alex Yat-Man HO, Kam-Tong Yip, Kwok-Cheung LUNG, Raymond Wai-To LIU, Eugene Yuk-Keung TSO, Wai-Shing LEUNG, Man-Chun CHAN, Yuk-Yung NG, Kit-Man SIN, Kitty Sau-Chun FUNG, Sandy Ka-Yee CHAU, Wing-Kin TO, Tak-Lun Que, David Ho-Keung SHUM, Shea Ping YIP, Wing Cheong YAM, Gilman Kit-Hang SIU
EPI_ISL_417181  hCoV-19/Hong Kong/HKPU19_0402/2020      Asia / Hong Kong        2020-02-05      Department of Pathology, United Christian Hospital    Department of Health Technology and Informatics, Faculty of Health and Social Science, The Hong Kong Polytechnic University    Kenneth Siu-Sing LEUNG, Timothy Ting-Leung NG, Alan Ka-Lun WU, Miranda Chong-Yee YAU, Hiu-Yin LAO, Ming-Pan CHOI, Kingsley King-Gee TAM, Lam-Kwong LEE, Barry Kin-Chung WONG, Alex Yat-Man HO, Kam-Tong Yip, Kwok-Cheung LUNG, Raymond Wai-To LIU, Eugene Yuk-Keung TSO, Wai-Shing LEUNG, Man-Chun CHAN, Yuk-Yung NG, Kit-Man SIN, Kitty Sau-Chun FUNG, Sandy Ka-Yee CHAU, Wing-Kin TO, Tak-Lun Que, David Ho-Keung SHUM, Shea Ping YIP, Wing Cheong YAM, Gilman Kit-Hang SIU
EPI_ISL_417185  hCoV-19/Hong Kong/HKPU28_3001/2020      Asia / Hong Kong        2020-02-09      Department of Pathology, United Christian Hospital    Department of Health Technology and Informatics, Faculty of Health and Social Science, The Hong Kong Polytechnic University    Kenneth Siu-Sing LEUNG, Timothy Ting-Leung NG, Alan Ka-Lun WU, Miranda Chong-Yee YAU, Hiu-Yin LAO, Ming-Pan CHOI, Kingsley King-Gee TAM, Lam-Kwong LEE, Barry Kin-Chung WONG, Alex Yat-Man HO, Kam-Tong Yip, Kwok-Cheung LUNG, Raymond Wai-To LIU, Eugene Yuk-Keung TSO, Wai-Shing LEUNG, Man-Chun CHAN, Yuk-Yung NG, Kit-Man SIN, Kitty Sau-Chun FUNG, Sandy Ka-Yee CHAU, Wing-Kin TO, Tak-Lun Que, David Ho-Keung SHUM, Shea Ping YIP, Wing Cheong YAM, Gilman Kit-Hang SIU
EPI_ISL_417186  hCoV-19/South Africa/R03006/2020        Africa / South Africa / KZN     2020-03-07      National Institute for Communicable Diseases of the National Health Laboratory Service  National Institute for Communicable Diseases of the National Health Laboratory Service Allam M, Kwenda S, van Heusden P, Khumalo Z, Mohale T, Subramoney K, von Gottberg, A, Ismail A, Bhiman JN
EPI_ISL_417187  hCoV-19/Hong Kong/HKPU29_0102/2020      Asia / Hong Kong        2020-02-08      Department of Clinical Pathology, Pamela Youde Nethersole Eastern Hospital    Department of Health Technology and Informatics, Faculty of Health and Social Science, The Hong Kong Polytechnic University Kenneth Siu-Sing LEUNG, Timothy Ting-Leung NG, Alan Ka-Lun WU, Miranda Chong-Yee YAU, Hiu-Yin LAO, Ming-Pan CHOI, Kingsley King-Gee TAM, Lam-Kwong LEE, Barry Kin-Chung WONG, Alex Yat-Man HO, Kam-Tong Yip, Kwok-Cheung LUNG, Raymond Wai-To LIU, Eugene Yuk-Keung TSO, Wai-Shing LEUNG, Man-Chun CHAN, Yuk-Yung NG, Kit-Man SIN, Kitty Sau-Chun FUNG, Sandy Ka-Yee CHAU, Wing-Kin TO, Tak-Lun Que, David Ho-Keung SHUM, Shea Ping YIP, Wing Cheong YAM, Gilman Kit-Hang SIU
EPI_ISL_417188  hCoV-19/Hong Kong/HKPU30_2901/2020      Asia / Hong Kong        2020-02-08      Department of Clinical Pathology, Pamela Youde Nethersole Eastern Hospital    Department of Health Technology and Informatics, Faculty of Health and Social Science, The Hong Kong Polytechnic University Kenneth Siu-Sing LEUNG, Timothy Ting-Leung NG, Alan Ka-Lun WU, Miranda Chong-Yee YAU, Hiu-Yin LAO, Ming-Pan CHOI, Kingsley King-Gee TAM, Lam-Kwong LEE, Barry Kin-Chung WONG, Alex Yat-Man HO, Kam-Tong Yip, Kwok-Cheung LUNG, Raymond Wai-To LIU, Eugene Yuk-Keung TSO, Wai-Shing LEUNG, Man-Chun CHAN, Yuk-Yung NG, Kit-Man SIN, Kitty Sau-Chun FUNG, Sandy Ka-Yee CHAU, Wing-Kin TO, Tak-Lun Que, David Ho-Keung SHUM, Shea Ping YIP, Wing Cheong YAM, Gilman Kit-Hang SIU
EPI_ISL_417191  hCoV-19/USA/MN5-MDH5/2020       North America / USA / Minnesota 2020-03-10      Minnesota Department of Health, Public Health Laboratory  Minnesota Department of Health, Public Health Laboratory Matt Plumb, Jake Garfin and Xiong Wang
EPI_ISL_417192  hCoV-19/USA/MN25-MDH25/2020     North America / USA / Minnesota 2020-03-12      Minnesota Department of Health, Public Health Laboratory  Minnesota Department of Health, Public Health Laboratory Matt Plumb, Jake Garfin and Xiong Wang
EPI_ISL_417193  hCoV-19/Hong Kong/HKPU32_0402/2020      Asia / Hong Kong        2020-02-09      Department of Clinical Pathology, Pamela Youde Nethersole Eastern Hospital    Department of Health Technology and Informatics, Faculty of Health and Social Science, The Hong Kong Polytechnic University Kenneth Siu-Sing LEUNG, Timothy Ting-Leung NG, Alan Ka-Lun WU, Miranda Chong-Yee YAU, Hiu-Yin LAO, Ming-Pan CHOI, Kingsley King-Gee TAM, Lam-Kwong LEE, Barry Kin-Chung WONG, Alex Yat-Man HO, Kam-Tong Yip, Kwok-Cheung LUNG, Raymond Wai-To LIU, Eugene Yuk-Keung TSO, Wai-Shing LEUNG, Man-Chun CHAN, Yuk-Yung NG, Kit-Man SIN, Kitty Sau-Chun FUNG, Sandy Ka-Yee CHAU, Wing-Kin TO, Tak-Lun Que, David Ho-Keung SHUM, Shea Ping YIP, Wing Cheong YAM, Gilman Kit-Hang SIU
EPI_ISL_417194  hCoV-19/USA/MN26-MDH26/2020     North America / USA / Minnesota 2020-03-12      Minnesota Department of Health, Public Health Laboratory  Minnesota Department of Health, Public Health Laboratory Matt Plumb, Jake Garfin and Xiong Wang
EPI_ISL_417196  hCoV-19/USA/MN29-MDH29/2020     North America / USA / Minnesota 2020-03-11      Minnesota

Department of Health, Public Health Laboratory  Minnesota Department of Health, Public Health Laboratory  Matt Plumb, Jake Garfin and Xiong Wang
EPI_ISL_417197  hCoV-19/Hong Kong/HKPU34_3001/2020  Asia / Hong Kong  2020-02-09  Department of Clinical Pathology, Pamela Youde Nethersole Eastern Hospital  Department of Health Technology and Informatics, Faculty of Health and Social Science, The Hong Kong Polytechnic University Kenneth Siu-Sing LEUNG, Timothy Ting-Leung NG, Alan Ka-Lun WU, Miranda Chong-Yee YAU, Hiu-Yin LAO, Ming-Pan CHOI, Kingsley King-Gee TAM, Lam-Kwong LEE, Barry Kin-Chung WONG, Alex Yat-Man HO, Kam-Tong Yip, Kwok-Cheung LUNG, Raymond Wai-To LIU, Eugene Yuk-Keung TSO, Wai-Shing LEUNG, Man-Chun CHAN, Yuk-Yung NG, Kit-Man SIN, Kitty Sau-Chun FUNG, Sandy Ka-Yee CHAU, Wing-Kin TO, Tak-Lun Que, David Ho-Keung SHUM, Shea Ping YIP, Wing Cheong YAM, Gilman Kit-Hang SIU
EPI_ISL_417250  hCoV-19/England/20108004702/2020  Europe / United Kingdom / England  2020-03-02  Respiratory Virus Unit, Microbiology Services Colindale, Public Health England  Respiratory Virus Unit, Microbiology Services Colindale, Public Health England  Monica Galiano, Shahjahan Miah, Angie Lackenby, Omolola Akinbami, Tiina Talts, Leena Bhaw, Richard Myers, Steven Platt, Kirstin Edwards, Jonathan Hubb, Joanna Ellis, Maria Zambon
EPI_ISL_417252  hCoV-19/England/20108006003/2020  Europe / United Kingdom / England  2020-03-05  Respiratory Virus Unit, Microbiology Services Colindale, Public Health England  Respiratory Virus Unit, Microbiology Services Colindale, Public Health England  Monica Galiano, Shahjahan Miah, Angie Lackenby, Omolola Akinbami, Tiina Talts, Leena Bhaw, Richard Myers, Steven Platt, Kirstin Edwards, Jonathan Hubb, Joanna Ellis, Maria Zambon
EPI_ISL_417254  hCoV-19/England/20108006802/2020  Europe / United Kingdom / England  2020-03-04  Respiratory Virus Unit, Microbiology Services Colindale, Public Health England  Respiratory Virus Unit, Microbiology Services Colindale, Public Health England  Monica Galiano, Shahjahan Miah, Angie Lackenby, Omolola Akinbami, Tiina Talts, Leena Bhaw, Richard Myers, Steven Platt, Kirstin Edwards, Jonathan Hubb, Joanna Ellis, Maria Zambon
EPI_ISL_417255  hCoV-19/England/20108007002/2020  Europe / United Kingdom / England  2020-03-04  Respiratory Virus Unit, Microbiology Services Colindale, Public Health England  Respiratory Virus Unit, Microbiology Services Colindale, Public Health England  Monica Galiano, Shahjahan Miah, Angie Lackenby, Omolola Akinbami, Tiina Talts, Leena Bhaw, Richard Myers, Steven Platt, Kirstin Edwards, Jonathan Hubb, Joanna Ellis, Maria Zambon
EPI_ISL_417256  hCoV-19/England/20108007302/2020  Europe / United Kingdom / England  2020-03-03  Respiratory Virus Unit, Microbiology Services Colindale, Public Health England  Respiratory Virus Unit, Microbiology Services Colindale, Public Health England  Monica Galiano, Shahjahan Miah, Angie Lackenby, Omolola Akinbami, Tiina Talts, Leena Bhaw, Richard Myers, Steven Platt, Kirstin Edwards, Jonathan Hubb, Joanna Ellis, Maria Zambon
EPI_ISL_417258  hCoV-19/England/20108034006/2020  Europe / United Kingdom / England  2020-03-06  Respiratory Virus Unit, Microbiology Services Colindale, Public Health England  Respiratory Virus Unit, Microbiology Services Colindale, Public Health England  Monica Galiano, Shahjahan Miah, Angie Lackenby, Omolola Akinbami, Tiina Talts, Leena Bhaw, Richard Myers, Steven Platt, Kirstin Edwards, Jonathan Hubb, Joanna Ellis, Maria Zambon
EPI_ISL_417260  hCoV-19/England/20109035906/2020  Europe / United Kingdom / England  2020-03-05  Respiratory Virus Unit, Microbiology Services Colindale, Public Health England  Respiratory Virus Unit, Microbiology Services Colindale, Public Health England  Monica Galiano, Shahjahan Miah, Angie Lackenby, Omolola Akinbami, Tiina Talts, Leena Bhaw, Richard Myers, Steven Platt, Kirstin Edwards, Jonathan Hubb, Joanna Ellis, Maria Zambon
EPI_ISL_417262  hCoV-19/England/20109039306/2020  Europe / United Kingdom / England  2020-03-06  Respiratory Virus Unit, Microbiology Services Colindale, Public Health England  Respiratory Virus Unit, Microbiology Services Colindale, Public Health England  Monica Galiano, Shahjahan Miah, Angie Lackenby, Omolola Akinbami, Tiina Talts, Leena Bhaw, Richard Myers, Steven Platt, Kirstin Edwards, Jonathan Hubb, Joanna Ellis, Maria Zambon
EPI_ISL_417263  hCoV-19/England/20109050106/2020  Europe / United Kingdom / England  2020-03-06  Respiratory Virus Unit, Microbiology Services Colindale, Public Health England  Respiratory Virus Unit, Microbiology Services Colindale, Public Health England  Monica Galiano, Shahjahan Miah, Angie Lackenby, Omolola Akinbami, Tiina Talts, Leena Bhaw, Richard Myers, Steven Platt, Kirstin Edwards, Jonathan Hubb, Joanna Ellis, Maria Zambon
EPI_ISL_417264  hCoV-19/England/20109050306/2020  Europe / United Kingdom / England  2020-03-04  Respiratory Virus Unit, Microbiology Services Colindale, Public Health England  Respiratory Virus Unit, Microbiology Services Colindale, Public Health England  Monica Galiano, Shahjahan Miah, Angie Lackenby, Omolola Akinbami, Tiina Talts, Leena Bhaw, Richard Myers, Steven Platt, Kirstin Edwards, Jonathan Hubb, Joanna Ellis, Maria Zambon
EPI_ISL_417265  hCoV-19/England/20109050406/2020  Europe / United Kingdom / England  2020-03-08  Respiratory Virus Unit, Microbiology Services Colindale, Public Health England  Respiratory Virus Unit, Microbiology Services Colindale, Public Health England  Monica Galiano, Shahjahan Miah, Angie Lackenby, Omolola Akinbami, Tiina Talts, Leena Bhaw, Richard Myers, Steven Platt, Kirstin Edwards, Jonathan Hubb, Joanna Ellis, Maria Zambon
EPI_ISL_417266  hCoV-19/England/20109050506/2020  Europe / United Kingdom / England  2020-03-05  Respiratory Virus Unit, Microbiology Services Colindale, Public Health England  Respiratory Virus Unit, Microbiology Services Colindale, Public Health England  Monica Galiano, Shahjahan Miah, Angie Lackenby, Omolola Akinbami, Tiina Talts, Leena Bhaw, Richard Myers, Steven Platt, Kirstin Edwards, Jonathan Hubb, Joanna Ellis, Maria Zambon
EPI_ISL_417267  hCoV-19/England/20109050606/2020  Europe / United Kingdom / England  2020-03-05

Respiratory Virus Unit, Microbiology Services Colindale, Public Health England  Respiratory Virus Unit, Microbiology Services Colindale, Public Health England  Monica Galiano, Shahjahan Miah, Angie Lackenby, Omolola Akinbami, Tiina Talts, Leena Bhaw, Richard Myers, Steven Platt, Kirstin Edwards, Jonathan Hubb, Joanna Ellis, Maria Zambon
EPI_ISL_417268  hCoV-19/England/20109050706/2020  Europe / United Kingdom / England  2020-03-05  Respiratory Virus Unit, Microbiology Services Colindale, Public Health England  Respiratory Virus Unit, Microbiology Services Colindale, Public Health England  Monica Galiano, Shahjahan Miah, Angie Lackenby, Omolola Akinbami, Tiina Talts, Leena Bhaw, Richard Myers, Steven Platt, Kirstin Edwards, Jonathan Hubb, Joanna Ellis, Maria Zambon
EPI_ISL_417269  hCoV-19/England/20109050806/2020  Europe / United Kingdom / England  2020-03-05  Respiratory Virus Unit, Microbiology Services Colindale, Public Health England  Respiratory Virus Unit, Microbiology Services Colindale, Public Health England  Monica Galiano, Shahjahan Miah, Angie Lackenby, Omolola Akinbami, Tiina Talts, Leena Bhaw, Richard Myers, Steven Platt, Kirstin Edwards, Jonathan Hubb, Joanna Ellis, Maria Zambon
EPI_ISL_417270  hCoV-19/England/20109051806/2020  Europe / United Kingdom / England  2020-03-06  Respiratory Virus Unit, Microbiology Services Colindale, Public Health England  Respiratory Virus Unit, Microbiology Services Colindale, Public Health England  Monica Galiano, Shahjahan Miah, Angie Lackenby, Omolola Akinbami, Tiina Talts, Leena Bhaw, Richard Myers, Steven Platt, Kirstin Edwards, Jonathan Hubb, Joanna Ellis, Maria Zambon
EPI_ISL_417272  hCoV-19/England/20109052006/2020  Europe / United Kingdom / England  2020-03-06  Respiratory Virus Unit, Microbiology Services Colindale, Public Health England  Respiratory Virus Unit, Microbiology Services Colindale, Public Health England  Monica Galiano, Shahjahan Miah, Angie Lackenby, Omolola Akinbami, Tiina Talts, Leena Bhaw, Richard Myers, Steven Platt, Kirstin Edwards, Jonathan Hubb, Joanna Ellis, Maria Zambon
EPI_ISL_417276  hCoV-19/England/20109052506/2020  Europe / United Kingdom / England  2020-03-07  Respiratory Virus Unit, Microbiology Services Colindale, Public Health England  Respiratory Virus Unit, Microbiology Services Colindale, Public Health England  Monica Galiano, Shahjahan Miah, Angie Lackenby, Omolola Akinbami, Tiina Talts, Leena Bhaw, Richard Myers, Steven Platt, Kirstin Edwards, Jonathan Hubb, Joanna Ellis, Maria Zambon
EPI_ISL_417278  hCoV-19/England/20109053406/2020  Europe / United Kingdom / England  2020-03-07  Respiratory Virus Unit, Microbiology Services Colindale, Public Health England  Respiratory Virus Unit, Microbiology Services Colindale, Public Health England  Monica Galiano, Shahjahan Miah, Angie Lackenby, Omolola Akinbami, Tiina Talts, Leena Bhaw, Richard Myers, Steven Platt, Kirstin Edwards, Jonathan Hubb, Joanna Ellis, Maria Zambon
EPI_ISL_417279  hCoV-19/England/20109053606/2020  Europe / United Kingdom / England  2020-03-07  Respiratory Virus Unit, Microbiology Services Colindale, Public Health England  Respiratory Virus Unit, Microbiology Services Colindale, Public Health England  Monica Galiano, Shahjahan Miah, Angie Lackenby, Omolola Akinbami, Tiina Talts, Leena Bhaw, Richard Myers, Steven Platt, Kirstin Edwards, Jonathan Hubb, Joanna Ellis, Maria Zambon
EPI_ISL_417280  hCoV-19/England/20109054806/2020  Europe / United Kingdom / England  2020-03-07  Respiratory Virus Unit, Microbiology Services Colindale, Public Health England  Respiratory Virus Unit, Microbiology Services Colindale, Public Health England  Monica Galiano, Shahjahan Miah, Angie Lackenby, Omolola Akinbami, Tiina Talts, Leena Bhaw, Richard Myers, Steven Platt, Kirstin Edwards, Jonathan Hubb, Joanna Ellis, Maria Zambon
EPI_ISL_417282  hCoV-19/England/20109056906/2020  Europe / United Kingdom / England  2020-03-05  Respiratory Virus Unit, Microbiology Services Colindale, Public Health England  Respiratory Virus Unit, Microbiology Services Colindale, Public Health England  Monica Galiano, Shahjahan Miah, Angie Lackenby, Omolola Akinbami, Tiina Talts, Leena Bhaw, Richard Myers, Steven Platt, Kirstin Edwards, Jonathan Hubb, Joanna Ellis, Maria Zambon
EPI_ISL_417283  hCoV-19/England/20109058906/2020  Europe / United Kingdom / England  2020-03-06  Respiratory Virus Unit, Microbiology Services Colindale, Public Health England  Respiratory Virus Unit, Microbiology Services Colindale, Public Health England  Monica Galiano, Shahjahan Miah, Angie Lackenby, Omolola Akinbami, Tiina Talts, Leena Bhaw, Richard Myers, Steven Platt, Kirstin Edwards, Jonathan Hubb, Joanna Ellis, Maria Zambon
EPI_ISL_417285  hCoV-19/England/20109093606/2020  Europe / United Kingdom / England  2020-03-06  Respiratory Virus Unit, Microbiology Services Colindale, Public Health England  Respiratory Virus Unit, Microbiology Services Colindale, Public Health England  Monica Galiano, Shahjahan Miah, Angie Lackenby, Omolola Akinbami, Tiina Talts, Leena Bhaw, Richard Myers, Steven Platt, Kirstin Edwards, Jonathan Hubb, Joanna Ellis, Maria Zambon
EPI_ISL_417286  hCoV-19/England/20109093706/2020  Europe / United Kingdom / England  2020-03-05  Respiratory Virus Unit, Microbiology Services Colindale, Public Health England  Respiratory Virus Unit, Microbiology Services Colindale, Public Health England  Monica Galiano, Shahjahan Miah, Angie Lackenby, Omolola Akinbami, Tiina Talts, Leena Bhaw, Richard Myers, Steven Platt, Kirstin Edwards, Jonathan Hubb, Joanna Ellis, Maria Zambon
EPI_ISL_417287  hCoV-19/England/20109093806/2020  Europe / United Kingdom / England  2020-03-06  Respiratory Virus Unit, Microbiology Services Colindale, Public Health England  Respiratory Virus Unit, Microbiology Services Colindale, Public Health England  Monica Galiano, Shahjahan Miah, Angie Lackenby, Omolola Akinbami, Tiina Talts, Leena Bhaw, Richard Myers, Steven Platt, Kirstin Edwards, Jonathan Hubb, Joanna Ellis, Maria Zambon
EPI_ISL_417289  hCoV-19/England/20109094006/2020  Europe / United Kingdom / England  2020-03-06

Respiratory Virus Unit, Microbiology Services Colindale, Public Health England  Respiratory Virus Unit, Microbiology Services Colindale, Public Health England  Monica Galiano, Shahjahan Miah, Angie Lackenby, Omolola Akinbami, Tiina Talts, Leena Bhaw, Richard Myers, Steven Platt, Kirstin Edwards, Jonathan Hubb, Joanna Ellis, Maria Zambon
EPI_ISL_417290  hCoV-19/England/20109094106/2020  Europe / United Kingdom / England  2020-03-06  Respiratory Virus Unit, Microbiology Services Colindale, Public Health England  Respiratory Virus Unit, Microbiology Services Colindale, Public Health England  Monica Galiano, Shahjahan Miah, Angie Lackenby, Omolola Akinbami, Tiina Talts, Leena Bhaw, Richard Myers, Steven Platt, Kirstin Edwards, Jonathan Hubb, Joanna Ellis, Maria Zambon
EPI_ISL_417291  hCoV-19/England/20109094206/2020  Europe / United Kingdom / England  2020-03-08  Respiratory Virus Unit, Microbiology Services Colindale, Public Health England  Respiratory Virus Unit, Microbiology Services Colindale, Public Health England  Monica Galiano, Shahjahan Miah, Angie Lackenby, Omolola Akinbami, Tiina Talts, Leena Bhaw, Richard Myers, Steven Platt, Kirstin Edwards, Jonathan Hubb, Joanna Ellis, Maria Zambon
EPI_ISL_417292  hCoV-19/England/20109094506/2020  Europe / United Kingdom / England  2020-03-08  Respiratory Virus Unit, Microbiology Services Colindale, Public Health England  Respiratory Virus Unit, Microbiology Services Colindale, Public Health England  Monica Galiano, Shahjahan Miah, Angie Lackenby, Omolola Akinbami, Tiina Talts, Leena Bhaw, Richard Myers, Steven Platt, Kirstin Edwards, Jonathan Hubb, Joanna Ellis, Maria Zambon
EPI_ISL_417293  hCoV-19/England/20109097506/2020  Europe / United Kingdom / England  2020-03-08  Respiratory Virus Unit, Microbiology Services Colindale, Public Health England  Respiratory Virus Unit, Microbiology Services Colindale, Public Health England  Monica Galiano, Shahjahan Miah, Angie Lackenby, Omolola Akinbami, Tiina Talts, Leena Bhaw, Richard Myers, Steven Platt, Kirstin Edwards, Jonathan Hubb, Joanna Ellis, Maria Zambon
EPI_ISL_417295  hCoV-19/England/20109098806/2020  Europe / United Kingdom / England  2020-03-08  Respiratory Virus Unit, Microbiology Services Colindale, Public Health England  Respiratory Virus Unit, Microbiology Services Colindale, Public Health England  Monica Galiano, Shahjahan Miah, Angie Lackenby, Omolola Akinbami, Tiina Talts, Leena Bhaw, Richard Myers, Steven Platt, Kirstin Edwards, Jonathan Hubb, Joanna Ellis, Maria Zambon
EPI_ISL_417297  hCoV-19/England/20109099006/2020  Europe / United Kingdom / England  2020-03-06  Respiratory Virus Unit, Microbiology Services Colindale, Public Health England  Respiratory Virus Unit, Microbiology Services Colindale, Public Health England  Monica Galiano, Shahjahan Miah, Angie Lackenby, Omolola Akinbami, Tiina Talts, Leena Bhaw, Richard Myers, Steven Platt, Kirstin Edwards, Jonathan Hubb, Joanna Ellis, Maria Zambon
EPI_ISL_417298  hCoV-19/England/20109099106/2020  Europe / United Kingdom / England  2020-03-08  Respiratory Virus Unit, Microbiology Services Colindale, Public Health England  Respiratory Virus Unit, Microbiology Services Colindale, Public Health England  Monica Galiano, Shahjahan Miah, Angie Lackenby, Omolola Akinbami, Tiina Talts, Leena Bhaw, Richard Myers, Steven Platt, Kirstin Edwards, Jonathan Hubb, Joanna Ellis, Maria Zambon
EPI_ISL_417299  hCoV-19/England/20109099206/2020  Europe / United Kingdom / England  2020-03-08  Respiratory Virus Unit, Microbiology Services Colindale, Public Health England  Respiratory Virus Unit, Microbiology Services Colindale, Public Health England  Monica Galiano, Shahjahan Miah, Angie Lackenby, Omolola Akinbami, Tiina Talts, Leena Bhaw, Richard Myers, Steven Platt, Kirstin Edwards, Jonathan Hubb, Joanna Ellis, Maria Zambon
EPI_ISL_418400  hCoV-19/Finland/13M83/2020  Europe / Finland  2020-03-13  Department of Virology and Immunology, University of Helsinki and Helsinki University Hospital, Huslab Finland Department of Virology, Faculty of Medicine, University of Helsinki, Helsinki, Finland  Teemu Smura, Hannimari Kallio-Kokko, Olli Vapalahti
EPI_ISL_418401  hCoV-19/Finland/14M12/2020  Europe / Finland  2020-03-14  Department of Virology and Immunology, University of Helsinki and Helsinki University Hospital, Huslab Finland Department of Virology, Faculty of Medicine, University of Helsinki, Helsinki, Finland  Teemu Smura, Hannimari Kallio-Kokko, Olli Vapalahti
EPI_ISL_418402  hCoV-19/Finland/14M13/2020  Europe / Finland  2020-03-14  Department of Virology and Immunology, University of Helsinki and Helsinki University Hospital, Huslab Finland Department of Virology, Faculty of Medicine, University of Helsinki, Helsinki, Finland  Teemu Smura, Hannimari Kallio-Kokko, Olli Vapalahti
EPI_ISL_418403  hCoV-19/Finland/14M14/2020  Europe / Finland  2020-03-14  Department of Virology and Immunology, University of Helsinki and Helsinki University Hospital, Huslab Finland Department of Virology, Faculty of Medicine, University of Helsinki, Helsinki, Finland  Teemu Smura, Hannimari Kallio-Kokko, Olli Vapalahti
EPI_ISL_418404  hCoV-19/Finland/14M16/2020  Europe / Finland  2020-03-14  Department of Virology and Immunology, University of Helsinki and Helsinki University Hospital, Huslab Finland Department of Virology, Faculty of Medicine, University of Helsinki, Helsinki, Finland  Teemu Smura, Hannimari Kallio-Kokko, Olli Vapalahti
EPI_ISL_418405  hCoV-19/Finland/14M20/2020  Europe / Finland  2020-03-14  Department of Virology and Immunology, University of Helsinki and Helsinki University Hospital, Huslab Finland Department of Virology, Faculty of Medicine, University of Helsinki, Helsinki, Finland  Teemu Smura, Hannimari Kallio-Kokko, Olli Vapalahti
EPI_ISL_418406  hCoV-19/Finland/14M26/2020  Europe / Finland  2020-03-14  Department of Virology and Immunology, University of Helsinki and Helsinki University Hospital, Huslab Finland Department of Virology,

Faculty of Medicine, University of Helsinki, Helsinki, Finland  Teemu Smura, Hannimari Kallio-Kokko, Olli Vapalahti
EPI_ISL_418408  hCoV-19/Finland/14M32/2020  Europe / Finland / Helsinki  2020-03-14  Department of Virology and Immunology, University of Helsinki and Helsinki University Hospital, Huslab Finland  Department of Virology, Faculty of Medicine, University of Helsinki, Helsinki, Finland  Teemu Smura, Hannimari Kallio-Kokko, Olli Vapalahti
EPI_ISL_418409  hCoV-19/Finland/14M74/2020  Europe / Finland / Helsinki  2020-03-14  Department of Virology and Immunology, University of Helsinki and Helsinki University Hospital, Huslab Finland  Department of Virology, Faculty of Medicine, University of Helsinki, Helsinki, Finland  Teemu Smura, Hannimari Kallio-Kokko, Olli Vapalahti
EPI_ISL_418410  hCoV-19/Finland/14M77/2020  Europe / Finland / Helsinki  2020-03-14  Department of Virology and Immunology, University of Helsinki and Helsinki University Hospital, Huslab Finland  Department of Virology, Faculty of Medicine, University of Helsinki, Helsinki, Finland  Teemu Smura, Hannimari Kallio-Kokko, Olli Vapalahti
EPI_ISL_418411  hCoV-19/Finland/14M82/2020  Europe / Finland / Helsinki  2020-03-14  Department of Virology and Immunology, University of Helsinki and Helsinki University Hospital, Huslab Finland  Department of Virology, Faculty of Medicine, University of Helsinki, Helsinki, Finland  Teemu Smura, Hannimari Kallio-Kokko, Olli Vapalahti
EPI_ISL_418412  hCoV-19/France/ARA094100/2020  Europe / France / ARA / Privas  2020-03-15  Centre Hospitalier des Vals d'Ardeche  CNR Virus des Infections Respiratoires - France SUD  Antonin Bal, Gregory Destras, Gwendolyne Burfin, Solenne Brun, Carine Moustaud, Raphaelle Lamy, Alexandre Gaymard, Maude Bouscambert-Duchamp, Florence Morfin-Sherpa, Martine Valette, Bruno Lina, Laurence Josset
EPI_ISL_418413  hCoV-19/France/ARA09428/2020  Europe / France / ARA / Macon  2020-03-15  Centre Hospitalier de Macon  CNR Virus des Infections Respiratoires - France SUD  Antonin Bal, Gregory Destras, Gwendolyne Burfin, Solenne Brun, Carine Moustaud, Raphaelle Lamy, Alexandre Gaymard, Maude Bouscambert-Duchamp, Florence Morfin-Sherpa, Martine Valette, Bruno Lina, Laurence Josset
EPI_ISL_418414  hCoV-19/France/ARA09434/2020  Europe / France / ARA / Valence 2020-03-15  Centre Hospitalier de Valence  CNR Virus des Infections Respiratoires - France SUD  Antonin Bal, Gregory Destras, Gwendolyne Burfin, Solenne Brun, Carine Moustaud, Raphaelle Lamy, Alexandre Gaymard, Maude Bouscambert-Duchamp, Florence Morfin-Sherpa, Martine Valette, Bruno Lina, Laurence Josset
EPI_ISL_418416  hCoV-19/France/ARA09588/2020  Europe / France / ARA / Venissieux  2020-03-16  GH Les Portes du Sud  CNR Virus des Infections Respiratoires - France SUD  Antonin Bal, Gregory Destras, Gwendolyne Burfin, Solenne Brun, Carine Moustaud, Raphaelle Lamy, Alexandre Gaymard, Maude Bouscambert-Duchamp, Florence Morfin-Sherpa, Martine Valette, Bruno Lina, Laurence Josset
EPI_ISL_418417  hCoV-19/France/ARA09686/2020  Europe / France / ARA / Valence 2020-03-16  Centre Hospitalier de Valence  CNR Virus des Infections Respiratoires - France SUD  Antonin Bal, Gregory Destras, Gwendolyne Burfin, Solenne Brun, Carine Moustaud, Raphaelle Lamy, Alexandre Gaymard, Maude Bouscambert-Duchamp, Florence Morfin-Sherpa, Martine Valette, Bruno Lina, Laurence Josset
EPI_ISL_418418  hCoV-19/France/ARA10163/2020  Europe / France / ARA / Lyon  2020-03-16  Centre Hospitalier Saint Joseph Saint Luc  CNR Virus des Infections Respiratoires - France SUD  Antonin Bal, Gregory Destras, Gwendolyne Burfin, Solenne Brun, Carine Moustaud, Raphaelle Lamy, Alexandre Gaymard, Maude Bouscambert-Duchamp, Florence Morfin-Sherpa, Martine Valette, Bruno Lina, Laurence Josset
EPI_ISL_418419  hCoV-19/France/ARA10165/2020  Europe / France / ARA / Lyon  2020-03-16  Centre Hospitalier Saint Joseph Saint Luc  CNR Virus des Infections Respiratoires - France SUD  Antonin Bal, Gregory Destras, Gwendolyne Burfin, Solenne Brun, Carine Moustaud, Raphaelle Lamy, Alexandre Gaymard, Maude Bouscambert-Duchamp, Florence Morfin-Sherpa, Martine Valette, Bruno Lina, Laurence Josset
EPI_ISL_418420  hCoV-19/France/ARA10170/2020  Europe / France / ARA / Lyon  2020-03-17  Institut des Agents Infectieux (IAI), Hospices Civils de Lyon  CNR Virus des Infections Respiratoires - France SUD  Antonin Bal, Gregory Destras, Gwendolyne Burfin, Solenne Brun, Carine Moustaud, Raphaelle Lamy, Alexandre Gaymard, Maude Bouscambert-Duchamp, Florence Morfin-Sherpa, Martine Valette, Bruno Lina, Laurence Josset
EPI_ISL_418422  hCoV-19/France/ARA10184/2020  Europe / France / ARA / Lyon  2020-03-17  Institut des Agents Infectieux (IAI), Hospices Civils de Lyon  CNR Virus des Infections Respiratoires - France SUD  Antonin Bal, Gregory Destras, Gwendolyne Burfin, Solenne Brun, Carine Moustaud, Raphaelle Lamy, Alexandre Gaymard, Maude Bouscambert-Duchamp, Florence Morfin-Sherpa, Martine Valette, Bruno Lina, Laurence Josset
EPI_ISL_418423  hCoV-19/France/ARA10188/2020  Europe / France / ARA / Lyon  2020-03-17  Institut des Agents Infectieux (IAI), Hospices Civils de Lyon  CNR Virus des Infections Respiratoires - France SUD  Antonin Bal, Gregory Destras, Gwendolyne Burfin, Solenne Brun, Carine Moustaud, Raphaelle Lamy, Alexandre Gaymard, Maude Bouscambert-Duchamp, Florence Morfin-Sherpa, Martine Valette, Bruno Lina, Laurence Josset
EPI_ISL_418424  hCoV-19/France/ARA10189/2020  Europe / France / ARA / Lyon  2020-03-17  Institut des Agents Infectieux (IAI), Hospices Civils de Lyon  CNR Virus des Infections Respiratoires - France SUD  Antonin Bal, Gregory Destras, Gwendolyne Burfin, Solenne Brun, Carine Moustaud, Raphaelle Lamy, Alexandre Gaymard, Maude Bouscambert-Duchamp, Florence Morfin-Sherpa, Martine Valette, Bruno Lina, Laurence Josset
EPI_ISL_418425  hCoV-19/France/ARA10192/2020  Europe / France / ARA / Lyon  2020-03-17  Institut des Agents Infectieux (IAI), Hospices Civils de Lyon  CNR Virus des Infections Respiratoires - France SUD  Antonin Bal, Gregory Destras, Gwendolyne Burfin, Solenne Brun, Carine Moustaud, Raphaelle Lamy, Alexandre Gaymard, Maude Bouscambert-Duchamp, Florence Morfin-Sherpa, Martine Valette, Bruno Lina, Laurence Josset
EPI_ISL_418426  hCoV-19/France/ARA10251/2020  Europe / France / ARA / Bourg-en-Bresse 2020-03-17  Centre Hospitalier de Bourg en Bresse  CNR Virus des Infections Respiratoires - France SUD  Antonin Bal, Gregory Destras, Gwendolyne Burfin, Solenne Brun, Carine Moustaud, Raphaelle Lamy, Alexandre Gaymard, Maude Bouscambert-Duchamp, Florence Morfin-Sherpa, Martine Valette, Bruno Lina, Laurence Josset

EPI_ISL_418427 hCoV-19/France/ARA10257/2020 Europe / France / ARA / Saint-Priest 2020-03-17 Hopital Privé de l'Est Lyonnais CNR Virus des Infections Respiratoires - France SUD Antonin Bal, Gregory Destras, Gwendolyne Burfin, Solenne Brun, Carine Moustaud, Raphaelle Lamy, Alexandre Gaymard, Maude Bouscambert-Duchamp, Florence Morfin-Sherpa, Martine Valette, Bruno Lina, Laurence Josset
EPI_ISL_418428 hCoV-19/France/ARA10282/2020 Europe / France / ARA / Vienne 2020-03-17 Centre Hospitalier Lucien Hussel CNR Virus des Infections Respiratoires - France SUD Antonin Bal, Gregory Destras, Gwendolyne Burfin, Solenne Brun, Carine Moustaud, Raphaelle Lamy, Alexandre Gaymard, Maude Bouscambert-Duchamp, Florence Morfin-Sherpa, Martine Valette, Bruno Lina, Laurence Josset
EPI_ISL_418429 hCoV-19/France/ARA10876/2020 Europe / France / ARA / Lyon 2020-03-18 Institut des Agents Infectieux (IAI), Hospices Civils de Lyon CNR Virus des Infections Respiratoires - France SUD Antonin Bal, Gregory Destras, Gwendolyne Burfin, Solenne Brun, Carine Moustaud, Raphaelle Lamy, Alexandre Gaymard, Maude Bouscambert-Duchamp, Florence Morfin-Sherpa, Martine Valette, Bruno Lina, Laurence Josset
EPI_ISL_418430 hCoV-19/France/ARA10910/2020 Europe / France / ARA / Lyon 2020-03-18 Institut des Agents Infectieux (IAI), Hospices Civils de Lyon CNR Virus des Infections Respiratoires - France SUD Antonin Bal, Gregory Destras, Gwendolyne Burfin, Solenne Brun, Carine Moustaud, Raphaelle Lamy, Alexandre Gaymard, Maude Bouscambert-Duchamp, Florence Morfin-Sherpa, Martine Valette, Bruno Lina, Laurence Josset
EPI_ISL_418431 hCoV-19/France/ARA10968/2020 Europe / France / ARA / Lyon 2020-03-18 Institut des Agents Infectieux (IAI), Hospices Civils de Lyon CNR Virus des Infections Respiratoires - France SUD Antonin Bal, Gregory Destras, Gwendolyne Burfin, Solenne Brun, Carine Moustaud, Raphaelle Lamy, Alexandre Gaymard, Maude Bouscambert-Duchamp, Florence Morfin-Sherpa, Martine Valette, Bruno Lina, Laurence Josset
EPI_ISL_418432 hCoV-19/France/ARA11036/2020 Europe / France / ARA / Lyon 2020-03-18 Institut des Agents Infectieux (IAI), Hospices Civils de Lyon CNR Virus des Infections Respiratoires - France SUD Antonin Bal, Gregory Destras, Gwendolyne Burfin, Solenne Brun, Carine Moustaud, Raphaelle Lamy, Alexandre Gaymard, Maude Bouscambert-Duchamp, Florence Morfin-Sherpa, Martine Valette, Bruno Lina, Laurence Josset
EPI_ISL_418441 hCoV-19/Hangzhou/HZ48/2020 Asia / China / Hangzhou 2020-01-21 Hangzhou Center for Disease Control and Prevention Inspection Center of Hangzhou Center for Disease Control and Prevention Yu hua, Wang haoqiu, Li jun, Yu xinfeng, Pan jingcao
EPI_ISL_418442 hCoV-19/Hangzhou/HZ49/2020 Asia / China / Hangzhou 2020-01-21 Hangzhou Center for Disease Control and Prevention Inspection Center of Hangzhou Center for Disease Control and Prevention Yu hua, Wang haoqiu, Li jun, Yu xinfeng, Pan jingcao
EPI_ISL_418502 hCoV-19/Hangzhou/HZ60/2020 Asia / China / Hangzhou 2020-01-22 Hangzhou Center for Disease Control and Prevention Inspection Center of Hangzhou Center for Disease Control and Prevention Yu hua, Wang haoqiu, Li jun, Yu xinfeng, Pan jingcao
EPI_ISL_418503 hCoV-19/Hangzhou/HZ62/2020 Asia / China / Hangzhou 2020-01-22 Hangzhou Center for Disease Control and Prevention Inspection Center of Hangzhou Center for Disease Control and Prevention Yu hua, Wang haoqiu, Li jun, Yu xinfeng, Pan jingcao
EPI_ISL_418504 hCoV-19/Hangzhou/HZ79/2020 Asia / China / Hangzhou 2020-01-21 Hangzhou Center for Disease Control and Prevention Inspection Center of Hangzhou Center for Disease Control and Prevention Yu hua, Wang haoqiu, Li jun, Yu xinfeng, Pan jingcao
EPI_ISL_418506 hCoV-19/Hangzhou/HZ90/2020 Asia / China / Hangzhou 2020-01-21 Hangzhou Center for Disease Control and Prevention Inspection Center of Hanghzou Center for Disease Control and Prevention Yu hua, Wang haoqiu, Li jun, Yu xinfeng, Pan jingcao
EPI_ISL_418507 hCoV-19/Hangzhou/HZ91/2020 Asia/China/Zhejiang/Hangzhou 2020-01-21 Hangzhou Center for Disease Control and Prevention Inspection Center of Hangzhou Center for Disease Control and Prevention Yu hua, Wang haoqiu, Li jun, Yu xinfeng, Pan jingcao
EPI_ISL_418508 hCoV-19/Hangzhou/HZ162/2020 Asia / China / Zhejiang / Hangzhou 2020-01-23 Hangzhou Center for Disease Control and Prevention Inspection Center of Hangzhou Center for Disease Control and Prevention Yu hua, Wang haoqiu, Li jun, Yu xinfeng, Pan jingcao
EPI_ISL_418509 hCoV-19/Hangzhou/HZ178/2020 Asia / China / Zhejiang / Hangzhou 2020-01-23 Hangzhou Center for Disease Control and Prevention Inspection Center of Hangzhou Center for Disease Control and Prevention Yu hua, Wang haoqiu, Li jun, Yu xinfeng, Pan jingcao
EPI_ISL_418510 hCoV-19/Hangzhou/HZ185/2020 Asia / China / Hangzhou 2020-01-23 Hangzhou Center for Disease Control and Prevention Insepction Center of Hangzhou Center for Disease Control and Prevention Yu hua, Wang haoqiu, Li jun, Yu xinfeng, Pan jingcao
EPI_ISL_418511 hCoV-19/Hangzhou/HZ477/2020 Asia / China / Zhejiang / Hangzhou 2020-01-24 Hangzhou Center for Disease Control and Prevention Inspection Center of Hangzhou Center for Disease Control and Prevention Yu hua, Wang haoqiu, Li jun, Yu xinfeng, Pan jingcao
EPI_ISL_418512 hCoV-19/Hangzhou/HZ481/2020 Asia / China / Zhejiang / Hangzhou 2020-01-25 Hangzhou Center for Disease Control and Prevention Inspection Center of Hangzhou Center for Disease Control and Prevention Yu hua, Wang haoqiu, Li jun, Yu xinfeng, Pan jingcao
EPI_ISL_418513 hCoV-19/Hangzhou/HZ551/2020 Asia / China / Hangzhou 2020-01-25 Hangzhou Center for Disease Control and Prevention Inspection Center of Hangzhou Center for Disease Control and Prevention Yu hua, Wang haoqiu, Li jun, Yu xinfeng, Pan jingcao
EPI_ISL_418514 hCoV-19/Hangzhou/HZ576/2020 Asia / China / Zhejiang / Hangzhou 2020-01-25 Hangzhou Center for Disease Control and Prevention Inspection Center of Hangzhou Center for Disease Control and Prevention Yu hua, Wang haoqiu, Li jun, Yu xinfeng, Pan jingcao
EPI_ISL_418515 hCoV-19/Hangzhou/HZ638/2020 Asia / China / Hangzhou 2020-01-25 Hangzhou Center for Disease Control and Prevention Inspection Center of Hangzhou Center for Disease Control and Prevention Yu hua, Wang haoqiu, Li jun, Yu xinfeng, Pan jingcao
EPI_ISL_418516 hCoV-19/Ireland/21023/2020 Europe / Ireland / Tipperary 2020-03-06 UCD National

Virus Reference Laboratory	UCD National Virus Reference Laboratory	Michael Carr, Gabriel Gonzalez, Jonathan Dean, Suzie Coughlan, Alison Murphy, Kevin Byrne, Ken Wolfe, Jeff Connell, Brendan Loftus, Cillian F De Gascun
EPI_ISL_418580	hCoV-19/Ireland/Dublin-22361/2020	Europe / Ireland / Cork	2020-03-08	UCD National Virus Reference Laboratory	UCD National Virus Reference Laboratory	Michael Carr, Gabriel Gonzalez, Jonathan Dean, Suzie Coughlan, Alison Murphy, Kevin Byrne, Ken Wolfe, Jeff Connell, Brendan Loftus, Cillian F De Gascun
EPI_ISL_418581	hCoV-19/Ireland/Dublin-22428/2020	Europe / Ireland / Dublin	2020-03-08	UCD National Virus Reference Laboratory	UCD National Virus Reference Laboratory	Michael Carr, Gabriel Gonzalez, Jonathan Dean, Suzie Coughlan, Alison Murphy, Kevin Byrne, Ken Wolfe, Jeff Connell, Brendan Loftus, Cillian F De Gascun
EPI_ISL_418582	hCoV-19/Ireland/22901/2020	Europe / Ireland / Louth	2020-03-10	UCD National Virus Reference Laboratory	UCD National Virus Reference Laboratory	Michael Carr, Gabriel Gonzalez, Jonathan Dean, Suzie Coughlan, Alison Murphy, Kevin Byrne, Ken Wolfe, Jeff Connell, Brendan Loftus, Cillian F De Gascun
EPI_ISL_418583	hCoV-19/Ireland/24042/2020	Europe / Ireland / Dublin	2020-03-10	UCD National Virus Reference Laboratory	UCD National Virus Reference Laboratory	Michael Carr, Gabriel Gonzalez, Jonathan Dean, Suzie Coughlan, Alison Murphy, Kevin Byrne, Ken Wolfe, Jeff Connell, Brendan Loftus, Cillian F De Gascun
EPI_ISL_418584	hCoV-19/Ireland/24052/2020	Europe / Ireland / Wicklow	2020-03-10	UCD National Virus Reference Laboratory	UCD National Virus Reference Laboratory	Michael Carr, Gabriel Gonzalez, Jonathan Dean, Suzie Coughlan, Alison Murphy, Kevin Byrne, Ken Wolfe, Jeff Connell, Brendan Loftus, Cillian F De Gascun
EPI_ISL_419700	hCoV-19/USA/NY-NYUMC39/2020	North America / USA / New York / Manhattan	2020-03-18	NYU Langone Health	Departments of Pathology and Medicine, New York University School of Medicine	Maria Aguero-Rosenfeld, Margaret Black, John Cadley, Paolo Cotzia, John Chen, Dacia Dimartino, Xiaojun Feng, Adriana Heguy, Megan Hogan, Emily Huang, George Jour, Christian Marier, Matthew T. Maurano, Mark J. Mulligan, Peter Meyn, Jared Pinnell, Sitharam Ramaswami, Amy Rapkiewicz, Marie Samanovic-Golden, Antonio Serrano, Guomiao Shen, Matija Snuderl, Nick Vulpescu, Gael Westby, Paul Zappile, Yutong Zhang
EPI_ISL_419702	hCoV-19/USA/NY-NYUMC41/2020	North America / USA / New York / Brooklyn	2020-03-18	NYU Langone Health	Departments of Pathology and Medicine, New York University School of Medicine	Maria Aguero-Rosenfeld, Margaret Black, John Cadley, Paolo Cotzia, John Chen, Dacia Dimartino, Xiaojun Feng, Adriana Heguy, Megan Hogan, Emily Huang, George Jour, Christian Marier, Matthew T. Maurano, Mark J. Mulligan, Peter Meyn, Jared Pinnell, Sitharam Ramaswami, Amy Rapkiewicz, Marie Samanovic-Golden, Antonio Serrano, Guomiao Shen, Matija Snuderl, Nick Vulpescu, Gael Westby, Paul Zappile, Yutong Zhang
EPI_ISL_419703	hCoV-19/USA/NY-NYUMC42/2020	North America / USA / New York / Manhattan	2020-03-18	NYU Langone Health	Departments of Pathology and Medicine, New York University School of Medicine	Maria Aguero-Rosenfeld, Margaret Black, John Cadley, Paolo Cotzia, John Chen, Dacia Dimartino, Xiaojun Feng, Adriana Heguy, Megan Hogan, Emily Huang, George Jour, Christian Marier, Matthew T. Maurano, Mark J. Mulligan, Peter Meyn, Jared Pinnell, Sitharam Ramaswami, Amy Rapkiewicz, Marie Samanovic-Golden, Antonio Serrano, Guomiao Shen, Matija Snuderl, Nick Vulpescu, Gael Westby, Paul Zappile, Yutong Zhang
EPI_ISL_419704	hCoV-19/USA/NY-NYUMC43/2020	North America / USA / New York / Manhattan	2020-03-18	NYU Langone Health	Departments of Pathology and Medicine, New York University School of Medicine	Maria Aguero-Rosenfeld, Margaret Black, John Cadley, Paolo Cotzia, John Chen, Dacia Dimartino, Xiaojun Feng, Adriana Heguy, Megan Hogan, Emily Huang, George Jour, Christian Marier, Matthew T. Maurano, Mark J. Mulligan, Peter Meyn, Jared Pinnell, Sitharam Ramaswami, Amy Rapkiewicz, Marie Samanovic-Golden, Antonio Serrano, Guomiao Shen, Matija Snuderl, Nick Vulpescu, Gael Westby, Paul Zappile, Yutong Zhang
EPI_ISL_419705	hCoV-19/USA/NY-NYUMC44/2020	North America / USA / New York / Manhattan	2020-03-18	NYU Langone Health	Departments of Pathology and Medicine, New York University School of Medicine	Maria Aguero-Rosenfeld, Margaret Black, John Cadley, Paolo Cotzia, John Chen, Dacia Dimartino, Xiaojun Feng, Adriana Heguy, Megan Hogan, Emily Huang, George Jour, Christian Marier, Matthew T. Maurano, Mark J. Mulligan, Peter Meyn, Jared Pinnell, Sitharam Ramaswami, Amy Rapkiewicz, Marie Samanovic-Golden, Antonio Serrano, Guomiao Shen, Matija Snuderl, Nick Vulpescu, Gael Westby, Paul Zappile, Yutong Zhang
EPI_ISL_419706	hCoV-19/USA/VA-DCLS-0012/2020	North America / USA / Virginia	2020-03-10	Virginia DCLS	Virginia DCLS	Virginia DCLS
EPI_ISL_419707	hCoV-19/Spain/Cataluna201397/2020	Europe / Spain / Catalonia	2020	HOSPITAL CLINIC	Instituto de Salud Carlos III	Iglesias-Caballero, M. Molinero Calamita, M. González-Esguevillas, M. Camarero S. Pozo F. Casas I. Jiménez, P. Jiménez, M. Zaballos, A. Monzón, S. Varona, S. Juliá, M. Cuesta, I. Marcos, M.A
EPI_ISL_419708	hCoV-19/USA/VA-DCLS-0014/2020	North America / USA / Virginia	2020-03-12	Virginia DCLS	Virginia DCLS	Virginia DCLS
EPI_ISL_419709	hCoV-19/Spain/PaisVasco201602/2020	Europe / Spain / BasqueCountry	2020-03-04	HOSPITAL TXAGORRITXU	Instituto de Salud Carlos III	Iglesias-Caballero, M. Molinero Calamita, M. González-Esguevillas, M. Camarero S. Pozo F. Casas I. Jiménez, P. Jiménez, M. Zaballos, A. Monzón, S. Varona, S. Juliá, M. Cuesta, I. Gómez, C.
EPI_ISL_419710	hCoV-19/USA/VA-DCLS-0016/2020	North America / USA / Virginia	2020-03-12	Virginia DCLS	Virginia DCLS	Virginia DCLS
EPI_ISL_419711	hCoV-19/USA/VA-DCLS-0017/2020	North America / USA / Virginia	2020-03-11	Virginia DCLS	Virginia DCLS	Virginia DCLS
EPI_ISL_419712	hCoV-19/USA/VA-DCLS-0020/2020	North America / USA / Virginia	2020-03-10	Virginia DCLS	Virginia DCLS	Virginia DCLS

EPI_ISL_419713 hCoV-19/USA/VA-DCLS-0021/2020 North America / USA / Virginia 2020-03-11 Virginia DCLS Virginia DCLS Virginia DCLS
EPI_ISL_419714 hCoV-19/Australia/VIC108/2020 Oceania / Australia / Victoria 2020-03-18 Microbiological Diagnostic Unit Public Health Laboratory Microbiological Diagnostic Unit Public Health Laboratory Seemann T., Schultz M., Sait, M., Sherry, N.
EPI_ISL_419715 hCoV-19/Australia/VIC111/2020 Oceania / Australia / Victoria 2020-03-18 Microbiological Diagnostic Unit Public Health Laboratory Microbiological Diagnostic Unit Public Health Laboratory Seemann T., Schultz M., Sait, M., Sherry, N.
EPI_ISL_419716 hCoV-19/Australia/VIC110/2020 Oceania / Australia / Victoria 2020-03-18 Microbiological Diagnostic Unit Public Health Laboratory Microbiological Diagnostic Unit Public Health Laboratory Seemann T., Schultz M., Sait, M., Sherry, N.
EPI_ISL_419717 hCoV-19/Australia/VIC115/2020 Oceania / Australia / Victoria 2020-03-19 Microbiological Diagnostic Unit Public Health Laboratory Microbiological Diagnostic Unit Public Health Laboratory Seemann T., Schultz M., Sait, M., Sherry, N.
EPI_ISL_419718 hCoV-19/Australia/VIC109/2020 Oceania / Australia / Victoria 2020-03-18 Microbiological Diagnostic Unit Public Health Laboratory Microbiological Diagnostic Unit Public Health Laboratory Seemann T., Schultz M., Sait, M., Sherry, N.
EPI_ISL_419719 hCoV-19/Australia/VIC118/2020 Oceania / Australia / Victoria 2020-03-20 Microbiological Diagnostic Unit Public Health Laboratory Microbiological Diagnostic Unit Public Health Laboratory Seemann T., Schultz M., Sait, M., Sherry, N.
EPI_ISL_419720 hCoV-19/Australia/VIC107/2020 Oceania / Australia / Victoria 2020-03-17 Microbiological Diagnostic Unit Public Health Laboratory Microbiological Diagnostic Unit Public Health Laboratory Seemann T., Schultz M., Sait, M., Sherry, N.
EPI_ISL_419721 hCoV-19/Australia/VIC116/2020 Oceania / Australia / Victoria 2020-03-19 Microbiological Diagnostic Unit Public Health Laboratory Microbiological Diagnostic Unit Public Health Laboratory Seemann T., Schultz M., Sait, M., Sherry, N.
EPI_ISL_419722 hCoV-19/Australia/VIC120/2020 Oceania / Australia / Victoria 2020-03-19 Microbiological Diagnostic Unit Public Health Laboratory Microbiological Diagnostic Unit Public Health Laboratory Seemann T., Schultz M., Sait, M., Sherry, N.
EPI_ISL_419723 hCoV-19/Australia/VIC121/2020 Oceania / Australia / Victoria 2020-03-18 Microbiological Diagnostic Unit Public Health Laboratory Microbiological Diagnostic Unit Public Health Laboratory Seemann T., Schultz M., Sait, M., Sherry, N.
EPI_ISL_419725 hCoV-19/Australia/VIC114/2020 Oceania / Australia / Victoria 2020-03-18 Microbiological Diagnostic Unit Public Health Laboratory Microbiological Diagnostic Unit Public Health Laboratory Seemann T., Schultz M., Sait, M., Sherry, N.
EPI_ISL_419726 hCoV-19/Australia/VIC119/2020 Oceania / Australia / Victoria 2020-03-19 Microbiological Diagnostic Unit Public Health Laboratory Microbiological Diagnostic Unit Public Health Laboratory Seemann T., Schultz M., Sait, M., Sherry, N.
EPI_ISL_419727 hCoV-19/Australia/VIC117/2020 Oceania / Australia / Victoria 2020-03-19 Microbiological Diagnostic Unit Public Health Laboratory Microbiological Diagnostic Unit Public Health Laboratory Seemann T., Schultz M., Sait, M., Sherry, N.
EPI_ISL_419728 hCoV-19/Australia/VIC126/2020 Oceania / Australia / Victoria 2020-03-20 Microbiological Diagnostic Unit Public Health Laboratory Microbiological Diagnostic Unit Public Health Laboratory Seemann T., Schultz M., Sait, M., Sherry, N.
EPI_ISL_419729 hCoV-19/Australia/VIC127/2020 Oceania / Australia / Victoria 2020-03-20 Microbiological Diagnostic Unit Public Health Laboratory Microbiological Diagnostic Unit Public Health Laboratory Seemann T., Schultz M., Sait, M., Sherry, N.
EPI_ISL_419730 hCoV-19/Australia/VIC128/2020 Oceania / Australia / Victoria 2020-03-20 Microbiological Diagnostic Unit Public Health Laboratory Microbiological Diagnostic Unit Public Health Laboratory Seemann T., Schultz M., Sait, M., Sherry, N.
EPI_ISL_419731 hCoV-19/Australia/VIC123/2020 Oceania / Australia / Victoria 2020-03-20 Microbiological Diagnostic Unit Public Health Laboratory Microbiological Diagnostic Unit Public Health Laboratory Seemann T., Schultz M., Sait, M., Sherry, N.
EPI_ISL_419732 hCoV-19/Australia/VIC129/2020 Oceania / Australia / Victoria 2020-03-21 Microbiological Diagnostic Unit Public Health Laboratory Microbiological Diagnostic Unit Public Health Laboratory Seemann T., Schultz M., Sait, M., Sherry, N.
EPI_ISL_419733 hCoV-19/Australia/VIC13/2020 Oceania / Australia / Victoria 2020-01-31 Victorian Infectious Diseases Reference Laboratory (VIDRL) Victorian Infectious Diseases Reference Laboratory and Microbiological Diagnostic Unit Public Health Laboratory, Doherty Institute Caly L., Seemann T., Sait, M., Schultz M., Druce J., Sherry, N.
EPI_ISL_419736 hCoV-19/Australia/VIC17/2020 Oceania / Australia / Victoria 2020-03-07 Victorian Infectious Diseases Reference Laboratory (VIDRL) Victorian Infectious Diseases Reference Laboratory and Microbiological Diagnostic Unit Public Health Laboratory, Doherty Institute Caly L., Seemann T., Sait, M., Schultz M., Druce J., Sherry, N.
EPI_ISL_419737 hCoV-19/Australia/VIC18/2020 Oceania / Australia / Victoria 2020-03-07 Victorian Infectious Diseases Reference Laboratory (VIDRL) Victorian Infectious Diseases Reference Laboratory and Microbiological Diagnostic Unit Public Health Laboratory, Doherty Institute Caly L., Seemann T., Sait, M., Schultz M., Druce J., Sherry, N.
EPI_ISL_419739 hCoV-19/Australia/VIC24/2020 Oceania / Australia / Victoria 2020-03-09 Victorian Infectious Diseases Reference Laboratory (VIDRL) Victorian Infectious Diseases Reference Laboratory and

Microbiological Diagnostic Unit Public Health Laboratory, Doherty Institute	Caly L., Seemann T., Sait, M., Schultz M., Druce J., Sherry, N.
EPI_ISL_419740	hCoV-19/Australia/VIC21/2020	Oceania / Australia / Victoria	2020-03-09	Victorian Infectious Diseases Reference Laboratory (VIDRL)	Victorian Infectious Diseases Reference Laboratory and Microbiological Diagnostic Unit Public Health Laboratory, Doherty Institute	Caly L., Seemann T., Sait, M., Schultz M., Druce J., Sherry, N.
EPI_ISL_419741	hCoV-19/Australia/VIC20/2020	Oceania / Australia / Victoria	2020-03-08	Victorian Infectious Diseases Reference Laboratory (VIDRL)	Victorian Infectious Diseases Reference Laboratory and Microbiological Diagnostic Unit Public Health Laboratory, Doherty Institute	Caly L., Seemann T., Sait, M., Schultz M., Druce J., Sherry, N.
EPI_ISL_419742	hCoV-19/Australia/VIC22/2020	Oceania / Australia / Victoria	2020-03-09	Victorian Infectious Diseases Reference Laboratory (VIDRL)	Victorian Infectious Diseases Reference Laboratory and Microbiological Diagnostic Unit Public Health Laboratory, Doherty Institute	Caly L., Seemann T., Sait, M., Schultz M., Druce J., Sherry, N.
EPI_ISL_419743	hCoV-19/Australia/VIC23/2020	Oceania / Australia / Victoria	2020-03-09	Victorian Infectious Diseases Reference Laboratory (VIDRL)	Victorian Infectious Diseases Reference Laboratory and Microbiological Diagnostic Unit Public Health Laboratory, Doherty Institute	Caly L., Seemann T., Sait, M., Schultz M., Druce J., Sherry, N.
EPI_ISL_419744	hCoV-19/Australia/VIC25/2020	Oceania / Australia / Victoria	2020-03-10	Victorian Infectious Diseases Reference Laboratory (VIDRL)	Victorian Infectious Diseases Reference Laboratory and Microbiological Diagnostic Unit Public Health Laboratory, Doherty Institute	Caly L., Seemann T., Sait, M., Schultz M., Druce J., Sherry, N.
EPI_ISL_419745	hCoV-19/Australia/VIC26/2020	Oceania / Australia / Victoria	2020-03-10	Victorian Infectious Diseases Reference Laboratory (VIDRL)	Victorian Infectious Diseases Reference Laboratory and Microbiological Diagnostic Unit Public Health Laboratory, Doherty Institute	Caly L., Seemann T., Sait, M., Schultz M., Druce J., Sherry, N.
EPI_ISL_419746	hCoV-19/Australia/VIC27/2020	Oceania / Australia / Victoria	2020-03-10	Victorian Infectious Diseases Reference Laboratory (VIDRL)	Victorian Infectious Diseases Reference Laboratory and Microbiological Diagnostic Unit Public Health Laboratory, Doherty Institute	Caly L., Seemann T., Sait, M., Schultz M., Druce J., Sherry, N.
EPI_ISL_419747	hCoV-19/Australia/VIC28/2020	Oceania / Australia / Victoria	2020-03-10	Victorian Infectious Diseases Reference Laboratory (VIDRL)	Victorian Infectious Diseases Reference Laboratory and Microbiological Diagnostic Unit Public Health Laboratory, Doherty Institute	Caly L., Seemann T., Sait, M., Schultz M., Druce J., Sherry, N.
EPI_ISL_419748	hCoV-19/Australia/VIC29/2020	Oceania / Australia / Victoria	2020-03-10	Victorian Infectious Diseases Reference Laboratory (VIDRL)	Victorian Infectious Diseases Reference Laboratory and Microbiological Diagnostic Unit Public Health Laboratory, Doherty Institute	Caly L., Seemann T., Sait, M., Schultz M., Druce J., Sherry, N.
EPI_ISL_419749	hCoV-19/Australia/VIC30/2020	Oceania / Australia / Victoria	2020-03-10	Victorian Infectious Diseases Reference Laboratory (VIDRL)	Victorian Infectious Diseases Reference Laboratory and Microbiological Diagnostic Unit Public Health Laboratory, Doherty Institute	Caly L., Seemann T., Sait, M., Schultz M., Druce J., Sherry, N.
EPI_ISL_419750	hCoV-19/Australia/VIC31/2020	Oceania / Australia / Victoria	2020-03-10	Victorian Infectious Diseases Reference Laboratory (VIDRL)	Victorian Infectious Diseases Reference Laboratory and Microbiological Diagnostic Unit Public Health Laboratory, Doherty Institute	Caly L., Seemann T., Sait, M., Schultz M., Druce J., Sherry, N.
EPI_ISL_419751	hCoV-19/Australia/VIC32/2020	Oceania / Australia / Victoria	2020-03-10	Victorian Infectious Diseases Reference Laboratory (VIDRL)	Victorian Infectious Diseases Reference Laboratory and Microbiological Diagnostic Unit Public Health Laboratory, Doherty Institute	Caly L., Seemann T., Sait, M., Schultz M., Druce J., Sherry, N.
EPI_ISL_419752	hCoV-19/Australia/VIC33/2020	Oceania / Australia / Victoria	2020-03-10	Victorian Infectious Diseases Reference Laboratory (VIDRL)	Victorian Infectious Diseases Reference Laboratory and Microbiological Diagnostic Unit Public Health Laboratory, Doherty Institute	Caly L., Seemann T., Sait, M., Schultz M., Druce J., Sherry, N.
EPI_ISL_419753	hCoV-19/Australia/VIC34/2020	Oceania / Australia / Victoria	2020-03-10	Victorian Infectious Diseases Reference Laboratory (VIDRL)	Victorian Infectious Diseases Reference Laboratory and Microbiological Diagnostic Unit Public Health Laboratory, Doherty Institute	Caly L., Seemann T., Sait, M., Schultz M., Druce J., Sherry, N.
EPI_ISL_419754	hCoV-19/Australia/VIC37/2020	Oceania / Australia / Victoria	2020-03-11	Victorian Infectious Diseases Reference Laboratory (VIDRL)	Victorian Infectious Diseases Reference Laboratory and Microbiological Diagnostic Unit Public Health Laboratory, Doherty Institute	Caly L., Seemann T., Sait, M., Schultz M., Druce J., Sherry, N.
EPI_ISL_419755	hCoV-19/Australia/VIC35/2020	Oceania / Australia / Victoria	2020-03-10	Victorian Infectious Diseases Reference Laboratory (VIDRL)	Victorian Infectious Diseases Reference Laboratory and Microbiological Diagnostic Unit Public Health Laboratory, Doherty Institute	Caly L., Seemann T., Sait, M., Schultz M., Druce J., Sherry, N.
EPI_ISL_419756	hCoV-19/Australia/VIC38/2020	Oceania / Australia / Victoria	2020-03-11	Victorian Infectious Diseases Reference Laboratory (VIDRL)	Victorian Infectious Diseases Reference Laboratory and Microbiological Diagnostic Unit Public Health Laboratory, Doherty Institute	Caly L., Seemann T., Sait, M., Schultz M., Druce J., Sherry, N.

EPI_ISL_419757 hCoV-19/Australia/VIC39/2020 Oceania / Australia / Victoria 2020-03-11 Victorian Infectious Diseases Reference Laboratory (VIDRL) Victorian Infectious Diseases Reference Laboratory and Microbiological Diagnostic Unit Public Health Laboratory, Doherty Institute Caly L., Seemann T., Sait, M., Schultz M., Druce J., Sherry, N.
EPI_ISL_419758 hCoV-19/Australia/VIC40/2020 Oceania / Australia / Victoria 2020-03-11 Victorian Infectious Diseases Reference Laboratory (VIDRL) Victorian Infectious Diseases Reference Laboratory and Microbiological Diagnostic Unit Public Health Laboratory, Doherty Institute Caly L., Seemann T., Sait, M., Schultz M., Druce J., Sherry, N.
EPI_ISL_419759 hCoV-19/Australia/VIC47/2020 Oceania / Australia / Victoria 2020-03-12 Victorian Infectious Diseases Reference Laboratory (VIDRL) Victorian Infectious Diseases Reference Laboratory and Microbiological Diagnostic Unit Public Health Laboratory, Doherty Institute Caly L., Seemann T., Sait, M., Schultz M., Druce J., Sherry, N.
EPI_ISL_419760 hCoV-19/Australia/VIC41/2020 Oceania / Australia / Victoria 2020-03-11 Victorian Infectious Diseases Reference Laboratory (VIDRL) Victorian Infectious Diseases Reference Laboratory and Microbiological Diagnostic Unit Public Health Laboratory, Doherty Institute Caly L., Seemann T., Sait, M., Schultz M., Druce J., Sherry, N.
EPI_ISL_419761 hCoV-19/Australia/VIC42/2020 Oceania / Australia / Victoria 2020-03-11 Victorian Infectious Diseases Reference Laboratory (VIDRL) Victorian Infectious Diseases Reference Laboratory and Microbiological Diagnostic Unit Public Health Laboratory, Doherty Institute Caly L., Seemann T., Sait, M., Schultz M., Druce J., Sherry, N.
EPI_ISL_419762 hCoV-19/Australia/VIC48/2020 Oceania / Australia / Victoria 2020-03-12 Victorian Infectious Diseases Reference Laboratory (VIDRL) Victorian Infectious Diseases Reference Laboratory and Microbiological Diagnostic Unit Public Health Laboratory, Doherty Institute Caly L., Seemann T., Sait, M., Schultz M., Druce J., Sherry, N.
EPI_ISL_419763 hCoV-19/Australia/VIC43/2020 Oceania / Australia / Victoria 2020-03-11 Victorian Infectious Diseases Reference Laboratory (VIDRL) Victorian Infectious Diseases Reference Laboratory and Microbiological Diagnostic Unit Public Health Laboratory, Doherty Institute Caly L., Seemann T., Sait, M., Schultz M., Druce J., Sherry, N.
EPI_ISL_419764 hCoV-19/Australia/VIC49/2020 Oceania / Australia / Victoria 2020-03-12 Victorian Infectious Diseases Reference Laboratory (VIDRL) Victorian Infectious Diseases Reference Laboratory and Microbiological Diagnostic Unit Public Health Laboratory, Doherty Institute Caly L., Seemann T., Sait, M., Schultz M., Druce J., Sherry, N.
EPI_ISL_419765 hCoV-19/Australia/VIC36/2020 Oceania / Australia / Victoria 2020-03-10 Victorian Infectious Diseases Reference Laboratory (VIDRL) Victorian Infectious Diseases Reference Laboratory and Microbiological Diagnostic Unit Public Health Laboratory, Doherty Institute Caly L., Seemann T., Sait, M., Schultz M., Druce J., Sherry, N.
EPI_ISL_419766 hCoV-19/Australia/VIC50/2020 Oceania / Australia / Victoria 2020-03-12 Victorian Infectious Diseases Reference Laboratory (VIDRL) Victorian Infectious Diseases Reference Laboratory and Microbiological Diagnostic Unit Public Health Laboratory, Doherty Institute Caly L., Seemann T., Sait, M., Schultz M., Druce J., Sherry, N.
EPI_ISL_419767 hCoV-19/Australia/VIC44/2020 Oceania / Australia / Victoria 2020-03-11 Victorian Infectious Diseases Reference Laboratory (VIDRL) Victorian Infectious Diseases Reference Laboratory and Microbiological Diagnostic Unit Public Health Laboratory, Doherty Institute Caly L., Seemann T., Sait, M., Schultz M., Druce J., Sherry, N.
EPI_ISL_419768 hCoV-19/Australia/VIC52/2020 Oceania / Australia / Victoria 2020-03-12 Victorian Infectious Diseases Reference Laboratory (VIDRL) Victorian Infectious Diseases Reference Laboratory and Microbiological Diagnostic Unit Public Health Laboratory, Doherty Institute Caly L., Seemann T., Sait, M., Schultz M., Druce J., Sherry, N.
EPI_ISL_419769 hCoV-19/Australia/VIC53/2020 Oceania / Australia / Victoria 2020-03-12 Victorian Infectious Diseases Reference Laboratory (VIDRL) Victorian Infectious Diseases Reference Laboratory and Microbiological Diagnostic Unit Public Health Laboratory, Doherty Institute Caly L., Seemann T., Sait, M., Schultz M., Druce J., Sherry, N.
EPI_ISL_419770 hCoV-19/Australia/VIC54/2020 Oceania / Australia / Victoria 2020-03-12 Victorian Infectious Diseases Reference Laboratory (VIDRL) Victorian Infectious Diseases Reference Laboratory and Microbiological Diagnostic Unit Public Health Laboratory, Doherty Institute Caly L., Seemann T., Sait, M., Schultz M., Druce J., Sherry, N.
EPI_ISL_419771 hCoV-19/Australia/VIC59/2020 Oceania / Australia / Victoria 2020-03-13 Victorian Infectious Diseases Reference Laboratory (VIDRL) Victorian Infectious Diseases Reference Laboratory and Microbiological Diagnostic Unit Public Health Laboratory, Doherty Institute Caly L., Seemann T., Sait, M., Schultz M., Druce J., Sherry, N.
EPI_ISL_419772 hCoV-19/Australia/VIC60/2020 Oceania / Australia / Victoria 2020-03-13 Victorian Infectious Diseases Reference Laboratory (VIDRL) Victorian Infectious Diseases Reference Laboratory and Microbiological Diagnostic Unit Public Health Laboratory, Doherty Institute Caly L., Seemann T., Sait, M., Schultz M., Druce J., Sherry, N.
EPI_ISL_419773 hCoV-19/Australia/VIC61/2020 Oceania / Australia / Victoria 2020-03-13 Victorian Infectious Diseases Reference Laboratory (VIDRL) Victorian Infectious Diseases Reference Laboratory and Microbiological Diagnostic Unit Public Health Laboratory, Doherty Institute Caly L., Seemann T., Sait, M., Schultz M., Druce J., Sherry, N.
EPI_ISL_419774 hCoV-19/Australia/VIC62/2020 Oceania / Australia / Victoria 2020-03-13 Victorian Infectious Diseases Reference Laboratory (VIDRL) Victorian Infectious Diseases Reference Laboratory and

Microbiological Diagnostic Unit Public Health Laboratory, Doherty Institute	Caly L., Seemann T., Sait, M., Schultz M., Druce J., Sherry, N.
EPI_ISL_419775	hCoV-19/Australia/VIC63/2020	Oceania / Australia / Victoria	2020-03-13	Victorian Infectious Diseases Reference Laboratory (VIDRL)	Victorian Infectious Diseases Reference Laboratory and Microbiological Diagnostic Unit Public Health Laboratory, Doherty Institute	Caly L., Seemann T., Sait, M., Schultz M., Druce J., Sherry, N.
EPI_ISL_419776	hCoV-19/Australia/VIC55/2020	Oceania / Australia / Victoria	2020-03-12	Victorian Infectious Diseases Reference Laboratory (VIDRL)	Victorian Infectious Diseases Reference Laboratory and Microbiological Diagnostic Unit Public Health Laboratory, Doherty Institute	Caly L., Seemann T., Sait, M., Schultz M., Druce J., Sherry, N.
EPI_ISL_419777	hCoV-19/Australia/VIC56/2020	Oceania / Australia / Victoria	2020-03-12	Victorian Infectious Diseases Reference Laboratory (VIDRL)	Victorian Infectious Diseases Reference Laboratory and Microbiological Diagnostic Unit Public Health Laboratory, Doherty Institute	Caly L., Seemann T., Sait, M., Schultz M., Druce J., Sherry, N.
EPI_ISL_419778	hCoV-19/Australia/VIC64/2020	Oceania / Australia / Victoria	2020-03-13	Victorian Infectious Diseases Reference Laboratory (VIDRL)	Victorian Infectious Diseases Reference Laboratory and Microbiological Diagnostic Unit Public Health Laboratory, Doherty Institute	Caly L., Seemann T., Sait, M., Schultz M., Druce J., Sherry, N.
EPI_ISL_419779	hCoV-19/Australia/VIC57/2020	Oceania / Australia / Victoria	2020-03-12	Victorian Infectious Diseases Reference Laboratory (VIDRL)	Victorian Infectious Diseases Reference Laboratory and Microbiological Diagnostic Unit Public Health Laboratory, Doherty Institute	Caly L., Seemann T., Sait, M., Schultz M., Druce J., Sherry, N.
EPI_ISL_419780	hCoV-19/Australia/VIC66/2020	Oceania / Australia / Victoria	2020-03-13	Victorian Infectious Diseases Reference Laboratory (VIDRL)	Victorian Infectious Diseases Reference Laboratory and Microbiological Diagnostic Unit Public Health Laboratory, Doherty Institute	Caly L., Seemann T., Sait, M., Schultz M., Druce J., Sherry, N.
EPI_ISL_419781	hCoV-19/Australia/VIC67/2020	Oceania / Australia / Victoria	2020-03-13	Victorian Infectious Diseases Reference Laboratory (VIDRL)	Victorian Infectious Diseases Reference Laboratory and Microbiological Diagnostic Unit Public Health Laboratory, Doherty Institute	Caly L., Seemann T., Sait, M., Schultz M., Druce J., Sherry, N.
EPI_ISL_419782	hCoV-19/Australia/VIC68/2020	Oceania / Australia / Victoria	2020-03-13	Victorian Infectious Diseases Reference Laboratory (VIDRL)	Victorian Infectious Diseases Reference Laboratory and Microbiological Diagnostic Unit Public Health Laboratory, Doherty Institute	Caly L., Seemann T., Sait, M., Schultz M., Druce J., Sherry, N.
EPI_ISL_419783	hCoV-19/Australia/VIC69/2020	Oceania / Australia / Victoria	2020-03-13	Victorian Infectious Diseases Reference Laboratory (VIDRL)	Victorian Infectious Diseases Reference Laboratory and Microbiological Diagnostic Unit Public Health Laboratory, Doherty Institute	Caly L., Seemann T., Sait, M., Schultz M., Druce J., Sherry, N.
EPI_ISL_419784	hCoV-19/Australia/VIC58/2020	Oceania / Australia / Victoria	2020-03-12	Victorian Infectious Diseases Reference Laboratory (VIDRL)	Victorian Infectious Diseases Reference Laboratory and Microbiological Diagnostic Unit Public Health Laboratory, Doherty Institute	Caly L., Seemann T., Sait, M., Schultz M., Druce J., Sherry, N.
EPI_ISL_419785	hCoV-19/Australia/VIC73/2020	Oceania / Australia / Victoria	2020-03-14	Victorian Infectious Diseases Reference Laboratory (VIDRL)	Victorian Infectious Diseases Reference Laboratory and Microbiological Diagnostic Unit Public Health Laboratory, Doherty Institute	Caly L., Seemann T., Sait, M., Schultz M., Druce J., Sherry, N.
EPI_ISL_419786	hCoV-19/Australia/VIC70/2020	Oceania / Australia / Victoria	2020-03-13	Victorian Infectious Diseases Reference Laboratory (VIDRL)	Victorian Infectious Diseases Reference Laboratory and Microbiological Diagnostic Unit Public Health Laboratory, Doherty Institute	Caly L., Seemann T., Sait, M., Schultz M., Druce J., Sherry, N.
EPI_ISL_419787	hCoV-19/Australia/VIC75/2020	Oceania / Australia / Victoria	2020-03-14	Victorian Infectious Diseases Reference Laboratory (VIDRL)	Victorian Infectious Diseases Reference Laboratory and Microbiological Diagnostic Unit Public Health Laboratory, Doherty Institute	Caly L., Seemann T., Sait, M., Schultz M., Druce J., Sherry, N.
EPI_ISL_419788	hCoV-19/Australia/VIC76/2020	Oceania / Australia / Victoria	2020-03-14	Victorian Infectious Diseases Reference Laboratory (VIDRL)	Victorian Infectious Diseases Reference Laboratory and Microbiological Diagnostic Unit Public Health Laboratory, Doherty Institute	Caly L., Seemann T., Sait, M., Schultz M., Druce J., Sherry, N.
EPI_ISL_419789	hCoV-19/Australia/VIC77/2020	Oceania / Australia / Victoria	2020-03-14	Victorian Infectious Diseases Reference Laboratory (VIDRL)	Victorian Infectious Diseases Reference Laboratory and Microbiological Diagnostic Unit Public Health Laboratory, Doherty Institute	Caly L., Seemann T., Sait, M., Schultz M., Druce J., Sherry, N.
EPI_ISL_419790	hCoV-19/Australia/VIC78/2020	Oceania / Australia / Victoria	2020-03-14	Victorian Infectious Diseases Reference Laboratory (VIDRL)	Victorian Infectious Diseases Reference Laboratory and Microbiological Diagnostic Unit Public Health Laboratory, Doherty Institute	Caly L., Seemann T., Sait, M., Schultz M., Druce J., Sherry, N.
EPI_ISL_419791	hCoV-19/Australia/VIC79/2020	Oceania / Australia / Victoria	2020-03-14	Victorian Infectious Diseases Reference Laboratory (VIDRL)	Victorian Infectious Diseases Reference Laboratory and Microbiological Diagnostic Unit Public Health Laboratory, Doherty Institute	Caly L., Seemann T., Sait, M., Schultz M., Druce J., Sherry, N.

EPI_ISL_419792 hCoV-19/Australia/VIC80/2020 Oceania / Australia / Victoria 2020-03-14 Victorian Infectious Diseases Reference Laboratory (VIDRL) Victorian Infectious Diseases Reference Laboratory and Microbiological Diagnostic Unit Public Health Laboratory, Doherty Institute Caly L., Seemann T., Sait, M., Schultz M., Druce J., Sherry, N.
EPI_ISL_419793 hCoV-19/Australia/VIC81/2020 Oceania / Australia / Victoria 2020-03-14 Victorian Infectious Diseases Reference Laboratory (VIDRL) Victorian Infectious Diseases Reference Laboratory and Microbiological Diagnostic Unit Public Health Laboratory, Doherty Institute Caly L., Seemann T., Sait, M., Schultz M., Druce J., Sherry, N.
EPI_ISL_419794 hCoV-19/Australia/VIC82/2020 Oceania / Australia / Victoria 2020-03-14 Victorian Infectious Diseases Reference Laboratory (VIDRL) Victorian Infectious Diseases Reference Laboratory and Microbiological Diagnostic Unit Public Health Laboratory, Doherty Institute Caly L., Seemann T., Sait, M., Schultz M., Druce J., Sherry, N.
EPI_ISL_419795 hCoV-19/Australia/VIC85/2020 Oceania / Australia / Victoria 2020-03-15 Victorian Infectious Diseases Reference Laboratory (VIDRL) Victorian Infectious Diseases Reference Laboratory and Microbiological Diagnostic Unit Public Health Laboratory, Doherty Institute Caly L., Seemann T., Sait, M., Schultz M., Druce J., Sherry, N.
EPI_ISL_419796 hCoV-19/Australia/VIC86/2020 Oceania / Australia / Victoria 2020-03-15 Victorian Infectious Diseases Reference Laboratory (VIDRL) Victorian Infectious Diseases Reference Laboratory and Microbiological Diagnostic Unit Public Health Laboratory, Doherty Institute Caly L., Seemann T., Sait, M., Schultz M., Druce J., Sherry, N.
EPI_ISL_419797 hCoV-19/Australia/VIC87/2020 Oceania / Australia / Victoria 2020-03-15 Victorian Infectious Diseases Reference Laboratory (VIDRL) Victorian Infectious Diseases Reference Laboratory and Microbiological Diagnostic Unit Public Health Laboratory, Doherty Institute Caly L., Seemann T., Sait, M., Schultz M., Druce J., Sherry, N.
EPI_ISL_419798 hCoV-19/Australia/VIC88/2020 Oceania / Australia / Victoria 2020-03-15 Victorian Infectious Diseases Reference Laboratory (VIDRL) Victorian Infectious Diseases Reference Laboratory and Microbiological Diagnostic Unit Public Health Laboratory, Doherty Institute Caly L., Seemann T., Sait, M., Schultz M., Druce J., Sherry, N.
EPI_ISL_419829 hCoV-19/Australia/VIC136/2020 Oceania / Australia / Victoria 2020-03-22 Microbiological Diagnostic Unit Public Health Laboratory Microbiological Diagnostic Unit Public Health Laboratory Seemann T., Schultz M., Sait, M., Sherry, N.
EPI_ISL_419830 hCoV-19/Australia/VIC137/2020 Oceania / Australia / Victoria 2020-03-22 Microbiological Diagnostic Unit Public Health Laboratory Microbiological Diagnostic Unit Public Health Laboratory Seemann T., Schultz M., Sait, M., Sherry, N.
EPI_ISL_419831 hCoV-19/Australia/NT01/2020 Oceania / Australia / Northern Territory 2020-02-21 Royal Darwin Hospital Victorian Infectious Diseases Reference Laboratory and Microbiological Diagnostic Unit Public Health Laboratory, Doherty Institute Meumann, E., Seemann T., Sait, M., Schultz M., Caly L., Druce J.
EPI_ISL_419832 hCoV-19/Australia/NT02/2020 Oceania / Australia / Northern Territory 2020-02-21 Royal Darwin Hospital Victorian Infectious Diseases Reference Laboratory and Microbiological Diagnostic Unit Public Health Laboratory, Doherty Institute Meumann, E., Seemann T., Sait, M., Schultz M., Caly L., Druce J.
EPI_ISL_419834 hCoV-19/Australia/VIC138/2020 Oceania / Australia / Victoria 2020-02-23 Victorian Infectious Diseases Reference Laboratory (VIDRL) Victorian Infectious Diseases Reference Laboratory and Microbiological Diagnostic Unit Public Health Laboratory, Doherty Institute Caly L., Seemann T., Sait, M., Schultz M., Druce J., Sherry, N.
EPI_ISL_419835 hCoV-19/Australia/NT05/2020 Oceania / Australia / Northern Territory 2020-02-24 Royal Darwin Hospital Victorian Infectious Diseases Reference Laboratory and Microbiological Diagnostic Unit Public Health Laboratory, Doherty Institute Meumann, E., Seemann T., Sait, M., Schultz M., Caly L., Druce J.
EPI_ISL_419836 hCoV-19/Australia/VIC139/2020 Oceania / Australia / Victoria 2020-03-15 Victorian Infectious Diseases Reference Laboratory (VIDRL) Victorian Infectious Diseases Reference Laboratory and Microbiological Diagnostic Unit Public Health Laboratory, Doherty Institute Caly L., Seemann T., Sait, M., Schultz M., Druce J., Sherry, N.
EPI_ISL_419837 hCoV-19/Australia/VIC140/2020 Oceania / Australia / Victoria 2020-03-16 Victorian Infectious Diseases Reference Laboratory (VIDRL) Victorian Infectious Diseases Reference Laboratory and Microbiological Diagnostic Unit Public Health Laboratory, Doherty Institute Caly L., Seemann T., Sait, M., Schultz M., Druce J., Sherry, N.
EPI_ISL_419838 hCoV-19/Australia/VIC141/2020 Oceania / Australia / Victoria 2020-03-16 Victorian Infectious Diseases Reference Laboratory (VIDRL) Victorian Infectious Diseases Reference Laboratory and Microbiological Diagnostic Unit Public Health Laboratory, Doherty Institute Caly L., Seemann T., Sait, M., Schultz M., Druce J., Sherry, N.
EPI_ISL_419839 hCoV-19/Australia/VIC142/2020 Oceania / Australia / Victoria 2020-03-16 Victorian Infectious Diseases Reference Laboratory (VIDRL) Victorian Infectious Diseases Reference Laboratory and Microbiological Diagnostic Unit Public Health Laboratory, Doherty Institute Caly L., Seemann T., Sait, M., Schultz M., Druce J., Sherry, N.
EPI_ISL_419840 hCoV-19/Australia/VIC143/2020 Oceania / Australia / Victoria 2020-03-16 Victorian Infectious Diseases Reference Laboratory (VIDRL) Victorian Infectious Diseases Reference Laboratory and Microbiological Diagnostic Unit Public Health Laboratory, Doherty Institute Caly L., Seemann T., Sait, M., Schultz M., Druce J., Sherry, N.

EPI_ISL_419841 hCoV-19/Australia/VIC144/2020 Oceania / Australia / Victoria 2020-03-17 Victorian Infectious Diseases Reference Laboratory (VIDRL) Victorian Infectious Diseases Reference Laboratory and Microbiological Diagnostic Unit Public Health Laboratory, Doherty Institute Caly L., Seemann T., Sait, M., Schultz M., Druce J., Sherry, N.
EPI_ISL_419842 hCoV-19/Australia/VIC145/2020 Oceania / Australia / Victoria 2020-03-17 Victorian Infectious Diseases Reference Laboratory (VIDRL) Victorian Infectious Diseases Reference Laboratory and Microbiological Diagnostic Unit Public Health Laboratory, Doherty Institute Caly L., Seemann T., Sait, M., Schultz M., Druce J., Sherry, N.
EPI_ISL_419843 hCoV-19/Australia/VIC146/2020 Oceania / Australia / Victoria 2020-03-17 Victorian Infectious Diseases Reference Laboratory (VIDRL) Victorian Infectious Diseases Reference Laboratory and Microbiological Diagnostic Unit Public Health Laboratory, Doherty Institute Caly L., Seemann T., Sait, M., Schultz M., Druce J., Sherry, N.
EPI_ISL_419844 hCoV-19/Australia/VIC147/2020 Oceania / Australia / Victoria 2020-03-17 Victorian Infectious Diseases Reference Laboratory (VIDRL) Victorian Infectious Diseases Reference Laboratory and Microbiological Diagnostic Unit Public Health Laboratory, Doherty Institute Caly L., Seemann T., Sait, M., Schultz M., Druce J., Sherry, N.
EPI_ISL_419845 hCoV-19/Australia/VIC148/2020 Oceania / Australia / Victoria 2020-03-17 Victorian Infectious Diseases Reference Laboratory (VIDRL) Victorian Infectious Diseases Reference Laboratory and Microbiological Diagnostic Unit Public Health Laboratory, Doherty Institute Caly L., Seemann T., Sait, M., Schultz M., Druce J., Sherry, N.
EPI_ISL_419846 hCoV-19/Australia/VIC149/2020 Oceania / Australia / Victoria 2020-03-17 Victorian Infectious Diseases Reference Laboratory (VIDRL) Victorian Infectious Diseases Reference Laboratory and Microbiological Diagnostic Unit Public Health Laboratory, Doherty Institute Caly L., Seemann T., Sait, M., Schultz M., Druce J., Sherry, N.
EPI_ISL_419847 hCoV-19/Australia/VIC150/2020 Oceania / Australia / Victoria 2020-03-17 Victorian Infectious Diseases Reference Laboratory (VIDRL) Victorian Infectious Diseases Reference Laboratory and Microbiological Diagnostic Unit Public Health Laboratory, Doherty Institute Caly L., Seemann T., Sait, M., Schultz M., Druce J., Sherry, N.
EPI_ISL_419848 hCoV-19/Australia/VIC151/2020 Oceania / Australia / Victoria 2020-03-17 Victorian Infectious Diseases Reference Laboratory (VIDRL) Victorian Infectious Diseases Reference Laboratory and Microbiological Diagnostic Unit Public Health Laboratory, Doherty Institute Caly L., Seemann T., Sait, M., Schultz M., Druce J., Sherry, N.
EPI_ISL_419849 hCoV-19/Australia/VIC152/2020 Oceania / Australia / Victoria 2020-03-17 Victorian Infectious Diseases Reference Laboratory (VIDRL) Victorian Infectious Diseases Reference Laboratory and Microbiological Diagnostic Unit Public Health Laboratory, Doherty Institute Caly L., Seemann T., Sait, M., Schultz M., Druce J., Sherry, N.
EPI_ISL_420714 hCoV-19/England/20139055204/2020 Europe / United Kingdom / England 2020-03-28 Respiratory Virus Unit, Microbiology Services Colindale, Public Health England Respiratory Virus Unit, Microbiology Services Colindale, Public Health England Monica Galiano, Shahjahan Miah, Angie Lackenby, Omolola Akinbami, Tiina Talts, Leena Bhaw, Richard Myers, Steven Platt, Kirstin Edwards, Jonathan Hubb, Joanna Ellis, Maria Zambon
EPI_ISL_420715 hCoV-19/England/20139055304/2020 Europe / United Kingdom / England 2020-03-29 Respiratory Virus Unit, Microbiology Services Colindale, Public Health England Respiratory Virus Unit, Microbiology Services Colindale, Public Health England Monica Galiano, Shahjahan Miah, Angie Lackenby, Omolola Akinbami, Tiina Talts, Leena Bhaw, Richard Myers, Steven Platt, Kirstin Edwards, Jonathan Hubb, Joanna Ellis, Maria Zambon
EPI_ISL_420716 hCoV-19/England/20139056904/2020 Europe / United Kingdom / England 2020-03-26 Respiratory Virus Unit, Microbiology Services Colindale, Public Health England Respiratory Virus Unit, Microbiology Services Colindale, Public Health England Monica Galiano, Shahjahan Miah, Angie Lackenby, Omolola Akinbami, Tiina Talts, Leena Bhaw, Richard Myers, Steven Platt, Kirstin Edwards, Jonathan Hubb, Joanna Ellis, Maria Zambon
EPI_ISL_420717 hCoV-19/England/20139057004/2020 Europe / United Kingdom / England 2020-03-26 Respiratory Virus Unit, Microbiology Services Colindale, Public Health England Respiratory Virus Unit, Microbiology Services Colindale, Public Health England Monica Galiano, Shahjahan Miah, Angie Lackenby, Omolola Akinbami, Tiina Talts, Leena Bhaw, Richard Myers, Steven Platt, Kirstin Edwards, Jonathan Hubb, Joanna Ellis, Maria Zambon
EPI_ISL_420718 hCoV-19/England/20139057204/2020 Europe / United Kingdom / England 2020-03-27 Respiratory Virus Unit, Microbiology Services Colindale, Public Health England Respiratory Virus Unit, Microbiology Services Colindale, Public Health England Monica Galiano, Shahjahan Miah, Angie Lackenby, Omolola Akinbami, Tiina Talts, Leena Bhaw, Richard Myers, Steven Platt, Kirstin Edwards, Jonathan Hubb, Joanna Ellis, Maria Zambon
EPI_ISL_420719 hCoV-19/England/20139057404/2020 Europe / United Kingdom / England 2020-03-27 Respiratory Virus Unit, Microbiology Services Colindale, Public Health England Respiratory Virus Unit, Microbiology Services Colindale, Public Health England Monica Galiano, Shahjahan Miah, Angie Lackenby, Omolola Akinbami, Tiina Talts, Leena Bhaw, Richard Myers, Steven Platt, Kirstin Edwards, Jonathan Hubb, Joanna Ellis, Maria Zambon
EPI_ISL_420720 hCoV-19/England/20139057504/2020 Europe / United Kingdom / England 2020-03-28 Respiratory Virus Unit, Microbiology Services Colindale, Public Health England Respiratory Virus Unit, Microbiology Services Colindale, Public Health England Monica Galiano, Shahjahan Miah, Angie Lackenby, Omolola Akinbami, Tiina Talts, Leena Bhaw, Richard Myers, Steven Platt, Kirstin Edwards, Jonathan Hubb, Joanna Ellis,

Maria Zambon
EPI_ISL_420721  hCoV-19/England/20139057604/2020        Europe / United Kingdom / England       2020-03-28
Respiratory Virus Unit, Microbiology Services Colindale, Public Health England  Respiratory Virus Unit, Microbiology Services Colindale, Public Health England  Monica Galiano, Shahjahan Miah, Angie Lackenby, Omolola Akinbami, Tiina Talts, Leena Bhaw, Richard Myers, Steven Platt, Kirstin Edwards, Jonathan Hubb, Joanna Ellis, Maria Zambon
EPI_ISL_420722  hCoV-19/England/20139057704/2020        Europe / United Kingdom / England       2020-03-28
Respiratory Virus Unit, Microbiology Services Colindale, Public Health England  Respiratory Virus Unit, Microbiology Services Colindale, Public Health England  Monica Galiano, Shahjahan Miah, Angie Lackenby, Omolola Akinbami, Tiina Talts, Leena Bhaw, Richard Myers, Steven Platt, Kirstin Edwards, Jonathan Hubb, Joanna Ellis, Maria Zambon
EPI_ISL_420723  hCoV-19/England/20139058104/2020        Europe / United Kingdom / England       2020-03-28
Respiratory Virus Unit, Microbiology Services Colindale, Public Health England  Respiratory Virus Unit, Microbiology Services Colindale, Public Health England  Monica Galiano, Shahjahan Miah, Angie Lackenby, Omolola Akinbami, Tiina Talts, Leena Bhaw, Richard Myers, Steven Platt, Kirstin Edwards, Jonathan Hubb, Joanna Ellis, Maria Zambon
EPI_ISL_420724  hCoV-19/England/20139058304/2020        Europe / United Kingdom / England       2020-03-27
Respiratory Virus Unit, Microbiology Services Colindale, Public Health England  Respiratory Virus Unit, Microbiology Services Colindale, Public Health England  Monica Galiano, Shahjahan Miah, Angie Lackenby, Omolola Akinbami, Tiina Talts, Leena Bhaw, Richard Myers, Steven Platt, Kirstin Edwards, Jonathan Hubb, Joanna Ellis, Maria Zambon
EPI_ISL_420727  hCoV-19/England/20139059004/2020        Europe / United Kingdom / England       2020-03-28
Respiratory Virus Unit, Microbiology Services Colindale, Public Health England  Respiratory Virus Unit, Microbiology Services Colindale, Public Health England  Monica Galiano, Shahjahan Miah, Angie Lackenby, Omolola Akinbami, Tiina Talts, Leena Bhaw, Richard Myers, Steven Platt, Kirstin Edwards, Jonathan Hubb, Joanna Ellis, Maria Zambon
EPI_ISL_420728  hCoV-19/England/20139059504/2020        Europe / United Kingdom / England       2020-03-28
Respiratory Virus Unit, Microbiology Services Colindale, Public Health England  Respiratory Virus Unit, Microbiology Services Colindale, Public Health England  Monica Galiano, Shahjahan Miah, Angie Lackenby, Omolola Akinbami, Tiina Talts, Leena Bhaw, Richard Myers, Steven Platt, Kirstin Edwards, Jonathan Hubb, Joanna Ellis, Maria Zambon
EPI_ISL_420729  hCoV-19/England/20139059804/2020        Europe / United Kingdom / England       2020-03-28
Respiratory Virus Unit, Microbiology Services Colindale, Public Health England  Respiratory Virus Unit, Microbiology Services Colindale, Public Health England  Monica Galiano, Shahjahan Miah, Angie Lackenby, Omolola Akinbami, Tiina Talts, Leena Bhaw, Richard Myers, Steven Platt, Kirstin Edwards, Jonathan Hubb, Joanna Ellis, Maria Zambon
EPI_ISL_420730  hCoV-19/England/20139059904/2020        Europe / United Kingdom / England       2020-03-27
Respiratory Virus Unit, Microbiology Services Colindale, Public Health England  Respiratory Virus Unit, Microbiology Services Colindale, Public Health England  Monica Galiano, Shahjahan Miah, Angie Lackenby, Omolola Akinbami, Tiina Talts, Leena Bhaw, Richard Myers, Steven Platt, Kirstin Edwards, Jonathan Hubb, Joanna Ellis, Maria Zambon
EPI_ISL_420731  hCoV-19/England/20139060004/2020        Europe / United Kingdom / England       2020-03-28
Respiratory Virus Unit, Microbiology Services Colindale, Public Health England  Respiratory Virus Unit, Microbiology Services Colindale, Public Health England  Monica Galiano, Shahjahan Miah, Angie Lackenby, Omolola Akinbami, Tiina Talts, Leena Bhaw, Richard Myers, Steven Platt, Kirstin Edwards, Jonathan Hubb, Joanna Ellis, Maria Zambon
EPI_ISL_420732  hCoV-19/England/20139060404/2020        Europe / United Kingdom / England       2020-03-28
Respiratory Virus Unit, Microbiology Services Colindale, Public Health England  Respiratory Virus Unit, Microbiology Services Colindale, Public Health England  Monica Galiano, Shahjahan Miah, Angie Lackenby, Omolola Akinbami, Tiina Talts, Leena Bhaw, Richard Myers, Steven Platt, Kirstin Edwards, Jonathan Hubb, Joanna Ellis, Maria Zambon
EPI_ISL_420733  hCoV-19/England/20139060504/2020        Europe / United Kingdom / England       2020-03-28
Respiratory Virus Unit, Microbiology Services Colindale, Public Health England  Respiratory Virus Unit, Microbiology Services Colindale, Public Health England  Monica Galiano, Shahjahan Miah, Angie Lackenby, Omolola Akinbami, Tiina Talts, Leena Bhaw, Richard Myers, Steven Platt, Kirstin Edwards, Jonathan Hubb, Joanna Ellis, Maria Zambon
EPI_ISL_420734  hCoV-19/England/20139060804/2020        Europe / United Kingdom / England       2020-03-27
Respiratory Virus Unit, Microbiology Services Colindale, Public Health England  Respiratory Virus Unit, Microbiology Services Colindale, Public Health England  Monica Galiano, Shahjahan Miah, Angie Lackenby, Omolola Akinbami, Tiina Talts, Leena Bhaw, Richard Myers, Steven Platt, Kirstin Edwards, Jonathan Hubb, Joanna Ellis, Maria Zambon
EPI_ISL_420735  hCoV-19/England/20139061104/2020        Europe / United Kingdom / England       2020-03-27
Respiratory Virus Unit, Microbiology Services Colindale, Public Health England  Respiratory Virus Unit, Microbiology Services Colindale, Public Health England  Monica Galiano, Shahjahan Miah, Angie Lackenby, Omolola Akinbami, Tiina Talts, Leena Bhaw, Richard Myers, Steven Platt, Kirstin Edwards, Jonathan Hubb, Joanna Ellis, Maria Zambon
EPI_ISL_420741  hCoV-19/England/20139062504/2020        Europe / United Kingdom / England       2020-03-27
Respiratory Virus Unit, Microbiology Services Colindale, Public Health England  Respiratory Virus Unit, Microbiology Services Colindale, Public Health England  Monica Galiano, Shahjahan Miah, Angie Lackenby, Omolola Akinbami, Tiina Talts, Leena Bhaw, Richard Myers, Steven Platt, Kirstin Edwards, Jonathan Hubb, Joanna Ellis,

Maria Zambon
EPI_ISL_420742 hCoV-19/England/20139062704/2020 Europe / United Kingdom / England 2020-03-28 Respiratory Virus Unit, Microbiology Services Colindale, Public Health England Respiratory Virus Unit, Microbiology Services Colindale, Public Health England Monica Galiano, Shahjahan Miah, Angie Lackenby, Omolola Akinbami, Tiina Talts, Leena Bhaw, Richard Myers, Steven Platt, Kirstin Edwards, Jonathan Hubb, Joanna Ellis, Maria Zambon
EPI_ISL_420743 hCoV-19/England/20139062804/2020 Europe / United Kingdom / England 2020-03-27 Respiratory Virus Unit, Microbiology Services Colindale, Public Health England Respiratory Virus Unit, Microbiology Services Colindale, Public Health England Monica Galiano, Shahjahan Miah, Angie Lackenby, Omolola Akinbami, Tiina Talts, Leena Bhaw, Richard Myers, Steven Platt, Kirstin Edwards, Jonathan Hubb, Joanna Ellis, Maria Zambon
EPI_ISL_420744 hCoV-19/England/20139063204/2020 Europe / United Kingdom / England 2020-03-28 Respiratory Virus Unit, Microbiology Services Colindale, Public Health England Respiratory Virus Unit, Microbiology Services Colindale, Public Health England Monica Galiano, Shahjahan Miah, Angie Lackenby, Omolola Akinbami, Tiina Talts, Leena Bhaw, Richard Myers, Steven Platt, Kirstin Edwards, Jonathan Hubb, Joanna Ellis, Maria Zambon
EPI_ISL_420745 hCoV-19/England/20139063504/2020 Europe / United Kingdom / England 2020-03-28 Respiratory Virus Unit, Microbiology Services Colindale, Public Health England Respiratory Virus Unit, Microbiology Services Colindale, Public Health England Monica Galiano, Shahjahan Miah, Angie Lackenby, Omolola Akinbami, Tiina Talts, Leena Bhaw, Richard Myers, Steven Platt, Kirstin Edwards, Jonathan Hubb, Joanna Ellis, Maria Zambon
EPI_ISL_420746 hCoV-19/England/20139063604/2020 Europe / United Kingdom / England 2020-03-25 Respiratory Virus Unit, Microbiology Services Colindale, Public Health England Respiratory Virus Unit, Microbiology Services Colindale, Public Health England Monica Galiano, Shahjahan Miah, Angie Lackenby, Omolola Akinbami, Tiina Talts, Leena Bhaw, Richard Myers, Steven Platt, Kirstin Edwards, Jonathan Hubb, Joanna Ellis, Maria Zambon
EPI_ISL_420747 hCoV-19/England/20139063704/2020 Europe / United Kingdom / England 2020-03-28 Respiratory Virus Unit, Microbiology Services Colindale, Public Health England Respiratory Virus Unit, Microbiology Services Colindale, Public Health England Monica Galiano, Shahjahan Miah, Angie Lackenby, Omolola Akinbami, Tiina Talts, Leena Bhaw, Richard Myers, Steven Platt, Kirstin Edwards, Jonathan Hubb, Joanna Ellis, Maria Zambon
EPI_ISL_420748 hCoV-19/England/20139063804/2020 Europe / United Kingdom / England 2020-03-28 Respiratory Virus Unit, Microbiology Services Colindale, Public Health England Respiratory Virus Unit, Microbiology Services Colindale, Public Health England Monica Galiano, Shahjahan Miah, Angie Lackenby, Omolola Akinbami, Tiina Talts, Leena Bhaw, Richard Myers, Steven Platt, Kirstin Edwards, Jonathan Hubb, Joanna Ellis, Maria Zambon
EPI_ISL_420749 hCoV-19/England/20140037504/2020 Europe / United Kingdom / England 2020-03-25 Respiratory Virus Unit, Microbiology Services Colindale, Public Health England Respiratory Virus Unit, Microbiology Services Colindale, Public Health England Monica Galiano, Shahjahan Miah, Angie Lackenby, Omolola Akinbami, Tiina Talts, Leena Bhaw, Richard Myers, Steven Platt, Kirstin Edwards, Jonathan Hubb, Joanna Ellis, Maria Zambon
EPI_ISL_420750 hCoV-19/England/20140038604/2020 Europe / United Kingdom / England 2020-03-23 Respiratory Virus Unit, Microbiology Services Colindale, Public Health England Respiratory Virus Unit, Microbiology Services Colindale, Public Health England Monica Galiano, Shahjahan Miah, Angie Lackenby, Omolola Akinbami, Tiina Talts, Leena Bhaw, Richard Myers, Steven Platt, Kirstin Edwards, Jonathan Hubb, Joanna Ellis, Maria Zambon
EPI_ISL_420751 hCoV-19/England/20140038704/2020 Europe / United Kingdom / England 2020-03-30 Respiratory Virus Unit, Microbiology Services Colindale, Public Health England Respiratory Virus Unit, Microbiology Services Colindale, Public Health England Monica Galiano, Shahjahan Miah, Angie Lackenby, Omolola Akinbami, Tiina Talts, Leena Bhaw, Richard Myers, Steven Platt, Kirstin Edwards, Jonathan Hubb, Joanna Ellis, Maria Zambon
EPI_ISL_420752 hCoV-19/England/20140039004/2020 Europe / United Kingdom / England 2020-03-26 Respiratory Virus Unit, Microbiology Services Colindale, Public Health England Respiratory Virus Unit, Microbiology Services Colindale, Public Health England Monica Galiano, Shahjahan Miah, Angie Lackenby, Omolola Akinbami, Tiina Talts, Leena Bhaw, Richard Myers, Steven Platt, Kirstin Edwards, Jonathan Hubb, Joanna Ellis, Maria Zambon
EPI_ISL_420753 hCoV-19/England/20140039104/2020 Europe / United Kingdom / England 2020-03-25 Respiratory Virus Unit, Microbiology Services Colindale, Public Health England Respiratory Virus Unit, Microbiology Services Colindale, Public Health England Monica Galiano, Shahjahan Miah, Angie Lackenby, Omolola Akinbami, Tiina Talts, Leena Bhaw, Richard Myers, Steven Platt, Kirstin Edwards, Jonathan Hubb, Joanna Ellis, Maria Zambon
EPI_ISL_420755 hCoV-19/England/20140039304/2020 Europe / United Kingdom / England 2020-03-27 Respiratory Virus Unit, Microbiology Services Colindale, Public Health England Respiratory Virus Unit, Microbiology Services Colindale, Public Health England Monica Galiano, Shahjahan Miah, Angie Lackenby, Omolola Akinbami, Tiina Talts, Leena Bhaw, Richard Myers, Steven Platt, Kirstin Edwards, Jonathan Hubb, Joanna Ellis, Maria Zambon
EPI_ISL_420756 hCoV-19/England/20140039404/2020 Europe / United Kingdom / England 2020-03-27 Respiratory Virus Unit, Microbiology Services Colindale, Public Health England Respiratory Virus Unit, Microbiology Services Colindale, Public Health England Monica Galiano, Shahjahan Miah, Angie Lackenby, Omolola Akinbami, Tiina Talts, Leena Bhaw, Richard Myers, Steven Platt, Kirstin Edwards, Jonathan Hubb, Joanna Ellis,

Maria Zambon
EPI_ISL_420758	hCoV-19/England/20140039604/2020	Europe / United Kingdom / England	2020-03-27	Respiratory Virus Unit, Microbiology Services Colindale, Public Health England	Respiratory Virus Unit, Microbiology Services Colindale, Public Health England	Monica Galiano, Shahjahan Miah, Angie Lackenby, Omolola Akinbami, Tiina Talts, Leena Bhaw, Richard Myers, Steven Platt, Kirstin Edwards, Jonathan Hubb, Joanna Ellis, Maria Zambon
EPI_ISL_420761	hCoV-19/England/20140048704/2020	Europe / United Kingdom / England	2020-03-25	Respiratory Virus Unit, Microbiology Services Colindale, Public Health England	Respiratory Virus Unit, Microbiology Services Colindale, Public Health England	Monica Galiano, Shahjahan Miah, Angie Lackenby, Omolola Akinbami, Tiina Talts, Leena Bhaw, Richard Myers, Steven Platt, Kirstin Edwards, Jonathan Hubb, Joanna Ellis, Maria Zambon
EPI_ISL_420765	hCoV-19/England/20142022504/2020	Europe / United Kingdom / England	2020-03-27	Respiratory Virus Unit, Microbiology Services Colindale, Public Health England	Respiratory Virus Unit, Microbiology Services Colindale, Public Health England	Monica Galiano, Shahjahan Miah, Angie Lackenby, Omolola Akinbami, Tiina Talts, Leena Bhaw, Richard Myers, Steven Platt, Kirstin Edwards, Jonathan Hubb, Joanna Ellis, Maria Zambon
EPI_ISL_420766	hCoV-19/England/20142060404/2020	Europe / United Kingdom / England	2020-03-27	Respiratory Virus Unit, Microbiology Services Colindale, Public Health England	Respiratory Virus Unit, Microbiology Services Colindale, Public Health England	Monica Galiano, Shahjahan Miah, Angie Lackenby, Omolola Akinbami, Tiina Talts, Leena Bhaw, Richard Myers, Steven Platt, Kirstin Edwards, Jonathan Hubb, Joanna Ellis, Maria Zambon
EPI_ISL_420767	hCoV-19/England/20142060804/2020	Europe / United Kingdom / England	2020-03-30	Respiratory Virus Unit, Microbiology Services Colindale, Public Health England	Respiratory Virus Unit, Microbiology Services Colindale, Public Health England	Monica Galiano, Shahjahan Miah, Angie Lackenby, Omolola Akinbami, Tiina Talts, Leena Bhaw, Richard Myers, Steven Platt, Kirstin Edwards, Jonathan Hubb, Joanna Ellis, Maria Zambon
EPI_ISL_420772	hCoV-19/England/20142061804/2020	Europe / United Kingdom / England	2020-03-30	Respiratory Virus Unit, Microbiology Services Colindale, Public Health England	Respiratory Virus Unit, Microbiology Services Colindale, Public Health England	Monica Galiano, Shahjahan Miah, Angie Lackenby, Omolola Akinbami, Tiina Talts, Leena Bhaw, Richard Myers, Steven Platt, Kirstin Edwards, Jonathan Hubb, Joanna Ellis, Maria Zambon
EPI_ISL_420773	hCoV-19/England/20142062204/2020	Europe / United Kingdom / England	2020-03-30	Respiratory Virus Unit, Microbiology Services Colindale, Public Health England	Respiratory Virus Unit, Microbiology Services Colindale, Public Health England	Monica Galiano, Shahjahan Miah, Angie Lackenby, Omolola Akinbami, Tiina Talts, Leena Bhaw, Richard Myers, Steven Platt, Kirstin Edwards, Jonathan Hubb, Joanna Ellis, Maria Zambon
EPI_ISL_420774	hCoV-19/England/20142062604/2020	Europe / United Kingdom / England	2020-03-28	Respiratory Virus Unit, Microbiology Services Colindale, Public Health England	Respiratory Virus Unit, Microbiology Services Colindale, Public Health England	Monica Galiano, Shahjahan Miah, Angie Lackenby, Omolola Akinbami, Tiina Talts, Leena Bhaw, Richard Myers, Steven Platt, Kirstin Edwards, Jonathan Hubb, Joanna Ellis, Maria Zambon
EPI_ISL_420775	hCoV-19/England/20142062904/2020	Europe / United Kingdom / England	2020-03-30	Respiratory Virus Unit, Microbiology Services Colindale, Public Health England	Respiratory Virus Unit, Microbiology Services Colindale, Public Health England	Monica Galiano, Shahjahan Miah, Angie Lackenby, Omolola Akinbami, Tiina Talts, Leena Bhaw, Richard Myers, Steven Platt, Kirstin Edwards, Jonathan Hubb, Joanna Ellis, Maria Zambon
EPI_ISL_420784	hCoV-19/USA/AZ_4811/2020	North America / USA / Arizona	2020-03-02	AZ Department of Health Services	Pathogen Discovery, Respiratory Viruses Branch, Division of Viral Diseases, Centers for Disease Control and Prevention	Krista Queen, Yan Li, Ying Tao, Jing Zhang, Anne Uehara, Clinton R. Paden, Haibin Wang, Rachel Marine, Mary S. Keckler, Alison S. Laufer Halpin, Jasmine Padilla, Justin Lee, Christopher A. Elkins, Suxiang Tong
EPI_ISL_420785	hCoV-19/USA/FL_6318/2020	North America / USA / Florida	2020-03-02	FL Bureau of Health Laboratories Tampa	Pathogen Discovery, Respiratory Viruses Branch, Division of Viral Diseases, Centers for Disease Control and Prevention	Krista Queen, Yan Li, Ying Tao, Jing Zhang, Anne Uehara, Clinton R. Paden, Haibin Wang, Rachel Marine, Mary S. Keckler, Alison S. Laufer Halpin, Jasmine Padilla, Justin Lee, Christopher A. Elkins, Suxiang Tong
EPI_ISL_420786	hCoV-19/USA/GA_1320/2020	North America / USA / Georgia	2020-03-03	GA Department of Public Health	Pathogen Discovery, Respiratory Viruses Branch, Division of Viral Diseases, Centers for Disease Control and Prevention	Krista Queen, Yan Li, Ying Tao, Jing Zhang, Anne Uehara, Clinton R. Paden, Haibin Wang, Rachel Marine, Mary S. Keckler, Alison S. Laufer Halpin, Jasmine Padilla, Justin Lee, Christopher A. Elkins, Suxiang Tong
EPI_ISL_420787	hCoV-19/USA/GA_1299/2020	North America / USA / Georgia	2020-03-03	GA Department of Public Health	Pathogen Discovery, Respiratory Viruses Branch, Division of Viral Diseases, Centers for Disease Control and Prevention	Krista Queen, Yan Li, Ying Tao, Jing Zhang, Anne Uehara, Clinton R. Paden, Haibin Wang, Rachel Marine, Mary S. Keckler, Alison S. Laufer Halpin, Jasmine Padilla, Justin Lee, Christopher A. Elkins, Suxiang Tong
EPI_ISL_420788	hCoV-19/USA/GA_1445/2020	North America / USA / Georgia	2020-03-04	GA Department of Public Health	Pathogen Discovery, Respiratory Viruses Branch, Division of Viral Diseases, Centers for Disease Control and Prevention	Krista Queen, Yan Li, Ying Tao, Jing Zhang, Anne Uehara, Clinton R. Paden, Haibin Wang, Rachel Marine, Mary S. Keckler, Alison S. Laufer Halpin, Jasmine Padilla, Justin Lee, Christopher

A. Elkins, Suxiang Tong
EPI_ISL_420789 hCoV-19/USA/IL_1375/2020 North America / USA / Illinois 2020-03-01 Illinois Department of Public Health Chicago Laboratory Pathogen Discovery, Respiratory Viruses Branch, Division of Viral Diseases, Centers for Disease Control and Prevention Krista Queen, Yan Li, Ying Tao, Jing Zhang, Anne Uehara, Clinton R. Paden, Haibin Wang, Rachel Marine, Mary S. Keckler, Alison S. Laufer Halpin, Jasmine Padilla, Justin Lee, Christopher A. Elkins, Suxiang Tong
EPI_ISL_420790 hCoV-19/USA/IL_1293/2020 North America / USA / Illinois 2020-03-01 Illinois Department of Public Health Chicago Laboratory Pathogen Discovery, Respiratory Viruses Branch, Division of Viral Diseases, Centers for Disease Control and Prevention Krista Queen, Yan Li, Ying Tao, Jing Zhang, Anne Uehara, Clinton R. Paden, Haibin Wang, Rachel Marine, Mary S. Keckler, Alison S. Laufer Halpin, Jasmine Padilla, Justin Lee, Christopher A. Elkins, Suxiang Tong
EPI_ISL_420791 hCoV-19/USA/NH_0004/2020 North America / USA / New Hampshire 2020-02-29 NH Department of Health and Human Services Public Health Labs Pathogen Discovery, Respiratory Viruses Branch, Division of Viral Diseases, Centers for Disease Control and Prevention Krista Queen, Yan Li, Ying Tao, Jing Zhang, Anne Uehara, Clinton R. Paden, Haibin Wang, Rachel Marine, Mary S. Keckler, Alison S. Laufer Halpin, Jasmine Padilla, Justin Lee, Christopher A. Elkins, Suxiang Tong
EPI_ISL_420792 hCoV-19/USA/NH_0008/2020 North America / USA / New Hampshire 2020-03-02 NH Department of Health and Human Services Public Health Labs Pathogen Discovery, Respiratory Viruses Branch, Division of Viral Diseases, Centers for Disease Control and Prevention Krista Queen, Yan Li, Ying Tao, Jing Zhang, Anne Uehara, Clinton R. Paden, Haibin Wang, Rachel Marine, Mary S. Keckler, Alison S. Laufer Halpin, Jasmine Padilla, Justin Lee, Christopher A. Elkins, Suxiang Tong
EPI_ISL_420793 hCoV-19/USA/NY_2929/2020 North America / USA / New York 2020-03-02 NYC Department of Health and Mental Hygiene Pathogen Discovery, Respiratory Viruses Branch, Division of Viral Diseases, Centers for Disease Control and Prevention Krista Queen, Yan Li, Ying Tao, Jing Zhang, Anne Uehara, Clinton R. Paden, Haibin Wang, Rachel Marine, Mary S. Keckler, Alison S. Laufer Halpin, Jasmine Padilla, Justin Lee, Christopher A. Elkins, Suxiang Tong
EPI_ISL_420794 hCoV-19/USA/OR_5430/2020 North America / USA / Oregon 2020-03-01 Oregon State Public Health- Virology section Pathogen Discovery, Respiratory Viruses Branch, Division of Viral Diseases, Centers for Disease Control and Prevention Krista Queen, Yan Li, Ying Tao, Jing Zhang, Anne Uehara, Clinton R. Paden, Haibin Wang, Rachel Marine, Mary S. Keckler, Alison S. Laufer Halpin, Jasmine Padilla, Justin Lee, Christopher A. Elkins, Suxiang Tong
EPI_ISL_420795 hCoV-19/USA/RI_0556/2020 North America / USA / Rhode Island 2020-03-01 RI State Health Laboratory Pathogen Discovery, Respiratory Viruses Branch, Division of Viral Diseases, Centers for Disease Control and Prevention Krista Queen, Yan Li, Ying Tao, Jing Zhang, Anne Uehara, Clinton R. Paden, Haibin Wang, Rachel Marine, Mary S. Keckler, Alison S. Laufer Halpin, Jasmine Padilla, Justin Lee, Christopher A. Elkins, Suxiang Tong
EPI_ISL_420796 hCoV-19/USA/TX_2039/2020 North America / USA / Texas 2020-02-29 Texas DSHS Lab Services Pathogen Discovery, Respiratory Viruses Branch, Division of Viral Diseases, Centers for Disease Control and Prevention Krista Queen, Yan Li, Ying Tao, Jing Zhang, Anne Uehara, Clinton R. Paden, Haibin Wang, Rachel Marine, Mary S. Keckler, Alison S. Laufer Halpin, Jasmine Padilla, Justin Lee, Christopher A. Elkins, Suxiang Tong
EPI_ISL_420797 hCoV-19/USA/TX_2817/2020 North America / USA / Texas 2020-03-01 Texas DSHS Lab Services Pathogen Discovery, Respiratory Viruses Branch, Division of Viral Diseases, Centers for Disease Control and Prevention Krista Queen, Yan Li, Ying Tao, Jing Zhang, Anne Uehara, Clinton R. Paden, Haibin Wang, Rachel Marine, Mary S. Keckler, Alison S. Laufer Halpin, Jasmine Padilla, Justin Lee, Christopher A. Elkins, Suxiang Tong
EPI_ISL_420798 hCoV-19/USA/TX_2967/2020 North America / USA / Texas 2020-03-01 Texas DSHS Lab Services Pathogen Discovery, Respiratory Viruses Branch, Division of Viral Diseases, Centers for Disease Control and Prevention Krista Queen, Yan Li, Ying Tao, Jing Zhang, Anne Uehara, Clinton R. Paden, Haibin Wang, Rachel Marine, Mary S. Keckler, Alison S. Laufer Halpin, Jasmine Padilla, Justin Lee, Christopher A. Elkins, Suxiang Tong
EPI_ISL_420799 hCoV-19/Korea/BA-ACH_2604/2020 Asia / Korea 2020-02-27 Brian D. Allgood Army Community Hospital Pathogen Discovery, Respiratory Viruses Branch, Division of Viral Diseases, Centers for Disease Control and Prevention Krista Queen, Yan Li, Ying Tao, Jing Zhang, Anne Uehara, Clinton R. Paden, Haibin Wang, Rachel Marine, Mary S. Keckler, Alison S. Laufer Halpin, Jasmine Padilla, Justin Lee, Christopher A. Elkins, Suxiang Tong
EPI_ISL_420822 hCoV-19/USA/UT-0349/2020 North America / USA / Utah 2020-03-25 Utah Public Health Laboratory Utah Public Health Laboratory Erin Young, Kelly Oakeson
EPI_ISL_420899 hCoV-19/Germany/BAV-V2010492/2020 Europe / Germany / Munich 2020-03-11 Max von Pettenkofer Institute, Virology, National Reference Center for Retroviruses, LMU Munich Laboratory for Functional Genome Analysis, Dept. Genomics, Gene Center of the LMU Munich Max Muenchhoff, Stefan Krebs, Alexander Graf, Ashok Varadharajan, Oliver Keppler, Helmut Blum
EPI_ISL_423091 hCoV-19/England/20134016802/2020 Europe / United Kingdom / England 2020-03-24 Respiratory Virus Unit, Microbiology Services Colindale, Public Health England Respiratory Virus Unit, Microbiology Services Colindale, Public Health England Monica Galiano, Shahjahan Miah, Angie Lackenby, Omolola Akinbami, Tiina Talts, Leena Bhaw, Richard Myers, Steven Platt, Kirstin Edwards, Jonathan Hubb, Joanna Ellis, Maria Zambon
EPI_ISL_423092 hCoV-19/England/20134016902/2020 Europe / United Kingdom / England 2020-03-24 Respiratory Virus Unit, Microbiology Services Colindale, Public Health England Respiratory Virus Unit, Microbiology Services Colindale, Public Health England Monica Galiano, Shahjahan Miah, Angie Lackenby, Omolola

Akinbami, Tiina Talts, Leena Bhaw, Richard Myers, Steven Platt, Kirstin Edwards, Jonathan Hubb, Joanna Ellis, Maria Zambon
EPI_ISL_423094 hCoV-19/England/20134017404/2020 Europe / United Kingdom / England 2020-03-24 Respiratory Virus Unit, Microbiology Services Colindale, Public Health England Respiratory Virus Unit, Microbiology Services Colindale, Public Health England Monica Galiano, Shahjahan Miah, Angie Lackenby, Omolola Akinbami, Tiina Talts, Leena Bhaw, Richard Myers, Steven Platt, Kirstin Edwards, Jonathan Hubb, Joanna Ellis, Maria Zambon
EPI_ISL_423095 hCoV-19/England/20134017504/2020 Europe / United Kingdom / England 2020-03-24 Respiratory Virus Unit, Microbiology Services Colindale, Public Health England Respiratory Virus Unit, Microbiology Services Colindale, Public Health England Monica Galiano, Shahjahan Miah, Angie Lackenby, Omolola Akinbami, Tiina Talts, Leena Bhaw, Richard Myers, Steven Platt, Kirstin Edwards, Jonathan Hubb, Joanna Ellis, Maria Zambon
EPI_ISL_423096 hCoV-19/England/20134017904/2020 Europe / United Kingdom / England 2020-03-23 Respiratory Virus Unit, Microbiology Services Colindale, Public Health England Respiratory Virus Unit, Microbiology Services Colindale, Public Health England Monica Galiano, Shahjahan Miah, Angie Lackenby, Omolola Akinbami, Tiina Talts, Leena Bhaw, Richard Myers, Steven Platt, Kirstin Edwards, Jonathan Hubb, Joanna Ellis, Maria Zambon
EPI_ISL_423097 hCoV-19/England/20134018004/2020 Europe / United Kingdom / England 2020-03-23 Respiratory Virus Unit, Microbiology Services Colindale, Public Health England Respiratory Virus Unit, Microbiology Services Colindale, Public Health England Monica Galiano, Shahjahan Miah, Angie Lackenby, Omolola Akinbami, Tiina Talts, Leena Bhaw, Richard Myers, Steven Platt, Kirstin Edwards, Jonathan Hubb, Joanna Ellis, Maria Zambon
EPI_ISL_423098 hCoV-19/England/20134018402/2020 Europe / United Kingdom / England 2020-03-23 Respiratory Virus Unit, Microbiology Services Colindale, Public Health England Respiratory Virus Unit, Microbiology Services Colindale, Public Health England Monica Galiano, Shahjahan Miah, Angie Lackenby, Omolola Akinbami, Tiina Talts, Leena Bhaw, Richard Myers, Steven Platt, Kirstin Edwards, Jonathan Hubb, Joanna Ellis, Maria Zambon
EPI_ISL_423099 hCoV-19/England/20134018602/2020 Europe / United Kingdom / England 2020-03-23 Respiratory Virus Unit, Microbiology Services Colindale, Public Health England Respiratory Virus Unit, Microbiology Services Colindale, Public Health England Monica Galiano, Shahjahan Miah, Angie Lackenby, Omolola Akinbami, Tiina Talts, Leena Bhaw, Richard Myers, Steven Platt, Kirstin Edwards, Jonathan Hubb, Joanna Ellis, Maria Zambon
EPI_ISL_424200 hCoV-19/USA/WA-UW-1723/2020 North America / USA / Washington 2020-03-20 UW Virology Lab UW Virology Lab Pavitra Roychoudhury, Hong Xie, Keith Jerome, Alexander Greninger
EPI_ISL_424202 hCoV-19/USA/WA-UW-1651/2020 North America / USA / Washington 2020-03-20 UW Virology Lab UW Virology Lab Pavitra Roychoudhury, Hong Xie, Keith Jerome, Alexander Greninger
EPI_ISL_424203 hCoV-19/USA/WA-UW-1652/2020 North America / USA / Washington 2020-03-20 UW Virology Lab UW Virology Lab Pavitra Roychoudhury, Hong Xie, Keith Jerome, Alexander Greninger
EPI_ISL_424204 hCoV-19/USA/WA-UW-1698/2020 North America / USA / Washington 2020-03-20 UW Virology Lab UW Virology Lab Pavitra Roychoudhury, Hong Xie, Keith Jerome, Alexander Greninger
EPI_ISL_424205 hCoV-19/USA/WA-UW-1697/2020 North America / USA / Washington 2020-03-20 UW Virology Lab UW Virology Lab Pavitra Roychoudhury, Hong Xie, Keith Jerome, Alexander Greninger
EPI_ISL_424207 hCoV-19/USA/WA-UW-1716/2020 North America / USA / Washington 2020-03-20 UW Virology Lab UW Virology Lab Pavitra Roychoudhury, Hong Xie, Keith Jerome, Alexander Greninger
EPI_ISL_424208 hCoV-19/USA/WA-UW-1712/2020 North America / USA / Washington 2020-03-20 UW Virology Lab UW Virology Lab Pavitra Roychoudhury, Hong Xie, Keith Jerome, Alexander Greninger
EPI_ISL_424209 hCoV-19/USA/WA-UW-1765/2020 North America / USA / Washington 2020-03-20 UW Virology Lab UW Virology Lab Pavitra Roychoudhury, Hong Xie, Keith Jerome, Alexander Greninger
EPI_ISL_424210 hCoV-19/USA/WA-UW-1718/2020 North America / USA / Washington 2020-03-20 UW Virology Lab UW Virology Lab Pavitra Roychoudhury, Hong Xie, Keith Jerome, Alexander Greninger
EPI_ISL_424211 hCoV-19/USA/WA-UW-1689/2020 North America / USA / Washington 2020-03-19 UW Virology Lab UW Virology Lab Pavitra Roychoudhury, Hong Xie, Keith Jerome, Alexander Greninger
EPI_ISL_424212 hCoV-19/USA/WA-UW-1715/2020 North America / USA / Washington 2020-03-19 UW Virology Lab UW Virology Lab Pavitra Roychoudhury, Hong Xie, Keith Jerome, Alexander Greninger
EPI_ISL_424213 hCoV-19/USA/WA-UW-1670/2020 North America / USA / Washington 2020-03-20 UW Virology Lab UW Virology Lab Pavitra Roychoudhury, Hong Xie, Keith Jerome, Alexander Greninger
EPI_ISL_424214 hCoV-19/USA/WA-UW-1677/2020 North America / USA / Washington 2020-03-20 UW Virology Lab UW Virology Lab Pavitra Roychoudhury, Hong Xie, Keith Jerome, Alexander Greninger
EPI_ISL_424215 hCoV-19/USA/WA-UW-1680/2020 North America / USA / Washington 2020-03-20 UW Virology Lab UW Virology Lab Pavitra Roychoudhury, Hong Xie, Keith Jerome, Alexander Greninger
EPI_ISL_424216 hCoV-19/USA/WA-UW-1690/2020 North America / USA / Washington 2020-03-19 UW Virology Lab UW Virology Lab Pavitra Roychoudhury, Hong Xie, Keith Jerome, Alexander Greninger
EPI_ISL_424218 hCoV-19/USA/WA-UW-1682/2020 North America / USA / Washington 2020-03-20 UW Virology Lab UW Virology Lab Pavitra Roychoudhury, Hong Xie, Keith Jerome, Alexander Greninger
EPI_ISL_424219 hCoV-19/USA/WA-UW-1695/2020 North America / USA / Washington 2020-03-20 UW Virology Lab UW Virology Lab Pavitra Roychoudhury, Hong Xie, Keith Jerome, Alexander Greninger
EPI_ISL_424220 hCoV-19/USA/WA-UW-1729/2020 North America / USA / Washington 2020-03-21 UW Virology Lab UW Virology Lab Pavitra Roychoudhury, Hong Xie, Keith Jerome, Alexander Greninger
EPI_ISL_424221 hCoV-19/USA/WA-UW-1748/2020 North America / USA / Washington 2020-03-21 UW Virology Lab UW Virology Lab Pavitra Roychoudhury, Hong Xie, Keith Jerome, Alexander Greninger

EPI_ISL_424222 hCoV-19/USA/WA-UW-1700/2020 North America / USA / Washington 2020-03-20 UW Virology Lab UW Virology Lab Pavitra Roychoudhury, Hong Xie, Keith Jerome, Alexander Greninger
EPI_ISL_424223 hCoV-19/USA/WA-UW-1744/2020 North America / USA / Washington 2020-03-21 UW Virology Lab UW Virology Lab Pavitra Roychoudhury, Hong Xie, Keith Jerome, Alexander Greninger
EPI_ISL_424224 hCoV-19/USA/WA-UW-1745/2020 North America / USA / Washington 2020-03-21 UW Virology Lab UW Virology Lab Pavitra Roychoudhury, Hong Xie, Keith Jerome, Alexander Greninger
EPI_ISL_424225 hCoV-19/USA/WA-UW-1707/2020 North America / USA / Washington 2020-03-21 UW Virology Lab UW Virology Lab Pavitra Roychoudhury, Hong Xie, Keith Jerome, Alexander Greninger
EPI_ISL_424226 hCoV-19/USA/WA-UW-1705/2020 North America / USA / Washington 2020-03-21 UW Virology Lab UW Virology Lab Pavitra Roychoudhury, Hong Xie, Keith Jerome, Alexander Greninger
EPI_ISL_424227 hCoV-19/USA/WA-UW-1706/2020 North America / USA / Washington 2020-03-21 UW Virology Lab UW Virology Lab Pavitra Roychoudhury, Hong Xie, Keith Jerome, Alexander Greninger
EPI_ISL_424228 hCoV-19/USA/WA-UW-1702/2020 North America / USA / Washington 2020-03-21 UW Virology Lab UW Virology Lab Pavitra Roychoudhury, Hong Xie, Keith Jerome, Alexander Greninger
EPI_ISL_424229 hCoV-19/USA/WA-UW-1724/2020 North America / USA / Washington 2020-03-21 UW Virology Lab UW Virology Lab Pavitra Roychoudhury, Hong Xie, Keith Jerome, Alexander Greninger
EPI_ISL_424230 hCoV-19/USA/WA-UW-1709/2020 North America / USA / Washington 2020-03-21 UW Virology Lab UW Virology Lab Pavitra Roychoudhury, Hong Xie, Keith Jerome, Alexander Greninger
EPI_ISL_424231 hCoV-19/USA/WA-UW-1731/2020 North America / USA / Washington 2020-03-21 UW Virology Lab UW Virology Lab Pavitra Roychoudhury, Hong Xie, Keith Jerome, Alexander Greninger
EPI_ISL_424232 hCoV-19/USA/WA-UW-1708/2020 North America / USA / Washington 2020-03-21 UW Virology Lab UW Virology Lab Pavitra Roychoudhury, Hong Xie, Keith Jerome, Alexander Greninger
EPI_ISL_424233 hCoV-19/USA/WA-UW-1772/2020 North America / USA / Washington 2020-03-21 UW Virology Lab UW Virology Lab Pavitra Roychoudhury, Hong Xie, Keith Jerome, Alexander Greninger
EPI_ISL_424234 hCoV-19/USA/WA-UW-1722/2020 North America / USA / Washington 2020-03-21 UW Virology Lab UW Virology Lab Pavitra Roychoudhury, Hong Xie, Keith Jerome, Alexander Greninger
EPI_ISL_424235 hCoV-19/USA/WA-UW-1733/2020 North America / USA / Washington 2020-03-21 UW Virology Lab UW Virology Lab Pavitra Roychoudhury, Hong Xie, Keith Jerome, Alexander Greninger
EPI_ISL_424236 hCoV-19/USA/WA-UW-1732/2020 North America / USA / Washington 2020-03-21 UW Virology Lab UW Virology Lab Pavitra Roychoudhury, Hong Xie, Keith Jerome, Alexander Greninger
EPI_ISL_424237 hCoV-19/USA/WA-UW-1739/2020 North America / USA / Washington 2020-03-21 UW Virology Lab UW Virology Lab Pavitra Roychoudhury, Hong Xie, Keith Jerome, Alexander Greninger
EPI_ISL_424238 hCoV-19/USA/WA-UW-1741/2020 North America / USA / Washington 2020-03-21 UW Virology Lab UW Virology Lab Pavitra Roychoudhury, Hong Xie, Keith Jerome, Alexander Greninger
EPI_ISL_424239 hCoV-19/USA/WA-UW-1750/2020 North America / USA / Washington 2020-03-21 UW Virology Lab UW Virology Lab Pavitra Roychoudhury, Hong Xie, Keith Jerome, Alexander Greninger
EPI_ISL_424240 hCoV-19/USA/WA-UW-1730/2020 North America / USA / Washington 2020-03-21 UW Virology Lab UW Virology Lab Pavitra Roychoudhury, Hong Xie, Keith Jerome, Alexander Greninger
EPI_ISL_424241 hCoV-19/USA/WA-UW-1735/2020 North America / USA / Washington 2020-03-21 UW Virology Lab UW Virology Lab Pavitra Roychoudhury, Hong Xie, Keith Jerome, Alexander Greninger
EPI_ISL_424242 hCoV-19/USA/WA-UW-1753/2020 North America / USA / Washington 2020-03-21 UW Virology Lab UW Virology Lab Pavitra Roychoudhury, Hong Xie, Keith Jerome, Alexander Greninger
EPI_ISL_424243 hCoV-19/USA/WA-UW-1756/2020 North America / USA / Washington 2020-03-21 UW Virology Lab UW Virology Lab Pavitra Roychoudhury, Hong Xie, Keith Jerome, Alexander Greninger
EPI_ISL_424244 hCoV-19/USA/WA-UW-1770/2020 North America / USA / Washington 2020-03-22 UW Virology Lab UW Virology Lab Pavitra Roychoudhury, Hong Xie, Keith Jerome, Alexander Greninger
EPI_ISL_424245 hCoV-19/USA/WA-UW-1762/2020 North America / USA / Washington 2020-03-21 UW Virology Lab UW Virology Lab Pavitra Roychoudhury, Hong Xie, Keith Jerome, Alexander Greninger
EPI_ISL_424246 hCoV-19/USA/WA-UW-1758/2020 North America / USA / Washington 2020-03-21 UW Virology Lab UW Virology Lab Pavitra Roychoudhury, Hong Xie, Keith Jerome, Alexander Greninger
EPI_ISL_424247 hCoV-19/USA/WA-UW-1760/2020 North America / USA / Washington 2020-03-21 UW Virology Lab UW Virology Lab Pavitra Roychoudhury, Hong Xie, Keith Jerome, Alexander Greninger
EPI_ISL_424248 hCoV-19/USA/UN-UW-1773/2020 North America / USA 2020-03 UW Virology Lab UW Virology Lab Pavitra Roychoudhury, Hong Xie, Keith Jerome, Alexander Greninger
EPI_ISL_424249 hCoV-19/USA/UN-UW-1791/2020 North America / USA 2020-03-20 UW Virology Lab UW Virology Lab Pavitra Roychoudhury, Hong Xie, Keith Jerome, Alexander Greninger
EPI_ISL_424250 hCoV-19/USA/WA-UW-1785/2020 North America / USA / Washington 2020-03-19 UW Virology Lab UW Virology Lab Pavitra Roychoudhury, Hong Xie, Keith Jerome, Alexander Greninger
EPI_ISL_424251 hCoV-19/USA/WA-UW-1801/2020 North America / USA / Washington 2020-03-23 UW Virology Lab UW Virology Lab Pavitra Roychoudhury, Hong Xie, Keith Jerome, Alexander Greninger
EPI_ISL_424252 hCoV-19/USA/WA-UW-1840/2020 North America / USA / Washington 2020-03-23 UW Virology Lab UW Virology Lab Pavitra Roychoudhury, Hong Xie, Keith Jerome, Alexander Greninger
EPI_ISL_424253 hCoV-19/USA/WA-UW-1792/2020 North America / USA / Washington 2020-03-21 UW Virology Lab UW Virology Lab Pavitra Roychoudhury, Hong Xie, Keith Jerome, Alexander Greninger
EPI_ISL_424254 hCoV-19/USA/WA-UW-1796/2020 North America / USA / Washington 2020-03-21 UW Virology Lab UW Virology Lab Pavitra Roychoudhury, Hong Xie, Keith Jerome, Alexander Greninger
EPI_ISL_424255 hCoV-19/USA/UN-UW-1832/2020 North America / USA 2020-03-16 UW Virology Lab UW Virology Lab Pavitra Roychoudhury, Hong Xie, Keith Jerome, Alexander Greninger
EPI_ISL_424256 hCoV-19/USA/WA-UW-1774/2020 North America / USA / Washington 2020-03-22 UW Virology Lab UW Virology Lab Pavitra Roychoudhury, Hong Xie, Keith Jerome, Alexander Greninger

EPI_ISL_424257 hCoV-19/USA/WA-UW-1775/2020 North America / USA / Washington 2020-03-22 UW Virology Lab UW Virology Lab Pavitra Roychoudhury, Hong Xie, Keith Jerome, Alexander Greninger
EPI_ISL_424258 hCoV-19/USA/WA-UW-1778/2020 North America / USA / Washington 2020-03-22 UW Virology Lab UW Virology Lab Pavitra Roychoudhury, Hong Xie, Keith Jerome, Alexander Greninger
EPI_ISL_424259 hCoV-19/USA/WA-UW-1797/2020 North America / USA / Washington 2020-03-22 UW Virology Lab UW Virology Lab Pavitra Roychoudhury, Hong Xie, Keith Jerome, Alexander Greninger
EPI_ISL_424260 hCoV-19/USA/WA-UW-1779/2020 North America / USA / Washington 2020-03-22 UW Virology Lab UW Virology Lab Pavitra Roychoudhury, Hong Xie, Keith Jerome, Alexander Greninger
EPI_ISL_424261 hCoV-19/USA/WA-UW-1784/2020 North America / USA / Washington 2020-03-21 UW Virology Lab UW Virology Lab Pavitra Roychoudhury, Hong Xie, Keith Jerome, Alexander Greninger
EPI_ISL_424262 hCoV-19/USA/WA-UW-1782/2020 North America / USA / Washington 2020-03-22 UW Virology Lab UW Virology Lab Pavitra Roychoudhury, Hong Xie, Keith Jerome, Alexander Greninger
EPI_ISL_424263 hCoV-19/USA/WA-UW-1799/2020 North America / USA / Washington 2020-03-22 UW Virology Lab UW Virology Lab Pavitra Roychoudhury, Hong Xie, Keith Jerome, Alexander Greninger
EPI_ISL_424264 hCoV-19/USA/WA-UW-1805/2020 North America / USA / Washington 2020-03-22 UW Virology Lab UW Virology Lab Pavitra Roychoudhury, Hong Xie, Keith Jerome, Alexander Greninger
EPI_ISL_424266 hCoV-19/USA/UN-UW-1818/2020 North America / USA 2020-03-22 UW Virology Lab UW Virology Lab Pavitra Roychoudhury, Hong Xie, Keith Jerome, Alexander Greninger
EPI_ISL_424267 hCoV-19/USA/UN-UW-1817/2020 North America / USA 2020-03-22 UW Virology Lab UW Virology Lab Pavitra Roychoudhury, Hong Xie, Keith Jerome, Alexander Greninger
EPI_ISL_424268 hCoV-19/USA/UN-UW-1816/2020 North America / USA 2020-03-22 UW Virology Lab UW Virology Lab Pavitra Roychoudhury, Hong Xie, Keith Jerome, Alexander Greninger
EPI_ISL_424269 hCoV-19/USA/WA-UW-1828/2020 North America / USA / Washington 2020-03-22 UW Virology Lab UW Virology Lab Pavitra Roychoudhury, Hong Xie, Keith Jerome, Alexander Greninger
EPI_ISL_424270 hCoV-19/USA/WA-UW-1831/2020 North America / USA / Washington 2020-03-22 UW Virology Lab UW Virology Lab Pavitra Roychoudhury, Hong Xie, Keith Jerome, Alexander Greninger
EPI_ISL_424271 hCoV-19/USA/WA-UW-1827/2020 North America / USA / Washington 2020-03-22 UW Virology Lab UW Virology Lab Pavitra Roychoudhury, Hong Xie, Keith Jerome, Alexander Greninger
EPI_ISL_424273 hCoV-19/USA/WA-UW-1826/2020 North America / USA / Washington 2020-03-22 UW Virology Lab UW Virology Lab Pavitra Roychoudhury, Hong Xie, Keith Jerome, Alexander Greninger
EPI_ISL_424274 hCoV-19/USA/WA-UW-1824/2020 North America / USA / Washington 2020-03-22 UW Virology Lab UW Virology Lab Pavitra Roychoudhury, Hong Xie, Keith Jerome, Alexander Greninger
EPI_ISL_424275 hCoV-19/USA/WA-UW-1821/2020 North America / USA / Washington 2020-03-22 UW Virology Lab UW Virology Lab Pavitra Roychoudhury, Hong Xie, Keith Jerome, Alexander Greninger
EPI_ISL_424276 hCoV-19/USA/WA-UW-1822/2020 North America / USA / Washington 2020-03-22 UW Virology Lab UW Virology Lab Pavitra Roychoudhury, Hong Xie, Keith Jerome, Alexander Greninger
EPI_ISL_424278 hCoV-19/USA/WA-UW-1866/2020 North America / USA / Washington 2020-03-23 UW Virology Lab UW Virology Lab Pavitra Roychoudhury, Hong Xie, Keith Jerome, Alexander Greninger
EPI_ISL_424279 hCoV-19/USA/WA-UW-1850/2020 North America / USA / Washington 2020-03-23 UW Virology Lab UW Virology Lab Pavitra Roychoudhury, Hong Xie, Keith Jerome, Alexander Greninger
EPI_ISL_424280 hCoV-19/USA/WA-UW-1868/2020 North America / USA / Washington 2020-03-23 UW Virology Lab UW Virology Lab Pavitra Roychoudhury, Hong Xie, Keith Jerome, Alexander Greninger
EPI_ISL_424281 hCoV-19/USA/WA-UW-1864/2020 North America / USA / Washington 2020-03-23 UW Virology Lab UW Virology Lab Pavitra Roychoudhury, Hong Xie, Keith Jerome, Alexander Greninger
EPI_ISL_424282 hCoV-19/USA/WA-UW-1835/2020 North America / USA / Washington 2020-03-23 UW Virology Lab UW Virology Lab Pavitra Roychoudhury, Hong Xie, Keith Jerome, Alexander Greninger
EPI_ISL_424284 hCoV-19/USA/WA-UW-1863/2020 North America / USA / Washington 2020-03-23 UW Virology Lab UW Virology Lab Pavitra Roychoudhury, Hong Xie, Keith Jerome, Alexander Greninger
EPI_ISL_424285 hCoV-19/USA/OR-UW-1851/2020 North America / USA / Oregon 2020-03-21 UW Virology Lab UW Virology Lab Pavitra Roychoudhury, Hong Xie, Keith Jerome, Alexander Greninger
EPI_ISL_424286 hCoV-19/USA/OR-UW-1849/2020 North America / USA / Oregon 2020-03-20 UW Virology Lab UW Virology Lab Pavitra Roychoudhury, Hong Xie, Keith Jerome, Alexander Greninger
EPI_ISL_424287 hCoV-19/USA/WA-UW-1843/2020 North America / USA / Washington 2020-03-23 UW Virology Lab UW Virology Lab Pavitra Roychoudhury, Hong Xie, Keith Jerome, Alexander Greninger
EPI_ISL_424288 hCoV-19/USA/WA-UW-1846/2020 North America / USA / Washington 2020-03-22 UW Virology Lab UW Virology Lab Pavitra Roychoudhury, Hong Xie, Keith Jerome, Alexander Greninger
EPI_ISL_424289 hCoV-19/USA/WA-UW-1860/2020 North America / USA / Washington 2020-03-23 UW Virology Lab UW Virology Lab Pavitra Roychoudhury, Hong Xie, Keith Jerome, Alexander Greninger
EPI_ISL_424290 hCoV-19/USA/WA-UW-1853/2020 North America / USA / Washington 2020-03-23 UW Virology Lab UW Virology Lab Pavitra Roychoudhury, Hong Xie, Keith Jerome, Alexander Greninger
EPI_ISL_424291 hCoV-19/USA/WA-UW-1854/2020 North America / USA / Washington 2020-03-23 UW Virology Lab UW Virology Lab Pavitra Roychoudhury, Hong Xie, Keith Jerome, Alexander Greninger
EPI_ISL_424292 hCoV-19/USA/WA-UW-1855/2020 North America / USA / Washington 2020-03-23 UW Virology Lab UW Virology Lab Pavitra Roychoudhury, Hong Xie, Keith Jerome, Alexander Greninger
EPI_ISL_424293 hCoV-19/USA/WA-UW-1870/2020 North America / USA / Washington 2020-03-23 UW Virology Lab UW Virology Lab Pavitra Roychoudhury, Hong Xie, Keith Jerome, Alexander Greninger
EPI_ISL_424294 hCoV-19/USA/WA-UW-1872/2020 North America / USA / Washington 2020-03-23 UW Virology Lab UW Virology Lab Pavitra Roychoudhury, Hong Xie, Keith Jerome, Alexander Greninger
EPI_ISL_424295 hCoV-19/USA/WA-UW-1905/2020 North America / USA / Washington 2020-03-23 UW Virology Lab UW Virology Lab Pavitra Roychoudhury, Hong Xie, Keith Jerome, Alexander Greninger

EPI_ISL_424296 hCoV-19/USA/WA-UW-1919/2020 North America / USA / Washington 2020-03-23 UW Virology Lab UW Virology Lab Pavitra Roychoudhury, Hong Xie, Keith Jerome, Alexander Greninger
EPI_ISL_424297 hCoV-19/USA/WA-UW-1913/2020 North America / USA / Washington 2020-03-23 UW Virology Lab UW Virology Lab Pavitra Roychoudhury, Hong Xie, Keith Jerome, Alexander Greninger
EPI_ISL_424298 hCoV-19/USA/WA-UW-1920/2020 North America / USA / Washington 2020-03-23 UW Virology Lab UW Virology Lab Pavitra Roychoudhury, Hong Xie, Keith Jerome, Alexander Greninger
EPI_ISL_424299 hCoV-19/USA/WA-UW-1904/2020 North America / USA / Washington 2020-03-23 UW Virology Lab UW Virology Lab Pavitra Roychoudhury, Hong Xie, Keith Jerome, Alexander Greninger
EPI_ISL_425501 hCoV-19/England/NOTT-10E12C/2020 Europe / United Kingdom / England 2020-03-15 Queens Medical Centre, Clinical Microbiology Department / DeepSeq Nottingham COVID-19 Genomics UK (COG-UK) Consortium Gemma Clark, Wendy Smith, Manjinder Khakh, Hannah Howson-Wells, Jonathan Ball, Patrick McClure, Joseph Chappell, Theocharis Tsoleridis, Nadine Holmes, Matthew Carlisle, Christopher Moore, Fei Sang, Johnny Debebe, Victoria Wright, Matthew Loose
EPI_ISL_425502 hCoV-19/England/NOTT-10E14A/2020 Europe / United Kingdom / England 2020-03-17 Queens Medical Centre, Clinical Microbiology Department / DeepSeq Nottingham COVID-19 Genomics UK (COG-UK) Consortium Gemma Clark, Wendy Smith, Manjinder Khakh, Hannah Howson-Wells, Jonathan Ball, Patrick McClure, Joseph Chappell, Theocharis Tsoleridis, Nadine Holmes, Matthew Carlisle, Christopher Moore, Fei Sang, Johnny Debebe, Victoria Wright, Matthew Loose
EPI_ISL_425504 hCoV-19/England/NOTT-10E186/2020 Europe / United Kingdom / England 2020-03-18 Queens Medical Centre, Clinical Microbiology Department / DeepSeq Nottingham COVID-19 Genomics UK (COG-UK) Consortium Gemma Clark, Wendy Smith, Manjinder Khakh, Hannah Howson-Wells, Jonathan Ball, Patrick McClure, Joseph Chappell, Theocharis Tsoleridis, Nadine Holmes, Matthew Carlisle, Christopher Moore, Fei Sang, Johnny Debebe, Victoria Wright, Matthew Loose
EPI_ISL_425505 hCoV-19/England/NOTT-10E195/2020 Europe / United Kingdom / England 2020-03-18 Queens Medical Centre, Clinical Microbiology Department / DeepSeq Nottingham COVID-19 Genomics UK (COG-UK) Consortium Gemma Clark, Wendy Smith, Manjinder Khakh, Hannah Howson-Wells, Jonathan Ball, Patrick McClure, Joseph Chappell, Theocharis Tsoleridis, Nadine Holmes, Matthew Carlisle, Christopher Moore, Fei Sang, Johnny Debebe, Victoria Wright, Matthew Loose
EPI_ISL_425510 hCoV-19/England/NOTT-10E1E0/2020 Europe / United Kingdom / England 2020-03-19 Queens Medical Centre, Clinical Microbiology Department / DeepSeq Nottingham COVID-19 Genomics UK (COG-UK) Consortium Gemma Clark, Wendy Smith, Manjinder Khakh, Hannah Howson-Wells, Jonathan Ball, Patrick McClure, Joseph Chappell, Theocharis Tsoleridis, Nadine Holmes, Matthew Carlisle, Christopher Moore, Fei Sang, Johnny Debebe, Victoria Wright, Matthew Loose
EPI_ISL_425513 hCoV-19/England/NOTT-10E21A/2020 Europe / United Kingdom / England 2020-03-20 Queens Medical Centre, Clinical Microbiology Department / DeepSeq Nottingham COVID-19 Genomics UK (COG-UK) Consortium Gemma Clark, Wendy Smith, Manjinder Khakh, Hannah Howson-Wells, Jonathan Ball, Patrick McClure, Joseph Chappell, Theocharis Tsoleridis, Nadine Holmes, Matthew Carlisle, Christopher Moore, Fei Sang, Johnny Debebe, Victoria Wright, Matthew Loose
EPI_ISL_425514 hCoV-19/England/NOTT-10E229/2020 Europe / United Kingdom / England 2020-03-20 Queens Medical Centre, Clinical Microbiology Department / DeepSeq Nottingham COVID-19 Genomics UK (COG-UK) Consortium Gemma Clark, Wendy Smith, Manjinder Khakh, Hannah Howson-Wells, Jonathan Ball, Patrick McClure, Joseph Chappell, Theocharis Tsoleridis, Nadine Holmes, Matthew Carlisle, Christopher Moore, Fei Sang, Johnny Debebe, Victoria Wright, Matthew Loose
EPI_ISL_425515 hCoV-19/England/NOTT-10E238/2020 Europe / United Kingdom / England 2020-03-20 Queens Medical Centre, Clinical Microbiology Department / DeepSeq Nottingham COVID-19 Genomics UK (COG-UK) Consortium Gemma Clark, Wendy Smith, Manjinder Khakh, Hannah Howson-Wells, Jonathan Ball, Patrick McClure, Joseph Chappell, Theocharis Tsoleridis, Nadine Holmes, Matthew Carlisle, Christopher Moore, Fei Sang, Johnny Debebe, Victoria Wright, Matthew Loose
EPI_ISL_425517 hCoV-19/England/NOTT-10E256/2020 Europe / United Kingdom / England 2020-03-20 Queens Medical Centre, Clinical Microbiology Department / DeepSeq Nottingham COVID-19 Genomics UK (COG-UK) Consortium Gemma Clark, Wendy Smith, Manjinder Khakh, Hannah Howson-Wells, Jonathan Ball, Patrick McClure, Joseph Chappell, Theocharis Tsoleridis, Nadine Holmes, Matthew Carlisle, Christopher Moore, Fei Sang, Johnny Debebe, Victoria Wright, Matthew Loose
EPI_ISL_425518 hCoV-19/England/NOTT-10E265/2020 Europe / United Kingdom / England 2020-03-20 Queens Medical Centre, Clinical Microbiology Department / DeepSeq Nottingham COVID-19 Genomics UK (COG-UK) Consortium Gemma Clark, Wendy Smith, Manjinder Khakh, Hannah Howson-Wells, Jonathan Ball, Patrick McClure, Joseph Chappell, Theocharis Tsoleridis, Nadine Holmes, Matthew Carlisle, Christopher Moore, Fei Sang, Johnny Debebe, Victoria Wright, Matthew Loose
EPI_ISL_425519 hCoV-19/England/NOTT-10E283/2020 Europe / United Kingdom / England 2020-03-20 Queens Medical Centre, Clinical Microbiology Department / DeepSeq Nottingham COVID-19 Genomics UK (COG-UK) Consortium Gemma Clark, Wendy Smith, Manjinder Khakh, Hannah Howson-Wells, Jonathan Ball, Patrick McClure, Joseph Chappell, Theocharis Tsoleridis, Nadine Holmes, Matthew Carlisle, Christopher Moore, Fei Sang, Johnny Debebe, Victoria Wright, Matthew Loose
EPI_ISL_425522 hCoV-19/England/NOTT-10E2B0/2020 Europe / United Kingdom / England 2020-03-20 Queens Medical Centre, Clinical Microbiology Department / DeepSeq Nottingham COVID-19 Genomics UK (COG-UK) Consortium Gemma Clark, Wendy Smith, Manjinder Khakh, Hannah Howson-Wells, Jonathan Ball, Patrick McClure, Joseph Chappell, Theocharis Tsoleridis, Nadine Holmes, Matthew Carlisle, Christopher Moore, Fei Sang, Johnny Debebe, Victoria Wright, Matthew Loose
EPI_ISL_425523 hCoV-19/England/NOTT-10E2CF/2020 Europe / United Kingdom / England 2020-03-20 Queens Medical Centre, Clinical Microbiology Department / DeepSeq Nottingham COVID-19 Genomics UK (COG-UK)

Consortium	Gemma Clark, Wendy Smith, Manjinder Khakh, Hannah Howson-Wells, Jonathan Ball, Patrick McClure, Joseph Chappell, Theocharis Tsoleridis, Nadine Holmes, Matthew Carlisle, Christopher Moore, Fei Sang, Johnny Debebe, Victoria Wright, Matthew Loose
EPI_ISL_425525	hCoV-19/England/NOTT-10E2ED/2020	Europe / United Kingdom / England	2020-03-21	Queens Medical Centre, Clinical Microbiology Department / DeepSeq Nottingham	COVID-19 Genomics UK (COG-UK) Consortium	Gemma Clark, Wendy Smith, Manjinder Khakh, Hannah Howson-Wells, Jonathan Ball, Patrick McClure, Joseph Chappell, Theocharis Tsoleridis, Nadine Holmes, Matthew Carlisle, Christopher Moore, Fei Sang, Johnny Debebe, Victoria Wright, Matthew Loose
EPI_ISL_425526	hCoV-19/England/NOTT-10E2FC/2020	Europe / United Kingdom / England	2020-03-21	Queens Medical Centre, Clinical Microbiology Department / DeepSeq Nottingham	COVID-19 Genomics UK (COG-UK) Consortium	Gemma Clark, Wendy Smith, Manjinder Khakh, Hannah Howson-Wells, Jonathan Ball, Patrick McClure, Joseph Chappell, Theocharis Tsoleridis, Nadine Holmes, Matthew Carlisle, Christopher Moore, Fei Sang, Johnny Debebe, Victoria Wright, Matthew Loose
EPI_ISL_425527	hCoV-19/England/NOTT-10E308/2020	Europe / United Kingdom / England	2020-03-19	Queens Medical Centre, Clinical Microbiology Department / DeepSeq Nottingham	COVID-19 Genomics UK (COG-UK) Consortium	Gemma Clark, Wendy Smith, Manjinder Khakh, Hannah Howson-Wells, Jonathan Ball, Patrick McClure, Joseph Chappell, Theocharis Tsoleridis, Nadine Holmes, Matthew Carlisle, Christopher Moore, Fei Sang, Johnny Debebe, Victoria Wright, Matthew Loose
EPI_ISL_425530	hCoV-19/England/NOTT-10E335/2020	Europe / United Kingdom / England	2020-03-21	Queens Medical Centre, Clinical Microbiology Department / DeepSeq Nottingham	COVID-19 Genomics UK (COG-UK) Consortium	Gemma Clark, Wendy Smith, Manjinder Khakh, Hannah Howson-Wells, Jonathan Ball, Patrick McClure, Joseph Chappell, Theocharis Tsoleridis, Nadine Holmes, Matthew Carlisle, Christopher Moore, Fei Sang, Johnny Debebe, Victoria Wright, Matthew Loose
EPI_ISL_425532	hCoV-19/England/NOTT-10E362/2020	Europe / United Kingdom / England	2020-03-21	Queens Medical Centre, Clinical Microbiology Department / DeepSeq Nottingham	COVID-19 Genomics UK (COG-UK) Consortium	Gemma Clark, Wendy Smith, Manjinder Khakh, Hannah Howson-Wells, Jonathan Ball, Patrick McClure, Joseph Chappell, Theocharis Tsoleridis, Nadine Holmes, Matthew Carlisle, Christopher Moore, Fei Sang, Johnny Debebe, Victoria Wright, Matthew Loose
EPI_ISL_425533	hCoV-19/England/NOTT-10E371/2020	Europe / United Kingdom / England	2020-03-21	Queens Medical Centre, Clinical Microbiology Department / DeepSeq Nottingham	COVID-19 Genomics UK (COG-UK) Consortium	Gemma Clark, Wendy Smith, Manjinder Khakh, Hannah Howson-Wells, Jonathan Ball, Patrick McClure, Joseph Chappell, Theocharis Tsoleridis, Nadine Holmes, Matthew Carlisle, Christopher Moore, Fei Sang, Johnny Debebe, Victoria Wright, Matthew Loose
EPI_ISL_425535	hCoV-19/England/NOTT-10E39F/2020	Europe / United Kingdom / England	2020-03-21	Queens Medical Centre, Clinical Microbiology Department / DeepSeq Nottingham	COVID-19 Genomics UK (COG-UK) Consortium	Gemma Clark, Wendy Smith, Manjinder Khakh, Hannah Howson-Wells, Jonathan Ball, Patrick McClure, Joseph Chappell, Theocharis Tsoleridis, Nadine Holmes, Matthew Carlisle, Christopher Moore, Fei Sang, Johnny Debebe, Victoria Wright, Matthew Loose
EPI_ISL_425536	hCoV-19/England/NOTT-10E3AE/2020	Europe / United Kingdom / England	2020-03-21	Queens Medical Centre, Clinical Microbiology Department / DeepSeq Nottingham	COVID-19 Genomics UK (COG-UK) Consortium	Gemma Clark, Wendy Smith, Manjinder Khakh, Hannah Howson-Wells, Jonathan Ball, Patrick McClure, Joseph Chappell, Theocharis Tsoleridis, Nadine Holmes, Matthew Carlisle, Christopher Moore, Fei Sang, Johnny Debebe, Victoria Wright, Matthew Loose
EPI_ISL_425537	hCoV-19/England/NOTT-10E3BD/2020	Europe / United Kingdom / England	2020-03-21	Queens Medical Centre, Clinical Microbiology Department / DeepSeq Nottingham	COVID-19 Genomics UK (COG-UK) Consortium	Gemma Clark, Wendy Smith, Manjinder Khakh, Hannah Howson-Wells, Jonathan Ball, Patrick McClure, Joseph Chappell, Theocharis Tsoleridis, Nadine Holmes, Matthew Carlisle, Christopher Moore, Fei Sang, Johnny Debebe, Victoria Wright, Matthew Loose
EPI_ISL_425548	hCoV-19/England/NOTT-10E46F/2020	Europe / United Kingdom / England	2020-03-22	Queens Medical Centre, Clinical Microbiology Department / DeepSeq Nottingham	COVID-19 Genomics UK (COG-UK) Consortium	Gemma Clark, Wendy Smith, Manjinder Khakh, Hannah Howson-Wells, Jonathan Ball, Patrick McClure, Joseph Chappell, Theocharis Tsoleridis, Nadine Holmes, Matthew Carlisle, Christopher Moore, Fei Sang, Johnny Debebe, Victoria Wright, Matthew Loose
EPI_ISL_425549	hCoV-19/England/NOTT-10E47E/2020	Europe / United Kingdom / England	2020-03-22	Queens Medical Centre, Clinical Microbiology Department / DeepSeq Nottingham	COVID-19 Genomics UK (COG-UK) Consortium	Gemma Clark, Wendy Smith, Manjinder Khakh, Hannah Howson-Wells, Jonathan Ball, Patrick McClure, Joseph Chappell, Theocharis Tsoleridis, Nadine Holmes, Matthew Carlisle, Christopher Moore, Fei Sang, Johnny Debebe, Victoria Wright, Matthew Loose
EPI_ISL_425551	hCoV-19/England/NOTT-10E49C/2020	Europe / United Kingdom / England	2020-03-22	Queens Medical Centre, Clinical Microbiology Department / DeepSeq Nottingham	COVID-19 Genomics UK (COG-UK) Consortium	Gemma Clark, Wendy Smith, Manjinder Khakh, Hannah Howson-Wells, Jonathan Ball, Patrick McClure, Joseph Chappell, Theocharis Tsoleridis, Nadine Holmes, Matthew Carlisle, Christopher Moore, Fei Sang, Johnny Debebe, Victoria Wright, Matthew Loose
EPI_ISL_425554	hCoV-19/England/NOTT-10E4C9/2020	Europe / United Kingdom / England	2020-03-22	Queens Medical Centre, Clinical Microbiology Department / DeepSeq Nottingham	COVID-19 Genomics UK (COG-UK) Consortium	Gemma Clark, Wendy Smith, Manjinder Khakh, Hannah Howson-Wells, Jonathan Ball, Patrick McClure, Joseph Chappell, Theocharis Tsoleridis, Nadine Holmes, Matthew Carlisle, Christopher Moore, Fei Sang, Johnny Debebe, Victoria Wright, Matthew Loose
EPI_ISL_425561	hCoV-19/England/NOTT-10E55D/2020	Europe / United Kingdom / England	2020-03-25	Queens Medical Centre, Clinical Microbiology Department / DeepSeq Nottingham	COVID-19 Genomics UK (COG-UK)

Consortium	Gemma Clark, Wendy Smith, Manjinder Khakh, Hannah Howson-Wells, Jonathan Ball, Patrick McClure, Joseph Chappell, Theocharis Tsoleridis, Nadine Holmes, Matthew Carlisle, Christopher Moore, Fei Sang, Johnny Debebe, Victoria Wright, Matthew Loose
EPI_ISL_425562	hCoV-19/England/NOTT-10E57B/2020	Europe / United Kingdom / England	2020-03-25	Queens Medical Centre, Clinical Microbiology Department / DeepSeq Nottingham	COVID-19 Genomics UK (COG-UK) Consortium	Gemma Clark, Wendy Smith, Manjinder Khakh, Hannah Howson-Wells, Jonathan Ball, Patrick McClure, Joseph Chappell, Theocharis Tsoleridis, Nadine Holmes, Matthew Carlisle, Christopher Moore, Fei Sang, Johnny Debebe, Victoria Wright, Matthew Loose
EPI_ISL_425563	hCoV-19/England/NOTT-10E58A/2020	Europe / United Kingdom / England	2020-03-26	Queens Medical Centre, Clinical Microbiology Department / DeepSeq Nottingham	COVID-19 Genomics UK (COG-UK) Consortium	Gemma Clark, Wendy Smith, Manjinder Khakh, Hannah Howson-Wells, Jonathan Ball, Patrick McClure, Joseph Chappell, Theocharis Tsoleridis, Nadine Holmes, Matthew Carlisle, Christopher Moore, Fei Sang, Johnny Debebe, Victoria Wright, Matthew Loose
EPI_ISL_425565	hCoV-19/England/NOTT-10E5A8/2020	Europe / United Kingdom / England	2020-03-26	Queens Medical Centre, Clinical Microbiology Department / DeepSeq Nottingham	COVID-19 Genomics UK (COG-UK) Consortium	Gemma Clark, Wendy Smith, Manjinder Khakh, Hannah Howson-Wells, Jonathan Ball, Patrick McClure, Joseph Chappell, Theocharis Tsoleridis, Nadine Holmes, Matthew Carlisle, Christopher Moore, Fei Sang, Johnny Debebe, Victoria Wright, Matthew Loose
EPI_ISL_425570	hCoV-19/England/NOTT-10E5F3/2020	Europe / United Kingdom / England	2020-03-23	Queens Medical Centre, Clinical Microbiology Department / DeepSeq Nottingham	COVID-19 Genomics UK (COG-UK) Consortium	Gemma Clark, Wendy Smith, Manjinder Khakh, Hannah Howson-Wells, Jonathan Ball, Patrick McClure, Joseph Chappell, Theocharis Tsoleridis, Nadine Holmes, Matthew Carlisle, Christopher Moore, Fei Sang, Johnny Debebe, Victoria Wright, Matthew Loose
EPI_ISL_425571	hCoV-19/England/NOTT-10E61E/2020	Europe / United Kingdom / England	2020-03-23	Queens Medical Centre, Clinical Microbiology Department / DeepSeq Nottingham	COVID-19 Genomics UK (COG-UK) Consortium	Gemma Clark, Wendy Smith, Manjinder Khakh, Hannah Howson-Wells, Jonathan Ball, Patrick McClure, Joseph Chappell, Theocharis Tsoleridis, Nadine Holmes, Matthew Carlisle, Christopher Moore, Fei Sang, Johnny Debebe, Victoria Wright, Matthew Loose
EPI_ISL_425573	hCoV-19/England/NOTT-10E64B/2020	Europe / United Kingdom / England	2020-03-23	Queens Medical Centre, Clinical Microbiology Department / DeepSeq Nottingham	COVID-19 Genomics UK (COG-UK) Consortium	Gemma Clark, Wendy Smith, Manjinder Khakh, Hannah Howson-Wells, Jonathan Ball, Patrick McClure, Joseph Chappell, Theocharis Tsoleridis, Nadine Holmes, Matthew Carlisle, Christopher Moore, Fei Sang, Johnny Debebe, Victoria Wright, Matthew Loose
EPI_ISL_425575	hCoV-19/England/NOTT-10E669/2020	Europe / United Kingdom / England	2020-03-24	Queens Medical Centre, Clinical Microbiology Department / DeepSeq Nottingham	COVID-19 Genomics UK (COG-UK) Consortium	Gemma Clark, Wendy Smith, Manjinder Khakh, Hannah Howson-Wells, Jonathan Ball, Patrick McClure, Joseph Chappell, Theocharis Tsoleridis, Nadine Holmes, Matthew Carlisle, Christopher Moore, Fei Sang, Johnny Debebe, Victoria Wright, Matthew Loose
EPI_ISL_425576	hCoV-19/England/NOTT-10E678/2020	Europe / United Kingdom / England	2020-03-24	Queens Medical Centre, Clinical Microbiology Department / DeepSeq Nottingham	COVID-19 Genomics UK (COG-UK) Consortium	Gemma Clark, Wendy Smith, Manjinder Khakh, Hannah Howson-Wells, Jonathan Ball, Patrick McClure, Joseph Chappell, Theocharis Tsoleridis, Nadine Holmes, Matthew Carlisle, Christopher Moore, Fei Sang, Johnny Debebe, Victoria Wright, Matthew Loose
EPI_ISL_425581	hCoV-19/England/NOTT-10E6C3/2020	Europe / United Kingdom / England	2020-03-24	Queens Medical Centre, Clinical Microbiology Department / DeepSeq Nottingham	COVID-19 Genomics UK (COG-UK) Consortium	Gemma Clark, Wendy Smith, Manjinder Khakh, Hannah Howson-Wells, Jonathan Ball, Patrick McClure, Joseph Chappell, Theocharis Tsoleridis, Nadine Holmes, Matthew Carlisle, Christopher Moore, Fei Sang, Johnny Debebe, Victoria Wright, Matthew Loose
EPI_ISL_425584	hCoV-19/England/NOTT-10E71B/2020	Europe / United Kingdom / England	2020-03-24	Queens Medical Centre, Clinical Microbiology Department / DeepSeq Nottingham	COVID-19 Genomics UK (COG-UK) Consortium	Gemma Clark, Wendy Smith, Manjinder Khakh, Hannah Howson-Wells, Jonathan Ball, Patrick McClure, Joseph Chappell, Theocharis Tsoleridis, Nadine Holmes, Matthew Carlisle, Christopher Moore, Fei Sang, Johnny Debebe, Victoria Wright, Matthew Loose
EPI_ISL_425586	hCoV-19/England/NOTT-10E739/2020	Europe / United Kingdom / England	2020-03-24	Queens Medical Centre, Clinical Microbiology Department / DeepSeq Nottingham	COVID-19 Genomics UK (COG-UK) Consortium	Gemma Clark, Wendy Smith, Manjinder Khakh, Hannah Howson-Wells, Jonathan Ball, Patrick McClure, Joseph Chappell, Theocharis Tsoleridis, Nadine Holmes, Matthew Carlisle, Christopher Moore, Fei Sang, Johnny Debebe, Victoria Wright, Matthew Loose
EPI_ISL_425587	hCoV-19/England/NOTT-10E748/2020	Europe / United Kingdom / England	2020-03-24	Queens Medical Centre, Clinical Microbiology Department / DeepSeq Nottingham	COVID-19 Genomics UK (COG-UK) Consortium	Gemma Clark, Wendy Smith, Manjinder Khakh, Hannah Howson-Wells, Jonathan Ball, Patrick McClure, Joseph Chappell, Theocharis Tsoleridis, Nadine Holmes, Matthew Carlisle, Christopher Moore, Fei Sang, Johnny Debebe, Victoria Wright, Matthew Loose
EPI_ISL_425593	hCoV-19/England/NOTT-10E7A2/2020	Europe / United Kingdom / England	2020-03-25	Queens Medical Centre, Clinical Microbiology Department / DeepSeq Nottingham	COVID-19 Genomics UK (COG-UK) Consortium	Gemma Clark, Wendy Smith, Manjinder Khakh, Hannah Howson-Wells, Jonathan Ball, Patrick McClure, Joseph Chappell, Theocharis Tsoleridis, Nadine Holmes, Matthew Carlisle, Christopher Moore, Fei Sang, Johnny Debebe, Victoria Wright, Matthew Loose
EPI_ISL_425700	hCoV-19/Scotland/CVR161/2020	Europe / United Kingdom / Scotland	2020-03-17	West of Scotland Specialist Virology Centre, NHSGGC / MRC-University of Glasgow Centre for Virus Research	COVID-

19 Genomics UK (COG-UK) Consortium	Ana da Silva Filipe, Kathy Smollett, Stephen Carmichael, Natasha Johnson, Daniel Mair, Lily Tong, Jenna Nichols; Sarah McDonald; Richard Orton, Joseph Hughes, Sreenu Vattipally, David L Robertson; Kathy Li, Natasha Jesudason, Rajiv Shah, James Shepherd, Antonia Ho, Emma Thomson; Alasdair MacLean, Rory Gunson.
EPI_ISL_425701	hCoV-19/Scotland/CVR162/2020	Europe / United Kingdom / Scotland	2020-03-17	West of Scotland Specialist Virology Centre, NHSGGC / MRC-University of Glasgow Centre for Virus Research	COVID-19 Genomics UK (COG-UK) Consortium	Ana da Silva Filipe, Kathy Smollett, Stephen Carmichael, Natasha Johnson, Daniel Mair, Lily Tong, Jenna Nichols; Sarah McDonald; Richard Orton, Joseph Hughes, Sreenu Vattipally, David L Robertson; Kathy Li, Natasha Jesudason, Rajiv Shah, James Shepherd, Antonia Ho, Emma Thomson; Alasdair MacLean, Rory Gunson.
EPI_ISL_426800	hCoV-19/Australia/VIC534/2020	Oceania / Australia / Victoria	2020-03-25	Victorian Infectious Diseases Reference Laboratory (VIDRL)	Microbiological Diagnostic Unit Public Health Laboratory and Victorian Infectious Diseases Reference Laboratory, Doherty Institute	Caly L., Seemann T., Sait, M., Schultz M., Druce J., Sherry, N.
EPI_ISL_426801	hCoV-19/Australia/VIC535/2020	Oceania / Australia / Victoria	2020-03-25	Victorian Infectious Diseases Reference Laboratory (VIDRL)	Microbiological Diagnostic Unit Public Health Laboratory and Victorian Infectious Diseases Reference Laboratory, Doherty Institute	Caly L., Seemann T., Sait, M., Schultz M., Druce J., Sherry, N.
EPI_ISL_426802	hCoV-19/Australia/VIC536/2020	Oceania / Australia / Victoria	2020-03-24	Victorian Infectious Diseases Reference Laboratory (VIDRL)	Microbiological Diagnostic Unit Public Health Laboratory and Victorian Infectious Diseases Reference Laboratory, Doherty Institute	Caly L., Seemann T., Sait, M., Schultz M., Druce J., Sherry, N.
EPI_ISL_426803	hCoV-19/Australia/VIC537/2020	Oceania / Australia / Victoria	2020-03-24	Victorian Infectious Diseases Reference Laboratory (VIDRL)	Microbiological Diagnostic Unit Public Health Laboratory and Victorian Infectious Diseases Reference Laboratory, Doherty Institute	Caly L., Seemann T., Sait, M., Schultz M., Druce J., Sherry, N.
EPI_ISL_426804	hCoV-19/Australia/VIC538/2020	Oceania / Australia / Victoria	2020-03-23	Victorian Infectious Diseases Reference Laboratory (VIDRL)	Microbiological Diagnostic Unit Public Health Laboratory and Victorian Infectious Diseases Reference Laboratory, Doherty Institute	Caly L., Seemann T., Sait, M., Schultz M., Druce J., Sherry, N.
EPI_ISL_426805	hCoV-19/Australia/VIC540/2020	Oceania / Australia / Victoria	2020-03-24	Victorian Infectious Diseases Reference Laboratory (VIDRL)	Microbiological Diagnostic Unit Public Health Laboratory and Victorian Infectious Diseases Reference Laboratory, Doherty Institute	Caly L., Seemann T., Sait, M., Schultz M., Druce J., Sherry, N.
EPI_ISL_426806	hCoV-19/Australia/VIC541/2020	Oceania / Australia / Victoria	2020-03-24	Victorian Infectious Diseases Reference Laboratory (VIDRL)	Microbiological Diagnostic Unit Public Health Laboratory and Victorian Infectious Diseases Reference Laboratory, Doherty Institute	Caly L., Seemann T., Sait, M., Schultz M., Druce J., Sherry, N.
EPI_ISL_426807	hCoV-19/Australia/VIC544/2020	Oceania / Australia / Victoria	2020-03-23	Victorian Infectious Diseases Reference Laboratory (VIDRL)	Microbiological Diagnostic Unit Public Health Laboratory and Victorian Infectious Diseases Reference Laboratory, Doherty Institute	Caly L., Seemann T., Sait, M., Schultz M., Druce J., Sherry, N.
EPI_ISL_426809	hCoV-19/Australia/VIC546/2020	Oceania / Australia / Victoria	2020-03-24	Victorian Infectious Diseases Reference Laboratory (VIDRL)	Microbiological Diagnostic Unit Public Health Laboratory and Victorian Infectious Diseases Reference Laboratory, Doherty Institute	Caly L., Seemann T., Sait, M., Schultz M., Druce J., Sherry, N.
EPI_ISL_426810	hCoV-19/Australia/VIC547/2020	Oceania / Australia / Victoria	2020-03-25	Victorian Infectious Diseases Reference Laboratory (VIDRL)	Microbiological Diagnostic Unit Public Health Laboratory and Victorian Infectious Diseases Reference Laboratory, Doherty Institute	Caly L., Seemann T., Sait, M., Schultz M., Druce J., Sherry, N.
EPI_ISL_426811	hCoV-19/Australia/VIC548/2020	Oceania / Australia / Victoria	2020-03-25	Victorian Infectious Diseases Reference Laboratory (VIDRL)	Microbiological Diagnostic Unit Public Health Laboratory and Victorian Infectious Diseases Reference Laboratory, Doherty Institute	Caly L., Seemann T., Sait, M., Schultz M., Druce J., Sherry, N.
EPI_ISL_426812	hCoV-19/Australia/VIC549/2020	Oceania / Australia / Victoria	2020-03-23	Victorian Infectious Diseases Reference Laboratory (VIDRL)	Microbiological Diagnostic Unit Public Health Laboratory and Victorian Infectious Diseases Reference Laboratory, Doherty Institute	Caly L., Seemann T., Sait, M., Schultz M., Druce J., Sherry, N.
EPI_ISL_426813	hCoV-19/Australia/VIC551/2020	Oceania / Australia / Victoria	2020-03-25	Victorian Infectious Diseases Reference Laboratory (VIDRL)	Microbiological Diagnostic Unit Public Health Laboratory and Victorian Infectious Diseases Reference Laboratory, Doherty Institute	Caly L., Seemann T., Sait, M., Schultz M., Druce J., Sherry, N.
EPI_ISL_426816	hCoV-19/Australia/VIC555/2020	Oceania / Australia / Victoria	2020-03-25	Victorian Infectious Diseases Reference Laboratory (VIDRL)	Microbiological Diagnostic Unit Public Health Laboratory and Victorian Infectious Diseases Reference Laboratory, Doherty Institute	Caly L., Seemann T., Sait, M., Schultz M., Druce J., Sherry, N.
EPI_ISL_426817	hCoV-19/Australia/VIC556/2020	Oceania / Australia / Victoria	2020-03-25	Victorian Infectious Diseases Reference Laboratory (VIDRL)	Microbiological Diagnostic Unit Public Health Laboratory and Victorian Infectious Diseases Reference Laboratory, Doherty Institute	Caly L., Seemann T., Sait, M., Schultz M., Druce J., Sherry, N.

EPI_ISL_426819 hCoV-19/Australia/VIC558/2020 Oceania / Australia / Victoria 2020-03-25 Victorian Infectious Diseases Reference Laboratory (VIDRL) Microbiological Diagnostic Unit Public Health Laboratory and Victorian Infectious Diseases Reference Laboratory, Doherty Institute Caly L., Seemann T., Sait, M., Schultz M., Druce J., Sherry, N.
EPI_ISL_426821 hCoV-19/Australia/VIC560/2020 Oceania / Australia / Victoria 2020-03-25 Victorian Infectious Diseases Reference Laboratory (VIDRL) Microbiological Diagnostic Unit Public Health Laboratory and Victorian Infectious Diseases Reference Laboratory, Doherty Institute Caly L., Seemann T., Sait, M., Schultz M., Druce J., Sherry, N.
EPI_ISL_426822 hCoV-19/Australia/VIC561/2020 Oceania / Australia / Victoria 2020-03-25 Victorian Infectious Diseases Reference Laboratory (VIDRL) Microbiological Diagnostic Unit Public Health Laboratory and Victorian Infectious Diseases Reference Laboratory, Doherty Institute Caly L., Seemann T., Sait, M., Schultz M., Druce J., Sherry, N.
EPI_ISL_426824 hCoV-19/Australia/VIC563/2020 Oceania / Australia / Victoria 2020-03-24 Victorian Infectious Diseases Reference Laboratory (VIDRL) Microbiological Diagnostic Unit Public Health Laboratory and Victorian Infectious Diseases Reference Laboratory, Doherty Institute Caly L., Seemann T., Sait, M., Schultz M., Druce J., Sherry, N.
EPI_ISL_426825 hCoV-19/Australia/VIC565/2020 Oceania / Australia / Victoria 2020-03-25 Victorian Infectious Diseases Reference Laboratory (VIDRL) Microbiological Diagnostic Unit Public Health Laboratory and Victorian Infectious Diseases Reference Laboratory, Doherty Institute Caly L., Seemann T., Sait, M., Schultz M., Druce J., Sherry, N.
EPI_ISL_426826 hCoV-19/Australia/VIC566/2020 Oceania / Australia / Victoria 2020-03-25 Victorian Infectious Diseases Reference Laboratory (VIDRL) Microbiological Diagnostic Unit Public Health Laboratory and Victorian Infectious Diseases Reference Laboratory, Doherty Institute Caly L., Seemann T., Sait, M., Schultz M., Druce J., Sherry, N.
EPI_ISL_426827 hCoV-19/Australia/VIC567/2020 Oceania / Australia / Victoria 2020-03-25 Victorian Infectious Diseases Reference Laboratory (VIDRL) Microbiological Diagnostic Unit Public Health Laboratory and Victorian Infectious Diseases Reference Laboratory, Doherty Institute Caly L., Seemann T., Sait, M., Schultz M., Druce J., Sherry, N.
EPI_ISL_426828 hCoV-19/Australia/VIC568/2020 Oceania / Australia / Victoria 2020-03-25 Victorian Infectious Diseases Reference Laboratory (VIDRL) Microbiological Diagnostic Unit Public Health Laboratory and Victorian Infectious Diseases Reference Laboratory, Doherty Institute Caly L., Seemann T., Sait, M., Schultz M., Druce J., Sherry, N.
EPI_ISL_426831 hCoV-19/Australia/VIC571/2020 Oceania / Australia / Victoria 2020-03-25 Victorian Infectious Diseases Reference Laboratory (VIDRL) Microbiological Diagnostic Unit Public Health Laboratory and Victorian Infectious Diseases Reference Laboratory, Doherty Institute Caly L., Seemann T., Sait, M., Schultz M., Druce J., Sherry, N.
EPI_ISL_426832 hCoV-19/Australia/VIC572/2020 Oceania / Australia / Victoria 2020-03-25 Victorian Infectious Diseases Reference Laboratory (VIDRL) Microbiological Diagnostic Unit Public Health Laboratory and Victorian Infectious Diseases Reference Laboratory, Doherty Institute Caly L., Seemann T., Sait, M., Schultz M., Druce J., Sherry, N.
EPI_ISL_426833 hCoV-19/Australia/VIC574/2020 Oceania / Australia / Victoria 2020-03-26 Victorian Infectious Diseases Reference Laboratory (VIDRL) Microbiological Diagnostic Unit Public Health Laboratory and Victorian Infectious Diseases Reference Laboratory, Doherty Institute Caly L., Seemann T., Sait, M., Schultz M., Druce J., Sherry, N.
EPI_ISL_426835 hCoV-19/Australia/VIC576/2020 Oceania / Australia / Victoria 2020-03-26 Victorian Infectious Diseases Reference Laboratory (VIDRL) Microbiological Diagnostic Unit Public Health Laboratory and Victorian Infectious Diseases Reference Laboratory, Doherty Institute Caly L., Seemann T., Sait, M., Schultz M., Druce J., Sherry, N.
EPI_ISL_426836 hCoV-19/Australia/VIC578/2020 Oceania / Australia / Victoria 2020-03-26 Victorian Infectious Diseases Reference Laboratory (VIDRL) Microbiological Diagnostic Unit Public Health Laboratory and Victorian Infectious Diseases Reference Laboratory, Doherty Institute Caly L., Seemann T., Sait, M., Schultz M., Druce J., Sherry, N.
EPI_ISL_426839 hCoV-19/Australia/VIC582/2020 Oceania / Australia / Victoria 2020-03-25 Victorian Infectious Diseases Reference Laboratory (VIDRL) Microbiological Diagnostic Unit Public Health Laboratory and Victorian Infectious Diseases Reference Laboratory, Doherty Institute Caly L., Seemann T., Sait, M., Schultz M., Druce J., Sherry, N.
EPI_ISL_426840 hCoV-19/Australia/VIC583/2020 Oceania / Australia / Victoria 2020-03-25 Victorian Infectious Diseases Reference Laboratory (VIDRL) Microbiological Diagnostic Unit Public Health Laboratory and Victorian Infectious Diseases Reference Laboratory, Doherty Institute Caly L., Seemann T., Sait, M., Schultz M., Druce J., Sherry, N.
EPI_ISL_426841 hCoV-19/Australia/VIC584/2020 Oceania / Australia / Victoria 2020-03-25 Victorian Infectious Diseases Reference Laboratory (VIDRL) Microbiological Diagnostic Unit Public Health Laboratory and Victorian Infectious Diseases Reference Laboratory, Doherty Institute Caly L., Seemann T., Sait, M., Schultz M., Druce J., Sherry, N.
EPI_ISL_426842 hCoV-19/Australia/VIC585/2020 Oceania / Australia / Victoria 2020-03-25 Victorian Infectious Diseases Reference Laboratory (VIDRL) Microbiological Diagnostic Unit Public Health Laboratory and Victorian Infectious Diseases Reference Laboratory, Doherty Institute Caly L., Seemann T., Sait, M., Schultz M., Druce J., Sherry, N.
EPI_ISL_426844 hCoV-19/Australia/VIC587/2020 Oceania / Australia / Victoria 2020-03-25 Victorian Infectious Diseases Reference Laboratory (VIDRL) Microbiological Diagnostic Unit Public Health

Laboratory and Victorian Infectious Diseases Reference Laboratory, Doherty Institute    Caly L., Seemann T., Sait, M., Schultz M., Druce J., Sherry, N.
EPI_ISL_426846  hCoV-19/Australia/VIC589/2020   Oceania / Australia / Victoria  2020-03-22      Victorian Infectious Diseases Reference Laboratory (VIDRL)        Microbiological Diagnostic Unit Public Health Laboratory and Victorian Infectious Diseases Reference Laboratory, Doherty Institute    Caly L., Seemann T., Sait, M., Schultz M., Druce J., Sherry, N.
EPI_ISL_426847  hCoV-19/Australia/VIC590/2020   Oceania / Australia / Victoria  2020-03-22      Victorian Infectious Diseases Reference Laboratory (VIDRL)        Microbiological Diagnostic Unit Public Health Laboratory and Victorian Infectious Diseases Reference Laboratory, Doherty Institute    Caly L., Seemann T., Sait, M., Schultz M., Druce J., Sherry, N.
EPI_ISL_426848  hCoV-19/Australia/VIC591/2020   Oceania / Australia / Victoria  2020-03-22      Victorian Infectious Diseases Reference Laboratory (VIDRL)        Microbiological Diagnostic Unit Public Health Laboratory and Victorian Infectious Diseases Reference Laboratory, Doherty Institute    Caly L., Seemann T., Sait, M., Schultz M., Druce J., Sherry, N.
EPI_ISL_426850  hCoV-19/Australia/VIC593/2020   Oceania / Australia / Victoria  2020-03-23      Victorian Infectious Diseases Reference Laboratory (VIDRL)        Microbiological Diagnostic Unit Public Health Laboratory and Victorian Infectious Diseases Reference Laboratory, Doherty Institute    Caly L., Seemann T., Sait, M., Schultz M., Druce J., Sherry, N.
EPI_ISL_426851  hCoV-19/Australia/VIC594/2020   Oceania / Australia / Victoria  2020-03-25      Victorian Infectious Diseases Reference Laboratory (VIDRL)        Microbiological Diagnostic Unit Public Health Laboratory and Victorian Infectious Diseases Reference Laboratory, Doherty Institute    Caly L., Seemann T., Sait, M., Schultz M., Druce J., Sherry, N.
EPI_ISL_426852  hCoV-19/Australia/VIC595/2020   Oceania / Australia / Victoria  2020-03-25      Victorian Infectious Diseases Reference Laboratory (VIDRL)        Microbiological Diagnostic Unit Public Health Laboratory and Victorian Infectious Diseases Reference Laboratory, Doherty Institute    Caly L., Seemann T., Sait, M., Schultz M., Druce J., Sherry, N.
EPI_ISL_426853  hCoV-19/Australia/VIC596/2020   Oceania / Australia / Victoria  2020-03-25      Victorian Infectious Diseases Reference Laboratory (VIDRL)        Microbiological Diagnostic Unit Public Health Laboratory and Victorian Infectious Diseases Reference Laboratory, Doherty Institute    Caly L., Seemann T., Sait, M., Schultz M., Druce J., Sherry, N.
EPI_ISL_426854  hCoV-19/Australia/VIC597/2020   Oceania / Australia / Victoria  2020-03-25      Victorian Infectious Diseases Reference Laboratory (VIDRL)        Microbiological Diagnostic Unit Public Health Laboratory and Victorian Infectious Diseases Reference Laboratory, Doherty Institute    Caly L., Seemann T., Sait, M., Schultz M., Druce J., Sherry, N.
EPI_ISL_426855  hCoV-19/Australia/VIC598/2020   Oceania / Australia / Victoria  2020-03-25      Victorian Infectious Diseases Reference Laboratory (VIDRL)        Microbiological Diagnostic Unit Public Health Laboratory and Victorian Infectious Diseases Reference Laboratory, Doherty Institute    Caly L., Seemann T., Sait, M., Schultz M., Druce J., Sherry, N.
EPI_ISL_426856  hCoV-19/Australia/VIC599/2020   Oceania / Australia / Victoria  2020-03-25      Victorian Infectious Diseases Reference Laboratory (VIDRL)        Microbiological Diagnostic Unit Public Health Laboratory and Victorian Infectious Diseases Reference Laboratory, Doherty Institute    Caly L., Seemann T., Sait, M., Schultz M., Druce J., Sherry, N.
EPI_ISL_426858  hCoV-19/Australia/VIC603/2020   Oceania / Australia / Victoria  2020-03-26      Victorian Infectious Diseases Reference Laboratory (VIDRL)        Microbiological Diagnostic Unit Public Health Laboratory and Victorian Infectious Diseases Reference Laboratory, Doherty Institute    Caly L., Seemann T., Sait, M., Schultz M., Druce J., Sherry, N.
EPI_ISL_426860  hCoV-19/Australia/VIC605/2020   Oceania / Australia / Victoria  2020-03-27      Victorian Infectious Diseases Reference Laboratory (VIDRL)        Microbiological Diagnostic Unit Public Health Laboratory and Victorian Infectious Diseases Reference Laboratory, Doherty Institute    Caly L., Seemann T., Sait, M., Schultz M., Druce J., Sherry, N.
EPI_ISL_426861  hCoV-19/Australia/VIC606/2020   Oceania / Australia / Victoria  2020-03-27      Victorian Infectious Diseases Reference Laboratory (VIDRL)        Microbiological Diagnostic Unit Public Health Laboratory and Victorian Infectious Diseases Reference Laboratory, Doherty Institute    Caly L., Seemann T., Sait, M., Schultz M., Druce J., Sherry, N.
EPI_ISL_426862  hCoV-19/Australia/VIC607/2020   Oceania / Australia / Victoria  2020-03-27      Victorian Infectious Diseases Reference Laboratory (VIDRL)        Microbiological Diagnostic Unit Public Health Laboratory and Victorian Infectious Diseases Reference Laboratory, Doherty Institute    Caly L., Seemann T., Sait, M., Schultz M., Druce J., Sherry, N.
EPI_ISL_426864  hCoV-19/Australia/VIC610/2020   Oceania / Australia / Victoria  2020-03-26      Victorian Infectious Diseases Reference Laboratory (VIDRL)        Microbiological Diagnostic Unit Public Health Laboratory and Victorian Infectious Diseases Reference Laboratory, Doherty Institute    Caly L., Seemann T., Sait, M., Schultz M., Druce J., Sherry, N.
EPI_ISL_426865  hCoV-19/Australia/VIC611/2020   Oceania / Australia / Victoria  2020-03-27      Victorian Infectious Diseases Reference Laboratory (VIDRL)        Microbiological Diagnostic Unit Public Health Laboratory and Victorian Infectious Diseases Reference Laboratory, Doherty Institute    Caly L., Seemann T., Sait, M., Schultz M., Druce J., Sherry, N.
EPI_ISL_426866  hCoV-19/Australia/VIC612/2020   Oceania / Australia / Victoria  2020-03-26      Victorian Infectious Diseases Reference Laboratory (VIDRL)        Microbiological Diagnostic Unit Public Health Laboratory and Victorian Infectious Diseases Reference Laboratory, Doherty Institute    Caly L., Seemann T., Sait, M., Schultz M., Druce J., Sherry, N.

EPI_ISL_426870 hCoV-19/Australia/VIC615/2020 Oceania / Australia / Victoria 2020-03-28 Victorian Infectious Diseases Reference Laboratory (VIDRL) Microbiological Diagnostic Unit Public Health Laboratory and Victorian Infectious Diseases Reference Laboratory, Doherty Institute Caly L., Seemann T., Sait, M., Schultz M., Druce J., Sherry, N.
EPI_ISL_426872 hCoV-19/Australia/VIC617/2020 Oceania / Australia / Victoria 2020-03-26 Victorian Infectious Diseases Reference Laboratory (VIDRL) Microbiological Diagnostic Unit Public Health Laboratory and Victorian Infectious Diseases Reference Laboratory, Doherty Institute Caly L., Seemann T., Sait, M., Schultz M., Druce J., Sherry, N.
EPI_ISL_426873 hCoV-19/Australia/VIC629/2020 Oceania / Australia / Victoria 2020-03-25 Victorian Infectious Diseases Reference Laboratory (VIDRL) Microbiological Diagnostic Unit Public Health Laboratory and Victorian Infectious Diseases Reference Laboratory, Doherty Institute Caly L., Seemann T., Sait, M., Schultz M., Druce J., Sherry, N.
EPI_ISL_426874 hCoV-19/Australia/VIC630/2020 Oceania / Australia / Victoria 2020-03-26 Victorian Infectious Diseases Reference Laboratory (VIDRL) Microbiological Diagnostic Unit Public Health Laboratory and Victorian Infectious Diseases Reference Laboratory, Doherty Institute Caly L., Seemann T., Sait, M., Schultz M., Druce J., Sherry, N.
EPI_ISL_426875 hCoV-19/Australia/VIC633/2020 Oceania / Australia / Victoria 2020-03-27 Victorian Infectious Diseases Reference Laboratory (VIDRL) Microbiological Diagnostic Unit Public Health Laboratory and Victorian Infectious Diseases Reference Laboratory, Doherty Institute Caly L., Seemann T., Sait, M., Schultz M., Druce J., Sherry, N.
EPI_ISL_426876 hCoV-19/Australia/VIC635/2020 Oceania / Australia / Victoria 2020-03-28 Victorian Infectious Diseases Reference Laboratory (VIDRL) Microbiological Diagnostic Unit Public Health Laboratory and Victorian Infectious Diseases Reference Laboratory, Doherty Institute Caly L., Seemann T., Sait, M., Schultz M., Druce J., Sherry, N.
EPI_ISL_426877 hCoV-19/Australia/VIC636/2020 Oceania / Australia / Victoria 2020-03-28 Victorian Infectious Diseases Reference Laboratory (VIDRL) Microbiological Diagnostic Unit Public Health Laboratory and Victorian Infectious Diseases Reference Laboratory, Doherty Institute Caly L., Seemann T., Sait, M., Schultz M., Druce J., Sherry, N.
EPI_ISL_426878 hCoV-19/Australia/VIC701/2020 Oceania / Australia / Victoria 2020-03-21 Victorian Infectious Diseases Reference Laboratory (VIDRL) Microbiological Diagnostic Unit Public Health Laboratory and Victorian Infectious Diseases Reference Laboratory, Doherty Institute Caly L., Seemann T., Sait, M., Schultz M., Druce J., Sherry, N.
EPI_ISL_426881 hCoV-19/Australia/VIC912/2020 Oceania / Australia / Victoria 2020-03-24 Victorian Infectious Diseases Reference Laboratory (VIDRL) Microbiological Diagnostic Unit Public Health Laboratory and Victorian Infectious Diseases Reference Laboratory, Doherty Institute Caly L., Seemann T., Sait, M., Schultz M., Druce J., Sherry, N.
EPI_ISL_426882 hCoV-19/Australia/VIC913/2020 Oceania / Australia / Victoria 2020-03-24 Victorian Infectious Diseases Reference Laboratory (VIDRL) Microbiological Diagnostic Unit Public Health Laboratory and Victorian Infectious Diseases Reference Laboratory, Doherty Institute Caly L., Seemann T., Sait, M., Schultz M., Druce J., Sherry, N.
EPI_ISL_426883 hCoV-19/Czech Republic/IAB_1/2020 Europe / Czech Republic 2020-03-27 Motol University Hospital Institute of Applied Biotechnologies a.s. Petr Brož, Jan Geryk, Petr Klempt, Martin Kašný, Adam Novotný, Kateřina Kvapilová, Pavel Dřevínek, Petr Kvapil, Milan Macek
EPI_ISL_426884 hCoV-19/Czech Republic/IAB_4/2020 Europe / Czech Republic 2020-03-27 Motol University Hospital Institute of Applied Biotechnologies a.s. Petr Brož, Jan Geryk, Petr Klempt, Martin Kašný, Adam Novotný, Kateřina Kvapilová, Pavel Dřevínek, Petr Kvapil, Milan Macek
EPI_ISL_426885 hCoV-19/Czech Republic/IAB_8/2020 Europe / Czech Republic 2020-03-27 Motol University Hospital Institute of Applied Biotechnologies a.s. Petr Brož, Jan Geryk, Petr Klempt, Martin Kašný, Adam Novotný, Kateřina Kvapilová, Pavel Dřevínek, Petr Kvapil, Milan Macek
EPI_ISL_426886 hCoV-19/Czech Republic/IAB_9/2020 Europe / Czech Republic 2020-03-28 Motol University Hospital Institute of Applied Biotechnologies a.s. Petr Brož, Jan Geryk, Petr Klempt, Martin Kašný, Adam Novotný, Kateřina Kvapilová, Pavel Dřevínek, Petr Kvapil, Milan Macek
EPI_ISL_426887 hCoV-19/Czech Republic/IAB_10/2020 Europe / Czech Republic 2020-03-31 Motol University Hospital Institute of Applied Biotechnologies a.s. Petr Brož, Jan Geryk, Petr Klempt, Martin Kašný, Adam Novotný, Kateřina Kvapilová, Pavel Dřevínek, Petr Kvapil, Milan Macek
EPI_ISL_426888 hCoV-19/Czech Republic/IAB_12/2020 Europe / Czech Republic 2020-03-31 Motol University Hospital Institute of Applied Biotechnologies a.s. Petr Brož, Jan Geryk, Petr Klempt, Martin Kašný, Adam Novotný, Kateřina Kvapilová, Pavel Dřevínek, Petr Kvapil, Milan Macek
EPI_ISL_426889 hCoV-19/Czech Republic/IAB_14/2020 Europe / Czech Republic 2020-04-02 Motol University Hospital Institute of Applied Biotechnologies a.s. Petr Brož, Jan Geryk, Petr Klempt, Martin Kašný, Adam Novotný, Kateřina Kvapilová, Pavel Dřevínek, Petr Kvapil, Milan Macek
EPI_ISL_426890 hCoV-19/Czech Republic/IAB_15/2020 Europe / Czech Republic 2020-03-26 Motol University Hospital Institute of Applied Biotechnologies a.s. Petr Brož, Jan Geryk, Petr Klempt, Martin Kašný, Adam Novotný, Kateřina Kvapilová, Pavel Dřevínek, Petr Kvapil, Milan Macek
EPI_ISL_426891 hCoV-19/Czech Republic/IAB_16/2020 Europe / Czech Republic 2020-03-26 Motol University Hospital Institute of Applied Biotechnologies a.s. Petr Brož, Jan Geryk, Petr Klempt, Martin Kašný, Adam Novotný, Kateřina Kvapilová, Pavel Dřevínek, Petr Kvapil, Milan Macek
EPI_ISL_426892 hCoV-19/Czech Republic/IAB_17/2020 Europe / Czech Republic 2020-03-27 Motol University Hospital Institute of Applied Biotechnologies a.s. Petr Brož, Jan Geryk, Petr Klempt, Martin Kašný, Adam Novotný, Kateřina Kvapilová, Pavel Dřevínek, Petr Kvapil, Milan Macek

EPI_ISL_426893 hCoV-19/Czech Republic/IAB_18/2020 Europe / Czech Republic 2020-03-27 Motol University Hospital Institute of Applied Biotechnologies a.s. Petr Brož, Jan Geryk, Petr Klempt, Martin Kašný, Adam Novotný, Kateřina Kvapilová, Pavel Dřevínek, Petr Kvapil, Milan Macek
EPI_ISL_426894 hCoV-19/Czech Republic/IAB_20/2020 Europe / Czech Republic 2020-03-27 Motol University Hospital Institute of Applied Biotechnologies a.s. Petr Brož, Jan Geryk, Petr Klempt, Martin Kašný, Adam Novotný, Kateřina Kvapilová, Pavel Dřevínek, Petr Kvapil, Milan Macek
EPI_ISL_426895 hCoV-19/Czech Republic/IAB_21/2020 Europe / Czech Republic 2020-03-27 Motol University Hospital Institute of Applied Biotechnologies a.s. Petr Brož, Jan Geryk, Petr Klempt, Martin Kašný, Adam Novotný, Kateřina Kvapilová, Pavel Dřevínek, Petr Kvapil, Milan Macek
EPI_ISL_426896 hCoV-19/Czech Republic/IAB_22/2020 Europe / Czech Republic 2020-03-28 Motol University Hospital Institute of Applied Biotechnologies a.s. Petr Brož, Jan Geryk, Petr Klempt, Martin Kašný, Adam Novotný, Kateřina Kvapilová, Pavel Dřevínek, Petr Kvapil, Milan Macek
EPI_ISL_426897 hCoV-19/Czech Republic/IAB_23/2020 Europe / Czech Republic 2020-03-29 Motol University Hospital Institute of Applied Biotechnologies a.s. Petr Brož, Jan Geryk, Petr Klempt, Martin Kašný, Adam Novotný, Kateřina Kvapilová, Pavel Dřevínek, Petr Kvapil, Milan Macek
EPI_ISL_426898 hCoV-19/Australia/NT10/2020 Oceania / Australia / Northern territory 2020-03-26 Royal Darwin Hospital Pathology Microbiological Diagnostic Unit Public Health Laboratory and Victorian Infectious Diseases Reference Laboratory, Doherty Institute Meumann, E., Caly L., Seemann T., Sait, M., Schultz M., Druce J., Sherry, N.
EPI_ISL_426899 hCoV-19/Australia/NT11/2020 Oceania / Australia / Northern territory 2020-03-27 Royal Darwin Hospital Pathology Microbiological Diagnostic Unit Public Health Laboratory and Victorian Infectious Diseases Reference Laboratory, Doherty Institute Meumann, E., Caly L., Seemann T., Sait, M., Schultz M., Druce J., Sherry, N.
EPI_ISL_426900 hCoV-19/Australia/NT12/2020 Oceania / Australia / Northern territory 2020-03-25 Royal Darwin Hospital Pathology Microbiological Diagnostic Unit Public Health Laboratory and Victorian Infectious Diseases Reference Laboratory, Doherty Institute Meumann, E., Caly L., Seemann T., Sait, M., Schultz M., Druce J., Sherry, N.
EPI_ISL_426901 hCoV-19/Australia/NT13/2020 Oceania / Australia / Northern territory 2020-03-25 Royal Darwin Hospital Pathology Microbiological Diagnostic Unit Public Health Laboratory and Victorian Infectious Diseases Reference Laboratory, Doherty Institute Meumann, E., Caly L., Seemann T., Sait, M., Schultz M., Druce J., Sherry, N.
EPI_ISL_426904 hCoV-19/Australia/NT17/2020 Oceania / Australia / Northern territory 2020-03-24 Royal Darwin Hospital Pathology Microbiological Diagnostic Unit Public Health Laboratory and Victorian Infectious Diseases Reference Laboratory, Doherty Institute Meumann, E., Caly L., Seemann T., Sait, M., Schultz M., Druce J., Sherry, N.
EPI_ISL_426906 hCoV-19/Australia/VIC473/2020 Oceania / Australia / Victoria 2020-03-28 Microbiological Diagnostic Unit Public Health Laboratory Microbiological Diagnostic Unit Public Health Laboratory Seemann T., Schultz M., Sait, M., Sherry, N.
EPI_ISL_426907 hCoV-19/Australia/VIC475/2020 Oceania / Australia / Victoria 2020-03-28 Microbiological Diagnostic Unit Public Health Laboratory Microbiological Diagnostic Unit Public Health Laboratory Seemann T., Schultz M., Sait, M., Sherry, N.
EPI_ISL_426910 hCoV-19/Australia/VIC478/2020 Oceania / Australia / Victoria 2020-03-28 Microbiological Diagnostic Unit Public Health Laboratory Microbiological Diagnostic Unit Public Health Laboratory Seemann T., Schultz M., Sait, M., Sherry, N.
EPI_ISL_429074 hCoV-19/Guangzhou/GZMU0003/2020 Asia / China / Guangzhou 2020-02-07 The First Affiliated Hospital of Guangzhou Medical University BGI-shenzhen & The First Affiliated Hospital of Guangzhou Medical University Yanqun Wang, Daxi Wang, Lu Zhang, Wanying Sun, Zhaoyong Zhang et al.
EPI_ISL_429075 hCoV-19/Guangzhou/GZMU0008/2020 Asia / China / Guangzhou 2020-02-07 The First Affiliated Hospital of Guangzhou Medical University BGI-shenzhen & The First Affiliated Hospital of Guangzhou Medical University Yanqun Wang, Daxi Wang, Lu Zhang, Wanying Sun, Zhaoyong Zhang et al.
EPI_ISL_429076 hCoV-19/Guangzhou/GZMU0009/2020 Asia / China / Guangzhou 2020-02-07 The First Affiliated Hospital of Guangzhou Medical University BGI-shenzhen & The First Affiliated Hospital of Guangzhou Medical University
EPI_ISL_429077 hCoV-19/Guangzhou/GZMU0010/2020 Asia / China / Guangzhou 2020-02-07 The First Affiliated Hospital of Guangzhou Medical University BGI-shenzhen & The First Affiliated Hospital of Guangzhou Medical University Yanqun Wang, Daxi Wang, Lu Zhang, Wanying Sun, Zhaoyong Zhang et al.
EPI_ISL_429078 hCoV-19/Guangzhou/GZMU0013/2020 Asia / China / Guangzhou 2020-02-07 The First Affiliated Hospital of Guangzhou Medical University BGI-shenzhen & The First Affiliated Hospital of Guangzhou Medical University
EPI_ISL_429079 hCoV-19/Guangzhou/GZMU0017/2020 Asia / China / Guangzhou 2020-02-05 The First Affiliated Hospital of Guangzhou Medical University BGI-shenzhen & The First Affiliated Hospital of Guangzhou Medical University
EPI_ISL_429080 hCoV-19/Guangzhou/GZMU0019/2020 Asia / China / Guangzhou 2020-02-05 The First Affiliated Hospital of Guangzhou Medical University BGI-shenzhen & The First Affiliated Hospital of Guangzhou Medical University
EPI_ISL_429081 hCoV-19/Guangzhou/GZMU0020/2020 Asia / China / Guangzhou 2020-02-05 The First Affiliated Hospital of Guangzhou Medical University BGI-shenzhen & The First Affiliated Hospital of Guangzhou Medical University
EPI_ISL_429082 hCoV-19/Guangzhou/GZMU0023/2020 Asia / China / Guangzhou 2020-02-05 The First Affiliated Hospital of Guangzhou Medical University BGI-shenzhen & The First Affiliated Hospital of

Guangzhou Medical University	Yanqun Wang, Daxi Wang, Lu Zhang, Wanying Sun, Zhaoyong Zhang et al.
EPI_ISL_429083	hCoV-19/Guangzhou/GZMU0024/2020 Asia / China / Guangzhou	2020-02-05	The First Affiliated Hospital of Guangzhou Medical University	BGI-shenzhen & The First Affiliated Hospital of Guangzhou Medical University	Yanqun Wang, Daxi Wang, Lu Zhang, Wanying Sun, Zhaoyong Zhang et al.
EPI_ISL_429084	hCoV-19/Guangzhou/GZMU0025/2020 Asia / China / Guangzhou	2020-02-05	The First Affiliated Hospital of Guangzhou Medical University	BGI-shenzhen & The First Affiliated Hospital of Guangzhou Medical University
EPI_ISL_429085	hCoV-19/Guangzhou/GZMU0033/2020 Asia / China / Guangzhou	2020-02-01	The First Affiliated Hospital of Guangzhou Medical University	BGI-shenzhen & The First Affiliated Hospital of Guangzhou Medical University	Yanqun Wang, Daxi Wang, Lu Zhang, Wanying Sun, Zhaoyong Zhang et al.
EPI_ISL_429086	hCoV-19/Guangzhou/GZMU0034/2020 Asia / China / Guangzhou	2020-02-01	The First Affiliated Hospital of Guangzhou Medical University	BGI-shenzhen & The First Affiliated Hospital of Guangzhou Medical University
EPI_ISL_429088	hCoV-19/Guangzhou/GZMU0035/2020 Asia / China / Guangzhou	2020-02-01	The First Affiliated Hospital of Guangzhou Medical University	BGI-shenzhen & The First Affiliated Hospital of Guangzhou Medical University	Yanqun Wang, Daxi Wang, Lu Zhang, Wanying Sun, Zhaoyong Zhang et al.
EPI_ISL_429089	hCoV-19/Guangzhou/GZMU0037/2020 Asia / China / Guangzhou	2020-02-08	The First Affiliated Hospital of Guangzhou Medical University	BGI-shenzhen & The First Affiliated Hospital of Guangzhou Medical University	Yanqun Wang, Daxi Wang, Lu Zhang, Wanying Sun, Zhaoyong Zhang et al.
EPI_ISL_429090	hCoV-19/Guangzhou/GZMU0039/2020 Asia / China / Guangzhou	2020-02-08	The First Affiliated Hospital of Guangzhou Medical University	BGI-shenzhen & The First Affiliated Hospital of Guangzhou Medical University	Yanqun Wang, Daxi Wang, Lu Zhang, Wanying Sun, Zhaoyong Zhang et al.
EPI_ISL_429091	hCoV-19/Guangzhou/GZMU0041/2020 Asia / China / Guangzhou	2020-02-08	The First Affiliated Hospital of Guangzhou Medical University	BGI-shenzhen & The First Affiliated Hospital of Guangzhou Medical University	Yanqun Wang, Daxi Wang, Lu Zhang, Wanying Sun, Zhaoyong Zhang et al.
EPI_ISL_429092	hCoV-19/Guangzhou/GZMU0045/2020 Asia / China / Guangzhou	2020-02-08	The First Affiliated Hospital of Guangzhou Medical University	BGI-shenzhen & The First Affiliated Hospital of Guangzhou Medical University	Yanqun Wang, Daxi Wang, Lu Zhang, Wanying Sun, Zhaoyong Zhang et al.
EPI_ISL_429093	hCoV-19/Guangzhou/GZMU0046/2020 Asia / China / Guangzhou	2020-02-08	The First Affiliated Hospital of Guangzhou Medical University	BGI-shenzhen & The First Affiliated Hospital of Guangzhou Medical University	Yanqun Wang, Daxi Wang, Lu Zhang, Wanying Sun, Zhaoyong Zhang et al.
EPI_ISL_429094	hCoV-19/Guangzhou/GZMU0049/2020 Asia / China / Guangzhou	2020-02-08	The First Affiliated Hospital of Guangzhou Medical University	BGI-shenzhen & The First Affiliated Hospital of Guangzhou Medical University
EPI_ISL_429095	hCoV-19/Guangzhou/GZMU0050/2020 Asia / China / Guangzhou	2020-02-08	The First Affiliated Hospital of Guangzhou Medical University	BGI-shenzhen & The First Affiliated Hospital of Guangzhou Medical University
EPI_ISL_429096	hCoV-19/Guangzhou/GZMU0051/2020 Asia / China / Guangzhou	2020-01-29	The First Affiliated Hospital of Guangzhou Medical University	BGI-shenzhen & The First Affiliated Hospital of Guangzhou Medical University	Yanqun Wang, Daxi Wang, Lu Zhang, Wanying Sun, Zhaoyong Zhang et al.
EPI_ISL_429097	hCoV-19/Guangzhou/GZMU0052/2020 Asia / China / Guangzhou	2020-01-29	The First Affiliated Hospital of Guangzhou Medical University	BGI-shenzhen & The First Affiliated Hospital of Guangzhou Medical University	Yanqun Wang, Daxi Wang, Lu Zhang, Wanying Sun, Zhaoyong Zhang et al.
EPI_ISL_429098	hCoV-19/Guangzhou/GZMU0054/2020 Asia / China / Guangzhou	2020-01-29	The First Affiliated Hospital of Guangzhou Medical University	BGI-shenzhen & The First Affiliated Hospital of Guangzhou Medical University	Yanqun Wang, Daxi Wang, Lu Zhang, Wanying Sun, Zhaoyong Zhang et al.
EPI_ISL_429100	hCoV-19/Guangzhou/GZMU0057/2020 Asia / China / Guangzhou	2020-01-27	The First Affiliated Hospital of Guangzhou Medical University	BGI-shenzhen & The First Affiliated Hospital of Guangzhou Medical University
EPI_ISL_429101	hCoV-19/Guangzhou/GZMU0058/2020 Asia / China / Guangzhou	2020-01-27	The First Affiliated Hospital of Guangzhou Medical University	BGI-shenzhen & The First Affiliated Hospital of Guangzhou Medical University
EPI_ISL_429102	hCoV-19/Guangzhou/GZMU0059/2020 Asia / China / Guangzhou	2020-01-27	The First Affiliated Hospital of Guangzhou Medical University	BGI-shenzhen & The First Affiliated Hospital of Guangzhou Medical University
EPI_ISL_429103	hCoV-19/Guangzhou/GZMU0060/2020 Asia / China / Guangzhou	2020-02-09	The First Affiliated Hospital of Guangzhou Medical University	BGI-shenzhen & The First Affiliated Hospital of Guangzhou Medical University
EPI_ISL_429104	hCoV-19/Guangzhou/GZMU0036/2020 Asia / China / Guangzhou	2020-02-01	The First Affiliated Hospital of Guangzhou Medical University	BGI-shenzhen & The First Affiliated Hospital of Guangzhou Medical University
EPI_ISL_429105	hCoV-19/Guangzhou/GZMU0055/2020 Asia / China / Guangzhou	2020-01-29	The First Affiliated Hospital of Guangzhou Medical University	BGI-shenzhen & The First Affiliated Hospital of Guangzhou Medical University
EPI_ISL_429115	hCoV-19/Sweden/20-08142/2020	Europe / Sweden / Halland	2020-03-11	Klinisk mikrobiologi och vardhygien Halmstad	The Public Health Agency of Sweden	Arne Kotz, Olov Svartstrom, Maria Lind Karlberg, Anna-Malin Linde, Oskar Karlsson Lindsjo, Anna Risberg, Shaman Muradrasoli, Karin Tegmark-Wisell
EPI_ISL_429116	hCoV-19/Sweden/20-08143/2020	Europe / Sweden / Halland	2020-03-15	Klinisk mikrobiologi och vardhygien Halmstad	The Public Health Agency of Sweden	Arne Kotz, Olov Svartstrom,

Maria Lind Karlberg, Anna-Malin Linde, Oskar Karlsson Lindsjo, Anna Risberg, Shaman Muradrasoli, Karin Tegmark-Wisell
EPI_ISL_429117 hCoV-19/Sweden/20-08145/2020 Europe / Sweden / Halland 2020-03-11 Klinisk mikrobiologi och vardhygien Halmstad The Public Health Agency of Sweden Arne Kotz, Olov Svartstrom, Maria Lind Karlberg, Anna-Malin Linde, Oskar Karlsson Lindsjo, Anna Risberg, Shaman Muradrasoli, Karin Tegmark-Wisell
EPI_ISL_429118 hCoV-19/Sweden/20-08146/2020 Europe / Sweden / Halland 2020-03-11 Klinisk mikrobiologi och vardhygien Halmstad The Public Health Agency of Sweden Arne Kotz, Olov Svartstrom, Maria Lind Karlberg, Anna-Malin Linde, Oskar Karlsson Lindsjo, Anna Risberg, Shaman Muradrasoli, Karin Tegmark-Wisell
EPI_ISL_429119 hCoV-19/Sweden/20-08148/2020 Europe / Sweden / Halland 2020-03-11 Klinisk mikrobiologi och vardhygien Halmstad The Public Health Agency of Sweden Arne Kotz, Olov Svartstrom, Maria Lind Karlberg, Anna-Malin Linde, Oskar Karlsson Lindsjo, Anna Risberg, Shaman Muradrasoli, Karin Tegmark-Wisell
EPI_ISL_429120 hCoV-19/Sweden/20-08149/2020 Europe / Sweden / Halland 2020-03-11 Klinisk mikrobiologi och vardhygien Halmstad The Public Health Agency of Sweden Arne Kotz, Olov Svartstrom, Maria Lind Karlberg, Anna-Malin Linde, Oskar Karlsson Lindsjo, Anna Risberg, Shaman Muradrasoli, Karin Tegmark-Wisell
EPI_ISL_429121 hCoV-19/Sweden/20-08151/2020 Europe / Sweden / Halland 2020-03-11 Klinisk mikrobiologi och vardhygien Halmstad The Public Health Agency of Sweden Arne Kotz, Olov Svartstrom, Maria Lind Karlberg, Anna-Malin Linde, Oskar Karlsson Lindsjo, Anna Risberg, Shaman Muradrasoli, Karin Tegmark-Wisell
EPI_ISL_429122 hCoV-19/Sweden/20-08155/2020 Europe / Sweden / Halland 2020-03-11 Klinisk mikrobiologi och vardhygien Halmstad The Public Health Agency of Sweden Arne Kotz, Olov Svartstrom, Maria Lind Karlberg, Anna-Malin Linde, Oskar Karlsson Lindsjo, Anna Risberg, Shaman Muradrasoli, Karin Tegmark-Wisell
EPI_ISL_429123 hCoV-19/Sweden/20-08169/2020 Europe / Sweden / Halland 2020-03-12 Klinisk mikrobiologi och vardhygien Halmstad The Public Health Agency of Sweden Arne Kotz, Olov Svartstrom, Maria Lind Karlberg, Anna-Malin Linde, Oskar Karlsson Lindsjo, Anna Risberg, Shaman Muradrasoli, Karin Tegmark-Wisell
EPI_ISL_429124 hCoV-19/Sweden/20-08170/2020 Europe / Sweden / Halland 2020-03-12 Klinisk mikrobiologi och vardhygien Halmstad The Public Health Agency of Sweden Arne Kotz, Olov Svartstrom, Maria Lind Karlberg, Anna-Malin Linde, Oskar Karlsson Lindsjo, Anna Risberg, Shaman Muradrasoli, Karin Tegmark-Wisell
EPI_ISL_429125 hCoV-19/Sweden/20-08173/2020 Europe / Sweden / Halland 2020-03-12 Klinisk mikrobiologi och vardhygien Halmstad The Public Health Agency of Sweden Arne Kotz, Olov Svartstrom, Maria Lind Karlberg, Anna-Malin Linde, Oskar Karlsson Lindsjo, Anna Risberg, Shaman Muradrasoli, Karin Tegmark-Wisell
EPI_ISL_429126 hCoV-19/Sweden/20-08320/2020 Europe / Sweden / Sormland 2020-03-21 Unilabs Skovde The Public Health Agency of Sweden Tobias Kollberg, Helena Enroth, Olov Svartstrom, Maria Lind Karlberg, Anna-Malin Linde, Oskar Karlsson Lindsjo, Anna Risberg, Shaman Muradrasoli, Karin Tegmark-Wisell
EPI_ISL_429127 hCoV-19/Sweden/20-08332/2020 Europe / Sweden / Sormland 2020-03-20 Unilabs Skovde The Public Health Agency of Sweden Tobias Kollberg, Helena Enroth, Olov Svartstrom, Maria Lind Karlberg, Anna-Malin Linde, Oskar Karlsson Lindsjo, Anna Risberg, Shaman Muradrasoli, Karin Tegmark-Wisell
EPI_ISL_429129 hCoV-19/Sweden/20-08681/2020 Europe / Sweden / Norrbotten 2020-03-27 Laboratoriemedicin The Public Health Agency of Sweden Olov Svartstrom, Maria Lind Karlberg, Anna-Malin Linde, Oskar Karlsson Lindsjo, Anna Risberg, Shaman Muradrasoli, Karin Tegmark-Wisell
EPI_ISL_429130 hCoV-19/Sweden/20-08683/2020 Europe / Sweden / Norrbotten 2020-03-23 Laboratoriemedicin The Public Health Agency of Sweden Olov Svartstrom, Maria Lind Karlberg, Anna-Malin Linde, Oskar Karlsson Lindsjo, Anna Risberg, Shaman Muradrasoli, Karin Tegmark-Wisell
EPI_ISL_429131 hCoV-19/Sweden/20-08684/2020 Europe / Sweden / Norrbotten 2020-03-27 Laboratoriemedicin The Public Health Agency of Sweden Olov Svartstrom, Maria Lind Karlberg, Anna-Malin Linde, Oskar Karlsson Lindsjo, Anna Risberg, Shaman Muradrasoli, Karin Tegmark-Wisell
EPI_ISL_429132 hCoV-19/Sweden/20-08685/2020 Europe / Sweden / Norrbotten 2020-03-27 Laboratoriemedicin The Public Health Agency of Sweden Olov Svartstrom, Maria Lind Karlberg, Anna-Malin Linde, Oskar Karlsson Lindsjo, Anna Risberg, Shaman Muradrasoli, Karin Tegmark-Wisell
EPI_ISL_429133 hCoV-19/Sweden/20-08690/2020 Europe / Sweden / Norrbotten 2020-03-26 Laboratoriemedicin The Public Health Agency of Sweden Olov Svartstrom, Maria Lind Karlberg, Anna-Malin Linde, Oskar Karlsson Lindsjo, Anna Risberg, Shaman Muradrasoli, Karin Tegmark-Wisell
EPI_ISL_429134 hCoV-19/Sweden/20-08693/2020 Europe / Sweden / Norrbotten 2020-03-24 Laboratoriemedicin The Public Health Agency of Sweden Olov Svartstrom, Maria Lind Karlberg, Anna-Malin Linde, Oskar Karlsson Lindsjo, Anna Risberg, Shaman Muradrasoli, Karin Tegmark-Wisell
EPI_ISL_429135 hCoV-19/Sweden/20-08719/2020 Europe / Sweden / Norrbotten 2020-04-02 Laboratoriemedicin The Public Health Agency of Sweden Olov Svartstrom, Maria Lind Karlberg, Anna-Malin Linde, Oskar Karlsson Lindsjo, Anna Risberg, Shaman Muradrasoli, Karin Tegmark-Wisell
EPI_ISL_429136 hCoV-19/Sweden/20-50048/2020 Europe / Sweden / Varmland 2020-03-04 Klinisk mikrobiologi Orebro The Public Health Agency of Sweden Martin Sundqvist, Olov Svartstrom, Maria Lind Karlberg, Anna-Malin Linde, Oskar Karlsson Lindsjo, Anna Risberg, Shaman Muradrasoli, Karin Tegmark-Wisell
EPI_ISL_429137 hCoV-19/Sweden/20-50076/2020 Europe / Sweden / Orebro lan 2020-03-04 Klinisk mikrobiologi Orebro The Public Health Agency of Sweden Martin Sundqvist, Olov Svartstrom, Maria Lind

Karlberg, Anna-Malin Linde, Oskar Karlsson Lindsjo, Anna Risberg, Shaman Muradrasoli, Karin Tegmark-Wisell
EPI_ISL_429138 hCoV-19/Sweden/20-50087/2020 Europe / Sweden / Varmland 2020-03-05 Klinisk mikrobiologi Orebro The Public Health Agency of Sweden Martin Sundqvist, Olov Svartstrom, Maria Lind Karlberg, Anna-Malin Linde, Oskar Karlsson Lindsjo, Anna Risberg, Shaman Muradrasoli, Karin Tegmark-Wisell
EPI_ISL_429139 hCoV-19/Sweden/20-50123/2020 Europe / Sweden / Varmland 2020-03-06 Klinisk mikrobiologi Orebro The Public Health Agency of Sweden Martin Sundqvist, Olov Svartstrom, Maria Lind Karlberg, Anna-Malin Linde, Oskar Karlsson Lindsjo, Anna Risberg, Shaman Muradrasoli, Karin Tegmark-Wisell
EPI_ISL_429140 hCoV-19/Sweden/20-50125/2020 Europe / Sweden / Varmland 2020-03-06 Klinisk mikrobiologi Orebro The Public Health Agency of Sweden Martin Sundqvist, Olov Svartstrom, Maria Lind Karlberg, Anna-Malin Linde, Oskar Karlsson Lindsjo, Anna Risberg, Shaman Muradrasoli, Karin Tegmark-Wisell
EPI_ISL_429141 hCoV-19/Sweden/20-50130/2020 Europe / Sweden / Varmland 2020-03-06 Klinisk mikrobiologi Orebro The Public Health Agency of Sweden Martin Sundqvist, Olov Svartstrom, Maria Lind Karlberg, Anna-Malin Linde, Oskar Karlsson Lindsjo, Anna Risberg, Shaman Muradrasoli, Karin Tegmark-Wisell
EPI_ISL_429142 hCoV-19/Sweden/20-50131/2020 Europe / Sweden / Varmland 2020-03-06 Klinisk mikrobiologi Orebro The Public Health Agency of Sweden Martin Sundqvist, Olov Svartstrom, Maria Lind Karlberg, Anna-Malin Linde, Oskar Karlsson Lindsjo, Anna Risberg, Shaman Muradrasoli, Karin Tegmark-Wisell
EPI_ISL_429143 hCoV-19/Sweden/20-50132/2020 Europe / Sweden / Varmland 2020-03-06 Klinisk mikrobiologi Orebro The Public Health Agency of Sweden Martin Sundqvist, Olov Svartstrom, Maria Lind Karlberg, Anna-Malin Linde, Oskar Karlsson Lindsjo, Anna Risberg, Shaman Muradrasoli, Karin Tegmark-Wisell
EPI_ISL_429144 hCoV-19/Sweden/20-50133/2020 Europe / Sweden / Varmland 2020-03-06 Klinisk mikrobiologi Orebro The Public Health Agency of Sweden Martin Sundqvist, Olov Svartstrom, Maria Lind Karlberg, Anna-Malin Linde, Oskar Karlsson Lindsjo, Anna Risberg, Shaman Muradrasoli, Karin Tegmark-Wisell
EPI_ISL_429145 hCoV-19/Sweden/20-50170/2020 Europe / Sweden / Varmland 2020-03-08 Klinisk mikrobiologi Orebro The Public Health Agency of Sweden Martin Sundqvist, Olov Svartstrom, Maria Lind Karlberg, Anna-Malin Linde, Oskar Karlsson Lindsjo, Anna Risberg, Shaman Muradrasoli, Karin Tegmark-Wisell
EPI_ISL_429146 hCoV-19/Sweden/20-50171/2020 Europe / Sweden / Varmland 2020-03-08 Klinisk mikrobiologi Orebro The Public Health Agency of Sweden Martin Sundqvist, Olov Svartstrom, Maria Lind Karlberg, Anna-Malin Linde, Oskar Karlsson Lindsjo, Anna Risberg, Shaman Muradrasoli, Karin Tegmark-Wisell
EPI_ISL_429147 hCoV-19/Sweden/20-50172/2020 Europe / Sweden / Varmland 2020-03-08 Klinisk mikrobiologi Orebro The Public Health Agency of Sweden Martin Sundqvist, Olov Svartstrom, Maria Lind Karlberg, Anna-Malin Linde, Oskar Karlsson Lindsjo, Anna Risberg, Shaman Muradrasoli, Karin Tegmark-Wisell
EPI_ISL_429148 hCoV-19/Sweden/20-50173/2020 Europe / Sweden / Varmland 2020-03-08 Klinisk mikrobiologi Orebro The Public Health Agency of Sweden Martin Sundqvist, Olov Svartstrom, Maria Lind Karlberg, Anna-Malin Linde, Oskar Karlsson Lindsjo, Anna Risberg, Shaman Muradrasoli, Karin Tegmark-Wisell
EPI_ISL_429150 hCoV-19/Sweden/20-50175/2020 Europe / Sweden / Varmland 2020-03-08 Klinisk mikrobiologi Orebro The Public Health Agency of Sweden Martin Sundqvist, Olov Svartstrom, Maria Lind Karlberg, Anna-Malin Linde, Oskar Karlsson Lindsjo, Anna Risberg, Shaman Muradrasoli, Karin Tegmark-Wisell
EPI_ISL_429151 hCoV-19/Sweden/20-50176/2020 Europe / Sweden / Varmland 2020-03-08 Klinisk mikrobiologi Orebro The Public Health Agency of Sweden Martin Sundqvist, Olov Svartstrom, Maria Lind Karlberg, Anna-Malin Linde, Oskar Karlsson Lindsjo, Anna Risberg, Shaman Muradrasoli, Karin Tegmark-Wisell
EPI_ISL_429152 hCoV-19/Sweden/20-50179/2020 Europe / Sweden / Orebro lan 2020-03-08 Klinisk mikrobiologi Orebro The Public Health Agency of Sweden Martin Sundqvist, Olov Svartstrom, Maria Lind Karlberg, Anna-Malin Linde, Oskar Karlsson Lindsjo, Anna Risberg, Shaman Muradrasoli, Karin Tegmark-Wisell
EPI_ISL_429153 hCoV-19/Sweden/20-50180/2020 Europe / Sweden / Varmland 2020-03-08 Klinisk mikrobiologi Orebro The Public Health Agency of Sweden Martin Sundqvist, Olov Svartstrom, Maria Lind Karlberg, Anna-Malin Linde, Oskar Karlsson Lindsjo, Anna Risberg, Shaman Muradrasoli, Karin Tegmark-Wisell
EPI_ISL_429154 hCoV-19/Sweden/20-50181/2020 Europe / Sweden / Varmland 2020-03-08 Klinisk mikrobiologi Orebro The Public Health Agency of Sweden Martin Sundqvist, Olov Svartstrom, Maria Lind Karlberg, Anna-Malin Linde, Oskar Karlsson Lindsjo, Anna Risberg, Shaman Muradrasoli, Karin Tegmark-Wisell
EPI_ISL_429155 hCoV-19/Sweden/20-50230/2020 Europe / Sweden / Orebro lan 2020-03-09 Klinisk mikrobiologi Orebro The Public Health Agency of Sweden Martin Sundqvist, Olov Svartstrom, Maria Lind Karlberg, Anna-Malin Linde, Oskar Karlsson Lindsjo, Anna Risberg, Shaman Muradrasoli, Karin Tegmark-Wisell
EPI_ISL_429156 hCoV-19/Sweden/20-50232/2020 Europe / Sweden / Varmland 2020-03-09 Klinisk mikrobiologi Orebro The Public Health Agency of Sweden Martin Sundqvist, Olov Svartstrom, Maria Lind Karlberg, Anna-Malin Linde, Oskar Karlsson Lindsjo, Anna Risberg, Shaman Muradrasoli, Karin Tegmark-Wisell
EPI_ISL_429157 hCoV-19/Sweden/20-50234/2020 Europe / Sweden / Orebro lan 2020-03-10 Klinisk mikrobiologi Orebro The Public Health Agency of Sweden Martin Sundqvist, Olov Svartstrom, Maria Lind Karlberg, Anna-Malin Linde, Oskar Karlsson Lindsjo, Anna Risberg, Shaman Muradrasoli, Karin Tegmark-Wisell
EPI_ISL_429158 hCoV-19/Sweden/20-50235/2020 Europe / Sweden / Varmland 2020-03-10 Klinisk mikrobiologi Orebro The Public Health Agency of Sweden Martin Sundqvist, Olov Svartstrom, Maria Lind Karlberg, Anna-Malin Linde, Oskar Karlsson Lindsjo, Anna Risberg, Shaman Muradrasoli, Karin Tegmark-Wisell
EPI_ISL_429160 hCoV-19/Sweden/20-50312/2020 Europe / Sweden / Ostergotland 2020-03-12 Klinisk mikrobiologi Orebro The Public Health Agency of Sweden Martin Sundqvist, Olov Svartstrom, Maria Lind Karlberg, Anna-Malin Linde, Oskar Karlsson Lindsjo, Anna Risberg, Shaman Muradrasoli, Karin Tegmark-Wisell
EPI_ISL_429161 hCoV-19/Sweden/20-50315/2020 Europe / Sweden / Stockholm 2020-03-12 The Public Health Agency of Sweden The Public Health Agency of Sweden Olov Svartstrom, Maria Lind Karlberg, Anna-Malin Linde, Oskar Karlsson Lindsjo, Anna Risberg, Shaman Muradrasoli, Karin Tegmark-Wisell
EPI_ISL_429162 hCoV-19/Sweden/20-50357/2020 Europe / Sweden / Stockholm 2020-03-15 The Public Health Agency of Sweden The Public Health Agency of Sweden Olov Svartstrom, Maria Lind Karlberg, Anna-Malin Linde, Oskar Karlsson Lindsjo, Anna Risberg, Shaman Muradrasoli, Karin Tegmark-Wisell

EPI_ISL_429163  hCoV-19/Sweden/20-50361/2020    Europe / Sweden / Gavleborgs lan        2020-03-16      The Public Health Agency of Sweden  The Public Health Agency of Sweden      Olov Svartstrom, Maria Lind Karlberg, Anna-Malin Linde, Oskar Karlsson Lindsjo, Anna Risberg, Shaman Muradrasoli, Karin Tegmark-Wisell
EPI_ISL_429164  hCoV-19/Thailand/Bangkok-0025/2020      Asia / Thailand / Bangkok       2020-03-13      Ramathibodi Hospital    COVID-19 Network Investigations (CONI) Alliance Elizabeth Batty, Wasun Chantratita, Thanat Chookajorn, Stefan Fernandez, Angkana Huang, Poramate Jiaranai, Anthony R. Jones, Khajohn Joonsalak, Chonticha Klungtong, Theerarat Kochakarn, Namfon Kotanan, Krittikorn Kumpornsin, Wudtichai Manasatienkij, Bhakbhoom Panthan, Ekawat Pasomsub, Kingkan Rakmanee, Insee Sensorn, Janjira Thaipadungpanit, Arporn Wangwiwatsin,Treewat Watthanachockchai
EPI_ISL_429165  hCoV-19/Thailand/Bangkok-0026/2020      Asia / Thailand / Bangkok       2020-03-13      Ramathibodi Hospital    COVID-19 Network Investigations (CONI) Alliance Elizabeth Batty, Wasun Chantratita, Thanat Chookajorn, Stefan Fernandez, Angkana Huang, Poramate Jiaranai, Anthony R. Jones, Khajohn Joonsalak, Chonticha Klungtong, Theerarat Kochakarn, Namfon Kotanan, Krittikorn Kumpornsin, Wudtichai Manasatienkij, Bhakbhoom Panthan, Ekawat Pasomsub, Kingkan Rakmanee, Insee Sensorn, Janjira Thaipadungpanit, Arporn Wangwiwatsin,Treewat Watthanachockchai
EPI_ISL_429166  hCoV-19/Thailand/Bangkok-0028/2020      Asia / Thailand / Bangkok       2020-03-13      Ramathibodi Hospital    COVID-19 Network Investigations (CONI) Alliance Elizabeth Batty, Wasun Chantratita, Thanat Chookajorn, Stefan Fernandez, Angkana Huang, Poramate Jiaranai, Anthony R. Jones, Khajohn Joonsalak, Chonticha Klungtong, Theerarat Kochakarn, Namfon Kotanan, Krittikorn Kumpornsin, Wudtichai Manasatienkij, Bhakbhoom Panthan, Ekawat Pasomsub, Kingkan Rakmanee, Insee Sensorn, Janjira Thaipadungpanit, Arporn Wangwiwatsin,Treewat Watthanachockchai
EPI_ISL_429167  hCoV-19/Thailand/Bangkok-0029/2020      Asia / Thailand / Bangkok       2020-03-13      Ramathibodi Hospital    COVID-19 Network Investigations (CONI) Alliance Elizabeth Batty, Wasun Chantratita, Thanat Chookajorn, Stefan Fernandez, Angkana Huang, Poramate Jiaranai, Anthony R. Jones, Khajohn Joonsalak, Chonticha Klungtong, Theerarat Kochakarn, Namfon Kotanan, Krittikorn Kumpornsin, Wudtichai Manasatienkij, Bhakbhoom Panthan, Ekawat Pasomsub, Kingkan Rakmanee, Insee Sensorn, Janjira Thaipadungpanit, Arporn Wangwiwatsin,Treewat Watthanachockchai
EPI_ISL_429169  hCoV-19/Thailand/Bangkok-0033/2020      Asia / Thailand / Bangkok       2020-03-17      Ramathibodi Hospital    COVID-19 Network Investigations (CONI) Alliance Elizabeth Batty, Wasun Chantratita, Thanat Chookajorn, Stefan Fernandez, Angkana Huang, Poramate Jiaranai, Anthony R. Jones, Khajohn Joonsalak, Chonticha Klungtong, Theerarat Kochakarn, Namfon Kotanan, Krittikorn Kumpornsin, Wudtichai Manasatienkij, Bhakbhoom Panthan, Ekawat Pasomsub, Kingkan Rakmanee, Insee Sensorn, Janjira Thaipadungpanit, Arporn Wangwiwatsin,Treewat Watthanachockchai
EPI_ISL_429170  hCoV-19/Thailand/Bangkok-0034/2020      Asia / Thailand / Bangkok       2020-03-17      Ramathibodi Hospital    COVID-19 Network Investigations (CONI) Alliance Elizabeth Batty, Wasun Chantratita, Thanat Chookajorn, Stefan Fernandez, Angkana Huang, Poramate Jiaranai, Anthony R. Jones, Khajohn Joonsalak, Chonticha Klungtong, Theerarat Kochakarn, Namfon Kotanan, Krittikorn Kumpornsin, Wudtichai Manasatienkij, Bhakbhoom Panthan, Ekawat Pasomsub, Kingkan Rakmanee, Insee Sensorn, Janjira Thaipadungpanit, Arporn Wangwiwatsin,Treewat Watthanachockchai
EPI_ISL_429171  hCoV-19/Thailand/Bangkok-0035/2020      Asia / Thailand / Bangkok       2020-03-18      Ramathibodi Hospital    COVID-19 Network Investigations (CONI) Alliance Elizabeth Batty, Wasun Chantratita, Thanat Chookajorn, Stefan Fernandez, Angkana Huang, Poramate Jiaranai, Anthony R. Jones, Khajohn Joonsalak, Chonticha Klungtong, Theerarat Kochakarn, Namfon Kotanan, Krittikorn Kumpornsin, Wudtichai Manasatienkij, Bhakbhoom Panthan, Ekawat Pasomsub, Kingkan Rakmanee, Insee Sensorn, Janjira Thaipadungpanit, Arporn Wangwiwatsin,Treewat Watthanachockchai
EPI_ISL_429172  hCoV-19/Thailand/Bangkok-0036/2020      Asia / Thailand / Bangkok       2020-03-21      Ramathibodi Hospital    COVID-19 Network Investigations (CONI) Alliance Elizabeth Batty, Wasun Chantratita, Thanat Chookajorn, Stefan Fernandez, Angkana Huang, Poramate Jiaranai, Anthony R. Jones, Khajohn Joonsalak, Chonticha Klungtong, Theerarat Kochakarn, Namfon Kotanan, Krittikorn Kumpornsin, Wudtichai Manasatienkij, Bhakbhoom Panthan, Ekawat Pasomsub, Kingkan Rakmanee, Insee Sensorn, Janjira Thaipadungpanit, Arporn Wangwiwatsin,Treewat Watthanachockchai
EPI_ISL_429173  hCoV-19/Thailand/Bangkok-0037/2020      Asia / Thailand / Bangkok       2020-03-21      Ramathibodi Hospital    COVID-19 Network Investigations (CONI) Alliance Elizabeth Batty, Wasun Chantratita, Thanat Chookajorn, Stefan Fernandez, Angkana Huang, Poramate Jiaranai, Anthony R. Jones, Khajohn Joonsalak, Chonticha Klungtong, Theerarat Kochakarn, Namfon Kotanan, Krittikorn Kumpornsin, Wudtichai Manasatienkij, Bhakbhoom Panthan, Ekawat Pasomsub, Kingkan Rakmanee, Insee Sensorn, Janjira Thaipadungpanit, Arporn Wangwiwatsin,Treewat Watthanachockchai
EPI_ISL_429174  hCoV-19/Thailand/Bangkok-0038/2020      Asia / Thailand / Bangkok       2020-03-25      Ramathibodi Hospital    COVID-19 Network Investigations (CONI) Alliance Elizabeth Batty, Wasun Chantratita, Thanat Chookajorn, Stefan Fernandez, Angkana Huang, Poramate Jiaranai, Anthony R. Jones, Khajohn Joonsalak, Chonticha Klungtong, Theerarat Kochakarn, Namfon Kotanan, Krittikorn Kumpornsin, Wudtichai Manasatienkij, Bhakbhoom Panthan, Ekawat Pasomsub, Kingkan Rakmanee, Insee Sensorn, Janjira Thaipadungpanit, Arporn Wangwiwatsin,Treewat Watthanachockchai
EPI_ISL_429175  hCoV-19/Thailand/Bangkok-0039/2020      Asia / Thailand / Bangkok       2020-03-28      Ramathibodi Hospital    COVID-19 Network Investigations (CONI) Alliance Elizabeth Batty, Wasun Chantratita, Thanat Chookajorn, Stefan Fernandez, Angkana Huang, Poramate Jiaranai, Anthony R. Jones, Khajohn Joonsalak, Chonticha Klungtong, Theerarat Kochakarn, Namfon Kotanan, Krittikorn Kumpornsin, Wudtichai Manasatienkij, Bhakbhoom Panthan, Ekawat Pasomsub, Kingkan Rakmanee, Insee Sensorn, Janjira Thaipadungpanit, Arporn Wangwiwatsin,Treewat Watthanachockchai
EPI_ISL_429178  hCoV-19/Thailand/Bangkok-0042/2020      Asia / Thailand / Bangkok       2020-03-28

Ramathibodi Hospital    COVID-19 Network Investigations (CONI) Alliance Elizabeth Batty, Wasun Chantratita, Thanat Chookajorn, Stefan Fernandez, Angkana Huang, Poramate Jiaranai, Anthony R. Jones, Khajohn Joonsalak, Chonticha Klungtong, Theerarat Kochakarn, Namfon Kotanan, Krittikorn Kumpornsin, Wudtichai Manasatienkij, Bhakbhoom Panthan, Ekawat Pasomsub, Kingkan Rakmanee, Insee Sensorn, Janjira Thaipadungpanit, Arporn Wangwiwatsin,Treewat Watthanachockchai
EPI_ISL_429179  hCoV-19/Thailand/Bangkok-0043/2020      Asia / Thailand / Bangkok       2020-03-28      Ramathibodi Hospital    COVID-19 Network Investigations (CONI) Alliance Elizabeth Batty, Wasun Chantratita, Thanat Chookajorn, Stefan Fernandez, Angkana Huang, Poramate Jiaranai, Anthony R. Jones, Khajohn Joonsalak, Chonticha Klungtong, Theerarat Kochakarn, Namfon Kotanan, Krittikorn Kumpornsin, Wudtichai Manasatienkij, Bhakbhoom Panthan, Ekawat Pasomsub, Kingkan Rakmanee, Insee Sensorn, Janjira Thaipadungpanit, Arporn Wangwiwatsin,Treewat Watthanachockchai
EPI_ISL_429181  hCoV-19/Thailand/Bangkok-0045/2020      Asia / Thailand / Bangkok       2020-03-28      Ramathibodi Hospital    COVID-19 Network Investigations (CONI) Alliance Elizabeth Batty, Wasun Chantratita, Thanat Chookajorn, Stefan Fernandez, Angkana Huang, Poramate Jiaranai, Anthony R. Jones, Khajohn Joonsalak, Chonticha Klungtong, Theerarat Kochakarn, Namfon Kotanan, Krittikorn Kumpornsin, Wudtichai Manasatienkij, Bhakbhoom Panthan, Ekawat Pasomsub, Kingkan Rakmanee, Insee Sensorn, Janjira Thaipadungpanit, Arporn Wangwiwatsin,Treewat Watthanachockchai
EPI_ISL_429182  hCoV-19/Thailand/Bangkok-0046/2020      Asia / Thailand / Bangkok       2020-03-28      Ramathibodi Hospital    COVID-19 Network Investigations (CONI) Alliance Elizabeth Batty, Wasun Chantratita, Thanat Chookajorn, Stefan Fernandez, Angkana Huang, Poramate Jiaranai, Anthony R. Jones, Khajohn Joonsalak, Chonticha Klungtong, Theerarat Kochakarn, Namfon Kotanan, Krittikorn Kumpornsin, Wudtichai Manasatienkij, Bhakbhoom Panthan, Ekawat Pasomsub, Kingkan Rakmanee, Insee Sensorn, Janjira Thaipadungpanit, Arporn Wangwiwatsin,Treewat Watthanachockchai
EPI_ISL_429183  hCoV-19/Thailand/Bangkok-0047/2020      Asia / Thailand / Bangkok       2020-03-28      Ramathibodi Hospital    COVID-19 Network Investigations (CONI) Alliance Elizabeth Batty, Wasun Chantratita, Thanat Chookajorn, Stefan Fernandez, Angkana Huang, Poramate Jiaranai, Anthony R. Jones, Khajohn Joonsalak, Chonticha Klungtong, Theerarat Kochakarn, Namfon Kotanan, Krittikorn Kumpornsin, Wudtichai Manasatienkij, Bhakbhoom Panthan, Ekawat Pasomsub, Kingkan Rakmanee, Insee Sensorn, Janjira Thaipadungpanit, Arporn Wangwiwatsin,Treewat Watthanachockchai
EPI_ISL_429184  hCoV-19/Thailand/Bangkok-0048/2020      Asia / Thailand / Bangkok       2020-03-28      Ramathibodi Hospital    COVID-19 Network Investigations (CONI) Alliance Elizabeth Batty, Wasun Chantratita, Thanat Chookajorn, Stefan Fernandez, Angkana Huang, Poramate Jiaranai, Anthony R. Jones, Khajohn Joonsalak, Chonticha Klungtong, Theerarat Kochakarn, Namfon Kotanan, Krittikorn Kumpornsin, Wudtichai Manasatienkij, Bhakbhoom Panthan, Ekawat Pasomsub, Kingkan Rakmanee, Insee Sensorn, Janjira Thaipadungpanit, Arporn Wangwiwatsin,Treewat Watthanachockchai
EPI_ISL_429231  hCoV-19/Italy/TE6571/2020       Europe / Italy / Abruzzo        2020-03-24      Ospedale Civile Giuseppe Mazzini        "Istituto Zooprofilattico Sperimentale dell'Abruzzo e Molise ""G. Caporale"""   Lorusso A, Marcacci M, Di Domenico M, Ancora M, Curini V, Mangone I, Rinaldi A, Di Pasquale A, Camma C, Puglia I, Savini G
EPI_ISL_429232  hCoV-19/Italy/TE6572/2020       Europe / Italy / Abruzzo        2020-03-24      Ospedale Civile Giuseppe Mazzini        "Istituto Zooprofilattico Sperimentale dell'Abruzzo e Molise ""G. Caporale"""   Lorusso A, Marcacci M, Di Domenico M, Ancora M, Curini V, Mangone I, Rinaldi A, Di Pasquale A, Camma C, Puglia I, Savini G
EPI_ISL_429235  hCoV-19/Italy/TE6580/2020       Europe / Italy / Abruzzo        2020-03-24      Ospedale Civile Giuseppe Mazzini        "Istituto Zooprofilattico Sperimentale dell'Abruzzo e Molise ""G. Caporale"""   Lorusso A, Marcacci M, Di Domenico M, Ancora M, Curini V, Mangone I, Rinaldi A, Di Pasquale A, Camma C, Puglia I, Savini G
EPI_ISL_429236  hCoV-19/Italy/TE7859/2020       Europe / Italy / Abruzzo        2020-03-28      Ospedale Civile S. Liberatore di Atri   "Istituto Zooprofilattico Sperimentale dell'Abruzzo e Molise ""G. Caporale"""   Lorusso A, Marcacci M, Di Domenico M, Ancora M, Curini V, Mangone I, Rinaldi A, Di Pasquale A, Camma C, Puglia I, Savini G
EPI_ISL_429239  hCoV-19/Yunnan/0306-466/2020    Asia / China / Yunnan   2020-03-06      Department of Clinical Laboratory, the First People's Hospital of Yunnan Province      Department of Clinical Laboratory, the First People's Hospital of Yunnan Province    Yi Sun,Ziqin Dian,Ya Xu,Guiqian Zhang,Xin Fan,Yu Zhang
EPI_ISL_429255  hCoV-19/DRC/1131/2020   Africa / Democratic Republic of the Congo / Kinshasa    2020-04-05      Viral Respiratory Lab, National Institute for Biomedical Research (INRB)        Pathogen Sequencing Lab, National Institute for Biomedical Research (INRB)       Placide Mbala-Kingebeni, Edith Nkwembe, Eddy Kinganda-Lusamaki, Amuri Aziza, Catherine Pratt, Matthias Pauthner, Josh Quick, Allison Black, James Hadfield, Trevor Bedford, Ian Goodfellow, Nick Loman, Kristian Andersen, Michael Wiley, Steve Ahuka-Mundeke, Jean-Jacques Muyembe Tamfum
EPI_ISL_429256  hCoV-19/Spain/Vircell001/2020   Europe / Spain / Madrid 2020-03-12      Health Sciences Technology Park, Avicena, 8, 18016 Granada. Spain       Sequencing and Bioinformatics Service FISABIO-Public Health  Joaquín Mendoza, Almudena Rojas, Pablo Mendoza
EPI_ISL_429262  hCoV-19/Denmark/ALAB-HH01/2020  Europe / Denmark / Unknown      2020-03-10      Department of Clinical Microbiology, Copenhagen University Hospital, Hvidovre, Kettegaard Alle 30, 2650 Hvidovre.     Albertsen lab, Department of Chemistry and Bioscience, Aalborg University, Denmark      Rasmus Kirkegaard
EPI_ISL_429263  hCoV-19/Denmark/ALAB-HH02/2020  Europe / Denmark / Unknown      2020-03-10      Department of Clinical Microbiology, Copenhagen University Hospital, Hvidovre, Kettegaard Alle 30, 2650 Hvidovre.     Albertsen lab, Department of Chemistry and Bioscience, Aalborg University, Denmark      Rasmus Kirkegaard
EPI_ISL_429264  hCoV-19/Denmark/ALAB-HH03/2020  Europe / Denmark / Unknown      2020-03-10      Department of

Clinical Microbiology, Copenhagen University Hospital, Hvidovre, Kettegaard Alle 30, 2650 Hvidovre.
Albertsen lab, Department of Chemistry and Bioscience, Aalborg University, Denmark Rasmus Kirkegaard
EPI_ISL_429265 hCoV-19/Denmark/ALAB-HH04/2020 Europe / Denmark / Unknown 2020-03-10 Department of Clinical Microbiology, Copenhagen University Hospital, Hvidovre, Kettegaard Alle 30, 2650 Hvidovre.
Albertsen lab, Department of Chemistry and Bioscience, Aalborg University, Denmark Rasmus Kirkegaard
EPI_ISL_429266 hCoV-19/Denmark/ALAB-HH05/2020 Europe / Denmark / Unknown 2020-03-10 Department of Clinical Microbiology, Copenhagen University Hospital, Hvidovre, Kettegaard Alle 30, 2650 Hvidovre.
Albertsen lab, Department of Chemistry and Bioscience, Aalborg University, Denmark Rasmus Kirkegaard
EPI_ISL_429267 hCoV-19/Denmark/ALAB-HH06/2020 Europe / Denmark / Unknown 2020-03-10 Department of Clinical Microbiology, Copenhagen University Hospital, Hvidovre, Kettegaard Alle 30, 2650 Hvidovre.
Albertsen lab, Department of Chemistry and Bioscience, Aalborg University, Denmark Rasmus Kirkegaard
EPI_ISL_429268 hCoV-19/Denmark/ALAB-HH07/2020 Europe / Denmark / Unknown 2020-03-10 Department of Clinical Microbiology, Copenhagen University Hospital, Hvidovre, Kettegaard Alle 30, 2650 Hvidovre.
Albertsen lab, Department of Chemistry and Bioscience, Aalborg University, Denmark Rasmus Kirkegaard
EPI_ISL_429269 hCoV-19/Denmark/ALAB-HH08/2020 Europe / Denmark / Unknown 2020-03-10 Department of Clinical Microbiology, Copenhagen University Hospital, Hvidovre, Kettegaard Alle 30, 2650 Hvidovre.
Albertsen lab, Department of Chemistry and Bioscience, Aalborg University, Denmark Rasmus Kirkegaard
EPI_ISL_429270 hCoV-19/Denmark/ALAB-HH11/2020 Europe / Denmark / Unknown 2020-03-10 Department of Clinical Microbiology, Copenhagen University Hospital, Hvidovre, Kettegaard Alle 30, 2650 Hvidovre.
Albertsen lab, Department of Chemistry and Bioscience, Aalborg University, Denmark Rasmus Kirkegaard
EPI_ISL_429271 hCoV-19/Denmark/ALAB-HH13/2020 Europe / Denmark / Unknown 2020-03-10 Department of Clinical Microbiology, Copenhagen University Hospital, Hvidovre, Kettegaard Alle 30, 2650 Hvidovre.
Albertsen lab, Department of Chemistry and Bioscience, Aalborg University, Denmark Rasmus Kirkegaard
EPI_ISL_429272 hCoV-19/Denmark/ALAB-HH14/2020 Europe / Denmark / Unknown 2020-03-10 Department of Clinical Microbiology, Copenhagen University Hospital, Hvidovre, Kettegaard Alle 30, 2650 Hvidovre.
Albertsen lab, Department of Chemistry and Bioscience, Aalborg University, Denmark Rasmus Kirkegaard
EPI_ISL_429273 hCoV-19/Denmark/ALAB-HH15/2020 Europe / Denmark / Unknown 2020-03-10 Department of Clinical Microbiology, Copenhagen University Hospital, Hvidovre, Kettegaard Alle 30, 2650 Hvidovre.
Albertsen lab, Department of Chemistry and Bioscience, Aalborg University, Denmark Rasmus Kirkegaard
EPI_ISL_429274 hCoV-19/Denmark/ALAB-HH16/2020 Europe / Denmark / Unknown 2020-03-10 Department of Clinical Microbiology, Copenhagen University Hospital, Hvidovre, Kettegaard Alle 30, 2650 Hvidovre.
Albertsen lab, Department of Chemistry and Bioscience, Aalborg University, Denmark Rasmus Kirkegaard
EPI_ISL_429275 hCoV-19/Denmark/ALAB-HH17/2020 Europe / Denmark / Unknown 2020-03-10 Department of Clinical Microbiology, Copenhagen University Hospital, Hvidovre, Kettegaard Alle 30, 2650 Hvidovre.
Albertsen lab, Department of Chemistry and Bioscience, Aalborg University, Denmark Rasmus Kirkegaard
EPI_ISL_429276 hCoV-19/Denmark/ALAB-HH18/2020 Europe / Denmark / Unknown 2020-03-10 Department of Clinical Microbiology, Copenhagen University Hospital, Hvidovre, Kettegaard Alle 30, 2650 Hvidovre.
Albertsen lab, Department of Chemistry and Bioscience, Aalborg University, Denmark Rasmus Kirkegaard
EPI_ISL_429277 hCoV-19/Denmark/ALAB-HH19/2020 Europe / Denmark / Unknown 2020-03-10 Department of Clinical Microbiology, Copenhagen University Hospital, Hvidovre, Kettegaard Alle 30, 2650 Hvidovre.
Albertsen lab, Department of Chemistry and Bioscience, Aalborg University, Denmark Rasmus Kirkegaard
EPI_ISL_429278 hCoV-19/Denmark/ALAB-HH20/2020 Europe / Denmark / Unknown 2020-03-10 Department of Clinical Microbiology, Copenhagen University Hospital, Hvidovre, Kettegaard Alle 30, 2650 Hvidovre.
Albertsen lab, Department of Chemistry and Bioscience, Aalborg University, Denmark Rasmus Kirkegaard
EPI_ISL_429279 hCoV-19/Denmark/ALAB-HH21/2020 Europe / Denmark / Unknown 2020-03-10 Department of Clinical Microbiology, Copenhagen University Hospital, Hvidovre, Kettegaard Alle 30, 2650 Hvidovre.
Albertsen lab, Department of Chemistry and Bioscience, Aalborg University, Denmark Rasmus Kirkegaard
EPI_ISL_429280 hCoV-19/Denmark/ALAB-HH22/2020 Europe / Denmark / Unknown 2020-03-10 Department of Clinical Microbiology, Copenhagen University Hospital, Hvidovre, Kettegaard Alle 30, 2650 Hvidovre.
Albertsen lab, Department of Chemistry and Bioscience, Aalborg University, Denmark Rasmus Kirkegaard
EPI_ISL_429281 hCoV-19/Denmark/ALAB-HH23/2020 Europe / Denmark / Unknown 2020-03-10 Department of Clinical Microbiology, Copenhagen University Hospital, Hvidovre, Kettegaard Alle 30, 2650 Hvidovre.
Albertsen lab, Department of Chemistry and Bioscience, Aalborg University, Denmark Rasmus Kirkegaard
EPI_ISL_429282 hCoV-19/Denmark/ALAB-HH24/2020 Europe / Denmark / Unknown 2020-03-11 Department of Clinical Microbiology, Copenhagen University Hospital, Hvidovre, Kettegaard Alle 30, 2650 Hvidovre.
Albertsen lab, Department of Chemistry and Bioscience, Aalborg University, Denmark Rasmus Kirkegaard
EPI_ISL_429283 hCoV-19/Denmark/ALAB-HH25/2020 Europe / Denmark / Unknown 2020-03-11 Department of Clinical Microbiology, Copenhagen University Hospital, Hvidovre, Kettegaard Alle 30, 2650 Hvidovre.
Albertsen lab, Department of Chemistry and Bioscience, Aalborg University, Denmark Rasmus Kirkegaard
EPI_ISL_429284 hCoV-19/Denmark/ALAB-HH26/2020 Europe / Denmark / Unknown 2020-03-11 Department of Clinical Microbiology, Copenhagen University Hospital, Hvidovre, Kettegaard Alle 30, 2650 Hvidovre.
Albertsen lab, Department of Chemistry and Bioscience, Aalborg University, Denmark Rasmus Kirkegaard
EPI_ISL_429285 hCoV-19/Denmark/ALAB-HH27/2020 Europe / Denmark / Unknown 2020-03-11 Department of Clinical Microbiology, Copenhagen University Hospital, Hvidovre, Kettegaard Alle 30, 2650 Hvidovre.
Albertsen lab, Department of Chemistry and Bioscience, Aalborg University, Denmark Rasmus Kirkegaard
EPI_ISL_429287 hCoV-19/Denmark/ALAB-HH29/2020 Europe / Denmark / Unknown 2020-03-11 Department of Clinical Microbiology, Copenhagen University Hospital, Hvidovre, Kettegaard Alle 30, 2650 Hvidovre.
Albertsen lab, Department of Chemistry and Bioscience, Aalborg University, Denmark Rasmus Kirkegaard
EPI_ISL_429289 hCoV-19/Denmark/ALAB-HH31/2020 Europe / Denmark / Unknown 2020-03-12 Department of Clinical Microbiology, Copenhagen University Hospital, Hvidovre, Kettegaard Alle 30, 2650 Hvidovre.

Albertsen lab, Department of Chemistry and Bioscience, Aalborg University, Denmark Rasmus Kirkegaard
EPI_ISL_429293 hCoV-19/Denmark/ALAB-HH37/2020 Europe / Denmark / Unknown 2020-03-13 Department of Clinical Microbiology, Copenhagen University Hospital, Hvidovre, Kettegaard Alle 30, 2650 Hvidovre. Albertsen lab, Department of Chemistry and Bioscience, Aalborg University, Denmark Rasmus Kirkegaard
EPI_ISL_429295 hCoV-19/Denmark/ALAB-HH39/2020 Europe / Denmark / Unknown 2020-03-13 Department of Clinical Microbiology, Copenhagen University Hospital, Hvidovre, Kettegaard Alle 30, 2650 Hvidovre. Albertsen lab, Department of Chemistry and Bioscience, Aalborg University, Denmark Rasmus Kirkegaard
EPI_ISL_429299 hCoV-19/Denmark/ALAB-HH43/2020 Europe / Denmark / Unknown 2020-03-14 Department of Clinical Microbiology, Copenhagen University Hospital, Hvidovre, Kettegaard Alle 30, 2650 Hvidovre. Albertsen lab, Department of Chemistry and Bioscience, Aalborg University, Denmark Rasmus Kirkegaard
EPI_ISL_430059 hCoV-19/USA/UT-0513/2020 North America / USA / Utah 2020-04-03 Utah Public Health Laboratory Utah Public Health Laboratory Erin Young, Kelly Oakeson
EPI_ISL_430061 hCoV-19/USA/UT-0519/2020 North America / USA / Utah 2020-04-03 Utah Public Health Laboratory Utah Public Health Laboratory Erin Young, Kelly Oakeson
EPI_ISL_430064 hCoV-19/Australia/VIC-CBA4/2020 Oceania / Australia / Victoria 2020-03-07 Geelong Centre for Emerging Infectious Diseases Geelong Centre for Emerging Infectious Diseases Chamings A., Bhatta T.R., Alexandersen S.
EPI_ISL_430065 hCoV-19/Australia/VIC-CBA5/2020 Oceania / Australia / Victoria 2020-04-08 Geelong Centre for Emerging Infectious Diseases Geelong Centre for Emerging Infectious Diseases Chamings A., Bhatta T.R., Alexandersen S.
EPI_ISL_430067 hCoV-19/Russia/StPetersburg-RII4705S/2020 Europe / Russia / St.Petersburg 2020-04-07 WHO National Influenza Centre Russian Federation WHO National Influenza Centre Russian Federation Andrey Komissarov, Artem Fadeev, Mariia Sergeeva, Anna Ivanova, Daria Danilenko
EPI_ISL_430068 hCoV-19/Russia/StPetersburg-RII4712V/2020 Europe / Russia / St.Petersburg 2020-04-07 WHO National Influenza Centre Russian Federation WHO National Influenza Centre Russian Federation Andrey Komissarov, Artem Fadeev, Mariia Sergeeva, Anna Ivanova, Daria Danilenko
EPI_ISL_430069 hCoV-19/Russia/StPetersburg-RII4780S/2020 Europe / Russia / St.Petersburg 2020-04-07 WHO National Influenza Centre Russian Federation WHO National Influenza Centre Russian Federation Andrey Komissarov, Artem Fadeev, Mariia Sergeeva, Anna Ivanova, Daria Danilenko
EPI_ISL_430070 hCoV-19/Russia/StPetersburg-RII4917S/2020 Europe / Russia / St.Petersburg 2020-04-07 WHO National Influenza Centre Russian Federation WHO National Influenza Centre Russian Federation Andrey Komissarov, Artem Fadeev, Mariia Sergeeva, Anna Ivanova, Daria Danilenko
EPI_ISL_430071 hCoV-19/Russia/StPetersburg-RII4936S/2020 Europe / Russia / St.Petersburg 2020-04-08 WHO National Influenza Centre Russian Federation WHO National Influenza Centre Russian Federation Andrey Komissarov, Artem Fadeev, Mariia Sergeeva, Anna Ivanova, Daria Danilenko
EPI_ISL_430072 hCoV-19/Russia/StPetersburg-RII4937S/2020 Europe / Russia / St.Petersburg 2020-04-08 WHO National Influenza Centre Russian Federation WHO National Influenza Centre Russian Federation Andrey Komissarov, Artem Fadeev, Mariia Sergeeva, Anna Ivanova, Daria Danilenko
EPI_ISL_430073 hCoV-19/Russia/StPetersburg-RII4938S/2020 Europe / Russia / St.Petersburg 2020-04-08 WHO National Influenza Centre Russian Federation WHO National Influenza Centre Russian Federation Andrey Komissarov, Artem Fadeev, Mariia Sergeeva, Anna Ivanova, Daria Danilenko
EPI_ISL_430074 hCoV-19/Russia/StPetersburg-RII4944S/2020 Europe / Russia / St.Petersburg 2020-04-08 WHO National Influenza Centre Russian Federation WHO National Influenza Centre Russian Federation Andrey Komissarov, Artem Fadeev, Mariia Sergeeva, Anna Ivanova, Daria Danilenko
EPI_ISL_430075 hCoV-19/Russia/StPetersburg-RII4949S/2020 Europe / Russia / St.Petersburg 2020-04-09 WHO National Influenza Centre Russian Federation WHO National Influenza Centre Russian Federation Andrey Komissarov, Artem Fadeev, Mariia Sergeeva, Anna Ivanova, Daria Danilenko
EPI_ISL_430076 hCoV-19/Russia/StPetersburg-RII4969S/2020 Europe / Russia / St.Petersburg 2020-04-10 WHO National Influenza Centre Russian Federation WHO National Influenza Centre Russian Federation Andrey Komissarov, Artem Fadeev, Mariia Sergeeva, Anna Ivanova, Daria Danilenko
EPI_ISL_430077 hCoV-19/Russia/StPetersburg-RII4970S/2020 Europe / Russia / St.Petersburg 2020-04-10 WHO National Influenza Centre Russian Federation WHO National Influenza Centre Russian Federation Andrey Komissarov, Artem Fadeev, Mariia Sergeeva, Anna Ivanova, Daria Danilenko
EPI_ISL_430078 hCoV-19/Russia/StPetersburg-RII4972S/2020 Europe / Russia / St.Petersburg 2020-04-10 WHO National Influenza Centre Russian Federation WHO National Influenza Centre Russian Federation Andrey Komissarov, Artem Fadeev, Mariia Sergeeva, Anna Ivanova, Daria Danilenko
EPI_ISL_430079 hCoV-19/Russia/StPetersburg-RII4980S/2020 Europe / Russia / St.Petersburg 2020-04-10 WHO National Influenza Centre Russian Federation WHO National Influenza Centre Russian Federation Andrey Komissarov, Artem Fadeev, Mariia Sergeeva, Anna Ivanova, Daria Danilenko
EPI_ISL_430080 hCoV-19/Russia/StPetersburg-RII4983S/2020 Europe / Russia / St.Petersburg 2020-04-10 WHO National Influenza Centre Russian Federation WHO National Influenza Centre Russian Federation Andrey Komissarov, Artem Fadeev, Mariia Sergeeva, Anna Ivanova, Daria Danilenko
EPI_ISL_430081 hCoV-19/Russia/StPetersburg-RII4984S/2020 Europe / Russia / St.Petersburg 2020-04-10 WHO National Influenza Centre Russian Federation WHO National Influenza Centre Russian Federation Andrey Komissarov, Artem Fadeev, Mariia Sergeeva, Anna Ivanova, Daria Danilenko
EPI_ISL_430082 hCoV-19/Russia/StPetersburg-RII4985S/2020 Europe / Russia / St.Petersburg 2020-04-10 WHO National Influenza Centre Russian Federation WHO National Influenza Centre Russian Federation Andrey Komissarov, Artem Fadeev, Mariia Sergeeva, Anna Ivanova, Daria Danilenko
EPI_ISL_430083 hCoV-19/Russia/StPetersburg-RII4988S/2020 Europe / Russia / St.Petersburg 2020-04-10 WHO National Influenza Centre Russian Federation WHO National Influenza Centre Russian Federation

Andrey Komissarov, Artem Fadeev, Mariia Sergeeva, Anna Ivanova, Daria Danilenko
EPI_ISL_430084 hCoV-19/Russia/StPetersburg-RII5017V/2020 Europe / Russia / St.Petersburg 2020-04-09
WHO National Influenza Centre Russian Federation WHO National Influenza Centre Russian Federation
Andrey Komissarov, Artem Fadeev, Mariia Sergeeva, Anna Ivanova, Daria Danilenko
EPI_ISL_430085 hCoV-19/Russia/StPetersburg-RII5029S/2020 Europe / Russia / St.Petersburg 2020-04-10
WHO National Influenza Centre Russian Federation WHO National Influenza Centre Russian Federation
Andrey Komissarov, Artem Fadeev, Mariia Sergeeva, Anna Ivanova, Daria Danilenko
EPI_ISL_430086 hCoV-19/Russia/StPetersburg-RII5033S/2020 Europe / Russia / St.Petersburg 2020-04-10
WHO National Influenza Centre Russian Federation WHO National Influenza Centre Russian Federation
Andrey Komissarov, Artem Fadeev, Mariia Sergeeva, Anna Ivanova, Daria Danilenko
EPI_ISL_430087 hCoV-19/Russia/StPetersburg-RII5047S/2020 Europe / Russia / St.Petersburg 2020-04-10
WHO National Influenza Centre Russian Federation WHO National Influenza Centre Russian Federation
Andrey Komissarov, Artem Fadeev, Mariia Sergeeva, Anna Ivanova, Daria Danilenko
EPI_ISL_430088 hCoV-19/Russia/StPetersburg-RII5169S/2020 Europe / Russia / St.Petersburg 2020-04-09
WHO National Influenza Centre Russian Federation WHO National Influenza Centre Russian Federation
Andrey Komissarov, Artem Fadeev, Mariia Sergeeva, Anna Ivanova, Daria Danilenko
EPI_ISL_430089 hCoV-19/Russia/StPetersburg-RII5170S/2020 Europe / Russia / St.Petersburg 2020-04-09
WHO National Influenza Centre Russian Federation WHO National Influenza Centre Russian Federation
Andrey Komissarov, Artem Fadeev, Mariia Sergeeva, Anna Ivanova, Daria Danilenko
EPI_ISL_430090 hCoV-19/Russia/StPetersburg-RII5244S/2020 Europe / Russia / St.Petersburg 2020-04-10
WHO National Influenza Centre Russian Federation WHO National Influenza Centre Russian Federation
Andrey Komissarov, Artem Fadeev, Mariia Sergeeva, Anna Ivanova, Daria Danilenko
EPI_ISL_430091 hCoV-19/Russia/StPetersburg-RII5247S/2020 Europe / Russia / St.Petersburg 2020-04-10
WHO National Influenza Centre Russian Federation WHO National Influenza Centre Russian Federation
Andrey Komissarov, Artem Fadeev, Mariia Sergeeva, Anna Ivanova, Daria Danilenko
EPI_ISL_430092 hCoV-19/Russia/StPetersburg-RII5633S/2020 Europe / Russia / St.Petersburg 2020-04-14
WHO National Influenza Centre Russian Federation WHO National Influenza Centre Russian Federation
Andrey Komissarov, Artem Fadeev, Mariia Sergeeva, Anna Ivanova, Daria Danilenko
EPI_ISL_430093 hCoV-19/Russia/StPetersburg-RII5634S/2020 Europe / Russia / St.Petersburg 2020-04-14
WHO National Influenza Centre Russian Federation WHO National Influenza Centre Russian Federation
Andrey Komissarov, Artem Fadeev, Mariia Sergeeva, Anna Ivanova, Daria Danilenko
EPI_ISL_430094 hCoV-19/Russia/StPetersburg-RII5635S/2020 Europe / Russia / St.Petersburg 2020-04-14
WHO National Influenza Centre Russian Federation WHO National Influenza Centre Russian Federation
Andrey Komissarov, Artem Fadeev, Mariia Sergeeva, Anna Ivanova, Daria Danilenko
EPI_ISL_430095 hCoV-19/Russia/StPetersburg-RII5642S/2020 Europe / Russia / St.Petersburg 2020-04-14
WHO National Influenza Centre Russian Federation WHO National Influenza Centre Russian Federation
Andrey Komissarov, Artem Fadeev, Mariia Sergeeva, Anna Ivanova, Daria Danilenko
EPI_ISL_430096 hCoV-19/Russia/StPetersburg-RII5643S/2020 Europe / Russia / St.Petersburg 2020-04-14
WHO National Influenza Centre Russian Federation WHO National Influenza Centre Russian Federation
Andrey Komissarov, Artem Fadeev, Mariia Sergeeva, Anna Ivanova, Daria Danilenko
EPI_ISL_430097 hCoV-19/Russia/StPetersburg-RII5644S/2020 Europe / Russia / St.Petersburg 2020-04-14
WHO National Influenza Centre Russian Federation WHO National Influenza Centre Russian Federation
Andrey Komissarov, Artem Fadeev, Mariia Sergeeva, Anna Ivanova, Daria Danilenko
EPI_ISL_430098 hCoV-19/Russia/StPetersburg-RII5654S/2020 Europe / Russia / St.Petersburg 2020-04-14
WHO National Influenza Centre Russian Federation WHO National Influenza Centre Russian Federation
Andrey Komissarov, Artem Fadeev, Mariia Sergeeva, Anna Ivanova, Daria Danilenko
EPI_ISL_430099 hCoV-19/Russia/StPetersburg-RII5942S/2020 Europe / Russia / St.Petersburg 2020-04-13
WHO National Influenza Centre Russian Federation WHO National Influenza Centre Russian Federation
Andrey Komissarov, Artem Fadeev, Mariia Sergeeva, Anna Ivanova, Daria Danilenko
EPI_ISL_430100 hCoV-19/Russia/StPetersburg-RII6027S/2020 Europe / Russia / St.Petersburg 2020-04-14
WHO National Influenza Centre Russian Federation WHO National Influenza Centre Russian Federation
Andrey Komissarov, Artem Fadeev, Mariia Sergeeva, Anna Ivanova, Daria Danilenko
EPI_ISL_430101 hCoV-19/Russia/StPetersburg-RII6031S/2020 Europe / Russia / St.Petersburg 2020-04-14
WHO National Influenza Centre Russian Federation WHO National Influenza Centre Russian Federation
Andrey Komissarov, Artem Fadeev, Mariia Sergeeva, Anna Ivanova, Daria Danilenko
EPI_ISL_430102 hCoV-19/Russia/StPetersburg-RII6039S/2020 Europe / Russia / St.Petersburg 2020-04-14
WHO National Influenza Centre Russian Federation WHO National Influenza Centre Russian Federation
Andrey Komissarov, Artem Fadeev, Mariia Sergeeva, Anna Ivanova, Daria Danilenko
EPI_ISL_430103 hCoV-19/Russia/StPetersburg-RII6043S/2020 Europe / Russia / St.Petersburg 2020-04-14
WHO National Influenza Centre Russian Federation WHO National Influenza Centre Russian Federation
Andrey Komissarov, Artem Fadeev, Mariia Sergeeva, Anna Ivanova, Daria Danilenko
EPI_ISL_430104 hCoV-19/Russia/StPetersburg-RII6053S/2020 Europe / Russia / St.Petersburg 2020-04-15
WHO National Influenza Centre Russian Federation WHO National Influenza Centre Russian Federation
Andrey Komissarov, Artem Fadeev, Mariia Sergeeva, Anna Ivanova, Daria Danilenko
EPI_ISL_430105 hCoV-19/Russia/StPetersburg-RII6055S/2020 Europe / Russia / St.Petersburg 2020-04-15
WHO National Influenza Centre Russian Federation WHO National Influenza Centre Russian Federation
Andrey Komissarov, Artem Fadeev, Mariia Sergeeva, Anna Ivanova, Daria Danilenko
EPI_ISL_430106 hCoV-19/Russia/StPetersburg-RII6056S/2020 Europe / Russia / St.Petersburg 2020-04-15
WHO National Influenza Centre Russian Federation WHO National Influenza Centre Russian Federation
Andrey Komissarov, Artem Fadeev, Mariia Sergeeva, Anna Ivanova, Daria Danilenko

EPI_ISL_430107 hCoV-19/Russia/StPetersburg-RII6060S/2020 Europe / Russia / St.Petersburg 2020-04-15 WHO National Influenza Centre Russian Federation WHO National Influenza Centre Russian Federation Andrey Komissarov, Artem Fadeev, Mariia Sergeeva, Anna Ivanova, Daria Danilenko
EPI_ISL_430108 hCoV-19/Russia/StPetersburg-RII6061S/2020 Europe / Russia / St.Petersburg 2020-04-15 WHO National Influenza Centre Russian Federation WHO National Influenza Centre Russian Federation Andrey Komissarov, Artem Fadeev, Mariia Sergeeva, Anna Ivanova, Daria Danilenko
EPI_ISL_430109 hCoV-19/Russia/StPetersburg-RII6063S/2020 Europe / Russia / St.Petersburg 2020-04-15 WHO National Influenza Centre Russian Federation WHO National Influenza Centre Russian Federation Andrey Komissarov, Artem Fadeev, Mariia Sergeeva, Anna Ivanova, Daria Danilenko
EPI_ISL_430110 hCoV-19/Russia/StPetersburg-RII6065S/2020 Europe / Russia / St.Petersburg 2020-04-15 WHO National Influenza Centre Russian Federation WHO National Influenza Centre Russian Federation Andrey Komissarov, Artem Fadeev, Mariia Sergeeva, Anna Ivanova, Daria Danilenko
EPI_ISL_430111 hCoV-19/Russia/StPetersburg-RII6086S/2020 Europe / Russia / St.Petersburg 2020-04-15 WHO National Influenza Centre Russian Federation WHO National Influenza Centre Russian Federation Andrey Komissarov, Artem Fadeev, Mariia Sergeeva, Anna Ivanova, Daria Danilenko
EPI_ISL_430112 hCoV-19/Russia/Ulan-Ude-RII4562V/2020 Europe / Russia / Buryat Republic / Ulan-Ude 2020-03-25 WHO National Influenza Centre Russian Federation WHO National Influenza Centre Russian Federation Andrey Komissarov, Artem Fadeev, Mariia Sergeeva, Anna Ivanova, Daria Danilenko
EPI_ISL_430113 hCoV-19/USA/WA-S259/2020 North America / USA / Washington / King County 2020-03-04 Seattle Flu Study Seattle Flu Study Chu et al
EPI_ISL_430114 hCoV-19/USA/WA-S260/2020 North America / USA / Washington / King County 2020-03-12 Seattle Flu Study Seattle Flu Study Chu et al
EPI_ISL_430115 hCoV-19/USA/WA-S261/2020 North America / USA / Washington 2020-03-30 Seattle Flu Study Seattle Flu Study Chu et al
EPI_ISL_430116 hCoV-19/USA/WA-S262/2020 North America / USA / Washington 2020-03-16 Seattle Flu Study Seattle Flu Study Chu et al
EPI_ISL_430117 hCoV-19/USA/WA-S263/2020 North America / USA / Washington / King County 2020-03-05 Seattle Flu Study Seattle Flu Study Chu et al
EPI_ISL_430118 hCoV-19/USA/WA-S264/2020 North America / USA / Washington / King County 2020-03-03 Seattle Flu Study Seattle Flu Study Chu et al
EPI_ISL_430119 hCoV-19/USA/WA-S265/2020 North America / USA / Washington / Snohomish County 2020-03-05 Seattle Flu Study Seattle Flu Study Chu et al
EPI_ISL_430121 hCoV-19/USA/WA-S267/2020 North America / USA / Washington / King County 2020-03-31 Seattle Flu Study Seattle Flu Study Chu et al
EPI_ISL_430122 hCoV-19/USA/WA-S268/2020 North America / USA / Washington / King County 2020-03-05 Seattle Flu Study Seattle Flu Study Chu et al
EPI_ISL_430123 hCoV-19/USA/WA-S269/2020 North America / USA / Washington / King County 2020-03-10 Seattle Flu Study Seattle Flu Study Chu et al
EPI_ISL_430124 hCoV-19/USA/WA-S270/2020 North America / USA / Washington / King County 2020-03-06 Seattle Flu Study Seattle Flu Study Chu et al
EPI_ISL_430125 hCoV-19/USA/WA-S271/2020 North America / USA / Washington / King County 2020-03-31 Seattle Flu Study Seattle Flu Study Chu et al
EPI_ISL_430126 hCoV-19/USA/WA-S272/2020 North America / USA / Washington / King County 2020-02-26 Seattle Flu Study Seattle Flu Study Chu et al
EPI_ISL_430127 hCoV-19/USA/WA-S273/2020 North America / USA / Washington / King County 2020-03-12 Seattle Flu Study Seattle Flu Study Chu et al
EPI_ISL_430128 hCoV-19/USA/WA-S274/2020 North America / USA / Washington / King County 2020-03-08 Seattle Flu Study Seattle Flu Study Chu et al
EPI_ISL_430129 hCoV-19/USA/WA-S275/2020 North America / USA / Washington / King County 2020-03-06 Seattle Flu Study Seattle Flu Study Chu et al
EPI_ISL_430130 hCoV-19/USA/WA-S276/2020 North America / USA / Washington 2020-04-01 Seattle Flu Study Seattle Flu Study Chu et al
EPI_ISL_430131 hCoV-19/USA/WA-S277/2020 North America / USA / Washington / King County 2020-02-22 Seattle Flu Study Seattle Flu Study Chu et al
EPI_ISL_430132 hCoV-19/USA/WA-S278/2020 North America / USA / Washington 2020-03-17 Seattle Flu Study Seattle Flu Study Chu et al
EPI_ISL_430133 hCoV-19/USA/WA-S279/2020 North America / USA / Washington / King County 2020-03-04 Seattle Flu Study Seattle Flu Study Chu et al
EPI_ISL_430134 hCoV-19/USA/WA-S280/2020 North America / USA / Washington / King County 2020-03-09 Seattle Flu Study Seattle Flu Study Chu et al
EPI_ISL_430135 hCoV-19/USA/WA-S281/2020 North America / USA / Washington / King County 2020-02-29 Seattle Flu Study Seattle Flu Study Chu et al
EPI_ISL_430136 hCoV-19/USA/WA-S282/2020 North America / USA / Washington / King County 2020-03-10 Seattle Flu Study Seattle Flu Study Chu et al
EPI_ISL_430137 hCoV-19/USA/WA-S283/2020 North America / USA / Washington / King County 2020-03-21 Seattle Flu Study Seattle Flu Study Chu et al
EPI_ISL_430140 hCoV-19/USA/WA-S286/2020 North America / USA / Washington / Pierce County 2020-03-15 Seattle Flu Study Seattle Flu Study Chu et al
EPI_ISL_430141 hCoV-19/USA/WA-S287/2020 North America / USA / Washington / King County 2020-03-06 Seattle Flu Study Seattle Flu Study Chu et al

EPI_ISL_430142 hCoV-19/USA/WA-S288/2020 North America / USA / Washington / King County 2020-03-06 Seattle Flu Study Seattle Flu Study Chu et al
EPI_ISL_430143 hCoV-19/USA/WA-S289/2020 North America / USA / Washington / King County 2020-03-23 Seattle Flu Study Seattle Flu Study Chu et al
EPI_ISL_430146 hCoV-19/USA/WA-S292/2020 North America / USA / Washington / King County 2020-03-05 Seattle Flu Study Seattle Flu Study Chu et al
EPI_ISL_430147 hCoV-19/USA/WA-S293/2020 North America / USA / Washington 2020-04-01 Seattle Flu Study Seattle Flu Study Chu et al
EPI_ISL_430148 hCoV-19/USA/WA-S294/2020 North America / USA / Washington / King County 2020-03-23 Seattle Flu Study Seattle Flu Study Chu et al
EPI_ISL_430149 hCoV-19/USA/WA-S295/2020 North America / USA / Washington / King County 2020-03-31 Seattle Flu Study Seattle Flu Study Chu et al
EPI_ISL_430150 hCoV-19/USA/WA-S296/2020 North America / USA / Washington / King County 2020-03-13 Seattle Flu Study Seattle Flu Study Chu et al
EPI_ISL_430151 hCoV-19/USA/WA-S297/2020 North America / USA / Washington / King County 2020-03-13 Seattle Flu Study Seattle Flu Study Chu et al
EPI_ISL_430152 hCoV-19/USA/WA-S298/2020 North America / USA / Washington / King County 2020-03-05 Seattle Flu Study Seattle Flu Study Chu et al
EPI_ISL_430153 hCoV-19/USA/WA-S299/2020 North America / USA / Washington / King County 2020-03-31 Seattle Flu Study Seattle Flu Study Chu et al
EPI_ISL_430154 hCoV-19/USA/WA-S300/2020 North America / USA / Washington / King County 2020-03-25 Seattle Flu Study Seattle Flu Study Chu et al
EPI_ISL_430155 hCoV-19/USA/WA-S301/2020 North America / USA / Washington / Pierce County 2020-03-14 Seattle Flu Study Seattle Flu Study Chu et al
EPI_ISL_430160 hCoV-19/USA/WA-S123/2020 North America / USA / Washington 2020-02-29 Washington State Department of Health Seattle Flu Study Chu et al
EPI_ISL_430161 hCoV-19/USA/WA-S124/2020 North America / USA / Washington / King County 2020-03-13 Washington State Department of Health Seattle Flu Study Chu et al
EPI_ISL_430162 hCoV-19/USA/WA-S125/2020 North America / USA / Washington / King County 2020-03-13 Washington State Department of Health Seattle Flu Study Chu et al
EPI_ISL_430163 hCoV-19/USA/WA-S126/2020 North America / USA / Washington / King County 2020-03-13 Washington State Department of Health Seattle Flu Study Chu et al
EPI_ISL_430164 hCoV-19/USA/WA-S127/2020 North America / USA / Washington / King County 2020-03-13 Washington State Department of Health Seattle Flu Study Chu et al
EPI_ISL_430165 hCoV-19/USA/WA-S128/2020 North America / USA / Washington / King County 2020-03-13 Washington State Department of Health Seattle Flu Study Chu et al
EPI_ISL_430166 hCoV-19/USA/WA-S129/2020 North America / USA / Washington / King County 2020-03-13 Washington State Department of Health Seattle Flu Study Chu et al
EPI_ISL_430167 hCoV-19/USA/WA-S130/2020 North America / USA / Washington / King County 2020-03-13 Washington State Department of Health Seattle Flu Study Chu et al
EPI_ISL_430168 hCoV-19/USA/WA-S131/2020 North America / USA / Washington / King County 2020-03-13 Washington State Department of Health Seattle Flu Study Chu et al
EPI_ISL_430169 hCoV-19/USA/WA-S132/2020 North America / USA / Washington / King County 2020-03-13 Washington State Department of Health Seattle Flu Study Chu et al
EPI_ISL_430170 hCoV-19/USA/WA-S133/2020 North America / USA / Washington / King County 2020-03-13 Washington State Department of Health Seattle Flu Study Chu et al
EPI_ISL_430171 hCoV-19/USA/WA-S134/2020 North America / USA / Washington / King County 2020-03-13 Washington State Department of Health Seattle Flu Study Chu et al
EPI_ISL_430172 hCoV-19/USA/WA-S135/2020 North America / USA / Washington / King County 2020-03-13 Washington State Department of Health Seattle Flu Study Chu et al
EPI_ISL_430173 hCoV-19/USA/WA-S136/2020 North America / USA / Washington / King County 2020-03-13 Washington State Department of Health Seattle Flu Study Chu et al
EPI_ISL_430174 hCoV-19/USA/WA-S137/2020 North America / USA / Washington / King County 2020-03-13 Washington State Department of Health Seattle Flu Study Chu et al
EPI_ISL_430175 hCoV-19/USA/WA-S138/2020 North America / USA / Washington / Snohomish County 2020-03-12 Washington State Department of Health Seattle Flu Study Chu et al
EPI_ISL_430176 hCoV-19/USA/WA-S139/2020 North America / USA / Washington / King County 2020-03-12 Washington State Department of Health Seattle Flu Study Chu et al
EPI_ISL_430177 hCoV-19/USA/WA-S140/2020 North America / USA / Washington / King County 2020-03-12 Washington State Department of Health Seattle Flu Study Chu et al
EPI_ISL_430178 hCoV-19/USA/WA-S141/2020 North America / USA / Washington / King County 2020-03-13 Washington State Department of Health Seattle Flu Study Chu et al
EPI_ISL_430212 hCoV-19/USA/WA-S174/2020 North America / USA / Washington / King County 2020-03-19 Washington State Department of Health Seattle Flu Study Chu et al
EPI_ISL_430213 hCoV-19/USA/WA-S175/2020 North America / USA / Washington / King County 2020-03-19 Washington State Department of Health Seattle Flu Study Chu et al
EPI_ISL_430214 hCoV-19/USA/WA-S176/2020 North America / USA / Washington / King County 2020-03-19 Washington State Department of Health Seattle Flu Study Chu et al
EPI_ISL_430215 hCoV-19/USA/WA-S177/2020 North America / USA / Washington / King County 2020-03-19 Washington State Department of Health Seattle Flu Study Chu et al

EPI_ISL_430216  hCoV-19/USA/WA-S178/2020  North America / USA / Washington / King County  2020-03-20
Washington State Department of Health  Seattle Flu Study  Chu et al
EPI_ISL_430217  hCoV-19/USA/WA-S179/2020  North America / USA / Washington / Whatcom County  2020-
03-20  Washington State Department of Health  Seattle Flu Study  Chu et al
EPI_ISL_430219  hCoV-19/USA/WA-S181/2020  North America / USA / Washington  2020-03-19
Washington State Department of Health  Seattle Flu Study  Chu et al
EPI_ISL_430220  hCoV-19/USA/WA-S182/2020  North America / USA / Washington / Whatcom County  2020-
03-20  Washington State Department of Health  Seattle Flu Study  Chu et al
EPI_ISL_430221  hCoV-19/USA/WA-S183/2020  North America / USA / Washington / King County  2020-03-19
Washington State Department of Health  Seattle Flu Study  Chu et al
EPI_ISL_430222  hCoV-19/USA/WA-S184/2020  North America / USA / Washington / King County  2020-03-19
Washington State Department of Health  Seattle Flu Study  Chu et al
EPI_ISL_430223  hCoV-19/USA/WA-S185/2020  North America / USA / Washington  2020-03-19
Washington State Department of Health  Seattle Flu Study  Chu et al
EPI_ISL_430224  hCoV-19/USA/WA-S186/2020  North America / USA / Washington / Whatcom County  2020-
03-20  Washington State Department of Health  Seattle Flu Study  Chu et al
EPI_ISL_430225  hCoV-19/USA/WA-S187/2020  North America / USA / Washington / King County  2020-03-20
Washington State Department of Health  Seattle Flu Study  Chu et al
EPI_ISL_430226  hCoV-19/USA/WA-S188/2020  North America / USA / Washington / Whatcom County  2020-
03-20  Washington State Department of Health  Seattle Flu Study  Chu et al
EPI_ISL_430227  hCoV-19/USA/WA-S189/2020  North America / USA / Washington / Whatcom County  2020-
03-20  Washington State Department of Health  Seattle Flu Study  Chu et al
EPI_ISL_430228  hCoV-19/USA/WA-S190/2020  North America / USA / Washington / King County  2020-03-23
Washington State Department of Health  Seattle Flu Study  Chu et al
EPI_ISL_430229  hCoV-19/USA/WA-S191/2020  North America / USA / Washington / Whatcom County  2020-
03-20  Washington State Department of Health  Seattle Flu Study  Chu et al
EPI_ISL_430230  hCoV-19/USA/WA-S192/2020  North America / USA / Washington / Whatcom County  2020-
03-20  Washington State Department of Health  Seattle Flu Study  Chu et al
EPI_ISL_430231  hCoV-19/USA/WA-S193/2020  North America / USA / Washington / Grant County 2020-03-19
Washington State Department of Health  Seattle Flu Study  Chu et al
EPI_ISL_430232  hCoV-19/USA/WA-S194/2020  North America / USA / Washington / King County  2020-03-20
Washington State Department of Health  Seattle Flu Study  Chu et al
EPI_ISL_430233  hCoV-19/USA/WA-S195/2020  North America / USA / Washington / Whatcom County  2020-
03-20  Washington State Department of Health  Seattle Flu Study  Chu et al
EPI_ISL_430234  hCoV-19/USA/WA-S196/2020  North America / USA / Washington / King County  2020-03-19
Washington State Department of Health  Seattle Flu Study  Chu et al
EPI_ISL_430235  hCoV-19/USA/WA-S197/2020  North America / USA / Washington / Whatcom County  2020-
03-20  Washington State Department of Health  Seattle Flu Study  Chu et al
EPI_ISL_430236  hCoV-19/USA/WA-S198/2020  North America / USA / Washington / Whatcom County  2020-
03-20  Washington State Department of Health  Seattle Flu Study  Chu et al
EPI_ISL_430237  hCoV-19/USA/WA-S199/2020  North America / USA / Washington / Whatcom County  2020-
03-20  Washington State Department of Health  Seattle Flu Study  Chu et al
EPI_ISL_430238  hCoV-19/USA/WA-S200/2020  North America / USA / Washington / Yakima County  2020-
03-20  Washington State Department of Health  Seattle Flu Study  Chu et al
EPI_ISL_430239  hCoV-19/USA/WA-S201/2020  North America / USA / Washington / King County  2020-03-20
Washington State Department of Health  Seattle Flu Study  Chu et al
EPI_ISL_430240  hCoV-19/USA/WA-S202/2020  North America / USA / Washington / Whatcom County  2020-
03-20  Washington State Department of Health  Seattle Flu Study  Chu et al
EPI_ISL_430241  hCoV-19/USA/WA-S203/2020  North America / USA / Washington / Whatcom County  2020-
03-20  Washington State Department of Health  Seattle Flu Study  Chu et al
EPI_ISL_430242  hCoV-19/USA/WA-S204/2020  North America / USA / Washington / Whatcom County  2020-
03-22  Washington State Department of Health  Seattle Flu Study  Chu et al
EPI_ISL_430243  hCoV-19/USA/WA-S205/2020  North America / USA / Washington / Whatcom County  2020-
03-20  Washington State Department of Health  Seattle Flu Study  Chu et al
EPI_ISL_430244  hCoV-19/USA/WA-S206/2020  North America / USA / Washington / Whatcom County  2020-
03-20  Washington State Department of Health  Seattle Flu Study  Chu et al
EPI_ISL_430245  hCoV-19/USA/WA-S207/2020  North America / USA / Washington  2020-03-20
Washington State Department of Health  Seattle Flu Study  Chu et al
EPI_ISL_430246  hCoV-19/USA/WA-S208/2020  North America / USA / Washington / Whatcom County  2020-
03-23  Washington State Department of Health  Seattle Flu Study  Chu et al
EPI_ISL_430247  hCoV-19/USA/WA-S209/2020  North America / USA / Washington / Whatcom County  2020-
03-23  Washington State Department of Health  Seattle Flu Study  Chu et al
EPI_ISL_430248  hCoV-19/USA/WA-S210/2020  North America / USA / Washington / King County  2020-03-19
Washington State Department of Health  Seattle Flu Study  Chu et al
EPI_ISL_430249  hCoV-19/USA/WA-S211/2020  North America / USA / Washington / King County  2020-03-20
Washington State Department of Health  Seattle Flu Study  Chu et al
EPI_ISL_430250  hCoV-19/USA/WA-S212/2020  North America / USA / Washington  2020-03-19
Washington State Department of Health  Seattle Flu Study  Chu et al
EPI_ISL_430252  hCoV-19/USA/WA-S214/2020  North America / USA / Washington / Unknown County  2020-
03-20  Washington State Department of Health  Seattle Flu Study  Chu et al

EPI_ISL_430253 hCoV-19/USA/WA-S215/2020 North America / USA / Washington 2020-03-20
Washington State Department of Health Seattle Flu Study Chu et al
EPI_ISL_430254 hCoV-19/USA/WA-S216/2020 North America / USA / Washington / Whatcom County 2020-
03-20 Washington State Department of Health Seattle Flu Study Chu et al
EPI_ISL_430255 hCoV-19/USA/WA-S217/2020 North America / USA / Washington / Whatcom County 2020-
03-20 Washington State Department of Health Seattle Flu Study Chu et al
EPI_ISL_430256 hCoV-19/USA/WA-S218/2020 North America / USA / Washington / Whatcom County 2020-
03-23 Washington State Department of Health Seattle Flu Study Chu et al
EPI_ISL_430257 hCoV-19/USA/WA-S219/2020 North America / USA / Washington / Pierce County 2020-
03-20 Washington State Department of Health Seattle Flu Study Chu et al
EPI_ISL_430258 hCoV-19/USA/WA-S220/2020 North America / USA / Washington / Whatcom County 2020-
03-20 Washington State Department of Health Seattle Flu Study Chu et al
EPI_ISL_430259 hCoV-19/USA/WA-S221/2020 North America / USA / Washington / King County 2020-03-19
Washington State Department of Health Seattle Flu Study Chu et al
EPI_ISL_430261 hCoV-19/USA/WA-S223/2020 North America / USA / Washington / Whatcom County 2020-
03-20 Washington State Department of Health Seattle Flu Study Chu et al
EPI_ISL_430262 hCoV-19/USA/WA-S224/2020 North America / USA / Washington / King County 2020-03-19
Washington State Department of Health Seattle Flu Study Chu et al
EPI_ISL_430263 hCoV-19/USA/WA-S225/2020 North America / USA / Washington / Whatcom County 2020-
03-20 Washington State Department of Health Seattle Flu Study Chu et al
EPI_ISL_430264 hCoV-19/USA/WA-S226/2020 North America / USA / Washington / Whatcom County 2020-
03-21 Washington State Department of Health Seattle Flu Study Chu et al
EPI_ISL_430265 hCoV-19/USA/WA-S227/2020 North America / USA / Washington 2020-03-20
Washington State Department of Health Seattle Flu Study Chu et al
EPI_ISL_430266 hCoV-19/USA/WA-S228/2020 North America / USA / Washington / Whatcom County 2020-
03-20 Washington State Department of Health Seattle Flu Study Chu et al
EPI_ISL_430267 hCoV-19/USA/WA-S229/2020 North America / USA / Washington / King County 2020-03-19
Washington State Department of Health Seattle Flu Study Chu et al
EPI_ISL_430268 hCoV-19/USA/WA-S230/2020 North America / USA / Washington / King County 2020-03-19
Washington State Department of Health Seattle Flu Study Chu et al
EPI_ISL_430269 hCoV-19/USA/WA-S231/2020 North America / USA / Washington / Whatcom County 2020-
03-20 Washington State Department of Health Seattle Flu Study Chu et al
EPI_ISL_430270 hCoV-19/USA/WA-S232/2020 North America / USA / Washington / Clark County 2020-03-08
Washington State Department of Health Seattle Flu Study Chu et al
EPI_ISL_430271 hCoV-19/USA/WA-S233/2020 North America / USA / Washington / Snohomish County 2020-
03-03 Washington State Department of Health Seattle Flu Study Chu et al
EPI_ISL_430272 hCoV-19/USA/WA-S234/2020 North America / USA / Washington / King County 2020-02-28
Washington State Department of Health Seattle Flu Study Chu et al
EPI_ISL_430273 hCoV-19/USA/WA-S235/2020 North America / USA / Washington / King County 2020-02-29
Washington State Department of Health Seattle Flu Study Chu et al
EPI_ISL_430274 hCoV-19/USA/WA-S236/2020 North America / USA / Washington / King County 2020-03-01
Washington State Department of Health Seattle Flu Study Chu et al
EPI_ISL_430275 hCoV-19/USA/WA-S237/2020 North America / USA / Washington 2020-03-06
Washington State Department of Health Seattle Flu Study Chu et al
EPI_ISL_430276 hCoV-19/USA/WA-S238/2020 North America / USA / Washington / King County 2020-03-02
Washington State Department of Health Seattle Flu Study Chu et al
EPI_ISL_430277 hCoV-19/USA/WA-S239/2020 North America / USA / Washington / King County 2020-03-07
Washington State Department of Health Seattle Flu Study Chu et al
EPI_ISL_430278 hCoV-19/USA/WA-S240/2020 North America / USA / Washington / King County 2020-03-06
Washington State Department of Health Seattle Flu Study Chu et al
EPI_ISL_430279 hCoV-19/USA/WA-S241/2020 North America / USA / Washington / King County 2020-03-04
Washington State Department of Health Seattle Flu Study Chu et al
EPI_ISL_430280 hCoV-19/USA/WA-S242/2020 North America / USA / Washington / King County 2020-03-01
Washington State Department of Health Seattle Flu Study Chu et al
EPI_ISL_430281 hCoV-19/USA/WA-S243/2020 North America / USA / Washington / King County 2020-02-29
Washington State Department of Health Seattle Flu Study Chu et al
EPI_ISL_430282 hCoV-19/USA/WA-S244/2020 North America / USA / Washington / Thurston County 2020-
03-11 Washington State Department of Health Seattle Flu Study Chu et al
EPI_ISL_430283 hCoV-19/USA/WA-S245/2020 North America / USA / Washington / Snohomish County 2020-
03-02 Washington State Department of Health Seattle Flu Study Chu et al
EPI_ISL_430285 hCoV-19/USA/WA-S247/2020 North America / USA / Washington / King County 2020-03-01
Washington State Department of Health Seattle Flu Study Chu et al
EPI_ISL_430286 hCoV-19/USA/WA-S248/2020 North America / USA / Washington / King County 2020-03-10
Washington State Department of Health Seattle Flu Study Chu et al
EPI_ISL_430287 hCoV-19/USA/WA-S249/2020 North America / USA / Washington / King County 2020-03-01
Washington State Department of Health Seattle Flu Study Chu et al
EPI_ISL_430288 hCoV-19/USA/WA-S250/2020 North America / USA / Washington / Snohomish County 2020-
03-03 Washington State Department of Health Seattle Flu Study Chu et al
EPI_ISL_430289 hCoV-19/USA/WA-S251/2020 North America / USA / Washington / Skagit County 2020-
03-11 Washington State Department of Health Seattle Flu Study Chu et al

EPI_ISL_430290 hCoV-19/USA/WA-S252/2020 North America / USA / Washington / King County 2020-03-02 Washington State Department of Health Seattle Flu Study Chu et al
EPI_ISL_430291 hCoV-19/USA/WA-S253/2020 North America / USA / Washington / Snohomish County 2020-03-07 Washington State Department of Health Seattle Flu Study Chu et al
EPI_ISL_430292 hCoV-19/USA/WA-S254/2020 North America / USA / Washington / Snohomish County 2020-02-24 Washington State Department of Health Seattle Flu Study Chu et al
EPI_ISL_430293 hCoV-19/USA/WA-S255/2020 North America / USA / Washington / Clark County 2020-03-08 Washington State Department of Health Seattle Flu Study Chu et al
EPI_ISL_430294 hCoV-19/USA/WA-S256/2020 North America / USA / Washington / King County 2020-03-02 Washington State Department of Health Seattle Flu Study Chu et al
EPI_ISL_430295 hCoV-19/USA/WA-S257/2020 North America / USA / Washington / King County 2020-02-25 Washington State Department of Health Seattle Flu Study Chu et al
EPI_ISL_430297 hCoV-19/South Africa/R02827/2020 Africa / South Africa / GP 2020-03-06 National Institute for Communicable Diseases of the National Health Laboratory Service National Institute for Communicable Diseases of the National Health Laboratory Service Allam M, Kwenda S, van Heusden P, Khumalo Z, Mohale T, Subramoney K, von Gottberg, A, Ismail A, Bhiman JN
EPI_ISL_450500 hCoV-19/Wuhan/OS52/2020 Asia / China / Hubei / Wuhan 2020-01-10 unknown CAS Key Laboratory of Special Pathogens and Biosafety and Center for Emerging Infectious Diseases Si,H., Zhu,Y., Lin,H., Xie,S., Shi,Z., Zhou,P.
EPI_ISL_450501 hCoV-19/Wuhan/YB012611/2020 Asia / China / Hubei / Wuhan 2020-01-26 unknown CAS Key Laboratory of Special Pathogens and Biosafety and Center for Emerging Infectious Diseases Si,H., Zhu,Y., Lin,H., Xie,S., Shi,Z., Zhou,P.
EPI_ISL_450502 hCoV-19/Wuhan/YB012605/2020 Asia / China / Hubei / Wuhan 2020-01-26 unknown CAS Key Laboratory of Special Pathogens and Biosafety and Center for Emerging Infectious Diseases Si,H., Zhu,Y., Lin,H., Xie,S., Shi,Z., Zhou,P.
EPI_ISL_450503 hCoV-19/Wuhan/YB012602/2020 Asia / China / Hubei / Wuhan 2020-01-26 unknown CAS Key Laboratory of Special Pathogens and Biosafety and Center for Emerging Infectious Diseases Si,H., Zhu,Y., Lin,H., Xie,S., Shi,Z., Zhou,P.
EPI_ISL_450504 hCoV-19/Wuhan/YB012506/2020 Asia / China / Hubei / Wuhan 2020-01-25 unknown CAS Key Laboratory of Special Pathogens and Biosafety and Center for Emerging Infectious Diseases Si,H., Zhu,Y., Lin,H., Xie,S., Shi,Z., Zhou,P.
EPI_ISL_450506 hCoV-19/Brazil/SP02cc/2020 South America / Brazil 2020-03-18 unknown Clinical Laboratory Malta,F., Amgarten,D., de Oliveira,D.B.L., Araujo,D.B., Machado,R.R.G., Santana,R.A.F., Mangueira,C.L.P., Durigon,E.L. and Pinho,J.R.R.
EPI_ISL_403928 hCoV-19/Wuhan/IPBCAMS-WH-05/2020 Asia / China / Hubei / Wuhan 2020-01-01 Institute of Pathogen Biology, Chinese Academy of Medical Sciences & Peking Union Medical College Institute of Pathogen Biology, Chinese Academy of Medical Sciences & Peking Union Medical College Lili Ren, Jianwei Wang, Qi Jin, Zichun Xiang, Zhiqiang Wu, Chao Wu, Yiwei Liu
EPI_ISL_403929 hCoV-19/Wuhan/IPBCAMS-WH-04/2019 Asia / China / Hubei / Wuhan 2019-12-30 Institute of Pathogen Biology, Chinese Academy of Medical Sciences & Peking Union Medical College Institute of Pathogen Biology, Chinese Academy of Medical Sciences & Peking Union Medical College Lili Ren, Jianwei Wang, Qi Jin, Zichun Xiang, Zhiqiang Wu, Chao Wu, Yiwei Liu
EPI_ISL_403930 hCoV-19/Wuhan/IPBCAMS-WH-03/2019 Asia / China / Hubei / Wuhan 2019-12-30 Institute of Pathogen Biology, Chinese Academy of Medical Sciences & Peking Union Medical College Institute of Pathogen Biology, Chinese Academy of Medical Sciences & Peking Union Medical College Lili Ren, Jianwei Wang, Qi Jin, Zichun Xiang, Zhiqiang Wu, Chao Wu, Yiwei Liu
EPI_ISL_403931 hCoV-19/Wuhan/IPBCAMS-WH-02/2019 Asia / China / Hubei / Wuhan 2019-12-30 Institute of Pathogen Biology, Chinese Academy of Medical Sciences & Peking Union Medical College Institute of Pathogen Biology, Chinese Academy of Medical Sciences & Peking Union Medical College Lili Ren, Jianwei Wang, Qi Jin, Zichun Xiang, Zhiqiang Wu, Chao Wu, Yiwei Liu
EPI_ISL_403932 hCoV-19/Guangdong/20SF012/2020 Asia / China / Guangdong / Shenzhen 2020-01-14 Guangdong Provincial Center for Diseases Control and Prevention; Guangdong Provincial Public Health Department of Microbiology, Guangdong Provincial Center for Diseases Control and Prevention Min Kang, Jie Wu, Jing Lu, Tao Liu, Baisheng Li, Shujiang Mei, Feng Ruan, Lifeng Lin, Changwen Ke, Haojie Zhong, Yingtao Zhang, Lirong Zou, Xuguang Chen, Qi Zhu, Jianpeng Xiao, Jianxiang Geng, Zhe Liu, Jianxiong Hu, Weilin Zeng, Xing Li, Yuhuang Liao, Xiujuan Tang, Songjian Xiao, Ying Wang, Yingchao Song, Xue Zhuang, Lijun Liang, Guanhao He, Huihong Deng, Tie Song, Jianfeng He, Wenjun Ma
EPI_ISL_403933 hCoV-19/Guangdong/20SF013/2020 Asia / China / Guangdong / Shenzhen 2020-01-15 Guangdong Provincial Center for Diseases Control and Prevention; Guangdong Provincial Public Health Department of Microbiology, Guangdong Provincial Center for Diseases Control and Prevention Min Kang, Jie Wu, Jing Lu, Tao Liu, Baisheng Li, Shujiang Mei, Feng Ruan, Lifeng Lin, Changwen Ke, Haojie Zhong, Yingtao Zhang, Lirong Zou, Xuguang Chen, Qi Zhu, Jianpeng Xiao, Jianxiang Geng, Zhe Liu, Jianxiong Hu, Weilin Zeng, Xing Li, Yuhuang Liao, Xiujuan Tang, Songjian Xiao, Ying Wang, Yingchao Song, Xue Zhuang, Lijun Liang, Guanhao He, Huihong Deng, Tie Song, Jianfeng He, Wenjun Ma
EPI_ISL_403934 hCoV-19/Guangdong/20SF014/2020 Asia / China / Guangdong / Shenzhen 2020-01-15 Guangdong Provincial Center for Diseases Control and Prevention; Guangdong Provincial Public Health Department of Microbiology, Guangdong Provincial Center for Diseases Control and Prevention Min Kang, Jie Wu, Jing Lu, Tao Liu, Baisheng Li, Shujiang Mei, Feng Ruan, Lifeng Lin, Changwen Ke, Haojie Zhong, Yingtao Zhang, Lirong Zou, Xuguang Chen, Qi Zhu, Jianpeng Xiao, Jianxiang Geng, Zhe Liu, Jianxiong Hu, Weilin Zeng, Xing Li, Yuhuang Liao, Xiujuan Tang, Songjian Xiao, Ying Wang, Yingchao Song, Xue Zhuang, Lijun Liang, Guanhao

He, Huihong Deng, Tie Song, Jianfeng He, Wenjun Ma
EPI_ISL_403935 hCoV-19/Guangdong/20SF025/2020 Asia / China / Guangdong / Shenzhen 2020-01-15 Guangdong Provincial Center for Diseases Control and Prevention; Guangdong Provincial Public Health Department of Microbiology, Guangdong Provincial Center for Diseases Control and Prevention Min Kang, Jie Wu, Jing Lu, Tao Liu, Baisheng Li, Shujiang Mei, Feng Ruan, Lifeng Lin, Changwen Ke, Haojie Zhong, Yingtao Zhang, Lirong Zou, Xuguang Chen, Qi Zhu, Jianpeng Xiao, Jianxiang Geng, Zhe Liu, Jianxiong Hu, Weilin Zeng, Xing Li, Yuhuang Liao, Xiujuan Tang, Songjian Xiao, Ying Wang, Yingchao Song, Xue Zhuang, Lijun Liang, Guanhao He, Huihong Deng, Tie Song, Jianfeng He, Wenjun Ma
EPI_ISL_403936 hCoV-19/Guangdong/20SF028/2020 Asia / China / Guangdong / Zhuhai 2020-01-17 Guangdong Provincial Center for Diseases Control and Prevention; Guangdong Provincial Public Health Department of Microbiology, Guangdong Provincial Center for Diseases Control and Prevention Min Kang, Jie Wu, Jing Lu, Tao Liu, Baisheng Li, Shujiang Mei, Feng Ruan, Lifeng Lin, Changwen Ke, Haojie Zhong, Yingtao Zhang, Lirong Zou, Xuguang Chen, Qi Zhu, Jianpeng Xiao, Jianxiang Geng, Zhe Liu, Jianxiong Hu, Weilin Zeng, Xing Li, Yuhuang Liao, Xiujuan Tang, Songjian Xiao, Ying Wang, Yingchao Song, Xue Zhuang, Lijun Liang, Guanhao He, Huihong Deng, Tie Song, Jianfeng He, Wenjun Ma
EPI_ISL_403937 hCoV-19/Guangdong/20SF040/2020 Asia / China / Guangdong / Zhuhai 2020-01-18 Guangdong Provincial Center for Diseases Control and Prevention; Guangdong Provincial Public Health Department of Microbiology, Guangdong Provincial Center for Diseases Control and Prevention Min Kang, Jie Wu, Jing Lu, Tao Liu, Baisheng Li, Shujiang Mei, Feng Ruan, Lifeng Lin, Changwen Ke, Haojie Zhong, Yingtao Zhang, Lirong Zou, Xuguang Chen, Qi Zhu, Jianpeng Xiao, Jianxiang Geng, Zhe Liu, Jianxiong Hu, Weilin Zeng, Xing Li, Yuhuang Liao, Xiujuan Tang, Songjian Xiao, Ying Wang, Yingchao Song, Xue Zhuang, Lijun Liang, Guanhao He, Huihong Deng, Tie Song, Jianfeng He, Wenjun Ma
EPI_ISL_403962 hCoV-19/Thailand/61/2020 Asia / Thailand / Nonthaburi 2020-01-08 Bamrasnaradura Hospital 1. Department of Medical Sciences, Ministry of Public Health, Thailand 2. Thai Red Cross Emerging Infectious Diseases - Health Science Centre 3. Department of Disease Control, Ministry of Public Health, Thailand Pilailuk,Okada; Siripaporn,Phuygun; Thanutsapa,Thanadachakul; Supaporn,Wacharapluesadee; Sittiporn,Parnmen; Warawan,Wongboot; Sunthareeya,Waicharoen; Rome,Buathong; Malinee,Chittaganpitch; Nanthawan,Mekha
EPI_ISL_403963 hCoV-19/Thailand/74/2020 Asia / Thailand / Nonthaburi 2020-01-13 Bamrasnaradura Hospital 1. Department of Medical Sciences, Ministry of Public Health, Thailand 2. Thai Red Cross Emerging Infectious Diseases - Health Science Centre 3. Department of Disease Control, Ministry of Public Health, Thailand Pilailuk,Okada; Siripaporn,Phuygun; Thanutsapa,Thanadachakul; Supaporn,Wacharapluesadee; Sittiporn,Parnmen; Warawan,Wongboot; Sunthareeya,Waicharoen; Rome,Buathong; Malinee,Chittaganpitch; Nanthawan,Mekha
EPI_ISL_406223 hCoV-19/USA/AZ1/2020 North America / USA / Arizona / Phoenix 2020-01-22 Arizona Department of Health Services Pathogen Discovery, Respiratory Viruses Branch, Division of Viral Diseases, Centers for Disease Control and Prevention Ying Tao, Clinton R. Paden, Krista Queen, Anna Uehara, Yan Li, Jing Zhang, Xiaoyan Lu, Brian Lynch, Senthil Kumar K. Sakthivel, Brett L. Whitaker, Shifaq Kamili, Lijuan Wang, Janna' R. Murray, Susan I. Gerber, Stephen Lindstrom, Suxiang Tong
EPI_ISL_406592 hCoV-19/Shenzhen/SZTH-001/2020 Asia / China / Guangdong / Shenzhen 2020-01-13 Shenzhen Third People's Hospital Shenzhen Key Laboratory of Pathogen and Immunity, National Clinical Research Center for Infectious Disease,Shenzhen Third People's Hospital Yang Yang, Chenguang Shen, Li Xing, Zhixiang Xu, Haixia Zheng, Yingxia Liu
EPI_ISL_406593 hCoV-19/Shenzhen/SZTH-002/2020 Asia / China / Guangdong / Shenzhen 2020-01-13 Shenzhen Key Laboratory of Pathogen and Immunity, National Clinical Research Center for Infectious Disease, Shenzhen Third People's Hospital Shenzhen Key Laboratory of Pathogen and Immunity, National Clinical Research Center for Infectious Disease, Shenzhen Third People's Hospital Yang Yang, Chenguang Shen, Li Xing, Zhixiang Xu, Haixia Zheng, Yingxia Liu
EPI_ISL_406594 hCoV-19/Shenzhen/SZTH-003/2020 Asia / China / Guangdong / Shenzhen 2020-01-16 Shenzhen Key Laboratory of Pathogen and Immunity, National Clinical Research Center for Infectious Disease, Shenzhen Third People's Hospital Shenzhen Key Laboratory of Pathogen and Immunity, National Clinical Research Center for Infectious Disease, Shenzhen Third People's Hospital Yang Yang, Chenguang Shen, Li Xing, Zhixiang Xu, Haixia Zheng, Yingxia Liu
EPI_ISL_406595 hCoV-19/Shenzhen/SZTH-004/2020 Asia / China / Guangdong / Shenzhen 2020-01-16 Shenzhen Key Laboratory of Pathogen and Immunity, National Clinical Research Center for Infectious Disease, Shenzhen Third People's Hospital Shenzhen Key Laboratory of Pathogen and Immunity, National Clinical Research Center for Infectious Disease, Shenzhen Third People's Hospital Yang Yang, Chenguang Shen, Li Xing, Zhixiang Xu, Haixia Zheng, Yingxia Liu
EPI_ISL_406596 hCoV-19/France/IDF0372/2020 Europe / France / Ile-de-France / Paris 2020-01-23 Department of Infectious and Tropical Diseases, Bichat Claude Bernard Hospital, Paris National Reference Center for Viruses of Respiratory Infections, Institut Pasteur, Paris Mélanie Albert, Marion Barbet, Sylvie Behillil, Méline Bizard, Angela Brisebarre, Flora Donati, Vincent Enouf, Maud Vanpeene, Sylvie van der Werf, Yazdan Yazdanpanah, Xavier Lescure.
EPI_ISL_406597 hCoV-19/France/IDF0373/2020 Europe / France / Ile-de-France / Paris 2020-01-23 Department of Infectious and Tropical Diseases, Bichat Claude Bernard Hospital, Paris National Reference Center for Viruses of Respiratory Infections, Institut Pasteur, Paris Mélanie Albert, Marion Barbet, Sylvie Behillil, Méline Bizard, Angela Brisebarre, Flora Donati, Vincent Enouf, Maud Vanpeene, Sylvie van der Werf, Yazdan Yazdanpanah, Xavier Lescure.
EPI_ISL_407893 hCoV-19/Australia/NSW01/2020 Oceania / Australia / New South Wales / Sydney 2020-01-24 Centre for Infectious Diseases and Microbiology Laboratory Services NSW Health Pathology - Institute of

Clinical Pathology and Medical Research; Westmead Hospital; University of Sydney Eden J-S, Carter I, Rahman H, Holmes EC, Rockett R, O'Sullivan MV, Sintchenko V, Chen SC, Maddocks S, Kok J and Dwyer DE for the 2019-nCoV Study Group
EPI_ISL_407894 hCoV-19/Australia/QLD01/2020 Oceania / Australia / Queensland / Gold Coast 2020-01-28 Pathology Queensland Public Health Virology Laboratory Ben Huang, Alyssa Pyke, Amanda De Jong, Andrew Van Den Hurk, Carmel Taylor, David Warrilow, Doris Genge, Elisabeth Gamez, Glen Hewitson, Ian Maxwell Mackay, Inga Sultana, Jamie McMahon, Jean Barcelon, Judy Northill, Mitchell Finger, Natalie Simpson, Neelima Nair, Peter Burtonclay, Peter Moore, Sarah Wheatley, Sean Moody, Sonja Hall-Mendelin, Timothy Gardam, and Frederick Moore.
EPI_ISL_407896 hCoV-19/Australia/QLD02/2020 Oceania / Australia / Queensland / Gold Coast 2020-01-30 Pathology Queensland Public Health Virology Laboratory Ben Huang, Alyssa Pyke, Amanda De Jong, Andrew Van Den Hurk, Carmel Taylor, David Warrilow, Doris Genge, Elisabeth Gamez, Glen Hewitson, Ian Maxwell Mackay, Inga Sultana, Jamie McMahon, Jean Barcelon, Judy Northill, Mitchell Finger, Natalie Simpson, Neelima Nair, Peter Burtonclay, Peter Moore, Sarah Wheatley, Sean Moody, Sonja Hall-Mendelin, Timothy Gardam, and Frederick Moore.
EPI_ISL_411060 hCoV-19/Fujian/8/2020 Asia / China / Fujian 2020-01-21 Fujian Center for Disease Control and Prevention Fujian Center for Disease Control and Prevention Chen Wei, Zhang Yanhua, He Wenxiang, Weng Yuwei
EPI_ISL_411066 hCoV-19/Fujian/13/2020 Asia / China / Fujian 2020-01-22 Fujian Center for Disease Control and Prevention Fujian Center for Disease Control and Prevention Chen Wei, Zhang Yanhua, He Wenxiang, Weng Yuwei
EPI_ISL_412026 hCoV-19/Hefei/2/2020 Asia / China / Anhui / Hefei 2020-02-23 Second Hospital of Anhui Medical University Second Hospital of Anhui Medical University Changtai Wang, Zhongping Liua, Zixiang Chen, Xin Huang, Mengyuan Xua, Tengfei He, Mengji Lu, Zhenhua Zhang
EPI_ISL_412028 hCoV-19/Hong Kong/VM20001061-2/2020 Asia / Hong Kong 2020-01-22 Hong Kong Department of Health School of Public Health, The University of Hon g Kong Dominic N.C. Tsang, Daniel K.W. Chu, Leo L.M. Poon, Malik Peiris
EPI_ISL_412029 hCoV-19/Hong Kong/VB20024950-2/2020 Asia / Hong Kong 2020-01-30 Hong Kong Department of Health The University of Hong Kong Dominic N.C. Tsang, Daniel K.W. Chu, Leo L.M. Poon, Malik Peiris
EPI_ISL_412030 hCoV-19/Hong Kong/VB20026565-2/2020 Asia / Hong Kong 2020-02-01 Hong Kong Department of Health School of Public Health, The University of Hon g Kong Dominic N.C. Tsang, Daniel K.W. Chu, Leo L.M. Poon, Malik Peiris
EPI_ISL_413602 hCoV-19/Finland/FIN03032020A/2020 Europe / Finland / Helsinki 2020-03-03 Department of Virology and Immunology, University of Helsinki and Helsinki University Hospital, Huslab Finland Department of Virology, Faculty of Medicine, University of Helsinki, Helsinki, Finland Teemu Smura, Hannimari Kallio-Kokko, Olli Vapalahti
EPI_ISL_413603 hCoV-19/Finland/FIN03032020B/2020 Europe / Finland / Helsinki 2020-03-03 Department of Virology and Immunology, University of Helsinki and Helsinki University Hospital, Huslab Finland Department of Virology, Faculty of Medicine, University of Helsinki, Helsinki, Finland Teemu Smura, Hannimari Kallio-Kokko, Olli Vapalahti
EPI_ISL_413604 hCoV-19/Finland/FIN03032020C/2020 Europe / Finland / Helsinki 2020-03-03 Department of Virology and Immunology, University of Helsinki and Helsinki University Hospital, Huslab Finland Department of Virology, Faculty of Medicine, University of Helsinki, Helsinki, Finland Teemu Smura, Hannimari Kallio-Kokko, Olli Vapalahti
EPI_ISL_413606 hCoV-19/USA/CruiseA-1/2020 North America / USA 2020-02-17 unknown Pathogen Discovery, Respiratory Viruses Branch, Division of Viral Diseases, Centers for Dieases Control and Prevention Anna Uehara, Ying Tao, Clinton R. Paden, Krista Queen, Jing Zhang, Yan Li, Mary S. Keckler, Alison S Laufer Halpin, Haibin Wang, Jasmine Padilla, Justin Lee, Christopher A. Elkins, Susan I. Gerber, Suxiang Tong
EPI_ISL_413607 hCoV-19/USA/CruiseA-2/2020 North America / USA 2020-02-18 unknown Pathogen Discovery, Respiratory Viruses Branch, Division of Viral Diseases, Centers for Dieases Control and Prevention Anna Uehara, Ying Tao, Clinton R. Paden, Krista Queen, Jing Zhang, Yan Li, Mary S. Keckler, Alison S Laufer Halpin, Haibin Wang, Jasmine Padilla, Justin Lee, Christopher A. Elkins, Susan I. Gerber, Suxiang Tong
EPI_ISL_413608 hCoV-19/USA/CruiseA-3/2020 North America / USA 2020-02-18 unknown Pathogen Discovery, Respiratory Viruses Branch, Division of Viral Diseases, Centers for Dieases Control and Prevention Anna Uehara, Ying Tao, Clinton R. Paden, Krista Queen, Jing Zhang, Yan Li, Mary S. Keckler, Alison S Laufer Halpin, Haibin Wang, Jasmine Padilla, Justin Lee, Christopher A. Elkins, Susan I. Gerber, Suxiang Tong
EPI_ISL_413609 hCoV-19/USA/CruiseA-4/2020 North America / USA 2020-02-21 unknown Pathogen Discovery, Respiratory Viruses Branch, Division of Viral Diseases, Centers for Dieases Control and Prevention Anna Uehara, Ying Tao, Clinton R. Paden, Krista Queen, Jing Zhang, Yan Li, Mary S. Keckler, Alison S Laufer Halpin, Haibin Wang, Jasmine Padilla, Justin Lee, Christopher A. Elkins, Susan I. Gerber, Suxiang Tong
EPI_ISL_413610 hCoV-19/USA/CruiseA-5/2020 North America / USA 2020-02-21 unknown Pathogen Discovery, Respiratory Viruses Branch, Division of Viral Diseases, Centers for Dieases Control and Prevention Anna Uehara, Ying Tao, Clinton R. Paden, Krista Queen, Jing Zhang, Yan Li, Mary S. Keckler, Alison S Laufer Halpin, Haibin Wang, Jasmine Padilla, Justin Lee, Christopher A. Elkins, Susan I. Gerber, Suxiang Tong
EPI_ISL_413611 hCoV-19/USA/CruiseA-6/2020 North America / USA 2020-02-21 unknown Pathogen Discovery, Respiratory Viruses Branch, Division of Viral Diseases, Centers for Dieases Control and Prevention Anna Uehara, Ying Tao, Clinton R. Paden, Krista Queen, Jing Zhang, Yan Li, Mary S. Keckler, Alison S Laufer Halpin, Haibin Wang, Jasmine Padilla, Justin Lee, Christopher A. Elkins, Susan I. Gerber, Suxiang Tong
EPI_ISL_413612 hCoV-19/USA/CruiseA-7/2020 North America / USA 2020-02-17 unknown Pathogen

Discovery, Respiratory Viruses Branch, Division of Viral Diseases, Centers for Dieases Control and Prevention
Ying Tao, Clinton R. Paden, Krista Queen, Anna Uehara, Jing Zhang, Yan Li, Haibin Wang, Shifaq Kamili, Xiaoyan Lu, Brian Lynch, Senthil Kumar K. Sakthivel, Brett L. Whitaker, Lijuan Wang, Janna' R. Murray, Jasmine Padilla, Justin Lee, Susan I. Gerber, Stephen Lindstrom, Suxiang Tong
EPI_ISL_413613 hCoV-19/USA/CruiseA-8/2020 North America / USA 2020-02-17 unknown Pathogen Discovery, Respiratory Viruses Branch, Division of Viral Diseases, Centers for Dieases Control and Prevention
Ying Tao, Clinton R. Paden, Krista Queen, Anna Uehara, Jing Zhang, Yan Li, Haibin Wang, Shifaq Kamili, Xiaoyan Lu, Brian Lynch, Senthil Kumar K. Sakthivel, Brett L. Whitaker, Lijuan Wang, Janna' R. Murray, Jasmine Padilla, Justin Lee, Susan I. Gerber, Stephen Lindstrom, Suxiang Tong
EPI_ISL_413614 hCoV-19/USA/CruiseA-9/2020 North America / USA 2020-02-17 unknown Pathogen Discovery, Respiratory Viruses Branch, Division of Viral Diseases, Centers for Dieases Control and Prevention
Ying Tao, Clinton R. Paden, Krista Queen, Anna Uehara, Jing Zhang, Yan Li, Haibin Wang, Shifaq Kamili, Xiaoyan Lu, Brian Lynch, Senthil Kumar K. Sakthivel, Brett L. Whitaker, Lijuan Wang, Janna' R. Murray, Jasmine Padilla, Justin Lee, Susan I. Gerber, Stephen Lindstrom, Suxiang Tong
EPI_ISL_413615 hCoV-19/USA/CruiseA-10/2020 North America / USA 2020-02-17 unknown Pathogen Discovery, Respiratory Viruses Branch, Division of Viral Diseases, Centers for Dieases Control and Prevention
Ying Tao, Clinton R. Paden, Krista Queen, Anna Uehara, Jing Zhang, Yan Li, Haibin Wang, Shifaq Kamili, Xiaoyan Lu, Brian Lynch, Senthil Kumar K. Sakthivel, Brett L. Whitaker, Lijuan Wang, Janna' R. Murray, Jasmine Padilla, Justin Lee, Susan I. Gerber, Stephen Lindstrom, Suxiang Tong
EPI_ISL_413616 hCoV-19/USA/CruiseA-11/2020 North America / USA 2020-02-17 unknown Pathogen Discovery, Respiratory Viruses Branch, Division of Viral Diseases, Centers for Dieases Control and Prevention
Ying Tao, Clinton R. Paden, Krista Queen, Anna Uehara, Jing Zhang, Yan Li, Haibin Wang, Shifaq Kamili, Xiaoyan Lu, Brian Lynch, Senthil Kumar K. Sakthivel, Brett L. Whitaker, Lijuan Wang, Janna' R. Murray, Jasmine Padilla, Justin Lee, Susan I. Gerber, Stephen Lindstrom, Suxiang Tong
EPI_ISL_413617 hCoV-19/USA/CruiseA-12/2020 North America / USA 2020-02-20 unknown Pathogen Discovery, Respiratory Viruses Branch, Division of Viral Diseases, Centers for Dieases Control and Prevention
Ying Tao, Clinton R. Paden, Krista Queen, Anna Uehara, Jing Zhang, Yan Li, Haibin Wang, Shifaq Kamili, Xiaoyan Lu, Brian Lynch, Senthil Kumar K. Sakthivel, Brett L. Whitaker, Lijuan Wang, Janna' R. Murray, Jasmine Padilla, Justin Lee, Susan I. Gerber, Stephen Lindstrom, Suxiang Tong
EPI_ISL_413618 hCoV-19/USA/CruiseA-13/2020 North America / USA 2020-02-20 unknown Pathogen Discovery, Respiratory Viruses Branch, Division of Viral Diseases, Centers for Dieases Control and Prevention
Clinton R. Paden, Ying Tao, Krista Queen, Anna Uehara, Jing Zhang, Yan Li, Haibin Wang, Shifaq Kamili, Xiaoyan Lu, Brian Lynch, Senthil Kumar K. Sakthivel, Brett L. Whitaker, Lijuan Wang, Janna' R. Murray, Jasmine Padilla, Justin Lee, Susan I. Gerber, Stephen Lindstrom, Suxiang Tong
EPI_ISL_413619 hCoV-19/USA/CruiseA-14/2020 North America / USA 2020-02-25 unknown Pathogen Discovery, Respiratory Viruses Branch, Division of Viral Diseases, Centers for Dieases Control and Prevention
Clinton R. Paden, Ying Tao, Krista Queen, Anna Uehara, Jing Zhang, Yan Li, Haibin Wang, Shifaq Kamili, Xiaoyan Lu, Brian Lynch, Senthil Kumar K. Sakthivel, Brett L. Whitaker, Lijuan Wang, Janna' R. Murray, Jasmine Padilla, Justin Lee, Susan I. Gerber, Stephen Lindstrom, Suxiang Tong
EPI_ISL_413620 hCoV-19/USA/CruiseA-15/2020 North America / USA 2020-02-18 unknown Pathogen Discovery, Respiratory Viruses Branch, Division of Viral Diseases, Centers for Dieases Control and Prevention
Clinton R. Paden, Ying Tao, Krista Queen, Anna Uehara, Jing Zhang, Yan Li, Haibin Wang, Shifaq Kamili, Xiaoyan Lu, Brian Lynch, Senthil Kumar K. Sakthivel, Brett L. Whitaker, Lijuan Wang, Janna' R. Murray, Jasmine Padilla, Justin Lee, Susan I. Gerber, Stephen Lindstrom, Suxiang Tong
EPI_ISL_413621 hCoV-19/USA/CruiseA-16/2020 North America / USA 2020-02-18 unknown Pathogen Discovery, Respiratory Viruses Branch, Division of Viral Diseases, Centers for Dieases Control and Prevention
Clinton R. Paden, Ying Tao, Krista Queen, Anna Uehara, Jing Zhang, Yan Li, Haibin Wang, Shifaq Kamili, Xiaoyan Lu, Brian Lynch, Senthil Kumar K. Sakthivel, Brett L. Whitaker, Lijuan Wang, Janna' R. Murray, Jasmine Padilla, Justin Lee, Susan I. Gerber, Stephen Lindstrom, Suxiang Tong
EPI_ISL_413622 hCoV-19/USA/CruiseA-17/2020 North America / USA 2020-02-24 unknown Pathogen Discovery, Respiratory Viruses Branch, Division of Viral Diseases, Centers for Dieases Control and Prevention
Clinton R. Paden, Ying Tao, Krista Queen, Anna Uehara, Jing Zhang, Yan Li, Haibin Wang, Shifaq Kamili, Xiaoyan Lu, Brian Lynch, Senthil Kumar K. Sakthivel, Brett L. Whitaker, Lijuan Wang, Janna' R. Murray, Jasmine Padilla, Justin Lee, Susan I. Gerber, Stephen Lindstrom, Suxiang Tong
EPI_ISL_413623 hCoV-19/USA/CruiseA-18/2020 North America / USA 2020-02-24 unknown Pathogen Discovery, Respiratory Viruses Branch, Division of Viral Diseases, Centers for Dieases Control and Prevention
Clinton R. Paden, Ying Tao, Krista Queen, Anna Uehara, Jing Zhang, Yan Li, Haibin Wang, Shifaq Kamili, Xiaoyan Lu, Brian Lynch, Senthil Kumar K. Sakthivel, Brett L. Whitaker, Lijuan Wang, Janna' R. Murray, Jasmine Padilla, Justin Lee, Susan I. Gerber, Stephen Lindstrom, Suxiang Tong
EPI_ISL_413647 hCoV-19/Portugal/CV62/2020 Europe / Portugal 2020-03-01 Centro Hospital do Porto, E.P.E. - H. Geral de Santo Antonio Instituto Nacional de Saude (INSA) Raquel Guiomar, Inês Costa, Pedro Pechirra, Joana Mendonça, Luís Vieira, Helena Ramos, Joana Isidro, Vítor Borges, João Paulo Gomes
EPI_ISL_413648 hCoV-19/Portugal/CV63/2020 Europe / Portugal 2020-03-01 Centro Hospitalar e Universitário de Sao Joao, Porto Instituto Nacional de Saude (INSA) Raquel Guiomar, Inês Costa, Pedro Pechirra, Joana Mendonça, Luís Vieira, João Tiago Guimarães, Joana Isidro, Vítor Borges, João Paulo Gomes
EPI_ISL_413649 hCoV-19/USA/WA14-UW10/2020 North America / USA / Washington 2020-03-05 UW Virology Lab UW Virology Lab Pavitra Roychoudhury, Hong Xie, Keith Jerome, Alexander Greninger
EPI_ISL_413650 hCoV-19/USA/WA15-UW11/2020 North America / USA / Washington 2020-03-05 UW Virology Lab UW Virology Lab Pavitra Roychoudhury, Hong Xie, Keith Jerome, Alexander Greninger
EPI_ISL_413651 hCoV-19/USA/WA16-UW12/2020 North America / USA / Washington 2020-03-05 UW

Virology Lab	UW Virology Lab Pavitra Roychoudhury, Hong Xie, Keith Jerome, Alexander Greninger
EPI_ISL_413652	hCoV-19/USA/WA17-UW13/2020	North America / USA / Washington	2020-03-05	UW Virology Lab	UW Virology Lab Pavitra Roychoudhury, Hong Xie, Keith Jerome, Alexander Greninger
EPI_ISL_413653	hCoV-19/USA/WA18-UW14/2020	North America / USA / Washington	2020-03-05	UW Virology Lab	UW Virology Lab Pavitra Roychoudhury, Hong Xie, Keith Jerome, Alexander Greninger
EPI_ISL_413691	hCoV-19/China/WF0001/2020	Asia / China	2020-01 Weifang Center for Disease Control and Prevention	Weifang Center for Disease Control and Prevention & BGI-Shenzhen	Qing Nie, Xingguang Li, Erik M Volz, Han Fu, Haowei Wang, Xiaoyue Xi, Wei Chen, Dehui Liu, Yingying Chen, Mengmeng Tian, Wei Tan, Junjie Zai, Wanying Sun, Jiandong Li, Junhua Li
EPI_ISL_413692	hCoV-19/China/WF0002/2020	Asia / China	2020-01 Weifang Center for Disease Control and Prevention	Weifang Center for Disease Control and Prevention & BGI-Shenzhen	Qing Nie, Xingguang Li, Erik M Volz, Han Fu, Haowei Wang, Xiaoyue Xi, Wei Chen, Dehui Liu, Yingying Chen, Mengmeng Tian, Wei Tan, Junjie Zai, Wanying Sun, Jiandong Li, Junhua Li
EPI_ISL_413693	hCoV-19/China/WF0003/2020	Asia / China	2020-01 Weifang Center for Disease Control and Prevention	Weifang Center for Disease Control and Prevention & BGI-Shenzhen	Qing Nie, Xingguang Li, Erik M Volz, Han Fu, Haowei Wang, Xiaoyue Xi, Wei Chen, Dehui Liu, Yingying Chen, Mengmeng Tian, Wei Tan, Junjie Zai, Wanying Sun, Jiandong Li, Junhua Li
EPI_ISL_413694	hCoV-19/China/WF0004/2020	Asia / China	2020-01 Weifang Center for Disease Control and Prevention	Weifang Center for Disease Control and Prevention & BGI-Shenzhen	Qing Nie, Xingguang Li, Erik M Volz, Han Fu, Haowei Wang, Xiaoyue Xi, Wei Chen, Dehui Liu, Yingying Chen, Mengmeng Tian, Wei Tan, Junjie Zai, Wanying Sun, Jiandong Li, Junhua Li
EPI_ISL_413697	hCoV-19/China/WF0012/2020	Asia / China	2020-02 Weifang Center for Disease Control and Prevention	Weifang Center for Disease Control and Prevention & BGI-Shenzhen	Qing Nie, Xingguang Li, Erik M Volz, Han Fu, Haowei Wang, Xiaoyue Xi, Wei Chen, Dehui Liu, Yingying Chen, Mengmeng Tian, Wei Tan, Junjie Zai, Wanying Sun, Jiandong Li, Junhua Li
EPI_ISL_414600	hCoV-19/France/GE1583/2020	Europe / France / Grand-Est / Strasbourg	2020-02-26	Laboratoire de Virologie Institut de Virologie - INSERM U 1109 Hôpitaux Universitaires de Strasbourg	National Reference Center for Viruses of Respiratory Infections, Institut Pasteur, Paris	Mélnie Albert, Marion Barbet, Sylvie Behillil, Méline Bizard, Angela Brisebarre, Flora Donati Vincent Enouf, Maud Vanpeene, Sylvie van der Werf, Samira Fafi-Kremer
EPI_ISL_414620	hCoV-19/USA/WA-UW33/2020	North America / USA	2020-03-08	UW Virology Lab UW Virology Lab	Pavitra Roychoudhury, Hong Xie, Keith Jerome, Alexander Greninger
EPI_ISL_414625	hCoV-19/France/PL1643/2020	Europe / France / Pays de la Loire / Nantes	2020-02-26	Centre Hospitalier Régional Universitaire de Nantes Laboratoire de Virologie	National Reference Center for Viruses of Respiratory Infections, Institut Pasteur, Paris	Mélnie Albert, Marion Barbet, Sylvie Behillil, Méline Bizard, Angela Brisebarre, Flora Donati Vincent Enouf, Maud Vanpeene, Sylvie van der Werf, Marianne Coste-Burel
EPI_ISL_414626	hCoV-19/France/HF1684/2020	Europe / France / Hauts de France / Crépy en Valois	2020-02-29	unknown National Reference Center for Viruses of Respiratory Infections, Institut Pasteur, Paris	Mélnie Albert, Marion Barbet, Sylvie Behillil, Méline Bizard, Angela Brisebarre, Flora Donati Vincent Enouf, Maud Vanpeene, Sylvie van der Werf
EPI_ISL_414627	hCoV-19/France/HF1795/2020	Europe / France / Hauts de France / Compiègne	2020-03-02	Centre Hospitalier Compiègne Laboratoire de Biologie	National Reference Center for Viruses of Respiratory Infections, Institut Pasteur, Paris	Mélnie Albert, Marion Barbet, Sylvie Behillil, Méline Bizard, Angela Brisebarre, Flora Donati Vincent Enouf, Maud Vanpeene, Sylvie van der Werf, Raulin Olivia
EPI_ISL_414628	hCoV-19/France/HF1805/2020	Europe / France / Hauts de France / Compiègne	2020-03-02	Centre Hospitalier Compiègne Laboratoire de Biologie	National Reference Center for Viruses of Respiratory Infections, Institut Pasteur, Paris	Mélnie Albert, Marion Barbet, Sylvie Behillil, Méline Bizard, Angela Brisebarre, Flora Donati Vincent Enouf, Maud Vanpeene, Sylvie van der Werf, Raulin Olivia
EPI_ISL_414629	hCoV-19/France/HF1870/2020	Europe / France / Hauts de France / Compiègne	2020-03-03	Centre Hospitalier Compiègne Laboratoire de Biologie	National Reference Center for Viruses of Respiratory Infections, Institut Pasteur, Paris	Mélnie Albert, Marion Barbet, Sylvie Behillil, Méline Bizard, Angela Brisebarre, Flora Donati Vincent Enouf, Maud Vanpeene, Sylvie van der Werf, Raulin Olivia
EPI_ISL_414630	hCoV-19/France/HF1871/2020	Europe / France / Hauts de France / Compiègne	2020-03-03	Centre Hospitalier Compiègne Laboratoire de Biologie	National Reference Center for Viruses of Respiratory Infections, Institut Pasteur, Paris	Mélnie Albert, Marion Barbet, Sylvie Behillil, Méline Bizard, Angela Brisebarre, Flora Donati Vincent Enouf, Maud Vanpeene, Sylvie van der Werf, Raulin Olivia
EPI_ISL_414631	hCoV-19/France/GE1973/2020	Europe / France / Grand-Est / Reims	2020-03-04	Hôpital Robert Debré Laboratoire de Virologie	National Reference Center for Viruses of Respiratory Infections, Institut Pasteur, Paris Mélnie Albert, Marion Barbet, Sylvie Behillil, Méline Bizard, Angela Brisebarre, Flora Donati Vincent Enouf, Maud Vanpeene, Sylvie van der Werf, Laurent Andreoletti
EPI_ISL_414632	hCoV-19/France/GE1977/2020	Europe / France / Grand-Est / Reims	2020-03-04	Hôpital Robert Debré Laboratoire de Virologie	National Reference Center for Viruses of Respiratory Infections, Institut Pasteur, Paris Mélnie Albert, Marion Barbet, Sylvie Behillil, Méline Bizard, Angela Brisebarre, Flora Donati Vincent Enouf, Maud Vanpeene, Sylvie van der Werf, Laurent Andreoletti
EPI_ISL_414633	hCoV-19/France/IDF1980/2020	Europe / France / Ile-de-France / Pontoise	2020-03-04	Centre Hospitalier René Dubois Laboratoire de Microbiologie - Bât A	National Reference Center for Viruses of Respiratory Infections, Institut Pasteur, Paris	Mélnie Albert, Marion Barbet, Sylvie Behillil, Méline Bizard, Angela Brisebarre, Flora Donati Vincent Enouf, Maud Vanpeene, Sylvie van der Werf, Pascale Martres
EPI_ISL_414634	hCoV-19/France/HF1986/2020	Europe / France / Hauts de France / Compiègne	2020-03-04

Centre Hospitalier Compiègne Laboratoire de Biologie National Reference Center for Viruses of Respiratory Infections, Institut Pasteur, Paris Mélnie Albert, Marion Barbet, Sylvie Behillil, Méline Bizard, Angela Brisebarre, Flora Donati Vincent Enouf, Maud Vanpeene, Sylvie van der Werf, Raulin Olivia
EPI_ISL_414635 hCoV-19/France/HF1988/2020 Europe / France / Hauts de France / Compiègne 2020-03-04 Centre Hospitalier Compiègne Laboratoire de Biologie National Reference Center for Viruses of Respiratory Infections, Institut Pasteur, Paris Mélnie Albert, Marion Barbet, Sylvie Behillil, Méline Bizard, Angela Brisebarre, Flora Donati Vincent Enouf, Maud Vanpeene, Sylvie van der Werf, Raulin Olivia
EPI_ISL_414663 hCoV-19/Guangzhou/GZMU0016/2020 Asia / China / Guangdong / Guangzhou 2020-02-25 State Key Laboratory of Respiratory Disease, National Clinical Research Center for Respiratory Disease, Guangzhou Institute of Respiratory Health, the First Affiliated Hospital of Guangzhou Medical University The First Affiliated Hospital of Guangzhou Medical University & BGI-Shenzhen Zhao et al
EPI_ISL_414686 hCoV-19/Guangzhou/GZMU0030/2020 Asia / China / Guangdong / Guangzhou 2020-02-27 State Key Laboratory of Respiratory Disease, National Clinical Research Center for Respiratory Disease, Guangzhou Institute of Respiratory Health, the First Affiliated Hospital of Guangzhou Medical University The First Affiliated Hospital of Guangzhou Medical University & BGI-Shenzhen Zhao et al
EPI_ISL_414687 hCoV-19/Guangzhou/GZMU0031/2020 Asia / China / Guangdong / Guangzhou 2020-02-25 State Key Laboratory of Respiratory Disease, National Clinical Research Center for Respiratory Disease, Guangzhou Institute of Respiratory Health, the First Affiliated Hospital of Guangzhou Medical University the First Affiliated Hospital of Guangzhou Medical University & BGI-Shenzhen Zhao et al
EPI_ISL_414689 hCoV-19/Guangzhou/GZMU0044/2020 Asia / China / Guangdong / Guangzhou 2020-02-25 State Key Laboratory of Respiratory Disease, National Clinical Research Center for Respiratory Disease, Guangzhou Institute of Respiratory Health, the First Affiliated Hospital of Guangzhou Medical University The First Affiliated Hospital of Guangzhou Medical University & BGI-Shenzhen Zhao et al
EPI_ISL_414690 hCoV-19/Guangzhou/GZMU0047/2020 Asia / China / Guangdong / Guangzhou 2020-02-25 State Key Laboratory of Respiratory Disease, National Clinical Research Center for Respiratory Disease, Guangzhou Institute of Respiratory Health, the First Affiliated Hospital of Guangzhou Medical University The First Affiliated Hospital of Guangzhou Medical University & BGI-Shenzhen Zhao et al
EPI_ISL_414691 hCoV-19/Guangzhou/GZMU0048/2020 Asia / China / Guangdong / Guangzhou 2020-02-25 State Key Laboratory of Respiratory Disease, National Clinical Research Center for Respiratory Disease, Guangzhou Institute of Respiratory Health, the First Affiliated Hospital of Guangzhou Medical University The First Affiliated Hospital of Guangzhou Medical University & BGI-Shenzhen Zhao et al
EPI_ISL_414692 hCoV-19/Guangzhou/GZMU0014/2020 Asia / China / Guangdong / Guangzhou 2020-02-25 State Key Laboratory of Respiratory Disease, National Clinical Research Center for Respiratory Disease, Guangzhou Institute of Respiratory Health, the First Affiliated Hospital of Guangzhou Medical University The First Affiliated Hospital of Guangzhou Medical University & BGI-Shenzhen Zhao et al
EPI_ISL_414936 hCoV-19/Shandong/LY003-2/2020 Asia / China / Shandong 2020-01-23 Shandong Provincial Center for Disease Control and Prevention Beijing Institute of Microbiology and Epidemiology Xiao-Lin Jiang, Xiao-Li Zhang, Xiang-Na Zhao, Cun-Bao Li, Jie Lei, Zeng-Qiang Kou, Wen-Kui Sun, Yang Hang, Feng Gao, Sheng-Xiang Ji, Can-Fang Lin, Bo Pang, Ming-Xiao Yao, Guo-Lin Wang, Lin Yao, Li-Jun Duan, Xiao Wei, Dian-Ming Kang, Mai-Juan Ma
EPI_ISL_414937 hCoV-19/Shandong/LY004-2/2020 Asia / China / Shandong 2020-01-26 Shandong Provincial Center for Disease Control and Prevention Beijing Institute of Microbiology and Epidemiology Xiao-Lin Jiang, Xiao-Li Zhang, Xiang-Na Zhao, Cun-Bao Li, Jie Lei, Zeng-Qiang Kou, Wen-Kui Sun, Yang Hang, Feng Gao, Sheng-Xiang Ji, Can-Fang Lin, Bo Pang, Ming-Xiao Yao, Guo-Lin Wang, Lin Yao, Li-Jun Duan, Xiao Wei, Dian-Ming Kang, Mai-Juan Ma
EPI_ISL_414938 hCoV-19/Shandong/LY005-2/2020 Asia / China / Shandong 2020-01-24 Shandong Provincial Center for Disease Control and Prevention Beijing Institute of Microbiology and Epidemiology Xiao-Lin Jiang, Xiao-Li Zhang, Xiang-Na Zhao, Cun-Bao Li, Jie Lei, Zeng-Qiang Kou, Wen-Kui Sun, Yang Hang, Feng Gao, Sheng-Xiang Ji, Can-Fang Lin, Bo Pang, Ming-Xiao Yao, Guo-Lin Wang, Lin Yao, Li-Jun Duan, Xiao Wei, Dian-Ming Kang, Mai-Juan Ma
EPI_ISL_414940 hCoV-19/Shandong/LY007-2/2020 Asia / China / Shandong 2020-01-25 Shandong Provincial Center for Disease Control and Prevention Beijing Institute of Microbiology and Epidemiology Xiao-Lin Jiang, Xiao-Li Zhang, Xiang-Na Zhao, Cun-Bao Li, Jie Lei, Zeng-Qiang Kou, Wen-Kui Sun, Yang Hang, Feng Gao, Sheng-Xiang Ji, Can-Fang Lin, Bo Pang, Ming-Xiao Yao, Guo-Lin Wang, Lin Yao, Li-Jun Duan, Xiao Wei, Dian-Ming Kang, Mai-Juan Ma
EPI_ISL_414941 hCoV-19/Shandong/LY008-2/2020 Asia / China / Shandong 2020-01-30 Shandong Provincial Center for Disease Control and Prevention Beijing Institute of Microbiology and Epidemiology Xiao-Lin Jiang, Xiao-Li Zhang, Xiang-Na Zhao, Cun-Bao Li, Jie Lei, Zeng-Qiang Kou, Wen-Kui Sun, Yang Hang, Feng Gao, Sheng-Xiang Ji, Can-Fang Lin, Bo Pang, Ming-Xiao Yao, Guo-Lin Wang, Lin Yao, Li-Jun Duan, Xiao Wei, Dian-Ming Kang, Mai-Juan Ma
EPI_ISL_415920 hCoV-19/Wales/PHW32/2020 Europe / United Kingdom / Wales 2020-03-12 Wales Specialist Virology Centre Public Health Wales Microbiology Cardiff Catherine Moore, Joanne Watkins, Sally Corden, Tom Connor
EPI_ISL_417200 hCoV-19/USA/WI-UW-06/2020 North America / USA / Wisconsin / Cottage Grove 2020-03-21 University of Wisconsin-Madison AIDS Vaccine Research Laboratories University of Wisconsin-Madison AIDS Vaccine Research Laboratories Gage Moreno, Katarina Braun, et al. AIDS Vaccine Research Laboratories
EPI_ISL_417201 hCoV-19/USA/WI-UW-07/2020 North America / USA / Wisconsin / Madison 2020-03-21 University of Wisconsin-Madison AIDS Vaccine Research Laboratories University of Wisconsin-Madison AIDS Vaccine Research Laboratories Gage Moreno, Katarina Braun, et al. AIDS Vaccine Research Laboratories
EPI_ISL_417202 hCoV-19/USA/WI-UW-08/2020 North America / USA / Wisconsin / Stoughton 2020-03-21

University of Wisconsin-Madison AIDS Vaccine Research Laboratories	University of Wisconsin-Madison AIDS Vaccine Research Laboratories	Gage Moreno, Katarina Braun, et al. AIDS Vaccine Research Laboratories
EPI_ISL_417203	hCoV-19/USA/WI-UW-09/2020	North America / USA / Wisconsin / Madison	2020-03-21	University of Wisconsin-Madison AIDS Vaccine Research Laboratories	University of Wisconsin-Madison AIDS Vaccine Research Laboratories	Gage Moreno, Katarina Braun, et al. AIDS Vaccine Research Laboratories
EPI_ISL_417204	hCoV-19/USA/WI-UW-10/2020	North America / USA / Wisconsin / Cambridge	2020-03-21	University of Wisconsin-Madison AIDS Vaccine Research Laboratories	University of Wisconsin-Madison AIDS Vaccine Research Laboratories	Gage Moreno, Katarina Braun, et al. AIDS Vaccine Research Laboratories
EPI_ISL_417211	hCoV-19/NewZealand/CoV001/2020	Oceania / New Zealand / Otago	2020-03-11	Dunedin Hospital	University of Otago	M.E. Quiñones-Mateu, B. Lawley, J. Grant, R. Harfoot, J. Ussher
EPI_ISL_417213	hCoV-19/England/20102068502/2020	Europe / United Kingdom / England	2020-03-01	Respiratory Virus Unit, Microbiology Services Colindale, Public Health England	Respiratory Virus Unit, Microbiology Services Colindale, Public Health England	Monica Galiano, Shahjahan Miah, Angie Lackenby, Omolola Akinbami, Tiina Talts, Leena Bhaw, Richard Myers, Steven Platt, Kirstin Edwards, Jonathan Hubb, Joanna Ellis, Maria Zambon
EPI_ISL_417217	hCoV-19/England/20102098802/2020	Europe / United Kingdom / England	2020-03-01	Respiratory Virus Unit, Microbiology Services Colindale, Public Health England	Respiratory Virus Unit, Microbiology Services Colindale, Public Health England	Monica Galiano, Shahjahan Miah, Angie Lackenby, Omolola Akinbami, Tiina Talts, Leena Bhaw, Richard Myers, Steven Platt, Kirstin Edwards, Jonathan Hubb, Joanna Ellis, Maria Zambon
EPI_ISL_417220	hCoV-19/England/20104002606/2020	Europe / United Kingdom / England	2020-03-03	Respiratory Virus Unit, Microbiology Services Colindale, Public Health England	Respiratory Virus Unit, Microbiology Services Colindale, Public Health England	Monica Galiano, Shahjahan Miah, Angie Lackenby, Omolola Akinbami, Tiina Talts, Leena Bhaw, Richard Myers, Steven Platt, Kirstin Edwards, Jonathan Hubb, Joanna Ellis, Maria Zambon
EPI_ISL_417222	hCoV-19/England/20104003002/2020	Europe / United Kingdom / England	2020-03-04	Respiratory Virus Unit, Microbiology Services Colindale, Public Health England	Respiratory Virus Unit, Microbiology Services Colindale, Public Health England	Monica Galiano, Shahjahan Miah, Angie Lackenby, Omolola Akinbami, Tiina Talts, Leena Bhaw, Richard Myers, Steven Platt, Kirstin Edwards, Jonathan Hubb, Joanna Ellis, Maria Zambon
EPI_ISL_417226	hCoV-19/England/20104004902/2020	Europe / United Kingdom / England	2020-03-02	Respiratory Virus Unit, Microbiology Services Colindale, Public Health England	Respiratory Virus Unit, Microbiology Services Colindale, Public Health England	Monica Galiano, Shahjahan Miah, Angie Lackenby, Omolola Akinbami, Tiina Talts, Leena Bhaw, Richard Myers, Steven Platt, Kirstin Edwards, Jonathan Hubb, Joanna Ellis, Maria Zambon
EPI_ISL_417227	hCoV-19/England/20104007503/2020	Europe / United Kingdom / England	2020-03-03	Respiratory Virus Unit, Microbiology Services Colindale, Public Health England	Respiratory Virus Unit, Microbiology Services Colindale, Public Health England	Monica Galiano, Shahjahan Miah, Angie Lackenby, Omolola Akinbami, Tiina Talts, Leena Bhaw, Richard Myers, Steven Platt, Kirstin Edwards, Jonathan Hubb, Joanna Ellis, Maria Zambon
EPI_ISL_417228	hCoV-19/England/20104008402/2020	Europe / United Kingdom / England	2020-03-03	Respiratory Virus Unit, Microbiology Services Colindale, Public Health England	Respiratory Virus Unit, Microbiology Services Colindale, Public Health England	Monica Galiano, Shahjahan Miah, Angie Lackenby, Omolola Akinbami, Tiina Talts, Leena Bhaw, Richard Myers, Steven Platt, Kirstin Edwards, Jonathan Hubb, Joanna Ellis, Maria Zambon
EPI_ISL_417230	hCoV-19/England/20104008702/2020	Europe / United Kingdom / England	2020-03-02	Respiratory Virus Unit, Microbiology Services Colindale, Public Health England	Respiratory Virus Unit, Microbiology Services Colindale, Public Health England	Monica Galiano, Shahjahan Miah, Angie Lackenby, Omolola Akinbami, Tiina Talts, Leena Bhaw, Richard Myers, Steven Platt, Kirstin Edwards, Jonathan Hubb, Joanna Ellis, Maria Zambon
EPI_ISL_417231	hCoV-19/England/20104008802/2020	Europe / United Kingdom / England	2020-03-03	Respiratory Virus Unit, Microbiology Services Colindale, Public Health England	Respiratory Virus Unit, Microbiology Services Colindale, Public Health England	Monica Galiano, Shahjahan Miah, Angie Lackenby, Omolola Akinbami, Tiina Talts, Leena Bhaw, Richard Myers, Steven Platt, Kirstin Edwards, Jonathan Hubb, Joanna Ellis, Maria Zambon
EPI_ISL_417232	hCoV-19/England/20104008902/2020	Europe / United Kingdom / England	2020-03-04	Respiratory Virus Unit, Microbiology Services Colindale, Public Health England	Respiratory Virus Unit, Microbiology Services Colindale, Public Health England	Monica Galiano, Shahjahan Miah, Angie Lackenby, Omolola Akinbami, Tiina Talts, Leena Bhaw, Richard Myers, Steven Platt, Kirstin Edwards, Jonathan Hubb, Joanna Ellis, Maria Zambon
EPI_ISL_417233	hCoV-19/England/20104009002/2020	Europe / United Kingdom / England	2020-03-02	Respiratory Virus Unit, Microbiology Services Colindale, Public Health England	Respiratory Virus Unit, Microbiology Services Colindale, Public Health England	Monica Galiano, Shahjahan Miah, Angie Lackenby, Omolola Akinbami, Tiina Talts, Leena Bhaw, Richard Myers, Steven Platt, Kirstin Edwards, Jonathan Hubb, Joanna Ellis, Maria Zambon
EPI_ISL_417234	hCoV-19/England/20104009102/2020	Europe / United Kingdom / England	2020-03-02	Respiratory Virus Unit, Microbiology Services Colindale, Public Health England	Respiratory Virus Unit, Microbiology Services Colindale, Public Health England	Monica Galiano, Shahjahan Miah, Angie Lackenby, Omolola Akinbami, Tiina Talts, Leena Bhaw, Richard Myers, Steven Platt, Kirstin Edwards, Jonathan Hubb, Joanna Ellis, Maria Zambon

EPI_ISL_417235 hCoV-19/England/20104013703/2020 Europe / United Kingdom / England 2020-03-02 Respiratory Virus Unit, Microbiology Services Colindale, Public Health England Respiratory Virus Unit, Microbiology Services Colindale, Public Health England Monica Galiano, Shahjahan Miah, Angie Lackenby, Omolola Akinbami, Tiina Talts, Leena Bhaw, Richard Myers, Steven Platt, Kirstin Edwards, Jonathan Hubb, Joanna Ellis, Maria Zambon
EPI_ISL_417236 hCoV-19/England/20104015302/2020 Europe / United Kingdom / England 2020-03-01 Respiratory Virus Unit, Microbiology Services Colindale, Public Health England Respiratory Virus Unit, Microbiology Services Colindale, Public Health England Monica Galiano, Shahjahan Miah, Angie Lackenby, Omolola Akinbami, Tiina Talts, Leena Bhaw, Richard Myers, Steven Platt, Kirstin Edwards, Jonathan Hubb, Joanna Ellis, Maria Zambon
EPI_ISL_417238 hCoV-19/England/20104035803/2020 Europe / United Kingdom / England 2020-03-03 Respiratory Virus Unit, Microbiology Services Colindale, Public Health England Respiratory Virus Unit, Microbiology Services Colindale, Public Health England Monica Galiano, Shahjahan Miah, Angie Lackenby, Omolola Akinbami, Tiina Talts, Leena Bhaw, Richard Myers, Steven Platt, Kirstin Edwards, Jonathan Hubb, Joanna Ellis, Maria Zambon
EPI_ISL_417239 hCoV-19/England/20106003303/2020 Europe / United Kingdom / England 2020-03-03 Respiratory Virus Unit, Microbiology Services Colindale, Public Health England Respiratory Virus Unit, Microbiology Services Colindale, Public Health England Monica Galiano, Shahjahan Miah, Angie Lackenby, Omolola Akinbami, Tiina Talts, Leena Bhaw, Richard Myers, Steven Platt, Kirstin Edwards, Jonathan Hubb, Joanna Ellis, Maria Zambon
EPI_ISL_417240 hCoV-19/England/20106004803/2020 Europe / United Kingdom / England 2020-03-03 Respiratory Virus Unit, Microbiology Services Colindale, Public Health England Respiratory Virus Unit, Microbiology Services Colindale, Public Health England Monica Galiano, Shahjahan Miah, Angie Lackenby, Omolola Akinbami, Tiina Talts, Leena Bhaw, Richard Myers, Steven Platt, Kirstin Edwards, Jonathan Hubb, Joanna Ellis, Maria Zambon
EPI_ISL_417244 hCoV-19/England/20106005403/2020 Europe / United Kingdom / England 2020-03-03 Respiratory Virus Unit, Microbiology Services Colindale, Public Health England Respiratory Virus Unit, Microbiology Services Colindale, Public Health England Monica Galiano, Shahjahan Miah, Angie Lackenby, Omolola Akinbami, Tiina Talts, Leena Bhaw, Richard Myers, Steven Platt, Kirstin Edwards, Jonathan Hubb, Joanna Ellis, Maria Zambon
EPI_ISL_417246 hCoV-19/England/20106145903/2020 Europe / United Kingdom / England 2020-03-03 Respiratory Virus Unit, Microbiology Services Colindale, Public Health England Respiratory Virus Unit, Microbiology Services Colindale, Public Health England Monica Galiano, Shahjahan Miah, Angie Lackenby, Omolola Akinbami, Tiina Talts, Leena Bhaw, Richard Myers, Steven Platt, Kirstin Edwards, Jonathan Hubb, Joanna Ellis, Maria Zambon
EPI_ISL_417248 hCoV-19/England/20108003302/2020 Europe / United Kingdom / England 2020-03-03 Respiratory Virus Unit, Microbiology Services Colindale, Public Health England Respiratory Virus Unit, Microbiology Services Colindale, Public Health England Monica Galiano, Shahjahan Miah, Angie Lackenby, Omolola Akinbami, Tiina Talts, Leena Bhaw, Richard Myers, Steven Platt, Kirstin Edwards, Jonathan Hubb, Joanna Ellis, Maria Zambon
EPI_ISL_418182 hCoV-19/Spain/Madrid_H8_37/2020 Europe / Spain / Madrid 2020-03-12 Hospital Universitario 12 de Octubre Hospital Universitario La Paz Elias Dahdouh, Sara González, Fernando Lázaro, Esther Viedma, Natalia Stella, Julio García, Juan Carlos Galán, Rafael Cantón, Mª Dolores Folgueira, Rafael Delgado, Jesús Mingorance
EPI_ISL_418183 hCoV-19/Hungary/mbl2/2020 Europe / Hungary / Baranya 2020-03-17 Virological Research Group, Szentágothai Research Centre Bioinformatics Research Group, Szentágothai Research Centre Péter Urbán, Endre Gábor Tóth, Gábor Kemenesi, Róbert Herczeg, Attila Gyenesei, Ferenc Jakab
EPI_ISL_418184 hCoV-19/USA/WI-GMF-00018/2020 North America / USA / Wisconsin / La Crosse County 2020-03-18 Gundersen Molecular Diagnostics Laboratory Kabara Cancer Research Institute Craig S. Richmond & Paraic A. Kenny
EPI_ISL_418186 hCoV-19/USA/WI-GMF-00227/2020 North America / USA / Wisconsin / La Crosse County 2020-03-23 Gundersen Molecular Diagnostic Laboratory Kabara Cancer Research Institute Craig S. Richmond & Paraic A. Kenny
EPI_ISL_418187 hCoV-19/USA/WI-GMF-00228/2020 North America / USA / Wisconsin / La Crosse County 2020-03-23 Gundersen Molecular Diagnostics Laboratory Kabara Cancer Research Institute Craig S. Richmond & Paraic A. Kenny
EPI_ISL_418188 hCoV-19/USA/WI-GMF-00232/2020 North America / USA / Wisconsin / Monroe County 2020-03-23 Gundersen Molecular Diagnostics Laboratory Kabara Cancer Research Institute Craig S. Richmond & Paraic A. Kenny
EPI_ISL_418192 hCoV-19/USA/NY-NYUMC7/2020 North America / USA / New York / Manhattan 2020-03-16 NYU Langone Health Department of Pathology and Medicine, New York University School of Medicine Margaret Black, John Cadley, Paolo Cotzia, John Chen, Dacia Dimartino, Xiaojun Feng, Adriana Heguy, Megan Hogan, Emily Huang, George Jour, Christian Marier, Matthew T. Maurano, Mark J. Mulligan, Peter Meyn, Jared Pinnell, Amy Rapkiewicz, Marie Samanovic-Golden, Antonio Serrano, Guomiao Shen, Matija Snuderl, Nick Vulpescu, Gael Westby, Paul Zappile
EPI_ISL_418193 hCoV-19/USA/NY-NYUMC8/2020 North America / USA / New York / Manhattan 2020-03-17 NYU Langone Health Department of Pathology and Medicine, New York University School of Medicine Margaret Black, John Cadley, Paolo Cotzia, John Chen, Dacia Dimartino, Xiaojun Feng, Adriana Heguy, Megan Hogan, Emily Huang, George Jour, Christian Marier, Matthew T. Maurano, Mark J. Mulligan, Peter Meyn, Jared Pinnell, Amy Rapkiewicz, Marie Samanovic-Golden, Antonio Serrano, Guomiao Shen, Matija Snuderl, Nick Vulpescu,

Gael Westby, Paul Zappile
EPI_ISL_418196 hCoV-19/USA/NY-NYUMC11/2020 North America / USA / New York / Manhattan 2020-03-14
NYU Langone Health Department of Pathology and Medicine, New York University School of Medicine
Margaret Black, John Cadley, Paolo Cotzia, John Chen, Dacia Dimartino, Xiaojun Feng, Adriana Heguy, Megan Hogan, Emily Huang, George Jour, Christian Marier, Matthew T. Maurano, Mark J. Mulligan, Peter Meyn, Jared Pinnell, Amy Rapkiewicz, Marie Samanovic-Golden, Antonio Serrano, Guomiao Shen, Matija Snuderl, Nick Vulpescu, Gael Westby, Paul Zappile
EPI_ISL_418197 hCoV-19/USA/NY-NYUMC12/2020 North America / USA / New York / Manhattan 2020-03-15
NYU Langone Health Department of Pathology and Medicine, New York University School of Medicine
Margaret Black, John Cadley, Paolo Cotzia, John Chen, Dacia Dimartino, Xiaojun Feng, Adriana Heguy, Megan Hogan, Emily Huang, George Jour, Christian Marier, Matthew T. Maurano, Mark J. Mulligan, Peter Meyn, Jared Pinnell, Amy Rapkiewicz, Marie Samanovic-Golden, Antonio Serrano, Guomiao Shen, Matija Snuderl, Nick Vulpescu, Gael Westby, Paul Zappile
EPI_ISL_418198 hCoV-19/USA/NY-NYUMC13/2020 North America / USA / New York / Manhattan 2020-03-17
NYU Langone Health Department of Pathology and Medicine, New York University School of Medicine
Margaret Black, John Cadley, Paolo Cotzia, John Chen, Dacia Dimartino, Xiaojun Feng, Adriana Heguy, Megan Hogan, Emily Huang, George Jour, Christian Marier, Matthew T. Maurano, Mark J. Mulligan, Peter Meyn, Jared Pinnell, Amy Rapkiewicz, Marie Samanovic-Golden, Antonio Serrano, Guomiao Shen, Matija Snuderl, Nick Vulpescu, Gael Westby, Paul Zappile
EPI_ISL_418199 hCoV-19/USA/NY-NYUMC14/2020 North America / USA / New York / Manhattan 2020-03-17
NYU Langone Health Department of Pathology and Medicine, New York University School of Medicine
Margaret Black, John Cadley, Paolo Cotzia, John Chen, Dacia Dimartino, Xiaojun Feng, Adriana Heguy, Megan Hogan, Emily Huang, George Jour, Christian Marier, Matthew T. Maurano, Mark J. Mulligan, Peter Meyn, Jared Pinnell, Amy Rapkiewicz, Marie Samanovic-Golden, Antonio Serrano, Guomiao Shen, Matija Snuderl, Nick Vulpescu, Gael Westby, Paul Zappile
EPI_ISL_418200 hCoV-19/USA/NY-NYUMC15/2020 North America / USA / New York / Manhattan 2020-03-17
NYU Langone Health Department of Pathology and Medicine, New York University School of Medicine
Margaret Black, John Cadley, Paolo Cotzia, John Chen, Dacia Dimartino, Xiaojun Feng, Adriana Heguy, Megan Hogan, Emily Huang, George Jour, Christian Marier, Matthew T. Maurano, Mark J. Mulligan, Peter Meyn, Jared Pinnell, Amy Rapkiewicz, Marie Samanovic-Golden, Antonio Serrano, Guomiao Shen, Matija Snuderl, Nick Vulpescu, Gael Westby, Paul Zappile
EPI_ISL_418203 hCoV-19/USA/NY-NYUMC18/2020 North America / USA / New York / Manhattan 2020-03-17
NYU Langone Health Department of Pathology and Medicine, New York University School of Medicine
Margaret Black, John Cadley, Paolo Cotzia, John Chen, Dacia Dimartino, Xiaojun Feng, Adriana Heguy, Megan Hogan, Emily Huang, George Jour, Christian Marier, Matthew T. Maurano, Mark J. Mulligan, Peter Meyn, Jared Pinnell, Amy Rapkiewicz, Marie Samanovic-Golden, Antonio Serrano, Guomiao Shen, Matija Snuderl, Nick Vulpescu, Gael Westby, Paul Zappile
EPI_ISL_418204 hCoV-19/USA/NY-NYUMC19/2020 North America / USA / New York / Manhattan 2020-03-17
NYU Langone Health Department of Pathology and Medicine, New York University School of Medicine
Margaret Black, John Cadley, Paolo Cotzia, John Chen, Dacia Dimartino, Xiaojun Feng, Adriana Heguy, Megan Hogan, Emily Huang, George Jour, Christian Marier, Matthew T. Maurano, Mark J. Mulligan, Peter Meyn, Jared Pinnell, Amy Rapkiewicz, Marie Samanovic-Golden, Antonio Serrano, Guomiao Shen, Matija Snuderl, Nick Vulpescu, Gael Westby, Paul Zappile
EPI_ISL_418206 hCoV-19/Senegal/003/2020 Africa / Senegal / Dakar 2020-02-28 Institut Pasteur Dakar Institut Pasteur de Dakar Ndongo Dia, Ousmane Faye, Amadou Alpha Sall
EPI_ISL_418207 hCoV-19/Senegal/016/2020 Africa / Senegal / Dakar 2020-03-02 Institut Pasteur Dakar Institut Pasteur de Dakar Ndongo Dia, Ousmane Faye, Amadou Alpha Sall
EPI_ISL_418208 hCoV-19/Senegal/020/2020 Africa / Senegal / Dakar 2020-03-04 Institut Pasteur Dakar Institut Pasteur de Dakar Ndongo Dia, Ousmane Faye, Amadou Alpha Sall
EPI_ISL_418209 hCoV-19/Senegal/026/2020 Africa / Senegal / Dakar 2020-03-03 Institut Pasteur Dakar Institut Pasteur de Dakar Ndongo Dia, Ousmane Faye, Amadou Alpha Sall
EPI_ISL_418210 hCoV-19/Senegal/073/2020 Africa / Senegal / Touba 2020-03-10 Institut Pasteur Dakar Institut Pasteur de Dakar Ndongo Dia, Ousmane Faye, Amadou Alpha Sall
EPI_ISL_418211 hCoV-19/Senegal/082/2020 Africa / Senegal / Touba 2020-03-11 Institut Pasteur Dakar Institut Pasteur de Dakar Ndongo Dia, Ousmane Faye, Amadou Alpha Sall
EPI_ISL_418212 hCoV-19/Senegal/087/2020 Africa / Senegal / Touba 2020-03-11 Institut Pasteur Dakar Institut Pasteur de Dakar Ndongo Dia, Ousmane Faye, Amadou Alpha sall
EPI_ISL_418213 hCoV-19/Senegal/094/2020 Africa / Senegal / Touba 2020-03-12 Institut Pasteur Dakar Institut Pasteur de Dakar Ndongo Dia, Ousmane Faye, Amadou Alpha Sall
EPI_ISL_418216 hCoV-19/Senegal/136/2020 Africa / Senegal / Dakar 2020-03-13 Institut Pasteur Dakar Institut Pasteur de Dakar Ndongo Dia, Ousmane Faye, Amadou Alpha Sall
EPI_ISL_418217 hCoV-19/Senegal/139/2020 Africa / Senegal / Mbour 2020-03-13 Institut Pasteur Dakar Institut Pasteur de Dakar Ndongo Dia, Ousmane Faye, Amadou Alpha Sall
EPI_ISL_418218 hCoV-19/France/HF1465/2020 Europe / France / Hauts de France / Compiègne 2020-02-21
Centre Hospitalier Compiègne Laboratoire de Biologie National Reference Center for Viruses of Respiratory Infections, Institut Pasteur, Paris Mélanie Albert, Marion Barbet, Sylvie Behillil, Méline Bizard, Angela Brisebarre, Flora Donati, Fabiana Gambaro, Etienne Simon-Lorière, Vincent Enouf, Maud Vanpeene, Sylvie van der Werf, Raulin Olivia
EPI_ISL_418219 hCoV-19/France/B1623/2020 Europe / France / Bretagne / Brest 2020-02-26 CHU - Hôpital Cavale Blanche - Labo. de Virologie National Reference Center for Viruses of Respiratory

Infections, Institut Pasteur, Paris     Mélanie Albert, Marion Barbet, Sylvie Behillil, Méline Bizard, Angela Brisebarre, Flora Donati, Fabiana Gambaro, Etienne Simon-Lorière, Vincent Enouf, Maud Vanpeene, Sylvie van der Werf, Léa Pilorge
EPI_ISL_418220  hCoV-19/France/HF1645/2020      Europe / France / Hauts de France / Compiègne   2020-02-28 Centre Hospitalier Compiègne Laboratoire de Biologie    National Reference Center for Viruses of Respiratory Infections, Institut Pasteur, Paris     Mélanie Albert, Marion Barbet, Sylvie Behillil, Méline Bizard, Angela Brisebarre, Flora Donati, Fabiana Gambaro, Etienne Simon-Lorière, Vincent Enouf, Maud Vanpeene, Sylvie van der Werf, Raulin Olivia
EPI_ISL_418222  hCoV-19/France/CVL2000/2020     Europe / France / Centre-Val de Loire / Tours   2020-03-04 CHRU Bretonneau - Serv. Bacterio-Virol. National Reference Center for Viruses of Respiratory Infections, Institut Pasteur, Paris Mélanie Albert, Marion Barbet, Sylvie Behillil, Méline Bizard, Angela Brisebarre, Flora Donati, Fabiana Gambaro, Etienne Simon-Lorière, Vincent Enouf, Maud Vanpeene, Sylvie van der Werf, Julien Marlet
EPI_ISL_418223  hCoV-19/France/HF2060/2020      Europe / France / Hauts de France / Compiègne   2020-03-05 Centre Hospitalier Compiègne Laboratoire de Biologie    National Reference Center for Viruses of Respiratory Infections, Institut Pasteur, Paris     Mélanie Albert, Marion Barbet, Sylvie Behillil, Méline Bizard, Angela Brisebarre, Flora Donati, Fabiana Gambaro, Etienne Simon-Lorière, Vincent Enouf, Maud Vanpeene, Sylvie van der Werf, Raulin Olivia
EPI_ISL_418224  hCoV-19/France/HF2150/2020      Europe / France / Hauts de France / Compiègne   2020-03-08 Centre Hospitalier Compiègne Laboratoire de Biologie    National Reference Center for Viruses of Respiratory Infections, Institut Pasteur, Paris     Mélanie Albert, Marion Barbet, Sylvie Behillil, Méline Bizard, Angela Brisebarre, Flora Donati, Fabiana Gambaro, Etienne Simon-Lorière, Vincent Enouf, Maud Vanpeene, Sylvie van der Werf, Raulin Olivia
EPI_ISL_418225  hCoV-19/France/HF2155/2020      Europe / France / Hauts de France / Compiègne   2020-03-08 Centre Hospitalier Compiègne Laboratoire de Biologie    National Reference Center for Viruses of Respiratory Infections, Institut Pasteur, Paris     Mélanie Albert, Marion Barbet, Sylvie Behillil, Méline Bizard, Angela Brisebarre, Flora Donati, Fabiana Gambaro, Etienne Simon-Lorière, Vincent Enouf, Maud Vanpeene, Sylvie van der Werf, Raulin Olivia
EPI_ISL_418226  hCoV-19/France/HF2381/2020      Europe / France / Hauts de France / Crouy en Thelle     2020-03-09   EHPAD - Résidences les Cèdres   National Reference Center for Viruses of Respiratory Infections, Institut Pasteur, Paris Mélanie Albert, Marion Barbet, Sylvie Behillil, Méline Bizard, Angela Brisebarre, Flora Donati, Etienne Simon-Lorière, Vincent Enouf, Maud Vanpeene, Sylvie van der Werf
EPI_ISL_418227  hCoV-19/France/HF2393/2020      Europe / France / Hauts de France / Compiègne   2020-03-12 Centre Hospitalier Compiègne Laboratoire de Biologie    National Reference Center for Viruses of Respiratory Infections, Institut Pasteur, Paris     Mélanie Albert, Marion Barbet, Sylvie Behillil, Méline Bizard, Angela Brisebarre, Flora Donati, Etienne Simon-Lorière, Vincent Enouf, Maud Vanpeene, Sylvie van der Werf, Raulin Olivia
EPI_ISL_418228  hCoV-19/France/HF2405/2020      Europe / France / Hauts de France / Compiègne   2020-03-12 Centre Hospitalier Compiègne Laboratoire de Biologie    National Reference Center for Viruses of Respiratory Infections, Institut Pasteur, Paris     Mélanie Albert, Marion Barbet, Sylvie Behillil, Méline Bizard, Angela Brisebarre, Flora Donati, Etienne Simon-Lorière, Vincent Enouf, Maud Vanpeene, Sylvie van der Werf, Raulin Olivia
EPI_ISL_418229  hCoV-19/France/IDF2410/2020     Europe / France / Ile de France / Levallois-Perret      2020-03-12   Hopital franco britannique - Laboratoire        National Reference Center for Viruses of Respiratory Infections, Institut Pasteur, Paris     Mélanie Albert, Marion Barbet, Sylvie Behillil, Méline Bizard, Angela Brisebarre, Flora Donati, Etienne Simon-Lorière, Vincent Enouf, Maud Vanpeene, Sylvie van der Werf, Marianne Asso Bonnet
EPI_ISL_418230  hCoV-19/France/IDF2420/2020     Europe / France / Ile de France / Paris 2020-03-13 Clinique AVERAY LA BROUSTE, Med. Polyvalente    National Reference Center for Viruses of Respiratory Infections, Institut Pasteur, Paris     Mélanie Albert, Marion Barbet, Sylvie Behillil, Méline Bizard, Angela Brisebarre, Flora Donati, Etienne Simon-Lorière, Vincent Enouf, Maud Vanpeene, Sylvie van der Werf, Elsa Ngwem
EPI_ISL_418231  hCoV-19/France/HF2496/2020      Europe / France / Hauts de France / Compiègne   2020-03-15 Centre Hospitalier Compiègne Laboratoire de Biologie    National Reference Center for Viruses of Respiratory Infections, Institut Pasteur, Paris     Mélanie Albert, Marion Barbet, Sylvie Behillil, Méline Bizard, Angela Brisebarre, Flora Donati, Etienne Simon-Lorière, Vincent Enouf, Maud Vanpeene, Sylvie van der Werf, Raulin Olivia
EPI_ISL_418232  hCoV-19/France/IDF2532/2020     Europe / France / Ile de France / Meudon la Forêt       2020-03-15   Service des Urgences    National Reference Center for Viruses of Respiratory Infections, Institut Pasteur, Paris  Mélanie Albert, Marion Barbet, Sylvie Behillil, Méline Bizard, Angela Brisebarre, Flora Donati, Etienne Simon-Lorière, Vincent Enouf, Maud Vanpeene, Sylvie van der Werf, Boubkeur
EPI_ISL_418233  hCoV-19/France/IDF2533/2020     Europe / France / Ile de France / Meudon la Forêt       2020-03-15   Service des Urgences    National Reference Center for Viruses of Respiratory Infections, Institut Pasteur, Paris  Mélanie Albert, Marion Barbet, Sylvie Behillil, Méline Bizard, Angela Brisebarre, Flora Donati, Etienne Simon-Lorière, Vincent Enouf, Maud Vanpeene, Sylvie van der Werf, Boubkeur
EPI_ISL_418234  hCoV-19/France/IDF2534/2020     Europe / France / Ile de France / Longjumeau    2020-03-14 LABM GH nord Essonne    National Reference Center for Viruses of Respiratory Infections, Institut Pasteur, Paris   Mélanie Albert, Marion Barbet, Sylvie Behillil, Méline Bizard, Angela Brisebarre, Flora Donati, Etienne Simon-Lorière, Vincent Enouf, Maud Vanpeene, Sylvie van der Werf, Christine Lambert
EPI_ISL_418235  hCoV-19/France/IDF2561/2020     Europe / France / Ile de France / Vanves        2020-03-16 Cabinet médical National Reference Center for Viruses of Respiratory Infections, Institut Pasteur, Paris

Mélanie Albert, Marion Barbet, Sylvie Behillil, Méline Bizard, Angela Brisebarre, Flora Donati, Etienne Simon-Lorière, Vincent Enouf, Maud Vanpeene, Sylvie van der Werf
EPI_ISL_418236 hCoV-19/France/HF2586/2020 Europe / France / Hauts de France / Compiègne 2020-03-16 Centre Hospitalier Compiègne Laboratoire de Biologie National Reference Center for Viruses of Respiratory Infections, Institut Pasteur, Paris Mélanie Albert, Marion Barbet, Sylvie Behillil, Méline Bizard, Angela Brisebarre, Flora Donati, Etienne Simon-Lorière, Vincent Enouf, Maud Vanpeene, Sylvie van der Werf, Raulin Olivia
EPI_ISL_418237 hCoV-19/France/HF2595/2020 Europe / France / Hauts de France / Compiègne 2020-03-16 Centre Hospitalier Compiègne Laboratoire de Biologie National Reference Center for Viruses of Respiratory Infections, Institut Pasteur, Paris Mélanie Albert, Marion Barbet, Sylvie Behillil, Méline Bizard, Angela Brisebarre, Flora Donati, Etienne Simon-Lorière, Vincent Enouf, Maud Vanpeene, Sylvie van der Werf, Raulin Olivia
EPI_ISL_418238 hCoV-19/France/HF2597/2020 Europe / France / Hauts de France / Compiègne 2020-03-16 Centre Hospitalier Compiègne Laboratoire de Biologie National Reference Center for Viruses of Respiratory Infections, Institut Pasteur, Paris Mélanie Albert, Marion Barbet, Sylvie Behillil, Méline Bizard, Angela Brisebarre, Flora Donati, Etienne Simon-Lorière, Vincent Enouf, Maud Vanpeene, Sylvie van der Werf, Raulin Olivia
EPI_ISL_418239 hCoV-19/France/HF2601/2020 Europe / France / Hauts de France / Compiègne 2020-03-16 Centre Hospitalier Compiègne Laboratoire de Biologie National Reference Center for Viruses of Respiratory Infections, Institut Pasteur, Paris Mélanie Albert, Marion Barbet, Sylvie Behillil, Méline Bizard, Angela Brisebarre, Flora Donati, Etienne Simon-Lorière, Vincent Enouf, Maud Vanpeene, Sylvie van der Werf, Raulin Olivia
EPI_ISL_418240 hCoV-19/France/IDF2684/2020 Europe / France / Ile de France / Longjumeau 2020-03-16 LABM GH nord Essonne National Reference Center for Viruses of Respiratory Infections, Institut Pasteur, Paris Mélanie Albert, Marion Barbet, Sylvie Behillil, Méline Bizard, Angela Brisebarre, Flora Donati, Etienne Simon-Lorière, Vincent Enouf, Maud Vanpeene, Sylvie van der Werf, Christine Lambert
EPI_ISL_418241 hCoV-19/Algeria/G0638_2264/2020 Africa / Algeria / Boufarik 2020-03-02 NIC Viral Respiratory Unit - Institut Pasteur of Algeria National Reference Center for Viruses of Respiratory Infections, Institut Pasteur, Paris Mélanie Albert, Marion Barbet, Sylvie Behillil, Méline Bizard, Angela Brisebarre, Flora Donati, Etienne Simon-Lorière, Vincent Enouf, Maud Vanpeene, Sylvie van der Werf, Fawzi Derrar
EPI_ISL_418242 hCoV-19/Algeria/G0640_2265/2020 Africa / Algeria / Blida 2020-03-08 NIC Viral Respiratory Unit - Institut Pasteur of Algeria National Reference Center for Viruses of Respiratory Infections, Institut Pasteur, Paris Mélanie Albert, Marion Barbet, Sylvie Behillil, Méline Bizard, Angela Brisebarre, Flora Donati, Etienne Simon-Lorière, Vincent Enouf, Maud Vanpeene, Sylvie van der Werf, Fawzi Derrar
EPI_ISL_418243 hCoV-19/Spain/Andalucia201272/2020 Europe / Spain / Andalusia 2020-02-28 HOSPITAL UNIVERSITARIO VIRGEN DE LAS NIEVES Instituto de Salud Carlos III Iglesias-Caballero, M. Molinero Calamita, M. González-Esguevillas, M. Camarero, S. Pozo, F. Casas, I. Jiménez, P. Jiménez, M. Zaballos, A. Monzón, S. Varona, S. Juliá, M. Cuesta, I. Sanbonmatsu S.
EPI_ISL_418244 hCoV-19/Spain/Andalucia201373/2020 Europe / Spain / Andalusia 2020-03-02 HOSPITAL UNIVERSITARIO VIRGEN DE LAS NIEVES Instituto de Salud Carlos III Iglesias-Caballero, M. Molinero Calamita, M. González-Esguevillas, M. Camarero, S. Pozo, F. Casas, I. Jiménez, P. Jiménez, M. Zaballos, A. Monzón, S. Varona, S. Juliá, M. Cuesta, I. Sanbonmatsu S.
EPI_ISL_418245 hCoV-19/Spain/CastillaLaMancha201328/2020 Europe / Spain / Castilla La Mancha 2020-03-01 Hospital General y Universitario de Guadalajara Instituto de Salud Carlos III Iglesias-Caballero, M. Molinero Calamita, M. González-Esguevillas, M. Camarero, S. Pozo, F. Casas, I. Jiménez, P. Jiménez, M. Zaballos, A. Monzón, S. Varona, S. Juliá, M. Cuesta, I. Gonzalez-Praetorius A.
EPI_ISL_418246 hCoV-19/Spain/CastillaLaMancha201329/2020 Europe / Spain / Castilla-La Mancha 2020-03-01 Hospital General y Universitario de Guadalajara Instituto de Salud Carlos III Iglesias-Caballero, M. Molinero Calamita, M. González-Esguevillas, M. Camarero, S. Pozo, F. Casas, I. Jiménez, P. Jiménez, M. Zaballos, A. Monzón, S. Varona, S. Juliá, M. Cuesta, I. Gonzalez-Praetorius A.
EPI_ISL_418247 hCoV-19/Spain/CastillayLeon201061/2020 Europe / Spain / Castilla y Leon 2020-02-26 HOSPITAL GENERAL DE SEGOVIA Instituto de Salud Carlos III Iglesias-Caballero, M. Molinero Calamita, M. González-Esguevillas, M. Camarero, S. Pozo, F. Casas, I. Jiménez, P. Jiménez, M. Zaballos, A. Monzón, S. Varona, S. Juliá, M. Cuesta, I. Hernando-Real S.
EPI_ISL_418248 hCoV-19/Spain/CastillayLeon201323/2020 Europe / Spain / Castilla y Leon 2020-03-01 COMPLEJO ASISTENCIAL UNIVERSITARIO DE BURGOS Instituto de Salud Carlos III Iglesias-Caballero, M. Molinero Calamita, M. González-Esguevillas, M. Camarero, S. Pozo, F. Casas, I. Jiménez, P. Jiménez, M. Zaballos, A. Monzón, S. Varona, S. Juliá, M. Cuesta, I. Megias-Lobon G.
EPI_ISL_418249 hCoV-19/Spain/CastillayLeon201372/2020 Europe / Spain / Castilla y Leon 2020-03-03 COMPLEJO ASISTENCIAL UNIVERSITARIO DE BURGOS Instituto de Salud Carlos III Iglesias-Caballero, M. Molinero Calamita, M. González-Esguevillas, M. Camarero, S. Pozo, F. Casas, I. Jiménez, P. Jiménez, M. Zaballos, A. Monzón, S. Varona, S. Juliá, M. Cuesta, I. Megias-Lobon G.
EPI_ISL_418250 hCoV-19/Spain/Cataluna201396/2020 Europe / Spain / Catalonia 2020 HOSPITAL CLINIC Instituto de Salud Carlos III Iglesias-Caballero, M. Molinero Calamita, M. González-Esguevillas, M. Camarero, S. Pozo, F. Casas, I. Jiménez, P. Jiménez, M. Zaballos, A. Monzón, S. Varona, S. Juliá, M. Cuesta, I. Marcos M.A
EPI_ISL_418251 hCoV-19/Spain/Madrid201105/2020 Europe / Spain / Madrid 2020-02-25 HOSPITAL UNIVERSITARIO LA PAZ Instituto de Salud Carlos III Iglesias-Caballero, M. Molinero Calamita, M. González-Esguevillas, M.

Camarero, S. Pozo, F. Casas, I. Jiménez, P. Jiménez, M. Zaballos, A. Monzón, S. Varona, S. Juliá, M. Cuesta, I. Romero P.
EPI_ISL_418252 hCoV-19/Spain/Madrid201449/2020 Europe / Spain / Madrid 2020-03-04 FUNDACION JIMENEZ DIAZ Instituto de Salud Carlos III Iglesias-Caballero, M. Molinero Calamita, M. González-Esguevillas, M. Camarero, S. Pozo, F. Casas, I. Jiménez, P. Jiménez, M. Zaballos, A. Monzón, S. Varona, S. Juliá, M. Cuesta, I. Fernández Roblas, R.
EPI_ISL_418253 hCoV-19/Spain/PaisVasco201382/2020 Europe / Spain / Basque Country 2020-03-02 HOSPITAL TXAGORRITXU Instituto de Salud Carlos III Iglesias-Caballero, M. Molinero Calamita, M. González-Esguevillas, M. Camarero, S. Pozo, F. Casas, I. Jiménez, P. Jiménez, M. Zaballos, A. Monzón, S. Varona, S. Juliá, M. Cuesta, I. Gomez-Gonzalez C.
EPI_ISL_418255 hCoV-19/Italy/TE4925/2020 Europe / Italy / Abruzzo 2020-03-14 "Presidio Ospedaliero ""S. Spirito"" - PESCARA" "Istituto Zooprofilattico Sperimentale dell'Abruzzo e Molise ""G. Caporale""" Lorusso A, Marcacci M, Cammà C, Monaco F, Puglia I, Di Pasquale A, Rinaldi A, Mangone I, Savini G
EPI_ISL_418256 hCoV-19/Italy/TE4880/2020 Europe / Italy / Abruzzo 2020-03-14 Ospedale “San Liberatore” di Atri "Istituto Zooprofilattico Sperimentale dell'Abruzzo e Molise ""G. Caporale""" Lorusso A, Marcacci M, Di Domenico M, Puglia I, Curini V, Ancora M, Di Pasquale A, Rinaldi A, Mangone I, Cammà C, Savini G.
EPI_ISL_418257 hCoV-19/Italy/TE5056/2020 Europe / Italy / Abruzzo 2020-03-17 Ospedale Civile Giuseppe Mazzini, Teramo "Istituto Zooprofilattico Sperimentale dell'Abruzzo e Molise ""G.Caporale""" Lorusso A, Marcacci M, Di Domenico M, Puglia I, Curini V, Ancora M, Di Pasquale A, Rinaldi A, Mangone I, Cammà C, Savini G.
EPI_ISL_418258 hCoV-19/Italy/TE4953/2020 Europe / Italy / Abruzzo 2020-03-14 "Presidio ospedaliero ""Santo Spirito""" "Istituto Zooprofilattico Sperimentale dell'Abruzzo e Molise ""G. Caporale""" Lorusso A, Marcacci M, Di Domenico M, Puglia I, Curini V, Ancora M, Di Pasquale A, Rinaldi A, Mangone I, Cammà C, Savini G.
EPI_ISL_418259 hCoV-19/Italy/TE4959/2020 Europe / Italy / Abruzzo 2020-03-14 "Presidio ospedaliero ""Santo Spirito""" "Istituto Zooprofilattico Sperimentale dell'Abruzzo e Molise ""G. Caporale""" Lorusso A, Marcacci M, Di Domenico M, Puglia I, Curini V, Ancora M, Di Pasquale A, Rinaldi A, Mangone I, Cammà C, Savini G.
EPI_ISL_418260 hCoV-19/Italy/TE4836/2020 Europe / Italy / Abruzzo 2020-03-16 Ospedale Civile Giuseppe Mazzini "Istituto Zooprofilattico Sperimentale dell'Abruzzo e Molise ""G. Caporale""" Lorusso A, Marcacci M, Di Domenico M, Puglia I, Curini V, Ancora M, Di Pasquale A, Rinaldi A, Mangone I, Cammà C, Savini G.
EPI_ISL_418263 hCoV-19/Greece/10/2020 Europe / Greece / Athens 2020-03-18 Laboratory of Microbiology, Department of Medicine, National and Kapodistrian University of Athens, Greece Laboratory of Biology, Department of Medicine, Democritus University of Thrace, Greece Maria Bampali, Elisavet Gatzidou, Nikolaos Dovrolis, Stavroula Veletza, Nikolaos Spanakis, Ioannis Karakasiliotis
EPI_ISL_418264 hCoV-19/Greece/12/2020 Europe / Greece / Athens 2020-03-18 Laboratory of Microbiology, Department of Medicine, National and Kapodistrian University of Athens, Greece Laboratory of Biology, Department of Medicine, Democritus University of Thrace, Greece Maria Bampali, Elisavet Gatzidou, Nikolaos Dovrolis, Stavroula Veletza, Nikolaos Spanakis, Ioannis Karakasiliotis
EPI_ISL_418265 hCoV-19/Greece/16/2020 Europe / Greece / Athens 2020-03-18 Laboratory of Microbiology, Department of Medicine, National and Kapodistrian University of Athens, Greece Laboratory of Biology, Department of Medicine, Democritus University of Thrace, Greece Maria Bampali, Elisavet Gatzidou, Nikolaos Dovrolis, Stavroula Veletza, Nikolaos Spanakis, Ioannis Karakasiliotis
EPI_ISL_418267 hCoV-19/Vietnam/19-02S/2020 Asia / Vietnam / Ho Chi Minh City 2020-01-22 unknown Microbiology and Immunology department Nguyen,H.T., Cao,T.M., Pham,H.T.T., Vu,N.P.H., Dao,M.H., Huynh,L.T.K., Nguyen,L.T., Nguyen,N.T., Nguyen,T.T.N., Nguyen,A.H., Luong,Q.C., Nguyen,T.V., Tran,K.C., Pham,Q.D., Tran,T., Hoang,C.Q., Nguyen,T.T., Le,H.Q., Phung,T.M., Vo,T.N.A., Nguyen,S.N., Pham,D.T., Nguyen,T.V. and Phan,L.T.
EPI_ISL_418269 hCoV-19/Vietnam/19-01S/2020 Asia / Vietnam / Ho Chi Minh City 2020-01-22 unknown Microbiology and Immunology department Cao,T.M., Nguyen,H.T., Pham,H.T.T., Vu,N.P.H., Dao,M.H., Huynh,L.T.K., Nguyen,L.T., Nguyen,N.T., Nguyen,T.T.N., Nguyen,A.H., Luong,Q.C., Nguyen,T.V., Tran,K.C., Pham,Q.D., Tran,T., Hoang,C.Q., Nguyen,T.T., Le,H.Q., Phung,T.M., Vo,T.N.A., Nguyen,S.N., Pham,D.T., Phan,L.T. and Nguyen,T.V.
EPI_ISL_418270 hCoV-19/Belgium/JL-03044/2020 Europe / Belgium / Antwerp 2020-03-04 KU Leuven, Clinical and Epidemiological Virology KU Leuven, Clinical and Epidemiological Virology Tony Wawina, Joan Marti-Carreras, Bert Vanmechelen, Piet Maes
EPI_ISL_418294 hCoV-19/England/SHEF-BFDFA/2020 Europe / United Kingdom / England / South Yorkshire 2020-03-18 Virology Department, Sheffield Teaching Hospitals NHS Foundation Trust Department of Infection, Immunity and Cardiovascular Disease, The Florey Institute, The Medical School, University of Sheffield Thushan de Silva, Matthew Parker, Adri Angyal, Rebecca Brown, Rachel Tucker, Paul Parsons, Danielle Groves, Alex Keeley, Dave Partridge, Matthew Wyles, Benjamin Lindsey, Mehmet Yavuz, Mohammad Raza, Cariad Evans
EPI_ISL_418295 hCoV-19/England/SHEF-BFE06/2020 Europe / United Kingdom / England / South Yorkshire 2020-03-17 Virology Department, Sheffield Teaching Hospitals NHS Foundation Trust Department of Infection, Immunity and Cardiovascular Disease, The Florey Institute, The Medical School, University of Sheffield Thushan de Silva, Matthew Parker, Adri Angyal, Rebecca Brown, Rachel Tucker, Paul Parsons, Danielle Groves, Alex Keeley, Dave Partridge, Matthew Wyles, Benjamin Lindsey, Mehmet Yavuz, Mohammad Raza, Cariad Evans
EPI_ISL_418548 hCoV-19/Ireland/21145/2020 Europe / Ireland / Dublin 2020-03-06 UCD National Virus Reference Laboratory UCD National Virus Reference Laboratory Michael Carr, Gabriel Gonzalez, Jonathan Dean, Suzie Coughlan, Alison Murphy, Kevin Byrne, Ken Wolfe, Jeff Connell, Brendan Loftus, Cillian F

De Gascun
EPI_ISL_419508 hCoV-19/Wales/PHWC-24D87/2020 Europe / United Kingdom / Wales 2020-03-20 Wales Specialist Virology Centre Public Health Wales Microbiology Cardiff Catherine Moore, Joanne Watkins, Sally Corden, Sara Rey, Matt Bull, Tom Connor
EPI_ISL_419541 hCoV-19/Germany/NRW-24/2020 Europe / Germany / North Rhine Westphalia / Duesseldorf 2020-03-14 Center of Medical Microbiology, Virology, and Hospital Hygiene, University of Duesseldorf Center of Medical Microbiology, Virology, and Hospital Hygiene, University of Duesseldorf Ortwin Adams, Marcel Andree, Alexander Dilthey, Torsten Feldt, Sandra Hauka, Torsten Houwaart, Björn-Erik Jensen, Detlef Kindgen-Milles, Malte Kohns Vasconcelos, Klaus Pfeffer, Tina Senff, Daniel Strelow, Jörg Timm, Andreas Walker, Tobias Wienemann
EPI_ISL_419542 hCoV-19/Germany/NRW-25/2020 Europe / Germany / North Rhine Westphalia / Duesseldorf 2020-03-15 Center of Medical Microbiology, Virology, and Hospital Hygiene, University of Duesseldorf Center of Medical Microbiology, Virology, and Hospital Hygiene, University of Duesseldorf Ortwin Adams, Marcel Andree, Alexander Dilthey, Torsten Feldt, Sandra Hauka, Torsten Houwaart, Björn-Erik Jensen, Detlef Kindgen-Milles, Malte Kohns Vasconcelos, Klaus Pfeffer, Tina Senff, Daniel Strelow, Jörg Timm, Andreas Walker, Tobias Wienemann
EPI_ISL_419543 hCoV-19/Germany/NRW-26/2020 Europe / Germany / North Rhine Westphalia / Duesseldorf 2020-03-15 Center of Medical Microbiology, Virology, and Hospital Hygiene, University of Duesseldorf Center of Medical Microbiology, Virology, and Hospital Hygiene, University of Duesseldorf Ortwin Adams, Marcel Andree, Alexander Dilthey, Torsten Feldt, Sandra Hauka, Torsten Houwaart, Björn-Erik Jensen, Detlef Kindgen-Milles, Malte Kohns Vasconcelos, Klaus Pfeffer, Tina Senff, Daniel Strelow, Jörg Timm, Andreas Walker, Tobias Wienemann
EPI_ISL_419544 hCoV-19/Germany/NRW-27/2020 Europe / Germany / North Rhine Westphalia / Duesseldorf 2020-03-15 Center of Medical Microbiology, Virology, and Hospital Hygiene, University of Duesseldorf Center of Medical Microbiology, Virology, and Hospital Hygiene, University of Duesseldorf Ortwin Adams, Marcel Andree, Alexander Dilthey, Torsten Feldt, Sandra Hauka, Torsten Houwaart, Björn-Erik Jensen, Detlef Kindgen-Milles, Malte Kohns Vasconcelos, Klaus Pfeffer, Tina Senff, Daniel Strelow, Jörg Timm, Andreas Walker, Tobias Wienemann
EPI_ISL_419545 hCoV-19/Germany/NRW-28/2020 Europe / Germany / North Rhine Westphalia / Duesseldorf 2020-03-15 Center of Medical Microbiology, Virology, and Hospital Hygiene, University of Duesseldorf Center of Medical Microbiology, Virology, and Hospital Hygiene, University of Duesseldorf Ortwin Adams, Marcel Andree, Alexander Dilthey, Torsten Feldt, Sandra Hauka, Torsten Houwaart, Björn-Erik Jensen, Detlef Kindgen-Milles, Malte Kohns Vasconcelos, Klaus Pfeffer, Tina Senff, Daniel Strelow, Jörg Timm, Andreas Walker, Tobias Wienemann
EPI_ISL_419546 hCoV-19/Germany/NRW-29/2020 Europe / Germany / North Rhine Westphalia / Duesseldorf 2020-03-15 Center of Medical Microbiology, Virology, and Hospital Hygiene, University of Duesseldorf Center of Medical Microbiology, Virology, and Hospital Hygiene, University of Duesseldorf Ortwin Adams, Marcel Andree, Alexander Dilthey, Torsten Feldt, Sandra Hauka, Torsten Houwaart, Björn-Erik Jensen, Detlef Kindgen-Milles, Malte Kohns Vasconcelos, Klaus Pfeffer, Tina Senff, Daniel Strelow, Jörg Timm, Andreas Walker, Tobias Wienemann
EPI_ISL_419548 hCoV-19/Germany/NRW-31/2020 Europe / Germany / North Rhine Westphalia / Duesseldorf 2020-03-15 Center of Medical Microbiology, Virology, and Hospital Hygiene, University of Duesseldorf Center of Medical Microbiology, Virology, and Hospital Hygiene, University of Duesseldorf Ortwin Adams, Marcel Andree, Alexander Dilthey, Torsten Feldt, Sandra Hauka, Torsten Houwaart, Björn-Erik Jensen, Detlef Kindgen-Milles, Malte Kohns Vasconcelos, Klaus Pfeffer, Tina Senff, Daniel Strelow, Jörg Timm, Andreas Walker, Tobias Wienemann
EPI_ISL_419549 hCoV-19/Germany/NRW-32/2020 Europe / Germany / North Rhine Westphalia / Duesseldorf 2020-03-15 Center of Medical Microbiology, Virology, and Hospital Hygiene, University of Duesseldorf Center of Medical Microbiology, Virology, and Hospital Hygiene, University of Duesseldorf Ortwin Adams, Marcel Andree, Alexander Dilthey, Torsten Feldt, Sandra Hauka, Torsten Houwaart, Björn-Erik Jensen, Detlef Kindgen-Milles, Malte Kohns Vasconcelos, Klaus Pfeffer, Tina Senff, Daniel Strelow, Jörg Timm, Andreas Walker, Tobias Wienemann
EPI_ISL_419550 hCoV-19/Germany/NRW-33/2020 Europe / Germany / North Rhine Westphalia / Duesseldorf 2020-03-16 Center of Medical Microbiology, Virology, and Hospital Hygiene, University of Duesseldorf Center of Medical Microbiology, Virology, and Hospital Hygiene, University of Duesseldorf Ortwin Adams, Marcel Andree, Alexander Dilthey, Torsten Feldt, Sandra Hauka, Torsten Houwaart, Björn-Erik Jensen, Detlef Kindgen-Milles, Malte Kohns Vasconcelos, Klaus Pfeffer, Tina Senff, Daniel Strelow, Jörg Timm, Andreas Walker, Tobias Wienemann
EPI_ISL_419551 hCoV-19/Germany/NRW-34/2020 Europe / Germany / North Rhine Westphalia / Duesseldorf 2020-03-16 Center of Medical Microbiology, Virology, and Hospital Hygiene, University of Duesseldorf Center of Medical Microbiology, Virology, and Hospital Hygiene, University of Duesseldorf Ortwin Adams, Marcel Andree, Alexander Dilthey, Torsten Feldt, Sandra Hauka, Torsten Houwaart, Björn-Erik Jensen, Detlef Kindgen-Milles, Malte Kohns Vasconcelos, Klaus Pfeffer, Tina Senff, Daniel Strelow, Jörg Timm, Andreas Walker, Tobias Wienemann
EPI_ISL_419552 hCoV-19/Germany/NRW-35/2020 Europe / Germany / North Rhine Westphalia / Duesseldorf 2020-03-16 Center of Medical Microbiology, Virology, and Hospital Hygiene, University of Duesseldorf Center of Medical Microbiology, Virology, and Hospital Hygiene, University of Duesseldorf Ortwin Adams, Marcel Andree, Alexander Dilthey, Torsten Feldt, Sandra Hauka, Torsten Houwaart, Björn-Erik Jensen, Detlef Kindgen-Milles, Malte Kohns Vasconcelos, Klaus Pfeffer, Tina Senff, Daniel Strelow, Jörg Timm, Andreas Walker, Tobias Wienemann

EPI_ISL_419553 hCoV-19/USA/RI_0520/2020 North America / USA / Rhode Island 2020-02-28 RI State Health Laboratories Pathogen Discovery, Respiratory Viruses Branch, Division of Viral Diseases, Centers for Disease Control and Prevention Ying Tao, Jing Zhang, Krista Queen, Anna Uehara, Clinton R. Paden, Yan Li, Haibin Wang, Jasmine Padilla, Justin Lee, Suxiang Tong
EPI_ISL_419554 hCoV-19/USA/CA_2602/2020 North America / USA / California 2020-02-26 California Department of Public Health Pathogen Discovery, Respiratory Viruses Branch, Division of Viral Diseases, Centers for Disease Control and Prevention Ying Tao, Jing Zhang, Krista Queen, Anna Uehara, Clinton R. Paden, Yan Li, Haibin Wang, Jasmine Padilla, Justin Lee, Suxiang Tong
EPI_ISL_419555 hCoV-19/USA/WA_5030/2020 North America / USA / Washington 2020-02-27 WA State Department of Health Pathogen Discovery, Respiratory Viruses Branch, Division of Viral Diseases, Centers for Disease Control and Prevention Ying Tao, Jing Zhang, Krista Queen, Anna Uehara, Clinton R. Paden, Yan Li, Haibin Wang, Jasmine Padilla, Justin Lee, Suxiang Tong
EPI_ISL_419556 hCoV-19/USA/GA_2741/2020 North America / USA / Georgia 2020-02-29 GA Department of Public Health Laboratory Pathogen Discovery, Respiratory Viruses Branch, Division of Viral Diseases, Centers for Disease Control and Prevention Ying Tao, Jing Zhang, Krista Queen, Anna Uehara, Clinton R. Paden, Yan Li, Haibin Wang, Jasmine Padilla, Justin Lee, Suxiang Tong
EPI_ISL_419557 hCoV-19/USA/GA_2742/2020 North America / USA / Georgia 2020-02-29 GA Department of Public Health Laboratory Pathogen Discovery, Respiratory Viruses Branch, Division of Viral Diseases, Centers for Disease Control and Prevention Ying Tao, Jing Zhang, Krista Queen, Anna Uehara, Clinton R. Paden, Yan Li, Haibin Wang, Jasmine Padilla, Justin Lee, Suxiang Tong
EPI_ISL_419558 hCoV-19/USA/OR_2656/2020 North America / USA / Oregon 2020-02-27 OR State PHL-Virology/Immunology Section Pathogen Discovery, Respiratory Viruses Branch, Division of Viral Diseases, Centers for Disease Control and Prevention Ying Tao, Jing Zhang, Krista Queen, Anna Uehara, Clinton R. Paden, Yan Li, Haibin Wang, Jasmine Padilla, Justin Lee, Suxiang Tong
EPI_ISL_419559 hCoV-19/USA/FL_5125/2020 North America / USA / Florida 2020-02-28 FL Bureau of Public Health Laboratories-Tampa Pathogen Discovery, Respiratory Viruses Branch, Division of Viral Diseases, Centers for Disease Control and Prevention Anna Uehara, Ying Tao, Jing Zhang, Krista Queen, Clinton R. Paden, Yan Li, Haibin Wang, Jasmine Padilla, Justin Lee, Suxiang Tong
EPI_ISL_419560 hCoV-19/USA/FL_5091/2020 North America / USA / Florida 2020-02-28 FL Bureau of Public Health Laboratories-Tampa Pathogen Discovery, Respiratory Viruses Branch, Division of Viral Diseases, Centers for Disease Control and Prevention Anna Uehara, Ying Tao, Jing Zhang, Krista Queen, Clinton R. Paden, Yan Li, Haibin Wang, Jasmine Padilla, Justin Lee, Suxiang Tong
EPI_ISL_419562 hCoV-19/Luxembourg/LNS0000001/2020 Europe / Luxembourg 2020-02-29 Laboratoire National de Santé, Microbiology, Virology Laboratoire National de Santé, Microbiology, Epidemiology and Microbial Genomics Anke Wienecke-Baldacchino, Ardashel Latsuzbaia, Jessica Tapp, Catherine Ragimbeau, Guillaume Fournier, Tamir Abdelrahman, Trung Nguyen Nguyen, Joel Mossong
EPI_ISL_419563 hCoV-19/Luxembourg/LNS0156959/2020 Europe / Luxembourg 2020-03-12 Laboratoire National de Santé, Microbiology, Virology Laboratoire National de Santé, Microbiology, Epidemiology and Microbial Genomics Anke Wienecke-Baldacchino, Ardashel Latsuzbaia, Jessica Tapp, Catherine Ragimbeau, Guillaume Fournier, Tamir Abdelrahman, Trung Nguyen Nguyen, Joel Mossong
EPI_ISL_419564 hCoV-19/Luxembourg/LNS0366116/2020 Europe / Luxembourg 2020-03-15 Laboratoire National de Santé, Microbiology, Virology Laboratoire National de Santé, Microbiology, Epidemiology and Microbial Genomics Anke Wienecke-Baldacchino, Ardashel Latsuzbaia, Jessica Tapp, Catherine Ragimbeau, Guillaume Fournier, Tamir Abdelrahman, Trung Nguyen Nguyen, Joel Mossong
EPI_ISL_419566 hCoV-19/Luxembourg/LNS0641910/2020 Europe / Luxembourg 2020-03-05 Laboratoire National de Santé, Microbiology, Virology Laboratoire National de Santé, Microbiology, Epidemiology and Microbial Genomics Anke Wienecke-Baldacchino, Ardashel Latsuzbaia, Jessica Tapp, Catherine Ragimbeau, Guillaume Fournier, Tamir Abdelrahman, Trung Nguyen Nguyen, Joel Mossong
EPI_ISL_419568 hCoV-19/Luxembourg/LNS0756270/2020 Europe / Luxembourg 2020-03-14 Laboratoire National de Santé, Microbiology, Virology Laboratoire National de Santé, Microbiology, Epidemiology and Microbial Genomics Anke Wienecke-Baldacchino, Ardashel Latsuzbaia, Jessica Tapp, Catherine Ragimbeau, Guillaume Fournier, Tamir Abdelrahman, Trung Nguyen Nguyen, Joel Mossong
EPI_ISL_419569 hCoV-19/Luxembourg/LNS0945359/2020 Europe / Luxembourg 2020-03-07 Laboratoire National de Santé, Microbiology, Virology Laboratoire National de Santé, Microbiology, Epidemiology and Microbial Genomics Anke Wienecke-Baldacchino, Ardashel Latsuzbaia, Jessica Tapp, Catherine Ragimbeau, Guillaume Fournier, Tamir Abdelrahman, Trung Nguyen Nguyen, Joel Mossong
EPI_ISL_419570 hCoV-19/Luxembourg/LNS1234709/2020 Europe / Luxembourg 2020-03-12 Laboratoire National de Santé, Microbiology, Virology Laboratoire National de Santé, Microbiology, Epidemiology and Microbial Genomics Anke Wienecke-Baldacchino, Ardashel Latsuzbaia, Jessica Tapp, Catherine Ragimbeau, Guillaume Fournier, Tamir Abdelrahman, Trung Nguyen Nguyen, Joel Mossong
EPI_ISL_419573 hCoV-19/Luxembourg/LNS1874423/2020 Europe / Luxembourg 2020-03-11 Laboratoire National de Santé, Microbiology, Virology Laboratoire National de Santé, Microbiology, Epidemiology and Microbial Genomics Anke Wienecke-Baldacchino, Ardashel Latsuzbaia, Jessica Tapp, Catherine Ragimbeau, Guillaume Fournier, Tamir Abdelrahman, Trung Nguyen Nguyen, Joel Mossong
EPI_ISL_419578 hCoV-19/Luxembourg/LNS2614631/2020 Europe / Luxembourg 2020-03-08 Laboratoire National de Santé, Microbiology, Virology Laboratoire National de Santé, Microbiology, Epidemiology and Microbial Genomics Anke Wienecke-Baldacchino, Ardashel Latsuzbaia, Jessica Tapp, Catherine Ragimbeau, Guillaume Fournier, Tamir Abdelrahman, Trung Nguyen Nguyen, Joel Mossong
EPI_ISL_419579 hCoV-19/Luxembourg/LNS2886370/2020 Europe / Luxembourg 2020-03-14 Laboratoire National de Santé, Microbiology, Virology Laboratoire National de Santé, Microbiology, Epidemiology and

Microbial Genomics	Anke Wienecke-Baldacchino, Ardashel Latsuzbaia, Jessica Tapp, Catherine Ragimbeau, Guillaume Fournier, Tamir Abdelrahman, Trung Nguyen Nguyen, Joel Mossong
EPI_ISL_419580	hCoV-19/Luxembourg/LNS2907333/2020	Europe / Luxembourg	2020-03-12	Laboratoire National de Santé, Microbiology, Virology	Laboratoire National de Santé, Microbiology, Epidemiology and Microbial Genomics	Anke Wienecke-Baldacchino, Ardashel Latsuzbaia, Jessica Tapp, Catherine Ragimbeau, Guillaume Fournier, Tamir Abdelrahman, Trung Nguyen Nguyen, Joel Mossong
EPI_ISL_419582	hCoV-19/Luxembourg/LNS3156434/2020	Europe / Luxembourg	2020-03-13	Laboratoire National de Santé, Microbiology, Virology	Laboratoire National de Santé, Microbiology, Epidemiology and Microbial Genomics	Anke Wienecke-Baldacchino, Ardashel Latsuzbaia, Jessica Tapp, Catherine Ragimbeau, Guillaume Fournier, Tamir Abdelrahman, Trung Nguyen Nguyen, Joel Mossong
EPI_ISL_419583	hCoV-19/Luxembourg/LNS3588186/2020	Europe / Luxembourg	2020-03-15	Laboratoire National de Santé, Microbiology, Virology	Laboratoire National de Santé, Microbiology, Epidemiology and Microbial Genomics	Anke Wienecke-Baldacchino, Ardashel Latsuzbaia, Jessica Tapp, Catherine Ragimbeau, Guillaume Fournier, Tamir Abdelrahman, Trung Nguyen Nguyen, Joel Mossong
EPI_ISL_419584	hCoV-19/Luxembourg/LNS3711853/2020	Europe / Luxembourg	2020-03-13	Laboratoire National de Santé, Microbiology, Virology	Laboratoire National de Santé, Microbiology, Epidemiology and Microbial Genomics	Anke Wienecke-Baldacchino, Ardashel Latsuzbaia, Jessica Tapp, Catherine Ragimbeau, Guillaume Fournier, Tamir Abdelrahman, Trung Nguyen Nguyen, Joel Mossong
EPI_ISL_419585	hCoV-19/Luxembourg/LNS3879580/2020	Europe / Luxembourg	2020-03-14	Laboratoire National de Santé, Microbiology, Virology	Laboratoire National de Santé, Microbiology, Epidemiology and Microbial Genomics	Anke Wienecke-Baldacchino, Ardashel Latsuzbaia, Jessica Tapp, Catherine Ragimbeau, Guillaume Fournier, Tamir Abdelrahman, Trung Nguyen Nguyen, Joel Mossong
EPI_ISL_419586	hCoV-19/Luxembourg/LNS4134806/2020	Europe / Luxembourg	2020-03-14	Laboratoire National de Santé, Microbiology, Virology	Laboratoire National de Santé, Microbiology, Epidemiology and Microbial Genomics	Anke Wienecke-Baldacchino, Ardashel Latsuzbaia, Jessica Tapp, Catherine Ragimbeau, Guillaume Fournier, Tamir Abdelrahman, Trung Nguyen Nguyen, Joel Mossong
EPI_ISL_419587	hCoV-19/Luxembourg/LNS4691488/2020	Europe / Luxembourg	2020-03-11	Laboratoire National de Santé, Microbiology, Virology	Laboratoire National de Santé, Microbiology, Epidemiology and Microbial Genomics	Anke Wienecke-Baldacchino, Ardashel Latsuzbaia, Jessica Tapp, Catherine Ragimbeau, Guillaume Fournier, Tamir Abdelrahman, Trung Nguyen Nguyen, Joel Mossong
EPI_ISL_419588	hCoV-19/Luxembourg/LNS4836560/2020	Europe / Luxembourg	2020-03-16	Laboratoire National de Santé, Microbiology, Virology	Laboratoire National de Santé, Microbiology, Epidemiology and Microbial Genomics	Anke Wienecke-Baldacchino, Ardashel Latsuzbaia, Jessica Tapp, Catherine Ragimbeau, Guillaume Fournier, Tamir Abdelrahman, Trung Nguyen Nguyen, Joel Mossong
EPI_ISL_419589	hCoV-19/Luxembourg/LNS4845603/2020	Europe / Luxembourg	2020-03-15	Laboratoire National de Santé, Microbiology, Virology	Laboratoire National de Santé, Microbiology, Epidemiology and Microbial Genomics	Anke Wienecke-Baldacchino, Ardashel Latsuzbaia, Jessica Tapp, Catherine Ragimbeau, Guillaume Fournier, Tamir Abdelrahman, Trung Nguyen Nguyen, Joel Mossong
EPI_ISL_419590	hCoV-19/Luxembourg/LNS5032431/2020	Europe / Luxembourg	2020-03-11	Laboratoire National de Santé, Microbiology, Virology	Laboratoire National de Santé, Microbiology, Epidemiology and Microbial Genomics	Anke Wienecke-Baldacchino, Ardashel Latsuzbaia, Jessica Tapp, Catherine Ragimbeau, Guillaume Fournier, Tamir Abdelrahman, Trung Nguyen Nguyen, Joel Mossong
EPI_ISL_419591	hCoV-19/Luxembourg/LNS5228153/2020	Europe / Luxembourg	2020-03-14	Laboratoire National de Santé, Microbiology, Virology	Laboratoire National de Santé, Microbiology, Epidemiology and Microbial Genomics	Anke Wienecke-Baldacchino, Ardashel Latsuzbaia, Jessica Tapp, Catherine Ragimbeau, Guillaume Fournier, Tamir Abdelrahman, Trung Nguyen Nguyen, Joel Mossong
EPI_ISL_419592	hCoV-19/Luxembourg/LNS5440244/2020	Europe / Luxembourg	2020-03-15	Laboratoire National de Santé, Microbiology, Virology	Laboratoire National de Santé, Microbiology, Epidemiology and Microbial Genomics	Anke Wienecke-Baldacchino, Ardashel Latsuzbaia, Jessica Tapp, Catherine Ragimbeau, Guillaume Fournier, Tamir Abdelrahman, Trung Nguyen Nguyen, Joel Mossong
EPI_ISL_419593	hCoV-19/Luxembourg/LNS6603085/2020	Europe / Luxembourg	2020-03-16	Laboratoire National de Santé, Microbiology, Virology	Laboratoire National de Santé, Microbiology, Epidemiology and Microbial Genomics	Anke Wienecke-Baldacchino, Ardashel Latsuzbaia, Jessica Tapp, Catherine Ragimbeau, Guillaume Fournier, Tamir Abdelrahman, Trung Nguyen Nguyen, Joel Mossong
EPI_ISL_419594	hCoV-19/Luxembourg/LNS7537751/2020	Europe / Luxembourg	2020-03-16	Laboratoire National de Santé, Microbiology, Virology	Laboratoire National de Santé, Microbiology, Epidemiology and Microbial Genomics	Anke Wienecke-Baldacchino, Ardashel Latsuzbaia, Jessica Tapp, Catherine Ragimbeau, Guillaume Fournier, Tamir Abdelrahman, Trung Nguyen Nguyen, Joel Mossong
EPI_ISL_419595	hCoV-19/Luxembourg/LNS7753431/2020	Europe / Luxembourg	2020-03-12	Laboratoire National de Santé, Microbiology, Virology	Laboratoire National de Santé, Microbiology, Epidemiology and Microbial Genomics	Anke Wienecke-Baldacchino, Ardashel Latsuzbaia, Jessica Tapp, Catherine Ragimbeau, Guillaume Fournier, Tamir Abdelrahman, Trung Nguyen Nguyen, Joel Mossong
EPI_ISL_419596	hCoV-19/Luxembourg/LNS7866283/2020	Europe / Luxembourg	2020-03-12	Laboratoire National de Santé, Microbiology, Virology	Laboratoire National de Santé, Microbiology, Epidemiology and Microbial Genomics	Anke Wienecke-Baldacchino, Ardashel Latsuzbaia, Jessica Tapp, Catherine Ragimbeau, Guillaume Fournier, Tamir Abdelrahman, Trung Nguyen Nguyen, Joel Mossong
EPI_ISL_419597	hCoV-19/Luxembourg/LNS7928872/2020	Europe / Luxembourg	2020-03-15	Laboratoire National de Santé, Microbiology, Virology	Laboratoire National de Santé, Microbiology, Epidemiology and Microbial Genomics	Anke Wienecke-Baldacchino, Ardashel Latsuzbaia, Jessica Tapp, Catherine Ragimbeau, Guillaume Fournier, Tamir Abdelrahman, Trung Nguyen Nguyen, Joel Mossong

EPI_ISL_419598 hCoV-19/Luxembourg/LNS8188502/2020 Europe / Luxembourg 2020-03-14 Laboratoire National de Santé, Microbiology, Virology Laboratoire National de Santé, Microbiology, Epidemiology and Microbial Genomics Anke Wienecke-Baldacchino, Ardashel Latsuzbaia, Jessica Tapp, Catherine Ragimbeau, Guillaume Fournier, Tamir Abdelrahman, Trung Nguyen Nguyen, Joel Mossong
EPI_ISL_419599 hCoV-19/Luxembourg/LNS8639502/2020 Europe / Luxembourg 2020-03-16 Laboratoire National de Santé, Microbiology, Virology Laboratoire National de Santé, Microbiology, Epidemiology and Microbial Genomics Anke Wienecke-Baldacchino, Ardashel Latsuzbaia, Jessica Tapp, Catherine Ragimbeau, Guillaume Fournier, Tamir Abdelrahman, Trung Nguyen Nguyen, Joel Mossong
EPI_ISL_419800 hCoV-19/Australia/VIC89/2020 Oceania / Australia / Victoria 2020-03-15 Victorian Infectious Diseases Reference Laboratory (VIDRL) Victorian Infectious Diseases Reference Laboratory and Microbiological Diagnostic Unit Public Health Laboratory, Doherty Institute Caly L., Seemann T., Sait, M., Schultz M., Druce J., Sherry, N.
EPI_ISL_419801 hCoV-19/Australia/VIC90/2020 Oceania / Australia / Victoria 2020-03-15 Victorian Infectious Diseases Reference Laboratory (VIDRL) Victorian Infectious Diseases Reference Laboratory and Microbiological Diagnostic Unit Public Health Laboratory, Doherty Institute Caly L., Seemann T., Sait, M., Schultz M., Druce J., Sherry, N.
EPI_ISL_419802 hCoV-19/Australia/VIC45/2020 Oceania / Australia / Victoria 2020-03-11 Victorian Infectious Diseases Reference Laboratory (VIDRL) Victorian Infectious Diseases Reference Laboratory and Microbiological Diagnostic Unit Public Health Laboratory, Doherty Institute Caly L., Seemann T., Sait, M., Schultz M., Druce J., Sherry, N.
EPI_ISL_419803 hCoV-19/Australia/VIC84/2020 Oceania / Australia / Victoria 2020-03-14 Victorian Infectious Diseases Reference Laboratory (VIDRL) Victorian Infectious Diseases Reference Laboratory and Microbiological Diagnostic Unit Public Health Laboratory, Doherty Institute Caly L., Seemann T., Sait, M., Schultz M., Druce J., Sherry, N.
EPI_ISL_419804 hCoV-19/Australia/VIC91/2020 Oceania / Australia / Victoria 2020-03-15 Victorian Infectious Diseases Reference Laboratory (VIDRL) Victorian Infectious Diseases Reference Laboratory and Microbiological Diagnostic Unit Public Health Laboratory, Doherty Institute Caly L., Seemann T., Sait, M., Schultz M., Druce J., Sherry, N.
EPI_ISL_419805 hCoV-19/Australia/VIC94/2020 Oceania / Australia / Victoria 2020-03-16 Victorian Infectious Diseases Reference Laboratory (VIDRL) Victorian Infectious Diseases Reference Laboratory and Microbiological Diagnostic Unit Public Health Laboratory, Doherty Institute Caly L., Seemann T., Sait, M., Schultz M., Druce J., Sherry, N.
EPI_ISL_419806 hCoV-19/Australia/VIC95/2020 Oceania / Australia / Victoria 2020-03-16 Victorian Infectious Diseases Reference Laboratory (VIDRL) Victorian Infectious Diseases Reference Laboratory and Microbiological Diagnostic Unit Public Health Laboratory, Doherty Institute Caly L., Seemann T., Sait, M., Schultz M., Druce J., Sherry, N.
EPI_ISL_419807 hCoV-19/Australia/VIC96/2020 Oceania / Australia / Victoria 2020-03-16 Victorian Infectious Diseases Reference Laboratory (VIDRL) Victorian Infectious Diseases Reference Laboratory and Microbiological Diagnostic Unit Public Health Laboratory, Doherty Institute Caly L., Seemann T., Sait, M., Schultz M., Druce J., Sherry, N.
EPI_ISL_419808 hCoV-19/Australia/VIC97/2020 Oceania / Australia / Victoria 2020-03-16 Victorian Infectious Diseases Reference Laboratory (VIDRL) Victorian Infectious Diseases Reference Laboratory and Microbiological Diagnostic Unit Public Health Laboratory, Doherty Institute Caly L., Seemann T., Sait, M., Schultz M., Druce J., Sherry, N.
EPI_ISL_419809 hCoV-19/Australia/VIC92/2020 Oceania / Australia / Victoria 2020-03-15 Victorian Infectious Diseases Reference Laboratory (VIDRL) Victorian Infectious Diseases Reference Laboratory and Microbiological Diagnostic Unit Public Health Laboratory, Doherty Institute Caly L., Seemann T., Sait, M., Schultz M., Druce J., Sherry, N.
EPI_ISL_419810 hCoV-19/Australia/VIC98/2020 Oceania / Australia / Victoria 2020-03-16 Victorian Infectious Diseases Reference Laboratory (VIDRL) Victorian Infectious Diseases Reference Laboratory and Microbiological Diagnostic Unit Public Health Laboratory, Doherty Institute Caly L., Seemann T., Sait, M., Schultz M., Druce J., Sherry, N.
EPI_ISL_419811 hCoV-19/Australia/VIC99/2020 Oceania / Australia / Victoria 2020-03-16 Victorian Infectious Diseases Reference Laboratory (VIDRL) Victorian Infectious Diseases Reference Laboratory and Microbiological Diagnostic Unit Public Health Laboratory, Doherty Institute Caly L., Seemann T., Sait, M., Schultz M., Druce J., Sherry, N.
EPI_ISL_419812 hCoV-19/Australia/VIC100/2020 Oceania / Australia / Victoria 2020-03-16 Victorian Infectious Diseases Reference Laboratory (VIDRL) Victorian Infectious Diseases Reference Laboratory and Microbiological Diagnostic Unit Public Health Laboratory, Doherty Institute Caly L., Seemann T., Sait, M., Schultz M., Druce J., Sherry, N.
EPI_ISL_419813 hCoV-19/Australia/VIC101/2020 Oceania / Australia / Victoria 2020-03-16 Victorian Infectious Diseases Reference Laboratory (VIDRL) Victorian Infectious Diseases Reference Laboratory and Microbiological Diagnostic Unit Public Health Laboratory, Doherty Institute Caly L., Seemann T., Sait, M., Schultz M., Druce J., Sherry, N.
EPI_ISL_419814 hCoV-19/Australia/VIC103/2020 Oceania / Australia / Victoria 2020-03-17 Victorian Infectious Diseases Reference Laboratory (VIDRL) Victorian Infectious Diseases Reference Laboratory and Microbiological Diagnostic Unit Public Health Laboratory, Doherty Institute Caly L., Seemann T., Sait, M., Schultz M., Druce J., Sherry, N.
EPI_ISL_419816 hCoV-19/Australia/VIC46/2020 Oceania / Australia / Victoria 2020-03-11 Victorian Infectious Diseases Reference Laboratory (VIDRL) Victorian Infectious Diseases Reference Laboratory and

Microbiological Diagnostic Unit Public Health Laboratory, Doherty Institute     Caly L., Seemann T., Sait, M., Schultz M., Druce J., Sherry, N.
EPI_ISL_419817  hCoV-19/Australia/VIC102/2020   Oceania / Australia / Victoria  2020-03-16      Victorian Infectious Diseases Reference Laboratory (VIDRL)        Victorian Infectious Diseases Reference Laboratory and Microbiological Diagnostic Unit Public Health Laboratory, Doherty Institute     Caly L., Seemann T., Sait, M., Schultz M., Druce J., Sherry, N.
EPI_ISL_419818  hCoV-19/Australia/VIC104/2020   Oceania / Australia / Victoria  2020-03-17      Victorian Infectious Diseases Reference Laboratory (VIDRL)        Victorian Infectious Diseases Reference Laboratory and Microbiological Diagnostic Unit Public Health Laboratory, Doherty Institute     Caly L., Seemann T., Sait, M., Schultz M., Druce J., Sherry, N.
EPI_ISL_419819  hCoV-19/Australia/VIC105/2020   Oceania / Australia / Victoria  2020-03-17      Victorian Infectious Diseases Reference Laboratory (VIDRL)        Victorian Infectious Diseases Reference Laboratory and Microbiological Diagnostic Unit Public Health Laboratory, Doherty Institute     Caly L., Seemann T., Sait, M., Schultz M., Druce J., Sherry, N.
EPI_ISL_419820  hCoV-19/Australia/VIC106/2020   Oceania / Australia / Victoria  2020-03-17      Victorian Infectious Diseases Reference Laboratory (VIDRL)        Victorian Infectious Diseases Reference Laboratory and Microbiological Diagnostic Unit Public Health Laboratory, Doherty Institute     Caly L., Seemann T., Sait, M., Schultz M., Druce J., Sherry, N.
EPI_ISL_419821  hCoV-19/Australia/VIC112/2020   Oceania / Australia / Victoria  2020-03-18      Victorian Infectious Diseases Reference Laboratory (VIDRL)        Victorian Infectious Diseases Reference Laboratory and Microbiological Diagnostic Unit Public Health Laboratory, Doherty Institute     Caly L., Seemann T., Sait, M., Schultz M., Druce J., Sherry, N.
EPI_ISL_419822  hCoV-19/Australia/VIC113/2020   Oceania / Australia / Victoria  2020-03-18      Victorian Infectious Diseases Reference Laboratory (VIDRL)        Victorian Infectious Diseases Reference Laboratory and Microbiological Diagnostic Unit Public Health Laboratory, Doherty Institute     Caly L., Seemann T., Sait, M., Schultz M., Druce J., Sherry, N.
EPI_ISL_419823  hCoV-19/Australia/VIC130/2020   Oceania / Australia / Victoria  2020-03-21      Microbiological Diagnostic Unit Public Health Laboratory       Microbiological Diagnostic Unit Public Health Laboratory     Seemann T., Schultz M., Sait, M., Sherry, N.
EPI_ISL_419824  hCoV-19/Australia/VIC131/2020   Oceania / Australia / Victoria  2020-03-21      Microbiological Diagnostic Unit Public Health Laboratory       Microbiological Diagnostic Unit Public Health Laboratory     Seemann T., Schultz M., Sait, M., Sherry, N.
EPI_ISL_419825  hCoV-19/Australia/VIC132/2020   Oceania / Australia / Victoria  2020-03-22      Microbiological Diagnostic Unit Public Health Laboratory       Microbiological Diagnostic Unit Public Health Laboratory     Seemann T., Schultz M., Sait, M., Sherry, N.
EPI_ISL_419826  hCoV-19/Australia/VIC51/2020    Oceania / Australia / Victoria  2020-03-12      Victorian Infectious Diseases Reference Laboratory (VIDRL)        Victorian Infectious Diseases Reference Laboratory and Microbiological Diagnostic Unit Public Health Laboratory, Doherty Institute     Caly L., Seemann T., Sait, M., Schultz M., Druce J., Sherry, N.
EPI_ISL_419827  hCoV-19/Australia/VIC134/2020   Oceania / Australia / Victoria  2020-03-22      Microbiological Diagnostic Unit Public Health Laboratory       Microbiological Diagnostic Unit Public Health Laboratory     Seemann T., Schultz M., Sait, M., Sherry, N.
EPI_ISL_419828  hCoV-19/Australia/VIC135/2020   Oceania / Australia / Victoria  2020-03-22      Microbiological Diagnostic Unit Public Health Laboratory       Microbiological Diagnostic Unit Public Health Laboratory     Seemann T., Schultz M., Sait, M., Sherry, N.
EPI_ISL_419850  hCoV-19/Australia/VIC153/2020   Oceania / Australia / Victoria  2020-03-17      Victorian Infectious Diseases Reference Laboratory (VIDRL)        Victorian Infectious Diseases Reference Laboratory and Microbiological Diagnostic Unit Public Health Laboratory, Doherty Institute     Caly L., Seemann T., Sait, M., Schultz M., Druce J., Sherry, N.
EPI_ISL_419851  hCoV-19/Australia/VIC154/2020   Oceania / Australia / Victoria  2020-03-17      Victorian Infectious Diseases Reference Laboratory (VIDRL)        Victorian Infectious Diseases Reference Laboratory and Microbiological Diagnostic Unit Public Health Laboratory, Doherty Institute     Caly L., Seemann T., Sait, M., Schultz M., Druce J., Sherry, N.
EPI_ISL_419852  hCoV-19/Australia/VIC155/2020   Oceania / Australia / Victoria  2020-03-17      Victorian Infectious Diseases Reference Laboratory (VIDRL)        Victorian Infectious Diseases Reference Laboratory and Microbiological Diagnostic Unit Public Health Laboratory, Doherty Institute     Caly L., Seemann T., Sait, M., Schultz M., Druce J., Sherry, N.
EPI_ISL_419853  hCoV-19/Australia/VIC156/2020   Oceania / Australia / Victoria  2020-03-17      Victorian Infectious Diseases Reference Laboratory (VIDRL)        Victorian Infectious Diseases Reference Laboratory and Microbiological Diagnostic Unit Public Health Laboratory, Doherty Institute     Caly L., Seemann T., Sait, M., Schultz M., Druce J., Sherry, N.
EPI_ISL_419854  hCoV-19/Australia/VIC157/2020   Oceania / Australia / Victoria  2020-03-17      Victorian Infectious Diseases Reference Laboratory (VIDRL)        Victorian Infectious Diseases Reference Laboratory and Microbiological Diagnostic Unit Public Health Laboratory, Doherty Institute     Caly L., Seemann T., Sait, M., Schultz M., Druce J., Sherry, N.
EPI_ISL_419855  hCoV-19/Australia/VIC158/2020   Oceania / Australia / Victoria  2020-03-18      Victorian Infectious Diseases Reference Laboratory (VIDRL)        Victorian Infectious Diseases Reference Laboratory and Microbiological Diagnostic Unit Public Health Laboratory, Doherty Institute     Caly L., Seemann T., Sait, M., Schultz M., Druce J., Sherry, N.
EPI_ISL_419856  hCoV-19/Australia/VIC159/2020   Oceania / Australia / Victoria  2020-03-18      Victorian

Infectious Diseases Reference Laboratory (VIDRL)	Victorian Infectious Diseases Reference Laboratory and Microbiological Diagnostic Unit Public Health Laboratory, Doherty Institute	Caly L., Seemann T., Sait, M., Schultz M., Druce J., Sherry, N.
EPI_ISL_419857	hCoV-19/Australia/VIC160/2020	Oceania / Australia / Victoria	2020-03-18	Victorian Infectious Diseases Reference Laboratory (VIDRL)	Victorian Infectious Diseases Reference Laboratory and Microbiological Diagnostic Unit Public Health Laboratory, Doherty Institute	Caly L., Seemann T., Sait, M., Schultz M., Druce J., Sherry, N.
EPI_ISL_419859	hCoV-19/Australia/VIC162/2020	Oceania / Australia / Victoria	2020-03-18	Victorian Infectious Diseases Reference Laboratory (VIDRL)	Victorian Infectious Diseases Reference Laboratory and Microbiological Diagnostic Unit Public Health Laboratory, Doherty Institute	Caly L., Seemann T., Sait, M., Schultz M., Druce J., Sherry, N.
EPI_ISL_419860	hCoV-19/Australia/VIC163/2020	Oceania / Australia / Victoria	2020-03-18	Victorian Infectious Diseases Reference Laboratory (VIDRL)	Victorian Infectious Diseases Reference Laboratory and Microbiological Diagnostic Unit Public Health Laboratory, Doherty Institute	Caly L., Seemann T., Sait, M., Schultz M., Druce J., Sherry, N.
EPI_ISL_419861	hCoV-19/Australia/VIC164/2020	Oceania / Australia / Victoria	2020-03-18	Victorian Infectious Diseases Reference Laboratory (VIDRL)	Victorian Infectious Diseases Reference Laboratory and Microbiological Diagnostic Unit Public Health Laboratory, Doherty Institute	Caly L., Seemann T., Sait, M., Schultz M., Druce J., Sherry, N.
EPI_ISL_419862	hCoV-19/Australia/VIC165/2020	Oceania / Australia / Victoria	2020-03-18	Victorian Infectious Diseases Reference Laboratory (VIDRL)	Victorian Infectious Diseases Reference Laboratory and Microbiological Diagnostic Unit Public Health Laboratory, Doherty Institute	Caly L., Seemann T., Sait, M., Schultz M., Druce J., Sherry, N.
EPI_ISL_419863	hCoV-19/Australia/VIC166/2020	Oceania / Australia / Victoria	2020-03-18	Victorian Infectious Diseases Reference Laboratory (VIDRL)	Victorian Infectious Diseases Reference Laboratory and Microbiological Diagnostic Unit Public Health Laboratory, Doherty Institute	Caly L., Seemann T., Sait, M., Schultz M., Druce J., Sherry, N.
EPI_ISL_419864	hCoV-19/Australia/VIC167/2020	Oceania / Australia / Victoria	2020-03-18	Victorian Infectious Diseases Reference Laboratory (VIDRL)	Victorian Infectious Diseases Reference Laboratory and Microbiological Diagnostic Unit Public Health Laboratory, Doherty Institute	Caly L., Seemann T., Sait, M., Schultz M., Druce J., Sherry, N.
EPI_ISL_419865	hCoV-19/Australia/VIC168/2020	Oceania / Australia / Victoria	2020-03-18	Victorian Infectious Diseases Reference Laboratory (VIDRL)	Victorian Infectious Diseases Reference Laboratory and Microbiological Diagnostic Unit Public Health Laboratory, Doherty Institute	Caly L., Seemann T., Sait, M., Schultz M., Druce J., Sherry, N.
EPI_ISL_419866	hCoV-19/Australia/VIC169/2020	Oceania / Australia / Victoria	2020-03-18	Victorian Infectious Diseases Reference Laboratory (VIDRL)	Victorian Infectious Diseases Reference Laboratory and Microbiological Diagnostic Unit Public Health Laboratory, Doherty Institute	Caly L., Seemann T., Sait, M., Schultz M., Druce J., Sherry, N.
EPI_ISL_419867	hCoV-19/Australia/VIC170/2020	Oceania / Australia / Victoria	2020-03-18	Victorian Infectious Diseases Reference Laboratory (VIDRL)	Victorian Infectious Diseases Reference Laboratory and Microbiological Diagnostic Unit Public Health Laboratory, Doherty Institute	Caly L., Seemann T., Sait, M., Schultz M., Druce J., Sherry, N.
EPI_ISL_419868	hCoV-19/Australia/VIC171/2020	Oceania / Australia / Victoria	2020-03-18	Victorian Infectious Diseases Reference Laboratory (VIDRL)	Victorian Infectious Diseases Reference Laboratory and Microbiological Diagnostic Unit Public Health Laboratory, Doherty Institute	Caly L., Seemann T., Sait, M., Schultz M., Druce J., Sherry, N.
EPI_ISL_419869	hCoV-19/Australia/VIC172/2020	Oceania / Australia / Victoria	2020-03-18	Victorian Infectious Diseases Reference Laboratory (VIDRL)	Victorian Infectious Diseases Reference Laboratory and Microbiological Diagnostic Unit Public Health Laboratory, Doherty Institute	Caly L., Seemann T., Sait, M., Schultz M., Druce J., Sherry, N.
EPI_ISL_419870	hCoV-19/Australia/VIC173/2020	Oceania / Australia / Victoria	2020-03-18	Victorian Infectious Diseases Reference Laboratory (VIDRL)	Victorian Infectious Diseases Reference Laboratory and Microbiological Diagnostic Unit Public Health Laboratory, Doherty Institute	Caly L., Seemann T., Sait, M., Schultz M., Druce J., Sherry, N.
EPI_ISL_419871	hCoV-19/Australia/VIC174/2020	Oceania / Australia / Victoria	2020-03-18	Victorian Infectious Diseases Reference Laboratory (VIDRL)	Victorian Infectious Diseases Reference Laboratory and Microbiological Diagnostic Unit Public Health Laboratory, Doherty Institute	Caly L., Seemann T., Sait, M., Schultz M., Druce J., Sherry, N.
EPI_ISL_419872	hCoV-19/Australia/VIC175/2020	Oceania / Australia / Victoria	2020-03-18	Victorian Infectious Diseases Reference Laboratory (VIDRL)	Victorian Infectious Diseases Reference Laboratory and Microbiological Diagnostic Unit Public Health Laboratory, Doherty Institute	Caly L., Seemann T., Sait, M., Schultz M., Druce J., Sherry, N.
EPI_ISL_419873	hCoV-19/Australia/VIC176/2020	Oceania / Australia / Victoria	2020-03-18	Victorian Infectious Diseases Reference Laboratory (VIDRL)	Victorian Infectious Diseases Reference Laboratory and Microbiological Diagnostic Unit Public Health Laboratory, Doherty Institute	Caly L., Seemann T., Sait, M., Schultz M., Druce J., Sherry, N.
EPI_ISL_419874	hCoV-19/Australia/VIC177/2020	Oceania / Australia / Victoria	2020-03-18	Victorian Infectious Diseases Reference Laboratory (VIDRL)	Victorian Infectious Diseases Reference Laboratory and Microbiological Diagnostic Unit Public Health Laboratory, Doherty Institute	Caly L., Seemann T., Sait, M.,

Schultz M., Druce J., Sherry, N.
EPI_ISL_419875 hCoV-19/Australia/VIC178/2020 Oceania / Australia / Victoria 2020-03-18 Victorian Infectious Diseases Reference Laboratory (VIDRL) Victorian Infectious Diseases Reference Laboratory and Microbiological Diagnostic Unit Public Health Laboratory, Doherty Institute Caly L., Seemann T., Sait, M., Schultz M., Druce J., Sherry, N.
EPI_ISL_419876 hCoV-19/Australia/VIC179/2020 Oceania / Australia / Victoria 2020-03-18 Victorian Infectious Diseases Reference Laboratory (VIDRL) Victorian Infectious Diseases Reference Laboratory and Microbiological Diagnostic Unit Public Health Laboratory, Doherty Institute Caly L., Seemann T., Sait, M., Schultz M., Druce J., Sherry, N.
EPI_ISL_419877 hCoV-19/Australia/VIC180/2020 Oceania / Australia / Victoria 2020-03-19 Victorian Infectious Diseases Reference Laboratory (VIDRL) Victorian Infectious Diseases Reference Laboratory and Microbiological Diagnostic Unit Public Health Laboratory, Doherty Institute Caly L., Seemann T., Sait, M., Schultz M., Druce J., Sherry, N.
EPI_ISL_419878 hCoV-19/Australia/VIC181/2020 Oceania / Australia / Victoria 2020-03-19 Victorian Infectious Diseases Reference Laboratory (VIDRL) Victorian Infectious Diseases Reference Laboratory and Microbiological Diagnostic Unit Public Health Laboratory, Doherty Institute Caly L., Seemann T., Sait, M., Schultz M., Druce J., Sherry, N.
EPI_ISL_419879 hCoV-19/Australia/VIC183/2020 Oceania / Australia / Victoria 2020-03-19 Victorian Infectious Diseases Reference Laboratory (VIDRL) Victorian Infectious Diseases Reference Laboratory and Microbiological Diagnostic Unit Public Health Laboratory, Doherty Institute Caly L., Seemann T., Sait, M., Schultz M., Druce J., Sherry, N.
EPI_ISL_419880 hCoV-19/Australia/VIC184/2020 Oceania / Australia / Victoria 2020-03-19 Victorian Infectious Diseases Reference Laboratory (VIDRL) Victorian Infectious Diseases Reference Laboratory and Microbiological Diagnostic Unit Public Health Laboratory, Doherty Institute Caly L., Seemann T., Sait, M., Schultz M., Druce J., Sherry, N.
EPI_ISL_419881 hCoV-19/Australia/VIC185/2020 Oceania / Australia / Victoria 2020-03-19 Victorian Infectious Diseases Reference Laboratory (VIDRL) Victorian Infectious Diseases Reference Laboratory and Microbiological Diagnostic Unit Public Health Laboratory, Doherty Institute Caly L., Seemann T., Sait, M., Schultz M., Druce J., Sherry, N.
EPI_ISL_419882 hCoV-19/Australia/VIC186/2020 Oceania / Australia / Victoria 2020-03-19 Victorian Infectious Diseases Reference Laboratory (VIDRL) Victorian Infectious Diseases Reference Laboratory and Microbiological Diagnostic Unit Public Health Laboratory, Doherty Institute Caly L., Seemann T., Sait, M., Schultz M., Druce J., Sherry, N.
EPI_ISL_419883 hCoV-19/Australia/VIC187/2020 Oceania / Australia / Victoria 2020-03-19 Victorian Infectious Diseases Reference Laboratory (VIDRL) Victorian Infectious Diseases Reference Laboratory and Microbiological Diagnostic Unit Public Health Laboratory, Doherty Institute Caly L., Seemann T., Sait, M., Schultz M., Druce J., Sherry, N.
EPI_ISL_419884 hCoV-19/Australia/VIC188/2020 Oceania / Australia / Victoria 2020-03-19 Victorian Infectious Diseases Reference Laboratory (VIDRL) Victorian Infectious Diseases Reference Laboratory and Microbiological Diagnostic Unit Public Health Laboratory, Doherty Institute Caly L., Seemann T., Sait, M., Schultz M., Druce J., Sherry, N.
EPI_ISL_419885 hCoV-19/Australia/VIC189/2020 Oceania / Australia / Victoria 2020-03-19 Victorian Infectious Diseases Reference Laboratory (VIDRL) Victorian Infectious Diseases Reference Laboratory and Microbiological Diagnostic Unit Public Health Laboratory, Doherty Institute Caly L., Seemann T., Sait, M., Schultz M., Druce J., Sherry, N.
EPI_ISL_419886 hCoV-19/Australia/VIC190/2020 Oceania / Australia / Victoria 2020-03-19 Victorian Infectious Diseases Reference Laboratory (VIDRL) Victorian Infectious Diseases Reference Laboratory and Microbiological Diagnostic Unit Public Health Laboratory, Doherty Institute Caly L., Seemann T., Sait, M., Schultz M., Druce J., Sherry, N.
EPI_ISL_419887 hCoV-19/Australia/VIC191/2020 Oceania / Australia / Victoria 2020-03-19 Victorian Infectious Diseases Reference Laboratory (VIDRL) Victorian Infectious Diseases Reference Laboratory and Microbiological Diagnostic Unit Public Health Laboratory, Doherty Institute Caly L., Seemann T., Sait, M., Schultz M., Druce J., Sherry, N.
EPI_ISL_419888 hCoV-19/Australia/VIC192/2020 Oceania / Australia / Victoria 2020-03-19 Victorian Infectious Diseases Reference Laboratory (VIDRL) Victorian Infectious Diseases Reference Laboratory and Microbiological Diagnostic Unit Public Health Laboratory, Doherty Institute Caly L., Seemann T., Sait, M., Schultz M., Druce J., Sherry, N.
EPI_ISL_419889 hCoV-19/Australia/VIC193/2020 Oceania / Australia / Victoria 2020-03-19 Victorian Infectious Diseases Reference Laboratory (VIDRL) Victorian Infectious Diseases Reference Laboratory and Microbiological Diagnostic Unit Public Health Laboratory, Doherty Institute Caly L., Seemann T., Sait, M., Schultz M., Druce J., Sherry, N.
EPI_ISL_419890 hCoV-19/Australia/VIC194/2020 Oceania / Australia / Victoria 2020-03-19 Victorian Infectious Diseases Reference Laboratory (VIDRL) Victorian Infectious Diseases Reference Laboratory and Microbiological Diagnostic Unit Public Health Laboratory, Doherty Institute Caly L., Seemann T., Sait, M., Schultz M., Druce J., Sherry, N.
EPI_ISL_419891 hCoV-19/Australia/VIC195/2020 Oceania / Australia / Victoria 2020-03-19 Victorian Infectious Diseases Reference Laboratory (VIDRL) Victorian Infectious Diseases Reference Laboratory and Microbiological Diagnostic Unit Public Health Laboratory, Doherty Institute Caly L., Seemann T., Sait, M., Schultz M., Druce J., Sherry, N.
EPI_ISL_419892 hCoV-19/Australia/VIC196/2020 Oceania / Australia / Victoria 2020-03-19 Victorian

Infectious Diseases Reference Laboratory (VIDRL) Victorian Infectious Diseases Reference Laboratory and Microbiological Diagnostic Unit Public Health Laboratory, Doherty Institute Caly L., Seemann T., Sait, M., Schultz M., Druce J., Sherry, N.
EPI_ISL_419893 hCoV-19/Australia/VIC197/2020 Oceania / Australia / Victoria 2020-03-19 Victorian Infectious Diseases Reference Laboratory (VIDRL) Victorian Infectious Diseases Reference Laboratory and Microbiological Diagnostic Unit Public Health Laboratory, Doherty Institute Caly L., Seemann T., Sait, M., Schultz M., Druce J., Sherry, N.
EPI_ISL_419894 hCoV-19/Australia/VIC198/2020 Oceania / Australia / Victoria 2020-03-19 Victorian Infectious Diseases Reference Laboratory (VIDRL) Victorian Infectious Diseases Reference Laboratory and Microbiological Diagnostic Unit Public Health Laboratory, Doherty Institute Caly L., Seemann T., Sait, M., Schultz M., Druce J., Sherry, N.
EPI_ISL_419895 hCoV-19/Australia/VIC199/2020 Oceania / Australia / Victoria 2020-03-19 Victorian Infectious Diseases Reference Laboratory (VIDRL) Victorian Infectious Diseases Reference Laboratory and Microbiological Diagnostic Unit Public Health Laboratory, Doherty Institute Caly L., Seemann T., Sait, M., Schultz M., Druce J., Sherry, N.
EPI_ISL_419896 hCoV-19/Australia/VIC201/2020 Oceania / Australia / Victoria 2020-03-19 Victorian Infectious Diseases Reference Laboratory (VIDRL) Victorian Infectious Diseases Reference Laboratory and Microbiological Diagnostic Unit Public Health Laboratory, Doherty Institute Caly L., Seemann T., Sait, M., Schultz M., Druce J., Sherry, N.
EPI_ISL_419897 hCoV-19/Australia/VIC202/2020 Oceania / Australia / Victoria 2020-03-19 Victorian Infectious Diseases Reference Laboratory (VIDRL) Victorian Infectious Diseases Reference Laboratory and Microbiological Diagnostic Unit Public Health Laboratory, Doherty Institute Caly L., Seemann T., Sait, M., Schultz M., Druce J., Sherry, N.
EPI_ISL_419898 hCoV-19/Australia/VIC203/2020 Oceania / Australia / Victoria 2020-03-19 Victorian Infectious Diseases Reference Laboratory (VIDRL) Victorian Infectious Diseases Reference Laboratory and Microbiological Diagnostic Unit Public Health Laboratory, Doherty Institute Caly L., Seemann T., Sait, M., Schultz M., Druce J., Sherry, N.
EPI_ISL_419899 hCoV-19/Australia/VIC204/2020 Oceania / Australia / Victoria 2020-03-19 Victorian Infectious Diseases Reference Laboratory (VIDRL) Victorian Infectious Diseases Reference Laboratory and Microbiological Diagnostic Unit Public Health Laboratory, Doherty Institute Caly L., Seemann T., Sait, M., Schultz M., Druce J., Sherry, N.
EPI_ISL_420508 hCoV-19/England/20132072802/2020 Europe / United Kingdom / England 2020-03-23 Respiratory Virus Unit, Microbiology Services Colindale, Public Health England Respiratory Virus Unit, Microbiology Services Colindale, Public Health England Monica Galiano, Shahjahan Miah, Angie Lackenby, Omolola Akinbami, Tiina Talts, Leena Bhaw, Richard Myers, Steven Platt, Kirstin Edwards, Jonathan Hubb, Joanna Ellis, Maria Zambon
EPI_ISL_420520 hCoV-19/England/20136015502/2020 Europe / United Kingdom / England 2020-03-24 Respiratory Virus Unit, Microbiology Services Colindale, Public Health England Respiratory Virus Unit, Microbiology Services Colindale, Public Health England Monica Galiano, Shahjahan Miah, Angie Lackenby, Omolola Akinbami, Tiina Talts, Leena Bhaw, Richard Myers, Steven Platt, Kirstin Edwards, Jonathan Hubb, Joanna Ellis, Maria Zambon
EPI_ISL_420521 hCoV-19/England/20138012802/2020 Europe / United Kingdom / England 2020-03-25 Respiratory Virus Unit, Microbiology Services Colindale, Public Health England Respiratory Virus Unit, Microbiology Services Colindale, Public Health England Monica Galiano, Shahjahan Miah, Angie Lackenby, Omolola Akinbami, Tiina Talts, Leena Bhaw, Richard Myers, Steven Platt, Kirstin Edwards, Jonathan Hubb, Joanna Ellis, Maria Zambon
EPI_ISL_420522 hCoV-19/England/20138014402/2020 Europe / United Kingdom / England 2020-03-25 Respiratory Virus Unit, Microbiology Services Colindale, Public Health England Respiratory Virus Unit, Microbiology Services Colindale, Public Health England Monica Galiano, Shahjahan Miah, Angie Lackenby, Omolola Akinbami, Tiina Talts, Leena Bhaw, Richard Myers, Steven Platt, Kirstin Edwards, Jonathan Hubb, Joanna Ellis, Maria Zambon
EPI_ISL_420524 hCoV-19/England/20139018302/2020 Europe / United Kingdom / England 2020-03-26 Respiratory Virus Unit, Microbiology Services Colindale, Public Health England Respiratory Virus Unit, Microbiology Services Colindale, Public Health England Monica Galiano, Shahjahan Miah, Angie Lackenby, Omolola Akinbami, Tiina Talts, Leena Bhaw, Richard Myers, Steven Platt, Kirstin Edwards, Jonathan Hubb, Joanna Ellis, Maria Zambon
EPI_ISL_420531 hCoV-19/Australia/WA01/2020 Oceania / Australia 2020-03-14 Department of Microbiology, PathWest QEII Medical Centre Department of Microbiology, PathWest QEII Medical Centre Chisha Sikazwe, Jurissa Lang, Avram Levy, David Speers and David Smith
EPI_ISL_420532 hCoV-19/Australia/WA02/2020 Oceania / Australia / Western Australia / Perth 2020-03-13 Department of Microbiology, PathWest QEII Medical Centre Department of Microbiology, PathWest QEII Medical Centre Chisha Sikazwe, Jurissa Lang, Avram Levy, David Speers and David Smith
EPI_ISL_420533 hCoV-19/Australia/WA03/2020 Oceania / Australia / Western Australia / Perth 2020-03-14 Department of Microbiology, PathWest QEII Medical Centre Department of Microbiology, PathWest QEII Medical Centre Chisha Sikazwe, Jurissa Lang, Avram Levy, David Speers and David Smith
EPI_ISL_420534 hCoV-19/Australia/WA06/2020 Oceania / Australia / Western Australia / Perth 2020-03-04 Department of Microbiology, PathWest QEII Medical Centre Department of Microbiology, PathWest QEII Medical Centre Chisha Sikazwe, Jurissa Lang, Avram Levy, David Speers and David Smith
EPI_ISL_420536 hCoV-19/Australia/WA04/2020 Oceania / Australia / Western Australia / Perth 2020-03-15 Department of Microbiology, PathWest QEII Medical Centre Department of Microbiology, PathWest QEII

Medical Centre		Chisha Sikazwe, Jurissa Lang, Avram Levy, David Speers and David Smith
EPI_ISL_420537	hCoV-19/Australia/WA07/2020	Oceania / Australia / Western Australia / Perth 2020-03-13	Department of Microbiology, PathWest QEII Medical Centre	Department of Microbiology, PathWest QEII Medical Centre		Chisha Sikazwe, Jurissa Lang, Avram Levy, David Speers and David Smith
EPI_ISL_420538	hCoV-19/Australia/WA09/2020	Oceania / Australia / Western Australia / Perth 2020-03-14	Department of Microbiology, PathWest QEII Medical Centre	Department of Microbiology, PathWest QEII Medical Centre		Chisha Sikazwe, Jurissa Lang, Avram Levy, David Speers and David Smith
EPI_ISL_420539	hCoV-19/Australia/WA10/2020	Oceania / Australia / Western Australia / Perth 2020-03-14	Department of Microbiology, PathWest QEII Medical Centre	Department of Microbiology, PathWest QEII Medical Centre		Chisha Sikazwe, Jurissa Lang, Avram Levy, David Speers and David Smith
EPI_ISL_420540	hCoV-19/Estonia/ChVir2148/2020	Europe / Estonia	2020-03 SYNLAB Eesti OÜ Charité Universitätsmedizin Berlin, Institute of Virology	Victor M Corman, Jörn Beheim-Schwarzbach, Barbara Mühlemann, Talitha Veith, Julia Schneider, Paul Naaber, Terry Jones, Christian Drosten
EPI_ISL_420541	hCoV-19/Slovenia/808/2020	Europe / Slovenia	2020-03-05	Institute of Microbiology and Immunology, Faculty of Medicine, University of Ljubljana	Institute of Microbiology and Immunology, Faculty of Medicine, University of Ljubljana	Tomaž Mark Zorec, Samo Zakotnik, Lucijan Skubic, Miša Korva, Tatjana Avšič - Županc, Mario Poljak
EPI_ISL_420543	hCoV-19/India/763/2020	Asia / India	2020-03-03	National Influenza Center, Indian Council of Medical Research - National Institute of Virology	Indian Council of Medical Research-National Institute of Virology, Microbial Containment Complex	Pragya D. Yadav. Savita Patil, Varsha Potdar, Prasad Sarkale, Dimpal A. Nyayanit, Gajanan Sapkal, Anita M. Shete, Atanu Basu, Lalit Dar, M Choudhary, Amita Jain, Bharati Malhotra, Pranita Gawande, Sarah Cherian, Priya Abraham
EPI_ISL_420544	hCoV-19/India/2020763/2020	Asia / India	2020	Indian Council of Medical Research-National Institute of Virology, Microbial Containment Complex	Indian Council of Medical Research-National Institute of Virology, Microbial Containment Complex	Pragya D. Yadav. Savita Patil, Varsha Potdar, Prasad Sarkale, Dimpal A. Nyayanit, Gajanan Sapkal, Anita M. Shete, Atanu Basu, Lalit Dar, M Choudhary, Amita Jain, Bharati Malhotra, Pranita Gawande, Sarah Cherian, Priya Abraham
EPI_ISL_420545	hCoV-19/India/770/2020	Asia / India	2020-03-03	National Influenza Center, Indian Council of Medical Research - National Institute of Virology	Indian Council of Medical Research-National Institute of Virology, Microbial Containment Complex	Pragya D. Yadav. Savita Patil, Varsha Potdar, Prasad Sarkale, Dimpal A. Nyayanit, Gajanan Sapkal, Anita M. Shete, Atanu Basu, Lalit Dar, M Choudhary, Amita Jain, Bharati Malhotra, Pranita Gawande, Sarah Cherian, Priya Abraham
EPI_ISL_420546	hCoV-19/India/2020770/2020	Asia / India	2020	Indian Council of Medical Research-National Institute of Virology, Microbial Containment Complex	Indian Council of Medical Research-National Institute of Virology, Microbial Containment Complex	Pragya D. Yadav. Savita Patil, Varsha Potdar, Prasad Sarkale, Dimpal A. Nyayanit, Gajanan Sapkal, Anita M. Shete, Atanu Basu, Lalit Dar, M Choudhary, Amita Jain, Bharati Malhotra, Pranita Gawande, Sarah Cherian, Priya Abraham
EPI_ISL_420547	hCoV-19/India/772/2020	Asia / India	2020-03-03	National Influenza Center, Indian Council of Medical Research - National Institute of Virology	Indian Council of Medical Research-National Institute of Virology, Microbial Containment Complex	Pragya D. Yadav. Savita Patil, Varsha Potdar, Prasad Sarkale, Dimpal A. Nyayanit, Gajanan Sapkal, Anita M. Shete, Atanu Basu, Lalit Dar, M Choudhary, Amita Jain, Bharati Malhotra, Pranita Gawande, Sarah Cherian, Priya Abraham
EPI_ISL_420548	hCoV-19/India/2020772/2020	Asia / India	2020	Indian Council of Medical Research-National Institute of Virology, Microbial Containment Complex	Indian Council of Medical Research-National Institute of Virology, Microbial Containment Complex	Pragya D. Yadav. Savita Patil, Varsha Potdar, Prasad Sarkale, Dimpal A. Nyayanit, Gajanan Sapkal, Anita M. Shete, Atanu Basu, Lalit Dar, M Choudhary, Amita Jain, Bharati Malhotra, Pranita Gawande, Sarah Cherian, Priya Abraham
EPI_ISL_420549	hCoV-19/India/773/2020	Asia / India	2020-03-03	National Influenza Center, Indian Council of Medical Research - National Institute of Virology	Indian Council of Medical Research-National Institute of Virology, Microbial Containment Complex	Pragya D. Yadav. Savita Patil, Varsha Potdar, Prasad Sarkale, Dimpal A. Nyayanit, Gajanan Sapkal, Anita M. Shete, Atanu Basu, Lalit Dar, M Choudhary, Amita Jain, Bharati Malhotra, Pranita Gawande, Sarah Cherian, Priya Abraham
EPI_ISL_420550	hCoV-19/India/2020773/2020	Asia / India	2020	Indian Council of Medical Research-National Institute of Virology, Microbial Containment Complex	Indian Council of Medical Research-National Institute of Virology, Microbial Containment Complex	Pragya D. Yadav. Savita Patil, Varsha Potdar, Prasad Sarkale, Dimpal A. Nyayanit, Gajanan Sapkal, Anita M. Shete, Atanu Basu, Lalit Dar, M Choudhary, Amita Jain, Bharati Malhotra, Pranita Gawande, Sarah Cherian, Priya Abraham
EPI_ISL_420551	hCoV-19/India/777/2020	Asia / India	2020-03-03	National Influenza Center, Indian Council of Medical Research - National Institute of Virology	Indian Council of Medical Research-National Institute of Virology, Microbial Containment Complex	Pragya D. Yadav. Savita Patil, Varsha Potdar, Prasad Sarkale, Dimpal A. Nyayanit, Gajanan Sapkal, Anita M. Shete, Atanu Basu, Lalit Dar, M Choudhary, Amita Jain, Bharati Malhotra, Pranita Gawande, Sarah Cherian, Priya Abraham
EPI_ISL_420552	hCoV-19/India/2020777/2020	Asia / India	2020	Indian Council of Medical Research-National Institute of Virology, Microbial Containment Complex	Indian Council of Medical Research-National Institute of Virology, Microbial Containment Complex	Pragya D. Yadav. Savita Patil, Varsha Potdar, Prasad Sarkale, Dimpal A. Nyayanit, Gajanan Sapkal, Anita M. Shete, Atanu Basu, Lalit Dar, M Choudhary, Amita Jain, Bharati Malhotra, Pranita Gawande, Sarah Cherian, Priya Abraham
EPI_ISL_420553	hCoV-19/India/781/2020	Asia / India	2020-03-03	National Influenza Center, Indian Council of Medical Research - National Institute of Virology	Indian Council of Medical Research-National Institute of Virology, Microbial Containment Complex	Pragya D. Yadav. Savita Patil, Varsha Potdar, Prasad

Sarkale, Dimpal A. Nyayanit, Gajanan Sapkal, Anita M. Shete, Atanu Basu, Lalit Dar, M Choudhary, Amita Jain, Bharati Malhotra, Pranita Gawande, Sarah Cherian, Priya Abraham
EPI_ISL_420554 hCoV-19/India/2020781/2020 Asia / India 2020 Indian Council of Medical Research-National Institute of Virology, Microbial Containment Complex Indian Council of Medical Research-National Institute of Virology, Microbial Containment Complex Pragya D. Yadav. Savita Patil, Varsha Potdar, Prasad Sarkale, Dimpal A. Nyayanit, Gajanan Sapkal, Anita M. Shete, Atanu Basu, Lalit Dar, M Choudhary, Amita Jain, Bharati Malhotra, Pranita Gawande, Sarah Cherian, Priya Abraham
EPI_ISL_420555 hCoV-19/India/c32/2020 Asia / India 2020-03-03 National Influenza Center, Indian Council of Medical Research - National Institute of Virology Indian Council of Medical Research-National Institute of Virology, Microbial Containment Complex Pragya D. Yadav. Savita Patil, Varsha Potdar, Prasad Sarkale, Dimpal A. Nyayanit, Gajanan Sapkal, Anita M. Shete, Atanu Basu, Lalit Dar, M Choudhary, Amita Jain, Bharati Malhotra, Pranita Gawande, Sarah Cherian, Priya Abraham
EPI_ISL_420556 hCoV-19/India/2020c32/2020 Asia / India 2020 Indian Council of Medical Research-National Institute of Virology, Microbial Containment Complex Indian Council of Medical Research-National Institute of Virology, Microbial Containment Complex Pragya D. Yadav. Savita Patil, Varsha Potdar, Prasad Sarkale, Dimpal A. Nyayanit, Gajanan Sapkal, Anita M. Shete, Atanu Basu, Lalit Dar, M Choudhary, Amita Jain, Bharati Malhotra, Pranita Gawande, Sarah Cherian, Priya Abraham
EPI_ISL_420563 hCoV-19/Italy/TE5166/2020 Europe / Italy / Teramo 2020-03-18 Ospedale Civile Giuseppe Mazzini "Istituto Zooprofilattico Sperimentale dell'Abruzzo e Molise ""G. Caporale""" Lorusso A, Marcacci M, Di Domenico M, Ancora M, Curini V, Mangone I, Rinaldi A, Di Pasquale A, Cammà C, Puglia I, Savini G
EPI_ISL_420564 hCoV-19/Italy/TE5472/2020 Europe / Italy / Castel di Sangro 2020-03-19 Ospedale Civile Castel Di Sangro Istituto Zooprofilattico Sperimentale dell’Abruzzo e Molise “G. Caporale” Lorusso A, Marcacci M, Di Domenico M, Ancora M, Curini V, Mangone I, Rinaldi A, Di Pasquale A, Cammà C, Puglia I, Savini G
EPI_ISL_420565 hCoV-19/Italy/TE5476/2020 Europe / Italy / Abruzzo / Teramo 2020-03-19 Ospedale Civile Giuseppe Mazzini Istituto Zooprofilattico Sperimentale dell’Abruzzo e Molise “G. Caporale” Lorusso A, Marcacci M, Di Domenico M, Ancora M, Curini V, Mangone I, Rinaldi A, Di Pasquale A, Cammà C, Puglia I, Savini G
EPI_ISL_420566 hCoV-19/Italy/TE5512/2020 Europe / Italy / Abruzzo 2020-03-19 Ospedale Regionale San Salvatore Istituto Zooprofilattico Sperimentale dell’Abruzzo e Molise “G. Caporale” Lorusso A, Marcacci M, Di Domenico M, Ancora M, Curini V, Mangone I, Rinaldi A, Di Pasquale A, Cammà C, Puglia I, Savini G
EPI_ISL_420567 hCoV-19/Italy/TE5780/2020 Europe / Italy / Abruzzo / L'Aquila 2020-03-21 Ospedale Regionale San Salvatore Istituto Zooprofilattico Sperimentale dell’Abruzzo e Molise “G. Caporale” Lorusso A, Marcacci M, Di Domenico M, Ancora M, Curini V, Mangone I, Rinaldi A, Di Pasquale A, Cammà C, Puglia I, Savini G
EPI_ISL_420568 hCoV-19/Italy/6193/2020 Europe / Italy / Abruzzo 2020-03-23 Ospedale Civile Giuseppe Mazzini Istituto Zooprofilattico Sperimentale dell’Abruzzo e Molise “G. Caporale” Lorusso A, Marcacci M, Di Domenico M, Ancora M, Curini V, Mangone I, Rinaldi A, Di Pasquale A, Cammà C, Puglia I, Savini G
EPI_ISL_420569 hCoV-19/Italy/TE6195/2020 Europe / Italy / Abruzzo 2020-03-23 Ospedale Civile Giuseppe Mazzini Istituto Zooprofilattico Sperimentale dell’Abruzzo e Molise “G. Caporale” Lorusso A, Marcacci M, Di Domenico M, Ancora M, Curini V, Mangone I, Rinaldi A, Di Pasquale A, Cammà C, Puglia I, Savini G
EPI_ISL_420570 hCoV-19/USA/NY-NYUMC55/2020 North America / USA / New York / Nassau County 2020-03-17 NYU Langone Health Departments of Pathology and Medicine, New York University School of Medicine Maria Aguero-Rosenfeld, Brendan Belovarac, Margaret Black, Ludovic Boytard, John Cadley, Paolo Cotzia, John Chen, Dacia Dimartino, Xiaojun Feng, Tatyana Gindin, Adriana Heguy, Megan Hogan, Emily Huang, George Jour, Andrew Lytle, Christian Marier, Matthew T. Maurano, Mark J. Mulligan, Peter Meyn, Iman Osman, Jared Pinnell, Sitharam Ramaswami, Amy Rapkiewicz, Marie Samanovic-Golden, Antonio Serrano, Guomiao Shen, Matija Snuderl, Theodore Vougiouklakis, Nick Vulpescu, Gael Westby, Paul Zappile, Yutong Zhang
EPI_ISL_420571 hCoV-19/USA/NY-NYUMC56/2020 North America / USA / New York / Nassau County 2020-03-18 NYU Langone Health Departments of Pathology and Medicine, New York University School of Medicine Maria Aguero-Rosenfeld, Brendan Belovarac, Margaret Black, Ludovic Boytard, John Cadley, Paolo Cotzia, John Chen, Dacia Dimartino, Xiaojun Feng, Tatyana Gindin, Adriana Heguy, Megan Hogan, Emily Huang, George Jour, Andrew Lytle, Christian Marier, Matthew T. Maurano, Mark J. Mulligan, Peter Meyn, Iman Osman, Jared Pinnell, Sitharam Ramaswami, Amy Rapkiewicz, Marie Samanovic-Golden, Antonio Serrano, Guomiao Shen, Matija Snuderl, Theodore Vougiouklakis, Nick Vulpescu, Gael Westby, Paul Zappile, Yutong Zhang
EPI_ISL_420572 hCoV-19/USA/NY-NYUMC57/2020 North America / USA / New York / Nassau County 2020-03-17 NYU Langone Health Departments of Pathology and Medicine, New York University School of Medicine Maria Aguero-Rosenfeld, Brendan Belovarac, Margaret Black, Ludovic Boytard, John Cadley, Paolo Cotzia, John Chen, Dacia Dimartino, Xiaojun Feng, Tatyana Gindin, Adriana Heguy, Megan Hogan, Emily Huang, George Jour, Andrew Lytle, Christian Marier, Matthew T. Maurano, Mark J. Mulligan, Peter Meyn, Iman Osman, Jared Pinnell, Sitharam Ramaswami, Amy Rapkiewicz, Marie Samanovic-Golden, Antonio Serrano, Guomiao Shen, Matija Snuderl, Theodore Vougiouklakis, Nick Vulpescu, Gael Westby, Paul Zappile, Yutong Zhang
EPI_ISL_420573 hCoV-19/USA/NY-NYUMC58/2020 North America / USA / New York / Manhattan 2020-03-18 NYU Langone Health Departments of Pathology and Medicine, New York University School of Medicine Maria Aguero-Rosenfeld, Brendan Belovarac, Margaret Black, Ludovic Boytard, John Cadley, Paolo Cotzia, John Chen, Dacia Dimartino, Xiaojun Feng, Tatyana Gindin, Adriana Heguy, Megan Hogan, Emily Huang, George Jour, Andrew

Lytle, Christian Marier, Matthew T. Maurano, Mark J. Mulligan, Peter Meyn, Iman Osman, Jared Pinnell, Sitharam Ramaswami, Amy Rapkiewicz, Marie Samanovic-Golden, Antonio Serrano, Guomiao Shen, Matija Snuderl, Theodore Vougiouklakis, Nick Vulpescu, Gael Westby, Paul Zappile, Yutong Zhang
EPI_ISL_420575 hCoV-19/USA/NY-NYUMC60/2020 North America / USA / New York / Nassau County 2020-03-18 NYU Langone Health Departments of Pathology and Medicine, New York University School of Medicine Maria Aguero-Rosenfeld, Brendan Belovarac, Margaret Black, Ludovic Boytard, John Cadley, Paolo Cotzia, John Chen, Dacia Dimartino, Xiaojun Feng, Tatyana Gindin, Adriana Heguy, Megan Hogan, Emily Huang, George Jour, Andrew Lytle, Christian Marier, Matthew T. Maurano, Mark J. Mulligan, Peter Meyn, Iman Osman, Jared Pinnell, Sitharam Ramaswami, Amy Rapkiewicz, Marie Samanovic-Golden, Antonio Serrano, Guomiao Shen, Matija Snuderl, Theodore Vougiouklakis, Nick Vulpescu, Gael Westby, Paul Zappile, Yutong Zhang
EPI_ISL_420576 hCoV-19/USA/NY-NYUMC61/2020 North America / USA / New York 2020-03-18 NYU Langone Health Departments of Pathology and Medicine, New York University School of Medicine Maria Aguero-Rosenfeld, Brendan Belovarac, Margaret Black, Ludovic Boytard, John Cadley, Paolo Cotzia, John Chen, Dacia Dimartino, Xiaojun Feng, Tatyana Gindin, Adriana Heguy, Megan Hogan, Emily Huang, George Jour, Andrew Lytle, Christian Marier, Matthew T. Maurano, Mark J. Mulligan, Peter Meyn, Iman Osman, Jared Pinnell, Sitharam Ramaswami, Amy Rapkiewicz, Marie Samanovic-Golden, Antonio Serrano, Guomiao Shen, Matija Snuderl, Theodore Vougiouklakis, Nick Vulpescu, Gael Westby, Paul Zappile, Yutong Zhang
EPI_ISL_420577 hCoV-19/USA/NY-NYUMC62/2020 North America / USA / New York / Manhattan 2020-03-18 NYU Langone Health Departments of Pathology and Medicine, New York University School of Medicine Maria Aguero-Rosenfeld, Brendan Belovarac, Margaret Black, Ludovic Boytard, John Cadley, Paolo Cotzia, John Chen, Dacia Dimartino, Xiaojun Feng, Tatyana Gindin, Adriana Heguy, Megan Hogan, Emily Huang, George Jour, Andrew Lytle, Christian Marier, Matthew T. Maurano, Mark J. Mulligan, Peter Meyn, Iman Osman, Jared Pinnell, Sitharam Ramaswami, Amy Rapkiewicz, Marie Samanovic-Golden, Antonio Serrano, Guomiao Shen, Matija Snuderl, Theodore Vougiouklakis, Nick Vulpescu, Gael Westby, Paul Zappile, Yutong Zhang
EPI_ISL_420578 hCoV-19/USA/NY-NYUMC63/2020 North America / USA / New York / Manhattan 2020-03-18 NYU Langone Health Departments of Pathology and Medicine, New York University School of Medicine Maria Aguero-Rosenfeld, Brendan Belovarac, Margaret Black, Ludovic Boytard, John Cadley, Paolo Cotzia, John Chen, Dacia Dimartino, Xiaojun Feng, Tatyana Gindin, Adriana Heguy, Megan Hogan, Emily Huang, George Jour, Andrew Lytle, Christian Marier, Matthew T. Maurano, Mark J. Mulligan, Peter Meyn, Iman Osman, Jared Pinnell, Sitharam Ramaswami, Amy Rapkiewicz, Marie Samanovic-Golden, Antonio Serrano, Guomiao Shen, Matija Snuderl, Theodore Vougiouklakis, Nick Vulpescu, Gael Westby, Paul Zappile, Yutong Zhang
EPI_ISL_420579 hCoV-19/USA/NY-NYUMC64/2020 North America / USA / New York / Manhattan 2020-03-18 NYU Langone Health Departments of Pathology and Medicine, New York University School of Medicine Maria Aguero-Rosenfeld, Brendan Belovarac, Margaret Black, Ludovic Boytard, John Cadley, Paolo Cotzia, John Chen, Dacia Dimartino, Xiaojun Feng, Tatyana Gindin, Adriana Heguy, Megan Hogan, Emily Huang, George Jour, Andrew Lytle, Christian Marier, Matthew T. Maurano, Mark J. Mulligan, Peter Meyn, Iman Osman, Jared Pinnell, Sitharam Ramaswami, Amy Rapkiewicz, Marie Samanovic-Golden, Antonio Serrano, Guomiao Shen, Matija Snuderl, Theodore Vougiouklakis, Nick Vulpescu, Gael Westby, Paul Zappile, Yutong Zhang
EPI_ISL_420580 hCoV-19/USA/NY-NYUMC65/2020 North America / USA / New York / Manhattan 2020-03-18 NYU Langone Health Departments of Pathology and Medicine, New York University School of Medicine Maria Aguero-Rosenfeld, Brendan Belovarac, Margaret Black, Ludovic Boytard, John Cadley, Paolo Cotzia, John Chen, Dacia Dimartino, Xiaojun Feng, Tatyana Gindin, Adriana Heguy, Megan Hogan, Emily Huang, George Jour, Andrew Lytle, Christian Marier, Matthew T. Maurano, Mark J. Mulligan, Peter Meyn, Iman Osman, Jared Pinnell, Sitharam Ramaswami, Amy Rapkiewicz, Marie Samanovic-Golden, Antonio Serrano, Guomiao Shen, Matija Snuderl, Theodore Vougiouklakis, Nick Vulpescu, Gael Westby, Paul Zappile, Yutong Zhang
EPI_ISL_420581 hCoV-19/USA/NY-NYUMC66/2020 North America / USA / New York / Manhattan 2020-03-18 NYU Langone Health Departments of Pathology and Medicine, New York University School of Medicine Maria Aguero-Rosenfeld, Brendan Belovarac, Margaret Black, Ludovic Boytard, John Cadley, Paolo Cotzia, John Chen, Dacia Dimartino, Xiaojun Feng, Tatyana Gindin, Adriana Heguy, Megan Hogan, Emily Huang, George Jour, Andrew Lytle, Christian Marier, Matthew T. Maurano, Mark J. Mulligan, Peter Meyn, Iman Osman, Jared Pinnell, Sitharam Ramaswami, Amy Rapkiewicz, Marie Samanovic-Golden, Antonio Serrano, Guomiao Shen, Matija Snuderl, Theodore Vougiouklakis, Nick Vulpescu, Gael Westby, Paul Zappile, Yutong Zhang
EPI_ISL_420583 hCoV-19/Italy/TE6222/2020 Europe / Italy / Abruzzo 2020-03-23 Ospedale Civile Giuseppe Mazzini Istituto Zooprofilattico Sperimentale dell’Abruzzo e Molise “G. Caporale” Lorusso A, Marcacci M, Di Domenico M, Ancora M, Curini V, Mangone I, Rinaldi A, Di Pasquale A, Cammà C, Puglia I, Savini G
EPI_ISL_420585 hCoV-19/USA/NY-NYUMC69/2020 North America / USA / New York / Manhattan 2020-03-18 NYU Langone Health Departments of Pathology and Medicine, New York University School of Medicine Maria Aguero-Rosenfeld, Brendan Belovarac, Margaret Black, Ludovic Boytard, John Cadley, Paolo Cotzia, John Chen, Dacia Dimartino, Xiaojun Feng, Tatyana Gindin, Adriana Heguy, Megan Hogan, Emily Huang, George Jour, Andrew Lytle, Christian Marier, Matthew T. Maurano, Mark J. Mulligan, Peter Meyn, Iman Osman, Jared Pinnell, Sitharam Ramaswami, Amy Rapkiewicz, Marie Samanovic-Golden, Antonio Serrano, Guomiao Shen, Matija Snuderl, Theodore Vougiouklakis, Nick Vulpescu, Gael Westby, Paul Zappile, Yutong Zhang
EPI_ISL_420586 hCoV-19/USA/NY-NYUMC70/2020 North America / USA / New York / Brooklyn 2020-03-19 NYU Langone Health Departments of Pathology and Medicine, New York University School of Medicine Maria Aguero-Rosenfeld, Brendan Belovarac, Margaret Black, Ludovic Boytard, John Cadley, Paolo Cotzia, John Chen, Dacia Dimartino, Xiaojun Feng, Tatyana Gindin, Adriana Heguy, Megan Hogan, Emily Huang, George Jour, Andrew Lytle, Christian Marier, Matthew T. Maurano, Mark J. Mulligan, Peter Meyn, Iman Osman, Jared Pinnell, Sitharam Ramaswami, Amy Rapkiewicz, Marie Samanovic-Golden, Antonio Serrano, Guomiao Shen, Matija Snuderl, Theodore Vougiouklakis, Nick Vulpescu, Gael Westby, Paul Zappile, Yutong Zhang

EPI_ISL_420587 hCoV-19/USA/NY-NYUMC71/2020 North America / USA / New York / Manhattan 2020-03-18 NYU Langone Health Departments of Pathology and Medicine, New York University School of Medicine Maria Aguero-Rosenfeld, Brendan Belovarac, Margaret Black, Ludovic Boytard, John Cadley, Paolo Cotzia, John Chen, Dacia Dimartino, Xiaojun Feng, Tatyana Gindin, Adriana Heguy, Megan Hogan, Emily Huang, George Jour, Andrew Lytle, Christian Marier, Matthew T. Maurano, Mark J. Mulligan, Peter Meyn, Iman Osman, Jared Pinnell, Sitharam Ramaswami, Amy Rapkiewicz, Marie Samanovic-Golden, Antonio Serrano, Guomiao Shen, Matija Snuderl, Theodore Vougiouklakis, Nick Vulpescu, Gael Westby, Paul Zappile, Yutong Zhang
EPI_ISL_420588 hCoV-19/USA/NY-NYUMC72/2020 North America / USA / New York / Manhattan 2020-03-18 NYU Langone Health Departments of Pathology and Medicine, New York University School of Medicine Maria Aguero-Rosenfeld, Brendan Belovarac, Margaret Black, Ludovic Boytard, John Cadley, Paolo Cotzia, John Chen, Dacia Dimartino, Xiaojun Feng, Tatyana Gindin, Adriana Heguy, Megan Hogan, Emily Huang, George Jour, Andrew Lytle, Christian Marier, Matthew T. Maurano, Mark J. Mulligan, Peter Meyn, Iman Osman, Jared Pinnell, Sitharam Ramaswami, Amy Rapkiewicz, Marie Samanovic-Golden, Antonio Serrano, Guomiao Shen, Matija Snuderl, Theodore Vougiouklakis, Nick Vulpescu, Gael Westby, Paul Zappile, Yutong Zhang
EPI_ISL_420589 hCoV-19/USA/NY-NYUMC73/2020 North America / USA / New York / Manhattan 2020-03-18 NYU Langone Health Departments of Pathology and Medicine, New York University School of Medicine Maria Aguero-Rosenfeld, Brendan Belovarac, Margaret Black, Ludovic Boytard, John Cadley, Paolo Cotzia, John Chen, Dacia Dimartino, Xiaojun Feng, Tatyana Gindin, Adriana Heguy, Megan Hogan, Emily Huang, George Jour, Andrew Lytle, Christian Marier, Matthew T. Maurano, Mark J. Mulligan, Peter Meyn, Iman Osman, Jared Pinnell, Sitharam Ramaswami, Amy Rapkiewicz, Marie Samanovic-Golden, Antonio Serrano, Guomiao Shen, Matija Snuderl, Theodore Vougiouklakis, Nick Vulpescu, Gael Westby, Paul Zappile, Yutong Zhang
EPI_ISL_420590 hCoV-19/USA/NY-NYUMC74/2020 North America / USA / New York / Nassau County 2020-03-18 NYU Langone Health Departments of Pathology and Medicine, New York University School of Medicine Maria Aguero-Rosenfeld, Brendan Belovarac, Margaret Black, Ludovic Boytard, John Cadley, Paolo Cotzia, John Chen, Dacia Dimartino, Xiaojun Feng, Tatyana Gindin, Adriana Heguy, Megan Hogan, Emily Huang, George Jour, Andrew Lytle, Christian Marier, Matthew T. Maurano, Mark J. Mulligan, Peter Meyn, Iman Osman, Jared Pinnell, Sitharam Ramaswami, Amy Rapkiewicz, Marie Samanovic-Golden, Antonio Serrano, Guomiao Shen, Matija Snuderl, Theodore Vougiouklakis, Nick Vulpescu, Gael Westby, Paul Zappile, Yutong Zhang
EPI_ISL_420591 hCoV-19/USA/NY-NYUMC75/2020 North America / USA / New York / Manhattan 2020-03-18 NYU Langone Health Departments of Pathology and Medicine, New York University School of Medicine Maria Aguero-Rosenfeld, Brendan Belovarac, Margaret Black, Ludovic Boytard, John Cadley, Paolo Cotzia, John Chen, Dacia Dimartino, Xiaojun Feng, Tatyana Gindin, Adriana Heguy, Megan Hogan, Emily Huang, George Jour, Andrew Lytle, Christian Marier, Matthew T. Maurano, Mark J. Mulligan, Peter Meyn, Iman Osman, Jared Pinnell, Sitharam Ramaswami, Amy Rapkiewicz, Marie Samanovic-Golden, Antonio Serrano, Guomiao Shen, Matija Snuderl, Theodore Vougiouklakis, Nick Vulpescu, Gael Westby, Paul Zappile, Yutong Zhang
EPI_ISL_420598 hCoV-19/Argentina/C3013/2020 South America / Argentina 2020-03-22 Servicio Virosis Respiratorias-Departamento Virología-INEI Instituto Nacional Enfermedades Infecciosas C.G.Malbran Baumeister E., Avaro M., Benedetti E., Russo M., Dattero ME, Pontoriero A., Cisterna D., Molina V., Perandones C., Tuduri E., Lorenzo F., Poklepovich T., Campos J.
EPI_ISL_420599 hCoV-19/Argentina/C1374/2020 South America / Argentina 2020-03-18 Servicio Virosis Respiratorias-Departamento Virología-INEI Instituto Nacional Enfermedades Infecciosas C.G.Malbran Baumeister E., Avaro M., Benedetti E., Russo M., Dattero ME, Pontoriero A., Cisterna D., Molina V., Perandones C., Tuduri E., Lorenzo F., Poklepovich T., Campos J.
EPI_ISL_420800 hCoV-19/Korea/BA-ACH_2718/2020 Asia / Korea 2020-02-29 Brian D. Allgood Army Community Hospital Pathogen Discovery, Respiratory Viruses Branch, Division of Viral Diseases, Centers for Disease Control and Prevention Krista Queen, Yan Li, Ying Tao, Jing Zhang, Anne Uehara, Clinton R. Paden, Haibin Wang, Rachel Marine, Mary S. Keckler, Alison S. Laufer Halpin, Jasmine Padilla, Justin Lee, Christopher A. Elkins, Suxiang Tong
EPI_ISL_420801 hCoV-19/Korea/BA-ACH_2719/2020 Asia / Korea 2020-02-29 Brian D. Allgood Army Community Hospital Pathogen Discovery, Respiratory Viruses Branch, Division of Viral Diseases, Centers for Disease Control and Prevention Krista Queen, Yan Li, Ying Tao, Jing Zhang, Anne Uehara, Clinton R. Paden, Haibin Wang, Rachel Marine, Mary S. Keckler, Alison S. Laufer Halpin, Jasmine Padilla, Justin Lee, Christopher A. Elkins, Suxiang Tong
EPI_ISL_420803 hCoV-19/USA/UT-089/2020 North America / USA / Utah 2020-03-12 Utah Public Health Laboratory Utah Public Health Laboratory Erin Young, Kelly Oakeson
EPI_ISL_420810 hCoV-19/USA/UT-0290/2020 North America / USA / Utah 2020-03-27 Utah Public Health Laboratory Utah Public Health Laboratory Erin Young, Kelly Oakeson
EPI_ISL_420812 hCoV-19/USA/UT-0297/2020 North America / USA / Utah 2020-03-27 Utah Public Health Laboratory Utah Public Health Laboratory Erin Young, Kelly Oakeson
EPI_ISL_420814 hCoV-19/USA/UT-0301/2020 North America / USA / Utah 2020-03-27 Utah Public Health Laboratory Utah Public Health Laboratory Erin Young, Kelly Oakeson
EPI_ISL_420838 hCoV-19/DRC/253/2020 Africa / Democratic Republic of the Congo / Kinshasa 2020-03-22 Viral Respiratory Lab, National Institute for Biomedical Research (INRB) Pathogen Sequencing Lab, National Institute for Biomedical Research (INRB) Placide Mbala-Kingebeni, Edith Nkwembe, Eddy Kinganda-Lusamaki, Amuri Aziza, Catherine Pratt, Matthias Pauthner, Josh Quick, Allison Black, James Hadfield, Trevor Bedford, Ian Goodfellow, Nick Loman, Kristian Andersen, Michael Wiley, Steve Ahuka-Mundeke, Jean-Jacques Muyembe Tamfum
EPI_ISL_420841 hCoV-19/DRC/300/2020 Africa / Democratic Republic of the Congo / Sud Kivu 2020-03-22 Viral Respiratory Lab, National Institute for Biomedical Research (INRB) Pathogen Sequencing Lab, National Institute for Biomedical Research (INRB) Placide Mbala-Kingebeni, Edith Nkwembe, Eddy Kinganda-

Lusamaki, Amuri Aziza, Catherine Pratt, Matthias Pauthner, Josh Quick, Allison Black, James Hadfield, Trevor Bedford, Ian Goodfellow, Nick Loman, Kristian Andersen, Michael Wiley, Steve Ahuka-Mundeke, Jean-Jacques Muyembe Tamfum
EPI_ISL_420844 hCoV-19/DRC/355/2020 Africa / Democratic Republic of the Congo / Kinshasa 2020-03 Viral Respiratory Lab, National Institute for Biomedical Research (INRB) Pathogen Sequencing Lab, National Institute for Biomedical Research (INRB) Placide Mbala-Kingebeni, Edith Nkwembe, Eddy Kinganda-Lusamaki, Amuri Aziza, Catherine Pratt, Matthias Pauthner, Josh Quick, Allison Black, James Hadfield, Trevor Bedford, Ian Goodfellow, Nick Loman, Kristian Andersen, Michael Wiley, Steve Ahuka-Mundeke, Jean-Jacques Muyembe Tamfum
EPI_ISL_420845 hCoV-19/DRC/397/2020 Africa / Democratic Republic of the Congo / Kinshasa 2020-03-26 Viral Respiratory Lab, National Institute for Biomedical Research (INRB) Pathogen Sequencing Lab, National Institute for Biomedical Research (INRB) Placide Mbala-Kingebeni, Edith Nkwembe, Eddy Kinganda-Lusamaki, Amuri Aziza, Catherine Pratt, Matthias Pauthner, Josh Quick, Allison Black, James Hadfield, Trevor Bedford, Ian Goodfellow, Nick Loman, Kristian Andersen, Michael Wiley, Steve Ahuka-Mundeke, Jean-Jacques Muyembe Tamfum
EPI_ISL_420846 hCoV-19/DRC/400/2020 Africa / Democratic Republic of the Congo / Kinshasa 2020-03 Viral Respiratory Lab, National Institute for Biomedical Research (INRB) Pathogen Sequencing Lab, National Institute for Biomedical Research (INRB) Placide Mbala-Kingebeni, Edith Nkwembe, Eddy Kinganda-Lusamaki, Amuri Aziza, Catherine Pratt, Matthias Pauthner, Josh Quick, Allison Black, James Hadfield, Trevor Bedford, Ian Goodfellow, Nick Loman, Kristian Andersen, Michael Wiley, Steve Ahuka-Mundeke, Jean-Jacques Muyembe Tamfum
EPI_ISL_420847 hCoV-19/DRC/431/2020 Africa / Democratic Republic of the Congo 2020-03-26 Viral Respiratory Lab, National Institute for Biomedical Research (INRB) Pathogen Sequencing Lab, National Institute for Biomedical Research (INRB) Placide Mbala-Kingebeni, Edith Nkwembe, Eddy Kinganda-Lusamaki, Amuri Aziza, Catherine Pratt, Matthias Pauthner, Josh Quick, Allison Black, James Hadfield, Trevor Bedford, Ian Goodfellow, Nick Loman, Kristian Andersen, Michael Wiley, Steve Ahuka-Mundeke, Jean-Jacques Muyembe Tamfum
EPI_ISL_420849 hCoV-19/DRC/523/2020 Africa / Democratic Republic of the Congo / Kinshasa 2020-03-28 Viral Respiratory Lab, National Institute for Biomedical Research (INRB) Pathogen Sequencing Lab, National Institute for Biomedical Research (INRB) Placide Mbala-Kingebeni, Edith Nkwembe, Eddy Kinganda-Lusamaki, Amuri Aziza, Catherine Pratt, Matthias Pauthner, Josh Quick, Allison Black, James Hadfield, Trevor Bedford, Ian Goodfellow, Nick Loman, Kristian Andersen, Michael Wiley, Steve Ahuka-Mundeke, Jean-Jacques Muyembe Tamfum
EPI_ISL_420851 hCoV-19/DRC/376/2020 Africa / Democratic Republic of the Congo / Kinshasa 2020-03-25 Viral Respiratory Lab, National Institute for Biomedical Research (INRB) Pathogen Sequencing Lab, National Institute for Biomedical Research (INRB) Placide Mbala-Kingebeni, Edith Nkwembe, Eddy Kinganda-Lusamaki, Amuri Aziza, Catherine Pratt, Matthias Pauthner, Josh Quick, Allison Black, James Hadfield, Trevor Bedford, Ian Goodfellow, Nick Loman, Kristian Andersen, Michael Wiley, Steve Ahuka-Mundeke, Jean-Jacques Muyembe Tamfum
EPI_ISL_420852 hCoV-19/DRC/396/2020 Africa / Democratic Republic of the Congo / Kinshasa 2020-03-26 Viral Respiratory Lab, National Institute for Biomedical Research (INRB) Pathogen Sequencing Lab, National Institute for Biomedical Research (INRB) Placide Mbala-Kingebeni, Edith Nkwembe, Eddy Kinganda-Lusamaki, Amuri Aziza, Catherine Pratt, Matthias Pauthner, Josh Quick, Allison Black, James Hadfield, Trevor Bedford, Ian Goodfellow, Nick Loman, Kristian Andersen, Michael Wiley, Steve Ahuka-Mundeke, Jean-Jacques Muyembe Tamfum
EPI_ISL_420854 hCoV-19/DRC/521/2020 Africa / Democratic Republic of the Congo / Kinshasa 2020-03-25 Viral Respiratory Lab, National Institute for Biomedical Research (INRB) Pathogen Sequencing Lab, National Institute for Biomedical Research (INRB) Placide Mbala-Kingebeni, Edith Nkwembe, Eddy Kinganda-Lusamaki, Amuri Aziza, Catherine Pratt, Matthias Pauthner, Josh Quick, Allison Black, James Hadfield, Trevor Bedford, Ian Goodfellow, Nick Loman, Kristian Andersen, Michael Wiley, Steve Ahuka-Mundeke, Jean-Jacques Muyembe Tamfum
EPI_ISL_420855 hCoV-19/Australia/VIC-CBA1/2020 Oceania / Australia / Victoria 2020-03-23 Geelong Centre for Emerging Infectious Diseases Geelong Centre for Emerging Infectious Diseases Chamings,A., Raj Bhatta T., Alexandersen S.
EPI_ISL_420876 hCoV-19/Australia/VIC-CBA2/2020 Oceania / Australia / Victoria 2020-03-24 Geelong Centre for Emerging Infectious Diseases Geelong Centre for Emerging Infectious Diseases Chamings,A., Raj Bhatta T., Alexandersen S.
EPI_ISL_420877 hCoV-19/Australia/VIC-CBA3/2020 Oceania / Australia / Victoria 2020-03-28 Geelong Centre for Emerging Infectious Diseases Geelong Centre for Emerging Infectious Diseases Chamings,A., Raj Bhatta T., Alexandersen S.
EPI_ISL_420878 hCoV-19/Australia/QLDID929/2020 Oceania / Australia / Queensland / Brisbane 2020-03-24 Mater Pathology Public Health Virology Laboratory Bixing Huang, Alyssa Pyke, Amanda De Jong, Andrew Van Den Hurk, Carmel Taylor, David Warrilow, Doris Genge, Elisabeth Gamez, Glen Hewitson, Ian Maxwell Mackay, Inga Sultana, Jamie McMahon, Jean Barcelon, Judy Northill, Mitchell Finger, Natalie Simpson, Neelima Nair, Peter Burtonclay, Peter Moore, Sarah Wheatley, Sean Moody, Sonja Hall-Mendelin, Timothy Gardam, and Frederick Moore
EPI_ISL_420879 hCoV-19/Australia/QLDID931/2020 Oceania / Australia / Queensland / Brisbane 2020-03-25 Mater Pathology Public Health Virology Laboratory Bixing Huang, Alyssa Pyke, Amanda De Jong, Andrew Van Den Hurk, Carmel Taylor, David Warrilow, Doris Genge, Elisabeth Gamez, Glen Hewitson, Ian Maxwell Mackay, Inga Sultana, Jamie McMahon, Jean Barcelon, Judy Northill, Mitchell Finger, Natalie Simpson, Neelima Nair, Peter Burtonclay, Peter Moore, Sarah Wheatley, Sean Moody, Sonja Hall-Mendelin, Timothy Gardam, and Frederick Moore
EPI_ISL_420889 hCoV-19/Japan/Hu_DP_Kng_19-031/2020 Asia / Japan 2020-02-14 unknown Takayuki Hishiki Kanagawa Prefectural Institute of Public Health Hishiki,T., Suzuki,R., Sakuragi,J., Usui,K., Tanaka,Y., Kawai,J., Kogo,Y., Matsuki,Y., An,T., Hayashizaki,Y. and Takasaki,T.

EPI_ISL_420890  hCoV-19/Japan/Hu_Kng_19-437/2020        Asia / Japan    2020-03-09      unknown Takayuki Hishiki Kanagawa Prefectural Institute of Public Health Hishiki,T., Suzuki,R., Sakuragi,J., Usui,K., Tanaka,Y., Kawai,J., Kogo,Y., Matsuki,Y., An,T., Hayashizaki,Y. and Takasaki,T.
EPI_ISL_421800  hCoV-19/England/201380036/2020  Europe / United Kingdom / England       2020-03-26      Respiratory Virus Unit, Microbiology Services Colindale, Public Health England  Respiratory Virus Unit, Microbiology Services Colindale, Public Health England  Monica Galiano, Shahjahan Miah, Angie Lackenby, Omolola Akinbami, Tiina Talts, Leena Bhaw, Richard Myers, Steven Platt, Kirstin Edwards, Jonathan Hubb, Joanna Ellis, Maria Zambon
EPI_ISL_421801  hCoV-19/England/201380037/2020  Europe / United Kingdom / England       2020-03-26      Respiratory Virus Unit, Microbiology Services Colindale, Public Health England  Respiratory Virus Unit, Microbiology Services Colindale, Public Health England  Monica Galiano, Shahjahan Miah, Angie Lackenby, Omolola Akinbami, Tiina Talts, Leena Bhaw, Richard Myers, Steven Platt, Kirstin Edwards, Jonathan Hubb, Joanna Ellis, Maria Zambon
EPI_ISL_421802  hCoV-19/England/201380040/2020  Europe / United Kingdom / England       2020-03-27      Respiratory Virus Unit, Microbiology Services Colindale, Public Health England  Respiratory Virus Unit, Microbiology Services Colindale, Public Health England  Monica Galiano, Shahjahan Miah, Angie Lackenby, Omolola Akinbami, Tiina Talts, Leena Bhaw, Richard Myers, Steven Platt, Kirstin Edwards, Jonathan Hubb, Joanna Ellis, Maria Zambon
EPI_ISL_421803  hCoV-19/England/201380041/2020  Europe / United Kingdom / England       2020-03-26      Respiratory Virus Unit, Microbiology Services Colindale, Public Health England  Respiratory Virus Unit, Microbiology Services Colindale, Public Health England  Monica Galiano, Shahjahan Miah, Angie Lackenby, Omolola Akinbami, Tiina Talts, Leena Bhaw, Richard Myers, Steven Platt, Kirstin Edwards, Jonathan Hubb, Joanna Ellis, Maria Zambon
EPI_ISL_421804  hCoV-19/England/201380042/2020  Europe / United Kingdom / England       2020-03-25      Respiratory Virus Unit, Microbiology Services Colindale, Public Health England  Respiratory Virus Unit, Microbiology Services Colindale, Public Health England  Monica Galiano, Shahjahan Miah, Angie Lackenby, Omolola Akinbami, Tiina Talts, Leena Bhaw, Richard Myers, Steven Platt, Kirstin Edwards, Jonathan Hubb, Joanna Ellis, Maria Zambon
EPI_ISL_421805  hCoV-19/England/201380044/2020  Europe / United Kingdom / England       2020-03-26      Respiratory Virus Unit, Microbiology Services Colindale, Public Health England  Respiratory Virus Unit, Microbiology Services Colindale, Public Health England  Monica Galiano, Shahjahan Miah, Angie Lackenby, Omolola Akinbami, Tiina Talts, Leena Bhaw, Richard Myers, Steven Platt, Kirstin Edwards, Jonathan Hubb, Joanna Ellis, Maria Zambon
EPI_ISL_421806  hCoV-19/England/201380049/2020  Europe / United Kingdom / England       2020-03-25      Respiratory Virus Unit, Microbiology Services Colindale, Public Health England  Respiratory Virus Unit, Microbiology Services Colindale, Public Health England  Monica Galiano, Shahjahan Miah, Angie Lackenby, Omolola Akinbami, Tiina Talts, Leena Bhaw, Richard Myers, Steven Platt, Kirstin Edwards, Jonathan Hubb, Joanna Ellis, Maria Zambon
EPI_ISL_421807  hCoV-19/England/201380050/2020  Europe / United Kingdom / England       2020-03-25      Respiratory Virus Unit, Microbiology Services Colindale, Public Health England  Respiratory Virus Unit, Microbiology Services Colindale, Public Health England  Monica Galiano, Shahjahan Miah, Angie Lackenby, Omolola Akinbami, Tiina Talts, Leena Bhaw, Richard Myers, Steven Platt, Kirstin Edwards, Jonathan Hubb, Joanna Ellis, Maria Zambon
EPI_ISL_421809  hCoV-19/England/201380054/2020  Europe / United Kingdom / England       2020-03-25      Respiratory Virus Unit, Microbiology Services Colindale, Public Health England  Respiratory Virus Unit, Microbiology Services Colindale, Public Health England  Monica Galiano, Shahjahan Miah, Angie Lackenby, Omolola Akinbami, Tiina Talts, Leena Bhaw, Richard Myers, Steven Platt, Kirstin Edwards, Jonathan Hubb, Joanna Ellis, Maria Zambon
EPI_ISL_421810  hCoV-19/England/201380055/2020  Europe / United Kingdom / England       2020-03-25      Respiratory Virus Unit, Microbiology Services Colindale, Public Health England  Respiratory Virus Unit, Microbiology Services Colindale, Public Health England  Monica Galiano, Shahjahan Miah, Angie Lackenby, Omolola Akinbami, Tiina Talts, Leena Bhaw, Richard Myers, Steven Platt, Kirstin Edwards, Jonathan Hubb, Joanna Ellis, Maria Zambon
EPI_ISL_421811  hCoV-19/England/201380056/2020  Europe / United Kingdom / England       2020-03-25      Respiratory Virus Unit, Microbiology Services Colindale, Public Health England  Respiratory Virus Unit, Microbiology Services Colindale, Public Health England  Monica Galiano, Shahjahan Miah, Angie Lackenby, Omolola Akinbami, Tiina Talts, Leena Bhaw, Richard Myers, Steven Platt, Kirstin Edwards, Jonathan Hubb, Joanna Ellis, Maria Zambon
EPI_ISL_421813  hCoV-19/England/201380061/2020  Europe / United Kingdom / England       2020-03-25      Respiratory Virus Unit, Microbiology Services Colindale, Public Health England  Respiratory Virus Unit, Microbiology Services Colindale, Public Health England  Monica Galiano, Shahjahan Miah, Angie Lackenby, Omolola Akinbami, Tiina Talts, Leena Bhaw, Richard Myers, Steven Platt, Kirstin Edwards, Jonathan Hubb, Joanna Ellis, Maria Zambon
EPI_ISL_421815  hCoV-19/England/201380063/2020  Europe / United Kingdom / England       2020-03-25      Respiratory Virus Unit, Microbiology Services Colindale, Public Health England  Respiratory Virus Unit, Microbiology Services Colindale, Public Health England  Monica Galiano, Shahjahan Miah, Angie Lackenby, Omolola Akinbami, Tiina Talts, Leena Bhaw, Richard Myers, Steven Platt, Kirstin Edwards, Jonathan Hubb, Joanna Ellis, Maria Zambon
EPI_ISL_421816  hCoV-19/England/201380064/2020  Europe / United Kingdom / England       2020-03-25      Respiratory Virus Unit, Microbiology Services Colindale, Public Health England  Respiratory Virus Unit,

Microbiology Services Colindale, Public Health England  Monica Galiano, Shahjahan Miah, Angie Lackenby, Omolola Akinbami, Tiina Talts, Leena Bhaw, Richard Myers, Steven Platt, Kirstin Edwards, Jonathan Hubb, Joanna Ellis, Maria Zambon
EPI_ISL_421817  hCoV-19/England/201380069/2020  Europe / United Kingdom / England  2020-03-25  Respiratory Virus Unit, Microbiology Services Colindale, Public Health England  Respiratory Virus Unit, Microbiology Services Colindale, Public Health England  Monica Galiano, Shahjahan Miah, Angie Lackenby, Omolola Akinbami, Tiina Talts, Leena Bhaw, Richard Myers, Steven Platt, Kirstin Edwards, Jonathan Hubb, Joanna Ellis, Maria Zambon
EPI_ISL_421818  hCoV-19/England/201380070/2020  Europe / United Kingdom / England  2020-03-25  Respiratory Virus Unit, Microbiology Services Colindale, Public Health England  Respiratory Virus Unit, Microbiology Services Colindale, Public Health England  Monica Galiano, Shahjahan Miah, Angie Lackenby, Omolola Akinbami, Tiina Talts, Leena Bhaw, Richard Myers, Steven Platt, Kirstin Edwards, Jonathan Hubb, Joanna Ellis, Maria Zambon
EPI_ISL_421822  hCoV-19/England/201380217/2020  Europe / United Kingdom / England  2020-03-25  Respiratory Virus Unit, Microbiology Services Colindale, Public Health England  Respiratory Virus Unit, Microbiology Services Colindale, Public Health England  Monica Galiano, Shahjahan Miah, Angie Lackenby, Omolola Akinbami, Tiina Talts, Leena Bhaw, Richard Myers, Steven Platt, Kirstin Edwards, Jonathan Hubb, Joanna Ellis, Maria Zambon
EPI_ISL_421823  hCoV-19/England/201380221/2020  Europe / United Kingdom / England  2020-03-25  Respiratory Virus Unit, Microbiology Services Colindale, Public Health England  Respiratory Virus Unit, Microbiology Services Colindale, Public Health England  Monica Galiano, Shahjahan Miah, Angie Lackenby, Omolola Akinbami, Tiina Talts, Leena Bhaw, Richard Myers, Steven Platt, Kirstin Edwards, Jonathan Hubb, Joanna Ellis, Maria Zambon
EPI_ISL_421824  hCoV-19/England/201380228/2020  Europe / United Kingdom / England  2020-03-25  Respiratory Virus Unit, Microbiology Services Colindale, Public Health England  Respiratory Virus Unit, Microbiology Services Colindale, Public Health England  Monica Galiano, Shahjahan Miah, Angie Lackenby, Omolola Akinbami, Tiina Talts, Leena Bhaw, Richard Myers, Steven Platt, Kirstin Edwards, Jonathan Hubb, Joanna Ellis, Maria Zambon
EPI_ISL_421825  hCoV-19/England/201380229/2020  Europe / United Kingdom / England  2020-03-25  Respiratory Virus Unit, Microbiology Services Colindale, Public Health England  Respiratory Virus Unit, Microbiology Services Colindale, Public Health England  Monica Galiano, Shahjahan Miah, Angie Lackenby, Omolola Akinbami, Tiina Talts, Leena Bhaw, Richard Myers, Steven Platt, Kirstin Edwards, Jonathan Hubb, Joanna Ellis, Maria Zambon
EPI_ISL_421826  hCoV-19/England/201380276/2020  Europe / United Kingdom / England  2020-03-23  Respiratory Virus Unit, Microbiology Services Colindale, Public Health England  Respiratory Virus Unit, Microbiology Services Colindale, Public Health England  Monica Galiano, Shahjahan Miah, Angie Lackenby, Omolola Akinbami, Tiina Talts, Leena Bhaw, Richard Myers, Steven Platt, Kirstin Edwards, Jonathan Hubb, Joanna Ellis, Maria Zambon
EPI_ISL_421827  hCoV-19/England/201380277/2020  Europe / United Kingdom / England  2020-03-24  Respiratory Virus Unit, Microbiology Services Colindale, Public Health England  Respiratory Virus Unit, Microbiology Services Colindale, Public Health England  Monica Galiano, Shahjahan Miah, Angie Lackenby, Omolola Akinbami, Tiina Talts, Leena Bhaw, Richard Myers, Steven Platt, Kirstin Edwards, Jonathan Hubb, Joanna Ellis, Maria Zambon
EPI_ISL_421828  hCoV-19/England/201380280/2020  Europe / United Kingdom / England  2020-03-23  Respiratory Virus Unit, Microbiology Services Colindale, Public Health England  Respiratory Virus Unit, Microbiology Services Colindale, Public Health England  Monica Galiano, Shahjahan Miah, Angie Lackenby, Omolola Akinbami, Tiina Talts, Leena Bhaw, Richard Myers, Steven Platt, Kirstin Edwards, Jonathan Hubb, Joanna Ellis, Maria Zambon
EPI_ISL_421829  hCoV-19/England/201380281/2020  Europe / United Kingdom / England  2020-03-25  Respiratory Virus Unit, Microbiology Services Colindale, Public Health England  Respiratory Virus Unit, Microbiology Services Colindale, Public Health England  Monica Galiano, Shahjahan Miah, Angie Lackenby, Omolola Akinbami, Tiina Talts, Leena Bhaw, Richard Myers, Steven Platt, Kirstin Edwards, Jonathan Hubb, Joanna Ellis, Maria Zambon
EPI_ISL_421830  hCoV-19/England/201380282/2020  Europe / United Kingdom / England  2020-03-25  Respiratory Virus Unit, Microbiology Services Colindale, Public Health England  Respiratory Virus Unit, Microbiology Services Colindale, Public Health England  Monica Galiano, Shahjahan Miah, Angie Lackenby, Omolola Akinbami, Tiina Talts, Leena Bhaw, Richard Myers, Steven Platt, Kirstin Edwards, Jonathan Hubb, Joanna Ellis, Maria Zambon
EPI_ISL_421831  hCoV-19/England/201380283/2020  Europe / United Kingdom / England  2020-03-23  Respiratory Virus Unit, Microbiology Services Colindale, Public Health England  Respiratory Virus Unit, Microbiology Services Colindale, Public Health England  Monica Galiano, Shahjahan Miah, Angie Lackenby, Omolola Akinbami, Tiina Talts, Leena Bhaw, Richard Myers, Steven Platt, Kirstin Edwards, Jonathan Hubb, Joanna Ellis, Maria Zambon
EPI_ISL_421834  hCoV-19/England/20138082004/2020  Europe / United Kingdom / England  2020-03-26  Respiratory Virus Unit, Microbiology Services Colindale, Public Health England  Respiratory Virus Unit, Microbiology Services Colindale, Public Health England  Monica Galiano, Shahjahan Miah, Angie Lackenby, Omolola Akinbami, Tiina Talts, Leena Bhaw, Richard Myers, Steven Platt, Kirstin Edwards, Jonathan Hubb, Joanna Ellis, Maria Zambon
EPI_ISL_421836  hCoV-19/England/20138082704/2020  Europe / United Kingdom / England  2020-03-26  Respiratory Virus Unit, Microbiology Services Colindale, Public Health England  Respiratory Virus Unit,

Microbiology Services Colindale, Public Health England  Monica Galiano, Shahjahan Miah, Angie Lackenby, Omolola Akinbami, Tiina Talts, Leena Bhaw, Richard Myers, Steven Platt, Kirstin Edwards, Jonathan Hubb, Joanna Ellis, Maria Zambon
EPI_ISL_421837  hCoV-19/England/20138082804/2020  Europe / United Kingdom / England  2020-03-27  Respiratory Virus Unit, Microbiology Services Colindale, Public Health England  Respiratory Virus Unit, Microbiology Services Colindale, Public Health England  Monica Galiano, Shahjahan Miah, Angie Lackenby, Omolola Akinbami, Tiina Talts, Leena Bhaw, Richard Myers, Steven Platt, Kirstin Edwards, Jonathan Hubb, Joanna Ellis, Maria Zambon
EPI_ISL_421839  hCoV-19/England/20138083204/2020  Europe / United Kingdom / England  2020-03-27  Respiratory Virus Unit, Microbiology Services Colindale, Public Health England  Respiratory Virus Unit, Microbiology Services Colindale, Public Health England  Monica Galiano, Shahjahan Miah, Angie Lackenby, Omolola Akinbami, Tiina Talts, Leena Bhaw, Richard Myers, Steven Platt, Kirstin Edwards, Jonathan Hubb, Joanna Ellis, Maria Zambon
EPI_ISL_421842  hCoV-19/England/20138086904/2020  Europe / United Kingdom / England  2020-03-23  Respiratory Virus Unit, Microbiology Services Colindale, Public Health England  Respiratory Virus Unit, Microbiology Services Colindale, Public Health England  Monica Galiano, Shahjahan Miah, Angie Lackenby, Omolola Akinbami, Tiina Talts, Leena Bhaw, Richard Myers, Steven Platt, Kirstin Edwards, Jonathan Hubb, Joanna Ellis, Maria Zambon
EPI_ISL_421845  hCoV-19/England/20138088004/2020  Europe / United Kingdom / England  2020-03-24  Respiratory Virus Unit, Microbiology Services Colindale, Public Health England  Respiratory Virus Unit, Microbiology Services Colindale, Public Health England  Monica Galiano, Shahjahan Miah, Angie Lackenby, Omolola Akinbami, Tiina Talts, Leena Bhaw, Richard Myers, Steven Platt, Kirstin Edwards, Jonathan Hubb, Joanna Ellis, Maria Zambon
EPI_ISL_421847  hCoV-19/England/20138088204/2020  Europe / United Kingdom / England  2020-03-26  Respiratory Virus Unit, Microbiology Services Colindale, Public Health England  Respiratory Virus Unit, Microbiology Services Colindale, Public Health England  Monica Galiano, Shahjahan Miah, Angie Lackenby, Omolola Akinbami, Tiina Talts, Leena Bhaw, Richard Myers, Steven Platt, Kirstin Edwards, Jonathan Hubb, Joanna Ellis, Maria Zambon
EPI_ISL_421862  hCoV-19/England/20139008204/2020  Europe / United Kingdom / England  2020-03-26  Respiratory Virus Unit, Microbiology Services Colindale, Public Health England  Respiratory Virus Unit, Microbiology Services Colindale, Public Health England  Monica Galiano, Shahjahan Miah, Angie Lackenby, Omolola Akinbami, Tiina Talts, Leena Bhaw, Richard Myers, Steven Platt, Kirstin Edwards, Jonathan Hubb, Joanna Ellis, Maria Zambon
EPI_ISL_421863  hCoV-19/England/20139008304/2020  Europe / United Kingdom / England  2020-03-26  Respiratory Virus Unit, Microbiology Services Colindale, Public Health England  Respiratory Virus Unit, Microbiology Services Colindale, Public Health England  Monica Galiano, Shahjahan Miah, Angie Lackenby, Omolola Akinbami, Tiina Talts, Leena Bhaw, Richard Myers, Steven Platt, Kirstin Edwards, Jonathan Hubb, Joanna Ellis, Maria Zambon
EPI_ISL_421864  hCoV-19/England/20139008404/2020  Europe / United Kingdom / England  2020-03-27  Respiratory Virus Unit, Microbiology Services Colindale, Public Health England  Respiratory Virus Unit, Microbiology Services Colindale, Public Health England  Monica Galiano, Shahjahan Miah, Angie Lackenby, Omolola Akinbami, Tiina Talts, Leena Bhaw, Richard Myers, Steven Platt, Kirstin Edwards, Jonathan Hubb, Joanna Ellis, Maria Zambon
EPI_ISL_421865  hCoV-19/England/20139008704/2020  Europe / United Kingdom / England  2020-03-27  Respiratory Virus Unit, Microbiology Services Colindale, Public Health England  Respiratory Virus Unit, Microbiology Services Colindale, Public Health England  Monica Galiano, Shahjahan Miah, Angie Lackenby, Omolola Akinbami, Tiina Talts, Leena Bhaw, Richard Myers, Steven Platt, Kirstin Edwards, Jonathan Hubb, Joanna Ellis, Maria Zambon
EPI_ISL_421866  hCoV-19/England/20139009004/2020  Europe / United Kingdom / England  2020-03-27  Respiratory Virus Unit, Microbiology Services Colindale, Public Health England  Respiratory Virus Unit, Microbiology Services Colindale, Public Health England  Monica Galiano, Shahjahan Miah, Angie Lackenby, Omolola Akinbami, Tiina Talts, Leena Bhaw, Richard Myers, Steven Platt, Kirstin Edwards, Jonathan Hubb, Joanna Ellis, Maria Zambon
EPI_ISL_421867  hCoV-19/England/20139009204/2020  Europe / United Kingdom / England  2020-03-27  Respiratory Virus Unit, Microbiology Services Colindale, Public Health England  Respiratory Virus Unit, Microbiology Services Colindale, Public Health England  Monica Galiano, Shahjahan Miah, Angie Lackenby, Omolola Akinbami, Tiina Talts, Leena Bhaw, Richard Myers, Steven Platt, Kirstin Edwards, Jonathan Hubb, Joanna Ellis, Maria Zambon
EPI_ISL_421868  hCoV-19/England/20139009504/2020  Europe / United Kingdom / England  2020-03-27  Respiratory Virus Unit, Microbiology Services Colindale, Public Health England  Respiratory Virus Unit, Microbiology Services Colindale, Public Health England  Monica Galiano, Shahjahan Miah, Angie Lackenby, Omolola Akinbami, Tiina Talts, Leena Bhaw, Richard Myers, Steven Platt, Kirstin Edwards, Jonathan Hubb, Joanna Ellis, Maria Zambon
EPI_ISL_421869  hCoV-19/England/20139009604/2020  Europe / United Kingdom / England  2020-03-27  Respiratory Virus Unit, Microbiology Services Colindale, Public Health England  Respiratory Virus Unit, Microbiology Services Colindale, Public Health England  Monica Galiano, Shahjahan Miah, Angie Lackenby, Omolola Akinbami, Tiina Talts, Leena Bhaw, Richard Myers, Steven Platt, Kirstin Edwards, Jonathan Hubb, Joanna Ellis, Maria Zambon
EPI_ISL_421870  hCoV-19/England/20139009704/2020  Europe / United Kingdom / England  2020-03-27  Respiratory Virus Unit, Microbiology Services Colindale, Public Health England  Respiratory Virus Unit,

Microbiology Services Colindale, Public Health England  Monica Galiano, Shahjahan Miah, Angie Lackenby, Omolola Akinbami, Tiina Talts, Leena Bhaw, Richard Myers, Steven Platt, Kirstin Edwards, Jonathan Hubb, Joanna Ellis, Maria Zambon
EPI_ISL_421875  hCoV-19/England/20139011304/2020        Europe / United Kingdom / England       2020-03-26
Respiratory Virus Unit, Microbiology Services Colindale, Public Health England  Respiratory Virus Unit, Microbiology Services Colindale, Public Health England  Monica Galiano, Shahjahan Miah, Angie Lackenby, Omolola Akinbami, Tiina Talts, Leena Bhaw, Richard Myers, Steven Platt, Kirstin Edwards, Jonathan Hubb, Joanna Ellis, Maria Zambon
EPI_ISL_421877  hCoV-19/England/20139014404/2020        Europe / United Kingdom / England       2020-03-27
Respiratory Virus Unit, Microbiology Services Colindale, Public Health England  Respiratory Virus Unit, Microbiology Services Colindale, Public Health England  Monica Galiano, Shahjahan Miah, Angie Lackenby, Omolola Akinbami, Tiina Talts, Leena Bhaw, Richard Myers, Steven Platt, Kirstin Edwards, Jonathan Hubb, Joanna Ellis, Maria Zambon
EPI_ISL_421879  hCoV-19/England/20139020204/2020        Europe / United Kingdom / England       2020-03-27
Respiratory Virus Unit, Microbiology Services Colindale, Public Health England  Respiratory Virus Unit, Microbiology Services Colindale, Public Health England  Monica Galiano, Shahjahan Miah, Angie Lackenby, Omolola Akinbami, Tiina Talts, Leena Bhaw, Richard Myers, Steven Platt, Kirstin Edwards, Jonathan Hubb, Joanna Ellis, Maria Zambon
EPI_ISL_421880  hCoV-19/England/20139021104/2020        Europe / United Kingdom / England       2020-03-26
Respiratory Virus Unit, Microbiology Services Colindale, Public Health England  Respiratory Virus Unit, Microbiology Services Colindale, Public Health England  Monica Galiano, Shahjahan Miah, Angie Lackenby, Omolola Akinbami, Tiina Talts, Leena Bhaw, Richard Myers, Steven Platt, Kirstin Edwards, Jonathan Hubb, Joanna Ellis, Maria Zambon
EPI_ISL_421882  hCoV-19/England/20139021604/2020        Europe / United Kingdom / England       2020-03-27
Respiratory Virus Unit, Microbiology Services Colindale, Public Health England  Respiratory Virus Unit, Microbiology Services Colindale, Public Health England  Monica Galiano, Shahjahan Miah, Angie Lackenby, Omolola Akinbami, Tiina Talts, Leena Bhaw, Richard Myers, Steven Platt, Kirstin Edwards, Jonathan Hubb, Joanna Ellis, Maria Zambon
EPI_ISL_421883  hCoV-19/England/20139022004/2020        Europe / United Kingdom / England       2020-03-27
Respiratory Virus Unit, Microbiology Services Colindale, Public Health England  Respiratory Virus Unit, Microbiology Services Colindale, Public Health England  Monica Galiano, Shahjahan Miah, Angie Lackenby, Omolola Akinbami, Tiina Talts, Leena Bhaw, Richard Myers, Steven Platt, Kirstin Edwards, Jonathan Hubb, Joanna Ellis, Maria Zambon
EPI_ISL_421885  hCoV-19/England/20139022504/2020        Europe / United Kingdom / England       2020-03-27
Respiratory Virus Unit, Microbiology Services Colindale, Public Health England  Respiratory Virus Unit, Microbiology Services Colindale, Public Health England  Monica Galiano, Shahjahan Miah, Angie Lackenby, Omolola Akinbami, Tiina Talts, Leena Bhaw, Richard Myers, Steven Platt, Kirstin Edwards, Jonathan Hubb, Joanna Ellis, Maria Zambon
EPI_ISL_421886  hCoV-19/England/20139022704/2020        Europe / United Kingdom / England       2020-03-27
Respiratory Virus Unit, Microbiology Services Colindale, Public Health England  Respiratory Virus Unit, Microbiology Services Colindale, Public Health England  Monica Galiano, Shahjahan Miah, Angie Lackenby, Omolola Akinbami, Tiina Talts, Leena Bhaw, Richard Myers, Steven Platt, Kirstin Edwards, Jonathan Hubb, Joanna Ellis, Maria Zambon
EPI_ISL_421887  hCoV-19/England/20139023504/2020        Europe / United Kingdom / England       2020-03-27
Respiratory Virus Unit, Microbiology Services Colindale, Public Health England  Respiratory Virus Unit, Microbiology Services Colindale, Public Health England  Monica Galiano, Shahjahan Miah, Angie Lackenby, Omolola Akinbami, Tiina Talts, Leena Bhaw, Richard Myers, Steven Platt, Kirstin Edwards, Jonathan Hubb, Joanna Ellis, Maria Zambon
EPI_ISL_421888  hCoV-19/England/20139023604/2020        Europe / United Kingdom / England       2020-03-27
Respiratory Virus Unit, Microbiology Services Colindale, Public Health England  Respiratory Virus Unit, Microbiology Services Colindale, Public Health England  Monica Galiano, Shahjahan Miah, Angie Lackenby, Omolola Akinbami, Tiina Talts, Leena Bhaw, Richard Myers, Steven Platt, Kirstin Edwards, Jonathan Hubb, Joanna Ellis, Maria Zambon
EPI_ISL_421890  hCoV-19/England/20139024004/2020        Europe / United Kingdom / England       2020-03-27
Respiratory Virus Unit, Microbiology Services Colindale, Public Health England  Respiratory Virus Unit, Microbiology Services Colindale, Public Health England  Monica Galiano, Shahjahan Miah, Angie Lackenby, Omolola Akinbami, Tiina Talts, Leena Bhaw, Richard Myers, Steven Platt, Kirstin Edwards, Jonathan Hubb, Joanna Ellis, Maria Zambon
EPI_ISL_421891  hCoV-19/England/20139024204/2020        Europe / United Kingdom / England       2020-03-27
Respiratory Virus Unit, Microbiology Services Colindale, Public Health England  Respiratory Virus Unit, Microbiology Services Colindale, Public Health England  Monica Galiano, Shahjahan Miah, Angie Lackenby, Omolola Akinbami, Tiina Talts, Leena Bhaw, Richard Myers, Steven Platt, Kirstin Edwards, Jonathan Hubb, Joanna Ellis, Maria Zambon
EPI_ISL_421894  hCoV-19/England/20139024804/2020        Europe / United Kingdom / England       2020-03-27
Respiratory Virus Unit, Microbiology Services Colindale, Public Health England  Respiratory Virus Unit, Microbiology Services Colindale, Public Health England  Monica Galiano, Shahjahan Miah, Angie Lackenby, Omolola Akinbami, Tiina Talts, Leena Bhaw, Richard Myers, Steven Platt, Kirstin Edwards, Jonathan Hubb, Joanna Ellis, Maria Zambon
EPI_ISL_421895  hCoV-19/England/20139024904/2020        Europe / United Kingdom / England       2020-03-27
Respiratory Virus Unit, Microbiology Services Colindale, Public Health England  Respiratory Virus Unit,

Microbiology Services Colindale, Public Health England	Monica Galiano, Shahjahan Miah, Angie Lackenby, Omolola Akinbami, Tiina Talts, Leena Bhaw, Richard Myers, Steven Platt, Kirstin Edwards, Jonathan Hubb, Joanna Ellis, Maria Zambon
EPI_ISL_421899	hCoV-19/England/20139026904/2020	Europe / United Kingdom / England	2020-03-26	Respiratory Virus Unit, Microbiology Services Colindale, Public Health England	Respiratory Virus Unit, Microbiology Services Colindale, Public Health England	Monica Galiano, Shahjahan Miah, Angie Lackenby, Omolola Akinbami, Tiina Talts, Leena Bhaw, Richard Myers, Steven Platt, Kirstin Edwards, Jonathan Hubb, Joanna Ellis, Maria Zambon
EPI_ISL_423000	hCoV-19/USA/WA-UW-1420/2020	North America / USA / Washington	2020-03-18	UW Virology Lab	UW Virology Lab	Pavitra Roychoudhury, Hong Xie, Keith Jerome, Alexander Greninger
EPI_ISL_423001	hCoV-19/USA/WA-UW-1567/2020	North America / USA / Washington	2020-03-19	UW Virology Lab	UW Virology Lab	Pavitra Roychoudhury, Hong Xie, Keith Jerome, Alexander Greninger
EPI_ISL_423002	hCoV-19/USA/WA-UW-1593/2020	North America / USA / Washington	2020-03-19	UW Virology Lab	UW Virology Lab	Pavitra Roychoudhury, Hong Xie, Keith Jerome, Alexander Greninger
EPI_ISL_423003	hCoV-19/USA/WA-UW-1583/2020	North America / USA / Washington	2020-03-19	UW Virology Lab	UW Virology Lab	Pavitra Roychoudhury, Hong Xie, Keith Jerome, Alexander Greninger
EPI_ISL_423004	hCoV-19/USA/WA-UW-1581/2020	North America / USA / Washington	2020-03-19	UW Virology Lab	UW Virology Lab	Pavitra Roychoudhury, Hong Xie, Keith Jerome, Alexander Greninger
EPI_ISL_423005	hCoV-19/USA/WA-UW-1603/2020	North America / USA / Washington	2020-03-19	UW Virology Lab	UW Virology Lab	Pavitra Roychoudhury, Hong Xie, Keith Jerome, Alexander Greninger
EPI_ISL_423006	hCoV-19/USA/WA-UW-1611/2020	North America / USA / Washington	2020-03-19	UW Virology Lab	UW Virology Lab	Pavitra Roychoudhury, Hong Xie, Keith Jerome, Alexander Greninger
EPI_ISL_423007	hCoV-19/USA/WA-UW-1448/2020	North America / USA / Washington	2020-03-19	UW Virology Lab	UW Virology Lab	Pavitra Roychoudhury, Hong Xie, Keith Jerome, Alexander Greninger
EPI_ISL_423008	hCoV-19/USA/WA-UW-1426/2020	North America / USA / Washington	2020-03-18	UW Virology Lab	UW Virology Lab	Pavitra Roychoudhury, Hong Xie, Keith Jerome, Alexander Greninger
EPI_ISL_423010	hCoV-19/USA/WA-UW-1446/2020	North America / USA / Washington	2020-03-18	UW Virology Lab	UW Virology Lab	Pavitra Roychoudhury, Hong Xie, Keith Jerome, Alexander Greninger
EPI_ISL_423011	hCoV-19/USA/WA-UW-1561/2020	North America / USA / Washington	2020-03-19	UW Virology Lab	UW Virology Lab	Pavitra Roychoudhury, Hong Xie, Keith Jerome, Alexander Greninger
EPI_ISL_423012	hCoV-19/USA/WA-UW-1588/2020	North America / USA / Washington	2020-03-19	UW Virology Lab	UW Virology Lab	Pavitra Roychoudhury, Hong Xie, Keith Jerome, Alexander Greninger
EPI_ISL_423013	hCoV-19/USA/WA-UW-1586/2020	North America / USA / Washington	2020-03-19	UW Virology Lab	UW Virology Lab	Pavitra Roychoudhury, Hong Xie, Keith Jerome, Alexander Greninger
EPI_ISL_423014	hCoV-19/USA/WA-UW-1587/2020	North America / USA / Washington	2020-03-19	UW Virology Lab	UW Virology Lab	Pavitra Roychoudhury, Hong Xie, Keith Jerome, Alexander Greninger
EPI_ISL_423015	hCoV-19/USA/WA-UW-1552/2020	North America / USA / Washington	2020-03-19	UW Virology Lab	UW Virology Lab	Pavitra Roychoudhury, Hong Xie, Keith Jerome, Alexander Greninger
EPI_ISL_423016	hCoV-19/USA/WA-UW-1578/2020	North America / USA / Washington	2020-03-19	UW Virology Lab	UW Virology Lab	Pavitra Roychoudhury, Hong Xie, Keith Jerome, Alexander Greninger
EPI_ISL_423017	hCoV-19/USA/WA-UW-1591/2020	North America / USA / Washington	2020-03-19	UW Virology Lab	UW Virology Lab	Pavitra Roychoudhury, Hong Xie, Keith Jerome, Alexander Greninger
EPI_ISL_423018	hCoV-19/USA/WA-UW-1576/2020	North America / USA / Washington	2020-03-19	UW Virology Lab	UW Virology Lab	Pavitra Roychoudhury, Hong Xie, Keith Jerome, Alexander Greninger
EPI_ISL_423019	hCoV-19/USA/WA-UW-1574/2020	North America / USA / Washington	2020-03-19	UW Virology Lab	UW Virology Lab	Pavitra Roychoudhury, Hong Xie, Keith Jerome, Alexander Greninger
EPI_ISL_423020	hCoV-19/USA/WA-UW-1570/2020	North America / USA / Washington	2020-03-19	UW Virology Lab	UW Virology Lab	Pavitra Roychoudhury, Hong Xie, Keith Jerome, Alexander Greninger
EPI_ISL_423021	hCoV-19/USA/WA-UW-1569/2020	North America / USA / Washington	2020-03-19	UW Virology Lab	UW Virology Lab	Pavitra Roychoudhury, Hong Xie, Keith Jerome, Alexander Greninger
EPI_ISL_423022	hCoV-19/USA/WA-UW-1562/2020	North America / USA / Washington	2020-03-19	UW Virology Lab	UW Virology Lab	Pavitra Roychoudhury, Hong Xie, Keith Jerome, Alexander Greninger
EPI_ISL_423023	hCoV-19/USA/UN-UW-1449/2020	North America / USA	2020-03	UW Virology Lab	UW Virology Lab	Pavitra Roychoudhury, Hong Xie, Keith Jerome, Alexander Greninger
EPI_ISL_423024	hCoV-19/USA/WA-UW-1435/2020	North America / USA / Washington	2020-03-17	UW Virology Lab	UW Virology Lab	Pavitra Roychoudhury, Hong Xie, Keith Jerome, Alexander Greninger
EPI_ISL_423025	hCoV-19/USA/WA-UW-1441/2020	North America / USA / Washington	2020-03-18	UW Virology Lab	UW Virology Lab	Pavitra Roychoudhury, Hong Xie, Keith Jerome, Alexander Greninger
EPI_ISL_423026	hCoV-19/USA/WA-UW-1440/2020	North America / USA / Washington	2020-03-18	UW Virology Lab	UW Virology Lab	Pavitra Roychoudhury, Hong Xie, Keith Jerome, Alexander Greninger
EPI_ISL_423027	hCoV-19/USA/WA-UW-1473/2020	North America / USA / Washington	2020-03-18	UW Virology Lab	UW Virology Lab	Pavitra Roychoudhury, Hong Xie, Keith Jerome, Alexander Greninger
EPI_ISL_423028	hCoV-19/USA/WA-UW-1470/2020	North America / USA / Washington	2020-03-18	UW Virology Lab	UW Virology Lab	Pavitra Roychoudhury, Hong Xie, Keith Jerome, Alexander Greninger
EPI_ISL_423029	hCoV-19/USA/WA-UW-1471/2020	North America / USA / Washington	2020-03-18	UW Virology Lab	UW Virology Lab	Pavitra Roychoudhury, Hong Xie, Keith Jerome, Alexander Greninger
EPI_ISL_423030	hCoV-19/USA/WA-UW-1433/2020	North America / USA / Washington	2020-03-18	UW Virology Lab	UW Virology Lab	Pavitra Roychoudhury, Hong Xie, Keith Jerome, Alexander Greninger
EPI_ISL_423031	hCoV-19/USA/WA-UW-1425/2020	North America / USA / Washington	2020-03-18	UW Virology Lab	UW Virology Lab	Pavitra Roychoudhury, Hong Xie, Keith Jerome, Alexander Greninger

EPI_ISL_423032 hCoV-19/USA/WA-UW-1608/2020 North America / USA / Washington 2020-03-18 UW Virology Lab UW Virology Lab Pavitra Roychoudhury, Hong Xie, Keith Jerome, Alexander Greninger
EPI_ISL_423033 hCoV-19/USA/WA-UW-1599/2020 North America / USA / Washington 2020-03-18 UW Virology Lab UW Virology Lab Pavitra Roychoudhury, Hong Xie, Keith Jerome, Alexander Greninger
EPI_ISL_423039 hCoV-19/Thailand/Bangkok-0017/2020 Asia / Thailand / Bangkok 2020-03-21 Ramathibodi Hospital COVID-19 Network Investigations (CONI) Alliance Elizabeth Batty, Wasun Chantratita, Thanat Chookajorn, Stefan Fernandez, Angkana Huang, Anthony R. Jones, Khajohn Joonsalak, Chonticha Klungtong, Theerarat Kochakarn, Namfon Kotanan, Krittikorn Kumpornsin, Wudtichai Manasatienkij, Bhakbhoom Panthan, Ekawat Pasomsub, Insee Sensorn, Arporn Wangwiwatsin
EPI_ISL_423040 hCoV-19/Thailand/Bangkok-0019/2020 Asia / Thailand / Bangkok 2020-03-20 Ramathibodi Hospital COVID-19 Network Investigations (CONI) Alliance Elizabeth Batty, Wasun Chantratita, Thanat Chookajorn, Stefan Fernandez, Angkana Huang, Anthony R. Jones, Khajohn Joonsalak, Chonticha Klungtong, Theerarat Kochakarn, Namfon Kotanan, Krittikorn Kumpornsin, Wudtichai Manasatienkij, Bhakbhoom Panthan, Ekawat Pasomsub, Insee Sensorn, Arporn Wangwiwatsin
EPI_ISL_423041 hCoV-19/Thailand/Bangkok-0020/2020 Asia / Thailand / Bangkok 2020-03-21 Ramathibodi Hospital COVID-19 Network Investigations (CONI) Alliance Elizabeth Batty, Wasun Chantratita, Thanat Chookajorn, Stefan Fernandez, Angkana Huang, Anthony R. Jones, Khajohn Joonsalak, Chonticha Klungtong, Theerarat Kochakarn, Namfon Kotanan, Krittikorn Kumpornsin, Wudtichai Manasatienkij, Bhakbhoom Panthan, Ekawat Pasomsub, Insee Sensorn, Arporn Wangwiwatsin
EPI_ISL_423042 hCoV-19/Thailand/Bangkok-0021/2020 Asia / Thailand / Bangkok 2020-03-24 Ramathibodi Hospital COVID-19 Network Investigations (CONI) Alliance Elizabeth Batty, Wasun Chantratita, Thanat Chookajorn, Stefan Fernandez, Angkana Huang, Anthony R. Jones, Khajohn Joonsalak, Chonticha Klungtong, Theerarat Kochakarn, Namfon Kotanan, Krittikorn Kumpornsin, Wudtichai Manasatienkij, Bhakbhoom Panthan, Ekawat Pasomsub, Insee Sensorn, Arporn Wangwiwatsin
EPI_ISL_423043 hCoV-19/Thailand/Bangkok-0022/2020 Asia / Thailand / Bangkok 2020-03-18 Ramathibodi Hospital COVID-19 Network Investigations (CONI) Alliance Elizabeth Batty, Wasun Chantratita, Thanat Chookajorn, Stefan Fernandez, Angkana Huang, Anthony R. Jones, Khajohn Joonsalak, Chonticha Klungtong, Theerarat Kochakarn, Namfon Kotanan, Krittikorn Kumpornsin, Wudtichai Manasatienkij, Bhakbhoom Panthan, Ekawat Pasomsub, Insee Sensorn, Arporn Wangwiwatsin
EPI_ISL_423044 hCoV-19/England/20134000704/2020 Europe / United Kingdom / England 2020-03-25 Respiratory Virus Unit, Microbiology Services Colindale, Public Health England Respiratory Virus Unit, Microbiology Services Colindale, Public Health England Monica Galiano, Shahjahan Miah, Angie Lackenby, Omolola Akinbami, Tiina Talts, Leena Bhaw, Richard Myers, Steven Platt, Kirstin Edwards, Jonathan Hubb, Joanna Ellis, Maria Zambon
EPI_ISL_423045 hCoV-19/England/20134000804/2020 Europe / United Kingdom / England 2020-03-24 Respiratory Virus Unit, Microbiology Services Colindale, Public Health England Respiratory Virus Unit, Microbiology Services Colindale, Public Health England Monica Galiano, Shahjahan Miah, Angie Lackenby, Omolola Akinbami, Tiina Talts, Leena Bhaw, Richard Myers, Steven Platt, Kirstin Edwards, Jonathan Hubb, Joanna Ellis, Maria Zambon
EPI_ISL_423047 hCoV-19/England/20134001104/2020 Europe / United Kingdom / England 2020-03-24 Respiratory Virus Unit, Microbiology Services Colindale, Public Health England Respiratory Virus Unit, Microbiology Services Colindale, Public Health England Monica Galiano, Shahjahan Miah, Angie Lackenby, Omolola Akinbami, Tiina Talts, Leena Bhaw, Richard Myers, Steven Platt, Kirstin Edwards, Jonathan Hubb, Joanna Ellis, Maria Zambon
EPI_ISL_423052 hCoV-19/England/20134001904/2020 Europe / United Kingdom / England 2020-03-19 Respiratory Virus Unit, Microbiology Services Colindale, Public Health England Respiratory Virus Unit, Microbiology Services Colindale, Public Health England Monica Galiano, Shahjahan Miah, Angie Lackenby, Omolola Akinbami, Tiina Talts, Leena Bhaw, Richard Myers, Steven Platt, Kirstin Edwards, Jonathan Hubb, Joanna Ellis, Maria Zambon
EPI_ISL_423053 hCoV-19/England/20134002104/2020 Europe / United Kingdom / England 2020-03-18 Respiratory Virus Unit, Microbiology Services Colindale, Public Health England Respiratory Virus Unit, Microbiology Services Colindale, Public Health England Monica Galiano, Shahjahan Miah, Angie Lackenby, Omolola Akinbami, Tiina Talts, Leena Bhaw, Richard Myers, Steven Platt, Kirstin Edwards, Jonathan Hubb, Joanna Ellis, Maria Zambon
EPI_ISL_423055 hCoV-19/England/20134003004/2020 Europe / United Kingdom / England 2020-03-19 Respiratory Virus Unit, Microbiology Services Colindale, Public Health England Respiratory Virus Unit, Microbiology Services Colindale, Public Health England Monica Galiano, Shahjahan Miah, Angie Lackenby, Omolola Akinbami, Tiina Talts, Leena Bhaw, Richard Myers, Steven Platt, Kirstin Edwards, Jonathan Hubb, Joanna Ellis, Maria Zambon
EPI_ISL_423057 hCoV-19/England/20134003204/2020 Europe / United Kingdom / England 2020-03-23 Respiratory Virus Unit, Microbiology Services Colindale, Public Health England Respiratory Virus Unit, Microbiology Services Colindale, Public Health England Monica Galiano, Shahjahan Miah, Angie Lackenby, Omolola Akinbami, Tiina Talts, Leena Bhaw, Richard Myers, Steven Platt, Kirstin Edwards, Jonathan Hubb, Joanna Ellis, Maria Zambon
EPI_ISL_423059 hCoV-19/England/20134003404/2020 Europe / United Kingdom / England 2020-03-24 Respiratory Virus Unit, Microbiology Services Colindale, Public Health England Respiratory Virus Unit, Microbiology Services Colindale, Public Health England Monica Galiano, Shahjahan Miah, Angie Lackenby, Omolola Akinbami, Tiina Talts, Leena Bhaw, Richard Myers, Steven Platt, Kirstin Edwards, Jonathan Hubb, Joanna Ellis, Maria Zambon
EPI_ISL_423063 hCoV-19/England/20134004704/2020 Europe / United Kingdom / England 2020-03-24

Respiratory Virus Unit, Microbiology Services Colindale, Public Health England  Respiratory Virus Unit, Microbiology Services Colindale, Public Health England  Monica Galiano, Shahjahan Miah, Angie Lackenby, Omolola Akinbami, Tiina Talts, Leena Bhaw, Richard Myers, Steven Platt, Kirstin Edwards, Jonathan Hubb, Joanna Ellis, Maria Zambon
EPI_ISL_423065  hCoV-19/England/20134004904/2020  Europe / United Kingdom / England  2020-03-24  Respiratory Virus Unit, Microbiology Services Colindale, Public Health England  Respiratory Virus Unit, Microbiology Services Colindale, Public Health England  Monica Galiano, Shahjahan Miah, Angie Lackenby, Omolola Akinbami, Tiina Talts, Leena Bhaw, Richard Myers, Steven Platt, Kirstin Edwards, Jonathan Hubb, Joanna Ellis, Maria Zambon
EPI_ISL_423066  hCoV-19/England/20134005004/2020  Europe / United Kingdom / England  2020-03-24  Respiratory Virus Unit, Microbiology Services Colindale, Public Health England  Respiratory Virus Unit, Microbiology Services Colindale, Public Health England  Monica Galiano, Shahjahan Miah, Angie Lackenby, Omolola Akinbami, Tiina Talts, Leena Bhaw, Richard Myers, Steven Platt, Kirstin Edwards, Jonathan Hubb, Joanna Ellis, Maria Zambon
EPI_ISL_423067  hCoV-19/England/20134005104/2020  Europe / United Kingdom / England  2020-03-24  Respiratory Virus Unit, Microbiology Services Colindale, Public Health England  Respiratory Virus Unit, Microbiology Services Colindale, Public Health England  Monica Galiano, Shahjahan Miah, Angie Lackenby, Omolola Akinbami, Tiina Talts, Leena Bhaw, Richard Myers, Steven Platt, Kirstin Edwards, Jonathan Hubb, Joanna Ellis, Maria Zambon
EPI_ISL_423068  hCoV-19/England/20134005304/2020  Europe / United Kingdom / England  2020-03-23  Respiratory Virus Unit, Microbiology Services Colindale, Public Health England  Respiratory Virus Unit, Microbiology Services Colindale, Public Health England  Monica Galiano, Shahjahan Miah, Angie Lackenby, Omolola Akinbami, Tiina Talts, Leena Bhaw, Richard Myers, Steven Platt, Kirstin Edwards, Jonathan Hubb, Joanna Ellis, Maria Zambon
EPI_ISL_423069  hCoV-19/England/20134006404/2020  Europe / United Kingdom / England  2020-03-24  Respiratory Virus Unit, Microbiology Services Colindale, Public Health England  Respiratory Virus Unit, Microbiology Services Colindale, Public Health England  Monica Galiano, Shahjahan Miah, Angie Lackenby, Omolola Akinbami, Tiina Talts, Leena Bhaw, Richard Myers, Steven Platt, Kirstin Edwards, Jonathan Hubb, Joanna Ellis, Maria Zambon
EPI_ISL_423070  hCoV-19/England/20134006504/2020  Europe / United Kingdom / England  2020-03-24  Respiratory Virus Unit, Microbiology Services Colindale, Public Health England  Respiratory Virus Unit, Microbiology Services Colindale, Public Health England  Monica Galiano, Shahjahan Miah, Angie Lackenby, Omolola Akinbami, Tiina Talts, Leena Bhaw, Richard Myers, Steven Platt, Kirstin Edwards, Jonathan Hubb, Joanna Ellis, Maria Zambon
EPI_ISL_423071  hCoV-19/England/20134006604/2020  Europe / United Kingdom / England  2020-03-23  Respiratory Virus Unit, Microbiology Services Colindale, Public Health England  Respiratory Virus Unit, Microbiology Services Colindale, Public Health England  Monica Galiano, Shahjahan Miah, Angie Lackenby, Omolola Akinbami, Tiina Talts, Leena Bhaw, Richard Myers, Steven Platt, Kirstin Edwards, Jonathan Hubb, Joanna Ellis, Maria Zambon
EPI_ISL_423072  hCoV-19/England/20134006704/2020  Europe / United Kingdom / England  2020-03-24  Respiratory Virus Unit, Microbiology Services Colindale, Public Health England  Respiratory Virus Unit, Microbiology Services Colindale, Public Health England  Monica Galiano, Shahjahan Miah, Angie Lackenby, Omolola Akinbami, Tiina Talts, Leena Bhaw, Richard Myers, Steven Platt, Kirstin Edwards, Jonathan Hubb, Joanna Ellis, Maria Zambon
EPI_ISL_423073  hCoV-19/England/20134007004/2020  Europe / United Kingdom / England  2020-03-24  Respiratory Virus Unit, Microbiology Services Colindale, Public Health England  Respiratory Virus Unit, Microbiology Services Colindale, Public Health England  Monica Galiano, Shahjahan Miah, Angie Lackenby, Omolola Akinbami, Tiina Talts, Leena Bhaw, Richard Myers, Steven Platt, Kirstin Edwards, Jonathan Hubb, Joanna Ellis, Maria Zambon
EPI_ISL_423074  hCoV-19/England/20134007104/2020  Europe / United Kingdom / England  2020-03-24  Respiratory Virus Unit, Microbiology Services Colindale, Public Health England  Respiratory Virus Unit, Microbiology Services Colindale, Public Health England  Monica Galiano, Shahjahan Miah, Angie Lackenby, Omolola Akinbami, Tiina Talts, Leena Bhaw, Richard Myers, Steven Platt, Kirstin Edwards, Jonathan Hubb, Joanna Ellis, Maria Zambon
EPI_ISL_423075  hCoV-19/England/20134007204/2020  Europe / United Kingdom / England  2020-03-23  Respiratory Virus Unit, Microbiology Services Colindale, Public Health England  Respiratory Virus Unit, Microbiology Services Colindale, Public Health England  Monica Galiano, Shahjahan Miah, Angie Lackenby, Omolola Akinbami, Tiina Talts, Leena Bhaw, Richard Myers, Steven Platt, Kirstin Edwards, Jonathan Hubb, Joanna Ellis, Maria Zambon
EPI_ISL_423076  hCoV-19/England/20134009504/2020  Europe / United Kingdom / England  2020-03-24  Respiratory Virus Unit, Microbiology Services Colindale, Public Health England  Respiratory Virus Unit, Microbiology Services Colindale, Public Health England  Monica Galiano, Shahjahan Miah, Angie Lackenby, Omolola Akinbami, Tiina Talts, Leena Bhaw, Richard Myers, Steven Platt, Kirstin Edwards, Jonathan Hubb, Joanna Ellis, Maria Zambon
EPI_ISL_423078  hCoV-19/England/20134010904/2020  Europe / United Kingdom / England  2020-03-25  Respiratory Virus Unit, Microbiology Services Colindale, Public Health England  Respiratory Virus Unit, Microbiology Services Colindale, Public Health England  Monica Galiano, Shahjahan Miah, Angie Lackenby, Omolola Akinbami, Tiina Talts, Leena Bhaw, Richard Myers, Steven Platt, Kirstin Edwards, Jonathan Hubb, Joanna Ellis, Maria Zambon
EPI_ISL_423081  hCoV-19/England/201340120/2020  Europe / United Kingdom / England  2020-03-24

Respiratory Virus Unit, Microbiology Services Colindale, Public Health England Respiratory Virus Unit, Microbiology Services Colindale, Public Health England Monica Galiano, Shahjahan Miah, Angie Lackenby, Omolola Akinbami, Tiina Talts, Leena Bhaw, Richard Myers, Steven Platt, Kirstin Edwards, Jonathan Hubb, Joanna Ellis, Maria Zambon
EPI_ISL_423084 hCoV-19/England/201340127/2020 Europe / United Kingdom / England 2020-03-24 Respiratory Virus Unit, Microbiology Services Colindale, Public Health England Respiratory Virus Unit, Microbiology Services Colindale, Public Health England Monica Galiano, Shahjahan Miah, Angie Lackenby, Omolola Akinbami, Tiina Talts, Leena Bhaw, Richard Myers, Steven Platt, Kirstin Edwards, Jonathan Hubb, Joanna Ellis, Maria Zambon
EPI_ISL_423085 hCoV-19/England/20134015504/2020 Europe / United Kingdom / England 2020-03-25 Respiratory Virus Unit, Microbiology Services Colindale, Public Health England Respiratory Virus Unit, Microbiology Services Colindale, Public Health England Monica Galiano, Shahjahan Miah, Angie Lackenby, Omolola Akinbami, Tiina Talts, Leena Bhaw, Richard Myers, Steven Platt, Kirstin Edwards, Jonathan Hubb, Joanna Ellis, Maria Zambon
EPI_ISL_423086 hCoV-19/England/20134015704/2020 Europe / United Kingdom / England 2020-03-24 Respiratory Virus Unit, Microbiology Services Colindale, Public Health England Respiratory Virus Unit, Microbiology Services Colindale, Public Health England Monica Galiano, Shahjahan Miah, Angie Lackenby, Omolola Akinbami, Tiina Talts, Leena Bhaw, Richard Myers, Steven Platt, Kirstin Edwards, Jonathan Hubb, Joanna Ellis, Maria Zambon
EPI_ISL_423087 hCoV-19/England/20134015902/2020 Europe / United Kingdom / England 2020-03-23 Respiratory Virus Unit, Microbiology Services Colindale, Public Health England Respiratory Virus Unit, Microbiology Services Colindale, Public Health England Monica Galiano, Shahjahan Miah, Angie Lackenby, Omolola Akinbami, Tiina Talts, Leena Bhaw, Richard Myers, Steven Platt, Kirstin Edwards, Jonathan Hubb, Joanna Ellis, Maria Zambon
EPI_ISL_424001 hCoV-19/England/20129016104/2020 Europe / United Kingdom / England 2020-03-19 Respiratory Virus Unit, Microbiology Services Colindale, Public Health England Respiratory Virus Unit, Microbiology Services Colindale, Public Health England Monica Galiano, Shahjahan Miah, Angie Lackenby, Omolola Akinbami, Tiina Talts, Leena Bhaw, Richard Myers, Steven Platt, Kirstin Edwards, Jonathan Hubb, Joanna Ellis, Maria Zambon
EPI_ISL_424003 hCoV-19/England/20129017104/2020 Europe / United Kingdom / England 2020-03-20 Respiratory Virus Unit, Microbiology Services Colindale, Public Health England Respiratory Virus Unit, Microbiology Services Colindale, Public Health England Monica Galiano, Shahjahan Miah, Angie Lackenby, Omolola Akinbami, Tiina Talts, Leena Bhaw, Richard Myers, Steven Platt, Kirstin Edwards, Jonathan Hubb, Joanna Ellis, Maria Zambon
EPI_ISL_424006 hCoV-19/England/20129018604/2020 Europe / United Kingdom / England 2020-03-20 Respiratory Virus Unit, Microbiology Services Colindale, Public Health England Respiratory Virus Unit, Microbiology Services Colindale, Public Health England Monica Galiano, Shahjahan Miah, Angie Lackenby, Omolola Akinbami, Tiina Talts, Leena Bhaw, Richard Myers, Steven Platt, Kirstin Edwards, Jonathan Hubb, Joanna Ellis, Maria Zambon
EPI_ISL_424007 hCoV-19/England/20129018904/2020 Europe / United Kingdom / England 2020-03-20 Respiratory Virus Unit, Microbiology Services Colindale, Public Health England Respiratory Virus Unit, Microbiology Services Colindale, Public Health England Monica Galiano, Shahjahan Miah, Angie Lackenby, Omolola Akinbami, Tiina Talts, Leena Bhaw, Richard Myers, Steven Platt, Kirstin Edwards, Jonathan Hubb, Joanna Ellis, Maria Zambon
EPI_ISL_424009 hCoV-19/England/20129020004/2020 Europe / United Kingdom / England 2020-03-20 Respiratory Virus Unit, Microbiology Services Colindale, Public Health England Respiratory Virus Unit, Microbiology Services Colindale, Public Health England Monica Galiano, Shahjahan Miah, Angie Lackenby, Omolola Akinbami, Tiina Talts, Leena Bhaw, Richard Myers, Steven Platt, Kirstin Edwards, Jonathan Hubb, Joanna Ellis, Maria Zambon
EPI_ISL_424010 hCoV-19/England/20129020304/2020 Europe / United Kingdom / England 2020-03-20 Respiratory Virus Unit, Microbiology Services Colindale, Public Health England Respiratory Virus Unit, Microbiology Services Colindale, Public Health England Monica Galiano, Shahjahan Miah, Angie Lackenby, Omolola Akinbami, Tiina Talts, Leena Bhaw, Richard Myers, Steven Platt, Kirstin Edwards, Jonathan Hubb, Joanna Ellis, Maria Zambon
EPI_ISL_424012 hCoV-19/England/20129020604/2020 Europe / United Kingdom / England 2020-03-20 Respiratory Virus Unit, Microbiology Services Colindale, Public Health England Respiratory Virus Unit, Microbiology Services Colindale, Public Health England Monica Galiano, Shahjahan Miah, Angie Lackenby, Omolola Akinbami, Tiina Talts, Leena Bhaw, Richard Myers, Steven Platt, Kirstin Edwards, Jonathan Hubb, Joanna Ellis, Maria Zambon
EPI_ISL_424015 hCoV-19/England/20129023104/2020 Europe / United Kingdom / England 2020-03-19 Respiratory Virus Unit, Microbiology Services Colindale, Public Health England Respiratory Virus Unit, Microbiology Services Colindale, Public Health England Monica Galiano, Shahjahan Miah, Angie Lackenby, Omolola Akinbami, Tiina Talts, Leena Bhaw, Richard Myers, Steven Platt, Kirstin Edwards, Jonathan Hubb, Joanna Ellis, Maria Zambon
EPI_ISL_424017 hCoV-19/England/20129025404/2020 Europe / United Kingdom / England 2020-03-12 Respiratory Virus Unit, Microbiology Services Colindale, Public Health England Respiratory Virus Unit, Microbiology Services Colindale, Public Health England Monica Galiano, Shahjahan Miah, Angie Lackenby, Omolola Akinbami, Tiina Talts, Leena Bhaw, Richard Myers, Steven Platt, Kirstin Edwards, Jonathan Hubb, Joanna Ellis, Maria Zambon
EPI_ISL_424019 hCoV-19/England/20129031804/2020 Europe / United Kingdom / England 2020-03-19

Respiratory Virus Unit, Microbiology Services Colindale, Public Health England  Respiratory Virus Unit, Microbiology Services Colindale, Public Health England  Monica Galiano, Shahjahan Miah, Angie Lackenby, Omolola Akinbami, Tiina Talts, Leena Bhaw, Richard Myers, Steven Platt, Kirstin Edwards, Jonathan Hubb, Joanna Ellis, Maria Zambon
EPI_ISL_424021  hCoV-19/England/20129032404/2020  Europe / United Kingdom / England  2020-03-20  Respiratory Virus Unit, Microbiology Services Colindale, Public Health England  Respiratory Virus Unit, Microbiology Services Colindale, Public Health England  Monica Galiano, Shahjahan Miah, Angie Lackenby, Omolola Akinbami, Tiina Talts, Leena Bhaw, Richard Myers, Steven Platt, Kirstin Edwards, Jonathan Hubb, Joanna Ellis, Maria Zambon
EPI_ISL_424024  hCoV-19/England/20129033804/2020  Europe / United Kingdom / England  2020-03-20  Respiratory Virus Unit, Microbiology Services Colindale, Public Health England  Respiratory Virus Unit, Microbiology Services Colindale, Public Health England  Monica Galiano, Shahjahan Miah, Angie Lackenby, Omolola Akinbami, Tiina Talts, Leena Bhaw, Richard Myers, Steven Platt, Kirstin Edwards, Jonathan Hubb, Joanna Ellis, Maria Zambon
EPI_ISL_424025  hCoV-19/England/20129033904/2020  Europe / United Kingdom / England  2020-03-20  Respiratory Virus Unit, Microbiology Services Colindale, Public Health England  Respiratory Virus Unit, Microbiology Services Colindale, Public Health England  Monica Galiano, Shahjahan Miah, Angie Lackenby, Omolola Akinbami, Tiina Talts, Leena Bhaw, Richard Myers, Steven Platt, Kirstin Edwards, Jonathan Hubb, Joanna Ellis, Maria Zambon
EPI_ISL_424028  hCoV-19/England/20129035304/2020  Europe / United Kingdom / England  2020-03-21  Respiratory Virus Unit, Microbiology Services Colindale, Public Health England  Respiratory Virus Unit, Microbiology Services Colindale, Public Health England  Monica Galiano, Shahjahan Miah, Angie Lackenby, Omolola Akinbami, Tiina Talts, Leena Bhaw, Richard Myers, Steven Platt, Kirstin Edwards, Jonathan Hubb, Joanna Ellis, Maria Zambon
EPI_ISL_424029  hCoV-19/England/20129035404/2020  Europe / United Kingdom / England  2020-03-20  Respiratory Virus Unit, Microbiology Services Colindale, Public Health England  Respiratory Virus Unit, Microbiology Services Colindale, Public Health England  Monica Galiano, Shahjahan Miah, Angie Lackenby, Omolola Akinbami, Tiina Talts, Leena Bhaw, Richard Myers, Steven Platt, Kirstin Edwards, Jonathan Hubb, Joanna Ellis, Maria Zambon
EPI_ISL_424031  hCoV-19/England/20129035804/2020  Europe / United Kingdom / England  2020-03-20  Respiratory Virus Unit, Microbiology Services Colindale, Public Health England  Respiratory Virus Unit, Microbiology Services Colindale, Public Health England  Monica Galiano, Shahjahan Miah, Angie Lackenby, Omolola Akinbami, Tiina Talts, Leena Bhaw, Richard Myers, Steven Platt, Kirstin Edwards, Jonathan Hubb, Joanna Ellis, Maria Zambon
EPI_ISL_424032  hCoV-19/England/20129036004/2020  Europe / United Kingdom / England  2020-03-20  Respiratory Virus Unit, Microbiology Services Colindale, Public Health England  Respiratory Virus Unit, Microbiology Services Colindale, Public Health England  Monica Galiano, Shahjahan Miah, Angie Lackenby, Omolola Akinbami, Tiina Talts, Leena Bhaw, Richard Myers, Steven Platt, Kirstin Edwards, Jonathan Hubb, Joanna Ellis, Maria Zambon
EPI_ISL_424034  hCoV-19/England/20129036404/2020  Europe / United Kingdom / England  2020-03-20  Respiratory Virus Unit, Microbiology Services Colindale, Public Health England  Respiratory Virus Unit, Microbiology Services Colindale, Public Health England  Monica Galiano, Shahjahan Miah, Angie Lackenby, Omolola Akinbami, Tiina Talts, Leena Bhaw, Richard Myers, Steven Platt, Kirstin Edwards, Jonathan Hubb, Joanna Ellis, Maria Zambon
EPI_ISL_424035  hCoV-19/England/20129036504/2020  Europe / United Kingdom / England  2020-03-20  Respiratory Virus Unit, Microbiology Services Colindale, Public Health England  Respiratory Virus Unit, Microbiology Services Colindale, Public Health England  Monica Galiano, Shahjahan Miah, Angie Lackenby, Omolola Akinbami, Tiina Talts, Leena Bhaw, Richard Myers, Steven Platt, Kirstin Edwards, Jonathan Hubb, Joanna Ellis, Maria Zambon
EPI_ISL_424037  hCoV-19/England/20129037204/2020  Europe / United Kingdom / England  2020-03-20  Respiratory Virus Unit, Microbiology Services Colindale, Public Health England  Respiratory Virus Unit, Microbiology Services Colindale, Public Health England  Monica Galiano, Shahjahan Miah, Angie Lackenby, Omolola Akinbami, Tiina Talts, Leena Bhaw, Richard Myers, Steven Platt, Kirstin Edwards, Jonathan Hubb, Joanna Ellis, Maria Zambon
EPI_ISL_424038  hCoV-19/England/20129037304/2020  Europe / United Kingdom / England  2020-03-20  Respiratory Virus Unit, Microbiology Services Colindale, Public Health England  Respiratory Virus Unit, Microbiology Services Colindale, Public Health England  Monica Galiano, Shahjahan Miah, Angie Lackenby, Omolola Akinbami, Tiina Talts, Leena Bhaw, Richard Myers, Steven Platt, Kirstin Edwards, Jonathan Hubb, Joanna Ellis, Maria Zambon
EPI_ISL_424052  hCoV-19/England/20129056804/2020  Europe / United Kingdom / England  2020-03-21  Respiratory Virus Unit, Microbiology Services Colindale, Public Health England  Respiratory Virus Unit, Microbiology Services Colindale, Public Health England  Monica Galiano, Shahjahan Miah, Angie Lackenby, Omolola Akinbami, Tiina Talts, Leena Bhaw, Richard Myers, Steven Platt, Kirstin Edwards, Jonathan Hubb, Joanna Ellis, Maria Zambon
EPI_ISL_424054  hCoV-19/England/20129059204/2020  Europe / United Kingdom / England  2020-03-09  Respiratory Virus Unit, Microbiology Services Colindale, Public Health England  Respiratory Virus Unit, Microbiology Services Colindale, Public Health England  Monica Galiano, Shahjahan Miah, Angie Lackenby, Omolola Akinbami, Tiina Talts, Leena Bhaw, Richard Myers, Steven Platt, Kirstin Edwards, Jonathan Hubb, Joanna Ellis, Maria Zambon
EPI_ISL_424062  hCoV-19/England/20129063804/2020  Europe / United Kingdom / England  2020-03-11

Respiratory Virus Unit, Microbiology Services Colindale, Public Health England  Respiratory Virus Unit, Microbiology Services Colindale, Public Health England  Monica Galiano, Shahjahan Miah, Angie Lackenby, Omolola Akinbami, Tiina Talts, Leena Bhaw, Richard Myers, Steven Platt, Kirstin Edwards, Jonathan Hubb, Joanna Ellis, Maria Zambon
EPI_ISL_424076  hCoV-19/England/20129085704/2020        Europe / United Kingdom / England       2020-03-21      Respiratory Virus Unit, Microbiology Services Colindale, Public Health England  Respiratory Virus Unit, Microbiology Services Colindale, Public Health England  Monica Galiano, Shahjahan Miah, Angie Lackenby, Omolola Akinbami, Tiina Talts, Leena Bhaw, Richard Myers, Steven Platt, Kirstin Edwards, Jonathan Hubb, Joanna Ellis, Maria Zambon
EPI_ISL_424078  hCoV-19/England/20129089504/2020        Europe / United Kingdom / England       2020-03-11      Respiratory Virus Unit, Microbiology Services Colindale, Public Health England  Respiratory Virus Unit, Microbiology Services Colindale, Public Health England  Monica Galiano, Shahjahan Miah, Angie Lackenby, Omolola Akinbami, Tiina Talts, Leena Bhaw, Richard Myers, Steven Platt, Kirstin Edwards, Jonathan Hubb, Joanna Ellis, Maria Zambon
EPI_ISL_424082  hCoV-19/England/20129096604/2020        Europe / United Kingdom / England       2020-03-11      Respiratory Virus Unit, Microbiology Services Colindale, Public Health England  Respiratory Virus Unit, Microbiology Services Colindale, Public Health England  Monica Galiano, Shahjahan Miah, Angie Lackenby, Omolola Akinbami, Tiina Talts, Leena Bhaw, Richard Myers, Steven Platt, Kirstin Edwards, Jonathan Hubb, Joanna Ellis, Maria Zambon
EPI_ISL_424084  hCoV-19/England/20129096804/2020        Europe / United Kingdom / England       2020-03-11      Respiratory Virus Unit, Microbiology Services Colindale, Public Health England  Respiratory Virus Unit, Microbiology Services Colindale, Public Health England  Monica Galiano, Shahjahan Miah, Angie Lackenby, Omolola Akinbami, Tiina Talts, Leena Bhaw, Richard Myers, Steven Platt, Kirstin Edwards, Jonathan Hubb, Joanna Ellis, Maria Zambon
EPI_ISL_424300  hCoV-19/USA/WA-UW-1879/2020     North America / USA / Washington        2020-03-21      UW Virology Lab    UW Virology Lab Pavitra Roychoudhury, Hong Xie, Keith Jerome, Alexander Greninger
EPI_ISL_424301  hCoV-19/USA/WA-UW-1948/2020     North America / USA / Washington        2020-03-24      UW Virology Lab    UW Virology Lab Pavitra Roychoudhury, Hong Xie, Keith Jerome, Alexander Greninger
EPI_ISL_424302  hCoV-19/USA/WA-UW-1878/2020     North America / USA / Washington        2020-03-24      UW Virology Lab    UW Virology Lab Pavitra Roychoudhury, Hong Xie, Keith Jerome, Alexander Greninger
EPI_ISL_424304  hCoV-19/USA/WA-UW-1930/2020     North America / USA / Washington        2020-03-24      UW Virology Lab    UW Virology Lab Pavitra Roychoudhury, Hong Xie, Keith Jerome, Alexander Greninger
EPI_ISL_424305  hCoV-19/USA/WA-UW-1877/2020     North America / USA / Washington        2020-03-24      UW Virology Lab    UW Virology Lab Pavitra Roychoudhury, Hong Xie, Keith Jerome, Alexander Greninger
EPI_ISL_424306  hCoV-19/USA/WA-UW-1874/2020     North America / USA / Washington        2020-03-23      UW Virology Lab    UW Virology Lab Pavitra Roychoudhury, Hong Xie, Keith Jerome, Alexander Greninger
EPI_ISL_424307  hCoV-19/USA/OR-UW-1885/2020     North America / USA / Oregon    2020-03-23      UW Virology Lab UW Virology Lab Pavitra Roychoudhury, Hong Xie, Keith Jerome, Alexander Greninger
EPI_ISL_424308  hCoV-19/USA/OR-UW-1887/2020     North America / USA / Oregon    2020-03-22      UW Virology Lab UW Virology Lab Pavitra Roychoudhury, Hong Xie, Keith Jerome, Alexander Greninger
EPI_ISL_424309  hCoV-19/USA/WA-UW-1881/2020     North America / USA / Washington        2020-03-24      UW Virology Lab    UW Virology Lab Pavitra Roychoudhury, Hong Xie, Keith Jerome, Alexander Greninger
EPI_ISL_424310  hCoV-19/USA/WA-UW-1888/2020     North America / USA / Washington        2020-03-23      UW Virology Lab    UW Virology Lab Pavitra Roychoudhury, Hong Xie, Keith Jerome, Alexander Greninger
EPI_ISL_424311  hCoV-19/USA/WA-UW-1880/2020     North America / USA / Washington        2020-03-24      UW Virology Lab    UW Virology Lab Pavitra Roychoudhury, Hong Xie, Keith Jerome, Alexander Greninger
EPI_ISL_424312  hCoV-19/USA/WA-UW-1955/2020     North America / USA / Washington        2020-03-24      UW Virology Lab    UW Virology Lab Pavitra Roychoudhury, Hong Xie, Keith Jerome, Alexander Greninger
EPI_ISL_424313  hCoV-19/USA/WA-UW-1956/2020     North America / USA / Washington        2020-03-24      UW Virology Lab    UW Virology Lab Pavitra Roychoudhury, Hong Xie, Keith Jerome, Alexander Greninger
EPI_ISL_424314  hCoV-19/USA/WA-UW-1921/2020     North America / USA / Washington        2020-03-24      UW Virology Lab    UW Virology Lab Pavitra Roychoudhury, Hong Xie, Keith Jerome, Alexander Greninger
EPI_ISL_424315  hCoV-19/USA/ID-UW-1957/2020     North America / USA / Idaho     2020-03-20      UW Virology Lab UW Virology Lab Pavitra Roychoudhury, Hong Xie, Keith Jerome, Alexander Greninger
EPI_ISL_424316  hCoV-19/USA/WA-UW-1890/2020     North America / USA / Washington        2020-03-24      UW Virology Lab    UW Virology Lab Pavitra Roychoudhury, Hong Xie, Keith Jerome, Alexander Greninger
EPI_ISL_424317  hCoV-19/USA/WA-UW-1928/2020     North America / USA / Washington        2020-03-24      UW Virology Lab    UW Virology Lab Pavitra Roychoudhury, Hong Xie, Keith Jerome, Alexander Greninger
EPI_ISL_424318  hCoV-19/USA/ID-UW-1938/2020     North America / USA / Idaho     2020-03-22      UW Virology Lab UW Virology Lab Pavitra Roychoudhury, Hong Xie, Keith Jerome, Alexander Greninger
EPI_ISL_424319  hCoV-19/USA/ID-UW-1935/2020     North America / USA / Idaho     2020-03-22      UW Virology Lab UW Virology Lab Pavitra Roychoudhury, Hong Xie, Keith Jerome, Alexander Greninger
EPI_ISL_424321  hCoV-19/USA/ID-UW-1925/2020     North America / USA / Idaho     2020-03-20      UW Virology Lab UW Virology Lab Pavitra Roychoudhury, Hong Xie, Keith Jerome, Alexander Greninger
EPI_ISL_424322  hCoV-19/USA/ID-UW-1934/2020     North America / USA / Idaho     2020-03-20      UW Virology Lab UW Virology Lab Pavitra Roychoudhury, Hong Xie, Keith Jerome, Alexander Greninger
EPI_ISL_424323  hCoV-19/USA/WA-UW-1944/2020     North America / USA / Washington        2020-03-24      UW Virology Lab    UW Virology Lab Pavitra Roychoudhury, Hong Xie, Keith Jerome, Alexander Greninger
EPI_ISL_424324  hCoV-19/USA/WA-UW-1965/2020     North America / USA / Washington        2020-03-24      UW Virology Lab    UW Virology Lab Pavitra Roychoudhury, Hong Xie, Keith Jerome, Alexander Greninger

EPI_ISL_424325 hCoV-19/USA/WA-UW-1954/2020 North America / USA / Washington 2020-03-24 UW Virology Lab UW Virology Lab Pavitra Roychoudhury, Hong Xie, Keith Jerome, Alexander Greninger
EPI_ISL_424326 hCoV-19/USA/WA-UW-1968/2020 North America / USA / Washington 2020-03-20 UW Virology Lab UW Virology Lab Pavitra Roychoudhury, Hong Xie, Keith Jerome, Alexander Greninger
EPI_ISL_424327 hCoV-19/USA/WA-UW-1947/2020 North America / USA / Washington 2020-03-23 UW Virology Lab UW Virology Lab Pavitra Roychoudhury, Hong Xie, Keith Jerome, Alexander Greninger
EPI_ISL_424329 hCoV-19/USA/WA-UW-1984/2020 North America / USA / Washington 2020-03-24 UW Virology Lab UW Virology Lab Pavitra Roychoudhury, Hong Xie, Keith Jerome, Alexander Greninger
EPI_ISL_424330 hCoV-19/USA/WA-UW-1865/2020 North America / USA / Washington 2020-03-23 UW Virology Lab UW Virology Lab Pavitra Roychoudhury, Hong Xie, Keith Jerome, Alexander Greninger
EPI_ISL_424331 hCoV-19/USA/WA-UW-1907/2020 North America / USA / Washington 2020-03-22 UW Virology Lab UW Virology Lab Pavitra Roychoudhury, Hong Xie, Keith Jerome, Alexander Greninger
EPI_ISL_424332 hCoV-19/USA/WA-UW-1917/2020 North America / USA / Washington 2020-03-22 UW Virology Lab UW Virology Lab Pavitra Roychoudhury, Hong Xie, Keith Jerome, Alexander Greninger
EPI_ISL_424333 hCoV-19/USA/WA-UW-2006/2020 North America / USA / Washington 2020-03-24 UW Virology Lab UW Virology Lab Pavitra Roychoudhury, Hong Xie, Keith Jerome, Alexander Greninger
EPI_ISL_424334 hCoV-19/USA/WA-UW-1994/2020 North America / USA / Washington 2020-03-24 UW Virology Lab UW Virology Lab Pavitra Roychoudhury, Hong Xie, Keith Jerome, Alexander Greninger
EPI_ISL_424335 hCoV-19/USA/WA-UW-1998/2020 North America / USA / Washington 2020-03-24 UW Virology Lab UW Virology Lab Pavitra Roychoudhury, Hong Xie, Keith Jerome, Alexander Greninger
EPI_ISL_424336 hCoV-19/USA/WA-UW-1988/2020 North America / USA / Washington 2020-03-24 UW Virology Lab UW Virology Lab Pavitra Roychoudhury, Hong Xie, Keith Jerome, Alexander Greninger
EPI_ISL_424337 hCoV-19/USA/WA-UW-1977/2020 North America / USA / Washington 2020-03-24 UW Virology Lab UW Virology Lab Pavitra Roychoudhury, Hong Xie, Keith Jerome, Alexander Greninger
EPI_ISL_424338 hCoV-19/USA/WA-UW-1986/2020 North America / USA / Washington 2020-03-24 UW Virology Lab UW Virology Lab Pavitra Roychoudhury, Hong Xie, Keith Jerome, Alexander Greninger
EPI_ISL_424339 hCoV-19/USA/WA-UW-2016/2020 North America / USA / Washington 2020-03-24 UW Virology Lab UW Virology Lab Pavitra Roychoudhury, Hong Xie, Keith Jerome, Alexander Greninger
EPI_ISL_424340 hCoV-19/USA/WA-UW-2009/2020 North America / USA / Washington 2020-03-24 UW Virology Lab UW Virology Lab Pavitra Roychoudhury, Hong Xie, Keith Jerome, Alexander Greninger
EPI_ISL_424341 hCoV-19/USA/WA-UW-2008/2020 North America / USA / Washington 2020-03-24 UW Virology Lab UW Virology Lab Pavitra Roychoudhury, Hong Xie, Keith Jerome, Alexander Greninger
EPI_ISL_424342 hCoV-19/Italy/INMI8/2020 Europe / Italy / Rome 2020-03-07 INMI Lazzaro Spallanzani IRCCS Laboratory of Virology, INMI Lazzaro Spallanzani IRCCS Concetta Castilletti, Barbara Bartolini, Martina Rueca, Cesare Ernesto Maria Gruber, Francesco Messina, Fabrizio Carletti, Eleonora Lalle, Licia Bordi, Giulia Matusali, Francesca Colavita, Maria Rosaria Capobianchi, Francesco Vairo, Giuseppe Ippolito, Antonino Di Caro
EPI_ISL_424343 hCoV-19/Italy/INMI9/2020 Europe / Italy / Rome 2020-03-23 INMI Lazzaro Spallanzani IRCCS Laboratory of Virology, INMI Lazzaro Spallanzani IRCCS Fabrizio Carletti, Barbara Bartolini, Martina Rueca, Cesare Ernesto Maria Gruber, Francesco Messina, Eleonora Lalle, Licia Bordi, Giulia Matusali, Francesca Colavita, Maria Rosaria Capobianchi, Concetta Castilletti, Francesco Vairo, Giuseppe Ippolito, Antonino Di Caro
EPI_ISL_424344 hCoV-19/Italy/INMI10/2020 Europe / Italy / Rome 2020-03-04 INMI Lazzaro Spallanzani IRCCS Laboratory of Virology, INMI Lazzaro Spallanzani IRCCS Eleonora Lalle, Barbara Bartolini, Martina Rueca, Cesare Ernesto Maria Gruber, Francesco Messina, Fabrizio Carletti, Licia Bordi, Giulia Matusali, Francesca Colavita, Maria Rosaria Capobianchi, Concetta Castilletti, Francesco Vairo, Giuseppe Ippolito, Antonino Di Caro
EPI_ISL_424345 hCoV-19/Mexico/CDMX-INER_01/2020 North America / Mexico / Mexico City 2020-03-12 Instituto Nacional de Enfermedades Respiratorias Instituto Nacional de Enfermedades Respiratorias Joel Armando Vázquez Pérez, Celia Boukadida, Santiago Avila Ríos, Mario Mújica Sánchez, José Arturo Martínez Orozco, Eduardo Becerril Vargas, Jorge Salas Hernández, Irma López Martínez, Lucia Hernández Rivas, Gisela Barrera Badillo, Edgar Mendieta Condado, Fabiola Garcés Ayala, Adnan Araiza Rodríguez, José Ernesto Ramírez González, Victor Hugo Borja Aburto, Concepción Grajales Muñiz, Cesar Raúl González Bonilla, Carolina González Torres, Francisco Javier Gaytán Cervantes, José Esteban Muñoz Medina, Guillermo M. Ruiz-Palacios, Pilar Ramos Cervantes, Violeta Ibarra Gonzalez, Fernando Ledesma Barrientos, Luis Alberto García Andrade, Alfredo Ponce de León Garduño, Blanca Taboada, Alejandro Sánchez, Pavel Isa, Ricardo Grande, Gloria Vázquez, Francisco Pulido, Carlos F. Arias.
EPI_ISL_424346 hCoV-19/USA/AK-PHL037/2020 North America / USA / Alaska / Southeast 2020-03-21 Alaska State Virology Laboratory Alaska State Virology Laboratory Chen, J.
EPI_ISL_424347 hCoV-19/USA/AK-PHL077/2020 North America / USA / Alaska / Interior 2020-03-29 Alaska State Virology Laboratory Alaska State Virology Laboratory Chen, J.
EPI_ISL_424352 hCoV-19/Fuyang/FY002/2020 Asia / China / Anhui / Fuyang 2020-03-10 unknown Clinical Laboratory Ge,B.
EPI_ISL_424353 hCoV-19/USA/NC_200173/2020 North America / USA / North Carolina 2020-03 Dirk Dittmer Dirk Dittmer Bailey,A.G., Caro-Vegas,C., Thompson,C., Dittmer,D., Eason,A.B., Juarez,A., Landis,J.T., McNamara,R.P., Miller,M.B., Moorad,R., Pluta,L.J., Seltzer,T.A., Villamor,F. and Vahrson,W.
EPI_ISL_424354 hCoV-19/USA/NC_200189/2020 North America / USA / North Carolina 2020-03 Dirk Dittmer Dirk Dittmer Bailey,A.G., Caro-Vegas,C.P., Dittmer,D., Eason,A.B., Juarez,A., Landis,J.T., McNamara,R.P., Miller,M.B., Moorad,R., Pluta,L.J., Seltzer,T.A., Thompson,C., Vahrson,W. and Villamor,F.
EPI_ISL_424355 hCoV-19/Beijing/Wuhan_IME-BJ01/2020 Asia / China / Beijing 2020-01-24 unknown Beijing

Institute of Microbiology and Epidemiology	Fan,H., Qin,E., Wu,Y., Guo,Y., Zhang,X., Yong,Y., Hou,J., Xu,Z., Mu,J., Teng,Y., Mi,Z., Yang,R., Song,Y., Li,B. and Cui,Y.
EPI_ISL_424356	hCoV-19/Beijing/Wuhan_IME-BJ02/2020	Asia / China / Beijing	2020-01-25	unknown	Beijing Institute of Microbiology and Epidemiology	Fan,H., Qin,E., Wu,Y., Guo,Y., Zhang,X., Yong,Y., Hou,J., Xu,Z., Mu,J., Teng,Y., Mi,Z., Yang,R., Song,Y., Li,B. and Cui,Y.
EPI_ISL_424357	hCoV-19/Beijing/Wuhan_IME-BJ03/2020	Asia / China / Beijing	2020-01-28	unknown	Beijing Institute of Microbiology and Epidemiology	Fan,H., Qin,E., Wu,Y., Guo,Y., Zhang,X., Yong,Y., Hou,J., Xu,Z., Mu,J., Teng,Y., Mi,Z., Yang,R., Song,Y., Li,B. and Cui,Y.
EPI_ISL_424358	hCoV-19/Beijing/Wuhan_IME-BJ04/2020	Asia / China / Beijing	2020-01-28	unknown	Beijing Institute of Microbiology and Epidemiology	Fan,H., Qin,E., Wu,Y., Guo,Y., Zhang,X., Yong,Y., Hou,J., Xu,Z., Mu,J., Teng,Y., Mi,Z., Yang,R., Song,Y., Li,B. and Cui,Y.
EPI_ISL_424359	hCoV-19/Beijing/Wuhan_IME-BJ05/2020	Asia / China / Beijing	2020-01-27	unknown	Beijing Institute of Microbiology and Epidemiology	Fan,H., Qin,E., Wu,Y., Guo,Y., Zhang,X., Yong,Y., Hou,J., Xu,Z., Mu,J., Teng,Y., Mi,Z., Yang,R., Song,Y., Li,B. and Cui,Y.
EPI_ISL_424360	hCoV-19/Beijing/Wuhan_IME-BJ07/2020	Asia / China / Beijing	2020-01-29	unknown	Beijing Institute of Microbiology and Epidemiology	Fan,H., Qin,E., Wu,Y., Guo,Y., Zhang,X., Yong,Y., Hou,J., Xu,Z., Mu,J., Teng,Y., Mi,Z., Yang,R., Song,Y., Li,B. and Cui,Y.
EPI_ISL_424361	hCoV-19/India/1063/2020	Asia / India	2020-03-10	National Influenza Center, Indian Council of Medical Research - National Institute of Virology	Indian Council of Medical Research-National Institute of Virology, Microbial Containment Complex	Pragya D. Yadav, Varsha Potdar, Savita Patil, Dimpal A. Nyayanit, Triparna Majumdar, Manohar. L. Chaudhary, Gururaj Deshpande, Padinjaremattathil Thankappan Ullas, Anita Shete-Aich, Hitesh Dighe, Sreelekshmy Mohandas, Gajanan Sapkal, Atanu Basu, Amita Jain, Bharti Malhotra, Deepika Chaudhary, Sarah Cherian, Priya Abraham
EPI_ISL_424362	hCoV-19/India/1135/2020	Asia / India	2020-03-10	National Influenza Center, Indian Council of Medical Research - National Institute of Virology	Indian Council of Medical Research-National Institute of Virology, Microbial Containment Complex	Pragya D. Yadav, Varsha Potdar, Savita Patil, Dimpal A. Nyayanit, Triparna Majumdar, Manohar. L. Chaudhary, Gururaj Deshpande, Padinjaremattathil Thankappan Ullas, Anita Shete-Aich, Hitesh Dighe, Sreelekshmy Mohandas, Gajanan Sapkal, Atanu Basu, Amita Jain, Bharti Malhotra, Deepika Chaudhary, Sarah Cherian, Priya Abraham
EPI_ISL_424363	hCoV-19/India/1652/2020	Asia / India	2020-03-12	National Influenza Center, Indian Council of Medical Research - National Institute of Virology	Indian Council of Medical Research-National Institute of Virology, Microbial Containment Complex	Pragya D. Yadav, Varsha Potdar, Savita Patil, Dimpal A. Nyayanit, Triparna Majumdar, Manohar. L. Chaudhary, Gururaj Deshpande, Padinjaremattathil Thankappan Ullas, Anita Shete-Aich, Hitesh Dighe, Sreelekshmy Mohandas, Gajanan Sapkal, Atanu Basu, Amita Jain, Bharti Malhotra, Deepika Chaudhary, Sarah Cherian, Priya Abraham
EPI_ISL_424364	hCoV-19/India/3118/2020	Asia / India	2020-03-17	National Influenza Center, Indian Council of Medical Research - National Institute of Virology	Indian Council of Medical Research-National Institute of Virology, Microbial Containment Complex	Pragya D. Yadav, Varsha Potdar, Savita Patil, Dimpal A. Nyayanit, Triparna Majumdar, Manohar. L. Chaudhary, Gururaj Deshpande, Padinjaremattathil Thankappan Ullas, Anita Shete-Aich, Hitesh Dighe, Sreelekshmy Mohandas, Gajanan Sapkal, Atanu Basu, Amita Jain, Bharti Malhotra, Deepika Chaudhary, Sarah Cherian, Priya Abraham
EPI_ISL_424365	hCoV-19/India/3239/2020	Asia / India	2020-03-17	National Influenza Center, Indian Council of Medical Research - National Institute of Virology	Indian Council of Medical Research-National Institute of Virology, Microbial Containment Complex	Pragya D. Yadav, Varsha Potdar, Savita Patil, Dimpal A. Nyayanit, Triparna Majumdar, Manohar. L. Chaudhary, Gururaj Deshpande, Padinjaremattathil Thankappan Ullas, Anita Shete-Aich, Hitesh Dighe, Sreelekshmy Mohandas, Gajanan Sapkal, Atanu Basu, Amita Jain, Bharti Malhotra, Deepika Chaudhary, Sarah Cherian, Priya Abraham
EPI_ISL_424366	hCoV-19/Turkey/ERAGEM-001/2020	Europe / Turkey / Kayseri	2020-03-17	Vaccine Research, Development and Application Center, Erciyes University	Gen Era Diagnostics Inc.	Shaikh Terkis Islam Pavel, Hazel Yetiskin, Gunsu Aydin, Can Holyavkin, Muhammet Ali Uygut, Zehra B Dursun, İlhami Celik, Alper Iseri, Aykut Ozdarendeli
EPI_ISL_424367	hCoV-19/Iceland/343/2020	Europe / Iceland / Reykjavik	2020-03	The National University Hospital of Iceland	deCODE genetics	Daniel F Gudbjartsson; Agnar Helgason; Hakon Jonsson; Olafur T Magnusson; Pall Melsted; Gudmundur L Norddahl; Jona Saemundsdottir; Asgeir Sigurdsson; Patrick Sulem; Arna B Agustsdottir; Berglind Eiriksdottir; Run Fridriksdottir; Elisabet E Gardarsdottir; Gudmundur Georgsson; Olafia S Gretarsdottir; Kjartan R Gudmundsson; Thora R Gunnarsdottir; Arnaldur Gylfason; Hilma Holm; Brynjar O Jensson; Aslaug Jonasdottir; Kamilla S Josefsdottir; Thordur Kristjansson; Droplaug N Magnusdottir; Louise le Roux; Gudrun Sigmundsdottir; Gardar Sveinbjornsson; Kristin E Sveinsdottir; Maney Sveinsdottir; Emil A Thorarensen; Bjarni Thorbjornsson; Gisli Masson; Ingileif Jonsdottir; Alma Moller; Thorolfur Gudnason; Karl G Kristinsson; Unnur Thorsteinsdottir; Kari Stefansson
EPI_ISL_424368	hCoV-19/Iceland/344/2020	Europe / Iceland / Reykjavik	2020-03	The National University Hospital of Iceland	deCODE genetics	Daniel F Gudbjartsson; Agnar Helgason; Hakon Jonsson; Olafur T Magnusson; Pall Melsted; Gudmundur L Norddahl; Jona Saemundsdottir; Asgeir Sigurdsson; Patrick Sulem; Arna B Agustsdottir; Berglind Eiriksdottir; Run Fridriksdottir; Elisabet E Gardarsdottir; Gudmundur Georgsson; Olafia S Gretarsdottir; Kjartan R Gudmundsson; Thora R Gunnarsdottir; Arnaldur Gylfason; Hilma Holm; Brynjar O Jensson; Aslaug Jonasdottir; Kamilla S Josefsdottir; Thordur Kristjansson; Droplaug N Magnusdottir; Louise le Roux; Gudrun Sigmundsdottir; Gardar Sveinbjornsson; Kristin E Sveinsdottir; Maney Sveinsdottir; Emil A Thorarensen; Bjarni Thorbjornsson; Gisli Masson; Ingileif Jonsdottir; Alma Moller; Thorolfur Gudnason; Karl G Kristinsson; Unnur Thorsteinsdottir; Kari Stefansson
EPI_ISL_424369	hCoV-19/Iceland/345/2020	Europe / Iceland / Reykjavik	2020-03	The National University

Hospital of Iceland	deCODE genetics Daniel F Gudbjartsson; Agnar Helgason; Hakon Jonsson; Olafur T Magnusson; Pall Melsted; Gudmundur L Norddahl; Jona Saemundsdottir; Asgeir Sigurdsson; Patrick Sulem; Arna B Agustsdottir; Berglind Eiriksdottir; Run Fridriksdottir; Elisabet E Gardarsdottir; Gudmundur Georgsson; Olafia S Gretarsdottir; Kjartan R Gudmundsson; Thora R Gunnarsdottir; Arnaldur Gylfason; Hilma Holm; Brynjar O Jensson; Aslaug Jonasdottir; Kamilla S Josefsdottir; Thordur Kristjansson; Droplaug N Magnusdottir; Louise le Roux; Gudrun Sigmundsdottir; Gardar Sveinbjornsson; Kristin E Sveinsdottir; Maney Sveinsdottir; Emil A Thorarensen; Bjarni Thorbjornsson; Gisli Masson; Ingileif Jonsdottir; Alma Moller; Thorolfur Gudnason; Karl G Kristinsson; Unnur Thorsteinsdottir; Kari Stefansson
EPI_ISL_424370	hCoV-19/Iceland/346/2020	Europe / Iceland / Reykjavik	2020-03 The National University Hospital of Iceland	deCODE genetics Daniel F Gudbjartsson; Agnar Helgason; Hakon Jonsson; Olafur T Magnusson; Pall Melsted; Gudmundur L Norddahl; Jona Saemundsdottir; Asgeir Sigurdsson; Patrick Sulem; Arna B Agustsdottir; Berglind Eiriksdottir; Run Fridriksdottir; Elisabet E Gardarsdottir; Gudmundur Georgsson; Olafia S Gretarsdottir; Kjartan R Gudmundsson; Thora R Gunnarsdottir; Arnaldur Gylfason; Hilma Holm; Brynjar O Jensson; Aslaug Jonasdottir; Kamilla S Josefsdottir; Thordur Kristjansson; Droplaug N Magnusdottir; Louise le Roux; Gudrun Sigmundsdottir; Gardar Sveinbjornsson; Kristin E Sveinsdottir; Maney Sveinsdottir; Emil A Thorarensen; Bjarni Thorbjornsson; Gisli Masson; Ingileif Jonsdottir; Alma Moller; Thorolfur Gudnason; Karl G Kristinsson; Unnur Thorsteinsdottir; Kari Stefansson
EPI_ISL_424371	hCoV-19/Iceland/347/2020	Europe / Iceland / Reykjavik	2020-03 The National University Hospital of Iceland	deCODE genetics Daniel F Gudbjartsson; Agnar Helgason; Hakon Jonsson; Olafur T Magnusson; Pall Melsted; Gudmundur L Norddahl; Jona Saemundsdottir; Asgeir Sigurdsson; Patrick Sulem; Arna B Agustsdottir; Berglind Eiriksdottir; Run Fridriksdottir; Elisabet E Gardarsdottir; Gudmundur Georgsson; Olafia S Gretarsdottir; Kjartan R Gudmundsson; Thora R Gunnarsdottir; Arnaldur Gylfason; Hilma Holm; Brynjar O Jensson; Aslaug Jonasdottir; Kamilla S Josefsdottir; Thordur Kristjansson; Droplaug N Magnusdottir; Louise le Roux; Gudrun Sigmundsdottir; Gardar Sveinbjornsson; Kristin E Sveinsdottir; Maney Sveinsdottir; Emil A Thorarensen; Bjarni Thorbjornsson; Gisli Masson; Ingileif Jonsdottir; Alma Moller; Thorolfur Gudnason; Karl G Kristinsson; Unnur Thorsteinsdottir; Kari Stefansson
EPI_ISL_424377	hCoV-19/Iceland/353/2020	Europe / Iceland / Reykjavik	2020-03-18	deCODE genetics deCODE genetics Daniel F Gudbjartsson; Agnar Helgason; Hakon Jonsson; Olafur T Magnusson; Pall Melsted; Gudmundur L Norddahl; Jona Saemundsdottir; Asgeir Sigurdsson; Patrick Sulem; Arna B Agustsdottir; Berglind Eiriksdottir; Run Fridriksdottir; Elisabet E Gardarsdottir; Gudmundur Georgsson; Olafia S Gretarsdottir; Kjartan R Gudmundsson; Thora R Gunnarsdottir; Arnaldur Gylfason; Hilma Holm; Brynjar O Jensson; Aslaug Jonasdottir; Kamilla S Josefsdottir; Thordur Kristjansson; Droplaug N Magnusdottir; Louise le Roux; Gudrun Sigmundsdottir; Gardar Sveinbjornsson; Kristin E Sveinsdottir; Maney Sveinsdottir; Emil A Thorarensen; Bjarni Thorbjornsson; Gisli Masson; Ingileif Jonsdottir; Alma Moller; Thorolfur Gudnason; Karl G Kristinsson; Unnur Thorsteinsdottir; Kari Stefansson
EPI_ISL_424379	hCoV-19/Iceland/355/2020	Europe / Iceland / Reykjavik	2020-03-19	The National University Hospital of Iceland	deCODE genetics Daniel F Gudbjartsson; Agnar Helgason; Hakon Jonsson; Olafur T Magnusson; Pall Melsted; Gudmundur L Norddahl; Jona Saemundsdottir; Asgeir Sigurdsson; Patrick Sulem; Arna B Agustsdottir; Berglind Eiriksdottir; Run Fridriksdottir; Elisabet E Gardarsdottir; Gudmundur Georgsson; Olafia S Gretarsdottir; Kjartan R Gudmundsson; Thora R Gunnarsdottir; Arnaldur Gylfason; Hilma Holm; Brynjar O Jensson; Aslaug Jonasdottir; Kamilla S Josefsdottir; Thordur Kristjansson; Droplaug N Magnusdottir; Louise le Roux; Gudrun Sigmundsdottir; Gardar Sveinbjornsson; Kristin E Sveinsdottir; Maney Sveinsdottir; Emil A Thorarensen; Bjarni Thorbjornsson; Gisli Masson; Ingileif Jonsdottir; Alma Moller; Thorolfur Gudnason; Karl G Kristinsson; Unnur Thorsteinsdottir; Kari Stefansson
EPI_ISL_424380	hCoV-19/Iceland/356/2020	Europe / Iceland / Reykjavik	2020-03-19	The National University Hospital of Iceland	deCODE genetics Daniel F Gudbjartsson; Agnar Helgason; Hakon Jonsson; Olafur T Magnusson; Pall Melsted; Gudmundur L Norddahl; Jona Saemundsdottir; Asgeir Sigurdsson; Patrick Sulem; Arna B Agustsdottir; Berglind Eiriksdottir; Run Fridriksdottir; Elisabet E Gardarsdottir; Gudmundur Georgsson; Olafia S Gretarsdottir; Kjartan R Gudmundsson; Thora R Gunnarsdottir; Arnaldur Gylfason; Hilma Holm; Brynjar O Jensson; Aslaug Jonasdottir; Kamilla S Josefsdottir; Thordur Kristjansson; Droplaug N Magnusdottir; Louise le Roux; Gudrun Sigmundsdottir; Gardar Sveinbjornsson; Kristin E Sveinsdottir; Maney Sveinsdottir; Emil A Thorarensen; Bjarni Thorbjornsson; Gisli Masson; Ingileif Jonsdottir; Alma Moller; Thorolfur Gudnason; Karl G Kristinsson; Unnur Thorsteinsdottir; Kari Stefansson
EPI_ISL_424381	hCoV-19/Iceland/357/2020	Europe / Iceland / Reykjavik	2020-03-19	The National University Hospital of Iceland	deCODE genetics Daniel F Gudbjartsson; Agnar Helgason; Hakon Jonsson; Olafur T Magnusson; Pall Melsted; Gudmundur L Norddahl; Jona Saemundsdottir; Asgeir Sigurdsson; Patrick Sulem; Arna B Agustsdottir; Berglind Eiriksdottir; Run Fridriksdottir; Elisabet E Gardarsdottir; Gudmundur Georgsson; Olafia S Gretarsdottir; Kjartan R Gudmundsson; Thora R Gunnarsdottir; Arnaldur Gylfason; Hilma Holm; Brynjar O Jensson; Aslaug Jonasdottir; Kamilla S Josefsdottir; Thordur Kristjansson; Droplaug N Magnusdottir; Louise le Roux; Gudrun Sigmundsdottir; Gardar Sveinbjornsson; Kristin E Sveinsdottir; Maney Sveinsdottir; Emil A Thorarensen; Bjarni Thorbjornsson; Gisli Masson; Ingileif Jonsdottir; Alma Moller; Thorolfur Gudnason; Karl G Kristinsson; Unnur Thorsteinsdottir; Kari Stefansson
EPI_ISL_424382	hCoV-19/Iceland/358/2020	Europe / Iceland / Reykjavik	2020-03-19	The National University Hospital of Iceland	deCODE genetics Daniel F Gudbjartsson; Agnar Helgason; Hakon Jonsson; Olafur T Magnusson; Pall Melsted; Gudmundur L Norddahl; Jona Saemundsdottir; Asgeir Sigurdsson; Patrick Sulem; Arna B Agustsdottir; Berglind Eiriksdottir; Run Fridriksdottir; Elisabet E Gardarsdottir; Gudmundur Georgsson; Olafia S Gretarsdottir; Kjartan R Gudmundsson; Thora R Gunnarsdottir; Arnaldur Gylfason; Hilma Holm; Brynjar O Jensson; Aslaug Jonasdottir; Kamilla S Josefsdottir; Thordur Kristjansson; Droplaug N Magnusdottir; Louise le Roux; Gudrun Sigmundsdottir; Gardar Sveinbjornsson; Kristin E Sveinsdottir; Maney Sveinsdottir; Emil A Thorarensen; Bjarni Thorbjornsson; Gisli Masson; Ingileif Jonsdottir; Alma Moller; Thorolfur Gudnason; Karl G

Kristinsson; Unnur Thorsteinsdottir; Kari Stefansson
EPI_ISL_424383 hCoV-19/Iceland/359/2020 Europe / Iceland / Reykjavik 2020-03-19 The National University Hospital of Iceland deCODE genetics Daniel F Gudbjartsson; Agnar Helgason; Hakon Jonsson; Olafur T Magnusson; Pall Melsted; Gudmundur L Norddahl; Jona Saemundsdottir; Asgeir Sigurdsson; Patrick Sulem; Arna B Agustsdottir; Berglind Eiriksdottir; Run Fridriksdottir; Elisabet E Gardarsdottir; Gudmundur Georgsson; Olafia S Gretarsdottir; Kjartan R Gudmundsson; Thora R Gunnarsdottir; Arnaldur Gylfason; Hilma Holm; Brynjar O Jensson; Aslaug Jonasdottir; Kamilla S Josefsdottir; Thordur Kristjansson; Droplaug N Magnusdottir; Louise le Roux; Gudrun Sigmundsdottir; Gardar Sveinbjornsson; Kristin E Sveinsdottir; Maney Sveinsdottir; Emil A Thorarensen; Bjarni Thorbjornsson; Gisli Masson; Ingileif Jonsdottir; Alma Moller; Thorolfur Gudnason; Karl G Kristinsson; Unnur Thorsteinsdottir; Kari Stefansson
EPI_ISL_424384 hCoV-19/Iceland/360/2020 Europe / Iceland / Reykjavik 2020-03-19 The National University Hospital of Iceland deCODE genetics Daniel F Gudbjartsson; Agnar Helgason; Hakon Jonsson; Olafur T Magnusson; Pall Melsted; Gudmundur L Norddahl; Jona Saemundsdottir; Asgeir Sigurdsson; Patrick Sulem; Arna B Agustsdottir; Berglind Eiriksdottir; Run Fridriksdottir; Elisabet E Gardarsdottir; Gudmundur Georgsson; Olafia S Gretarsdottir; Kjartan R Gudmundsson; Thora R Gunnarsdottir; Arnaldur Gylfason; Hilma Holm; Brynjar O Jensson; Aslaug Jonasdottir; Kamilla S Josefsdottir; Thordur Kristjansson; Droplaug N Magnusdottir; Louise le Roux; Gudrun Sigmundsdottir; Gardar Sveinbjornsson; Kristin E Sveinsdottir; Maney Sveinsdottir; Emil A Thorarensen; Bjarni Thorbjornsson; Gisli Masson; Ingileif Jonsdottir; Alma Moller; Thorolfur Gudnason; Karl G Kristinsson; Unnur Thorsteinsdottir; Kari Stefansson
EPI_ISL_424385 hCoV-19/Iceland/361/2020 Europe / Iceland / Reykjavik 2020-03-19 The National University Hospital of Iceland deCODE genetics Daniel F Gudbjartsson; Agnar Helgason; Hakon Jonsson; Olafur T Magnusson; Pall Melsted; Gudmundur L Norddahl; Jona Saemundsdottir; Asgeir Sigurdsson; Patrick Sulem; Arna B Agustsdottir; Berglind Eiriksdottir; Run Fridriksdottir; Elisabet E Gardarsdottir; Gudmundur Georgsson; Olafia S Gretarsdottir; Kjartan R Gudmundsson; Thora R Gunnarsdottir; Arnaldur Gylfason; Hilma Holm; Brynjar O Jensson; Aslaug Jonasdottir; Kamilla S Josefsdottir; Thordur Kristjansson; Droplaug N Magnusdottir; Louise le Roux; Gudrun Sigmundsdottir; Gardar Sveinbjornsson; Kristin E Sveinsdottir; Maney Sveinsdottir; Emil A Thorarensen; Bjarni Thorbjornsson; Gisli Masson; Ingileif Jonsdottir; Alma Moller; Thorolfur Gudnason; Karl G Kristinsson; Unnur Thorsteinsdottir; Kari Stefansson
EPI_ISL_424388 hCoV-19/Iceland/364/2020 Europe / Iceland / Reykjavik 2020-03-19 The National University Hospital of Iceland deCODE genetics Daniel F Gudbjartsson; Agnar Helgason; Hakon Jonsson; Olafur T Magnusson; Pall Melsted; Gudmundur L Norddahl; Jona Saemundsdottir; Asgeir Sigurdsson; Patrick Sulem; Arna B Agustsdottir; Berglind Eiriksdottir; Run Fridriksdottir; Elisabet E Gardarsdottir; Gudmundur Georgsson; Olafia S Gretarsdottir; Kjartan R Gudmundsson; Thora R Gunnarsdottir; Arnaldur Gylfason; Hilma Holm; Brynjar O Jensson; Aslaug Jonasdottir; Kamilla S Josefsdottir; Thordur Kristjansson; Droplaug N Magnusdottir; Louise le Roux; Gudrun Sigmundsdottir; Gardar Sveinbjornsson; Kristin E Sveinsdottir; Maney Sveinsdottir; Emil A Thorarensen; Bjarni Thorbjornsson; Gisli Masson; Ingileif Jonsdottir; Alma Moller; Thorolfur Gudnason; Karl G Kristinsson; Unnur Thorsteinsdottir; Kari Stefansson
EPI_ISL_424389 hCoV-19/Iceland/365/2020 Europe / Iceland / Reykjavik 2020-03-19 The National University Hospital of Iceland deCODE genetics Daniel F Gudbjartsson; Agnar Helgason; Hakon Jonsson; Olafur T Magnusson; Pall Melsted; Gudmundur L Norddahl; Jona Saemundsdottir; Asgeir Sigurdsson; Patrick Sulem; Arna B Agustsdottir; Berglind Eiriksdottir; Run Fridriksdottir; Elisabet E Gardarsdottir; Gudmundur Georgsson; Olafia S Gretarsdottir; Kjartan R Gudmundsson; Thora R Gunnarsdottir; Arnaldur Gylfason; Hilma Holm; Brynjar O Jensson; Aslaug Jonasdottir; Kamilla S Josefsdottir; Thordur Kristjansson; Droplaug N Magnusdottir; Louise le Roux; Gudrun Sigmundsdottir; Gardar Sveinbjornsson; Kristin E Sveinsdottir; Maney Sveinsdottir; Emil A Thorarensen; Bjarni Thorbjornsson; Gisli Masson; Ingileif Jonsdottir; Alma Moller; Thorolfur Gudnason; Karl G Kristinsson; Unnur Thorsteinsdottir; Kari Stefansson
EPI_ISL_424390 hCoV-19/Iceland/366/2020 Europe / Iceland / Reykjavik 2020-03-19 The National University Hospital of Iceland deCODE genetics Daniel F Gudbjartsson; Agnar Helgason; Hakon Jonsson; Olafur T Magnusson; Pall Melsted; Gudmundur L Norddahl; Jona Saemundsdottir; Asgeir Sigurdsson; Patrick Sulem; Arna B Agustsdottir; Berglind Eiriksdottir; Run Fridriksdottir; Elisabet E Gardarsdottir; Gudmundur Georgsson; Olafia S Gretarsdottir; Kjartan R Gudmundsson; Thora R Gunnarsdottir; Arnaldur Gylfason; Hilma Holm; Brynjar O Jensson; Aslaug Jonasdottir; Kamilla S Josefsdottir; Thordur Kristjansson; Droplaug N Magnusdottir; Louise le Roux; Gudrun Sigmundsdottir; Gardar Sveinbjornsson; Kristin E Sveinsdottir; Maney Sveinsdottir; Emil A Thorarensen; Bjarni Thorbjornsson; Gisli Masson; Ingileif Jonsdottir; Alma Moller; Thorolfur Gudnason; Karl G Kristinsson; Unnur Thorsteinsdottir; Kari Stefansson
EPI_ISL_424391 hCoV-19/Iceland/367/2020 Europe / Iceland / Reykjavik 2020-03-19 The National University Hospital of Iceland deCODE genetics Daniel F Gudbjartsson; Agnar Helgason; Hakon Jonsson; Olafur T Magnusson; Pall Melsted; Gudmundur L Norddahl; Jona Saemundsdottir; Asgeir Sigurdsson; Patrick Sulem; Arna B Agustsdottir; Berglind Eiriksdottir; Run Fridriksdottir; Elisabet E Gardarsdottir; Gudmundur Georgsson; Olafia S Gretarsdottir; Kjartan R Gudmundsson; Thora R Gunnarsdottir; Arnaldur Gylfason; Hilma Holm; Brynjar O Jensson; Aslaug Jonasdottir; Kamilla S Josefsdottir; Thordur Kristjansson; Droplaug N Magnusdottir; Louise le Roux; Gudrun Sigmundsdottir; Gardar Sveinbjornsson; Kristin E Sveinsdottir; Maney Sveinsdottir; Emil A Thorarensen; Bjarni Thorbjornsson; Gisli Masson; Ingileif Jonsdottir; Alma Moller; Thorolfur Gudnason; Karl G Kristinsson; Unnur Thorsteinsdottir; Kari Stefansson
EPI_ISL_424392 hCoV-19/Iceland/368/2020 Europe / Iceland / Reykjavik 2020-03-19 The National University Hospital of Iceland deCODE genetics Daniel F Gudbjartsson; Agnar Helgason; Hakon Jonsson; Olafur T Magnusson; Pall Melsted; Gudmundur L Norddahl; Jona Saemundsdottir; Asgeir Sigurdsson; Patrick Sulem; Arna B Agustsdottir; Berglind Eiriksdottir; Run Fridriksdottir; Elisabet E Gardarsdottir; Gudmundur Georgsson; Olafia S Gretarsdottir; Kjartan R Gudmundsson; Thora R Gunnarsdottir; Arnaldur Gylfason; Hilma Holm; Brynjar O Jensson; Aslaug Jonasdottir; Kamilla S Josefsdottir; Thordur Kristjansson; Droplaug N Magnusdottir; Louise le

Roux; Gudrun Sigmundsdottir; Gardar Sveinbjornsson; Kristin E Sveinsdottir; Maney Sveinsdottir; Emil A Thorarensen; Bjarni Thorbjornsson; Gisli Masson; Ingileif Jonsdottir; Alma Moller; Thorolfur Gudnason; Karl G Kristinsson; Unnur Thorsteinsdottir; Kari Stefansson
EPI_ISL_424393 hCoV-19/Iceland/369/2020 Europe / Iceland / Reykjavik 2020-03-19 The National University Hospital of Iceland deCODE genetics Daniel F Gudbjartsson; Agnar Helgason; Hakon Jonsson; Olafur T Magnusson; Pall Melsted; Gudmundur L Norddahl; Jona Saemundsdottir; Asgeir Sigurdsson; Patrick Sulem; Arna B Agustsdottir; Berglind Eiriksdottir; Run Fridriksdottir; Elisabet E Gardarsdottir; Gudmundur Georgsson; Olafia S Gretarsdottir; Kjartan R Gudmundsson; Thora R Gunnarsdottir; Arnaldur Gylfason; Hilma Holm; Brynjar O Jensson; Aslaug Jonasdottir; Kamilla S Josefsdottir; Thordur Kristjansson; Droplaug N Magnusdottir; Louise le Roux; Gudrun Sigmundsdottir; Gardar Sveinbjornsson; Kristin E Sveinsdottir; Maney Sveinsdottir; Emil A Thorarensen; Bjarni Thorbjornsson; Gisli Masson; Ingileif Jonsdottir; Alma Moller; Thorolfur Gudnason; Karl G Kristinsson; Unnur Thorsteinsdottir; Kari Stefansson
EPI_ISL_424394 hCoV-19/Iceland/370/2020 Europe / Iceland / Reykjavik 2020-03-19 The National University Hospital of Iceland deCODE genetics Daniel F Gudbjartsson; Agnar Helgason; Hakon Jonsson; Olafur T Magnusson; Pall Melsted; Gudmundur L Norddahl; Jona Saemundsdottir; Asgeir Sigurdsson; Patrick Sulem; Arna B Agustsdottir; Berglind Eiriksdottir; Run Fridriksdottir; Elisabet E Gardarsdottir; Gudmundur Georgsson; Olafia S Gretarsdottir; Kjartan R Gudmundsson; Thora R Gunnarsdottir; Arnaldur Gylfason; Hilma Holm; Brynjar O Jensson; Aslaug Jonasdottir; Kamilla S Josefsdottir; Thordur Kristjansson; Droplaug N Magnusdottir; Louise le Roux; Gudrun Sigmundsdottir; Gardar Sveinbjornsson; Kristin E Sveinsdottir; Maney Sveinsdottir; Emil A Thorarensen; Bjarni Thorbjornsson; Gisli Masson; Ingileif Jonsdottir; Alma Moller; Thorolfur Gudnason; Karl G Kristinsson; Unnur Thorsteinsdottir; Kari Stefansson
EPI_ISL_424396 hCoV-19/Iceland/372/2020 Europe / Iceland / Reykjavik 2020-03-19 The National University Hospital of Iceland deCODE genetics Daniel F Gudbjartsson; Agnar Helgason; Hakon Jonsson; Olafur T Magnusson; Pall Melsted; Gudmundur L Norddahl; Jona Saemundsdottir; Asgeir Sigurdsson; Patrick Sulem; Arna B Agustsdottir; Berglind Eiriksdottir; Run Fridriksdottir; Elisabet E Gardarsdottir; Gudmundur Georgsson; Olafia S Gretarsdottir; Kjartan R Gudmundsson; Thora R Gunnarsdottir; Arnaldur Gylfason; Hilma Holm; Brynjar O Jensson; Aslaug Jonasdottir; Kamilla S Josefsdottir; Thordur Kristjansson; Droplaug N Magnusdottir; Louise le Roux; Gudrun Sigmundsdottir; Gardar Sveinbjornsson; Kristin E Sveinsdottir; Maney Sveinsdottir; Emil A Thorarensen; Bjarni Thorbjornsson; Gisli Masson; Ingileif Jonsdottir; Alma Moller; Thorolfur Gudnason; Karl G Kristinsson; Unnur Thorsteinsdottir; Kari Stefansson
EPI_ISL_424398 hCoV-19/Iceland/374/2020 Europe / Iceland / Reykjavik 2020-03-19 The National University Hospital of Iceland deCODE genetics Daniel F Gudbjartsson; Agnar Helgason; Hakon Jonsson; Olafur T Magnusson; Pall Melsted; Gudmundur L Norddahl; Jona Saemundsdottir; Asgeir Sigurdsson; Patrick Sulem; Arna B Agustsdottir; Berglind Eiriksdottir; Run Fridriksdottir; Elisabet E Gardarsdottir; Gudmundur Georgsson; Olafia S Gretarsdottir; Kjartan R Gudmundsson; Thora R Gunnarsdottir; Arnaldur Gylfason; Hilma Holm; Brynjar O Jensson; Aslaug Jonasdottir; Kamilla S Josefsdottir; Thordur Kristjansson; Droplaug N Magnusdottir; Louise le Roux; Gudrun Sigmundsdottir; Gardar Sveinbjornsson; Kristin E Sveinsdottir; Maney Sveinsdottir; Emil A Thorarensen; Bjarni Thorbjornsson; Gisli Masson; Ingileif Jonsdottir; Alma Moller; Thorolfur Gudnason; Karl G Kristinsson; Unnur Thorsteinsdottir; Kari Stefansson
EPI_ISL_424399 hCoV-19/Iceland/375/2020 Europe / Iceland / Reykjavik 2020-03-19 The National University Hospital of Iceland deCODE genetics Daniel F Gudbjartsson; Agnar Helgason; Hakon Jonsson; Olafur T Magnusson; Pall Melsted; Gudmundur L Norddahl; Jona Saemundsdottir; Asgeir Sigurdsson; Patrick Sulem; Arna B Agustsdottir; Berglind Eiriksdottir; Run Fridriksdottir; Elisabet E Gardarsdottir; Gudmundur Georgsson; Olafia S Gretarsdottir; Kjartan R Gudmundsson; Thora R Gunnarsdottir; Arnaldur Gylfason; Hilma Holm; Brynjar O Jensson; Aslaug Jonasdottir; Kamilla S Josefsdottir; Thordur Kristjansson; Droplaug N Magnusdottir; Louise le Roux; Gudrun Sigmundsdottir; Gardar Sveinbjornsson; Kristin E Sveinsdottir; Maney Sveinsdottir; Emil A Thorarensen; Bjarni Thorbjornsson; Gisli Masson; Ingileif Jonsdottir; Alma Moller; Thorolfur Gudnason; Karl G Kristinsson; Unnur Thorsteinsdottir; Kari Stefansson
EPI_ISL_425308 hCoV-19/England/CAMB-74580/2020 Europe / United Kingdom / England 2020-03-28 Department of Pathology, University of Cambridge COVID-19 Genomics UK (COG-UK) Consortium Luke W Meredith, M. Estee Torok , Myra Hosmillo, William L. Hamilton, Martin D. Curran, Theresa Feltwell, Anna Yakovleva, Charlotte J. Houldcroft, Aminu S. Jahun, Sarah L. Caddy, Ian Goodfellow
EPI_ISL_425309 hCoV-19/England/CAMB-7459F/2020 Europe / United Kingdom / England 2020-03-30 Department of Pathology, University of Cambridge COVID-19 Genomics UK (COG-UK) Consortium Luke W Meredith, M. Estee Torok , Myra Hosmillo, William L. Hamilton, Martin D. Curran, Theresa Feltwell, Anna Yakovleva, Charlotte J. Houldcroft, Aminu S. Jahun, Sarah L. Caddy, Ian Goodfellow
EPI_ISL_425310 hCoV-19/England/CAMB-745AE/2020 Europe / United Kingdom / England 2020-03-27 Department of Pathology, University of Cambridge COVID-19 Genomics UK (COG-UK) Consortium Luke W Meredith, M. Estee Torok , Myra Hosmillo, William L. Hamilton, Martin D. Curran, Theresa Feltwell, Anna Yakovleva, Charlotte J. Houldcroft, Aminu S. Jahun, Sarah L. Caddy, Ian Goodfellow
EPI_ISL_425311 hCoV-19/England/CAMB-745BD/2020 Europe / United Kingdom / England 2020-03-27 Department of Pathology, University of Cambridge COVID-19 Genomics UK (COG-UK) Consortium Luke W Meredith, M. Estee Torok , Myra Hosmillo, William L. Hamilton, Martin D. Curran, Theresa Feltwell, Anna Yakovleva, Charlotte J. Houldcroft, Aminu S. Jahun, Sarah L. Caddy, Ian Goodfellow
EPI_ISL_425312 hCoV-19/England/CAMB-745CC/2020 Europe / United Kingdom / England 2020-03-29 Department of Pathology, University of Cambridge COVID-19 Genomics UK (COG-UK) Consortium Luke W Meredith, M. Estee Torok , Myra Hosmillo, William L. Hamilton, Martin D. Curran, Theresa Feltwell, Anna Yakovleva, Charlotte J. Houldcroft, Aminu S. Jahun, Sarah L. Caddy, Ian Goodfellow
EPI_ISL_425313 hCoV-19/England/CAMB-745DB/2020 Europe / United Kingdom / England 2020-03-28 Department of Pathology, University of Cambridge COVID-19 Genomics UK (COG-UK) Consortium Luke W

Meredith, M. Estee Torok , Myra Hosmillo, William L. Hamilton, Martin D. Curran, Theresa Feltwell, Anna Yakovleva, Charlotte J. Houldcroft, Aminu S. Jahun, Sarah L. Caddy, Ian Goodfellow
EPI_ISL_425314 hCoV-19/England/CAMB-745EA/2020 Europe / United Kingdom / England 2020-03-26 Department of Pathology, University of Cambridge COVID-19 Genomics UK (COG-UK) Consortium Luke W Meredith, M. Estee Torok , Myra Hosmillo, William L. Hamilton, Martin D. Curran, Theresa Feltwell, Anna Yakovleva, Charlotte J. Houldcroft, Aminu S. Jahun, Sarah L. Caddy, Ian Goodfellow
EPI_ISL_425315 hCoV-19/England/CAMB-74605/2020 Europe / United Kingdom / England 2020-03-24 Department of Pathology, University of Cambridge COVID-19 Genomics UK (COG-UK) Consortium Luke W Meredith, M. Estee Torok , Myra Hosmillo, William L. Hamilton, Martin D. Curran, Theresa Feltwell, Anna Yakovleva, Charlotte J. Houldcroft, Aminu S. Jahun, Sarah L. Caddy, Ian Goodfellow
EPI_ISL_425316 hCoV-19/England/CAMB-74614/2020 Europe / United Kingdom / England 2020-03-22 Department of Pathology, University of Cambridge COVID-19 Genomics UK (COG-UK) Consortium Luke W Meredith, M. Estee Torok , Myra Hosmillo, William L. Hamilton, Martin D. Curran, Theresa Feltwell, Anna Yakovleva, Charlotte J. Houldcroft, Aminu S. Jahun, Sarah L. Caddy, Ian Goodfellow
EPI_ISL_425317 hCoV-19/England/CAMB-74623/2020 Europe / United Kingdom / England 2020-03-22 Department of Pathology, University of Cambridge COVID-19 Genomics UK (COG-UK) Consortium Luke W Meredith, M. Estee Torok , Myra Hosmillo, William L. Hamilton, Martin D. Curran, Theresa Feltwell, Anna Yakovleva, Charlotte J. Houldcroft, Aminu S. Jahun, Sarah L. Caddy, Ian Goodfellow
EPI_ISL_425318 hCoV-19/England/CAMB-74641/2020 Europe / United Kingdom / England 2020-03-20 Department of Pathology, University of Cambridge COVID-19 Genomics UK (COG-UK) Consortium Luke W Meredith, M. Estee Torok , Myra Hosmillo, William L. Hamilton, Martin D. Curran, Theresa Feltwell, Anna Yakovleva, Charlotte J. Houldcroft, Aminu S. Jahun, Sarah L. Caddy, Ian Goodfellow
EPI_ISL_425319 hCoV-19/England/CAMB-74650/2020 Europe / United Kingdom / England 2020-03-28 Department of Pathology, University of Cambridge COVID-19 Genomics UK (COG-UK) Consortium Luke W Meredith, M. Estee Torok , Myra Hosmillo, William L. Hamilton, Martin D. Curran, Theresa Feltwell, Anna Yakovleva, Charlotte J. Houldcroft, Aminu S. Jahun, Sarah L. Caddy, Ian Goodfellow
EPI_ISL_425323 hCoV-19/England/CAMB-746AB/2020 Europe / United Kingdom / England 2020-03-29 Department of Pathology, University of Cambridge COVID-19 Genomics UK (COG-UK) Consortium Luke W Meredith, M. Estee Torok , Myra Hosmillo, William L. Hamilton, Martin D. Curran, Theresa Feltwell, Anna Yakovleva, Charlotte J. Houldcroft, Aminu S. Jahun, Sarah L. Caddy, Ian Goodfellow
EPI_ISL_425324 hCoV-19/England/CAMB-746BA/2020 Europe / United Kingdom / England 2020-03-29 Department of Pathology, University of Cambridge COVID-19 Genomics UK (COG-UK) Consortium Luke W Meredith, M. Estee Torok , Myra Hosmillo, William L. Hamilton, Martin D. Curran, Theresa Feltwell, Anna Yakovleva, Charlotte J. Houldcroft, Aminu S. Jahun, Sarah L. Caddy, Ian Goodfellow
EPI_ISL_425325 hCoV-19/England/CAMB-746D8/2020 Europe / United Kingdom / England 2020-03-29 Department of Pathology, University of Cambridge COVID-19 Genomics UK (COG-UK) Consortium Luke W Meredith, M. Estee Torok , Myra Hosmillo, William L. Hamilton, Martin D. Curran, Theresa Feltwell, Anna Yakovleva, Charlotte J. Houldcroft, Aminu S. Jahun, Sarah L. Caddy, Ian Goodfellow
EPI_ISL_425326 hCoV-19/England/CAMB-74702/2020 Europe / United Kingdom / England 2020-03-29 Department of Pathology, University of Cambridge COVID-19 Genomics UK (COG-UK) Consortium Luke W Meredith, M. Estee Torok , Myra Hosmillo, William L. Hamilton, Martin D. Curran, Theresa Feltwell, Anna Yakovleva, Charlotte J. Houldcroft, Aminu S. Jahun, Sarah L. Caddy, Ian Goodfellow
EPI_ISL_425327 hCoV-19/England/CAMB-74711/2020 Europe / United Kingdom / England 2020-03-29 Department of Pathology, University of Cambridge COVID-19 Genomics UK (COG-UK) Consortium Luke W Meredith, M. Estee Torok , Myra Hosmillo, William L. Hamilton, Martin D. Curran, Theresa Feltwell, Anna Yakovleva, Charlotte J. Houldcroft, Aminu S. Jahun, Sarah L. Caddy, Ian Goodfellow
EPI_ISL_425331 hCoV-19/England/CAMB-7475D/2020 Europe / United Kingdom / England 2020-03-29 Department of Pathology, University of Cambridge COVID-19 Genomics UK (COG-UK) Consortium Luke W Meredith, M. Estee Torok , Myra Hosmillo, William L. Hamilton, Martin D. Curran, Theresa Feltwell, Anna Yakovleva, Charlotte J. Houldcroft, Aminu S. Jahun, Sarah L. Caddy, Ian Goodfellow
EPI_ISL_425332 hCoV-19/England/CAMB-7476C/2020 Europe / United Kingdom / England 2020-03-29 Department of Pathology, University of Cambridge COVID-19 Genomics UK (COG-UK) Consortium Luke W Meredith, M. Estee Torok , Myra Hosmillo, William L. Hamilton, Martin D. Curran, Theresa Feltwell, Anna Yakovleva, Charlotte J. Houldcroft, Aminu S. Jahun, Sarah L. Caddy, Ian Goodfellow
EPI_ISL_425333 hCoV-19/England/CAMB-7477B/2020 Europe / United Kingdom / England 2020-03-29 Department of Pathology, University of Cambridge COVID-19 Genomics UK (COG-UK) Consortium Luke W Meredith, M. Estee Torok , Myra Hosmillo, William L. Hamilton, Martin D. Curran, Theresa Feltwell, Anna Yakovleva, Charlotte J. Houldcroft, Aminu S. Jahun, Sarah L. Caddy, Ian Goodfellow
EPI_ISL_425335 hCoV-19/England/CAMB-748E1/2020 Europe / United Kingdom / England 2020-03-28 Department of Pathology, University of Cambridge COVID-19 Genomics UK (COG-UK) Consortium Luke W Meredith, M. Estee Torok , Myra Hosmillo, William L. Hamilton, Martin D. Curran, Theresa Feltwell, Anna Yakovleva, Charlotte J. Houldcroft, Aminu S. Jahun, Sarah L. Caddy, Ian Goodfellow
EPI_ISL_425338 hCoV-19/England/CAMB-7491B/2020 Europe / United Kingdom / England 2020-03-28 Department of Pathology, University of Cambridge COVID-19 Genomics UK (COG-UK) Consortium Luke W Meredith, M. Estee Torok , Myra Hosmillo, William L. Hamilton, Martin D. Curran, Theresa Feltwell, Anna Yakovleva, Charlotte J. Houldcroft, Aminu S. Jahun, Sarah L. Caddy, Ian Goodfellow
EPI_ISL_425348 hCoV-19/England/CAMB-74B42/2020 Europe / United Kingdom / England 2020-03-18 Department of Pathology, University of Cambridge COVID-19 Genomics UK (COG-UK) Consortium Luke W Meredith, M. Estee Torok , Myra Hosmillo, William L. Hamilton, Martin D. Curran, Theresa Feltwell, Anna Yakovleva, Charlotte J. Houldcroft, Aminu S. Jahun, Sarah L. Caddy, Ian Goodfellow

EPI_ISL_425359  hCoV-19/England/CAMB-74C30/2020 Europe / United Kingdom / England       2020-03-17
Department of Pathology, University of Cambridge        COVID-19 Genomics UK (COG-UK) Consortium        Luke W Meredith, M. Estee Torok , Myra Hosmillo, William L. Hamilton, Martin D. Curran, Theresa Feltwell, Anna Yakovleva, Charlotte J. Houldcroft, Aminu S. Jahun, Sarah L. Caddy, Ian Goodfellow
EPI_ISL_425361  hCoV-19/England/CAMB-74C5E/2020 Europe / United Kingdom / England       2020-03-19
Department of Pathology, University of Cambridge        COVID-19 Genomics UK (COG-UK) Consortium        Luke W Meredith, M. Estee Torok , Myra Hosmillo, William L. Hamilton, Martin D. Curran, Theresa Feltwell, Anna Yakovleva, Charlotte J. Houldcroft, Aminu S. Jahun, Sarah L. Caddy, Ian Goodfellow
EPI_ISL_425362  hCoV-19/England/CAMB-74C8B/2020 Europe / United Kingdom / England       2020-03-17
Department of Pathology, University of Cambridge        COVID-19 Genomics UK (COG-UK) Consortium        Luke W Meredith, M. Estee Torok , Myra Hosmillo, William L. Hamilton, Martin D. Curran, Theresa Feltwell, Anna Yakovleva, Charlotte J. Houldcroft, Aminu S. Jahun, Sarah L. Caddy, Ian Goodfellow
EPI_ISL_425371  hCoV-19/England/CAMB-74D4C/2020 Europe / United Kingdom / England       2020-03-19
Department of Pathology, University of Cambridge        COVID-19 Genomics UK (COG-UK) Consortium        Luke W Meredith, M. Estee Torok , Myra Hosmillo, William L. Hamilton, Martin D. Curran, Theresa Feltwell, Anna Yakovleva, Charlotte J. Houldcroft, Aminu S. Jahun, Sarah L. Caddy, Ian Goodfellow
EPI_ISL_425373  hCoV-19/England/CAMB-74D5B/2020 Europe / United Kingdom / England       2020-03-19
Department of Pathology, University of Cambridge        COVID-19 Genomics UK (COG-UK) Consortium        Luke W Meredith, M. Estee Torok , Myra Hosmillo, William L. Hamilton, Martin D. Curran, Theresa Feltwell, Anna Yakovleva, Charlotte J. Houldcroft, Aminu S. Jahun, Sarah L. Caddy, Ian Goodfellow
EPI_ISL_425374  hCoV-19/England/CAMB-74D6A/2020 Europe / United Kingdom / England       2020-03-19
Department of Pathology, University of Cambridge        COVID-19 Genomics UK (COG-UK) Consortium        Luke W Meredith, M. Estee Torok , Myra Hosmillo, William L. Hamilton, Martin D. Curran, Theresa Feltwell, Anna Yakovleva, Charlotte J. Houldcroft, Aminu S. Jahun, Sarah L. Caddy, Ian Goodfellow
EPI_ISL_425382  hCoV-19/England/CAMB-74E85/2020 Europe / United Kingdom / England       2020-03-18
Department of Pathology, University of Cambridge        COVID-19 Genomics UK (COG-UK) Consortium        Luke W Meredith, M. Estee Torok , Myra Hosmillo, William L. Hamilton, Martin D. Curran, Theresa Feltwell, Anna Yakovleva, Charlotte J. Houldcroft, Aminu S. Jahun, Sarah L. Caddy, Ian Goodfellow
EPI_ISL_425611  hCoV-19/England/NOTT-10E8FA/2020        Europe / United Kingdom / England       2020-03-29
Queens Medical Centre, Clinical Microbiology Department / DeepSeq Nottingham    COVID-19 Genomics UK (COG-UK) Consortium      Gemma Clark, Wendy Smith, Manjinder Khakh, Hannah Howson-Wells, Jonathan Ball, Patrick McClure, Joseph Chappell, Theocharis Tsoleridis, Nadine Holmes, Matthew Carlisle, Christopher Moore, Fei Sang, Johnny Debebe, Victoria Wright, Matthew Loose
EPI_ISL_425614  hCoV-19/England/NOTT-10E933/2020        Europe / United Kingdom / England       2020-03-29
Queens Medical Centre, Clinical Microbiology Department / DeepSeq Nottingham    COVID-19 Genomics UK (COG-UK) Consortium      Gemma Clark, Wendy Smith, Manjinder Khakh, Hannah Howson-Wells, Jonathan Ball, Patrick McClure, Joseph Chappell, Theocharis Tsoleridis, Nadine Holmes, Matthew Carlisle, Christopher Moore, Fei Sang, Johnny Debebe, Victoria Wright, Matthew Loose
EPI_ISL_425621  hCoV-19/England/NOTT-10E9BB/2020        Europe / United Kingdom / England       2020-03-29
Queens Medical Centre, Clinical Microbiology Department / DeepSeq Nottingham    COVID-19 Genomics UK (COG-UK) Consortium      Gemma Clark, Wendy Smith, Manjinder Khakh, Hannah Howson-Wells, Jonathan Ball, Patrick McClure, Joseph Chappell, Theocharis Tsoleridis, Nadine Holmes, Matthew Carlisle, Christopher Moore, Fei Sang, Johnny Debebe, Victoria Wright, Matthew Loose
EPI_ISL_425628  hCoV-19/England/NOTT-10EA30/2020        Europe / United Kingdom / England       2020-03-30
Queens Medical Centre, Clinical Microbiology Department / DeepSeq Nottingham    COVID-19 Genomics UK (COG-UK) Consortium      Gemma Clark, Wendy Smith, Manjinder Khakh, Hannah Howson-Wells, Jonathan Ball, Patrick McClure, Joseph Chappell, Theocharis Tsoleridis, Nadine Holmes, Matthew Carlisle, Christopher Moore, Fei Sang, Johnny Debebe, Victoria Wright, Matthew Loose
EPI_ISL_425630  hCoV-19/England/NOTT-10EA5E/2020        Europe / United Kingdom / England       2020-03-29
Queens Medical Centre, Clinical Microbiology Department / DeepSeq Nottingham    COVID-19 Genomics UK (COG-UK) Consortium      Gemma Clark, Wendy Smith, Manjinder Khakh, Hannah Howson-Wells, Jonathan Ball, Patrick McClure, Joseph Chappell, Theocharis Tsoleridis, Nadine Holmes, Matthew Carlisle, Christopher Moore, Fei Sang, Johnny Debebe, Victoria Wright, Matthew Loose
EPI_ISL_425631  hCoV-19/England/NOTT-10EA6D/2020        Europe / United Kingdom / England       2020-03-29
Queens Medical Centre, Clinical Microbiology Department / DeepSeq Nottingham    COVID-19 Genomics UK (COG-UK) Consortium      Gemma Clark, Wendy Smith, Manjinder Khakh, Hannah Howson-Wells, Jonathan Ball, Patrick McClure, Joseph Chappell, Theocharis Tsoleridis, Nadine Holmes, Matthew Carlisle, Christopher Moore, Fei Sang, Johnny Debebe, Victoria Wright, Matthew Loose
EPI_ISL_425632  hCoV-19/England/NOTT-10EA7C/2020        Europe / United Kingdom / England       2020-03-30
Queens Medical Centre, Clinical Microbiology Department / DeepSeq Nottingham    COVID-19 Genomics UK (COG-UK) Consortium      Gemma Clark, Wendy Smith, Manjinder Khakh, Hannah Howson-Wells, Jonathan Ball, Patrick McClure, Joseph Chappell, Theocharis Tsoleridis, Nadine Holmes, Matthew Carlisle, Christopher Moore, Fei Sang, Johnny Debebe, Victoria Wright, Matthew Loose
EPI_ISL_425633  hCoV-19/England/NOTT-10EA8B/2020        Europe / United Kingdom / England       2020-03-29
Queens Medical Centre, Clinical Microbiology Department / DeepSeq Nottingham    COVID-19 Genomics UK (COG-UK) Consortium      Gemma Clark, Wendy Smith, Manjinder Khakh, Hannah Howson-Wells, Jonathan Ball, Patrick McClure, Joseph Chappell, Theocharis Tsoleridis, Nadine Holmes, Matthew Carlisle, Christopher Moore, Fei Sang, Johnny Debebe, Victoria Wright, Matthew Loose
EPI_ISL_425635  hCoV-19/England/NOTT-10EAA9/2020        Europe / United Kingdom / England       2020-03-30
Queens Medical Centre, Clinical Microbiology Department / DeepSeq Nottingham    COVID-19 Genomics UK (COG-UK)

Consortium	Gemma Clark, Wendy Smith, Manjinder Khakh, Hannah Howson-Wells, Jonathan Ball, Patrick McClure, Joseph Chappell, Theocharis Tsoleridis, Nadine Holmes, Matthew Carlisle, Christopher Moore, Fei Sang, Johnny Debebe, Victoria Wright, Matthew Loose
EPI_ISL_425638	hCoV-19/England/NOTT-10EAD6/2020	Europe / United Kingdom / England	2020-03-30	Queens Medical Centre, Clinical Microbiology Department / DeepSeq Nottingham	COVID-19 Genomics UK (COG-UK) Consortium	Gemma Clark, Wendy Smith, Manjinder Khakh, Hannah Howson-Wells, Jonathan Ball, Patrick McClure, Joseph Chappell, Theocharis Tsoleridis, Nadine Holmes, Matthew Carlisle, Christopher Moore, Fei Sang, Johnny Debebe, Victoria Wright, Matthew Loose
EPI_ISL_425639	hCoV-19/England/NOTT-10EAE5/2020	Europe / United Kingdom / England	2020-03-30	Queens Medical Centre, Clinical Microbiology Department / DeepSeq Nottingham	COVID-19 Genomics UK (COG-UK) Consortium	Gemma Clark, Wendy Smith, Manjinder Khakh, Hannah Howson-Wells, Jonathan Ball, Patrick McClure, Joseph Chappell, Theocharis Tsoleridis, Nadine Holmes, Matthew Carlisle, Christopher Moore, Fei Sang, Johnny Debebe, Victoria Wright, Matthew Loose
EPI_ISL_425640	hCoV-19/England/NOTT-10EAF4/2020	Europe / United Kingdom / England	2020-03-30	Queens Medical Centre, Clinical Microbiology Department / DeepSeq Nottingham	COVID-19 Genomics UK (COG-UK) Consortium	Gemma Clark, Wendy Smith, Manjinder Khakh, Hannah Howson-Wells, Jonathan Ball, Patrick McClure, Joseph Chappell, Theocharis Tsoleridis, Nadine Holmes, Matthew Carlisle, Christopher Moore, Fei Sang, Johnny Debebe, Victoria Wright, Matthew Loose
EPI_ISL_425642	hCoV-19/England/NOTT-10EB2E/2020	Europe / United Kingdom / England	2020-03-30	Queens Medical Centre, Clinical Microbiology Department / DeepSeq Nottingham	COVID-19 Genomics UK (COG-UK) Consortium	Gemma Clark, Wendy Smith, Manjinder Khakh, Hannah Howson-Wells, Jonathan Ball, Patrick McClure, Joseph Chappell, Theocharis Tsoleridis, Nadine Holmes, Matthew Carlisle, Christopher Moore, Fei Sang, Johnny Debebe, Victoria Wright, Matthew Loose
EPI_ISL_425644	hCoV-19/England/NOTT-10EB6A/2020	Europe / United Kingdom / England	2020-03-17	Queens Medical Centre, Clinical Microbiology Department / DeepSeq Nottingham	COVID-19 Genomics UK (COG-UK) Consortium	Gemma Clark, Wendy Smith, Manjinder Khakh, Hannah Howson-Wells, Jonathan Ball, Patrick McClure, Joseph Chappell, Theocharis Tsoleridis, Nadine Holmes, Matthew Carlisle, Christopher Moore, Fei Sang, Johnny Debebe, Victoria Wright, Matthew Loose
EPI_ISL_425645	hCoV-19/England/NOTT-10EB79/2020	Europe / United Kingdom / England	2020-03-18	Queens Medical Centre, Clinical Microbiology Department / DeepSeq Nottingham	COVID-19 Genomics UK (COG-UK) Consortium	Gemma Clark, Wendy Smith, Manjinder Khakh, Hannah Howson-Wells, Jonathan Ball, Patrick McClure, Joseph Chappell, Theocharis Tsoleridis, Nadine Holmes, Matthew Carlisle, Christopher Moore, Fei Sang, Johnny Debebe, Victoria Wright, Matthew Loose
EPI_ISL_425647	hCoV-19/Scotland/CVR100/2020	Europe / United Kingdom / Scotland	2020-03-14	West of Scotland Specialist Virology Centre, NHSGGC / MRC-University of Glasgow Centre for Virus Research	COVID-19 Genomics UK (COG-UK) Consortium	Ana da Silva Filipe, Kathy Smollett, Stephen Carmichael, Natasha Johnson, Daniel Mair, Lily Tong, Jenna Nichols; Sarah McDonald; Richard Orton, Joseph Hughes, Sreenu Vattipally, David L Robertson; Kathy Li, Natasha Jesudason, Rajiv Shah, James Shepherd, Antonia Ho, Emma Thomson; Alasdair MacLean, Rory Gunson.
EPI_ISL_425648	hCoV-19/Scotland/CVR101/2020	Europe / United Kingdom / Scotland	2020-03-14	West of Scotland Specialist Virology Centre, NHSGGC / MRC-University of Glasgow Centre for Virus Research	COVID-19 Genomics UK (COG-UK) Consortium	Ana da Silva Filipe, Kathy Smollett, Stephen Carmichael, Natasha Johnson, Daniel Mair, Lily Tong, Jenna Nichols; Sarah McDonald; Richard Orton, Joseph Hughes, Sreenu Vattipally, David L Robertson; Kathy Li, Natasha Jesudason, Rajiv Shah, James Shepherd, Antonia Ho, Emma Thomson; Alasdair MacLean, Rory Gunson.
EPI_ISL_425650	hCoV-19/Scotland/CVR103/2020	Europe / United Kingdom / Scotland	2020-03-13	West of Scotland Specialist Virology Centre, NHSGGC / MRC-University of Glasgow Centre for Virus Research	COVID-19 Genomics UK (COG-UK) Consortium	Ana da Silva Filipe, Kathy Smollett, Stephen Carmichael, Natasha Johnson, Daniel Mair, Lily Tong, Jenna Nichols; Sarah McDonald; Richard Orton, Joseph Hughes, Sreenu Vattipally, David L Robertson; Kathy Li, Natasha Jesudason, Rajiv Shah, James Shepherd, Antonia Ho, Emma Thomson; Alasdair MacLean, Rory Gunson.
EPI_ISL_425651	hCoV-19/Scotland/CVR104/2020	Europe / United Kingdom / Scotland	2020-03-13	West of Scotland Specialist Virology Centre, NHSGGC / MRC-University of Glasgow Centre for Virus Research	COVID-19 Genomics UK (COG-UK) Consortium	Ana da Silva Filipe, Kathy Smollett, Stephen Carmichael, Natasha Johnson, Daniel Mair, Lily Tong, Jenna Nichols; Sarah McDonald; Richard Orton, Joseph Hughes, Sreenu Vattipally, David L Robertson; Kathy Li, Natasha Jesudason, Rajiv Shah, James Shepherd, Antonia Ho, Emma Thomson; Alasdair MacLean, Rory Gunson.
EPI_ISL_425652	hCoV-19/Scotland/CVR106/2020	Europe / United Kingdom / Scotland	2020-03-13	West of Scotland Specialist Virology Centre, NHSGGC / MRC-University of Glasgow Centre for Virus Research	COVID-19 Genomics UK (COG-UK) Consortium	Ana da Silva Filipe, Kathy Smollett, Stephen Carmichael, Natasha Johnson, Daniel Mair, Lily Tong, Jenna Nichols; Sarah McDonald; Richard Orton, Joseph Hughes, Sreenu Vattipally, David L Robertson; Kathy Li, Natasha Jesudason, Rajiv Shah, James Shepherd, Antonia Ho, Emma Thomson; Alasdair MacLean, Rory Gunson.
EPI_ISL_425653	hCoV-19/Scotland/CVR107/2020	Europe / United Kingdom / Scotland	2020-03-13	West of Scotland Specialist Virology Centre, NHSGGC / MRC-University of Glasgow Centre for Virus Research	COVID-19 Genomics UK (COG-UK) Consortium	Ana da Silva Filipe, Kathy Smollett, Stephen Carmichael, Natasha Johnson, Daniel Mair, Lily Tong, Jenna Nichols; Sarah McDonald; Richard Orton, Joseph Hughes, Sreenu Vattipally, David L Robertson; Kathy Li, Natasha Jesudason, Rajiv Shah, James Shepherd, Antonia Ho, Emma Thomson; Alasdair MacLean, Rory Gunson.
EPI_ISL_425654	hCoV-19/Scotland/CVR108/2020	Europe / United Kingdom / Scotland	2020-03-13	West of

Scotland Specialist Virology Centre, NHSGGC / MRC-University of Glasgow Centre for Virus Research	COVID-19 Genomics UK (COG-UK) Consortium	Ana da Silva Filipe, Kathy Smollett, Stephen Carmichael, Natasha Johnson, Daniel Mair, Lily Tong, Jenna Nichols; Sarah McDonald; Richard Orton, Joseph Hughes, Sreenu Vattipally, David L Robertson; Kathy Li, Natasha Jesudason, Rajiv Shah, James Shepherd, Antonia Ho, Emma Thomson; Alasdair MacLean, Rory Gunson.
EPI_ISL_425656	hCoV-19/Scotland/CVR110/2020	Europe / United Kingdom / Scotland	2020-03-14	West of Scotland Specialist Virology Centre, NHSGGC / MRC-University of Glasgow Centre for Virus Research	COVID-19 Genomics UK (COG-UK) Consortium	Ana da Silva Filipe, Kathy Smollett, Stephen Carmichael, Natasha Johnson, Daniel Mair, Lily Tong, Jenna Nichols; Sarah McDonald; Richard Orton, Joseph Hughes, Sreenu Vattipally, David L Robertson; Kathy Li, Natasha Jesudason, Rajiv Shah, James Shepherd, Antonia Ho, Emma Thomson; Alasdair MacLean, Rory Gunson.
EPI_ISL_425657	hCoV-19/Scotland/CVR112/2020	Europe / United Kingdom / Scotland	2020-03-15	West of Scotland Specialist Virology Centre, NHSGGC / MRC-University of Glasgow Centre for Virus Research	COVID-19 Genomics UK (COG-UK) Consortium	Ana da Silva Filipe, Kathy Smollett, Stephen Carmichael, Natasha Johnson, Daniel Mair, Lily Tong, Jenna Nichols; Sarah McDonald; Richard Orton, Joseph Hughes, Sreenu Vattipally, David L Robertson; Kathy Li, Natasha Jesudason, Rajiv Shah, James Shepherd, Antonia Ho, Emma Thomson; Alasdair MacLean, Rory Gunson.
EPI_ISL_425658	hCoV-19/Scotland/CVR113/2020	Europe / United Kingdom / Scotland	2020-03-15	West of Scotland Specialist Virology Centre, NHSGGC / MRC-University of Glasgow Centre for Virus Research	COVID-19 Genomics UK (COG-UK) Consortium	Ana da Silva Filipe, Kathy Smollett, Stephen Carmichael, Natasha Johnson, Daniel Mair, Lily Tong, Jenna Nichols; Sarah McDonald; Richard Orton, Joseph Hughes, Sreenu Vattipally, David L Robertson; Kathy Li, Natasha Jesudason, Rajiv Shah, James Shepherd, Antonia Ho, Emma Thomson; Alasdair MacLean, Rory Gunson.
EPI_ISL_425659	hCoV-19/Scotland/CVR114/2020	Europe / United Kingdom / Scotland	2020-03-13	West of Scotland Specialist Virology Centre, NHSGGC / MRC-University of Glasgow Centre for Virus Research	COVID-19 Genomics UK (COG-UK) Consortium	Ana da Silva Filipe, Kathy Smollett, Stephen Carmichael, Natasha Johnson, Daniel Mair, Lily Tong, Jenna Nichols; Sarah McDonald; Richard Orton, Joseph Hughes, Sreenu Vattipally, David L Robertson; Kathy Li, Natasha Jesudason, Rajiv Shah, James Shepherd, Antonia Ho, Emma Thomson; Alasdair MacLean, Rory Gunson.
EPI_ISL_425660	hCoV-19/Scotland/CVR115/2020	Europe / United Kingdom / Scotland	2020-03-13	West of Scotland Specialist Virology Centre, NHSGGC / MRC-University of Glasgow Centre for Virus Research	COVID-19 Genomics UK (COG-UK) Consortium	Ana da Silva Filipe, Kathy Smollett, Stephen Carmichael, Natasha Johnson, Daniel Mair, Lily Tong, Jenna Nichols; Sarah McDonald; Richard Orton, Joseph Hughes, Sreenu Vattipally, David L Robertson; Kathy Li, Natasha Jesudason, Rajiv Shah, James Shepherd, Antonia Ho, Emma Thomson; Alasdair MacLean, Rory Gunson.
EPI_ISL_425662	hCoV-19/Scotland/CVR118/2020	Europe / United Kingdom / Scotland	2020-03-12	West of Scotland Specialist Virology Centre, NHSGGC / MRC-University of Glasgow Centre for Virus Research	COVID-19 Genomics UK (COG-UK) Consortium	Ana da Silva Filipe, Kathy Smollett, Stephen Carmichael, Natasha Johnson, Daniel Mair, Lily Tong, Jenna Nichols; Sarah McDonald; Richard Orton, Joseph Hughes, Sreenu Vattipally, David L Robertson; Kathy Li, Natasha Jesudason, Rajiv Shah, James Shepherd, Antonia Ho, Emma Thomson; Alasdair MacLean, Rory Gunson.
EPI_ISL_425663	hCoV-19/Scotland/CVR12/2020	Europe / United Kingdom / Scotland	2020-03-10	West of Scotland Specialist Virology Centre, NHSGGC / MRC-University of Glasgow Centre for Virus Research	COVID-19 Genomics UK (COG-UK) Consortium	Ana da Silva Filipe, Kathy Smollett, Stephen Carmichael, Natasha Johnson, Daniel Mair, Lily Tong, Jenna Nichols; Sarah McDonald; Richard Orton, Joseph Hughes, Sreenu Vattipally, David L Robertson; Kathy Li, Natasha Jesudason, Rajiv Shah, James Shepherd, Antonia Ho, Emma Thomson; Alasdair MacLean, Rory Gunson.
EPI_ISL_425668	hCoV-19/Scotland/CVR125/2020	Europe / United Kingdom / Scotland	2020-03-14	West of Scotland Specialist Virology Centre, NHSGGC / MRC-University of Glasgow Centre for Virus Research	COVID-19 Genomics UK (COG-UK) Consortium	Ana da Silva Filipe, Kathy Smollett, Stephen Carmichael, Natasha Johnson, Daniel Mair, Lily Tong, Jenna Nichols; Sarah McDonald; Richard Orton, Joseph Hughes, Sreenu Vattipally, David L Robertson; Kathy Li, Natasha Jesudason, Rajiv Shah, James Shepherd, Antonia Ho, Emma Thomson; Alasdair MacLean, Rory Gunson.
EPI_ISL_425669	hCoV-19/Scotland/CVR126/2020	Europe / United Kingdom / Scotland	2020-03-15	West of Scotland Specialist Virology Centre, NHSGGC / MRC-University of Glasgow Centre for Virus Research	COVID-19 Genomics UK (COG-UK) Consortium	Ana da Silva Filipe, Kathy Smollett, Stephen Carmichael, Natasha Johnson, Daniel Mair, Lily Tong, Jenna Nichols; Sarah McDonald; Richard Orton, Joseph Hughes, Sreenu Vattipally, David L Robertson; Kathy Li, Natasha Jesudason, Rajiv Shah, James Shepherd, Antonia Ho, Emma Thomson; Alasdair MacLean, Rory Gunson.
EPI_ISL_425670	hCoV-19/Scotland/CVR128/2020	Europe / United Kingdom / Scotland	2020-03-13	West of Scotland Specialist Virology Centre, NHSGGC / MRC-University of Glasgow Centre for Virus Research	COVID-19 Genomics UK (COG-UK) Consortium	Ana da Silva Filipe, Kathy Smollett, Stephen Carmichael, Natasha Johnson, Daniel Mair, Lily Tong, Jenna Nichols; Sarah McDonald; Richard Orton, Joseph Hughes, Sreenu Vattipally, David L Robertson; Kathy Li, Natasha Jesudason, Rajiv Shah, James Shepherd, Antonia Ho, Emma Thomson; Alasdair MacLean, Rory Gunson.
EPI_ISL_425671	hCoV-19/Scotland/CVR129/2020	Europe / United Kingdom / Scotland	2020-03-13	West of Scotland Specialist Virology Centre, NHSGGC / MRC-University of Glasgow Centre for Virus Research	COVID-19 Genomics UK (COG-UK) Consortium	Ana da Silva Filipe, Kathy Smollett, Stephen Carmichael, Natasha Johnson, Daniel Mair, Lily Tong, Jenna Nichols; Sarah McDonald; Richard Orton, Joseph Hughes, Sreenu Vattipally, David L Robertson; Kathy Li, Natasha Jesudason, Rajiv Shah, James Shepherd, Antonia Ho, Emma

Thomson; Alasdair MacLean, Rory Gunson.
EPI_ISL_425672 hCoV-19/Scotland/CVR13/2020 Europe / United Kingdom / Scotland 2020-03-11 West of Scotland Specialist Virology Centre, NHSGGC / MRC-University of Glasgow Centre for Virus Research COVID-19 Genomics UK (COG-UK) Consortium Ana da Silva Filipe, Kathy Smollett, Stephen Carmichael, Natasha Johnson, Daniel Mair, Lily Tong, Jenna Nichols; Sarah McDonald; Richard Orton, Joseph Hughes, Sreenu Vattipally, David L Robertson; Kathy Li, Natasha Jesudason, Rajiv Shah, James Shepherd, Antonia Ho, Emma Thomson; Alasdair MacLean, Rory Gunson.
EPI_ISL_425674 hCoV-19/Scotland/CVR131/2020 Europe / United Kingdom / Scotland 2020-03-12 West of Scotland Specialist Virology Centre, NHSGGC / MRC-University of Glasgow Centre for Virus Research COVID-19 Genomics UK (COG-UK) Consortium Ana da Silva Filipe, Kathy Smollett, Stephen Carmichael, Natasha Johnson, Daniel Mair, Lily Tong, Jenna Nichols; Sarah McDonald; Richard Orton, Joseph Hughes, Sreenu Vattipally, David L Robertson; Kathy Li, Natasha Jesudason, Rajiv Shah, James Shepherd, Antonia Ho, Emma Thomson; Alasdair MacLean, Rory Gunson.
EPI_ISL_425675 hCoV-19/Scotland/CVR132/2020 Europe / United Kingdom / Scotland 2020-03-16 West of Scotland Specialist Virology Centre, NHSGGC / MRC-University of Glasgow Centre for Virus Research COVID-19 Genomics UK (COG-UK) Consortium Ana da Silva Filipe, Kathy Smollett, Stephen Carmichael, Natasha Johnson, Daniel Mair, Lily Tong, Jenna Nichols; Sarah McDonald; Richard Orton, Joseph Hughes, Sreenu Vattipally, David L Robertson; Kathy Li, Natasha Jesudason, Rajiv Shah, James Shepherd, Antonia Ho, Emma Thomson; Alasdair MacLean, Rory Gunson.
EPI_ISL_425676 hCoV-19/Scotland/CVR133/2020 Europe / United Kingdom / Scotland 2020-03-16 West of Scotland Specialist Virology Centre, NHSGGC / MRC-University of Glasgow Centre for Virus Research COVID-19 Genomics UK (COG-UK) Consortium Ana da Silva Filipe, Kathy Smollett, Stephen Carmichael, Natasha Johnson, Daniel Mair, Lily Tong, Jenna Nichols; Sarah McDonald; Richard Orton, Joseph Hughes, Sreenu Vattipally, David L Robertson; Kathy Li, Natasha Jesudason, Rajiv Shah, James Shepherd, Antonia Ho, Emma Thomson; Alasdair MacLean, Rory Gunson.
EPI_ISL_425677 hCoV-19/Scotland/CVR134/2020 Europe / United Kingdom / Scotland 2020-03-16 West of Scotland Specialist Virology Centre, NHSGGC / MRC-University of Glasgow Centre for Virus Research COVID-19 Genomics UK (COG-UK) Consortium Ana da Silva Filipe, Kathy Smollett, Stephen Carmichael, Natasha Johnson, Daniel Mair, Lily Tong, Jenna Nichols; Sarah McDonald; Richard Orton, Joseph Hughes, Sreenu Vattipally, David L Robertson; Kathy Li, Natasha Jesudason, Rajiv Shah, James Shepherd, Antonia Ho, Emma Thomson; Alasdair MacLean, Rory Gunson.
EPI_ISL_425678 hCoV-19/Scotland/CVR135/2020 Europe / United Kingdom / Scotland 2020-03-16 West of Scotland Specialist Virology Centre, NHSGGC / MRC-University of Glasgow Centre for Virus Research COVID-19 Genomics UK (COG-UK) Consortium Ana da Silva Filipe, Kathy Smollett, Stephen Carmichael, Natasha Johnson, Daniel Mair, Lily Tong, Jenna Nichols; Sarah McDonald; Richard Orton, Joseph Hughes, Sreenu Vattipally, David L Robertson; Kathy Li, Natasha Jesudason, Rajiv Shah, James Shepherd, Antonia Ho, Emma Thomson; Alasdair MacLean, Rory Gunson.
EPI_ISL_425679 hCoV-19/Scotland/CVR136/2020 Europe / United Kingdom / Scotland 2020-03-16 West of Scotland Specialist Virology Centre, NHSGGC / MRC-University of Glasgow Centre for Virus Research COVID-19 Genomics UK (COG-UK) Consortium Ana da Silva Filipe, Kathy Smollett, Stephen Carmichael, Natasha Johnson, Daniel Mair, Lily Tong, Jenna Nichols; Sarah McDonald; Richard Orton, Joseph Hughes, Sreenu Vattipally, David L Robertson; Kathy Li, Natasha Jesudason, Rajiv Shah, James Shepherd, Antonia Ho, Emma Thomson; Alasdair MacLean, Rory Gunson.
EPI_ISL_425682 hCoV-19/Scotland/CVR139/2020 Europe / United Kingdom / Scotland 2020-03-16 West of Scotland Specialist Virology Centre, NHSGGC / MRC-University of Glasgow Centre for Virus Research COVID-19 Genomics UK (COG-UK) Consortium Ana da Silva Filipe, Kathy Smollett, Stephen Carmichael, Natasha Johnson, Daniel Mair, Lily Tong, Jenna Nichols; Sarah McDonald; Richard Orton, Joseph Hughes, Sreenu Vattipally, David L Robertson; Kathy Li, Natasha Jesudason, Rajiv Shah, James Shepherd, Antonia Ho, Emma Thomson; Alasdair MacLean, Rory Gunson.
EPI_ISL_425684 hCoV-19/Scotland/CVR140/2020 Europe / United Kingdom / Scotland 2020-03-16 West of Scotland Specialist Virology Centre, NHSGGC / MRC-University of Glasgow Centre for Virus Research COVID-19 Genomics UK (COG-UK) Consortium Ana da Silva Filipe, Kathy Smollett, Stephen Carmichael, Natasha Johnson, Daniel Mair, Lily Tong, Jenna Nichols; Sarah McDonald; Richard Orton, Joseph Hughes, Sreenu Vattipally, David L Robertson; Kathy Li, Natasha Jesudason, Rajiv Shah, James Shepherd, Antonia Ho, Emma Thomson; Alasdair MacLean, Rory Gunson.
EPI_ISL_425685 hCoV-19/Scotland/CVR141/2020 Europe / United Kingdom / Scotland 2020-03-16 West of Scotland Specialist Virology Centre, NHSGGC / MRC-University of Glasgow Centre for Virus Research COVID-19 Genomics UK (COG-UK) Consortium Ana da Silva Filipe, Kathy Smollett, Stephen Carmichael, Natasha Johnson, Daniel Mair, Lily Tong, Jenna Nichols; Sarah McDonald; Richard Orton, Joseph Hughes, Sreenu Vattipally, David L Robertson; Kathy Li, Natasha Jesudason, Rajiv Shah, James Shepherd, Antonia Ho, Emma Thomson; Alasdair MacLean, Rory Gunson.
EPI_ISL_425687 hCoV-19/Scotland/CVR143/2020 Europe / United Kingdom / Scotland 2020-03-17 West of Scotland Specialist Virology Centre, NHSGGC / MRC-University of Glasgow Centre for Virus Research COVID-19 Genomics UK (COG-UK) Consortium Ana da Silva Filipe, Kathy Smollett, Stephen Carmichael, Natasha Johnson, Daniel Mair, Lily Tong, Jenna Nichols; Sarah McDonald; Richard Orton, Joseph Hughes, Sreenu Vattipally, David L Robertson; Kathy Li, Natasha Jesudason, Rajiv Shah, James Shepherd, Antonia Ho, Emma Thomson; Alasdair MacLean, Rory Gunson.
EPI_ISL_425688 hCoV-19/Scotland/CVR144/2020 Europe / United Kingdom / Scotland 2020-03-17 West of Scotland Specialist Virology Centre, NHSGGC / MRC-University of Glasgow Centre for Virus Research COVID-19 Genomics UK (COG-UK) Consortium Ana da Silva Filipe, Kathy Smollett, Stephen Carmichael, Natasha

Johnson, Daniel Mair, Lily Tong, Jenna Nichols; Sarah McDonald; Richard Orton, Joseph Hughes, Sreenu Vattipally, David L Robertson; Kathy Li, Natasha Jesudason, Rajiv Shah, James Shepherd, Antonia Ho, Emma Thomson; Alasdair MacLean, Rory Gunson.
EPI_ISL_425692 hCoV-19/Scotland/CVR15/2020 Europe / United Kingdom / Scotland 2020-03-12 West of Scotland Specialist Virology Centre, NHSGGC / MRC-University of Glasgow Centre for Virus Research COVID-19 Genomics UK (COG-UK) Consortium Ana da Silva Filipe, Kathy Smollett, Stephen Carmichael, Natasha Johnson, Daniel Mair, Lily Tong, Jenna Nichols; Sarah McDonald; Richard Orton, Joseph Hughes, Sreenu Vattipally, David L Robertson; Kathy Li, Natasha Jesudason, Rajiv Shah, James Shepherd, Antonia Ho, Emma Thomson; Alasdair MacLean, Rory Gunson.
EPI_ISL_425693 hCoV-19/Scotland/CVR150/2020 Europe / United Kingdom / Scotland 2020-03-16 West of Scotland Specialist Virology Centre, NHSGGC / MRC-University of Glasgow Centre for Virus Research COVID-19 Genomics UK (COG-UK) Consortium Ana da Silva Filipe, Kathy Smollett, Stephen Carmichael, Natasha Johnson, Daniel Mair, Lily Tong, Jenna Nichols; Sarah McDonald; Richard Orton, Joseph Hughes, Sreenu Vattipally, David L Robertson; Kathy Li, Natasha Jesudason, Rajiv Shah, James Shepherd, Antonia Ho, Emma Thomson; Alasdair MacLean, Rory Gunson.
EPI_ISL_425695 hCoV-19/Scotland/CVR152/2020 Europe / United Kingdom / Scotland 2020-03-16 West of Scotland Specialist Virology Centre, NHSGGC / MRC-University of Glasgow Centre for Virus Research COVID-19 Genomics UK (COG-UK) Consortium Ana da Silva Filipe, Kathy Smollett, Stephen Carmichael, Natasha Johnson, Daniel Mair, Lily Tong, Jenna Nichols; Sarah McDonald; Richard Orton, Joseph Hughes, Sreenu Vattipally, David L Robertson; Kathy Li, Natasha Jesudason, Rajiv Shah, James Shepherd, Antonia Ho, Emma Thomson; Alasdair MacLean, Rory Gunson.
EPI_ISL_425696 hCoV-19/Scotland/CVR153/2020 Europe / United Kingdom / Scotland 2020-03-17 West of Scotland Specialist Virology Centre, NHSGGC / MRC-University of Glasgow Centre for Virus Research COVID-19 Genomics UK (COG-UK) Consortium Ana da Silva Filipe, Kathy Smollett, Stephen Carmichael, Natasha Johnson, Daniel Mair, Lily Tong, Jenna Nichols; Sarah McDonald; Richard Orton, Joseph Hughes, Sreenu Vattipally, David L Robertson; Kathy Li, Natasha Jesudason, Rajiv Shah, James Shepherd, Antonia Ho, Emma Thomson; Alasdair MacLean, Rory Gunson.
EPI_ISL_425697 hCoV-19/Scotland/CVR155/2020 Europe / United Kingdom / Scotland 2020-03-17 West of Scotland Specialist Virology Centre, NHSGGC / MRC-University of Glasgow Centre for Virus Research COVID-19 Genomics UK (COG-UK) Consortium Ana da Silva Filipe, Kathy Smollett, Stephen Carmichael, Natasha Johnson, Daniel Mair, Lily Tong, Jenna Nichols; Sarah McDonald; Richard Orton, Joseph Hughes, Sreenu Vattipally, David L Robertson; Kathy Li, Natasha Jesudason, Rajiv Shah, James Shepherd, Antonia Ho, Emma Thomson; Alasdair MacLean, Rory Gunson.
EPI_ISL_425698 hCoV-19/Scotland/CVR159/2020 Europe / United Kingdom / Scotland 2020-03-17 West of Scotland Specialist Virology Centre, NHSGGC / MRC-University of Glasgow Centre for Virus Research COVID-19 Genomics UK (COG-UK) Consortium Ana da Silva Filipe, Kathy Smollett, Stephen Carmichael, Natasha Johnson, Daniel Mair, Lily Tong, Jenna Nichols; Sarah McDonald; Richard Orton, Joseph Hughes, Sreenu Vattipally, David L Robertson; Kathy Li, Natasha Jesudason, Rajiv Shah, James Shepherd, Antonia Ho, Emma Thomson; Alasdair MacLean, Rory Gunson.
EPI_ISL_425699 hCoV-19/Scotland/CVR16/2020 Europe / United Kingdom / Scotland 2020-03-12 West of Scotland Specialist Virology Centre, NHSGGC / MRC-University of Glasgow Centre for Virus Research COVID-19 Genomics UK (COG-UK) Consortium Ana da Silva Filipe, Kathy Smollett, Stephen Carmichael, Natasha Johnson, Daniel Mair, Lily Tong, Jenna Nichols; Sarah McDonald; Richard Orton, Joseph Hughes, Sreenu Vattipally, David L Robertson; Kathy Li, Natasha Jesudason, Rajiv Shah, James Shepherd, Antonia Ho, Emma Thomson; Alasdair MacLean, Rory Gunson.
EPI_ISL_426617 hCoV-19/USA/NY-NYUMC162/2020 North America / USA / New York / Suffolk county 2020-04-01 NYU Langone Health Departments of Pathology and Medicine, New York University School of Medicine Maria Aguero-Rosenfeld, Brendan Belovarac, Margaret Black, Ludovic Boytard, John Cadley, Paolo Cotzia, John Chen, Dacia Dimartino, Xiaojun Feng, Tatyana Gindin, Emily Guzman, Adriana Heguy, Megan Hogan, Emily Huang, George Jour, Andrew Lytle, Christian Marier, Matthew T. Maurano, Mark J. Mulligan, Peter Meyn, Iman Osman, Jared Pinnell, Vanessa Raabe, Sitharam Ramaswami, Amy Rapkiewicz, Marie Samanovic-Golden, Antonio Serrano, Guomiao Shen, Matija Snuderl, Theodore Vougiouklakis, Nick Vulpescu, Gael Westby, Paul Zappile, Yutong Zhang
EPI_ISL_426618 hCoV-19/USA/NY-NYUMC163/2020 North America / USA / New York / Nassau county 2020-04-01 NYU Langone Health Departments of Pathology and Medicine, New York University School of Medicine Maria Aguero-Rosenfeld, Brendan Belovarac, Margaret Black, Ludovic Boytard, John Cadley, Paolo Cotzia, John Chen, Dacia Dimartino, Xiaojun Feng, Tatyana Gindin, Emily Guzman, Adriana Heguy, Megan Hogan, Emily Huang, George Jour, Andrew Lytle, Christian Marier, Matthew T. Maurano, Mark J. Mulligan, Peter Meyn, Iman Osman, Jared Pinnell, Vanessa Raabe, Sitharam Ramaswami, Amy Rapkiewicz, Marie Samanovic-Golden, Antonio Serrano, Guomiao Shen, Matija Snuderl, Theodore Vougiouklakis, Nick Vulpescu, Gael Westby, Paul Zappile, Yutong Zhang
EPI_ISL_426619 hCoV-19/USA/NY-NYUMC164/2020 North America / USA / New York / Nassau county 2020-04-01 NYU Langone Health Departments of Pathology and Medicine, New York University School of Medicine Maria Aguero-Rosenfeld, Brendan Belovarac, Margaret Black, Ludovic Boytard, John Cadley, Paolo Cotzia, John Chen, Dacia Dimartino, Xiaojun Feng, Tatyana Gindin, Emily Guzman, Adriana Heguy, Megan Hogan, Emily Huang, George Jour, Andrew Lytle, Christian Marier, Matthew T. Maurano, Mark J. Mulligan, Peter Meyn, Iman Osman, Jared Pinnell, Vanessa Raabe, Sitharam Ramaswami, Amy Rapkiewicz, Marie Samanovic-Golden, Antonio Serrano, Guomiao Shen, Matija Snuderl, Theodore Vougiouklakis, Nick Vulpescu, Gael Westby, Paul Zappile, Yutong Zhang
EPI_ISL_426620 hCoV-19/USA/NY-NYUMC165/2020 North America / USA / New York / Brooklyn 2020-04-01 NYU Langone Health Departments of Pathology and Medicine, New York University School of Medicine Maria Aguero-Rosenfeld, Brendan Belovarac, Margaret Black, Ludovic Boytard, John Cadley, Paolo Cotzia, John Chen, Dacia Dimartino, Xiaojun Feng, Tatyana Gindin, Emily Guzman, Adriana Heguy, Megan Hogan, Emily Huang, George

Jour, Andrew Lytle, Christian Marier, Matthew T. Maurano, Mark J. Mulligan, Peter Meyn, Iman Osman, Jared Pinnell, Vanessa Raabe, Sitharam Ramaswami, Amy Rapkiewicz, Marie Samanovic-Golden, Antonio Serrano, Guomiao Shen, Matija Snuderl, Theodore Vougiouklakis, Nick Vulpescu, Gael Westby, Paul Zappile, Yutong Zhang
EPI_ISL_426621 hCoV-19/USA/NY-NYUMC166/2020 North America / USA / New York / Brooklyn 2020-04-01 NYU Langone Health Departments of Pathology and Medicine, New York University School of Medicine Maria Aguero-Rosenfeld, Brendan Belovarac, Margaret Black, Ludovic Boytard, John Cadley, Paolo Cotzia, John Chen, Dacia Dimartino, Xiaojun Feng, Tatyana Gindin, Emily Guzman, Adriana Heguy, Megan Hogan, Emily Huang, George Jour, Andrew Lytle, Christian Marier, Matthew T. Maurano, Mark J. Mulligan, Peter Meyn, Iman Osman, Jared Pinnell, Vanessa Raabe, Sitharam Ramaswami, Amy Rapkiewicz, Marie Samanovic-Golden, Antonio Serrano, Guomiao Shen, Matija Snuderl, Theodore Vougiouklakis, Nick Vulpescu, Gael Westby, Paul Zappile, Yutong Zhang
EPI_ISL_426622 hCoV-19/USA/NY-NYUMC167/2020 North America / USA / New York / Manhattan 2020-04-01 NYU Langone Health Departments of Pathology and Medicine, New York University School of Medicine Maria Aguero-Rosenfeld, Brendan Belovarac, Margaret Black, Ludovic Boytard, John Cadley, Paolo Cotzia, John Chen, Dacia Dimartino, Xiaojun Feng, Tatyana Gindin, Emily Guzman, Adriana Heguy, Megan Hogan, Emily Huang, George Jour, Andrew Lytle, Christian Marier, Matthew T. Maurano, Mark J. Mulligan, Peter Meyn, Iman Osman, Jared Pinnell, Vanessa Raabe, Sitharam Ramaswami, Amy Rapkiewicz, Marie Samanovic-Golden, Antonio Serrano, Guomiao Shen, Matija Snuderl, Theodore Vougiouklakis, Nick Vulpescu, Gael Westby, Paul Zappile, Yutong Zhang
EPI_ISL_426623 hCoV-19/USA/NY-NYUMC168/2020 North America / USA / New York / Manhattan 2020-04-01 NYU Langone Health Departments of Pathology and Medicine, New York University School of Medicine Maria Aguero-Rosenfeld, Brendan Belovarac, Margaret Black, Ludovic Boytard, John Cadley, Paolo Cotzia, John Chen, Dacia Dimartino, Xiaojun Feng, Tatyana Gindin, Emily Guzman, Adriana Heguy, Megan Hogan, Emily Huang, George Jour, Andrew Lytle, Christian Marier, Matthew T. Maurano, Mark J. Mulligan, Peter Meyn, Iman Osman, Jared Pinnell, Vanessa Raabe, Sitharam Ramaswami, Amy Rapkiewicz, Marie Samanovic-Golden, Antonio Serrano, Guomiao Shen, Matija Snuderl, Theodore Vougiouklakis, Nick Vulpescu, Gael Westby, Paul Zappile, Yutong Zhang
EPI_ISL_426624 hCoV-19/USA/NY-NYUMC169/2020 North America / USA / New York / Brooklyn 2020-04-01 NYU Langone Health Departments of Pathology and Medicine, New York University School of Medicine Maria Aguero-Rosenfeld, Brendan Belovarac, Margaret Black, Ludovic Boytard, John Cadley, Paolo Cotzia, John Chen, Dacia Dimartino, Xiaojun Feng, Tatyana Gindin, Emily Guzman, Adriana Heguy, Megan Hogan, Emily Huang, George Jour, Andrew Lytle, Christian Marier, Matthew T. Maurano, Mark J. Mulligan, Peter Meyn, Iman Osman, Jared Pinnell, Vanessa Raabe, Sitharam Ramaswami, Amy Rapkiewicz, Marie Samanovic-Golden, Antonio Serrano, Guomiao Shen, Matija Snuderl, Theodore Vougiouklakis, Nick Vulpescu, Gael Westby, Paul Zappile, Yutong Zhang
EPI_ISL_426625 hCoV-19/USA/NY-NYUMC170/2020 North America / USA / New York / Brooklyn 2020-04-01 NYU Langone Health Departments of Pathology and Medicine, New York University School of Medicine Maria Aguero-Rosenfeld, Brendan Belovarac, Margaret Black, Ludovic Boytard, John Cadley, Paolo Cotzia, John Chen, Dacia Dimartino, Xiaojun Feng, Tatyana Gindin, Emily Guzman, Adriana Heguy, Megan Hogan, Emily Huang, George Jour, Andrew Lytle, Christian Marier, Matthew T. Maurano, Mark J. Mulligan, Peter Meyn, Iman Osman, Jared Pinnell, Vanessa Raabe, Sitharam Ramaswami, Amy Rapkiewicz, Marie Samanovic-Golden, Antonio Serrano, Guomiao Shen, Matija Snuderl, Theodore Vougiouklakis, Nick Vulpescu, Gael Westby, Paul Zappile, Yutong Zhang
EPI_ISL_426626 hCoV-19/USA/NY-NYUMC171/2020 North America / USA / New York / Brooklyn 2020-04-01 NYU Langone Health Departments of Pathology and Medicine, New York University School of Medicine Maria Aguero-Rosenfeld, Brendan Belovarac, Margaret Black, Ludovic Boytard, John Cadley, Paolo Cotzia, John Chen, Dacia Dimartino, Xiaojun Feng, Tatyana Gindin, Emily Guzman, Adriana Heguy, Megan Hogan, Emily Huang, George Jour, Andrew Lytle, Christian Marier, Matthew T. Maurano, Mark J. Mulligan, Peter Meyn, Iman Osman, Jared Pinnell, Vanessa Raabe, Sitharam Ramaswami, Amy Rapkiewicz, Marie Samanovic-Golden, Antonio Serrano, Guomiao Shen, Matija Snuderl, Theodore Vougiouklakis, Nick Vulpescu, Gael Westby, Paul Zappile, Yutong Zhang
EPI_ISL_426627 hCoV-19/USA/LA-BIE-096/2020 North America / USA / Louisiana 2020-04-06 Ochsner Health BioInfoExperts, LLC Amy Feehan, David Nolan, Rebecca Rose, Susanna Lamers, Sissy Cross, Julia-Garcia-Diaz, Tong Yang, Luke Caruso, David Moraga Amador, Wayra Navia, Lydia Von Borstel, Xiao Hui Zhou
EPI_ISL_426628 hCoV-19/USA/LA-BIE-097/2020 North America / USA / Louisiana 2020-04-06 Ochsner Health BioInfoExperts, LLC Amy Feehan, David Nolan, Rebecca Rose, Susanna Lamers, Sissy Cross, Julia-Garcia-Diaz, Tong Yang, Luke Caruso, David Moraga Amador, Wayra Navia, Lydia Von Borstel, Xiao Hui Zhou
EPI_ISL_426629 hCoV-19/Taiwan/TSGH-01/2020 Asia / Taiwan / Taipei 2020-02-07 TSGH-CP molecular lab TSGH-CP molecular lab Cherng-Lih Perng, Ming-Jr Jian, Chih-Kai Chang, Jung-Chung Lin, Kuo-Ming Yeh, Chien-Wen Chen, Sheng-Kang Chiu, Hsing-Yi Chung, Shih-Hung Tsai, Kuo-Sheng Hung, Feng-Yee Chang, Hung-Sheng Shang
EPI_ISL_426637 hCoV-19/Australia/VIC327/2020 Oceania / Australia / Victoria 2020-03-17 Victorian Infectious Diseases Reference Laboratory (VIDRL) Microbiological Diagnostic Unit Public Health Laboratory and Victorian Infectious Diseases Reference Laboratory, Doherty Institute Caly L., Seemann T., Sait, M., Schultz M., Druce J., Sherry, N.
EPI_ISL_426638 hCoV-19/Australia/VIC328/2020 Oceania / Australia / Victoria 2020-03-17 Victorian Infectious Diseases Reference Laboratory (VIDRL) Microbiological Diagnostic Unit Public Health Laboratory and Victorian Infectious Diseases Reference Laboratory, Doherty Institute Caly L., Seemann T., Sait, M., Schultz M., Druce J., Sherry, N.
EPI_ISL_426639 hCoV-19/Australia/VIC329/2020 Oceania / Australia / Victoria 2020-03-17 Victorian Infectious Diseases Reference Laboratory (VIDRL) Microbiological Diagnostic Unit Public Health Laboratory and Victorian Infectious Diseases Reference Laboratory, Doherty Institute Caly L., Seemann T., Sait, M., Schultz M., Druce J., Sherry, N.
EPI_ISL_426640 hCoV-19/Australia/VIC330/2020 Oceania / Australia / Victoria 2020-03-18 Victorian Infectious Diseases Reference Laboratory (VIDRL) Microbiological Diagnostic Unit Public Health Laboratory and Victorian Infectious Diseases Reference Laboratory, Doherty Institute Caly L., Seemann T., Sait, M., Schultz M., Druce J., Sherry, N.

EPI_ISL_426641 hCoV-19/Australia/VIC332/2020 Oceania / Australia / Victoria 2020-03-18 Victorian Infectious Diseases Reference Laboratory (VIDRL) Microbiological Diagnostic Unit Public Health Laboratory and Victorian Infectious Diseases Reference Laboratory, Doherty Institute Caly L., Seemann T., Sait, M., Schultz M., Druce J., Sherry, N.
EPI_ISL_426642 hCoV-19/Australia/VIC333/2020 Oceania / Australia / Victoria 2020-03-18 Victorian Infectious Diseases Reference Laboratory (VIDRL) Microbiological Diagnostic Unit Public Health Laboratory and Victorian Infectious Diseases Reference Laboratory, Doherty Institute Caly L., Seemann T., Sait, M., Schultz M., Druce J., Sherry, N.
EPI_ISL_426643 hCoV-19/Australia/VIC334/2020 Oceania / Australia / Victoria 2020-03-19 Victorian Infectious Diseases Reference Laboratory (VIDRL) Microbiological Diagnostic Unit Public Health Laboratory and Victorian Infectious Diseases Reference Laboratory, Doherty Institute Caly L., Seemann T., Sait, M., Schultz M., Druce J., Sherry, N.
EPI_ISL_426644 hCoV-19/Australia/VIC335/2020 Oceania / Australia / Victoria 2020-03-20 Victorian Infectious Diseases Reference Laboratory (VIDRL) Microbiological Diagnostic Unit Public Health Laboratory and Victorian Infectious Diseases Reference Laboratory, Doherty Institute Caly L., Seemann T., Sait, M., Schultz M., Druce J., Sherry, N.
EPI_ISL_426645 hCoV-19/Australia/VIC336/2020 Oceania / Australia / Victoria 2020-03-20 Victorian Infectious Diseases Reference Laboratory (VIDRL) Microbiological Diagnostic Unit Public Health Laboratory and Victorian Infectious Diseases Reference Laboratory, Doherty Institute Caly L., Seemann T., Sait, M., Schultz M., Druce J., Sherry, N.
EPI_ISL_426646 hCoV-19/Australia/VIC337/2020 Oceania / Australia / Victoria 2020-03-20 Victorian Infectious Diseases Reference Laboratory (VIDRL) Microbiological Diagnostic Unit Public Health Laboratory and Victorian Infectious Diseases Reference Laboratory, Doherty Institute Caly L., Seemann T., Sait, M., Schultz M., Druce J., Sherry, N.
EPI_ISL_426647 hCoV-19/Australia/VIC338/2020 Oceania / Australia / Victoria 2020-03-20 Victorian Infectious Diseases Reference Laboratory (VIDRL) Microbiological Diagnostic Unit Public Health Laboratory and Victorian Infectious Diseases Reference Laboratory, Doherty Institute Caly L., Seemann T., Sait, M., Schultz M., Druce J., Sherry, N.
EPI_ISL_426648 hCoV-19/Australia/VIC339/2020 Oceania / Australia / Victoria 2020-03-20 Victorian Infectious Diseases Reference Laboratory (VIDRL) Microbiological Diagnostic Unit Public Health Laboratory and Victorian Infectious Diseases Reference Laboratory, Doherty Institute Caly L., Seemann T., Sait, M., Schultz M., Druce J., Sherry, N.
EPI_ISL_426649 hCoV-19/Australia/VIC340/2020 Oceania / Australia / Victoria 2020-03-20 Victorian Infectious Diseases Reference Laboratory (VIDRL) Microbiological Diagnostic Unit Public Health Laboratory and Victorian Infectious Diseases Reference Laboratory, Doherty Institute Caly L., Seemann T., Sait, M., Schultz M., Druce J., Sherry, N.
EPI_ISL_426650 hCoV-19/Australia/VIC341/2020 Oceania / Australia / Victoria 2020-03-20 Victorian Infectious Diseases Reference Laboratory (VIDRL) Microbiological Diagnostic Unit Public Health Laboratory and Victorian Infectious Diseases Reference Laboratory, Doherty Institute Caly L., Seemann T., Sait, M., Schultz M., Druce J., Sherry, N.
EPI_ISL_426651 hCoV-19/Australia/VIC342/2020 Oceania / Australia / Victoria 2020-03-20 Victorian Infectious Diseases Reference Laboratory (VIDRL) Microbiological Diagnostic Unit Public Health Laboratory and Victorian Infectious Diseases Reference Laboratory, Doherty Institute Caly L., Seemann T., Sait, M., Schultz M., Druce J., Sherry, N.
EPI_ISL_426652 hCoV-19/Australia/VIC343/2020 Oceania / Australia / Victoria 2020-03-20 Victorian Infectious Diseases Reference Laboratory (VIDRL) Microbiological Diagnostic Unit Public Health Laboratory and Victorian Infectious Diseases Reference Laboratory, Doherty Institute Caly L., Seemann T., Sait, M., Schultz M., Druce J., Sherry, N.
EPI_ISL_426653 hCoV-19/Australia/VIC344/2020 Oceania / Australia / Victoria 2020-03-20 Victorian Infectious Diseases Reference Laboratory (VIDRL) Microbiological Diagnostic Unit Public Health Laboratory and Victorian Infectious Diseases Reference Laboratory, Doherty Institute Caly L., Seemann T., Sait, M., Schultz M., Druce J., Sherry, N.
EPI_ISL_426654 hCoV-19/Australia/VIC345/2020 Oceania / Australia / Victoria 2020-03-20 Victorian Infectious Diseases Reference Laboratory (VIDRL) Microbiological Diagnostic Unit Public Health Laboratory and Victorian Infectious Diseases Reference Laboratory, Doherty Institute Caly L., Seemann T., Sait, M., Schultz M., Druce J., Sherry, N.
EPI_ISL_426655 hCoV-19/Australia/VIC346/2020 Oceania / Australia / Victoria 2020-03-20 Victorian Infectious Diseases Reference Laboratory (VIDRL) Microbiological Diagnostic Unit Public Health Laboratory and Victorian Infectious Diseases Reference Laboratory, Doherty Institute Caly L., Seemann T., Sait, M., Schultz M., Druce J., Sherry, N.
EPI_ISL_426656 hCoV-19/Australia/VIC347/2020 Oceania / Australia / Victoria 2020-03-20 Victorian Infectious Diseases Reference Laboratory (VIDRL) Microbiological Diagnostic Unit Public Health Laboratory and Victorian Infectious Diseases Reference Laboratory, Doherty Institute Caly L., Seemann T., Sait, M., Schultz M., Druce J., Sherry, N.
EPI_ISL_426657 hCoV-19/Australia/VIC348/2020 Oceania / Australia / Victoria 2020-03-20 Victorian Infectious Diseases Reference Laboratory (VIDRL) Microbiological Diagnostic Unit Public Health Laboratory and Victorian Infectious Diseases Reference Laboratory, Doherty Institute Caly L., Seemann T., Sait, M., Schultz M., Druce J., Sherry, N.
EPI_ISL_426658 hCoV-19/Australia/VIC350/2020 Oceania / Australia / Victoria 2020-03-20 Victorian Infectious Diseases Reference Laboratory (VIDRL) Microbiological Diagnostic Unit Public Health

Laboratory and Victorian Infectious Diseases Reference Laboratory, Doherty Institute    Caly L., Seemann T., Sait, M., Schultz M., Druce J., Sherry, N.
EPI_ISL_426659  hCoV-19/Australia/VIC351/2020   Oceania / Australia / Victoria  2020-03-20      Victorian Infectious Diseases Reference Laboratory (VIDRL)        Microbiological Diagnostic Unit Public Health Laboratory and Victorian Infectious Diseases Reference Laboratory, Doherty Institute    Caly L., Seemann T., Sait, M., Schultz M., Druce J., Sherry, N.
EPI_ISL_426661  hCoV-19/Australia/VIC353/2020   Oceania / Australia / Victoria  2020-03-21      Victorian Infectious Diseases Reference Laboratory (VIDRL)        Microbiological Diagnostic Unit Public Health Laboratory and Victorian Infectious Diseases Reference Laboratory, Doherty Institute    Caly L., Seemann T., Sait, M., Schultz M., Druce J., Sherry, N.
EPI_ISL_426662  hCoV-19/Australia/VIC354/2020   Oceania / Australia / Victoria  2020-03-21      Victorian Infectious Diseases Reference Laboratory (VIDRL)        Microbiological Diagnostic Unit Public Health Laboratory and Victorian Infectious Diseases Reference Laboratory, Doherty Institute    Caly L., Seemann T., Sait, M., Schultz M., Druce J., Sherry, N.
EPI_ISL_426663  hCoV-19/Australia/VIC355/2020   Oceania / Australia / Victoria  2020-03-21      Victorian Infectious Diseases Reference Laboratory (VIDRL)        Microbiological Diagnostic Unit Public Health Laboratory and Victorian Infectious Diseases Reference Laboratory, Doherty Institute    Caly L., Seemann T., Sait, M., Schultz M., Druce J., Sherry, N.
EPI_ISL_426664  hCoV-19/Australia/VIC356/2020   Oceania / Australia / Victoria  2020-03-21      Victorian Infectious Diseases Reference Laboratory (VIDRL)        Microbiological Diagnostic Unit Public Health Laboratory and Victorian Infectious Diseases Reference Laboratory, Doherty Institute    Caly L., Seemann T., Sait, M., Schultz M., Druce J., Sherry, N.
EPI_ISL_426665  hCoV-19/Australia/VIC357/2020   Oceania / Australia / Victoria  2020-03-21      Victorian Infectious Diseases Reference Laboratory (VIDRL)        Microbiological Diagnostic Unit Public Health Laboratory and Victorian Infectious Diseases Reference Laboratory, Doherty Institute    Caly L., Seemann T., Sait, M., Schultz M., Druce J., Sherry, N.
EPI_ISL_426666  hCoV-19/Australia/VIC358/2020   Oceania / Australia / Victoria  2020-03-21      Victorian Infectious Diseases Reference Laboratory (VIDRL)        Microbiological Diagnostic Unit Public Health Laboratory and Victorian Infectious Diseases Reference Laboratory, Doherty Institute    Caly L., Seemann T., Sait, M., Schultz M., Druce J., Sherry, N.
EPI_ISL_426667  hCoV-19/Australia/VIC359/2020   Oceania / Australia / Victoria  2020-03-22      Victorian Infectious Diseases Reference Laboratory (VIDRL)        Microbiological Diagnostic Unit Public Health Laboratory and Victorian Infectious Diseases Reference Laboratory, Doherty Institute    Caly L., Seemann T., Sait, M., Schultz M., Druce J., Sherry, N.
EPI_ISL_426669  hCoV-19/Australia/VIC361/2020   Oceania / Australia / Victoria  2020-03-22      Victorian Infectious Diseases Reference Laboratory (VIDRL)        Microbiological Diagnostic Unit Public Health Laboratory and Victorian Infectious Diseases Reference Laboratory, Doherty Institute    Caly L., Seemann T., Sait, M., Schultz M., Druce J., Sherry, N.
EPI_ISL_426670  hCoV-19/Australia/VIC362/2020   Oceania / Australia / Victoria  2020-03-22      Victorian Infectious Diseases Reference Laboratory (VIDRL)        Microbiological Diagnostic Unit Public Health Laboratory and Victorian Infectious Diseases Reference Laboratory, Doherty Institute    Caly L., Seemann T., Sait, M., Schultz M., Druce J., Sherry, N.
EPI_ISL_426671  hCoV-19/Australia/VIC363/2020   Oceania / Australia / Victoria  2020-03-22      Victorian Infectious Diseases Reference Laboratory (VIDRL)        Microbiological Diagnostic Unit Public Health Laboratory and Victorian Infectious Diseases Reference Laboratory, Doherty Institute    Caly L., Seemann T., Sait, M., Schultz M., Druce J., Sherry, N.
EPI_ISL_426672  hCoV-19/Australia/VIC364/2020   Oceania / Australia / Victoria  2020-03-22      Victorian Infectious Diseases Reference Laboratory (VIDRL)        Microbiological Diagnostic Unit Public Health Laboratory and Victorian Infectious Diseases Reference Laboratory, Doherty Institute    Caly L., Seemann T., Sait, M., Schultz M., Druce J., Sherry, N.
EPI_ISL_426673  hCoV-19/Australia/VIC365/2020   Oceania / Australia / Victoria  2020-03-22      Victorian Infectious Diseases Reference Laboratory (VIDRL)        Microbiological Diagnostic Unit Public Health Laboratory and Victorian Infectious Diseases Reference Laboratory, Doherty Institute    Caly L., Seemann T., Sait, M., Schultz M., Druce J., Sherry, N.
EPI_ISL_426674  hCoV-19/Australia/VIC366/2020   Oceania / Australia / Victoria  2020-03-22      Victorian Infectious Diseases Reference Laboratory (VIDRL)        Microbiological Diagnostic Unit Public Health Laboratory and Victorian Infectious Diseases Reference Laboratory, Doherty Institute    Caly L., Seemann T., Sait, M., Schultz M., Druce J., Sherry, N.
EPI_ISL_426675  hCoV-19/Australia/VIC367/2020   Oceania / Australia / Victoria  2020-03-22      Victorian Infectious Diseases Reference Laboratory (VIDRL)        Microbiological Diagnostic Unit Public Health Laboratory and Victorian Infectious Diseases Reference Laboratory, Doherty Institute    Caly L., Seemann T., Sait, M., Schultz M., Druce J., Sherry, N.
EPI_ISL_426676  hCoV-19/Australia/VIC368/2020   Oceania / Australia / Victoria  2020-03-22      Victorian Infectious Diseases Reference Laboratory (VIDRL)        Microbiological Diagnostic Unit Public Health Laboratory and Victorian Infectious Diseases Reference Laboratory, Doherty Institute    Caly L., Seemann T., Sait, M., Schultz M., Druce J., Sherry, N.
EPI_ISL_426677  hCoV-19/Australia/VIC369/2020   Oceania / Australia / Victoria  2020-03-22      Victorian Infectious Diseases Reference Laboratory (VIDRL)        Microbiological Diagnostic Unit Public Health Laboratory and Victorian Infectious Diseases Reference Laboratory, Doherty Institute    Caly L., Seemann T., Sait, M., Schultz M., Druce J., Sherry, N.

EPI_ISL_426679 hCoV-19/Australia/VIC372/2020 Oceania / Australia / Victoria 2020-03-22 Victorian Infectious Diseases Reference Laboratory (VIDRL) Microbiological Diagnostic Unit Public Health Laboratory and Victorian Infectious Diseases Reference Laboratory, Doherty Institute Caly L., Seemann T., Sait, M., Schultz M., Druce J., Sherry, N.
EPI_ISL_426680 hCoV-19/Australia/VIC373/2020 Oceania / Australia / Victoria 2020-03-22 Victorian Infectious Diseases Reference Laboratory (VIDRL) Microbiological Diagnostic Unit Public Health Laboratory and Victorian Infectious Diseases Reference Laboratory, Doherty Institute Caly L., Seemann T., Sait, M., Schultz M., Druce J., Sherry, N.
EPI_ISL_426682 hCoV-19/Australia/VIC375/2020 Oceania / Australia / Victoria 2020-03-22 Victorian Infectious Diseases Reference Laboratory (VIDRL) Microbiological Diagnostic Unit Public Health Laboratory and Victorian Infectious Diseases Reference Laboratory, Doherty Institute Caly L., Seemann T., Sait, M., Schultz M., Druce J., Sherry, N.
EPI_ISL_426683 hCoV-19/Australia/VIC376/2020 Oceania / Australia / Victoria 2020-03-22 Victorian Infectious Diseases Reference Laboratory (VIDRL) Microbiological Diagnostic Unit Public Health Laboratory and Victorian Infectious Diseases Reference Laboratory, Doherty Institute Caly L., Seemann T., Sait, M., Schultz M., Druce J., Sherry, N.
EPI_ISL_426684 hCoV-19/Australia/VIC377/2020 Oceania / Australia / Victoria 2020-03-22 Victorian Infectious Diseases Reference Laboratory (VIDRL) Microbiological Diagnostic Unit Public Health Laboratory and Victorian Infectious Diseases Reference Laboratory, Doherty Institute Caly L., Seemann T., Sait, M., Schultz M., Druce J., Sherry, N.
EPI_ISL_426685 hCoV-19/Australia/VIC378/2020 Oceania / Australia / Victoria 2020-03-22 Victorian Infectious Diseases Reference Laboratory (VIDRL) Microbiological Diagnostic Unit Public Health Laboratory and Victorian Infectious Diseases Reference Laboratory, Doherty Institute Caly L., Seemann T., Sait, M., Schultz M., Druce J., Sherry, N.
EPI_ISL_426686 hCoV-19/Australia/VIC379/2020 Oceania / Australia / Victoria 2020-03-22 Victorian Infectious Diseases Reference Laboratory (VIDRL) Microbiological Diagnostic Unit Public Health Laboratory and Victorian Infectious Diseases Reference Laboratory, Doherty Institute Caly L., Seemann T., Sait, M., Schultz M., Druce J., Sherry, N.
EPI_ISL_426687 hCoV-19/Australia/VIC380/2020 Oceania / Australia / Victoria 2020-03-23 Victorian Infectious Diseases Reference Laboratory (VIDRL) Microbiological Diagnostic Unit Public Health Laboratory and Victorian Infectious Diseases Reference Laboratory, Doherty Institute Caly L., Seemann T., Sait, M., Schultz M., Druce J., Sherry, N.
EPI_ISL_426688 hCoV-19/Australia/VIC381/2020 Oceania / Australia / Victoria 2020-03-23 Victorian Infectious Diseases Reference Laboratory (VIDRL) Microbiological Diagnostic Unit Public Health Laboratory and Victorian Infectious Diseases Reference Laboratory, Doherty Institute Caly L., Seemann T., Sait, M., Schultz M., Druce J., Sherry, N.
EPI_ISL_426689 hCoV-19/Australia/VIC382/2020 Oceania / Australia / Victoria 2020-03-23 Victorian Infectious Diseases Reference Laboratory (VIDRL) Microbiological Diagnostic Unit Public Health Laboratory and Victorian Infectious Diseases Reference Laboratory, Doherty Institute Caly L., Seemann T., Sait, M., Schultz M., Druce J., Sherry, N.
EPI_ISL_426690 hCoV-19/Australia/VIC383/2020 Oceania / Australia / Victoria 2020-03-23 Victorian Infectious Diseases Reference Laboratory (VIDRL) Microbiological Diagnostic Unit Public Health Laboratory and Victorian Infectious Diseases Reference Laboratory, Doherty Institute Caly L., Seemann T., Sait, M., Schultz M., Druce J., Sherry, N.
EPI_ISL_426691 hCoV-19/Australia/VIC384/2020 Oceania / Australia / Victoria 2020-03-23 Victorian Infectious Diseases Reference Laboratory (VIDRL) Microbiological Diagnostic Unit Public Health Laboratory and Victorian Infectious Diseases Reference Laboratory, Doherty Institute Caly L., Seemann T., Sait, M., Schultz M., Druce J., Sherry, N.
EPI_ISL_426692 hCoV-19/Australia/VIC385/2020 Oceania / Australia / Victoria 2020-03-23 Victorian Infectious Diseases Reference Laboratory (VIDRL) Microbiological Diagnostic Unit Public Health Laboratory and Victorian Infectious Diseases Reference Laboratory, Doherty Institute Caly L., Seemann T., Sait, M., Schultz M., Druce J., Sherry, N.
EPI_ISL_426693 hCoV-19/Australia/VIC386/2020 Oceania / Australia / Victoria 2020-03-23 Victorian Infectious Diseases Reference Laboratory (VIDRL) Microbiological Diagnostic Unit Public Health Laboratory and Victorian Infectious Diseases Reference Laboratory, Doherty Institute Caly L., Seemann T., Sait, M., Schultz M., Druce J., Sherry, N.
EPI_ISL_426694 hCoV-19/Australia/VIC387/2020 Oceania / Australia / Victoria 2020-03-23 Victorian Infectious Diseases Reference Laboratory (VIDRL) Microbiological Diagnostic Unit Public Health Laboratory and Victorian Infectious Diseases Reference Laboratory, Doherty Institute Caly L., Seemann T., Sait, M., Schultz M., Druce J., Sherry, N.
EPI_ISL_426696 hCoV-19/Australia/VIC389/2020 Oceania / Australia / Victoria 2020-03-23 Victorian Infectious Diseases Reference Laboratory (VIDRL) Microbiological Diagnostic Unit Public Health Laboratory and Victorian Infectious Diseases Reference Laboratory, Doherty Institute Caly L., Seemann T., Sait, M., Schultz M., Druce J., Sherry, N.
EPI_ISL_426697 hCoV-19/Australia/VIC394/2020 Oceania / Australia / Victoria 2020-03-24 Victorian Infectious Diseases Reference Laboratory (VIDRL) Microbiological Diagnostic Unit Public Health Laboratory and Victorian Infectious Diseases Reference Laboratory, Doherty Institute Caly L., Seemann T., Sait, M., Schultz M., Druce J., Sherry, N.
EPI_ISL_426698 hCoV-19/Australia/VIC395/2020 Oceania / Australia / Victoria 2020-03-24 Victorian Infectious Diseases Reference Laboratory (VIDRL) Microbiological Diagnostic Unit Public Health

Laboratory and Victorian Infectious Diseases Reference Laboratory, Doherty Institute Caly L., Seemann T., Sait, M., Schultz M., Druce J., Sherry, N.
EPI_ISL_426914 hCoV-19/Australia/VIC483/2020 Oceania / Australia / Victoria 2020-03-29 Microbiological Diagnostic Unit Public Health Laboratory Microbiological Diagnostic Unit Public Health Laboratory Seemann T., Schultz M., Sait, M., Sherry, N.
EPI_ISL_426917 hCoV-19/Australia/VIC486/2020 Oceania / Australia / Victoria 2020-03-29 Microbiological Diagnostic Unit Public Health Laboratory Microbiological Diagnostic Unit Public Health Laboratory Seemann T., Schultz M., Sait, M., Sherry, N.
EPI_ISL_426918 hCoV-19/Australia/VIC487/2020 Oceania / Australia / Victoria 2020-03-29 Microbiological Diagnostic Unit Public Health Laboratory Microbiological Diagnostic Unit Public Health Laboratory Seemann T., Schultz M., Sait, M., Sherry, N.
EPI_ISL_426919 hCoV-19/Australia/VIC488/2020 Oceania / Australia / Victoria 2020-03-29 Microbiological Diagnostic Unit Public Health Laboratory Microbiological Diagnostic Unit Public Health Laboratory Seemann T., Schultz M., Sait, M., Sherry, N.
EPI_ISL_426920 hCoV-19/Australia/VIC489/2020 Oceania / Australia / Victoria 2020-03-29 Microbiological Diagnostic Unit Public Health Laboratory Microbiological Diagnostic Unit Public Health Laboratory Seemann T., Schultz M., Sait, M., Sherry, N.
EPI_ISL_426921 hCoV-19/Australia/VIC490/2020 Oceania / Australia / Victoria 2020-03-30 Microbiological Diagnostic Unit Public Health Laboratory Microbiological Diagnostic Unit Public Health Laboratory Seemann T., Schultz M., Sait, M., Sherry, N.
EPI_ISL_426922 hCoV-19/Australia/VIC491/2020 Oceania / Australia / Victoria 2020-03-30 Microbiological Diagnostic Unit Public Health Laboratory Microbiological Diagnostic Unit Public Health Laboratory Seemann T., Schultz M., Sait, M., Sherry, N.
EPI_ISL_426923 hCoV-19/Australia/VIC619/2020 Oceania / Australia / Victoria 2020-03-28 Victorian Infectious Diseases Reference Laboratory (VIDRL) Microbiological Diagnostic Unit Public Health Laboratory and Victorian Infectious Diseases Reference Laboratory, Doherty Institute Caly L., Seemann T., Sait, M., Schultz M., Druce J., Sherry, N.
EPI_ISL_426924 hCoV-19/Australia/VIC620/2020 Oceania / Australia / Victoria 2020-03-28 Victorian Infectious Diseases Reference Laboratory (VIDRL) Microbiological Diagnostic Unit Public Health Laboratory and Victorian Infectious Diseases Reference Laboratory, Doherty Institute Caly L., Seemann T., Sait, M., Schultz M., Druce J., Sherry, N.
EPI_ISL_426926 hCoV-19/Australia/VIC622/2020 Oceania / Australia / Victoria 2020-03-29 Victorian Infectious Diseases Reference Laboratory (VIDRL) Microbiological Diagnostic Unit Public Health Laboratory and Victorian Infectious Diseases Reference Laboratory, Doherty Institute Caly L., Seemann T., Sait, M., Schultz M., Druce J., Sherry, N.
EPI_ISL_426927 hCoV-19/Australia/VIC623/2020 Oceania / Australia / Victoria 2020-03-28 Victorian Infectious Diseases Reference Laboratory (VIDRL) Microbiological Diagnostic Unit Public Health Laboratory and Victorian Infectious Diseases Reference Laboratory, Doherty Institute Caly L., Seemann T., Sait, M., Schultz M., Druce J., Sherry, N.
EPI_ISL_426929 hCoV-19/Australia/VIC625/2020 Oceania / Australia / Victoria 2020-03-28 Victorian Infectious Diseases Reference Laboratory (VIDRL) Microbiological Diagnostic Unit Public Health Laboratory and Victorian Infectious Diseases Reference Laboratory, Doherty Institute Caly L., Seemann T., Sait, M., Schultz M., Druce J., Sherry, N.
EPI_ISL_426930 hCoV-19/Australia/VIC626/2020 Oceania / Australia / Victoria 2020-03-28 Victorian Infectious Diseases Reference Laboratory (VIDRL) Microbiological Diagnostic Unit Public Health Laboratory and Victorian Infectious Diseases Reference Laboratory, Doherty Institute Caly L., Seemann T., Sait, M., Schultz M., Druce J., Sherry, N.
EPI_ISL_426931 hCoV-19/Australia/VIC627/2020 Oceania / Australia / Victoria 2020-03-29 Victorian Infectious Diseases Reference Laboratory (VIDRL) Microbiological Diagnostic Unit Public Health Laboratory and Victorian Infectious Diseases Reference Laboratory, Doherty Institute Caly L., Seemann T., Sait, M., Schultz M., Druce J., Sherry, N.
EPI_ISL_426932 hCoV-19/Australia/VIC628/2020 Oceania / Australia / Victoria 2020-03-29 Victorian Infectious Diseases Reference Laboratory (VIDRL) Microbiological Diagnostic Unit Public Health Laboratory and Victorian Infectious Diseases Reference Laboratory, Doherty Institute Caly L., Seemann T., Sait, M., Schultz M., Druce J., Sherry, N.
EPI_ISL_426933 hCoV-19/Australia/VIC637/2020 Oceania / Australia / Victoria 2020-03-28 Victorian Infectious Diseases Reference Laboratory (VIDRL) Microbiological Diagnostic Unit Public Health Laboratory and Victorian Infectious Diseases Reference Laboratory, Doherty Institute Caly L., Seemann T., Sait, M., Schultz M., Druce J., Sherry, N.
EPI_ISL_426934 hCoV-19/Australia/VIC638/2020 Oceania / Australia / Victoria 2020-03-29 Victorian Infectious Diseases Reference Laboratory (VIDRL) Microbiological Diagnostic Unit Public Health Laboratory and Victorian Infectious Diseases Reference Laboratory, Doherty Institute Caly L., Seemann T., Sait, M., Schultz M., Druce J., Sherry, N.
EPI_ISL_426935 hCoV-19/Australia/VIC639/2020 Oceania / Australia / Victoria 2020-03-29 Victorian Infectious Diseases Reference Laboratory (VIDRL) Microbiological Diagnostic Unit Public Health Laboratory and Victorian Infectious Diseases Reference Laboratory, Doherty Institute Caly L., Seemann T., Sait, M., Schultz M., Druce J., Sherry, N.
EPI_ISL_426940 hCoV-19/Australia/VIC644/2020 Oceania / Australia / Victoria 2020-03-28 Victorian Infectious Diseases Reference Laboratory (VIDRL) Microbiological Diagnostic Unit Public Health Laboratory and Victorian Infectious Diseases Reference Laboratory, Doherty Institute Caly L., Seemann T.,

Sait, M., Schultz M., Druce J., Sherry, N.
EPI_ISL_426941 hCoV-19/Australia/VIC645/2020 Oceania / Australia / Victoria 2020-03-28 Victorian Infectious Diseases Reference Laboratory (VIDRL) Microbiological Diagnostic Unit Public Health Laboratory and Victorian Infectious Diseases Reference Laboratory, Doherty Institute Caly L., Seemann T., Sait, M., Schultz M., Druce J., Sherry, N.
EPI_ISL_426942 hCoV-19/Australia/VIC646/2020 Oceania / Australia / Victoria 2020-03-24 Victorian Infectious Diseases Reference Laboratory (VIDRL) Microbiological Diagnostic Unit Public Health Laboratory and Victorian Infectious Diseases Reference Laboratory, Doherty Institute Caly L., Seemann T., Sait, M., Schultz M., Druce J., Sherry, N.
EPI_ISL_426943 hCoV-19/Australia/VIC647/2020 Oceania / Australia / Victoria 2020-03-28 Victorian Infectious Diseases Reference Laboratory (VIDRL) Microbiological Diagnostic Unit Public Health Laboratory and Victorian Infectious Diseases Reference Laboratory, Doherty Institute Caly L., Seemann T., Sait, M., Schultz M., Druce J., Sherry, N.
EPI_ISL_426947 hCoV-19/Australia/VIC651/2020 Oceania / Australia / Victoria 2020-03-28 Victorian Infectious Diseases Reference Laboratory (VIDRL) Microbiological Diagnostic Unit Public Health Laboratory and Victorian Infectious Diseases Reference Laboratory, Doherty Institute Caly L., Seemann T., Sait, M., Schultz M., Druce J., Sherry, N.
EPI_ISL_426948 hCoV-19/Australia/VIC652/2020 Oceania / Australia / Victoria 2020-03-28 Victorian Infectious Diseases Reference Laboratory (VIDRL) Microbiological Diagnostic Unit Public Health Laboratory and Victorian Infectious Diseases Reference Laboratory, Doherty Institute Caly L., Seemann T., Sait, M., Schultz M., Druce J., Sherry, N.
EPI_ISL_426951 hCoV-19/Australia/VIC655/2020 Oceania / Australia / Victoria 2020-03-28 Victorian Infectious Diseases Reference Laboratory (VIDRL) Microbiological Diagnostic Unit Public Health Laboratory and Victorian Infectious Diseases Reference Laboratory, Doherty Institute Caly L., Seemann T., Sait, M., Schultz M., Druce J., Sherry, N.
EPI_ISL_426952 hCoV-19/Australia/VIC656/2020 Oceania / Australia / Victoria 2020-03-28 Victorian Infectious Diseases Reference Laboratory (VIDRL) Microbiological Diagnostic Unit Public Health Laboratory and Victorian Infectious Diseases Reference Laboratory, Doherty Institute Caly L., Seemann T., Sait, M., Schultz M., Druce J., Sherry, N.
EPI_ISL_426953 hCoV-19/Australia/VIC657/2020 Oceania / Australia / Victoria 2020-03-28 Victorian Infectious Diseases Reference Laboratory (VIDRL) Microbiological Diagnostic Unit Public Health Laboratory and Victorian Infectious Diseases Reference Laboratory, Doherty Institute Caly L., Seemann T., Sait, M., Schultz M., Druce J., Sherry, N.
EPI_ISL_426954 hCoV-19/Australia/VIC658/2020 Oceania / Australia / Victoria 2020-03-28 Victorian Infectious Diseases Reference Laboratory (VIDRL) Microbiological Diagnostic Unit Public Health Laboratory and Victorian Infectious Diseases Reference Laboratory, Doherty Institute Caly L., Seemann T., Sait, M., Schultz M., Druce J., Sherry, N.
EPI_ISL_426955 hCoV-19/Australia/VIC659/2020 Oceania / Australia / Victoria 2020-03-27 Victorian Infectious Diseases Reference Laboratory (VIDRL) Microbiological Diagnostic Unit Public Health Laboratory and Victorian Infectious Diseases Reference Laboratory, Doherty Institute Caly L., Seemann T., Sait, M., Schultz M., Druce J., Sherry, N.
EPI_ISL_426956 hCoV-19/Australia/VIC660/2020 Oceania / Australia / Victoria 2020-03-28 Victorian Infectious Diseases Reference Laboratory (VIDRL) Microbiological Diagnostic Unit Public Health Laboratory and Victorian Infectious Diseases Reference Laboratory, Doherty Institute Caly L., Seemann T., Sait, M., Schultz M., Druce J., Sherry, N.
EPI_ISL_426957 hCoV-19/Australia/VIC661/2020 Oceania / Australia / Victoria 2020-03-27 Victorian Infectious Diseases Reference Laboratory (VIDRL) Microbiological Diagnostic Unit Public Health Laboratory and Victorian Infectious Diseases Reference Laboratory, Doherty Institute Caly L., Seemann T., Sait, M., Schultz M., Druce J., Sherry, N.
EPI_ISL_426958 hCoV-19/Australia/VIC662/2020 Oceania / Australia / Victoria 2020-03-27 Victorian Infectious Diseases Reference Laboratory (VIDRL) Microbiological Diagnostic Unit Public Health Laboratory and Victorian Infectious Diseases Reference Laboratory, Doherty Institute Caly L., Seemann T., Sait, M., Schultz M., Druce J., Sherry, N.
EPI_ISL_426959 hCoV-19/Australia/VIC663/2020 Oceania / Australia / Victoria 2020-03-28 Victorian Infectious Diseases Reference Laboratory (VIDRL) Microbiological Diagnostic Unit Public Health Laboratory and Victorian Infectious Diseases Reference Laboratory, Doherty Institute Caly L., Seemann T., Sait, M., Schultz M., Druce J., Sherry, N.
EPI_ISL_426960 hCoV-19/Australia/VIC664/2020 Oceania / Australia / Victoria 2020-03-27 Victorian Infectious Diseases Reference Laboratory (VIDRL) Microbiological Diagnostic Unit Public Health Laboratory and Victorian Infectious Diseases Reference Laboratory, Doherty Institute Caly L., Seemann T., Sait, M., Schultz M., Druce J., Sherry, N.
EPI_ISL_426961 hCoV-19/Australia/VIC665/2020 Oceania / Australia / Victoria 2020-03-27 Victorian Infectious Diseases Reference Laboratory (VIDRL) Microbiological Diagnostic Unit Public Health Laboratory and Victorian Infectious Diseases Reference Laboratory, Doherty Institute Caly L., Seemann T., Sait, M., Schultz M., Druce J., Sherry, N.
EPI_ISL_426962 hCoV-19/Australia/VIC666/2020 Oceania / Australia / Victoria 2020-03-27 Victorian Infectious Diseases Reference Laboratory (VIDRL) Microbiological Diagnostic Unit Public Health Laboratory and Victorian Infectious Diseases Reference Laboratory, Doherty Institute Caly L., Seemann T., Sait, M., Schultz M., Druce J., Sherry, N.
EPI_ISL_426963 hCoV-19/Australia/VIC667/2020 Oceania / Australia / Victoria 2020-03-27 Victorian

Infectious Diseases Reference Laboratory (VIDRL)	Microbiological Diagnostic Unit Public Health Laboratory and Victorian Infectious Diseases Reference Laboratory, Doherty Institute	Caly L., Seemann T., Sait, M., Schultz M., Druce J., Sherry, N.
EPI_ISL_426964	hCoV-19/Australia/VIC668/2020	Oceania / Australia / Victoria	2020-03-27	Victorian Infectious Diseases Reference Laboratory (VIDRL)	Microbiological Diagnostic Unit Public Health Laboratory and Victorian Infectious Diseases Reference Laboratory, Doherty Institute	Caly L., Seemann T., Sait, M., Schultz M., Druce J., Sherry, N.
EPI_ISL_426968	hCoV-19/Australia/VIC672/2020	Oceania / Australia / Victoria	2020-03-27	Victorian Infectious Diseases Reference Laboratory (VIDRL)	Microbiological Diagnostic Unit Public Health Laboratory and Victorian Infectious Diseases Reference Laboratory, Doherty Institute	Caly L., Seemann T., Sait, M., Schultz M., Druce J., Sherry, N.
EPI_ISL_426970	hCoV-19/Australia/VIC674/2020	Oceania / Australia / Victoria	2020-03-27	Victorian Infectious Diseases Reference Laboratory (VIDRL)	Microbiological Diagnostic Unit Public Health Laboratory and Victorian Infectious Diseases Reference Laboratory, Doherty Institute	Caly L., Seemann T., Sait, M., Schultz M., Druce J., Sherry, N.
EPI_ISL_426971	hCoV-19/Australia/VIC676/2020	Oceania / Australia / Victoria	2020-03-27	Victorian Infectious Diseases Reference Laboratory (VIDRL)	Microbiological Diagnostic Unit Public Health Laboratory and Victorian Infectious Diseases Reference Laboratory, Doherty Institute	Caly L., Seemann T., Sait, M., Schultz M., Druce J., Sherry, N.
EPI_ISL_426972	hCoV-19/Australia/VIC677/2020	Oceania / Australia / Victoria	2020-03-27	Victorian Infectious Diseases Reference Laboratory (VIDRL)	Microbiological Diagnostic Unit Public Health Laboratory and Victorian Infectious Diseases Reference Laboratory, Doherty Institute	Caly L., Seemann T., Sait, M., Schultz M., Druce J., Sherry, N.
EPI_ISL_426973	hCoV-19/Australia/VIC678/2020	Oceania / Australia / Victoria	2020-03-27	Victorian Infectious Diseases Reference Laboratory (VIDRL)	Microbiological Diagnostic Unit Public Health Laboratory and Victorian Infectious Diseases Reference Laboratory, Doherty Institute	Caly L., Seemann T., Sait, M., Schultz M., Druce J., Sherry, N.
EPI_ISL_426974	hCoV-19/Australia/VIC679/2020	Oceania / Australia / Victoria	2020-03-27	Victorian Infectious Diseases Reference Laboratory (VIDRL)	Microbiological Diagnostic Unit Public Health Laboratory and Victorian Infectious Diseases Reference Laboratory, Doherty Institute	Caly L., Seemann T., Sait, M., Schultz M., Druce J., Sherry, N.
EPI_ISL_426975	hCoV-19/Australia/VIC680/2020	Oceania / Australia / Victoria	2020-03-27	Victorian Infectious Diseases Reference Laboratory (VIDRL)	Microbiological Diagnostic Unit Public Health Laboratory and Victorian Infectious Diseases Reference Laboratory, Doherty Institute	Caly L., Seemann T., Sait, M., Schultz M., Druce J., Sherry, N.
EPI_ISL_426976	hCoV-19/Australia/VIC681/2020	Oceania / Australia / Victoria	2020-03-27	Victorian Infectious Diseases Reference Laboratory (VIDRL)	Microbiological Diagnostic Unit Public Health Laboratory and Victorian Infectious Diseases Reference Laboratory, Doherty Institute	Caly L., Seemann T., Sait, M., Schultz M., Druce J., Sherry, N.
EPI_ISL_426977	hCoV-19/Australia/VIC682/2020	Oceania / Australia / Victoria	2020-03-29	Victorian Infectious Diseases Reference Laboratory (VIDRL)	Microbiological Diagnostic Unit Public Health Laboratory and Victorian Infectious Diseases Reference Laboratory, Doherty Institute	Caly L., Seemann T., Sait, M., Schultz M., Druce J., Sherry, N.
EPI_ISL_426978	hCoV-19/Australia/VIC683/2020	Oceania / Australia / Victoria	2020-03-29	Victorian Infectious Diseases Reference Laboratory (VIDRL)	Microbiological Diagnostic Unit Public Health Laboratory and Victorian Infectious Diseases Reference Laboratory, Doherty Institute	Caly L., Seemann T., Sait, M., Schultz M., Druce J., Sherry, N.
EPI_ISL_426980	hCoV-19/Australia/VIC685/2020	Oceania / Australia / Victoria	2020-03-28	Victorian Infectious Diseases Reference Laboratory (VIDRL)	Microbiological Diagnostic Unit Public Health Laboratory and Victorian Infectious Diseases Reference Laboratory, Doherty Institute	Caly L., Seemann T., Sait, M., Schultz M., Druce J., Sherry, N.
EPI_ISL_426981	hCoV-19/Australia/VIC687/2020	Oceania / Australia / Victoria	2020-03-29	Victorian Infectious Diseases Reference Laboratory (VIDRL)	Microbiological Diagnostic Unit Public Health Laboratory and Victorian Infectious Diseases Reference Laboratory, Doherty Institute	Caly L., Seemann T., Sait, M., Schultz M., Druce J., Sherry, N.
EPI_ISL_426986	hCoV-19/Australia/VIC694/2020	Oceania / Australia / Victoria	2020-03-28	Victorian Infectious Diseases Reference Laboratory (VIDRL)	Microbiological Diagnostic Unit Public Health Laboratory and Victorian Infectious Diseases Reference Laboratory, Doherty Institute	Caly L., Seemann T., Sait, M., Schultz M., Druce J., Sherry, N.
EPI_ISL_426988	hCoV-19/Australia/VIC696/2020	Oceania / Australia / Victoria	2020-03-28	Victorian Infectious Diseases Reference Laboratory (VIDRL)	Microbiological Diagnostic Unit Public Health Laboratory and Victorian Infectious Diseases Reference Laboratory, Doherty Institute	Caly L., Seemann T., Sait, M., Schultz M., Druce J., Sherry, N.
EPI_ISL_426989	hCoV-19/Australia/VIC697/2020	Oceania / Australia / Victoria	2020-03-28	Victorian Infectious Diseases Reference Laboratory (VIDRL)	Microbiological Diagnostic Unit Public Health Laboratory and Victorian Infectious Diseases Reference Laboratory, Doherty Institute	Caly L., Seemann T., Sait, M., Schultz M., Druce J., Sherry, N.
EPI_ISL_426993	hCoV-19/Australia/VIC706/2020	Oceania / Australia / Victoria	2020-03-30	Victorian Infectious Diseases Reference Laboratory (VIDRL)	Microbiological Diagnostic Unit Public Health Laboratory and Victorian Infectious Diseases Reference Laboratory, Doherty Institute	Caly L., Seemann T.,

Sait, M., Schultz M., Druce J., Sherry, N.
EPI_ISL_426994 hCoV-19/Australia/VIC707/2020 Oceania / Australia / Victoria 2020-03-29 Victorian Infectious Diseases Reference Laboratory (VIDRL) Microbiological Diagnostic Unit Public Health Laboratory and Victorian Infectious Diseases Reference Laboratory, Doherty Institute Caly L., Seemann T., Sait, M., Schultz M., Druce J., Sherry, N.
EPI_ISL_426995 hCoV-19/Australia/VIC708/2020 Oceania / Australia / Victoria 2020-03-29 Victorian Infectious Diseases Reference Laboratory (VIDRL) Microbiological Diagnostic Unit Public Health Laboratory and Victorian Infectious Diseases Reference Laboratory, Doherty Institute Caly L., Seemann T., Sait, M., Schultz M., Druce J., Sherry, N.
EPI_ISL_426997 hCoV-19/Australia/VIC710/2020 Oceania / Australia / Victoria 2020-03-29 Victorian Infectious Diseases Reference Laboratory (VIDRL) Microbiological Diagnostic Unit Public Health Laboratory and Victorian Infectious Diseases Reference Laboratory, Doherty Institute Caly L., Seemann T., Sait, M., Schultz M., Druce J., Sherry, N.
EPI_ISL_426999 hCoV-19/Australia/VIC712/2020 Oceania / Australia / Victoria 2020-03-29 Victorian Infectious Diseases Reference Laboratory (VIDRL) Microbiological Diagnostic Unit Public Health Laboratory and Victorian Infectious Diseases Reference Laboratory, Doherty Institute Caly L., Seemann T., Sait, M., Schultz M., Druce J., Sherry, N.
EPI_ISL_429196 hCoV-19/Switzerland/GE3144/2020 Europe / Switzerland 2020-03-18 University Hospitals of Geneva Laboratory of Virology University Hospitals of Geneva Laboratory of Virology Laubscher F.
EPI_ISL_429197 hCoV-19/Switzerland/GE0199-2/2020 Europe / Switzerland 2020-03-21 University Hospitals of Geneva Laboratory of Virology University Hospitals of Geneva Laboratory of Virology Laubscher F.
EPI_ISL_429199 hCoV-19/Switzerland/GE0636/2020 Europe / Switzerland 2020-03-22 University Hospitals of Geneva Laboratory of Virology University Hospitals of Geneva Laboratory of Virology Laubscher F.
EPI_ISL_429200 hCoV-19/Switzerland/GE5737/2020 Europe / Switzerland 2020-03-19 University Hospitals of Geneva Laboratory of Virology University Hospitals of Geneva Laboratory of Virology Laubscher F.
EPI_ISL_429201 hCoV-19/Switzerland/GE2297/2020 Europe / Switzerland 2020-03-29 University Hospitals of Geneva Laboratory of Virology University Hospitals of Geneva Laboratory of Virology Laubscher F.
EPI_ISL_429203 hCoV-19/Switzerland/GE1736/2020 Europe / Switzerland 2020-03-21 University Hospitals of Geneva Laboratory of Virology University Hospitals of Geneva Laboratory of Virology Laubscher F.
EPI_ISL_429204 hCoV-19/Switzerland/GE7617/2020 Europe / Switzerland 2020-04-03 University Hospitals of Geneva Laboratory of Virology University Hospitals of Geneva Laboratory of Virology Laubscher F.
EPI_ISL_429206 hCoV-19/Switzerland/GE5007/2020 Europe / Switzerland 2020-04-07 University Hospitals of Geneva Laboratory of Virology University Hospitals of Geneva Laboratory of Virology Laubscher F.
EPI_ISL_429207 hCoV-19/Switzerland/GE1184/2020 Europe / Switzerland 2020-04-03 University Hospitals of Geneva Laboratory of Virology University Hospitals of Geneva Laboratory of Virology Laubscher F.
EPI_ISL_429208 hCoV-19/Switzerland/GE2164/2020 Europe / Switzerland 2020-03-17 University Hospitals of Geneva Laboratory of Virology University Hospitals of Geneva Laboratory of Virology Laubscher F.
EPI_ISL_429209 hCoV-19/Switzerland/GE3650/2020 Europe / Switzerland 2020-03-16 University Hospitals of Geneva Laboratory of Virology University Hospitals of Geneva Laboratory of Virology Laubscher F.
EPI_ISL_429210 hCoV-19/Switzerland/GE8147/2020 Europe / Switzerland 2020-03-16 University Hospitals of Geneva Laboratory of Virology University Hospitals of Geneva Laboratory of Virology Laubscher F.
EPI_ISL_429211 hCoV-19/Switzerland/GE5708/2020 Europe / Switzerland 2020-03-24 University Hospitals of Geneva Laboratory of Virology University Hospitals of Geneva Laboratory of Virology Laubscher F.
EPI_ISL_429212 hCoV-19/Switzerland/GE0056/2020 Europe / Switzerland 2020-03-24 University Hospitals of Geneva Laboratory of Virology University Hospitals of Geneva Laboratory of Virology Laubscher F.
EPI_ISL_429213 hCoV-19/Switzerland/GE8086/2020 Europe / Switzerland 2020-03-16 University Hospitals of Geneva Laboratory of Virology University Hospitals of Geneva Laboratory of Virology Laubscher F.
EPI_ISL_429214 hCoV-19/Switzerland/GE2759/2020 Europe / Switzerland 2020-03-23 University Hospitals of Geneva Laboratory of Virology University Hospitals of Geneva Laboratory of Virology Laubscher F.
EPI_ISL_429215 hCoV-19/Switzerland/GE4807/2020 Europe / Switzerland 2020-03-23 University Hospitals of Geneva Laboratory of Virology University Hospitals of Geneva Laboratory of Virology Laubscher F.
EPI_ISL_429217 hCoV-19/Switzerland/GE6414/2020 Europe / Switzerland 2020-04-06 University Hospitals of Geneva Laboratory of Virology University Hospitals of Geneva Laboratory of Virology Laubscher F.
EPI_ISL_429218 hCoV-19/Switzerland/GE0304/2020 Europe / Switzerland 2020-04-02 University Hospitals of Geneva Laboratory of Virology University Hospitals of Geneva Laboratory of Virology Laubscher F.
EPI_ISL_429219 hCoV-19/Switzerland/GE2453/2020 Europe / Switzerland 2020-04-03 University Hospitals of Geneva Laboratory of Virology University Hospitals of Geneva Laboratory of Virology Laubscher F.
EPI_ISL_429220 hCoV-19/Switzerland/GE6065/2020 Europe / Switzerland 2020-04-06 University Hospitals of Geneva Laboratory of Virology University Hospitals of Geneva Laboratory of Virology Laubscher F.
EPI_ISL_429221 hCoV-19/Switzerland/GE6099/2020 Europe / Switzerland 2020-04-06 University Hospitals of Geneva Laboratory of Virology University Hospitals of Geneva Laboratory of Virology Laubscher F.
EPI_ISL_429222 hCoV-19/Switzerland/GE3655/2020 Europe / Switzerland 2020-04-07 University Hospitals of Geneva Laboratory of Virology University Hospitals of Geneva Laboratory of Virology Laubscher F.
EPI_ISL_429223 hCoV-19/Switzerland/GE4644/2020 Europe / Switzerland 2020-04-07 University Hospitals of Geneva Laboratory of Virology University Hospitals of Geneva Laboratory of Virology Laubscher F.
EPI_ISL_429226 hCoV-19/Italy/TE5551/2020 Europe / Italy / Abruzzo 2020-03-17 Presidio Ospedaliero Santo Spirito "Istituto Zooprofilattico Sperimentale dell'Abruzzo e Molise ""G. Caporale""" Lorusso A, Marcacci M, Di Domenico M, Ancora M, Curini V, Mangone I, Rinaldi A, Di Pasquale A, Camma C, Puglia I, Savini G

EPI_ISL_429227  hCoV-19/Italy/TE5553/2020       Europe / Italy / Abruzzo        2020-03-17      Presidio Ospedaliero Santo Spirito       "Istituto Zooprofilattico Sperimentale dell'Abruzzo e Molise ""G. Caporale""" Lorusso A, Marcacci M, Di Domenico M, Ancora M, Curini V, Mangone I, Rinaldi A, Di Pasquale A, Camma C, Puglia I, Savini G
EPI_ISL_429228  hCoV-19/Italy/TE5674/2020       Europe / Italy / Abruzzo        2020-03-20      Ospedale Civile Giuseppe Mazzini        "Istituto Zooprofilattico Sperimentale dell'Abruzzo e Molise ""G. Caporale"""   Lorusso A, Marcacci M, Di Domenico M, Ancora M, Curini V, Mangone I, Rinaldi A, Di Pasquale A, Camma C, Puglia I, Savini G
EPI_ISL_430179  hCoV-19/USA/WA-S142/2020        North America / USA / Washington / King County  2020-03-13 Washington State Department of Health   Seattle Flu Study       Chu et al
EPI_ISL_430180  hCoV-19/USA/WA-S143/2020        North America / USA / Washington / King County  2020-03-15 Washington State Department of Health   Seattle Flu Study       Chu et al
EPI_ISL_430181  hCoV-19/USA/WA-S144/2020        North America / USA / Washington / King County  2020-03-15 Washington State Department of Health   Seattle Flu Study       Chu et al
EPI_ISL_430182  hCoV-19/USA/WA-S145/2020        North America / USA / Washington / King County  2020-03-15 Washington State Department of Health   Seattle Flu Study       Chu et al
EPI_ISL_430183  hCoV-19/USA/WA-S146/2020        North America / USA / Washington / King County  2020-03-14 Washington State Department of Health   Seattle Flu Study       Chu et al
EPI_ISL_430184  hCoV-19/USA/WA-S147/2020        North America / USA / Washington / King County  2020-03-14 Washington State Department of Health   Seattle Flu Study       Chu et al
EPI_ISL_430185  hCoV-19/USA/WA-S148/2020        North America / USA / Washington / King County  2020-03-13 Washington State Department of Health   Seattle Flu Study       Chu et al
EPI_ISL_430186  hCoV-19/USA/WA-S149/2020        North America / USA / Washington / King County  2020-03-14 Washington State Department of Health   Seattle Flu Study       Chu et al
EPI_ISL_430187  hCoV-19/USA/WA-S150/2020        North America / USA / Washington / King County  2020-03-14 Washington State Department of Health   Seattle Flu Study       Chu et al
EPI_ISL_430188  hCoV-19/USA/WA-S151/2020        North America / USA / Washington / Snohomish County     2020-03-16   Washington State Department of Health   Seattle Flu Study       Chu et al
EPI_ISL_430189  hCoV-19/USA/WA-S152/2020        North America / USA / Washington / King County  2020-03-16 Washington State Department of Health   Seattle Flu Study       Chu et al
EPI_ISL_430190  hCoV-19/USA/WA-S153/2020        North America / USA / Washington / King County  2020-03-16 Washington State Department of Health   Seattle Flu Study       Chu et al
EPI_ISL_430191  hCoV-19/USA/WA-S154/2020        North America / USA / Washington / King County  2020-03-16 Washington State Department of Health   Seattle Flu Study       Chu et al
EPI_ISL_430193  hCoV-19/USA/WA-S156/2020        North America / USA / Washington / King County  2020-03-16 Washington State Department of Health   Seattle Flu Study       Chu et al
EPI_ISL_430196  hCoV-19/USA/WA-S158/2020        North America / USA / Washington / Snohomish County     2020-03-16   Washington State Department of Health   Seattle Flu Study       Chu et al
EPI_ISL_430197  hCoV-19/USA/WA-S159/2020        North America / USA / Washington / Snohomish County     2020-03-16   Washington State Department of Health   Seattle Flu Study       Chu et al
EPI_ISL_430198  hCoV-19/USA/WA-S160/2020        North America / USA / Washington / King County  2020-03-17 Washington State Department of Health   Seattle Flu Study       Chu et al
EPI_ISL_430199  hCoV-19/USA/WA-S161/2020        North America / USA / Washington / King County  2020-03-17 Washington State Department of Health   Seattle Flu Study       Chu et al
EPI_ISL_430200  hCoV-19/USA/WA-S162/2020        North America / USA / Washington        2020-03-04 Washington State Department of Health   Seattle Flu Study       Chu et al
EPI_ISL_430201  hCoV-19/USA/WA-S163/2020        North America / USA / Washington        2020-03-04 Washington State Department of Health   Seattle Flu Study       Chu et al
EPI_ISL_430202  hCoV-19/USA/WA-S164/2020        North America / USA / Washington        2020-03-02 Washington State Department of Health   Seattle Flu Study       Chu et al
EPI_ISL_430204  hCoV-19/USA/WA-S166/2020        North America / USA / Washington / King County  2020-03-18 Washington State Department of Health   Seattle Flu Study       Chu et al
EPI_ISL_430205  hCoV-19/USA/WA-S167/2020        North America / USA / Washington / Whatcom County       2020-03-18   Washington State Department of Health   Seattle Flu Study       Chu et al
EPI_ISL_430206  hCoV-19/USA/WA-S168/2020        North America / USA / Washington / Whatcom County       2020-03-18   Washington State Department of Health   Seattle Flu Study       Chu et al
EPI_ISL_430207  hCoV-19/USA/WA-S169/2020        North America / USA / Washington / Whatcom County       2020-03-18   Washington State Department of Health   Seattle Flu Study       Chu et al
EPI_ISL_430208  hCoV-19/USA/WA-S170/2020        North America / USA / Washington / Yakima County        2020-03-17   Washington State Department of Health   Seattle Flu Study       Chu et al
EPI_ISL_430209  hCoV-19/USA/WA-S171/2020        North America / USA / Washington        2020-03-19 Washington State Department of Health   Seattle Flu Study       Chu et al
EPI_ISL_430210  hCoV-19/USA/WA-S172/2020        North America / USA / Washington / King County  2020-03-19 Washington State Department of Health   Seattle Flu Study       Chu et al
EPI_ISL_430211  hCoV-19/USA/WA-S173/2020        North America / USA / Washington        2020-03-19 Washington State Department of Health   Seattle Flu Study       Chu et al
EPI_ISL_404227  hCoV-19/Zhejiang/WZ-01/2020     Asia / China / Zhejiang 2020-01-16      Zhejiang Provincial Center for Disease Control and Prevention       Department of Microbiology, Zhejiang Provincial Center for Disease Control and Prevention  Yin Chen, Yanjun Zhang, Haiyan Mao, Junhang Pan, Xiuyu Lou, Yiyu Lu, Juying Yan, Hanping Zhu, Jian Gao, Yan Feng, Yi Sun, Hao Yan, Zhen Li, Yisheng Sun, Liming Gong, Qiong Ge, Wen Shi,

Xinying Wang, Wenwu Yao, Zhangnv Yang, Fang Xu, Chen Chen, Enfu Chen, Zhen Wang, Zhiping Chen, Jianmin Jiang, Chonggao Hu
EPI_ISL_404228 hCoV-19/Zhejiang/WZ-02/2020 Asia / China / Zhejiang 2020-01-17 Zhejiang Provincial Center for Disease Control and Prevention Department of Microbiology, Zhejiang Provincial Center for Disease Control and Prevention Yanjun Zhang, Yin Chen, Haiyan Mao, Junhang Pan, Xiuyu Lou, Yiyu Lu, Juying Yan, Hanping Zhu, Jian Gao, Yan Feng, Yi Sun, Hao Yan, Zhen Li, Yisheng Sun, Liming Gong, Qiong Ge, Wen Shi, Xinying Wang, Wenwu Yao, Zhangnv Yang, Fang Xu, Chen Chen, Enfu Chen, Zhen Wang, Zhiping Chen, Jianmin Jiang, Chonggao Hu
EPI_ISL_404253 hCoV-19/USA/IL1/2020 North America / USA / Illinois / Chicago 2020-01-21 IL Department of Public Health Chicago Laboratory Pathogen Discovery, Respiratory Viruses Branch, Division of Viral Diseases, Centers for Dieases Control and Prevention Ying Tao, Krista Queen, Clinton R. Paden, Jing Zhang, Yan Li, Anna Uehara, Xiaoyan Lu, Brian Lynch, Senthil Kumar K. Sakthivel, Brett L. Whitaker, Shifaq Kamili, Lijuan Wang, Janna' R. Murray, Susan I. Gerber, Stephen Lindstrom, Suxiang Tong
EPI_ISL_406800 hCoV-19/Wuhan/WH03/2020 Asia / China / Hubei / Wuhan 2020-01-01 General Hospital of Central Theater Command of People's Liberation Army of China BGI & Institute of Microbiology, Chinese Academy of Sciences & Shandong First Medical University & Shandong Academy of Medical Sciences & General Hospital of Central Theater Command of People's Liberation Army of China Weijun Chen, Yuhai Bi, Weifeng Shi and Zhenhong Hu
EPI_ISL_406801 hCoV-19/Wuhan/WH04/2020 Asia / China / Hubei / Wuhan 2020-01-05 General Hospital of Central Theater Command of People's Liberation Army of China BGI & Institute of Microbiology, Chinese Academy of Sciences & Shandong First Medical University & Shandong Academy of Medical Sciences & General Hospital of Central Theater Command of People's Liberation Army of China Weijun Chen, Yuhai Bi, Weifeng Shi and Zhenhong Hu
EPI_ISL_408976 hCoV-19/Australia/NSW02/2020 Oceania / Australia / New South Wales / Sydney 2020-01-22 Centre for Infectious Diseases and Microbiology Laboratory Services NSW Health Pathology - Institute of Clinical Pathology and Medical Research; Westmead Hospital; University of Sydney Rockett R, Sadsad R, Eden J-S, Carter I, Rahman H, Holmes EC, O’Sullivan MV, Sintchenko V, Chen SC, Maddocks S, Kok J and Dwyer DE for the 2019-nCoV Study Group*
EPI_ISL_408977 hCoV-19/Australia/NSW03/2020 Oceania / Australia / New South Wales / Sydney 2020-01-25 Serology, Virology and OTDS Laboratories (SAViD), NSW Health Pathology Randwick NSW Health Pathology - Institute of Clinical Pathology and Medical Research; Centre for Infectious Diseases and Microbiology Laboratory Services; Westmead Hospital; University of Sydney Eden J-S, Carter I, Rahman H, Rawlinson W, Holmes EC, Rockett R, O’Sullivan MV, Sintchenko V, Chen SC, Maddocks S, Kok J and Dwyer DE for the 2019-nCoV Study Group*
EPI_ISL_412116 hCoV-19/England/09c/2020 Europe / United Kingdom / England 2020-02-09 Respiratory Virus Unit, Microbiology Services Colindale, Public Health England Respiratory Virus Unit, Microbiology Services Colindale, Public Health England Monica Galiano, Shahjahan Miah, Angie Lackenby, Omolola Akinbami, Tiina Talts, Leena Bhaw, Richard Myers, Steven Platt, Kirstin Edwards, Jonathan Hubb, Joanna Ellis, Maria Zambon
EPI_ISL_413455 hCoV-19/USA/WA4-UW2/2020 North America / USA / Washington 2020-02-28 Washington State Public Health Lab University of Washington Virology Lab Pavitra Roychoudhury, Arun Nalla, Hong Xie, Keith Jerome, Alexander Greninger
EPI_ISL_413456 hCoV-19/USA/WA-S2/2020 North America / USA / Washington / King County 2020-02-20 Seattle Flu Study Seattle Flu Study Chu et al
EPI_ISL_413457 hCoV-19/USA/WA6-UW3/2020 North America / USA / Washington 2020-02-29 Washington State Public Health Lab UW Virology Lab Pavitra Roychoudhury, Arun Nalla, Hong Xie, Keith Jerome, Alexander Greninger
EPI_ISL_413458 hCoV-19/USA/WA7-UW4/2020 North America / USA / Washington 2020-03-01 Washington State Public Health Lab UW Virology Lab Pavitra Roychoudhury, Arun Nalla, Hong Xie, Keith Jerome, Alexander Greninger
EPI_ISL_413459 hCoV-19/Japan/TK-20-31-3/2020 Asia / Japan / Tokyo 2020-02-20 Department of Pathology, Toshima Hospital Pathogen Genomics Center, National Institute of Infectious Diseases Tsuyoshi Sekizuka, Kentaro Itokawa, Takuya Adachi, Masahiro Sano, Jun Yamazaki, Ippei Miyamoto, Haruka Nishioka, Ja-Mun Chong, Noriko Nakajima, Yuko Sato, Minoru Tobiume, Harutaka Katano, Tadaki Suzuki, Makoto Kuroda
EPI_ISL_413486 hCoV-19/USA/WA8-UW5/2020 North America / USA / Washington 2020-03-01 Valley Medical Center University of Washington Virology Lab Pavitra Roychoudhury, Arun Nalla, Hong Xie, Keith Jerome, Alexander Greninger
EPI_ISL_413487 hCoV-19/USA/WA9-UW6/2020 North America / USA / Washington 2020-03-01 Harborview Medical Center University of Washington Virology Lab Pavitra Roychoudhury, Arun Nalla, Hong Xie, Keith Jerome, Alexander Greninger
EPI_ISL_413488 hCoV-19/Germany/NRW-01/2020 Europe / Germany / North Rhine Westphalia / Heinsberg District 2020-02-28 Center of Medical Microbiology, Virology, and Hospital Hygiene, University of Duesseldorf Center of Medical Microbiology, Virology, and Hospital Hygiene, University of Duesseldorf Ortwin Adams, Marcel Andree, Alexander Dilthey, Torsten Feldt, Sandra Hauka, Torsten Houwaart, Björn-Erik Jensen, Detlef Kindgen-Milles, Malte Kohns Vasconcelos, Klaus Pfeffer, Tina Senff, Daniel Strelow, Jörg Timm, Andreas Walker, Tobias Wienemann
EPI_ISL_413489 hCoV-19/Italy/UniSR1/2020 Europe / Italy / Lombardy / Milan 2020-03-03 Laboratorio di Microbiologia e Virologia, Università Vita-Salute San Raffaele, Milano Laboratorio di Microbiologia e Virologia, Università Vita-Salute San Raffaele, Milano R.A Diotti, E. Criscuolo, M. Castelli,

V. Caputo, R. Ferrarese, M. Sampaolo, E. Boeri, I. Negri, V. Amato, G. Lo Raso, C. Di Resta, R. Burioni, M. Clementi, N. Mancini & N. Clementi
EPI_ISL_413490 hCoV-19/New Zealand/01/2020 Oceania / New Zealand / Auckland 2020-02-27 Auckland Hospital Institute of Environmental Science and Research (ESR) Matt Storey, Xiaoyun Ren, Gary McAuliffe, Sally Roberts, Matthew Blakiston, Erasmus Smit, Lauren Jelly, Joep de Ligt
EPI_ISL_414616 hCoV-19/USA/WA-UW29/2020 North America / USA / Washington / Kirkland 2020-03-08 UW Virology Lab UW Virology Lab Pavitra Roychoudhury, Hong Xie, Keith Jerome, Alexander Greninger
EPI_ISL_414617 hCoV-19/USA/WA-UW30/2020 North America / USA 2020-03-08 UW Virology Lab UW Virology Lab Pavitra Roychoudhury, Hong Xie, Keith Jerome, Alexander Greninger
EPI_ISL_414618 hCoV-19/USA/WA-UW31/2020 North America / USA 2020-03-08 UW Virology Lab UW Virology Lab Pavitra Roychoudhury, Hong Xie, Keith Jerome, Alexander Greninger
EPI_ISL_414619 hCoV-19/USA/WA-UW32/2020 North America / USA 2020-03-07 UW Virology Lab UW Virology Lab Pavitra Roychoudhury, Hong Xie, Keith Jerome, Alexander Greninger
EPI_ISL_414621 hCoV-19/USA/WA-UW34/2020 North America / USA / Washington 2020-03-07 UW Virology Lab UW Virology Lab Pavitra Roychoudhury, Hong Xie, Keith Jerome, Alexander Greninger
EPI_ISL_414622 hCoV-19/USA/WA-UW35/2020 North America / USA / Washington / Seattle 2020-03-08 UW Virology Lab UW Virology Lab Pavitra Roychoudhury, Hong Xie, Keith Jerome, Alexander Greninger
EPI_ISL_414623 hCoV-19/France/GE1583/2020 Europe / France / Grand-Est / Strasbourg 2020-02-25 Laboratoire de Virologie Institut de Virologie - INSERM U 1109 Hôpitaux Universitaires de Strasbourg National Reference Center for Viruses of Respiratory Infections, Institut Pasteur, Paris Mélnie Albert, Marion Barbet, Sylvie Behillil, Méline Bizard, Angela Brisebarre, Flora Donati Vincent Enouf, Maud Vanpeene, Sylvie van der Werf, Samira Fafi-Kremer
EPI_ISL_414624 hCoV-19/France/N1620/2020 Europe / France / Normandie / Rouen 2020-02-26 Centre Hositalier Universitaire de Rouen Laboratoire de Virologie National Reference Center for Viruses of Respiratory Infections, Institut Pasteur, Paris Mélnie Albert, Marion Barbet, Sylvie Behillil, Méline Bizard, Angela Brisebarre, Flora Donati Vincent Enouf, Maud Vanpeene, Sylvie van der Werf, Jean-Christophe Plantier
EPI_ISL_414636 hCoV-19/France/HF1989/2020 Europe / France / Hauts de France / Compiègne 2020-03-04 Centre Hospitalier Compiègne Laboratoire de Biologie National Reference Center for Viruses of Respiratory Infections, Institut Pasteur, Paris Mélnie Albert, Marion Barbet, Sylvie Behillil, Méline Bizard, Angela Brisebarre, Flora Donati Vincent Enouf, Maud Vanpeene, Sylvie van der Werf, Raulin Olivia
EPI_ISL_414637 hCoV-19/France/HF1993/2020 Europe / France / Hauts de France / Compiègne 2020-03-04 Centre Hospitalier Compiègne Laboratoire de Biologie National Reference Center for Viruses of Respiratory Infections, Institut Pasteur, Paris Mélnie Albert, Marion Barbet, Sylvie Behillil, Méline Bizard, Angela Brisebarre, Flora Donati Vincent Enouf, Maud Vanpeene, Sylvie van der Werf, Raulin Olivia
EPI_ISL_414638 hCoV-19/France/HF1995/2020 Europe / France / Hauts de France / Compiègne 2020-03-04 Centre Hospitalier Compiègne Laboratoire de Biologie National Reference Center for Viruses of Respiratory Infections, Institut Pasteur, Paris Mélnie Albert, Marion Barbet, Sylvie Behillil, Méline Bizard, Angela Brisebarre, Flora Donati Vincent Enouf, Maud Vanpeene, Sylvie van der Werf, Raulin Olivia
EPI_ISL_414641 hCoV-19/Finland/FIN-313/2020 Europe / Finland 2020-03-05 Department of Virology and Immunology, University of Helsinki and Helsinki University Hospital, Huslab Finland Department of Virology, Faculty of Medicine, University of Helsinki, Helsinki, Finland Teemu Smura, Hannimari Kallio-Kokko, Olli Vapalahti
EPI_ISL_414642 hCoV-19/Finland/FIN-455/2020 Europe / Finland 2020-03-08 Department of Virology and Immunology, University of Helsinki and Helsinki University Hospital, Huslab Finland Department of Virology, Faculty of Medicine, University of Helsinki, Helsinki, Finland Teemu Smura, Hannimari Kallio-Kokko, Olli Vapalahti
EPI_ISL_414643 hCoV-19/Finland/FIN-508/2020 Europe / Finland 2020-03-07 Department of Virology and Immunology, University of Helsinki and Helsinki University Hospital, Huslab Finland Department of Virology, Faculty of Medicine, University of Helsinki, Helsinki, Finland Teemu Smura, Hannimari Kallio-Kokko, Olli Vapalahti
EPI_ISL_414646 hCoV-19/Finland/FIN-266/2020 Europe / Finland 2020-03-04 Department of Virology and Immunology, University of Helsinki and Helsinki University Hospital, Huslab Finland Department of Virology, Faculty of Medicine, University of Helsinki, Helsinki, Finland Teemu Smura, Hannimari Kallio-Kokko, Olli Vapalahti
EPI_ISL_414648 hCoV-19/USA/CA-PC101P/2020 North America / USA / California / San Diego County 2020-03-11 Andersen Lab, The Scripps Research Institute Andersen Lab, The Scripps Research Institute Mark Zeller, Catie Anderson, Emily Spender, Sarah Topol, Raphaelle Klitting, Refugio Robles-Sikisaka, Karthik Gangavarapu, Laura Nicholson, Kristian Andersen
EPI_ISL_417004 hCoV-19/Belgium/ULG-3000/2020 Europe / Belgium / Liège 2020-03-05 Department of Clinical Microbiology GIGA Medical Genomics Durkin Keith, Artesi Maria, Bontems Sébastien, Boreux Raphaël, Meex Cécile, Melin Pierrette, Hayette Marie-Pierre, Bours Vincent.
EPI_ISL_417006 hCoV-19/Belgium/ULG-3163/2020 Europe / Belgium / Liège 2020-03-05 Department of Clinical Microbiology GIGA Medical Genomics Durkin Keith, Artesi Maria, Bontems Sébastien, Boreux Raphaël, Meex Cécile, Melin Pierrette, Hayette Marie-Pierre, Bours Vincent.
EPI_ISL_417007 hCoV-19/Spain/Galicia201663/2020 Europe / Spain / Galicia 2020-03-07 HOSPITAL SANTA MARIA NAI Instituto de Salud Carlos III Iglesias-Caballero, M. Molinero Calamita, M. González-Esguevillas, M. Camarero S. Pozo F. Casas I. Jiménez, P. Jiménez, M. Zaballos, A. Monzón, S. Varona, S. Juliá, M. Cuesta, I. García Costa, J.
EPI_ISL_417008 hCoV-19/Belgium/ULG-3662/2020 Europe / Belgium / Liège 2020-03-07 Department of Clinical Microbiology GIGA Medical Genomics Durkin Keith, Artesi Maria, Bontems Sébastien, Boreux Raphaël,

Meex Cécile, Melin Pierrette, Hayette Marie-Pierre, Bours Vincent.
EPI_ISL_417009 hCoV-19/Belgium/ULG-3665/2020 Europe / Belgium / Liège 2020-03-07 Department of Clinical Microbiology GIGA Medical Genomics Durkin Keith, Artesi Maria, Bontems Sébastien, Boreux Raphaël, Meex Cécile, Melin Pierrette, Hayette Marie-Pierre, Bours Vincent.
EPI_ISL_417010 hCoV-19/Spain/Madrid201442/2020 Europe / Spain / Madrid 2020-03-04 FUNDACION JIMENEZ DIAZ Instituto de Salud Carlos III Iglesias-Caballero, M. Molinero Calamita, M. González-Esguevillas, M. Camarero, S. Pozo, F. Casas, I. Jiménez, P. Jiménez, M. Zaballos, A. Monzón, S. Varona, S. Juliá, M. Cuesta, I. Fernández Roblas, R.
EPI_ISL_417012 hCoV-19/Belgium/ULG-3843/2020 Europe / Belgium / Liège 2020-03-08 Department of Clinical Microbiology GIGA Medical Genomics Durkin Keith, Artesi Maria, Bontems Sébastien, Boreux Raphaël, Meex Cécile, Melin Pierrette, Hayette Marie-Pierre, Bours Vincent.
EPI_ISL_417013 hCoV-19/Belgium/ULG-4163/2020 Europe / Belgium / Liège 2020-03-09 Department of Clinical Microbiology GIGA Medical Genomics Durkin Keith, Artesi Maria, Bontems Sébastien, Boreux Raphaël, Meex Cécile, Melin Pierrette, Hayette Marie-Pierre, Bours Vincent.
EPI_ISL_417014 hCoV-19/Belgium/ULG-6216/2020 Europe / Belgium / Liège 2020-03-13 Department of Clinical Microbiology GIGA Medical Genomics Durkin Keith, Artesi Maria, Bontems Sébastien, Boreux Raphaël, Meex Cécile, Melin Pierrette, Hayette Marie-Pierre, Bours Vincent.
EPI_ISL_417015 hCoV-19/Belgium/ULG-6457/2020 Europe / Belgium / Liège 2020-03-13 Department of Clinical Microbiology GIGA Medical Genomics Durkin Keith, Artesi Maria, Bontems Sébastien, Boreux Raphaël, Meex Cécile, Melin Pierrette, Hayette Marie-Pierre, Bours Vincent.
EPI_ISL_417016 hCoV-19/Belgium/ULG-6503/2020 Europe / Belgium / Liège 2020-03-13 Department of Clinical Microbiology GIGA Medical Genomics Durkin Keith, Artesi Maria, Bontems Sébastien, Boreux Raphaël, Meex Cécile, Melin Pierrette, Hayette Marie-Pierre, Bours Vincent.
EPI_ISL_417017 hCoV-19/Belgium/ULG-6638/2020 Europe / Belgium / Liège 2020-03-14 Department of Clinical Microbiology GIGA Medical Genomics Durkin Keith, Artesi Maria, Bontems Sébastien, Boreux Raphaël, Meex Cécile, Melin Pierrette, Hayette Marie-Pierre, Bours Vincent.
EPI_ISL_417018 hCoV-19/Belgium/ULG-6670/2020 Europe / Belgium / Liège 2020-03-14 Department of Clinical Microbiology GIGA Medical Genomics Durkin Keith, Artesi Maria, Bontems Sébastien, Boreux Raphaël, Meex Cécile, Melin Pierrette, Hayette Marie-Pierre, Bours Vincent.
EPI_ISL_417019 hCoV-19/Belgium/ULG-6754/2020 Europe / Belgium / Liege 2020-03-14 Department of Clinical Microbiology GIGA Medical Genomics Durkin Keith, Artesi Maria, Bontems Sébastien, Boreux Raphaël, Meex Cécile, Melin Pierrette, Hayette Marie-Pierre, Bours Vincent.
EPI_ISL_417020 hCoV-19/Belgium/ULG-6939/2020 Europe / Belgium / Liège 2020-03-15 Department of Clinical Microbiology GIGA Medical Genomics Durkin Keith, Artesi Maria, Bontems Sébastien, Boreux Raphaël, Meex Cécile, Melin Pierrette, Hayette Marie-Pierre, Bours Vincent.
EPI_ISL_417021 hCoV-19/Belgium/ULG-6942/2020 Europe / Belgium / Liège 2020-03-15 Department of Clinical Microbiology GIGA Medical Genomics Durkin Keith, Artesi Maria, Bontems Sébastien, Boreux Raphaël, Meex Cécile, Melin Pierrette, Hayette Marie-Pierre, Bours Vincent.
EPI_ISL_417022 hCoV-19/Belgium/ULG-6948/2020 Europe / Belgium / Liège 2020-03-15 Department of Clinical Microbiology GIGA Medical Genomics Durkin Keith, Artesi Maria, Bontems Sébastien, Boreux Raphaël, Meex Cécile, Melin Pierrette, Hayette Marie-Pierre, Bours Vincent.
EPI_ISL_417023 hCoV-19/Belgium/ULG-6950/2020 Europe / Belgium / Liège 2020-03-15 Department of Clinical Microbiology GIGA Medical Genomics Durkin Keith, Artesi Maria, Bontems Sébastien, Boreux Raphaël, Meex Cécile, Melin Pierrette, Hayette Marie-Pierre, Bours Vincent.
EPI_ISL_417025 hCoV-19/Belgium/ULG-7019/2020 Europe / Belgium / Liège 2020-03-15 Department of Clinical Microbiology GIGA Medical Genomics Durkin Keith, Artesi Maria, Bontems Sébastien, Boreux Raphaël, Meex Cécile, Melin Pierrette, Hayette Marie-Pierre, Bours Vincent.
EPI_ISL_417026 hCoV-19/USA/UT-08/2020 North America / USA / Utah 2020-03-20 Utah Public Health Laboratory Utah Public Health Laboratory Erin Young, Kelly Oakeson
EPI_ISL_417028 hCoV-19/USA/UT-020/2020 North America / USA / Utah 2020-03-20 Utah Public Health Laboratory Utah Public Health Laboratory Erin Young, Kelly Oakeson
EPI_ISL_417030 hCoV-19/Australia/NSW04/2020 Oceania / Australia / New South Wales / Sydney 2020-01-24 Centre for Infectious Diseases and Microbiology Laboratory Services NSW Health Pathology - Institute of Clinical Pathology and Medical Research; Westmead Hospital; University of Sydney Eden J-S, Rockett R, Carter I, Rahman H, Holmes EC, O'Sullivan MV, Sintchenko V, Chen SC, Maddocks S, Kok J and Dwyer DE for the 2019-nCoV Study Group*
EPI_ISL_417031 hCoV-19/Australia/QLDID919/2020 Oceania / Australia / Queensland / Gold Coast 2020-03-11 Pathology Queensland Public Health Virology Laboratory Bixing Huang, Alyssa Pyke, Amanda De Jong, Andrew Van Den Hurk, Carmel Taylor, David Warrilow, Doris Genge, Elisabeth Gamez, Glen Hewitson, Ian Maxwell Mackay, Inga Sultana, Jamie McMahon, Jean Barcelon, Judy Northill, Mitchell Finger, Natalie Simpson, Neelima Nair, Peter Burtonclay, Peter Moore, Sarah Wheatley, Sean Moody, Sonja Hall-Mendelin, Timothy Gardam, and Frederick Moore
EPI_ISL_417032 hCoV-19/Australia/QLDID920/2020 Oceania / Australia / Queensland / Rockhampton 2020-03-11 Rockhampton Base Hospital Public Health Virology Laboratory Bixing Huang, Alyssa Pyke, Amanda De Jong, Andrew Van Den Hurk, Carmel Taylor, David Warrilow, Doris Genge, Elisabeth Gamez, Glen Hewitson, Ian Maxwell Mackay, Inga Sultana, Jamie McMahon, Jean Barcelon, Judy Northill, Mitchell Finger, Natalie Simpson, Neelima Nair, Peter Burtonclay, Peter Moore, Sarah Wheatley, Sean Moody, Sonja Hall-Mendelin, Timothy Gardam, and Frederick Moore
EPI_ISL_417033 hCoV-19/Australia/QLDID921/2020 Oceania / Australia / Queensland / Brisbane 2020-03-11 Sullivan Nicolaides Pathology Public Health Virology Laboratory Bixing Huang, Alyssa Pyke, Amanda De

Jong, Andrew Van Den Hurk, Carmel Taylor, David Warrilow, Doris Genge, Elisabeth Gamez, Glen Hewitson, Ian Maxwell Mackay, Inga Sultana, Jamie McMahon, Jean Barcelon, Judy Northill, Mitchell Finger, Natalie Simpson, Neelima Nair, Peter Burtonclay, Peter Moore, Sarah Wheatley, Sean Moody, Sonja Hall-Mendelin, Timothy Gardam, and Frederick Moore
EPI_ISL_417034 hCoV-19/Brazil/AMBR-02/2020 South America / Brazil / Amazonas State / Manaus 2020-03-16 Laboratorio de Ecologia de Doencas Transmissiveis na Amazonia, Instituto Leonidas e Maria Deane - Fiocruz Amazonia Laboratorio de Ecologia de Doencas Transmissiveis na Amazonia, Instituto Leonidas e Maria Deane - Fiocruz Amazonia Valdinete Nascimento, André Corado, Fernanda Nascimento, Ágatha Costa, Debora Duarte, Luciana Gonçalves, Michele Jesus, Sérgio Luz, Felipe Naveca
EPI_ISL_417064 hCoV-19/Hong Kong/VB20017970-2/2020 Asia / Hong Kong 2020-01-21 Prince of Wales Hospital Hong Kong Department of Health Alan K.L. Tsang, Peter C.W. Yip, Edman T.K. Lam, Rickjason C.W. Chan, Dominic N.C. Tsang
EPI_ISL_417065 hCoV-19/USA/WA-S12/2020 North America / USA / Washington 2020-03-03 Washington State Department of Health Seattle Flu Study Chu etl al
EPI_ISL_417066 hCoV-19/USA/WA-S13/2020 North America / USA / Washington / King County 2020-03-03 Washington State Department of Health Seattle Flu Study Chu etl al
EPI_ISL_417068 hCoV-19/USA/WA-S15/2020 North America / USA / Washington / King County 2020-03-02 Washington State Department of Health Seattle Flu Study Chu etl al
EPI_ISL_417069 hCoV-19/USA/WA-S16/2020 North America / USA / Washington 2020-03-03 Washington State Department of Health Seattle Flu Study Chu etl al
EPI_ISL_417070 hCoV-19/USA/WA-S17/2020 North America / USA / Washington / King County 2020-03-03 Washington State Department of Health Seattle Flu Study Chu etl al
EPI_ISL_417071 hCoV-19/USA/WA-S18/2020 North America / USA / Washington / King County 2020-03-03 Washington State Department of Health Seattle Flu Study Chu etl al
EPI_ISL_417072 hCoV-19/USA/WA-S19/2020 North America / USA / Washington / King County 2020-03-02 Washington State Department of Health Seattle Flu Study Chu etl al
EPI_ISL_417073 hCoV-19/USA/WA-S20/2020 North America / USA / Washington 2020-03-02 Washington State Department of Health Seattle Flu Study Chu etl al
EPI_ISL_417074 hCoV-19/USA/WA-S21/2020 North America / USA / Washington / King County 2020-03-02 Washington State Department of Health Seattle Flu Study Chu etl al
EPI_ISL_417075 hCoV-19/USA/WA-S22/2020 North America / USA / Washington / Snohomish County 2020-03-02 Washington State Department of Health Seattle Flu Study Chu etl al
EPI_ISL_417076 hCoV-19/USA/WA-S23/2020 North America / USA / Washington / Snohomish County 2020-03-02 Washington State Department of Health Seattle Flu Study Chu etl al
EPI_ISL_417077 hCoV-19/USA/WA-S24/2020 North America / USA / Washington / Snohomish County 2020-03-02 Washington State Department of Health Seattle Flu Study Chu etl al
EPI_ISL_417079 hCoV-19/USA/WA-S26/2020 North America / USA / Washington / Snohomish County 2020-03-02 Washington State Department of Health Seattle Flu Study Chu etl al
EPI_ISL_417081 hCoV-19/USA/WA-S28/2020 North America / USA / Washington / Snohomish County 2020-03-02 Washington State Department of Health Seattle Flu Study Chu etl al
EPI_ISL_417082 hCoV-19/USA/WA-S29/2020 North America / USA / Washington / Grant County 2020-03-02 Washington State Department of Health Seattle Flu Study Chu etl al
EPI_ISL_417085 hCoV-19/USA/WA-S32/2020 North America / USA / Washington / Snohomish County 2020-03-02 Washington State Department of Health Seattle Flu Study Chu etl al
EPI_ISL_417086 hCoV-19/USA/WA-S33/2020 North America / USA / Washington / Clark County 2020-03-01 Washington State Department of Health Seattle Flu Study Chu etl al
EPI_ISL_417087 hCoV-19/USA/WA-S34/2020 North America / USA / Washington 2020-03-02 Washington State Department of Health Seattle Flu Study Chu etl al
EPI_ISL_417088 hCoV-19/USA/WA-S35/2020 North America / USA / Washington 2020-03-02 Washington State Department of Health Seattle Flu Study Chu etl al
EPI_ISL_417089 hCoV-19/USA/WA-S36/2020 North America / USA / Washington 2020-03-02 Washington State Department of Health Seattle Flu Study Chu etl al
EPI_ISL_417500 hCoV-19/USA/MN56-MDH56/2020 North America / USA / Minnesota 2020-03-14 Minnesota Department of Health, Public Health Laboratory Minnesota Department of Health, Public Health Laboratory Matt Plumb, Jake Garfin and Xiong Wang
EPI_ISL_417501 hCoV-19/USA/MN57-MDH57/2020 North America / USA / Minnesota 2020-03-14 Minnesota Department of Health, Public Health Laboratory Minnesota Department of Health, Public Health Laboratory Matt Plumb, Jake Garfin and Xiong Wang
EPI_ISL_417502 hCoV-19/USA/MN58-MDH58/2020 North America / USA / Minnesota 2020-03-14 Minnesota Department of Health, Public Health Laboratory Minnesota Department of Health, Public Health Laboratory Matt Plumb, Jake Garfin and Xiong Wang
EPI_ISL_417503 hCoV-19/USA/MN59-MDH59/2020 North America / USA / Minnesota 2020-03-14 Minnesota Department of Health, Public Health Laboratory Minnesota Department of Health, Public Health Laboratory Matt Plumb, Jake Garfin and Xiong Wang
EPI_ISL_417504 hCoV-19/USA/WI-UW-15/2020 North America / USA / Wisconsin 2020-03-16 University of Wisconsin-Madison AIDS Vaccine Research Laboratories University of Wisconsin-Madison AIDS Vaccine Research Laboratories Gage Moreno, Katarina Braun, et al. AIDS Vaccine Research Laboratories
EPI_ISL_417505 hCoV-19/USA/WI-UW-11/2020 North America / USA / Wisconsin 2020-03-15 University of Wisconsin-Madison AIDS Vaccine Research Laboratories University of Wisconsin-Madison AIDS Vaccine Research Laboratories Gage Moreno, Katarina Braun, et al. AIDS Vaccine Research Laboratories

EPI_ISL_417506 hCoV-19/USA/WI-UW-12/2020 North America / USA / Wisconsin 2020-03-16 University of Wisconsin-Madison AIDS Vaccine Research Laboratories University of Wisconsin-Madison AIDS Vaccine Research Laboratories Gage Moreno, Katarina Braun, et al. AIDS Vaccine Research Laboratories
EPI_ISL_417529 hCoV-19/Luxembourg/LNS5731562/2020 Europe / Luxembourg 2020-03-18 Laboratoire Nationale de Santé, Microbiology, Virology Laboratoire Nationale de Santé, Microbiology, Epidemiology and Microbial Genomics Anke Wienecke-Baldacchino, Ardashel Latsuzbaia, Jessica Tapp, Catherine Ragimbeau, Guillaume Fournier, Tamir Abdelrahman, Trung Nguyen Nguyen, Joel Mossong
EPI_ISL_417530 hCoV-19/Luxembourg/LNS2128808/2020 Europe / Luxembourg 2020-03-18 Laboratoire Nationale de Santé, Microbiology, Virology Laboratoire Nationale de Santé, Microbiology, Epidemiology and Microbial Genomics Anke Wienecke-Baldacchino, Ardashel Latsuzbaia, Jessica Tapp, Catherine Ragimbeau, Guillaume Fournier, Tamir Abdelrahman, Trung Nguyen Nguyen, Joel Mossong
EPI_ISL_417531 hCoV-19/Luxembourg/LNS2013896/2020 Europe / Luxembourg 2020-03-18 Laboratoire Nationale de Santé, Microbiology, Virology Laboratoire Nationale de Santé, Microbiology, Epidemiology and Microbial Genomics Anke Wienecke-Baldacchino, Ardashel Latsuzbaia, Jessica Tapp, Catherine Ragimbeau, Guillaume Fournier, Tamir Abdelrahman, Trung Nguyen Nguyen, Joel Mossong
EPI_ISL_417532 hCoV-19/Luxembourg/LNS8489624/2020 Europe / Luxembourg 2020-03-18 Laboratoire Nationale de Santé, Microbiology, Virology Laboratoire Nationale de Santé, Microbiology, Epidemiology and Microbial Genomics Anke Wienecke-Baldacchino, Ardashel Latsuzbaia, Jessica Tapp, Catherine Ragimbeau, Guillaume Fournier, Tamir Abdelrahman, Trung Nguyen Nguyen, Joel Mossong
EPI_ISL_417533 hCoV-19/Luxembourg/LNS6282845/2020 Europe / Luxembourg 2020-03-18 Laboratoire Nationale de Santé, Microbiology, Virology Laboratoire Nationale de Santé, Microbiology, Epidemiology and Microbial Genomics Anke Wienecke-Baldacchino, Ardashel Latsuzbaia, Jessica Tapp, Catherine Ragimbeau, Guillaume Fournier, Tamir Abdelrahman, Trung Nguyen Nguyen, Joel Mossong
EPI_ISL_417534 hCoV-19/Luxembourg/LNS0158952/2020 Europe / Luxembourg 2020-03-18 Laboratoire Nationale de Santé, Microbiology, Virology Laboratoire Nationale de Santé, Microbiology, Epidemiology and Microbial Genomics Anke Wienecke-Baldacchino, Ardashel Latsuzbaia, Jessica Tapp, Catherine Ragimbeau, Guillaume Fournier, Tamir Abdelrahman, Trung Nguyen Nguyen, Joel Mossong
EPI_ISL_417535 hCoV-19/Iceland/1/2020 Europe / Iceland / Reykjavik 2020-03-13 deCODE genetics deCODE genetics Daniel F Gudbjartsson; Agnar Helgason; Hakon Jonsson; Olafur T Magnusson; Pall Melsted; Gudmundur L Norddahl; Jona Saemundsdottir; Asgeir Sigurdsson; Patrick Sulem; Arna B Agustsdottir; Berglind Eiriksdottir; Run Fridriksdottir; Elisabet E Gardarsdottir; Gudmundur Georgsson; Olafia S Gretarsdottir; Kjartan R Gudmundsson; Thora R Gunnarsdottir; Arnaldur Gylfason; Hilma Holm; Brynjar O Jensson; Aslaug Jonasdottir; Kamilla S Josefsdottir; Thordur Kristjansson; Droplaug N Magnusdottir; Louise le Roux; Gudrun Sigmundsdottir; Gardar Sveinbjornsson; Kristin E Sveinsdottir; Maney Sveinsdottir; Emil A Thorarensen; Bjarni Thorbjornsson; Gisli Masson; Ingileif Jonsdottir; Alma Moller; Thorolfur Gudnason; Karl G Kristinsson; Unnur Thorsteinsdottir; Kari Stefansson
EPI_ISL_417536 hCoV-19/Iceland/10/2020 Europe / Iceland / Reykjavik 2020-03-16 deCODE genetics deCODE genetics Daniel F Gudbjartsson; Agnar Helgason; Hakon Jonsson; Olafur T Magnusson; Pall Melsted; Gudmundur L Norddahl; Jona Saemundsdottir; Asgeir Sigurdsson; Patrick Sulem; Arna B Agustsdottir; Berglind Eiriksdottir; Run Fridriksdottir; Elisabet E Gardarsdottir; Gudmundur Georgsson; Olafia S Gretarsdottir; Kjartan R Gudmundsson; Thora R Gunnarsdottir; Arnaldur Gylfason; Hilma Holm; Brynjar O Jensson; Aslaug Jonasdottir; Kamilla S Josefsdottir; Thordur Kristjansson; Droplaug N Magnusdottir; Louise le Roux; Gudrun Sigmundsdottir; Gardar Sveinbjornsson; Kristin E Sveinsdottir; Maney Sveinsdottir; Emil A Thorarensen; Bjarni Thorbjornsson; Gisli Masson; Ingileif Jonsdottir; Alma Moller; Thorolfur Gudnason; Karl G Kristinsson; Unnur Thorsteinsdottir; Kari Stefansson
EPI_ISL_417537 hCoV-19/Iceland/100/2020 Europe / Iceland / Reykjavik 2020-03-16 deCODE genetics deCODE genetics Daniel F Gudbjartsson; Agnar Helgason; Hakon Jonsson; Olafur T Magnusson; Pall Melsted; Gudmundur L Norddahl; Jona Saemundsdottir; Asgeir Sigurdsson; Patrick Sulem; Arna B Agustsdottir; Berglind Eiriksdottir; Run Fridriksdottir; Elisabet E Gardarsdottir; Gudmundur Georgsson; Olafia S Gretarsdottir; Kjartan R Gudmundsson; Thora R Gunnarsdottir; Arnaldur Gylfason; Hilma Holm; Brynjar O Jensson; Aslaug Jonasdottir; Kamilla S Josefsdottir; Thordur Kristjansson; Droplaug N Magnusdottir; Louise le Roux; Gudrun Sigmundsdottir; Gardar Sveinbjornsson; Kristin E Sveinsdottir; Maney Sveinsdottir; Emil A Thorarensen; Bjarni Thorbjornsson; Gisli Masson; Ingileif Jonsdottir; Alma Moller; Thorolfur Gudnason; Karl G Kristinsson; Unnur Thorsteinsdottir; Kari Stefansson
EPI_ISL_417538 hCoV-19/Iceland/104/2020 Europe / Iceland / Reykjavik 2020-03-17 deCODE genetics deCODE genetics Daniel F Gudbjartsson; Agnar Helgason; Hakon Jonsson; Olafur T Magnusson; Pall Melsted; Gudmundur L Norddahl; Jona Saemundsdottir; Asgeir Sigurdsson; Patrick Sulem; Arna B Agustsdottir; Berglind Eiriksdottir; Run Fridriksdottir; Elisabet E Gardarsdottir; Gudmundur Georgsson; Olafia S Gretarsdottir; Kjartan R Gudmundsson; Thora R Gunnarsdottir; Arnaldur Gylfason; Hilma Holm; Brynjar O Jensson; Aslaug Jonasdottir; Kamilla S Josefsdottir; Thordur Kristjansson; Droplaug N Magnusdottir; Louise le Roux; Gudrun Sigmundsdottir; Gardar Sveinbjornsson; Kristin E Sveinsdottir; Maney Sveinsdottir; Emil A Thorarensen; Bjarni Thorbjornsson; Gisli Masson; Ingileif Jonsdottir; Alma Moller; Thorolfur Gudnason; Karl G Kristinsson; Unnur Thorsteinsdottir; Kari Stefansson
EPI_ISL_417539 hCoV-19/Iceland/11/2020 Europe / Iceland / Reykjavik 2020-03-16 deCODE genetics deCODE genetics Daniel F Gudbjartsson; Agnar Helgason; Hakon Jonsson; Olafur T Magnusson; Pall Melsted; Gudmundur L Norddahl; Jona Saemundsdottir; Asgeir Sigurdsson; Patrick Sulem; Arna B Agustsdottir; Berglind Eiriksdottir; Run Fridriksdottir; Elisabet E Gardarsdottir; Gudmundur Georgsson; Olafia S Gretarsdottir; Kjartan R Gudmundsson; Thora R Gunnarsdottir; Arnaldur Gylfason; Hilma Holm; Brynjar O Jensson; Aslaug Jonasdottir; Kamilla S Josefsdottir; Thordur Kristjansson; Droplaug N Magnusdottir; Louise le Roux; Gudrun Sigmundsdottir; Gardar Sveinbjornsson; Kristin E Sveinsdottir; Maney Sveinsdottir; Emil A Thorarensen; Bjarni

Thorbjornsson; Gisli Masson; Ingileif Jonsdottir; Alma Moller; Thorolfur Gudnason; Karl G Kristinsson; Unnur Thorsteinsdottir; Kari Stefansson
EPI_ISL_417540 hCoV-19/Iceland/115/2020 Europe / Iceland / Reykjavik 2020-03-17 deCODE genetics deCODE genetics Daniel F Gudbjartsson; Agnar Helgason; Hakon Jonsson; Olafur T Magnusson; Pall Melsted; Gudmundur L Norddahl; Jona Saemundsdottir; Asgeir Sigurdsson; Patrick Sulem; Arna B Agustsdottir; Berglind Eiriksdottir; Run Fridriksdottir; Elisabet E Gardarsdottir; Gudmundur Georgsson; Olafia S Gretarsdottir; Kjartan R Gudmundsson; Thora R Gunnarsdottir; Arnaldur Gylfason; Hilma Holm; Brynjar O Jensson; Aslaug Jonasdottir; Kamilla S Josefsdottir; Thordur Kristjansson; Droplaug N Magnusdottir; Louise le Roux; Gudrun Sigmundsdottir; Gardar Sveinbjornsson; Kristin E Sveinsdottir; Maney Sveinsdottir; Emil A Thorarensen; Bjarni Thorbjornsson; Gisli Masson; Ingileif Jonsdottir; Alma Moller; Thorolfur Gudnason; Karl G Kristinsson; Unnur Thorsteinsdottir; Kari Stefansson
EPI_ISL_417541 hCoV-19/Iceland/116/2020 Europe / Iceland / Reykjavik 2020-03-17 deCODE genetics deCODE genetics Daniel F Gudbjartsson; Agnar Helgason; Hakon Jonsson; Olafur T Magnusson; Pall Melsted; Gudmundur L Norddahl; Jona Saemundsdottir; Asgeir Sigurdsson; Patrick Sulem; Arna B Agustsdottir; Berglind Eiriksdottir; Run Fridriksdottir; Elisabet E Gardarsdottir; Gudmundur Georgsson; Olafia S Gretarsdottir; Kjartan R Gudmundsson; Thora R Gunnarsdottir; Arnaldur Gylfason; Hilma Holm; Brynjar O Jensson; Aslaug Jonasdottir; Kamilla S Josefsdottir; Thordur Kristjansson; Droplaug N Magnusdottir; Louise le Roux; Gudrun Sigmundsdottir; Gardar Sveinbjornsson; Kristin E Sveinsdottir; Maney Sveinsdottir; Emil A Thorarensen; Bjarni Thorbjornsson; Gisli Masson; Ingileif Jonsdottir; Alma Moller; Thorolfur Gudnason; Karl G Kristinsson; Unnur Thorsteinsdottir; Kari Stefansson
EPI_ISL_417542 hCoV-19/Iceland/117/2020 Europe / Iceland / Reykjavik 2020-03-17 deCODE genetics deCODE genetics Daniel F Gudbjartsson; Agnar Helgason; Hakon Jonsson; Olafur T Magnusson; Pall Melsted; Gudmundur L Norddahl; Jona Saemundsdottir; Asgeir Sigurdsson; Patrick Sulem; Arna B Agustsdottir; Berglind Eiriksdottir; Run Fridriksdottir; Elisabet E Gardarsdottir; Gudmundur Georgsson; Olafia S Gretarsdottir; Kjartan R Gudmundsson; Thora R Gunnarsdottir; Arnaldur Gylfason; Hilma Holm; Brynjar O Jensson; Aslaug Jonasdottir; Kamilla S Josefsdottir; Thordur Kristjansson; Droplaug N Magnusdottir; Louise le Roux; Gudrun Sigmundsdottir; Gardar Sveinbjornsson; Kristin E Sveinsdottir; Maney Sveinsdottir; Emil A Thorarensen; Bjarni Thorbjornsson; Gisli Masson; Ingileif Jonsdottir; Alma Moller; Thorolfur Gudnason; Karl G Kristinsson; Unnur Thorsteinsdottir; Kari Stefansson
EPI_ISL_417543 hCoV-19/Iceland/118/2020 Europe / Iceland / Reykjavik 2020-03-17 deCODE genetics deCODE genetics Daniel F Gudbjartsson; Agnar Helgason; Hakon Jonsson; Olafur T Magnusson; Pall Melsted; Gudmundur L Norddahl; Jona Saemundsdottir; Asgeir Sigurdsson; Patrick Sulem; Arna B Agustsdottir; Berglind Eiriksdottir; Run Fridriksdottir; Elisabet E Gardarsdottir; Gudmundur Georgsson; Olafia S Gretarsdottir; Kjartan R Gudmundsson; Thora R Gunnarsdottir; Arnaldur Gylfason; Hilma Holm; Brynjar O Jensson; Aslaug Jonasdottir; Kamilla S Josefsdottir; Thordur Kristjansson; Droplaug N Magnusdottir; Louise le Roux; Gudrun Sigmundsdottir; Gardar Sveinbjornsson; Kristin E Sveinsdottir; Maney Sveinsdottir; Emil A Thorarensen; Bjarni Thorbjornsson; Gisli Masson; Ingileif Jonsdottir; Alma Moller; Thorolfur Gudnason; Karl G Kristinsson; Unnur Thorsteinsdottir; Kari Stefansson
EPI_ISL_417544 hCoV-19/Iceland/119/2020 Europe / Iceland / Reykjavik 2020-03-17 deCODE genetics deCODE genetics Daniel F Gudbjartsson; Agnar Helgason; Hakon Jonsson; Olafur T Magnusson; Pall Melsted; Gudmundur L Norddahl; Jona Saemundsdottir; Asgeir Sigurdsson; Patrick Sulem; Arna B Agustsdottir; Berglind Eiriksdottir; Run Fridriksdottir; Elisabet E Gardarsdottir; Gudmundur Georgsson; Olafia S Gretarsdottir; Kjartan R Gudmundsson; Thora R Gunnarsdottir; Arnaldur Gylfason; Hilma Holm; Brynjar O Jensson; Aslaug Jonasdottir; Kamilla S Josefsdottir; Thordur Kristjansson; Droplaug N Magnusdottir; Louise le Roux; Gudrun Sigmundsdottir; Gardar Sveinbjornsson; Kristin E Sveinsdottir; Maney Sveinsdottir; Emil A Thorarensen; Bjarni Thorbjornsson; Gisli Masson; Ingileif Jonsdottir; Alma Moller; Thorolfur Gudnason; Karl G Kristinsson; Unnur Thorsteinsdottir; Kari Stefansson
EPI_ISL_417545 hCoV-19/Iceland/120/2020 Europe / Iceland / Reykjavik 2020-03-17 deCODE genetics deCODE genetics Daniel F Gudbjartsson; Agnar Helgason; Hakon Jonsson; Olafur T Magnusson; Pall Melsted; Gudmundur L Norddahl; Jona Saemundsdottir; Asgeir Sigurdsson; Patrick Sulem; Arna B Agustsdottir; Berglind Eiriksdottir; Run Fridriksdottir; Elisabet E Gardarsdottir; Gudmundur Georgsson; Olafia S Gretarsdottir; Kjartan R Gudmundsson; Thora R Gunnarsdottir; Arnaldur Gylfason; Hilma Holm; Brynjar O Jensson; Aslaug Jonasdottir; Kamilla S Josefsdottir; Thordur Kristjansson; Droplaug N Magnusdottir; Louise le Roux; Gudrun Sigmundsdottir; Gardar Sveinbjornsson; Kristin E Sveinsdottir; Maney Sveinsdottir; Emil A Thorarensen; Bjarni Thorbjornsson; Gisli Masson; Ingileif Jonsdottir; Alma Moller; Thorolfur Gudnason; Karl G Kristinsson; Unnur Thorsteinsdottir; Kari Stefansson
EPI_ISL_417548 hCoV-19/Iceland/170/2020 Europe / Iceland / Reykjavik 2020-03-17 deCODE genetics deCODE genetics Daniel F Gudbjartsson; Agnar Helgason; Hakon Jonsson; Olafur T Magnusson; Pall Melsted; Gudmundur L Norddahl; Jona Saemundsdottir; Asgeir Sigurdsson; Patrick Sulem; Arna B Agustsdottir; Berglind Eiriksdottir; Run Fridriksdottir; Elisabet E Gardarsdottir; Gudmundur Georgsson; Olafia S Gretarsdottir; Kjartan R Gudmundsson; Thora R Gunnarsdottir; Arnaldur Gylfason; Hilma Holm; Brynjar O Jensson; Aslaug Jonasdottir; Kamilla S Josefsdottir; Thordur Kristjansson; Droplaug N Magnusdottir; Louise le Roux; Gudrun Sigmundsdottir; Gardar Sveinbjornsson; Kristin E Sveinsdottir; Maney Sveinsdottir; Emil A Thorarensen; Bjarni Thorbjornsson; Gisli Masson; Ingileif Jonsdottir; Alma Moller; Thorolfur Gudnason; Karl G Kristinsson; Unnur Thorsteinsdottir; Kari Stefansson
EPI_ISL_417549 hCoV-19/Iceland/171/2020 Europe / Iceland / Reykjavik 2020-03-14 deCODE genetics deCODE genetics Daniel F Gudbjartsson; Agnar Helgason; Hakon Jonsson; Olafur T Magnusson; Pall Melsted; Gudmundur L Norddahl; Jona Saemundsdottir; Asgeir Sigurdsson; Patrick Sulem; Arna B Agustsdottir; Berglind Eiriksdottir; Run Fridriksdottir; Elisabet E Gardarsdottir; Gudmundur Georgsson; Olafia S Gretarsdottir; Kjartan R Gudmundsson; Thora R Gunnarsdottir; Arnaldur Gylfason; Hilma Holm; Brynjar O Jensson; Aslaug

Jonasdottir; Kamilla S Josefsdottir; Thordur Kristjansson; Droplaug N Magnusdottir; Louise le Roux; Gudrun Sigmundsdottir; Gardar Sveinbjornsson; Kristin E Sveinsdottir; Maney Sveinsdottir; Emil A Thorarensen; Bjarni Thorbjornsson; Gisli Masson; Ingileif Jonsdottir; Alma Moller; Thorolfur Gudnason; Karl G Kristinsson; Unnur Thorsteinsdottir; Kari Stefansson
EPI_ISL_417550 hCoV-19/Iceland/184/2020 Europe / Iceland / Reykjavik 2020-03-16 The National University Hospital of Iceland deCODE genetics Daniel F Gudbjartsson; Agnar Helgason; Hakon Jonsson; Olafur T Magnusson; Pall Melsted; Gudmundur L Norddahl; Jona Saemundsdottir; Asgeir Sigurdsson; Patrick Sulem; Arna B Agustsdottir; Berglind Eiriksdottir; Run Fridriksdottir; Elisabet E Gardarsdottir; Gudmundur Georgsson; Olafia S Gretarsdottir; Kjartan R Gudmundsson; Thora R Gunnarsdottir; Arnaldur Gylfason; Hilma Holm; Brynjar O Jensson; Aslaug Jonasdottir; Kamilla S Josefsdottir; Thordur Kristjansson; Droplaug N Magnusdottir; Louise le Roux; Gudrun Sigmundsdottir; Gardar Sveinbjornsson; Kristin E Sveinsdottir; Maney Sveinsdottir; Emil A Thorarensen; Bjarni Thorbjornsson; Gisli Masson; Ingileif Jonsdottir; Alma Moller; Thorolfur Gudnason; Karl G Kristinsson; Unnur Thorsteinsdottir; Kari Stefansson
EPI_ISL_417551 hCoV-19/Iceland/187/2020 Europe / Iceland / Reykjavik 2020-03-16 The National University Hospital of Iceland deCODE genetics Daniel F Gudbjartsson; Agnar Helgason; Hakon Jonsson; Olafur T Magnusson; Pall Melsted; Gudmundur L Norddahl; Jona Saemundsdottir; Asgeir Sigurdsson; Patrick Sulem; Arna B Agustsdottir; Berglind Eiriksdottir; Run Fridriksdottir; Elisabet E Gardarsdottir; Gudmundur Georgsson; Olafia S Gretarsdottir; Kjartan R Gudmundsson; Thora R Gunnarsdottir; Arnaldur Gylfason; Hilma Holm; Brynjar O Jensson; Aslaug Jonasdottir; Kamilla S Josefsdottir; Thordur Kristjansson; Droplaug N Magnusdottir; Louise le Roux; Gudrun Sigmundsdottir; Gardar Sveinbjornsson; Kristin E Sveinsdottir; Maney Sveinsdottir; Emil A Thorarensen; Bjarni Thorbjornsson; Gisli Masson; Ingileif Jonsdottir; Alma Moller; Thorolfur Gudnason; Karl G Kristinsson; Unnur Thorsteinsdottir; Kari Stefansson
EPI_ISL_417552 hCoV-19/Iceland/2/2020 Europe / Iceland / Reykjavik 2020-03-16 deCODE genetics deCODE genetics Daniel F Gudbjartsson; Agnar Helgason; Hakon Jonsson; Olafur T Magnusson; Pall Melsted; Gudmundur L Norddahl; Jona Saemundsdottir; Asgeir Sigurdsson; Patrick Sulem; Arna B Agustsdottir; Berglind Eiriksdottir; Run Fridriksdottir; Elisabet E Gardarsdottir; Gudmundur Georgsson; Olafia S Gretarsdottir; Kjartan R Gudmundsson; Thora R Gunnarsdottir; Arnaldur Gylfason; Hilma Holm; Brynjar O Jensson; Aslaug Jonasdottir; Kamilla S Josefsdottir; Thordur Kristjansson; Droplaug N Magnusdottir; Louise le Roux; Gudrun Sigmundsdottir; Gardar Sveinbjornsson; Kristin E Sveinsdottir; Maney Sveinsdottir; Emil A Thorarensen; Bjarni Thorbjornsson; Gisli Masson; Ingileif Jonsdottir; Alma Moller; Thorolfur Gudnason; Karl G Kristinsson; Unnur Thorsteinsdottir; Kari Stefansson
EPI_ISL_417553 hCoV-19/Iceland/220/2020 Europe / Iceland / Reykjavik 2020-03-16 The National University Hospital of Iceland deCODE genetics Daniel F Gudbjartsson; Agnar Helgason; Hakon Jonsson; Olafur T Magnusson; Pall Melsted; Gudmundur L Norddahl; Jona Saemundsdottir; Asgeir Sigurdsson; Patrick Sulem; Arna B Agustsdottir; Berglind Eiriksdottir; Run Fridriksdottir; Elisabet E Gardarsdottir; Gudmundur Georgsson; Olafia S Gretarsdottir; Kjartan R Gudmundsson; Thora R Gunnarsdottir; Arnaldur Gylfason; Hilma Holm; Brynjar O Jensson; Aslaug Jonasdottir; Kamilla S Josefsdottir; Thordur Kristjansson; Droplaug N Magnusdottir; Louise le Roux; Gudrun Sigmundsdottir; Gardar Sveinbjornsson; Kristin E Sveinsdottir; Maney Sveinsdottir; Emil A Thorarensen; Bjarni Thorbjornsson; Gisli Masson; Ingileif Jonsdottir; Alma Moller; Thorolfur Gudnason; Karl G Kristinsson; Unnur Thorsteinsdottir; Kari Stefansson
EPI_ISL_417555 hCoV-19/Iceland/227/2020 Europe / Iceland / Reykjavik 2020-03-17 The National University Hospital of Iceland deCODE genetics Daniel F Gudbjartsson; Agnar Helgason; Hakon Jonsson; Olafur T Magnusson; Pall Melsted; Gudmundur L Norddahl; Jona Saemundsdottir; Asgeir Sigurdsson; Patrick Sulem; Arna B Agustsdottir; Berglind Eiriksdottir; Run Fridriksdottir; Elisabet E Gardarsdottir; Gudmundur Georgsson; Olafia S Gretarsdottir; Kjartan R Gudmundsson; Thora R Gunnarsdottir; Arnaldur Gylfason; Hilma Holm; Brynjar O Jensson; Aslaug Jonasdottir; Kamilla S Josefsdottir; Thordur Kristjansson; Droplaug N Magnusdottir; Louise le Roux; Gudrun Sigmundsdottir; Gardar Sveinbjornsson; Kristin E Sveinsdottir; Maney Sveinsdottir; Emil A Thorarensen; Bjarni Thorbjornsson; Gisli Masson; Ingileif Jonsdottir; Alma Moller; Thorolfur Gudnason; Karl G Kristinsson; Unnur Thorsteinsdottir; Kari Stefansson
EPI_ISL_417556 hCoV-19/Iceland/228/2020 Europe / Iceland / Reykjavik 2020-03-17 The National University Hospital of Iceland deCODE genetics Daniel F Gudbjartsson; Agnar Helgason; Hakon Jonsson; Olafur T Magnusson; Pall Melsted; Gudmundur L Norddahl; Jona Saemundsdottir; Asgeir Sigurdsson; Patrick Sulem; Arna B Agustsdottir; Berglind Eiriksdottir; Run Fridriksdottir; Elisabet E Gardarsdottir; Gudmundur Georgsson; Olafia S Gretarsdottir; Kjartan R Gudmundsson; Thora R Gunnarsdottir; Arnaldur Gylfason; Hilma Holm; Brynjar O Jensson; Aslaug Jonasdottir; Kamilla S Josefsdottir; Thordur Kristjansson; Droplaug N Magnusdottir; Louise le Roux; Gudrun Sigmundsdottir; Gardar Sveinbjornsson; Kristin E Sveinsdottir; Maney Sveinsdottir; Emil A Thorarensen; Bjarni Thorbjornsson; Gisli Masson; Ingileif Jonsdottir; Alma Moller; Thorolfur Gudnason; Karl G Kristinsson; Unnur Thorsteinsdottir; Kari Stefansson
EPI_ISL_417557 hCoV-19/Iceland/229/2020 Europe / Iceland / Reykjavik 2020-03-17 The National University Hospital of Iceland deCODE genetics Daniel F Gudbjartsson; Agnar Helgason; Hakon Jonsson; Olafur T Magnusson; Pall Melsted; Gudmundur L Norddahl; Jona Saemundsdottir; Asgeir Sigurdsson; Patrick Sulem; Arna B Agustsdottir; Berglind Eiriksdottir; Run Fridriksdottir; Elisabet E Gardarsdottir; Gudmundur Georgsson; Olafia S Gretarsdottir; Kjartan R Gudmundsson; Thora R Gunnarsdottir; Arnaldur Gylfason; Hilma Holm; Brynjar O Jensson; Aslaug Jonasdottir; Kamilla S Josefsdottir; Thordur Kristjansson; Droplaug N Magnusdottir; Louise le Roux; Gudrun Sigmundsdottir; Gardar Sveinbjornsson; Kristin E Sveinsdottir; Maney Sveinsdottir; Emil A Thorarensen; Bjarni Thorbjornsson; Gisli Masson; Ingileif Jonsdottir; Alma Moller; Thorolfur Gudnason; Karl G Kristinsson; Unnur Thorsteinsdottir; Kari Stefansson
EPI_ISL_417558 hCoV-19/Iceland/230/2020 Europe / Iceland / Reykjavik 2020-03-17 The National University Hospital of Iceland deCODE genetics Daniel F Gudbjartsson; Agnar Helgason; Hakon Jonsson; Olafur T Magnusson; Pall Melsted; Gudmundur L Norddahl; Jona Saemundsdottir; Asgeir Sigurdsson; Patrick Sulem; Arna B

Agustsdottir; Berglind Eiriksdottir; Run Fridriksdottir; Elisabet E Gardarsdottir; Gudmundur Georgsson; Olafia S Gretarsdottir; Kjartan R Gudmundsson; Thora R Gunnarsdottir; Arnaldur Gylfason; Hilma Holm; Brynjar O Jensson; Aslaug Jonasdottir; Kamilla S Josefsdottir; Thordur Kristjansson; Droplaug N Magnusdottir; Louise le Roux; Gudrun Sigmundsdottir; Gardar Sveinbjornsson; Kristin E Sveinsdottir; Maney Sveinsdottir; Emil A Thorarensen; Bjarni Thorbjornsson; Gisli Masson; Ingileif Jonsdottir; Alma Moller; Thorolfur Gudnason; Karl G Kristinsson; Unnur Thorsteinsdottir; Kari Stefansson
EPI_ISL_417560 hCoV-19/Iceland/232/2020 Europe / Iceland / Reykjavik 2020-03-17 The National University Hospital of Iceland deCODE genetics Daniel F Gudbjartsson; Agnar Helgason; Hakon Jonsson; Olafur T Magnusson; Pall Melsted; Gudmundur L Norddahl; Jona Saemundsdottir; Asgeir Sigurdsson; Patrick Sulem; Arna B Agustsdottir; Berglind Eiriksdottir; Run Fridriksdottir; Elisabet E Gardarsdottir; Gudmundur Georgsson; Olafia S Gretarsdottir; Kjartan R Gudmundsson; Thora R Gunnarsdottir; Arnaldur Gylfason; Hilma Holm; Brynjar O Jensson; Aslaug Jonasdottir; Kamilla S Josefsdottir; Thordur Kristjansson; Droplaug N Magnusdottir; Louise le Roux; Gudrun Sigmundsdottir; Gardar Sveinbjornsson; Kristin E Sveinsdottir; Maney Sveinsdottir; Emil A Thorarensen; Bjarni Thorbjornsson; Gisli Masson; Ingileif Jonsdottir; Alma Moller; Thorolfur Gudnason; Karl G Kristinsson; Unnur Thorsteinsdottir; Kari Stefansson
EPI_ISL_417561 hCoV-19/Iceland/233/2020 Europe / Iceland / Reykjavik 2020-03-17 The National University Hospital of Iceland deCODE genetics Daniel F Gudbjartsson; Agnar Helgason; Hakon Jonsson; Olafur T Magnusson; Pall Melsted; Gudmundur L Norddahl; Jona Saemundsdottir; Asgeir Sigurdsson; Patrick Sulem; Arna B Agustsdottir; Berglind Eiriksdottir; Run Fridriksdottir; Elisabet E Gardarsdottir; Gudmundur Georgsson; Olafia S Gretarsdottir; Kjartan R Gudmundsson; Thora R Gunnarsdottir; Arnaldur Gylfason; Hilma Holm; Brynjar O Jensson; Aslaug Jonasdottir; Kamilla S Josefsdottir; Thordur Kristjansson; Droplaug N Magnusdottir; Louise le Roux; Gudrun Sigmundsdottir; Gardar Sveinbjornsson; Kristin E Sveinsdottir; Maney Sveinsdottir; Emil A Thorarensen; Bjarni Thorbjornsson; Gisli Masson; Ingileif Jonsdottir; Alma Moller; Thorolfur Gudnason; Karl G Kristinsson; Unnur Thorsteinsdottir; Kari Stefansson
EPI_ISL_417562 hCoV-19/Iceland/234/2020 Europe / Iceland / Reykjavik 2020-03-17 The National University Hospital of Iceland deCODE genetics Daniel F Gudbjartsson; Agnar Helgason; Hakon Jonsson; Olafur T Magnusson; Pall Melsted; Gudmundur L Norddahl; Jona Saemundsdottir; Asgeir Sigurdsson; Patrick Sulem; Arna B Agustsdottir; Berglind Eiriksdottir; Run Fridriksdottir; Elisabet E Gardarsdottir; Gudmundur Georgsson; Olafia S Gretarsdottir; Kjartan R Gudmundsson; Thora R Gunnarsdottir; Arnaldur Gylfason; Hilma Holm; Brynjar O Jensson; Aslaug Jonasdottir; Kamilla S Josefsdottir; Thordur Kristjansson; Droplaug N Magnusdottir; Louise le Roux; Gudrun Sigmundsdottir; Gardar Sveinbjornsson; Kristin E Sveinsdottir; Maney Sveinsdottir; Emil A Thorarensen; Bjarni Thorbjornsson; Gisli Masson; Ingileif Jonsdottir; Alma Moller; Thorolfur Gudnason; Karl G Kristinsson; Unnur Thorsteinsdottir; Kari Stefansson
EPI_ISL_417563 hCoV-19/Iceland/235/2020 Europe / Iceland / Reykjavik 2020-03-16 The National University Hospital of Iceland deCODE genetics Daniel F Gudbjartsson; Agnar Helgason; Hakon Jonsson; Olafur T Magnusson; Pall Melsted; Gudmundur L Norddahl; Jona Saemundsdottir; Asgeir Sigurdsson; Patrick Sulem; Arna B Agustsdottir; Berglind Eiriksdottir; Run Fridriksdottir; Elisabet E Gardarsdottir; Gudmundur Georgsson; Olafia S Gretarsdottir; Kjartan R Gudmundsson; Thora R Gunnarsdottir; Arnaldur Gylfason; Hilma Holm; Brynjar O Jensson; Aslaug Jonasdottir; Kamilla S Josefsdottir; Thordur Kristjansson; Droplaug N Magnusdottir; Louise le Roux; Gudrun Sigmundsdottir; Gardar Sveinbjornsson; Kristin E Sveinsdottir; Maney Sveinsdottir; Emil A Thorarensen; Bjarni Thorbjornsson; Gisli Masson; Ingileif Jonsdottir; Alma Moller; Thorolfur Gudnason; Karl G Kristinsson; Unnur Thorsteinsdottir; Kari Stefansson
EPI_ISL_417564 hCoV-19/Iceland/236/2020 Europe / Iceland / Reykjavik 2020-03-17 The National University Hospital of Iceland deCODE genetics Daniel F Gudbjartsson; Agnar Helgason; Hakon Jonsson; Olafur T Magnusson; Pall Melsted; Gudmundur L Norddahl; Jona Saemundsdottir; Asgeir Sigurdsson; Patrick Sulem; Arna B Agustsdottir; Berglind Eiriksdottir; Run Fridriksdottir; Elisabet E Gardarsdottir; Gudmundur Georgsson; Olafia S Gretarsdottir; Kjartan R Gudmundsson; Thora R Gunnarsdottir; Arnaldur Gylfason; Hilma Holm; Brynjar O Jensson; Aslaug Jonasdottir; Kamilla S Josefsdottir; Thordur Kristjansson; Droplaug N Magnusdottir; Louise le Roux; Gudrun Sigmundsdottir; Gardar Sveinbjornsson; Kristin E Sveinsdottir; Maney Sveinsdottir; Emil A Thorarensen; Bjarni Thorbjornsson; Gisli Masson; Ingileif Jonsdottir; Alma Moller; Thorolfur Gudnason; Karl G Kristinsson; Unnur Thorsteinsdottir; Kari Stefansson
EPI_ISL_417565 hCoV-19/Iceland/237/2020 Europe / Iceland / Reykjavik 2020-03-17 The National University Hospital of Iceland deCODE genetics Daniel F Gudbjartsson; Agnar Helgason; Hakon Jonsson; Olafur T Magnusson; Pall Melsted; Gudmundur L Norddahl; Jona Saemundsdottir; Asgeir Sigurdsson; Patrick Sulem; Arna B Agustsdottir; Berglind Eiriksdottir; Run Fridriksdottir; Elisabet E Gardarsdottir; Gudmundur Georgsson; Olafia S Gretarsdottir; Kjartan R Gudmundsson; Thora R Gunnarsdottir; Arnaldur Gylfason; Hilma Holm; Brynjar O Jensson; Aslaug Jonasdottir; Kamilla S Josefsdottir; Thordur Kristjansson; Droplaug N Magnusdottir; Louise le Roux; Gudrun Sigmundsdottir; Gardar Sveinbjornsson; Kristin E Sveinsdottir; Maney Sveinsdottir; Emil A Thorarensen; Bjarni Thorbjornsson; Gisli Masson; Ingileif Jonsdottir; Alma Moller; Thorolfur Gudnason; Karl G Kristinsson; Unnur Thorsteinsdottir; Kari Stefansson
EPI_ISL_417566 hCoV-19/Iceland/238/2020 Europe / Iceland / Reykjavik 2020-03-17 The National University Hospital of Iceland deCODE genetics Daniel F Gudbjartsson; Agnar Helgason; Hakon Jonsson; Olafur T Magnusson; Pall Melsted; Gudmundur L Norddahl; Jona Saemundsdottir; Asgeir Sigurdsson; Patrick Sulem; Arna B Agustsdottir; Berglind Eiriksdottir; Run Fridriksdottir; Elisabet E Gardarsdottir; Gudmundur Georgsson; Olafia S Gretarsdottir; Kjartan R Gudmundsson; Thora R Gunnarsdottir; Arnaldur Gylfason; Hilma Holm; Brynjar O Jensson; Aslaug Jonasdottir; Kamilla S Josefsdottir; Thordur Kristjansson; Droplaug N Magnusdottir; Louise le Roux; Gudrun Sigmundsdottir; Gardar Sveinbjornsson; Kristin E Sveinsdottir; Maney Sveinsdottir; Emil A Thorarensen; Bjarni Thorbjornsson; Gisli Masson; Ingileif Jonsdottir; Alma Moller; Thorolfur Gudnason; Karl G Kristinsson; Unnur Thorsteinsdottir; Kari Stefansson
EPI_ISL_417567 hCoV-19/Iceland/239/2020 Europe / Iceland / Reykjavik 2020-03-17 The National

University Hospital of Iceland  deCODE genetics Daniel F Gudbjartsson; Agnar Helgason; Hakon Jonsson; Olafur T Magnusson; Pall Melsted; Gudmundur L Norddahl; Jona Saemundsdottir; Asgeir Sigurdsson; Patrick Sulem; Arna B Agustsdottir; Berglind Eiriksdottir; Run Fridriksdottir; Elisabet E Gardarsdottir; Gudmundur Georgsson; Olafia S Gretarsdottir; Kjartan R Gudmundsson; Thora R Gunnarsdottir; Arnaldur Gylfason; Hilma Holm; Brynjar O Jensson; Aslaug Jonasdottir; Kamilla S Josefsdottir; Thordur Kristjansson; Droplaug N Magnusdottir; Louise le Roux; Gudrun Sigmundsdottir; Gardar Sveinbjornsson; Kristin E Sveinsdottir; Maney Sveinsdottir; Emil A Thorarensen; Bjarni Thorbjornsson; Gisli Masson; Ingileif Jonsdottir; Alma Moller; Thorolfur Gudnason; Karl G Kristinsson; Unnur Thorsteinsdottir; Kari Stefansson
EPI_ISL_417568  hCoV-19/Iceland/240/2020  Europe / Iceland / Reykjavik  2020-03-17  The National University Hospital of Iceland  deCODE genetics Daniel F Gudbjartsson; Agnar Helgason; Hakon Jonsson; Olafur T Magnusson; Pall Melsted; Gudmundur L Norddahl; Jona Saemundsdottir; Asgeir Sigurdsson; Patrick Sulem; Arna B Agustsdottir; Berglind Eiriksdottir; Run Fridriksdottir; Elisabet E Gardarsdottir; Gudmundur Georgsson; Olafia S Gretarsdottir; Kjartan R Gudmundsson; Thora R Gunnarsdottir; Arnaldur Gylfason; Hilma Holm; Brynjar O Jensson; Aslaug Jonasdottir; Kamilla S Josefsdottir; Thordur Kristjansson; Droplaug N Magnusdottir; Louise le Roux; Gudrun Sigmundsdottir; Gardar Sveinbjornsson; Kristin E Sveinsdottir; Maney Sveinsdottir; Emil A Thorarensen; Bjarni Thorbjornsson; Gisli Masson; Ingileif Jonsdottir; Alma Moller; Thorolfur Gudnason; Karl G Kristinsson; Unnur Thorsteinsdottir; Kari Stefansson
EPI_ISL_417569  hCoV-19/Iceland/241/2020  Europe / Iceland / Reykjavik  2020-03-17  The National University Hospital of Iceland  deCODE genetics Daniel F Gudbjartsson; Agnar Helgason; Hakon Jonsson; Olafur T Magnusson; Pall Melsted; Gudmundur L Norddahl; Jona Saemundsdottir; Asgeir Sigurdsson; Patrick Sulem; Arna B Agustsdottir; Berglind Eiriksdottir; Run Fridriksdottir; Elisabet E Gardarsdottir; Gudmundur Georgsson; Olafia S Gretarsdottir; Kjartan R Gudmundsson; Thora R Gunnarsdottir; Arnaldur Gylfason; Hilma Holm; Brynjar O Jensson; Aslaug Jonasdottir; Kamilla S Josefsdottir; Thordur Kristjansson; Droplaug N Magnusdottir; Louise le Roux; Gudrun Sigmundsdottir; Gardar Sveinbjornsson; Kristin E Sveinsdottir; Maney Sveinsdottir; Emil A Thorarensen; Bjarni Thorbjornsson; Gisli Masson; Ingileif Jonsdottir; Alma Moller; Thorolfur Gudnason; Karl G Kristinsson; Unnur Thorsteinsdottir; Kari Stefansson
EPI_ISL_417570  hCoV-19/Iceland/242/2020  Europe / Iceland / Reykjavik  2020-03-17  The National University Hospital of Iceland  deCODE genetics Daniel F Gudbjartsson; Agnar Helgason; Hakon Jonsson; Olafur T Magnusson; Pall Melsted; Gudmundur L Norddahl; Jona Saemundsdottir; Asgeir Sigurdsson; Patrick Sulem; Arna B Agustsdottir; Berglind Eiriksdottir; Run Fridriksdottir; Elisabet E Gardarsdottir; Gudmundur Georgsson; Olafia S Gretarsdottir; Kjartan R Gudmundsson; Thora R Gunnarsdottir; Arnaldur Gylfason; Hilma Holm; Brynjar O Jensson; Aslaug Jonasdottir; Kamilla S Josefsdottir; Thordur Kristjansson; Droplaug N Magnusdottir; Louise le Roux; Gudrun Sigmundsdottir; Gardar Sveinbjornsson; Kristin E Sveinsdottir; Maney Sveinsdottir; Emil A Thorarensen; Bjarni Thorbjornsson; Gisli Masson; Ingileif Jonsdottir; Alma Moller; Thorolfur Gudnason; Karl G Kristinsson; Unnur Thorsteinsdottir; Kari Stefansson
EPI_ISL_417571  hCoV-19/Iceland/243/2020  Europe / Iceland / Reykjavik  2020-03-17  The National University Hospital of Iceland  deCODE genetics Daniel F Gudbjartsson; Agnar Helgason; Hakon Jonsson; Olafur T Magnusson; Pall Melsted; Gudmundur L Norddahl; Jona Saemundsdottir; Asgeir Sigurdsson; Patrick Sulem; Arna B Agustsdottir; Berglind Eiriksdottir; Run Fridriksdottir; Elisabet E Gardarsdottir; Gudmundur Georgsson; Olafia S Gretarsdottir; Kjartan R Gudmundsson; Thora R Gunnarsdottir; Arnaldur Gylfason; Hilma Holm; Brynjar O Jensson; Aslaug Jonasdottir; Kamilla S Josefsdottir; Thordur Kristjansson; Droplaug N Magnusdottir; Louise le Roux; Gudrun Sigmundsdottir; Gardar Sveinbjornsson; Kristin E Sveinsdottir; Maney Sveinsdottir; Emil A Thorarensen; Bjarni Thorbjornsson; Gisli Masson; Ingileif Jonsdottir; Alma Moller; Thorolfur Gudnason; Karl G Kristinsson; Unnur Thorsteinsdottir; Kari Stefansson
EPI_ISL_417572  hCoV-19/Iceland/244/2020  Europe / Iceland / Reykjavik  2020-03-17  The National University Hospital of Iceland  deCODE genetics Daniel F Gudbjartsson; Agnar Helgason; Hakon Jonsson; Olafur T Magnusson; Pall Melsted; Gudmundur L Norddahl; Jona Saemundsdottir; Asgeir Sigurdsson; Patrick Sulem; Arna B Agustsdottir; Berglind Eiriksdottir; Run Fridriksdottir; Elisabet E Gardarsdottir; Gudmundur Georgsson; Olafia S Gretarsdottir; Kjartan R Gudmundsson; Thora R Gunnarsdottir; Arnaldur Gylfason; Hilma Holm; Brynjar O Jensson; Aslaug Jonasdottir; Kamilla S Josefsdottir; Thordur Kristjansson; Droplaug N Magnusdottir; Louise le Roux; Gudrun Sigmundsdottir; Gardar Sveinbjornsson; Kristin E Sveinsdottir; Maney Sveinsdottir; Emil A Thorarensen; Bjarni Thorbjornsson; Gisli Masson; Ingileif Jonsdottir; Alma Moller; Thorolfur Gudnason; Karl G Kristinsson; Unnur Thorsteinsdottir; Kari Stefansson
EPI_ISL_417573  hCoV-19/Iceland/245/2020  Europe / Iceland / Reykjavik  2020-03-16  The National University Hospital of Iceland  deCODE genetics Daniel F Gudbjartsson; Agnar Helgason; Hakon Jonsson; Olafur T Magnusson; Pall Melsted; Gudmundur L Norddahl; Jona Saemundsdottir; Asgeir Sigurdsson; Patrick Sulem; Arna B Agustsdottir; Berglind Eiriksdottir; Run Fridriksdottir; Elisabet E Gardarsdottir; Gudmundur Georgsson; Olafia S Gretarsdottir; Kjartan R Gudmundsson; Thora R Gunnarsdottir; Arnaldur Gylfason; Hilma Holm; Brynjar O Jensson; Aslaug Jonasdottir; Kamilla S Josefsdottir; Thordur Kristjansson; Droplaug N Magnusdottir; Louise le Roux; Gudrun Sigmundsdottir; Gardar Sveinbjornsson; Kristin E Sveinsdottir; Maney Sveinsdottir; Emil A Thorarensen; Bjarni Thorbjornsson; Gisli Masson; Ingileif Jonsdottir; Alma Moller; Thorolfur Gudnason; Karl G Kristinsson; Unnur Thorsteinsdottir; Kari Stefansson
EPI_ISL_417574  hCoV-19/Iceland/246/2020  Europe / Iceland / Reykjavik  2020-03-17  The National University Hospital of Iceland  deCODE genetics Daniel F Gudbjartsson; Agnar Helgason; Hakon Jonsson; Olafur T Magnusson; Pall Melsted; Gudmundur L Norddahl; Jona Saemundsdottir; Asgeir Sigurdsson; Patrick Sulem; Arna B Agustsdottir; Berglind Eiriksdottir; Run Fridriksdottir; Elisabet E Gardarsdottir; Gudmundur Georgsson; Olafia S Gretarsdottir; Kjartan R Gudmundsson; Thora R Gunnarsdottir; Arnaldur Gylfason; Hilma Holm; Brynjar O Jensson; Aslaug Jonasdottir; Kamilla S Josefsdottir; Thordur Kristjansson; Droplaug N Magnusdottir; Louise le Roux; Gudrun Sigmundsdottir; Gardar Sveinbjornsson; Kristin E Sveinsdottir; Maney Sveinsdottir; Emil A Thorarensen; Bjarni Thorbjornsson; Gisli Masson; Ingileif Jonsdottir; Alma Moller; Thorolfur Gudnason; Karl G

Kristinsson; Unnur Thorsteinsdottir; Kari Stefansson
EPI_ISL_417575 hCoV-19/Iceland/247/2020 Europe / Iceland / Reykjavik 2020-03-17 The National University Hospital of Iceland deCODE genetics Daniel F Gudbjartsson; Agnar Helgason; Hakon Jonsson; Olafur T Magnusson; Pall Melsted; Gudmundur L Norddahl; Jona Saemundsdottir; Asgeir Sigurdsson; Patrick Sulem; Arna B Agustsdottir; Berglind Eiriksdottir; Run Fridriksdottir; Elisabet E Gardarsdottir; Gudmundur Georgsson; Olafia S Gretarsdottir; Kjartan R Gudmundsson; Thora R Gunnarsdottir; Arnaldur Gylfason; Hilma Holm; Brynjar O Jensson; Aslaug Jonasdottir; Kamilla S Josefsdottir; Thordur Kristjansson; Droplaug N Magnusdottir; Louise le Roux; Gudrun Sigmundsdottir; Gardar Sveinbjornsson; Kristin E Sveinsdottir; Maney Sveinsdottir; Emil A Thorarensen; Bjarni Thorbjornsson; Gisli Masson; Ingileif Jonsdottir; Alma Moller; Thorolfur Gudnason; Karl G Kristinsson; Unnur Thorsteinsdottir; Kari Stefansson
EPI_ISL_417576 hCoV-19/Iceland/248/2020 Europe / Iceland / Reykjavik 2020-03-17 The National University Hospital of Iceland deCODE genetics Daniel F Gudbjartsson; Agnar Helgason; Hakon Jonsson; Olafur T Magnusson; Pall Melsted; Gudmundur L Norddahl; Jona Saemundsdottir; Asgeir Sigurdsson; Patrick Sulem; Arna B Agustsdottir; Berglind Eiriksdottir; Run Fridriksdottir; Elisabet E Gardarsdottir; Gudmundur Georgsson; Olafia S Gretarsdottir; Kjartan R Gudmundsson; Thora R Gunnarsdottir; Arnaldur Gylfason; Hilma Holm; Brynjar O Jensson; Aslaug Jonasdottir; Kamilla S Josefsdottir; Thordur Kristjansson; Droplaug N Magnusdottir; Louise le Roux; Gudrun Sigmundsdottir; Gardar Sveinbjornsson; Kristin E Sveinsdottir; Maney Sveinsdottir; Emil A Thorarensen; Bjarni Thorbjornsson; Gisli Masson; Ingileif Jonsdottir; Alma Moller; Thorolfur Gudnason; Karl G Kristinsson; Unnur Thorsteinsdottir; Kari Stefansson
EPI_ISL_417577 hCoV-19/Iceland/249/2020 Europe / Iceland / Reykjavik 2020-03-17 The National University Hospital of Iceland deCODE genetics Daniel F Gudbjartsson; Agnar Helgason; Hakon Jonsson; Olafur T Magnusson; Pall Melsted; Gudmundur L Norddahl; Jona Saemundsdottir; Asgeir Sigurdsson; Patrick Sulem; Arna B Agustsdottir; Berglind Eiriksdottir; Run Fridriksdottir; Elisabet E Gardarsdottir; Gudmundur Georgsson; Olafia S Gretarsdottir; Kjartan R Gudmundsson; Thora R Gunnarsdottir; Arnaldur Gylfason; Hilma Holm; Brynjar O Jensson; Aslaug Jonasdottir; Kamilla S Josefsdottir; Thordur Kristjansson; Droplaug N Magnusdottir; Louise le Roux; Gudrun Sigmundsdottir; Gardar Sveinbjornsson; Kristin E Sveinsdottir; Maney Sveinsdottir; Emil A Thorarensen; Bjarni Thorbjornsson; Gisli Masson; Ingileif Jonsdottir; Alma Moller; Thorolfur Gudnason; Karl G Kristinsson; Unnur Thorsteinsdottir; Kari Stefansson
EPI_ISL_417580 hCoV-19/Iceland/252/2020 Europe / Iceland / Reykjavik 2020-03-17 The National University Hospital of Iceland deCODE genetics Daniel F Gudbjartsson; Agnar Helgason; Hakon Jonsson; Olafur T Magnusson; Pall Melsted; Gudmundur L Norddahl; Jona Saemundsdottir; Asgeir Sigurdsson; Patrick Sulem; Arna B Agustsdottir; Berglind Eiriksdottir; Run Fridriksdottir; Elisabet E Gardarsdottir; Gudmundur Georgsson; Olafia S Gretarsdottir; Kjartan R Gudmundsson; Thora R Gunnarsdottir; Arnaldur Gylfason; Hilma Holm; Brynjar O Jensson; Aslaug Jonasdottir; Kamilla S Josefsdottir; Thordur Kristjansson; Droplaug N Magnusdottir; Louise le Roux; Gudrun Sigmundsdottir; Gardar Sveinbjornsson; Kristin E Sveinsdottir; Maney Sveinsdottir; Emil A Thorarensen; Bjarni Thorbjornsson; Gisli Masson; Ingileif Jonsdottir; Alma Moller; Thorolfur Gudnason; Karl G Kristinsson; Unnur Thorsteinsdottir; Kari Stefansson
EPI_ISL_417581 hCoV-19/Iceland/253/2020 Europe / Iceland / Reykjavik 2020-03-17 The National University Hospital of Iceland deCODE genetics Daniel F Gudbjartsson; Agnar Helgason; Hakon Jonsson; Olafur T Magnusson; Pall Melsted; Gudmundur L Norddahl; Jona Saemundsdottir; Asgeir Sigurdsson; Patrick Sulem; Arna B Agustsdottir; Berglind Eiriksdottir; Run Fridriksdottir; Elisabet E Gardarsdottir; Gudmundur Georgsson; Olafia S Gretarsdottir; Kjartan R Gudmundsson; Thora R Gunnarsdottir; Arnaldur Gylfason; Hilma Holm; Brynjar O Jensson; Aslaug Jonasdottir; Kamilla S Josefsdottir; Thordur Kristjansson; Droplaug N Magnusdottir; Louise le Roux; Gudrun Sigmundsdottir; Gardar Sveinbjornsson; Kristin E Sveinsdottir; Maney Sveinsdottir; Emil A Thorarensen; Bjarni Thorbjornsson; Gisli Masson; Ingileif Jonsdottir; Alma Moller; Thorolfur Gudnason; Karl G Kristinsson; Unnur Thorsteinsdottir; Kari Stefansson
EPI_ISL_417582 hCoV-19/Iceland/254/2020 Europe / Iceland / Reykjavik 2020-03-17 The National University Hospital of Iceland deCODE genetics Daniel F Gudbjartsson; Agnar Helgason; Hakon Jonsson; Olafur T Magnusson; Pall Melsted; Gudmundur L Norddahl; Jona Saemundsdottir; Asgeir Sigurdsson; Patrick Sulem; Arna B Agustsdottir; Berglind Eiriksdottir; Run Fridriksdottir; Elisabet E Gardarsdottir; Gudmundur Georgsson; Olafia S Gretarsdottir; Kjartan R Gudmundsson; Thora R Gunnarsdottir; Arnaldur Gylfason; Hilma Holm; Brynjar O Jensson; Aslaug Jonasdottir; Kamilla S Josefsdottir; Thordur Kristjansson; Droplaug N Magnusdottir; Louise le Roux; Gudrun Sigmundsdottir; Gardar Sveinbjornsson; Kristin E Sveinsdottir; Maney Sveinsdottir; Emil A Thorarensen; Bjarni Thorbjornsson; Gisli Masson; Ingileif Jonsdottir; Alma Moller; Thorolfur Gudnason; Karl G Kristinsson; Unnur Thorsteinsdottir; Kari Stefansson
EPI_ISL_417583 hCoV-19/Iceland/255/2020 Europe / Iceland / Reykjavik 2020-03-17 The National University Hospital of Iceland deCODE genetics Daniel F Gudbjartsson; Agnar Helgason; Hakon Jonsson; Olafur T Magnusson; Pall Melsted; Gudmundur L Norddahl; Jona Saemundsdottir; Asgeir Sigurdsson; Patrick Sulem; Arna B Agustsdottir; Berglind Eiriksdottir; Run Fridriksdottir; Elisabet E Gardarsdottir; Gudmundur Georgsson; Olafia S Gretarsdottir; Kjartan R Gudmundsson; Thora R Gunnarsdottir; Arnaldur Gylfason; Hilma Holm; Brynjar O Jensson; Aslaug Jonasdottir; Kamilla S Josefsdottir; Thordur Kristjansson; Droplaug N Magnusdottir; Louise le Roux; Gudrun Sigmundsdottir; Gardar Sveinbjornsson; Kristin E Sveinsdottir; Maney Sveinsdottir; Emil A Thorarensen; Bjarni Thorbjornsson; Gisli Masson; Ingileif Jonsdottir; Alma Moller; Thorolfur Gudnason; Karl G Kristinsson; Unnur Thorsteinsdottir; Kari Stefansson
EPI_ISL_417584 hCoV-19/Iceland/256/2020 Europe / Iceland / Reykjavik 2020-03-17 The National University Hospital of Iceland deCODE genetics Daniel F Gudbjartsson; Agnar Helgason; Hakon Jonsson; Olafur T Magnusson; Pall Melsted; Gudmundur L Norddahl; Jona Saemundsdottir; Asgeir Sigurdsson; Patrick Sulem; Arna B Agustsdottir; Berglind Eiriksdottir; Run Fridriksdottir; Elisabet E Gardarsdottir; Gudmundur Georgsson; Olafia S Gretarsdottir; Kjartan R Gudmundsson; Thora R Gunnarsdottir; Arnaldur Gylfason; Hilma Holm; Brynjar O Jensson; Aslaug Jonasdottir; Kamilla S Josefsdottir; Thordur Kristjansson; Droplaug N Magnusdottir; Louise le

Roux; Gudrun Sigmundsdottir; Gardar Sveinbjornsson; Kristin E Sveinsdottir; Maney Sveinsdottir; Emil A Thorarensen; Bjarni Thorbjornsson; Gisli Masson; Ingileif Jonsdottir; Alma Moller; Thorolfur Gudnason; Karl G Kristinsson; Unnur Thorsteinsdottir; Kari Stefansson
EPI_ISL_417585 hCoV-19/Iceland/257/2020 Europe / Iceland / Reykjavik 2020-03-17 The National University Hospital of Iceland deCODE genetics Daniel F Gudbjartsson; Agnar Helgason; Hakon Jonsson; Olafur T Magnusson; Pall Melsted; Gudmundur L Norddahl; Jona Saemundsdottir; Asgeir Sigurdsson; Patrick Sulem; Arna B Agustsdottir; Berglind Eiriksdottir; Run Fridriksdottir; Elisabet E Gardarsdottir; Gudmundur Georgsson; Olafia S Gretarsdottir; Kjartan R Gudmundsson; Thora R Gunnarsdottir; Arnaldur Gylfason; Hilma Holm; Brynjar O Jensson; Aslaug Jonasdottir; Kamilla S Josefsdottir; Thordur Kristjansson; Droplaug N Magnusdottir; Louise le Roux; Gudrun Sigmundsdottir; Gardar Sveinbjornsson; Kristin E Sveinsdottir; Maney Sveinsdottir; Emil A Thorarensen; Bjarni Thorbjornsson; Gisli Masson; Ingileif Jonsdottir; Alma Moller; Thorolfur Gudnason; Karl G Kristinsson; Unnur Thorsteinsdottir; Kari Stefansson
EPI_ISL_417586 hCoV-19/Iceland/258/2020 Europe / Iceland / Reykjavik 2020-03-17 The National University Hospital of Iceland deCODE genetics Daniel F Gudbjartsson; Agnar Helgason; Hakon Jonsson; Olafur T Magnusson; Pall Melsted; Gudmundur L Norddahl; Jona Saemundsdottir; Asgeir Sigurdsson; Patrick Sulem; Arna B Agustsdottir; Berglind Eiriksdottir; Run Fridriksdottir; Elisabet E Gardarsdottir; Gudmundur Georgsson; Olafia S Gretarsdottir; Kjartan R Gudmundsson; Thora R Gunnarsdottir; Arnaldur Gylfason; Hilma Holm; Brynjar O Jensson; Aslaug Jonasdottir; Kamilla S Josefsdottir; Thordur Kristjansson; Droplaug N Magnusdottir; Louise le Roux; Gudrun Sigmundsdottir; Gardar Sveinbjornsson; Kristin E Sveinsdottir; Maney Sveinsdottir; Emil A Thorarensen; Bjarni Thorbjornsson; Gisli Masson; Ingileif Jonsdottir; Alma Moller; Thorolfur Gudnason; Karl G Kristinsson; Unnur Thorsteinsdottir; Kari Stefansson
EPI_ISL_417587 hCoV-19/Iceland/259/2020 Europe / Iceland / Reykjavik 2020-03-17 The National University Hospital of Iceland deCODE genetics Daniel F Gudbjartsson; Agnar Helgason; Hakon Jonsson; Olafur T Magnusson; Pall Melsted; Gudmundur L Norddahl; Jona Saemundsdottir; Asgeir Sigurdsson; Patrick Sulem; Arna B Agustsdottir; Berglind Eiriksdottir; Run Fridriksdottir; Elisabet E Gardarsdottir; Gudmundur Georgsson; Olafia S Gretarsdottir; Kjartan R Gudmundsson; Thora R Gunnarsdottir; Arnaldur Gylfason; Hilma Holm; Brynjar O Jensson; Aslaug Jonasdottir; Kamilla S Josefsdottir; Thordur Kristjansson; Droplaug N Magnusdottir; Louise le Roux; Gudrun Sigmundsdottir; Gardar Sveinbjornsson; Kristin E Sveinsdottir; Maney Sveinsdottir; Emil A Thorarensen; Bjarni Thorbjornsson; Gisli Masson; Ingileif Jonsdottir; Alma Moller; Thorolfur Gudnason; Karl G Kristinsson; Unnur Thorsteinsdottir; Kari Stefansson
EPI_ISL_417588 hCoV-19/Iceland/260/2020 Europe / Iceland / Reykjavik 2020-03-17 The National University Hospital of Iceland deCODE genetics Daniel F Gudbjartsson; Agnar Helgason; Hakon Jonsson; Olafur T Magnusson; Pall Melsted; Gudmundur L Norddahl; Jona Saemundsdottir; Asgeir Sigurdsson; Patrick Sulem; Arna B Agustsdottir; Berglind Eiriksdottir; Run Fridriksdottir; Elisabet E Gardarsdottir; Gudmundur Georgsson; Olafia S Gretarsdottir; Kjartan R Gudmundsson; Thora R Gunnarsdottir; Arnaldur Gylfason; Hilma Holm; Brynjar O Jensson; Aslaug Jonasdottir; Kamilla S Josefsdottir; Thordur Kristjansson; Droplaug N Magnusdottir; Louise le Roux; Gudrun Sigmundsdottir; Gardar Sveinbjornsson; Kristin E Sveinsdottir; Maney Sveinsdottir; Emil A Thorarensen; Bjarni Thorbjornsson; Gisli Masson; Ingileif Jonsdottir; Alma Moller; Thorolfur Gudnason; Karl G Kristinsson; Unnur Thorsteinsdottir; Kari Stefansson
EPI_ISL_417589 hCoV-19/Iceland/261/2020 Europe / Iceland / Reykjavik 2020-03-17 The National University Hospital of Iceland deCODE genetics Daniel F Gudbjartsson; Agnar Helgason; Hakon Jonsson; Olafur T Magnusson; Pall Melsted; Gudmundur L Norddahl; Jona Saemundsdottir; Asgeir Sigurdsson; Patrick Sulem; Arna B Agustsdottir; Berglind Eiriksdottir; Run Fridriksdottir; Elisabet E Gardarsdottir; Gudmundur Georgsson; Olafia S Gretarsdottir; Kjartan R Gudmundsson; Thora R Gunnarsdottir; Arnaldur Gylfason; Hilma Holm; Brynjar O Jensson; Aslaug Jonasdottir; Kamilla S Josefsdottir; Thordur Kristjansson; Droplaug N Magnusdottir; Louise le Roux; Gudrun Sigmundsdottir; Gardar Sveinbjornsson; Kristin E Sveinsdottir; Maney Sveinsdottir; Emil A Thorarensen; Bjarni Thorbjornsson; Gisli Masson; Ingileif Jonsdottir; Alma Moller; Thorolfur Gudnason; Karl G Kristinsson; Unnur Thorsteinsdottir; Kari Stefansson
EPI_ISL_417590 hCoV-19/Iceland/262/2020 Europe / Iceland / Reykjavik 2020-03-18 deCODE genetics deCODE genetics Daniel F Gudbjartsson; Agnar Helgason; Hakon Jonsson; Olafur T Magnusson; Pall Melsted; Gudmundur L Norddahl; Jona Saemundsdottir; Asgeir Sigurdsson; Patrick Sulem; Arna B Agustsdottir; Berglind Eiriksdottir; Run Fridriksdottir; Elisabet E Gardarsdottir; Gudmundur Georgsson; Olafia S Gretarsdottir; Kjartan R Gudmundsson; Thora R Gunnarsdottir; Arnaldur Gylfason; Hilma Holm; Brynjar O Jensson; Aslaug Jonasdottir; Kamilla S Josefsdottir; Thordur Kristjansson; Droplaug N Magnusdottir; Louise le Roux; Gudrun Sigmundsdottir; Gardar Sveinbjornsson; Kristin E Sveinsdottir; Maney Sveinsdottir; Emil A Thorarensen; Bjarni Thorbjornsson; Gisli Masson; Ingileif Jonsdottir; Alma Moller; Thorolfur Gudnason; Karl G Kristinsson; Unnur Thorsteinsdottir; Kari Stefansson
EPI_ISL_417591 hCoV-19/Iceland/263/2020 Europe / Iceland / Reykjavik 2020-03-17 The National University Hospital of Iceland deCODE genetics Daniel F Gudbjartsson; Agnar Helgason; Hakon Jonsson; Olafur T Magnusson; Pall Melsted; Gudmundur L Norddahl; Jona Saemundsdottir; Asgeir Sigurdsson; Patrick Sulem; Arna B Agustsdottir; Berglind Eiriksdottir; Run Fridriksdottir; Elisabet E Gardarsdottir; Gudmundur Georgsson; Olafia S Gretarsdottir; Kjartan R Gudmundsson; Thora R Gunnarsdottir; Arnaldur Gylfason; Hilma Holm; Brynjar O Jensson; Aslaug Jonasdottir; Kamilla S Josefsdottir; Thordur Kristjansson; Droplaug N Magnusdottir; Louise le Roux; Gudrun Sigmundsdottir; Gardar Sveinbjornsson; Kristin E Sveinsdottir; Maney Sveinsdottir; Emil A Thorarensen; Bjarni Thorbjornsson; Gisli Masson; Ingileif Jonsdottir; Alma Moller; Thorolfur Gudnason; Karl G Kristinsson; Unnur Thorsteinsdottir; Kari Stefansson
EPI_ISL_417592 hCoV-19/Iceland/264/2020 Europe / Iceland / Reykjavik 2020-03-17 The National University Hospital of Iceland deCODE genetics Daniel F Gudbjartsson; Agnar Helgason; Hakon Jonsson; Olafur T Magnusson; Pall Melsted; Gudmundur L Norddahl; Jona Saemundsdottir; Asgeir Sigurdsson; Patrick Sulem; Arna B Agustsdottir; Berglind Eiriksdottir; Run Fridriksdottir; Elisabet E Gardarsdottir; Gudmundur Georgsson; Olafia

S Gretarsdottir; Kjartan R Gudmundsson; Thora R Gunnarsdottir; Arnaldur Gylfason; Hilma Holm; Brynjar O Jensson; Aslaug Jonasdottir; Kamilla S Josefsdottir; Thordur Kristjansson; Droplaug N Magnusdottir; Louise le Roux; Gudrun Sigmundsdottir; Gardar Sveinbjornsson; Kristin E Sveinsdottir; Maney Sveinsdottir; Emil A Thorarensen; Bjarni Thorbjornsson; Gisli Masson; Ingileif Jonsdottir; Alma Moller; Thorolfur Gudnason; Karl G Kristinsson; Unnur Thorsteinsdottir; Kari Stefansson
EPI_ISL_417595 hCoV-19/Iceland/267/2020 Europe / Iceland / Reykjavik 2020-03-17 The National University Hospital of Iceland deCODE genetics Daniel F Gudbjartsson; Agnar Helgason; Hakon Jonsson; Olafur T Magnusson; Pall Melsted; Gudmundur L Norddahl; Jona Saemundsdottir; Asgeir Sigurdsson; Patrick Sulem; Arna B Agustsdottir; Berglind Eiriksdottir; Run Fridriksdottir; Elisabet E Gardarsdottir; Gudmundur Georgsson; Olafia S Gretarsdottir; Kjartan R Gudmundsson; Thora R Gunnarsdottir; Arnaldur Gylfason; Hilma Holm; Brynjar O Jensson; Aslaug Jonasdottir; Kamilla S Josefsdottir; Thordur Kristjansson; Droplaug N Magnusdottir; Louise le Roux; Gudrun Sigmundsdottir; Gardar Sveinbjornsson; Kristin E Sveinsdottir; Maney Sveinsdottir; Emil A Thorarensen; Bjarni Thorbjornsson; Gisli Masson; Ingileif Jonsdottir; Alma Moller; Thorolfur Gudnason; Karl G Kristinsson; Unnur Thorsteinsdottir; Kari Stefansson
EPI_ISL_417597 hCoV-19/Iceland/269/2020 Europe / Iceland / Reykjavik 2020-03-17 The National University Hospital of Iceland deCODE genetics Daniel F Gudbjartsson; Agnar Helgason; Hakon Jonsson; Olafur T Magnusson; Pall Melsted; Gudmundur L Norddahl; Jona Saemundsdottir; Asgeir Sigurdsson; Patrick Sulem; Arna B Agustsdottir; Berglind Eiriksdottir; Run Fridriksdottir; Elisabet E Gardarsdottir; Gudmundur Georgsson; Olafia S Gretarsdottir; Kjartan R Gudmundsson; Thora R Gunnarsdottir; Arnaldur Gylfason; Hilma Holm; Brynjar O Jensson; Aslaug Jonasdottir; Kamilla S Josefsdottir; Thordur Kristjansson; Droplaug N Magnusdottir; Louise le Roux; Gudrun Sigmundsdottir; Gardar Sveinbjornsson; Kristin E Sveinsdottir; Maney Sveinsdottir; Emil A Thorarensen; Bjarni Thorbjornsson; Gisli Masson; Ingileif Jonsdottir; Alma Moller; Thorolfur Gudnason; Karl G Kristinsson; Unnur Thorsteinsdottir; Kari Stefansson
EPI_ISL_417598 hCoV-19/Iceland/270/2020 Europe / Iceland / Reykjavik 2020-03-17 The National University Hospital of Iceland deCODE genetics Daniel F Gudbjartsson; Agnar Helgason; Hakon Jonsson; Olafur T Magnusson; Pall Melsted; Gudmundur L Norddahl; Jona Saemundsdottir; Asgeir Sigurdsson; Patrick Sulem; Arna B Agustsdottir; Berglind Eiriksdottir; Run Fridriksdottir; Elisabet E Gardarsdottir; Gudmundur Georgsson; Olafia S Gretarsdottir; Kjartan R Gudmundsson; Thora R Gunnarsdottir; Arnaldur Gylfason; Hilma Holm; Brynjar O Jensson; Aslaug Jonasdottir; Kamilla S Josefsdottir; Thordur Kristjansson; Droplaug N Magnusdottir; Louise le Roux; Gudrun Sigmundsdottir; Gardar Sveinbjornsson; Kristin E Sveinsdottir; Maney Sveinsdottir; Emil A Thorarensen; Bjarni Thorbjornsson; Gisli Masson; Ingileif Jonsdottir; Alma Moller; Thorolfur Gudnason; Karl G Kristinsson; Unnur Thorsteinsdottir; Kari Stefansson
EPI_ISL_418302 hCoV-19/England/SHEF-BFE9D/2020 Europe / United Kingdom / England / South Yorkshire 2020-03-24 Virology Department, Sheffield Teaching Hospitals NHS Foundation Trust Department of Infection, Immunity and Cardiovascular Disease, The Florey Institute, The Medical School, University of Sheffield Thushan de Silva, Matthew Parker, Adri Angyal, Rebecca Brown, Rachel Tucker, Paul Parsons, Danielle Groves, Alex Keeley, Dave Partridge, Matthew Wyles, Benjamin Lindsey, Mehmet Yavuz, Mohammad Raza, Cariad Evans
EPI_ISL_418303 hCoV-19/England/SHEF-BFEAC/2020 Europe / United Kingdom / England / Derbyshire 2020-03-25 Virology Department, Sheffield Teaching Hospitals NHS Foundation Trust Department of Infection, Immunity and Cardiovascular Disease, The Florey Institute, The Medical School, University of Sheffield Thushan de Silva, Matthew Parker, Adri Angyal, Rebecca Brown, Rachel Tucker, Paul Parsons, Danielle Groves, Alex Keeley, Dave Partridge, Matthew Wyles, Benjamin Lindsey, Mehmet Yavuz, Mohammad Raza, Cariad Evans
EPI_ISL_418304 hCoV-19/England/SHEF-BFEBB/2020 Europe / United Kingdom / England / Derbyshire 2020-03-24 Virology Department, Sheffield Teaching Hospitals NHS Foundation Trust Department of Infection, Immunity and Cardiovascular Disease, The Florey Institute, The Medical School, University of Sheffield Thushan de Silva, Matthew Parker, Adri Angyal, Rebecca Brown, Rachel Tucker, Paul Parsons, Danielle Groves, Alex Keeley, Dave Partridge, Matthew Wyles, Benjamin Lindsey, Mehmet Yavuz, Mohammad Raza, Cariad Evans
EPI_ISL_418305 hCoV-19/England/SHEF-BFED9/2020 Europe / United Kingdom / England / South Yorkshire 2020-03-25 Virology Department, Sheffield Teaching Hospitals NHS Foundation Trust Department of Infection, Immunity and Cardiovascular Disease, The Florey Institute, The Medical School, University of Sheffield Thushan de Silva, Matthew Parker, Adri Angyal, Rebecca Brown, Rachel Tucker, Paul Parsons, Danielle Groves, Alex Keeley, Dave Partridge, Matthew Wyles, Benjamin Lindsey, Mehmet Yavuz, Mohammad Raza, Cariad Evans
EPI_ISL_418306 hCoV-19/England/SHEF-BFEE8/2020 Europe / United Kingdom / England / South Yorkshire 2020-03-25 Virology Department, Sheffield Teaching Hospitals NHS Foundation Trust Department of Infection, Immunity and Cardiovascular Disease, The Florey Institute, The Medical School, University of Sheffield Thushan de Silva, Matthew Parker, Adri Angyal, Rebecca Brown, Rachel Tucker, Paul Parsons, Danielle Groves, Alex Keeley, Dave Partridge, Matthew Wyles, Benjamin Lindsey, Mehmet Yavuz, Mohammad Raza, Cariad Evans
EPI_ISL_418307 hCoV-19/England/SHEF-BFF03/2020 Europe / United Kingdom / England / Derbyshire 2020-03-24 Virology Department, Sheffield Teaching Hospitals NHS Foundation Trust Department of Infection, Immunity and Cardiovascular Disease, The Florey Institute, The Medical School, University of Sheffield Thushan de Silva, Matthew Parker, Adri Angyal, Rebecca Brown, Rachel Tucker, Paul Parsons, Danielle Groves, Alex Keeley, Dave Partridge, Matthew Wyles, Benjamin Lindsey, Mehmet Yavuz, Mohammad Raza, Cariad Evans
EPI_ISL_418313 hCoV-19/England/SHEF-BFF6D/2020 Europe / United Kingdom / England / Derbyshire 2020-03-24 Virology Department, Sheffield Teaching Hospitals NHS Foundation Trust Department of Infection, Immunity and Cardiovascular Disease, The Florey Institute, The Medical School, University of Sheffield Thushan de Silva, Matthew Parker, Adri Angyal, Rebecca Brown, Rachel Tucker, Paul Parsons, Danielle Groves, Alex Keeley, Dave Partridge, Matthew Wyles, Benjamin Lindsey, Mehmet Yavuz, Mohammad Raza, Cariad Evans
EPI_ISL_418317 hCoV-19/England/SHEF-BFFA9/2020 Europe / United Kingdom / England / South Yorkshire 2020-03-23 Virology Department, Sheffield Teaching Hospitals NHS Foundation Trust Department of Infection, Immunity and Cardiovascular Disease, The Florey Institute, The Medical School, University of Sheffield Thushan

de Silva, Matthew Parker, Adri Angyal, Rebecca Brown, Rachel Tucker, Paul Parsons, Danielle Groves, Alex Keeley, Dave Partridge, Matthew Wyles, Benjamin Lindsey, Mehmet Yavuz, Mohammad Raza, Cariad Evans
EPI_ISL_418318 hCoV-19/England/SHEF-BFFB8/2020 Europe / United Kingdom / England / South Yorkshire 2020-03-25 Virology Department, Sheffield Teaching Hospitals NHS Foundation Trust Department of Infection, Immunity and Cardiovascular Disease, The Florey Institute, The Medical School, University of Sheffield Thushan de Silva, Matthew Parker, Adri Angyal, Rebecca Brown, Rachel Tucker, Paul Parsons, Danielle Groves, Alex Keeley, Dave Partridge, Matthew Wyles, Benjamin Lindsey, Mehmet Yavuz, Mohammad Raza, Cariad Evans
EPI_ISL_418320 hCoV-19/England/SHEF-BFFD6/2020 Europe / United Kingdom / England / South Yorkshire 2020-03-25 Virology Department, Sheffield Teaching Hospitals NHS Foundation Trust Department of Infection, Immunity and Cardiovascular Disease, The Florey Institute, The Medical School, University of Sheffield Thushan de Silva, Matthew Parker, Adri Angyal, Rebecca Brown, Rachel Tucker, Paul Parsons, Danielle Groves, Alex Keeley, Dave Partridge, Matthew Wyles, Benjamin Lindsey, Mehmet Yavuz, Mohammad Raza, Cariad Evans
EPI_ISL_418322 hCoV-19/Canada/ON_PHL3917/2020 North America / Canada / Ontario 2020-03-08 Public Health Ontario Laboratories Public Health Ontario Laboratories Alireza Eshaghi, Samir N Patel, Jonathan B Gubbay, Vanessa G Allen, Christine Frantz, Aimin Li, Sandeep Nagra
EPI_ISL_418323 hCoV-19/Canada/ON_PHL3919/2020 North America / Canada / Ontario 2020-03-14 Public Health Ontario Laboratories Public Health Ontario Laboratories Alireza Eshaghi, Samir N Patel, Jonathan B Gubbay, Vanessa G Allen, Christine Frantz, Aimin Li, Sandeep Nagra
EPI_ISL_418324 hCoV-19/Canada/ON_PHL3318/2020 North America / Canada / Ontario 2020-03-11 Public Health Ontario Laboratories Public Health Ontario Laboratories Alireza Eshaghi, Samir N Patel, Jonathan B Gubbay, Vanessa G Allen, Christine Frantz, Aimin Li, Sandeep Nagra
EPI_ISL_418325 hCoV-19/Canada/ON_PHL5472/2020 North America / Canada / Ontario 2020-02-20 Public Health Ontario Laboratories Public Health Ontario Laboratories Alireza Eshaghi, Samir N Patel, Jonathan B Gubbay, Vanessa G Allen, Christine Frantz, Aimin Li, Sandeep Nagra
EPI_ISL_418326 hCoV-19/Canada/ON_PHL7972/2020 North America / Canada / Ontario 2020-03-12 Public Health Ontario Laboratories Public Health Ontario Laboratories Alireza Eshaghi, Samir N Patel, Jonathan B Gubbay, Vanessa G Allen, Christine Frantz, Aimin Li, Sandeep Nagra
EPI_ISL_418327 hCoV-19/Canada/ON_PHL4181/2020 North America / Canada / Ontario 2020-01-25 Public Health Ontario Laboratories Public Health Ontario Laboratories Alireza Eshaghi, Samir N Patel, Jonathan B Gubbay, Vanessa G Allen, Christine Frantz, Aimin Li, Sandeep Nagra
EPI_ISL_418328 hCoV-19/Canada/ON_PHL3670/2020 North America / Canada / Ontario 2020-03-20 Public Health Ontario Laboratories Public Health Ontario Laboratories Alireza Eshaghi, Samir N Patel, Jonathan B Gubbay, Vanessa G Allen, Christine Frantz, Aimin Li, Sandeep Nagra
EPI_ISL_418329 hCoV-19/Canada/ON_PHL5694/2020 North America / Canada / Ontario 2020-03-12 Public Health Ontario Laboratories Public Health Ontario Laboratories Alireza Eshaghi, Samir N Patel, Jonathan B Gubbay, Vanessa G Allen, Christine Frantz, Aimin Li, Sandeep Nagra
EPI_ISL_418330 hCoV-19/Canada/ON_PHL8580/2020 North America / Canada / Ontario 2020-03-05 Public Health Ontario Laboratories Public Health Ontario Laboratories Alireza Eshaghi, Samir N Patel, Jonathan B Gubbay, Vanessa G Allen, Christine Frantz, Aimin Li, Sandeep Nagra
EPI_ISL_418331 hCoV-19/Canada/ON_PHL6883/2020 North America / Canada / Ontario 2020-03-08 Public Health Ontario Laboratories Public Health Ontario Laboratories Alireza Eshaghi, Samir N Patel, Jonathan B Gubbay, Vanessa G Allen, Christine Frantz, Aimin Li, Sandeep Nagra
EPI_ISL_418332 hCoV-19/Canada/ON_PHL0052/2020 North America / Canada / Ontario 2020-03-11 Public Health Ontario Laboratories Public Health Ontario Laboratories Alireza Eshaghi, Samir N Patel, Jonathan B Gubbay, Vanessa G Allen, Christine Frantz, Aimin Li, Sandeep Nagra
EPI_ISL_418333 hCoV-19/Canada/ON_PHL3501/2020 North America / Canada / Ontario 2020-03-10 Public Health Ontario Laboratories Public Health Ontario Laboratories Alireza Eshaghi, Samir N Patel, Jonathan B Gubbay, Vanessa G Allen, Christine Frantz, Aimin Li, Sandeep Nagra
EPI_ISL_418334 hCoV-19/Canada/ON_PHL4069/2020 North America / Canada / Ontario 2020-03-07 Public Health Ontario Laboratories Public Health Ontario Laboratories Alireza Eshaghi, Samir N Patel, Jonathan B Gubbay, Vanessa G Allen, Christine Frantz, Aimin Li, Sandeep Nagra
EPI_ISL_418335 hCoV-19/Canada/ON_PHL4464/2020 North America / Canada / Ontario 2020-03-14 Public Health Ontario Laboratories Public Health Ontario Laboratories Alireza Eshaghi, Samir N Patel, Jonathan B Gubbay, Vanessa G Allen, Christine Frantz, Aimin Li, Sandeep Nagra
EPI_ISL_418336 hCoV-19/Canada/ON_PHL5705/2020 North America / Canada / Ontario 2020-03-12 Public Health Ontario Laboratories Public Health Ontario Laboratories Alireza Eshaghi, Samir N Patel, Jonathan B Gubbay, Vanessa G Allen, Christine Frantz, Aimin Li, Sandeep Nagra
EPI_ISL_418337 hCoV-19/Canada/ON_PHL3802/2020 North America / Canada / Ontario 2020-03-12 Public Health Ontario Laboratories Public Health Ontario Laboratories Alireza Eshaghi, Samir N Patel, Jonathan B Gubbay, Vanessa G Allen, Christine Frantz, Aimin Li, Sandeep Nagra
EPI_ISL_418338 hCoV-19/Canada/ON_PHL5710/2020 North America / Canada / Ontario 2020-03-12 Public Health Ontario Laboratories Public Health Ontario Laboratories Alireza Eshaghi, Samir N Patel, Jonathan B Gubbay, Vanessa G Allen, Christine Frantz, Aimin Li, Sandeep Nagra
EPI_ISL_418339 hCoV-19/Canada/ON_PHL3877/2020 North America / Canada / Ontario 2020-03-09 Public Health Ontario Laboratories Public Health Ontario Laboratories Alireza Eshaghi, Samir N Patel, Jonathan B Gubbay, Vanessa G Allen, Christine Frantz, Aimin Li, Sandeep Nagra
EPI_ISL_418340 hCoV-19/Canada/ON_PHL1083/2020 North America / Canada / Ontario 2020-03-09 Public Health Ontario Laboratories Public Health Ontario Laboratories Alireza Eshaghi, Samir N Patel, Jonathan B Gubbay, Vanessa G Allen, Christine Frantz, Aimin Li, Sandeep Nagra
EPI_ISL_418341 hCoV-19/Canada/ON_PHL0142/2020 North America / Canada / Ontario 2020-03-12 Public

Health Ontario Laboratories	Public Health Ontario Laboratories	Alireza Eshaghi, Samir N Patel, Jonathan B Gubbay, Vanessa G Allen, Christine Frantz, Aimin Li, Sandeep Nagra
EPI_ISL_418342	hCoV-19/Canada/ON_PHL0178/2020	North America / Canada / Ontario	2020-03-11	Public Health Ontario Laboratories	Public Health Ontario Laboratories	Alireza Eshaghi, Samir N Patel, Jonathan B Gubbay, Vanessa G Allen, Christine Frantz, Aimin Li, Sandeep Nagra
EPI_ISL_418343	hCoV-19/Canada/ON_PHL6884/2020	North America / Canada / Ontario	2020-03-10	Public Health Ontario Laboratories	Public Health Ontario Laboratories	Alireza Eshaghi, Samir N Patel, Jonathan B Gubbay, Vanessa G Allen, Christine Frantz, Aimin Li, Sandeep Nagra
EPI_ISL_418344	hCoV-19/Canada/ON_PHL2259/2020	North America / Canada / Ontario	2020-02-29	Public Health Ontario Laboratories	Public Health Ontario Laboratories	Alireza Eshaghi, Samir N Patel, Jonathan B Gubbay, Vanessa G Allen, Christine Frantz, Aimin Li, Sandeep Nagra
EPI_ISL_418345	hCoV-19/Canada/ON_PHL8751/2020	North America / Canada / Ontario	2020-02-29	Public Health Ontario Laboratories	Public Health Ontario Laboratories	Alireza Eshaghi, Samir N Patel, Jonathan B Gubbay, Vanessa G Allen, Christine Frantz, Aimin Li, Sandeep Nagra
EPI_ISL_418346	hCoV-19/Canada/ON_PHL0743/2020	North America / Canada / Ontario	2020-03-07	Public Health Ontario Laboratories	Public Health Ontario Laboratories	Alireza Eshaghi, Samir N Patel, Jonathan B Gubbay, Vanessa G Allen, Christine Frantz, Aimin Li, Sandeep Nagra
EPI_ISL_418347	hCoV-19/Canada/ON_PHL3741/2020	North America / Canada / Ontario	2020-03-11	Public Health Ontario Laboratories	Public Health Ontario Laboratories	Alireza Eshaghi, Samir N Patel, Jonathan B Gubbay, Vanessa G Allen, Christine Frantz, Aimin Li, Sandeep Nagra
EPI_ISL_418348	hCoV-19/Canada/ON_PHL3680/2020	North America / Canada / Ontario	2020-03-08	Public Health Ontario Laboratories	Public Health Ontario Laboratories	Alireza Eshaghi, Samir N Patel, Jonathan B Gubbay, Vanessa G Allen, Christine Frantz, Aimin Li, Sandeep Nagra
EPI_ISL_418349	hCoV-19/Canada/ON_PHL3650/2020	North America / Canada / Ontario	2020-03-07	Public Health Ontario Laboratories	Public Health Ontario Laboratories	Alireza Eshaghi, Samir N Patel, Jonathan B Gubbay, Vanessa G Allen, Christine Frantz, Aimin Li, Sandeep Nagra
EPI_ISL_418350	hCoV-19/Canada/ON_PHL3380/2020	North America / Canada / Ontario	2020-03-20	Public Health Ontario Laboratories	Public Health Ontario Laboratories	Alireza Eshaghi, Samir N Patel, Jonathan B Gubbay, Vanessa G Allen, Christine Frantz, Aimin Li, Sandeep Nagra
EPI_ISL_418351	hCoV-19/Canada/ON_PHL3575/2020	North America / Canada / Ontario	2020-03-12	Public Health Ontario Laboratories	Public Health Ontario Laboratories	Alireza Eshaghi, Samir N Patel, Jonathan B Gubbay, Vanessa G Allen, Christine Frantz, Aimin Li, Sandeep Nagra
EPI_ISL_418352	hCoV-19/Canada/ON_PHL5672/2020	North America / Canada / Ontario	2020-03-11	Public Health Ontario Laboratories	Public Health Ontario Laboratories	Alireza Eshaghi, Samir N Patel, Jonathan B Gubbay, Vanessa G Allen, Christine Frantz, Aimin Li, Sandeep Nagra
EPI_ISL_418353	hCoV-19/Canada/ON_PHL0539/2020	North America / Canada / Ontario	2020-03-09	Public Health Ontario Laboratories	Public Health Ontario Laboratories	Alireza Eshaghi, Samir N Patel, Jonathan B Gubbay, Vanessa G Allen, Christine Frantz, Aimin Li, Sandeep Nagra
EPI_ISL_418354	hCoV-19/Canada/ON_PHL4088/2020	North America / Canada / Ontario	2020-03-15	Public Health Ontario Laboratories	Public Health Ontario Laboratories	Alireza Eshaghi, Samir N Patel, Jonathan B Gubbay, Vanessa G Allen, Christine Frantz, Aimin Li, Sandeep Nagra
EPI_ISL_418355	hCoV-19/Canada/ON_PHLU8150/2020	North America / Canada / Ontario	2020-03-08	Public Health Ontario Laboratories	Public Health Ontario Laboratories	Alireza Eshaghi, Samir N Patel, Jonathan B Gubbay, Vanessa G Allen, Christine Frantz, Aimin Li, Sandeep Nagra
EPI_ISL_418356	hCoV-19/Canada/ON_PHL6980/2020	North America / Canada / Ontario	2020-03-12	Public Health Ontario Laboratories	Public Health Ontario Laboratories	Alireza Eshaghi, Samir N Patel, Jonathan B Gubbay, Vanessa G Allen, Christine Frantz, Aimin Li, Sandeep Nagra
EPI_ISL_418357	hCoV-19/Canada/ON_PHLH6415/2020	North America / Canada / Ontario	2020-03-12	Public Health Ontario Laboratories	Public Health Ontario Laboratories	Alireza Eshaghi, Samir N Patel, Jonathan B Gubbay, Vanessa G Allen, Christine Frantz, Aimin Li, Sandeep Nagra
EPI_ISL_418358	hCoV-19/Canada/ON_PHL5756/2020	North America / Canada / Ontario	2020-03-12	Public Health Ontario Laboratories	Public Health Ontario Laboratories	Alireza Eshaghi, Samir N Patel, Jonathan B Gubbay, Vanessa G Allen, Christine Frantz, Aimin Li, Sandeep Nagra
EPI_ISL_418359	hCoV-19/Canada/ON_PHL5757/2020	North America / Canada / Ontario	2020-03-12	Public Health Ontario Laboratories	Public Health Ontario Laboratories	Alireza Eshaghi, Samir N Patel, Jonathan B Gubbay, Vanessa G Allen, Christine Frantz, Aimin Li, Sandeep Nagra
EPI_ISL_418360	hCoV-19/Canada/ON_PHL3692/2020	North America / Canada / Ontario	2020-03-11	Public Health Ontario Laboratories	Public Health Ontario Laboratories	Alireza Eshaghi, Samir N Patel, Jonathan B Gubbay, Vanessa G Allen, Christine Frantz, Aimin Li, Sandeep Nagra
EPI_ISL_418361	hCoV-19/Canada/ON_PHL1095/2020	North America / Canada / Ontario	2020-03-12	Public Health Ontario Laboratories	Public Health Ontario Laboratories	Alireza Eshaghi, Samir N Patel, Jonathan B Gubbay, Vanessa G Allen, Christine Frantz, Aimin Li, Sandeep Nagra
EPI_ISL_418362	hCoV-19/Canada/ON_PHL3536/2020	North America / Canada / Ontario	2020-03-10	Public Health Ontario Laboratories	Public Health Ontario Laboratories	Alireza Eshaghi, Samir N Patel, Jonathan B Gubbay, Vanessa G Allen, Christine Frantz, Aimin Li, Sandeep Nagra
EPI_ISL_418363	hCoV-19/Canada/ON_PHL3695/2020	North America / Canada / Ontario	2020-03-11	Public Health Ontario Laboratories	Public Health Ontario Laboratories	Alireza Eshaghi, Samir N Patel, Jonathan B Gubbay, Vanessa G Allen, Christine Frantz, Aimin Li, Sandeep Nagra
EPI_ISL_418364	hCoV-19/Canada/ON_PHL6922/2020	North America / Canada / Ontario	2020-03-13	Public Health Ontario Laboratories	Public Health Ontario Laboratories	Alireza Eshaghi, Samir N Patel,

Jonathan B Gubbay, Vanessa G Allen, Christine Frantz, Aimin Li, Sandeep Nagra
EPI_ISL_418365 hCoV-19/Canada/ON_PHL0977/2020 North America / Canada / Ontario 2020-03-10 Public Health Ontario Laboratories Public Health Ontario Laboratories Alireza Eshaghi, Samir N Patel, Jonathan B Gubbay, Vanessa G Allen, Christine Frantz, Aimin Li, Sandeep Nagra
EPI_ISL_418366 hCoV-19/Canada/ON_PHL3350/2020 North America / Canada / Ontario 2020-03-05 Public Health Ontario Laboratories Public Health Ontario Laboratories Alireza Eshaghi, Samir N Patel, Jonathan B Gubbay, Vanessa G Allen, Christine Frantz, Aimin Li, Sandeep Nagra
EPI_ISL_418367 hCoV-19/Canada/ON_PHL7513/2020 North America / Canada / Ontario 2020-03-12 Public Health Ontario Laboratories Public Health Ontario Laboratories Alireza Eshaghi, Samir N Patel, Jonathan B Gubbay, Vanessa G Allen, Christine Frantz, Aimin Li, Sandeep Nagra
EPI_ISL_418368 hCoV-19/Canada/ON_PHL3458/2020 North America / Canada / Ontario 2020-03-12 Public Health Ontario Laboratories Public Health Ontario Laboratories Alireza Eshaghi, Samir N Patel, Jonathan B Gubbay, Vanessa G Allen, Christine Frantz, Aimin Li, Sandeep Nagra
EPI_ISL_418369 hCoV-19/Canada/ON_PHL8539/2020 North America / Canada / Ontario 2020-03-10 Public Health Ontario Laboratories Public Health Ontario Laboratories Alireza Eshaghi, Samir N Patel, Jonathan B Gubbay, Vanessa G Allen, Christine Frantz, Aimin Li, Sandeep Nagra
EPI_ISL_418370 hCoV-19/Canada/ON_PHL0654/2020 North America / Canada / Ontario 2020-03-11 Public Health Ontario Laboratories Public Health Ontario Laboratories Alireza Eshaghi, Samir N Patel, Jonathan B Gubbay, Vanessa G Allen, Christine Frantz, Aimin Li, Sandeep Nagra
EPI_ISL_418371 hCoV-19/Canada/ON_PHL2653/2020 North America / Canada / Ontario 2020-03-07 Public Health Ontario Laboratories Public Health Ontario Laboratories Alireza Eshaghi, Samir N Patel, Jonathan B Gubbay, Vanessa G Allen, Christine Frantz, Aimin Li, Sandeep Nagra
EPI_ISL_418372 hCoV-19/Canada/ON_PHL8458/2020 North America / Canada / Ontario 2020-03-12 Public Health Ontario Laboratories Public Health Ontario Laboratories Alireza Eshaghi, Samir N Patel, Jonathan B Gubbay, Vanessa G Allen, Christine Frantz, Aimin Li, Sandeep Nagra
EPI_ISL_418373 hCoV-19/Canada/ON_PHL7590/2020 North America / Canada / Ontario 2020-03-14 Public Health Ontario Laboratories Public Health Ontario Laboratories Alireza Eshaghi, Samir N Patel, Jonathan B Gubbay, Vanessa G Allen, Christine Frantz, Aimin Li, Sandeep Nagra
EPI_ISL_418374 hCoV-19/Canada/ON_PHL4232/2020 North America / Canada / Ontario 2020-03-11 Public Health Ontario Laboratories Public Health Ontario Laboratories Alireza Eshaghi, Samir N Patel, Jonathan B Gubbay, Vanessa G Allen, Christine Frantz, Aimin Li, Sandeep Nagra
EPI_ISL_418375 hCoV-19/Canada/ON_PHL0141/2020 North America / Canada / Ontario 2020-03-12 Public Health Ontario Laboratories Public Health Ontario Laboratories Alireza Eshaghi, Samir N Patel, Jonathan B Gubbay, Vanessa G Allen, Christine Frantz, Aimin Li, Sandeep Nagra
EPI_ISL_418376 hCoV-19/Canada/ON_PHL3459/2020 North America / Canada / Ontario 2020-03-12 Public Health Ontario Laboratories Public Health Ontario Laboratories Alireza Eshaghi, Samir N Patel, Jonathan B Gubbay, Vanessa G Allen, Christine Frantz, Aimin Li, Sandeep Nagra
EPI_ISL_418377 hCoV-19/Canada/ON_PHL1898/2020 North America / Canada / Ontario 2020-03-13 Public Health Ontario Laboratories Public Health Ontario Laboratories Alireza Eshaghi, Samir N Patel, Jonathan B Gubbay, Vanessa G Allen, Christine Frantz, Aimin Li, Sandeep Nagra
EPI_ISL_418378 hCoV-19/Canada/ON_PHL0976/2020 North America / Canada / Ontario 2020-03-13 Public Health Ontario Laboratories Public Health Ontario Laboratories Alireza Eshaghi, Samir N Patel, Jonathan B Gubbay, Vanessa G Allen, Christine Frantz, Aimin Li, Sandeep Nagra
EPI_ISL_418379 hCoV-19/Canada/ON_PHL7512/2020 North America / Canada / Ontario 2020-03-13 Public Health Ontario Laboratories Public Health Ontario Laboratories Alireza Eshaghi, Samir N Patel, Jonathan B Gubbay, Vanessa G Allen, Christine Frantz, Aimin Li, Sandeep Nagra
EPI_ISL_418380 hCoV-19/Canada/ON_PHL3476/2020 North America / Canada / Ontario 2020-03-13 Public Health Ontario Laboratories Public Health Ontario Laboratories Alireza Eshaghi, Samir N Patel, Jonathan B Gubbay, Vanessa G Allen, Christine Frantz, Aimin Li, Sandeep Nagra
EPI_ISL_418381 hCoV-19/Canada/ON_PHL2223/2020 North America / Canada / Ontario 2020-03-19 Public Health Ontario Laboratories Public Health Ontario Laboratories Alireza Eshaghi, Samir N Patel, Jonathan B Gubbay, Vanessa G Allen, Christine Frantz, Aimin Li, Sandeep Nagra
EPI_ISL_418382 hCoV-19/Canada/ON_PHL5930/2020 North America / Canada / Ontario 2020-02-28 Public Health Ontario Laboratories Public Health Ontario Laboratories Alireza Eshaghi, Samir N Patel, Jonathan B Gubbay, Vanessa G Allen, Christine Frantz, Aimin Li, Sandeep Nagra
EPI_ISL_418383 hCoV-19/Canada/ON_PHL2273/2020 North America / Canada / Ontario 2020-02-29 Public Health Ontario Laboratories Public Health Ontario Laboratories Alireza Eshaghi, Samir N Patel, Jonathan B Gubbay, Vanessa G Allen, Christine Frantz, Aimin Li, Sandeep Nagra
EPI_ISL_418385 hCoV-19/Finland/13M19/2020 Europe / Finland 2020-03-13 Department of Virology and Immunology, University of Helsinki and Helsinki University Hospital, Huslab Finland Department of Virology, Faculty of Medicine, University of Helsinki, Helsinki, Finland Teemu Smura, Hannimari Kallio-Kokko, Olli Vapalahti
EPI_ISL_418386 hCoV-19/Finland/13M26/2020 Europe / Finland 2020-03-13 Department of Virology and Immunology, University of Helsinki and Helsinki University Hospital, Huslab Finland Department of Virology, Faculty of Medicine, University of Helsinki, Helsinki, Finland Teemu Smura, Hannimari Kallio-Kokko, Olli Vapalahti
EPI_ISL_418387 hCoV-19/Finland/13M27/2020 Europe / Finland 2020-03-13 Department of Virology and Immunology, University of Helsinki and Helsinki University Hospital, Huslab Finland Department of Virology, Faculty of Medicine, University of Helsinki, Helsinki, Finland Teemu Smura, Hannimari Kallio-Kokko, Olli Vapalahti

EPI_ISL_418388 hCoV-19/Finland/13M29/2020 Europe / Finland 2020-03-13 Department of Virology and Immunology, University of Helsinki and Helsinki University Hospital, Huslab Finland Department of Virology, Faculty of Medicine, University of Helsinki, Helsinki, Finland Teemu Smura, Hannimari Kallio-Kokko, Olli Vapalahti
EPI_ISL_418389 hCoV-19/Finland/13M3/2020 Europe / Finland 2020-03-13 Department of Virology and Immunology, University of Helsinki and Helsinki University Hospital, Huslab Finland Department of Virology, Faculty of Medicine, University of Helsinki, Helsinki, Finland Teemu Smura, Hannimari Kallio-Kokko, Olli Vapalahti
EPI_ISL_418390 hCoV-19/Finland/13M33/2020 Europe / Finland 2020-03-13 Department of Virology and Immunology, University of Helsinki and Helsinki University Hospital, Huslab Finland Department of Virology, Faculty of Medicine, University of Helsinki, Helsinki, Finland Teemu Smura, Hannimari Kallio-Kokko, Olli Vapalahti
EPI_ISL_418391 hCoV-19/Finland/13M57/2020 Europe / Finland 2020-03-13 Department of Virology and Immunology, University of Helsinki and Helsinki University Hospital, Huslab Finland Department of Virology, Faculty of Medicine, University of Helsinki, Helsinki, Finland Teemu Smura, Hannimari Kallio-Kokko, Olli Vapalahti
EPI_ISL_418392 hCoV-19/Finland/13M58/2020 Europe / Finland 2020-03-13 Department of Virology and Immunology, University of Helsinki and Helsinki University Hospital, Huslab Finland Department of Virology, Faculty of Medicine, University of Helsinki, Helsinki, Finland Teemu Smura, Hannimari Kallio-Kokko, Olli Vapalahti
EPI_ISL_418393 hCoV-19/Finland/13M60/2020 Europe / Finland 2020-03-13 Department of Virology and Immunology, University of Helsinki and Helsinki University Hospital, Huslab Finland Department of Virology, Faculty of Medicine, University of Helsinki, Helsinki, Finland Teemu Smura, Hannimari Kallio-Kokko, Olli Vapalahti
EPI_ISL_418394 hCoV-19/Finland/13M64/2020 Europe / Finland 2020-03-13 Department of Virology and Immunology, University of Helsinki and Helsinki University Hospital, Huslab Finland Department of Virology, Faculty of Medicine, University of Helsinki, Helsinki, Finland Teemu Smura, Hannimari Kallio-Kokko, Olli Vapalahti
EPI_ISL_418395 hCoV-19/Finland/13M65/2020 Europe / Finland 2020-03-13 Department of Virology and Immunology, University of Helsinki and Helsinki University Hospital, Huslab Finland Department of Virology, Faculty of Medicine, University of Helsinki, Helsinki, Finland Teemu Smura, Hannimari Kallio-Kokko, Olli Vapalahti
EPI_ISL_418396 hCoV-19/Finland/13M69/2020 Europe / Finland 2020-03-13 Department of Virology and Immunology, University of Helsinki and Helsinki University Hospital, Huslab Finland Department of Virology, Faculty of Medicine, University of Helsinki, Helsinki, Finland Teemu Smura, Hannimari Kallio-Kokko, Olli Vapalahti
EPI_ISL_418397 hCoV-19/Finland/13M77/2020 Europe / Finland 2020-03-13 Department of Virology and Immunology, University of Helsinki and Helsinki University Hospital, Huslab Finland Department of Virology, Faculty of Medicine, University of Helsinki, Helsinki, Finland Teemu Smura, Hannimari Kallio-Kokko, Olli Vapalahti
EPI_ISL_418399 hCoV-19/Finland/13M82/2020 Europe / Finland 2020-03-13 Department of Virology and Immunology, University of Helsinki and Helsinki University Hospital, Huslab Finland Department of Virology, Faculty of Medicine, University of Helsinki, Helsinki, Finland Teemu Smura, Hannimari Kallio-Kokko, Olli Vapalahti
EPI_ISL_418800 hCoV-19/Belgium/CS-031052/2020 Europe / Belgium / Gierle 2020-03-10 KU Leuven, Clinical and Epidemiological Virology KU Leuven, Clinical and Epidemiological Virology Bert Vanmechelen, Joan Marti-Carreras, Tony Wawina, Piet Maes
EPI_ISL_418801 hCoV-19/Australia/QLDID923/2020 Oceania / Australia / Queensland / Brisbane 2020-03-14 Mater Pathology Public Health Virology Laboratory Bixing Huang, Alyssa Pyke, Amanda De Jong, Andrew Van Den Hurk, Carmel Taylor, David Warrilow, Doris Genge, Elisabeth Gamez, Glen Hewitson, Ian Maxwell Mackay, Inga Sultana, Jamie McMahon, Jean Barcelon, Judy Northill, Mitchell Finger, Natalie Simpson, Neelima Nair, Peter Burtonclay, Peter Moore, Sarah Wheatley, Sean Moody, Sonja Hall-Mendelin, Timothy Gardam, and Frederick Moore
EPI_ISL_418802 hCoV-19/Australia/QLDID924/2020 Oceania / Australia / Queensland / Gold Coast 2020-03-18 Pathology Queensland Public Health Virology Laboratory Bixing Huang, Alyssa Pyke, Amanda De Jong, Andrew Van Den Hurk, Carmel Taylor, David Warrilow, Doris Genge, Elisabeth Gamez, Glen Hewitson, Ian Maxwell Mackay, Inga Sultana, Jamie McMahon, Jean Barcelon, Judy Northill, Mitchell Finger, Natalie Simpson, Neelima Nair, Peter Burtonclay, Peter Moore, Sarah Wheatley, Sean Moody, Sonja Hall-Mendelin, Timothy Gardam, and Frederick Moore
EPI_ISL_418803 hCoV-19/Australia/QLDID925/2020 Oceania / Australia / Queensland / Sunshine Coast 2020-03-18 Pathology Queensland Public Health Virology Laboratory Bixing Huang, Alyssa Pyke, Amanda De Jong, Andrew Van Den Hurk, Carmel Taylor, David Warrilow, Doris Genge, Elisabeth Gamez, Glen Hewitson, Ian Maxwell Mackay, Inga Sultana, Jamie McMahon, Jean Barcelon, Judy Northill, Mitchell Finger, Natalie Simpson, Neelima Nair, Peter Burtonclay, Peter Moore, Sarah Wheatley, Sean Moody, Sonja Hall-Mendelin, Timothy Gardam, and Frederick Moore
EPI_ISL_418804 hCoV-19/Australia/QLDID926/2020 Oceania / Australia / Queensland / Sunshine Coast 2020-03-18 Pathology Queensland Public Health Virology Laboratory Bixing Huang, Alyssa Pyke, Amanda De Jong, Andrew Van Den Hurk, Carmel Taylor, David Warrilow, Doris Genge, Elisabeth Gamez, Glen Hewitson, Ian Maxwell Mackay, Inga Sultana, Jamie McMahon, Jean Barcelon, Judy Northill, Mitchell Finger, Natalie Simpson, Neelima Nair, Peter Burtonclay, Peter Moore, Sarah Wheatley, Sean Moody, Sonja Hall-Mendelin, Timothy Gardam, and Frederick Moore

EPI_ISL_418805  hCoV-19/Belgium/VPE-030650/2020 Europe / Belgium / Deinze       2020-03-06      KU Leuven, Clinical and Epidemiological Virology   KU Leuven, Clinical and Epidemiological Virology        Bert Vanmechelen, Joan Marti-Carreras, Tony Wawina, Piet Maes
EPI_ISL_418806  hCoV-19/Belgium/BG-030551/2020  Europe / Belgium / Beersel      2020-03-05      KU Leuven, Clinical and Epidemiological Virology   KU Leuven, Clinical and Epidemiological Virology        Bert Vanmechelen, Joan Marti-Carreras, Tony Wawina, Piet Maes
EPI_ISL_418807  hCoV-19/Australia/QLDID927/2020 Oceania / Australia / Queensland / Brisbane     2020-03-19      Pathology Queensland    Public Health Virology Laboratory       Bixing Huang, Alyssa Pyke, Amanda De Jong, Andrew Van Den Hurk, Carmel Taylor, David Warrilow, Doris Genge, Elisabeth Gamez, Glen Hewitson, Ian Maxwell Mackay, Inga Sultana, Jamie McMahon, Jean Barcelon, Judy Northill, Mitchell Finger, Natalie Simpson, Neelima Nair, Peter Burtonclay, Peter Moore, Sarah Wheatley, Sean Moody, Sonja Hall-Mendelin, Timothy Gardam, and Frederick Moore
EPI_ISL_418808  hCoV-19/Australia/QLDID928/2020 Oceania / Australia / Queensland / Brisbane     2020-03-19      Pathology Queensland    Public Health Virology Laboratory       Bixing Huang, Alyssa Pyke, Amanda De Jong, Andrew Van Den Hurk, Carmel Taylor, David Warrilow, Doris Genge, Elisabeth Gamez, Glen Hewitson, Ian Maxwell Mackay, Inga Sultana, Jamie McMahon, Jean Barcelon, Judy Northill, Mitchell Finger, Natalie Simpson, Neelima Nair, Peter Burtonclay, Peter Moore, Sarah Wheatley, Sean Moody, Sonja Hall-Mendelin, Timothy Gardam, and Frederick Moore
EPI_ISL_418809  hCoV-19/Japan/UT-NCGM02/2020    Asia / Japan / Tokyo    2020-02-01      University of Wisconsin - Madison: Influenza Research Institute University of Wisconsin Madison, AIDS Vaccine Research Laboratories Katarina Braun, Gage Moreno, Peter Halfmann, et al.
EPI_ISL_418811  hCoV-19/Canada/NB_6/2020        North America / Canada / New Brunswick  2020-03-12      Dr. Georges-L.-Dumont University Hospital Centre    National Microbiology Laboratory        Anna Majer, Shari Tyson, Grace Seo, Philip Mabon, Natalie Knox, Morag Graham, Richard Garceau, Guillaume Desnoyers, Nathalie Bastien, Yan Li, Matthew Gilmour, Timothy Booth
EPI_ISL_418812  hCoV-19/Canada/MB_8/2020        North America / Canada / Manitoba       2020-03-12      Cadham Provincial Laboratory   National Microbiology Laboratory        Anna Majer, Shari Tyson, Grace Seo, Philip Mabon, Natalie Knox, Morag Graham, Paul Van Caeseele,  Jared Bullard, David Alexander, Kerry Dust, Nathalie Bastien, Yan Li, Matthew Gilmour, Timothy Booth
EPI_ISL_418813  hCoV-19/Canada/MB_10/2020       North America / Canada / Manitoba       2020-03-13      Cadham Provincial Laboratory   National Microbiology Laboratory        Anna Majer, Shari Tyson, Grace Seo, Philip Mabon, Natalie Knox, Morag Graham, Paul Van Caeseele,  Jared Bullard, David Alexander, Kerry Dust, Nathalie Bastien, Yan Li, Matthew Gilmour, Timothy Booth
EPI_ISL_418814  hCoV-19/Canada/NS_13/2020       North America / Canada / Nova Scotia    2020-03-13      Queen Elizabeth II Health Science Centre      National Microbiology Laboratory        Anna Majer, Shari Tyson, Grace Seo, Philip Mabon, Natalie Knox, Morag Graham, Todd Hatchette, Jason LeBlanc, Nathalie Bastien, Yan Li, Matthew Gilmour, Timothy Booth
EPI_ISL_418815  hCoV-19/Hong Kong/HKPU36-0702/2020      Asia / Hong Kong        2020-02-09      Department of Clinical Pathology, Pamela Youde Nethersole Eastern Hospital    Department of Health Technology and Informatics, Faculty of Health and Social Science, The Hong Kong Polytechnic University Kenneth Siu-Sing LEUNG, Timothy Ting-Leung NG, Alan Ka-Lun WU, Miranda Chong-Yee YAU, Hiu-Yin LAO, Ming-Pan CHOI, Kingsley King-Gee TAM, Lam-Kwong LEE, Barry Kin-Chung WONG, Alex Yat-Man HO, Kam-Tong YIP, Kwok-Cheung LUNG, Raymond Wai-To LIU, Eugene Yuk-Keung TSO, Wai-Shing LEUNG, Man-Chun CHAN, Yuk-Yung NG, Kit-Man SIN, Kitty Sau-Chun FUNG, Sandy Ka-Yee CHAU, Wing-Kin TO, Tak-Lun QUE, David Ho-Keung SHUM, Shea Ping YIP, Wing Cheong YAM, Gilman Kit-Hang SIU
EPI_ISL_418819  hCoV-19/Canada/BC_3808524/2020  North America / Canada / British Columbia       2020-03-09      BCCDC Public Health Laboratory  BCCDC Public Health Laboratory  Harrigan, Prystajecky, Krajden, Lee, Kamelian, Lapointe, Choi, Hoang, Sekirov, Levett, Tyson, Snutch, Loman, Quick, Li, Gilmour
EPI_ISL_418820  hCoV-19/Canada/BC_3842755/2020  North America / Canada / British Columbia       2020-03-09      BCCDC Public Health Laboratory  BCCDC Public Health Laboratory  Harrigan, Prystajecky, Krajden, Lee, Kamelian, Lapointe, Choi, Hoang, Sekirov, Levett, Tyson, Snutch, Loman, Quick, Li, Gilmour
EPI_ISL_418821  hCoV-19/Canada/BC_3968175/2020  North America / Canada / British Columbia       2020-03-10      BCCDC Public Health Laboratory  BCCDC Public Health Laboratory  Harrigan, Prystajecky, Krajden, Lee, Kamelian, Lapointe, Choi, Hoang, Sekirov, Levett, Tyson, Snutch, Loman, Quick, Li, Gilmour
EPI_ISL_418822  hCoV-19/Canada/BC_3972884/2020  North America / Canada / British Columbia       2020-03-10      BCCDC Public Health Laboratory  BCCDC Public Health Laboratory  Harrigan, Prystajecky, Krajden, Lee, Kamelian, Lapointe, Choi, Hoang, Sekirov, Levett, Tyson, Snutch, Loman, Quick, Li, Gilmour
EPI_ISL_418823  hCoV-19/Canada/BC_3989992/2020  North America / Canada / British Columbia       2020-03-09      BCCDC Public Health Laboratory  BCCDC Public Health Laboratory  Harrigan, Prystajecky, Krajden, Lee, Kamelian, Lapointe, Choi, Hoang, Sekirov, Levett, Tyson, Snutch, Loman, Quick, Li, Gilmour
EPI_ISL_418824  hCoV-19/Canada/BC_4078583/2020  North America / Canada / British Columbia       2020-03-03      BCCDC Public Health Laboratory  BCCDC Public Health Laboratory  Harrigan, Prystajecky, Krajden, Lee, Kamelian, Lapointe, Choi, Hoang, Sekirov, Levett, Tyson, Snutch, Loman, Quick, Li, Gilmour
EPI_ISL_418825  hCoV-19/Canada/BC_4118226/2020  North America / Canada / British Columbia       2020-03-10      BCCDC Public Health Laboratory  BCCDC Public Health Laboratory  Harrigan, Prystajecky, Krajden, Lee, Kamelian, Lapointe, Choi, Hoang, Sekirov, Levett, Tyson, Snutch, Loman, Quick, Li, Gilmour
EPI_ISL_418826  hCoV-19/Canada/BC_4122951/2020  North America / Canada / British Columbia       2020-03-10      BCCDC Public Health Laboratory  BCCDC Public Health Laboratory  Harrigan, Prystajecky, Krajden, Lee, Kamelian, Lapointe, Choi, Hoang, Sekirov, Levett, Tyson, Snutch, Loman, Quick, Li, Gilmour
EPI_ISL_418829  hCoV-19/Canada/BC_4540462/2020  North America / Canada / British Columbia       2020-03-10      BCCDC Public Health Laboratory  BCCDC Public Health Laboratory  Harrigan, Prystajecky, Krajden, Lee, Kamelian,

Lapointe, Choi, Hoang, Sekirov, Levett, Tyson, Snutch, Loman, Quick, Li, Gilmour
EPI_ISL_418830 hCoV-19/Canada/BC_4799711/2020 North America / Canada / British Columbia 2020-03-09 BCCDC Public Health Laboratory BCCDC Public Health Laboratory Harrigan, Prystajecky, Krajden, Lee, Kamelian, Lapointe, Choi, Hoang, Sekirov, Levett, Tyson, Snutch, Loman, Quick, Li, Gilmour
EPI_ISL_418831 hCoV-19/Canada/BC_5275718/2020 North America / Canada / British Columbia 2020-03-11 BCCDC Public Health Laboratory BCCDC Public Health Laboratory Harrigan, Prystajecky, Krajden, Lee, Kamelian, Lapointe, Choi, Hoang, Sekirov, Levett, Tyson, Snutch, Loman, Quick, Li, Gilmour
EPI_ISL_418832 hCoV-19/Canada/BC_5282984/2020 North America / Canada / British Columbia 2020-03-11 BCCDC Public Health Laboratory BCCDC Public Health Laboratory Harrigan, Prystajecky, Krajden, Lee, Kamelian, Lapointe, Choi, Hoang, Sekirov, Levett, Tyson, Snutch, Loman, Quick, Li, Gilmour
EPI_ISL_418833 hCoV-19/Canada/BC_5306970/2020 North America / Canada / British Columbia 2020-03-11 BCCDC Public Health Laboratory BCCDC Public Health Laboratory Harrigan, Prystajecky, Krajden, Lee, Kamelian, Lapointe, Choi, Hoang, Sekirov, Levett, Tyson, Snutch, Loman, Quick, Li, Gilmour
EPI_ISL_418834 hCoV-19/Canada/BC_5522039/2020 North America / Canada / British Columbia 2020-03-11 BCCDC Public Health Laboratory BCCDC Public Health Laboratory Harrigan, Prystajecky, Krajden, Lee, Kamelian, Lapointe, Choi, Hoang, Sekirov, Levett, Tyson, Snutch, Loman, Quick, Li, Gilmour
EPI_ISL_418835 hCoV-19/Canada/BC_5979789/2020 North America / Canada / British Columbia 2020-03-11 BCCDC Public Health Laboratory BCCDC Public Health Laboratory Harrigan, Prystajecky, Krajden, Lee, Kamelian, Lapointe, Choi, Hoang, Sekirov, Levett, Tyson, Snutch, Loman, Quick, Li, Gilmour
EPI_ISL_418836 hCoV-19/Canada/BC_6004567/2020 North America / Canada / British Columbia 2020-03-10 BCCDC Public Health Laboratory BCCDC Public Health Laboratory Harrigan, Prystajecky, Krajden, Lee, Kamelian, Lapointe, Choi, Hoang, Sekirov, Levett, Tyson, Snutch, Loman, Quick, Li, Gilmour
EPI_ISL_418837 hCoV-19/Canada/BC_6129127/2020 North America / Canada / British Columbia 2020-03-10 BCCDC Public Health Laboratory BCCDC Public Health Laboratory Harrigan, Prystajecky, Krajden, Lee, Kamelian, Lapointe, Choi, Hoang, Sekirov, Levett, Tyson, Snutch, Loman, Quick, Li, Gilmour
EPI_ISL_418860 hCoV-19/Spain/VH000001133/2020 Europe / Spain / Barcelona 2020-03-15 Hospital Universitari Vall d'Hebron (HUVH) - Vall d'Hebron Research Institute (VHIR) Hospital Universitari Vall d'Hebron (HUVH) - Vall d'Hebron Research Institute (VHIR) Cristina Andrés, Dàmir Garcia-Cehic, Maria Piñana, Mercedes Guerrero-Murillo, Ariadna Rando, Tomàs Pumarola, Maria Gema Codina, Andrés Antón, Josep Quer
EPI_ISL_418861 hCoV-19/Spain/VH198152683/2020 Europe / Spain / Barcelona 2020-03-24 Hospital Universitari Vall d'Hebron (HUVH) - Vall d'Hebron Research Institute (VHIR) Hospital Universitari Vall d'Hebron (HUVH) - Vall d'Hebron Research Institute (VHIR) Cristina Andrés, Dàmir Garcia-Cehic, Maria Piñana, Mercedes Guerrero-Murillo, Ariadna Rando, Tomàs Pumarola, Maria Gema Codina, Andrés Antón, Josep Quer
EPI_ISL_418863 hCoV-19/Belgium/VS-030542/2020 Europe / Belgium / Komen 2020-03-05 KU Leuven, Clinical and Epidemiological Virology KU Leuven, Clinical and Epidemiological Virology Bert Vanmechelen, Joan Marti-Carreras, Tony Wawina, Piet Maes
EPI_ISL_418864 hCoV-19/USA/VA-DCLS-0001/2020 North America / USA / Virginia 2020-03-17 Virginia DCLS Virginia DCLS Virginia DCLS
EPI_ISL_418866 hCoV-19/USA/WA-UW298/2020 North America / USA / Washington 2020-03-13 UW Virology Lab UW Virology Lab Pavitra Roychoudhury, Hong Xie, Keith Jerome, Alexander Greninger
EPI_ISL_418867 hCoV-19/USA/WA-UW299/2020 North America / USA / Washington 2020-03-13 UW Virology Lab UW Virology Lab Pavitra Roychoudhury, Hong Xie, Keith Jerome, Alexander Greninger
EPI_ISL_418868 hCoV-19/USA/WA-UW300/2020 North America / USA / Washington 2020-03-13 UW Virology Lab UW Virology Lab Pavitra Roychoudhury, Hong Xie, Keith Jerome, Alexander Greninger
EPI_ISL_418869 hCoV-19/USA/WA-UW301/2020 North America / USA / Washington 2020-03-16 UW Virology Lab UW Virology Lab Pavitra Roychoudhury, Hong Xie, Keith Jerome, Alexander Greninger
EPI_ISL_418870 hCoV-19/USA/WA-UW302/2020 North America / USA / Washington 2020-03-16 UW Virology Lab UW Virology Lab Pavitra Roychoudhury, Hong Xie, Keith Jerome, Alexander Greninger
EPI_ISL_418871 hCoV-19/USA/CT-UW303/2020 North America / USA / Connecticut 2020-03-14 UW Virology Lab UW Virology Lab Pavitra Roychoudhury, Hong Xie, Keith Jerome, Alexander Greninger
EPI_ISL_418872 hCoV-19/USA/WA-UW304/2020 North America / USA / Washington 2020-03-16 UW Virology Lab UW Virology Lab Pavitra Roychoudhury, Hong Xie, Keith Jerome, Alexander Greninger
EPI_ISL_418873 hCoV-19/USA/WA-UW305/2020 North America / USA / Washington 2020-03-16 UW Virology Lab UW Virology Lab Pavitra Roychoudhury, Hong Xie, Keith Jerome, Alexander Greninger
EPI_ISL_418875 hCoV-19/USA/WA-UW307/2020 North America / USA / Washington 2020-03-16 UW Virology Lab UW Virology Lab Pavitra Roychoudhury, Hong Xie, Keith Jerome, Alexander Greninger
EPI_ISL_418876 hCoV-19/USA/WA-UW308/2020 North America / USA / Washington 2020-03-16 UW Virology Lab UW Virology Lab Pavitra Roychoudhury, Hong Xie, Keith Jerome, Alexander Greninger
EPI_ISL_418877 hCoV-19/USA/WA-UW309/2020 North America / USA / Washington 2020-03-24 UW Virology Lab UW Virology Lab Pavitra Roychoudhury, Hong Xie, Keith Jerome, Alexander Greninger
EPI_ISL_418878 hCoV-19/USA/WA-UW310/2020 North America / USA / Washington 2020-03-14 UW Virology Lab UW Virology Lab Pavitra Roychoudhury, Hong Xie, Keith Jerome, Alexander Greninger
EPI_ISL_418880 hCoV-19/USA/WA-UW313/2020 North America / USA / Washington 2020-03-14 UW Virology Lab UW Virology Lab Pavitra Roychoudhury, Hong Xie, Keith Jerome, Alexander Greninger
EPI_ISL_418881 hCoV-19/USA/WA-UW314/2020 North America / USA / Washington 2020-03-14 UW Virology Lab UW Virology Lab Pavitra Roychoudhury, Hong Xie, Keith Jerome, Alexander Greninger
EPI_ISL_418882 hCoV-19/USA/WA-UW315/2020 North America / USA / Washington 2020-03-13 UW Virology Lab UW Virology Lab Pavitra Roychoudhury, Hong Xie, Keith Jerome, Alexander Greninger
EPI_ISL_418883 hCoV-19/USA/WA-UW316/2020 North America / USA / Washington 2020-03-14 UW Virology Lab UW Virology Lab Pavitra Roychoudhury, Hong Xie, Keith Jerome, Alexander Greninger

EPI_ISL_418884	hCoV-19/USA/WA-UW317/2020	North America / USA / Washington	2020-03-15	UW Virology Lab	UW Virology Lab	Pavitra Roychoudhury, Hong Xie, Keith Jerome, Alexander Greninger
EPI_ISL_418886	hCoV-19/USA/WA-UW319/2020	North America / USA / Washington	2020-03-15	UW Virology Lab	UW Virology Lab	Pavitra Roychoudhury, Hong Xie, Keith Jerome, Alexander Greninger
EPI_ISL_418887	hCoV-19/USA/WA-UW320/2020	North America / USA / Washington	2020-03-14	UW Virology Lab	UW Virology Lab	Pavitra Roychoudhury, Hong Xie, Keith Jerome, Alexander Greninger
EPI_ISL_418888	hCoV-19/USA/WA-UW321/2020	North America / USA / Washington	2020-03-15	UW Virology Lab	UW Virology Lab	Pavitra Roychoudhury, Hong Xie, Keith Jerome, Alexander Greninger
EPI_ISL_418889	hCoV-19/USA/WA-UW322/2020	North America / USA / Washington	2020-03-16	UW Virology Lab	UW Virology Lab	Pavitra Roychoudhury, Hong Xie, Keith Jerome, Alexander Greninger
EPI_ISL_418890	hCoV-19/USA/WA-UW323/2020	North America / USA / Washington	2020-03-16	UW Virology Lab	UW Virology Lab	Pavitra Roychoudhury, Hong Xie, Keith Jerome, Alexander Greninger
EPI_ISL_418891	hCoV-19/USA/WA-UW324/2020	North America / USA / Washington	2020-03-24	UW Virology Lab	UW Virology Lab	Pavitra Roychoudhury, Hong Xie, Keith Jerome, Alexander Greninger
EPI_ISL_418892	hCoV-19/USA/WA-UW325/2020	North America / USA / Washington	2020-03-16	UW Virology Lab	UW Virology Lab	Pavitra Roychoudhury, Hong Xie, Keith Jerome, Alexander Greninger
EPI_ISL_418893	hCoV-19/USA/MN-UW326/2020	North America / USA / Minnesota	2020-03-13	UW Virology Lab	UW Virology Lab	Pavitra Roychoudhury, Hong Xie, Keith Jerome, Alexander Greninger
EPI_ISL_418895	hCoV-19/USA/CT-UW328/2020	North America / USA / Connecticut	2020-03-14	UW Virology Lab	UW Virology Lab	Pavitra Roychoudhury, Hong Xie, Keith Jerome, Alexander Greninger
EPI_ISL_418897	hCoV-19/USA/WA-UW330/2020	North America / USA / Washington	2020-03-16	UW Virology Lab	UW Virology Lab	Pavitra Roychoudhury, Hong Xie, Keith Jerome, Alexander Greninger
EPI_ISL_418898	hCoV-19/USA/CT-UW331/2020	North America / USA / Connecticut	2020-03-14	UW Virology Lab	UW Virology Lab	Pavitra Roychoudhury, Hong Xie, Keith Jerome, Alexander Greninger
EPI_ISL_418899	hCoV-19/USA/WA-UW332/2020	North America / USA / Washington	2020-03-16	UW Virology Lab	UW Virology Lab	Pavitra Roychoudhury, Hong Xie, Keith Jerome, Alexander Greninger
EPI_ISL_419600	hCoV-19/Luxembourg/LNS9080444/2020	Europe / Luxembourg	2020-03-13	Laboratoire National de Santé, Microbiology, Virology	Laboratoire National de Santé, Microbiology, Epidemiology and Microbial Genomics	Anke Wienecke-Baldacchino, Ardashel Latsuzbaia, Jessica Tapp, Catherine Ragimbeau, Guillaume Fournier, Tamir Abdelrahman, Trung Nguyen Nguyen, Joel Mossong
EPI_ISL_419601	hCoV-19/Luxembourg/LNS9086704/2020	Europe / Luxembourg	2020-03-15	Laboratoire National de Santé, Microbiology, Virology	Laboratoire National de Santé, Microbiology, Epidemiology and Microbial Genomics	Anke Wienecke-Baldacchino, Ardashel Latsuzbaia, Jessica Tapp, Catherine Ragimbeau, Guillaume Fournier, Tamir Abdelrahman, Trung Nguyen Nguyen, Joel Mossong
EPI_ISL_419602	hCoV-19/Luxembourg/LNS9324837/2020	Europe / Luxembourg	2020-03-12	Laboratoire National de Santé, Microbiology, Virology	Laboratoire National de Santé, Microbiology, Epidemiology and Microbial Genomics	Anke Wienecke-Baldacchino, Ardashel Latsuzbaia, Jessica Tapp, Catherine Ragimbeau, Guillaume Fournier, Tamir Abdelrahman, Trung Nguyen Nguyen, Joel Mossong
EPI_ISL_419603	hCoV-19/Luxembourg/LNS9512434/2020	Europe / Luxembourg	2020-03-16	Laboratoire National de Santé, Microbiology, Virology	Laboratoire National de Santé, Microbiology, Epidemiology and Microbial Genomics	Anke Wienecke-Baldacchino, Ardashel Latsuzbaia, Jessica Tapp, Catherine Ragimbeau, Guillaume Fournier, Tamir Abdelrahman, Trung Nguyen Nguyen, Joel Mossong
EPI_ISL_419604	hCoV-19/Luxembourg/LNS9627078/2020	Europe / Luxembourg	2020-03-15	Laboratoire National de Santé, Microbiology, Virology	Laboratoire National de Santé, Microbiology, Epidemiology and Microbial Genomics	Anke Wienecke-Baldacchino, Ardashel Latsuzbaia, Jessica Tapp, Catherine Ragimbeau, Guillaume Fournier, Tamir Abdelrahman, Trung Nguyen Nguyen, Joel Mossong
EPI_ISL_419606	hCoV-19/Luxembourg/LNS9862689/2020	Europe / Luxembourg	2020-03-11	Laboratoire National de Santé, Microbiology, Virology	Laboratoire National de Santé, Microbiology, Epidemiology and Microbial Genomics	Anke Wienecke-Baldacchino, Ardashel Latsuzbaia, Jessica Tapp, Catherine Ragimbeau, Guillaume Fournier, Tamir Abdelrahman, Trung Nguyen Nguyen, Joel Mossong
EPI_ISL_419607	hCoV-19/Luxembourg/LNS9982497/2020	Europe / Luxembourg	2020-03-14	Laboratoire National de Santé, Microbiology, Virology	Laboratoire National de Santé, Microbiology, Epidemiology and Microbial Genomics	Anke Wienecke-Baldacchino, Ardashel Latsuzbaia, Jessica Tapp, Catherine Ragimbeau, Guillaume Fournier, Tamir Abdelrahman, Trung Nguyen Nguyen, Joel Mossong
EPI_ISL_419651	hCoV-19/USA/WI-GMF-00466/2020	North America / USA / Minnesota / Winona County	2020-03-29	Gundersen Molecular Diagnostics Laboratory	Kabara Cancer Research Institute	Craig S. Richmond & Paraic A. Kenny
EPI_ISL_419652	hCoV-19/USA/WI-GMF-00281/2020	North America / USA / Wisconsin / La Crosse County	2020-03-25	Gundersen Molecular Diagnostics Laboratory	Kabara Cancer Research Institute	Craig S. Richmond & Paraic A. Kenny
EPI_ISL_419654	hCoV-19/Austria/CeMM0001/2020	Europe / Austria	2020-03-03	Center for Virology, Medical University of Vienna	Bergthaler laboratory, CeMM Research Center for Molecular Medicine of the Austrian Academy of Sciences	Alexandra Popa, Benedikt Agerer, Henrique Colaco, Lukas Endler, Jakob-Wendelin Genger, Alexander Lercher, Mark Smyth, Thomas Penz, Michael Schuster, Judith Aberle, Stephan Aberle, Elisabeth Puchhammer-Stöckl, Christoph Bock, Andreas Bergthaler
EPI_ISL_419655	hCoV-19/Austria/CeMM0002/2020	Europe / Austria / Vienna	2020-02-26	Center for Virology, Medical University of Vienna	Bergthaler laboratory, CeMM Research Center for Molecular Medicine of the Austrian Academy of Sciences	Alexandra Popa, Benedikt Agerer, Henrique Colaco, Lukas Endler, Jakob-Wendelin Genger, Alexander Lercher, Mark Smyth, Thomas Penz, Michael Schuster, Judith Aberle, Stephan Aberle, Elisabeth Puchhammer-Stöckl, Christoph Bock, Andreas Bergthaler

EPI_ISL_419656 hCoV-19/Austria/CeMM0003/2020 Europe / Austria 2020-02-26 Center for Virology, Medical University of Vienna Bergthaler laboratory, CeMM Research Center for Molecular Medicine of the Austrian Academy of Sciences Alexandra Popa, Benedikt Agerer, Henrique Colaco, Lukas Endler, Jakob-Wendelin Genger, Alexander Lercher, Mark Smyth, Thomas Penz, Michael Schuster, Judith Aberle, Stephan Aberle, Elisabeth Puchhammer-Stöckl, Christoph Bock, Andreas Bergthaler
EPI_ISL_419658 hCoV-19/Austria/CeMM0005/2020 Europe / Austria 2020-03-06 Center for Virology, Medical University of Vienna Bergthaler laboratory, CeMM Research Center for Molecular Medicine of the Austrian Academy of Sciences Alexandra Popa, Benedikt Agerer, Henrique Colaco, Lukas Endler, Jakob-Wendelin Genger, Alexander Lercher, Mark Smyth, Thomas Penz, Michael Schuster, Judith Aberle, Stephan Aberle, Elisabeth Puchhammer-Stöckl, Christoph Bock, Andreas Bergthaler
EPI_ISL_419659 hCoV-19/Austria/CeMM0006/2020 Europe / Austria 2020-03-10 Center for Virology, Medical University of Vienna Bergthaler laboratory, CeMM Research Center for Molecular Medicine of the Austrian Academy of Sciences Alexandra Popa, Benedikt Agerer, Henrique Colaco, Lukas Endler, Jakob-Wendelin Genger, Alexander Lercher, Mark Smyth, Thomas Penz, Michael Schuster, Judith Aberle, Stephan Aberle, Elisabeth Puchhammer-Stöckl, Christoph Bock, Andreas Bergthaler
EPI_ISL_419660 hCoV-19/Austria/CeMM0007/2020 Europe / Austria 2020-03-13 Center for Virology, Medical University of Vienna Bergthaler laboratory, CeMM Research Center for Molecular Medicine of the Austrian Academy of Sciences Alexandra Popa, Benedikt Agerer, Henrique Colaco, Lukas Endler, Jakob-Wendelin Genger, Alexander Lercher, Mark Smyth, Thomas Penz, Michael Schuster, Judith Aberle, Stephan Aberle, Elisabeth Puchhammer-Stöckl, Christoph Bock, Andreas Bergthaler
EPI_ISL_419661 hCoV-19/Austria/CeMM0008/2020 Europe / Austria 2020-03-10 Center for Virology, Medical University of Vienna Bergthaler laboratory, CeMM Research Center for Molecular Medicine of the Austrian Academy of Sciences Alexandra Popa, Benedikt Agerer, Henrique Colaco, Lukas Endler, Jakob-Wendelin Genger, Alexander Lercher, Mark Smyth, Thomas Penz, Michael Schuster, Judith Aberle, Stephan Aberle, Elisabeth Puchhammer-Stöckl, Christoph Bock, Andreas Bergthaler
EPI_ISL_419662 hCoV-19/Austria/CeMM0009/2020 Europe / Austria 2020-03-14 Center for Virology, Medical University of Vienna Bergthaler laboratory, CeMM Research Center for Molecular Medicine of the Austrian Academy of Sciences Alexandra Popa, Benedikt Agerer, Henrique Colaco, Lukas Endler, Jakob-Wendelin Genger, Alexander Lercher, Mark Smyth, Thomas Penz, Michael Schuster, Judith Aberle, Stephan Aberle, Elisabeth Puchhammer-Stöckl, Christoph Bock, Andreas Bergthaler
EPI_ISL_419664 hCoV-19/Austria/CeMM0011/2020 Europe / Austria 2020-03-23 Center for Virology, Medical University of Vienna Bergthaler laboratory, CeMM Research Center for Molecular Medicine of the Austrian Academy of Sciences Alexandra Popa, Benedikt Agerer, Henrique Colaco, Lukas Endler, Jakob-Wendelin Genger, Alexander Lercher, Mark Smyth, Thomas Penz, Michael Schuster, Judith Aberle, Stephan Aberle, Elisabeth Puchhammer-Stöckl, Christoph Bock, Andreas Bergthaler
EPI_ISL_419665 hCoV-19/Austria/CeMM0012/2020 Europe / Austria 2020-03-10 Center for Virology, Medical University of Vienna Bergthaler laboratory, CeMM Research Center for Molecular Medicine of the Austrian Academy of Sciences Alexandra Popa, Benedikt Agerer, Henrique Colaco, Lukas Endler, Jakob-Wendelin Genger, Alexander Lercher, Mark Smyth, Thomas Penz, Michael Schuster, Judith Aberle, Stephan Aberle, Elisabeth Puchhammer-Stöckl, Christoph Bock, Andreas Bergthaler
EPI_ISL_419666 hCoV-19/Austria/CeMM0013/2020 Europe / Austria 2020-03-11 Center for Virology, Medical University of Vienna Bergthaler laboratory, CeMM Research Center for Molecular Medicine of the Austrian Academy of Sciences Alexandra Popa, Benedikt Agerer, Henrique Colaco, Lukas Endler, Jakob-Wendelin Genger, Alexander Lercher, Mark Smyth, Thomas Penz, Michael Schuster, Judith Aberle, Stephan Aberle, Elisabeth Puchhammer-Stöckl, Christoph Bock, Andreas Bergthaler
EPI_ISL_419667 hCoV-19/Austria/CeMM0014/2020 Europe / Austria 2020-03-13 Center for Virology, Medical University of Vienna Bergthaler laboratory, CeMM Research Center for Molecular Medicine of the Austrian Academy of Sciences Alexandra Popa, Benedikt Agerer, Henrique Colaco, Lukas Endler, Jakob-Wendelin Genger, Alexander Lercher, Mark Smyth, Thomas Penz, Michael Schuster, Judith Aberle, Stephan Aberle, Elisabeth Puchhammer-Stöckl, Christoph Bock, Andreas Bergthaler
EPI_ISL_419669 hCoV-19/Austria/CeMM0016/2020 Europe / Austria 2020-03-16 Center for Virology, Medical University of Vienna Bergthaler laboratory, CeMM Research Center for Molecular Medicine of the Austrian Academy of Sciences Alexandra Popa, Benedikt Agerer, Henrique Colaco, Lukas Endler, Jakob-Wendelin Genger, Alexander Lercher, Mark Smyth, Thomas Penz, Michael Schuster, Judith Aberle, Stephan Aberle, Elisabeth Puchhammer-Stöckl, Christoph Bock, Andreas Bergthaler
EPI_ISL_419670 hCoV-19/Austria/CeMM0017/2020 Europe / Austria 2020-03-16 Center for Virology, Medical University of Vienna Bergthaler laboratory, CeMM Research Center for Molecular Medicine of the Austrian Academy of Sciences Alexandra Popa, Benedikt Agerer, Henrique Colaco, Lukas Endler, Jakob-Wendelin Genger, Alexander Lercher, Mark Smyth, Thomas Penz, Michael Schuster, Judith Aberle, Stephan Aberle, Elisabeth Puchhammer-Stöckl, Christoph Bock, Andreas Bergthaler
EPI_ISL_419671 hCoV-19/Austria/CeMM0018/2020 Europe / Austria 2020-03-19 Center for Virology, Medical University of Vienna Bergthaler laboratory, CeMM Research Center for Molecular Medicine of the Austrian Academy of Sciences Alexandra Popa, Benedikt Agerer, Henrique Colaco, Lukas Endler, Jakob-Wendelin Genger, Alexander Lercher, Mark Smyth, Thomas Penz, Michael Schuster, Judith Aberle, Stephan Aberle, Elisabeth Puchhammer-Stöckl, Christoph Bock, Andreas Bergthaler
EPI_ISL_419672 hCoV-19/Austria/CeMM0019/2020 Europe / Austria 2020-03-19 Center for Virology, Medical University of Vienna Bergthaler laboratory, CeMM Research Center for Molecular Medicine of the Austrian Academy of Sciences Alexandra Popa, Benedikt Agerer, Henrique Colaco, Lukas Endler, Jakob-Wendelin Genger, Alexander Lercher, Mark Smyth, Thomas Penz, Michael Schuster, Judith Aberle, Stephan Aberle, Elisabeth Puchhammer-Stöckl, Christoph Bock, Andreas Bergthaler

EPI_ISL_419673 hCoV-19/Austria/CeMM0020/2020 Europe / Austria 2020-03-22 Center for Virology, Medical University of Vienna Bergthaler laboratory, CeMM Research Center for Molecular Medicine of the Austrian Academy of Sciences Alexandra Popa, Benedikt Agerer, Henrique Colaco, Lukas Endler, Jakob-Wendelin Genger, Alexander Lercher, Mark Smyth, Thomas Penz, Michael Schuster, Judith Aberle, Stephan Aberle, Elisabeth Puchhammer-Stöckl, Christoph Bock, Andreas Bergthaler
EPI_ISL_419674 hCoV-19/Austria/CeMM0021/2020 Europe / Austria 2020-03-24 Center for Virology, Medical University of Vienna Bergthaler laboratory, CeMM Research Center for Molecular Medicine of the Austrian Academy of Sciences Alexandra Popa, Benedikt Agerer, Henrique Colaco, Lukas Endler, Jakob-Wendelin Genger, Alexander Lercher, Mark Smyth, Thomas Penz, Michael Schuster, Judith Aberle, Stephan Aberle, Elisabeth Puchhammer-Stöckl, Christoph Bock, Andreas Bergthaler
EPI_ISL_419675 hCoV-19/Spain/Valencia11/2020 Europe / Spain / Comunitat Valenciana / Valencia 2020-03-20 Servicio de Microbiología. Consorcio Hospital General Universitario de Valencia Sequencing and Bioinformatics Service and Molecular Epidemiology Research Group. FISABIO-Public Health Maria Alma Bracho, Maria Dolores Ocete, Giuseppe D'Auria, Griselda De Marco, Neris Garcia-Gonzalez, Concepcion Gimeno, Fernando Gonzalez-Candelas
EPI_ISL_419676 hCoV-19/Spain/Valencia12/2020 Europe / Spain / Comunitat Valenciana / Valencia 2020-03-09 Servicio de Microbiología. Consorcio Hospital General Universitario de Valencia Sequencing and Bioinformatics Service and Molecular Epidemiology Research Group. FISABIO-Public Health Maria Dolores Ocete, Giuseppe D'Auria, Griselda De Marco, Neris Garcia-Gonzalez, Maria Alma Bracho, Concepcion Gimeno, Fernando Gonzalez-Candelas
EPI_ISL_419677 hCoV-19/Spain/Valencia13/2020 Europe / Spain / Comunitat Valenciana / Valencia 2020-03-09 Servicio de Microbiología. Consorcio Hospital General Universitario de Valencia Sequencing and Bioinformatics Service and Molecular Epidemiology Research Group. FISABIO-Public Health Giuseppe D'Auria, Griselda De Marco, Neris Garcia-Gonzalez, Maria Alma Bracho, Maria Dolores Ocete, Concepcion Gimeno, Fernando Gonzalez-Candelas
EPI_ISL_419678 hCoV-19/Spain/Valencia14/2020 Europe / Spain / Comunitat Valenciana / Valencia 2020-03-09 Servicio de Microbiología. Consorcio Hospital General Universitario de Valencia Sequencing and Bioinformatics Service and Molecular Epidemiology Research Group. FISABIO-Public Health Griselda De Marco, Neris Garcia-Gonzalez, Maria Alma Bracho, Maria Dolores Ocete, Giuseppe D'Auria, Concepcion Gimeno, Fernando Gonzalez-Candelas
EPI_ISL_419679 hCoV-19/Spain/Valencia15/2020 Europe / Spain / Comunitat Valenciana / Valencia 2020-03-02 Servicio de Microbiología. Consorcio Hospital General Universitario de Valencia Sequencing and Bioinformatics Service and Molecular Epidemiology Research Group. FISABIO-Public Health Neris Garcia-Gonzalez, Maria Alma Bracho, Maria Dolores Ocete, Giuseppe D'Auria, Griselda De Marco, Concepcion Gimeno, Fernando Gonzalez-Candelas
EPI_ISL_419680 hCoV-19/Spain/Valencia16/2020 Europe / Spain / Comunitat Valenciana / Valencia 2020-03-10 Servicio de Microbiología. Consorcio Hospital General Universitario de Valencia Sequencing and Bioinformatics Service and Molecular Epidemiology Research Group. FISABIO-Public Health Maria Alma Bracho, Maria Dolores Ocete, Giuseppe D'Auria, Griselda De Marco, Neris Garcia-Gonzalez, Concepcion Gimeno, Fernando Gonzalez-Candelas
EPI_ISL_419681 hCoV-19/Spain/Valencia17/2020 Europe / Spain / Comunitat Valenciana / Valencia 2020-03-10 Servicio de Microbiología. Consorcio Hospital General Universitario de Valencia Sequencing and Bioinformatics Service and Molecular Epidemiology Research Group. FISABIO-Public Health Maria Dolores Ocete, Giuseppe D'Auria, Griselda De Marco, Neris Garcia-Gonzalez, Maria Alma Bracho, Concepcion Gimeno, Fernando Gonzalez-Candelas
EPI_ISL_419682 hCoV-19/Spain/Valencia18/2020 Europe / Spain / Comunitat Valenciana / Valencia 2020-03-10 Servicio de Microbiología. Consorcio Hospital General Universitario de Valencia Sequencing and Bioinformatics Service and Molecular Epidemiology Research Group. FISABIO-Public Health Giuseppe D'Auria, Griselda De Marco, Neris Garcia-Gonzalez, Maria Alma Bracho, Maria Dolores Ocete, Concepcion Gimeno, Fernando Gonzalez-Candelas
EPI_ISL_419683 hCoV-19/Spain/Valencia19/2020 Europe / Spain / Comunitat Valenciana / Valencia 2020-03-08 Servicio de Microbiología. Consorcio Hospital General Universitario de Valencia Sequencing and Bioinformatics Service and Molecular Epidemiology Research Group. FISABIO-Public Health Griselda De Marco, Neris Garcia-Gonzalez, Maria Alma Bracho, Maria Dolores Ocete, Giuseppe D'Auria, Concepcion Gimeno, Fernando Gonzalez-Candelas
EPI_ISL_419691 hCoV-19/Latvia/ChVir2025/2020 Europe / Latvia / Riga 2020-03 E. Gulbja Laboratorija Charité Universitätsmedizin Berlin, Institute of Virology Victor M Corman, Julia Schneider, Barbara Mühlemann, Talitha Veith, Jörn Beheim-Schwarzbach, Terry Jones, Dr. Didzis Gavars, Mikus Gavars, Dmitrijs Perminovs, Christian Drosten
EPI_ISL_419692 hCoV-19/Belarus/ChVir2072/2020 Europe / Belarus 2020-03 The Republican Research and Practical Center for Epidemiology and Microbiology Charité Universitätsmedizin Berlin, Institute of Virology Victor M Corman, Julia Schneider, Barbara Mühlemann, Talitha Veith, Jörn Beheim-Schwarzbach, Terry Jones, Natallia Shmialiova, Natallia Sivets, Christian Drosten
EPI_ISL_419693 hCoV-19/Belarus/ChVir2073/2020 Europe / Belarus 2020-03 The Republican Research and Practical Center for Epidemiology and Microbiology Charité Universitätsmedizin Berlin, Institute of Virology Victor M Corman, Julia Schneider, Barbara Mühlemann, Talitha Veith, Jörn Beheim-Schwarzbach, Terry Jones, Natallia Shmialiova, Natallia Sivets, Christian Drosten
EPI_ISL_419696 hCoV-19/USA/NY-NYUMC35/2020 North America / USA / New York / Manhattan 2020-03-18 NYU Langone Health Departments of Pathology and Medicine, New York University School of Medicine Maria Aguero-Rosenfeld, Margaret Black, John Cadley, Paolo Cotzia, John Chen, Dacia Dimartino, Xiaojun Feng, Adriana

Heguy, Megan Hogan, Emily Huang, George Jour, Christian Marier, Matthew T. Maurano, Mark J. Mulligan, Peter Meyn, Jared Pinnell, Sitharam Ramaswami, Amy Rapkiewicz, Marie Samanovic-Golden, Antonio Serrano, Guomiao Shen, Matija Snuderl, Nick Vulpescu, Gael Westby, Paul Zappile, Yutong Zhang
EPI_ISL_419697 hCoV-19/USA/NY-NYUMC36/2020 North America / USA / New York / Manhattan 2020-03-18 NYU Langone Health Departments of Pathology and Medicine, New York University School of Medicine Maria Aguero-Rosenfeld, Margaret Black, John Cadley, Paolo Cotzia, John Chen, Dacia Dimartino, Xiaojun Feng, Adriana Heguy, Megan Hogan, Emily Huang, George Jour, Christian Marier, Matthew T. Maurano, Mark J. Mulligan, Peter Meyn, Jared Pinnell, Sitharam Ramaswami, Amy Rapkiewicz, Marie Samanovic-Golden, Antonio Serrano, Guomiao Shen, Matija Snuderl, Nick Vulpescu, Gael Westby, Paul Zappile, Yutong Zhang
EPI_ISL_419698 hCoV-19/USA/NY-NYUMC37/2020 North America / USA / New York / Manhattan 2020-03-18 NYU Langone Health Departments of Pathology and Medicine, New York University School of Medicine Maria Aguero-Rosenfeld, Margaret Black, John Cadley, Paolo Cotzia, John Chen, Dacia Dimartino, Xiaojun Feng, Adriana Heguy, Megan Hogan, Emily Huang, George Jour, Christian Marier, Matthew T. Maurano, Mark J. Mulligan, Peter Meyn, Jared Pinnell, Sitharam Ramaswami, Amy Rapkiewicz, Marie Samanovic-Golden, Antonio Serrano, Guomiao Shen, Matija Snuderl, Nick Vulpescu, Gael Westby, Paul Zappile, Yutong Zhang
EPI_ISL_420600 hCoV-19/Argentina/C121/2020 South America / Argentina 2020-03-07 Servicio Virosis Respiratorias-Departamento Virología-INEI Instituto Nacional Enfermedades Infecciosas C.G.Malbran Baumeister E., Avaro M., Benedetti E., Russo M., Dattero ME, Pontoriero A., Cisterna D., Molina V., Perandones C., Tuduri E., Lorenzo F., Poklepovich T., Campos J.
EPI_ISL_420604 hCoV-19/France/ARA12371/2020 Europe / France / ARA / Lyon 2020-03-23 Institut des Agents Infectieux (IAI), Hospices Civils de Lyon CNR Virus des Infections Respiratoires - France SUD Antonin Bal, Gregory Destras, Gwendolyne Burfin, Solenne Brun, Carine Moustaud, Raphaelle Lamy, Alexandre Gaymard, Maude Bouscambert-Duchamp, Florence Morfin-Sherpa, Martine Valette, Bruno Lina, Laurence Josset
EPI_ISL_420605 hCoV-19/France/ARA12384/2020 Europe / France / ARA / Lyon 2020-03-22 Institut des Agents Infectieux (IAI), Hospices Civils de Lyon CNR Virus des Infections Respiratoires - France SUD Antonin Bal, Gregory Destras, Gwendolyne Burfin, Solenne Brun, Carine Moustaud, Raphaelle Lamy, Alexandre Gaymard, Maude Bouscambert-Duchamp, Florence Morfin-Sherpa, Martine Valette, Bruno Lina, Laurence Josset
EPI_ISL_420606 hCoV-19/France/ARA12388/2020 Europe / France / ARA / Lyon 2020-03-22 Institut des Agents Infectieux (IAI), Hospices Civils de Lyon CNR Virus des Infections Respiratoires - France SUD Antonin Bal, Gregory Destras, Gwendolyne Burfin, Solenne Brun, Carine Moustaud, Raphaelle Lamy, Alexandre Gaymard, Maude Bouscambert-Duchamp, Florence Morfin-Sherpa, Martine Valette, Bruno Lina, Laurence Josset
EPI_ISL_420607 hCoV-19/France/ARA12485/2020 Europe / France / ARA / Lyon 2020-03-23 Institut des Agents Infectieux (IAI), Hospices Civils de Lyon CNR Virus des Infections Respiratoires - France SUD Antonin Bal, Gregory Destras, Gwendolyne Burfin, Solenne Brun, Carine Moustaud, Raphaelle Lamy, Alexandre Gaymard, Maude Bouscambert-Duchamp, Florence Morfin-Sherpa, Martine Valette, Bruno Lina, Laurence Josset
EPI_ISL_420608 hCoV-19/France/ARA12499/2020 Europe / France / ARA / Lyon 2020-03-23 Institut des Agents Infectieux (IAI), Hospices Civils de Lyon CNR Virus des Infections Respiratoires - France SUD Antonin Bal, Gregory Destras, Gwendolyne Burfin, Solenne Brun, Carine Moustaud, Raphaelle Lamy, Alexandre Gaymard, Maude Bouscambert-Duchamp, Florence Morfin-Sherpa, Martine Valette, Bruno Lina, Laurence Josset
EPI_ISL_420609 hCoV-19/France/ARA12524/2020 Europe / France / ARA / Lyon 2020-03-23 Institut des Agents Infectieux (IAI), Hospices Civils de Lyon CNR Virus des Infections Respiratoires - France SUD Antonin Bal, Gregory Destras, Gwendolyne Burfin, Solenne Brun, Carine Moustaud, Raphaelle Lamy, Alexandre Gaymard, Maude Bouscambert-Duchamp, Florence Morfin-Sherpa, Martine Valette, Bruno Lina, Laurence Josset
EPI_ISL_420610 hCoV-19/France/ARA12558/2020 Europe / France / ARA / Lyon 2020-03-23 Institut des Agents Infectieux (IAI), Hospices Civils de Lyon CNR Virus des Infections Respiratoires - France SUD Antonin Bal, Gregory Destras, Gwendolyne Burfin, Solenne Brun, Carine Moustaud, Raphaelle Lamy, Alexandre Gaymard, Maude Bouscambert-Duchamp, Florence Morfin-Sherpa, Martine Valette, Bruno Lina, Laurence Josset
EPI_ISL_420611 hCoV-19/France/ARA12576/2020 Europe / France / ARA / Lyon 2020-03-23 Institut des Agents Infectieux (IAI), Hospices Civils de Lyon CNR Virus des Infections Respiratoires - France SUD Antonin Bal, Gregory Destras, Gwendolyne Burfin, Solenne Brun, Carine Moustaud, Raphaelle Lamy, Alexandre Gaymard, Maude Bouscambert-Duchamp, Florence Morfin-Sherpa, Martine Valette, Bruno Lina, Laurence Josset
EPI_ISL_420613 hCoV-19/France/ARA12630/2020 Europe / France / ARA / Macon 2020-03-23 Centre Hospitalier de Macon CNR Virus des Infections Respiratoires - France SUD Antonin Bal, Gregory Destras, Gwendolyne Burfin, Solenne Brun, Carine Moustaud, Raphaelle Lamy, Alexandre Gaymard, Maude Bouscambert-Duchamp, Florence Morfin-Sherpa, Martine Valette, Bruno Lina, Laurence Josset
EPI_ISL_420614 hCoV-19/France/ARA12632/2020 Europe / France / ARA / Macon 2020-03-23 Centre Hospitalier de Macon CNR Virus des Infections Respiratoires - France SUD Antonin Bal, Gregory Destras, Gwendolyne Burfin, Solenne Brun, Carine Moustaud, Raphaelle Lamy, Alexandre Gaymard, Maude Bouscambert-Duchamp, Florence Morfin-Sherpa, Martine Valette, Bruno Lina, Laurence Josset
EPI_ISL_420615 hCoV-19/France/ARA12759/2020 Europe / France / ARA / Lyon 2020-03-23 Institut des Agents Infectieux (IAI), Hospices Civils de Lyon CNR Virus des Infections Respiratoires - France SUD Antonin Bal, Gregory Destras, Gwendolyne Burfin, Solenne Brun, Carine Moustaud, Raphaelle Lamy, Alexandre Gaymard, Maude Bouscambert-Duchamp, Florence Morfin-Sherpa, Martine Valette, Bruno Lina, Laurence Josset
EPI_ISL_420616 hCoV-19/France/ARA1284/2020 Europe / France / ARA / Lyon 2020-03-23 Institut des Agents Infectieux (IAI), Hospices Civils de Lyon CNR Virus des Infections Respiratoires - France SUD Antonin Bal, Gregory Destras, Gwendolyne Burfin, Solenne Brun, Carine Moustaud, Raphaelle Lamy, Alexandre Gaymard, Maude Bouscambert-Duchamp, Florence Morfin-Sherpa, Martine Valette, Bruno Lina, Laurence Josset
EPI_ISL_420617 hCoV-19/France/ARA12877/2020 Europe / France / ARA / Lyon 2020-03-23 Centre Hospitalier Saint Joseph Saint Luc CNR Virus des Infections Respiratoires - France SUD Antonin Bal, Gregory Destras, Gwendolyne Burfin, Solenne Brun, Carine Moustaud, Raphaelle Lamy, Alexandre Gaymard, Maude

Bouscambert-Duchamp, Florence Morfin-Sherpa, Martine Valette, Bruno Lina, Laurence Josset
EPI_ISL_420618 hCoV-19/France/ARA12915/2020 Europe / France / ARA / Lyon 2020-03-23 Institut des Agents Infectieux (IAI), Hospices Civils de Lyon CNR Virus des Infections Respiratoires - France SUD Antonin Bal, Gregory Destras, Gwendolyne Burfin, Solenne Brun, Carine Moustaud, Raphaelle Lamy, Alexandre Gaymard, Maude Bouscambert-Duchamp, Florence Morfin-Sherpa, Martine Valette, Bruno Lina, Laurence Josset
EPI_ISL_420619 hCoV-19/France/ARA12973/2020 Europe / France / ARA / Lyon 2020-03-23 Institut des Agents Infectieux (IAI), Hospices Civils de Lyon CNR Virus des Infections Respiratoires - France SUD Antonin Bal, Gregory Destras, Gwendolyne Burfin, Solenne Brun, Carine Moustaud, Raphaelle Lamy, Alexandre Gaymard, Maude Bouscambert-Duchamp, Florence Morfin-Sherpa, Martine Valette, Bruno Lina, Laurence Josset
EPI_ISL_420620 hCoV-19/France/ARA12996/2020 Europe / France / ARA / Bourg-en-Bresse 2020-03-23 Centre Hospitalier de Bourg en Bresse CNR Virus des Infections Respiratoires - France SUD Antonin Bal, Gregory Destras, Gwendolyne Burfin, Solenne Brun, Carine Moustaud, Raphaelle Lamy, Alexandre Gaymard, Maude Bouscambert-Duchamp, Florence Morfin-Sherpa, Martine Valette, Bruno Lina, Laurence Josset
EPI_ISL_420621 hCoV-19/France/ARA1307/2020 Europe / France / ARA / Lyon 2020-03-24 Institut des Agents Infectieux (IAI), Hospices Civils de Lyon CNR Virus des Infections Respiratoires - France SUD Antonin Bal, Gregory Destras, Gwendolyne Burfin, Solenne Brun, Carine Moustaud, Raphaelle Lamy, Alexandre Gaymard, Maude Bouscambert-Duchamp, Florence Morfin-Sherpa, Martine Valette, Bruno Lina, Laurence Josset
EPI_ISL_420623 hCoV-19/France/ARA13095/2020 Europe / France / ARA / Lyon 2020-03-24 Institut des Agents Infectieux (IAI), Hospices Civils de Lyon CNR Virus des Infections Respiratoires - France SUD Antonin Bal, Gregory Destras, Gwendolyne Burfin, Solenne Brun, Carine Moustaud, Raphaelle Lamy, Alexandre Gaymard, Maude Bouscambert-Duchamp, Florence Morfin-Sherpa, Martine Valette, Bruno Lina, Laurence Josset
EPI_ISL_420624 hCoV-19/France/ARA13160/2020 Europe / France / ARA / Lyon 2020-03-24 Institut des Agents Infectieux (IAI), Hospices Civils de Lyon CNR Virus des Infections Respiratoires - France SUD Antonin Bal, Gregory Destras, Gwendolyne Burfin, Solenne Brun, Carine Moustaud, Raphaelle Lamy, Alexandre Gaymard, Maude Bouscambert-Duchamp, Florence Morfin-Sherpa, Martine Valette, Bruno Lina, Laurence Josset
EPI_ISL_420625 hCoV-19/France/ARA1322/2020 Europe / France / ARA / Lyon 2020-03-24 Institut des Agents Infectieux (IAI), Hospices Civils de Lyon CNR Virus des Infections Respiratoires - France SUD Antonin Bal, Gregory Destras, Gwendolyne Burfin, Solenne Brun, Carine Moustaud, Raphaelle Lamy, Alexandre Gaymard, Maude Bouscambert-Duchamp, Florence Morfin-Sherpa, Martine Valette, Bruno Lina, Laurence Josset
EPI_ISL_420628 hCoV-19/USA/VA-DCLS-0024/2020 North America / USA / Virginia 2020-03-13 Virginia DCLS Virginia DCLS Virginia DCLS
EPI_ISL_420629 hCoV-19/USA/VA-DCLS-0034/2020 North America / USA / Virginia 2020-03-20 Virginia DCLS Virginia DCLS Virginia DCLS
EPI_ISL_420630 hCoV-19/USA/VA-DCLS-0038/2020 North America / USA / Virginia 2020-03-23 Virginia DCLS Virginia DCLS Virginia DCLS
EPI_ISL_420633 hCoV-19/England/20130055502/2020 Europe / United Kingdom / England 2020-03-19 Respiratory Virus Unit, Microbiology Services Colindale, Public Health England Respiratory Virus Unit, Microbiology Services Colindale, Public Health England Monica Galiano, Shahjahan Miah, Angie Lackenby, Omolola Akinbami, Tiina Talts, Leena Bhaw, Richard Myers, Steven Platt, Kirstin Edwards, Jonathan Hubb, Joanna Ellis, Maria Zambon
EPI_ISL_420636 hCoV-19/England/20136085404/2020 Europe / United Kingdom / England 2020-03-24 Respiratory Virus Unit, Microbiology Services Colindale, Public Health England Respiratory Virus Unit, Microbiology Services Colindale, Public Health England Monica Galiano, Shahjahan Miah, Angie Lackenby, Omolola Akinbami, Tiina Talts, Leena Bhaw, Richard Myers, Steven Platt, Kirstin Edwards, Jonathan Hubb, Joanna Ellis, Maria Zambon
EPI_ISL_420640 hCoV-19/England/20136088602/2020 Europe / United Kingdom / England 2020-03-23 Respiratory Virus Unit, Microbiology Services Colindale, Public Health England Respiratory Virus Unit, Microbiology Services Colindale, Public Health England Monica Galiano, Shahjahan Miah, Angie Lackenby, Omolola Akinbami, Tiina Talts, Leena Bhaw, Richard Myers, Steven Platt, Kirstin Edwards, Jonathan Hubb, Joanna Ellis, Maria Zambon
EPI_ISL_420642 hCoV-19/England/20136089402/2020 Europe / United Kingdom / England 2020-03-23 Respiratory Virus Unit, Microbiology Services Colindale, Public Health England Respiratory Virus Unit, Microbiology Services Colindale, Public Health England Monica Galiano, Shahjahan Miah, Angie Lackenby, Omolola Akinbami, Tiina Talts, Leena Bhaw, Richard Myers, Steven Platt, Kirstin Edwards, Jonathan Hubb, Joanna Ellis, Maria Zambon
EPI_ISL_420643 hCoV-19/England/20136094704/2020 Europe / United Kingdom / England 2020-03-26 Respiratory Virus Unit, Microbiology Services Colindale, Public Health England Respiratory Virus Unit, Microbiology Services Colindale, Public Health England Monica Galiano, Shahjahan Miah, Angie Lackenby, Omolola Akinbami, Tiina Talts, Leena Bhaw, Richard Myers, Steven Platt, Kirstin Edwards, Jonathan Hubb, Joanna Ellis, Maria Zambon
EPI_ISL_420648 hCoV-19/England/20136096504/2020 Europe / United Kingdom / England 2020-03-23 Respiratory Virus Unit, Microbiology Services Colindale, Public Health England Respiratory Virus Unit, Microbiology Services Colindale, Public Health England Monica Galiano, Shahjahan Miah, Angie Lackenby, Omolola Akinbami, Tiina Talts, Leena Bhaw, Richard Myers, Steven Platt, Kirstin Edwards, Jonathan Hubb, Joanna Ellis, Maria Zambon
EPI_ISL_420652 hCoV-19/England/20136097204/2020 Europe / United Kingdom / England 2020-03-23 Respiratory Virus Unit, Microbiology Services Colindale, Public Health England Respiratory Virus Unit, Microbiology Services Colindale, Public Health England Monica Galiano, Shahjahan Miah, Angie Lackenby, Omolola Akinbami, Tiina Talts, Leena Bhaw, Richard Myers, Steven Platt, Kirstin Edwards, Jonathan Hubb, Joanna Ellis, Maria Zambon

EPI_ISL_420661  hCoV-19/England/20139013406/2020        Europe / United Kingdom / England       2020-03-27
Respiratory Virus Unit, Microbiology Services Colindale, Public Health England  Respiratory Virus Unit, Microbiology Services Colindale, Public Health England  Monica Galiano, Shahjahan Miah, Angie Lackenby, Omolola Akinbami, Tiina Talts, Leena Bhaw, Richard Myers, Steven Platt, Kirstin Edwards, Jonathan Hubb, Joanna Ellis, Maria Zambon
EPI_ISL_420670  hCoV-19/England/20139037504/2020        Europe / United Kingdom / England       2020-03-27
Respiratory Virus Unit, Microbiology Services Colindale, Public Health England  Respiratory Virus Unit, Microbiology Services Colindale, Public Health England  Monica Galiano, Shahjahan Miah, Angie Lackenby, Omolola Akinbami, Tiina Talts, Leena Bhaw, Richard Myers, Steven Platt, Kirstin Edwards, Jonathan Hubb, Joanna Ellis, Maria Zambon
EPI_ISL_420683  hCoV-19/England/20139044504/2020        Europe / United Kingdom / England       2020-03-28
Respiratory Virus Unit, Microbiology Services Colindale, Public Health England  Respiratory Virus Unit, Microbiology Services Colindale, Public Health England  Monica Galiano, Shahjahan Miah, Angie Lackenby, Omolola Akinbami, Tiina Talts, Leena Bhaw, Richard Myers, Steven Platt, Kirstin Edwards, Jonathan Hubb, Joanna Ellis, Maria Zambon
EPI_ISL_420686  hCoV-19/England/20139045804/2020        Europe / United Kingdom / England       2020-03-28
Respiratory Virus Unit, Microbiology Services Colindale, Public Health England  Respiratory Virus Unit, Microbiology Services Colindale, Public Health England  Monica Galiano, Shahjahan Miah, Angie Lackenby, Omolola Akinbami, Tiina Talts, Leena Bhaw, Richard Myers, Steven Platt, Kirstin Edwards, Jonathan Hubb, Joanna Ellis, Maria Zambon
EPI_ISL_420687  hCoV-19/England/20139045904/2020        Europe / United Kingdom / England       2020-03-28
Respiratory Virus Unit, Microbiology Services Colindale, Public Health England  Respiratory Virus Unit, Microbiology Services Colindale, Public Health England  Monica Galiano, Shahjahan Miah, Angie Lackenby, Omolola Akinbami, Tiina Talts, Leena Bhaw, Richard Myers, Steven Platt, Kirstin Edwards, Jonathan Hubb, Joanna Ellis, Maria Zambon
EPI_ISL_420688  hCoV-19/England/20139046504/2020        Europe / United Kingdom / England       2020-03-28
Respiratory Virus Unit, Microbiology Services Colindale, Public Health England  Respiratory Virus Unit, Microbiology Services Colindale, Public Health England  Monica Galiano, Shahjahan Miah, Angie Lackenby, Omolola Akinbami, Tiina Talts, Leena Bhaw, Richard Myers, Steven Platt, Kirstin Edwards, Jonathan Hubb, Joanna Ellis, Maria Zambon
EPI_ISL_420689  hCoV-19/England/20139046604/2020        Europe / United Kingdom / England       2020-03-28
Respiratory Virus Unit, Microbiology Services Colindale, Public Health England  Respiratory Virus Unit, Microbiology Services Colindale, Public Health England  Monica Galiano, Shahjahan Miah, Angie Lackenby, Omolola Akinbami, Tiina Talts, Leena Bhaw, Richard Myers, Steven Platt, Kirstin Edwards, Jonathan Hubb, Joanna Ellis, Maria Zambon
EPI_ISL_420690  hCoV-19/England/20139046704/2020        Europe / United Kingdom / England       2020-03-28
Respiratory Virus Unit, Microbiology Services Colindale, Public Health England  Respiratory Virus Unit, Microbiology Services Colindale, Public Health England  Monica Galiano, Shahjahan Miah, Angie Lackenby, Omolola Akinbami, Tiina Talts, Leena Bhaw, Richard Myers, Steven Platt, Kirstin Edwards, Jonathan Hubb, Joanna Ellis, Maria Zambon
EPI_ISL_420692  hCoV-19/England/20139047104/2020        Europe / United Kingdom / England       2020-03-29
Respiratory Virus Unit, Microbiology Services Colindale, Public Health England  Respiratory Virus Unit, Microbiology Services Colindale, Public Health England  Monica Galiano, Shahjahan Miah, Angie Lackenby, Omolola Akinbami, Tiina Talts, Leena Bhaw, Richard Myers, Steven Platt, Kirstin Edwards, Jonathan Hubb, Joanna Ellis, Maria Zambon
EPI_ISL_420693  hCoV-19/England/20139047404/2020        Europe / United Kingdom / England       2020-03-28
Respiratory Virus Unit, Microbiology Services Colindale, Public Health England  Respiratory Virus Unit, Microbiology Services Colindale, Public Health England  Monica Galiano, Shahjahan Miah, Angie Lackenby, Omolola Akinbami, Tiina Talts, Leena Bhaw, Richard Myers, Steven Platt, Kirstin Edwards, Jonathan Hubb, Joanna Ellis, Maria Zambon
EPI_ISL_420694  hCoV-19/England/20139047504/2020        Europe / United Kingdom / England       2020-03-28
Respiratory Virus Unit, Microbiology Services Colindale, Public Health England  Respiratory Virus Unit, Microbiology Services Colindale, Public Health England  Monica Galiano, Shahjahan Miah, Angie Lackenby, Omolola Akinbami, Tiina Talts, Leena Bhaw, Richard Myers, Steven Platt, Kirstin Edwards, Jonathan Hubb, Joanna Ellis, Maria Zambon
EPI_ISL_420697  hCoV-19/England/20139047804/2020        Europe / United Kingdom / England       2020-03-29
Respiratory Virus Unit, Microbiology Services Colindale, Public Health England  Respiratory Virus Unit, Microbiology Services Colindale, Public Health England  Monica Galiano, Shahjahan Miah, Angie Lackenby, Omolola Akinbami, Tiina Talts, Leena Bhaw, Richard Myers, Steven Platt, Kirstin Edwards, Jonathan Hubb, Joanna Ellis, Maria Zambon
EPI_ISL_420698  hCoV-19/England/20139048804/2020        Europe / United Kingdom / England       2020-03-28
Respiratory Virus Unit, Microbiology Services Colindale, Public Health England  Respiratory Virus Unit, Microbiology Services Colindale, Public Health England  Monica Galiano, Shahjahan Miah, Angie Lackenby, Omolola Akinbami, Tiina Talts, Leena Bhaw, Richard Myers, Steven Platt, Kirstin Edwards, Jonathan Hubb, Joanna Ellis, Maria Zambon
EPI_ISL_421900  hCoV-19/England/20139027004/2020        Europe / United Kingdom / England       2020-03-26
Respiratory Virus Unit, Microbiology Services Colindale, Public Health England  Respiratory Virus Unit, Microbiology Services Colindale, Public Health England  Monica Galiano, Shahjahan Miah, Angie Lackenby, Omolola Akinbami, Tiina Talts, Leena Bhaw, Richard Myers, Steven Platt, Kirstin Edwards, Jonathan Hubb, Joanna Ellis, Maria Zambon

EPI_ISL_421903	hCoV-19/England/20139027304/2020	Europe / United Kingdom / England	2020-03-26	Respiratory Virus Unit, Microbiology Services Colindale, Public Health England	Respiratory Virus Unit, Microbiology Services Colindale, Public Health England	Monica Galiano, Shahjahan Miah, Angie Lackenby, Omolola Akinbami, Tiina Talts, Leena Bhaw, Richard Myers, Steven Platt, Kirstin Edwards, Jonathan Hubb, Joanna Ellis, Maria Zambon
EPI_ISL_421904	hCoV-19/England/20139027404/2020	Europe / United Kingdom / England	2020-03-26	Respiratory Virus Unit, Microbiology Services Colindale, Public Health England	Respiratory Virus Unit, Microbiology Services Colindale, Public Health England	Monica Galiano, Shahjahan Miah, Angie Lackenby, Omolola Akinbami, Tiina Talts, Leena Bhaw, Richard Myers, Steven Platt, Kirstin Edwards, Jonathan Hubb, Joanna Ellis, Maria Zambon
EPI_ISL_421907	hCoV-19/England/20139027904/2020	Europe / United Kingdom / England	2020-03-26	Respiratory Virus Unit, Microbiology Services Colindale, Public Health England	Respiratory Virus Unit, Microbiology Services Colindale, Public Health England	Monica Galiano, Shahjahan Miah, Angie Lackenby, Omolola Akinbami, Tiina Talts, Leena Bhaw, Richard Myers, Steven Platt, Kirstin Edwards, Jonathan Hubb, Joanna Ellis, Maria Zambon
EPI_ISL_421909	hCoV-19/England/20139029104/2020	Europe / United Kingdom / England	2020-03-26	Respiratory Virus Unit, Microbiology Services Colindale, Public Health England	Respiratory Virus Unit, Microbiology Services Colindale, Public Health England	Monica Galiano, Shahjahan Miah, Angie Lackenby, Omolola Akinbami, Tiina Talts, Leena Bhaw, Richard Myers, Steven Platt, Kirstin Edwards, Jonathan Hubb, Joanna Ellis, Maria Zambon
EPI_ISL_421910	hCoV-19/England/20139029304/2020	Europe / United Kingdom / England	2020-03-26	Respiratory Virus Unit, Microbiology Services Colindale, Public Health England	Respiratory Virus Unit, Microbiology Services Colindale, Public Health England	Monica Galiano, Shahjahan Miah, Angie Lackenby, Omolola Akinbami, Tiina Talts, Leena Bhaw, Richard Myers, Steven Platt, Kirstin Edwards, Jonathan Hubb, Joanna Ellis, Maria Zambon
EPI_ISL_421911	hCoV-19/England/20139029404/2020	Europe / United Kingdom / England	2020-03-26	Respiratory Virus Unit, Microbiology Services Colindale, Public Health England	Respiratory Virus Unit, Microbiology Services Colindale, Public Health England	Monica Galiano, Shahjahan Miah, Angie Lackenby, Omolola Akinbami, Tiina Talts, Leena Bhaw, Richard Myers, Steven Platt, Kirstin Edwards, Jonathan Hubb, Joanna Ellis, Maria Zambon
EPI_ISL_421912	hCoV-19/England/20139030404/2020	Europe / United Kingdom / England	2020-03-26	Respiratory Virus Unit, Microbiology Services Colindale, Public Health England	Respiratory Virus Unit, Microbiology Services Colindale, Public Health England	Monica Galiano, Shahjahan Miah, Angie Lackenby, Omolola Akinbami, Tiina Talts, Leena Bhaw, Richard Myers, Steven Platt, Kirstin Edwards, Jonathan Hubb, Joanna Ellis, Maria Zambon
EPI_ISL_421914	hCoV-19/England/20139031404/2020	Europe / United Kingdom / England	2020-03-27	Respiratory Virus Unit, Microbiology Services Colindale, Public Health England	Respiratory Virus Unit, Microbiology Services Colindale, Public Health England	Monica Galiano, Shahjahan Miah, Angie Lackenby, Omolola Akinbami, Tiina Talts, Leena Bhaw, Richard Myers, Steven Platt, Kirstin Edwards, Jonathan Hubb, Joanna Ellis, Maria Zambon
EPI_ISL_421915	hCoV-19/England/20139031504/2020	Europe / United Kingdom / England	2020-03-27	Respiratory Virus Unit, Microbiology Services Colindale, Public Health England	Respiratory Virus Unit, Microbiology Services Colindale, Public Health England	Monica Galiano, Shahjahan Miah, Angie Lackenby, Omolola Akinbami, Tiina Talts, Leena Bhaw, Richard Myers, Steven Platt, Kirstin Edwards, Jonathan Hubb, Joanna Ellis, Maria Zambon
EPI_ISL_421916	hCoV-19/England/20139031804/2020	Europe / United Kingdom / England	2020-03-27	Respiratory Virus Unit, Microbiology Services Colindale, Public Health England	Respiratory Virus Unit, Microbiology Services Colindale, Public Health England	Monica Galiano, Shahjahan Miah, Angie Lackenby, Omolola Akinbami, Tiina Talts, Leena Bhaw, Richard Myers, Steven Platt, Kirstin Edwards, Jonathan Hubb, Joanna Ellis, Maria Zambon
EPI_ISL_421918	hCoV-19/England/20139032404/2020	Europe / United Kingdom / England	2020-03-27	Respiratory Virus Unit, Microbiology Services Colindale, Public Health England	Respiratory Virus Unit, Microbiology Services Colindale, Public Health England	Monica Galiano, Shahjahan Miah, Angie Lackenby, Omolola Akinbami, Tiina Talts, Leena Bhaw, Richard Myers, Steven Platt, Kirstin Edwards, Jonathan Hubb, Joanna Ellis, Maria Zambon
EPI_ISL_421919	hCoV-19/England/20139033004/2020	Europe / United Kingdom / England	2020-03-27	Respiratory Virus Unit, Microbiology Services Colindale, Public Health England	Respiratory Virus Unit, Microbiology Services Colindale, Public Health England	Monica Galiano, Shahjahan Miah, Angie Lackenby, Omolola Akinbami, Tiina Talts, Leena Bhaw, Richard Myers, Steven Platt, Kirstin Edwards, Jonathan Hubb, Joanna Ellis, Maria Zambon
EPI_ISL_421920	hCoV-19/England/20139033304/2020	Europe / United Kingdom / England	2020-03-27	Respiratory Virus Unit, Microbiology Services Colindale, Public Health England	Respiratory Virus Unit, Microbiology Services Colindale, Public Health England	Monica Galiano, Shahjahan Miah, Angie Lackenby, Omolola Akinbami, Tiina Talts, Leena Bhaw, Richard Myers, Steven Platt, Kirstin Edwards, Jonathan Hubb, Joanna Ellis, Maria Zambon
EPI_ISL_421921	hCoV-19/England/20139033404/2020	Europe / United Kingdom / England	2020-03-27	Respiratory Virus Unit, Microbiology Services Colindale, Public Health England	Respiratory Virus Unit, Microbiology Services Colindale, Public Health England	Monica Galiano, Shahjahan Miah, Angie Lackenby, Omolola Akinbami, Tiina Talts, Leena Bhaw, Richard Myers, Steven Platt, Kirstin Edwards, Jonathan Hubb, Joanna Ellis, Maria Zambon

EPI_ISL_421923 hCoV-19/England/20139036504/2020 Europe / United Kingdom / England 2020-03-27 Respiratory Virus Unit, Microbiology Services Colindale, Public Health England Respiratory Virus Unit, Microbiology Services Colindale, Public Health England Monica Galiano, Shahjahan Miah, Angie Lackenby, Omolola Akinbami, Tiina Talts, Leena Bhaw, Richard Myers, Steven Platt, Kirstin Edwards, Jonathan Hubb, Joanna Ellis, Maria Zambon
EPI_ISL_421932 hCoV-19/England/20144013504/2020 Europe / United Kingdom / England 2020-03-31 Respiratory Virus Unit, Microbiology Services Colindale, Public Health England Respiratory Virus Unit, Microbiology Services Colindale, Public Health England Monica Galiano, Shahjahan Miah, Angie Lackenby, Omolola Akinbami, Tiina Talts, Leena Bhaw, Richard Myers, Steven Platt, Kirstin Edwards, Jonathan Hubb, Joanna Ellis, Maria Zambon
EPI_ISL_421935 hCoV-19/England/20144014604/2020 Europe / United Kingdom / England 2020-03-31 Respiratory Virus Unit, Microbiology Services Colindale, Public Health England Respiratory Virus Unit, Microbiology Services Colindale, Public Health England Monica Galiano, Shahjahan Miah, Angie Lackenby, Omolola Akinbami, Tiina Talts, Leena Bhaw, Richard Myers, Steven Platt, Kirstin Edwards, Jonathan Hubb, Joanna Ellis, Maria Zambon
EPI_ISL_421936 hCoV-19/England/20144014704/2020 Europe / United Kingdom / England 2020-03-31 Respiratory Virus Unit, Microbiology Services Colindale, Public Health England Respiratory Virus Unit, Microbiology Services Colindale, Public Health England Monica Galiano, Shahjahan Miah, Angie Lackenby, Omolola Akinbami, Tiina Talts, Leena Bhaw, Richard Myers, Steven Platt, Kirstin Edwards, Jonathan Hubb, Joanna Ellis, Maria Zambon
EPI_ISL_421937 hCoV-19/England/20144016604/2020 Europe / United Kingdom / England 2020-03-30 Respiratory Virus Unit, Microbiology Services Colindale, Public Health England Respiratory Virus Unit, Microbiology Services Colindale, Public Health England Monica Galiano, Shahjahan Miah, Angie Lackenby, Omolola Akinbami, Tiina Talts, Leena Bhaw, Richard Myers, Steven Platt, Kirstin Edwards, Jonathan Hubb, Joanna Ellis, Maria Zambon
EPI_ISL_421938 hCoV-19/England/20144018204/2020 Europe / United Kingdom / England 2020-03-30 Respiratory Virus Unit, Microbiology Services Colindale, Public Health England Respiratory Virus Unit, Microbiology Services Colindale, Public Health England Monica Galiano, Shahjahan Miah, Angie Lackenby, Omolola Akinbami, Tiina Talts, Leena Bhaw, Richard Myers, Steven Platt, Kirstin Edwards, Jonathan Hubb, Joanna Ellis, Maria Zambon
EPI_ISL_421939 hCoV-19/England/20144018704/2020 Europe / United Kingdom / England 2020-04-01 Respiratory Virus Unit, Microbiology Services Colindale, Public Health England Respiratory Virus Unit, Microbiology Services Colindale, Public Health England Monica Galiano, Shahjahan Miah, Angie Lackenby, Omolola Akinbami, Tiina Talts, Leena Bhaw, Richard Myers, Steven Platt, Kirstin Edwards, Jonathan Hubb, Joanna Ellis, Maria Zambon
EPI_ISL_421940 hCoV-19/England/20144019304/2020 Europe / United Kingdom / England 2020-03-30 Respiratory Virus Unit, Microbiology Services Colindale, Public Health England Respiratory Virus Unit, Microbiology Services Colindale, Public Health England Monica Galiano, Shahjahan Miah, Angie Lackenby, Omolola Akinbami, Tiina Talts, Leena Bhaw, Richard Myers, Steven Platt, Kirstin Edwards, Jonathan Hubb, Joanna Ellis, Maria Zambon
EPI_ISL_421942 hCoV-19/England/20144020004/2020 Europe / United Kingdom / England 2020-03-31 Respiratory Virus Unit, Microbiology Services Colindale, Public Health England Respiratory Virus Unit, Microbiology Services Colindale, Public Health England Monica Galiano, Shahjahan Miah, Angie Lackenby, Omolola Akinbami, Tiina Talts, Leena Bhaw, Richard Myers, Steven Platt, Kirstin Edwards, Jonathan Hubb, Joanna Ellis, Maria Zambon
EPI_ISL_421945 hCoV-19/England/20144035904/2020 Europe / United Kingdom / England 2020-03-31 Respiratory Virus Unit, Microbiology Services Colindale, Public Health England Respiratory Virus Unit, Microbiology Services Colindale, Public Health England Monica Galiano, Shahjahan Miah, Angie Lackenby, Omolola Akinbami, Tiina Talts, Leena Bhaw, Richard Myers, Steven Platt, Kirstin Edwards, Jonathan Hubb, Joanna Ellis, Maria Zambon
EPI_ISL_421946 hCoV-19/England/20144037304/2020 Europe / United Kingdom / England 2020-03-31 Respiratory Virus Unit, Microbiology Services Colindale, Public Health England Respiratory Virus Unit, Microbiology Services Colindale, Public Health England Monica Galiano, Shahjahan Miah, Angie Lackenby, Omolola Akinbami, Tiina Talts, Leena Bhaw, Richard Myers, Steven Platt, Kirstin Edwards, Jonathan Hubb, Joanna Ellis, Maria Zambon
EPI_ISL_421947 hCoV-19/England/20144038204/2020 Europe / United Kingdom / England 2020-03-31 Respiratory Virus Unit, Microbiology Services Colindale, Public Health England Respiratory Virus Unit, Microbiology Services Colindale, Public Health England Monica Galiano, Shahjahan Miah, Angie Lackenby, Omolola Akinbami, Tiina Talts, Leena Bhaw, Richard Myers, Steven Platt, Kirstin Edwards, Jonathan Hubb, Joanna Ellis, Maria Zambon
EPI_ISL_421948 hCoV-19/England/20144049404/2020 Europe / United Kingdom / England 2020-03-31 Respiratory Virus Unit, Microbiology Services Colindale, Public Health England Respiratory Virus Unit, Microbiology Services Colindale, Public Health England Monica Galiano, Shahjahan Miah, Angie Lackenby, Omolola Akinbami, Tiina Talts, Leena Bhaw, Richard Myers, Steven Platt, Kirstin Edwards, Jonathan Hubb, Joanna Ellis, Maria Zambon
EPI_ISL_421951 hCoV-19/England/20144049704/2020 Europe / United Kingdom / England 2020-03-31 Respiratory Virus Unit, Microbiology Services Colindale, Public Health England Respiratory Virus Unit, Microbiology Services Colindale, Public Health England Monica Galiano, Shahjahan Miah, Angie Lackenby, Omolola Akinbami, Tiina Talts, Leena Bhaw, Richard Myers, Steven Platt, Kirstin Edwards, Jonathan Hubb, Joanna Ellis, Maria Zambon

EPI_ISL_421956 hCoV-19/England/20144065604/2020 Europe / United Kingdom / England 2020-03-31 Respiratory Virus Unit, Microbiology Services Colindale, Public Health England Respiratory Virus Unit, Microbiology Services Colindale, Public Health England Monica Galiano, Shahjahan Miah, Angie Lackenby, Omolola Akinbami, Tiina Talts, Leena Bhaw, Richard Myers, Steven Platt, Kirstin Edwards, Jonathan Hubb, Joanna Ellis, Maria Zambon
EPI_ISL_421957 hCoV-19/England/20144067504/2020 Europe / United Kingdom / England 2020-03-20 Respiratory Virus Unit, Microbiology Services Colindale, Public Health England Respiratory Virus Unit, Microbiology Services Colindale, Public Health England Monica Galiano, Shahjahan Miah, Angie Lackenby, Omolola Akinbami, Tiina Talts, Leena Bhaw, Richard Myers, Steven Platt, Kirstin Edwards, Jonathan Hubb, Joanna Ellis, Maria Zambon
EPI_ISL_421958 hCoV-19/England/20144071004/2020 Europe / United Kingdom / England 2020-04-01 Respiratory Virus Unit, Microbiology Services Colindale, Public Health England Respiratory Virus Unit, Microbiology Services Colindale, Public Health England Monica Galiano, Shahjahan Miah, Angie Lackenby, Omolola Akinbami, Tiina Talts, Leena Bhaw, Richard Myers, Steven Platt, Kirstin Edwards, Jonathan Hubb, Joanna Ellis, Maria Zambon
EPI_ISL_421962 hCoV-19/England/20144072804/2020 Europe / United Kingdom / England 2020-04-01 Respiratory Virus Unit, Microbiology Services Colindale, Public Health England Respiratory Virus Unit, Microbiology Services Colindale, Public Health England Monica Galiano, Shahjahan Miah, Angie Lackenby, Omolola Akinbami, Tiina Talts, Leena Bhaw, Richard Myers, Steven Platt, Kirstin Edwards, Jonathan Hubb, Joanna Ellis, Maria Zambon
EPI_ISL_421963 hCoV-19/England/20144074804/2020 Europe / United Kingdom / England 2020-04-01 Respiratory Virus Unit, Microbiology Services Colindale, Public Health England Respiratory Virus Unit, Microbiology Services Colindale, Public Health England Monica Galiano, Shahjahan Miah, Angie Lackenby, Omolola Akinbami, Tiina Talts, Leena Bhaw, Richard Myers, Steven Platt, Kirstin Edwards, Jonathan Hubb, Joanna Ellis, Maria Zambon
EPI_ISL_421964 hCoV-19/England/20144076304/2020 Europe / United Kingdom / England 2020-04-01 Respiratory Virus Unit, Microbiology Services Colindale, Public Health England Respiratory Virus Unit, Microbiology Services Colindale, Public Health England Monica Galiano, Shahjahan Miah, Angie Lackenby, Omolola Akinbami, Tiina Talts, Leena Bhaw, Richard Myers, Steven Platt, Kirstin Edwards, Jonathan Hubb, Joanna Ellis, Maria Zambon
EPI_ISL_421982 hCoV-19/England/20146019704/2020 Europe / United Kingdom / England 2020-04-01 Respiratory Virus Unit, Microbiology Services Colindale, Public Health England Respiratory Virus Unit, Microbiology Services Colindale, Public Health England Monica Galiano, Shahjahan Miah, Angie Lackenby, Omolola Akinbami, Tiina Talts, Leena Bhaw, Richard Myers, Steven Platt, Kirstin Edwards, Jonathan Hubb, Joanna Ellis, Maria Zambon
EPI_ISL_422039 hCoV-19/Wales/PHWC-25779/2020 Europe / United Kingdom / Wales 2020-03-24 Wales Specialist Virology Centre Public Health Wales Microbiology Cardiff Catherine Moore, Johnathan Evans, Malorie Perry, Simon Cottrell, Alec Birchley, Alexander Adams, Amy Gaskin, Bree Gatica-Wilcox, Jason Coombes, Lauren Gilbert, Lee Graham, Nicole Pacchiarini, Sara Kumziene-Summerhayes, Sarah Taylor, Sophie Jones, Sara Rey, Matthew Bull, Joanne Watkins, Sally Corden, Tom Connor
EPI_ISL_422040 hCoV-19/Wales/PHWC-26D1C/2020 Europe / United Kingdom / Wales 2020-03-30 Wales Specialist Virology Centre Public Health Wales Microbiology Cardiff Catherine Moore, Johnathan Evans, Malorie Perry, Simon Cottrell, Alec Birchley, Alexander Adams, Amy Gaskin, Bree Gatica-Wilcox, Jason Coombes, Lauren Gilbert, Lee Graham, Nicole Pacchiarini, Sara Kumziene-Summerhayes, Sarah Taylor, Sophie Jones, Sara Rey, Matthew Bull, Joanne Watkins, Sally Corden, Tom Connor
EPI_ISL_422041 hCoV-19/Wales/PHWC-25C3E/2020 Europe / United Kingdom / Wales 2020-03-25 Wales Specialist Virology Centre Public Health Wales Microbiology Cardiff Catherine Moore, Johnathan Evans, Malorie Perry, Simon Cottrell, Alec Birchley, Alexander Adams, Amy Gaskin, Bree Gatica-Wilcox, Jason Coombes, Lauren Gilbert, Lee Graham, Nicole Pacchiarini, Sara Kumziene-Summerhayes, Sarah Taylor, Sophie Jones, Sara Rey, Matthew Bull, Joanne Watkins, Sally Corden, Tom Connor
EPI_ISL_422042 hCoV-19/Wales/PHWC-264BD/2020 Europe / United Kingdom / Wales 2020-03-28 Wales Specialist Virology Centre Public Health Wales Microbiology Cardiff Catherine Moore, Johnathan Evans, Malorie Perry, Simon Cottrell, Alec Birchley, Alexander Adams, Amy Gaskin, Bree Gatica-Wilcox, Jason Coombes, Lauren Gilbert, Lee Graham, Nicole Pacchiarini, Sara Kumziene-Summerhayes, Sarah Taylor, Sophie Jones, Sara Rey, Matthew Bull, Joanne Watkins, Sally Corden, Tom Connor
EPI_ISL_422043 hCoV-19/Wales/PHWC-269DC/2020 Europe / United Kingdom / Wales 2020-03-29 Wales Specialist Virology Centre Public Health Wales Microbiology Cardiff Catherine Moore, Johnathan Evans, Malorie Perry, Simon Cottrell, Alec Birchley, Alexander Adams, Amy Gaskin, Bree Gatica-Wilcox, Jason Coombes, Lauren Gilbert, Lee Graham, Nicole Pacchiarini, Sara Kumziene-Summerhayes, Sarah Taylor, Sophie Jones, Sara Rey, Matthew Bull, Joanne Watkins, Sally Corden, Tom Connor
EPI_ISL_422044 hCoV-19/Wales/PHWC-26E19/2020 Europe / United Kingdom / Wales 2020-03-29 Wales Specialist Virology Centre Public Health Wales Microbiology Cardiff Catherine Moore, Johnathan Evans, Malorie Perry, Simon Cottrell, Alec Birchley, Alexander Adams, Amy Gaskin, Bree Gatica-Wilcox, Jason Coombes, Lauren Gilbert, Lee Graham, Nicole Pacchiarini, Sara Kumziene-Summerhayes, Sarah Taylor, Sophie Jones, Sara Rey, Matthew Bull, Joanne Watkins, Sally Corden, Tom Connor
EPI_ISL_422045 hCoV-19/Wales/PHWC-25AF8/2020 Europe / United Kingdom / Wales 2020-03-24 Wales Specialist Virology Centre Public Health Wales Microbiology Cardiff Catherine Moore, Johnathan Evans, Malorie Perry, Simon Cottrell, Alec Birchley, Alexander Adams, Amy Gaskin, Bree Gatica-Wilcox, Jason Coombes, Lauren Gilbert, Lee Graham, Nicole Pacchiarini, Sara Kumziene-Summerhayes, Sarah Taylor, Sophie Jones, Sara Rey, Matthew Bull, Joanne Watkins, Sally Corden, Tom Connor

EPI_ISL_422046 hCoV-19/Wales/PHWC-2666C/2020 Europe / United Kingdom / Wales 2020-03-27 Wales Specialist Virology Centre Public Health Wales Microbiology Cardiff Catherine Moore, Johnathan Evans, Malorie Perry, Simon Cottrell, Alec Birchley, Alexander Adams, Amy Gaskin, Bree Gatica-Wilcox, Jason Coombes, Lauren Gilbert, Lee Graham, Nicole Pacchiarini, Sara Kumziene-Summerhayes, Sarah Taylor, Sophie Jones, Sara Rey, Matthew Bull, Joanne Watkins, Sally Corden, Tom Connor
EPI_ISL_422047 hCoV-19/Wales/PHWC-26F34/2020 Europe / United Kingdom / Wales 2020-03-30 Wales Specialist Virology Centre Public Health Wales Microbiology Cardiff Catherine Moore, Johnathan Evans, Malorie Perry, Simon Cottrell, Alec Birchley, Alexander Adams, Amy Gaskin, Bree Gatica-Wilcox, Jason Coombes, Lauren Gilbert, Lee Graham, Nicole Pacchiarini, Sara Kumziene-Summerhayes, Sarah Taylor, Sophie Jones, Sara Rey, Matthew Bull, Joanne Watkins, Sally Corden, Tom Connor
EPI_ISL_422048 hCoV-19/Wales/PHWC-26918/2020 Europe / United Kingdom / Wales 2020-03-27 Wales Specialist Virology Centre Public Health Wales Microbiology Cardiff Catherine Moore, Johnathan Evans, Malorie Perry, Simon Cottrell, Alec Birchley, Alexander Adams, Amy Gaskin, Bree Gatica-Wilcox, Jason Coombes, Lauren Gilbert, Lee Graham, Nicole Pacchiarini, Sara Kumziene-Summerhayes, Sarah Taylor, Sophie Jones, Sara Rey, Matthew Bull, Joanne Watkins, Sally Corden, Tom Connor
EPI_ISL_422049 hCoV-19/Wales/PHWC-2575B/2020 Europe / United Kingdom / Wales 2020-03-24 Wales Specialist Virology Centre Public Health Wales Microbiology Cardiff Catherine Moore, Johnathan Evans, Malorie Perry, Simon Cottrell, Alec Birchley, Alexander Adams, Amy Gaskin, Bree Gatica-Wilcox, Jason Coombes, Lauren Gilbert, Lee Graham, Nicole Pacchiarini, Sara Kumziene-Summerhayes, Sarah Taylor, Sophie Jones, Sara Rey, Matthew Bull, Joanne Watkins, Sally Corden, Tom Connor
EPI_ISL_422050 hCoV-19/Wales/PHWC-26DEF/2020 Europe / United Kingdom / Wales 2020-03-30 Wales Specialist Virology Centre Public Health Wales Microbiology Cardiff Catherine Moore, Johnathan Evans, Malorie Perry, Simon Cottrell, Alec Birchley, Alexander Adams, Amy Gaskin, Bree Gatica-Wilcox, Jason Coombes, Lauren Gilbert, Lee Graham, Nicole Pacchiarini, Sara Kumziene-Summerhayes, Sarah Taylor, Sophie Jones, Sara Rey, Matthew Bull, Joanne Watkins, Sally Corden, Tom Connor
EPI_ISL_422051 hCoV-19/Wales/PHWC-26A51/2020 Europe / United Kingdom / Wales 2020-03-30 Wales Specialist Virology Centre Public Health Wales Microbiology Cardiff Catherine Moore, Johnathan Evans, Malorie Perry, Simon Cottrell, Alec Birchley, Alexander Adams, Amy Gaskin, Bree Gatica-Wilcox, Jason Coombes, Lauren Gilbert, Lee Graham, Nicole Pacchiarini, Sara Kumziene-Summerhayes, Sarah Taylor, Sophie Jones, Sara Rey, Matthew Bull, Joanne Watkins, Sally Corden, Tom Connor
EPI_ISL_422052 hCoV-19/Wales/PHWC-26884/2020 Europe / United Kingdom / Wales 2020-03-27 Wales Specialist Virology Centre Public Health Wales Microbiology Cardiff Catherine Moore, Johnathan Evans, Malorie Perry, Simon Cottrell, Alec Birchley, Alexander Adams, Amy Gaskin, Bree Gatica-Wilcox, Jason Coombes, Lauren Gilbert, Lee Graham, Nicole Pacchiarini, Sara Kumziene-Summerhayes, Sarah Taylor, Sophie Jones, Sara Rey, Matthew Bull, Joanne Watkins, Sally Corden, Tom Connor
EPI_ISL_422053 hCoV-19/Wales/PHWC-25ABC/2020 Europe / United Kingdom / Wales 2020-03-24 Wales Specialist Virology Centre Public Health Wales Microbiology Cardiff Catherine Moore, Johnathan Evans, Malorie Perry, Simon Cottrell, Alec Birchley, Alexander Adams, Amy Gaskin, Bree Gatica-Wilcox, Jason Coombes, Lauren Gilbert, Lee Graham, Nicole Pacchiarini, Sara Kumziene-Summerhayes, Sarah Taylor, Sophie Jones, Sara Rey, Matthew Bull, Joanne Watkins, Sally Corden, Tom Connor
EPI_ISL_422054 hCoV-19/Wales/PHWC-26F9E/2020 Europe / United Kingdom / Wales 2020-03-29 Wales Specialist Virology Centre Public Health Wales Microbiology Cardiff Catherine Moore, Johnathan Evans, Malorie Perry, Simon Cottrell, Alec Birchley, Alexander Adams, Amy Gaskin, Bree Gatica-Wilcox, Jason Coombes, Lauren Gilbert, Lee Graham, Nicole Pacchiarini, Sara Kumziene-Summerhayes, Sarah Taylor, Sophie Jones, Sara Rey, Matthew Bull, Joanne Watkins, Sally Corden, Tom Connor
EPI_ISL_422055 hCoV-19/Wales/PHWC-25E47/2020 Europe / United Kingdom / Wales 2020-03-25 Wales Specialist Virology Centre Public Health Wales Microbiology Cardiff Catherine Moore, Johnathan Evans, Malorie Perry, Simon Cottrell, Alec Birchley, Alexander Adams, Amy Gaskin, Bree Gatica-Wilcox, Jason Coombes, Lauren Gilbert, Lee Graham, Nicole Pacchiarini, Sara Kumziene-Summerhayes, Sarah Taylor, Sophie Jones, Sara Rey, Matthew Bull, Joanne Watkins, Sally Corden, Tom Connor
EPI_ISL_422056 hCoV-19/Wales/PHWC-26CC4/2020 Europe / United Kingdom / Wales 2020-03-28 Wales Specialist Virology Centre Public Health Wales Microbiology Cardiff Catherine Moore, Johnathan Evans, Malorie Perry, Simon Cottrell, Alec Birchley, Alexander Adams, Amy Gaskin, Bree Gatica-Wilcox, Jason Coombes, Lauren Gilbert, Lee Graham, Nicole Pacchiarini, Sara Kumziene-Summerhayes, Sarah Taylor, Sophie Jones, Sara Rey, Matthew Bull, Joanne Watkins, Sally Corden, Tom Connor
EPI_ISL_422057 hCoV-19/Wales/PHWC-268A2/2020 Europe / United Kingdom / Wales 2020-03-28 Wales Specialist Virology Centre Public Health Wales Microbiology Cardiff Catherine Moore, Johnathan Evans, Malorie Perry, Simon Cottrell, Alec Birchley, Alexander Adams, Amy Gaskin, Bree Gatica-Wilcox, Jason Coombes, Lauren Gilbert, Lee Graham, Nicole Pacchiarini, Sara Kumziene-Summerhayes, Sarah Taylor, Sophie Jones, Sara Rey, Matthew Bull, Joanne Watkins, Sally Corden, Tom Connor
EPI_ISL_422058 hCoV-19/Wales/PHWC-26E91/2020 Europe / United Kingdom / Wales 2020-03-30 Wales Specialist Virology Centre Public Health Wales Microbiology Cardiff Catherine Moore, Johnathan Evans, Malorie Perry, Simon Cottrell, Alec Birchley, Alexander Adams, Amy Gaskin, Bree Gatica-Wilcox, Jason Coombes, Lauren Gilbert, Lee Graham, Nicole Pacchiarini, Sara Kumziene-Summerhayes, Sarah Taylor, Sophie Jones, Sara Rey, Matthew Bull, Joanne Watkins, Sally Corden, Tom Connor
EPI_ISL_422059 hCoV-19/Wales/PHWC-26FE9/2020 Europe / United Kingdom / Wales 2020-03-30 Wales Specialist Virology Centre Public Health Wales Microbiology Cardiff Catherine Moore, Johnathan Evans, Malorie Perry, Simon Cottrell, Alec Birchley, Alexander Adams, Amy Gaskin, Bree Gatica-Wilcox, Jason Coombes, Lauren Gilbert, Lee Graham, Nicole Pacchiarini, Sara Kumziene-Summerhayes, Sarah Taylor, Sophie Jones, Sara Rey, Matthew Bull, Joanne Watkins, Sally Corden, Tom Connor

EPI_ISL_422060 hCoV-19/Wales/PHWC-25FEA/2020 Europe / United Kingdom / Wales 2020-03-25 Wales Specialist Virology Centre Public Health Wales Microbiology Cardiff Catherine Moore, Johnathan Evans, Malorie Perry, Simon Cottrell, Alec Birchley, Alexander Adams, Amy Gaskin, Bree Gatica-Wilcox, Jason Coombes, Lauren Gilbert, Lee Graham, Nicole Pacchiarini, Sara Kumziene-Summerhayes, Sarah Taylor, Sophie Jones, Sara Rey, Matthew Bull, Joanne Watkins, Sally Corden, Tom Connor
EPI_ISL_422061 hCoV-19/Wales/PHWC-26E82/2020 Europe / United Kingdom / Wales 2020-03-30 Wales Specialist Virology Centre Public Health Wales Microbiology Cardiff Catherine Moore, Johnathan Evans, Malorie Perry, Simon Cottrell, Alec Birchley, Alexander Adams, Amy Gaskin, Bree Gatica-Wilcox, Jason Coombes, Lauren Gilbert, Lee Graham, Nicole Pacchiarini, Sara Kumziene-Summerhayes, Sarah Taylor, Sophie Jones, Sara Rey, Matthew Bull, Joanne Watkins, Sally Corden, Tom Connor
EPI_ISL_422062 hCoV-19/Wales/PHWC-269AF/2020 Europe / United Kingdom / Wales 2020-03-30 Wales Specialist Virology Centre Public Health Wales Microbiology Cardiff Catherine Moore, Johnathan Evans, Malorie Perry, Simon Cottrell, Alec Birchley, Alexander Adams, Amy Gaskin, Bree Gatica-Wilcox, Jason Coombes, Lauren Gilbert, Lee Graham, Nicole Pacchiarini, Sara Kumziene-Summerhayes, Sarah Taylor, Sophie Jones, Sara Rey, Matthew Bull, Joanne Watkins, Sally Corden, Tom Connor
EPI_ISL_422063 hCoV-19/Wales/PHWC-25B6E/2020 Europe / United Kingdom / Wales 2020-03-25 Wales Specialist Virology Centre Public Health Wales Microbiology Cardiff Catherine Moore, Johnathan Evans, Malorie Perry, Simon Cottrell, Alec Birchley, Alexander Adams, Amy Gaskin, Bree Gatica-Wilcox, Jason Coombes, Lauren Gilbert, Lee Graham, Nicole Pacchiarini, Sara Kumziene-Summerhayes, Sarah Taylor, Sophie Jones, Sara Rey, Matthew Bull, Joanne Watkins, Sally Corden, Tom Connor
EPI_ISL_422064 hCoV-19/Wales/PHWC-25C01/2020 Europe / United Kingdom / Wales 2020-03-25 Wales Specialist Virology Centre Public Health Wales Microbiology Cardiff Catherine Moore, Johnathan Evans, Malorie Perry, Simon Cottrell, Alec Birchley, Alexander Adams, Amy Gaskin, Bree Gatica-Wilcox, Jason Coombes, Lauren Gilbert, Lee Graham, Nicole Pacchiarini, Sara Kumziene-Summerhayes, Sarah Taylor, Sophie Jones, Sara Rey, Matthew Bull, Joanne Watkins, Sally Corden, Tom Connor
EPI_ISL_422065 hCoV-19/Wales/PHWC-2566D/2020 Europe / United Kingdom / Wales 2020-03-24 Wales Specialist Virology Centre Public Health Wales Microbiology Cardiff Catherine Moore, Johnathan Evans, Malorie Perry, Simon Cottrell, Alec Birchley, Alexander Adams, Amy Gaskin, Bree Gatica-Wilcox, Jason Coombes, Lauren Gilbert, Lee Graham, Nicole Pacchiarini, Sara Kumziene-Summerhayes, Sarah Taylor, Sophie Jones, Sara Rey, Matthew Bull, Joanne Watkins, Sally Corden, Tom Connor
EPI_ISL_422066 hCoV-19/Wales/PHWC-2573D/2020 Europe / United Kingdom / Wales 2020-03-24 Wales Specialist Virology Centre Public Health Wales Microbiology Cardiff Catherine Moore, Johnathan Evans, Malorie Perry, Simon Cottrell, Alec Birchley, Alexander Adams, Amy Gaskin, Bree Gatica-Wilcox, Jason Coombes, Lauren Gilbert, Lee Graham, Nicole Pacchiarini, Sara Kumziene-Summerhayes, Sarah Taylor, Sophie Jones, Sara Rey, Matthew Bull, Joanne Watkins, Sally Corden, Tom Connor
EPI_ISL_422067 hCoV-19/Wales/PHWC-25973/2020 Europe / United Kingdom / Wales 2020-03-24 Wales Specialist Virology Centre Public Health Wales Microbiology Cardiff Catherine Moore, Johnathan Evans, Malorie Perry, Simon Cottrell, Alec Birchley, Alexander Adams, Amy Gaskin, Bree Gatica-Wilcox, Jason Coombes, Lauren Gilbert, Lee Graham, Nicole Pacchiarini, Sara Kumziene-Summerhayes, Sarah Taylor, Sophie Jones, Sara Rey, Matthew Bull, Joanne Watkins, Sally Corden, Tom Connor
EPI_ISL_422068 hCoV-19/Wales/PHWC-26EFB/2020 Europe / United Kingdom / Wales 2020-03-30 Wales Specialist Virology Centre Public Health Wales Microbiology Cardiff Catherine Moore, Johnathan Evans, Malorie Perry, Simon Cottrell, Alec Birchley, Alexander Adams, Amy Gaskin, Bree Gatica-Wilcox, Jason Coombes, Lauren Gilbert, Lee Graham, Nicole Pacchiarini, Sara Kumziene-Summerhayes, Sarah Taylor, Sophie Jones, Sara Rey, Matthew Bull, Joanne Watkins, Sally Corden, Tom Connor
EPI_ISL_422069 hCoV-19/Wales/PHWC-270A9/2020 Europe / United Kingdom / Wales 2020-03-29 Wales Specialist Virology Centre Public Health Wales Microbiology Cardiff Catherine Moore, Johnathan Evans, Malorie Perry, Simon Cottrell, Alec Birchley, Alexander Adams, Amy Gaskin, Bree Gatica-Wilcox, Jason Coombes, Lauren Gilbert, Lee Graham, Nicole Pacchiarini, Sara Kumziene-Summerhayes, Sarah Taylor, Sophie Jones, Sara Rey, Matthew Bull, Joanne Watkins, Sally Corden, Tom Connor
EPI_ISL_422070 hCoV-19/Wales/PHWC-26839/2020 Europe / United Kingdom / Wales 2020-03-27 Wales Specialist Virology Centre Public Health Wales Microbiology Cardiff Catherine Moore, Johnathan Evans, Malorie Perry, Simon Cottrell, Alec Birchley, Alexander Adams, Amy Gaskin, Bree Gatica-Wilcox, Jason Coombes, Lauren Gilbert, Lee Graham, Nicole Pacchiarini, Sara Kumziene-Summerhayes, Sarah Taylor, Sophie Jones, Sara Rey, Matthew Bull, Joanne Watkins, Sally Corden, Tom Connor
EPI_ISL_422071 hCoV-19/Wales/PHWC-257B5/2020 Europe / United Kingdom / Wales 2020-03-24 Wales Specialist Virology Centre Public Health Wales Microbiology Cardiff Catherine Moore, Johnathan Evans, Malorie Perry, Simon Cottrell, Alec Birchley, Alexander Adams, Amy Gaskin, Bree Gatica-Wilcox, Jason Coombes, Lauren Gilbert, Lee Graham, Nicole Pacchiarini, Sara Kumziene-Summerhayes, Sarah Taylor, Sophie Jones, Sara Rey, Matthew Bull, Joanne Watkins, Sally Corden, Tom Connor
EPI_ISL_422072 hCoV-19/Wales/PHWC-26F52/2020 Europe / United Kingdom / Wales 2020-03-28 Wales Specialist Virology Centre Public Health Wales Microbiology Cardiff Catherine Moore, Johnathan Evans, Malorie Perry, Simon Cottrell, Alec Birchley, Alexander Adams, Amy Gaskin, Bree Gatica-Wilcox, Jason Coombes, Lauren Gilbert, Lee Graham, Nicole Pacchiarini, Sara Kumziene-Summerhayes, Sarah Taylor, Sophie Jones, Sara Rey, Matthew Bull, Joanne Watkins, Sally Corden, Tom Connor
EPI_ISL_422073 hCoV-19/Wales/PHWC-260AA/2020 Europe / United Kingdom / Wales 2020-03-24 Wales Specialist Virology Centre Public Health Wales Microbiology Cardiff Catherine Moore, Johnathan Evans, Malorie Perry, Simon Cottrell, Alec Birchley, Alexander Adams, Amy Gaskin, Bree Gatica-Wilcox, Jason Coombes, Lauren Gilbert, Lee Graham, Nicole Pacchiarini, Sara Kumziene-Summerhayes, Sarah Taylor, Sophie Jones, Sara Rey, Matthew Bull, Joanne Watkins, Sally Corden, Tom Connor

EPI_ISL_422074 hCoV-19/Wales/PHWC-26954/2020 Europe / United Kingdom / Wales 2020-03-27 Wales Specialist Virology Centre Public Health Wales Microbiology Cardiff Catherine Moore, Johnathan Evans, Malorie Perry, Simon Cottrell, Alec Birchley, Alexander Adams, Amy Gaskin, Bree Gatica-Wilcox, Jason Coombes, Lauren Gilbert, Lee Graham, Nicole Pacchiarini, Sara Kumziene-Summerhayes, Sarah Taylor, Sophie Jones, Sara Rey, Matthew Bull, Joanne Watkins, Sally Corden, Tom Connor
EPI_ISL_422075 hCoV-19/Wales/PHWC-26E46/2020 Europe / United Kingdom / Wales 2020-03-29 Wales Specialist Virology Centre Public Health Wales Microbiology Cardiff Catherine Moore, Johnathan Evans, Malorie Perry, Simon Cottrell, Alec Birchley, Alexander Adams, Amy Gaskin, Bree Gatica-Wilcox, Jason Coombes, Lauren Gilbert, Lee Graham, Nicole Pacchiarini, Sara Kumziene-Summerhayes, Sarah Taylor, Sophie Jones, Sara Rey, Matthew Bull, Joanne Watkins, Sally Corden, Tom Connor
EPI_ISL_422076 hCoV-19/Wales/PHWC-26D3A/2020 Europe / United Kingdom / Wales 2020-03-30 Wales Specialist Virology Centre Public Health Wales Microbiology Cardiff Catherine Moore, Johnathan Evans, Malorie Perry, Simon Cottrell, Alec Birchley, Alexander Adams, Amy Gaskin, Bree Gatica-Wilcox, Jason Coombes, Lauren Gilbert, Lee Graham, Nicole Pacchiarini, Sara Kumziene-Summerhayes, Sarah Taylor, Sophie Jones, Sara Rey, Matthew Bull, Joanne Watkins, Sally Corden, Tom Connor
EPI_ISL_422077 hCoV-19/Wales/PHWC-26A42/2020 Europe / United Kingdom / Wales 2020-03-29 Wales Specialist Virology Centre Public Health Wales Microbiology Cardiff Catherine Moore, Johnathan Evans, Malorie Perry, Simon Cottrell, Alec Birchley, Alexander Adams, Amy Gaskin, Bree Gatica-Wilcox, Jason Coombes, Lauren Gilbert, Lee Graham, Nicole Pacchiarini, Sara Kumziene-Summerhayes, Sarah Taylor, Sophie Jones, Sara Rey, Matthew Bull, Joanne Watkins, Sally Corden, Tom Connor
EPI_ISL_422078 hCoV-19/Wales/PHWC-26013/2020 Europe / United Kingdom / Wales 2020-03-25 Wales Specialist Virology Centre Public Health Wales Microbiology Cardiff Catherine Moore, Johnathan Evans, Malorie Perry, Simon Cottrell, Alec Birchley, Alexander Adams, Amy Gaskin, Bree Gatica-Wilcox, Jason Coombes, Lauren Gilbert, Lee Graham, Nicole Pacchiarini, Sara Kumziene-Summerhayes, Sarah Taylor, Sophie Jones, Sara Rey, Matthew Bull, Joanne Watkins, Sally Corden, Tom Connor
EPI_ISL_422079 hCoV-19/Wales/PHWC-26E37/2020 Europe / United Kingdom / Wales 2020-03-29 Wales Specialist Virology Centre Public Health Wales Microbiology Cardiff Catherine Moore, Johnathan Evans, Malorie Perry, Simon Cottrell, Alec Birchley, Alexander Adams, Amy Gaskin, Bree Gatica-Wilcox, Jason Coombes, Lauren Gilbert, Lee Graham, Nicole Pacchiarini, Sara Kumziene-Summerhayes, Sarah Taylor, Sophie Jones, Sara Rey, Matthew Bull, Joanne Watkins, Sally Corden, Tom Connor
EPI_ISL_422080 hCoV-19/Wales/PHWC-26E55/2020 Europe / United Kingdom / Wales 2020-03-29 Wales Specialist Virology Centre Public Health Wales Microbiology Cardiff Catherine Moore, Johnathan Evans, Malorie Perry, Simon Cottrell, Alec Birchley, Alexander Adams, Amy Gaskin, Bree Gatica-Wilcox, Jason Coombes, Lauren Gilbert, Lee Graham, Nicole Pacchiarini, Sara Kumziene-Summerhayes, Sarah Taylor, Sophie Jones, Sara Rey, Matthew Bull, Joanne Watkins, Sally Corden, Tom Connor
EPI_ISL_422081 hCoV-19/Wales/PHWC-26EBF/2020 Europe / United Kingdom / Wales 2020-03-30 Wales Specialist Virology Centre Public Health Wales Microbiology Cardiff Catherine Moore, Johnathan Evans, Malorie Perry, Simon Cottrell, Alec Birchley, Alexander Adams, Amy Gaskin, Bree Gatica-Wilcox, Jason Coombes, Lauren Gilbert, Lee Graham, Nicole Pacchiarini, Sara Kumziene-Summerhayes, Sarah Taylor, Sophie Jones, Sara Rey, Matthew Bull, Joanne Watkins, Sally Corden, Tom Connor
EPI_ISL_422082 hCoV-19/Wales/PHWC-26BD6/2020 Europe / United Kingdom / Wales 2020-03-28 Wales Specialist Virology Centre Public Health Wales Microbiology Cardiff Catherine Moore, Johnathan Evans, Malorie Perry, Simon Cottrell, Alec Birchley, Alexander Adams, Amy Gaskin, Bree Gatica-Wilcox, Jason Coombes, Lauren Gilbert, Lee Graham, Nicole Pacchiarini, Sara Kumziene-Summerhayes, Sarah Taylor, Sophie Jones, Sara Rey, Matthew Bull, Joanne Watkins, Sally Corden, Tom Connor
EPI_ISL_422083 hCoV-19/Wales/PHWC-2624A/2020 Europe / United Kingdom / Wales 2020-03-27 Wales Specialist Virology Centre Public Health Wales Microbiology Cardiff Catherine Moore, Johnathan Evans, Malorie Perry, Simon Cottrell, Alec Birchley, Alexander Adams, Amy Gaskin, Bree Gatica-Wilcox, Jason Coombes, Lauren Gilbert, Lee Graham, Nicole Pacchiarini, Sara Kumziene-Summerhayes, Sarah Taylor, Sophie Jones, Sara Rey, Matthew Bull, Joanne Watkins, Sally Corden, Tom Connor
EPI_ISL_422084 hCoV-19/Wales/PHWC-26DB2/2020 Europe / United Kingdom / Wales 2020-03-30 Wales Specialist Virology Centre Public Health Wales Microbiology Cardiff Catherine Moore, Johnathan Evans, Malorie Perry, Simon Cottrell, Alec Birchley, Alexander Adams, Amy Gaskin, Bree Gatica-Wilcox, Jason Coombes, Lauren Gilbert, Lee Graham, Nicole Pacchiarini, Sara Kumziene-Summerhayes, Sarah Taylor, Sophie Jones, Sara Rey, Matthew Bull, Joanne Watkins, Sally Corden, Tom Connor
EPI_ISL_422085 hCoV-19/Wales/PHWC-25FAE/2020 Europe / United Kingdom / Wales 2020-03-25 Wales Specialist Virology Centre Public Health Wales Microbiology Cardiff Catherine Moore, Johnathan Evans, Malorie Perry, Simon Cottrell, Alec Birchley, Alexander Adams, Amy Gaskin, Bree Gatica-Wilcox, Jason Coombes, Lauren Gilbert, Lee Graham, Nicole Pacchiarini, Sara Kumziene-Summerhayes, Sarah Taylor, Sophie Jones, Sara Rey, Matthew Bull, Joanne Watkins, Sally Corden, Tom Connor
EPI_ISL_422086 hCoV-19/Wales/PHWC-2708B/2020 Europe / United Kingdom / Wales 2020-03-29 Wales Specialist Virology Centre Public Health Wales Microbiology Cardiff Catherine Moore, Johnathan Evans, Malorie Perry, Simon Cottrell, Alec Birchley, Alexander Adams, Amy Gaskin, Bree Gatica-Wilcox, Jason Coombes, Lauren Gilbert, Lee Graham, Nicole Pacchiarini, Sara Kumziene-Summerhayes, Sarah Taylor, Sophie Jones, Sara Rey, Matthew Bull, Joanne Watkins, Sally Corden, Tom Connor
EPI_ISL_422087 hCoV-19/Wales/PHWC-26CA6/2020 Europe / United Kingdom / Wales 2020-03-28 Wales Specialist Virology Centre Public Health Wales Microbiology Cardiff Catherine Moore, Johnathan Evans, Malorie Perry, Simon Cottrell, Alec Birchley, Alexander Adams, Amy Gaskin, Bree Gatica-Wilcox, Jason Coombes, Lauren Gilbert, Lee Graham, Nicole Pacchiarini, Sara Kumziene-Summerhayes, Sarah Taylor, Sophie Jones, Sara Rey, Matthew Bull, Joanne Watkins, Sally Corden, Tom Connor

EPI_ISL_422088 hCoV-19/Wales/PHWC-26DC1/2020 Europe / United Kingdom / Wales 2020-03-30 Wales Specialist Virology Centre Public Health Wales Microbiology Cardiff Catherine Moore, Johnathan Evans, Malorie Perry, Simon Cottrell, Alec Birchley, Alexander Adams, Amy Gaskin, Bree Gatica-Wilcox, Jason Coombes, Lauren Gilbert, Lee Graham, Nicole Pacchiarini, Sara Kumziene-Summerhayes, Sarah Taylor, Sophie Jones, Sara Rey, Matthew Bull, Joanne Watkins, Sally Corden, Tom Connor
EPI_ISL_422089 hCoV-19/Wales/PHWC-26DA3/2020 Europe / United Kingdom / Wales 2020-03-30 Wales Specialist Virology Centre Public Health Wales Microbiology Cardiff Catherine Moore, Johnathan Evans, Malorie Perry, Simon Cottrell, Alec Birchley, Alexander Adams, Amy Gaskin, Bree Gatica-Wilcox, Jason Coombes, Lauren Gilbert, Lee Graham, Nicole Pacchiarini, Sara Kumziene-Summerhayes, Sarah Taylor, Sophie Jones, Sara Rey, Matthew Bull, Joanne Watkins, Sally Corden, Tom Connor
EPI_ISL_422090 hCoV-19/Wales/PHWC-26ECE/2020 Europe / United Kingdom / Wales 2020-03-30 Wales Specialist Virology Centre Public Health Wales Microbiology Cardiff Catherine Moore, Johnathan Evans, Malorie Perry, Simon Cottrell, Alec Birchley, Alexander Adams, Amy Gaskin, Bree Gatica-Wilcox, Jason Coombes, Lauren Gilbert, Lee Graham, Nicole Pacchiarini, Sara Kumziene-Summerhayes, Sarah Taylor, Sophie Jones, Sara Rey, Matthew Bull, Joanne Watkins, Sally Corden, Tom Connor
EPI_ISL_422091 hCoV-19/Wales/PHWC-264DB/2020 Europe / United Kingdom / Wales 2020-03-28 Wales Specialist Virology Centre Public Health Wales Microbiology Cardiff Catherine Moore, Johnathan Evans, Malorie Perry, Simon Cottrell, Alec Birchley, Alexander Adams, Amy Gaskin, Bree Gatica-Wilcox, Jason Coombes, Lauren Gilbert, Lee Graham, Nicole Pacchiarini, Sara Kumziene-Summerhayes, Sarah Taylor, Sophie Jones, Sara Rey, Matthew Bull, Joanne Watkins, Sally Corden, Tom Connor
EPI_ISL_422092 hCoV-19/Wales/PHWC-26857/2020 Europe / United Kingdom / Wales 2020-03-27 Wales Specialist Virology Centre Public Health Wales Microbiology Cardiff Catherine Moore, Johnathan Evans, Malorie Perry, Simon Cottrell, Alec Birchley, Alexander Adams, Amy Gaskin, Bree Gatica-Wilcox, Jason Coombes, Lauren Gilbert, Lee Graham, Nicole Pacchiarini, Sara Kumziene-Summerhayes, Sarah Taylor, Sophie Jones, Sara Rey, Matthew Bull, Joanne Watkins, Sally Corden, Tom Connor
EPI_ISL_422093 hCoV-19/Wales/PHWC-2707C/2020 Europe / United Kingdom / Wales 2020-03-29 Wales Specialist Virology Centre Public Health Wales Microbiology Cardiff Catherine Moore, Johnathan Evans, Malorie Perry, Simon Cottrell, Alec Birchley, Alexander Adams, Amy Gaskin, Bree Gatica-Wilcox, Jason Coombes, Lauren Gilbert, Lee Graham, Nicole Pacchiarini, Sara Kumziene-Summerhayes, Sarah Taylor, Sophie Jones, Sara Rey, Matthew Bull, Joanne Watkins, Sally Corden, Tom Connor
EPI_ISL_422094 hCoV-19/Wales/PHWC-2617A/2020 Europe / United Kingdom / Wales 2020-03-25 Wales Specialist Virology Centre Public Health Wales Microbiology Cardiff Catherine Moore, Johnathan Evans, Malorie Perry, Simon Cottrell, Alec Birchley, Alexander Adams, Amy Gaskin, Bree Gatica-Wilcox, Jason Coombes, Lauren Gilbert, Lee Graham, Nicole Pacchiarini, Sara Kumziene-Summerhayes, Sarah Taylor, Sophie Jones, Sara Rey, Matthew Bull, Joanne Watkins, Sally Corden, Tom Connor
EPI_ISL_422095 hCoV-19/Wales/PHWC-2705E/2020 Europe / United Kingdom / Wales 2020-03-28 Wales Specialist Virology Centre Public Health Wales Microbiology Cardiff Catherine Moore, Johnathan Evans, Malorie Perry, Simon Cottrell, Alec Birchley, Alexander Adams, Amy Gaskin, Bree Gatica-Wilcox, Jason Coombes, Lauren Gilbert, Lee Graham, Nicole Pacchiarini, Sara Kumziene-Summerhayes, Sarah Taylor, Sophie Jones, Sara Rey, Matthew Bull, Joanne Watkins, Sally Corden, Tom Connor
EPI_ISL_422096 hCoV-19/Wales/PHWC-2675A/2020 Europe / United Kingdom / Wales 2020-03-28 Wales Specialist Virology Centre Public Health Wales Microbiology Cardiff Catherine Moore, Johnathan Evans, Malorie Perry, Simon Cottrell, Alec Birchley, Alexander Adams, Amy Gaskin, Bree Gatica-Wilcox, Jason Coombes, Lauren Gilbert, Lee Graham, Nicole Pacchiarini, Sara Kumziene-Summerhayes, Sarah Taylor, Sophie Jones, Sara Rey, Matthew Bull, Joanne Watkins, Sally Corden, Tom Connor
EPI_ISL_422097 hCoV-19/Wales/PHWC-26D0D/2020 Europe / United Kingdom / Wales 2020-03-30 Wales Specialist Virology Centre Public Health Wales Microbiology Cardiff Catherine Moore, Johnathan Evans, Malorie Perry, Simon Cottrell, Alec Birchley, Alexander Adams, Amy Gaskin, Bree Gatica-Wilcox, Jason Coombes, Lauren Gilbert, Lee Graham, Nicole Pacchiarini, Sara Kumziene-Summerhayes, Sarah Taylor, Sophie Jones, Sara Rey, Matthew Bull, Joanne Watkins, Sally Corden, Tom Connor
EPI_ISL_422098 hCoV-19/Wales/PHWC-26848/2020 Europe / United Kingdom / Wales 2020-03-27 Wales Specialist Virology Centre Public Health Wales Microbiology Cardiff Catherine Moore, Johnathan Evans, Malorie Perry, Simon Cottrell, Alec Birchley, Alexander Adams, Amy Gaskin, Bree Gatica-Wilcox, Jason Coombes, Lauren Gilbert, Lee Graham, Nicole Pacchiarini, Sara Kumziene-Summerhayes, Sarah Taylor, Sophie Jones, Sara Rey, Matthew Bull, Joanne Watkins, Sally Corden, Tom Connor
EPI_ISL_422099 hCoV-19/Wales/PHWC-27021/2020 Europe / United Kingdom / Wales 2020-03-28 Wales Specialist Virology Centre Public Health Wales Microbiology Cardiff Catherine Moore, Johnathan Evans, Malorie Perry, Simon Cottrell, Alec Birchley, Alexander Adams, Amy Gaskin, Bree Gatica-Wilcox, Jason Coombes, Lauren Gilbert, Lee Graham, Nicole Pacchiarini, Sara Kumziene-Summerhayes, Sarah Taylor, Sophie Jones, Sara Rey, Matthew Bull, Joanne Watkins, Sally Corden, Tom Connor
EPI_ISL_423371 hCoV-19/England/20140002204/2020 Europe / United Kingdom / England 2020-03-29 Respiratory Virus Unit, Microbiology Services Colindale, Public Health England Respiratory Virus Unit, Microbiology Services Colindale, Public Health England Monica Galiano, Shahjahan Miah, Angie Lackenby, Omolola Akinbami, Tiina Talts, Leena Bhaw, Richard Myers, Steven Platt, Kirstin Edwards, Jonathan Hubb, Joanna Ellis, Maria Zambon
EPI_ISL_423372 hCoV-19/England/20140002604/2020 Europe / United Kingdom / England 2020-03-28 Respiratory Virus Unit, Microbiology Services Colindale, Public Health England Respiratory Virus Unit, Microbiology Services Colindale, Public Health England Monica Galiano, Shahjahan Miah, Angie Lackenby, Omolola Akinbami, Tiina Talts, Leena Bhaw, Richard Myers, Steven Platt, Kirstin Edwards, Jonathan Hubb, Joanna Ellis, Maria Zambon

EPI_ISL_423373  hCoV-19/England/20140002804/2020        Europe / United Kingdom / England       2020-03-28
Respiratory Virus Unit, Microbiology Services Colindale, Public Health England  Respiratory Virus Unit, Microbiology Services Colindale, Public Health England  Monica Galiano, Shahjahan Miah, Angie Lackenby, Omolola Akinbami, Tiina Talts, Leena Bhaw, Richard Myers, Steven Platt, Kirstin Edwards, Jonathan Hubb, Joanna Ellis, Maria Zambon
EPI_ISL_423375  hCoV-19/England/20140003402/2020        Europe / United Kingdom / England       2020-03-28
Respiratory Virus Unit, Microbiology Services Colindale, Public Health England  Respiratory Virus Unit, Microbiology Services Colindale, Public Health England  Monica Galiano, Shahjahan Miah, Angie Lackenby, Omolola Akinbami, Tiina Talts, Leena Bhaw, Richard Myers, Steven Platt, Kirstin Edwards, Jonathan Hubb, Joanna Ellis, Maria Zambon
EPI_ISL_423376  hCoV-19/England/20140003404/2020        Europe / United Kingdom / England       2020-03-28
Respiratory Virus Unit, Microbiology Services Colindale, Public Health England  Respiratory Virus Unit, Microbiology Services Colindale, Public Health England  Monica Galiano, Shahjahan Miah, Angie Lackenby, Omolola Akinbami, Tiina Talts, Leena Bhaw, Richard Myers, Steven Platt, Kirstin Edwards, Jonathan Hubb, Joanna Ellis, Maria Zambon
EPI_ISL_423379  hCoV-19/England/20140004004/2020        Europe / United Kingdom / England       2020-03-28
Respiratory Virus Unit, Microbiology Services Colindale, Public Health England  Respiratory Virus Unit, Microbiology Services Colindale, Public Health England  Monica Galiano, Shahjahan Miah, Angie Lackenby, Omolola Akinbami, Tiina Talts, Leena Bhaw, Richard Myers, Steven Platt, Kirstin Edwards, Jonathan Hubb, Joanna Ellis, Maria Zambon
EPI_ISL_423380  hCoV-19/England/20140005304/2020        Europe / United Kingdom / England       2020-03-29
Respiratory Virus Unit, Microbiology Services Colindale, Public Health England  Respiratory Virus Unit, Microbiology Services Colindale, Public Health England  Monica Galiano, Shahjahan Miah, Angie Lackenby, Omolola Akinbami, Tiina Talts, Leena Bhaw, Richard Myers, Steven Platt, Kirstin Edwards, Jonathan Hubb, Joanna Ellis, Maria Zambon
EPI_ISL_423382  hCoV-19/England/20140007004/2020        Europe / United Kingdom / England       2020-03-29
Respiratory Virus Unit, Microbiology Services Colindale, Public Health England  Respiratory Virus Unit, Microbiology Services Colindale, Public Health England  Monica Galiano, Shahjahan Miah, Angie Lackenby, Omolola Akinbami, Tiina Talts, Leena Bhaw, Richard Myers, Steven Platt, Kirstin Edwards, Jonathan Hubb, Joanna Ellis, Maria Zambon
EPI_ISL_423384  hCoV-19/England/20140007604/2020        Europe / United Kingdom / England       2020-03-29
Respiratory Virus Unit, Microbiology Services Colindale, Public Health England  Respiratory Virus Unit, Microbiology Services Colindale, Public Health England  Monica Galiano, Shahjahan Miah, Angie Lackenby, Omolola Akinbami, Tiina Talts, Leena Bhaw, Richard Myers, Steven Platt, Kirstin Edwards, Jonathan Hubb, Joanna Ellis, Maria Zambon
EPI_ISL_423385  hCoV-19/England/20140009204/2020        Europe / United Kingdom / England       2020-03-28
Respiratory Virus Unit, Microbiology Services Colindale, Public Health England  Respiratory Virus Unit, Microbiology Services Colindale, Public Health England  Monica Galiano, Shahjahan Miah, Angie Lackenby, Omolola Akinbami, Tiina Talts, Leena Bhaw, Richard Myers, Steven Platt, Kirstin Edwards, Jonathan Hubb, Joanna Ellis, Maria Zambon
EPI_ISL_423387  hCoV-19/England/20140009404/2020        Europe / United Kingdom / England       2020-03-28
Respiratory Virus Unit, Microbiology Services Colindale, Public Health England  Respiratory Virus Unit, Microbiology Services Colindale, Public Health England  Monica Galiano, Shahjahan Miah, Angie Lackenby, Omolola Akinbami, Tiina Talts, Leena Bhaw, Richard Myers, Steven Platt, Kirstin Edwards, Jonathan Hubb, Joanna Ellis, Maria Zambon
EPI_ISL_423389  hCoV-19/England/20140060304/2020        Europe / United Kingdom / England       2020-03-30
Respiratory Virus Unit, Microbiology Services Colindale, Public Health England  Respiratory Virus Unit, Microbiology Services Colindale, Public Health England  Monica Galiano, Shahjahan Miah, Angie Lackenby, Omolola Akinbami, Tiina Talts, Leena Bhaw, Richard Myers, Steven Platt, Kirstin Edwards, Jonathan Hubb, Joanna Ellis, Maria Zambon
EPI_ISL_423393  hCoV-19/England/20140064004/2020        Europe / United Kingdom / England       2020-03-30
Respiratory Virus Unit, Microbiology Services Colindale, Public Health England  Respiratory Virus Unit, Microbiology Services Colindale, Public Health England  Monica Galiano, Shahjahan Miah, Angie Lackenby, Omolola Akinbami, Tiina Talts, Leena Bhaw, Richard Myers, Steven Platt, Kirstin Edwards, Jonathan Hubb, Joanna Ellis, Maria Zambon
EPI_ISL_423394  hCoV-19/England/20140064504/2020        Europe / United Kingdom / England       2020-03-30
Respiratory Virus Unit, Microbiology Services Colindale, Public Health England  Respiratory Virus Unit, Microbiology Services Colindale, Public Health England  Monica Galiano, Shahjahan Miah, Angie Lackenby, Omolola Akinbami, Tiina Talts, Leena Bhaw, Richard Myers, Steven Platt, Kirstin Edwards, Jonathan Hubb, Joanna Ellis, Maria Zambon
EPI_ISL_423395  hCoV-19/England/20140064704/2020        Europe / United Kingdom / England       2020-03-28
Respiratory Virus Unit, Microbiology Services Colindale, Public Health England  Respiratory Virus Unit, Microbiology Services Colindale, Public Health England  Monica Galiano, Shahjahan Miah, Angie Lackenby, Omolola Akinbami, Tiina Talts, Leena Bhaw, Richard Myers, Steven Platt, Kirstin Edwards, Jonathan Hubb, Joanna Ellis, Maria Zambon
EPI_ISL_423396  hCoV-19/England/20140064804/2020        Europe / United Kingdom / England       2020-03-28
Respiratory Virus Unit, Microbiology Services Colindale, Public Health England  Respiratory Virus Unit, Microbiology Services Colindale, Public Health England  Monica Galiano, Shahjahan Miah, Angie Lackenby, Omolola Akinbami, Tiina Talts, Leena Bhaw, Richard Myers, Steven Platt, Kirstin Edwards, Jonathan Hubb, Joanna Ellis, Maria Zambon

EPI_ISL_423398 hCoV-19/England/20140066004/2020 Europe / United Kingdom / England 2020-03-29 Respiratory Virus Unit, Microbiology Services Colindale, Public Health England Respiratory Virus Unit, Microbiology Services Colindale, Public Health England Monica Galiano, Shahjahan Miah, Angie Lackenby, Omolola Akinbami, Tiina Talts, Leena Bhaw, Richard Myers, Steven Platt, Kirstin Edwards, Jonathan Hubb, Joanna Ellis, Maria Zambon
EPI_ISL_423399 hCoV-19/England/20140066204/2020 Europe / United Kingdom / England 2020-03-29 Respiratory Virus Unit, Microbiology Services Colindale, Public Health England Respiratory Virus Unit, Microbiology Services Colindale, Public Health England Monica Galiano, Shahjahan Miah, Angie Lackenby, Omolola Akinbami, Tiina Talts, Leena Bhaw, Richard Myers, Steven Platt, Kirstin Edwards, Jonathan Hubb, Joanna Ellis, Maria Zambon
EPI_ISL_423400 hCoV-19/England/20140068404/2020 Europe / United Kingdom / England 2020-03-30 Respiratory Virus Unit, Microbiology Services Colindale, Public Health England Respiratory Virus Unit, Microbiology Services Colindale, Public Health England Monica Galiano, Shahjahan Miah, Angie Lackenby, Omolola Akinbami, Tiina Talts, Leena Bhaw, Richard Myers, Steven Platt, Kirstin Edwards, Jonathan Hubb, Joanna Ellis, Maria Zambon
EPI_ISL_423401 hCoV-19/England/20140068904/2020 Europe / United Kingdom / England 2020-03-29 Respiratory Virus Unit, Microbiology Services Colindale, Public Health England Respiratory Virus Unit, Microbiology Services Colindale, Public Health England Monica Galiano, Shahjahan Miah, Angie Lackenby, Omolola Akinbami, Tiina Talts, Leena Bhaw, Richard Myers, Steven Platt, Kirstin Edwards, Jonathan Hubb, Joanna Ellis, Maria Zambon
EPI_ISL_424101 hCoV-19/England/20129126204/2020 Europe / United Kingdom / England 2020-03-12 Respiratory Virus Unit, Microbiology Services Colindale, Public Health England Respiratory Virus Unit, Microbiology Services Colindale, Public Health England Monica Galiano, Shahjahan Miah, Angie Lackenby, Omolola Akinbami, Tiina Talts, Leena Bhaw, Richard Myers, Steven Platt, Kirstin Edwards, Jonathan Hubb, Joanna Ellis, Maria Zambon
EPI_ISL_424117 hCoV-19/England/20129148904/2020 Europe / United Kingdom / England 2020-03-12 Respiratory Virus Unit, Microbiology Services Colindale, Public Health England Respiratory Virus Unit, Microbiology Services Colindale, Public Health England Monica Galiano, Shahjahan Miah, Angie Lackenby, Omolola Akinbami, Tiina Talts, Leena Bhaw, Richard Myers, Steven Platt, Kirstin Edwards, Jonathan Hubb, Joanna Ellis, Maria Zambon
EPI_ISL_424119 hCoV-19/England/20129158804/2020 Europe / United Kingdom / England 2020-03-21 Respiratory Virus Unit, Microbiology Services Colindale, Public Health England Respiratory Virus Unit, Microbiology Services Colindale, Public Health England Monica Galiano, Shahjahan Miah, Angie Lackenby, Omolola Akinbami, Tiina Talts, Leena Bhaw, Richard Myers, Steven Platt, Kirstin Edwards, Jonathan Hubb, Joanna Ellis, Maria Zambon
EPI_ISL_424121 hCoV-19/England/20129160704/2020 Europe / United Kingdom / England 2020-03-21 Respiratory Virus Unit, Microbiology Services Colindale, Public Health England Respiratory Virus Unit, Microbiology Services Colindale, Public Health England Monica Galiano, Shahjahan Miah, Angie Lackenby, Omolola Akinbami, Tiina Talts, Leena Bhaw, Richard Myers, Steven Platt, Kirstin Edwards, Jonathan Hubb, Joanna Ellis, Maria Zambon
EPI_ISL_424126 hCoV-19/England/20129163604/2020 Europe / United Kingdom / England 2020-03-21 Respiratory Virus Unit, Microbiology Services Colindale, Public Health England Respiratory Virus Unit, Microbiology Services Colindale, Public Health England Monica Galiano, Shahjahan Miah, Angie Lackenby, Omolola Akinbami, Tiina Talts, Leena Bhaw, Richard Myers, Steven Platt, Kirstin Edwards, Jonathan Hubb, Joanna Ellis, Maria Zambon
EPI_ISL_424140 hCoV-19/England/20129171604/2020 Europe / United Kingdom / England 2020-03-21 Respiratory Virus Unit, Microbiology Services Colindale, Public Health England Respiratory Virus Unit, Microbiology Services Colindale, Public Health England Monica Galiano, Shahjahan Miah, Angie Lackenby, Omolola Akinbami, Tiina Talts, Leena Bhaw, Richard Myers, Steven Platt, Kirstin Edwards, Jonathan Hubb, Joanna Ellis, Maria Zambon
EPI_ISL_424152 hCoV-19/England/20129174804/2020 Europe / United Kingdom / England 2020-03-20 Respiratory Virus Unit, Microbiology Services Colindale, Public Health England Respiratory Virus Unit, Microbiology Services Colindale, Public Health England Monica Galiano, Shahjahan Miah, Angie Lackenby, Omolola Akinbami, Tiina Talts, Leena Bhaw, Richard Myers, Steven Platt, Kirstin Edwards, Jonathan Hubb, Joanna Ellis, Maria Zambon
EPI_ISL_424166 hCoV-19/USA/WA-UW-1620/2020 North America / USA / Washington 2020-03-19 UW Virology Lab UW Virology Lab Pavitra Roychoudhury, Hong Xie, Keith Jerome, Alexander Greninger
EPI_ISL_424167 hCoV-19/USA/WA-UW-1616/2020 North America / USA / Washington 2020-03-19 UW Virology Lab UW Virology Lab Pavitra Roychoudhury, Hong Xie, Keith Jerome, Alexander Greninger
EPI_ISL_424169 hCoV-19/USA/WA-UW-1538/2020 North America / USA / Washington 2020-03-19 UW Virology Lab UW Virology Lab Pavitra Roychoudhury, Hong Xie, Keith Jerome, Alexander Greninger
EPI_ISL_424170 hCoV-19/USA/WA-UW-1595/2020 North America / USA / Washington 2020-03-19 UW Virology Lab UW Virology Lab Pavitra Roychoudhury, Hong Xie, Keith Jerome, Alexander Greninger
EPI_ISL_424172 hCoV-19/USA/WA-UW-1575/2020 North America / USA / Washington 2020-03-19 UW Virology Lab UW Virology Lab Pavitra Roychoudhury, Hong Xie, Keith Jerome, Alexander Greninger
EPI_ISL_424173 hCoV-19/USA/IL-UW-1311/2020 North America / USA / Illinois 2020-03-14 UW Virology Lab UW Virology Lab Pavitra Roychoudhury, Hong Xie, Keith Jerome, Alexander Greninger
EPI_ISL_424174 hCoV-19/USA/CT-UW-1365/2020 North America / USA / Connecticut 2020-03-13 UW Virology Lab UW Virology Lab Pavitra Roychoudhury, Hong Xie, Keith Jerome, Alexander Greninger
EPI_ISL_424175 hCoV-19/USA/WA-UW-1362/2020 North America / USA / Washington 2020-03-17 UW

Virology Lab	UW Virology Lab Pavitra Roychoudhury, Hong Xie, Keith Jerome, Alexander Greninger
EPI_ISL_424176	hCoV-19/USA/WA-UW-1472/2020	North America / USA / Washington	2020-03-17	UW Virology Lab	UW Virology Lab Pavitra Roychoudhury, Hong Xie, Keith Jerome, Alexander Greninger
EPI_ISL_424178	hCoV-19/USA/WA-UW-1342/2020	North America / USA / Washington	2020-03-17	UW Virology Lab	UW Virology Lab Pavitra Roychoudhury, Hong Xie, Keith Jerome, Alexander Greninger
EPI_ISL_424179	hCoV-19/USA/WA-UW-1341/2020	North America / USA / Washington	2020-03-17	UW Virology Lab	UW Virology Lab Pavitra Roychoudhury, Hong Xie, Keith Jerome, Alexander Greninger
EPI_ISL_424180	hCoV-19/USA/WA-UW-1294/2020	North America / USA / Washington	2020-03-18	UW Virology Lab	UW Virology Lab Pavitra Roychoudhury, Hong Xie, Keith Jerome, Alexander Greninger
EPI_ISL_424181	hCoV-19/USA/UN-UW-1402/2020	North America / USA	2020-03-16	UW Virology Lab UW Virology Lab	Pavitra Roychoudhury, Hong Xie, Keith Jerome, Alexander Greninger
EPI_ISL_424182	hCoV-19/USA/WA-UW-1478/2020	North America / USA / Washington	2020-03-18	UW Virology Lab	UW Virology Lab Pavitra Roychoudhury, Hong Xie, Keith Jerome, Alexander Greninger
EPI_ISL_424183	hCoV-19/USA/WA-UW-1436/2020	North America / USA / Washington	2020-03-18	UW Virology Lab	UW Virology Lab Pavitra Roychoudhury, Hong Xie, Keith Jerome, Alexander Greninger
EPI_ISL_424184	hCoV-19/USA/WA-UW-1422/2020	North America / USA / Washington	2020-03-18	UW Virology Lab	UW Virology Lab Pavitra Roychoudhury, Hong Xie, Keith Jerome, Alexander Greninger
EPI_ISL_424186	hCoV-19/USA/WA-UW-1644/2020	North America / USA / Washington	2020-03-20	UW Virology Lab	UW Virology Lab Pavitra Roychoudhury, Hong Xie, Keith Jerome, Alexander Greninger
EPI_ISL_424187	hCoV-19/USA/WA-UW-1646/2020	North America / USA / Washington	2020-03-20	UW Virology Lab	UW Virology Lab Pavitra Roychoudhury, Hong Xie, Keith Jerome, Alexander Greninger
EPI_ISL_424188	hCoV-19/USA/WA-UW-1645/2020	North America / USA / Washington	2020-03-20	UW Virology Lab	UW Virology Lab Pavitra Roychoudhury, Hong Xie, Keith Jerome, Alexander Greninger
EPI_ISL_424189	hCoV-19/USA/UN-UW-Not found/2020	North America / USA	2020-03 UW Virology Lab UW Virology Lab	Pavitra Roychoudhury, Hong Xie, Keith Jerome, Alexander Greninger
EPI_ISL_424191	hCoV-19/USA/WA-UW-1683/2020	North America / USA / Washington	2020-03-20	UW Virology Lab	UW Virology Lab Pavitra Roychoudhury, Hong Xie, Keith Jerome, Alexander Greninger
EPI_ISL_424192	hCoV-19/USA/WA-UW-1688/2020	North America / USA / Washington	2020-03-20	UW Virology Lab	UW Virology Lab Pavitra Roychoudhury, Hong Xie, Keith Jerome, Alexander Greninger
EPI_ISL_424193	hCoV-19/USA/WA-UW-1728/2020	North America / USA / Washington	2020-03-21	UW Virology Lab	UW Virology Lab Pavitra Roychoudhury, Hong Xie, Keith Jerome, Alexander Greninger
EPI_ISL_424194	hCoV-19/USA/WA-UW-1661/2020	North America / USA / Washington	2020-03-20	UW Virology Lab	UW Virology Lab Pavitra Roychoudhury, Hong Xie, Keith Jerome, Alexander Greninger
EPI_ISL_424195	hCoV-19/USA/WA-UW-1726/2020	North America / USA / Washington	2020-03-21	UW Virology Lab	UW Virology Lab Pavitra Roychoudhury, Hong Xie, Keith Jerome, Alexander Greninger
EPI_ISL_424196	hCoV-19/USA/WA-UW-1666/2020	North America / USA / Washington	2020-03-20	UW Virology Lab	UW Virology Lab Pavitra Roychoudhury, Hong Xie, Keith Jerome, Alexander Greninger
EPI_ISL_424197	hCoV-19/USA/WA-UW-1640/2020	North America / USA / Washington	2020-03-20	UW Virology Lab	UW Virology Lab Pavitra Roychoudhury, Hong Xie, Keith Jerome, Alexander Greninger
EPI_ISL_424198	hCoV-19/USA/WA-UW-1656/2020	North America / USA / Washington	2020-03-20	UW Virology Lab	UW Virology Lab Pavitra Roychoudhury, Hong Xie, Keith Jerome, Alexander Greninger
EPI_ISL_424703	hCoV-19/Sweden/RV-FOI-1/2020	Europe / Sweden / Vasterbotten	2020-03-15	Klinisk mikrobiologi, Region Västerbotten	Unit for Biological Agents, Department for CBRN Defence and Security, Swedish Defence Research Agency	FOI Bioinformatics team
EPI_ISL_425300	hCoV-19/England/CAMB-74508/2020 Europe / United Kingdom / England	2020-03-28	Department of Pathology, University of Cambridge	COVID-19 Genomics UK (COG-UK) Consortium	Luke W Meredith, M. Estee Torok , Myra Hosmillo, William L. Hamilton, Martin D. Curran, Theresa Feltwell, Anna Yakovleva, Charlotte J. Houldcroft, Aminu S. Jahun, Sarah L. Caddy, Ian Goodfellow
EPI_ISL_425304	hCoV-19/England/CAMB-74544/2020 Europe / United Kingdom / England	2020-03-28	Department of Pathology, University of Cambridge	COVID-19 Genomics UK (COG-UK) Consortium	Luke W Meredith, M. Estee Torok , Myra Hosmillo, William L. Hamilton, Martin D. Curran, Theresa Feltwell, Anna Yakovleva, Charlotte J. Houldcroft, Aminu S. Jahun, Sarah L. Caddy, Ian Goodfellow
EPI_ISL_425305	hCoV-19/England/CAMB-74553/2020 Europe / United Kingdom / England	2020-03-28	Department of Pathology, University of Cambridge	COVID-19 Genomics UK (COG-UK) Consortium	Luke W Meredith, M. Estee Torok , Myra Hosmillo, William L. Hamilton, Martin D. Curran, Theresa Feltwell, Anna Yakovleva, Charlotte J. Houldcroft, Aminu S. Jahun, Sarah L. Caddy, Ian Goodfellow
EPI_ISL_425423	hCoV-19/England/CAMB-76D1D/2020 Europe / United Kingdom / England	2020-03-30	Department of Pathology, University of Cambridge	COVID-19 Genomics UK (COG-UK) Consortium	Luke W Meredith, M. Estee Torok , Myra Hosmillo, William L. Hamilton, Martin D. Curran, Theresa Feltwell, Anna Yakovleva, Charlotte J. Houldcroft, Aminu S. Jahun, Sarah L. Caddy, Ian Goodfellow
EPI_ISL_425424	hCoV-19/England/CAMB-76D3B/2020 Europe / United Kingdom / England	2020-03-31	Department of Pathology, University of Cambridge	COVID-19 Genomics UK (COG-UK) Consortium	Luke W Meredith, M. Estee Torok , Myra Hosmillo, William L. Hamilton, Martin D. Curran, Theresa Feltwell, Anna Yakovleva, Charlotte J. Houldcroft, Aminu S. Jahun, Sarah L. Caddy, Ian Goodfellow
EPI_ISL_425425	hCoV-19/England/CAMB-76D59/2020 Europe / United Kingdom / England	2020-03-30	Department of Pathology, University of Cambridge	COVID-19 Genomics UK (COG-UK) Consortium	Luke W Meredith, M. Estee Torok , Myra Hosmillo, William L. Hamilton, Martin D. Curran, Theresa Feltwell, Anna Yakovleva, Charlotte J. Houldcroft, Aminu S. Jahun, Sarah L. Caddy, Ian Goodfellow
EPI_ISL_425427	hCoV-19/England/CAMB-76D95/2020 Europe / United Kingdom / England	2020-03-31	Department of Pathology, University of Cambridge	COVID-19 Genomics UK (COG-UK) Consortium	Luke W

Meredith, M. Estee Torok , Myra Hosmillo, William L. Hamilton, Martin D. Curran, Theresa Feltwell, Anna Yakovleva, Charlotte J. Houldcroft, Aminu S. Jahun, Sarah L. Caddy, Ian Goodfellow
EPI_ISL_425428 hCoV-19/England/CAMB-76DB3/2020 Europe / United Kingdom / England 2020-03-31 Department of Pathology, University of Cambridge COVID-19 Genomics UK (COG-UK) Consortium Luke W Meredith, M. Estee Torok , Myra Hosmillo, William L. Hamilton, Martin D. Curran, Theresa Feltwell, Anna Yakovleva, Charlotte J. Houldcroft, Aminu S. Jahun, Sarah L. Caddy, Ian Goodfellow
EPI_ISL_425430 hCoV-19/England/CAMB-76DFF/2020 Europe / United Kingdom / England 2020-03-30 Department of Pathology, University of Cambridge COVID-19 Genomics UK (COG-UK) Consortium Luke W Meredith, M. Estee Torok , Myra Hosmillo, William L. Hamilton, Martin D. Curran, Theresa Feltwell, Anna Yakovleva, Charlotte J. Houldcroft, Aminu S. Jahun, Sarah L. Caddy, Ian Goodfellow
EPI_ISL_425431 hCoV-19/England/CAMB-76E1A/2020 Europe / United Kingdom / England 2020-03-30 Department of Pathology, University of Cambridge COVID-19 Genomics UK (COG-UK) Consortium Luke W Meredith, M. Estee Torok , Myra Hosmillo, William L. Hamilton, Martin D. Curran, Theresa Feltwell, Anna Yakovleva, Charlotte J. Houldcroft, Aminu S. Jahun, Sarah L. Caddy, Ian Goodfellow
EPI_ISL_425433 hCoV-19/England/CAMB-76E56/2020 Europe / United Kingdom / England 2020-03-30 Department of Pathology, University of Cambridge COVID-19 Genomics UK (COG-UK) Consortium Luke W Meredith, M. Estee Torok , Myra Hosmillo, William L. Hamilton, Martin D. Curran, Theresa Feltwell, Anna Yakovleva, Charlotte J. Houldcroft, Aminu S. Jahun, Sarah L. Caddy, Ian Goodfellow
EPI_ISL_425434 hCoV-19/England/CAMB-76E74/2020 Europe / United Kingdom / England 2020-03-31 Department of Pathology, University of Cambridge COVID-19 Genomics UK (COG-UK) Consortium Luke W Meredith, M. Estee Torok , Myra Hosmillo, William L. Hamilton, Martin D. Curran, Theresa Feltwell, Anna Yakovleva, Charlotte J. Houldcroft, Aminu S. Jahun, Sarah L. Caddy, Ian Goodfellow
EPI_ISL_425435 hCoV-19/England/CAMB-76E92/2020 Europe / United Kingdom / England 2020-04-01 Department of Pathology, University of Cambridge COVID-19 Genomics UK (COG-UK) Consortium Luke W Meredith, M. Estee Torok , Myra Hosmillo, William L. Hamilton, Martin D. Curran, Theresa Feltwell, Anna Yakovleva, Charlotte J. Houldcroft, Aminu S. Jahun, Sarah L. Caddy, Ian Goodfellow
EPI_ISL_425436 hCoV-19/England/CAMB-76EB0/2020 Europe / United Kingdom / England 2020-04-01 Department of Pathology, University of Cambridge COVID-19 Genomics UK (COG-UK) Consortium Luke W Meredith, M. Estee Torok , Myra Hosmillo, William L. Hamilton, Martin D. Curran, Theresa Feltwell, Anna Yakovleva, Charlotte J. Houldcroft, Aminu S. Jahun, Sarah L. Caddy, Ian Goodfellow
EPI_ISL_425437 hCoV-19/England/CAMB-76EDE/2020 Europe / United Kingdom / England 2020-03-31 Department of Pathology, University of Cambridge COVID-19 Genomics UK (COG-UK) Consortium Luke W Meredith, M. Estee Torok , Myra Hosmillo, William L. Hamilton, Martin D. Curran, Theresa Feltwell, Anna Yakovleva, Charlotte J. Houldcroft, Aminu S. Jahun, Sarah L. Caddy, Ian Goodfellow
EPI_ISL_425439 hCoV-19/England/CAMB-76F17/2020 Europe / United Kingdom / England 2020 Department of Pathology, University of Cambridge COVID-19 Genomics UK (COG-UK) Consortium Luke W Meredith, M. Estee Torok , Myra Hosmillo, William L. Hamilton, Martin D. Curran, Theresa Feltwell, Anna Yakovleva, Charlotte J. Houldcroft, Aminu S. Jahun, Sarah L. Caddy, Ian Goodfellow
EPI_ISL_425440 hCoV-19/England/CAMB-76F35/2020 Europe / United Kingdom / England 2020 Department of Pathology, University of Cambridge COVID-19 Genomics UK (COG-UK) Consortium Luke W Meredith, M. Estee Torok , Myra Hosmillo, William L. Hamilton, Martin D. Curran, Theresa Feltwell, Anna Yakovleva, Charlotte J. Houldcroft, Aminu S. Jahun, Sarah L. Caddy, Ian Goodfellow
EPI_ISL_425441 hCoV-19/England/CAMB-76F62/2020 Europe / United Kingdom / England 2020-03-31 Department of Pathology, University of Cambridge COVID-19 Genomics UK (COG-UK) Consortium Luke W Meredith, M. Estee Torok , Myra Hosmillo, William L. Hamilton, Martin D. Curran, Theresa Feltwell, Anna Yakovleva, Charlotte J. Houldcroft, Aminu S. Jahun, Sarah L. Caddy, Ian Goodfellow
EPI_ISL_425443 hCoV-19/England/CAMB-76F9F/2020 Europe / United Kingdom / England 2020-04-01 Department of Pathology, University of Cambridge COVID-19 Genomics UK (COG-UK) Consortium Luke W Meredith, M. Estee Torok , Myra Hosmillo, William L. Hamilton, Martin D. Curran, Theresa Feltwell, Anna Yakovleva, Charlotte J. Houldcroft, Aminu S. Jahun, Sarah L. Caddy, Ian Goodfellow
EPI_ISL_425446 hCoV-19/England/CAMB-76FCC/2020 Europe / United Kingdom / England 2020-04-01 Department of Pathology, University of Cambridge COVID-19 Genomics UK (COG-UK) Consortium Luke W Meredith, M. Estee Torok , Myra Hosmillo, William L. Hamilton, Martin D. Curran, Theresa Feltwell, Anna Yakovleva, Charlotte J. Houldcroft, Aminu S. Jahun, Sarah L. Caddy, Ian Goodfellow
EPI_ISL_425449 hCoV-19/England/CAMB-76FF9/2020 Europe / United Kingdom / England 2020-04-01 Department of Pathology, University of Cambridge COVID-19 Genomics UK (COG-UK) Consortium Luke W Meredith, M. Estee Torok , Myra Hosmillo, William L. Hamilton, Martin D. Curran, Theresa Feltwell, Anna Yakovleva, Charlotte J. Houldcroft, Aminu S. Jahun, Sarah L. Caddy, Ian Goodfellow
EPI_ISL_425450 hCoV-19/England/CAMB-77004/2020 Europe / United Kingdom / England 2020-03-31 Department of Pathology, University of Cambridge COVID-19 Genomics UK (COG-UK) Consortium Luke W Meredith, M. Estee Torok , Myra Hosmillo, William L. Hamilton, Martin D. Curran, Theresa Feltwell, Anna Yakovleva, Charlotte J. Houldcroft, Aminu S. Jahun, Sarah L. Caddy, Ian Goodfellow
EPI_ISL_425451 hCoV-19/England/CAMB-77013/2020 Europe / United Kingdom / England 2020-04-01 Department of Pathology, University of Cambridge COVID-19 Genomics UK (COG-UK) Consortium Luke W Meredith, M. Estee Torok , Myra Hosmillo, William L. Hamilton, Martin D. Curran, Theresa Feltwell, Anna Yakovleva, Charlotte J. Houldcroft, Aminu S. Jahun, Sarah L. Caddy, Ian Goodfellow
EPI_ISL_425452 hCoV-19/England/CAMB-77022/2020 Europe / United Kingdom / England 2020-03-31 Department of Pathology, University of Cambridge COVID-19 Genomics UK (COG-UK) Consortium Luke W Meredith, M. Estee Torok , Myra Hosmillo, William L. Hamilton, Martin D. Curran, Theresa Feltwell, Anna Yakovleva, Charlotte J. Houldcroft, Aminu S. Jahun, Sarah L. Caddy, Ian Goodfellow

EPI_ISL_425453 hCoV-19/England/CAMB-77031/2020 Europe / United Kingdom / England 2020-04-01
Department of Pathology, University of Cambridge COVID-19 Genomics UK (COG-UK) Consortium Luke W Meredith, M. Estee Torok , Myra Hosmillo, William L. Hamilton, Martin D. Curran, Theresa Feltwell, Anna Yakovleva, Charlotte J. Houldcroft, Aminu S. Jahun, Sarah L. Caddy, Ian Goodfellow
EPI_ISL_425454 hCoV-19/England/CAMB-77040/2020 Europe / United Kingdom / England 2020-03-31
Department of Pathology, University of Cambridge COVID-19 Genomics UK (COG-UK) Consortium Luke W Meredith, M. Estee Torok , Myra Hosmillo, William L. Hamilton, Martin D. Curran, Theresa Feltwell, Anna Yakovleva, Charlotte J. Houldcroft, Aminu S. Jahun, Sarah L. Caddy, Ian Goodfellow
EPI_ISL_425455 hCoV-19/England/CAMB-7705F/2020 Europe / United Kingdom / England 2020-03-31
Department of Pathology, University of Cambridge COVID-19 Genomics UK (COG-UK) Consortium Luke W Meredith, M. Estee Torok , Myra Hosmillo, William L. Hamilton, Martin D. Curran, Theresa Feltwell, Anna Yakovleva, Charlotte J. Houldcroft, Aminu S. Jahun, Sarah L. Caddy, Ian Goodfellow
EPI_ISL_425457 hCoV-19/England/CAMB-7707D/2020 Europe / United Kingdom / England 2020-04-01
Department of Pathology, University of Cambridge COVID-19 Genomics UK (COG-UK) Consortium Luke W Meredith, M. Estee Torok , Myra Hosmillo, William L. Hamilton, Martin D. Curran, Theresa Feltwell, Anna Yakovleva, Charlotte J. Houldcroft, Aminu S. Jahun, Sarah L. Caddy, Ian Goodfellow
EPI_ISL_425458 hCoV-19/England/CAMB-7708C/2020 Europe / United Kingdom / England 2020-03-31
Department of Pathology, University of Cambridge COVID-19 Genomics UK (COG-UK) Consortium Luke W Meredith, M. Estee Torok , Myra Hosmillo, William L. Hamilton, Martin D. Curran, Theresa Feltwell, Anna Yakovleva, Charlotte J. Houldcroft, Aminu S. Jahun, Sarah L. Caddy, Ian Goodfellow
EPI_ISL_425460 hCoV-19/England/CAMB-770B9/2020 Europe / United Kingdom / England 2020-03-31
Department of Pathology, University of Cambridge COVID-19 Genomics UK (COG-UK) Consortium Luke W Meredith, M. Estee Torok , Myra Hosmillo, William L. Hamilton, Martin D. Curran, Theresa Feltwell, Anna Yakovleva, Charlotte J. Houldcroft, Aminu S. Jahun, Sarah L. Caddy, Ian Goodfellow
EPI_ISL_425466 hCoV-19/England/NOTT-10DEEA/2020 Europe / United Kingdom / England 2020-03-11
Queens Medical Centre, Clinical Microbiology Department / DeepSeq Nottingham COVID-19 Genomics UK (COG-UK) Consortium Gemma Clark, Wendy Smith, Manjinder Khakh, Hannah Howson-Wells, Jonathan Ball, Patrick McClure, Joseph Chappell, Theocharis Tsoleridis, Nadine Holmes, Matthew Carlisle, Christopher Moore, Fei Sang, Johnny Debebe, Victoria Wright, Matthew Loose
EPI_ISL_425468 hCoV-19/England/NOTT-10DF05/2020 Europe / United Kingdom / England 2020-03-12
Queens Medical Centre, Clinical Microbiology Department / DeepSeq Nottingham COVID-19 Genomics UK (COG-UK) Consortium Gemma Clark, Wendy Smith, Manjinder Khakh, Hannah Howson-Wells, Jonathan Ball, Patrick McClure, Joseph Chappell, Theocharis Tsoleridis, Nadine Holmes, Matthew Carlisle, Christopher Moore, Fei Sang, Johnny Debebe, Victoria Wright, Matthew Loose
EPI_ISL_425470 hCoV-19/England/NOTT-10DF23/2020 Europe / United Kingdom / England 2020-03-12
Queens Medical Centre, Clinical Microbiology Department / DeepSeq Nottingham COVID-19 Genomics UK (COG-UK) Consortium Gemma Clark, Wendy Smith, Manjinder Khakh, Hannah Howson-Wells, Jonathan Ball, Patrick McClure, Joseph Chappell, Theocharis Tsoleridis, Nadine Holmes, Matthew Carlisle, Christopher Moore, Fei Sang, Johnny Debebe, Victoria Wright, Matthew Loose
EPI_ISL_425471 hCoV-19/England/NOTT-10DF32/2020 Europe / United Kingdom / England 2020-03-13
Queens Medical Centre, Clinical Microbiology Department / DeepSeq Nottingham COVID-19 Genomics UK (COG-UK) Consortium Gemma Clark, Wendy Smith, Manjinder Khakh, Hannah Howson-Wells, Jonathan Ball, Patrick McClure, Joseph Chappell, Theocharis Tsoleridis, Nadine Holmes, Matthew Carlisle, Christopher Moore, Fei Sang, Johnny Debebe, Victoria Wright, Matthew Loose
EPI_ISL_425473 hCoV-19/England/NOTT-10DF50/2020 Europe / United Kingdom / England 2020-03-14
Queens Medical Centre, Clinical Microbiology Department / DeepSeq Nottingham COVID-19 Genomics UK (COG-UK) Consortium Gemma Clark, Wendy Smith, Manjinder Khakh, Hannah Howson-Wells, Jonathan Ball, Patrick McClure, Joseph Chappell, Theocharis Tsoleridis, Nadine Holmes, Matthew Carlisle, Christopher Moore, Fei Sang, Johnny Debebe, Victoria Wright, Matthew Loose
EPI_ISL_425474 hCoV-19/England/NOTT-10DF6F/2020 Europe / United Kingdom / England 2020-03-14
Queens Medical Centre, Clinical Microbiology Department / DeepSeq Nottingham COVID-19 Genomics UK (COG-UK) Consortium Gemma Clark, Wendy Smith, Manjinder Khakh, Hannah Howson-Wells, Jonathan Ball, Patrick McClure, Joseph Chappell, Theocharis Tsoleridis, Nadine Holmes, Matthew Carlisle, Christopher Moore, Fei Sang, Johnny Debebe, Victoria Wright, Matthew Loose
EPI_ISL_425475 hCoV-19/England/NOTT-10DF7E/2020 Europe / United Kingdom / England 2020-03-14
Queens Medical Centre, Clinical Microbiology Department / DeepSeq Nottingham COVID-19 Genomics UK (COG-UK) Consortium Gemma Clark, Wendy Smith, Manjinder Khakh, Hannah Howson-Wells, Jonathan Ball, Patrick McClure, Joseph Chappell, Theocharis Tsoleridis, Nadine Holmes, Matthew Carlisle, Christopher Moore, Fei Sang, Johnny Debebe, Victoria Wright, Matthew Loose
EPI_ISL_425476 hCoV-19/England/NOTT-10DF9C/2020 Europe / United Kingdom / England 2020-03-14
Queens Medical Centre, Clinical Microbiology Department / DeepSeq Nottingham COVID-19 Genomics UK (COG-UK) Consortium Gemma Clark, Wendy Smith, Manjinder Khakh, Hannah Howson-Wells, Jonathan Ball, Patrick McClure, Joseph Chappell, Theocharis Tsoleridis, Nadine Holmes, Matthew Carlisle, Christopher Moore, Fei Sang, Johnny Debebe, Victoria Wright, Matthew Loose
EPI_ISL_425477 hCoV-19/England/NOTT-10DFAB/2020 Europe / United Kingdom / England 2020-03-14
Queens Medical Centre, Clinical Microbiology Department / DeepSeq Nottingham COVID-19 Genomics UK (COG-UK) Consortium Gemma Clark, Wendy Smith, Manjinder Khakh, Hannah Howson-Wells, Jonathan Ball, Patrick McClure, Joseph Chappell, Theocharis Tsoleridis, Nadine Holmes, Matthew Carlisle, Christopher Moore, Fei Sang, Johnny Debebe, Victoria Wright, Matthew Loose
EPI_ISL_425480 hCoV-19/England/NOTT-10DFD8/2020 Europe / United Kingdom / England 2020-03-16

Queens Medical Centre, Clinical Microbiology Department / DeepSeq Nottingham    COVID-19 Genomics UK (COG-UK) Consortium      Gemma Clark, Wendy Smith, Manjinder Khakh, Hannah Howson-Wells, Jonathan Ball, Patrick McClure, Joseph Chappell, Theocharis Tsoleridis, Nadine Holmes, Matthew Carlisle, Christopher Moore, Fei Sang, Johnny Debebe, Victoria Wright, Matthew Loose
EPI_ISL_425481  hCoV-19/England/NOTT-10DFE7/2020        Europe / United Kingdom / England       2020-03-16
Queens Medical Centre, Clinical Microbiology Department / DeepSeq Nottingham    COVID-19 Genomics UK (COG-UK) Consortium      Gemma Clark, Wendy Smith, Manjinder Khakh, Hannah Howson-Wells, Jonathan Ball, Patrick McClure, Joseph Chappell, Theocharis Tsoleridis, Nadine Holmes, Matthew Carlisle, Christopher Moore, Fei Sang, Johnny Debebe, Victoria Wright, Matthew Loose
EPI_ISL_425482  hCoV-19/England/NOTT-10DFF6/2020        Europe / United Kingdom / England       2020-03-16
Queens Medical Centre, Clinical Microbiology Department / DeepSeq Nottingham    COVID-19 Genomics UK (COG-UK) Consortium      Gemma Clark, Wendy Smith, Manjinder Khakh, Hannah Howson-Wells, Jonathan Ball, Patrick McClure, Joseph Chappell, Theocharis Tsoleridis, Nadine Holmes, Matthew Carlisle, Christopher Moore, Fei Sang, Johnny Debebe, Victoria Wright, Matthew Loose
EPI_ISL_425483  hCoV-19/England/NOTT-10E001/2020        Europe / United Kingdom / England       2020-03-16
Queens Medical Centre, Clinical Microbiology Department / DeepSeq Nottingham    COVID-19 Genomics UK (COG-UK) Consortium      Gemma Clark, Wendy Smith, Manjinder Khakh, Hannah Howson-Wells, Jonathan Ball, Patrick McClure, Joseph Chappell, Theocharis Tsoleridis, Nadine Holmes, Matthew Carlisle, Christopher Moore, Fei Sang, Johnny Debebe, Victoria Wright, Matthew Loose
EPI_ISL_425484  hCoV-19/England/NOTT-10E010/2020        Europe / United Kingdom / England       2020-03-17
Queens Medical Centre, Clinical Microbiology Department / DeepSeq Nottingham    COVID-19 Genomics UK (COG-UK) Consortium      Gemma Clark, Wendy Smith, Manjinder Khakh, Hannah Howson-Wells, Jonathan Ball, Patrick McClure, Joseph Chappell, Theocharis Tsoleridis, Nadine Holmes, Matthew Carlisle, Christopher Moore, Fei Sang, Johnny Debebe, Victoria Wright, Matthew Loose
EPI_ISL_425486  hCoV-19/England/NOTT-10E03E/2020        Europe / United Kingdom / England       2020-03-17
Queens Medical Centre, Clinical Microbiology Department / DeepSeq Nottingham    COVID-19 Genomics UK (COG-UK) Consortium      Gemma Clark, Wendy Smith, Manjinder Khakh, Hannah Howson-Wells, Jonathan Ball, Patrick McClure, Joseph Chappell, Theocharis Tsoleridis, Nadine Holmes, Matthew Carlisle, Christopher Moore, Fei Sang, Johnny Debebe, Victoria Wright, Matthew Loose
EPI_ISL_425487  hCoV-19/England/NOTT-10E04D/2020        Europe / United Kingdom / England       2020-03-17
Queens Medical Centre, Clinical Microbiology Department / DeepSeq Nottingham    COVID-19 Genomics UK (COG-UK) Consortium      Gemma Clark, Wendy Smith, Manjinder Khakh, Hannah Howson-Wells, Jonathan Ball, Patrick McClure, Joseph Chappell, Theocharis Tsoleridis, Nadine Holmes, Matthew Carlisle, Christopher Moore, Fei Sang, Johnny Debebe, Victoria Wright, Matthew Loose
EPI_ISL_425488  hCoV-19/England/NOTT-10E05C/2020        Europe / United Kingdom / England       2020-03-18
Queens Medical Centre, Clinical Microbiology Department / DeepSeq Nottingham    COVID-19 Genomics UK (COG-UK) Consortium      Gemma Clark, Wendy Smith, Manjinder Khakh, Hannah Howson-Wells, Jonathan Ball, Patrick McClure, Joseph Chappell, Theocharis Tsoleridis, Nadine Holmes, Matthew Carlisle, Christopher Moore, Fei Sang, Johnny Debebe, Victoria Wright, Matthew Loose
EPI_ISL_425489  hCoV-19/England/NOTT-10E06B/2020        Europe / United Kingdom / England       2020-03-18
Queens Medical Centre, Clinical Microbiology Department / DeepSeq Nottingham    COVID-19 Genomics UK (COG-UK) Consortium      Gemma Clark, Wendy Smith, Manjinder Khakh, Hannah Howson-Wells, Jonathan Ball, Patrick McClure, Joseph Chappell, Theocharis Tsoleridis, Nadine Holmes, Matthew Carlisle, Christopher Moore, Fei Sang, Johnny Debebe, Victoria Wright, Matthew Loose
EPI_ISL_425491  hCoV-19/England/NOTT-10E089/2020        Europe / United Kingdom / England       2020-03-18
Queens Medical Centre, Clinical Microbiology Department / DeepSeq Nottingham    COVID-19 Genomics UK (COG-UK) Consortium      Gemma Clark, Wendy Smith, Manjinder Khakh, Hannah Howson-Wells, Jonathan Ball, Patrick McClure, Joseph Chappell, Theocharis Tsoleridis, Nadine Holmes, Matthew Carlisle, Christopher Moore, Fei Sang, Johnny Debebe, Victoria Wright, Matthew Loose
EPI_ISL_425492  hCoV-19/England/NOTT-10E098/2020        Europe / United Kingdom / England       2020-03-18
Queens Medical Centre, Clinical Microbiology Department / DeepSeq Nottingham    COVID-19 Genomics UK (COG-UK) Consortium      Gemma Clark, Wendy Smith, Manjinder Khakh, Hannah Howson-Wells, Jonathan Ball, Patrick McClure, Joseph Chappell, Theocharis Tsoleridis, Nadine Holmes, Matthew Carlisle, Christopher Moore, Fei Sang, Johnny Debebe, Victoria Wright, Matthew Loose
EPI_ISL_425494  hCoV-19/England/NOTT-10E0B6/2020        Europe / United Kingdom / England       2020-03-18
Queens Medical Centre, Clinical Microbiology Department / DeepSeq Nottingham    COVID-19 Genomics UK (COG-UK) Consortium      Gemma Clark, Wendy Smith, Manjinder Khakh, Hannah Howson-Wells, Jonathan Ball, Patrick McClure, Joseph Chappell, Theocharis Tsoleridis, Nadine Holmes, Matthew Carlisle, Christopher Moore, Fei Sang, Johnny Debebe, Victoria Wright, Matthew Loose
EPI_ISL_425495  hCoV-19/England/NOTT-10E0C5/2020        Europe / United Kingdom / England       2020-03-18
Queens Medical Centre, Clinical Microbiology Department / DeepSeq Nottingham    COVID-19 Genomics UK (COG-UK) Consortium      Gemma Clark, Wendy Smith, Manjinder Khakh, Hannah Howson-Wells, Jonathan Ball, Patrick McClure, Joseph Chappell, Theocharis Tsoleridis, Nadine Holmes, Matthew Carlisle, Christopher Moore, Fei Sang, Johnny Debebe, Victoria Wright, Matthew Loose
EPI_ISL_425496  hCoV-19/England/NOTT-10E0D4/2020        Europe / United Kingdom / England       2020-03-19
Queens Medical Centre, Clinical Microbiology Department / DeepSeq Nottingham    COVID-19 Genomics UK (COG-UK) Consortium      Gemma Clark, Wendy Smith, Manjinder Khakh, Hannah Howson-Wells, Jonathan Ball, Patrick McClure, Joseph Chappell, Theocharis Tsoleridis, Nadine Holmes, Matthew Carlisle, Christopher Moore, Fei Sang, Johnny Debebe, Victoria Wright, Matthew Loose
EPI_ISL_425497  hCoV-19/England/NOTT-10E0E3/2020        Europe / United Kingdom / England       2020-03-19

Queens Medical Centre, Clinical Microbiology Department / DeepSeq Nottingham COVID-19 Genomics UK (COG-UK) Consortium Gemma Clark, Wendy Smith, Manjinder Khakh, Hannah Howson-Wells, Jonathan Ball, Patrick McClure, Joseph Chappell, Theocharis Tsoleridis, Nadine Holmes, Matthew Carlisle, Christopher Moore, Fei Sang, Johnny Debebe, Victoria Wright, Matthew Loose
EPI_ISL_425498 hCoV-19/England/NOTT-10E0F2/2020 Europe / United Kingdom / England 2020-03-19 Queens Medical Centre, Clinical Microbiology Department / DeepSeq Nottingham COVID-19 Genomics UK (COG-UK) Consortium Gemma Clark, Wendy Smith, Manjinder Khakh, Hannah Howson-Wells, Jonathan Ball, Patrick McClure, Joseph Chappell, Theocharis Tsoleridis, Nadine Holmes, Matthew Carlisle, Christopher Moore, Fei Sang, Johnny Debebe, Victoria Wright, Matthew Loose
EPI_ISL_426630 hCoV-19/Taiwan/TSGH-02/2020 Asia / Taiwan / New Taipei city 2020-03-17 TSGH-CP molecular lab TSGH-CP molecular lab Cherng-Lih Perng, Ming-Jr Jian, Chih-Kai Chang, Jung-Chung Lin, Kuo-Ming Yeh, Chien-Wen Chen, Sheng-Kang Chiu, Hsing-Yi Chung, Shih-Hung Tsai, Kuo-Sheng Hung, Feng-Yee Chang, Hung-Sheng Shang
EPI_ISL_426631 hCoV-19/Taiwan/TSGH-03/2020 Asia / Taiwan / New Taipei City 2020-03-17 TSGH-CP molecular lab TSGH-CP molecular lab Cherng-Lih Perng, Ming-Jr Jian, Chih-Kai Chang, Jung-Chung Lin, Kuo-Ming Yeh, Chien-Wen Chen, Sheng-Kang Chiu, Hsing-Yi Chung, Shih-Hung Tsai, Kuo-Sheng Hung, Tien-Yao Chang, Feng-Yee Chang, Hung-Sheng Shang
EPI_ISL_426632 hCoV-19/Taiwan/TSGH-04/2020 Asia / Taiwan / New Taipei City 2020-03-17 TSGH-CP molecular lab TSGH-CP molecular lab Cherng-Lih Perng, Ming-Jr Jian, Chih-Kai Chang, Jung-Chung Lin, Kuo-Ming Yeh, Chien-Wen Chen, Sheng-Kang Chiu, Hsing-Yi Chung, Shih-Hung Tsai, Kuo-Sheng Hung, Tien-Yao Chang, Feng-Yee Chang, Hung-Sheng Shang
EPI_ISL_426634 hCoV-19/Australia/NT08/2020 Oceania / Australia / Northern territory 2020-03-20 Royal Darwin Hospital Pathology Microbiological Diagnostic Unit Public Health Laboratory and Victorian Infectious Diseases Reference Laboratory, Doherty Institute Meumann, E., Caly L., Seemann T., Sait, M., Schultz M., Druce J., Sherry, N.
EPI_ISL_426635 hCoV-19/Australia/NT18/2020 Oceania / Australia / Northern territory 2020-03-23 Royal Darwin Hospital Pathology Microbiological Diagnostic Unit Public Health Laboratory and Victorian Infectious Diseases Reference Laboratory, Doherty Institute Meumann, E., Caly L., Seemann T., Sait, M., Schultz M., Druce J., Sherry, N.
EPI_ISL_426636 hCoV-19/Australia/NT19/2020 Oceania / Australia / Northern territory 2020-03-25 Royal Darwin Hospital Pathology Microbiological Diagnostic Unit Public Health Laboratory and Victorian Infectious Diseases Reference Laboratory, Doherty Institute Meumann, E., Caly L., Seemann T., Sait, M., Schultz M., Druce J., Sherry, N.
EPI_ISL_426700 hCoV-19/Australia/VIC397/2020 Oceania / Australia / Victoria 2020-03-24 Victorian Infectious Diseases Reference Laboratory (VIDRL) Microbiological Diagnostic Unit Public Health Laboratory and Victorian Infectious Diseases Reference Laboratory, Doherty Institute Caly L., Seemann T., Sait, M., Schultz M., Druce J., Sherry, N.
EPI_ISL_426701 hCoV-19/Australia/VIC398/2020 Oceania / Australia / Victoria 2020-03-24 Victorian Infectious Diseases Reference Laboratory (VIDRL) Microbiological Diagnostic Unit Public Health Laboratory and Victorian Infectious Diseases Reference Laboratory, Doherty Institute Caly L., Seemann T., Sait, M., Schultz M., Druce J., Sherry, N.
EPI_ISL_426702 hCoV-19/Australia/VIC399/2020 Oceania / Australia / Victoria 2020-03-24 Victorian Infectious Diseases Reference Laboratory (VIDRL) Microbiological Diagnostic Unit Public Health Laboratory and Victorian Infectious Diseases Reference Laboratory, Doherty Institute Caly L., Seemann T., Sait, M., Schultz M., Druce J., Sherry, N.
EPI_ISL_426703 hCoV-19/Australia/VIC400/2020 Oceania / Australia / Victoria 2020-03-24 Victorian Infectious Diseases Reference Laboratory (VIDRL) Microbiological Diagnostic Unit Public Health Laboratory and Victorian Infectious Diseases Reference Laboratory, Doherty Institute Caly L., Seemann T., Sait, M., Schultz M., Druce J., Sherry, N.
EPI_ISL_426705 hCoV-19/Australia/VIC402/2020 Oceania / Australia / Victoria 2020-03-24 Victorian Infectious Diseases Reference Laboratory (VIDRL) Microbiological Diagnostic Unit Public Health Laboratory and Victorian Infectious Diseases Reference Laboratory, Doherty Institute Caly L., Seemann T., Sait, M., Schultz M., Druce J., Sherry, N.
EPI_ISL_426706 hCoV-19/Australia/VIC403/2020 Oceania / Australia / Victoria 2020-03-22 Victorian Infectious Diseases Reference Laboratory (VIDRL) Microbiological Diagnostic Unit Public Health Laboratory and Victorian Infectious Diseases Reference Laboratory, Doherty Institute Caly L., Seemann T., Sait, M., Schultz M., Druce J., Sherry, N.
EPI_ISL_426707 hCoV-19/Australia/VIC404/2020 Oceania / Australia / Victoria 2020-03-24 Victorian Infectious Diseases Reference Laboratory (VIDRL) Microbiological Diagnostic Unit Public Health Laboratory and Victorian Infectious Diseases Reference Laboratory, Doherty Institute Caly L., Seemann T., Sait, M., Schultz M., Druce J., Sherry, N.
EPI_ISL_426709 hCoV-19/Australia/VIC407/2020 Oceania / Australia / Victoria 2020-03-25 Victorian Infectious Diseases Reference Laboratory (VIDRL) Microbiological Diagnostic Unit Public Health Laboratory and Victorian Infectious Diseases Reference Laboratory, Doherty Institute Caly L., Seemann T., Sait, M., Schultz M., Druce J., Sherry, N.
EPI_ISL_426711 hCoV-19/Australia/VIC409/2020 Oceania / Australia / Victoria 2020-03-26 Victorian Infectious Diseases Reference Laboratory (VIDRL) Microbiological Diagnostic Unit Public Health Laboratory and Victorian Infectious Diseases Reference Laboratory, Doherty Institute Caly L., Seemann T., Sait, M., Schultz M., Druce J., Sherry, N.
EPI_ISL_426714 hCoV-19/Australia/VIC414/2020 Oceania / Australia / Victoria 2020-03-26 Victorian

Infectious Diseases Reference Laboratory (VIDRL)	Microbiological Diagnostic Unit Public Health Laboratory and Victorian Infectious Diseases Reference Laboratory, Doherty Institute	Caly L., Seemann T., Sait, M., Schultz M., Druce J., Sherry, N.
EPI_ISL_426715	hCoV-19/Australia/VIC415/2020	Oceania / Australia / Victoria	2020-03-25	Victorian Infectious Diseases Reference Laboratory (VIDRL)	Microbiological Diagnostic Unit Public Health Laboratory and Victorian Infectious Diseases Reference Laboratory, Doherty Institute	Caly L., Seemann T., Sait, M., Schultz M., Druce J., Sherry, N.
EPI_ISL_426717	hCoV-19/Australia/VIC417/2020	Oceania / Australia / Victoria	2020-03-25	Victorian Infectious Diseases Reference Laboratory (VIDRL)	Microbiological Diagnostic Unit Public Health Laboratory and Victorian Infectious Diseases Reference Laboratory, Doherty Institute	Caly L., Seemann T., Sait, M., Schultz M., Druce J., Sherry, N.
EPI_ISL_426718	hCoV-19/Australia/VIC418/2020	Oceania / Australia / Victoria	2020-03-25	Victorian Infectious Diseases Reference Laboratory (VIDRL)	Microbiological Diagnostic Unit Public Health Laboratory and Victorian Infectious Diseases Reference Laboratory, Doherty Institute	Caly L., Seemann T., Sait, M., Schultz M., Druce J., Sherry, N.
EPI_ISL_426721	hCoV-19/Australia/VIC421/2020	Oceania / Australia / Victoria	2020-03-25	Victorian Infectious Diseases Reference Laboratory (VIDRL)	Microbiological Diagnostic Unit Public Health Laboratory and Victorian Infectious Diseases Reference Laboratory, Doherty Institute	Caly L., Seemann T., Sait, M., Schultz M., Druce J., Sherry, N.
EPI_ISL_426725	hCoV-19/Australia/VIC425/2020	Oceania / Australia / Victoria	2020-03-25	Victorian Infectious Diseases Reference Laboratory (VIDRL)	Microbiological Diagnostic Unit Public Health Laboratory and Victorian Infectious Diseases Reference Laboratory, Doherty Institute	Caly L., Seemann T., Sait, M., Schultz M., Druce J., Sherry, N.
EPI_ISL_426727	hCoV-19/Australia/VIC427/2020	Oceania / Australia / Victoria	2020-03-25	Victorian Infectious Diseases Reference Laboratory (VIDRL)	Microbiological Diagnostic Unit Public Health Laboratory and Victorian Infectious Diseases Reference Laboratory, Doherty Institute	Caly L., Seemann T., Sait, M., Schultz M., Druce J., Sherry, N.
EPI_ISL_426729	hCoV-19/Australia/VIC429/2020	Oceania / Australia / Victoria	2020-03-26	Victorian Infectious Diseases Reference Laboratory (VIDRL)	Microbiological Diagnostic Unit Public Health Laboratory and Victorian Infectious Diseases Reference Laboratory, Doherty Institute	Caly L., Seemann T., Sait, M., Schultz M., Druce J., Sherry, N.
EPI_ISL_426730	hCoV-19/Australia/VIC430/2020	Oceania / Australia / Victoria	2020-03-26	Victorian Infectious Diseases Reference Laboratory (VIDRL)	Microbiological Diagnostic Unit Public Health Laboratory and Victorian Infectious Diseases Reference Laboratory, Doherty Institute	Caly L., Seemann T., Sait, M., Schultz M., Druce J., Sherry, N.
EPI_ISL_426732	hCoV-19/Australia/VIC433/2020	Oceania / Australia / Victoria	2020-03-25	Victorian Infectious Diseases Reference Laboratory (VIDRL)	Microbiological Diagnostic Unit Public Health Laboratory and Victorian Infectious Diseases Reference Laboratory, Doherty Institute	Caly L., Seemann T., Sait, M., Schultz M., Druce J., Sherry, N.
EPI_ISL_426733	hCoV-19/Australia/VIC434/2020	Oceania / Australia / Victoria	2020-03-25	Victorian Infectious Diseases Reference Laboratory (VIDRL)	Microbiological Diagnostic Unit Public Health Laboratory and Victorian Infectious Diseases Reference Laboratory, Doherty Institute	Caly L., Seemann T., Sait, M., Schultz M., Druce J., Sherry, N.
EPI_ISL_426734	hCoV-19/Australia/VIC437/2020	Oceania / Australia / Victoria	2020-03-26	Victorian Infectious Diseases Reference Laboratory (VIDRL)	Microbiological Diagnostic Unit Public Health Laboratory and Victorian Infectious Diseases Reference Laboratory, Doherty Institute	Caly L., Seemann T., Sait, M., Schultz M., Druce J., Sherry, N.
EPI_ISL_426735	hCoV-19/Australia/VIC438/2020	Oceania / Australia / Victoria	2020-03-26	Victorian Infectious Diseases Reference Laboratory (VIDRL)	Microbiological Diagnostic Unit Public Health Laboratory and Victorian Infectious Diseases Reference Laboratory, Doherty Institute	Caly L., Seemann T., Sait, M., Schultz M., Druce J., Sherry, N.
EPI_ISL_426736	hCoV-19/Australia/VIC439/2020	Oceania / Australia / Victoria	2020-03-26	Victorian Infectious Diseases Reference Laboratory (VIDRL)	Microbiological Diagnostic Unit Public Health Laboratory and Victorian Infectious Diseases Reference Laboratory, Doherty Institute	Caly L., Seemann T., Sait, M., Schultz M., Druce J., Sherry, N.
EPI_ISL_426737	hCoV-19/Australia/VIC440/2020	Oceania / Australia / Victoria	2020-03-26	Victorian Infectious Diseases Reference Laboratory (VIDRL)	Microbiological Diagnostic Unit Public Health Laboratory and Victorian Infectious Diseases Reference Laboratory, Doherty Institute	Caly L., Seemann T., Sait, M., Schultz M., Druce J., Sherry, N.
EPI_ISL_426740	hCoV-19/Australia/VIC443/2020	Oceania / Australia / Victoria	2020-03-27	Victorian Infectious Diseases Reference Laboratory (VIDRL)	Microbiological Diagnostic Unit Public Health Laboratory and Victorian Infectious Diseases Reference Laboratory, Doherty Institute	Caly L., Seemann T., Sait, M., Schultz M., Druce J., Sherry, N.
EPI_ISL_426741	hCoV-19/Australia/VIC444/2020	Oceania / Australia / Victoria	2020-03-27	Victorian Infectious Diseases Reference Laboratory (VIDRL)	Microbiological Diagnostic Unit Public Health Laboratory and Victorian Infectious Diseases Reference Laboratory, Doherty Institute	Caly L., Seemann T., Sait, M., Schultz M., Druce J., Sherry, N.
EPI_ISL_426743	hCoV-19/Australia/VIC447/2020	Oceania / Australia / Victoria	2020-03-27	Victorian Infectious Diseases Reference Laboratory (VIDRL)	Microbiological Diagnostic Unit Public Health Laboratory and Victorian Infectious Diseases Reference Laboratory, Doherty Institute	Caly L., Seemann T.,

Sait, M., Schultz M., Druce J., Sherry, N.
EPI_ISL_426745 hCoV-19/Australia/VIC449/2020 Oceania / Australia / Victoria 2020-03-24 Victorian Infectious Diseases Reference Laboratory (VIDRL) Microbiological Diagnostic Unit Public Health Laboratory and Victorian Infectious Diseases Reference Laboratory, Doherty Institute Caly L., Seemann T., Sait, M., Schultz M., Druce J., Sherry, N.
EPI_ISL_426746 hCoV-19/Australia/VIC450/2020 Oceania / Australia / Victoria 2020-03-25 Victorian Infectious Diseases Reference Laboratory (VIDRL) Microbiological Diagnostic Unit Public Health Laboratory and Victorian Infectious Diseases Reference Laboratory, Doherty Institute Caly L., Seemann T., Sait, M., Schultz M., Druce J., Sherry, N.
EPI_ISL_426747 hCoV-19/Australia/VIC451/2020 Oceania / Australia / Victoria 2020-03-25 Victorian Infectious Diseases Reference Laboratory (VIDRL) Microbiological Diagnostic Unit Public Health Laboratory and Victorian Infectious Diseases Reference Laboratory, Doherty Institute Caly L., Seemann T., Sait, M., Schultz M., Druce J., Sherry, N.
EPI_ISL_426751 hCoV-19/Australia/VIC456/2020 Oceania / Australia / Victoria 2020-03-25 Victorian Infectious Diseases Reference Laboratory (VIDRL) Microbiological Diagnostic Unit Public Health Laboratory and Victorian Infectious Diseases Reference Laboratory, Doherty Institute Caly L., Seemann T., Sait, M., Schultz M., Druce J., Sherry, N.
EPI_ISL_426752 hCoV-19/Australia/VIC458/2020 Oceania / Australia / Victoria 2020-03-26 Victorian Infectious Diseases Reference Laboratory (VIDRL) Microbiological Diagnostic Unit Public Health Laboratory and Victorian Infectious Diseases Reference Laboratory, Doherty Institute Caly L., Seemann T., Sait, M., Schultz M., Druce J., Sherry, N.
EPI_ISL_426754 hCoV-19/Australia/VIC461/2020 Oceania / Australia / Victoria 2020-03-27 Victorian Infectious Diseases Reference Laboratory (VIDRL) Microbiological Diagnostic Unit Public Health Laboratory and Victorian Infectious Diseases Reference Laboratory, Doherty Institute Caly L., Seemann T., Sait, M., Schultz M., Druce J., Sherry, N.
EPI_ISL_426756 hCoV-19/Australia/VIC463/2020 Oceania / Australia / Victoria 2020-03-26 Victorian Infectious Diseases Reference Laboratory (VIDRL) Microbiological Diagnostic Unit Public Health Laboratory and Victorian Infectious Diseases Reference Laboratory, Doherty Institute Caly L., Seemann T., Sait, M., Schultz M., Druce J., Sherry, N.
EPI_ISL_426761 hCoV-19/Australia/VIC468/2020 Oceania / Australia / Victoria 2020-03-27 Victorian Infectious Diseases Reference Laboratory (VIDRL) Microbiological Diagnostic Unit Public Health Laboratory and Victorian Infectious Diseases Reference Laboratory, Doherty Institute Caly L., Seemann T., Sait, M., Schultz M., Druce J., Sherry, N.
EPI_ISL_426763 hCoV-19/Australia/VIC471/2020 Oceania / Australia / Victoria 2020-03-26 Victorian Infectious Diseases Reference Laboratory (VIDRL) Microbiological Diagnostic Unit Public Health Laboratory and Victorian Infectious Diseases Reference Laboratory, Doherty Institute Caly L., Seemann T., Sait, M., Schultz M., Druce J., Sherry, N.
EPI_ISL_426764 hCoV-19/Australia/VIC472/2020 Oceania / Australia / Victoria 2020-03-26 Victorian Infectious Diseases Reference Laboratory (VIDRL) Microbiological Diagnostic Unit Public Health Laboratory and Victorian Infectious Diseases Reference Laboratory, Doherty Institute Caly L., Seemann T., Sait, M., Schultz M., Druce J., Sherry, N.
EPI_ISL_426765 hCoV-19/Australia/VIC492/2020 Oceania / Australia / Victoria 2020-03-23 Victorian Infectious Diseases Reference Laboratory (VIDRL) Microbiological Diagnostic Unit Public Health Laboratory and Victorian Infectious Diseases Reference Laboratory, Doherty Institute Caly L., Seemann T., Sait, M., Schultz M., Druce J., Sherry, N.
EPI_ISL_426766 hCoV-19/Australia/VIC493/2020 Oceania / Australia / Victoria 2020-03-22 Victorian Infectious Diseases Reference Laboratory (VIDRL) Microbiological Diagnostic Unit Public Health Laboratory and Victorian Infectious Diseases Reference Laboratory, Doherty Institute Caly L., Seemann T., Sait, M., Schultz M., Druce J., Sherry, N.
EPI_ISL_426767 hCoV-19/Australia/VIC494/2020 Oceania / Australia / Victoria 2020-03-23 Victorian Infectious Diseases Reference Laboratory (VIDRL) Microbiological Diagnostic Unit Public Health Laboratory and Victorian Infectious Diseases Reference Laboratory, Doherty Institute Caly L., Seemann T., Sait, M., Schultz M., Druce J., Sherry, N.
EPI_ISL_426768 hCoV-19/Australia/VIC495/2020 Oceania / Australia / Victoria 2020-03-23 Victorian Infectious Diseases Reference Laboratory (VIDRL) Microbiological Diagnostic Unit Public Health Laboratory and Victorian Infectious Diseases Reference Laboratory, Doherty Institute Caly L., Seemann T., Sait, M., Schultz M., Druce J., Sherry, N.
EPI_ISL_426769 hCoV-19/Australia/VIC496/2020 Oceania / Australia / Victoria 2020-03-23 Victorian Infectious Diseases Reference Laboratory (VIDRL) Microbiological Diagnostic Unit Public Health Laboratory and Victorian Infectious Diseases Reference Laboratory, Doherty Institute Caly L., Seemann T., Sait, M., Schultz M., Druce J., Sherry, N.
EPI_ISL_426770 hCoV-19/Australia/VIC497/2020 Oceania / Australia / Victoria 2020-03-23 Victorian Infectious Diseases Reference Laboratory (VIDRL) Microbiological Diagnostic Unit Public Health Laboratory and Victorian Infectious Diseases Reference Laboratory, Doherty Institute Caly L., Seemann T., Sait, M., Schultz M., Druce J., Sherry, N.
EPI_ISL_426771 hCoV-19/Australia/VIC498/2020 Oceania / Australia / Victoria 2020-03-23 Victorian Infectious Diseases Reference Laboratory (VIDRL) Microbiological Diagnostic Unit Public Health Laboratory and Victorian Infectious Diseases Reference Laboratory, Doherty Institute Caly L., Seemann T., Sait, M., Schultz M., Druce J., Sherry, N.
EPI_ISL_426772 hCoV-19/Australia/VIC500/2020 Oceania / Australia / Victoria 2020-03-24 Victorian

Infectious Diseases Reference Laboratory (VIDRL)	Microbiological Diagnostic Unit Public Health Laboratory and Victorian Infectious Diseases Reference Laboratory, Doherty Institute	Caly L., Seemann T., Sait, M., Schultz M., Druce J., Sherry, N.
EPI_ISL_426774	hCoV-19/Australia/VIC503/2020	Oceania / Australia / Victoria	2020-03-23	Victorian Infectious Diseases Reference Laboratory (VIDRL)	Microbiological Diagnostic Unit Public Health Laboratory and Victorian Infectious Diseases Reference Laboratory, Doherty Institute	Caly L., Seemann T., Sait, M., Schultz M., Druce J., Sherry, N.
EPI_ISL_426775	hCoV-19/Australia/VIC504/2020	Oceania / Australia / Victoria	2020-03-23	Victorian Infectious Diseases Reference Laboratory (VIDRL)	Microbiological Diagnostic Unit Public Health Laboratory and Victorian Infectious Diseases Reference Laboratory, Doherty Institute	Caly L., Seemann T., Sait, M., Schultz M., Druce J., Sherry, N.
EPI_ISL_426777	hCoV-19/Australia/VIC506/2020	Oceania / Australia / Victoria	2020-03-23	Victorian Infectious Diseases Reference Laboratory (VIDRL)	Microbiological Diagnostic Unit Public Health Laboratory and Victorian Infectious Diseases Reference Laboratory, Doherty Institute	Caly L., Seemann T., Sait, M., Schultz M., Druce J., Sherry, N.
EPI_ISL_426779	hCoV-19/Australia/VIC509/2020	Oceania / Australia / Victoria	2020-03-22	Victorian Infectious Diseases Reference Laboratory (VIDRL)	Microbiological Diagnostic Unit Public Health Laboratory and Victorian Infectious Diseases Reference Laboratory, Doherty Institute	Caly L., Seemann T., Sait, M., Schultz M., Druce J., Sherry, N.
EPI_ISL_426781	hCoV-19/Australia/VIC511/2020	Oceania / Australia / Victoria	2020-03-23	Victorian Infectious Diseases Reference Laboratory (VIDRL)	Microbiological Diagnostic Unit Public Health Laboratory and Victorian Infectious Diseases Reference Laboratory, Doherty Institute	Caly L., Seemann T., Sait, M., Schultz M., Druce J., Sherry, N.
EPI_ISL_426786	hCoV-19/Australia/VIC516/2020	Oceania / Australia / Victoria	2020-03-23	Victorian Infectious Diseases Reference Laboratory (VIDRL)	Microbiological Diagnostic Unit Public Health Laboratory and Victorian Infectious Diseases Reference Laboratory, Doherty Institute	Caly L., Seemann T., Sait, M., Schultz M., Druce J., Sherry, N.
EPI_ISL_426788	hCoV-19/Australia/VIC518/2020	Oceania / Australia / Victoria	2020-03-23	Victorian Infectious Diseases Reference Laboratory (VIDRL)	Microbiological Diagnostic Unit Public Health Laboratory and Victorian Infectious Diseases Reference Laboratory, Doherty Institute	Caly L., Seemann T., Sait, M., Schultz M., Druce J., Sherry, N.
EPI_ISL_426789	hCoV-19/Australia/VIC520/2020	Oceania / Australia / Victoria	2020-03-23	Victorian Infectious Diseases Reference Laboratory (VIDRL)	Microbiological Diagnostic Unit Public Health Laboratory and Victorian Infectious Diseases Reference Laboratory, Doherty Institute	Caly L., Seemann T., Sait, M., Schultz M., Druce J., Sherry, N.
EPI_ISL_426790	hCoV-19/Australia/VIC522/2020	Oceania / Australia / Victoria	2020-03-23	Victorian Infectious Diseases Reference Laboratory (VIDRL)	Microbiological Diagnostic Unit Public Health Laboratory and Victorian Infectious Diseases Reference Laboratory, Doherty Institute	Caly L., Seemann T., Sait, M., Schultz M., Druce J., Sherry, N.
EPI_ISL_426792	hCoV-19/Australia/VIC525/2020	Oceania / Australia / Victoria	2020-03-24	Victorian Infectious Diseases Reference Laboratory (VIDRL)	Microbiological Diagnostic Unit Public Health Laboratory and Victorian Infectious Diseases Reference Laboratory, Doherty Institute	Caly L., Seemann T., Sait, M., Schultz M., Druce J., Sherry, N.
EPI_ISL_426793	hCoV-19/Australia/VIC527/2020	Oceania / Australia / Victoria	2020-03-24	Victorian Infectious Diseases Reference Laboratory (VIDRL)	Microbiological Diagnostic Unit Public Health Laboratory and Victorian Infectious Diseases Reference Laboratory, Doherty Institute	Caly L., Seemann T., Sait, M., Schultz M., Druce J., Sherry, N.
EPI_ISL_426796	hCoV-19/Australia/VIC530/2020	Oceania / Australia / Victoria	2020-03-23	Victorian Infectious Diseases Reference Laboratory (VIDRL)	Microbiological Diagnostic Unit Public Health Laboratory and Victorian Infectious Diseases Reference Laboratory, Doherty Institute	Caly L., Seemann T., Sait, M., Schultz M., Druce J., Sherry, N.
EPI_ISL_426797	hCoV-19/Australia/VIC531/2020	Oceania / Australia / Victoria	2020-03-24	Victorian Infectious Diseases Reference Laboratory (VIDRL)	Microbiological Diagnostic Unit Public Health Laboratory and Victorian Infectious Diseases Reference Laboratory, Doherty Institute	Caly L., Seemann T., Sait, M., Schultz M., Druce J., Sherry, N.
EPI_ISL_426798	hCoV-19/Australia/VIC532/2020	Oceania / Australia / Victoria	2020-03-23	Victorian Infectious Diseases Reference Laboratory (VIDRL)	Microbiological Diagnostic Unit Public Health Laboratory and Victorian Infectious Diseases Reference Laboratory, Doherty Institute	Caly L., Seemann T., Sait, M., Schultz M., Druce J., Sherry, N.
EPI_ISL_426799	hCoV-19/Australia/VIC533/2020	Oceania / Australia / Victoria	2020-03-24	Victorian Infectious Diseases Reference Laboratory (VIDRL)	Microbiological Diagnostic Unit Public Health Laboratory and Victorian Infectious Diseases Reference Laboratory, Doherty Institute	Caly L., Seemann T., Sait, M., Schultz M., Druce J., Sherry, N.
EPI_ISL_428201	hCoV-19/Sweden/RV-FOI-6/2020	Europe / Sweden / Västerbotten	2020-03-20	Klinisk mikrobiologi, Region Västerbotten	Unit for Biological Agents, Department for CBRN Defence and Security, Swedish Defence Research Agency	FOI Bioinformatics team
EPI_ISL_428229	hCoV-19/Taiwan/TSGH-12/2020	Asia / Taiwan / Taipei	2020-03-18	TSGH-CP molecular lab	TSGH-CP molecular lab	Cherng-Lih Perng, Ming-Jr Jian, Chih-Kai Chang, Jung-Chung Lin, Kuo-Ming Yeh, Chien-Wen Chen, Sheng-Kang Chiu, Hsing-Yi Chung, Shih-Hung Tsai, Kuo-Sheng Hung, Tien-Yao Chang, Feng-Yee Chang, Hung-Sheng Shang

EPI_ISL_428230 hCoV-19/Taiwan/TSGH-13/2020 Asia / Taiwan / New Taipei City 2020-03-17 TSGH-CP molecular lab TSGH-CP molecular lab Cherng-Lih Perng, Ming-Jr Jian, Chih-Kai Chang, Jung-Chung Lin, Kuo-Ming Yeh, Chien-Wen Chen, Sheng-Kang Chiu, Hsing-Yi Chung, Shih-Hung Tsai, Kuo-Sheng Hung, Tien-Yao Chang, Feng-Yee Chang, Hung-Sheng Shang
EPI_ISL_428231 hCoV-19/Taiwan/TSGH-14/2020 Asia / Taiwan / New Taipei City 2020-03-24 TSGH-CP molecular lab TSGH-CP molecular lab Cherng-Lih Perng, Ming-Jr JIAN, Chih-Kai Chang, Jung-Chung Lin, Kuo-Ming Yeh, Chien-Wen Chen, Sheng-Kang Chiu, Hsing-Yi Chung, Shih-Hung Tsai, Kuo-Sheng Hung, Tien-Yao Chang, Feng-Yee Chang, Hung-Sheng Shang
EPI_ISL_428252 hCoV-19/USA/WI-UW-156/2020 North America / USA / Wisconsin / Wauwatosa 2020-03-25 University of Wisconsin-Madison AIDS Vaccine Research Laboratories University of Wisconsin-Madison AIDS Vaccine Research Laboratories Gage Moreno, Katarina Braun, et al. AIDS Vaccine Research Laboratories
EPI_ISL_428253 hCoV-19/USA/WI-UW-157/2020 North America / USA / Wisconsin / Milwaukee 2020-03-15 University of Wisconsin-Madison AIDS Vaccine Research Laboratories University of Wisconsin-Madison AIDS Vaccine Research Laboratories Gage Moreno, Katarina Braun, et al. AIDS Vaccine Research Laboratories
EPI_ISL_428254 hCoV-19/USA/WI-UW-158/2020 North America / USA / Wisconsin / Oak Creek 2020-03-15 University of Wisconsin-Madison AIDS Vaccine Research Laboratories University of Wisconsin-Madison AIDS Vaccine Research Laboratories Gage Moreno, Katarina Braun, et al. AIDS Vaccine Research Laboratories
EPI_ISL_428255 hCoV-19/USA/WI-UW-159/2020 North America / USA / Wisconsin / Oak Creek 2020-03-15 University of Wisconsin-Madison AIDS Vaccine Research Laboratories University of Wisconsin-Madison AIDS Vaccine Research Laboratories Gage Moreno, Katarina Braun, et al. AIDS Vaccine Research Laboratories
EPI_ISL_428256 hCoV-19/USA/WI-UW-160/2020 North America / USA / Wisconsin / Oak Creek 2020-03-15 University of Wisconsin-Madison AIDS Vaccine Research Laboratories University of Wisconsin-Madison AIDS Vaccine Research Laboratories Gage Moreno, Katarina Braun, et al. AIDS Vaccine Research Laboratories
EPI_ISL_428257 hCoV-19/USA/WI-UW-161/2020 North America / USA / Wisconsin / Milwaukee 2020-03-29 University of Wisconsin-Madison AIDS Vaccine Research Laboratories University of Wisconsin-Madison AIDS Vaccine Research Laboratories Gage Moreno, Katarina Braun, et al. AIDS Vaccine Research Laboratories
EPI_ISL_428258 hCoV-19/USA/WI-UW-162/2020 North America / USA / Wisconsin / Milwaukee 2020-03-29 University of Wisconsin-Madison AIDS Vaccine Research Laboratories University of Wisconsin-Madison AIDS Vaccine Research Laboratories Gage Moreno, Katarina Braun, et al. AIDS Vaccine Research Laboratories
EPI_ISL_428259 hCoV-19/USA/WI-UW-163/2020 North America / USA / Wisconsin / Elm Grove 2020-03-16 University of Wisconsin-Madison AIDS Vaccine Research Laboratories University of Wisconsin-Madison AIDS Vaccine Research Laboratories Gage Moreno, Katarina Braun, et al. AIDS Vaccine Research Laboratories
EPI_ISL_428260 hCoV-19/USA/WI-UW-164/2020 North America / USA / Wisconsin / Pewaukee 2020-03-16 University of Wisconsin-Madison AIDS Vaccine Research Laboratories University of Wisconsin-Madison AIDS Vaccine Research Laboratories Gage Moreno, Katarina Braun, et al. AIDS Vaccine Research Laboratories
EPI_ISL_428261 hCoV-19/USA/WI-UW-165/2020 North America / USA / Wisconsin / Milwaukee 2020-03-17 University of Wisconsin-Madison AIDS Vaccine Research Laboratories University of Wisconsin-Madison AIDS Vaccine Research Laboratories Gage Moreno, Katarina Braun, et al. AIDS Vaccine Research Laboratories
EPI_ISL_428262 hCoV-19/USA/WI-UW-166/2020 North America / USA / Wisconsin / Greenfield 2020-03-17 University of Wisconsin-Madison AIDS Vaccine Research Laboratories University of Wisconsin-Madison AIDS Vaccine Research Laboratories Gage Moreno, Katarina Braun, et al. AIDS Vaccine Research Laboratories
EPI_ISL_428263 hCoV-19/USA/WI-UW-167/2020 North America / USA / Wisconsin / New Berlin 2020-03-18 University of Wisconsin-Madison AIDS Vaccine Research Laboratories University of Wisconsin-Madison AIDS Vaccine Research Laboratories Gage Moreno, Katarina Braun, et al. AIDS Vaccine Research Laboratories
EPI_ISL_428264 hCoV-19/USA/WI-UW-168/2020 North America / USA / Wisconsin / Milwaukee 2020-03-19 University of Wisconsin-Madison AIDS Vaccine Research Laboratories University of Wisconsin-Madison AIDS Vaccine Research Laboratories Gage Moreno, Katarina Braun, et al. AIDS Vaccine Research Laboratories
EPI_ISL_428265 hCoV-19/USA/WI-UW-169/2020 North America / USA / Wisconsin / Grafton 2020-03-19 University of Wisconsin-Madison AIDS Vaccine Research Laboratories University of Wisconsin-Madison AIDS Vaccine Research Laboratories Gage Moreno, Katarina Braun, et al. AIDS Vaccine Research Laboratories
EPI_ISL_428266 hCoV-19/USA/WI-UW-170/2020 North America / USA / Wisconsin / Milwaukee 2020-03-19 University of Wisconsin-Madison AIDS Vaccine Research Laboratories University of Wisconsin-Madison AIDS Vaccine Research Laboratories Gage Moreno, Katarina Braun, et al. AIDS Vaccine Research Laboratories
EPI_ISL_428267 hCoV-19/USA/WI-UW-171/2020 North America / USA / Wisconsin / Milwaukee 2020-03-19 University of Wisconsin-Madison AIDS Vaccine Research Laboratories University of Wisconsin-Madison AIDS Vaccine Research Laboratories Gage Moreno, Katarina Braun, et al. AIDS Vaccine Research Laboratories
EPI_ISL_428268 hCoV-19/USA/WI-UW-172/2020 North America / USA / Wisconsin / Milwaukee 2020-03-19 University of Wisconsin-Madison AIDS Vaccine Research Laboratories University of Wisconsin-Madison AIDS Vaccine Research Laboratories Gage Moreno, Katarina Braun, et al. AIDS Vaccine Research Laboratories
EPI_ISL_428269 hCoV-19/USA/WI-UW-173/2020 North America / USA / Wisconsin / Milwaukee 2020-03-20 University of Wisconsin-Madison AIDS Vaccine Research Laboratories University of Wisconsin-Madison AIDS Vaccine Research Laboratories Gage Moreno, Katarina Braun, et al. AIDS Vaccine Research Laboratories
EPI_ISL_428270 hCoV-19/USA/WI-UW-174/2020 North America / USA / Wisconsin / Grafton 2020-03-20 University of Wisconsin-Madison AIDS Vaccine Research Laboratories University of Wisconsin-Madison AIDS Vaccine Research Laboratories Gage Moreno, Katarina Braun, et al. AIDS Vaccine Research Laboratories
EPI_ISL_428271 hCoV-19/USA/WI-UW-175/2020 North America / USA / Wisconsin / Grafton 2020-03-20 University of Wisconsin-Madison AIDS Vaccine Research Laboratories University of Wisconsin-Madison AIDS Vaccine Research Laboratories Gage Moreno, Katarina Braun, et al. AIDS Vaccine Research Laboratories
EPI_ISL_428272 hCoV-19/USA/WI-UW-176/2020 North America / USA / Wisconsin / Milwaukee 2020-03-20 University of Wisconsin-Madison AIDS Vaccine Research Laboratories University of Wisconsin-Madison AIDS

Vaccine Research Laboratories   Gage Moreno, Katarina Braun, et al. AIDS Vaccine Research Laboratories
EPI_ISL_428273  hCoV-19/USA/WI-UW-177/2020      North America / USA / Wisconsin / Milwaukee     2020-03-20
University of Wisconsin-Madison AIDS Vaccine Research Laboratories      University of Wisconsin-Madison AIDS Vaccine Research Laboratories   Gage Moreno, Katarina Braun, et al. AIDS Vaccine Research Laboratories
EPI_ISL_428274  hCoV-19/USA/WI-UW-178/2020      North America / USA / Wisconsin / Milwaukee     2020-03-20
University of Wisconsin-Madison AIDS Vaccine Research Laboratories      University of Wisconsin-Madison AIDS Vaccine Research Laboratories   Gage Moreno, Katarina Braun, et al. AIDS Vaccine Research Laboratories
EPI_ISL_428275  hCoV-19/USA/WI-UW-179/2020      North America / USA / Wisconsin / Milwaukee     2020-03-21
University of Wisconsin-Madison AIDS Vaccine Research Laboratories      University of Wisconsin-Madison AIDS Vaccine Research Laboratories   Gage Moreno, Katarina Braun, et al. AIDS Vaccine Research Laboratories
EPI_ISL_428276  hCoV-19/USA/WI-UW-180/2020      North America / USA / Wisconsin / Grafton       2020-03-21
University of Wisconsin-Madison AIDS Vaccine Research Laboratories      University of Wisconsin-Madison AIDS Vaccine Research Laboratories   Gage Moreno, Katarina Braun, et al. AIDS Vaccine Research Laboratories
EPI_ISL_428277  hCoV-19/USA/WI-UW-181/2020      North America / USA / Wisconsin / Milwaukee     2020-03-22
University of Wisconsin-Madison AIDS Vaccine Research Laboratories      University of Wisconsin-Madison AIDS Vaccine Research Laboratories   Gage Moreno, Katarina Braun, et al. AIDS Vaccine Research Laboratories
EPI_ISL_428278  hCoV-19/USA/WI-UW-182/2020      North America / USA / Wisconsin / Milwaukee     2020-03-22
University of Wisconsin-Madison AIDS Vaccine Research Laboratories      University of Wisconsin-Madison AIDS Vaccine Research Laboratories   Gage Moreno, Katarina Braun, et al. AIDS Vaccine Research Laboratories
EPI_ISL_428279  hCoV-19/USA/WI-UW-183/2020      North America / USA / Wisconsin / Milwaukee     2020-03-22
University of Wisconsin-Madison AIDS Vaccine Research Laboratories      University of Wisconsin-Madison AIDS Vaccine Research Laboratories   Gage Moreno, Katarina Braun, et al. AIDS Vaccine Research Laboratories
EPI_ISL_428280  hCoV-19/USA/WI-UW-184/2020      North America / USA / Wisconsin / Milwaukee     2020-03-22
University of Wisconsin-Madison AIDS Vaccine Research Laboratories      University of Wisconsin-Madison AIDS Vaccine Research Laboratories   Gage Moreno, Katarina Braun, et al. AIDS Vaccine Research Laboratories
EPI_ISL_428281  hCoV-19/USA/WI-UW-185/2020      North America / USA / Wisconsin / Milwaukee     2020-03-22
University of Wisconsin-Madison AIDS Vaccine Research Laboratories      University of Wisconsin-Madison AIDS Vaccine Research Laboratories   Gage Moreno, Katarina Braun, et al. AIDS Vaccine Research Laboratories
EPI_ISL_428282  hCoV-19/USA/WI-UW-186/2020      North America / USA / Wisconsin / Milwaukee     2020-03-23
University of Wisconsin-Madison AIDS Vaccine Research Laboratories      University of Wisconsin-Madison AIDS Vaccine Research Laboratories   Gage Moreno, Katarina Braun, et al. AIDS Vaccine Research Laboratories
EPI_ISL_428283  hCoV-19/USA/WI-UW-187/2020      North America / USA / Wisconsin / Milwaukee     2020-03-23
University of Wisconsin-Madison AIDS Vaccine Research Laboratories      University of Wisconsin-Madison AIDS Vaccine Research Laboratories   Gage Moreno, Katarina Braun, et al. AIDS Vaccine Research Laboratories
EPI_ISL_428284  hCoV-19/USA/WI-UW-188/2020      North America / USA / Wisconsin / Milwaukee     2020-03-23
University of Wisconsin-Madison AIDS Vaccine Research Laboratories      University of Wisconsin-Madison AIDS Vaccine Research Laboratories   Gage Moreno, Katarina Braun, et al. AIDS Vaccine Research Laboratories
EPI_ISL_428285  hCoV-19/USA/WI-UW-189/2020      North America / USA / Wisconsin / Milwaukee     2020-03-23
University of Wisconsin-Madison AIDS Vaccine Research Laboratories      University of Wisconsin-Madison AIDS Vaccine Research Laboratories   Gage Moreno, Katarina Braun, et al. AIDS Vaccine Research Laboratories
EPI_ISL_428286  hCoV-19/USA/WI-UW-190/2020      North America / USA / Wisconsin / Wauwatosa     2020-03-23
University of Wisconsin-Madison AIDS Vaccine Research Laboratories      University of Wisconsin-Madison AIDS Vaccine Research Laboratories   Gage Moreno, Katarina Braun, et al. AIDS Vaccine Research Laboratories
EPI_ISL_428287  hCoV-19/USA/WI-UW-191/2020      North America / USA / Wisconsin / Milwaukee     2020-03-23
University of Wisconsin-Madison AIDS Vaccine Research Laboratories      University of Wisconsin-Madison AIDS Vaccine Research Laboratories   Gage Moreno, Katarina Braun, et al. AIDS Vaccine Research Laboratories
EPI_ISL_428288  hCoV-19/USA/WI-UW-192/2020      North America / USA / Wisconsin / Milwaukee     2020-03-23
University of Wisconsin-Madison AIDS Vaccine Research Laboratories      University of Wisconsin-Madison AIDS Vaccine Research Laboratories   Gage Moreno, Katarina Braun, et al. AIDS Vaccine Research Laboratories
EPI_ISL_428289  hCoV-19/USA/WI-UW-193/2020      North America / USA / Wisconsin / Jackson       2020-03-23
University of Wisconsin-Madison AIDS Vaccine Research Laboratories      University of Wisconsin-Madison AIDS Vaccine Research Laboratories   Gage Moreno, Katarina Braun, et al. AIDS Vaccine Research Laboratories
EPI_ISL_428290  hCoV-19/USA/WI-UW-194/2020      North America / USA / Wisconsin / Grafton       2020-03-24
University of Wisconsin-Madison AIDS Vaccine Research Laboratories      University of Wisconsin-Madison AIDS Vaccine Research Laboratories   Gage Moreno, Katarina Braun, et al. AIDS Vaccine Research Laboratories
EPI_ISL_428291  hCoV-19/USA/WI-UW-195/2020      North America / USA / Wisconsin / Milwaukee     2020-03-24
University of Wisconsin-Madison AIDS Vaccine Research Laboratories      University of Wisconsin-Madison AIDS Vaccine Research Laboratories   Gage Moreno, Katarina Braun, et al. AIDS Vaccine Research Laboratories
EPI_ISL_428292  hCoV-19/USA/WI-UW-196/2020      North America / USA / Wisconsin / Milwaukee     2020-03-21
University of Wisconsin-Madison AIDS Vaccine Research Laboratories      University of Wisconsin-Madison AIDS Vaccine Research Laboratories   Gage Moreno, Katarina Braun, et al. AIDS Vaccine Research Laboratories
EPI_ISL_428293  hCoV-19/USA/WI-UW-197/2020      North America / USA / Wisconsin / Greenfield    2020-03-24
University of Wisconsin-Madison AIDS Vaccine Research Laboratories      University of Wisconsin-Madison AIDS Vaccine Research Laboratories   Gage Moreno, Katarina Braun, et al. AIDS Vaccine Research Laboratories
EPI_ISL_428294  hCoV-19/USA/WI-UW-198/2020      North America / USA / Wisconsin / Brookfield    2020-03-24
University of Wisconsin-Madison AIDS Vaccine Research Laboratories      University of Wisconsin-Madison AIDS Vaccine Research Laboratories   Gage Moreno, Katarina Braun, et al. AIDS Vaccine Research Laboratories
EPI_ISL_428295  hCoV-19/USA/WI-UW-199/2020      North America / USA / Wisconsin / Brookfield    2020-03-24
University of Wisconsin-Madison AIDS Vaccine Research Laboratories      University of Wisconsin-Madison AIDS Vaccine Research Laboratories   Gage Moreno, Katarina Braun, et al. AIDS Vaccine Research Laboratories

EPI_ISL_428296 hCoV-19/USA/WI-UW-200/2020 North America / USA / Wisconsin / Milwaukee 2020-03-24
University of Wisconsin-Madison AIDS Vaccine Research Laboratories University of Wisconsin-Madison AIDS Vaccine Research Laboratories Gage Moreno, Katarina Braun, et al. AIDS Vaccine Research Laboratories
EPI_ISL_428297 hCoV-19/USA/WI-UW-201/2020 North America / USA / Wisconsin / Grafton 2020-03-25
University of Wisconsin-Madison AIDS Vaccine Research Laboratories University of Wisconsin-Madison AIDS Vaccine Research Laboratories Gage Moreno, Katarina Braun, et al. AIDS Vaccine Research Laboratories
EPI_ISL_428298 hCoV-19/USA/WI-UW-202/2020 North America / USA / Wisconsin / Grafton 2020-03-25
University of Wisconsin-Madison AIDS Vaccine Research Laboratories University of Wisconsin-Madison AIDS Vaccine Research Laboratories Gage Moreno, Katarina Braun, et al. AIDS Vaccine Research Laboratories
EPI_ISL_428299 hCoV-19/USA/WI-UW-203/2020 North America / USA / Wisconsin / Grafton 2020-03-25
University of Wisconsin-Madison AIDS Vaccine Research Laboratories University of Wisconsin-Madison AIDS Vaccine Research Laboratories Gage Moreno, Katarina Braun, et al. AIDS Vaccine Research Laboratories
EPI_ISL_429000 hCoV-19/USA/CA-CZB0103/2020 North America / USA / California / San Francisco 2020-03-28 UCSF Clinical Microbiology Laboratory Chan-Zuckerberg Biohub CZB Cliahub Consortium
EPI_ISL_429001 hCoV-19/USA/CA-CZB046/2020 North America / USA / California / San Francisco 2020-03-25 UCSF Clinical Microbiology Laboratory Chan-Zuckerberg Biohub CZB Cliahub Consortium
EPI_ISL_429002 hCoV-19/USA/CA-CZB098/2020 North America / USA / California / San Francisco 2020-03-28 UCSF Clinical Microbiology Laboratory Chan-Zuckerberg Biohub CZB Cliahub Consortium
EPI_ISL_429003 hCoV-19/USA/CA-CZB0136/2020 North America / USA / California / San Francisco 2020-04-08 UCSF Clinical Microbiology Laboratory Chan-Zuckerberg Biohub CZB Cliahub Consortium
EPI_ISL_429004 hCoV-19/USA/CA-CZB085/2020 North America / USA / California / San Francisco 2020-03-28 UCSF Clinical Microbiology Laboratory Chan-Zuckerberg Biohub CZB Cliahub Consortium
EPI_ISL_429005 hCoV-19/USA/CA-CZB079/2020 North America / USA / California / San Francisco 2020-03-28 UCSF Clinical Microbiology Laboratory Chan-Zuckerberg Biohub CZB Cliahub Consortium
EPI_ISL_429006 hCoV-19/USA/CA-CZB095/2020 North America / USA / California / San Francisco 2020-03-28 UCSF Clinical Microbiology Laboratory Chan-Zuckerberg Biohub CZB Cliahub Consortium
EPI_ISL_429007 hCoV-19/USA/CA-CZB0458/2020 North America / USA / California / San Francisco 2020-03-18 UCSF Clinical Microbiology Laboratory Chan-Zuckerberg Biohub CZB Cliahub Consortium
EPI_ISL_429008 hCoV-19/USA/CA-CZB091/2020 North America / USA / California / San Francisco 2020-03-28 UCSF Clinical Microbiology Laboratory Chan-Zuckerberg Biohub CZB Cliahub Consortium
EPI_ISL_429009 hCoV-19/USA/CA-CZB0122/2020 North America / USA / California / San Francisco 2020-04-08 UCSF Clinical Microbiology Laboratory Chan-Zuckerberg Biohub CZB Cliahub Consortium
EPI_ISL_429010 hCoV-19/USA/CA-CZB0108/2020 North America / USA / California / San Francisco 2020-03-28 UCSF Clinical Microbiology Laboratory Chan-Zuckerberg Biohub CZB Cliahub Consortium
EPI_ISL_429011 hCoV-19/USA/CA-CZB0147/2020 North America / USA / California / San Francisco 2020-04-08 UCSF Clinical Microbiology Laboratory Chan-Zuckerberg Biohub CZB Cliahub Consortium
EPI_ISL_429012 hCoV-19/USA/CA-CZB016/2020 North America / USA / California / San Francisco 2020-03-25 UCSF Clinical Microbiology Laboratory Chan-Zuckerberg Biohub CZB Cliahub Consortium
EPI_ISL_429013 hCoV-19/USA/CA-CZB080/2020 North America / USA / California / San Francisco 2020-03-28 UCSF Clinical Microbiology Laboratory Chan-Zuckerberg Biohub CZB Cliahub Consortium
EPI_ISL_429014 hCoV-19/USA/CA-CZB0142/2020 North America / USA / California / San Francisco 2020-04-08 UCSF Clinical Microbiology Laboratory Chan-Zuckerberg Biohub CZB Cliahub Consortium
EPI_ISL_429015 hCoV-19/USA/CA-CZB0100/2020 North America / USA / California / San Francisco 2020-03-28 UCSF Clinical Microbiology Laboratory Chan-Zuckerberg Biohub CZB Cliahub Consortium
EPI_ISL_429016 hCoV-19/USA/CA-CZB0709/2020 North America / USA / California / San Francisco 2020-03-23 UCSF Clinical Microbiology Laboratory Chan-Zuckerberg Biohub CZB Cliahub Consortium
EPI_ISL_429017 hCoV-19/USA/CA-CZB030/2020 North America / USA / California / San Francisco 2020-03-25 UCSF Clinical Microbiology Laboratory Chan-Zuckerberg Biohub CZB Cliahub Consortium
EPI_ISL_429018 hCoV-19/USA/CA-CZB0750/2020 North America / USA / California / San Francisco 2020-04-03 UCSF Clinical Microbiology Laboratory Chan-Zuckerberg Biohub CZB Cliahub Consortium
EPI_ISL_429019 hCoV-19/USA/CA-CZB078/2020 North America / USA / California / San Francisco 2020-03-28 UCSF Clinical Microbiology Laboratory Chan-Zuckerberg Biohub CZB Cliahub Consortium
EPI_ISL_429020 hCoV-19/USA/CA-CZB0721/2020 North America / USA / California / San Francisco 2020-03-31 UCSF Clinical Microbiology Laboratory Chan-Zuckerberg Biohub CZB Cliahub Consortium
EPI_ISL_429021 hCoV-19/USA/CA-CZB045/2020 North America / USA / California / San Francisco 2020-03-25 UCSF Clinical Microbiology Laboratory Chan-Zuckerberg Biohub CZB Cliahub Consortium
EPI_ISL_429022 hCoV-19/USA/CA-CZB0713/2020 North America / USA / California / San Francisco 2020-03-23 UCSF Clinical Microbiology Laboratory Chan-Zuckerberg Biohub CZB Cliahub Consortium
EPI_ISL_429023 hCoV-19/USA/CA-CZB0740/2020 North America / USA / California / San Francisco 2020-03-31 UCSF Clinical Microbiology Laboratory Chan-Zuckerberg Biohub CZB Cliahub Consortium
EPI_ISL_429024 hCoV-19/USA/CA-CZB0705/2020 North America / USA / California / San Francisco 2020-03-23 UCSF Clinical Microbiology Laboratory Chan-Zuckerberg Biohub CZB Cliahub Consortium
EPI_ISL_429025 hCoV-19/USA/CA-CZB0737/2020 North America / USA / California / San Francisco 2020-03-31 UCSF Clinical Microbiology Laboratory Chan-Zuckerberg Biohub CZB Cliahub Consortium
EPI_ISL_429026 hCoV-19/USA/CA-CZB0731/2020 North America / USA / California / San Francisco 2020-03-31 UCSF Clinical Microbiology Laboratory Chan-Zuckerberg Biohub CZB Cliahub Consortium
EPI_ISL_429027 hCoV-19/USA/CA-CZB0137/2020 North America / USA / California / San Francisco 2020-04-08 UCSF Clinical Microbiology Laboratory Chan-Zuckerberg Biohub CZB Cliahub Consortium
EPI_ISL_429028 hCoV-19/USA/CA-CZB0746/2020 North America / USA / California / San Francisco 2020-04-03 UCSF Clinical Microbiology Laboratory Chan-Zuckerberg Biohub CZB Cliahub Consortium

EPI_ISL_429029  hCoV-19/USA/CA-CZB067/2020      North America / USA / California / San Francisco        2020-03-28   UCSF Clinical Microbiology Laboratory   Chan-Zuckerberg Biohub  CZB Cliahub Consortium
EPI_ISL_429030  hCoV-19/USA/CA-CZB072/2020      North America / USA / California / San Francisco        2020-03-28   UCSF Clinical Microbiology Laboratory   Chan-Zuckerberg Biohub  CZB Cliahub Consortium
EPI_ISL_429031  hCoV-19/USA/CA-CZB0139/2020      North America / USA / California / San Francisco        2020-04-08   UCSF Clinical Microbiology Laboratory   Chan-Zuckerberg Biohub  CZB Cliahub Consortium
EPI_ISL_429032  hCoV-19/USA/CA-CZB082/2020      North America / USA / California / San Francisco        2020-03-28   UCSF Clinical Microbiology Laboratory   Chan-Zuckerberg Biohub  CZB Cliahub Consortium
EPI_ISL_429033  hCoV-19/USA/CA-CZB086/2020      North America / USA / California / San Francisco        2020-03-28   UCSF Clinical Microbiology Laboratory   Chan-Zuckerberg Biohub  CZB Cliahub Consortium
EPI_ISL_429034  hCoV-19/USA/CA-CZB0706/2020      North America / USA / California / San Francisco        2020-03-23   UCSF Clinical Microbiology Laboratory   Chan-Zuckerberg Biohub  CZB Cliahub Consortium
EPI_ISL_429035  hCoV-19/USA/CA-CZB042/2020      North America / USA / California / San Francisco        2020-03-25   UCSF Clinical Microbiology Laboratory   Chan-Zuckerberg Biohub  CZB Cliahub Consortium
EPI_ISL_429036  hCoV-19/USA/CA-CZB0107/2020      North America / USA / California / San Francisco        2020-03-28   UCSF Clinical Microbiology Laboratory   Chan-Zuckerberg Biohub  CZB Cliahub Consortium
EPI_ISL_429037  hCoV-19/USA/CA-CZB09/2020      North America / USA / California / San Francisco        2020-03-25   UCSF Clinical Microbiology Laboratory   Chan-Zuckerberg Biohub  CZB Cliahub Consortium
EPI_ISL_429038  hCoV-19/USA/CA-CZB0707/2020      North America / USA / California / San Francisco        2020-03-23   UCSF Clinical Microbiology Laboratory   Chan-Zuckerberg Biohub  CZB Cliahub Consortium
EPI_ISL_429039  hCoV-19/USA/CA-CZB036/2020      North America / USA / California / San Francisco        2020-03-25   UCSF Clinical Microbiology Laboratory   Chan-Zuckerberg Biohub  CZB Cliahub Consortium
EPI_ISL_429040  hCoV-19/USA/CA-CZB0701/2020      North America / USA / California / San Francisco        2020-03-23   UCSF Clinical Microbiology Laboratory   Chan-Zuckerberg Biohub  CZB Cliahub Consortium
EPI_ISL_429041  hCoV-19/USA/CA-CZB096/2020      North America / USA / California / San Francisco        2020-03-28   UCSF Clinical Microbiology Laboratory   Chan-Zuckerberg Biohub  CZB Cliahub Consortium
EPI_ISL_429042  hCoV-19/USA/CA-CZB0710/2020      North America / USA / California / San Francisco        2020-03-23   UCSF Clinical Microbiology Laboratory   Chan-Zuckerberg Biohub  CZB Cliahub Consortium
EPI_ISL_429043  hCoV-19/USA/CA-CZB0156/2020      North America / USA / California / San Francisco        2020-04-08   UCSF Clinical Microbiology Laboratory   Chan-Zuckerberg Biohub  CZB Cliahub Consortium
EPI_ISL_429044  hCoV-19/USA/CA-CZB0158/2020      North America / USA / California / San Francisco        2020-04-08   UCSF Clinical Microbiology Laboratory   Chan-Zuckerberg Biohub  CZB Cliahub Consortium
EPI_ISL_429045  hCoV-19/USA/CA-CZB039/2020      North America / USA / California / San Francisco        2020-03-25   UCSF Clinical Microbiology Laboratory   Chan-Zuckerberg Biohub  CZB Cliahub Consortium
EPI_ISL_429046  hCoV-19/USA/CA-CZB0125/2020      North America / USA / California / San Francisco        2020-04-08   UCSF Clinical Microbiology Laboratory   Chan-Zuckerberg Biohub  CZB Cliahub Consortium
EPI_ISL_429047  hCoV-19/USA/CA-CZB0154/2020      North America / USA / California / San Francisco        2020-04-08   UCSF Clinical Microbiology Laboratory   Chan-Zuckerberg Biohub  CZB Cliahub Consortium
EPI_ISL_429048  hCoV-19/USA/CA-CZB043/2020      North America / USA / California / San Francisco        2020-03-25   UCSF Clinical Microbiology Laboratory   Chan-Zuckerberg Biohub  CZB Cliahub Consortium
EPI_ISL_429049  hCoV-19/USA/CA-CZB090/2020      North America / USA / California / San Francisco        2020-03-28   UCSF Clinical Microbiology Laboratory   Chan-Zuckerberg Biohub  CZB Cliahub Consortium
EPI_ISL_429050  hCoV-19/USA/CA-CZB089/2020      North America / USA / California / San Francisco        2020-03-28   UCSF Clinical Microbiology Laboratory   Chan-Zuckerberg Biohub  CZB Cliahub Consortium
EPI_ISL_429051  hCoV-19/USA/CA-CZB0741/2020      North America / USA / California / San Francisco        2020-04-03   UCSF Clinical Microbiology Laboratory   Chan-Zuckerberg Biohub  CZB Cliahub Consortium
EPI_ISL_429052  hCoV-19/USA/CA-CZB0714/2020      North America / USA / California / San Francisco        2020-03-23   UCSF Clinical Microbiology Laboratory   Chan-Zuckerberg Biohub  CZB Cliahub Consortium
EPI_ISL_429053  hCoV-19/USA/CA-CZB0167/2020      North America / USA / California / San Francisco        2020-04-08   UCSF Clinical Microbiology Laboratory   Chan-Zuckerberg Biohub  CZB Cliahub Consortium
EPI_ISL_429054  hCoV-19/USA/CA-CZB0141/2020      North America / USA / California / San Francisco        2020-04-08   UCSF Clinical Microbiology Laboratory   Chan-Zuckerberg Biohub  CZB Cliahub Consortium
EPI_ISL_429055  hCoV-19/USA/CA-CZB0726/2020      North America / USA / California / San Francisco        2020-03-31   UCSF Clinical Microbiology Laboratory   Chan-Zuckerberg Biohub  CZB Cliahub Consortium
EPI_ISL_429056  hCoV-19/USA/CA-CZB0101/2020      North America / USA / California / San Francisco        2020-03-28   UCSF Clinical Microbiology Laboratory   Chan-Zuckerberg Biohub  CZB Cliahub Consortium
EPI_ISL_429057  hCoV-19/USA/CA-CZB076/2020      North America / USA / California / San Francisco        2020-03-28   UCSF Clinical Microbiology Laboratory   Chan-Zuckerberg Biohub  CZB Cliahub Consortium
EPI_ISL_429058  hCoV-19/USA/CA-CZB0114/2020      North America / USA / California / San Francisco        2020-03-28   UCSF Clinical Microbiology Laboratory   Chan-Zuckerberg Biohub  CZB Cliahub Consortium
EPI_ISL_429059  hCoV-19/USA/CA-CZB0110/2020      North America / USA / California / San Francisco        2020-03-28   UCSF Clinical Microbiology Laboratory   Chan-Zuckerberg Biohub  CZB Cliahub Consortium
EPI_ISL_429508  hCoV-19/Denmark/ALAB-SSI412A/2020       Europe / Denmark / Unknown      2020-03-23   Department of Virus and Microbiological Special Diagnostics, Statens Serum Institut, Copenhagen, Denmark, Artillerivej 5, 2300 Copenahgen S       Albertsen lab, Department of Chemistry and Bioscience, Aalborg University, Denmark     Rasmus Kirkegaard
EPI_ISL_429509  hCoV-19/Denmark/ALAB-SSI413/2020        Europe / Denmark / Unknown      2020-03-10   Department of Virus and Microbiological Special Diagnostics, Statens Serum Institut, Copenhagen, Denmark, Artillerivej 5, 2300 Copenahgen S       Albertsen lab, Department of Chemistry and Bioscience, Aalborg University, Denmark     Rasmus Kirkegaard

EPI_ISL_429510 hCoV-19/Denmark/ALAB-SSI413A/2020 Europe / Denmark / Unknown 2020-03-23
Department of Virus and Microbiological Special Diagnostics, Statens Serum Institut, Copenhagen, Denmark, Artillerivej 5, 2300 Copenahgen S Albertsen lab, Department of Chemistry and Bioscience, Aalborg University, Denmark Rasmus Kirkegaard
EPI_ISL_429511 hCoV-19/Denmark/ALAB-SSI414/2020 Europe / Denmark / Unknown 2020-03-10
Department of Virus and Microbiological Special Diagnostics, Statens Serum Institut, Copenhagen, Denmark, Artillerivej 5, 2300 Copenahgen S Albertsen lab, Department of Chemistry and Bioscience, Aalborg University, Denmark Rasmus Kirkegaard
EPI_ISL_429512 hCoV-19/Denmark/ALAB-SSI414A/2020 Europe / Denmark / Unknown 2020-03-23
Department of Virus and Microbiological Special Diagnostics, Statens Serum Institut, Copenhagen, Denmark, Artillerivej 5, 2300 Copenahgen S Albertsen lab, Department of Chemistry and Bioscience, Aalborg University, Denmark Rasmus Kirkegaard
EPI_ISL_429513 hCoV-19/Denmark/ALAB-SSI416/2020 Europe / Denmark / Unknown 2020-03-10
Department of Virus and Microbiological Special Diagnostics, Statens Serum Institut, Copenhagen, Denmark, Artillerivej 5, 2300 Copenahgen S Albertsen lab, Department of Chemistry and Bioscience, Aalborg University, Denmark Rasmus Kirkegaard
EPI_ISL_429514 hCoV-19/Denmark/ALAB-SSI417/2020 Europe / Denmark / Unknown 2020-03-10
Department of Virus and Microbiological Special Diagnostics, Statens Serum Institut, Copenhagen, Denmark, Artillerivej 5, 2300 Copenahgen S Albertsen lab, Department of Chemistry and Bioscience, Aalborg University, Denmark Rasmus Kirkegaard
EPI_ISL_429516 hCoV-19/Denmark/ALAB-SSI418/2020 Europe / Denmark / Unknown 2020-03-10
Department of Virus and Microbiological Special Diagnostics, Statens Serum Institut, Copenhagen, Denmark, Artillerivej 5, 2300 Copenahgen S Albertsen lab, Department of Chemistry and Bioscience, Aalborg University, Denmark Rasmus Kirkegaard
EPI_ISL_429517 hCoV-19/Denmark/ALAB-SSI419/2020 Europe / Denmark / Unknown 2020-03-11
Department of Virus and Microbiological Special Diagnostics, Statens Serum Institut, Copenhagen, Denmark, Artillerivej 5, 2300 Copenahgen S Albertsen lab, Department of Chemistry and Bioscience, Aalborg University, Denmark Rasmus Kirkegaard
EPI_ISL_429518 hCoV-19/Denmark/ALAB-SSI420/2020 Europe / Denmark / Unknown 2020-03-11
Department of Virus and Microbiological Special Diagnostics, Statens Serum Institut, Copenhagen, Denmark, Artillerivej 5, 2300 Copenahgen S Albertsen lab, Department of Chemistry and Bioscience, Aalborg University, Denmark Rasmus Kirkegaard
EPI_ISL_429519 hCoV-19/Denmark/ALAB-SSI420A/2020 Europe / Denmark / Unknown 2020-03-22
Department of Virus and Microbiological Special Diagnostics, Statens Serum Institut, Copenhagen, Denmark, Artillerivej 5, 2300 Copenahgen S Albertsen lab, Department of Chemistry and Bioscience, Aalborg University, Denmark Rasmus Kirkegaard
EPI_ISL_429520 hCoV-19/Denmark/ALAB-SSI423/2020 Europe / Denmark / Unknown 2020-03-11
Department of Virus and Microbiological Special Diagnostics, Statens Serum Institut, Copenhagen, Denmark, Artillerivej 5, 2300 Copenahgen S Albertsen lab, Department of Chemistry and Bioscience, Aalborg University, Denmark Rasmus Kirkegaard
EPI_ISL_429521 hCoV-19/Denmark/ALAB-SSI424/2020 Europe / Denmark / Unknown 2020-03-11
Department of Virus and Microbiological Special Diagnostics, Statens Serum Institut, Copenhagen, Denmark, Artillerivej 5, 2300 Copenahgen S Albertsen lab, Department of Chemistry and Bioscience, Aalborg University, Denmark Rasmus Kirkegaard
EPI_ISL_429524 hCoV-19/Denmark/ALAB-SSI425A/2020 Europe / Denmark / Unknown 2020-03-24
Department of Virus and Microbiological Special Diagnostics, Statens Serum Institut, Copenhagen, Denmark, Artillerivej 5, 2300 Copenahgen S Albertsen lab, Department of Chemistry and Bioscience, Aalborg University, Denmark Rasmus Kirkegaard
EPI_ISL_429525 hCoV-19/Denmark/ALAB-SSI426/2020 Europe / Denmark / Unknown 2020-03-12
Department of Virus and Microbiological Special Diagnostics, Statens Serum Institut, Copenhagen, Denmark, Artillerivej 5, 2300 Copenahgen S Albertsen lab, Department of Chemistry and Bioscience, Aalborg University, Denmark Rasmus Kirkegaard
EPI_ISL_429526 hCoV-19/Denmark/ALAB-SSI430/2020 Europe / Denmark / Unknown 2020-03-24
Department of Virus and Microbiological Special Diagnostics, Statens Serum Institut, Copenhagen, Denmark, Artillerivej 5, 2300 Copenahgen S Albertsen lab, Department of Chemistry and Bioscience, Aalborg University, Denmark Rasmus Kirkegaard
EPI_ISL_429527 hCoV-19/Denmark/ALAB-SSI431/2020 Europe / Denmark / Unknown 2020-03-24
Department of Virus and Microbiological Special Diagnostics, Statens Serum Institut, Copenhagen, Denmark, Artillerivej 5, 2300 Copenahgen S Albertsen lab, Department of Chemistry and Bioscience, Aalborg University, Denmark Rasmus Kirkegaard
EPI_ISL_429528 hCoV-19/Denmark/ALAB-SSI432/2020 Europe / Denmark / Unknown 2020-03-24
Department of Virus and Microbiological Special Diagnostics, Statens Serum Institut, Copenhagen, Denmark, Artillerivej 5, 2300 Copenahgen S Albertsen lab, Department of Chemistry and Bioscience, Aalborg University, Denmark Rasmus Kirkegaard
EPI_ISL_429529 hCoV-19/Denmark/ALAB-SSI434/2020 Europe / Denmark / Unknown 2020-03-24
Department of Virus and Microbiological Special Diagnostics, Statens Serum Institut, Copenhagen, Denmark, Artillerivej 5, 2300 Copenahgen S Albertsen lab, Department of Chemistry and Bioscience, Aalborg University, Denmark Rasmus Kirkegaard
EPI_ISL_429530 hCoV-19/Denmark/ALAB-SSI437/2020 Europe / Denmark / Unknown 2020-03-25
Department of Virus and Microbiological Special Diagnostics, Statens Serum Institut, Copenhagen, Denmark,

Artillerivej 5, 2300 Copenahgen S	Albertsen lab, Department of Chemistry and Bioscience, Aalborg University, Denmark	Rasmus Kirkegaard
EPI_ISL_429533	hCoV-19/Denmark/ALAB-SSI444/2020	Europe / Denmark / Unknown	2020-03-25	Department of Virus and Microbiological Special Diagnostics, Statens Serum Institut, Copenhagen, Denmark, Artillerivej 5, 2300 Copenahgen S	Albertsen lab, Department of Chemistry and Bioscience, Aalborg University, Denmark	Rasmus Kirkegaard
EPI_ISL_429534	hCoV-19/Denmark/ALAB-SSI446/2020	Europe / Denmark / Unknown	2020-03-25	Department of Virus and Microbiological Special Diagnostics, Statens Serum Institut, Copenhagen, Denmark, Artillerivej 5, 2300 Copenahgen S	Albertsen lab, Department of Chemistry and Bioscience, Aalborg University, Denmark	Rasmus Kirkegaard
EPI_ISL_429535	hCoV-19/Denmark/ALAB-SSI448/2020	Europe / Denmark / Unknown	2020-03-25	Department of Virus and Microbiological Special Diagnostics, Statens Serum Institut, Copenhagen, Denmark, Artillerivej 5, 2300 Copenahgen S	Albertsen lab, Department of Chemistry and Bioscience, Aalborg University, Denmark	Rasmus Kirkegaard
EPI_ISL_429536	hCoV-19/Denmark/ALAB-SSI449/2020	Europe / Denmark / Unknown	2020-03-25	Department of Virus and Microbiological Special Diagnostics, Statens Serum Institut, Copenhagen, Denmark, Artillerivej 5, 2300 Copenahgen S	Albertsen lab, Department of Chemistry and Bioscience, Aalborg University, Denmark	Rasmus Kirkegaard
EPI_ISL_429537	hCoV-19/Denmark/ALAB-SSI450/2020	Europe / Denmark / Unknown	2020-03-24	Department of Virus and Microbiological Special Diagnostics, Statens Serum Institut, Copenhagen, Denmark, Artillerivej 5, 2300 Copenahgen S	Albertsen lab, Department of Chemistry and Bioscience, Aalborg University, Denmark	Rasmus Kirkegaard
EPI_ISL_429538	hCoV-19/Denmark/ALAB-SSI451/2020	Europe / Denmark / Unknown	2020-03-25	Department of Virus and Microbiological Special Diagnostics, Statens Serum Institut, Copenhagen, Denmark, Artillerivej 5, 2300 Copenahgen S	Albertsen lab, Department of Chemistry and Bioscience, Aalborg University, Denmark	Rasmus Kirkegaard
EPI_ISL_429539	hCoV-19/Denmark/ALAB-SSI454/2020	Europe / Denmark / Unknown	2020-03-25	Department of Virus and Microbiological Special Diagnostics, Statens Serum Institut, Copenhagen, Denmark, Artillerivej 5, 2300 Copenahgen S	Albertsen lab, Department of Chemistry and Bioscience, Aalborg University, Denmark	Rasmus Kirkegaard
EPI_ISL_429540	hCoV-19/Denmark/ALAB-SSI456/2020	Europe / Denmark / Unknown	2020-03-25	Department of Virus and Microbiological Special Diagnostics, Statens Serum Institut, Copenhagen, Denmark, Artillerivej 5, 2300 Copenahgen S	Albertsen lab, Department of Chemistry and Bioscience, Aalborg University, Denmark	Rasmus Kirkegaard
EPI_ISL_429542	hCoV-19/Denmark/ALAB-SSI461/2020	Europe / Denmark / Unknown	2020-03-25	Department of Virus and Microbiological Special Diagnostics, Statens Serum Institut, Copenhagen, Denmark, Artillerivej 5, 2300 Copenahgen S	Albertsen lab, Department of Chemistry and Bioscience, Aalborg University, Denmark	Rasmus Kirkegaard
EPI_ISL_429543	hCoV-19/Denmark/ALAB-SSI462/2020	Europe / Denmark / Unknown	2020-03-25	Department of Virus and Microbiological Special Diagnostics, Statens Serum Institut, Copenhagen, Denmark, Artillerivej 5, 2300 Copenahgen S	Albertsen lab, Department of Chemistry and Bioscience, Aalborg University, Denmark	Rasmus Kirkegaard
EPI_ISL_429544	hCoV-19/Denmark/ALAB-SSI463/2020	Europe / Denmark / Unknown	2020-03-25	Department of Virus and Microbiological Special Diagnostics, Statens Serum Institut, Copenhagen, Denmark, Artillerivej 5, 2300 Copenahgen S	Albertsen lab, Department of Chemistry and Bioscience, Aalborg University, Denmark	Rasmus Kirkegaard
EPI_ISL_429545	hCoV-19/Denmark/ALAB-SSI464/2020	Europe / Denmark / Unknown	2020-03-25	Department of Virus and Microbiological Special Diagnostics, Statens Serum Institut, Copenhagen, Denmark, Artillerivej 5, 2300 Copenahgen S	Albertsen lab, Department of Chemistry and Bioscience, Aalborg University, Denmark	Rasmus Kirkegaard
EPI_ISL_429546	hCoV-19/Denmark/ALAB-SSI466/2020	Europe / Denmark / Unknown	2020-03-25	Department of Virus and Microbiological Special Diagnostics, Statens Serum Institut, Copenhagen, Denmark, Artillerivej 5, 2300 Copenahgen S	Albertsen lab, Department of Chemistry and Bioscience, Aalborg University, Denmark	Rasmus Kirkegaard
EPI_ISL_429547	hCoV-19/Denmark/ALAB-SSI467/2020	Europe / Denmark / Unknown	2020-03-25	Department of Virus and Microbiological Special Diagnostics, Statens Serum Institut, Copenhagen, Denmark, Artillerivej 5, 2300 Copenahgen S	Albertsen lab, Department of Chemistry and Bioscience, Aalborg University, Denmark	Rasmus Kirkegaard
EPI_ISL_429548	hCoV-19/Denmark/ALAB-SSI468/2020	Europe / Denmark / Unknown	2020-03-25	Department of Virus and Microbiological Special Diagnostics, Statens Serum Institut, Copenhagen, Denmark, Artillerivej 5, 2300 Copenahgen S	Albertsen lab, Department of Chemistry and Bioscience, Aalborg University, Denmark	Rasmus Kirkegaard
EPI_ISL_429549	hCoV-19/Denmark/ALAB-SSI469/2020	Europe / Denmark / Unknown	2020-03-25	Department of Virus and Microbiological Special Diagnostics, Statens Serum Institut, Copenhagen, Denmark, Artillerivej 5, 2300 Copenahgen S	Albertsen lab, Department of Chemistry and Bioscience, Aalborg University, Denmark	Rasmus Kirkegaard
EPI_ISL_429550	hCoV-19/Denmark/ALAB-SSI470/2020	Europe / Denmark / Unknown	2020-03-25	Department of Virus and Microbiological Special Diagnostics, Statens Serum Institut, Copenhagen, Denmark, Artillerivej 5, 2300 Copenahgen S	Albertsen lab, Department of Chemistry and Bioscience, Aalborg University, Denmark	Rasmus Kirkegaard

EPI_ISL_429551	hCoV-19/Denmark/ALAB-SSI471/2020	Europe / Denmark / Unknown	2020-03-25
Department of Virus and Microbiological Special Diagnostics, Statens Serum Institut, Copenhagen, Denmark, Artillerivej 5, 2300 Copenahgen S	Albertsen lab, Department of Chemistry and Bioscience, Aalborg University, Denmark	Rasmus Kirkegaard
EPI_ISL_429552	hCoV-19/Denmark/ALAB-SSI472/2020	Europe / Denmark / Unknown	2020-03-25
Department of Virus and Microbiological Special Diagnostics, Statens Serum Institut, Copenhagen, Denmark, Artillerivej 5, 2300 Copenahgen S	Albertsen lab, Department of Chemistry and Bioscience, Aalborg University, Denmark	Rasmus Kirkegaard
EPI_ISL_429553	hCoV-19/Denmark/ALAB-SSI473/2020	Europe / Denmark / Unknown	2020-03-25
Department of Virus and Microbiological Special Diagnostics, Statens Serum Institut, Copenhagen, Denmark, Artillerivej 5, 2300 Copenahgen S	Albertsen lab, Department of Chemistry and Bioscience, Aalborg University, Denmark	Rasmus Kirkegaard
EPI_ISL_429554	hCoV-19/Denmark/ALAB-SSI474/2020	Europe / Denmark / Unknown	2020-03-25
Department of Virus and Microbiological Special Diagnostics, Statens Serum Institut, Copenhagen, Denmark, Artillerivej 5, 2300 Copenahgen S	Albertsen lab, Department of Chemistry and Bioscience, Aalborg University, Denmark	Rasmus Kirkegaard
EPI_ISL_429555	hCoV-19/Denmark/ALAB-SSI475/2020	Europe / Denmark / Unknown	2020-03-25
Department of Virus and Microbiological Special Diagnostics, Statens Serum Institut, Copenhagen, Denmark, Artillerivej 5, 2300 Copenahgen S	Albertsen lab, Department of Chemistry and Bioscience, Aalborg University, Denmark	Rasmus Kirkegaard
EPI_ISL_429556	hCoV-19/Denmark/ALAB-SSI477/2020	Europe / Denmark / Unknown	2020-03-26
Department of Virus and Microbiological Special Diagnostics, Statens Serum Institut, Copenhagen, Denmark, Artillerivej 5, 2300 Copenahgen S	Albertsen lab, Department of Chemistry and Bioscience, Aalborg University, Denmark	Rasmus Kirkegaard
EPI_ISL_429557	hCoV-19/Denmark/ALAB-SSI478/2020	Europe / Denmark / Unknown	2020-03-26
Department of Virus and Microbiological Special Diagnostics, Statens Serum Institut, Copenhagen, Denmark, Artillerivej 5, 2300 Copenahgen S	Albertsen lab, Department of Chemistry and Bioscience, Aalborg University, Denmark	Rasmus Kirkegaard
EPI_ISL_429558	hCoV-19/Denmark/ALAB-SSI479/2020	Europe / Denmark / Unknown	2020-03-25
Department of Virus and Microbiological Special Diagnostics, Statens Serum Institut, Copenhagen, Denmark, Artillerivej 5, 2300 Copenahgen S	Albertsen lab, Department of Chemistry and Bioscience, Aalborg University, Denmark	Rasmus Kirkegaard
EPI_ISL_429559	hCoV-19/Denmark/ALAB-SSI480/2020	Europe / Denmark / Unknown	2020-03-25
Department of Virus and Microbiological Special Diagnostics, Statens Serum Institut, Copenhagen, Denmark, Artillerivej 5, 2300 Copenahgen S	Albertsen lab, Department of Chemistry and Bioscience, Aalborg University, Denmark	Rasmus Kirkegaard
EPI_ISL_429560	hCoV-19/Denmark/ALAB-SSI481/2020	Europe / Denmark / Unknown	2020-03-25
Department of Virus and Microbiological Special Diagnostics, Statens Serum Institut, Copenhagen, Denmark, Artillerivej 5, 2300 Copenahgen S	Albertsen lab, Department of Chemistry and Bioscience, Aalborg University, Denmark	Rasmus Kirkegaard
EPI_ISL_429561	hCoV-19/Denmark/ALAB-SSI482/2020	Europe / Denmark / Unknown	2020-03-25
Department of Virus and Microbiological Special Diagnostics, Statens Serum Institut, Copenhagen, Denmark, Artillerivej 5, 2300 Copenahgen S	Albertsen lab, Department of Chemistry and Bioscience, Aalborg University, Denmark	Rasmus Kirkegaard
EPI_ISL_429562	hCoV-19/Denmark/ALAB-SSI483/2020	Europe / Denmark / Unknown	2020-03-26
Department of Virus and Microbiological Special Diagnostics, Statens Serum Institut, Copenhagen, Denmark, Artillerivej 5, 2300 Copenahgen S	Albertsen lab, Department of Chemistry and Bioscience, Aalborg University, Denmark	Rasmus Kirkegaard
EPI_ISL_429563	hCoV-19/Denmark/ALAB-SSI484/2020	Europe / Denmark / Unknown	2020-03-23
Department of Virus and Microbiological Special Diagnostics, Statens Serum Institut, Copenhagen, Denmark, Artillerivej 5, 2300 Copenahgen S	Albertsen lab, Department of Chemistry and Bioscience, Aalborg University, Denmark	Rasmus Kirkegaard
EPI_ISL_429564	hCoV-19/Denmark/ALAB-SSI486/2020	Europe / Denmark / Unknown	2020-03-26
Department of Virus and Microbiological Special Diagnostics, Statens Serum Institut, Copenhagen, Denmark, Artillerivej 5, 2300 Copenahgen S	Albertsen lab, Department of Chemistry and Bioscience, Aalborg University, Denmark	Rasmus Kirkegaard
EPI_ISL_429565	hCoV-19/Denmark/ALAB-SSI488/2020	Europe / Denmark / Unknown	2020-03-24
Department of Virus and Microbiological Special Diagnostics, Statens Serum Institut, Copenhagen, Denmark, Artillerivej 5, 2300 Copenahgen S	Albertsen lab, Department of Chemistry and Bioscience, Aalborg University, Denmark	Rasmus Kirkegaard
EPI_ISL_429566	hCoV-19/Denmark/ALAB-SSI489/2020	Europe / Denmark / Unknown	2020-03-23
Department of Virus and Microbiological Special Diagnostics, Statens Serum Institut, Copenhagen, Denmark, Artillerivej 5, 2300 Copenahgen S	Albertsen lab, Department of Chemistry and Bioscience, Aalborg University, Denmark	Rasmus Kirkegaard
EPI_ISL_429567	hCoV-19/Denmark/ALAB-SSI490/2020	Europe / Denmark / Unknown	2020-03-21
Department of Virus and Microbiological Special Diagnostics, Statens Serum Institut, Copenhagen, Denmark, Artillerivej 5, 2300 Copenahgen S	Albertsen lab, Department of Chemistry and Bioscience, Aalborg University, Denmark	Rasmus Kirkegaard
EPI_ISL_429569	hCoV-19/Denmark/ALAB-SSI494/2020	Europe / Denmark / Unknown	2020-03-24
Department of Virus and Microbiological Special Diagnostics, Statens Serum Institut, Copenhagen, Denmark,

Artillerivej 5, 2300 Copenahgen S	Albertsen lab, Department of Chemistry and Bioscience, Aalborg University, Denmark	Rasmus Kirkegaard
EPI_ISL_429570	hCoV-19/Denmark/ALAB-SSI495/2020	Europe / Denmark / Unknown	2020-03-24	Department of Virus and Microbiological Special Diagnostics, Statens Serum Institut, Copenhagen, Denmark, Artillerivej 5, 2300 Copenahgen S	Albertsen lab, Department of Chemistry and Bioscience, Aalborg University, Denmark	Rasmus Kirkegaard
EPI_ISL_429571	hCoV-19/Denmark/ALAB-SSI497/2020	Europe / Denmark / Unknown	2020-03-26	Department of Virus and Microbiological Special Diagnostics, Statens Serum Institut, Copenhagen, Denmark, Artillerivej 5, 2300 Copenahgen S	Albertsen lab, Department of Chemistry and Bioscience, Aalborg University, Denmark	Rasmus Kirkegaard
EPI_ISL_429572	hCoV-19/Denmark/ALAB-SSI501/2020	Europe / Denmark / Unknown	2020-03-24	Department of Virus and Microbiological Special Diagnostics, Statens Serum Institut, Copenhagen, Denmark, Artillerivej 5, 2300 Copenahgen S	Albertsen lab, Department of Chemistry and Bioscience, Aalborg University, Denmark	Rasmus Kirkegaard
EPI_ISL_429573	hCoV-19/Denmark/ALAB-SSI502/2020	Europe / Denmark / Unknown	2020-03-23	Department of Virus and Microbiological Special Diagnostics, Statens Serum Institut, Copenhagen, Denmark, Artillerivej 5, 2300 Copenahgen S	Albertsen lab, Department of Chemistry and Bioscience, Aalborg University, Denmark	Rasmus Kirkegaard
EPI_ISL_429574	hCoV-19/Denmark/ALAB-SSI506/2020	Europe / Denmark / Unknown	2020-03-26	Department of Virus and Microbiological Special Diagnostics, Statens Serum Institut, Copenhagen, Denmark, Artillerivej 5, 2300 Copenahgen S	Albertsen lab, Department of Chemistry and Bioscience, Aalborg University, Denmark	Rasmus Kirkegaard
EPI_ISL_429575	hCoV-19/Denmark/ALAB-SSI507/2020	Europe / Denmark / Unknown	2020-03-25	Department of Virus and Microbiological Special Diagnostics, Statens Serum Institut, Copenhagen, Denmark, Artillerivej 5, 2300 Copenahgen S	Albertsen lab, Department of Chemistry and Bioscience, Aalborg University, Denmark	Rasmus Kirkegaard
EPI_ISL_429577	hCoV-19/Denmark/ALAB-SSI509/2020	Europe / Denmark / Unknown	2020-03-25	Department of Virus and Microbiological Special Diagnostics, Statens Serum Institut, Copenhagen, Denmark, Artillerivej 5, 2300 Copenahgen S	Albertsen lab, Department of Chemistry and Bioscience, Aalborg University, Denmark	Rasmus Kirkegaard
EPI_ISL_429578	hCoV-19/Denmark/ALAB-SSI510/2020	Europe / Denmark / Unknown	2020-03-25	Department of Virus and Microbiological Special Diagnostics, Statens Serum Institut, Copenhagen, Denmark, Artillerivej 5, 2300 Copenahgen S	Albertsen lab, Department of Chemistry and Bioscience, Aalborg University, Denmark	Rasmus Kirkegaard
EPI_ISL_429579	hCoV-19/Denmark/ALAB-SSI511/2020	Europe / Denmark / Unknown	2020-03-26	Department of Virus and Microbiological Special Diagnostics, Statens Serum Institut, Copenhagen, Denmark, Artillerivej 5, 2300 Copenahgen S	Albertsen lab, Department of Chemistry and Bioscience, Aalborg University, Denmark	Rasmus Kirkegaard
EPI_ISL_429580	hCoV-19/Denmark/ALAB-SSI512/2020	Europe / Denmark / Unknown	2020-03-25	Department of Virus and Microbiological Special Diagnostics, Statens Serum Institut, Copenhagen, Denmark, Artillerivej 5, 2300 Copenahgen S	Albertsen lab, Department of Chemistry and Bioscience, Aalborg University, Denmark	Rasmus Kirkegaard
EPI_ISL_429581	hCoV-19/Denmark/ALAB-SSI513/2020	Europe / Denmark / Unknown	2020-03-24	Department of Virus and Microbiological Special Diagnostics, Statens Serum Institut, Copenhagen, Denmark, Artillerivej 5, 2300 Copenahgen S	Albertsen lab, Department of Chemistry and Bioscience, Aalborg University, Denmark	Rasmus Kirkegaard
EPI_ISL_429582	hCoV-19/Denmark/ALAB-SSI514/2020	Europe / Denmark / Unknown	2020-03-16	Department of Virus and Microbiological Special Diagnostics, Statens Serum Institut, Copenhagen, Denmark, Artillerivej 5, 2300 Copenahgen S	Albertsen lab, Department of Chemistry and Bioscience, Aalborg University, Denmark	Rasmus Kirkegaard
EPI_ISL_429583	hCoV-19/Denmark/ALAB-SSI515/2020	Europe / Denmark / Unknown	2020-03-23	Department of Virus and Microbiological Special Diagnostics, Statens Serum Institut, Copenhagen, Denmark, Artillerivej 5, 2300 Copenahgen S	Albertsen lab, Department of Chemistry and Bioscience, Aalborg University, Denmark	Rasmus Kirkegaard
EPI_ISL_429584	hCoV-19/Denmark/ALAB-SSI518/2020	Europe / Denmark / Unknown	2020-03-26	Department of Virus and Microbiological Special Diagnostics, Statens Serum Institut, Copenhagen, Denmark, Artillerivej 5, 2300 Copenahgen S	Albertsen lab, Department of Chemistry and Bioscience, Aalborg University, Denmark	Rasmus Kirkegaard
EPI_ISL_429585	hCoV-19/Denmark/ALAB-SSI520/2020	Europe / Denmark / Unknown	2020-03-26	Department of Virus and Microbiological Special Diagnostics, Statens Serum Institut, Copenhagen, Denmark, Artillerivej 5, 2300 Copenahgen S	Albertsen lab, Department of Chemistry and Bioscience, Aalborg University, Denmark	Rasmus Kirkegaard
EPI_ISL_429586	hCoV-19/Denmark/ALAB-SSI521/2020	Europe / Denmark / Unknown	2020-03-26	Department of Virus and Microbiological Special Diagnostics, Statens Serum Institut, Copenhagen, Denmark, Artillerivej 5, 2300 Copenahgen S	Albertsen lab, Department of Chemistry and Bioscience, Aalborg University, Denmark	Rasmus Kirkegaard
EPI_ISL_429587	hCoV-19/Denmark/ALAB-SSI522/2020	Europe / Denmark / Unknown	2020-03-26	Department of Virus and Microbiological Special Diagnostics, Statens Serum Institut, Copenhagen, Denmark, Artillerivej 5, 2300 Copenahgen S	Albertsen lab, Department of Chemistry and Bioscience, Aalborg University, Denmark	Rasmus Kirkegaard

EPI_ISL_429588 hCoV-19/Denmark/ALAB-SSI523/2020 Europe / Denmark / Unknown 2020-03-26 Department of Virus and Microbiological Special Diagnostics, Statens Serum Institut, Copenhagen, Denmark, Artillerivej 5, 2300 Copenahgen S Albertsen lab, Department of Chemistry and Bioscience, Aalborg University, Denmark Rasmus Kirkegaard
EPI_ISL_429589 hCoV-19/Denmark/ALAB-SSI592/2020 Europe / Denmark / Unknown 2020-03-28 Department of Virus and Microbiological Special Diagnostics, Statens Serum Institut, Copenhagen, Denmark, Artillerivej 5, 2300 Copenahgen S Albertsen lab, Department of Chemistry and Bioscience, Aalborg University, Denmark Rasmus Kirkegaard
EPI_ISL_429590 hCoV-19/Denmark/ALAB-SSI595/2020 Europe / Denmark / Unknown 2020-03-28 Department of Virus and Microbiological Special Diagnostics, Statens Serum Institut, Copenhagen, Denmark, Artillerivej 5, 2300 Copenahgen S Albertsen lab, Department of Chemistry and Bioscience, Aalborg University, Denmark Rasmus Kirkegaard
EPI_ISL_429598 hCoV-19/USA/WA-UW-4546/2020 North America / USA / Washington 2020-03-31 UW Virology Lab UW Virology Lab Pavitra Roychoudhury, Hong Xie, Keith Jerome, Alexander Greninger
EPI_ISL_430000 hCoV-19/Jordan/SR-042/2020 Asia / Jordan / Amman 2020-03-30 Biolab Diagnostic Laboratories Andersen lab at Scripps Research Issa Abu-Dayyeh, Ahmad Tibi, Lama Hussein, Lina Mohammad, Zein Naber, Amid Abdelnour with SEARCH Alliance San Diego
EPI_ISL_430001 hCoV-19/Jordan/SR-043/2020 Asia / Jordan / Amman 2020-03-17 Biolab Diagnostic Laboratories Andersen lab at Scripps Research Issa Abu-Dayyeh, Ahmad Tibi, Lama Hussein, Lina Mohammad, Zein Naber, Amid Abdelnour with SEARCH Alliance San Diego
EPI_ISL_430002 hCoV-19/Jordan/SR-044/2020 Asia / Jordan / Amman 2020-03-16 Biolab Diagnostic Laboratories Andersen lab at Scripps Research Issa Abu-Dayyeh, Ahmad Tibi, Lama Hussein, Lina Mohammad, Zein Naber, Amid Abdelnour with SEARCH Alliance San Diego
EPI_ISL_430003 hCoV-19/Jordan/SR-045/2020 Asia / Jordan / Amman 2020-03-16 Biolab Diagnostic Laboratories Andersen lab at Scripps Research Issa Abu-Dayyeh, Ahmad Tibi, Lama Hussein, Lina Mohammad, Zein Naber, Amid Abdelnour with SEARCH Alliance San Diego
EPI_ISL_430004 hCoV-19/Jordan/SR-046/2020 Asia / Jordan / Amman 2020-03-17 Biolab Diagnostic Laboratories Andersen lab at Scripps Research Issa Abu-Dayyeh, Ahmad Tibi, Lama Hussein, Lina Mohammad, Zein Naber, Amid Abdelnour with SEARCH Alliance San Diego
EPI_ISL_430006 hCoV-19/Jordan/SR-048/2020 Asia / Jordan / Amman 2020-03-23 Biolab Diagnostic Laboratories Andersen lab at Scripps Research Issa Abu-Dayyeh, Ahmad Tibi, Lama Hussein, Lina Mohammad, Zein Naber, Amid Abdelnour with SEARCH Alliance San Diego
EPI_ISL_430007 hCoV-19/Jordan/SR-049/2020 Asia / Jordan / Amman 2020-03-19 Biolab Diagnostic Laboratories Andersen lab at Scripps Research Issa Abu-Dayyeh, Ahmad Tibi, Lama Hussein, Lina Mohammad, Zein Naber, Amid Abdelnour with SEARCH Alliance San Diego
EPI_ISL_430009 hCoV-19/Jordan/SR-052/2020 Asia / Jordan / Irbid 2020-03-30 Biolab Diagnostic Laboratories Andersen lab at Scripps Research Issa Abu-Dayyeh, Ahmad Tibi, Lama Hussein, Lina Mohammad, Zein Naber, Amid Abdelnour with SEARCH Alliance San Diego
EPI_ISL_430011 hCoV-19/Jordan/SR-053/2020 Asia / Jordan / Amman 2020-03-19 Biolab Diagnostic Laboratories Andersen lab at Scripps Research Issa Abu-Dayyeh, Ahmad Tibi, Lama Hussein, Lina Mohammad, Zein Naber, Amid Abdelnour with SEARCH Alliance San Diego
EPI_ISL_430012 hCoV-19/Jordan/SR-054/2020 Asia / Jordan / Irbid 2020-03-30 Biolab Diagnostic Laboratories Andersen lab at Scripps Research Issa Abu-Dayyeh, Ahmad Tibi, Lama Hussein, Lina Mohammad, Zein Naber, Amid Abdelnour with SEARCH Alliance San Diego
EPI_ISL_430014 hCoV-19/Jordan/SR-056/2020 Asia / Jordan / Amman 2020-03-17 Biolab Diagnostic Laboratories Andersen lab at Scripps Research Issa Abu-Dayyeh, Ahmad Tibi, Lama Hussein, Lina Mohammad, Zein Naber, Amid Abdelnour with SEARCH Alliance San Diego
EPI_ISL_430015 hCoV-19/Jordan/SR-057/2020 Asia / Jordan / Amman 2020-03-24 Biolab Diagnostic Laboratories Andersen lab at Scripps Research Issa Abu-Dayyeh, Ahmad Tibi, Lama Hussein, Lina Mohammad, Zein Naber, Amid Abdelnour with SEARCH Alliance San Diego
EPI_ISL_430016 hCoV-19/USA/CA-SR016/2020 North America / USA / California / San Diego 2020-03-24 Andersen lab at Scripps Research Andersen lab at Scripps Research SEARCH Alliance San Diego
EPI_ISL_430039 hCoV-19/USA/UT-0370/2020 North America / USA / Utah 2020-03-30 Utah Public Health Laboratory Utah Public Health Laboratory Erin Young, Kelly Oakeson
EPI_ISL_430041 hCoV-19/USA/UT-0443/2020 North America / USA / Utah 2020-03-30 Utah Public Health Laboratory Utah Public Health Laboratory Erin Young, Kelly Oakeson
EPI_ISL_430043 hCoV-19/USA/UT-0445/2020 North America / USA / Utah 2020-03-30 Utah Public Health Laboratory Utah Public Health Laboratory Erin Young, Kelly Oakeson
EPI_ISL_430044 hCoV-19/USA/UT-0446/2020 North America / USA / Utah 2020-03-30 Utah Public Health Laboratory Utah Public Health Laboratory Erin Young, Kelly Oakeson
EPI_ISL_430045 hCoV-19/USA/UT-0447/2020 North America / USA / Utah 2020-03-31 Utah Public Health Laboratory Utah Public Health Laboratory Erin Young, Kelly Oakeson
EPI_ISL_430047 hCoV-19/USA/UT-0492/2020 North America / USA / Utah 2020-04-02 Utah Public Health Laboratory Utah Public Health Laboratory Erin Young, Kelly Oakeson
EPI_ISL_430050 hCoV-19/USA/UT-0500/2020 North America / USA / Utah 2020-03-31 Utah Public Health Laboratory Utah Public Health Laboratory Erin Young, Kelly Oakeson
EPI_ISL_430051 hCoV-19/USA/UT-0501/2020 North America / USA / Utah 2020-04-02 Utah Public Health Laboratory Utah Public Health Laboratory Erin Young, Kelly Oakeson
EPI_ISL_430053 hCoV-19/USA/UT-0503/2020 North America / USA / Utah 2020-04-02 Utah Public Health Laboratory Utah Public Health Laboratory Erin Young, Kelly Oakeson

EPI_ISL_430055 hCoV-19/USA/UT-0507/2020 North America / USA / Utah 2020-04-02 Utah Public Health Laboratory Utah Public Health Laboratory Erin Young, Kelly Oakeson
EPI_ISL_430057 hCoV-19/USA/UT-0510/2020 North America / USA / Utah 2020-04-03 Utah Public Health Laboratory Utah Public Health Laboratory Erin Young, Kelly Oakeson
EPI_ISL_430500 hCoV-19/Australia/VIC1210/2020 Oceania / Australia / Victoria 2020-04-01 Victorian Infectious Diseases Reference Laboratory (VIDRL) Microbiological Diagnostic Unit Public Health Laboratory and Victorian Infectious Diseases Reference Laboratory, The Peter Doherty Institute for Infection and Immunity Caly L., Seemann T., Sait, M., Schultz M., Druce J., Sherry, N.
EPI_ISL_430512 hCoV-19/Australia/VIC1235/2020 Oceania / Australia / Victoria 2020-04-02 Victorian Infectious Diseases Reference Laboratory (VIDRL) Microbiological Diagnostic Unit Public Health Laboratory and Victorian Infectious Diseases Reference Laboratory, The Peter Doherty Institute for Infection and Immunity Caly L., Seemann T., Sait, M., Schultz M., Druce J., Sherry, N.
EPI_ISL_430519 hCoV-19/Australia/VIC1222/2020 Oceania / Australia / Victoria 2020-04-01 Victorian Infectious Diseases Reference Laboratory (VIDRL) Microbiological Diagnostic Unit Public Health Laboratory and Victorian Infectious Diseases Reference Laboratory, The Peter Doherty Institute for Infection and Immunity Caly L., Seemann T., Sait, M., Schultz M., Druce J., Sherry, N.
EPI_ISL_430522 hCoV-19/Australia/VIC1224/2020 Oceania / Australia / Victoria 2020-04-01 Victorian Infectious Diseases Reference Laboratory (VIDRL) Microbiological Diagnostic Unit Public Health Laboratory and Victorian Infectious Diseases Reference Laboratory, The Peter Doherty Institute for Infection and Immunity Caly L., Seemann T., Sait, M., Schultz M., Druce J., Sherry, N.
EPI_ISL_430528 hCoV-19/Australia/VIC1192/2020 Oceania / Australia / Victoria 2020-03-30 Victorian Infectious Diseases Reference Laboratory (VIDRL) Microbiological Diagnostic Unit Public Health Laboratory and Victorian Infectious Diseases Reference Laboratory, The Peter Doherty Institute for Infection and Immunity Caly L., Seemann T., Sait, M., Schultz M., Druce J., Sherry, N.
EPI_ISL_430532 hCoV-19/Australia/VIC1025/2020 Oceania / Australia / Victoria 2020-04-02 Victorian Infectious Diseases Reference Laboratory (VIDRL) Microbiological Diagnostic Unit Public Health Laboratory and Victorian Infectious Diseases Reference Laboratory, The Peter Doherty Institute for Infection and Immunity Caly L., Seemann T., Sait, M., Schultz M., Druce J., Sherry, N.
EPI_ISL_430533 hCoV-19/Australia/VIC1037/2020 Oceania / Australia / Victoria 2020-04-03 Victorian Infectious Diseases Reference Laboratory (VIDRL) Microbiological Diagnostic Unit Public Health Laboratory and Victorian Infectious Diseases Reference Laboratory, The Peter Doherty Institute for Infection and Immunity Caly L., Seemann T., Sait, M., Schultz M., Druce J., Sherry, N.
EPI_ISL_430535 hCoV-19/Australia/VIC1039/2020 Oceania / Australia / Victoria 2020-04-03 Victorian Infectious Diseases Reference Laboratory (VIDRL) Microbiological Diagnostic Unit Public Health Laboratory and Victorian Infectious Diseases Reference Laboratory, The Peter Doherty Institute for Infection and Immunity Caly L., Seemann T., Sait, M., Schultz M., Druce J., Sherry, N.
EPI_ISL_430536 hCoV-19/Australia/VIC1041/2020 Oceania / Australia / Victoria 2020-04-03 Victorian Infectious Diseases Reference Laboratory (VIDRL) Microbiological Diagnostic Unit Public Health Laboratory and Victorian Infectious Diseases Reference Laboratory, The Peter Doherty Institute for Infection and Immunity Caly L., Seemann T., Sait, M., Schultz M., Druce J., Sherry, N.
EPI_ISL_430538 hCoV-19/Australia/VIC1042/2020 Oceania / Australia / Victoria 2020-04-03 Victorian Infectious Diseases Reference Laboratory (VIDRL) Microbiological Diagnostic Unit Public Health Laboratory and Victorian Infectious Diseases Reference Laboratory, The Peter Doherty Institute for Infection and Immunity Caly L., Seemann T., Sait, M., Schultz M., Druce J., Sherry, N.
EPI_ISL_430539 hCoV-19/Australia/VIC1043/2020 Oceania / Australia / Victoria 2020-04-03 Victorian Infectious Diseases Reference Laboratory (VIDRL) Microbiological Diagnostic Unit Public Health Laboratory and Victorian Infectious Diseases Reference Laboratory, The Peter Doherty Institute for Infection and Immunity Caly L., Seemann T., Sait, M., Schultz M., Druce J., Sherry, N.
EPI_ISL_430542 hCoV-19/Australia/VIC978/2020 Oceania / Australia / Victoria 2020-03-29 Victorian Infectious Diseases Reference Laboratory (VIDRL) Microbiological Diagnostic Unit Public Health Laboratory and Victorian Infectious Diseases Reference Laboratory, The Peter Doherty Institute for Infection and Immunity Caly L., Seemann T., Sait, M., Schultz M., Druce J., Sherry, N.
EPI_ISL_430543 hCoV-19/Australia/VIC982/2020 Oceania / Australia / Victoria 2020-03-30 Victorian Infectious Diseases Reference Laboratory (VIDRL) Microbiological Diagnostic Unit Public Health Laboratory and Victorian Infectious Diseases Reference Laboratory, The Peter Doherty Institute for Infection and Immunity Caly L., Seemann T., Sait, M., Schultz M., Druce J., Sherry, N.
EPI_ISL_430545 hCoV-19/Australia/VIC983/2020 Oceania / Australia / Victoria 2020-03-30 Victorian Infectious Diseases Reference Laboratory (VIDRL) Microbiological Diagnostic Unit Public Health Laboratory and Victorian Infectious Diseases Reference Laboratory, The Peter Doherty Institute for Infection and Immunity Caly L., Seemann T., Sait, M., Schultz M., Druce J., Sherry, N.
EPI_ISL_430547 hCoV-19/Australia/VIC1011/2020 Oceania / Australia / Victoria 2020-04-01 Victorian Infectious Diseases Reference Laboratory (VIDRL) Microbiological Diagnostic Unit Public Health Laboratory and Victorian Infectious Diseases Reference Laboratory, The Peter Doherty Institute for Infection and Immunity Caly L., Seemann T., Sait, M., Schultz M., Druce J., Sherry, N.
EPI_ISL_430550 hCoV-19/Australia/VIC1046/2020 Oceania / Australia / Victoria 2020-04-03 Victorian Infectious Diseases Reference Laboratory (VIDRL) Microbiological Diagnostic Unit Public Health Laboratory and Victorian Infectious Diseases Reference Laboratory, The Peter Doherty Institute for Infection and Immunity Caly L., Seemann T., Sait, M., Schultz M., Druce J., Sherry, N.
EPI_ISL_430551 hCoV-19/Australia/VIC989/2020 Oceania / Australia / Victoria 2020-03-31 Victorian Infectious Diseases Reference Laboratory (VIDRL) Microbiological Diagnostic Unit Public Health

Laboratory and Victorian Infectious Diseases Reference Laboratory, The Peter Doherty Institute for Infection and Immunity	Caly L., Seemann T., Sait, M., Schultz M., Druce J., Sherry, N.
EPI_ISL_430552	hCoV-19/Australia/VIC984/2020	Oceania / Australia / Victoria	2020-03-30	Victorian Infectious Diseases Reference Laboratory (VIDRL)	Microbiological Diagnostic Unit Public Health Laboratory and Victorian Infectious Diseases Reference Laboratory, The Peter Doherty Institute for Infection and Immunity	Caly L., Seemann T., Sait, M., Schultz M., Druce J., Sherry, N.
EPI_ISL_430553	hCoV-19/Australia/VIC1013/2020	Oceania / Australia / Victoria	2020-04-01	Victorian Infectious Diseases Reference Laboratory (VIDRL)	Microbiological Diagnostic Unit Public Health Laboratory and Victorian Infectious Diseases Reference Laboratory, The Peter Doherty Institute for Infection and Immunity	Caly L., Seemann T., Sait, M., Schultz M., Druce J., Sherry, N.
EPI_ISL_430556	hCoV-19/Australia/VIC990/2020	Oceania / Australia / Victoria	2020-03-31	Victorian Infectious Diseases Reference Laboratory (VIDRL)	Microbiological Diagnostic Unit Public Health Laboratory and Victorian Infectious Diseases Reference Laboratory, The Peter Doherty Institute for Infection and Immunity	Caly L., Seemann T., Sait, M., Schultz M., Druce J., Sherry, N.
EPI_ISL_430559	hCoV-19/Australia/VIC1015/2020	Oceania / Australia / Victoria	2020-04-01	Victorian Infectious Diseases Reference Laboratory (VIDRL)	Microbiological Diagnostic Unit Public Health Laboratory and Victorian Infectious Diseases Reference Laboratory, The Peter Doherty Institute for Infection and Immunity	Caly L., Seemann T., Sait, M., Schultz M., Druce J., Sherry, N.
EPI_ISL_430567	hCoV-19/Australia/VIC1119/2020	Oceania / Australia / Victoria	2020-04-08	Victorian Infectious Diseases Reference Laboratory (VIDRL)	Microbiological Diagnostic Unit Public Health Laboratory and Victorian Infectious Diseases Reference Laboratory, The Peter Doherty Institute for Infection and Immunity	Caly L., Seemann T., Sait, M., Schultz M., Druce J., Sherry, N.
EPI_ISL_430569	hCoV-19/Australia/VIC1110/2020	Oceania / Australia / Victoria	2020-04-07	Victorian Infectious Diseases Reference Laboratory (VIDRL)	Microbiological Diagnostic Unit Public Health Laboratory and Victorian Infectious Diseases Reference Laboratory, The Peter Doherty Institute for Infection and Immunity	Caly L., Seemann T., Sait, M., Schultz M., Druce J., Sherry, N.
EPI_ISL_430573	hCoV-19/Australia/VIC1112/2020	Oceania / Australia / Victoria	2020-04-07	Victorian Infectious Diseases Reference Laboratory (VIDRL)	Microbiological Diagnostic Unit Public Health Laboratory and Victorian Infectious Diseases Reference Laboratory, The Peter Doherty Institute for Infection and Immunity	Caly L., Seemann T., Sait, M., Schultz M., Druce J., Sherry, N.
EPI_ISL_430574	hCoV-19/Australia/VIC1087/2020	Oceania / Australia / Victoria	2020-04-06	Victorian Infectious Diseases Reference Laboratory (VIDRL)	Microbiological Diagnostic Unit Public Health Laboratory and Victorian Infectious Diseases Reference Laboratory, The Peter Doherty Institute for Infection and Immunity	Caly L., Seemann T., Sait, M., Schultz M., Druce J., Sherry, N.
EPI_ISL_430576	hCoV-19/Australia/VIC1114/2020	Oceania / Australia / Victoria	2020-04-07	Victorian Infectious Diseases Reference Laboratory (VIDRL)	Microbiological Diagnostic Unit Public Health Laboratory and Victorian Infectious Diseases Reference Laboratory, The Peter Doherty Institute for Infection and Immunity	Caly L., Seemann T., Sait, M., Schultz M., Druce J., Sherry, N.
EPI_ISL_430577	hCoV-19/Australia/VIC1115/2020	Oceania / Australia / Victoria	2020-04-07	Victorian Infectious Diseases Reference Laboratory (VIDRL)	Microbiological Diagnostic Unit Public Health Laboratory and Victorian Infectious Diseases Reference Laboratory, The Peter Doherty Institute for Infection and Immunity	Caly L., Seemann T., Sait, M., Schultz M., Druce J., Sherry, N.
EPI_ISL_430578	hCoV-19/Australia/VIC1062/2020	Oceania / Australia / Victoria	2020-04-04	Victorian Infectious Diseases Reference Laboratory (VIDRL)	Microbiological Diagnostic Unit Public Health Laboratory and Victorian Infectious Diseases Reference Laboratory, The Peter Doherty Institute for Infection and Immunity	Caly L., Seemann T., Sait, M., Schultz M., Druce J., Sherry, N.
EPI_ISL_430579	hCoV-19/Australia/VIC1080/2020	Oceania / Australia / Victoria	2020-04-06	Victorian Infectious Diseases Reference Laboratory (VIDRL)	Microbiological Diagnostic Unit Public Health Laboratory and Victorian Infectious Diseases Reference Laboratory, The Peter Doherty Institute for Infection and Immunity	Caly L., Seemann T., Sait, M., Schultz M., Druce J., Sherry, N.
EPI_ISL_430580	hCoV-19/Australia/VIC1081/2020	Oceania / Australia / Victoria	2020-04-06	Victorian Infectious Diseases Reference Laboratory (VIDRL)	Microbiological Diagnostic Unit Public Health Laboratory and Victorian Infectious Diseases Reference Laboratory, The Peter Doherty Institute for Infection and Immunity	Caly L., Seemann T., Sait, M., Schultz M., Druce J., Sherry, N.
EPI_ISL_430581	hCoV-19/Australia/VIC1071/2020	Oceania / Australia / Victoria	2020-04-05	Victorian Infectious Diseases Reference Laboratory (VIDRL)	Microbiological Diagnostic Unit Public Health Laboratory and Victorian Infectious Diseases Reference Laboratory, The Peter Doherty Institute for Infection and Immunity	Caly L., Seemann T., Sait, M., Schultz M., Druce J., Sherry, N.
EPI_ISL_430583	hCoV-19/Australia/VIC1094/2020	Oceania / Australia / Victoria	2020-04-07	Victorian Infectious Diseases Reference Laboratory (VIDRL)	Microbiological Diagnostic Unit Public Health Laboratory and Victorian Infectious Diseases Reference Laboratory, The Peter Doherty Institute for Infection and Immunity	Caly L., Seemann T., Sait, M., Schultz M., Druce J., Sherry, N.
EPI_ISL_430584	hCoV-19/Australia/VIC1103/2020	Oceania / Australia / Victoria	2020-04-07	Victorian Infectious Diseases Reference Laboratory (VIDRL)	Microbiological Diagnostic Unit Public Health Laboratory and Victorian Infectious Diseases Reference Laboratory, The Peter Doherty Institute for Infection and Immunity	Caly L., Seemann T., Sait, M., Schultz M., Druce J., Sherry, N.
EPI_ISL_430590	hCoV-19/Australia/VIC1107/2020	Oceania / Australia / Victoria	2020-04-07	Victorian Infectious Diseases Reference Laboratory (VIDRL)	Microbiological Diagnostic Unit Public Health Laboratory and Victorian Infectious Diseases Reference Laboratory, The Peter Doherty Institute for Infection and Immunity	Caly L., Seemann T., Sait, M., Schultz M., Druce J., Sherry, N.

EPI_ISL_430596 hCoV-19/Australia/VIC1133/2020 Oceania / Australia / Victoria 2020-04-08 Victorian Infectious Diseases Reference Laboratory (VIDRL) Microbiological Diagnostic Unit Public Health Laboratory and Victorian Infectious Diseases Reference Laboratory, The Peter Doherty Institute for Infection and Immunity Caly L., Seemann T., Sait, M., Schultz M., Druce J., Sherry, N.
EPI_ISL_430597 hCoV-19/Australia/VIC1086/2020 Oceania / Australia / Victoria 2020-04-06 Victorian Infectious Diseases Reference Laboratory (VIDRL) Microbiological Diagnostic Unit Public Health Laboratory and Victorian Infectious Diseases Reference Laboratory, The Peter Doherty Institute for Infection and Immunity Caly L., Seemann T., Sait, M., Schultz M., Druce J., Sherry, N.
EPI_ISL_430598 hCoV-19/Australia/VIC1157/2020 Oceania / Australia / Victoria 2020-04-09 Victorian Infectious Diseases Reference Laboratory (VIDRL) Microbiological Diagnostic Unit Public Health Laboratory and Victorian Infectious Diseases Reference Laboratory, The Peter Doherty Institute for Infection and Immunity Caly L., Seemann T., Sait, M., Schultz M., Druce J., Sherry, N.
EPI_ISL_450404 hCoV-19/Hong Kong/VM20009579/2020 Asia / Hong Kong 2020-03-16 unknown School of Public Health Sit,T.H.S., Brackman,C.J., Sims,L.D., Tsang,D.N.C., Chu,D.K.W., Perera,R.A.P.M., Poon,L.L.M. and Peiris,M.
EPI_ISL_450444 hCoV-19/Beijing/IME-HZ01/2020 Asia / China / Beijing 2020-01-22 unknown Dept. OPA Zhang,X.A., Fan,H., Qi,R.Z., Zheng,W., Zheng,K., Gong,J.H., Fang,L.Q. and Liu,W.
EPI_ISL_450489 hCoV-19/Wuhan/YB012504/2020 Asia / China / Wuhan / Hubei 2020-01-25 unknown CAS Key Laboratory of Special Pathogens and Biosafety and Center for Emerging Infectious Diseases Si,H., Zhu,Y., Lin,H., Xie,S., Shi,Z. and Zhou,P.
EPI_ISL_406862 hCoV-19/Germany/BavPat1/2020 Europe / Germany / Bavaria / Munich 2020-01-28 Charité Universitätsmedizin Berlin, Institute of Virology; Institut für Mikrobiologie der Bundeswehr, Munich Charité Universitätsmedizin Berlin, Institute of Virology Victor M Corman, Julia Schneider, Talitha Veith, Barbara Mühlemann, Markus Antwerpen, Christian Drosten, Roman Wölfel
EPI_ISL_406970 hCoV-19/Hangzhou/HZ-1/2020 Asia / China / Zhejiang / Hangzhou 2020-01-20 Hangzhou Center for Disease and Control Microbiology Lab Hangzhou Center for Disease and Control Microbiology Lab Yu Hua, Wang Haoqiu, Li Jun, Yu Xinfeng
EPI_ISL_406973 hCoV-19/Singapore/1/2020 Asia / Singapore 2020-01-23 Singapore General Hospital National Public Health Laboratory Mak, TM; Octavia S; Chavatte JM; Zhou, ZY; Cui, L; Lin, RTP
EPI_ISL_409067 hCoV-19/USA/MA1/2020 North America / USA / Massachusetts 2020-01-29 Massachusetts Department of Public Health Pathogen Discovery, Respiratory Viruses Branch, Division of Viral Diseases, Centers for Dieases Control and Prevention Clinton R. Paden, Jing Zhang, Krista Queen, Yan Li, Ying Tao, Anna Uehara, Xiaoyan Lu, Brian Lynch, Senthil Kumar K. Sakthivel, Brett L. Whitaker, Shifaq Kamili, Lijuan Wang, Janna' R. Murray, Susan I. Gerber, Stephen Lindstrom, Suxiang Tong
EPI_ISL_410044 hCoV-19/USA/CA6/2020 North America / USA / California 2020-01-27 California Department of Public Health Pathogen Discovery, Respiratory Viruses Branch, Division of Viral Diseases, Centers for Dieases Control and Prevention Jing Zhang, Krista Queen, Yan Li, Ying Tao, Anna Uehara, Clinton R. Paden, Xiaoyan Lu, Brian Lynch, Senthil Kumar K. Sakthivel, Brett L. Whitaker, Shifaq Kamili, Lijuan Wang, Janna' R. Murray, Susan I. Gerber, Stephen Lindstrom, Suxiang Tong
EPI_ISL_410045 hCoV-19/USA/IL2/2020 North America / USA / Illinois 2020-01-28 IL Department of Public Health Chicago Laboratory Pathogen Discovery, Respiratory Viruses Branch, Division of Viral Diseases, Centers for Dieases Control and Prevention Yan Li, Jing Zhang, Krista Queen, Ying Tao, Anna Uehara, Clinton R. Paden, Xiaoyan Lu, Brian Lynch, Senthil Kumar K. Sakthivel, Brett L. Whitaker, Shifaq Kamili, Lijuan Wang, Janna' R. Murray, Susan I. Gerber, Stephen Lindstrom, Suxiang Tong
EPI_ISL_412862 hCoV-19/USA/CA9/2020 North America / USA / California / Solano 2020-02-23 California Department of Public Health Pathogen Discovery, Respiratory Viruses Branch, Division of Viral Diseases, Centers for Disease Control and Prevention Krista Queen, Anna Uehara, Jing Zhang, Yan Li, Ying Tao, Clinton R. Paden, Haibin Wang, Shifaq Kamili, Xiaoyan Lu, Brian Lynch, Senthil Kumar K. Sakthivel, Brett L. Whitaker, Lijuan Wang, Janna' R. Murray, Jasmine Padilla, Justin Lee, Susan I. Gerber, Stephen Lindstrom, Suxiang Tong
EPI_ISL_412869 hCoV-19/South Korea/KCDC05/2020 Asia / South Korea / Seoul 2020-01-30 Division of Viral Diseases, Center for Laboratory Control of Infectious Diseases, Korea Centers for Diseases Control and Prevention Division of Viral Diseases, Center for Laboratory Control of Infectious Diseases, Korea Centers for Diseases Control and Prevention Jeong-Min Kim, Yoon-Seok Chung, Namjoo Lee, Mi-Seon Kim, Sang Hee Woo, Hye-Jun Jo, Sehee Park, Heui Man Kim, Myung Guk Han
EPI_ISL_412870 hCoV-19/South Korea/KCDC06/2020 Asia / South Korea / Seoul 2020-01-30 Division of Viral Diseases, Center for Laboratory Control of Infectious Diseases, Korea Centers for Diseases Control and Prevention Division of Viral Diseases, Center for Laboratory Control of Infectious Diseases, Korea Centers for Diseases Control and Prevention Jeong-Min Kim, Yoon-Seok Chung, Namjoo Lee, Mi-Seon Kim, Sang Hee Woo, Hye-Jun Jo, Sehee Park, Heui Man Kim, Myung Guk Han
EPI_ISL_412871 hCoV-19/South Korea/KCDC07/2020 Asia / South Korea / Seoul 2020-01-31 Division of Viral Diseases, Center for Laboratory Control of Infectious Diseases, Korea Centers for Diseases Control and Prevention Division of Viral Diseases, Center for Laboratory Control of Infectious Diseases, Korea Centers for Diseases Control and Prevention Jeong-Min Kim, Yoon-Seok Chung, Namjoo Lee, Mi-Seon Kim, Sang Hee Woo, Hye-Jun Jo, Sehee Park, Heui Man Kim, Myung Guk Han
EPI_ISL_412872 hCoV-19/South Korea/KCDC12/2020 Asia / South Korea / Gyeonggi-do 2020-02-01 Division of Viral Diseases, Center for Laboratory Control of Infectious Diseases, Korea Centers for Diseases Control and Prevention Division of Viral Diseases, Center for Laboratory Control of Infectious Diseases, Korea Centers for Diseases Control and Prevention Jeong-Min Kim, Yoon-Seok Chung, Namjoo Lee, Mi-Seon Kim, Sang

Hee Woo, Hye-Jun Jo, Sehee Park, Heui Man Kim, Myung Guk Han
EPI_ISL_412873 hCoV-19/South Korea/KCDC24/2020 Asia / South Korea / Chungcheongnam-do 2020-02-06 Division of Viral Diseases, Center for Laboratory Control of Infectious Diseases, Korea Centers for Diseases Control and Prevention Division of Viral Diseases, Center for Laboratory Control of Infectious Diseases, Korea Centers for Diseases Control and Prevention Jeong-Min Kim, Yoon-Seok Chung, Namjoo Lee, Mi-Seon Kim, Sang Hee Woo, Hye-Jun Jo, Sehee Park, Heui Man Kim, Myung Guk Han
EPI_ISL_413996 hCoV-19/Switzerland/TI9486/2020 Europe / Switzerland / Tessin 2020-02-24 Laboratoire de Virologie, HUG Swiss National Reference Centre for Influenza LAUBSCHER Florian et al.
EPI_ISL_413997 hCoV-19/Switzerland/GE3895/2020 Europe / Switzerland / Geneva 2020-02-26 Laboratoire de Virologie, HUG Swiss National Reference Centre for Influenza LAUBSCHER Florian et al.
EPI_ISL_413999 hCoV-19/Switzerland/AG0361/2020 Europe / Switzerland / Argovie 2020-02-27 Laboratoire de Virologie, HUG Swiss National Reference Centre for Influenza LAUBSCHER Florian et al.
EPI_ISL_415041 hCoV-19/Wales/PHW04/2020 Europe / United Kingdom / Wales 2020-03-07 Wales Specialist Virology Centre Public Health Wales Microbiology Cardiff Catherine Moore, Joanne Watkins, Sally Corden, Tom Connor
EPI_ISL_416042 hCoV-19/Hangzhou/ZJU-02/2020 Asia / China / Hangzhou 2020-01-26 State Key Laboratory for Diagnosis and Treatment of Infectious Diseases, National Clinical Research Center for Infectious Diseases, First Affiliated Hospital, Zhejiang University School of Medicine, Hangzhou, China. 310003 State Key Laboratory for Diagnosis and Treatment of Infectious Diseases, National Clinical Research Center for Infectious Diseases, First Affiliated Hospital, Zhejiang University School of Medicine, Hangzhou, China. 310003 Hangping Yao, Nanping Wu, Chao Jiang, Xiangyun Lu, Linfang Cheng, Fumin Liu, Zhigang Wu, Haibo Wu, Changzhong Jin, Min Zheng, Lanjuan Li
EPI_ISL_416044 hCoV-19/Hangzhou/ZJU-03/2020 Asia / China / Hangzhou 2020-01-25 State Key Laboratory for Diagnosis and Treatment of Infectious Diseases, National Clinical Research Center for Infectious Diseases, First Affiliated Hospital, Zhejiang University School of Medicine, Hangzhou, China 310003 State Key Laboratory for Diagnosis and Treatment of Infectious Diseases, National Clinical Research Center for Infectious Diseases, First Affiliated Hospital, Zhejiang University School of Medicine, Hangzhou, China 310003 Hangping Yao, Nanping Wu, Chao Jiang, Xiangyun Lu, Linfang Cheng, Fumin Liu, Zhigang Wu, Haibo Wu, Changzhong Jin, Min Zheng, Lanjuan Li
EPI_ISL_416046 hCoV-19/Hangzhou/ZJU-04/2020 Asia / China / Hangzhou 2020-01-24 State Key Laboratory for Diagnosis and Treatment of Infectious Diseases, National Clinical Research Center for Infectious Diseases, First Affiliated Hospital, Zhejiang University School of Medicine, Hangzhou, China 310003 State Key Laboratory for Diagnosis and Treatment of Infectious Diseases, National Clinical Research Center for Infectious Diseases, First Affiliated Hospital, Zhejiang University School of Medicine, Hangzhou, China 310003 Hangping Yao, Nanping Wu, Chao Jiang, Xiangyun Lu, Linfang Cheng, Fumin Liu, Zhigang Wu, Haibo Wu, Changzhong Jin, Min Zheng, Lanjuan Li
EPI_ISL_416047 hCoV-19/Hangzhou/ZJU-06/2020 Asia / China / Hangzhou 2020-02-02 State Key Laboratory for Diagnosis and Treatment of Infectious Diseases, National Clinical Research Center for Infectious Diseases, First Affiliated Hospital, Zhejiang University School of Medicine, Hangzhou, China 310003 State Key Laboratory for Diagnosis and Treatment of Infectious Diseases, National Clinical Research Center for Infectious Diseases, First Affiliated Hospital, Zhejiang University School of Medicine, Hangzhou, China 310003 Hangping Yao, Nanping Wu, Chao Jiang, Xiangyun Lu, Linfang Cheng, Fumin Liu, Zhigang Wu, Haibo Wu, Changzhong Jin, Min Zheng, Lanjuan Li
EPI_ISL_416314 hCoV-19/Hong Kong/CUHK1/2020 Asia / Hong Kong 2020-02-07 Department of Microbiology, Faculty of Medicine, The Chinese University of Hong Kong, Hong Kong SAR, China Department of Microbiology, Faculty of Medicine, Chinese University of Hong Kong, Hong Kong SAR, China Zigui Chen, Paul KS Chan
EPI_ISL_416316 hCoV-19/Shanghai/SH0002/2020 Asia / China / Shanghai 2020-01-25 Shanghai Public Health Clinical Center, Shanghai Medical College, Fudan University National Research Center for Translational Medicine (Shanghai), Ruijin Hospital affiliated to Shanghai Jiao Tong University School of Medicine & Shanghai Public Health Clinical Center Shengyue Wang, Xiaonan Zhang, Gang Lu, Yun Tan, Yun Ling, Hongzhou Lu, Saijuan Chen
EPI_ISL_416317 hCoV-19/Shanghai/SH0003/2020 Asia / China / Shanghai 2020-01-25 Shanghai Public Health Clinical Center, Shanghai Medical College, Fudan University National Research Center for Translational Medicine (Shanghai), Ruijin Hospital affiliated to Shanghai Jiao Tong University School of Medicine & Shanghai Public Health Clinical Center Shengyue Wang, Xiaonan Zhang, Gang Lu, Yun Tan, Yun Ling, Hongzhou Lu, Saijuan Chen
EPI_ISL_416318 hCoV-19/Shanghai/SH0004/2020 Asia / China / Shanghai 2020-01-28 Shanghai Public Health Clinical Center, Shanghai Medical College, Fudan University National Research Center for Translational Medicine (Shanghai), Ruijin Hospital affiliated to Shanghai Jiao Tong University School of Medicine & Shanghai Public Health Clinical Center Shengyue Wang, Xiaonan Zhang, Gang Lu, Yun Tan, Yun Ling, Hongzhou Lu, Saijuan Chen
EPI_ISL_416319 hCoV-19/Shanghai/SH0005/2020 Asia / China / Shanghai 2020-01-28 Shanghai Public Health Clinical Center, Shanghai Medical College, Fudan University National Research Center for Translational Medicine (Shanghai), Ruijin Hospital affiliated to Shanghai Jiao Tong University School of Medicine & Shanghai Public Health Clinical Center Shengyue Wang, Xiaonan Zhang, Gang Lu, Yun Tan, Yun Ling, Hongzhou Lu, Saijuan Chen
EPI_ISL_416320 hCoV-19/Shanghai/SH0007/2020 Asia / China / Shanghai 2020-01-28 Shanghai Public Health Clinical Center, Shanghai Medical College, Fudan University National Research Center for Translational Medicine (Shanghai), Ruijin Hospital affiliated to Shanghai Jiao Tong University School of Medicine & Shanghai

Public Health Clinical Center	Shengyue Wang, Xiaonan Zhang, Gang Lu, Yun Tan, Yun Ling, Hongzhou Lu, Saijuan Chen
EPI_ISL_416321	hCoV-19/Shanghai/SH0008/2020	Asia / China / Shanghai	2020-01-28	Shanghai Public Health Clinical Center, Shanghai Medical College, Fudan University	National Research Center for Translational Medicine (Shanghai), Ruijin Hospital affiliated to Shanghai Jiao Tong University School of Medicine & Shanghai Public Health Clinical Center	Shengyue Wang, Xiaonan Zhang, Gang Lu, Yun Tan, Yun Ling, Hongzhou Lu, Saijuan Chen
EPI_ISL_416322	hCoV-19/Shanghai/SH0009/2020	Asia / China / Shanghai	2020-01-29	Shanghai Public Health Clinical Center, Shanghai Medical College, Fudan University	National Research Center for Translational Medicine (Shanghai), Ruijin Hospital affiliated to Shanghai Jiao Tong University School of Medicine & Shanghai Public Health Clinical Center	Shengyue Wang, Xiaonan Zhang, Gang Lu, Yun Tan, Yun Ling, Hongzhou Lu, Saijuan Chen
EPI_ISL_416323	hCoV-19/Shanghai/SH0010/2020	Asia / China / Shanghai	2020-01-29	Shanghai Public Health Clinical Center, Shanghai Medical College, Fudan University	National Research Center for Translational Medicine (Shanghai), Ruijin Hospital affiliated to Shanghai Jiao Tong University School of Medicine & Shanghai Public Health Clinical Center	Shengyue Wang, Xiaonan Zhang, Gang Lu, Yun Tan, Yun Ling, Hongzhou Lu, Saijuan Chen
EPI_ISL_416324	hCoV-19/Shanghai/SH0011/2020	Asia / China / Shanghai	2020-01-29	Shanghai Public Health Clinical Center, Shanghai Medical College, Fudan University	National Research Center for Translational Medicine (Shanghai), Ruijin Hospital affiliated to Shanghai Jiao Tong University School of Medicine & Shanghai Public Health Clinical Center	Shengyue Wang, Xiaonan Zhang, Gang Lu, Yun Tan, Yun Ling, Hongzhou Lu, Saijuan Chen
EPI_ISL_416325	hCoV-19/Shanghai/SH0012/2020	Asia / China / Shanghai	2020-02-02	Shanghai Public Health Clinical Center, Shanghai Medical College, Fudan University	National Research Center for Translational Medicine (Shanghai), Ruijin Hospital affiliated to Shanghai Jiao Tong University School of Medicine & Shanghai Public Health Clinical Center	Shengyue Wang, Xiaonan Zhang, Gang Lu, Yun Tan, Yun Ling, Hongzhou Lu, Saijuan Chen
EPI_ISL_416326	hCoV-19/Shanghai/SH0013/2020	Asia / China / Shanghai	2020-01-30	Shanghai Public Health Clinical Center, Shanghai Medical College, Fudan University	National Research Center for Translational Medicine (Shanghai), Ruijin Hospital affiliated to Shanghai Jiao Tong University School of Medicine & Shanghai Public Health Clinical Center	Shengyue Wang, Xiaonan Zhang, Gang Lu, Yun Tan, Yun Ling, Hongzhou Lu, Saijuan Chen
EPI_ISL_416329	hCoV-19/Shanghai/SH0020/2020	Asia / China / Shanghai	2020-02-01	Shanghai Public Health Clinical Center, Shanghai Medical College, Fudan University	National Research Center for Translational Medicine (Shanghai), Ruijin Hospital affiliated to Shanghai Jiao Tong University School of Medicine & Shanghai Public Health Clinical Center	Shengyue Wang, Xiaonan Zhang, Gang Lu, Yun Tan, Yun Ling, Hongzhou Lu, Saijuan Chen
EPI_ISL_416330	hCoV-19/Shanghai/SH0021/2020	Asia / China / Shanghai	2020-02-02	Shanghai Public Health Clinical Center, Shanghai Medical College, Fudan University	National Research Center for Translational Medicine (Shanghai), Ruijin Hospital affiliated to Shanghai Jiao Tong University School of Medicine & Shanghai Public Health Clinical Center	Shengyue Wang, Xiaonan Zhang, Gang Lu, Yun Tan, Yun Ling, Hongzhou Lu, Saijuan Chen
EPI_ISL_416331	hCoV-19/Shanghai/SH0022/2020	Asia / China / Shanghai	2020-01-30	Shanghai Public Health Clinical Center, Shanghai Medical College, Fudan University	National Research Center for Translational Medicine (Shanghai), Ruijin Hospital affiliated to Shanghai Jiao Tong University School of Medicine & Shanghai Public Health Clinical Center	Shengyue Wang, Xiaonan Zhang, Gang Lu, Yun Tan, Yun Ling, Hongzhou Lu, Saijuan Chen
EPI_ISL_416332	hCoV-19/Shanghai/SH0023/2020	Asia / China / Shanghai	2020-01-30	Shanghai Public Health Clinical Center, Shanghai Medical College, Fudan University	National Research Center for Translational Medicine (Shanghai), Ruijin Hospital affiliated to Shanghai Jiao Tong University School of Medicine & Shanghai Public Health Clinical Center	Shengyue Wang, Xiaonan Zhang, Gang Lu, Yun Tan, Yun Ling, Hongzhou Lu, Saijuan Chen
EPI_ISL_416333	hCoV-19/Shanghai/SH0024/2020	Asia / China / Shanghai	2020-01-30	Shanghai Public Health Clinical Center, Shanghai Medical College, Fudan University	National Research Center for Translational Medicine (Shanghai), Ruijin Hospital affiliated to Shanghai Jiao Tong University School of Medicine & Shanghai Public Health Clinical Center	Shengyue Wang, Xiaonan Zhang, Gang Lu, Yun Tan, Yun Ling, Hongzhou Lu, Saijuan Chen
EPI_ISL_416334	hCoV-19/Shanghai/SH0025/2020	Asia / China / Shanghai	2020-02-06	Shanghai Public Health Clinical Center, Shanghai Medical College, Fudan University	National Research Center for Translational Medicine (Shanghai), Ruijin Hospital affiliated to Shanghai Jiao Tong University School of Medicine & Shanghai Public Health Clinical Center	Shengyue Wang, Xiaonan Zhang, Gang Lu, Yun Tan, Yun Ling, Hongzhou Lu, Saijuan Chen
EPI_ISL_416335	hCoV-19/Shanghai/SH0026/2020	Asia / China / Shanghai	2020-02-02	Shanghai Public Health Clinical Center, Shanghai Medical College, Fudan University	National Research Center for Translational Medicine (Shanghai), Ruijin Hospital affiliated to Shanghai Jiao Tong University School of Medicine & Shanghai Public Health Clinical Center	Shengyue Wang, Xiaonan Zhang, Gang Lu, Yun Tan, Yun Ling, Hongzhou Lu, Saijuan Chen
EPI_ISL_416336	hCoV-19/Shanghai/SH0027/2020	Asia / China / Shanghai	2020-02-01	Shanghai Public Health Clinical Center, Shanghai Medical College, Fudan University	National Research Center for Translational Medicine (Shanghai), Ruijin Hospital affiliated to Shanghai Jiao Tong University School of Medicine & Shanghai

Public Health Clinical Center	Shengyue Wang, Xiaonan Zhang, Gang Lu, Yun Tan, Yun Ling, Hongzhou Lu, Saijuan Chen
EPI_ISL_416337	hCoV-19/Shanghai/SH0028/2020	Asia / China / Shanghai	2020-02-04	Shanghai Public Health Clinical Center, Shanghai Medical College, Fudan University	National Research Center for Translational Medicine (Shanghai), Ruijin Hospital affiliated to Shanghai Jiao Tong University School of Medicine & Shanghai Public Health Clinical Center	Shengyue Wang, Xiaonan Zhang, Gang Lu, Yun Tan, Yun Ling, Hongzhou Lu, Saijuan Chen
EPI_ISL_416338	hCoV-19/Shanghai/SH0029/2020	Asia / China / Shanghai	2020-02-01	Shanghai Public Health Clinical Center, Shanghai Medical College, Fudan University	National Research Center for Translational Medicine (Shanghai), Ruijin Hospital affiliated to Shanghai Jiao Tong University School of Medicine & Shanghai Public Health Clinical Center	Shengyue Wang, Xiaonan Zhang, Gang Lu, Yun Tan, Yun Ling, Hongzhou Lu, Saijuan Chen
EPI_ISL_416339	hCoV-19/Shanghai/SH0030/2020	Asia / China / Shanghai	2020-02-03	Shanghai Public Health Clinical Center, Shanghai Medical College, Fudan University	National Research Center for Translational Medicine (Shanghai), Ruijin Hospital affiliated to Shanghai Jiao Tong University School of Medicine & Shanghai Public Health Clinical Center	Shengyue Wang, Xiaonan Zhang, Gang Lu, Yun Tan, Yun Ling, Hongzhou Lu, Saijuan Chen
EPI_ISL_416340	hCoV-19/Shanghai/SH0031/2020	Asia / China / Shanghai	2020-02-02	Shanghai Public Health Clinical Center, Shanghai Medical College, Fudan University	National Research Center for Translational Medicine (Shanghai), Ruijin Hospital affiliated to Shanghai Jiao Tong University School of Medicine & Shanghai Public Health Clinical Center	Shengyue Wang, Xiaonan Zhang, Gang Lu, Yun Tan, Yun Ling, Hongzhou Lu, Saijuan Chen
EPI_ISL_416341	hCoV-19/Shanghai/SH0032/2020	Asia / China / Shanghai	2020-02-01	Shanghai Public Health Clinical Center, Shanghai Medical College, Fudan University	National Research Center for Translational Medicine (Shanghai), Ruijin Hospital affiliated to Shanghai Jiao Tong University School of Medicine & Shanghai Public Health Clinical Center	Shengyue Wang, Xiaonan Zhang, Gang Lu, Yun Tan, Yun Ling, Hongzhou Lu, Saijuan Chen
EPI_ISL_416342	hCoV-19/Shanghai/SH0033/2020	Asia / China / Shanghai	2020-02-04	Shanghai Public Health Clinical Center, Shanghai Medical College, Fudan University	National Research Center for Translational Medicine (Shanghai), Ruijin Hospital affiliated to Shanghai Jiao Tong University School of Medicine & Shanghai Public Health Clinical Center	Shengyue Wang, Xiaonan Zhang, Gang Lu, Yun Tan, Yun Ling, Hongzhou Lu, Saijuan Chen
EPI_ISL_416348	hCoV-19/Shanghai/SH0039/2020	Asia / China / Shanghai	2020-02-06	Shanghai Public Health Clinical Center, Shanghai Medical College, Fudan University	National Research Center for Translational Medicine (Shanghai), Ruijin Hospital affiliated to Shanghai Jiao Tong University School of Medicine & Shanghai Public Health Clinical Center	Shengyue Wang, Xiaonan Zhang, Gang Lu, Yun Tan, Yun Ling, Hongzhou Lu, Saijuan Chen
EPI_ISL_416349	hCoV-19/Shanghai/SH0040/2020	Asia / China / Shanghai	2020-01-30	Shanghai Public Health Clinical Center, Shanghai Medical College, Fudan University	National Research Center for Translational Medicine (Shanghai), Ruijin Hospital affiliated to Shanghai Jiao Tong University School of Medicine & Shanghai Public Health Clinical Center	Shengyue Wang, Xiaonan Zhang, Gang Lu, Yun Tan, Yun Ling, Hongzhou Lu, Saijuan Chen
EPI_ISL_416350	hCoV-19/Shanghai/SH0041/2020	Asia / China / Shanghai	2020-01-29	Shanghai Public Health Clinical Center, Shanghai Medical College, Fudan University	National Research Center for Translational Medicine (Shanghai), Ruijin Hospital affiliated to Shanghai Jiao Tong University School of Medicine & Shanghai Public Health Clinical Center	Shengyue Wang, Xiaonan Zhang, Gang Lu, Yun Tan, Yun Ling, Hongzhou Lu, Saijuan Chen
EPI_ISL_416352	hCoV-19/Shanghai/SH0043/2020	Asia / China / Shanghai	2020-01-30	Shanghai Public Health Clinical Center, Shanghai Medical College, Fudan University	National Research Center for Translational Medicine (Shanghai), Ruijin Hospital affiliated to Shanghai Jiao Tong University School of Medicine & Shanghai Public Health Clinical Center	Shengyue Wang, Xiaonan Zhang, Gang Lu, Yun Tan, Yun Ling, Hongzhou Lu, Saijuan Chen
EPI_ISL_416353	hCoV-19/Shanghai/SH0044/2020	Asia / China / Shanghai	2020-02-04	Shanghai Public Health Clinical Center, Shanghai Medical College, Fudan University	National Research Center for Translational Medicine (Shanghai), Ruijin Hospital affiliated to Shanghai Jiao Tong University School of Medicine & Shanghai Public Health Clinical Center	Shengyue Wang, Xiaonan Zhang, Gang Lu, Yun Tan, Yun Ling, Hongzhou Lu, Saijuan Chen
EPI_ISL_416354	hCoV-19/Shanghai/SH0045/2020	Asia / China / Shanghai	2020-02-04	Shanghai Public Health Clinical Center, Shanghai Medical College, Fudan University	National Research Center for Translational Medicine (Shanghai), Ruijin Hospital affiliated to Shanghai Jiao Tong University School of Medicine & Shanghai Public Health Clinical Center	Shengyue Wang, Xiaonan Zhang, Gang Lu, Yun Tan, Yun Ling, Hongzhou Lu, Saijuan Chen
EPI_ISL_416355	hCoV-19/Shanghai/SH0047/2020	Asia / China / Shanghai	2020-02-04	Shanghai Public Health Clinical Center, Shanghai Medical College, Fudan University	National Research Center for Translational Medicine (Shanghai), Ruijin Hospital affiliated to Shanghai Jiao Tong University School of Medicine & Shanghai Public Health Clinical Center	Shengyue Wang, Xiaonan Zhang, Gang Lu, Yun Tan, Yun Ling, Hongzhou Lu, Saijuan Chen
EPI_ISL_416358	hCoV-19/Shanghai/SH0050/2020	Asia / China / Shanghai	2020-02-05	Shanghai Public Health Clinical Center, Shanghai Medical College, Fudan University	National Research Center for Translational Medicine (Shanghai), Ruijin Hospital affiliated to Shanghai Jiao Tong University School of Medicine & Shanghai

Public Health Clinical Center   Shengyue Wang, Xiaonan Zhang, Gang Lu, Yun Tan, Yun Ling, Hongzhou Lu, Saijuan Chen
EPI_ISL_416359  hCoV-19/Shanghai/SH0051/2020    Asia / China / Shanghai 2020-02-06      Shanghai Public Health Clinical Center, Shanghai Medical College, Fudan University     National Research Center for Translational Medicine (Shanghai), Ruijin Hospital affiliated to Shanghai Jiao Tong University School of Medicine & Shanghai Public Health Clinical Center   Shengyue Wang, Xiaonan Zhang, Gang Lu, Yun Tan, Yun Ling, Hongzhou Lu, Saijuan Chen
EPI_ISL_416361  hCoV-19/Shanghai/SH0053/2020    Asia / China / Shanghai 2020-02-01      Shanghai Public Health Clinical Center, Shanghai Medical College, Fudan University     National Research Center for Translational Medicine (Shanghai), Ruijin Hospital affiliated to Shanghai Jiao Tong University School of Medicine & Shanghai Public Health Clinical Center   Shengyue Wang, Xiaonan Zhang, Gang Lu, Yun Tan, Yun Ling, Hongzhou Lu, Saijuan Chen
EPI_ISL_416362  hCoV-19/Shanghai/SH0054/2020    Asia / China / Shanghai 2020-02-02      Shanghai Public Health Clinical Center, Shanghai Medical College, Fudan University     National Research Center for Translational Medicine (Shanghai), Ruijin Hospital affiliated to Shanghai Jiao Tong University School of Medicine & Shanghai Public Health Clinical Center   Shengyue Wang, Xiaonan Zhang, Gang Lu, Yun Tan, Yun Ling, Hongzhou Lu, Saijuan Chen
EPI_ISL_416363  hCoV-19/Shanghai/SH0055/2020    Asia / China / Shanghai 2020-02-08      Shanghai Public Health Clinical Center, Shanghai Medical College, Fudan University     National Research Center for Translational Medicine (Shanghai), Ruijin Hospital affiliated to Shanghai Jiao Tong University School of Medicine & Shanghai Public Health Clinical Center   Shengyue Wang, Xiaonan Zhang, Gang Lu, Yun Tan, Yun Ling, Hongzhou Lu, Saijuan Chen
EPI_ISL_416364  hCoV-19/Shanghai/SH0056/2020    Asia / China / Shanghai 2020-02-08      Shanghai Public Health Clinical Center, Shanghai Medical College, Fudan University     National Research Center for Translational Medicine (Shanghai), Ruijin Hospital affiliated to Shanghai Jiao Tong University School of Medicine & Shanghai Public Health Clinical Center   Shengyue Wang, Xiaonan Zhang, Gang Lu, Yun Tan, Yun Ling, Hongzhou Lu, Saijuan Chen
EPI_ISL_416365  hCoV-19/Shanghai/SH0058/2020    Asia / China / Shanghai 2020-01-30      Shanghai Public Health Clinical Center, Shanghai Medical College, Fudan University     National Research Center for Translational Medicine (Shanghai), Ruijin Hospital affiliated to Shanghai Jiao Tong University School of Medicine & Shanghai Public Health Clinical Center   Shengyue Wang, Xiaonan Zhang, Gang Lu, Yun Tan, Yun Ling, Hongzhou Lu, Saijuan Chen
EPI_ISL_416366  hCoV-19/Shanghai/SH0059/2020    Asia / China / Shanghai 2020-01-30      Shanghai Public Health Clinical Center, Shanghai Medical College, Fudan University     National Research Center for Translational Medicine (Shanghai), Ruijin Hospital affiliated to Shanghai Jiao Tong University School of Medicine & Shanghai Public Health Clinical Center   Shengyue Wang, Xiaonan Zhang, Gang Lu, Yun Tan, Yun Ling, Hongzhou Lu, Saijuan Chen
EPI_ISL_416367  hCoV-19/Shanghai/SH0060/2020    Asia / China / Shanghai 2020-02-01      Shanghai Public Health Clinical Center, Shanghai Medical College, Fudan University     National Research Center for Translational Medicine (Shanghai), Ruijin Hospital affiliated to Shanghai Jiao Tong University School of Medicine & Shanghai Public Health Clinical Center   Shengyue Wang, Xiaonan Zhang, Gang Lu, Yun Tan, Yun Ling, Hongzhou Lu, Saijuan Chen
EPI_ISL_416368  hCoV-19/Shanghai/SH0065/2020    Asia / China / Shanghai 2020-02-05      Shanghai Public Health Clinical Center, Shanghai Medical College, Fudan University     National Research Center for Translational Medicine (Shanghai), Ruijin Hospital affiliated to Shanghai Jiao Tong University School of Medicine & Shanghai Public Health Clinical Center   Shengyue Wang, Xiaonan Zhang, Gang Lu, Yun Tan, Yun Ling, Hongzhou Lu, Saijuan Chen
EPI_ISL_416369  hCoV-19/Shanghai/SH0066/2020    Asia / China / Shanghai 2020-02-05      Shanghai Public Health Clinical Center, Shanghai Medical College, Fudan University     National Research Center for Translational Medicine (Shanghai), Ruijin Hospital affiliated to Shanghai Jiao Tong University School of Medicine & Shanghai Public Health Clinical Center   Shengyue Wang, Xiaonan Zhang, Gang Lu, Yun Tan, Yun Ling, Hongzhou Lu, Saijuan Chen
EPI_ISL_416370  hCoV-19/Shanghai/SH0067/2020    Asia / China / Shanghai 2020-02-01      Shanghai Public Health Clinical Center, Shanghai Medical College, Fudan University     National Research Center for Translational Medicine (Shanghai), Ruijin Hospital affiliated to Shanghai Jiao Tong University School of Medicine & Shanghai Public Health Clinical Center   Shengyue Wang, Xiaonan Zhang, Gang Lu, Yun Tan, Yun Ling, Hongzhou Lu, Saijuan Chen
EPI_ISL_416372  hCoV-19/Shanghai/SH0069/2020    Asia / China / Shanghai 2020-02-01      Shanghai Public Health Clinical Center, Shanghai Medical College, Fudan University     National Research Center for Translational Medicine (Shanghai), Ruijin Hospital affiliated to Shanghai Jiao Tong University School of Medicine & Shanghai Public Health Clinical Center   Shengyue Wang, Xiaonan Zhang, Gang Lu, Yun Tan, Yun Ling, Hongzhou Lu, Saijuan Chen
EPI_ISL_416373  hCoV-19/Shanghai/SH0070/2020    Asia / China / Shanghai 2020-02-07      Shanghai Public Health Clinical Center, Shanghai Medical College, Fudan University     National Research Center for Translational Medicine (Shanghai), Ruijin Hospital affiliated to Shanghai Jiao Tong University School of Medicine & Shanghai Public Health Clinical Center   Shengyue Wang, Xiaonan Zhang, Gang Lu, Yun Tan, Yun Ling, Hongzhou Lu, Saijuan Chen
EPI_ISL_416376  hCoV-19/Shanghai/SH0073/2020    Asia / China / Shanghai 2020-02-05      Shanghai Public Health Clinical Center, Shanghai Medical College, Fudan University     National Research Center for Translational Medicine (Shanghai), Ruijin Hospital affiliated to Shanghai Jiao Tong University School of Medicine & Shanghai

Public Health Clinical Center   Shengyue Wang, Xiaonan Zhang, Gang Lu, Yun Tan, Yun Ling, Hongzhou Lu, Saijuan Chen
EPI_ISL_416377  hCoV-19/Shanghai/SH0074/2020    Asia / China / Shanghai 2020-02-07      Shanghai Public Health Clinical Center, Shanghai Medical College, Fudan University     National Research Center for Translational Medicine (Shanghai), Ruijin Hospital affiliated to Shanghai Jiao Tong University School of Medicine & Shanghai Public Health Clinical Center   Shengyue Wang, Xiaonan Zhang, Gang Lu, Yun Tan, Yun Ling, Hongzhou Lu, Saijuan Chen
EPI_ISL_416378  hCoV-19/Shanghai/SH0075/2020    Asia / China / Shanghai 2020-01-30      Shanghai Public Health Clinical Center, Shanghai Medical College, Fudan University     National Research Center for Translational Medicine (Shanghai), Ruijin Hospital affiliated to Shanghai Jiao Tong University School of Medicine & Shanghai Public Health Clinical Center   Shengyue Wang, Xiaonan Zhang, Gang Lu, Yun Tan, Yun Ling, Hongzhou Lu, Saijuan Chen
EPI_ISL_416379  hCoV-19/Shanghai/SH0076/2020    Asia / China / Shanghai 2020-02-01      Shanghai Public Health Clinical Center, Shanghai Medical College, Fudan University     National Research Center for Translational Medicine (Shanghai), Ruijin Hospital affiliated to Shanghai Jiao Tong University School of Medicine & Shanghai Public Health Clinical Center   Shengyue Wang, Xiaonan Zhang, Gang Lu, Yun Tan, Yun Ling, Hongzhou Lu, Saijuan Chen
EPI_ISL_416380  hCoV-19/Shanghai/SH0077/2020    Asia / China / Shanghai 2020-02-09      Shanghai Public Health Clinical Center, Shanghai Medical College, Fudan University     National Research Center for Translational Medicine (Shanghai), Ruijin Hospital affiliated to Shanghai Jiao Tong University School of Medicine & Shanghai Public Health Clinical Center   Shengyue Wang, Xiaonan Zhang, Gang Lu, Yun Tan, Yun Ling, Hongzhou Lu, Saijuan Chen
EPI_ISL_416381  hCoV-19/Shanghai/SH0079/2020    Asia / China / Shanghai 2020-02-01      Shanghai Public Health Clinical Center, Shanghai Medical College, Fudan University     National Research Center for Translational Medicine (Shanghai), Ruijin Hospital affiliated to Shanghai Jiao Tong University School of Medicine & Shanghai Public Health Clinical Center   Shengyue Wang, Xiaonan Zhang, Gang Lu, Yun Tan, Yun Ling, Hongzhou Lu, Saijuan Chen
EPI_ISL_416382  hCoV-19/Shanghai/SH0080/2020    Asia / China / Shanghai 2020-02-08      Shanghai Public Health Clinical Center, Shanghai Medical College, Fudan University     National Research Center for Translational Medicine (Shanghai), Ruijin Hospital affiliated to Shanghai Jiao Tong University School of Medicine & Shanghai Public Health Clinical Center   Shengyue Wang, Xiaonan Zhang, Gang Lu, Yun Tan, Yun Ling, Hongzhou Lu, Saijuan Chen
EPI_ISL_416384  hCoV-19/Shanghai/SH0083/2020    Asia / China / Shanghai 2020-02-04      Shanghai Public Health Clinical Center, Shanghai Medical College, Fudan University     National Research Center for Translational Medicine (Shanghai), Ruijin Hospital affiliated to Shanghai Jiao Tong University School of Medicine & Shanghai Public Health Clinical Center   Shengyue Wang, Xiaonan Zhang, Gang Lu, Yun Tan, Yun Ling, Hongzhou Lu, Saijuan Chen
EPI_ISL_416387  hCoV-19/Shanghai/SH0088/2020    Asia / China / Shanghai 2020-02-01      Shanghai Public Health Clinical Center, Shanghai Medical College, Fudan University     National Research Center for Translational Medicine (Shanghai), Ruijin Hospital affiliated to Shanghai Jiao Tong University School of Medicine & Shanghai Public Health Clinical Center   Shengyue Wang, Xiaonan Zhang, Gang Lu, Yun Tan, Yun Ling, Hongzhou Lu, Saijuan Chen
EPI_ISL_416389  hCoV-19/Shanghai/SH0093/2020    Asia / China / Shanghai 2020-01-21      Shanghai Public Health Clinical Center, Shanghai Medical College, Fudan University     National Research Center for Translational Medicine (Shanghai), Ruijin Hospital affiliated to Shanghai Jiao Tong University School of Medicine & Shanghai Public Health Clinical Center   Shengyue Wang, Xiaonan Zhang, Gang Lu, Yun Tan, Yun Ling, Hongzhou Lu, Saijuan Chen
EPI_ISL_416390  hCoV-19/Shanghai/SH0094/2020    Asia / China / Shanghai 2020-01-31      Shanghai Public Health Clinical Center, Shanghai Medical College, Fudan University     National Research Center for Translational Medicine (Shanghai), Ruijin Hospital affiliated to Shanghai Jiao Tong University School of Medicine & Shanghai Public Health Clinical Center   Shengyue Wang, Xiaonan Zhang, Gang Lu, Yun Tan, Yun Ling, Hongzhou Lu, Saijuan Chen
EPI_ISL_416393  hCoV-19/Shanghai/SH0101/2020    Asia / China / Shanghai 2020-02-09      Shanghai Public Health Clinical Center, Shanghai Medical College, Fudan University     National Research Center for Translational Medicine (Shanghai), Ruijin Hospital affiliated to Shanghai Jiao Tong University School of Medicine & Shanghai Public Health Clinical Center   Shengyue Wang, Xiaonan Zhang, Gang Lu, Yun Tan, Yun Ling, Hongzhou Lu, Saijuan Chen
EPI_ISL_416394  hCoV-19/Shanghai/SH0102/2020    Asia / China / Shanghai 2020-02-14      Shanghai Public Health Clinical Center, Shanghai Medical College, Fudan University     National Research Center for Translational Medicine (Shanghai), Ruijin Hospital affiliated to Shanghai Jiao Tong University School of Medicine & Shanghai Public Health Clinical Center   Shengyue Wang, Xiaonan Zhang, Gang Lu, Yun Tan, Yun Ling, Hongzhou Lu, Saijuan Chen
EPI_ISL_416396  hCoV-19/Shanghai/SH0106/2020    Asia / China / Shanghai 2020-02-10      Shanghai Public Health Clinical Center, Shanghai Medical College, Fudan University     National Research Center for Translational Medicine (Shanghai), Ruijin Hospital affiliated to Shanghai Jiao Tong University School of Medicine & Shanghai Public Health Clinical Center   Shengyue Wang, Xiaonan Zhang, Gang Lu, Yun Tan, Yun Ling, Hongzhou Lu, Saijuan Chen
EPI_ISL_416397  hCoV-19/Shanghai/SH0107/2020    Asia / China / Shanghai 2020-02-02      Shanghai Public Health Clinical Center, Shanghai Medical College, Fudan University     National Research Center for Translational Medicine (Shanghai), Ruijin Hospital affiliated to Shanghai Jiao Tong University School of Medicine & Shanghai

Public Health Clinical Center	Shengyue Wang, Xiaonan Zhang, Gang Lu, Yun Tan, Yun Ling, Hongzhou Lu, Saijuan Chen
EPI_ISL_416398	hCoV-19/Shanghai/SH0109/2020	Asia / China / Shanghai	2020-02-02	Shanghai Public Health Clinical Center, Shanghai Medical College, Fudan University	National Research Center for Translational Medicine (Shanghai), Ruijin Hospital affiliated to Shanghai Jiao Tong University School of Medicine & Shanghai Public Health Clinical Center	Shengyue Wang, Xiaonan Zhang, Gang Lu, Yun Tan, Yun Ling, Hongzhou Lu, Saijuan Chen
EPI_ISL_416399	hCoV-19/Shanghai/SH0110/2020	Asia / China / Shanghai	2020-02-13	Shanghai Public Health Clinical Center, Shanghai Medical College, Fudan University	National Research Center for Translational Medicine (Shanghai), Ruijin Hospital affiliated to Shanghai Jiao Tong University School of Medicine & Shanghai Public Health Clinical Center	Shengyue Wang, Xiaonan Zhang, Gang Lu, Yun Tan, Yun Ling, Hongzhou Lu, Saijuan Chen
EPI_ISL_417370	hCoV-19/USA/WA-UW221/2020	North America / USA / Washington	2020-03-14	UW Virology Lab	UW Virology Lab	Pavitra Roychoudhury, Hong Xie, Keith Jerome, Alexander Greninger
EPI_ISL_417371	hCoV-19/USA/WA-UW222/2020	North America / USA / Washington	2020-03-13	UW Virology Lab	UW Virology Lab	Pavitra Roychoudhury, Hong Xie, Keith Jerome, Alexander Greninger
EPI_ISL_417372	hCoV-19/USA/CT-UW223/2020	North America / USA / Connecticut	2020-03-13	UW Virology Lab	UW Virology Lab	Pavitra Roychoudhury, Hong Xie, Keith Jerome, Alexander Greninger
EPI_ISL_417373	hCoV-19/USA/CT-UW224/2020	North America / USA / Connecticut	2020-03-13	UW Virology Lab	UW Virology Lab	Pavitra Roychoudhury, Hong Xie, Keith Jerome, Alexander Greninger
EPI_ISL_417374	hCoV-19/USA/WA-UW225/2020	North America / USA / Washington	2020-03-15	UW Virology Lab	UW Virology Lab	Pavitra Roychoudhury, Hong Xie, Keith Jerome, Alexander Greninger
EPI_ISL_417375	hCoV-19/USA/WA-UW226/2020	North America / USA / Washington	2020-03-15	UW Virology Lab	UW Virology Lab	Pavitra Roychoudhury, Hong Xie, Keith Jerome, Alexander Greninger
EPI_ISL_417376	hCoV-19/USA/WA-UW227/2020	North America / USA / Washington	2020-03-15	UW Virology Lab	UW Virology Lab	Pavitra Roychoudhury, Hong Xie, Keith Jerome, Alexander Greninger
EPI_ISL_417377	hCoV-19/USA/WA-UW228/2020	North America / USA / Washington	2020-03-15	UW Virology Lab	UW Virology Lab	Pavitra Roychoudhury, Hong Xie, Keith Jerome, Alexander Greninger
EPI_ISL_417378	hCoV-19/USA/WA-UW229/2020	North America / USA / Washington	2020-03-14	UW Virology Lab	UW Virology Lab	Pavitra Roychoudhury, Hong Xie, Keith Jerome, Alexander Greninger
EPI_ISL_417379	hCoV-19/USA/WA-UW230/2020	North America / USA / Washington	2020-03-15	UW Virology Lab	UW Virology Lab	Pavitra Roychoudhury, Hong Xie, Keith Jerome, Alexander Greninger
EPI_ISL_417380	hCoV-19/USA/WA-UW231/2020	North America / USA / Washington	2020-03-14	UW Virology Lab	UW Virology Lab	Pavitra Roychoudhury, Hong Xie, Keith Jerome, Alexander Greninger
EPI_ISL_417381	hCoV-19/USA/WA-UW232/2020	North America / USA / Washington	2020-03-14	UW Virology Lab	UW Virology Lab	Pavitra Roychoudhury, Hong Xie, Keith Jerome, Alexander Greninger
EPI_ISL_417382	hCoV-19/USA/WA-UW233/2020	North America / USA / Washington	2020-03-15	UW Virology Lab	UW Virology Lab	Pavitra Roychoudhury, Hong Xie, Keith Jerome, Alexander Greninger
EPI_ISL_417383	hCoV-19/Australia/NSW15/2020	Oceania / Australia / New South Wales / Sydney	2020-03-03	Centre for Infectious Diseases and Microbiology Public Health	NSW Health Pathology - Institute of Clinical Pathology and Medical Research; Westmead Hospital; University of Sydney	Rockett R, Eden J-S, Lam C, Gray K, Timms V, Gall M, Arnott A, Sadsad R, Carter I, Rahman H, Holmes EC, O’Sullivan MV, Sintchenko V, Chen SC, Maddocks S, Kok J and Dwyer DE for the 2019-nCoV Study Group
EPI_ISL_417384	hCoV-19/Australia/NSW18/2020	Oceania / Australia / New South Wales / Sydney	2020-03-05	Centre for Infectious Diseases and Microbiology Public Health	NSW Health Pathology - Institute of Clinical Pathology and Medical Research; Westmead Hospital; University of Sydney	Eden J-S, Lam C, Gray K, Timms V, Gall M, Arnott A, Sadsad R, Carter I, Rahman H, Holmes EC, O’Sullivan MV, Sintchenko V, Chen SC, Maddocks S, Kok J, Dwyer DE and Rockett R for the 2019-nCoV Study Group
EPI_ISL_417385	hCoV-19/Australia/NSW19/2020	Oceania / Australia / New South Wales / Sydney	2020-03-06	Centre for Infectious Diseases and Microbiology Public Health	NSW Health Pathology - Institute of Clinical Pathology and Medical Research; Westmead Hospital; University of Sydney	Lam C, Gray K, Timms V, Gall M, Arnott A, Sadsad R, Carter I, Rahman H, Holmes EC, O’Sullivan MV, Sintchenko V, Chen SC, Maddocks S, Kok J, Dwyer DE, Rockett R and Eden J-S for the 2019-nCoV Study Group
EPI_ISL_417386	hCoV-19/Australia/NSW21/2020	Oceania / Australia / New South Wales / Sydney	2020-03-04	Centre for Infectious Diseases and Microbiology Public Health	NSW Health Pathology - Institute of Clinical Pathology and Medical Research; Westmead Hospital; University of Sydney	Gray K, Timms V, Gall M, Arnott A, Sadsad R, Carter I, Rahman H, Holmes EC, O’Sullivan MV, Sintchenko V, Chen SC, Maddocks S, Kok J, Dwyer DE, Rockett R, Eden J-S and Lam C for the 2019-nCoV Study Group
EPI_ISL_417387	hCoV-19/Australia/NSW24/2020	Oceania / Australia / New South Wales / Sydney	2020-03-06	Centre for Infectious Diseases and Microbiology Public Health	NSW Health Pathology - Institute of Clinical Pathology and Medical Research; Westmead Hospital; University of Sydney	Timms V, Gall M, Arnott A, Sadsad R, Carter I, Rahman H, Holmes EC, O’Sullivan MV, Sintchenko V, Chen SC, Maddocks S, Kok J, Dwyer DE, Rockett R, Eden J-S, Lam C and Gray K for the 2019-nCoV Study Group
EPI_ISL_417388	hCoV-19/Australia/NSW25/2020	Oceania / Australia / New South Wales / Sydney	2020-03-05	Centre for Infectious Diseases and Microbiology Public Health	NSW Health Pathology - Institute of Clinical Pathology and Medical Research; Westmead Hospital; University of Sydney	Gall M, Arnott A, Sadsad R, Carter I, Rahman H, Holmes EC, O’Sullivan MV, Sintchenko V, Chen SC, Maddocks S, Kok J, Dwyer DE, Rockett R, Eden J-S, Lam C, Gray K and Timms V for the 2019-nCoV Study Group
EPI_ISL_417389	hCoV-19/Australia/NSW26/2020	Oceania / Australia / New South Wales / Sydney	2020-03-09	Centre for Infectious Diseases and Microbiology Public Health	NSW Health Pathology - Institute of Clinical

Pathology and Medical Research; Westmead Hospital; University of Sydney Arnott A, Sadsad R, Carter I, Rahman H, Holmes EC, O'Sullivan MV, Sintchenko V, Chen SC, Maddocks S, Kok J, Dwyer DE, Rockett R, Eden J-S, Lam C, Gray K, Timms V and Gall M for the 2019-nCoV Study Group
EPI_ISL_417507 hCoV-19/USA/WI-UW-23/2020 North America / USA / Wisconsin 2020-03-17 University of Wisconsin-Madison AIDS Vaccine Research Laboratories University of Wisconsin-Madison AIDS Vaccine Research Laboratories Gage Moreno, Katarina Braun, et al. AIDS Vaccine Research Laboratories
EPI_ISL_417508 hCoV-19/USA/WI-UW-21/2020 North America / USA / Wisconsin 2020-03-16 University of Wisconsin-Madison AIDS Vaccine Research Laboratories University of Wisconsin-Madison AIDS Vaccine Research Laboratories Gage Moreno, Katarina Braun, et al. AIDS Vaccine Research Laboratories
EPI_ISL_417509 hCoV-19/USA/WI-UW-16/2020 North America / USA / Wisconsin 2020-03-14 University of Wisconsin-Madison AIDS Vaccine Research Laboratories University of Wisconsin-Madison AIDS Vaccine Research Laboratories Gage Moreno, Katarina Braun, et al. AIDS Vaccine Research Laboratories
EPI_ISL_417510 hCoV-19/USA/WI-UW-19/2020 North America / USA / Wisconsin 2020-03-16 University of Wisconsin-Madison AIDS Vaccine Research Laboratories University of Wisconsin-Madison AIDS Vaccine Research Laboratories Gage Moreno, Katarina Braun, et al. AIDS Vaccine Research Laboratories
EPI_ISL_417512 hCoV-19/USA/WI-UW-24/2020 North America / USA / Wisconsin 2020-03-15 University of Wisconsin-Madison AIDS Vaccine Research Laboratories University of Wisconsin-Madison AIDS Vaccine Research Laboratories Gage Moreno, Katarina Braun, et al. AIDS Vaccine Research Laboratories
EPI_ISL_417513 hCoV-19/USA/WI-UW-14/2020 North America / USA / Wisconsin 2020-03-16 University of Wisconsin-Madison AIDS Vaccine Research Laboratories University of Wisconsin-Madison AIDS Vaccine Research Laboratories Gage Moreno, Katarina Braun, et al. AIDS Vaccine Research Laboratories
EPI_ISL_417514 hCoV-19/USA/WI-UW-22/2020 North America / USA / Wisconsin 2020-03-13 University of Wisconsin-Madison AIDS Vaccine Research Laboratories University of Wisconsin-Madison AIDS Vaccine Research Laboratories Gage Moreno, Katarina Braun, et al. AIDS Vaccine Research Laboratories
EPI_ISL_417515 hCoV-19/USA/WI-UW-18/2020 North America / USA / Wisconsin 2020-03-17 University of Wisconsin-Madison AIDS Vaccine Research Laboratories University of Wisconsin-Madison AIDS Vaccine Research Laboratories Gage Moreno, Katarina Braun, et al. AIDS Vaccine Research Laboratories
EPI_ISL_417516 hCoV-19/USA/WI-UW-13/2020 North America / USA / Wisconsin 2020-03-15 University of Wisconsin-Madison AIDS Vaccine Research Laboratories University of Wisconsin-Madison AIDS Vaccine Research Laboratories Gage Moreno, Katarina Braun, et al. AIDS Vaccine Research Laboratories
EPI_ISL_417517 hCoV-19/USA/WI-UW-17/2020 North America / USA / Wisconsin 2020-03-13 University of Wisconsin-Madison AIDS Vaccine Research Laboratories University of Wisconsin-Madison AIDS Vaccine Research Laboratories Gage Moreno, Katarina Braun, et al. AIDS Vaccine Research Laboratories
EPI_ISL_417519 hCoV-19/Taiwan/CGMH-CGU-06/2020 Asia / Taiwan / Taoyuan 2020-03-05 Laboratory Medicine Department of Laboratory Medicine, Lin-Kou Chang Gung Memorial Hospital, Taoyuan, Taiwan Kuo-Chien Tsao, Yu-Nong Gong, Shu-Li Yang, Yi-Chun Liu, Chung-Guei Huang, Po-Wei Huang, Mei-Jen Hsiao, Cheng-Ta Yang, Cheng-Hsun Chiu, Peng-Nien Huang, Kuo-Ming Lee, Guang-Wu Chen , Shin-Ru Shih
EPI_ISL_417520 hCoV-19/Taiwan/CGMH-CGU-07/2020 Asia / Taiwan / Taoyuan 2020-03-09 Laboratory Medicine Department of Laboratory Medicine, Lin-Kou Chang Gung Memorial Hospital, Taoyuan, Taiwan Kuo-Chien Tsao, Yu-Nong Gong, Shu-Li Yang, Yi-Chun Liu, Chung-Guei Huang, Po-Wei Huang, Mei-Jen Hsiao, Cheng-Ta Yang, Cheng-Hsun Chiu, Peng-Nien Huang, Kuo-Ming Lee, Guang-Wu Chen , Shin-Ru Shih
EPI_ISL_417521 hCoV-19/Taiwan/CGMH-CGU-08/2020 Asia / Taiwan / Taoyuan 2020-03-10 Laboratory Medicine Department of Laboratory Medicine, Lin-Kou Chang Gung Memorial Hospital, Taoyuan, Taiwan Kuo-Chien Tsao, Yu-Nong Gong, Shu-Li Yang, Yi-Chun Liu, Chung-Guei Huang, Po-Wei Huang, Mei-Jen Hsiao, Cheng-Ta Yang, Cheng-Hsun Chiu, Peng-Nien Huang, Kuo-Ming Lee, Guang-Wu Chen , Shin-Ru Shih
EPI_ISL_417522 hCoV-19/Taiwan/CGMH-CGU-09/2020 Asia / Taiwan / Taoyuan 2020-03-13 Laboratory Medicine Department of Laboratory Medicine, Lin-Kou Chang Gung Memorial Hospital, Taoyuan, Taiwan Kuo-Chien Tsao, Yu-Nong Gong, Shu-Li Yang, Yi-Chun Liu, Chung-Guei Huang, Po-Wei Huang, Mei-Jen Hsiao, Cheng-Ta Yang, Cheng-Hsun Chiu, Peng-Nien Huang, Kuo-Ming Lee, Guang-Wu Chen , Shin-Ru Shih
EPI_ISL_417523 hCoV-19/Taiwan/CGMH-CGU-10/2020 Asia / Taiwan / Taoyuan 2020-03-13 Laboratory Medicine Department of Laboratory Medicine, Lin-Kou Chang Gung Memorial Hospital, Taoyuan, Taiwan Kuo-Chien Tsao, Yu-Nong Gong, Shu-Li Yang, Yi-Chun Liu, Chung-Guei Huang, Po-Wei Huang, Mei-Jen Hsiao, Cheng-Ta Yang, Cheng-Hsun Chiu, Peng-Nien Huang, Kuo-Ming Lee, Guang-Wu Chen , Shin-Ru Shih
EPI_ISL_417524 hCoV-19/Taiwan/CGMH-CGU-11/2020 Asia / Taiwan / Taoyuan 2020-03-14 Laboratory Medicine Department of Laboratory Medicine, Lin-Kou Chang Gung Memorial Hospital, Taoyuan, Taiwan Kuo-Chien Tsao, Yu-Nong Gong, Shu-Li Yang, Yi-Chun Liu, Chung-Guei Huang, Po-Wei Huang, Mei-Jen Hsiao, Cheng-Ta Yang, Cheng-Hsun Chiu, Peng-Nien Huang, Kuo-Ming Lee, Guang-Wu Chen , Shin-Ru Shih
EPI_ISL_417525 hCoV-19/Taiwan/CGMH-CGU-12/2020 Asia / Taiwan / Taoyuan 2020-03-14 Laboratory Medicine Department of Laboratory Medicine, Lin-Kou Chang Gung Memorial Hospital, Taoyuan, Taiwan Kuo-Chien Tsao, Yu-Nong Gong, Shu-Li Yang, Yi-Chun Liu, Chung-Guei Huang, Po-Wei Huang, Mei-Jen Hsiao, Cheng-Ta Yang, Cheng-Hsun Chiu, Peng-Nien Huang, Kuo-Ming Lee, Guang-Wu Chen , Shin-Ru Shih
EPI_ISL_417526 hCoV-19/Luxembourg/LNS9371718/2020 Europe / Luxembourg 2020-03-18 Laboratoire Nationale de Santé, Microbiology, Virology Laboratoire Nationale de Santé, Microbiology, Epidemiology and Microbial Genomics Anke Wienecke-Baldacchino, Ardashel Latsuzbaia, Jessica Tapp, Catherine Ragimbeau, Guillaume Fournier, Tamir Abdelrahman, Trung Nguyen Nguyen, Joel Mossong
EPI_ISL_417527 hCoV-19/Luxembourg/LNS2848109/2020 Europe / Luxembourg 2020-03-18 Laboratoire Nationale de Santé, Microbiology, Virology Laboratoire Nationale de Santé, Microbiology, Epidemiology and Microbial Genomics Anke Wienecke-Baldacchino, Ardashel Latsuzbaia, Jessica Tapp, Catherine Ragimbeau, Guillaume Fournier, Tamir Abdelrahman, Trung Nguyen Nguyen, Joel Mossong
EPI_ISL_417528 hCoV-19/Luxembourg/LNS3694003/2020 Europe / Luxembourg 2020-03-18 Laboratoire

Nationale de Santé, Microbiology, Virology	Laboratoire Nationale de Santé, Microbiology, Epidemiology and Microbial Genomics	Anke Wienecke-Baldacchino, Ardashel Latsuzbaia, Jessica Tapp, Catherine Ragimbeau, Guillaume Fournier, Tamir Abdelrahman, Trung Nguyen Nguyen, Joel Mossong
EPI_ISL_417600	hCoV-19/Iceland/272/2020	Europe / Iceland / Reykjavik	2020-03-17	The National University Hospital of Iceland	deCODE genetics Daniel F Gudbjartsson; Agnar Helgason; Hakon Jonsson; Olafur T Magnusson; Pall Melsted; Gudmundur L Norddahl; Jona Saemundsdottir; Asgeir Sigurdsson; Patrick Sulem; Arna B Agustsdottir; Berglind Eiriksdottir; Run Fridriksdottir; Elisabet E Gardarsdottir; Gudmundur Georgsson; Olafia S Gretarsdottir; Kjartan R Gudmundsson; Thora R Gunnarsdottir; Arnaldur Gylfason; Hilma Holm; Brynjar O Jensson; Aslaug Jonasdottir; Kamilla S Josefsdottir; Thordur Kristjansson; Droplaug N Magnusdottir; Louise le Roux; Gudrun Sigmundsdottir; Gardar Sveinbjornsson; Kristin E Sveinsdottir; Maney Sveinsdottir; Emil A Thorarensen; Bjarni Thorbjornsson; Gisli Masson; Ingileif Jonsdottir; Alma Moller; Thorolfur Gudnason; Karl G Kristinsson; Unnur Thorsteinsdottir; Kari Stefansson
EPI_ISL_417601	hCoV-19/Iceland/273/2020	Europe / Iceland / Reykjavik	2020-03-17	The National University Hospital of Iceland	deCODE genetics Daniel F Gudbjartsson; Agnar Helgason; Hakon Jonsson; Olafur T Magnusson; Pall Melsted; Gudmundur L Norddahl; Jona Saemundsdottir; Asgeir Sigurdsson; Patrick Sulem; Arna B Agustsdottir; Berglind Eiriksdottir; Run Fridriksdottir; Elisabet E Gardarsdottir; Gudmundur Georgsson; Olafia S Gretarsdottir; Kjartan R Gudmundsson; Thora R Gunnarsdottir; Arnaldur Gylfason; Hilma Holm; Brynjar O Jensson; Aslaug Jonasdottir; Kamilla S Josefsdottir; Thordur Kristjansson; Droplaug N Magnusdottir; Louise le Roux; Gudrun Sigmundsdottir; Gardar Sveinbjornsson; Kristin E Sveinsdottir; Maney Sveinsdottir; Emil A Thorarensen; Bjarni Thorbjornsson; Gisli Masson; Ingileif Jonsdottir; Alma Moller; Thorolfur Gudnason; Karl G Kristinsson; Unnur Thorsteinsdottir; Kari Stefansson
EPI_ISL_417602	hCoV-19/Iceland/274/2020	Europe / Iceland / Reykjavik	2020-03-17	The National University Hospital of Iceland	deCODE genetics Daniel F Gudbjartsson; Agnar Helgason; Hakon Jonsson; Olafur T Magnusson; Pall Melsted; Gudmundur L Norddahl; Jona Saemundsdottir; Asgeir Sigurdsson; Patrick Sulem; Arna B Agustsdottir; Berglind Eiriksdottir; Run Fridriksdottir; Elisabet E Gardarsdottir; Gudmundur Georgsson; Olafia S Gretarsdottir; Kjartan R Gudmundsson; Thora R Gunnarsdottir; Arnaldur Gylfason; Hilma Holm; Brynjar O Jensson; Aslaug Jonasdottir; Kamilla S Josefsdottir; Thordur Kristjansson; Droplaug N Magnusdottir; Louise le Roux; Gudrun Sigmundsdottir; Gardar Sveinbjornsson; Kristin E Sveinsdottir; Maney Sveinsdottir; Emil A Thorarensen; Bjarni Thorbjornsson; Gisli Masson; Ingileif Jonsdottir; Alma Moller; Thorolfur Gudnason; Karl G Kristinsson; Unnur Thorsteinsdottir; Kari Stefansson
EPI_ISL_417603	hCoV-19/Iceland/275/2020	Europe / Iceland / Reykjavik	2020-03-17	The National University Hospital of Iceland	deCODE genetics Daniel F Gudbjartsson; Agnar Helgason; Hakon Jonsson; Olafur T Magnusson; Pall Melsted; Gudmundur L Norddahl; Jona Saemundsdottir; Asgeir Sigurdsson; Patrick Sulem; Arna B Agustsdottir; Berglind Eiriksdottir; Run Fridriksdottir; Elisabet E Gardarsdottir; Gudmundur Georgsson; Olafia S Gretarsdottir; Kjartan R Gudmundsson; Thora R Gunnarsdottir; Arnaldur Gylfason; Hilma Holm; Brynjar O Jensson; Aslaug Jonasdottir; Kamilla S Josefsdottir; Thordur Kristjansson; Droplaug N Magnusdottir; Louise le Roux; Gudrun Sigmundsdottir; Gardar Sveinbjornsson; Kristin E Sveinsdottir; Maney Sveinsdottir; Emil A Thorarensen; Bjarni Thorbjornsson; Gisli Masson; Ingileif Jonsdottir; Alma Moller; Thorolfur Gudnason; Karl G Kristinsson; Unnur Thorsteinsdottir; Kari Stefansson
EPI_ISL_417604	hCoV-19/Iceland/276/2020	Europe / Iceland / Reykjavik	2020-03-17	The National University Hospital of Iceland	deCODE genetics Daniel F Gudbjartsson; Agnar Helgason; Hakon Jonsson; Olafur T Magnusson; Pall Melsted; Gudmundur L Norddahl; Jona Saemundsdottir; Asgeir Sigurdsson; Patrick Sulem; Arna B Agustsdottir; Berglind Eiriksdottir; Run Fridriksdottir; Elisabet E Gardarsdottir; Gudmundur Georgsson; Olafia S Gretarsdottir; Kjartan R Gudmundsson; Thora R Gunnarsdottir; Arnaldur Gylfason; Hilma Holm; Brynjar O Jensson; Aslaug Jonasdottir; Kamilla S Josefsdottir; Thordur Kristjansson; Droplaug N Magnusdottir; Louise le Roux; Gudrun Sigmundsdottir; Gardar Sveinbjornsson; Kristin E Sveinsdottir; Maney Sveinsdottir; Emil A Thorarensen; Bjarni Thorbjornsson; Gisli Masson; Ingileif Jonsdottir; Alma Moller; Thorolfur Gudnason; Karl G Kristinsson; Unnur Thorsteinsdottir; Kari Stefansson
EPI_ISL_417607	hCoV-19/Iceland/279/2020	Europe / Iceland / Reykjavik	2020-03-17	The National University Hospital of Iceland	deCODE genetics Daniel F Gudbjartsson; Agnar Helgason; Hakon Jonsson; Olafur T Magnusson; Pall Melsted; Gudmundur L Norddahl; Jona Saemundsdottir; Asgeir Sigurdsson; Patrick Sulem; Arna B Agustsdottir; Berglind Eiriksdottir; Run Fridriksdottir; Elisabet E Gardarsdottir; Gudmundur Georgsson; Olafia S Gretarsdottir; Kjartan R Gudmundsson; Thora R Gunnarsdottir; Arnaldur Gylfason; Hilma Holm; Brynjar O Jensson; Aslaug Jonasdottir; Kamilla S Josefsdottir; Thordur Kristjansson; Droplaug N Magnusdottir; Louise le Roux; Gudrun Sigmundsdottir; Gardar Sveinbjornsson; Kristin E Sveinsdottir; Maney Sveinsdottir; Emil A Thorarensen; Bjarni Thorbjornsson; Gisli Masson; Ingileif Jonsdottir; Alma Moller; Thorolfur Gudnason; Karl G Kristinsson; Unnur Thorsteinsdottir; Kari Stefansson
EPI_ISL_417608	hCoV-19/Iceland/280/2020	Europe / Iceland / Reykjavik	2020-03-17	The National University Hospital of Iceland	deCODE genetics Daniel F Gudbjartsson; Agnar Helgason; Hakon Jonsson; Olafur T Magnusson; Pall Melsted; Gudmundur L Norddahl; Jona Saemundsdottir; Asgeir Sigurdsson; Patrick Sulem; Arna B Agustsdottir; Berglind Eiriksdottir; Run Fridriksdottir; Elisabet E Gardarsdottir; Gudmundur Georgsson; Olafia S Gretarsdottir; Kjartan R Gudmundsson; Thora R Gunnarsdottir; Arnaldur Gylfason; Hilma Holm; Brynjar O Jensson; Aslaug Jonasdottir; Kamilla S Josefsdottir; Thordur Kristjansson; Droplaug N Magnusdottir; Louise le Roux; Gudrun Sigmundsdottir; Gardar Sveinbjornsson; Kristin E Sveinsdottir; Maney Sveinsdottir; Emil A Thorarensen; Bjarni Thorbjornsson; Gisli Masson; Ingileif Jonsdottir; Alma Moller; Thorolfur Gudnason; Karl G Kristinsson; Unnur Thorsteinsdottir; Kari Stefansson
EPI_ISL_417609	hCoV-19/Iceland/281/2020	Europe / Iceland / Reykjavik	2020-03-17	The National University Hospital of Iceland	deCODE genetics Daniel F Gudbjartsson; Agnar Helgason; Hakon Jonsson; Olafur T Magnusson; Pall Melsted; Gudmundur L Norddahl; Jona Saemundsdottir; Asgeir Sigurdsson; Patrick Sulem; Arna B Agustsdottir; Berglind Eiriksdottir; Run Fridriksdottir; Elisabet E Gardarsdottir; Gudmundur Georgsson; Olafia

S Gretarsdottir; Kjartan R Gudmundsson; Thora R Gunnarsdottir; Arnaldur Gylfason; Hilma Holm; Brynjar O Jensson; Aslaug Jonasdottir; Kamilla S Josefsdottir; Thordur Kristjansson; Droplaug N Magnusdottir; Louise le Roux; Gudrun Sigmundsdottir; Gardar Sveinbjornsson; Kristin E Sveinsdottir; Maney Sveinsdottir; Emil A Thorarensen; Bjarni Thorbjornsson; Gisli Masson; Ingileif Jonsdottir; Alma Moller; Thorolfur Gudnason; Karl G Kristinsson; Unnur Thorsteinsdottir; Kari Stefansson
EPI_ISL_417610 hCoV-19/Iceland/282/2020 Europe / Iceland / Reykjavik 2020-03-17 The National University Hospital of Iceland deCODE genetics Daniel F Gudbjartsson; Agnar Helgason; Hakon Jonsson; Olafur T Magnusson; Pall Melsted; Gudmundur L Norddahl; Jona Saemundsdottir; Asgeir Sigurdsson; Patrick Sulem; Arna B Agustsdottir; Berglind Eiriksdottir; Run Fridriksdottir; Elisabet E Gardarsdottir; Gudmundur Georgsson; Olafia S Gretarsdottir; Kjartan R Gudmundsson; Thora R Gunnarsdottir; Arnaldur Gylfason; Hilma Holm; Brynjar O Jensson; Aslaug Jonasdottir; Kamilla S Josefsdottir; Thordur Kristjansson; Droplaug N Magnusdottir; Louise le Roux; Gudrun Sigmundsdottir; Gardar Sveinbjornsson; Kristin E Sveinsdottir; Maney Sveinsdottir; Emil A Thorarensen; Bjarni Thorbjornsson; Gisli Masson; Ingileif Jonsdottir; Alma Moller; Thorolfur Gudnason; Karl G Kristinsson; Unnur Thorsteinsdottir; Kari Stefansson
EPI_ISL_417611 hCoV-19/Iceland/283/2020 Europe / Iceland / Reykjavik 2020-03-17 The National University Hospital of Iceland deCODE genetics Daniel F Gudbjartsson; Agnar Helgason; Hakon Jonsson; Olafur T Magnusson; Pall Melsted; Gudmundur L Norddahl; Jona Saemundsdottir; Asgeir Sigurdsson; Patrick Sulem; Arna B Agustsdottir; Berglind Eiriksdottir; Run Fridriksdottir; Elisabet E Gardarsdottir; Gudmundur Georgsson; Olafia S Gretarsdottir; Kjartan R Gudmundsson; Thora R Gunnarsdottir; Arnaldur Gylfason; Hilma Holm; Brynjar O Jensson; Aslaug Jonasdottir; Kamilla S Josefsdottir; Thordur Kristjansson; Droplaug N Magnusdottir; Louise le Roux; Gudrun Sigmundsdottir; Gardar Sveinbjornsson; Kristin E Sveinsdottir; Maney Sveinsdottir; Emil A Thorarensen; Bjarni Thorbjornsson; Gisli Masson; Ingileif Jonsdottir; Alma Moller; Thorolfur Gudnason; Karl G Kristinsson; Unnur Thorsteinsdottir; Kari Stefansson
EPI_ISL_417612 hCoV-19/Iceland/284/2020 Europe / Iceland / Reykjavik 2020-03-17 The National University Hospital of Iceland deCODE genetics Daniel F Gudbjartsson; Agnar Helgason; Hakon Jonsson; Olafur T Magnusson; Pall Melsted; Gudmundur L Norddahl; Jona Saemundsdottir; Asgeir Sigurdsson; Patrick Sulem; Arna B Agustsdottir; Berglind Eiriksdottir; Run Fridriksdottir; Elisabet E Gardarsdottir; Gudmundur Georgsson; Olafia S Gretarsdottir; Kjartan R Gudmundsson; Thora R Gunnarsdottir; Arnaldur Gylfason; Hilma Holm; Brynjar O Jensson; Aslaug Jonasdottir; Kamilla S Josefsdottir; Thordur Kristjansson; Droplaug N Magnusdottir; Louise le Roux; Gudrun Sigmundsdottir; Gardar Sveinbjornsson; Kristin E Sveinsdottir; Maney Sveinsdottir; Emil A Thorarensen; Bjarni Thorbjornsson; Gisli Masson; Ingileif Jonsdottir; Alma Moller; Thorolfur Gudnason; Karl G Kristinsson; Unnur Thorsteinsdottir; Kari Stefansson
EPI_ISL_417613 hCoV-19/Iceland/285/2020 Europe / Iceland / Reykjavik 2020-03-17 The National University Hospital of Iceland deCODE genetics Daniel F Gudbjartsson; Agnar Helgason; Hakon Jonsson; Olafur T Magnusson; Pall Melsted; Gudmundur L Norddahl; Jona Saemundsdottir; Asgeir Sigurdsson; Patrick Sulem; Arna B Agustsdottir; Berglind Eiriksdottir; Run Fridriksdottir; Elisabet E Gardarsdottir; Gudmundur Georgsson; Olafia S Gretarsdottir; Kjartan R Gudmundsson; Thora R Gunnarsdottir; Arnaldur Gylfason; Hilma Holm; Brynjar O Jensson; Aslaug Jonasdottir; Kamilla S Josefsdottir; Thordur Kristjansson; Droplaug N Magnusdottir; Louise le Roux; Gudrun Sigmundsdottir; Gardar Sveinbjornsson; Kristin E Sveinsdottir; Maney Sveinsdottir; Emil A Thorarensen; Bjarni Thorbjornsson; Gisli Masson; Ingileif Jonsdottir; Alma Moller; Thorolfur Gudnason; Karl G Kristinsson; Unnur Thorsteinsdottir; Kari Stefansson
EPI_ISL_417614 hCoV-19/Iceland/286/2020 Europe / Iceland / Reykjavik 2020-03-18 The National University Hospital of Iceland deCODE genetics Daniel F Gudbjartsson; Agnar Helgason; Hakon Jonsson; Olafur T Magnusson; Pall Melsted; Gudmundur L Norddahl; Jona Saemundsdottir; Asgeir Sigurdsson; Patrick Sulem; Arna B Agustsdottir; Berglind Eiriksdottir; Run Fridriksdottir; Elisabet E Gardarsdottir; Gudmundur Georgsson; Olafia S Gretarsdottir; Kjartan R Gudmundsson; Thora R Gunnarsdottir; Arnaldur Gylfason; Hilma Holm; Brynjar O Jensson; Aslaug Jonasdottir; Kamilla S Josefsdottir; Thordur Kristjansson; Droplaug N Magnusdottir; Louise le Roux; Gudrun Sigmundsdottir; Gardar Sveinbjornsson; Kristin E Sveinsdottir; Maney Sveinsdottir; Emil A Thorarensen; Bjarni Thorbjornsson; Gisli Masson; Ingileif Jonsdottir; Alma Moller; Thorolfur Gudnason; Karl G Kristinsson; Unnur Thorsteinsdottir; Kari Stefansson
EPI_ISL_417615 hCoV-19/Iceland/287/2020 Europe / Iceland / Reykjavik 2020-03-18 The National University Hospital of Iceland deCODE genetics Daniel F Gudbjartsson; Agnar Helgason; Hakon Jonsson; Olafur T Magnusson; Pall Melsted; Gudmundur L Norddahl; Jona Saemundsdottir; Asgeir Sigurdsson; Patrick Sulem; Arna B Agustsdottir; Berglind Eiriksdottir; Run Fridriksdottir; Elisabet E Gardarsdottir; Gudmundur Georgsson; Olafia S Gretarsdottir; Kjartan R Gudmundsson; Thora R Gunnarsdottir; Arnaldur Gylfason; Hilma Holm; Brynjar O Jensson; Aslaug Jonasdottir; Kamilla S Josefsdottir; Thordur Kristjansson; Droplaug N Magnusdottir; Louise le Roux; Gudrun Sigmundsdottir; Gardar Sveinbjornsson; Kristin E Sveinsdottir; Maney Sveinsdottir; Emil A Thorarensen; Bjarni Thorbjornsson; Gisli Masson; Ingileif Jonsdottir; Alma Moller; Thorolfur Gudnason; Karl G Kristinsson; Unnur Thorsteinsdottir; Kari Stefansson
EPI_ISL_417616 hCoV-19/Iceland/288/2020 Europe / Iceland / Reykjavik 2020-03-18 The National University Hospital of Iceland deCODE genetics Daniel F Gudbjartsson; Agnar Helgason; Hakon Jonsson; Olafur T Magnusson; Pall Melsted; Gudmundur L Norddahl; Jona Saemundsdottir; Asgeir Sigurdsson; Patrick Sulem; Arna B Agustsdottir; Berglind Eiriksdottir; Run Fridriksdottir; Elisabet E Gardarsdottir; Gudmundur Georgsson; Olafia S Gretarsdottir; Kjartan R Gudmundsson; Thora R Gunnarsdottir; Arnaldur Gylfason; Hilma Holm; Brynjar O Jensson; Aslaug Jonasdottir; Kamilla S Josefsdottir; Thordur Kristjansson; Droplaug N Magnusdottir; Louise le Roux; Gudrun Sigmundsdottir; Gardar Sveinbjornsson; Kristin E Sveinsdottir; Maney Sveinsdottir; Emil A Thorarensen; Bjarni Thorbjornsson; Gisli Masson; Ingileif Jonsdottir; Alma Moller; Thorolfur Gudnason; Karl G Kristinsson; Unnur Thorsteinsdottir; Kari Stefansson
EPI_ISL_417618 hCoV-19/Iceland/29/2020 Europe / Iceland / Reykjavik 2020-03-16 deCODE genetics deCODE genetics Daniel F Gudbjartsson; Agnar Helgason; Hakon Jonsson; Olafur T Magnusson; Pall Melsted;

Gudmundur L Norddahl; Jona Saemundsdottir; Asgeir Sigurdsson; Patrick Sulem; Arna B Agustsdottir; Berglind Eiriksdottir; Run Fridriksdottir; Elisabet E Gardarsdottir; Gudmundur Georgsson; Olafia S Gretarsdottir; Kjartan R Gudmundsson; Thora R Gunnarsdottir; Arnaldur Gylfason; Hilma Holm; Brynjar O Jensson; Aslaug Jonasdottir; Kamilla S Josefsdottir; Thordur Kristjansson; Droplaug N Magnusdottir; Louise le Roux; Gudrun Sigmundsdottir; Gardar Sveinbjornsson; Kristin E Sveinsdottir; Maney Sveinsdottir; Emil A Thorarensen; Bjarni Thorbjornsson; Gisli Masson; Ingileif Jonsdottir; Alma Moller; Thorolfur Gudnason; Karl G Kristinsson; Unnur Thorsteinsdottir; Kari Stefansson
EPI_ISL_417619 hCoV-19/Iceland/290/2020 Europe / Iceland / Reykjavik 2020-03-18 The National University Hospital of Iceland deCODE genetics Daniel F Gudbjartsson; Agnar Helgason; Hakon Jonsson; Olafur T Magnusson; Pall Melsted; Gudmundur L Norddahl; Jona Saemundsdottir; Asgeir Sigurdsson; Patrick Sulem; Arna B Agustsdottir; Berglind Eiriksdottir; Run Fridriksdottir; Elisabet E Gardarsdottir; Gudmundur Georgsson; Olafia S Gretarsdottir; Kjartan R Gudmundsson; Thora R Gunnarsdottir; Arnaldur Gylfason; Hilma Holm; Brynjar O Jensson; Aslaug Jonasdottir; Kamilla S Josefsdottir; Thordur Kristjansson; Droplaug N Magnusdottir; Louise le Roux; Gudrun Sigmundsdottir; Gardar Sveinbjornsson; Kristin E Sveinsdottir; Maney Sveinsdottir; Emil A Thorarensen; Bjarni Thorbjornsson; Gisli Masson; Ingileif Jonsdottir; Alma Moller; Thorolfur Gudnason; Karl G Kristinsson; Unnur Thorsteinsdottir; Kari Stefansson
EPI_ISL_417620 hCoV-19/Iceland/291/2020 Europe / Iceland / Reykjavik 2020-03-18 The National University Hospital of Iceland deCODE genetics Daniel F Gudbjartsson; Agnar Helgason; Hakon Jonsson; Olafur T Magnusson; Pall Melsted; Gudmundur L Norddahl; Jona Saemundsdottir; Asgeir Sigurdsson; Patrick Sulem; Arna B Agustsdottir; Berglind Eiriksdottir; Run Fridriksdottir; Elisabet E Gardarsdottir; Gudmundur Georgsson; Olafia S Gretarsdottir; Kjartan R Gudmundsson; Thora R Gunnarsdottir; Arnaldur Gylfason; Hilma Holm; Brynjar O Jensson; Aslaug Jonasdottir; Kamilla S Josefsdottir; Thordur Kristjansson; Droplaug N Magnusdottir; Louise le Roux; Gudrun Sigmundsdottir; Gardar Sveinbjornsson; Kristin E Sveinsdottir; Maney Sveinsdottir; Emil A Thorarensen; Bjarni Thorbjornsson; Gisli Masson; Ingileif Jonsdottir; Alma Moller; Thorolfur Gudnason; Karl G Kristinsson; Unnur Thorsteinsdottir; Kari Stefansson
EPI_ISL_417621 hCoV-19/Iceland/292/2020 Europe / Iceland / Reykjavik 2020-03-18 The National University Hospital of Iceland deCODE genetics Daniel F Gudbjartsson; Agnar Helgason; Hakon Jonsson; Olafur T Magnusson; Pall Melsted; Gudmundur L Norddahl; Jona Saemundsdottir; Asgeir Sigurdsson; Patrick Sulem; Arna B Agustsdottir; Berglind Eiriksdottir; Run Fridriksdottir; Elisabet E Gardarsdottir; Gudmundur Georgsson; Olafia S Gretarsdottir; Kjartan R Gudmundsson; Thora R Gunnarsdottir; Arnaldur Gylfason; Hilma Holm; Brynjar O Jensson; Aslaug Jonasdottir; Kamilla S Josefsdottir; Thordur Kristjansson; Droplaug N Magnusdottir; Louise le Roux; Gudrun Sigmundsdottir; Gardar Sveinbjornsson; Kristin E Sveinsdottir; Maney Sveinsdottir; Emil A Thorarensen; Bjarni Thorbjornsson; Gisli Masson; Ingileif Jonsdottir; Alma Moller; Thorolfur Gudnason; Karl G Kristinsson; Unnur Thorsteinsdottir; Kari Stefansson
EPI_ISL_417622 hCoV-19/Iceland/293/2020 Europe / Iceland / Reykjavik 2020-03-18 The National University Hospital of Iceland deCODE genetics Daniel F Gudbjartsson; Agnar Helgason; Hakon Jonsson; Olafur T Magnusson; Pall Melsted; Gudmundur L Norddahl; Jona Saemundsdottir; Asgeir Sigurdsson; Patrick Sulem; Arna B Agustsdottir; Berglind Eiriksdottir; Run Fridriksdottir; Elisabet E Gardarsdottir; Gudmundur Georgsson; Olafia S Gretarsdottir; Kjartan R Gudmundsson; Thora R Gunnarsdottir; Arnaldur Gylfason; Hilma Holm; Brynjar O Jensson; Aslaug Jonasdottir; Kamilla S Josefsdottir; Thordur Kristjansson; Droplaug N Magnusdottir; Louise le Roux; Gudrun Sigmundsdottir; Gardar Sveinbjornsson; Kristin E Sveinsdottir; Maney Sveinsdottir; Emil A Thorarensen; Bjarni Thorbjornsson; Gisli Masson; Ingileif Jonsdottir; Alma Moller; Thorolfur Gudnason; Karl G Kristinsson; Unnur Thorsteinsdottir; Kari Stefansson
EPI_ISL_417623 hCoV-19/Iceland/294/2020 Europe / Iceland / Reykjavik 2020-03-18 The National University Hospital of Iceland deCODE genetics Daniel F Gudbjartsson; Agnar Helgason; Hakon Jonsson; Olafur T Magnusson; Pall Melsted; Gudmundur L Norddahl; Jona Saemundsdottir; Asgeir Sigurdsson; Patrick Sulem; Arna B Agustsdottir; Berglind Eiriksdottir; Run Fridriksdottir; Elisabet E Gardarsdottir; Gudmundur Georgsson; Olafia S Gretarsdottir; Kjartan R Gudmundsson; Thora R Gunnarsdottir; Arnaldur Gylfason; Hilma Holm; Brynjar O Jensson; Aslaug Jonasdottir; Kamilla S Josefsdottir; Thordur Kristjansson; Droplaug N Magnusdottir; Louise le Roux; Gudrun Sigmundsdottir; Gardar Sveinbjornsson; Kristin E Sveinsdottir; Maney Sveinsdottir; Emil A Thorarensen; Bjarni Thorbjornsson; Gisli Masson; Ingileif Jonsdottir; Alma Moller; Thorolfur Gudnason; Karl G Kristinsson; Unnur Thorsteinsdottir; Kari Stefansson
EPI_ISL_417625 hCoV-19/Iceland/296/2020 Europe / Iceland / Reykjavik 2020-03-18 The National University Hospital of Iceland deCODE genetics Daniel F Gudbjartsson; Agnar Helgason; Hakon Jonsson; Olafur T Magnusson; Pall Melsted; Gudmundur L Norddahl; Jona Saemundsdottir; Asgeir Sigurdsson; Patrick Sulem; Arna B Agustsdottir; Berglind Eiriksdottir; Run Fridriksdottir; Elisabet E Gardarsdottir; Gudmundur Georgsson; Olafia S Gretarsdottir; Kjartan R Gudmundsson; Thora R Gunnarsdottir; Arnaldur Gylfason; Hilma Holm; Brynjar O Jensson; Aslaug Jonasdottir; Kamilla S Josefsdottir; Thordur Kristjansson; Droplaug N Magnusdottir; Louise le Roux; Gudrun Sigmundsdottir; Gardar Sveinbjornsson; Kristin E Sveinsdottir; Maney Sveinsdottir; Emil A Thorarensen; Bjarni Thorbjornsson; Gisli Masson; Ingileif Jonsdottir; Alma Moller; Thorolfur Gudnason; Karl G Kristinsson; Unnur Thorsteinsdottir; Kari Stefansson
EPI_ISL_417626 hCoV-19/Iceland/297/2020 Europe / Iceland / Reykjavik 2020-03-18 The National University Hospital of Iceland deCODE genetics Daniel F Gudbjartsson; Agnar Helgason; Hakon Jonsson; Olafur T Magnusson; Pall Melsted; Gudmundur L Norddahl; Jona Saemundsdottir; Asgeir Sigurdsson; Patrick Sulem; Arna B Agustsdottir; Berglind Eiriksdottir; Run Fridriksdottir; Elisabet E Gardarsdottir; Gudmundur Georgsson; Olafia S Gretarsdottir; Kjartan R Gudmundsson; Thora R Gunnarsdottir; Arnaldur Gylfason; Hilma Holm; Brynjar O Jensson; Aslaug Jonasdottir; Kamilla S Josefsdottir; Thordur Kristjansson; Droplaug N Magnusdottir; Louise le Roux; Gudrun Sigmundsdottir; Gardar Sveinbjornsson; Kristin E Sveinsdottir; Maney Sveinsdottir; Emil A Thorarensen; Bjarni Thorbjornsson; Gisli Masson; Ingileif Jonsdottir; Alma Moller; Thorolfur Gudnason; Karl G Kristinsson; Unnur Thorsteinsdottir; Kari Stefansson

EPI_ISL_417627 hCoV-19/Iceland/298/2020 Europe / Iceland / Reykjavik 2020-03-18 The National University Hospital of Iceland deCODE genetics Daniel F Gudbjartsson; Agnar Helgason; Hakon Jonsson; Olafur T Magnusson; Pall Melsted; Gudmundur L Norddahl; Jona Saemundsdottir; Asgeir Sigurdsson; Patrick Sulem; Arna B Agustsdottir; Berglind Eiriksdottir; Run Fridriksdottir; Elisabet E Gardarsdottir; Gudmundur Georgsson; Olafia S Gretarsdottir; Kjartan R Gudmundsson; Thora R Gunnarsdottir; Arnaldur Gylfason; Hilma Holm; Brynjar O Jensson; Aslaug Jonasdottir; Kamilla S Josefsdottir; Thordur Kristjansson; Droplaug N Magnusdottir; Louise le Roux; Gudrun Sigmundsdottir; Gardar Sveinbjornsson; Kristin E Sveinsdottir; Maney Sveinsdottir; Emil A Thorarensen; Bjarni Thorbjornsson; Gisli Masson; Ingileif Jonsdottir; Alma Moller; Thorolfur Gudnason; Karl G Kristinsson; Unnur Thorsteinsdottir; Kari Stefansson
EPI_ISL_417628 hCoV-19/Iceland/299/2020 Europe / Iceland / Reykjavik 2020-03-18 The National University Hospital of Iceland deCODE genetics Daniel F Gudbjartsson; Agnar Helgason; Hakon Jonsson; Olafur T Magnusson; Pall Melsted; Gudmundur L Norddahl; Jona Saemundsdottir; Asgeir Sigurdsson; Patrick Sulem; Arna B Agustsdottir; Berglind Eiriksdottir; Run Fridriksdottir; Elisabet E Gardarsdottir; Gudmundur Georgsson; Olafia S Gretarsdottir; Kjartan R Gudmundsson; Thora R Gunnarsdottir; Arnaldur Gylfason; Hilma Holm; Brynjar O Jensson; Aslaug Jonasdottir; Kamilla S Josefsdottir; Thordur Kristjansson; Droplaug N Magnusdottir; Louise le Roux; Gudrun Sigmundsdottir; Gardar Sveinbjornsson; Kristin E Sveinsdottir; Maney Sveinsdottir; Emil A Thorarensen; Bjarni Thorbjornsson; Gisli Masson; Ingileif Jonsdottir; Alma Moller; Thorolfur Gudnason; Karl G Kristinsson; Unnur Thorsteinsdottir; Kari Stefansson
EPI_ISL_417629 hCoV-19/Iceland/300/2020 Europe / Iceland / Reykjavik 2020-03-18 The National University Hospital of Iceland deCODE genetics Daniel F Gudbjartsson; Agnar Helgason; Hakon Jonsson; Olafur T Magnusson; Pall Melsted; Gudmundur L Norddahl; Jona Saemundsdottir; Asgeir Sigurdsson; Patrick Sulem; Arna B Agustsdottir; Berglind Eiriksdottir; Run Fridriksdottir; Elisabet E Gardarsdottir; Gudmundur Georgsson; Olafia S Gretarsdottir; Kjartan R Gudmundsson; Thora R Gunnarsdottir; Arnaldur Gylfason; Hilma Holm; Brynjar O Jensson; Aslaug Jonasdottir; Kamilla S Josefsdottir; Thordur Kristjansson; Droplaug N Magnusdottir; Louise le Roux; Gudrun Sigmundsdottir; Gardar Sveinbjornsson; Kristin E Sveinsdottir; Maney Sveinsdottir; Emil A Thorarensen; Bjarni Thorbjornsson; Gisli Masson; Ingileif Jonsdottir; Alma Moller; Thorolfur Gudnason; Karl G Kristinsson; Unnur Thorsteinsdottir; Kari Stefansson
EPI_ISL_417630 hCoV-19/Iceland/301/2020 Europe / Iceland / Reykjavik 2020-03-18 The National University Hospital of Iceland deCODE genetics Daniel F Gudbjartsson; Agnar Helgason; Hakon Jonsson; Olafur T Magnusson; Pall Melsted; Gudmundur L Norddahl; Jona Saemundsdottir; Asgeir Sigurdsson; Patrick Sulem; Arna B Agustsdottir; Berglind Eiriksdottir; Run Fridriksdottir; Elisabet E Gardarsdottir; Gudmundur Georgsson; Olafia S Gretarsdottir; Kjartan R Gudmundsson; Thora R Gunnarsdottir; Arnaldur Gylfason; Hilma Holm; Brynjar O Jensson; Aslaug Jonasdottir; Kamilla S Josefsdottir; Thordur Kristjansson; Droplaug N Magnusdottir; Louise le Roux; Gudrun Sigmundsdottir; Gardar Sveinbjornsson; Kristin E Sveinsdottir; Maney Sveinsdottir; Emil A Thorarensen; Bjarni Thorbjornsson; Gisli Masson; Ingileif Jonsdottir; Alma Moller; Thorolfur Gudnason; Karl G Kristinsson; Unnur Thorsteinsdottir; Kari Stefansson
EPI_ISL_417632 hCoV-19/Iceland/303/2020 Europe / Iceland / Reykjavik 2020-03-18 The National University Hospital of Iceland deCODE genetics Daniel F Gudbjartsson; Agnar Helgason; Hakon Jonsson; Olafur T Magnusson; Pall Melsted; Gudmundur L Norddahl; Jona Saemundsdottir; Asgeir Sigurdsson; Patrick Sulem; Arna B Agustsdottir; Berglind Eiriksdottir; Run Fridriksdottir; Elisabet E Gardarsdottir; Gudmundur Georgsson; Olafia S Gretarsdottir; Kjartan R Gudmundsson; Thora R Gunnarsdottir; Arnaldur Gylfason; Hilma Holm; Brynjar O Jensson; Aslaug Jonasdottir; Kamilla S Josefsdottir; Thordur Kristjansson; Droplaug N Magnusdottir; Louise le Roux; Gudrun Sigmundsdottir; Gardar Sveinbjornsson; Kristin E Sveinsdottir; Maney Sveinsdottir; Emil A Thorarensen; Bjarni Thorbjornsson; Gisli Masson; Ingileif Jonsdottir; Alma Moller; Thorolfur Gudnason; Karl G Kristinsson; Unnur Thorsteinsdottir; Kari Stefansson
EPI_ISL_417633 hCoV-19/Iceland/304/2020 Europe / Iceland / Reykjavik 2020-03-17 The National University Hospital of Iceland deCODE genetics Daniel F Gudbjartsson; Agnar Helgason; Hakon Jonsson; Olafur T Magnusson; Pall Melsted; Gudmundur L Norddahl; Jona Saemundsdottir; Asgeir Sigurdsson; Patrick Sulem; Arna B Agustsdottir; Berglind Eiriksdottir; Run Fridriksdottir; Elisabet E Gardarsdottir; Gudmundur Georgsson; Olafia S Gretarsdottir; Kjartan R Gudmundsson; Thora R Gunnarsdottir; Arnaldur Gylfason; Hilma Holm; Brynjar O Jensson; Aslaug Jonasdottir; Kamilla S Josefsdottir; Thordur Kristjansson; Droplaug N Magnusdottir; Louise le Roux; Gudrun Sigmundsdottir; Gardar Sveinbjornsson; Kristin E Sveinsdottir; Maney Sveinsdottir; Emil A Thorarensen; Bjarni Thorbjornsson; Gisli Masson; Ingileif Jonsdottir; Alma Moller; Thorolfur Gudnason; Karl G Kristinsson; Unnur Thorsteinsdottir; Kari Stefansson
EPI_ISL_417634 hCoV-19/Iceland/305/2020 Europe / Iceland / Reykjavik 2020-03-18 The National University Hospital of Iceland deCODE genetics Daniel F Gudbjartsson; Agnar Helgason; Hakon Jonsson; Olafur T Magnusson; Pall Melsted; Gudmundur L Norddahl; Jona Saemundsdottir; Asgeir Sigurdsson; Patrick Sulem; Arna B Agustsdottir; Berglind Eiriksdottir; Run Fridriksdottir; Elisabet E Gardarsdottir; Gudmundur Georgsson; Olafia S Gretarsdottir; Kjartan R Gudmundsson; Thora R Gunnarsdottir; Arnaldur Gylfason; Hilma Holm; Brynjar O Jensson; Aslaug Jonasdottir; Kamilla S Josefsdottir; Thordur Kristjansson; Droplaug N Magnusdottir; Louise le Roux; Gudrun Sigmundsdottir; Gardar Sveinbjornsson; Kristin E Sveinsdottir; Maney Sveinsdottir; Emil A Thorarensen; Bjarni Thorbjornsson; Gisli Masson; Ingileif Jonsdottir; Alma Moller; Thorolfur Gudnason; Karl G Kristinsson; Unnur Thorsteinsdottir; Kari Stefansson
EPI_ISL_417635 hCoV-19/Iceland/306/2020 Europe / Iceland / Reykjavik 2020-03-18 The National University Hospital of Iceland deCODE genetics Daniel F Gudbjartsson; Agnar Helgason; Hakon Jonsson; Olafur T Magnusson; Pall Melsted; Gudmundur L Norddahl; Jona Saemundsdottir; Asgeir Sigurdsson; Patrick Sulem; Arna B Agustsdottir; Berglind Eiriksdottir; Run Fridriksdottir; Elisabet E Gardarsdottir; Gudmundur Georgsson; Olafia S Gretarsdottir; Kjartan R Gudmundsson; Thora R Gunnarsdottir; Arnaldur Gylfason; Hilma Holm; Brynjar O Jensson; Aslaug Jonasdottir; Kamilla S Josefsdottir; Thordur Kristjansson; Droplaug N Magnusdottir; Louise le Roux; Gudrun Sigmundsdottir; Gardar Sveinbjornsson; Kristin E Sveinsdottir; Maney Sveinsdottir; Emil A

Thorarensen; Bjarni Thorbjornsson; Gisli Masson; Ingileif Jonsdottir; Alma Moller; Thorolfur Gudnason; Karl G Kristinsson; Unnur Thorsteinsdottir; Kari Stefansson
EPI_ISL_417636 hCoV-19/Iceland/307/2020 Europe / Iceland / Reykjavik 2020-03-18 The National University Hospital of Iceland deCODE genetics Daniel F Gudbjartsson; Agnar Helgason; Hakon Jonsson; Olafur T Magnusson; Pall Melsted; Gudmundur L Norddahl; Jona Saemundsdottir; Asgeir Sigurdsson; Patrick Sulem; Arna B Agustsdottir; Berglind Eiriksdottir; Run Fridriksdottir; Elisabet E Gardarsdottir; Gudmundur Georgsson; Olafia S Gretarsdottir; Kjartan R Gudmundsson; Thora R Gunnarsdottir; Arnaldur Gylfason; Hilma Holm; Brynjar O Jensson; Aslaug Jonasdottir; Kamilla S Josefsdottir; Thordur Kristjansson; Droplaug N Magnusdottir; Louise le Roux; Gudrun Sigmundsdottir; Gardar Sveinbjornsson; Kristin E Sveinsdottir; Maney Sveinsdottir; Emil A Thorarensen; Bjarni Thorbjornsson; Gisli Masson; Ingileif Jonsdottir; Alma Moller; Thorolfur Gudnason; Karl G Kristinsson; Unnur Thorsteinsdottir; Kari Stefansson
EPI_ISL_417637 hCoV-19/Iceland/308/2020 Europe / Iceland / Reykjavik 2020-03-18 The National University Hospital of Iceland deCODE genetics Daniel F Gudbjartsson; Agnar Helgason; Hakon Jonsson; Olafur T Magnusson; Pall Melsted; Gudmundur L Norddahl; Jona Saemundsdottir; Asgeir Sigurdsson; Patrick Sulem; Arna B Agustsdottir; Berglind Eiriksdottir; Run Fridriksdottir; Elisabet E Gardarsdottir; Gudmundur Georgsson; Olafia S Gretarsdottir; Kjartan R Gudmundsson; Thora R Gunnarsdottir; Arnaldur Gylfason; Hilma Holm; Brynjar O Jensson; Aslaug Jonasdottir; Kamilla S Josefsdottir; Thordur Kristjansson; Droplaug N Magnusdottir; Louise le Roux; Gudrun Sigmundsdottir; Gardar Sveinbjornsson; Kristin E Sveinsdottir; Maney Sveinsdottir; Emil A Thorarensen; Bjarni Thorbjornsson; Gisli Masson; Ingileif Jonsdottir; Alma Moller; Thorolfur Gudnason; Karl G Kristinsson; Unnur Thorsteinsdottir; Kari Stefansson
EPI_ISL_417639 hCoV-19/Iceland/310/2020 Europe / Iceland / Reykjavik 2020-03-18 The National University Hospital of Iceland deCODE genetics Daniel F Gudbjartsson; Agnar Helgason; Hakon Jonsson; Olafur T Magnusson; Pall Melsted; Gudmundur L Norddahl; Jona Saemundsdottir; Asgeir Sigurdsson; Patrick Sulem; Arna B Agustsdottir; Berglind Eiriksdottir; Run Fridriksdottir; Elisabet E Gardarsdottir; Gudmundur Georgsson; Olafia S Gretarsdottir; Kjartan R Gudmundsson; Thora R Gunnarsdottir; Arnaldur Gylfason; Hilma Holm; Brynjar O Jensson; Aslaug Jonasdottir; Kamilla S Josefsdottir; Thordur Kristjansson; Droplaug N Magnusdottir; Louise le Roux; Gudrun Sigmundsdottir; Gardar Sveinbjornsson; Kristin E Sveinsdottir; Maney Sveinsdottir; Emil A Thorarensen; Bjarni Thorbjornsson; Gisli Masson; Ingileif Jonsdottir; Alma Moller; Thorolfur Gudnason; Karl G Kristinsson; Unnur Thorsteinsdottir; Kari Stefansson
EPI_ISL_417641 hCoV-19/Iceland/312/2020 Europe / Iceland / Reykjavik 2020-03-18 The National University Hospital of Iceland deCODE genetics Daniel F Gudbjartsson; Agnar Helgason; Hakon Jonsson; Olafur T Magnusson; Pall Melsted; Gudmundur L Norddahl; Jona Saemundsdottir; Asgeir Sigurdsson; Patrick Sulem; Arna B Agustsdottir; Berglind Eiriksdottir; Run Fridriksdottir; Elisabet E Gardarsdottir; Gudmundur Georgsson; Olafia S Gretarsdottir; Kjartan R Gudmundsson; Thora R Gunnarsdottir; Arnaldur Gylfason; Hilma Holm; Brynjar O Jensson; Aslaug Jonasdottir; Kamilla S Josefsdottir; Thordur Kristjansson; Droplaug N Magnusdottir; Louise le Roux; Gudrun Sigmundsdottir; Gardar Sveinbjornsson; Kristin E Sveinsdottir; Maney Sveinsdottir; Emil A Thorarensen; Bjarni Thorbjornsson; Gisli Masson; Ingileif Jonsdottir; Alma Moller; Thorolfur Gudnason; Karl G Kristinsson; Unnur Thorsteinsdottir; Kari Stefansson
EPI_ISL_417642 hCoV-19/Iceland/313/2020 Europe / Iceland / Reykjavik 2020-03-18 The National University Hospital of Iceland deCODE genetics Daniel F Gudbjartsson; Agnar Helgason; Hakon Jonsson; Olafur T Magnusson; Pall Melsted; Gudmundur L Norddahl; Jona Saemundsdottir; Asgeir Sigurdsson; Patrick Sulem; Arna B Agustsdottir; Berglind Eiriksdottir; Run Fridriksdottir; Elisabet E Gardarsdottir; Gudmundur Georgsson; Olafia S Gretarsdottir; Kjartan R Gudmundsson; Thora R Gunnarsdottir; Arnaldur Gylfason; Hilma Holm; Brynjar O Jensson; Aslaug Jonasdottir; Kamilla S Josefsdottir; Thordur Kristjansson; Droplaug N Magnusdottir; Louise le Roux; Gudrun Sigmundsdottir; Gardar Sveinbjornsson; Kristin E Sveinsdottir; Maney Sveinsdottir; Emil A Thorarensen; Bjarni Thorbjornsson; Gisli Masson; Ingileif Jonsdottir; Alma Moller; Thorolfur Gudnason; Karl G Kristinsson; Unnur Thorsteinsdottir; Kari Stefansson
EPI_ISL_417643 hCoV-19/Iceland/314/2020 Europe / Iceland / Reykjavik 2020-03-18 The National University Hospital of Iceland deCODE genetics Daniel F Gudbjartsson; Agnar Helgason; Hakon Jonsson; Olafur T Magnusson; Pall Melsted; Gudmundur L Norddahl; Jona Saemundsdottir; Asgeir Sigurdsson; Patrick Sulem; Arna B Agustsdottir; Berglind Eiriksdottir; Run Fridriksdottir; Elisabet E Gardarsdottir; Gudmundur Georgsson; Olafia S Gretarsdottir; Kjartan R Gudmundsson; Thora R Gunnarsdottir; Arnaldur Gylfason; Hilma Holm; Brynjar O Jensson; Aslaug Jonasdottir; Kamilla S Josefsdottir; Thordur Kristjansson; Droplaug N Magnusdottir; Louise le Roux; Gudrun Sigmundsdottir; Gardar Sveinbjornsson; Kristin E Sveinsdottir; Maney Sveinsdottir; Emil A Thorarensen; Bjarni Thorbjornsson; Gisli Masson; Ingileif Jonsdottir; Alma Moller; Thorolfur Gudnason; Karl G Kristinsson; Unnur Thorsteinsdottir; Kari Stefansson
EPI_ISL_417644 hCoV-19/Iceland/315/2020 Europe / Iceland / Reykjavik 2020-03-18 The National University Hospital of Iceland deCODE genetics Daniel F Gudbjartsson; Agnar Helgason; Hakon Jonsson; Olafur T Magnusson; Pall Melsted; Gudmundur L Norddahl; Jona Saemundsdottir; Asgeir Sigurdsson; Patrick Sulem; Arna B Agustsdottir; Berglind Eiriksdottir; Run Fridriksdottir; Elisabet E Gardarsdottir; Gudmundur Georgsson; Olafia S Gretarsdottir; Kjartan R Gudmundsson; Thora R Gunnarsdottir; Arnaldur Gylfason; Hilma Holm; Brynjar O Jensson; Aslaug Jonasdottir; Kamilla S Josefsdottir; Thordur Kristjansson; Droplaug N Magnusdottir; Louise le Roux; Gudrun Sigmundsdottir; Gardar Sveinbjornsson; Kristin E Sveinsdottir; Maney Sveinsdottir; Emil A Thorarensen; Bjarni Thorbjornsson; Gisli Masson; Ingileif Jonsdottir; Alma Moller; Thorolfur Gudnason; Karl G Kristinsson; Unnur Thorsteinsdottir; Kari Stefansson
EPI_ISL_417645 hCoV-19/Iceland/316/2020 Europe / Iceland / Reykjavik 2020-03-18 The National University Hospital of Iceland deCODE genetics Daniel F Gudbjartsson; Agnar Helgason; Hakon Jonsson; Olafur T Magnusson; Pall Melsted; Gudmundur L Norddahl; Jona Saemundsdottir; Asgeir Sigurdsson; Patrick Sulem; Arna B Agustsdottir; Berglind Eiriksdottir; Run Fridriksdottir; Elisabet E Gardarsdottir; Gudmundur Georgsson; Olafia S Gretarsdottir; Kjartan R Gudmundsson; Thora R Gunnarsdottir; Arnaldur Gylfason; Hilma Holm; Brynjar O

Jensson; Aslaug Jonasdottir; Kamilla S Josefsdottir; Thordur Kristjansson; Droplaug N Magnusdottir; Louise le Roux; Gudrun Sigmundsdottir; Gardar Sveinbjornsson; Kristin E Sveinsdottir; Maney Sveinsdottir; Emil A Thorarensen; Bjarni Thorbjornsson; Gisli Masson; Ingileif Jonsdottir; Alma Moller; Thorolfur Gudnason; Karl G Kristinsson; Unnur Thorsteinsdottir; Kari Stefansson
EPI_ISL_417646 hCoV-19/Iceland/317/2020 Europe / Iceland / Reykjavik 2020-03-18 The National University Hospital of Iceland deCODE genetics Daniel F Gudbjartsson; Agnar Helgason; Hakon Jonsson; Olafur T Magnusson; Pall Melsted; Gudmundur L Norddahl; Jona Saemundsdottir; Asgeir Sigurdsson; Patrick Sulem; Arna B Agustsdottir; Berglind Eiriksdottir; Run Fridriksdottir; Elisabet E Gardarsdottir; Gudmundur Georgsson; Olafia S Gretarsdottir; Kjartan R Gudmundsson; Thora R Gunnarsdottir; Arnaldur Gylfason; Hilma Holm; Brynjar O Jensson; Aslaug Jonasdottir; Kamilla S Josefsdottir; Thordur Kristjansson; Droplaug N Magnusdottir; Louise le Roux; Gudrun Sigmundsdottir; Gardar Sveinbjornsson; Kristin E Sveinsdottir; Maney Sveinsdottir; Emil A Thorarensen; Bjarni Thorbjornsson; Gisli Masson; Ingileif Jonsdottir; Alma Moller; Thorolfur Gudnason; Karl G Kristinsson; Unnur Thorsteinsdottir; Kari Stefansson
EPI_ISL_417647 hCoV-19/Iceland/318/2020 Europe / Iceland / Reykjavik 2020-03-18 The National University Hospital of Iceland deCODE genetics Daniel F Gudbjartsson; Agnar Helgason; Hakon Jonsson; Olafur T Magnusson; Pall Melsted; Gudmundur L Norddahl; Jona Saemundsdottir; Asgeir Sigurdsson; Patrick Sulem; Arna B Agustsdottir; Berglind Eiriksdottir; Run Fridriksdottir; Elisabet E Gardarsdottir; Gudmundur Georgsson; Olafia S Gretarsdottir; Kjartan R Gudmundsson; Thora R Gunnarsdottir; Arnaldur Gylfason; Hilma Holm; Brynjar O Jensson; Aslaug Jonasdottir; Kamilla S Josefsdottir; Thordur Kristjansson; Droplaug N Magnusdottir; Louise le Roux; Gudrun Sigmundsdottir; Gardar Sveinbjornsson; Kristin E Sveinsdottir; Maney Sveinsdottir; Emil A Thorarensen; Bjarni Thorbjornsson; Gisli Masson; Ingileif Jonsdottir; Alma Moller; Thorolfur Gudnason; Karl G Kristinsson; Unnur Thorsteinsdottir; Kari Stefansson
EPI_ISL_417649 hCoV-19/Iceland/320/2020 Europe / Iceland / Reykjavik 2020-03-18 The National University Hospital of Iceland deCODE genetics Daniel F Gudbjartsson; Agnar Helgason; Hakon Jonsson; Olafur T Magnusson; Pall Melsted; Gudmundur L Norddahl; Jona Saemundsdottir; Asgeir Sigurdsson; Patrick Sulem; Arna B Agustsdottir; Berglind Eiriksdottir; Run Fridriksdottir; Elisabet E Gardarsdottir; Gudmundur Georgsson; Olafia S Gretarsdottir; Kjartan R Gudmundsson; Thora R Gunnarsdottir; Arnaldur Gylfason; Hilma Holm; Brynjar O Jensson; Aslaug Jonasdottir; Kamilla S Josefsdottir; Thordur Kristjansson; Droplaug N Magnusdottir; Louise le Roux; Gudrun Sigmundsdottir; Gardar Sveinbjornsson; Kristin E Sveinsdottir; Maney Sveinsdottir; Emil A Thorarensen; Bjarni Thorbjornsson; Gisli Masson; Ingileif Jonsdottir; Alma Moller; Thorolfur Gudnason; Karl G Kristinsson; Unnur Thorsteinsdottir; Kari Stefansson
EPI_ISL_417650 hCoV-19/Iceland/321/2020 Europe / Iceland / Reykjavik 2020-03-18 The National University Hospital of Iceland deCODE genetics Daniel F Gudbjartsson; Agnar Helgason; Hakon Jonsson; Olafur T Magnusson; Pall Melsted; Gudmundur L Norddahl; Jona Saemundsdottir; Asgeir Sigurdsson; Patrick Sulem; Arna B Agustsdottir; Berglind Eiriksdottir; Run Fridriksdottir; Elisabet E Gardarsdottir; Gudmundur Georgsson; Olafia S Gretarsdottir; Kjartan R Gudmundsson; Thora R Gunnarsdottir; Arnaldur Gylfason; Hilma Holm; Brynjar O Jensson; Aslaug Jonasdottir; Kamilla S Josefsdottir; Thordur Kristjansson; Droplaug N Magnusdottir; Louise le Roux; Gudrun Sigmundsdottir; Gardar Sveinbjornsson; Kristin E Sveinsdottir; Maney Sveinsdottir; Emil A Thorarensen; Bjarni Thorbjornsson; Gisli Masson; Ingileif Jonsdottir; Alma Moller; Thorolfur Gudnason; Karl G Kristinsson; Unnur Thorsteinsdottir; Kari Stefansson
EPI_ISL_417651 hCoV-19/Iceland/322/2020 Europe / Iceland / Reykjavik 2020-03-18 The National University Hospital of Iceland deCODE genetics Daniel F Gudbjartsson; Agnar Helgason; Hakon Jonsson; Olafur T Magnusson; Pall Melsted; Gudmundur L Norddahl; Jona Saemundsdottir; Asgeir Sigurdsson; Patrick Sulem; Arna B Agustsdottir; Berglind Eiriksdottir; Run Fridriksdottir; Elisabet E Gardarsdottir; Gudmundur Georgsson; Olafia S Gretarsdottir; Kjartan R Gudmundsson; Thora R Gunnarsdottir; Arnaldur Gylfason; Hilma Holm; Brynjar O Jensson; Aslaug Jonasdottir; Kamilla S Josefsdottir; Thordur Kristjansson; Droplaug N Magnusdottir; Louise le Roux; Gudrun Sigmundsdottir; Gardar Sveinbjornsson; Kristin E Sveinsdottir; Maney Sveinsdottir; Emil A Thorarensen; Bjarni Thorbjornsson; Gisli Masson; Ingileif Jonsdottir; Alma Moller; Thorolfur Gudnason; Karl G Kristinsson; Unnur Thorsteinsdottir; Kari Stefansson
EPI_ISL_417652 hCoV-19/Iceland/323/2020 Europe / Iceland / Reykjavik 2020-03-18 The National University Hospital of Iceland deCODE genetics Daniel F Gudbjartsson; Agnar Helgason; Hakon Jonsson; Olafur T Magnusson; Pall Melsted; Gudmundur L Norddahl; Jona Saemundsdottir; Asgeir Sigurdsson; Patrick Sulem; Arna B Agustsdottir; Berglind Eiriksdottir; Run Fridriksdottir; Elisabet E Gardarsdottir; Gudmundur Georgsson; Olafia S Gretarsdottir; Kjartan R Gudmundsson; Thora R Gunnarsdottir; Arnaldur Gylfason; Hilma Holm; Brynjar O Jensson; Aslaug Jonasdottir; Kamilla S Josefsdottir; Thordur Kristjansson; Droplaug N Magnusdottir; Louise le Roux; Gudrun Sigmundsdottir; Gardar Sveinbjornsson; Kristin E Sveinsdottir; Maney Sveinsdottir; Emil A Thorarensen; Bjarni Thorbjornsson; Gisli Masson; Ingileif Jonsdottir; Alma Moller; Thorolfur Gudnason; Karl G Kristinsson; Unnur Thorsteinsdottir; Kari Stefansson
EPI_ISL_417653 hCoV-19/Iceland/324/2020 Europe / Iceland / Reykjavik 2020-03-18 The National University Hospital of Iceland deCODE genetics Daniel F Gudbjartsson; Agnar Helgason; Hakon Jonsson; Olafur T Magnusson; Pall Melsted; Gudmundur L Norddahl; Jona Saemundsdottir; Asgeir Sigurdsson; Patrick Sulem; Arna B Agustsdottir; Berglind Eiriksdottir; Run Fridriksdottir; Elisabet E Gardarsdottir; Gudmundur Georgsson; Olafia S Gretarsdottir; Kjartan R Gudmundsson; Thora R Gunnarsdottir; Arnaldur Gylfason; Hilma Holm; Brynjar O Jensson; Aslaug Jonasdottir; Kamilla S Josefsdottir; Thordur Kristjansson; Droplaug N Magnusdottir; Louise le Roux; Gudrun Sigmundsdottir; Gardar Sveinbjornsson; Kristin E Sveinsdottir; Maney Sveinsdottir; Emil A Thorarensen; Bjarni Thorbjornsson; Gisli Masson; Ingileif Jonsdottir; Alma Moller; Thorolfur Gudnason; Karl G Kristinsson; Unnur Thorsteinsdottir; Kari Stefansson
EPI_ISL_417654 hCoV-19/Iceland/325/2020 Europe / Iceland / Reykjavik 2020-03-18 The National University Hospital of Iceland deCODE genetics Daniel F Gudbjartsson; Agnar Helgason; Hakon Jonsson; Olafur T Magnusson; Pall Melsted; Gudmundur L Norddahl; Jona Saemundsdottir; Asgeir Sigurdsson; Patrick Sulem; Arna B

Agustsdottir; Berglind Eiriksdottir; Run Fridriksdottir; Elisabet E Gardarsdottir; Gudmundur Georgsson; Olafia S Gretarsdottir; Kjartan R Gudmundsson; Thora R Gunnarsdottir; Arnaldur Gylfason; Hilma Holm; Brynjar O Jensson; Aslaug Jonasdottir; Kamilla S Josefsdottir; Thordur Kristjansson; Droplaug N Magnusdottir; Louise le Roux; Gudrun Sigmundsdottir; Gardar Sveinbjornsson; Kristin E Sveinsdottir; Maney Sveinsdottir; Emil A Thorarensen; Bjarni Thorbjornsson; Gisli Masson; Ingileif Jonsdottir; Alma Moller; Thorolfur Gudnason; Karl G Kristinsson; Unnur Thorsteinsdottir; Kari Stefansson
EPI_ISL_417655 hCoV-19/Iceland/326/2020 Europe / Iceland / Reykjavik 2020-03-14 deCODE genetics deCODE genetics Daniel F Gudbjartsson; Agnar Helgason; Hakon Jonsson; Olafur T Magnusson; Pall Melsted; Gudmundur L Norddahl; Jona Saemundsdottir; Asgeir Sigurdsson; Patrick Sulem; Arna B Agustsdottir; Berglind Eiriksdottir; Run Fridriksdottir; Elisabet E Gardarsdottir; Gudmundur Georgsson; Olafia S Gretarsdottir; Kjartan R Gudmundsson; Thora R Gunnarsdottir; Arnaldur Gylfason; Hilma Holm; Brynjar O Jensson; Aslaug Jonasdottir; Kamilla S Josefsdottir; Thordur Kristjansson; Droplaug N Magnusdottir; Louise le Roux; Gudrun Sigmundsdottir; Gardar Sveinbjornsson; Kristin E Sveinsdottir; Maney Sveinsdottir; Emil A Thorarensen; Bjarni Thorbjornsson; Gisli Masson; Ingileif Jonsdottir; Alma Moller; Thorolfur Gudnason; Karl G Kristinsson; Unnur Thorsteinsdottir; Kari Stefansson
EPI_ISL_417659 hCoV-19/Iceland/330/2020 Europe / Iceland / Reykjavik 2020-03-14 deCODE genetics deCODE genetics Daniel F Gudbjartsson; Agnar Helgason; Hakon Jonsson; Olafur T Magnusson; Pall Melsted; Gudmundur L Norddahl; Jona Saemundsdottir; Asgeir Sigurdsson; Patrick Sulem; Arna B Agustsdottir; Berglind Eiriksdottir; Run Fridriksdottir; Elisabet E Gardarsdottir; Gudmundur Georgsson; Olafia S Gretarsdottir; Kjartan R Gudmundsson; Thora R Gunnarsdottir; Arnaldur Gylfason; Hilma Holm; Brynjar O Jensson; Aslaug Jonasdottir; Kamilla S Josefsdottir; Thordur Kristjansson; Droplaug N Magnusdottir; Louise le Roux; Gudrun Sigmundsdottir; Gardar Sveinbjornsson; Kristin E Sveinsdottir; Maney Sveinsdottir; Emil A Thorarensen; Bjarni Thorbjornsson; Gisli Masson; Ingileif Jonsdottir; Alma Moller; Thorolfur Gudnason; Karl G Kristinsson; Unnur Thorsteinsdottir; Kari Stefansson
EPI_ISL_417660 hCoV-19/Iceland/331/2020 Europe / Iceland / Reykjavik 2020-03-14 deCODE genetics deCODE genetics Daniel F Gudbjartsson; Agnar Helgason; Hakon Jonsson; Olafur T Magnusson; Pall Melsted; Gudmundur L Norddahl; Jona Saemundsdottir; Asgeir Sigurdsson; Patrick Sulem; Arna B Agustsdottir; Berglind Eiriksdottir; Run Fridriksdottir; Elisabet E Gardarsdottir; Gudmundur Georgsson; Olafia S Gretarsdottir; Kjartan R Gudmundsson; Thora R Gunnarsdottir; Arnaldur Gylfason; Hilma Holm; Brynjar O Jensson; Aslaug Jonasdottir; Kamilla S Josefsdottir; Thordur Kristjansson; Droplaug N Magnusdottir; Louise le Roux; Gudrun Sigmundsdottir; Gardar Sveinbjornsson; Kristin E Sveinsdottir; Maney Sveinsdottir; Emil A Thorarensen; Bjarni Thorbjornsson; Gisli Masson; Ingileif Jonsdottir; Alma Moller; Thorolfur Gudnason; Karl G Kristinsson; Unnur Thorsteinsdottir; Kari Stefansson
EPI_ISL_417662 hCoV-19/Iceland/333/2020 Europe / Iceland / Reykjavik 2020-03-14 deCODE genetics deCODE genetics Daniel F Gudbjartsson; Agnar Helgason; Hakon Jonsson; Olafur T Magnusson; Pall Melsted; Gudmundur L Norddahl; Jona Saemundsdottir; Asgeir Sigurdsson; Patrick Sulem; Arna B Agustsdottir; Berglind Eiriksdottir; Run Fridriksdottir; Elisabet E Gardarsdottir; Gudmundur Georgsson; Olafia S Gretarsdottir; Kjartan R Gudmundsson; Thora R Gunnarsdottir; Arnaldur Gylfason; Hilma Holm; Brynjar O Jensson; Aslaug Jonasdottir; Kamilla S Josefsdottir; Thordur Kristjansson; Droplaug N Magnusdottir; Louise le Roux; Gudrun Sigmundsdottir; Gardar Sveinbjornsson; Kristin E Sveinsdottir; Maney Sveinsdottir; Emil A Thorarensen; Bjarni Thorbjornsson; Gisli Masson; Ingileif Jonsdottir; Alma Moller; Thorolfur Gudnason; Karl G Kristinsson; Unnur Thorsteinsdottir; Kari Stefansson
EPI_ISL_417663 hCoV-19/Iceland/334/2020 Europe / Iceland / Reykjavik 2020-03-14 deCODE genetics deCODE genetics Daniel F Gudbjartsson; Agnar Helgason; Hakon Jonsson; Olafur T Magnusson; Pall Melsted; Gudmundur L Norddahl; Jona Saemundsdottir; Asgeir Sigurdsson; Patrick Sulem; Arna B Agustsdottir; Berglind Eiriksdottir; Run Fridriksdottir; Elisabet E Gardarsdottir; Gudmundur Georgsson; Olafia S Gretarsdottir; Kjartan R Gudmundsson; Thora R Gunnarsdottir; Arnaldur Gylfason; Hilma Holm; Brynjar O Jensson; Aslaug Jonasdottir; Kamilla S Josefsdottir; Thordur Kristjansson; Droplaug N Magnusdottir; Louise le Roux; Gudrun Sigmundsdottir; Gardar Sveinbjornsson; Kristin E Sveinsdottir; Maney Sveinsdottir; Emil A Thorarensen; Bjarni Thorbjornsson; Gisli Masson; Ingileif Jonsdottir; Alma Moller; Thorolfur Gudnason; Karl G Kristinsson; Unnur Thorsteinsdottir; Kari Stefansson
EPI_ISL_417664 hCoV-19/Iceland/335/2020 Europe / Iceland / Reykjavik 2020-03-13 deCODE genetics deCODE genetics Daniel F Gudbjartsson; Agnar Helgason; Hakon Jonsson; Olafur T Magnusson; Pall Melsted; Gudmundur L Norddahl; Jona Saemundsdottir; Asgeir Sigurdsson; Patrick Sulem; Arna B Agustsdottir; Berglind Eiriksdottir; Run Fridriksdottir; Elisabet E Gardarsdottir; Gudmundur Georgsson; Olafia S Gretarsdottir; Kjartan R Gudmundsson; Thora R Gunnarsdottir; Arnaldur Gylfason; Hilma Holm; Brynjar O Jensson; Aslaug Jonasdottir; Kamilla S Josefsdottir; Thordur Kristjansson; Droplaug N Magnusdottir; Louise le Roux; Gudrun Sigmundsdottir; Gardar Sveinbjornsson; Kristin E Sveinsdottir; Maney Sveinsdottir; Emil A Thorarensen; Bjarni Thorbjornsson; Gisli Masson; Ingileif Jonsdottir; Alma Moller; Thorolfur Gudnason; Karl G Kristinsson; Unnur Thorsteinsdottir; Kari Stefansson
EPI_ISL_417665 hCoV-19/Iceland/336/2020 Europe / Iceland / Reykjavik 2020-03-14 deCODE genetics deCODE genetics Daniel F Gudbjartsson; Agnar Helgason; Hakon Jonsson; Olafur T Magnusson; Pall Melsted; Gudmundur L Norddahl; Jona Saemundsdottir; Asgeir Sigurdsson; Patrick Sulem; Arna B Agustsdottir; Berglind Eiriksdottir; Run Fridriksdottir; Elisabet E Gardarsdottir; Gudmundur Georgsson; Olafia S Gretarsdottir; Kjartan R Gudmundsson; Thora R Gunnarsdottir; Arnaldur Gylfason; Hilma Holm; Brynjar O Jensson; Aslaug Jonasdottir; Kamilla S Josefsdottir; Thordur Kristjansson; Droplaug N Magnusdottir; Louise le Roux; Gudrun Sigmundsdottir; Gardar Sveinbjornsson; Kristin E Sveinsdottir; Maney Sveinsdottir; Emil A Thorarensen; Bjarni Thorbjornsson; Gisli Masson; Ingileif Jonsdottir; Alma Moller; Thorolfur Gudnason; Karl G Kristinsson; Unnur Thorsteinsdottir; Kari Stefansson
EPI_ISL_417666 hCoV-19/Iceland/337/2020 Europe / Iceland / Reykjavik 2020-03-15 deCODE genetics

deCODE genetics Daniel F Gudbjartsson; Agnar Helgason; Hakon Jonsson; Olafur T Magnusson; Pall Melsted; Gudmundur L Norddahl; Jona Saemundsdottir; Asgeir Sigurdsson; Patrick Sulem; Arna B Agustsdottir; Berglind Eiriksdottir; Run Fridriksdottir; Elisabet E Gardarsdottir; Gudmundur Georgsson; Olafia S Gretarsdottir; Kjartan R Gudmundsson; Thora R Gunnarsdottir; Arnaldur Gylfason; Hilma Holm; Brynjar O Jensson; Aslaug Jonasdottir; Kamilla S Josefsdottir; Thordur Kristjansson; Droplaug N Magnusdottir; Louise le Roux; Gudrun Sigmundsdottir; Gardar Sveinbjornsson; Kristin E Sveinsdottir; Maney Sveinsdottir; Emil A Thorarensen; Bjarni Thorbjornsson; Gisli Masson; Ingileif Jonsdottir; Alma Moller; Thorolfur Gudnason; Karl G Kristinsson; Unnur Thorsteinsdottir; Kari Stefansson
EPI_ISL_417667 hCoV-19/Iceland/338/2020 Europe / Iceland / Reykjavik 2020-03-15 deCODE genetics deCODE genetics Daniel F Gudbjartsson; Agnar Helgason; Hakon Jonsson; Olafur T Magnusson; Pall Melsted; Gudmundur L Norddahl; Jona Saemundsdottir; Asgeir Sigurdsson; Patrick Sulem; Arna B Agustsdottir; Berglind Eiriksdottir; Run Fridriksdottir; Elisabet E Gardarsdottir; Gudmundur Georgsson; Olafia S Gretarsdottir; Kjartan R Gudmundsson; Thora R Gunnarsdottir; Arnaldur Gylfason; Hilma Holm; Brynjar O Jensson; Aslaug Jonasdottir; Kamilla S Josefsdottir; Thordur Kristjansson; Droplaug N Magnusdottir; Louise le Roux; Gudrun Sigmundsdottir; Gardar Sveinbjornsson; Kristin E Sveinsdottir; Maney Sveinsdottir; Emil A Thorarensen; Bjarni Thorbjornsson; Gisli Masson; Ingileif Jonsdottir; Alma Moller; Thorolfur Gudnason; Karl G Kristinsson; Unnur Thorsteinsdottir; Kari Stefansson
EPI_ISL_417668 hCoV-19/Iceland/339/2020 Europe / Iceland / Reykjavik 2020-03-15 deCODE genetics deCODE genetics Daniel F Gudbjartsson; Agnar Helgason; Hakon Jonsson; Olafur T Magnusson; Pall Melsted; Gudmundur L Norddahl; Jona Saemundsdottir; Asgeir Sigurdsson; Patrick Sulem; Arna B Agustsdottir; Berglind Eiriksdottir; Run Fridriksdottir; Elisabet E Gardarsdottir; Gudmundur Georgsson; Olafia S Gretarsdottir; Kjartan R Gudmundsson; Thora R Gunnarsdottir; Arnaldur Gylfason; Hilma Holm; Brynjar O Jensson; Aslaug Jonasdottir; Kamilla S Josefsdottir; Thordur Kristjansson; Droplaug N Magnusdottir; Louise le Roux; Gudrun Sigmundsdottir; Gardar Sveinbjornsson; Kristin E Sveinsdottir; Maney Sveinsdottir; Emil A Thorarensen; Bjarni Thorbjornsson; Gisli Masson; Ingileif Jonsdottir; Alma Moller; Thorolfur Gudnason; Karl G Kristinsson; Unnur Thorsteinsdottir; Kari Stefansson
EPI_ISL_417670 hCoV-19/Iceland/341/2020 Europe / Iceland / Reykjavik 2020-03-15 deCODE genetics deCODE genetics Daniel F Gudbjartsson; Agnar Helgason; Hakon Jonsson; Olafur T Magnusson; Pall Melsted; Gudmundur L Norddahl; Jona Saemundsdottir; Asgeir Sigurdsson; Patrick Sulem; Arna B Agustsdottir; Berglind Eiriksdottir; Run Fridriksdottir; Elisabet E Gardarsdottir; Gudmundur Georgsson; Olafia S Gretarsdottir; Kjartan R Gudmundsson; Thora R Gunnarsdottir; Arnaldur Gylfason; Hilma Holm; Brynjar O Jensson; Aslaug Jonasdottir; Kamilla S Josefsdottir; Thordur Kristjansson; Droplaug N Magnusdottir; Louise le Roux; Gudrun Sigmundsdottir; Gardar Sveinbjornsson; Kristin E Sveinsdottir; Maney Sveinsdottir; Emil A Thorarensen; Bjarni Thorbjornsson; Gisli Masson; Ingileif Jonsdottir; Alma Moller; Thorolfur Gudnason; Karl G Kristinsson; Unnur Thorsteinsdottir; Kari Stefansson
EPI_ISL_417672 hCoV-19/Iceland/38/2020 Europe / Iceland / Reykjavik 2020-03-16 deCODE genetics deCODE genetics Daniel F Gudbjartsson; Agnar Helgason; Hakon Jonsson; Olafur T Magnusson; Pall Melsted; Gudmundur L Norddahl; Jona Saemundsdottir; Asgeir Sigurdsson; Patrick Sulem; Arna B Agustsdottir; Berglind Eiriksdottir; Run Fridriksdottir; Elisabet E Gardarsdottir; Gudmundur Georgsson; Olafia S Gretarsdottir; Kjartan R Gudmundsson; Thora R Gunnarsdottir; Arnaldur Gylfason; Hilma Holm; Brynjar O Jensson; Aslaug Jonasdottir; Kamilla S Josefsdottir; Thordur Kristjansson; Droplaug N Magnusdottir; Louise le Roux; Gudrun Sigmundsdottir; Gardar Sveinbjornsson; Kristin E Sveinsdottir; Maney Sveinsdottir; Emil A Thorarensen; Bjarni Thorbjornsson; Gisli Masson; Ingileif Jonsdottir; Alma Moller; Thorolfur Gudnason; Karl G Kristinsson; Unnur Thorsteinsdottir; Kari Stefansson
EPI_ISL_417676 hCoV-19/Iceland/98/2020 Europe / Iceland / Reykjavik 2020-03-16 deCODE genetics deCODE genetics Daniel F Gudbjartsson; Agnar Helgason; Hakon Jonsson; Olafur T Magnusson; Pall Melsted; Gudmundur L Norddahl; Jona Saemundsdottir; Asgeir Sigurdsson; Patrick Sulem; Arna B Agustsdottir; Berglind Eiriksdottir; Run Fridriksdottir; Elisabet E Gardarsdottir; Gudmundur Georgsson; Olafia S Gretarsdottir; Kjartan R Gudmundsson; Thora R Gunnarsdottir; Arnaldur Gylfason; Hilma Holm; Brynjar O Jensson; Aslaug Jonasdottir; Kamilla S Josefsdottir; Thordur Kristjansson; Droplaug N Magnusdottir; Louise le Roux; Gudrun Sigmundsdottir; Gardar Sveinbjornsson; Kristin E Sveinsdottir; Maney Sveinsdottir; Emil A Thorarensen; Bjarni Thorbjornsson; Gisli Masson; Ingileif Jonsdottir; Alma Moller; Thorolfur Gudnason; Karl G Kristinsson; Unnur Thorsteinsdottir; Kari Stefansson
EPI_ISL_417677 hCoV-19/Iceland/39/2020 Europe / Iceland / Reykjavik 2020-03-04 The National University Hospital of Iceland deCODE genetics Daniel F Gudbjartsson; Agnar Helgason; Hakon Jonsson; Olafur T Magnusson; Pall Melsted; Gudmundur L Norddahl; Jona Saemundsdottir; Asgeir Sigurdsson; Patrick Sulem; Arna B Agustsdottir; Berglind Eiriksdottir; Run Fridriksdottir; Elisabet E Gardarsdottir; Gudmundur Georgsson; Olafia S Gretarsdottir; Kjartan R Gudmundsson; Thora R Gunnarsdottir; Arnaldur Gylfason; Hilma Holm; Brynjar O Jensson; Aslaug Jonasdottir; Kamilla S Josefsdottir; Thordur Kristjansson; Droplaug N Magnusdottir; Louise le Roux; Gudrun Sigmundsdottir; Gardar Sveinbjornsson; Kristin E Sveinsdottir; Maney Sveinsdottir; Emil A Thorarensen; Bjarni Thorbjornsson; Gisli Masson; Ingileif Jonsdottir; Alma Moller; Thorolfur Gudnason; Karl G Kristinsson; Unnur Thorsteinsdottir; Kari Stefansson
EPI_ISL_417678 hCoV-19/Iceland/185/2020 Europe / Iceland / Reykjavik 2020-03-16 The National University Hospital of Iceland deCODE genetics Daniel F Gudbjartsson; Agnar Helgason; Hakon Jonsson; Olafur T Magnusson; Pall Melsted; Gudmundur L Norddahl; Jona Saemundsdottir; Asgeir Sigurdsson; Patrick Sulem; Arna B Agustsdottir; Berglind Eiriksdottir; Run Fridriksdottir; Elisabet E Gardarsdottir; Gudmundur Georgsson; Olafia S Gretarsdottir; Kjartan R Gudmundsson; Thora R Gunnarsdottir; Arnaldur Gylfason; Hilma Holm; Brynjar O Jensson; Aslaug Jonasdottir; Kamilla S Josefsdottir; Thordur Kristjansson; Droplaug N Magnusdottir; Louise le Roux; Gudrun Sigmundsdottir; Gardar Sveinbjornsson; Kristin E Sveinsdottir; Maney Sveinsdottir; Emil A Thorarensen; Bjarni Thorbjornsson; Gisli Masson; Ingileif Jonsdottir; Alma Moller; Thorolfur Gudnason; Karl G

Kristinsson; Unnur Thorsteinsdottir; Kari Stefansson
EPI_ISL_417679 hCoV-19/Iceland/79/2020 Europe / Iceland / Reykjavik 2020-03-09 The National University Hospital of Iceland deCODE genetics Daniel F Gudbjartsson; Agnar Helgason; Hakon Jonsson; Olafur T Magnusson; Pall Melsted; Gudmundur L Norddahl; Jona Saemundsdottir; Asgeir Sigurdsson; Patrick Sulem; Arna B Agustsdottir; Berglind Eiriksdottir; Run Fridriksdottir; Elisabet E Gardarsdottir; Gudmundur Georgsson; Olafia S Gretarsdottir; Kjartan R Gudmundsson; Thora R Gunnarsdottir; Arnaldur Gylfason; Hilma Holm; Brynjar O Jensson; Aslaug Jonasdottir; Kamilla S Josefsdottir; Thordur Kristjansson; Droplaug N Magnusdottir; Louise le Roux; Gudrun Sigmundsdottir; Gardar Sveinbjornsson; Kristin E Sveinsdottir; Maney Sveinsdottir; Emil A Thorarensen; Bjarni Thorbjornsson; Gisli Masson; Ingileif Jonsdottir; Alma Moller; Thorolfur Gudnason; Karl G Kristinsson; Unnur Thorsteinsdottir; Kari Stefansson
EPI_ISL_417680 hCoV-19/Iceland/177/2020 Europe / Iceland / Reykjavik 2020-03-15 The National University Hospital of Iceland deCODE genetics Daniel F Gudbjartsson; Agnar Helgason; Hakon Jonsson; Olafur T Magnusson; Pall Melsted; Gudmundur L Norddahl; Jona Saemundsdottir; Asgeir Sigurdsson; Patrick Sulem; Arna B Agustsdottir; Berglind Eiriksdottir; Run Fridriksdottir; Elisabet E Gardarsdottir; Gudmundur Georgsson; Olafia S Gretarsdottir; Kjartan R Gudmundsson; Thora R Gunnarsdottir; Arnaldur Gylfason; Hilma Holm; Brynjar O Jensson; Aslaug Jonasdottir; Kamilla S Josefsdottir; Thordur Kristjansson; Droplaug N Magnusdottir; Louise le Roux; Gudrun Sigmundsdottir; Gardar Sveinbjornsson; Kristin E Sveinsdottir; Maney Sveinsdottir; Emil A Thorarensen; Bjarni Thorbjornsson; Gisli Masson; Ingileif Jonsdottir; Alma Moller; Thorolfur Gudnason; Karl G Kristinsson; Unnur Thorsteinsdottir; Kari Stefansson
EPI_ISL_417681 hCoV-19/Iceland/37/2020 Europe / Iceland / Reykjavik 2020-03-04 The National University Hospital of Iceland deCODE genetics Daniel F Gudbjartsson; Agnar Helgason; Hakon Jonsson; Olafur T Magnusson; Pall Melsted; Gudmundur L Norddahl; Jona Saemundsdottir; Asgeir Sigurdsson; Patrick Sulem; Arna B Agustsdottir; Berglind Eiriksdottir; Run Fridriksdottir; Elisabet E Gardarsdottir; Gudmundur Georgsson; Olafia S Gretarsdottir; Kjartan R Gudmundsson; Thora R Gunnarsdottir; Arnaldur Gylfason; Hilma Holm; Brynjar O Jensson; Aslaug Jonasdottir; Kamilla S Josefsdottir; Thordur Kristjansson; Droplaug N Magnusdottir; Louise le Roux; Gudrun Sigmundsdottir; Gardar Sveinbjornsson; Kristin E Sveinsdottir; Maney Sveinsdottir; Emil A Thorarensen; Bjarni Thorbjornsson; Gisli Masson; Ingileif Jonsdottir; Alma Moller; Thorolfur Gudnason; Karl G Kristinsson; Unnur Thorsteinsdottir; Kari Stefansson
EPI_ISL_417682 hCoV-19/Iceland/101/2020 Europe / Iceland / Reykjavik 2020-03-10 The National University Hospital of Iceland deCODE genetics Daniel F Gudbjartsson; Agnar Helgason; Hakon Jonsson; Olafur T Magnusson; Pall Melsted; Gudmundur L Norddahl; Jona Saemundsdottir; Asgeir Sigurdsson; Patrick Sulem; Arna B Agustsdottir; Berglind Eiriksdottir; Run Fridriksdottir; Elisabet E Gardarsdottir; Gudmundur Georgsson; Olafia S Gretarsdottir; Kjartan R Gudmundsson; Thora R Gunnarsdottir; Arnaldur Gylfason; Hilma Holm; Brynjar O Jensson; Aslaug Jonasdottir; Kamilla S Josefsdottir; Thordur Kristjansson; Droplaug N Magnusdottir; Louise le Roux; Gudrun Sigmundsdottir; Gardar Sveinbjornsson; Kristin E Sveinsdottir; Maney Sveinsdottir; Emil A Thorarensen; Bjarni Thorbjornsson; Gisli Masson; Ingileif Jonsdottir; Alma Moller; Thorolfur Gudnason; Karl G Kristinsson; Unnur Thorsteinsdottir; Kari Stefansson
EPI_ISL_417683 hCoV-19/Iceland/47/2020 Europe / Iceland / Reykjavik 2020-03-05 The National University Hospital of Iceland deCODE genetics Daniel F Gudbjartsson; Agnar Helgason; Hakon Jonsson; Olafur T Magnusson; Pall Melsted; Gudmundur L Norddahl; Jona Saemundsdottir; Asgeir Sigurdsson; Patrick Sulem; Arna B Agustsdottir; Berglind Eiriksdottir; Run Fridriksdottir; Elisabet E Gardarsdottir; Gudmundur Georgsson; Olafia S Gretarsdottir; Kjartan R Gudmundsson; Thora R Gunnarsdottir; Arnaldur Gylfason; Hilma Holm; Brynjar O Jensson; Aslaug Jonasdottir; Kamilla S Josefsdottir; Thordur Kristjansson; Droplaug N Magnusdottir; Louise le Roux; Gudrun Sigmundsdottir; Gardar Sveinbjornsson; Kristin E Sveinsdottir; Maney Sveinsdottir; Emil A Thorarensen; Bjarni Thorbjornsson; Gisli Masson; Ingileif Jonsdottir; Alma Moller; Thorolfur Gudnason; Karl G Kristinsson; Unnur Thorsteinsdottir; Kari Stefansson
EPI_ISL_417684 hCoV-19/Iceland/54/2020 Europe / Iceland / Reykjavik 2020-03-06 The National University Hospital of Iceland deCODE genetics Daniel F Gudbjartsson; Agnar Helgason; Hakon Jonsson; Olafur T Magnusson; Pall Melsted; Gudmundur L Norddahl; Jona Saemundsdottir; Asgeir Sigurdsson; Patrick Sulem; Arna B Agustsdottir; Berglind Eiriksdottir; Run Fridriksdottir; Elisabet E Gardarsdottir; Gudmundur Georgsson; Olafia S Gretarsdottir; Kjartan R Gudmundsson; Thora R Gunnarsdottir; Arnaldur Gylfason; Hilma Holm; Brynjar O Jensson; Aslaug Jonasdottir; Kamilla S Josefsdottir; Thordur Kristjansson; Droplaug N Magnusdottir; Louise le Roux; Gudrun Sigmundsdottir; Gardar Sveinbjornsson; Kristin E Sveinsdottir; Maney Sveinsdottir; Emil A Thorarensen; Bjarni Thorbjornsson; Gisli Masson; Ingileif Jonsdottir; Alma Moller; Thorolfur Gudnason; Karl G Kristinsson; Unnur Thorsteinsdottir; Kari Stefansson
EPI_ISL_417685 hCoV-19/Iceland/215/2020 Europe / Iceland / Reykjavik 2020-03-16 The National University Hospital of Iceland deCODE genetics Daniel F Gudbjartsson; Agnar Helgason; Hakon Jonsson; Olafur T Magnusson; Pall Melsted; Gudmundur L Norddahl; Jona Saemundsdottir; Asgeir Sigurdsson; Patrick Sulem; Arna B Agustsdottir; Berglind Eiriksdottir; Run Fridriksdottir; Elisabet E Gardarsdottir; Gudmundur Georgsson; Olafia S Gretarsdottir; Kjartan R Gudmundsson; Thora R Gunnarsdottir; Arnaldur Gylfason; Hilma Holm; Brynjar O Jensson; Aslaug Jonasdottir; Kamilla S Josefsdottir; Thordur Kristjansson; Droplaug N Magnusdottir; Louise le Roux; Gudrun Sigmundsdottir; Gardar Sveinbjornsson; Kristin E Sveinsdottir; Maney Sveinsdottir; Emil A Thorarensen; Bjarni Thorbjornsson; Gisli Masson; Ingileif Jonsdottir; Alma Moller; Thorolfur Gudnason; Karl G Kristinsson; Unnur Thorsteinsdottir; Kari Stefansson
EPI_ISL_417688 hCoV-19/Iceland/12/2020 Europe / Iceland / Reykjavik 2020-03-01 The National University Hospital of Iceland deCODE genetics Daniel F Gudbjartsson; Agnar Helgason; Hakon Jonsson; Olafur T Magnusson; Pall Melsted; Gudmundur L Norddahl; Jona Saemundsdottir; Asgeir Sigurdsson; Patrick Sulem; Arna B Agustsdottir; Berglind Eiriksdottir; Run Fridriksdottir; Elisabet E Gardarsdottir; Gudmundur Georgsson; Olafia S Gretarsdottir; Kjartan R Gudmundsson; Thora R Gunnarsdottir; Arnaldur Gylfason; Hilma Holm; Brynjar O Jensson; Aslaug Jonasdottir; Kamilla S Josefsdottir; Thordur Kristjansson; Droplaug N Magnusdottir; Louise le

Roux; Gudrun Sigmundsdottir; Gardar Sveinbjornsson; Kristin E Sveinsdottir; Maney Sveinsdottir; Emil A Thorarensen; Bjarni Thorbjornsson; Gisli Masson; Ingileif Jonsdottir; Alma Moller; Thorolfur Gudnason; Karl G Kristinsson; Unnur Thorsteinsdottir; Kari Stefansson
EPI_ISL_417690 hCoV-19/Iceland/125/2020 Europe / Iceland / Reykjavik 2020-03-11 The National University Hospital of Iceland deCODE genetics Daniel F Gudbjartsson; Agnar Helgason; Hakon Jonsson; Olafur T Magnusson; Pall Melsted; Gudmundur L Norddahl; Jona Saemundsdottir; Asgeir Sigurdsson; Patrick Sulem; Arna B Agustsdottir; Berglind Eiriksdottir; Run Fridriksdottir; Elisabet E Gardarsdottir; Gudmundur Georgsson; Olafia S Gretarsdottir; Kjartan R Gudmundsson; Thora R Gunnarsdottir; Arnaldur Gylfason; Hilma Holm; Brynjar O Jensson; Aslaug Jonasdottir; Kamilla S Josefsdottir; Thordur Kristjansson; Droplaug N Magnusdottir; Louise le Roux; Gudrun Sigmundsdottir; Gardar Sveinbjornsson; Kristin E Sveinsdottir; Maney Sveinsdottir; Emil A Thorarensen; Bjarni Thorbjornsson; Gisli Masson; Ingileif Jonsdottir; Alma Moller; Thorolfur Gudnason; Karl G Kristinsson; Unnur Thorsteinsdottir; Kari Stefansson
EPI_ISL_417691 hCoV-19/Iceland/111/2020 Europe / Iceland / Reykjavik 2020-03-11 The National University Hospital of Iceland deCODE genetics Daniel F Gudbjartsson; Agnar Helgason; Hakon Jonsson; Olafur T Magnusson; Pall Melsted; Gudmundur L Norddahl; Jona Saemundsdottir; Asgeir Sigurdsson; Patrick Sulem; Arna B Agustsdottir; Berglind Eiriksdottir; Run Fridriksdottir; Elisabet E Gardarsdottir; Gudmundur Georgsson; Olafia S Gretarsdottir; Kjartan R Gudmundsson; Thora R Gunnarsdottir; Arnaldur Gylfason; Hilma Holm; Brynjar O Jensson; Aslaug Jonasdottir; Kamilla S Josefsdottir; Thordur Kristjansson; Droplaug N Magnusdottir; Louise le Roux; Gudrun Sigmundsdottir; Gardar Sveinbjornsson; Kristin E Sveinsdottir; Maney Sveinsdottir; Emil A Thorarensen; Bjarni Thorbjornsson; Gisli Masson; Ingileif Jonsdottir; Alma Moller; Thorolfur Gudnason; Karl G Kristinsson; Unnur Thorsteinsdottir; Kari Stefansson
EPI_ISL_417692 hCoV-19/Iceland/57/2020 Europe / Iceland / Reykjavik 2020-03-06 The National University Hospital of Iceland deCODE genetics Daniel F Gudbjartsson; Agnar Helgason; Hakon Jonsson; Olafur T Magnusson; Pall Melsted; Gudmundur L Norddahl; Jona Saemundsdottir; Asgeir Sigurdsson; Patrick Sulem; Arna B Agustsdottir; Berglind Eiriksdottir; Run Fridriksdottir; Elisabet E Gardarsdottir; Gudmundur Georgsson; Olafia S Gretarsdottir; Kjartan R Gudmundsson; Thora R Gunnarsdottir; Arnaldur Gylfason; Hilma Holm; Brynjar O Jensson; Aslaug Jonasdottir; Kamilla S Josefsdottir; Thordur Kristjansson; Droplaug N Magnusdottir; Louise le Roux; Gudrun Sigmundsdottir; Gardar Sveinbjornsson; Kristin E Sveinsdottir; Maney Sveinsdottir; Emil A Thorarensen; Bjarni Thorbjornsson; Gisli Masson; Ingileif Jonsdottir; Alma Moller; Thorolfur Gudnason; Karl G Kristinsson; Unnur Thorsteinsdottir; Kari Stefansson
EPI_ISL_417693 hCoV-19/Iceland/25/2020 Europe / Iceland / Reykjavik 2020-03-02 The National University Hospital of Iceland deCODE genetics Daniel F Gudbjartsson; Agnar Helgason; Hakon Jonsson; Olafur T Magnusson; Pall Melsted; Gudmundur L Norddahl; Jona Saemundsdottir; Asgeir Sigurdsson; Patrick Sulem; Arna B Agustsdottir; Berglind Eiriksdottir; Run Fridriksdottir; Elisabet E Gardarsdottir; Gudmundur Georgsson; Olafia S Gretarsdottir; Kjartan R Gudmundsson; Thora R Gunnarsdottir; Arnaldur Gylfason; Hilma Holm; Brynjar O Jensson; Aslaug Jonasdottir; Kamilla S Josefsdottir; Thordur Kristjansson; Droplaug N Magnusdottir; Louise le Roux; Gudrun Sigmundsdottir; Gardar Sveinbjornsson; Kristin E Sveinsdottir; Maney Sveinsdottir; Emil A Thorarensen; Bjarni Thorbjornsson; Gisli Masson; Ingileif Jonsdottir; Alma Moller; Thorolfur Gudnason; Karl G Kristinsson; Unnur Thorsteinsdottir; Kari Stefansson
EPI_ISL_417694 hCoV-19/Iceland/3/2020 Europe / Iceland / Reykjavik 2020-03-13 The National University Hospital of Iceland deCODE genetics Daniel F Gudbjartsson; Agnar Helgason; Hakon Jonsson; Olafur T Magnusson; Pall Melsted; Gudmundur L Norddahl; Jona Saemundsdottir; Asgeir Sigurdsson; Patrick Sulem; Arna B Agustsdottir; Berglind Eiriksdottir; Run Fridriksdottir; Elisabet E Gardarsdottir; Gudmundur Georgsson; Olafia S Gretarsdottir; Kjartan R Gudmundsson; Thora R Gunnarsdottir; Arnaldur Gylfason; Hilma Holm; Brynjar O Jensson; Aslaug Jonasdottir; Kamilla S Josefsdottir; Thordur Kristjansson; Droplaug N Magnusdottir; Louise le Roux; Gudrun Sigmundsdottir; Gardar Sveinbjornsson; Kristin E Sveinsdottir; Maney Sveinsdottir; Emil A Thorarensen; Bjarni Thorbjornsson; Gisli Masson; Ingileif Jonsdottir; Alma Moller; Thorolfur Gudnason; Karl G Kristinsson; Unnur Thorsteinsdottir; Kari Stefansson
EPI_ISL_417695 hCoV-19/Iceland/20/2020 Europe / Iceland / Reykjavik 2020-03-02 The National University Hospital of Iceland deCODE genetics Daniel F Gudbjartsson; Agnar Helgason; Hakon Jonsson; Olafur T Magnusson; Pall Melsted; Gudmundur L Norddahl; Jona Saemundsdottir; Asgeir Sigurdsson; Patrick Sulem; Arna B Agustsdottir; Berglind Eiriksdottir; Run Fridriksdottir; Elisabet E Gardarsdottir; Gudmundur Georgsson; Olafia S Gretarsdottir; Kjartan R Gudmundsson; Thora R Gunnarsdottir; Arnaldur Gylfason; Hilma Holm; Brynjar O Jensson; Aslaug Jonasdottir; Kamilla S Josefsdottir; Thordur Kristjansson; Droplaug N Magnusdottir; Louise le Roux; Gudrun Sigmundsdottir; Gardar Sveinbjornsson; Kristin E Sveinsdottir; Maney Sveinsdottir; Emil A Thorarensen; Bjarni Thorbjornsson; Gisli Masson; Ingileif Jonsdottir; Alma Moller; Thorolfur Gudnason; Karl G Kristinsson; Unnur Thorsteinsdottir; Kari Stefansson
EPI_ISL_417696 hCoV-19/Iceland/135/2020 Europe / Iceland / Reykjavik 2020-03-12 The National University Hospital of Iceland deCODE genetics Daniel F Gudbjartsson; Agnar Helgason; Hakon Jonsson; Olafur T Magnusson; Pall Melsted; Gudmundur L Norddahl; Jona Saemundsdottir; Asgeir Sigurdsson; Patrick Sulem; Arna B Agustsdottir; Berglind Eiriksdottir; Run Fridriksdottir; Elisabet E Gardarsdottir; Gudmundur Georgsson; Olafia S Gretarsdottir; Kjartan R Gudmundsson; Thora R Gunnarsdottir; Arnaldur Gylfason; Hilma Holm; Brynjar O Jensson; Aslaug Jonasdottir; Kamilla S Josefsdottir; Thordur Kristjansson; Droplaug N Magnusdottir; Louise le Roux; Gudrun Sigmundsdottir; Gardar Sveinbjornsson; Kristin E Sveinsdottir; Maney Sveinsdottir; Emil A Thorarensen; Bjarni Thorbjornsson; Gisli Masson; Ingileif Jonsdottir; Alma Moller; Thorolfur Gudnason; Karl G Kristinsson; Unnur Thorsteinsdottir; Kari Stefansson
EPI_ISL_417698 hCoV-19/Iceland/103/2020 Europe / Iceland / Reykjavik 2020-03-10 The National University Hospital of Iceland deCODE genetics Daniel F Gudbjartsson; Agnar Helgason; Hakon Jonsson; Olafur T Magnusson; Pall Melsted; Gudmundur L Norddahl; Jona Saemundsdottir; Asgeir Sigurdsson; Patrick Sulem; Arna B Agustsdottir; Berglind Eiriksdottir; Run Fridriksdottir; Elisabet E Gardarsdottir; Gudmundur Georgsson; Olafia

S Gretarsdottir; Kjartan R Gudmundsson; Thora R Gunnarsdottir; Arnaldur Gylfason; Hilma Holm; Brynjar O Jensson; Aslaug Jonasdottir; Kamilla S Josefsdottir; Thordur Kristjansson; Droplaug N Magnusdottir; Louise le Roux; Gudrun Sigmundsdottir; Gardar Sveinbjornsson; Kristin E Sveinsdottir; Maney Sveinsdottir; Emil A Thorarensen; Bjarni Thorbjornsson; Gisli Masson; Ingileif Jonsdottir; Alma Moller; Thorolfur Gudnason; Karl G Kristinsson; Unnur Thorsteinsdottir; Kari Stefansson
EPI_ISL_417699 hCoV-19/Iceland/206/2020 Europe / Iceland / Reykjavik 2020-03-16 The National University Hospital of Iceland deCODE genetics Daniel F Gudbjartsson; Agnar Helgason; Hakon Jonsson; Olafur T Magnusson; Pall Melsted; Gudmundur L Norddahl; Jona Saemundsdottir; Asgeir Sigurdsson; Patrick Sulem; Arna B Agustsdottir; Berglind Eiriksdottir; Run Fridriksdottir; Elisabet E Gardarsdottir; Gudmundur Georgsson; Olafia S Gretarsdottir; Kjartan R Gudmundsson; Thora R Gunnarsdottir; Arnaldur Gylfason; Hilma Holm; Brynjar O Jensson; Aslaug Jonasdottir; Kamilla S Josefsdottir; Thordur Kristjansson; Droplaug N Magnusdottir; Louise le Roux; Gudrun Sigmundsdottir; Gardar Sveinbjornsson; Kristin E Sveinsdottir; Maney Sveinsdottir; Emil A Thorarensen; Bjarni Thorbjornsson; Gisli Masson; Ingileif Jonsdottir; Alma Moller; Thorolfur Gudnason; Karl G Kristinsson; Unnur Thorsteinsdottir; Kari Stefansson
EPI_ISL_418816 hCoV-19/Canada/BC_0443574/2020 North America / Canada / British Columbia 2020-03-11 BCCDC Public Health Laboratory BCCDC Public Health Laboratory Harrigan, Prystajecky, Krajden, Lee, Kamelian, Lapointe, Choi, Hoang, Sekirov, Levett, Tyson, Snutch, Loman, Quick, Li, Gilmour
EPI_ISL_418817 hCoV-19/Canada/BC_0554880/2020 North America / Canada / British Columbia 2020-03-13 BCCDC Public Health Laboratory BCCDC Public Health Laboratory Harrigan, Prystajecky, Krajden, Lee, Kamelian, Lapointe, Choi, Hoang, Sekirov, Levett, Tyson, Snutch, Loman, Quick, Li, Gilmour
EPI_ISL_418818 hCoV-19/Canada/BC_1318414/2020 North America / Canada / British Columbia 2020-03-06 BCCDC Public Health Laboratory BCCDC Public Health Laboratory Harrigan, Prystajecky, Krajden, Lee, Kamelian, Lapointe, Choi, Hoang, Sekirov, Levett, Tyson, Snutch, Loman, Quick, Li, Gilmour
EPI_ISL_418838 hCoV-19/Canada/BC_6404889/2020 North America / Canada / British Columbia 2020-03-12 BCCDC Public Health Laboratory BCCDC Public Health Laboratory Harrigan, Prystajecky, Krajden, Lee, Kamelian, Lapointe, Choi, Hoang, Sekirov, Levett, Tyson, Snutch, Loman, Quick, Li, Gilmour
EPI_ISL_418839 hCoV-19/Canada/BC_6489864/2020 North America / Canada / British Columbia 2020-03-10 BCCDC Public Health Laboratory BCCDC Public Health Laboratory Harrigan, Prystajecky, Krajden, Lee, Kamelian, Lapointe, Choi, Hoang, Sekirov, Levett, Tyson, Snutch, Loman, Quick, Li, Gilmour
EPI_ISL_418840 hCoV-19/Canada/BC_6502001/2020 North America / Canada / British Columbia 2020-03-12 BCCDC Public Health Laboratory BCCDC Public Health Laboratory Harrigan, Prystajecky, Krajden, Lee, Kamelian, Lapointe, Choi, Hoang, Sekirov, Levett, Tyson, Snutch, Loman, Quick, Li, Gilmour
EPI_ISL_418841 hCoV-19/Canada/BC_6981299/2020 North America / Canada / British Columbia 2020-03-11 BCCDC Public Health Laboratory BCCDC Public Health Laboratory Harrigan, Prystajecky, Krajden, Lee, Kamelian, Lapointe, Choi, Hoang, Sekirov, Levett, Tyson, Snutch, Loman, Quick, Li, Gilmour
EPI_ISL_418842 hCoV-19/Canada/BC_6997898/2020 North America / Canada / British Columbia 2020-03-11 BCCDC Public Health Laboratory BCCDC Public Health Laboratory Harrigan, Prystajecky, Krajden, Lee, Kamelian, Lapointe, Choi, Hoang, Sekirov, Levett, Tyson, Snutch, Loman, Quick, Li, Gilmour
EPI_ISL_418843 hCoV-19/Canada/BC_7277381/2020 North America / Canada / British Columbia 2020-03-12 BCCDC Public Health Laboratory BCCDC Public Health Laboratory Harrigan, Prystajecky, Krajden, Lee, Kamelian, Lapointe, Choi, Hoang, Sekirov, Levett, Tyson, Snutch, Loman, Quick, Li, Gilmour
EPI_ISL_418845 hCoV-19/Canada/BC_7515663/2020 North America / Canada / British Columbia 2020-03-11 BCCDC Public Health Laboratory BCCDC Public Health Laboratory Harrigan, Prystajecky, Krajden, Lee, Kamelian, Lapointe, Choi, Hoang, Sekirov, Levett, Tyson, Snutch, Loman, Quick, Li, Gilmour
EPI_ISL_418846 hCoV-19/Canada/BC_7553799/2020 North America / Canada / British Columbia 2020-03-12 BCCDC Public Health Laboratory BCCDC Public Health Laboratory Harrigan, Prystajecky, Krajden, Lee, Kamelian, Lapointe, Choi, Hoang, Sekirov, Levett, Tyson, Snutch, Loman, Quick, Li, Gilmour
EPI_ISL_418847 hCoV-19/Canada/BC_8150537/2020 North America / Canada / British Columbia 2020-03-12 BCCDC Public Health Laboratory BCCDC Public Health Laboratory Harrigan, Prystajecky, Krajden, Lee, Kamelian, Lapointe, Choi, Hoang, Sekirov, Levett, Tyson, Snutch, Loman, Quick, Li, Gilmour
EPI_ISL_418848 hCoV-19/Canada/BC_8159203/2020 North America / Canada / British Columbia 2020-03-05 BCCDC Public Health Laboratory BCCDC Public Health Laboratory Harrigan, Prystajecky, Krajden, Lee, Kamelian, Lapointe, Choi, Hoang, Sekirov, Levett, Tyson, Snutch, Loman, Quick, Li, Gilmour
EPI_ISL_418849 hCoV-19/Canada/BC_8486790/2020 North America / Canada / British Columbia 2020-03-13 BCCDC Public Health Laboratory BCCDC Public Health Laboratory Harrigan, Prystajecky, Krajden, Lee, Kamelian, Lapointe, Choi, Hoang, Sekirov, Levett, Tyson, Snutch, Loman, Quick, Li, Gilmour
EPI_ISL_418851 hCoV-19/Canada/BC_8622445/2020 North America / Canada / British Columbia 2020-03-13 BCCDC Public Health Laboratory BCCDC Public Health Laboratory Harrigan, Prystajecky, Krajden, Lee, Kamelian, Lapointe, Choi, Hoang, Sekirov, Levett, Tyson, Snutch, Loman, Quick, Li, Gilmour
EPI_ISL_418852 hCoV-19/Canada/BC_8718874/2020 North America / Canada / British Columbia 2020-03-07 BCCDC Public Health Laboratory BCCDC Public Health Laboratory Harrigan, Prystajecky, Krajden, Lee, Kamelian, Lapointe, Choi, Hoang, Sekirov, Levett, Tyson, Snutch, Loman, Quick, Li, Gilmour
EPI_ISL_418853 hCoV-19/Canada/BC_8894200/2020 North America / Canada / British Columbia 2020-03-11 BCCDC Public Health Laboratory BCCDC Public Health Laboratory Harrigan, Prystajecky, Krajden, Lee, Kamelian, Lapointe, Choi, Hoang, Sekirov, Levett, Tyson, Snutch, Loman, Quick, Li, Gilmour
EPI_ISL_418854 hCoV-19/Canada/BC_8896915/2020 North America / Canada / British Columbia 2020-03-11 BCCDC Public Health Laboratory BCCDC Public Health Laboratory Harrigan, Prystajecky, Krajden, Lee, Kamelian, Lapointe, Choi, Hoang, Sekirov, Levett, Tyson, Snutch, Loman, Quick, Li, Gilmour
EPI_ISL_418855 hCoV-19/Canada/BC_8897642/2020 North America / Canada / British Columbia 2020-03-11 BCCDC Public Health Laboratory BCCDC Public Health Laboratory Harrigan, Prystajecky, Krajden, Lee, Kamelian,

Lapointe, Choi, Hoang, Sekirov, Levett, Tyson, Snutch, Loman, Quick, Li, Gilmour
EPI_ISL_418856 hCoV-19/Canada/BC_9345042/2020 North America / Canada / British Columbia 2020-03-13 BCCDC Public Health Laboratory BCCDC Public Health Laboratory Harrigan, Prystajecky, Krajden, Lee, Kamelian, Lapointe, Choi, Hoang, Sekirov, Levett, Tyson, Snutch, Loman, Quick, Li, Gilmour
EPI_ISL_418857 hCoV-19/Canada/BC_9345715/2020 North America / Canada / British Columbia 2020-03-13 BCCDC Public Health Laboratory BCCDC Public Health Laboratory Harrigan, Prystajecky, Krajden, Lee, Kamelian, Lapointe, Choi, Hoang, Sekirov, Levett, Tyson, Snutch, Loman, Quick, Li, Gilmour
EPI_ISL_418858 hCoV-19/Canada/BC_9446031/2020 North America / Canada / British Columbia 2020-03-13 BCCDC Public Health Laboratory BCCDC Public Health Laboratory Harrigan, Prystajecky, Krajden, Lee, Kamelian, Lapointe, Choi, Hoang, Sekirov, Levett, Tyson, Snutch, Loman, Quick, Li, Gilmour
EPI_ISL_418900 hCoV-19/USA/WA-UW333/2020 North America / USA / Washington 2020-03-16 UW Virology Lab UW Virology Lab Pavitra Roychoudhury, Hong Xie, Keith Jerome, Alexander Greninger
EPI_ISL_418901 hCoV-19/USA/MN-UW334/2020 North America / USA / Minnesota 2020-03-13 UW Virology Lab UW Virology Lab Pavitra Roychoudhury, Hong Xie, Keith Jerome, Alexander Greninger
EPI_ISL_418902 hCoV-19/USA/MN-UW335/2020 North America / USA / Minnesota 2020-03-13 UW Virology Lab UW Virology Lab Pavitra Roychoudhury, Hong Xie, Keith Jerome, Alexander Greninger
EPI_ISL_418903 hCoV-19/USA/WA-UW336/2020 North America / USA / Washington 2020-03-16 UW Virology Lab UW Virology Lab Pavitra Roychoudhury, Hong Xie, Keith Jerome, Alexander Greninger
EPI_ISL_418904 hCoV-19/USA/WA-UW337/2020 North America / USA / Washington 2020-03-16 UW Virology Lab UW Virology Lab Pavitra Roychoudhury, Hong Xie, Keith Jerome, Alexander Greninger
EPI_ISL_418905 hCoV-19/USA/WA-UW338/2020 North America / USA / Washington 2020-03-16 UW Virology Lab UW Virology Lab Pavitra Roychoudhury, Hong Xie, Keith Jerome, Alexander Greninger
EPI_ISL_418906 hCoV-19/USA/WA-UW339/2020 North America / USA / Washington 2020-03-16 UW Virology Lab UW Virology Lab Pavitra Roychoudhury, Hong Xie, Keith Jerome, Alexander Greninger
EPI_ISL_418907 hCoV-19/USA/WA-UW340/2020 North America / USA / Washington 2020-03-16 UW Virology Lab UW Virology Lab Pavitra Roychoudhury, Hong Xie, Keith Jerome, Alexander Greninger
EPI_ISL_418908 hCoV-19/USA/WA-UW341/2020 North America / USA / Washington 2020-03-16 UW Virology Lab UW Virology Lab Pavitra Roychoudhury, Hong Xie, Keith Jerome, Alexander Greninger
EPI_ISL_418909 hCoV-19/USA/WA-UW342/2020 North America / USA / Washington 2020-03-16 UW Virology Lab UW Virology Lab Pavitra Roychoudhury, Hong Xie, Keith Jerome, Alexander Greninger
EPI_ISL_418910 hCoV-19/USA/WA-UW343/2020 North America / USA / Washington 2020-03-16 UW Virology Lab UW Virology Lab Pavitra Roychoudhury, Hong Xie, Keith Jerome, Alexander Greninger
EPI_ISL_418911 hCoV-19/USA/WA-UW344/2020 North America / USA / Washington 2020-03-16 UW Virology Lab UW Virology Lab Pavitra Roychoudhury, Hong Xie, Keith Jerome, Alexander Greninger
EPI_ISL_418912 hCoV-19/USA/WA-UW345/2020 North America / USA / Washington 2020-03-16 UW Virology Lab UW Virology Lab Pavitra Roychoudhury, Hong Xie, Keith Jerome, Alexander Greninger
EPI_ISL_418913 hCoV-19/USA/WA-UW346/2020 North America / USA / Washington 2020-03-16 UW Virology Lab UW Virology Lab Pavitra Roychoudhury, Hong Xie, Keith Jerome, Alexander Greninger
EPI_ISL_418915 hCoV-19/USA/IL-UW348/2020 North America / USA / Illinois 2020-03-13 UW Virology Lab UW Virology Lab Pavitra Roychoudhury, Hong Xie, Keith Jerome, Alexander Greninger
EPI_ISL_418916 hCoV-19/USA/IL-UW349/2020 North America / USA / Illinois 2020-03-13 UW Virology Lab UW Virology Lab Pavitra Roychoudhury, Hong Xie, Keith Jerome, Alexander Greninger
EPI_ISL_418917 hCoV-19/USA/IL-UW350/2020 North America / USA / Illinois 2020-03-14 UW Virology Lab UW Virology Lab Pavitra Roychoudhury, Hong Xie, Keith Jerome, Alexander Greninger
EPI_ISL_418918 hCoV-19/USA/IL-UW351/2020 North America / USA / Illinois 2020-03-13 UW Virology Lab UW Virology Lab Pavitra Roychoudhury, Hong Xie, Keith Jerome, Alexander Greninger
EPI_ISL_418919 hCoV-19/USA/IL-UW352/2020 North America / USA / Illinois 2020-03-13 UW Virology Lab UW Virology Lab Pavitra Roychoudhury, Hong Xie, Keith Jerome, Alexander Greninger
EPI_ISL_418920 hCoV-19/USA/IL-UW353/2020 North America / USA / Illinois 2020-03-13 UW Virology Lab UW Virology Lab Pavitra Roychoudhury, Hong Xie, Keith Jerome, Alexander Greninger
EPI_ISL_418921 hCoV-19/USA/UN-UW354/2020 North America / USA 2020-03-16 UW Virology Lab UW Virology Lab Pavitra Roychoudhury, Hong Xie, Keith Jerome, Alexander Greninger
EPI_ISL_418922 hCoV-19/USA/WA-UW355/2020 North America / USA / Washington 2020-03-16 UW Virology Lab UW Virology Lab Pavitra Roychoudhury, Hong Xie, Keith Jerome, Alexander Greninger
EPI_ISL_418923 hCoV-19/USA/WA-UW356/2020 North America / USA / Washington 2020-03-16 UW Virology Lab UW Virology Lab Pavitra Roychoudhury, Hong Xie, Keith Jerome, Alexander Greninger
EPI_ISL_418924 hCoV-19/USA/WA-UW357/2020 North America / USA / Washington 2020-03-14 UW Virology Lab UW Virology Lab Pavitra Roychoudhury, Hong Xie, Keith Jerome, Alexander Greninger
EPI_ISL_418925 hCoV-19/USA/UN-UW358/2020 North America / USA 2020-03 UW Virology Lab UW Virology Lab Pavitra Roychoudhury, Hong Xie, Keith Jerome, Alexander Greninger
EPI_ISL_418926 hCoV-19/USA/WA-UW359/2020 North America / USA / Washington 2020-03-14 UW Virology Lab UW Virology Lab Pavitra Roychoudhury, Hong Xie, Keith Jerome, Alexander Greninger
EPI_ISL_418927 hCoV-19/USA/WA-UW360/2020 North America / USA / Washington 2020-03-16 UW Virology Lab UW Virology Lab Pavitra Roychoudhury, Hong Xie, Keith Jerome, Alexander Greninger
EPI_ISL_418928 hCoV-19/USA/WA-UW361/2020 North America / USA / Washington 2020-03-15 UW Virology Lab UW Virology Lab Pavitra Roychoudhury, Hong Xie, Keith Jerome, Alexander Greninger
EPI_ISL_418929 hCoV-19/USA/WA-UW362/2020 North America / USA / Washington 2020-03-17 UW Virology Lab UW Virology Lab Pavitra Roychoudhury, Hong Xie, Keith Jerome, Alexander Greninger
EPI_ISL_418930 hCoV-19/USA/WA-UW363/2020 North America / USA / Washington 2020-03-17 UW Virology Lab UW Virology Lab Pavitra Roychoudhury, Hong Xie, Keith Jerome, Alexander Greninger

EPI_ISL_418931 hCoV-19/USA/WA-UW364/2020 North America / USA / Washington 2020-03-17 UW
Virology Lab UW Virology Lab Pavitra Roychoudhury, Hong Xie, Keith Jerome, Alexander Greninger
EPI_ISL_418932 hCoV-19/USA/WA-UW365/2020 North America / USA / Washington 2020-03-16 UW
Virology Lab UW Virology Lab Pavitra Roychoudhury, Hong Xie, Keith Jerome, Alexander Greninger
EPI_ISL_418933 hCoV-19/USA/WA-UW366/2020 North America / USA / Washington 2020-03-17 UW
Virology Lab UW Virology Lab Pavitra Roychoudhury, Hong Xie, Keith Jerome, Alexander Greninger
EPI_ISL_418934 hCoV-19/USA/WA-UW367/2020 North America / USA / Washington 2020-03-17 UW
Virology Lab UW Virology Lab Pavitra Roychoudhury, Hong Xie, Keith Jerome, Alexander Greninger
EPI_ISL_418935 hCoV-19/USA/WA-UW368/2020 North America / USA / Washington 2020-03-16 UW
Virology Lab UW Virology Lab Pavitra Roychoudhury, Hong Xie, Keith Jerome, Alexander Greninger
EPI_ISL_418936 hCoV-19/USA/WA-UW369/2020 North America / USA / Washington 2020-03-17 UW
Virology Lab UW Virology Lab Pavitra Roychoudhury, Hong Xie, Keith Jerome, Alexander Greninger
EPI_ISL_418937 hCoV-19/USA/WA-UW370/2020 North America / USA / Washington 2020-03-17 UW
Virology Lab UW Virology Lab Pavitra Roychoudhury, Hong Xie, Keith Jerome, Alexander Greninger
EPI_ISL_418938 hCoV-19/USA/WA-UW371/2020 North America / USA / Washington 2020-03-16 UW
Virology Lab UW Virology Lab Pavitra Roychoudhury, Hong Xie, Keith Jerome, Alexander Greninger
EPI_ISL_418939 hCoV-19/USA/WA-UW372/2020 North America / USA / Washington 2020-03-17 UW
Virology Lab UW Virology Lab Pavitra Roychoudhury, Hong Xie, Keith Jerome, Alexander Greninger
EPI_ISL_418940 hCoV-19/USA/WA-UW373/2020 North America / USA / Washington 2020-03-17 UW
Virology Lab UW Virology Lab Pavitra Roychoudhury, Hong Xie, Keith Jerome, Alexander Greninger
EPI_ISL_418941 hCoV-19/USA/WA-UW374/2020 North America / USA 2020-03 UW Virology Lab UW Virology Lab
Pavitra Roychoudhury, Hong Xie, Keith Jerome, Alexander Greninger
EPI_ISL_418942 hCoV-19/USA/WA-UW375/2020 North America / USA / Washington 2020-03-17 UW
Virology Lab UW Virology Lab Pavitra Roychoudhury, Hong Xie, Keith Jerome, Alexander Greninger
EPI_ISL_418943 hCoV-19/USA/WA-UW376/2020 North America / USA / Washington 2020-03-17 UW
Virology Lab UW Virology Lab Pavitra Roychoudhury, Hong Xie, Keith Jerome, Alexander Greninger
EPI_ISL_418944 hCoV-19/USA/WA-UW378/2020 North America / USA / Washington 2020-03-15 UW
Virology Lab UW Virology Lab Pavitra Roychoudhury, Hong Xie, Keith Jerome, Alexander Greninger
EPI_ISL_418945 hCoV-19/USA/WA-UW379/2020 North America / USA / Washington 2020-03-15 UW
Virology Lab UW Virology Lab Pavitra Roychoudhury, Hong Xie, Keith Jerome, Alexander Greninger
EPI_ISL_418948 hCoV-19/USA/WA-UW383/2020 North America / USA / Washington 2020-03-16 UW
Virology Lab UW Virology Lab Pavitra Roychoudhury, Hong Xie, Keith Jerome, Alexander Greninger
EPI_ISL_418949 hCoV-19/USA/WA-UW384/2020 North America / USA / Washington 2020-03-16 UW
Virology Lab UW Virology Lab Pavitra Roychoudhury, Hong Xie, Keith Jerome, Alexander Greninger
EPI_ISL_418950 hCoV-19/USA/WA-UW385/2020 North America / USA / Washington 2020-03-16 UW
Virology Lab UW Virology Lab Pavitra Roychoudhury, Hong Xie, Keith Jerome, Alexander Greninger
EPI_ISL_418951 hCoV-19/USA/WA-UW386/2020 North America / USA / Washington 2020-03-16 UW
Virology Lab UW Virology Lab Pavitra Roychoudhury, Hong Xie, Keith Jerome, Alexander Greninger
EPI_ISL_418952 hCoV-19/USA/WA-UW387/2020 North America / USA / Washington 2020-03-16 UW
Virology Lab UW Virology Lab Pavitra Roychoudhury, Hong Xie, Keith Jerome, Alexander Greninger
EPI_ISL_418953 hCoV-19/USA/WA-UW388/2020 North America / USA / Washington 2020-03-16 UW
Virology Lab UW Virology Lab Pavitra Roychoudhury, Hong Xie, Keith Jerome, Alexander Greninger
EPI_ISL_418954 hCoV-19/USA/WA-UW389/2020 North America / USA / Washington 2020-03-17 UW
Virology Lab UW Virology Lab Pavitra Roychoudhury, Hong Xie, Keith Jerome, Alexander Greninger
EPI_ISL_418955 hCoV-19/USA/WA-UW390/2020 North America / USA / Washington 2020-03-15 UW
Virology Lab UW Virology Lab Pavitra Roychoudhury, Hong Xie, Keith Jerome, Alexander Greninger
EPI_ISL_418956 hCoV-19/USA/VA-DCLS-0002/2020 North America / USA / Virginia 2020-03-17 Virginia DCLS
Virginia DCLS Virginia DCLS
EPI_ISL_418957 hCoV-19/USA/VA-DCLS-0003/2020 North America / USA / Virginia 2020-03-17 Virginia DCLS
Virginia DCLS Virginia DCLS
EPI_ISL_418958 hCoV-19/USA/VA-DCLS-0004/2020 North America / USA / Virginia 2020-03-17 Virginia DCLS
Virginia DCLS Virginia DCLS
EPI_ISL_418962 hCoV-19/USA/UT-019/2020 North America / USA / Utah 2020-03-13 Utah Public Health
Laboratory Utah Public Health Laboratory Erin Young, Kelly Oakeson
EPI_ISL_418963 hCoV-19/USA/UT-021/2020 North America / USA / Utah 2020-03-14 Utah Public Health
Laboratory Utah Public Health Laboratory Erin Young, Kelly Oakeson
EPI_ISL_418964 hCoV-19/USA/UT-022/2020 North America / USA / Utah 2020-03-14 Utah Public Health
Laboratory Utah Public Health Laboratory Erin Young, Kelly Oakeson
EPI_ISL_418965 hCoV-19/USA/UT-023/2020 North America / USA / Utah 2020-03-11 Utah Public Health
Laboratory Utah Public Health Laboratory Erin Young, Kelly Oakeson
EPI_ISL_418967 hCoV-19/USA/UT-025/2020 North America / USA / Utah 2020-03-11 Utah Public Health
Laboratory Utah Public Health Laboratory Erin Young, Kelly Oakeson
EPI_ISL_418972 hCoV-19/USA/NY-NYUMC26/2020 North America / USA / New York / Manhattan 2020-03-17
NYU Langone Health Department of Pathology and Medicine, New York University School of Medicine Maria
Aguero-Rosenfeld, Margaret Black, John Cadley, Paolo Cotzia, John Chen, Dacia Dimartino, Xiaojun Feng, Adriana
Heguy, Megan Hogan, Emily Huang, George Jour, Christian Marier, Matthew T. Maurano, Mark J. Mulligan, Peter
Meyn, Jared Pinnell, Sitharam Ramaswami, Amy Rapkiewicz, Marie Samanovic-Golden, Antonio Serrano, Guomiao Shen,
Matija Snuderl, Nick Vulpescu, Gael Westby, Paul Zappile, Yutong Zhang
EPI_ISL_418973 hCoV-19/USA/NY-NYUMC27/2020 North America / USA / New York / Manhattan 2020-03-17
NYU Langone Health Department of Pathology and Medicine, New York University School of Medicine Maria

Aguero-Rosenfeld, Margaret Black, John Cadley, Paolo Cotzia, John Chen, Dacia Dimartino, Xiaojun Feng, Adriana Heguy, Megan Hogan, Emily Huang, George Jour, Christian Marier, Matthew T. Maurano, Mark J. Mulligan, Peter Meyn, Jared Pinnell, Sitharam Ramaswami, Amy Rapkiewicz, Marie Samanovic-Golden, Antonio Serrano, Guomiao Shen, Matija Snuderl, Nick Vulpescu, Gael Westby, Paul Zappile, Yutong Zhang
EPI_ISL_418974 hCoV-19/USA/NY-NYUMC28/2020 North America / USA / New York / Manhattan 2020-03-17 NYU Langone Health Department of Pathology and Medicine, New York University School of Medicine Maria Aguero-Rosenfeld, Margaret Black, John Cadley, Paolo Cotzia, John Chen, Dacia Dimartino, Xiaojun Feng, Adriana Heguy, Megan Hogan, Emily Huang, George Jour, Christian Marier, Matthew T. Maurano, Mark J. Mulligan, Peter Meyn, Jared Pinnell, Sitharam Ramaswami, Amy Rapkiewicz, Marie Samanovic-Golden, Antonio Serrano, Guomiao Shen, Matija Snuderl, Nick Vulpescu, Gael Westby, Paul Zappile, Yutong Zhang
EPI_ISL_418975 hCoV-19/USA/NY-NYUMC29/2020 North America / USA / New York / Manhattan 2020-03-17 NYU Langone Health Department of Pathology and Medicine, New York University School of Medicine Maria Aguero-Rosenfeld, Margaret Black, John Cadley, Paolo Cotzia, John Chen, Dacia Dimartino, Xiaojun Feng, Adriana Heguy, Megan Hogan, Emily Huang, George Jour, Christian Marier, Matthew T. Maurano, Mark J. Mulligan, Peter Meyn, Jared Pinnell, Sitharam Ramaswami, Amy Rapkiewicz, Marie Samanovic-Golden, Antonio Serrano, Guomiao Shen, Matija Snuderl, Nick Vulpescu, Gael Westby, Paul Zappile, Yutong Zhang
EPI_ISL_418976 hCoV-19/USA/NY-NYUMC30/2020 North America / USA / New York / Manhattan 2020-03-17 NYU Langone Health Department of Pathology and Medicine, New York University School of Medicine Maria Aguero-Rosenfeld, Margaret Black, John Cadley, Paolo Cotzia, John Chen, Dacia Dimartino, Xiaojun Feng, Adriana Heguy, Megan Hogan, Emily Huang, George Jour, Christian Marier, Matthew T. Maurano, Mark J. Mulligan, Peter Meyn, Jared Pinnell, Sitharam Ramaswami, Amy Rapkiewicz, Marie Samanovic-Golden, Antonio Serrano, Guomiao Shen, Matija Snuderl, Nick Vulpescu, Gael Westby, Paul Zappile, Yutong Zhang
EPI_ISL_418977 hCoV-19/USA/NY-NYUMC31/2020 North America / USA / New York / Manhattan 2020-03-17 NYU Langone Health Department of Pathology and Medicine, New York University School of Medicine Maria Aguero-Rosenfeld, Margaret Black, John Cadley, Paolo Cotzia, John Chen, Dacia Dimartino, Xiaojun Feng, Adriana Heguy, Megan Hogan, Emily Huang, George Jour, Christian Marier, Matthew T. Maurano, Mark J. Mulligan, Peter Meyn, Jared Pinnell, Sitharam Ramaswami, Amy Rapkiewicz, Marie Samanovic-Golden, Antonio Serrano, Guomiao Shen, Matija Snuderl, Nick Vulpescu, Gael Westby, Paul Zappile, Yutong Zhang
EPI_ISL_418978 hCoV-19/USA/NY-NYUMC32/2020 North America / USA / New York / Manhattan 2020-03-17 NYU Langone Health Department of Pathology and Medicine, New York University School of Medicine Maria Aguero-Rosenfeld, Margaret Black, John Cadley, Paolo Cotzia, John Chen, Dacia Dimartino, Xiaojun Feng, Adriana Heguy, Megan Hogan, Emily Huang, George Jour, Christian Marier, Matthew T. Maurano, Mark J. Mulligan, Peter Meyn, Jared Pinnell, Sitharam Ramaswami, Amy Rapkiewicz, Marie Samanovic-Golden, Antonio Serrano, Guomiao Shen, Matija Snuderl, Nick Vulpescu, Gael Westby, Paul Zappile, Yutong Zhang
EPI_ISL_418979 hCoV-19/USA/NY-NYUMC33/2020 North America / USA / New York / Manhattan 2020-03-17 NYU Langone Health Department of Pathology and Medicine, New York University School of Medicine Maria Aguero-Rosenfeld, Margaret Black, John Cadley, Paolo Cotzia, John Chen, Dacia Dimartino, Xiaojun Feng, Adriana Heguy, Megan Hogan, Emily Huang, George Jour, Christian Marier, Matthew T. Maurano, Mark J. Mulligan, Peter Meyn, Jared Pinnell, Sitharam Ramaswami, Amy Rapkiewicz, Marie Samanovic-Golden, Antonio Serrano, Guomiao Shen, Matija Snuderl, Nick Vulpescu, Gael Westby, Paul Zappile, Yutong Zhang
EPI_ISL_418980 hCoV-19/USA/NY-NYUMC34/2020 North America / USA / New York / Manhattan 2020-03-17 NYU Langone Health Department of Pathology and Medicine, New York University School of Medicine Maria Aguero-Rosenfeld, Margaret Black, John Cadley, Paolo Cotzia, John Chen, Dacia Dimartino, Xiaojun Feng, Adriana Heguy, Megan Hogan, Emily Huang, George Jour, Christian Marier, Matthew T. Maurano, Mark J. Mulligan, Peter Meyn, Jared Pinnell, Sitharam Ramaswami, Amy Rapkiewicz, Marie Samanovic-Golden, Antonio Serrano, Guomiao Shen, Matija Snuderl, Nick Vulpescu, Gael Westby, Paul Zappile, Yutong Zhang
EPI_ISL_418981 hCoV-19/Belgium/VRAR-030643/2020 Europe / Belgium / Merelbeke 2020-03-06 KU Leuven, Clinical and Epidemiological Virology KU Leuven, Clinical and Epidemiological Virology Bert Vanmechelen, Joan Marti-Carreras, Tony Wawina, Piet Maes
EPI_ISL_418982 hCoV-19/Belgium/HAA-030953/2020 Europe / Belgium / Oevel 2020-03-09 KU Leuven, Clinical and Epidemiological Virology KU Leuven, Clinical and Epidemiological Virology Bert Vanmechelen, Joan Marti-Carreras, Tony Wawina, Piet Maes
EPI_ISL_418983 hCoV-19/Belgium/DWAEE-030954/2020 Europe / Belgium / Asse 2020-03-09 KU Leuven, Clinical and Epidemiological Virology KU Leuven, Clinical and Epidemiological Virology Bert Vanmechelen, Joan Marti-Carreras, Tony Wawina, Piet Maes
EPI_ISL_418984 hCoV-19/Belgium/BG-030955/2020 Europe / Belgium / Kaulile 2020-03-09 KU Leuven, Clinical and Epidemiological Virology KU Leuven, Clinical and Epidemiological Virology Bert Vanmechelen, Joan Marti-Carreras, Tony Wawina, Piet Maes
EPI_ISL_418985 hCoV-19/Belgium/LT-030956/2020 Europe / Belgium / Ellezelles 2020-03-09 KU Leuven, Clinical and Epidemiological Virology KU Leuven, Clinical and Epidemiological Virology Bert Vanmechelen, Joan Marti-Carreras, Tony Wawina, Piet Maes
EPI_ISL_418986 hCoV-19/Belgium/RS-030257/2020 Europe / Belgium / Hoegaarden 2020-03-02 KU Leuven, Clinical and Epidemiological Virology KU Leuven, Clinical and Epidemiological Virology Bert Vanmechelen, Joan Marti-Carreras, Tony Wawina, Piet Maes
EPI_ISL_418987 hCoV-19/Belgium/CG-030158/2020 Europe / Belgium / Eupen 2020-03-01 KU Leuven, Clinical and Epidemiological Virology KU Leuven, Clinical and Epidemiological Virology Bert Vanmechelen, Joan Marti-Carreras, Tony Wawina, Piet Maes
EPI_ISL_418988 hCoV-19/Belgium/JRH-030459/2020 Europe / Belgium / Zolder 2020-03-04 Institute information KU Leuven, Clinical and Epidemiological Virology Institute information KU Leuven, Clinical and Epidemiological Virology Bert Vanmechelen, Joan Marti-Carreras, Tony Wawina, Piet Maes

EPI_ISL_418989 hCoV-19/Belgium/RT-030460/2020 Europe / Belgium / Vorst 2020-03-04 KU Leuven, Clinical and Epidemiological Virology KU Leuven, Clinical and Epidemiological Virology Bert Vanmechelen, Joan Marti-Carreras, Tony Wawina, Piet Maes
EPI_ISL_418990 hCoV-19/Hangzhou/ZJU-010/2020 Asia / China / Hangzhou 2020-02-03 State Key Laboratory for Diagnosis and Treatment of Infectious Diseases, National Clinical Research Center for Infectious Diseases, First Affiliated Hospital, Zhejiang University School of Medicine, Hangzhou, China 310003 State Key Laboratory for Diagnosis and Treatment of Infectious Diseases, National Clinical Research Center for Infectious Diseases, First Affiliated Hospital, Zhejiang University School of Medicine, Hangzhou, China 310003 Hangping Yao, Nanping Wu, Chao Jiang, Xiangyun Lu, Linfang Cheng, Fumin Liu, Zhigang Wu, Haibo Wu, Changzhong Jin, Min Zheng, Lanjuan Li
EPI_ISL_418991 hCoV-19/Hangzhou/ZJU-011/2020 Asia / China / Hangzhou 2020-02-04 State Key Laboratory for Diagnosis and Treatment of Infectious Diseases, National Clinical Research Center for Infectious Diseases, First Affiliated Hospital, Zhejiang University School of Medicine, Hangzhou, China 310003 State Key Laboratory for Diagnosis and Treatment of Infectious Diseases, National Clinical Research Center for Infectious Diseases, First Affiliated Hospital, Zhejiang University School of Medicine, Hangzhou, China 310003 Hangping Yao, Nanping Wu, Chao Jiang, Xiangyun Lu, Linfang Cheng, Fumin Liu, Zhigang Wu, Haibo Wu, Changzhong Jin, Min Zheng, Lanjuan Li
EPI_ISL_418992 hCoV-19/Singapore/17/2020 Asia / Singapore 2020-02-10 National Public Health Laboratory, National Centre for Infectious Diseases National Public Health Laboratory, National Centre for Infectious Diseases Mak TM, Octavia S, Cui L, Lin RTP
EPI_ISL_418993 hCoV-19/Singapore/20/2020 Asia / Singapore 2020-01-29 National Public Health Laboratory, National Centre for Infectious Diseases National Public Health Laboratory, National Centre for Infectious Diseases Mak TM, Octavia S, Cui L, Lin RTP
EPI_ISL_418994 hCoV-19/Singapore/3Clin/2020 Asia / Singapore 2020-01-29 National Public Health Laboratory, National Centre for Infectious Diseases National Public Health Laboratory, National Centre for Infectious Diseases Mak TM, Octavia S, Cui L, Lin RTP
EPI_ISL_418995 hCoV-19/Singapore/12Clin/2020 Asia / Singapore 2020-02-28 National Public Health Laboratory, National Centre for Infectious Diseases National Public Health Laboratory, National Centre for Infectious Diseases Mak TM, Octavia S, Cui L, Lin RTP
EPI_ISL_418996 hCoV-19/Singapore/15/2020 Asia / Singapore 2020-01-27 National Public Health Laboratory, National Centre for Infectious Diseases National Public Health Laboratory, National Centre for Infectious Diseases Mak TM, Octavia S, Cui L, Lin RTP
EPI_ISL_418997 hCoV-19/Singapore/16/2020 Asia / Singapore 2020-02-06 National Public Health Laboratory, National Centre for Infectious Diseases National Public Health Laboratory, National Centre for Infectious Diseases Mak TM, Octavia S, Cui L, Lin RTP
EPI_ISL_418998 hCoV-19/Singapore/14Clin/2020 Asia / Singapore 2020-02-14 National Public Health Laboratory, National Centre for Infectious Diseases National Public Health Laboratory, National Centre for Infectious Diseases Mak TM, Octavia S, Cui L, Lin RTP
EPI_ISL_418999 hCoV-19/Singapore/18/2020 Asia / Singapore 2020-03-01 National Public Health Laboratory, National Centre for Infectious Diseases National Public Health Laboratory, National Centre for Infectious Diseases Mak TM, Octavia S, Cui L, Lin RTP
EPI_ISL_422001 hCoV-19/England/20148004904/2020 Europe / United Kingdom / England 2020-04-03 Respiratory Virus Unit, Microbiology Services Colindale, Public Health England Respiratory Virus Unit, Microbiology Services Colindale, Public Health England Monica Galiano, Shahjahan Miah, Angie Lackenby, Omolola Akinbami, Tiina Talts, Leena Bhaw, Richard Myers, Steven Platt, Kirstin Edwards, Jonathan Hubb, Joanna Ellis, Maria Zambon
EPI_ISL_422002 hCoV-19/England/20148006104/2020 Europe / United Kingdom / England 2020-03-30 Respiratory Virus Unit, Microbiology Services Colindale, Public Health England Respiratory Virus Unit, Microbiology Services Colindale, Public Health England Monica Galiano, Shahjahan Miah, Angie Lackenby, Omolola Akinbami, Tiina Talts, Leena Bhaw, Richard Myers, Steven Platt, Kirstin Edwards, Jonathan Hubb, Joanna Ellis, Maria Zambon
EPI_ISL_422009 hCoV-19/England/20148039404/2020 Europe / United Kingdom / England 2020-04-02 Respiratory Virus Unit, Microbiology Services Colindale, Public Health England Respiratory Virus Unit, Microbiology Services Colindale, Public Health England Monica Galiano, Shahjahan Miah, Angie Lackenby, Omolola Akinbami, Tiina Talts, Leena Bhaw, Richard Myers, Steven Platt, Kirstin Edwards, Jonathan Hubb, Joanna Ellis, Maria Zambon
EPI_ISL_422010 hCoV-19/England/20148041604/2020 Europe / United Kingdom / England 2020-04-02 Respiratory Virus Unit, Microbiology Services Colindale, Public Health England Respiratory Virus Unit, Microbiology Services Colindale, Public Health England Monica Galiano, Shahjahan Miah, Angie Lackenby, Omolola Akinbami, Tiina Talts, Leena Bhaw, Richard Myers, Steven Platt, Kirstin Edwards, Jonathan Hubb, Joanna Ellis, Maria Zambon
EPI_ISL_422016 hCoV-19/Wales/PHWC-26796/2020 Europe / United Kingdom / Wales 2020-03-28 Wales Specialist Virology Centre Public Health Wales Microbiology Cardiff Catherine Moore, Johnathan Evans, Malorie Perry, Simon Cottrell, Alec Birchley, Alexander Adams, Amy Gaskin, Bree Gatica-Wilcox, Jason Coombes, Lauren Gilbert, Lee Graham, Nicole Pacchiarini, Sara Kumziene-Summerhayes, Sarah Taylor, Sophie Jones, Sara Rey, Matthew Bull, Joanne Watkins, Sally Corden, Tom Connor
EPI_ISL_422017 hCoV-19/Wales/PHWC-2711F/2020 Europe / United Kingdom / Wales 2020-03-29 Wales Specialist Virology Centre Public Health Wales Microbiology Cardiff Catherine Moore, Johnathan Evans, Malorie Perry, Simon Cottrell, Alec Birchley, Alexander Adams, Amy Gaskin, Bree Gatica-Wilcox, Jason Coombes, Lauren Gilbert, Lee Graham, Nicole Pacchiarini, Sara Kumziene-Summerhayes, Sarah Taylor, Sophie Jones,

Sara Rey, Matthew Bull, Joanne Watkins, Sally Corden, Tom Connor
EPI_ISL_422018 hCoV-19/Wales/PHWC-2673C/2020 Europe / United Kingdom / Wales 2020-03-28 Wales Specialist Virology Centre Public Health Wales Microbiology Cardiff Catherine Moore, Johnathan Evans, Malorie Perry, Simon Cottrell, Alec Birchley, Alexander Adams, Amy Gaskin, Bree Gatica-Wilcox, Jason Coombes, Lauren Gilbert, Lee Graham, Nicole Pacchiarini, Sara Kumziene-Summerhayes, Sarah Taylor, Sophie Jones, Sara Rey, Matthew Bull, Joanne Watkins, Sally Corden, Tom Connor
EPI_ISL_422019 hCoV-19/Wales/PHWC-265E7/2020 Europe / United Kingdom / Wales 2020-03-27 Wales Specialist Virology Centre Public Health Wales Microbiology Cardiff Catherine Moore, Johnathan Evans, Malorie Perry, Simon Cottrell, Alec Birchley, Alexander Adams, Amy Gaskin, Bree Gatica-Wilcox, Jason Coombes, Lauren Gilbert, Lee Graham, Nicole Pacchiarini, Sara Kumziene-Summerhayes, Sarah Taylor, Sophie Jones, Sara Rey, Matthew Bull, Joanne Watkins, Sally Corden, Tom Connor
EPI_ISL_422020 hCoV-19/Wales/PHWC-2682A/2020 Europe / United Kingdom / Wales 2020-03-27 Wales Specialist Virology Centre Public Health Wales Microbiology Cardiff Catherine Moore, Johnathan Evans, Malorie Perry, Simon Cottrell, Alec Birchley, Alexander Adams, Amy Gaskin, Bree Gatica-Wilcox, Jason Coombes, Lauren Gilbert, Lee Graham, Nicole Pacchiarini, Sara Kumziene-Summerhayes, Sarah Taylor, Sophie Jones, Sara Rey, Matthew Bull, Joanne Watkins, Sally Corden, Tom Connor
EPI_ISL_422021 hCoV-19/Wales/PHWC-2665D/2020 Europe / United Kingdom / Wales 2020-03-27 Wales Specialist Virology Centre Public Health Wales Microbiology Cardiff Catherine Moore, Johnathan Evans, Malorie Perry, Simon Cottrell, Alec Birchley, Alexander Adams, Amy Gaskin, Bree Gatica-Wilcox, Jason Coombes, Lauren Gilbert, Lee Graham, Nicole Pacchiarini, Sara Kumziene-Summerhayes, Sarah Taylor, Sophie Jones, Sara Rey, Matthew Bull, Joanne Watkins, Sally Corden, Tom Connor
EPI_ISL_422022 hCoV-19/Wales/PHWC-26D49/2020 Europe / United Kingdom / Wales 2020-03-30 Wales Specialist Virology Centre Public Health Wales Microbiology Cardiff Catherine Moore, Johnathan Evans, Malorie Perry, Simon Cottrell, Alec Birchley, Alexander Adams, Amy Gaskin, Bree Gatica-Wilcox, Jason Coombes, Lauren Gilbert, Lee Graham, Nicole Pacchiarini, Sara Kumziene-Summerhayes, Sarah Taylor, Sophie Jones, Sara Rey, Matthew Bull, Joanne Watkins, Sally Corden, Tom Connor
EPI_ISL_422023 hCoV-19/Wales/PHWC-26981/2020 Europe / United Kingdom / Wales 2020-03-30 Wales Specialist Virology Centre Public Health Wales Microbiology Cardiff Catherine Moore, Johnathan Evans, Malorie Perry, Simon Cottrell, Alec Birchley, Alexander Adams, Amy Gaskin, Bree Gatica-Wilcox, Jason Coombes, Lauren Gilbert, Lee Graham, Nicole Pacchiarini, Sara Kumziene-Summerhayes, Sarah Taylor, Sophie Jones, Sara Rey, Matthew Bull, Joanne Watkins, Sally Corden, Tom Connor
EPI_ISL_422024 hCoV-19/Wales/PHWC-26B12/2020 Europe / United Kingdom / Wales 2020-03-29 Wales Specialist Virology Centre Public Health Wales Microbiology Cardiff Catherine Moore, Johnathan Evans, Malorie Perry, Simon Cottrell, Alec Birchley, Alexander Adams, Amy Gaskin, Bree Gatica-Wilcox, Jason Coombes, Lauren Gilbert, Lee Graham, Nicole Pacchiarini, Sara Kumziene-Summerhayes, Sarah Taylor, Sophie Jones, Sara Rey, Matthew Bull, Joanne Watkins, Sally Corden, Tom Connor
EPI_ISL_422025 hCoV-19/Wales/PHWC-263DE/2020 Europe / United Kingdom / Wales 2020-03-27 Wales Specialist Virology Centre Public Health Wales Microbiology Cardiff Catherine Moore, Johnathan Evans, Malorie Perry, Simon Cottrell, Alec Birchley, Alexander Adams, Amy Gaskin, Bree Gatica-Wilcox, Jason Coombes, Lauren Gilbert, Lee Graham, Nicole Pacchiarini, Sara Kumziene-Summerhayes, Sarah Taylor, Sophie Jones, Sara Rey, Matthew Bull, Joanne Watkins, Sally Corden, Tom Connor
EPI_ISL_422026 hCoV-19/Wales/PHWC-263FC/2020 Europe / United Kingdom / Wales 2020-03-27 Wales Specialist Virology Centre Public Health Wales Microbiology Cardiff Catherine Moore, Johnathan Evans, Malorie Perry, Simon Cottrell, Alec Birchley, Alexander Adams, Amy Gaskin, Bree Gatica-Wilcox, Jason Coombes, Lauren Gilbert, Lee Graham, Nicole Pacchiarini, Sara Kumziene-Summerhayes, Sarah Taylor, Sophie Jones, Sara Rey, Matthew Bull, Joanne Watkins, Sally Corden, Tom Connor
EPI_ISL_422027 hCoV-19/Wales/PHWC-2672D/2020 Europe / United Kingdom / Wales 2020-03-27 Wales Specialist Virology Centre Public Health Wales Microbiology Cardiff Catherine Moore, Johnathan Evans, Malorie Perry, Simon Cottrell, Alec Birchley, Alexander Adams, Amy Gaskin, Bree Gatica-Wilcox, Jason Coombes, Lauren Gilbert, Lee Graham, Nicole Pacchiarini, Sara Kumziene-Summerhayes, Sarah Taylor, Sophie Jones, Sara Rey, Matthew Bull, Joanne Watkins, Sally Corden, Tom Connor
EPI_ISL_422028 hCoV-19/Wales/PHWC-2631A/2020 Europe / United Kingdom / Wales 2020-03-27 Wales Specialist Virology Centre Public Health Wales Microbiology Cardiff Catherine Moore, Johnathan Evans, Malorie Perry, Simon Cottrell, Alec Birchley, Alexander Adams, Amy Gaskin, Bree Gatica-Wilcox, Jason Coombes, Lauren Gilbert, Lee Graham, Nicole Pacchiarini, Sara Kumziene-Summerhayes, Sarah Taylor, Sophie Jones, Sara Rey, Matthew Bull, Joanne Watkins, Sally Corden, Tom Connor
EPI_ISL_422029 hCoV-19/Wales/PHWC-25F44/2020 Europe / United Kingdom / Wales 2020-03-25 Wales Specialist Virology Centre Public Health Wales Microbiology Cardiff Catherine Moore, Johnathan Evans, Malorie Perry, Simon Cottrell, Alec Birchley, Alexander Adams, Amy Gaskin, Bree Gatica-Wilcox, Jason Coombes, Lauren Gilbert, Lee Graham, Nicole Pacchiarini, Sara Kumziene-Summerhayes, Sarah Taylor, Sophie Jones, Sara Rey, Matthew Bull, Joanne Watkins, Sally Corden, Tom Connor
EPI_ISL_422030 hCoV-19/Wales/PHWC-270B8/2020 Europe / United Kingdom / Wales 2020-03-29 Wales Specialist Virology Centre Public Health Wales Microbiology Cardiff Catherine Moore, Johnathan Evans, Malorie Perry, Simon Cottrell, Alec Birchley, Alexander Adams, Amy Gaskin, Bree Gatica-Wilcox, Jason Coombes, Lauren Gilbert, Lee Graham, Nicole Pacchiarini, Sara Kumziene-Summerhayes, Sarah Taylor, Sophie Jones, Sara Rey, Matthew Bull, Joanne Watkins, Sally Corden, Tom Connor
EPI_ISL_422031 hCoV-19/Wales/PHWC-26DD0/2020 Europe / United Kingdom / Wales 2020-03-30 Wales Specialist Virology Centre Public Health Wales Microbiology Cardiff Catherine Moore, Johnathan Evans, Malorie Perry, Simon Cottrell, Alec Birchley, Alexander Adams, Amy Gaskin, Bree Gatica-Wilcox, Jason Coombes, Lauren Gilbert, Lee Graham, Nicole Pacchiarini, Sara Kumziene-Summerhayes, Sarah Taylor, Sophie Jones,

Sara Rey, Matthew Bull, Joanne Watkins, Sally Corden, Tom Connor
EPI_ISL_422032 hCoV-19/Wales/PHWC-2649F/2020 Europe / United Kingdom / Wales 2020-03-27 Wales Specialist Virology Centre Public Health Wales Microbiology Cardiff Catherine Moore, Johnathan Evans, Malorie Perry, Simon Cottrell, Alec Birchley, Alexander Adams, Amy Gaskin, Bree Gatica-Wilcox, Jason Coombes, Lauren Gilbert, Lee Graham, Nicole Pacchiarini, Sara Kumziene-Summerhayes, Sarah Taylor, Sophie Jones, Sara Rey, Matthew Bull, Joanne Watkins, Sally Corden, Tom Connor
EPI_ISL_422033 hCoV-19/Wales/PHWC-2681B/2020 Europe / United Kingdom / Wales 2020-03-27 Wales Specialist Virology Centre Public Health Wales Microbiology Cardiff Catherine Moore, Johnathan Evans, Malorie Perry, Simon Cottrell, Alec Birchley, Alexander Adams, Amy Gaskin, Bree Gatica-Wilcox, Jason Coombes, Lauren Gilbert, Lee Graham, Nicole Pacchiarini, Sara Kumziene-Summerhayes, Sarah Taylor, Sophie Jones, Sara Rey, Matthew Bull, Joanne Watkins, Sally Corden, Tom Connor
EPI_ISL_422034 hCoV-19/Wales/PHWC-27012/2020 Europe / United Kingdom / Wales 2020-03-28 Wales Specialist Virology Centre Public Health Wales Microbiology Cardiff Catherine Moore, Johnathan Evans, Malorie Perry, Simon Cottrell, Alec Birchley, Alexander Adams, Amy Gaskin, Bree Gatica-Wilcox, Jason Coombes, Lauren Gilbert, Lee Graham, Nicole Pacchiarini, Sara Kumziene-Summerhayes, Sarah Taylor, Sophie Jones, Sara Rey, Matthew Bull, Joanne Watkins, Sally Corden, Tom Connor
EPI_ISL_422035 hCoV-19/Wales/PHWC-26D58/2020 Europe / United Kingdom / Wales 2020-03-30 Wales Specialist Virology Centre Public Health Wales Microbiology Cardiff Catherine Moore, Johnathan Evans, Malorie Perry, Simon Cottrell, Alec Birchley, Alexander Adams, Amy Gaskin, Bree Gatica-Wilcox, Jason Coombes, Lauren Gilbert, Lee Graham, Nicole Pacchiarini, Sara Kumziene-Summerhayes, Sarah Taylor, Sophie Jones, Sara Rey, Matthew Bull, Joanne Watkins, Sally Corden, Tom Connor
EPI_ISL_422036 hCoV-19/Wales/PHWC-26A15/2020 Europe / United Kingdom / Wales 2020-03-29 Wales Specialist Virology Centre Public Health Wales Microbiology Cardiff Catherine Moore, Johnathan Evans, Malorie Perry, Simon Cottrell, Alec Birchley, Alexander Adams, Amy Gaskin, Bree Gatica-Wilcox, Jason Coombes, Lauren Gilbert, Lee Graham, Nicole Pacchiarini, Sara Kumziene-Summerhayes, Sarah Taylor, Sophie Jones, Sara Rey, Matthew Bull, Joanne Watkins, Sally Corden, Tom Connor
EPI_ISL_422037 hCoV-19/Wales/PHWC-2663F/2020 Europe / United Kingdom / Wales 2020-03-27 Wales Specialist Virology Centre Public Health Wales Microbiology Cardiff Catherine Moore, Johnathan Evans, Malorie Perry, Simon Cottrell, Alec Birchley, Alexander Adams, Amy Gaskin, Bree Gatica-Wilcox, Jason Coombes, Lauren Gilbert, Lee Graham, Nicole Pacchiarini, Sara Kumziene-Summerhayes, Sarah Taylor, Sophie Jones, Sara Rey, Matthew Bull, Joanne Watkins, Sally Corden, Tom Connor
EPI_ISL_422038 hCoV-19/Wales/PHWC-2621D/2020 Europe / United Kingdom / Wales 2020-03-26 Wales Specialist Virology Centre Public Health Wales Microbiology Cardiff Catherine Moore, Johnathan Evans, Malorie Perry, Simon Cottrell, Alec Birchley, Alexander Adams, Amy Gaskin, Bree Gatica-Wilcox, Jason Coombes, Lauren Gilbert, Lee Graham, Nicole Pacchiarini, Sara Kumziene-Summerhayes, Sarah Taylor, Sophie Jones, Sara Rey, Matthew Bull, Joanne Watkins, Sally Corden, Tom Connor
EPI_ISL_422100 hCoV-19/Wales/PHWC-260B9/2020 Europe / United Kingdom / Wales 2020-03-25 Wales Specialist Virology Centre Public Health Wales Microbiology Cardiff Catherine Moore, Johnathan Evans, Malorie Perry, Simon Cottrell, Alec Birchley, Alexander Adams, Amy Gaskin, Bree Gatica-Wilcox, Jason Coombes, Lauren Gilbert, Lee Graham, Nicole Pacchiarini, Sara Kumziene-Summerhayes, Sarah Taylor, Sophie Jones, Sara Rey, Matthew Bull, Joanne Watkins, Sally Corden, Tom Connor
EPI_ISL_422102 hCoV-19/Wales/PHWC-26444/2020 Europe / United Kingdom / Wales 2020-03-27 Wales Specialist Virology Centre Public Health Wales Microbiology Cardiff Catherine Moore, Johnathan Evans, Malorie Perry, Simon Cottrell, Alec Birchley, Alexander Adams, Amy Gaskin, Bree Gatica-Wilcox, Jason Coombes, Lauren Gilbert, Lee Graham, Nicole Pacchiarini, Sara Kumziene-Summerhayes, Sarah Taylor, Sophie Jones, Sara Rey, Matthew Bull, Joanne Watkins, Sally Corden, Tom Connor
EPI_ISL_422103 hCoV-19/Wales/PHWC-26C00/2020 Europe / United Kingdom / Wales 2020-03-29 Wales Specialist Virology Centre Public Health Wales Microbiology Cardiff Catherine Moore, Johnathan Evans, Malorie Perry, Simon Cottrell, Alec Birchley, Alexander Adams, Amy Gaskin, Bree Gatica-Wilcox, Jason Coombes, Lauren Gilbert, Lee Graham, Nicole Pacchiarini, Sara Kumziene-Summerhayes, Sarah Taylor, Sophie Jones, Sara Rey, Matthew Bull, Joanne Watkins, Sally Corden, Tom Connor
EPI_ISL_422104 hCoV-19/Wales/PHWC-26022/2020 Europe / United Kingdom / Wales 2020-03-26 Wales Specialist Virology Centre Public Health Wales Microbiology Cardiff Catherine Moore, Johnathan Evans, Malorie Perry, Simon Cottrell, Alec Birchley, Alexander Adams, Amy Gaskin, Bree Gatica-Wilcox, Jason Coombes, Lauren Gilbert, Lee Graham, Nicole Pacchiarini, Sara Kumziene-Summerhayes, Sarah Taylor, Sophie Jones, Sara Rey, Matthew Bull, Joanne Watkins, Sally Corden, Tom Connor
EPI_ISL_422105 hCoV-19/Wales/PHWC-26F25/2020 Europe / United Kingdom / Wales 2020-03-31 Wales Specialist Virology Centre Public Health Wales Microbiology Cardiff Catherine Moore, Johnathan Evans, Malorie Perry, Simon Cottrell, Alec Birchley, Alexander Adams, Amy Gaskin, Bree Gatica-Wilcox, Jason Coombes, Lauren Gilbert, Lee Graham, Nicole Pacchiarini, Sara Kumziene-Summerhayes, Sarah Taylor, Sophie Jones, Sara Rey, Matthew Bull, Joanne Watkins, Sally Corden, Tom Connor
EPI_ISL_422106 hCoV-19/Wales/PHWC-260F5/2020 Europe / United Kingdom / Wales 2020-03-25 Wales Specialist Virology Centre Public Health Wales Microbiology Cardiff Catherine Moore, Johnathan Evans, Malorie Perry, Simon Cottrell, Alec Birchley, Alexander Adams, Amy Gaskin, Bree Gatica-Wilcox, Jason Coombes, Lauren Gilbert, Lee Graham, Nicole Pacchiarini, Sara Kumziene-Summerhayes, Sarah Taylor, Sophie Jones, Sara Rey, Matthew Bull, Joanne Watkins, Sally Corden, Tom Connor
EPI_ISL_422107 hCoV-19/Wales/PHWC-25FF9/2020 Europe / United Kingdom / Wales 2020-03-25 Wales Specialist Virology Centre Public Health Wales Microbiology Cardiff Catherine Moore, Johnathan Evans, Malorie Perry, Simon Cottrell, Alec Birchley, Alexander Adams, Amy Gaskin, Bree Gatica-Wilcox, Jason Coombes, Lauren Gilbert, Lee Graham, Nicole Pacchiarini, Sara Kumziene-Summerhayes, Sarah Taylor, Sophie Jones,

Sara Rey, Matthew Bull, Joanne Watkins, Sally Corden, Tom Connor
EPI_ISL_422108 hCoV-19/Wales/PHWC-2581C/2020 Europe / United Kingdom / Wales 2020-03-25 Wales Specialist Virology Centre Public Health Wales Microbiology Cardiff Catherine Moore, Johnathan Evans, Malorie Perry, Simon Cottrell, Alec Birchley, Alexander Adams, Amy Gaskin, Bree Gatica-Wilcox, Jason Coombes, Lauren Gilbert, Lee Graham, Nicole Pacchiarini, Sara Kumziene-Summerhayes, Sarah Taylor, Sophie Jones, Sara Rey, Matthew Bull, Joanne Watkins, Sally Corden, Tom Connor
EPI_ISL_422109 hCoV-19/Wales/PHWC-266F3/2020 Europe / United Kingdom / Wales 2020-03-27 Wales Specialist Virology Centre Public Health Wales Microbiology Cardiff Catherine Moore, Johnathan Evans, Malorie Perry, Simon Cottrell, Alec Birchley, Alexander Adams, Amy Gaskin, Bree Gatica-Wilcox, Jason Coombes, Lauren Gilbert, Lee Graham, Nicole Pacchiarini, Sara Kumziene-Summerhayes, Sarah Taylor, Sophie Jones, Sara Rey, Matthew Bull, Joanne Watkins, Sally Corden, Tom Connor
EPI_ISL_422110 hCoV-19/Wales/PHWC-260D7/2020 Europe / United Kingdom / Wales 2020-03-25 Wales Specialist Virology Centre Public Health Wales Microbiology Cardiff Catherine Moore, Johnathan Evans, Malorie Perry, Simon Cottrell, Alec Birchley, Alexander Adams, Amy Gaskin, Bree Gatica-Wilcox, Jason Coombes, Lauren Gilbert, Lee Graham, Nicole Pacchiarini, Sara Kumziene-Summerhayes, Sarah Taylor, Sophie Jones, Sara Rey, Matthew Bull, Joanne Watkins, Sally Corden, Tom Connor
EPI_ISL_422111 hCoV-19/Wales/PHWC-260C8/2020 Europe / United Kingdom / Wales 2020-03-26 Wales Specialist Virology Centre Public Health Wales Microbiology Cardiff Catherine Moore, Johnathan Evans, Malorie Perry, Simon Cottrell, Alec Birchley, Alexander Adams, Amy Gaskin, Bree Gatica-Wilcox, Jason Coombes, Lauren Gilbert, Lee Graham, Nicole Pacchiarini, Sara Kumziene-Summerhayes, Sarah Taylor, Sophie Jones, Sara Rey, Matthew Bull, Joanne Watkins, Sally Corden, Tom Connor
EPI_ISL_422112 hCoV-19/Wales/PHWC-26417/2020 Europe / United Kingdom / Wales 2020-03-27 Wales Specialist Virology Centre Public Health Wales Microbiology Cardiff Catherine Moore, Johnathan Evans, Malorie Perry, Simon Cottrell, Alec Birchley, Alexander Adams, Amy Gaskin, Bree Gatica-Wilcox, Jason Coombes, Lauren Gilbert, Lee Graham, Nicole Pacchiarini, Sara Kumziene-Summerhayes, Sarah Taylor, Sophie Jones, Sara Rey, Matthew Bull, Joanne Watkins, Sally Corden, Tom Connor
EPI_ISL_422113 hCoV-19/Wales/PHWC-26CB5/2020 Europe / United Kingdom / Wales 2020-03-28 Wales Specialist Virology Centre Public Health Wales Microbiology Cardiff Catherine Moore, Johnathan Evans, Malorie Perry, Simon Cottrell, Alec Birchley, Alexander Adams, Amy Gaskin, Bree Gatica-Wilcox, Jason Coombes, Lauren Gilbert, Lee Graham, Nicole Pacchiarini, Sara Kumziene-Summerhayes, Sarah Taylor, Sophie Jones, Sara Rey, Matthew Bull, Joanne Watkins, Sally Corden, Tom Connor
EPI_ISL_422114 hCoV-19/Wales/PHWC-25BC8/2020 Europe / United Kingdom / Wales 2020-03-25 Wales Specialist Virology Centre Public Health Wales Microbiology Cardiff Catherine Moore, Johnathan Evans, Malorie Perry, Simon Cottrell, Alec Birchley, Alexander Adams, Amy Gaskin, Bree Gatica-Wilcox, Jason Coombes, Lauren Gilbert, Lee Graham, Nicole Pacchiarini, Sara Kumziene-Summerhayes, Sarah Taylor, Sophie Jones, Sara Rey, Matthew Bull, Joanne Watkins, Sally Corden, Tom Connor
EPI_ISL_422115 hCoV-19/Wales/PHWC-26EDD/2020 Europe / United Kingdom / Wales 2020-03-30 Wales Specialist Virology Centre Public Health Wales Microbiology Cardiff Catherine Moore, Johnathan Evans, Malorie Perry, Simon Cottrell, Alec Birchley, Alexander Adams, Amy Gaskin, Bree Gatica-Wilcox, Jason Coombes, Lauren Gilbert, Lee Graham, Nicole Pacchiarini, Sara Kumziene-Summerhayes, Sarah Taylor, Sophie Jones, Sara Rey, Matthew Bull, Joanne Watkins, Sally Corden, Tom Connor
EPI_ISL_422116 hCoV-19/Wales/PHWC-25B04/2020 Europe / United Kingdom / Wales 2020-03-24 Wales Specialist Virology Centre Public Health Wales Microbiology Cardiff Catherine Moore, Johnathan Evans, Malorie Perry, Simon Cottrell, Alec Birchley, Alexander Adams, Amy Gaskin, Bree Gatica-Wilcox, Jason Coombes, Lauren Gilbert, Lee Graham, Nicole Pacchiarini, Sara Kumziene-Summerhayes, Sarah Taylor, Sophie Jones, Sara Rey, Matthew Bull, Joanne Watkins, Sally Corden, Tom Connor
EPI_ISL_422117 hCoV-19/Wales/PHWC-25F71/2020 Europe / United Kingdom / Wales 2020-03-25 Wales Specialist Virology Centre Public Health Wales Microbiology Cardiff Catherine Moore, Johnathan Evans, Malorie Perry, Simon Cottrell, Alec Birchley, Alexander Adams, Amy Gaskin, Bree Gatica-Wilcox, Jason Coombes, Lauren Gilbert, Lee Graham, Nicole Pacchiarini, Sara Kumziene-Summerhayes, Sarah Taylor, Sophie Jones, Sara Rey, Matthew Bull, Joanne Watkins, Sally Corden, Tom Connor
EPI_ISL_422118 hCoV-19/Wales/PHWC-25F9F/2020 Europe / United Kingdom / Wales 2020-03-25 Wales Specialist Virology Centre Public Health Wales Microbiology Cardiff Catherine Moore, Johnathan Evans, Malorie Perry, Simon Cottrell, Alec Birchley, Alexander Adams, Amy Gaskin, Bree Gatica-Wilcox, Jason Coombes, Lauren Gilbert, Lee Graham, Nicole Pacchiarini, Sara Kumziene-Summerhayes, Sarah Taylor, Sophie Jones, Sara Rey, Matthew Bull, Joanne Watkins, Sally Corden, Tom Connor
EPI_ISL_422119 hCoV-19/Wales/PHWC-26C5B/2020 Europe / United Kingdom / Wales 2020-03-27 Wales Specialist Virology Centre Public Health Wales Microbiology Cardiff Catherine Moore, Johnathan Evans, Malorie Perry, Simon Cottrell, Alec Birchley, Alexander Adams, Amy Gaskin, Bree Gatica-Wilcox, Jason Coombes, Lauren Gilbert, Lee Graham, Nicole Pacchiarini, Sara Kumziene-Summerhayes, Sarah Taylor, Sophie Jones, Sara Rey, Matthew Bull, Joanne Watkins, Sally Corden, Tom Connor
EPI_ISL_422120 hCoV-19/Wales/PHWC-26FBC/2020 Europe / United Kingdom / Wales 2020-03-30 Wales Specialist Virology Centre Public Health Wales Microbiology Cardiff Catherine Moore, Johnathan Evans, Malorie Perry, Simon Cottrell, Alec Birchley, Alexander Adams, Amy Gaskin, Bree Gatica-Wilcox, Jason Coombes, Lauren Gilbert, Lee Graham, Nicole Pacchiarini, Sara Kumziene-Summerhayes, Sarah Taylor, Sophie Jones, Sara Rey, Matthew Bull, Joanne Watkins, Sally Corden, Tom Connor
EPI_ISL_422121 hCoV-19/Wales/PHWC-26C97/2020 Europe / United Kingdom / Wales 2020-03-28 Wales Specialist Virology Centre Public Health Wales Microbiology Cardiff Catherine Moore, Johnathan Evans, Malorie Perry, Simon Cottrell, Alec Birchley, Alexander Adams, Amy Gaskin, Bree Gatica-Wilcox, Jason Coombes, Lauren Gilbert, Lee Graham, Nicole Pacchiarini, Sara Kumziene-Summerhayes, Sarah Taylor, Sophie Jones,

Sara Rey, Matthew Bull, Joanne Watkins, Sally Corden, Tom Connor
EPI_ISL_422122 hCoV-19/Wales/PHWC-26E73/2020 Europe / United Kingdom / Wales 2020-03-30 Wales Specialist Virology Centre Public Health Wales Microbiology Cardiff Catherine Moore, Johnathan Evans, Malorie Perry, Simon Cottrell, Alec Birchley, Alexander Adams, Amy Gaskin, Bree Gatica-Wilcox, Jason Coombes, Lauren Gilbert, Lee Graham, Nicole Pacchiarini, Sara Kumziene-Summerhayes, Sarah Taylor, Sophie Jones, Sara Rey, Matthew Bull, Joanne Watkins, Sally Corden, Tom Connor
EPI_ISL_422123 hCoV-19/Wales/PHWC-26FCB/2020 Europe / United Kingdom / Wales 2020-03-30 Wales Specialist Virology Centre Public Health Wales Microbiology Cardiff Catherine Moore, Johnathan Evans, Malorie Perry, Simon Cottrell, Alec Birchley, Alexander Adams, Amy Gaskin, Bree Gatica-Wilcox, Jason Coombes, Lauren Gilbert, Lee Graham, Nicole Pacchiarini, Sara Kumziene-Summerhayes, Sarah Taylor, Sophie Jones, Sara Rey, Matthew Bull, Joanne Watkins, Sally Corden, Tom Connor
EPI_ISL_422124 hCoV-19/Wales/PHWC-2715B/2020 Europe / United Kingdom / Wales 2020-03-29 Wales Specialist Virology Centre Public Health Wales Microbiology Cardiff Catherine Moore, Johnathan Evans, Malorie Perry, Simon Cottrell, Alec Birchley, Alexander Adams, Amy Gaskin, Bree Gatica-Wilcox, Jason Coombes, Lauren Gilbert, Lee Graham, Nicole Pacchiarini, Sara Kumziene-Summerhayes, Sarah Taylor, Sophie Jones, Sara Rey, Matthew Bull, Joanne Watkins, Sally Corden, Tom Connor
EPI_ISL_422125 hCoV-19/Wales/PHWC-27003/2020 Europe / United Kingdom / Wales 2020-03-28 Wales Specialist Virology Centre Public Health Wales Microbiology Cardiff Catherine Moore, Johnathan Evans, Malorie Perry, Simon Cottrell, Alec Birchley, Alexander Adams, Amy Gaskin, Bree Gatica-Wilcox, Jason Coombes, Lauren Gilbert, Lee Graham, Nicole Pacchiarini, Sara Kumziene-Summerhayes, Sarah Taylor, Sophie Jones, Sara Rey, Matthew Bull, Joanne Watkins, Sally Corden, Tom Connor
EPI_ISL_422126 hCoV-19/Wales/PHWC-267B4/2020 Europe / United Kingdom / Wales 2020-03-28 Wales Specialist Virology Centre Public Health Wales Microbiology Cardiff Catherine Moore, Johnathan Evans, Malorie Perry, Simon Cottrell, Alec Birchley, Alexander Adams, Amy Gaskin, Bree Gatica-Wilcox, Jason Coombes, Lauren Gilbert, Lee Graham, Nicole Pacchiarini, Sara Kumziene-Summerhayes, Sarah Taylor, Sophie Jones, Sara Rey, Matthew Bull, Joanne Watkins, Sally Corden, Tom Connor
EPI_ISL_422127 hCoV-19/Wales/PHWC-26699/2020 Europe / United Kingdom / Wales 2020-03-27 Wales Specialist Virology Centre Public Health Wales Microbiology Cardiff Catherine Moore, Johnathan Evans, Malorie Perry, Simon Cottrell, Alec Birchley, Alexander Adams, Amy Gaskin, Bree Gatica-Wilcox, Jason Coombes, Lauren Gilbert, Lee Graham, Nicole Pacchiarini, Sara Kumziene-Summerhayes, Sarah Taylor, Sophie Jones, Sara Rey, Matthew Bull, Joanne Watkins, Sally Corden, Tom Connor
EPI_ISL_422128 hCoV-19/Wales/PHWC-266A8/2020 Europe / United Kingdom / Wales 2020-03-27 Wales Specialist Virology Centre Public Health Wales Microbiology Cardiff Catherine Moore, Johnathan Evans, Malorie Perry, Simon Cottrell, Alec Birchley, Alexander Adams, Amy Gaskin, Bree Gatica-Wilcox, Jason Coombes, Lauren Gilbert, Lee Graham, Nicole Pacchiarini, Sara Kumziene-Summerhayes, Sarah Taylor, Sophie Jones, Sara Rey, Matthew Bull, Joanne Watkins, Sally Corden, Tom Connor
EPI_ISL_422129 hCoV-19/Wales/PHWC-267A5/2020 Europe / United Kingdom / Wales 2020-03-28 Wales Specialist Virology Centre Public Health Wales Microbiology Cardiff Catherine Moore, Johnathan Evans, Malorie Perry, Simon Cottrell, Alec Birchley, Alexander Adams, Amy Gaskin, Bree Gatica-Wilcox, Jason Coombes, Lauren Gilbert, Lee Graham, Nicole Pacchiarini, Sara Kumziene-Summerhayes, Sarah Taylor, Sophie Jones, Sara Rey, Matthew Bull, Joanne Watkins, Sally Corden, Tom Connor
EPI_ISL_422130 hCoV-19/Wales/PHWC-26A9D/2020 Europe / United Kingdom / Wales 2020-03-30 Wales Specialist Virology Centre Public Health Wales Microbiology Cardiff Catherine Moore, Johnathan Evans, Malorie Perry, Simon Cottrell, Alec Birchley, Alexander Adams, Amy Gaskin, Bree Gatica-Wilcox, Jason Coombes, Lauren Gilbert, Lee Graham, Nicole Pacchiarini, Sara Kumziene-Summerhayes, Sarah Taylor, Sophie Jones, Sara Rey, Matthew Bull, Joanne Watkins, Sally Corden, Tom Connor
EPI_ISL_422131 hCoV-19/Wales/PHWC-27030/2020 Europe / United Kingdom / Wales 2020-03-28 Wales Specialist Virology Centre Public Health Wales Microbiology Cardiff Catherine Moore, Johnathan Evans, Malorie Perry, Simon Cottrell, Alec Birchley, Alexander Adams, Amy Gaskin, Bree Gatica-Wilcox, Jason Coombes, Lauren Gilbert, Lee Graham, Nicole Pacchiarini, Sara Kumziene-Summerhayes, Sarah Taylor, Sophie Jones, Sara Rey, Matthew Bull, Joanne Watkins, Sally Corden, Tom Connor
EPI_ISL_422132 hCoV-19/Wales/PHWC-26F07/2020 Europe / United Kingdom / Wales 2020-03-30 Wales Specialist Virology Centre Public Health Wales Microbiology Cardiff Catherine Moore, Johnathan Evans, Malorie Perry, Simon Cottrell, Alec Birchley, Alexander Adams, Amy Gaskin, Bree Gatica-Wilcox, Jason Coombes, Lauren Gilbert, Lee Graham, Nicole Pacchiarini, Sara Kumziene-Summerhayes, Sarah Taylor, Sophie Jones, Sara Rey, Matthew Bull, Joanne Watkins, Sally Corden, Tom Connor
EPI_ISL_422133 hCoV-19/Wales/PHWC-270C7/2020 Europe / United Kingdom / Wales 2020-03-30 Wales Specialist Virology Centre Public Health Wales Microbiology Cardiff Catherine Moore, Johnathan Evans, Malorie Perry, Simon Cottrell, Alec Birchley, Alexander Adams, Amy Gaskin, Bree Gatica-Wilcox, Jason Coombes, Lauren Gilbert, Lee Graham, Nicole Pacchiarini, Sara Kumziene-Summerhayes, Sarah Taylor, Sophie Jones, Sara Rey, Matthew Bull, Joanne Watkins, Sally Corden, Tom Connor
EPI_ISL_422134 hCoV-19/Wales/PHWC-25DD1/2020 Europe / United Kingdom / Wales 2020-03-25 Wales Specialist Virology Centre Public Health Wales Microbiology Cardiff Catherine Moore, Johnathan Evans, Malorie Perry, Simon Cottrell, Alec Birchley, Alexander Adams, Amy Gaskin, Bree Gatica-Wilcox, Jason Coombes, Lauren Gilbert, Lee Graham, Nicole Pacchiarini, Sara Kumziene-Summerhayes, Sarah Taylor, Sophie Jones, Sara Rey, Matthew Bull, Joanne Watkins, Sally Corden, Tom Connor
EPI_ISL_422135 hCoV-19/Wales/PHWC-25E65/2020 Europe / United Kingdom / Wales 2020-03-25 Wales Specialist Virology Centre Public Health Wales Microbiology Cardiff Catherine Moore, Johnathan Evans, Malorie Perry, Simon Cottrell, Alec Birchley, Alexander Adams, Amy Gaskin, Bree Gatica-Wilcox, Jason Coombes, Lauren Gilbert, Lee Graham, Nicole Pacchiarini, Sara Kumziene-Summerhayes, Sarah Taylor, Sophie Jones,

Sara Rey, Matthew Bull, Joanne Watkins, Sally Corden, Tom Connor
EPI_ISL_422136 hCoV-19/Wales/PHWC-25EB0/2020 Europe / United Kingdom / Wales 2020-03-25 Wales Specialist Virology Centre Public Health Wales Microbiology Cardiff Catherine Moore, Johnathan Evans, Malorie Perry, Simon Cottrell, Alec Birchley, Alexander Adams, Amy Gaskin, Bree Gatica-Wilcox, Jason Coombes, Lauren Gilbert, Lee Graham, Nicole Pacchiarini, Sara Kumziene-Summerhayes, Sarah Taylor, Sophie Jones, Sara Rey, Matthew Bull, Joanne Watkins, Sally Corden, Tom Connor
EPI_ISL_422137 hCoV-19/Wales/PHWC-26EEC/2020 Europe / United Kingdom / Wales 2020-03-30 Wales Specialist Virology Centre Public Health Wales Microbiology Cardiff Catherine Moore, Johnathan Evans, Malorie Perry, Simon Cottrell, Alec Birchley, Alexander Adams, Amy Gaskin, Bree Gatica-Wilcox, Jason Coombes, Lauren Gilbert, Lee Graham, Nicole Pacchiarini, Sara Kumziene-Summerhayes, Sarah Taylor, Sophie Jones, Sara Rey, Matthew Bull, Joanne Watkins, Sally Corden, Tom Connor
EPI_ISL_422138 hCoV-19/Wales/PHWC-259A0/2020 Europe / United Kingdom / Wales 2020-03-24 Wales Specialist Virology Centre Public Health Wales Microbiology Cardiff Catherine Moore, Johnathan Evans, Malorie Perry, Simon Cottrell, Alec Birchley, Alexander Adams, Amy Gaskin, Bree Gatica-Wilcox, Jason Coombes, Lauren Gilbert, Lee Graham, Nicole Pacchiarini, Sara Kumziene-Summerhayes, Sarah Taylor, Sophie Jones, Sara Rey, Matthew Bull, Joanne Watkins, Sally Corden, Tom Connor
EPI_ISL_422139 hCoV-19/Wales/PHWC-25867/2020 Europe / United Kingdom / Wales 2020-03-25 Wales Specialist Virology Centre Public Health Wales Microbiology Cardiff Catherine Moore, Johnathan Evans, Malorie Perry, Simon Cottrell, Alec Birchley, Alexander Adams, Amy Gaskin, Bree Gatica-Wilcox, Jason Coombes, Lauren Gilbert, Lee Graham, Nicole Pacchiarini, Sara Kumziene-Summerhayes, Sarah Taylor, Sophie Jones, Sara Rey, Matthew Bull, Joanne Watkins, Sally Corden, Tom Connor
EPI_ISL_422140 hCoV-19/Wales/PHWC-26F8F/2020 Europe / United Kingdom / Wales 2020-03-29 Wales Specialist Virology Centre Public Health Wales Microbiology Cardiff Catherine Moore, Johnathan Evans, Malorie Perry, Simon Cottrell, Alec Birchley, Alexander Adams, Amy Gaskin, Bree Gatica-Wilcox, Jason Coombes, Lauren Gilbert, Lee Graham, Nicole Pacchiarini, Sara Kumziene-Summerhayes, Sarah Taylor, Sophie Jones, Sara Rey, Matthew Bull, Joanne Watkins, Sally Corden, Tom Connor
EPI_ISL_422141 hCoV-19/Wales/PHWC-26893/2020 Europe / United Kingdom / Wales 2020-03-27 Wales Specialist Virology Centre Public Health Wales Microbiology Cardiff Catherine Moore, Johnathan Evans, Malorie Perry, Simon Cottrell, Alec Birchley, Alexander Adams, Amy Gaskin, Bree Gatica-Wilcox, Jason Coombes, Lauren Gilbert, Lee Graham, Nicole Pacchiarini, Sara Kumziene-Summerhayes, Sarah Taylor, Sophie Jones, Sara Rey, Matthew Bull, Joanne Watkins, Sally Corden, Tom Connor
EPI_ISL_422142 hCoV-19/Wales/PHWC-2613E/2020 Europe / United Kingdom / Wales 2020-03-25 Wales Specialist Virology Centre Public Health Wales Microbiology Cardiff Catherine Moore, Johnathan Evans, Malorie Perry, Simon Cottrell, Alec Birchley, Alexander Adams, Amy Gaskin, Bree Gatica-Wilcox, Jason Coombes, Lauren Gilbert, Lee Graham, Nicole Pacchiarini, Sara Kumziene-Summerhayes, Sarah Taylor, Sophie Jones, Sara Rey, Matthew Bull, Joanne Watkins, Sally Corden, Tom Connor
EPI_ISL_422143 hCoV-19/Wales/PHWC-26E64/2020 Europe / United Kingdom / Wales 2020-03-29 Wales Specialist Virology Centre Public Health Wales Microbiology Cardiff Catherine Moore, Johnathan Evans, Malorie Perry, Simon Cottrell, Alec Birchley, Alexander Adams, Amy Gaskin, Bree Gatica-Wilcox, Jason Coombes, Lauren Gilbert, Lee Graham, Nicole Pacchiarini, Sara Kumziene-Summerhayes, Sarah Taylor, Sophie Jones, Sara Rey, Matthew Bull, Joanne Watkins, Sally Corden, Tom Connor
EPI_ISL_422144 hCoV-19/Wales/PHWC-26F43/2020 Europe / United Kingdom / Wales 2020-03-28 Wales Specialist Virology Centre Public Health Wales Microbiology Cardiff Catherine Moore, Johnathan Evans, Malorie Perry, Simon Cottrell, Alec Birchley, Alexander Adams, Amy Gaskin, Bree Gatica-Wilcox, Jason Coombes, Lauren Gilbert, Lee Graham, Nicole Pacchiarini, Sara Kumziene-Summerhayes, Sarah Taylor, Sophie Jones, Sara Rey, Matthew Bull, Joanne Watkins, Sally Corden, Tom Connor
EPI_ISL_422145 hCoV-19/Wales/PHWC-26EA0/2020 Europe / United Kingdom / Wales 2020-03-30 Wales Specialist Virology Centre Public Health Wales Microbiology Cardiff Catherine Moore, Johnathan Evans, Malorie Perry, Simon Cottrell, Alec Birchley, Alexander Adams, Amy Gaskin, Bree Gatica-Wilcox, Jason Coombes, Lauren Gilbert, Lee Graham, Nicole Pacchiarini, Sara Kumziene-Summerhayes, Sarah Taylor, Sophie Jones, Sara Rey, Matthew Bull, Joanne Watkins, Sally Corden, Tom Connor
EPI_ISL_422146 hCoV-19/Wales/PHWC-26D2B/2020 Europe / United Kingdom / Wales 2020-03-30 Wales Specialist Virology Centre Public Health Wales Microbiology Cardiff Catherine Moore, Johnathan Evans, Malorie Perry, Simon Cottrell, Alec Birchley, Alexander Adams, Amy Gaskin, Bree Gatica-Wilcox, Jason Coombes, Lauren Gilbert, Lee Graham, Nicole Pacchiarini, Sara Kumziene-Summerhayes, Sarah Taylor, Sophie Jones, Sara Rey, Matthew Bull, Joanne Watkins, Sally Corden, Tom Connor
EPI_ISL_422147 hCoV-19/Wales/PHWC-2706D/2020 Europe / United Kingdom / Wales 2020-03-28 Wales Specialist Virology Centre Public Health Wales Microbiology Cardiff Catherine Moore, Johnathan Evans, Malorie Perry, Simon Cottrell, Alec Birchley, Alexander Adams, Amy Gaskin, Bree Gatica-Wilcox, Jason Coombes, Lauren Gilbert, Lee Graham, Nicole Pacchiarini, Sara Kumziene-Summerhayes, Sarah Taylor, Sophie Jones, Sara Rey, Matthew Bull, Joanne Watkins, Sally Corden, Tom Connor
EPI_ISL_422148 hCoV-19/Wales/PHWC-26BC7/2020 Europe / United Kingdom / Wales 2020-03-27 Wales Specialist Virology Centre Public Health Wales Microbiology Cardiff Catherine Moore, Johnathan Evans, Malorie Perry, Simon Cottrell, Alec Birchley, Alexander Adams, Amy Gaskin, Bree Gatica-Wilcox, Jason Coombes, Lauren Gilbert, Lee Graham, Nicole Pacchiarini, Sara Kumziene-Summerhayes, Sarah Taylor, Sophie Jones, Sara Rey, Matthew Bull, Joanne Watkins, Sally Corden, Tom Connor
EPI_ISL_422149 hCoV-19/Wales/PHWC-26004/2020 Europe / United Kingdom / Wales 2020-03-25 Wales Specialist Virology Centre Public Health Wales Microbiology Cardiff Catherine Moore, Johnathan Evans, Malorie Perry, Simon Cottrell, Alec Birchley, Alexander Adams, Amy Gaskin, Bree Gatica-Wilcox, Jason Coombes, Lauren Gilbert, Lee Graham, Nicole Pacchiarini, Sara Kumziene-Summerhayes, Sarah Taylor, Sophie Jones,

Sara Rey, Matthew Bull, Joanne Watkins, Sally Corden, Tom Connor
EPI_ISL_422150 hCoV-19/Wales/PHWC-25E0B/2020 Europe / United Kingdom / Wales 2020-03-25 Wales Specialist Virology Centre Public Health Wales Microbiology Cardiff Catherine Moore, Johnathan Evans, Malorie Perry, Simon Cottrell, Alec Birchley, Alexander Adams, Amy Gaskin, Bree Gatica-Wilcox, Jason Coombes, Lauren Gilbert, Lee Graham, Nicole Pacchiarini, Sara Kumziene-Summerhayes, Sarah Taylor, Sophie Jones, Sara Rey, Matthew Bull, Joanne Watkins, Sally Corden, Tom Connor
EPI_ISL_422151 hCoV-19/Wales/PHWC-26C1F/2020 Europe / United Kingdom / Wales 2020-03-28 Wales Specialist Virology Centre Public Health Wales Microbiology Cardiff Catherine Moore, Johnathan Evans, Malorie Perry, Simon Cottrell, Alec Birchley, Alexander Adams, Amy Gaskin, Bree Gatica-Wilcox, Jason Coombes, Lauren Gilbert, Lee Graham, Nicole Pacchiarini, Sara Kumziene-Summerhayes, Sarah Taylor, Sophie Jones, Sara Rey, Matthew Bull, Joanne Watkins, Sally Corden, Tom Connor
EPI_ISL_422152 hCoV-19/Wales/PHWC-26FAD/2020 Europe / United Kingdom / Wales 2020-03-30 Wales Specialist Virology Centre Public Health Wales Microbiology Cardiff Catherine Moore, Johnathan Evans, Malorie Perry, Simon Cottrell, Alec Birchley, Alexander Adams, Amy Gaskin, Bree Gatica-Wilcox, Jason Coombes, Lauren Gilbert, Lee Graham, Nicole Pacchiarini, Sara Kumziene-Summerhayes, Sarah Taylor, Sophie Jones, Sara Rey, Matthew Bull, Joanne Watkins, Sally Corden, Tom Connor
EPI_ISL_422153 hCoV-19/Wales/PHWC-257C4/2020 Europe / United Kingdom / Wales 2020-03-24 Wales Specialist Virology Centre Public Health Wales Microbiology Cardiff Catherine Moore, Johnathan Evans, Malorie Perry, Simon Cottrell, Alec Birchley, Alexander Adams, Amy Gaskin, Bree Gatica-Wilcox, Jason Coombes, Lauren Gilbert, Lee Graham, Nicole Pacchiarini, Sara Kumziene-Summerhayes, Sarah Taylor, Sophie Jones, Sara Rey, Matthew Bull, Joanne Watkins, Sally Corden, Tom Connor
EPI_ISL_422154 hCoV-19/Wales/PHWC-25C10/2020 Europe / United Kingdom / Wales 2020-03-25 Wales Specialist Virology Centre Public Health Wales Microbiology Cardiff Catherine Moore, Johnathan Evans, Malorie Perry, Simon Cottrell, Alec Birchley, Alexander Adams, Amy Gaskin, Bree Gatica-Wilcox, Jason Coombes, Lauren Gilbert, Lee Graham, Nicole Pacchiarini, Sara Kumziene-Summerhayes, Sarah Taylor, Sophie Jones, Sara Rey, Matthew Bull, Joanne Watkins, Sally Corden, Tom Connor
EPI_ISL_422155 hCoV-19/Wales/PHWC-2713D/2020 Europe / United Kingdom / Wales 2020-03-29 Wales Specialist Virology Centre Public Health Wales Microbiology Cardiff Catherine Moore, Johnathan Evans, Malorie Perry, Simon Cottrell, Alec Birchley, Alexander Adams, Amy Gaskin, Bree Gatica-Wilcox, Jason Coombes, Lauren Gilbert, Lee Graham, Nicole Pacchiarini, Sara Kumziene-Summerhayes, Sarah Taylor, Sophie Jones, Sara Rey, Matthew Bull, Joanne Watkins, Sally Corden, Tom Connor
EPI_ISL_422156 hCoV-19/Wales/PHWC-26C88/2020 Europe / United Kingdom / Wales 2020-03-28 Wales Specialist Virology Centre Public Health Wales Microbiology Cardiff Catherine Moore, Johnathan Evans, Malorie Perry, Simon Cottrell, Alec Birchley, Alexander Adams, Amy Gaskin, Bree Gatica-Wilcox, Jason Coombes, Lauren Gilbert, Lee Graham, Nicole Pacchiarini, Sara Kumziene-Summerhayes, Sarah Taylor, Sophie Jones, Sara Rey, Matthew Bull, Joanne Watkins, Sally Corden, Tom Connor
EPI_ISL_422157 hCoV-19/Wales/PHWC-26787/2020 Europe / United Kingdom / Wales 2020-03-28 Wales Specialist Virology Centre Public Health Wales Microbiology Cardiff Catherine Moore, Johnathan Evans, Malorie Perry, Simon Cottrell, Alec Birchley, Alexander Adams, Amy Gaskin, Bree Gatica-Wilcox, Jason Coombes, Lauren Gilbert, Lee Graham, Nicole Pacchiarini, Sara Kumziene-Summerhayes, Sarah Taylor, Sophie Jones, Sara Rey, Matthew Bull, Joanne Watkins, Sally Corden, Tom Connor
EPI_ISL_422158 hCoV-19/Wales/PHWC-266D5/2020 Europe / United Kingdom / Wales 2020-03-27 Wales Specialist Virology Centre Public Health Wales Microbiology Cardiff Catherine Moore, Johnathan Evans, Malorie Perry, Simon Cottrell, Alec Birchley, Alexander Adams, Amy Gaskin, Bree Gatica-Wilcox, Jason Coombes, Lauren Gilbert, Lee Graham, Nicole Pacchiarini, Sara Kumziene-Summerhayes, Sarah Taylor, Sophie Jones, Sara Rey, Matthew Bull, Joanne Watkins, Sally Corden, Tom Connor
EPI_ISL_422159 hCoV-19/Wales/PHWC-26E28/2020 Europe / United Kingdom / Wales 2020-03-29 Wales Specialist Virology Centre Public Health Wales Microbiology Cardiff Catherine Moore, Johnathan Evans, Malorie Perry, Simon Cottrell, Alec Birchley, Alexander Adams, Amy Gaskin, Bree Gatica-Wilcox, Jason Coombes, Lauren Gilbert, Lee Graham, Nicole Pacchiarini, Sara Kumziene-Summerhayes, Sarah Taylor, Sophie Jones, Sara Rey, Matthew Bull, Joanne Watkins, Sally Corden, Tom Connor
EPI_ISL_422160 hCoV-19/Wales/PHWC-26D94/2020 Europe / United Kingdom / Wales 2020-03-30 Wales Specialist Virology Centre Public Health Wales Microbiology Cardiff Catherine Moore, Johnathan Evans, Malorie Perry, Simon Cottrell, Alec Birchley, Alexander Adams, Amy Gaskin, Bree Gatica-Wilcox, Jason Coombes, Lauren Gilbert, Lee Graham, Nicole Pacchiarini, Sara Kumziene-Summerhayes, Sarah Taylor, Sophie Jones, Sara Rey, Matthew Bull, Joanne Watkins, Sally Corden, Tom Connor
EPI_ISL_422161 hCoV-19/Wales/PHWC-26CE2/2020 Europe / United Kingdom / Wales 2020-03-29 Wales Specialist Virology Centre Public Health Wales Microbiology Cardiff Catherine Moore, Johnathan Evans, Malorie Perry, Simon Cottrell, Alec Birchley, Alexander Adams, Amy Gaskin, Bree Gatica-Wilcox, Jason Coombes, Lauren Gilbert, Lee Graham, Nicole Pacchiarini, Sara Kumziene-Summerhayes, Sarah Taylor, Sophie Jones, Sara Rey, Matthew Bull, Joanne Watkins, Sally Corden, Tom Connor
EPI_ISL_422162 hCoV-19/Wales/PHWC-26611/2020 Europe / United Kingdom / Wales 2020-03-27 Wales Specialist Virology Centre Public Health Wales Microbiology Cardiff Catherine Moore, Johnathan Evans, Malorie Perry, Simon Cottrell, Alec Birchley, Alexander Adams, Amy Gaskin, Bree Gatica-Wilcox, Jason Coombes, Lauren Gilbert, Lee Graham, Nicole Pacchiarini, Sara Kumziene-Summerhayes, Sarah Taylor, Sophie Jones, Sara Rey, Matthew Bull, Joanne Watkins, Sally Corden, Tom Connor
EPI_ISL_422163 hCoV-19/Wales/PHWC-26D85/2020 Europe / United Kingdom / Wales 2020-03-30 Wales Specialist Virology Centre Public Health Wales Microbiology Cardiff Catherine Moore, Johnathan Evans, Malorie Perry, Simon Cottrell, Alec Birchley, Alexander Adams, Amy Gaskin, Bree Gatica-Wilcox, Jason Coombes, Lauren Gilbert, Lee Graham, Nicole Pacchiarini, Sara Kumziene-Summerhayes, Sarah Taylor, Sophie Jones,

Sara Rey, Matthew Bull, Joanne Watkins, Sally Corden, Tom Connor
EPI_ISL_422164 hCoV-19/Wales/PHWC-26620/2020 Europe / United Kingdom / Wales 2020-03-26 Wales Specialist Virology Centre Public Health Wales Microbiology Cardiff Catherine Moore, Johnathan Evans, Malorie Perry, Simon Cottrell, Alec Birchley, Alexander Adams, Amy Gaskin, Bree Gatica-Wilcox, Jason Coombes, Lauren Gilbert, Lee Graham, Nicole Pacchiarini, Sara Kumziene-Summerhayes, Sarah Taylor, Sophie Jones, Sara Rey, Matthew Bull, Joanne Watkins, Sally Corden, Tom Connor
EPI_ISL_422165 hCoV-19/Wales/PHWC-26C3D/2020 Europe / United Kingdom / Wales 2020-03-30 Wales Specialist Virology Centre Public Health Wales Microbiology Cardiff Catherine Moore, Johnathan Evans, Malorie Perry, Simon Cottrell, Alec Birchley, Alexander Adams, Amy Gaskin, Bree Gatica-Wilcox, Jason Coombes, Lauren Gilbert, Lee Graham, Nicole Pacchiarini, Sara Kumziene-Summerhayes, Sarah Taylor, Sophie Jones, Sara Rey, Matthew Bull, Joanne Watkins, Sally Corden, Tom Connor
EPI_ISL_422166 hCoV-19/Wales/PHWC-26927/2020 Europe / United Kingdom / Wales 2020-03-28 Wales Specialist Virology Centre Public Health Wales Microbiology Cardiff Catherine Moore, Johnathan Evans, Malorie Perry, Simon Cottrell, Alec Birchley, Alexander Adams, Amy Gaskin, Bree Gatica-Wilcox, Jason Coombes, Lauren Gilbert, Lee Graham, Nicole Pacchiarini, Sara Kumziene-Summerhayes, Sarah Taylor, Sophie Jones, Sara Rey, Matthew Bull, Joanne Watkins, Sally Corden, Tom Connor
EPI_ISL_422167 hCoV-19/Wales/PHWC-266B7/2020 Europe / United Kingdom / Wales 2020-03-27 Wales Specialist Virology Centre Public Health Wales Microbiology Cardiff Catherine Moore, Johnathan Evans, Malorie Perry, Simon Cottrell, Alec Birchley, Alexander Adams, Amy Gaskin, Bree Gatica-Wilcox, Jason Coombes, Lauren Gilbert, Lee Graham, Nicole Pacchiarini, Sara Kumziene-Summerhayes, Sarah Taylor, Sophie Jones, Sara Rey, Matthew Bull, Joanne Watkins, Sally Corden, Tom Connor
EPI_ISL_422168 hCoV-19/Wales/PHWC-25F62/2020 Europe / United Kingdom / Wales 2020-03-25 Wales Specialist Virology Centre Public Health Wales Microbiology Cardiff Catherine Moore, Johnathan Evans, Malorie Perry, Simon Cottrell, Alec Birchley, Alexander Adams, Amy Gaskin, Bree Gatica-Wilcox, Jason Coombes, Lauren Gilbert, Lee Graham, Nicole Pacchiarini, Sara Kumziene-Summerhayes, Sarah Taylor, Sophie Jones, Sara Rey, Matthew Bull, Joanne Watkins, Sally Corden, Tom Connor
EPI_ISL_422169 hCoV-19/Wales/PHWC-25946/2020 Europe / United Kingdom / Wales 2020-03-25 Wales Specialist Virology Centre Public Health Wales Microbiology Cardiff Catherine Moore, Johnathan Evans, Malorie Perry, Simon Cottrell, Alec Birchley, Alexander Adams, Amy Gaskin, Bree Gatica-Wilcox, Jason Coombes, Lauren Gilbert, Lee Graham, Nicole Pacchiarini, Sara Kumziene-Summerhayes, Sarah Taylor, Sophie Jones, Sara Rey, Matthew Bull, Joanne Watkins, Sally Corden, Tom Connor
EPI_ISL_422170 hCoV-19/Wales/PHWC-26BA9/2020 Europe / United Kingdom / Wales 2020-03-27 Wales Specialist Virology Centre Public Health Wales Microbiology Cardiff Catherine Moore, Johnathan Evans, Malorie Perry, Simon Cottrell, Alec Birchley, Alexander Adams, Amy Gaskin, Bree Gatica-Wilcox, Jason Coombes, Lauren Gilbert, Lee Graham, Nicole Pacchiarini, Sara Kumziene-Summerhayes, Sarah Taylor, Sophie Jones, Sara Rey, Matthew Bull, Joanne Watkins, Sally Corden, Tom Connor
EPI_ISL_422171 hCoV-19/Wales/PHWC-26FDA/2020 Europe / United Kingdom / Wales 2020-03-30 Wales Specialist Virology Centre Public Health Wales Microbiology Cardiff Catherine Moore, Johnathan Evans, Malorie Perry, Simon Cottrell, Alec Birchley, Alexander Adams, Amy Gaskin, Bree Gatica-Wilcox, Jason Coombes, Lauren Gilbert, Lee Graham, Nicole Pacchiarini, Sara Kumziene-Summerhayes, Sarah Taylor, Sophie Jones, Sara Rey, Matthew Bull, Joanne Watkins, Sally Corden, Tom Connor
EPI_ISL_422172 hCoV-19/Wales/PHWC-25A52/2020 Europe / United Kingdom / Wales 2020-03-23 Wales Specialist Virology Centre Public Health Wales Microbiology Cardiff Catherine Moore, Johnathan Evans, Malorie Perry, Simon Cottrell, Alec Birchley, Alexander Adams, Amy Gaskin, Bree Gatica-Wilcox, Jason Coombes, Lauren Gilbert, Lee Graham, Nicole Pacchiarini, Sara Kumziene-Summerhayes, Sarah Taylor, Sophie Jones, Sara Rey, Matthew Bull, Joanne Watkins, Sally Corden, Tom Connor
EPI_ISL_422173 hCoV-19/Wales/PHWC-26C4C/2020 Europe / United Kingdom / Wales 2020-03-29 Wales Specialist Virology Centre Public Health Wales Microbiology Cardiff Catherine Moore, Johnathan Evans, Malorie Perry, Simon Cottrell, Alec Birchley, Alexander Adams, Amy Gaskin, Bree Gatica-Wilcox, Jason Coombes, Lauren Gilbert, Lee Graham, Nicole Pacchiarini, Sara Kumziene-Summerhayes, Sarah Taylor, Sophie Jones, Sara Rey, Matthew Bull, Joanne Watkins, Sally Corden, Tom Connor
EPI_ISL_422174 hCoV-19/Wales/PHWC-26F70/2020 Europe / United Kingdom / Wales 2020-03-29 Wales Specialist Virology Centre Public Health Wales Microbiology Cardiff Catherine Moore, Johnathan Evans, Malorie Perry, Simon Cottrell, Alec Birchley, Alexander Adams, Amy Gaskin, Bree Gatica-Wilcox, Jason Coombes, Lauren Gilbert, Lee Graham, Nicole Pacchiarini, Sara Kumziene-Summerhayes, Sarah Taylor, Sophie Jones, Sara Rey, Matthew Bull, Joanne Watkins, Sally Corden, Tom Connor
EPI_ISL_422175 hCoV-19/Wales/PHWC-257F1/2020 Europe / United Kingdom / Wales 2020-03-24 Wales Specialist Virology Centre Public Health Wales Microbiology Cardiff Catherine Moore, Johnathan Evans, Malorie Perry, Simon Cottrell, Alec Birchley, Alexander Adams, Amy Gaskin, Bree Gatica-Wilcox, Jason Coombes, Lauren Gilbert, Lee Graham, Nicole Pacchiarini, Sara Kumziene-Summerhayes, Sarah Taylor, Sophie Jones, Sara Rey, Matthew Bull, Joanne Watkins, Sally Corden, Tom Connor
EPI_ISL_422176 hCoV-19/Wales/PHWC-269CD/2020 Europe / United Kingdom / Wales 2020-03-30 Wales Specialist Virology Centre Public Health Wales Microbiology Cardiff Catherine Moore, Johnathan Evans, Malorie Perry, Simon Cottrell, Alec Birchley, Alexander Adams, Amy Gaskin, Bree Gatica-Wilcox, Jason Coombes, Lauren Gilbert, Lee Graham, Nicole Pacchiarini, Sara Kumziene-Summerhayes, Sarah Taylor, Sophie Jones, Sara Rey, Matthew Bull, Joanne Watkins, Sally Corden, Tom Connor
EPI_ISL_422177 hCoV-19/Wales/PHWC-2668A/2020 Europe / United Kingdom / Wales 2020-03-27 Wales Specialist Virology Centre Public Health Wales Microbiology Cardiff Catherine Moore, Johnathan Evans, Malorie Perry, Simon Cottrell, Alec Birchley, Alexander Adams, Amy Gaskin, Bree Gatica-Wilcox, Jason Coombes, Lauren Gilbert, Lee Graham, Nicole Pacchiarini, Sara Kumziene-Summerhayes, Sarah Taylor, Sophie Jones,

Sara Rey, Matthew Bull, Joanne Watkins, Sally Corden, Tom Connor
EPI_ISL_422178 hCoV-19/Wales/PHWC-26D67/2020 Europe / United Kingdom / Wales 2020-03-30 Wales Specialist Virology Centre Public Health Wales Microbiology Cardiff Catherine Moore, Johnathan Evans, Malorie Perry, Simon Cottrell, Alec Birchley, Alexander Adams, Amy Gaskin, Bree Gatica-Wilcox, Jason Coombes, Lauren Gilbert, Lee Graham, Nicole Pacchiarini, Sara Kumziene-Summerhayes, Sarah Taylor, Sophie Jones, Sara Rey, Matthew Bull, Joanne Watkins, Sally Corden, Tom Connor
EPI_ISL_422179 hCoV-19/Wales/PHWC-262A4/2020 Europe / United Kingdom / Wales 2020-03-27 Wales Specialist Virology Centre Public Health Wales Microbiology Cardiff Catherine Moore, Johnathan Evans, Malorie Perry, Simon Cottrell, Alec Birchley, Alexander Adams, Amy Gaskin, Bree Gatica-Wilcox, Jason Coombes, Lauren Gilbert, Lee Graham, Nicole Pacchiarini, Sara Kumziene-Summerhayes, Sarah Taylor, Sophie Jones, Sara Rey, Matthew Bull, Joanne Watkins, Sally Corden, Tom Connor
EPI_ISL_422180 hCoV-19/Wales/PHWC-25B7D/2020 Europe / United Kingdom / Wales 2020-03-25 Wales Specialist Virology Centre Public Health Wales Microbiology Cardiff Catherine Moore, Johnathan Evans, Malorie Perry, Simon Cottrell, Alec Birchley, Alexander Adams, Amy Gaskin, Bree Gatica-Wilcox, Jason Coombes, Lauren Gilbert, Lee Graham, Nicole Pacchiarini, Sara Kumziene-Summerhayes, Sarah Taylor, Sophie Jones, Sara Rey, Matthew Bull, Joanne Watkins, Sally Corden, Tom Connor
EPI_ISL_422181 hCoV-19/Wales/PHWC-261F2/2020 Europe / United Kingdom / Wales 2020-03-26 Wales Specialist Virology Centre Public Health Wales Microbiology Cardiff Catherine Moore, Johnathan Evans, Malorie Perry, Simon Cottrell, Alec Birchley, Alexander Adams, Amy Gaskin, Bree Gatica-Wilcox, Jason Coombes, Lauren Gilbert, Lee Graham, Nicole Pacchiarini, Sara Kumziene-Summerhayes, Sarah Taylor, Sophie Jones, Sara Rey, Matthew Bull, Joanne Watkins, Sally Corden, Tom Connor
EPI_ISL_422182 hCoV-19/Wales/PHWC-267D2/2020 Europe / United Kingdom / Wales 2020-03-25 Wales Specialist Virology Centre Public Health Wales Microbiology Cardiff Catherine Moore, Johnathan Evans, Malorie Perry, Simon Cottrell, Alec Birchley, Alexander Adams, Amy Gaskin, Bree Gatica-Wilcox, Jason Coombes, Lauren Gilbert, Lee Graham, Nicole Pacchiarini, Sara Kumziene-Summerhayes, Sarah Taylor, Sophie Jones, Sara Rey, Matthew Bull, Joanne Watkins, Sally Corden, Tom Connor
EPI_ISL_422183 hCoV-19/Wales/PHWC-25B5F/2020 Europe / United Kingdom / Wales 2020-03-24 Wales Specialist Virology Centre Public Health Wales Microbiology Cardiff Catherine Moore, Johnathan Evans, Malorie Perry, Simon Cottrell, Alec Birchley, Alexander Adams, Amy Gaskin, Bree Gatica-Wilcox, Jason Coombes, Lauren Gilbert, Lee Graham, Nicole Pacchiarini, Sara Kumziene-Summerhayes, Sarah Taylor, Sophie Jones, Sara Rey, Matthew Bull, Joanne Watkins, Sally Corden, Tom Connor
EPI_ISL_422184 hCoV-19/Wales/PHWC-26875/2020 Europe / United Kingdom / Wales 2020-03-27 Wales Specialist Virology Centre Public Health Wales Microbiology Cardiff Catherine Moore, Johnathan Evans, Malorie Perry, Simon Cottrell, Alec Birchley, Alexander Adams, Amy Gaskin, Bree Gatica-Wilcox, Jason Coombes, Lauren Gilbert, Lee Graham, Nicole Pacchiarini, Sara Kumziene-Summerhayes, Sarah Taylor, Sophie Jones, Sara Rey, Matthew Bull, Joanne Watkins, Sally Corden, Tom Connor
EPI_ISL_422185 hCoV-19/Wales/PHWC-2680C/2020 Europe / United Kingdom / Wales 2020-03-26 Wales Specialist Virology Centre Public Health Wales Microbiology Cardiff Catherine Moore, Johnathan Evans, Malorie Perry, Simon Cottrell, Alec Birchley, Alexander Adams, Amy Gaskin, Bree Gatica-Wilcox, Jason Coombes, Lauren Gilbert, Lee Graham, Nicole Pacchiarini, Sara Kumziene-Summerhayes, Sarah Taylor, Sophie Jones, Sara Rey, Matthew Bull, Joanne Watkins, Sally Corden, Tom Connor
EPI_ISL_422186 hCoV-19/Wales/PHWC-26B4F/2020 Europe / United Kingdom / Wales 2020-03-29 Wales Specialist Virology Centre Public Health Wales Microbiology Cardiff Catherine Moore, Johnathan Evans, Malorie Perry, Simon Cottrell, Alec Birchley, Alexander Adams, Amy Gaskin, Bree Gatica-Wilcox, Jason Coombes, Lauren Gilbert, Lee Graham, Nicole Pacchiarini, Sara Kumziene-Summerhayes, Sarah Taylor, Sophie Jones, Sara Rey, Matthew Bull, Joanne Watkins, Sally Corden, Tom Connor
EPI_ISL_422187 hCoV-19/Wales/PHWC-2709A/2020 Europe / United Kingdom / Wales 2020-03-29 Wales Specialist Virology Centre Public Health Wales Microbiology Cardiff Catherine Moore, Johnathan Evans, Malorie Perry, Simon Cottrell, Alec Birchley, Alexander Adams, Amy Gaskin, Bree Gatica-Wilcox, Jason Coombes, Lauren Gilbert, Lee Graham, Nicole Pacchiarini, Sara Kumziene-Summerhayes, Sarah Taylor, Sophie Jones, Sara Rey, Matthew Bull, Joanne Watkins, Sally Corden, Tom Connor
EPI_ISL_423321 hCoV-19/England/20138063804/2020 Europe / United Kingdom / England 2020-03-26 Respiratory Virus Unit, Microbiology Services Colindale, Public Health England Respiratory Virus Unit, Microbiology Services Colindale, Public Health England Monica Galiano, Shahjahan Miah, Angie Lackenby, Omolola Akinbami, Tiina Talts, Leena Bhaw, Richard Myers, Steven Platt, Kirstin Edwards, Jonathan Hubb, Joanna Ellis, Maria Zambon
EPI_ISL_423324 hCoV-19/England/20138078204/2020 Europe / United Kingdom / England 2020-03-26 Respiratory Virus Unit, Microbiology Services Colindale, Public Health England Respiratory Virus Unit, Microbiology Services Colindale, Public Health England Monica Galiano, Shahjahan Miah, Angie Lackenby, Omolola Akinbami, Tiina Talts, Leena Bhaw, Richard Myers, Steven Platt, Kirstin Edwards, Jonathan Hubb, Joanna Ellis, Maria Zambon
EPI_ISL_423328 hCoV-19/England/20138084904/2020 Europe / United Kingdom / England 2020-03-27 Respiratory Virus Unit, Microbiology Services Colindale, Public Health England Respiratory Virus Unit, Microbiology Services Colindale, Public Health England Monica Galiano, Shahjahan Miah, Angie Lackenby, Omolola Akinbami, Tiina Talts, Leena Bhaw, Richard Myers, Steven Platt, Kirstin Edwards, Jonathan Hubb, Joanna Ellis, Maria Zambon
EPI_ISL_423337 hCoV-19/England/20139002204/2020 Europe / United Kingdom / England 2020-03-27 Respiratory Virus Unit, Microbiology Services Colindale, Public Health England Respiratory Virus Unit, Microbiology Services Colindale, Public Health England Monica Galiano, Shahjahan Miah, Angie Lackenby, Omolola Akinbami, Tiina Talts, Leena Bhaw, Richard Myers, Steven Platt, Kirstin Edwards, Jonathan Hubb, Joanna Ellis,

Maria Zambon
EPI_ISL_423340  hCoV-19/England/20139015304/2020        Europe / United Kingdom / England       2020-03-27
Respiratory Virus Unit, Microbiology Services Colindale, Public Health England  Respiratory Virus Unit, Microbiology Services Colindale, Public Health England  Monica Galiano, Shahjahan Miah, Angie Lackenby, Omolola Akinbami, Tiina Talts, Leena Bhaw, Richard Myers, Steven Platt, Kirstin Edwards, Jonathan Hubb, Joanna Ellis, Maria Zambon
EPI_ISL_423346  hCoV-19/England/20139016704/2020        Europe / United Kingdom / England       2020-03-27
Respiratory Virus Unit, Microbiology Services Colindale, Public Health England  Respiratory Virus Unit, Microbiology Services Colindale, Public Health England  Monica Galiano, Shahjahan Miah, Angie Lackenby, Omolola Akinbami, Tiina Talts, Leena Bhaw, Richard Myers, Steven Platt, Kirstin Edwards, Jonathan Hubb, Joanna Ellis, Maria Zambon
EPI_ISL_423350  hCoV-19/England/20139060704/2020        Europe / United Kingdom / England       2020-03-28
Respiratory Virus Unit, Microbiology Services Colindale, Public Health England  Respiratory Virus Unit, Microbiology Services Colindale, Public Health England  Monica Galiano, Shahjahan Miah, Angie Lackenby, Omolola Akinbami, Tiina Talts, Leena Bhaw, Richard Myers, Steven Platt, Kirstin Edwards, Jonathan Hubb, Joanna Ellis, Maria Zambon
EPI_ISL_423351  hCoV-19/England/20139064404/2020        Europe / United Kingdom / England       2020-03-28
Respiratory Virus Unit, Microbiology Services Colindale, Public Health England  Respiratory Virus Unit, Microbiology Services Colindale, Public Health England  Monica Galiano, Shahjahan Miah, Angie Lackenby, Omolola Akinbami, Tiina Talts, Leena Bhaw, Richard Myers, Steven Platt, Kirstin Edwards, Jonathan Hubb, Joanna Ellis, Maria Zambon
EPI_ISL_423352  hCoV-19/England/20139064904/2020        Europe / United Kingdom / England       2020-03-29
Respiratory Virus Unit, Microbiology Services Colindale, Public Health England  Respiratory Virus Unit, Microbiology Services Colindale, Public Health England  Monica Galiano, Shahjahan Miah, Angie Lackenby, Omolola Akinbami, Tiina Talts, Leena Bhaw, Richard Myers, Steven Platt, Kirstin Edwards, Jonathan Hubb, Joanna Ellis, Maria Zambon
EPI_ISL_423353  hCoV-19/England/20139065204/2020        Europe / United Kingdom / England       2020-03-28
Respiratory Virus Unit, Microbiology Services Colindale, Public Health England  Respiratory Virus Unit, Microbiology Services Colindale, Public Health England  Monica Galiano, Shahjahan Miah, Angie Lackenby, Omolola Akinbami, Tiina Talts, Leena Bhaw, Richard Myers, Steven Platt, Kirstin Edwards, Jonathan Hubb, Joanna Ellis, Maria Zambon
EPI_ISL_423354  hCoV-19/England/20139065404/2020        Europe / United Kingdom / England       2020-03-29
Respiratory Virus Unit, Microbiology Services Colindale, Public Health England  Respiratory Virus Unit, Microbiology Services Colindale, Public Health England  Monica Galiano, Shahjahan Miah, Angie Lackenby, Omolola Akinbami, Tiina Talts, Leena Bhaw, Richard Myers, Steven Platt, Kirstin Edwards, Jonathan Hubb, Joanna Ellis, Maria Zambon
EPI_ISL_423355  hCoV-19/England/20139065604/2020        Europe / United Kingdom / England       2020-03-29
Respiratory Virus Unit, Microbiology Services Colindale, Public Health England  Respiratory Virus Unit, Microbiology Services Colindale, Public Health England  Monica Galiano, Shahjahan Miah, Angie Lackenby, Omolola Akinbami, Tiina Talts, Leena Bhaw, Richard Myers, Steven Platt, Kirstin Edwards, Jonathan Hubb, Joanna Ellis, Maria Zambon
EPI_ISL_423356  hCoV-19/England/20139067504/2020        Europe / United Kingdom / England       2020-03-28
Respiratory Virus Unit, Microbiology Services Colindale, Public Health England  Respiratory Virus Unit, Microbiology Services Colindale, Public Health England  Monica Galiano, Shahjahan Miah, Angie Lackenby, Omolola Akinbami, Tiina Talts, Leena Bhaw, Richard Myers, Steven Platt, Kirstin Edwards, Jonathan Hubb, Joanna Ellis, Maria Zambon
EPI_ISL_423357  hCoV-19/England/20139068504/2020        Europe / United Kingdom / England       2020-03-29
Respiratory Virus Unit, Microbiology Services Colindale, Public Health England  Respiratory Virus Unit, Microbiology Services Colindale, Public Health England  Monica Galiano, Shahjahan Miah, Angie Lackenby, Omolola Akinbami, Tiina Talts, Leena Bhaw, Richard Myers, Steven Platt, Kirstin Edwards, Jonathan Hubb, Joanna Ellis, Maria Zambon
EPI_ISL_423358  hCoV-19/England/20139068804/2020        Europe / United Kingdom / England       2020-03-28
Respiratory Virus Unit, Microbiology Services Colindale, Public Health England  Respiratory Virus Unit, Microbiology Services Colindale, Public Health England  Monica Galiano, Shahjahan Miah, Angie Lackenby, Omolola Akinbami, Tiina Talts, Leena Bhaw, Richard Myers, Steven Platt, Kirstin Edwards, Jonathan Hubb, Joanna Ellis, Maria Zambon
EPI_ISL_423359  hCoV-19/England/20139069104/2020        Europe / United Kingdom / England       2020-03-29
Respiratory Virus Unit, Microbiology Services Colindale, Public Health England  Respiratory Virus Unit, Microbiology Services Colindale, Public Health England  Monica Galiano, Shahjahan Miah, Angie Lackenby, Omolola Akinbami, Tiina Talts, Leena Bhaw, Richard Myers, Steven Platt, Kirstin Edwards, Jonathan Hubb, Joanna Ellis, Maria Zambon
EPI_ISL_423360  hCoV-19/England/20139069404/2020        Europe / United Kingdom / England       2020-03-29
Respiratory Virus Unit, Microbiology Services Colindale, Public Health England  Respiratory Virus Unit, Microbiology Services Colindale, Public Health England  Monica Galiano, Shahjahan Miah, Angie Lackenby, Omolola Akinbami, Tiina Talts, Leena Bhaw, Richard Myers, Steven Platt, Kirstin Edwards, Jonathan Hubb, Joanna Ellis, Maria Zambon
EPI_ISL_423361  hCoV-19/England/20139070204/2020        Europe / United Kingdom / England       2020-03-29
Respiratory Virus Unit, Microbiology Services Colindale, Public Health England  Respiratory Virus Unit, Microbiology Services Colindale, Public Health England  Monica Galiano, Shahjahan Miah, Angie Lackenby, Omolola Akinbami, Tiina Talts, Leena Bhaw, Richard Myers, Steven Platt, Kirstin Edwards, Jonathan Hubb, Joanna Ellis,

Maria Zambon
EPI_ISL_423362 hCoV-19/England/20139070504/2020 Europe / United Kingdom / England 2020-03-28 Respiratory Virus Unit, Microbiology Services Colindale, Public Health England Respiratory Virus Unit, Microbiology Services Colindale, Public Health England Monica Galiano, Shahjahan Miah, Angie Lackenby, Omolola Akinbami, Tiina Talts, Leena Bhaw, Richard Myers, Steven Platt, Kirstin Edwards, Jonathan Hubb, Joanna Ellis, Maria Zambon
EPI_ISL_423363 hCoV-19/England/20139070704/2020 Europe / United Kingdom / England 2020-03-29 Respiratory Virus Unit, Microbiology Services Colindale, Public Health England Respiratory Virus Unit, Microbiology Services Colindale, Public Health England Monica Galiano, Shahjahan Miah, Angie Lackenby, Omolola Akinbami, Tiina Talts, Leena Bhaw, Richard Myers, Steven Platt, Kirstin Edwards, Jonathan Hubb, Joanna Ellis, Maria Zambon
EPI_ISL_423364 hCoV-19/England/20139071704/2020 Europe / United Kingdom / England 2020-03-28 Respiratory Virus Unit, Microbiology Services Colindale, Public Health England Respiratory Virus Unit, Microbiology Services Colindale, Public Health England Monica Galiano, Shahjahan Miah, Angie Lackenby, Omolola Akinbami, Tiina Talts, Leena Bhaw, Richard Myers, Steven Platt, Kirstin Edwards, Jonathan Hubb, Joanna Ellis, Maria Zambon
EPI_ISL_423365 hCoV-19/England/20139072204/2020 Europe / United Kingdom / England 2020-03-29 Respiratory Virus Unit, Microbiology Services Colindale, Public Health England Respiratory Virus Unit, Microbiology Services Colindale, Public Health England Monica Galiano, Shahjahan Miah, Angie Lackenby, Omolola Akinbami, Tiina Talts, Leena Bhaw, Richard Myers, Steven Platt, Kirstin Edwards, Jonathan Hubb, Joanna Ellis, Maria Zambon
EPI_ISL_423366 hCoV-19/England/20140000304/2020 Europe / United Kingdom / England 2020-03-29 Respiratory Virus Unit, Microbiology Services Colindale, Public Health England Respiratory Virus Unit, Microbiology Services Colindale, Public Health England Monica Galiano, Shahjahan Miah, Angie Lackenby, Omolola Akinbami, Tiina Talts, Leena Bhaw, Richard Myers, Steven Platt, Kirstin Edwards, Jonathan Hubb, Joanna Ellis, Maria Zambon
EPI_ISL_423367 hCoV-19/England/20140000404/2020 Europe / United Kingdom / England 2020-03-29 Respiratory Virus Unit, Microbiology Services Colindale, Public Health England Respiratory Virus Unit, Microbiology Services Colindale, Public Health England Monica Galiano, Shahjahan Miah, Angie Lackenby, Omolola Akinbami, Tiina Talts, Leena Bhaw, Richard Myers, Steven Platt, Kirstin Edwards, Jonathan Hubb, Joanna Ellis, Maria Zambon
EPI_ISL_423368 hCoV-19/England/20140000604/2020 Europe / United Kingdom / England 2020-03-29 Respiratory Virus Unit, Microbiology Services Colindale, Public Health England Respiratory Virus Unit, Microbiology Services Colindale, Public Health England Monica Galiano, Shahjahan Miah, Angie Lackenby, Omolola Akinbami, Tiina Talts, Leena Bhaw, Richard Myers, Steven Platt, Kirstin Edwards, Jonathan Hubb, Joanna Ellis, Maria Zambon
EPI_ISL_423369 hCoV-19/England/20140000904/2020 Europe / United Kingdom / England 2020-03-29 Respiratory Virus Unit, Microbiology Services Colindale, Public Health England Respiratory Virus Unit, Microbiology Services Colindale, Public Health England Monica Galiano, Shahjahan Miah, Angie Lackenby, Omolola Akinbami, Tiina Talts, Leena Bhaw, Richard Myers, Steven Platt, Kirstin Edwards, Jonathan Hubb, Joanna Ellis, Maria Zambon
EPI_ISL_423403 hCoV-19/England/20140069104/2020 Europe / United Kingdom / England 2020-03-28 Respiratory Virus Unit, Microbiology Services Colindale, Public Health England Respiratory Virus Unit, Microbiology Services Colindale, Public Health England Monica Galiano, Shahjahan Miah, Angie Lackenby, Omolola Akinbami, Tiina Talts, Leena Bhaw, Richard Myers, Steven Platt, Kirstin Edwards, Jonathan Hubb, Joanna Ellis, Maria Zambon
EPI_ISL_423404 hCoV-19/England/20140069404/2020 Europe / United Kingdom / England 2020-03-30 Respiratory Virus Unit, Microbiology Services Colindale, Public Health England Respiratory Virus Unit, Microbiology Services Colindale, Public Health England Monica Galiano, Shahjahan Miah, Angie Lackenby, Omolola Akinbami, Tiina Talts, Leena Bhaw, Richard Myers, Steven Platt, Kirstin Edwards, Jonathan Hubb, Joanna Ellis, Maria Zambon
EPI_ISL_423405 hCoV-19/England/20140070004/2020 Europe / United Kingdom / England 2020-03-30 Respiratory Virus Unit, Microbiology Services Colindale, Public Health England Respiratory Virus Unit, Microbiology Services Colindale, Public Health England Monica Galiano, Shahjahan Miah, Angie Lackenby, Omolola Akinbami, Tiina Talts, Leena Bhaw, Richard Myers, Steven Platt, Kirstin Edwards, Jonathan Hubb, Joanna Ellis, Maria Zambon
EPI_ISL_423408 hCoV-19/England/20140070504/2020 Europe / United Kingdom / England 2020-03-30 Respiratory Virus Unit, Microbiology Services Colindale, Public Health England Respiratory Virus Unit, Microbiology Services Colindale, Public Health England Monica Galiano, Shahjahan Miah, Angie Lackenby, Omolola Akinbami, Tiina Talts, Leena Bhaw, Richard Myers, Steven Platt, Kirstin Edwards, Jonathan Hubb, Joanna Ellis, Maria Zambon
EPI_ISL_423409 hCoV-19/England/20140070604/2020 Europe / United Kingdom / England 2020-03-30 Respiratory Virus Unit, Microbiology Services Colindale, Public Health England Respiratory Virus Unit, Microbiology Services Colindale, Public Health England Monica Galiano, Shahjahan Miah, Angie Lackenby, Omolola Akinbami, Tiina Talts, Leena Bhaw, Richard Myers, Steven Platt, Kirstin Edwards, Jonathan Hubb, Joanna Ellis, Maria Zambon
EPI_ISL_423410 hCoV-19/England/20140070804/2020 Europe / United Kingdom / England 2020-03-30 Respiratory Virus Unit, Microbiology Services Colindale, Public Health England Respiratory Virus Unit, Microbiology Services Colindale, Public Health England Monica Galiano, Shahjahan Miah, Angie Lackenby, Omolola Akinbami, Tiina Talts, Leena Bhaw, Richard Myers, Steven Platt, Kirstin Edwards, Jonathan Hubb, Joanna Ellis,

Maria Zambon
EPI_ISL_423411  hCoV-19/England/20140071104/2020        Europe / United Kingdom / England       2020-03-28
Respiratory Virus Unit, Microbiology Services Colindale, Public Health England  Respiratory Virus Unit, Microbiology Services Colindale, Public Health England  Monica Galiano, Shahjahan Miah, Angie Lackenby, Omolola Akinbami, Tiina Talts, Leena Bhaw, Richard Myers, Steven Platt, Kirstin Edwards, Jonathan Hubb, Joanna Ellis, Maria Zambon
EPI_ISL_423412  hCoV-19/England/20140071404/2020        Europe / United Kingdom / England       2020-03-29
Respiratory Virus Unit, Microbiology Services Colindale, Public Health England  Respiratory Virus Unit, Microbiology Services Colindale, Public Health England  Monica Galiano, Shahjahan Miah, Angie Lackenby, Omolola Akinbami, Tiina Talts, Leena Bhaw, Richard Myers, Steven Platt, Kirstin Edwards, Jonathan Hubb, Joanna Ellis, Maria Zambon
EPI_ISL_423413  hCoV-19/England/20140071504/2020        Europe / United Kingdom / England       2020-03-29
Respiratory Virus Unit, Microbiology Services Colindale, Public Health England  Respiratory Virus Unit, Microbiology Services Colindale, Public Health England  Monica Galiano, Shahjahan Miah, Angie Lackenby, Omolola Akinbami, Tiina Talts, Leena Bhaw, Richard Myers, Steven Platt, Kirstin Edwards, Jonathan Hubb, Joanna Ellis, Maria Zambon
EPI_ISL_423414  hCoV-19/England/20140071604/2020        Europe / United Kingdom / England       2020-03-27
Respiratory Virus Unit, Microbiology Services Colindale, Public Health England  Respiratory Virus Unit, Microbiology Services Colindale, Public Health England  Monica Galiano, Shahjahan Miah, Angie Lackenby, Omolola Akinbami, Tiina Talts, Leena Bhaw, Richard Myers, Steven Platt, Kirstin Edwards, Jonathan Hubb, Joanna Ellis, Maria Zambon
EPI_ISL_423415  hCoV-19/England/20140071704/2020        Europe / United Kingdom / England       2020-03-29
Respiratory Virus Unit, Microbiology Services Colindale, Public Health England  Respiratory Virus Unit, Microbiology Services Colindale, Public Health England  Monica Galiano, Shahjahan Miah, Angie Lackenby, Omolola Akinbami, Tiina Talts, Leena Bhaw, Richard Myers, Steven Platt, Kirstin Edwards, Jonathan Hubb, Joanna Ellis, Maria Zambon
EPI_ISL_423417  hCoV-19/England/20140072304/2020        Europe / United Kingdom / England       2020-03-28
Respiratory Virus Unit, Microbiology Services Colindale, Public Health England  Respiratory Virus Unit, Microbiology Services Colindale, Public Health England  Monica Galiano, Shahjahan Miah, Angie Lackenby, Omolola Akinbami, Tiina Talts, Leena Bhaw, Richard Myers, Steven Platt, Kirstin Edwards, Jonathan Hubb, Joanna Ellis, Maria Zambon
EPI_ISL_423418  hCoV-19/England/20140072504/2020        Europe / United Kingdom / England       2020-03-28
Respiratory Virus Unit, Microbiology Services Colindale, Public Health England  Respiratory Virus Unit, Microbiology Services Colindale, Public Health England  Monica Galiano, Shahjahan Miah, Angie Lackenby, Omolola Akinbami, Tiina Talts, Leena Bhaw, Richard Myers, Steven Platt, Kirstin Edwards, Jonathan Hubb, Joanna Ellis, Maria Zambon
EPI_ISL_423419  hCoV-19/England/20140073904/2020        Europe / United Kingdom / England       2020-03-30
Respiratory Virus Unit, Microbiology Services Colindale, Public Health England  Respiratory Virus Unit, Microbiology Services Colindale, Public Health England  Monica Galiano, Shahjahan Miah, Angie Lackenby, Omolola Akinbami, Tiina Talts, Leena Bhaw, Richard Myers, Steven Platt, Kirstin Edwards, Jonathan Hubb, Joanna Ellis, Maria Zambon
EPI_ISL_423420  hCoV-19/England/20140074104/2020        Europe / United Kingdom / England       2020-03-29
Respiratory Virus Unit, Microbiology Services Colindale, Public Health England  Respiratory Virus Unit, Microbiology Services Colindale, Public Health England  Monica Galiano, Shahjahan Miah, Angie Lackenby, Omolola Akinbami, Tiina Talts, Leena Bhaw, Richard Myers, Steven Platt, Kirstin Edwards, Jonathan Hubb, Joanna Ellis, Maria Zambon
EPI_ISL_423421  hCoV-19/England/20142002004/2020        Europe / United Kingdom / England       2020-03-30
Respiratory Virus Unit, Microbiology Services Colindale, Public Health England  Respiratory Virus Unit, Microbiology Services Colindale, Public Health England  Monica Galiano, Shahjahan Miah, Angie Lackenby, Omolola Akinbami, Tiina Talts, Leena Bhaw, Richard Myers, Steven Platt, Kirstin Edwards, Jonathan Hubb, Joanna Ellis, Maria Zambon
EPI_ISL_423422  hCoV-19/England/20142002104/2020        Europe / United Kingdom / England       2020-03-30
Respiratory Virus Unit, Microbiology Services Colindale, Public Health England  Respiratory Virus Unit, Microbiology Services Colindale, Public Health England  Monica Galiano, Shahjahan Miah, Angie Lackenby, Omolola Akinbami, Tiina Talts, Leena Bhaw, Richard Myers, Steven Platt, Kirstin Edwards, Jonathan Hubb, Joanna Ellis, Maria Zambon
EPI_ISL_423423  hCoV-19/England/20142002204/2020        Europe / United Kingdom / England       2020-03-30
Respiratory Virus Unit, Microbiology Services Colindale, Public Health England  Respiratory Virus Unit, Microbiology Services Colindale, Public Health England  Monica Galiano, Shahjahan Miah, Angie Lackenby, Omolola Akinbami, Tiina Talts, Leena Bhaw, Richard Myers, Steven Platt, Kirstin Edwards, Jonathan Hubb, Joanna Ellis, Maria Zambon
EPI_ISL_423424  hCoV-19/England/20142002704/2020        Europe / United Kingdom / England       2020-03-30
Respiratory Virus Unit, Microbiology Services Colindale, Public Health England  Respiratory Virus Unit, Microbiology Services Colindale, Public Health England  Monica Galiano, Shahjahan Miah, Angie Lackenby, Omolola Akinbami, Tiina Talts, Leena Bhaw, Richard Myers, Steven Platt, Kirstin Edwards, Jonathan Hubb, Joanna Ellis, Maria Zambon
EPI_ISL_423425  hCoV-19/England/20142002904/2020        Europe / United Kingdom / England       2020-03-30
Respiratory Virus Unit, Microbiology Services Colindale, Public Health England  Respiratory Virus Unit, Microbiology Services Colindale, Public Health England  Monica Galiano, Shahjahan Miah, Angie Lackenby, Omolola Akinbami, Tiina Talts, Leena Bhaw, Richard Myers, Steven Platt, Kirstin Edwards, Jonathan Hubb, Joanna Ellis,

Maria Zambon
EPI_ISL_423426  hCoV-19/England/20142003904/2020        Europe / United Kingdom / England       2020-03-30
Respiratory Virus Unit, Microbiology Services Colindale, Public Health England  Respiratory Virus Unit,
Microbiology Services Colindale, Public Health England  Monica Galiano, Shahjahan Miah, Angie Lackenby, Omolola
Akinbami, Tiina Talts, Leena Bhaw, Richard Myers, Steven Platt, Kirstin Edwards, Jonathan Hubb, Joanna Ellis,
Maria Zambon
EPI_ISL_423428  hCoV-19/England/20142012304/2020        Europe / United Kingdom / England       2020-03-30
Respiratory Virus Unit, Microbiology Services Colindale, Public Health England  Respiratory Virus Unit,
Microbiology Services Colindale, Public Health England  Monica Galiano, Shahjahan Miah, Angie Lackenby, Omolola
Akinbami, Tiina Talts, Leena Bhaw, Richard Myers, Steven Platt, Kirstin Edwards, Jonathan Hubb, Joanna Ellis,
Maria Zambon
EPI_ISL_423430  hCoV-19/England/20142014504/2020        Europe / United Kingdom / England       2020-03-31
Respiratory Virus Unit, Microbiology Services Colindale, Public Health England  Respiratory Virus Unit,
Microbiology Services Colindale, Public Health England  Monica Galiano, Shahjahan Miah, Angie Lackenby, Omolola
Akinbami, Tiina Talts, Leena Bhaw, Richard Myers, Steven Platt, Kirstin Edwards, Jonathan Hubb, Joanna Ellis,
Maria Zambon
EPI_ISL_423432  hCoV-19/England/20142015104/2020        Europe / United Kingdom / England       2020-03-29
Respiratory Virus Unit, Microbiology Services Colindale, Public Health England  Respiratory Virus Unit,
Microbiology Services Colindale, Public Health England  Monica Galiano, Shahjahan Miah, Angie Lackenby, Omolola
Akinbami, Tiina Talts, Leena Bhaw, Richard Myers, Steven Platt, Kirstin Edwards, Jonathan Hubb, Joanna Ellis,
Maria Zambon
EPI_ISL_423436  hCoV-19/England/20142016504/2020        Europe / United Kingdom / England       2020-03-30
Respiratory Virus Unit, Microbiology Services Colindale, Public Health England  Respiratory Virus Unit,
Microbiology Services Colindale, Public Health England  Monica Galiano, Shahjahan Miah, Angie Lackenby, Omolola
Akinbami, Tiina Talts, Leena Bhaw, Richard Myers, Steven Platt, Kirstin Edwards, Jonathan Hubb, Joanna Ellis,
Maria Zambon
EPI_ISL_423437  hCoV-19/England/20142016704/2020        Europe / United Kingdom / England       2020-03-30
Respiratory Virus Unit, Microbiology Services Colindale, Public Health England  Respiratory Virus Unit,
Microbiology Services Colindale, Public Health England  Monica Galiano, Shahjahan Miah, Angie Lackenby, Omolola
Akinbami, Tiina Talts, Leena Bhaw, Richard Myers, Steven Platt, Kirstin Edwards, Jonathan Hubb, Joanna Ellis,
Maria Zambon
EPI_ISL_423438  hCoV-19/England/20142018004/2020        Europe / United Kingdom / England       2020-03-30
Respiratory Virus Unit, Microbiology Services Colindale, Public Health England  Respiratory Virus Unit,
Microbiology Services Colindale, Public Health England  Monica Galiano, Shahjahan Miah, Angie Lackenby, Omolola
Akinbami, Tiina Talts, Leena Bhaw, Richard Myers, Steven Platt, Kirstin Edwards, Jonathan Hubb, Joanna Ellis,
Maria Zambon
EPI_ISL_423440  hCoV-19/England/20142018604/2020        Europe / United Kingdom / England       2020-03-30
Respiratory Virus Unit, Microbiology Services Colindale, Public Health England  Respiratory Virus Unit,
Microbiology Services Colindale, Public Health England  Monica Galiano, Shahjahan Miah, Angie Lackenby, Omolola
Akinbami, Tiina Talts, Leena Bhaw, Richard Myers, Steven Platt, Kirstin Edwards, Jonathan Hubb, Joanna Ellis,
Maria Zambon
EPI_ISL_423441  hCoV-19/England/20142018804/2020        Europe / United Kingdom / England       2020-03-30
Respiratory Virus Unit, Microbiology Services Colindale, Public Health England  Respiratory Virus Unit,
Microbiology Services Colindale, Public Health England  Monica Galiano, Shahjahan Miah, Angie Lackenby, Omolola
Akinbami, Tiina Talts, Leena Bhaw, Richard Myers, Steven Platt, Kirstin Edwards, Jonathan Hubb, Joanna Ellis,
Maria Zambon
EPI_ISL_423442  hCoV-19/England/20142019004/2020        Europe / United Kingdom / England       2020-03-30
Respiratory Virus Unit, Microbiology Services Colindale, Public Health England  Respiratory Virus Unit,
Microbiology Services Colindale, Public Health England  Monica Galiano, Shahjahan Miah, Angie Lackenby, Omolola
Akinbami, Tiina Talts, Leena Bhaw, Richard Myers, Steven Platt, Kirstin Edwards, Jonathan Hubb, Joanna Ellis,
Maria Zambon
EPI_ISL_423443  hCoV-19/England/20142019504/2020        Europe / United Kingdom / England       2020-03-30
Respiratory Virus Unit, Microbiology Services Colindale, Public Health England  Respiratory Virus Unit,
Microbiology Services Colindale, Public Health England  Monica Galiano, Shahjahan Miah, Angie Lackenby, Omolola
Akinbami, Tiina Talts, Leena Bhaw, Richard Myers, Steven Platt, Kirstin Edwards, Jonathan Hubb, Joanna Ellis,
Maria Zambon
EPI_ISL_423444  hCoV-19/England/20142019704/2020        Europe / United Kingdom / England       2020-03-30
Respiratory Virus Unit, Microbiology Services Colindale, Public Health England  Respiratory Virus Unit,
Microbiology Services Colindale, Public Health England  Monica Galiano, Shahjahan Miah, Angie Lackenby, Omolola
Akinbami, Tiina Talts, Leena Bhaw, Richard Myers, Steven Platt, Kirstin Edwards, Jonathan Hubb, Joanna Ellis,
Maria Zambon
EPI_ISL_423446  hCoV-19/England/20142021604/2020        Europe / United Kingdom / England       2020-03-29
Respiratory Virus Unit, Microbiology Services Colindale, Public Health England  Respiratory Virus Unit,
Microbiology Services Colindale, Public Health England  Monica Galiano, Shahjahan Miah, Angie Lackenby, Omolola
Akinbami, Tiina Talts, Leena Bhaw, Richard Myers, Steven Platt, Kirstin Edwards, Jonathan Hubb, Joanna Ellis,
Maria Zambon
EPI_ISL_423447  hCoV-19/England/20142022104/2020        Europe / United Kingdom / England       2020-03-29
Respiratory Virus Unit, Microbiology Services Colindale, Public Health England  Respiratory Virus Unit,
Microbiology Services Colindale, Public Health England  Monica Galiano, Shahjahan Miah, Angie Lackenby, Omolola
Akinbami, Tiina Talts, Leena Bhaw, Richard Myers, Steven Platt, Kirstin Edwards, Jonathan Hubb, Joanna Ellis,

Maria Zambon
EPI_ISL_423448  hCoV-19/England/20142023704/2020        Europe / United Kingdom / England       2020-03-31
Respiratory Virus Unit, Microbiology Services Colindale, Public Health England  Respiratory Virus Unit, Microbiology Services Colindale, Public Health England  Monica Galiano, Shahjahan Miah, Angie Lackenby, Omolola Akinbami, Tiina Talts, Leena Bhaw, Richard Myers, Steven Platt, Kirstin Edwards, Jonathan Hubb, Joanna Ellis, Maria Zambon
EPI_ISL_423449  hCoV-19/England/20142023804/2020        Europe / United Kingdom / England       2020-03-29
Respiratory Virus Unit, Microbiology Services Colindale, Public Health England  Respiratory Virus Unit, Microbiology Services Colindale, Public Health England  Monica Galiano, Shahjahan Miah, Angie Lackenby, Omolola Akinbami, Tiina Talts, Leena Bhaw, Richard Myers, Steven Platt, Kirstin Edwards, Jonathan Hubb, Joanna Ellis, Maria Zambon
EPI_ISL_423450  hCoV-19/England/20142023904/2020        Europe / United Kingdom / England       2020-03-29
Respiratory Virus Unit, Microbiology Services Colindale, Public Health England  Respiratory Virus Unit, Microbiology Services Colindale, Public Health England  Monica Galiano, Shahjahan Miah, Angie Lackenby, Omolola Akinbami, Tiina Talts, Leena Bhaw, Richard Myers, Steven Platt, Kirstin Edwards, Jonathan Hubb, Joanna Ellis, Maria Zambon
EPI_ISL_423451  hCoV-19/England/20142024404/2020        Europe / United Kingdom / England       2020-03-29
Respiratory Virus Unit, Microbiology Services Colindale, Public Health England  Respiratory Virus Unit, Microbiology Services Colindale, Public Health England  Monica Galiano, Shahjahan Miah, Angie Lackenby, Omolola Akinbami, Tiina Talts, Leena Bhaw, Richard Myers, Steven Platt, Kirstin Edwards, Jonathan Hubb, Joanna Ellis, Maria Zambon
EPI_ISL_423452  hCoV-19/England/20142024704/2020        Europe / United Kingdom / England       2020-03-30
Respiratory Virus Unit, Microbiology Services Colindale, Public Health England  Respiratory Virus Unit, Microbiology Services Colindale, Public Health England  Monica Galiano, Shahjahan Miah, Angie Lackenby, Omolola Akinbami, Tiina Talts, Leena Bhaw, Richard Myers, Steven Platt, Kirstin Edwards, Jonathan Hubb, Joanna Ellis, Maria Zambon
EPI_ISL_423453  hCoV-19/England/20142026304/2020        Europe / United Kingdom / England       2020-03-29
Respiratory Virus Unit, Microbiology Services Colindale, Public Health England  Respiratory Virus Unit, Microbiology Services Colindale, Public Health England  Monica Galiano, Shahjahan Miah, Angie Lackenby, Omolola Akinbami, Tiina Talts, Leena Bhaw, Richard Myers, Steven Platt, Kirstin Edwards, Jonathan Hubb, Joanna Ellis, Maria Zambon
EPI_ISL_423455  hCoV-19/England/20142026804/2020        Europe / United Kingdom / England       2020-03-29
Respiratory Virus Unit, Microbiology Services Colindale, Public Health England  Respiratory Virus Unit, Microbiology Services Colindale, Public Health England  Monica Galiano, Shahjahan Miah, Angie Lackenby, Omolola Akinbami, Tiina Talts, Leena Bhaw, Richard Myers, Steven Platt, Kirstin Edwards, Jonathan Hubb, Joanna Ellis, Maria Zambon
EPI_ISL_423457  hCoV-19/England/20142053204/2020        Europe / United Kingdom / England       2020-03-28
Respiratory Virus Unit, Microbiology Services Colindale, Public Health England  Respiratory Virus Unit, Microbiology Services Colindale, Public Health England  Monica Galiano, Shahjahan Miah, Angie Lackenby, Omolola Akinbami, Tiina Talts, Leena Bhaw, Richard Myers, Steven Platt, Kirstin Edwards, Jonathan Hubb, Joanna Ellis, Maria Zambon
EPI_ISL_423458  hCoV-19/England/20142053804/2020        Europe / United Kingdom / England       2020-03-28
Respiratory Virus Unit, Microbiology Services Colindale, Public Health England  Respiratory Virus Unit, Microbiology Services Colindale, Public Health England  Monica Galiano, Shahjahan Miah, Angie Lackenby, Omolola Akinbami, Tiina Talts, Leena Bhaw, Richard Myers, Steven Platt, Kirstin Edwards, Jonathan Hubb, Joanna Ellis, Maria Zambon
EPI_ISL_423459  hCoV-19/England/20142078304/2020        Europe / United Kingdom / England       2020-03-30
Respiratory Virus Unit, Microbiology Services Colindale, Public Health England  Respiratory Virus Unit, Microbiology Services Colindale, Public Health England  Monica Galiano, Shahjahan Miah, Angie Lackenby, Omolola Akinbami, Tiina Talts, Leena Bhaw, Richard Myers, Steven Platt, Kirstin Edwards, Jonathan Hubb, Joanna Ellis, Maria Zambon
EPI_ISL_423460  hCoV-19/England/20142079504/2020        Europe / United Kingdom / England       2020-03-30
Respiratory Virus Unit, Microbiology Services Colindale, Public Health England  Respiratory Virus Unit, Microbiology Services Colindale, Public Health England  Monica Galiano, Shahjahan Miah, Angie Lackenby, Omolola Akinbami, Tiina Talts, Leena Bhaw, Richard Myers, Steven Platt, Kirstin Edwards, Jonathan Hubb, Joanna Ellis, Maria Zambon
EPI_ISL_423461  hCoV-19/England/20142079804/2020        Europe / United Kingdom / England       2020-03-30
Respiratory Virus Unit, Microbiology Services Colindale, Public Health England  Respiratory Virus Unit, Microbiology Services Colindale, Public Health England  Monica Galiano, Shahjahan Miah, Angie Lackenby, Omolola Akinbami, Tiina Talts, Leena Bhaw, Richard Myers, Steven Platt, Kirstin Edwards, Jonathan Hubb, Joanna Ellis, Maria Zambon
EPI_ISL_423463  hCoV-19/England/20142082704/2020        Europe / United Kingdom / England       2020-03-30
Respiratory Virus Unit, Microbiology Services Colindale, Public Health England  Respiratory Virus Unit, Microbiology Services Colindale, Public Health England  Monica Galiano, Shahjahan Miah, Angie Lackenby, Omolola Akinbami, Tiina Talts, Leena Bhaw, Richard Myers, Steven Platt, Kirstin Edwards, Jonathan Hubb, Joanna Ellis, Maria Zambon
EPI_ISL_423465  hCoV-19/England/20142083604/2020        Europe / United Kingdom / England       2020-03-30
Respiratory Virus Unit, Microbiology Services Colindale, Public Health England  Respiratory Virus Unit, Microbiology Services Colindale, Public Health England  Monica Galiano, Shahjahan Miah, Angie Lackenby, Omolola Akinbami, Tiina Talts, Leena Bhaw, Richard Myers, Steven Platt, Kirstin Edwards, Jonathan Hubb, Joanna Ellis,

Maria Zambon
EPI_ISL_423466 hCoV-19/England/20142083804/2020 Europe / United Kingdom / England 2020-03-30 Respiratory Virus Unit, Microbiology Services Colindale, Public Health England Respiratory Virus Unit, Microbiology Services Colindale, Public Health England Monica Galiano, Shahjahan Miah, Angie Lackenby, Omolola Akinbami, Tiina Talts, Leena Bhaw, Richard Myers, Steven Platt, Kirstin Edwards, Jonathan Hubb, Joanna Ellis, Maria Zambon
EPI_ISL_423467 hCoV-19/England/20142130404/2020 Europe / United Kingdom / England 2020-03-30 Respiratory Virus Unit, Microbiology Services Colindale, Public Health England Respiratory Virus Unit, Microbiology Services Colindale, Public Health England Monica Galiano, Shahjahan Miah, Angie Lackenby, Omolola Akinbami, Tiina Talts, Leena Bhaw, Richard Myers, Steven Platt, Kirstin Edwards, Jonathan Hubb, Joanna Ellis, Maria Zambon
EPI_ISL_423469 hCoV-19/England/20142131504/2020 Europe / United Kingdom / England 2020-03-31 Respiratory Virus Unit, Microbiology Services Colindale, Public Health England Respiratory Virus Unit, Microbiology Services Colindale, Public Health England Monica Galiano, Shahjahan Miah, Angie Lackenby, Omolola Akinbami, Tiina Talts, Leena Bhaw, Richard Myers, Steven Platt, Kirstin Edwards, Jonathan Hubb, Joanna Ellis, Maria Zambon
EPI_ISL_423472 hCoV-19/England/20144000204/2020 Europe / United Kingdom / England 2020-03-31 Respiratory Virus Unit, Microbiology Services Colindale, Public Health England Respiratory Virus Unit, Microbiology Services Colindale, Public Health England Monica Galiano, Shahjahan Miah, Angie Lackenby, Omolola Akinbami, Tiina Talts, Leena Bhaw, Richard Myers, Steven Platt, Kirstin Edwards, Jonathan Hubb, Joanna Ellis, Maria Zambon
EPI_ISL_423473 hCoV-19/England/20144000404/2020 Europe / United Kingdom / England 2020-03-31 Respiratory Virus Unit, Microbiology Services Colindale, Public Health England Respiratory Virus Unit, Microbiology Services Colindale, Public Health England Monica Galiano, Shahjahan Miah, Angie Lackenby, Omolola Akinbami, Tiina Talts, Leena Bhaw, Richard Myers, Steven Platt, Kirstin Edwards, Jonathan Hubb, Joanna Ellis, Maria Zambon
EPI_ISL_423476 hCoV-19/England/20144001204/2020 Europe / United Kingdom / England 2020-03-30 Respiratory Virus Unit, Microbiology Services Colindale, Public Health England Respiratory Virus Unit, Microbiology Services Colindale, Public Health England Monica Galiano, Shahjahan Miah, Angie Lackenby, Omolola Akinbami, Tiina Talts, Leena Bhaw, Richard Myers, Steven Platt, Kirstin Edwards, Jonathan Hubb, Joanna Ellis, Maria Zambon
EPI_ISL_423477 hCoV-19/England/20144001404/2020 Europe / United Kingdom / England 2020-03-30 Respiratory Virus Unit, Microbiology Services Colindale, Public Health England Respiratory Virus Unit, Microbiology Services Colindale, Public Health England Monica Galiano, Shahjahan Miah, Angie Lackenby, Omolola Akinbami, Tiina Talts, Leena Bhaw, Richard Myers, Steven Platt, Kirstin Edwards, Jonathan Hubb, Joanna Ellis, Maria Zambon
EPI_ISL_423479 hCoV-19/England/20144002904/2020 Europe / United Kingdom / England 2020-03-31 Respiratory Virus Unit, Microbiology Services Colindale, Public Health England Respiratory Virus Unit, Microbiology Services Colindale, Public Health England Monica Galiano, Shahjahan Miah, Angie Lackenby, Omolola Akinbami, Tiina Talts, Leena Bhaw, Richard Myers, Steven Platt, Kirstin Edwards, Jonathan Hubb, Joanna Ellis, Maria Zambon
EPI_ISL_423480 hCoV-19/England/20144003404/2020 Europe / United Kingdom / England 2020-03-31 Respiratory Virus Unit, Microbiology Services Colindale, Public Health England Respiratory Virus Unit, Microbiology Services Colindale, Public Health England Monica Galiano, Shahjahan Miah, Angie Lackenby, Omolola Akinbami, Tiina Talts, Leena Bhaw, Richard Myers, Steven Platt, Kirstin Edwards, Jonathan Hubb, Joanna Ellis, Maria Zambon
EPI_ISL_423481 hCoV-19/England/20144003604/2020 Europe / United Kingdom / England 2020-03-31 Respiratory Virus Unit, Microbiology Services Colindale, Public Health England Respiratory Virus Unit, Microbiology Services Colindale, Public Health England Monica Galiano, Shahjahan Miah, Angie Lackenby, Omolola Akinbami, Tiina Talts, Leena Bhaw, Richard Myers, Steven Platt, Kirstin Edwards, Jonathan Hubb, Joanna Ellis, Maria Zambon
EPI_ISL_423483 hCoV-19/England/20144003804/2020 Europe / United Kingdom / England 2020-04-01 Respiratory Virus Unit, Microbiology Services Colindale, Public Health England Respiratory Virus Unit, Microbiology Services Colindale, Public Health England Monica Galiano, Shahjahan Miah, Angie Lackenby, Omolola Akinbami, Tiina Talts, Leena Bhaw, Richard Myers, Steven Platt, Kirstin Edwards, Jonathan Hubb, Joanna Ellis, Maria Zambon
EPI_ISL_423486 hCoV-19/England/20144004504/2020 Europe / United Kingdom / England 2020-03-31 Respiratory Virus Unit, Microbiology Services Colindale, Public Health England Respiratory Virus Unit, Microbiology Services Colindale, Public Health England Monica Galiano, Shahjahan Miah, Angie Lackenby, Omolola Akinbami, Tiina Talts, Leena Bhaw, Richard Myers, Steven Platt, Kirstin Edwards, Jonathan Hubb, Joanna Ellis, Maria Zambon
EPI_ISL_423487 hCoV-19/England/20144004704/2020 Europe / United Kingdom / England 2020-03-31 Respiratory Virus Unit, Microbiology Services Colindale, Public Health England Respiratory Virus Unit, Microbiology Services Colindale, Public Health England Monica Galiano, Shahjahan Miah, Angie Lackenby, Omolola Akinbami, Tiina Talts, Leena Bhaw, Richard Myers, Steven Platt, Kirstin Edwards, Jonathan Hubb, Joanna Ellis, Maria Zambon
EPI_ISL_423488 hCoV-19/England/20144004804/2020 Europe / United Kingdom / England 2020-03-31 Respiratory Virus Unit, Microbiology Services Colindale, Public Health England Respiratory Virus Unit, Microbiology Services Colindale, Public Health England Monica Galiano, Shahjahan Miah, Angie Lackenby, Omolola Akinbami, Tiina Talts, Leena Bhaw, Richard Myers, Steven Platt, Kirstin Edwards, Jonathan Hubb, Joanna Ellis,

Maria Zambon
EPI_ISL_423491 hCoV-19/England/20144005904/2020 Europe / United Kingdom / England 2020-03-31 Respiratory Virus Unit, Microbiology Services Colindale, Public Health England Respiratory Virus Unit, Microbiology Services Colindale, Public Health England Monica Galiano, Shahjahan Miah, Angie Lackenby, Omolola Akinbami, Tiina Talts, Leena Bhaw, Richard Myers, Steven Platt, Kirstin Edwards, Jonathan Hubb, Joanna Ellis, Maria Zambon
EPI_ISL_423492 hCoV-19/England/20144006004/2020 Europe / United Kingdom / England 2020-03-31 Respiratory Virus Unit, Microbiology Services Colindale, Public Health England Respiratory Virus Unit, Microbiology Services Colindale, Public Health England Monica Galiano, Shahjahan Miah, Angie Lackenby, Omolola Akinbami, Tiina Talts, Leena Bhaw, Richard Myers, Steven Platt, Kirstin Edwards, Jonathan Hubb, Joanna Ellis, Maria Zambon
EPI_ISL_423493 hCoV-19/England/20144006304/2020 Europe / United Kingdom / England 2020-03-31 Respiratory Virus Unit, Microbiology Services Colindale, Public Health England Respiratory Virus Unit, Microbiology Services Colindale, Public Health England Monica Galiano, Shahjahan Miah, Angie Lackenby, Omolola Akinbami, Tiina Talts, Leena Bhaw, Richard Myers, Steven Platt, Kirstin Edwards, Jonathan Hubb, Joanna Ellis, Maria Zambon
EPI_ISL_423494 hCoV-19/England/20144006404/2020 Europe / United Kingdom / England 2020-03-31 Respiratory Virus Unit, Microbiology Services Colindale, Public Health England Respiratory Virus Unit, Microbiology Services Colindale, Public Health England Monica Galiano, Shahjahan Miah, Angie Lackenby, Omolola Akinbami, Tiina Talts, Leena Bhaw, Richard Myers, Steven Platt, Kirstin Edwards, Jonathan Hubb, Joanna Ellis, Maria Zambon
EPI_ISL_423495 hCoV-19/England/20144006704/2020 Europe / United Kingdom / England 2020-03-30 Respiratory Virus Unit, Microbiology Services Colindale, Public Health England Respiratory Virus Unit, Microbiology Services Colindale, Public Health England Monica Galiano, Shahjahan Miah, Angie Lackenby, Omolola Akinbami, Tiina Talts, Leena Bhaw, Richard Myers, Steven Platt, Kirstin Edwards, Jonathan Hubb, Joanna Ellis, Maria Zambon
EPI_ISL_423496 hCoV-19/England/20144006904/2020 Europe / United Kingdom / England 2020-03-30 Respiratory Virus Unit, Microbiology Services Colindale, Public Health England Respiratory Virus Unit, Microbiology Services Colindale, Public Health England Monica Galiano, Shahjahan Miah, Angie Lackenby, Omolola Akinbami, Tiina Talts, Leena Bhaw, Richard Myers, Steven Platt, Kirstin Edwards, Jonathan Hubb, Joanna Ellis, Maria Zambon
EPI_ISL_423497 hCoV-19/England/20144007604/2020 Europe / United Kingdom / England 2020-03-31 Respiratory Virus Unit, Microbiology Services Colindale, Public Health England Respiratory Virus Unit, Microbiology Services Colindale, Public Health England Monica Galiano, Shahjahan Miah, Angie Lackenby, Omolola Akinbami, Tiina Talts, Leena Bhaw, Richard Myers, Steven Platt, Kirstin Edwards, Jonathan Hubb, Joanna Ellis, Maria Zambon
EPI_ISL_423498 hCoV-19/England/20144008004/2020 Europe / United Kingdom / England 2020-03-31 Respiratory Virus Unit, Microbiology Services Colindale, Public Health England Respiratory Virus Unit, Microbiology Services Colindale, Public Health England Monica Galiano, Shahjahan Miah, Angie Lackenby, Omolola Akinbami, Tiina Talts, Leena Bhaw, Richard Myers, Steven Platt, Kirstin Edwards, Jonathan Hubb, Joanna Ellis, Maria Zambon
EPI_ISL_423499 hCoV-19/England/20144008204/2020 Europe / United Kingdom / England 2020-03-31 Respiratory Virus Unit, Microbiology Services Colindale, Public Health England Respiratory Virus Unit, Microbiology Services Colindale, Public Health England Monica Galiano, Shahjahan Miah, Angie Lackenby, Omolola Akinbami, Tiina Talts, Leena Bhaw, Richard Myers, Steven Platt, Kirstin Edwards, Jonathan Hubb, Joanna Ellis, Maria Zambon
EPI_ISL_423500 hCoV-19/England/20144008304/2020 Europe / United Kingdom / England 2020-03-31 Respiratory Virus Unit, Microbiology Services Colindale, Public Health England Respiratory Virus Unit, Microbiology Services Colindale, Public Health England Monica Galiano, Shahjahan Miah, Angie Lackenby, Omolola Akinbami, Tiina Talts, Leena Bhaw, Richard Myers, Steven Platt, Kirstin Edwards, Jonathan Hubb, Joanna Ellis, Maria Zambon
EPI_ISL_423502 hCoV-19/England/20144008904/2020 Europe / United Kingdom / England 2020-03-31 Respiratory Virus Unit, Microbiology Services Colindale, Public Health England Respiratory Virus Unit, Microbiology Services Colindale, Public Health England Monica Galiano, Shahjahan Miah, Angie Lackenby, Omolola Akinbami, Tiina Talts, Leena Bhaw, Richard Myers, Steven Platt, Kirstin Edwards, Jonathan Hubb, Joanna Ellis, Maria Zambon
EPI_ISL_423503 hCoV-19/England/20144009004/2020 Europe / United Kingdom / England 2020-03-31 Respiratory Virus Unit, Microbiology Services Colindale, Public Health England Respiratory Virus Unit, Microbiology Services Colindale, Public Health England Monica Galiano, Shahjahan Miah, Angie Lackenby, Omolola Akinbami, Tiina Talts, Leena Bhaw, Richard Myers, Steven Platt, Kirstin Edwards, Jonathan Hubb, Joanna Ellis, Maria Zambon
EPI_ISL_423504 hCoV-19/England/20144009104/2020 Europe / United Kingdom / England 2020-03-31 Respiratory Virus Unit, Microbiology Services Colindale, Public Health England Respiratory Virus Unit, Microbiology Services Colindale, Public Health England Monica Galiano, Shahjahan Miah, Angie Lackenby, Omolola Akinbami, Tiina Talts, Leena Bhaw, Richard Myers, Steven Platt, Kirstin Edwards, Jonathan Hubb, Joanna Ellis, Maria Zambon
EPI_ISL_423506 hCoV-19/England/20144011504/2020 Europe / United Kingdom / England 2020-03-30 Respiratory Virus Unit, Microbiology Services Colindale, Public Health England Respiratory Virus Unit, Microbiology Services Colindale, Public Health England Monica Galiano, Shahjahan Miah, Angie Lackenby, Omolola Akinbami, Tiina Talts, Leena Bhaw, Richard Myers, Steven Platt, Kirstin Edwards, Jonathan Hubb, Joanna Ellis,

Maria Zambon
EPI_ISL_423507 hCoV-19/England/20144012404/2020 Europe / United Kingdom / England 2020-03-30 Respiratory Virus Unit, Microbiology Services Colindale, Public Health England Respiratory Virus Unit, Microbiology Services Colindale, Public Health England Monica Galiano, Shahjahan Miah, Angie Lackenby, Omolola Akinbami, Tiina Talts, Leena Bhaw, Richard Myers, Steven Platt, Kirstin Edwards, Jonathan Hubb, Joanna Ellis, Maria Zambon
EPI_ISL_423508 hCoV-19/England/20144013704/2020 Europe / United Kingdom / England 2020-03-31 Respiratory Virus Unit, Microbiology Services Colindale, Public Health England Respiratory Virus Unit, Microbiology Services Colindale, Public Health England Monica Galiano, Shahjahan Miah, Angie Lackenby, Omolola Akinbami, Tiina Talts, Leena Bhaw, Richard Myers, Steven Platt, Kirstin Edwards, Jonathan Hubb, Joanna Ellis, Maria Zambon
EPI_ISL_423509 hCoV-19/England/20144014504/2020 Europe / United Kingdom / England 2020-03-31 Respiratory Virus Unit, Microbiology Services Colindale, Public Health England Respiratory Virus Unit, Microbiology Services Colindale, Public Health England Monica Galiano, Shahjahan Miah, Angie Lackenby, Omolola Akinbami, Tiina Talts, Leena Bhaw, Richard Myers, Steven Platt, Kirstin Edwards, Jonathan Hubb, Joanna Ellis, Maria Zambon
EPI_ISL_423512 hCoV-19/England/20144018904/2020 Europe / United Kingdom / England 2020-03-31 Respiratory Virus Unit, Microbiology Services Colindale, Public Health England Respiratory Virus Unit, Microbiology Services Colindale, Public Health England Monica Galiano, Shahjahan Miah, Angie Lackenby, Omolola Akinbami, Tiina Talts, Leena Bhaw, Richard Myers, Steven Platt, Kirstin Edwards, Jonathan Hubb, Joanna Ellis, Maria Zambon
EPI_ISL_424613 hCoV-19/Iceland/593/2020 Europe / Iceland / Reykjavik 2020-03-27 The National University Hospital of Iceland deCODE genetics Daniel F Gudbjartsson; Agnar Helgason; Hakon Jonsson; Olafur T Magnusson; Pall Melsted; Gudmundur L Norddahl; Jona Saemundsdottir; Asgeir Sigurdsson; Patrick Sulem; Arna B Agustsdottir; Berglind Eiriksdottir; Run Fridriksdottir; Elisabet E Gardarsdottir; Gudmundur Georgsson; Olafia S Gretarsdottir; Kjartan R Gudmundsson; Thora R Gunnarsdottir; Arnaldur Gylfason; Hilma Holm; Brynjar O Jensson; Aslaug Jonasdottir; Kamilla S Josefsdottir; Thordur Kristjansson; Droplaug N Magnusdottir; Louise le Roux; Gudrun Sigmundsdottir; Gardar Sveinbjornsson; Kristin E Sveinsdottir; Maney Sveinsdottir; Emil A Thorarensen; Bjarni Thorbjornsson; Gisli Masson; Ingileif Jonsdottir; Alma Moller; Thorolfur Gudnason; Karl G Kristinsson; Unnur Thorsteinsdottir; Kari Stefansson
EPI_ISL_424614 hCoV-19/Iceland/594/2020 Europe / Iceland / Reykjavik 2020-03-27 The National University Hospital of Iceland deCODE genetics Daniel F Gudbjartsson; Agnar Helgason; Hakon Jonsson; Olafur T Magnusson; Pall Melsted; Gudmundur L Norddahl; Jona Saemundsdottir; Asgeir Sigurdsson; Patrick Sulem; Arna B Agustsdottir; Berglind Eiriksdottir; Run Fridriksdottir; Elisabet E Gardarsdottir; Gudmundur Georgsson; Olafia S Gretarsdottir; Kjartan R Gudmundsson; Thora R Gunnarsdottir; Arnaldur Gylfason; Hilma Holm; Brynjar O Jensson; Aslaug Jonasdottir; Kamilla S Josefsdottir; Thordur Kristjansson; Droplaug N Magnusdottir; Louise le Roux; Gudrun Sigmundsdottir; Gardar Sveinbjornsson; Kristin E Sveinsdottir; Maney Sveinsdottir; Emil A Thorarensen; Bjarni Thorbjornsson; Gisli Masson; Ingileif Jonsdottir; Alma Moller; Thorolfur Gudnason; Karl G Kristinsson; Unnur Thorsteinsdottir; Kari Stefansson
EPI_ISL_424615 hCoV-19/Iceland/595/2020 Europe / Iceland / Reykjavik 2020-03-27 The National University Hospital of Iceland deCODE genetics Daniel F Gudbjartsson; Agnar Helgason; Hakon Jonsson; Olafur T Magnusson; Pall Melsted; Gudmundur L Norddahl; Jona Saemundsdottir; Asgeir Sigurdsson; Patrick Sulem; Arna B Agustsdottir; Berglind Eiriksdottir; Run Fridriksdottir; Elisabet E Gardarsdottir; Gudmundur Georgsson; Olafia S Gretarsdottir; Kjartan R Gudmundsson; Thora R Gunnarsdottir; Arnaldur Gylfason; Hilma Holm; Brynjar O Jensson; Aslaug Jonasdottir; Kamilla S Josefsdottir; Thordur Kristjansson; Droplaug N Magnusdottir; Louise le Roux; Gudrun Sigmundsdottir; Gardar Sveinbjornsson; Kristin E Sveinsdottir; Maney Sveinsdottir; Emil A Thorarensen; Bjarni Thorbjornsson; Gisli Masson; Ingileif Jonsdottir; Alma Moller; Thorolfur Gudnason; Karl G Kristinsson; Unnur Thorsteinsdottir; Kari Stefansson
EPI_ISL_424616 hCoV-19/Iceland/596/2020 Europe / Iceland / Reykjavik 2020-03-28 The National University Hospital of Iceland deCODE genetics Daniel F Gudbjartsson; Agnar Helgason; Hakon Jonsson; Olafur T Magnusson; Pall Melsted; Gudmundur L Norddahl; Jona Saemundsdottir; Asgeir Sigurdsson; Patrick Sulem; Arna B Agustsdottir; Berglind Eiriksdottir; Run Fridriksdottir; Elisabet E Gardarsdottir; Gudmundur Georgsson; Olafia S Gretarsdottir; Kjartan R Gudmundsson; Thora R Gunnarsdottir; Arnaldur Gylfason; Hilma Holm; Brynjar O Jensson; Aslaug Jonasdottir; Kamilla S Josefsdottir; Thordur Kristjansson; Droplaug N Magnusdottir; Louise le Roux; Gudrun Sigmundsdottir; Gardar Sveinbjornsson; Kristin E Sveinsdottir; Maney Sveinsdottir; Emil A Thorarensen; Bjarni Thorbjornsson; Gisli Masson; Ingileif Jonsdottir; Alma Moller; Thorolfur Gudnason; Karl G Kristinsson; Unnur Thorsteinsdottir; Kari Stefansson
EPI_ISL_424617 hCoV-19/Iceland/597/2020 Europe / Iceland / Reykjavik 2020-03-28 The National University Hospital of Iceland deCODE genetics Daniel F Gudbjartsson; Agnar Helgason; Hakon Jonsson; Olafur T Magnusson; Pall Melsted; Gudmundur L Norddahl; Jona Saemundsdottir; Asgeir Sigurdsson; Patrick Sulem; Arna B Agustsdottir; Berglind Eiriksdottir; Run Fridriksdottir; Elisabet E Gardarsdottir; Gudmundur Georgsson; Olafia S Gretarsdottir; Kjartan R Gudmundsson; Thora R Gunnarsdottir; Arnaldur Gylfason; Hilma Holm; Brynjar O Jensson; Aslaug Jonasdottir; Kamilla S Josefsdottir; Thordur Kristjansson; Droplaug N Magnusdottir; Louise le Roux; Gudrun Sigmundsdottir; Gardar Sveinbjornsson; Kristin E Sveinsdottir; Maney Sveinsdottir; Emil A Thorarensen; Bjarni Thorbjornsson; Gisli Masson; Ingileif Jonsdottir; Alma Moller; Thorolfur Gudnason; Karl G Kristinsson; Unnur Thorsteinsdottir; Kari Stefansson
EPI_ISL_424619 hCoV-19/Iceland/599/2020 Europe / Iceland / Reykjavik 2020-03-28 The National University Hospital of Iceland deCODE genetics Daniel F Gudbjartsson; Agnar Helgason; Hakon Jonsson; Olafur T Magnusson; Pall Melsted; Gudmundur L Norddahl; Jona Saemundsdottir; Asgeir Sigurdsson; Patrick Sulem; Arna B Agustsdottir; Berglind Eiriksdottir; Run Fridriksdottir; Elisabet E Gardarsdottir; Gudmundur Georgsson; Olafia

S Gretarsdottir; Kjartan R Gudmundsson; Thora R Gunnarsdottir; Arnaldur Gylfason; Hilma Holm; Brynjar O Jensson; Aslaug Jonasdottir; Kamilla S Josefsdottir; Thordur Kristjansson; Droplaug N Magnusdottir; Louise le Roux; Gudrun Sigmundsdottir; Gardar Sveinbjornsson; Kristin E Sveinsdottir; Maney Sveinsdottir; Emil A Thorarensen; Bjarni Thorbjornsson; Gisli Masson; Ingileif Jonsdottir; Alma Moller; Thorolfur Gudnason; Karl G Kristinsson; Unnur Thorsteinsdottir; Kari Stefansson
EPI_ISL_424621 hCoV-19/Iceland/601/2020 Europe / Iceland / Reykjavik 2020-03-28 The National University Hospital of Iceland deCODE genetics Daniel F Gudbjartsson; Agnar Helgason; Hakon Jonsson; Olafur T Magnusson; Pall Melsted; Gudmundur L Norddahl; Jona Saemundsdottir; Asgeir Sigurdsson; Patrick Sulem; Arna B Agustsdottir; Berglind Eiriksdottir; Run Fridriksdottir; Elisabet E Gardarsdottir; Gudmundur Georgsson; Olafia S Gretarsdottir; Kjartan R Gudmundsson; Thora R Gunnarsdottir; Arnaldur Gylfason; Hilma Holm; Brynjar O Jensson; Aslaug Jonasdottir; Kamilla S Josefsdottir; Thordur Kristjansson; Droplaug N Magnusdottir; Louise le Roux; Gudrun Sigmundsdottir; Gardar Sveinbjornsson; Kristin E Sveinsdottir; Maney Sveinsdottir; Emil A Thorarensen; Bjarni Thorbjornsson; Gisli Masson; Ingileif Jonsdottir; Alma Moller; Thorolfur Gudnason; Karl G Kristinsson; Unnur Thorsteinsdottir; Kari Stefansson
EPI_ISL_424622 hCoV-19/Iceland/602/2020 Europe / Iceland / Reykjavik 2020-03-28 The National University Hospital of Iceland deCODE genetics Daniel F Gudbjartsson; Agnar Helgason; Hakon Jonsson; Olafur T Magnusson; Pall Melsted; Gudmundur L Norddahl; Jona Saemundsdottir; Asgeir Sigurdsson; Patrick Sulem; Arna B Agustsdottir; Berglind Eiriksdottir; Run Fridriksdottir; Elisabet E Gardarsdottir; Gudmundur Georgsson; Olafia S Gretarsdottir; Kjartan R Gudmundsson; Thora R Gunnarsdottir; Arnaldur Gylfason; Hilma Holm; Brynjar O Jensson; Aslaug Jonasdottir; Kamilla S Josefsdottir; Thordur Kristjansson; Droplaug N Magnusdottir; Louise le Roux; Gudrun Sigmundsdottir; Gardar Sveinbjornsson; Kristin E Sveinsdottir; Maney Sveinsdottir; Emil A Thorarensen; Bjarni Thorbjornsson; Gisli Masson; Ingileif Jonsdottir; Alma Moller; Thorolfur Gudnason; Karl G Kristinsson; Unnur Thorsteinsdottir; Kari Stefansson
EPI_ISL_424624 hCoV-19/Iceland/604/2020 Europe / Iceland / Reykjavik 2020-03-28 The National University Hospital of Iceland deCODE genetics Daniel F Gudbjartsson; Agnar Helgason; Hakon Jonsson; Olafur T Magnusson; Pall Melsted; Gudmundur L Norddahl; Jona Saemundsdottir; Asgeir Sigurdsson; Patrick Sulem; Arna B Agustsdottir; Berglind Eiriksdottir; Run Fridriksdottir; Elisabet E Gardarsdottir; Gudmundur Georgsson; Olafia S Gretarsdottir; Kjartan R Gudmundsson; Thora R Gunnarsdottir; Arnaldur Gylfason; Hilma Holm; Brynjar O Jensson; Aslaug Jonasdottir; Kamilla S Josefsdottir; Thordur Kristjansson; Droplaug N Magnusdottir; Louise le Roux; Gudrun Sigmundsdottir; Gardar Sveinbjornsson; Kristin E Sveinsdottir; Maney Sveinsdottir; Emil A Thorarensen; Bjarni Thorbjornsson; Gisli Masson; Ingileif Jonsdottir; Alma Moller; Thorolfur Gudnason; Karl G Kristinsson; Unnur Thorsteinsdottir; Kari Stefansson
EPI_ISL_424626 hCoV-19/Mexico/CDMX-INER_04/2020 North America / Mexico / Mexico City 2020-03-15 Instituto Nacional de Enfermedades Respiratorias Instituto Nacional de Enfermedades Respiratorias Joel Armando Vázquez Pérez, Celia Boukadida, Santiago Avila Ríos, Mario Mújica Sánchez, José Arturo Martínez Orozco, Eduardo Becerril Vargas, Jorge Salas Hernández, Irma López Martínez, Lucia Hernández Rivas, Gisela Barrera Badillo, Edgar Mendieta Condado, Fabiola Garcés Ayala, Adnan Araiza Rodríguez, José Ernesto Ramírez González, Victor Hugo Borja Aburto, Concepción Grajales Muñiz, Cesar Raúl González Bonilla, Carolina González Torres, Francisco Javier Gaytán Cervantes, José Esteban Muñoz Medina, Guillermo M. Ruiz-Palacios, Pilar Ramos Cervantes, Violeta Ibarra Gonzalez, Fernando Ledesma Barrientos, Luis Alberto García Andrade, Alfredo Ponce de León Garduño, Blanca Taboada, Alejandro Sánchez, Pavel Isa, Ricardo Grande, Gloria Vázquez, Francisco Pulido, Carlos F. Arias.
EPI_ISL_424627 hCoV-19/Mexico/CDMX-INER_05/2020 North America / Mexico / Mexico City 2020-03-16 Instituto Nacional de Enfermedades Respiratorias Instituto Nacional de Enfermedades Respiratorias Joel Armando Vázquez Pérez, Celia Boukadida, Santiago Avila Ríos, Mario Mújica Sánchez, José Arturo Martínez Orozco, Eduardo Becerril Vargas, Jorge Salas Hernández, Irma López Martínez, Lucia Hernández Rivas, Gisela Barrera Badillo, Edgar Mendieta Condado, Fabiola Garcés Ayala, Adnan Araiza Rodríguez, José Ernesto Ramírez González, Victor Hugo Borja Aburto, Concepción Grajales Muñiz, Cesar Raúl González Bonilla, Carolina González Torres, Francisco Javier Gaytán Cervantes, José Esteban Muñoz Medina, Guillermo M. Ruiz-Palacios, Pilar Ramos Cervantes, Violeta Ibarra Gonzalez, Fernando Ledesma Barrientos, Luis Alberto García Andrade, Alfredo Ponce de León Garduño, Blanca Taboada, Alejandro Sánchez, Pavel Isa, Ricardo Grande, Gloria Vázquez, Francisco Pulido, Carlos F. Arias.
EPI_ISL_424629 hCoV-19/Belgium/ULG-10066/2020 Europe / Belgium / Liège 2020-04-03 Department of Clinical Microbiology GIGA Medical Genomics Keith Durkin, Maria Artesi, Sébastien Bontems, Raphaël Boreux, Cécile Meex, Pierrette Melin, Marie-Pierre Hayette, Vincent Bours.
EPI_ISL_424630 hCoV-19/Belgium/ULG-10067/2020 Europe / Belgium / Liège 2020-04-03 Department of Clinical Microbiology GIGA Medical Genomics Keith Durkin, Maria Artesi, Sébastien Bontems, Raphaël Boreux, Cécile Meex, Pierrette Melin, Marie-Pierre Hayette, Vincent Bours.
EPI_ISL_424631 hCoV-19/Belgium/ULG-10072/2020 Europe / Belgium / Liège 2020-04-03 Department of Clinical Microbiology GIGA Medical Genomics Keith Durkin, Maria Artesi, Sébastien Bontems, Raphaël Boreux, Cécile Meex, Pierrette Melin, Marie-Pierre Hayette, Vincent Bours.
EPI_ISL_424633 hCoV-19/Belgium/ULG-10069/2020 Europe / Belgium / Liège 2020-04-03 Department of Clinical Microbiology GIGA Medical Genomics Keith Durkin, Maria Artesi, Sébastien Bontems, Raphaël Boreux, Cécile Meex, Pierrette Melin, Marie-Pierre Hayette, Vincent Bours.
EPI_ISL_424636 hCoV-19/Belgium/ULG-10068/2020 Europe / Belgium / Liège 2020-04-03 Department of Clinical Microbiology GIGA Medical Genomics Keith Durkin, Maria Artesi, Sébastien Bontems, Raphaël Boreux, Cécile Meex, Pierrette Melin, Marie-Pierre Hayette, Vincent Bours.
EPI_ISL_424637 hCoV-19/Belgium/ULG-10078/2020 Europe / Belgium / Liège 2020-04-03 Department of Clinical Microbiology GIGA Medical Genomics Keith Durkin, Maria Artesi, Sébastien Bontems, Raphaël Boreux, Cécile Meex, Pierrette Melin, Marie-Pierre Hayette, Vincent Bours.

EPI_ISL_424640 hCoV-19/Belgium/ULG-10081/2020 Europe / Belgium / Liège 2020-04-03 Department of Clinical Microbiology GIGA Medical Genomics Keith Durkin, Maria Artesi, Sébastien Bontems, Raphaël Boreux, Cécile Meex, Pierrette Melin, Marie-Pierre Hayette, Vincent Bours.
EPI_ISL_424641 hCoV-19/Belgium/ULG-10080/2020 Europe / Belgium / Liège 2020-04-03 Department of Clinical Microbiology GIGA Medical Genomics Keith Durkin, Maria Artesi, Sébastien Bontems, Raphaël Boreux, Cécile Meex, Pierrette Melin, Marie-Pierre Hayette, Vincent Bours.
EPI_ISL_424642 hCoV-19/Belgium/ULG-10077/2020 Europe / Belgium / Liège 2020-04-04 Department of Clinical Microbiology GIGA Medical Genomics Keith Durkin, Maria Artesi, Sébastien Bontems, Raphaël Boreux, Cécile Meex, Pierrette Melin, Marie-Pierre Hayette, Vincent Bours.
EPI_ISL_424643 hCoV-19/Belgium/ULG-10089/2020 Europe / Belgium / Liège 2020-04-04 Department of Clinical Microbiology GIGA Medical Genomics Keith Durkin, Maria Artesi, Sébastien Bontems, Raphaël Boreux, Cécile Meex, Pierrette Melin, Marie-Pierre Hayette, Vincent Bours.
EPI_ISL_424644 hCoV-19/Belgium/ULG-10086/2020 Europe / Belgium / Liège 2020-04-04 Department of Clinical Microbiology GIGA Medical Genomics Keith Durkin, Maria Artesi, Sébastien Bontems, Raphaël Boreux, Cécile Meex, Pierrette Melin, Marie-Pierre Hayette, Vincent Bours.
EPI_ISL_424645 hCoV-19/Belgium/ULG-10085/2020 Europe / Belgium / Liège 2020-04-04 Department of Clinical Microbiology GIGA Medical Genomics Keith Durkin, Maria Artesi, Sébastien Bontems, Raphaël Boreux, Cécile Meex, Pierrette Melin, Marie-Pierre Hayette, Vincent Bours.
EPI_ISL_424646 hCoV-19/Belgium/ULG-10088/2020 Europe / Belgium / Liège 2020-04-04 Department of Clinical Microbiology GIGA Medical Genomics Keith Durkin, Maria Artesi, Sébastien Bontems, Raphaël Boreux, Cécile Meex, Pierrette Melin, Marie-Pierre Hayette, Vincent Bours.
EPI_ISL_424647 hCoV-19/Belgium/ULG-10092/2020 Europe / Belgium / Liège 2020-04-04 Department of Clinical Microbiology GIGA Medical Genomics Keith Durkin, Maria Artesi, Sébastien Bontems, Raphaël Boreux, Cécile Meex, Pierrette Melin, Marie-Pierre Hayette, Vincent Bours.
EPI_ISL_424649 hCoV-19/Belgium/ULG-10082/2020 Europe / Belgium / Liège 2020-04-04 Department of Clinical Microbiology GIGA Medical Genomics Keith Durkin, Maria Artesi, Sébastien Bontems, Raphaël Boreux, Cécile Meex, Pierrette Melin, Marie-Pierre Hayette, Vincent Bours.
EPI_ISL_424650 hCoV-19/Belgium/ULG-10094/2020 Europe / Belgium / Liège 2020-04-04 Department of Clinical Microbiology GIGA Medical Genomics Keith Durkin, Maria Artesi, Sébastien Bontems, Raphaël Boreux, Cécile Meex, Pierrette Melin, Marie-Pierre Hayette, Vincent Bours.
EPI_ISL_424651 hCoV-19/Belgium/ULG-10095/2020 Europe / Belgium / Liège 2020-04-05 Department of Clinical Microbiology GIGA Medical Genomics Keith Durkin, Maria Artesi, Sébastien Bontems, Raphaël Boreux, Cécile Meex, Pierrette Melin, Marie-Pierre Hayette, Vincent Bours.
EPI_ISL_424652 hCoV-19/Belgium/ULG-10055/2020 Europe / Belgium / Liège 2020-04-05 Department of Clinical Microbiology GIGA Medical Genomics Keith Durkin, Maria Artesi, Sébastien Bontems, Raphaël Boreux, Cécile Meex, Pierrette Melin, Marie-Pierre Hayette, Vincent Bours.
EPI_ISL_424653 hCoV-19/Belgium/ULG-10058/2020 Europe / Belgium / Liège 2020-04-05 Department of Clinical Microbiology GIGA Medical Genomics Keith Durkin, Maria Artesi, Sébastien Bontems, Raphaël Boreux, Cécile Meex, Pierrette Melin, Marie-Pierre Hayette, Vincent Bours.
EPI_ISL_424654 hCoV-19/Belgium/ULG-10057/2020 Europe / Belgium / Liège 2020-04-05 Department of Clinical Microbiology GIGA Medical Genomics Keith Durkin, Maria Artesi, Sébastien Bontems, Raphaël Boreux, Cécile Meex, Pierrette Melin, Marie-Pierre Hayette, Vincent Bours.
EPI_ISL_424655 hCoV-19/Belgium/ULG-10056/2020 Europe / Belgium / Liège 2020-04-05 Department of Clinical Microbiology GIGA Medical Genomics Keith Durkin, Maria Artesi, Sébastien Bontems, Raphaël Boreux, Cécile Meex, Pierrette Melin, Marie-Pierre Hayette, Vincent Bours.
EPI_ISL_424657 hCoV-19/Belgium/ULG-10052/2020 Europe / Belgium / Liège 2020-04-05 Department of Clinical Microbiology GIGA Medical Genomics Keith Durkin, Maria Artesi, Sébastien Bontems, Raphaël Boreux, Cécile Meex, Pierrette Melin, Marie-Pierre Hayette, Vincent Bours.
EPI_ISL_424658 hCoV-19/Belgium/ULG-10049/2020 Europe / Belgium / Liège 2020-04-05 Department of Clinical Microbiology GIGA Medical Genomics Keith Durkin, Maria Artesi, Sébastien Bontems, Raphaël Boreux, Cécile Meex, Pierrette Melin, Marie-Pierre Hayette, Vincent Bours.
EPI_ISL_424660 hCoV-19/Belgium/ULG-10051/2020 Europe / Belgium / Liège 2020-04-05 Department of Clinical Microbiology GIGA Medical Genomics Keith Durkin, Maria Artesi, Sébastien Bontems, Raphaël Boreux, Cécile Meex, Pierrette Melin, Marie-Pierre Hayette, Vincent Bours.
EPI_ISL_424661 hCoV-19/Belgium/ULG-10053/2020 Europe / Belgium / Liège 2020-04-05 Department of Clinical Microbiology GIGA Medical Genomics Keith Durkin, Maria Artesi, Sébastien Bontems, Raphaël Boreux, Cécile Meex, Pierrette Melin, Marie-Pierre Hayette, Vincent Bours.
EPI_ISL_424662 hCoV-19/Belgium/ULG-10061/2020 Europe / Belgium / Liège 2020-04-05 Department of Clinical Microbiology GIGA Medical Genomics Keith Durkin, Maria Artesi, Sébastien Bontems, Raphaël Boreux, Cécile Meex, Pierrette Melin, Marie-Pierre Hayette, Vincent Bours.
EPI_ISL_424664 hCoV-19/Belgium/ULG-10062/2020 Europe / Belgium / Liège 2020-04-05 Department of Clinical Microbiology GIGA Medical Genomics Keith Durkin, Maria Artesi, Sébastien Bontems, Raphaël Boreux, Cécile Meex, Pierrette Melin, Marie-Pierre Hayette, Vincent Bours.
EPI_ISL_424667 hCoV-19/Mexico/EdoMex-InDRE_03/2020 North America / Mexico / Estado de Mexico 2020-03-04 Laboratorio Estatal de Salud Publica del Estado de México Instituto de Diagnóstico y Referencia Epidemiológicos Irma López Martínez, José Ernesto Ramírez González, Lucia Hernández Rivas, Gisela Barrera Badillo, Edgar Mendieta Condado, Fabiola Garcés Ayala, Adnan Araiza Rodríguez, Celia Boukadida, Santiago Avila Ríos, Mario Mújica Sánchez, José Arturo Martínez Orozco,Eduardo Becerril Vargas, Joel Armando Vázquez Pérez, Victor Hugo Borja Aburto, Concepción Grajales Muñiz, Cesar Raúl González Bonilla, Carolina González Torres, Francisco Javier Gaytán Cervantes, José Esteban Muñoz Medina, Guillermo M. Ruiz-Palacios, Pilar Ramos

Cervantes, Violeta Ibarra Gonzalez, Fernando Ledesma Barrientos, Luis Alberto García Andrade, Alfredo Ponce de León Garduño, Blanca Taboada, Alejandro Sánchez, Pavel Isa, Ricardo Grande, Gloria Vázquez, Francisco Pulido, Carlos F. Arias.
EPI_ISL_424668 hCoV-19/USA/AZ-ASU2922/2020 North America / USA / Arizona 2020-03-16 Arizona State University Health Services Arizona State University Rabia Maqsood, LaRinda A. Holland, Emily A. Kaelin, Bereket Estifanos, Nicholas J. Mellor, Jason Steel, Lily I. Wu, Arvind Varsani, Rolf U. Halden, Brenda G. Hogue, Matthew Scotch, Efrem S. Lim
EPI_ISL_424669 hCoV-19/USA/AZ-ASU2923/2020 North America / USA / Arizona 2020-03-16 Arizona State University Health Services Arizona State University Rabia Maqsood, LaRinda A. Holland, Emily A. Kaelin, Bereket Estifanos, Nicholas J. Mellor, Jason Steel, Lily I. Wu, Arvind Varsani, Rolf U. Halden, Brenda G. Hogue, Matthew Scotch, Efrem S. Lim
EPI_ISL_424670 hCoV-19/Mexico/Queretaro-InDRE_04/2020 North America / Mexico / Queretaro 2020-03-10 Laboratorio Estatal de Salud Publica del Estado de Queretaro Instituto de Diagnóstico y Referencia Epidemiologicos Gisela Barrera Badillo, Irma López Martínez, Lucia Hernández Rivas, Edgar Mendieta Condado, Fabiola Garcés Ayala, Adnan Araiza Rodríguez, Celia Boukadida, Santiago Avila Ríos, Mario Mújica Sánchez, José Arturo Martínez Orozco,Eduardo Becerril Vargas, Joel Armando Vázquez Pérez, Victor Hugo Borja Aburto, Concepción Grajales Muñiz, Cesar Raúl González Bonilla, Carolina González Torres, Francisco Javier Gaytán Cervantes, José Esteban Muñoz Medina, Guillermo M. Ruiz-Palacios, Pilar Ramos Cervantes, Violeta Ibarra Gonzalez, Fernando Ledesma Barrientos, Luis Alberto García Andrade, Alfredo Ponce de León Garduño, Blanca Taboada, Alejandro Sánchez, Pavel Isa, Ricardo Grande, Gloria Vázquez, Francisco Pulido, Carlos F. Arias,José Ernesto Ramírez González
EPI_ISL_424671 hCoV-19/USA/AZ-ASU2936/2020 North America / USA / Arizona 2020-03-17 Arizona State University Health Services Arizona State University Rabia Maqsood, LaRinda A. Holland, Emily A. Kaelin, Bereket Estifanos, Nicholas J. Mellor, Jason Steel, Lily I. Wu, Arvind Varsani, Rolf U. Halden, Brenda G. Hogue, Matthew Scotch, Efrem S. Lim
EPI_ISL_424672 hCoV-19/Mexico/Puebla-InDRE_05/2020 North America / Mexico / Puebla 2020-03-11 Laboratorio Estatal de Salud Publica del Estado de Puebla Instituto de Diagnostico y Referencia Epidemiologicos Fabiola Garcés Ayala, Gisela Barrera Badillo, Irma López Martínez, Lucia Hernández Rivas, Edgar Mendieta Condado, Adnan Araiza Rodríguez, Celia Boukadida, Santiago Avila Ríos, Mario Mújica Sánchez, José Arturo Martínez Orozco,Eduardo Becerril Vargas, Joel Armando Vázquez Pérez, Victor Hugo Borja Aburto, Concepción Grajales Muñiz, Cesar Raúl González Bonilla, Carolina González Torres, Francisco Javier Gaytán Cervantes, José Esteban Muñoz Medina, Guillermo M. Ruiz-Palacios, Pilar Ramos Cervantes, Violeta Ibarra Gonzalez, Fernando Ledesma Barrientos, Luis Alberto García Andrade, Alfredo Ponce de León Garduño, Blanca Taboada, Alejandro Sánchez, Pavel Isa, Ricardo Grande, Gloria Vázquez, Francisco Pulido, Carlos F. Arias,José Ernesto Ramírez González
EPI_ISL_424673 hCoV-19/Mexico/CDMX-InDRE_06/2020 North America / Mexico / CDMX 2020-03-12 Instituto de Diagnostico y Referencia Epidemiologicos Instituto de Diagnostico y Referencia Epidemiologicos Adnan Araiza Rodríguez, Edgar Mendieta Condado,Fabiola Garcés Ayala, Gisela Barrera Badillo, Irma López Martínez, Lucia Hernández Rivas, Celia Boukadida, Santiago Avila Ríos, Mario Mújica Sánchez, José Arturo Martínez Orozco,Eduardo Becerril Vargas, Joel Armando Vázquez Pérez, Victor Hugo Borja Aburto, Concepción Grajales Muñiz, Cesar Raúl González Bonilla, Carolina González Torres, Francisco Javier Gaytán Cervantes, José Esteban Muñoz Medina, Guillermo M. Ruiz-Palacios, Pilar Ramos Cervantes, Violeta Ibarra Gonzalez, Fernando Ledesma Barrientos, Luis Alberto García Andrade, Alfredo Ponce de León Garduño, Blanca Taboada, Alejandro Sánchez, Pavel Isa, Ricardo Grande, Gloria Vázquez, Francisco Pulido, Carlos F. Arias,José Ernesto Ramírez González
EPI_ISL_424841 hCoV-19/USA/SC_3569/2020 North America / USA / South Carolina 2020-03-06 SC Dept of Health and Env. Control-Bureau of Laboratories Pathogen Discovery, Respiratory Viruses Branch, Division of Viral Diseases, Centers for Disease Control and Prevention Yan Li, Krista Queen, Clinton R. Paden, Rachel Marine, Anna Uehara, Ying Tao, Jing Zhang, Haibin Wang, Mary S. Keckler, Alison S. Laufer Halpin, Christopher A. Elkins, Suxiang Tong
EPI_ISL_424842 hCoV-19/USA/SC_3575/2020 North America / USA / South Carolina 2020-03-07 SC Dept of Health and Env. Control-Bureau of Laboratories Pathogen Discovery, Respiratory Viruses Branch, Division of Viral Diseases, Centers for Disease Control and Prevention Yan Li, Krista Queen, Clinton R. Paden, Rachel Marine, Anna Uehara, Ying Tao, Jing Zhang, Haibin Wang, Mary S. Keckler, Alison S. Laufer Halpin, Christopher A. Elkins, Suxiang Tong
EPI_ISL_424843 hCoV-19/USA/MA_0020/2020 North America / USA / Massachusetts 2020-03-05 MA State Public Health Laboratory Pathogen Discovery, Respiratory Viruses Branch, Division of Viral Diseases, Centers for Disease Control and Prevention Yan Li, Krista Queen, Clinton R. Paden, Rachel Marine, Anna Uehara, Ying Tao, Jing Zhang, Haibin Wang, Mary S. Keckler, Alison S. Laufer Halpin, Christopher A. Elkins, Suxiang Tong
EPI_ISL_424844 hCoV-19/USA/MA_3672/2020 North America / USA / Massachusetts 2020-03-06 MA State Public Health Laboratory Pathogen Discovery, Respiratory Viruses Branch, Division of Viral Diseases, Centers for Disease Control and Prevention Yan Li, Krista Queen, Clinton R. Paden, Rachel Marine, Anna Uehara, Ying Tao, Jing Zhang, Haibin Wang, Mary S. Keckler, Alison S. Laufer Halpin, Christopher A. Elkins, Suxiang Tong
EPI_ISL_424845 hCoV-19/USA/MA_3699/2020 North America / USA / Massachusetts 2020-03-06 MA State Public Health Laboratory Pathogen Discovery, Respiratory Viruses Branch, Division of Viral Diseases, Centers for Disease Control and Prevention Yan Li, Krista Queen, Clinton R. Paden, Rachel Marine, Anna Uehara, Ying Tao, Jing Zhang, Haibin Wang, Mary S. Keckler, Alison S. Laufer Halpin, Christopher A. Elkins, Suxiang Tong

EPI_ISL_424846 hCoV-19/USA/MA_3878/2020 North America / USA / Massachusetts 2020-03-06 MA State Public Health Laboratory Pathogen Discovery, Respiratory Viruses Branch, Division of Viral Diseases, Centers for Disease Control and Prevention Yan Li, Krista Queen, Clinton R. Paden, Rachel Marine, Anna Uehara, Ying Tao, Jing Zhang, Haibin Wang, Mary S. Keckler, Alison S. Laufer Halpin, Christopher A. Elkins, Suxiang Tong
EPI_ISL_424847 hCoV-19/USA/MA_4811/2020 North America / USA / Massachusetts 2020-03-07 MA State Public Health Laboratory Pathogen Discovery, Respiratory Viruses Branch, Division of Viral Diseases, Centers for Disease Control and Prevention Yan Li, Krista Queen, Clinton R. Paden, Rachel Marine, Anna Uehara, Ying Tao, Jing Zhang, Haibin Wang, Mary S. Keckler, Alison S. Laufer Halpin, Christopher A. Elkins, Suxiang Tong
EPI_ISL_424848 hCoV-19/USA/AZ_8132/2020 North America / USA / Arizona 2020-03-09 AZ SPHL, Arizona Department of Health Services Pathogen Discovery, Respiratory Viruses Branch, Division of Viral Diseases, Centers for Disease Control and Prevention Yan Li, Krista Queen, Clinton R. Paden, Rachel Marine, Anna Uehara, Ying Tao, Jing Zhang, Haibin Wang, Mary S. Keckler, Alison S. Laufer Halpin, Christopher A. Elkins, Suxiang Tong
EPI_ISL_424849 hCoV-19/USA/AZ_8135/2020 North America / USA / Arizona 2020-03-09 AZ SPHL, Arizona Department of Health Services Pathogen Discovery, Respiratory Viruses Branch, Division of Viral Diseases, Centers for Disease Control and Prevention Yan Li, Krista Queen, Clinton R. Paden, Rachel Marine, Anna Uehara, Ying Tao, Jing Zhang, Haibin Wang, Mary S. Keckler, Alison S. Laufer Halpin, Christopher A. Elkins, Suxiang Tong
EPI_ISL_424850 hCoV-19/USA/IL_0087/2020 North America / USA / Illinois 2020-03-07 IL Department of Public Health Chicago Laboratory Pathogen Discovery, Respiratory Viruses Branch, Division of Viral Diseases, Centers for Disease Control and Prevention Yan Li, Krista Queen, Clinton R. Paden, Rachel Marine, Anna Uehara, Ying Tao, Jing Zhang, Haibin Wang, Mary S. Keckler, Alison S. Laufer Halpin, Christopher A. Elkins, Suxiang Tong
EPI_ISL_424851 hCoV-19/USA/IL_0089/2020 North America / USA / Illinois 2020-03-07 IL Department of Public Health Chicago Laboratory Pathogen Discovery, Respiratory Viruses Branch, Division of Viral Diseases, Centers for Disease Control and Prevention Yan Li, Krista Queen, Clinton R. Paden, Rachel Marine, Anna Uehara, Ying Tao, Jing Zhang, Haibin Wang, Mary S. Keckler, Alison S. Laufer Halpin, Christopher A. Elkins, Suxiang Tong
EPI_ISL_424852 hCoV-19/USA/DC_0004/2020 North America / USA / District of Columbia 2020-03-06 DC Public Health Lab/ Dept. of Forensic Sciences Pathogen Discovery, Respiratory Viruses Branch, Division of Viral Diseases, Centers for Disease Control and Prevention Yan Li, Krista Queen, Clinton R. Paden, Rachel Marine, Anna Uehara, Ying Tao, Jing Zhang, Haibin Wang, Mary S. Keckler, Alison S. Laufer Halpin, Christopher A. Elkins, Suxiang Tong
EPI_ISL_424853 hCoV-19/USA/FL_9590/2020 North America / USA / Florida 2020-03-05 FL Bureau of Public Health Laboratories-Miami Pathogen Discovery, Respiratory Viruses Branch, Division of Viral Diseases, Centers for Disease Control and Prevention Yan Li, Krista Queen, Clinton R. Paden, Rachel Marine, Anna Uehara, Ying Tao, Jing Zhang, Haibin Wang, Mary S. Keckler, Alison S. Laufer Halpin, Christopher A. Elkins, Suxiang Tong
EPI_ISL_424854 hCoV-19/USA/FL_9655/2020 North America / USA / Florida 2020-03-05 FL Bureau of Public Health Laboratories-Miami Pathogen Discovery, Respiratory Viruses Branch, Division of Viral Diseases, Centers for Disease Control and Prevention Yan Li, Krista Queen, Clinton R. Paden, Rachel Marine, Anna Uehara, Ying Tao, Jing Zhang, Haibin Wang, Mary S. Keckler, Alison S. Laufer Halpin, Christopher A. Elkins, Suxiang Tong
EPI_ISL_425944 hCoV-19/Scotland/EDB214/2020 Europe / United Kingdom / Scotland 2020-03-26 Virology Department, Royal Infirmary of Edinburgh, NHS Lothian / School of Biological Sciences, University of Edinburgh / Institute of Genetics and Molecular Medicine, University of Edinburgh COVID-19 Genomics UK (COG-UK) Consortium McHugh M, Dewar R, Rooke S, Gallagher M, Balcaza C, O'Toole A, Hill V, McCrone JT, Colquhoun R, Yu X, Jackson B, Scher E, Rambaut A, Williams TC, Templeton K
EPI_ISL_425945 hCoV-19/Scotland/EDB216/2020 Europe / United Kingdom / Scotland 2020-03-26 Virology Department, Royal Infirmary of Edinburgh, NHS Lothian / School of Biological Sciences, University of Edinburgh / Institute of Genetics and Molecular Medicine, University of Edinburgh COVID-19 Genomics UK (COG-UK) Consortium McHugh M, Dewar R, Rooke S, Gallagher M, Balcaza C, O'Toole A, Hill V, McCrone JT, Colquhoun R, Yu X, Jackson B, Scher E, Rambaut A, Williams TC, Templeton K
EPI_ISL_425947 hCoV-19/Scotland/EDB219/2020 Europe / United Kingdom / Scotland 2020-03-26 Virology Department, Royal Infirmary of Edinburgh, NHS Lothian / School of Biological Sciences, University of Edinburgh / Institute of Genetics and Molecular Medicine, University of Edinburgh COVID-19 Genomics UK (COG-UK) Consortium McHugh M, Dewar R, Rooke S, Gallagher M, Balcaza C, O'Toole A, Hill V, McCrone JT, Colquhoun R, Yu X, Jackson B, Scher E, Rambaut A, Williams TC, Templeton K
EPI_ISL_425948 hCoV-19/Scotland/EDB222/2020 Europe / United Kingdom / Scotland 2020-03-26 Virology Department, Royal Infirmary of Edinburgh, NHS Lothian / School of Biological Sciences, University of Edinburgh / Institute of Genetics and Molecular Medicine, University of Edinburgh COVID-19 Genomics UK (COG-UK) Consortium McHugh M, Dewar R, Rooke S, Gallagher M, Balcaza C, O'Toole A, Hill V, McCrone JT, Colquhoun R, Yu X, Jackson B, Scher E, Rambaut A, Williams TC, Templeton K
EPI_ISL_425949 hCoV-19/Scotland/EDB226/2020 Europe / United Kingdom / Scotland 2020-03-26 Virology Department, Royal Infirmary of Edinburgh, NHS Lothian / School of Biological Sciences, University of Edinburgh / Institute of Genetics and Molecular Medicine, University of Edinburgh COVID-19 Genomics UK (COG-UK) Consortium McHugh M, Dewar R, Rooke S, Gallagher M, Balcaza C, O'Toole A, Hill V, McCrone JT, Colquhoun R, Yu X, Jackson B, Scher E, Rambaut A, Williams TC, Templeton K

EPI_ISL_425950 hCoV-19/Scotland/EDB227/2020 Europe / United Kingdom / Scotland 2020-03-26 Virology Department, Royal Infirmary of Edinburgh, NHS Lothian / School of Biological Sciences, University of Edinburgh / Institute of Genetics and Molecular Medicine, University of Edinburgh COVID-19 Genomics UK (COG-UK) Consortium McHugh M, Dewar R, Rooke S, Gallagher M, Balcaza C, O'Toole A, Hill V, McCrone JT, Colquhoun R, Yu X, Jackson B, Scher E, Rambaut A, Williams TC, Templeton K
EPI_ISL_425951 hCoV-19/Scotland/EDB232/2020 Europe / United Kingdom / Scotland 2020-03-27 Virology Department, Royal Infirmary of Edinburgh, NHS Lothian / School of Biological Sciences, University of Edinburgh / Institute of Genetics and Molecular Medicine, University of Edinburgh COVID-19 Genomics UK (COG-UK) Consortium McHugh M, Dewar R, Rooke S, Gallagher M, Balcaza C, O'Toole A, Hill V, McCrone JT, Colquhoun R, Yu X, Jackson B, Scher E, Rambaut A, Williams TC, Templeton K
EPI_ISL_425952 hCoV-19/Scotland/EDB234/2020 Europe / United Kingdom / Scotland 2020-03-27 Virology Department, Royal Infirmary of Edinburgh, NHS Lothian / School of Biological Sciences, University of Edinburgh / Institute of Genetics and Molecular Medicine, University of Edinburgh COVID-19 Genomics UK (COG-UK) Consortium McHugh M, Dewar R, Rooke S, Gallagher M, Balcaza C, O'Toole A, Hill V, McCrone JT, Colquhoun R, Yu X, Jackson B, Scher E, Rambaut A, Williams TC, Templeton K
EPI_ISL_425953 hCoV-19/Scotland/EDB245/2020 Europe / United Kingdom / Scotland 2020-03-26 Virology Department, Royal Infirmary of Edinburgh, NHS Lothian / School of Biological Sciences, University of Edinburgh / Institute of Genetics and Molecular Medicine, University of Edinburgh COVID-19 Genomics UK (COG-UK) Consortium McHugh M, Dewar R, Rooke S, Gallagher M, Balcaza C, O'Toole A, Hill V, McCrone JT, Colquhoun R, Yu X, Jackson B, Scher E, Rambaut A, Williams TC, Templeton K
EPI_ISL_425954 hCoV-19/Scotland/EDB248/2020 Europe / United Kingdom / Scotland 2020-03-27 Virology Department, Royal Infirmary of Edinburgh, NHS Lothian / School of Biological Sciences, University of Edinburgh / Institute of Genetics and Molecular Medicine, University of Edinburgh COVID-19 Genomics UK (COG-UK) Consortium McHugh M, Dewar R, Rooke S, Gallagher M, Balcaza C, O'Toole A, Hill V, McCrone JT, Colquhoun R, Yu X, Jackson B, Scher E, Rambaut A, Williams TC, Templeton K
EPI_ISL_425956 hCoV-19/Scotland/EDB254/2020 Europe / United Kingdom / Scotland 2020-03-26 Virology Department, Royal Infirmary of Edinburgh, NHS Lothian / School of Biological Sciences, University of Edinburgh / Institute of Genetics and Molecular Medicine, University of Edinburgh COVID-19 Genomics UK (COG-UK) Consortium McHugh M, Dewar R, Rooke S, Gallagher M, Balcaza C, O'Toole A, Hill V, McCrone JT, Colquhoun R, Yu X, Jackson B, Scher E, Rambaut A, Williams TC, Templeton K
EPI_ISL_425958 hCoV-19/Scotland/EDB270/2020 Europe / United Kingdom / Scotland 2020-03-27 Virology Department, Royal Infirmary of Edinburgh, NHS Lothian / School of Biological Sciences, University of Edinburgh / Institute of Genetics and Molecular Medicine, University of Edinburgh COVID-19 Genomics UK (COG-UK) Consortium McHugh M, Dewar R, Rooke S, Gallagher M, Balcaza C, O'Toole A, Hill V, McCrone JT, Colquhoun R, Yu X, Jackson B, Scher E, Rambaut A, Williams TC, Templeton K
EPI_ISL_425959 hCoV-19/Scotland/EDB273/2020 Europe / United Kingdom / Scotland 2020-03-26 Virology Department, Royal Infirmary of Edinburgh, NHS Lothian / School of Biological Sciences, University of Edinburgh / Institute of Genetics and Molecular Medicine, University of Edinburgh COVID-19 Genomics UK (COG-UK) Consortium McHugh M, Dewar R, Rooke S, Gallagher M, Balcaza C, O'Toole A, Hill V, McCrone JT, Colquhoun R, Yu X, Jackson B, Scher E, Rambaut A, Williams TC, Templeton K
EPI_ISL_425961 hCoV-19/Scotland/EDB275/2020 Europe / United Kingdom / Scotland 2020-03-26 Virology Department, Royal Infirmary of Edinburgh, NHS Lothian / School of Biological Sciences, University of Edinburgh / Institute of Genetics and Molecular Medicine, University of Edinburgh COVID-19 Genomics UK (COG-UK) Consortium McHugh M, Dewar R, Rooke S, Gallagher M, Balcaza C, O'Toole A, Hill V, McCrone JT, Colquhoun R, Yu X, Jackson B, Scher E, Rambaut A, Williams TC, Templeton K
EPI_ISL_425962 hCoV-19/Scotland/EDB278/2020 Europe / United Kingdom / Scotland 2020-03-27 Virology Department, Royal Infirmary of Edinburgh, NHS Lothian / School of Biological Sciences, University of Edinburgh / Institute of Genetics and Molecular Medicine, University of Edinburgh COVID-19 Genomics UK (COG-UK) Consortium McHugh M, Dewar R, Rooke S, Gallagher M, Balcaza C, O'Toole A, Hill V, McCrone JT, Colquhoun R, Yu X, Jackson B, Scher E, Rambaut A, Williams TC, Templeton K
EPI_ISL_425963 hCoV-19/Scotland/EDB283/2020 Europe / United Kingdom / Scotland 2020-03-27 Virology Department, Royal Infirmary of Edinburgh, NHS Lothian / School of Biological Sciences, University of Edinburgh / Institute of Genetics and Molecular Medicine, University of Edinburgh COVID-19 Genomics UK (COG-UK) Consortium McHugh M, Dewar R, Rooke S, Gallagher M, Balcaza C, O'Toole A, Hill V, McCrone JT, Colquhoun R, Yu X, Jackson B, Scher E, Rambaut A, Williams TC, Templeton K
EPI_ISL_425964 hCoV-19/Scotland/EDB287/2020 Europe / United Kingdom / Scotland 2020-03-27 Virology Department, Royal Infirmary of Edinburgh, NHS Lothian / School of Biological Sciences, University of Edinburgh / Institute of Genetics and Molecular Medicine, University of Edinburgh COVID-19 Genomics UK (COG-UK) Consortium McHugh M, Dewar R, Rooke S, Gallagher M, Balcaza C, O'Toole A, Hill V, McCrone JT, Colquhoun R, Yu X, Jackson B, Scher E, Rambaut A, Williams TC, Templeton K
EPI_ISL_425965 hCoV-19/Scotland/EDB288/2020 Europe / United Kingdom / Scotland 2020-03-27 Virology Department, Royal Infirmary of Edinburgh, NHS Lothian / School of Biological Sciences, University of Edinburgh / Institute of Genetics and Molecular Medicine, University of Edinburgh COVID-19 Genomics UK (COG-UK) Consortium McHugh M, Dewar R, Rooke S, Gallagher M, Balcaza C, O'Toole A, Hill V, McCrone JT, Colquhoun R, Yu X, Jackson B, Scher E, Rambaut A, Williams TC, Templeton K
EPI_ISL_425966 hCoV-19/Scotland/EDB297/2020 Europe / United Kingdom / Scotland 2020-03-28 Virology Department, Royal Infirmary of Edinburgh, NHS Lothian / School of Biological Sciences, University of Edinburgh / Institute of Genetics and Molecular Medicine, University of Edinburgh COVID-19 Genomics UK (COG-UK) Consortium McHugh M, Dewar R, Rooke S, Gallagher M, Balcaza C, O'Toole A, Hill V, McCrone JT, Colquhoun R, Yu X, Jackson B, Scher E, Rambaut A, Williams TC, Templeton K

EPI_ISL_425967 hCoV-19/Scotland/EDB300/2020 Europe / United Kingdom / Scotland 2020-03-29 Virology Department, Royal Infirmary of Edinburgh, NHS Lothian / School of Biological Sciences, University of Edinburgh / Institute of Genetics and Molecular Medicine, University of Edinburgh COVID-19 Genomics UK (COG-UK) Consortium McHugh M, Dewar R, Rooke S, Gallagher M, Balcaza C, O'Toole A, Hill V, McCrone JT, Colquhoun R, Yu X, Jackson B, Scher E, Rambaut A, Williams TC, Templeton K
EPI_ISL_425968 hCoV-19/Scotland/EDB314/2020 Europe / United Kingdom / Scotland 2020-03-28 Virology Department, Royal Infirmary of Edinburgh, NHS Lothian / School of Biological Sciences, University of Edinburgh / Institute of Genetics and Molecular Medicine, University of Edinburgh COVID-19 Genomics UK (COG-UK) Consortium McHugh M, Dewar R, Rooke S, Gallagher M, Balcaza C, O'Toole A, Hill V, McCrone JT, Colquhoun R, Yu X, Jackson B, Scher E, Rambaut A, Williams TC, Templeton K
EPI_ISL_425969 hCoV-19/Scotland/EDB318/2020 Europe / United Kingdom / Scotland 2020-03-28 Virology Department, Royal Infirmary of Edinburgh, NHS Lothian / School of Biological Sciences, University of Edinburgh / Institute of Genetics and Molecular Medicine, University of Edinburgh COVID-19 Genomics UK (COG-UK) Consortium McHugh M, Dewar R, Rooke S, Gallagher M, Balcaza C, O'Toole A, Hill V, McCrone JT, Colquhoun R, Yu X, Jackson B, Scher E, Rambaut A, Williams TC, Templeton K
EPI_ISL_425971 hCoV-19/Scotland/EDB325/2020 Europe / United Kingdom / Scotland 2020-03-28 Virology Department, Royal Infirmary of Edinburgh, NHS Lothian / School of Biological Sciences, University of Edinburgh / Institute of Genetics and Molecular Medicine, University of Edinburgh COVID-19 Genomics UK (COG-UK) Consortium McHugh M, Dewar R, Rooke S, Gallagher M, Balcaza C, O'Toole A, Hill V, McCrone JT, Colquhoun R, Yu X, Jackson B, Scher E, Rambaut A, Williams TC, Templeton K
EPI_ISL_425972 hCoV-19/Scotland/EDB326/2020 Europe / United Kingdom / Scotland 2020-03-28 Virology Department, Royal Infirmary of Edinburgh, NHS Lothian / School of Biological Sciences, University of Edinburgh / Institute of Genetics and Molecular Medicine, University of Edinburgh COVID-19 Genomics UK (COG-UK) Consortium McHugh M, Dewar R, Rooke S, Gallagher M, Balcaza C, O'Toole A, Hill V, McCrone JT, Colquhoun R, Yu X, Jackson B, Scher E, Rambaut A, Williams TC, Templeton K
EPI_ISL_425973 hCoV-19/Scotland/EDB327/2020 Europe / United Kingdom / Scotland 2020-03-28 Virology Department, Royal Infirmary of Edinburgh, NHS Lothian / School of Biological Sciences, University of Edinburgh / Institute of Genetics and Molecular Medicine, University of Edinburgh COVID-19 Genomics UK (COG-UK) Consortium McHugh M, Dewar R, Rooke S, Gallagher M, Balcaza C, O'Toole A, Hill V, McCrone JT, Colquhoun R, Yu X, Jackson B, Scher E, Rambaut A, Williams TC, Templeton K
EPI_ISL_425976 hCoV-19/Scotland/EDB342/2020 Europe / United Kingdom / Scotland 2020-03-30 Virology Department, Royal Infirmary of Edinburgh, NHS Lothian / School of Biological Sciences, University of Edinburgh / Institute of Genetics and Molecular Medicine, University of Edinburgh COVID-19 Genomics UK (COG-UK) Consortium McHugh M, Dewar R, Rooke S, Gallagher M, Balcaza C, O'Toole A, Hill V, McCrone JT, Colquhoun R, Yu X, Jackson B, Scher E, Rambaut A, Williams TC, Templeton K
EPI_ISL_425978 hCoV-19/Scotland/EDB354/2020 Europe / United Kingdom / Scotland 2020-03-30 Virology Department, Royal Infirmary of Edinburgh, NHS Lothian / School of Biological Sciences, University of Edinburgh / Institute of Genetics and Molecular Medicine, University of Edinburgh COVID-19 Genomics UK (COG-UK) Consortium McHugh M, Dewar R, Rooke S, Gallagher M, Balcaza C, O'Toole A, Hill V, McCrone JT, Colquhoun R, Yu X, Jackson B, Scher E, Rambaut A, Williams TC, Templeton K
EPI_ISL_425979 hCoV-19/Scotland/EDB355/2020 Europe / United Kingdom / Scotland 2020-03-29 Virology Department, Royal Infirmary of Edinburgh, NHS Lothian / School of Biological Sciences, University of Edinburgh / Institute of Genetics and Molecular Medicine, University of Edinburgh COVID-19 Genomics UK (COG-UK) Consortium McHugh M, Dewar R, Rooke S, Gallagher M, Balcaza C, O'Toole A, Hill V, McCrone JT, Colquhoun R, Yu X, Jackson B, Scher E, Rambaut A, Williams TC, Templeton K
EPI_ISL_425980 hCoV-19/Scotland/EDB356/2020 Europe / United Kingdom / Scotland 2020-03-30 Virology Department, Royal Infirmary of Edinburgh, NHS Lothian / School of Biological Sciences, University of Edinburgh / Institute of Genetics and Molecular Medicine, University of Edinburgh COVID-19 Genomics UK (COG-UK) Consortium McHugh M, Dewar R, Rooke S, Gallagher M, Balcaza C, O'Toole A, Hill V, McCrone JT, Colquhoun R, Yu X, Jackson B, Scher E, Rambaut A, Williams TC, Templeton K
EPI_ISL_425982 hCoV-19/Scotland/EDB363/2020 Europe / United Kingdom / Scotland 2020-03-29 Virology Department, Royal Infirmary of Edinburgh, NHS Lothian / School of Biological Sciences, University of Edinburgh / Institute of Genetics and Molecular Medicine, University of Edinburgh COVID-19 Genomics UK (COG-UK) Consortium McHugh M, Dewar R, Rooke S, Gallagher M, Balcaza C, O'Toole A, Hill V, McCrone JT, Colquhoun R, Yu X, Jackson B, Scher E, Rambaut A, Williams TC, Templeton K
EPI_ISL_425985 hCoV-19/Scotland/EDB373/2020 Europe / United Kingdom / Scotland 2020-03-30 Virology Department, Royal Infirmary of Edinburgh, NHS Lothian / School of Biological Sciences, University of Edinburgh / Institute of Genetics and Molecular Medicine, University of Edinburgh COVID-19 Genomics UK (COG-UK) Consortium McHugh M, Dewar R, Rooke S, Gallagher M, Balcaza C, O'Toole A, Hill V, McCrone JT, Colquhoun R, Yu X, Jackson B, Scher E, Rambaut A, Williams TC, Templeton K
EPI_ISL_425986 hCoV-19/Scotland/EDB374/2020 Europe / United Kingdom / Scotland 2020-03-30 Virology Department, Royal Infirmary of Edinburgh, NHS Lothian / School of Biological Sciences, University of Edinburgh / Institute of Genetics and Molecular Medicine, University of Edinburgh COVID-19 Genomics UK (COG-UK) Consortium McHugh M, Dewar R, Rooke S, Gallagher M, Balcaza C, O'Toole A, Hill V, McCrone JT, Colquhoun R, Yu X, Jackson B, Scher E, Rambaut A, Williams TC, Templeton K
EPI_ISL_425987 hCoV-19/Scotland/EDB378/2020 Europe / United Kingdom / Scotland 2020-03-30 Virology Department, Royal Infirmary of Edinburgh, NHS Lothian / School of Biological Sciences, University of Edinburgh / Institute of Genetics and Molecular Medicine, University of Edinburgh COVID-19 Genomics UK (COG-UK) Consortium McHugh M, Dewar R, Rooke S, Gallagher M, Balcaza C, O'Toole A, Hill V, McCrone JT, Colquhoun R, Yu X, Jackson B, Scher E, Rambaut A, Williams TC, Templeton K

EPI_ISL_425988 hCoV-19/Scotland/EDB379/2020 Europe / United Kingdom / Scotland 2020-03-30 Virology Department, Royal Infirmary of Edinburgh, NHS Lothian / School of Biological Sciences, University of Edinburgh / Institute of Genetics and Molecular Medicine, University of Edinburgh COVID-19 Genomics UK (COG-UK) Consortium McHugh M, Dewar R, Rooke S, Gallagher M, Balcaza C, O'Toole A, Hill V, McCrone JT, Colquhoun R, Yu X, Jackson B, Scher E, Rambaut A, Williams TC, Templeton K
EPI_ISL_425990 hCoV-19/Scotland/EDB383/2020 Europe / United Kingdom / Scotland 2020-03-30 Virology Department, Royal Infirmary of Edinburgh, NHS Lothian / School of Biological Sciences, University of Edinburgh / Institute of Genetics and Molecular Medicine, University of Edinburgh COVID-19 Genomics UK (COG-UK) Consortium McHugh M, Dewar R, Rooke S, Gallagher M, Balcaza C, O'Toole A, Hill V, McCrone JT, Colquhoun R, Yu X, Jackson B, Scher E, Rambaut A, Williams TC, Templeton K
EPI_ISL_425991 hCoV-19/Scotland/EDB386/2020 Europe / United Kingdom / Scotland 2020-03-30 Virology Department, Royal Infirmary of Edinburgh, NHS Lothian / School of Biological Sciences, University of Edinburgh / Institute of Genetics and Molecular Medicine, University of Edinburgh COVID-19 Genomics UK (COG-UK) Consortium McHugh M, Dewar R, Rooke S, Gallagher M, Balcaza C, O'Toole A, Hill V, McCrone JT, Colquhoun R, Yu X, Jackson B, Scher E, Rambaut A, Williams TC, Templeton K
EPI_ISL_425992 hCoV-19/Scotland/EDB389/2020 Europe / United Kingdom / Scotland 2020-03-30 Virology Department, Royal Infirmary of Edinburgh, NHS Lothian / School of Biological Sciences, University of Edinburgh / Institute of Genetics and Molecular Medicine, University of Edinburgh COVID-19 Genomics UK (COG-UK) Consortium McHugh M, Dewar R, Rooke S, Gallagher M, Balcaza C, O'Toole A, Hill V, McCrone JT, Colquhoun R, Yu X, Jackson B, Scher E, Rambaut A, Williams TC, Templeton K
EPI_ISL_425993 hCoV-19/Scotland/EDB390/2020 Europe / United Kingdom / Scotland 2020-03-30 Virology Department, Royal Infirmary of Edinburgh, NHS Lothian / School of Biological Sciences, University of Edinburgh / Institute of Genetics and Molecular Medicine, University of Edinburgh COVID-19 Genomics UK (COG-UK) Consortium McHugh M, Dewar R, Rooke S, Gallagher M, Balcaza C, O'Toole A, Hill V, McCrone JT, Colquhoun R, Yu X, Jackson B, Scher E, Rambaut A, Williams TC, Templeton K
EPI_ISL_425994 hCoV-19/Scotland/EDB391/2020 Europe / United Kingdom / Scotland 2020-03-30 Virology Department, Royal Infirmary of Edinburgh, NHS Lothian / School of Biological Sciences, University of Edinburgh / Institute of Genetics and Molecular Medicine, University of Edinburgh COVID-19 Genomics UK (COG-UK) Consortium McHugh M, Dewar R, Rooke S, Gallagher M, Balcaza C, O'Toole A, Hill V, McCrone JT, Colquhoun R, Yu X, Jackson B, Scher E, Rambaut A, Williams TC, Templeton K
EPI_ISL_425995 hCoV-19/Scotland/EDB392/2020 Europe / United Kingdom / Scotland 2020-03-30 Virology Department, Royal Infirmary of Edinburgh, NHS Lothian / School of Biological Sciences, University of Edinburgh / Institute of Genetics and Molecular Medicine, University of Edinburgh COVID-19 Genomics UK (COG-UK) Consortium McHugh M, Dewar R, Rooke S, Gallagher M, Balcaza C, O'Toole A, Hill V, McCrone JT, Colquhoun R, Yu X, Jackson B, Scher E, Rambaut A, Williams TC, Templeton K
EPI_ISL_425996 hCoV-19/Scotland/EDB396/2020 Europe / United Kingdom / Scotland 2020-03-30 Virology Department, Royal Infirmary of Edinburgh, NHS Lothian / School of Biological Sciences, University of Edinburgh / Institute of Genetics and Molecular Medicine, University of Edinburgh COVID-19 Genomics UK (COG-UK) Consortium McHugh M, Dewar R, Rooke S, Gallagher M, Balcaza C, O'Toole A, Hill V, McCrone JT, Colquhoun R, Yu X, Jackson B, Scher E, Rambaut A, Williams TC, Templeton K
EPI_ISL_425997 hCoV-19/Scotland/EDB397/2020 Europe / United Kingdom / Scotland 2020-03-30 Virology Department, Royal Infirmary of Edinburgh, NHS Lothian / School of Biological Sciences, University of Edinburgh / Institute of Genetics and Molecular Medicine, University of Edinburgh COVID-19 Genomics UK (COG-UK) Consortium McHugh M, Dewar R, Rooke S, Gallagher M, Balcaza C, O'Toole A, Hill V, McCrone JT, Colquhoun R, Yu X, Jackson B, Scher E, Rambaut A, Williams TC, Templeton K
EPI_ISL_425998 hCoV-19/Scotland/EDB399/2020 Europe / United Kingdom / Scotland 2020-03-30 Virology Department, Royal Infirmary of Edinburgh, NHS Lothian / School of Biological Sciences, University of Edinburgh / Institute of Genetics and Molecular Medicine, University of Edinburgh COVID-19 Genomics UK (COG-UK) Consortium McHugh M, Dewar R, Rooke S, Gallagher M, Balcaza C, O'Toole A, Hill V, McCrone JT, Colquhoun R, Yu X, Jackson B, Scher E, Rambaut A, Williams TC, Templeton K
EPI_ISL_425999 hCoV-19/Scotland/EDB401/2020 Europe / United Kingdom / Scotland 2020-03-29 Virology Department, Royal Infirmary of Edinburgh, NHS Lothian / School of Biological Sciences, University of Edinburgh / Institute of Genetics and Molecular Medicine, University of Edinburgh COVID-19 Genomics UK (COG-UK) Consortium McHugh M, Dewar R, Rooke S, Gallagher M, Balcaza C, O'Toole A, Hill V, McCrone JT, Colquhoun R, Yu X, Jackson B, Scher E, Rambaut A, Williams TC, Templeton K
EPI_ISL_427001 hCoV-19/Australia/VIC716/2020 Oceania / Australia / Victoria 2020-03-27 Victorian Infectious Diseases Reference Laboratory (VIDRL) Microbiological Diagnostic Unit Public Health Laboratory and Victorian Infectious Diseases Reference Laboratory, Doherty Institute Caly L., Seemann T., Sait, M., Schultz M., Druce J., Sherry, N.
EPI_ISL_427002 hCoV-19/Australia/VIC718/2020 Oceania / Australia / Victoria 2020-03-27 Victorian Infectious Diseases Reference Laboratory (VIDRL) Microbiological Diagnostic Unit Public Health Laboratory and Victorian Infectious Diseases Reference Laboratory, Doherty Institute Caly L., Seemann T., Sait, M., Schultz M., Druce J., Sherry, N.
EPI_ISL_427003 hCoV-19/Australia/VIC720/2020 Oceania / Australia / Victoria 2020-03-28 Victorian Infectious Diseases Reference Laboratory (VIDRL) Microbiological Diagnostic Unit Public Health Laboratory and Victorian Infectious Diseases Reference Laboratory, Doherty Institute Caly L., Seemann T., Sait, M., Schultz M., Druce J., Sherry, N.
EPI_ISL_427006 hCoV-19/Australia/VIC725/2020 Oceania / Australia / Victoria 2020-03-28 Victorian Infectious Diseases Reference Laboratory (VIDRL) Microbiological Diagnostic Unit Public Health Laboratory and Victorian Infectious Diseases Reference Laboratory, Doherty Institute Caly L., Seemann T.,

Sait, M., Schultz M., Druce J., Sherry, N.
EPI_ISL_427010 hCoV-19/Australia/VIC731/2020 Oceania / Australia / Victoria 2020-03-30 Victorian Infectious Diseases Reference Laboratory (VIDRL) Microbiological Diagnostic Unit Public Health Laboratory and Victorian Infectious Diseases Reference Laboratory, Doherty Institute Caly L., Seemann T., Sait, M., Schultz M., Druce J., Sherry, N.
EPI_ISL_427011 hCoV-19/Australia/VIC732/2020 Oceania / Australia / Victoria 2020-03-30 Victorian Infectious Diseases Reference Laboratory (VIDRL) Microbiological Diagnostic Unit Public Health Laboratory and Victorian Infectious Diseases Reference Laboratory, Doherty Institute Caly L., Seemann T., Sait, M., Schultz M., Druce J., Sherry, N.
EPI_ISL_427012 hCoV-19/Australia/VIC733/2020 Oceania / Australia / Victoria 2020-03-30 Victorian Infectious Diseases Reference Laboratory (VIDRL) Microbiological Diagnostic Unit Public Health Laboratory and Victorian Infectious Diseases Reference Laboratory, Doherty Institute Caly L., Seemann T., Sait, M., Schultz M., Druce J., Sherry, N.
EPI_ISL_427013 hCoV-19/Australia/VIC734/2020 Oceania / Australia / Victoria 2020-03-30 Victorian Infectious Diseases Reference Laboratory (VIDRL) Microbiological Diagnostic Unit Public Health Laboratory and Victorian Infectious Diseases Reference Laboratory, Doherty Institute Caly L., Seemann T., Sait, M., Schultz M., Druce J., Sherry, N.
EPI_ISL_427014 hCoV-19/Australia/VIC735/2020 Oceania / Australia / Victoria 2020-03-30 Victorian Infectious Diseases Reference Laboratory (VIDRL) Microbiological Diagnostic Unit Public Health Laboratory and Victorian Infectious Diseases Reference Laboratory, Doherty Institute Caly L., Seemann T., Sait, M., Schultz M., Druce J., Sherry, N.
EPI_ISL_427015 hCoV-19/Australia/VIC736/2020 Oceania / Australia / Victoria 2020-03-30 Victorian Infectious Diseases Reference Laboratory (VIDRL) Microbiological Diagnostic Unit Public Health Laboratory and Victorian Infectious Diseases Reference Laboratory, Doherty Institute Caly L., Seemann T., Sait, M., Schultz M., Druce J., Sherry, N.
EPI_ISL_427016 hCoV-19/Australia/VIC737/2020 Oceania / Australia / Victoria 2020-03-30 Victorian Infectious Diseases Reference Laboratory (VIDRL) Microbiological Diagnostic Unit Public Health Laboratory and Victorian Infectious Diseases Reference Laboratory, Doherty Institute Caly L., Seemann T., Sait, M., Schultz M., Druce J., Sherry, N.
EPI_ISL_427017 hCoV-19/Australia/VIC738/2020 Oceania / Australia / Victoria 2020-03-30 Victorian Infectious Diseases Reference Laboratory (VIDRL) Microbiological Diagnostic Unit Public Health Laboratory and Victorian Infectious Diseases Reference Laboratory, Doherty Institute Caly L., Seemann T., Sait, M., Schultz M., Druce J., Sherry, N.
EPI_ISL_427018 hCoV-19/Australia/VIC739/2020 Oceania / Australia / Victoria 2020-03-30 Victorian Infectious Diseases Reference Laboratory (VIDRL) Microbiological Diagnostic Unit Public Health Laboratory and Victorian Infectious Diseases Reference Laboratory, Doherty Institute Caly L., Seemann T., Sait, M., Schultz M., Druce J., Sherry, N.
EPI_ISL_427019 hCoV-19/Australia/VIC741/2020 Oceania / Australia / Victoria 2020-03-30 Victorian Infectious Diseases Reference Laboratory (VIDRL) Microbiological Diagnostic Unit Public Health Laboratory and Victorian Infectious Diseases Reference Laboratory, Doherty Institute Caly L., Seemann T., Sait, M., Schultz M., Druce J., Sherry, N.
EPI_ISL_427020 hCoV-19/Australia/VIC742/2020 Oceania / Australia / Victoria 2020-03-29 Victorian Infectious Diseases Reference Laboratory (VIDRL) Microbiological Diagnostic Unit Public Health Laboratory and Victorian Infectious Diseases Reference Laboratory, Doherty Institute Caly L., Seemann T., Sait, M., Schultz M., Druce J., Sherry, N.
EPI_ISL_427022 hCoV-19/Australia/VIC744/2020 Oceania / Australia / Victoria 2020-03-29 Victorian Infectious Diseases Reference Laboratory (VIDRL) Microbiological Diagnostic Unit Public Health Laboratory and Victorian Infectious Diseases Reference Laboratory, Doherty Institute Caly L., Seemann T., Sait, M., Schultz M., Druce J., Sherry, N.
EPI_ISL_427023 hCoV-19/Australia/VIC745/2020 Oceania / Australia / Victoria 2020-03-29 Victorian Infectious Diseases Reference Laboratory (VIDRL) Microbiological Diagnostic Unit Public Health Laboratory and Victorian Infectious Diseases Reference Laboratory, Doherty Institute Caly L., Seemann T., Sait, M., Schultz M., Druce J., Sherry, N.
EPI_ISL_427024 hCoV-19/Australia/VIC746/2020 Oceania / Australia / Victoria 2020-03-29 Victorian Infectious Diseases Reference Laboratory (VIDRL) Microbiological Diagnostic Unit Public Health Laboratory and Victorian Infectious Diseases Reference Laboratory, Doherty Institute Caly L., Seemann T., Sait, M., Schultz M., Druce J., Sherry, N.
EPI_ISL_427025 hCoV-19/Australia/VIC747/2020 Oceania / Australia / Victoria 2020-03-31 Victorian Infectious Diseases Reference Laboratory (VIDRL) Microbiological Diagnostic Unit Public Health Laboratory and Victorian Infectious Diseases Reference Laboratory, Doherty Institute Caly L., Seemann T., Sait, M., Schultz M., Druce J., Sherry, N.
EPI_ISL_427026 hCoV-19/Australia/VIC748/2020 Oceania / Australia / Victoria 2020-03-30 Victorian Infectious Diseases Reference Laboratory (VIDRL) Microbiological Diagnostic Unit Public Health Laboratory and Victorian Infectious Diseases Reference Laboratory, Doherty Institute Caly L., Seemann T., Sait, M., Schultz M., Druce J., Sherry, N.
EPI_ISL_427028 hCoV-19/Australia/VIC751/2020 Oceania / Australia / Victoria 2020-03-30 Victorian Infectious Diseases Reference Laboratory (VIDRL) Microbiological Diagnostic Unit Public Health Laboratory and Victorian Infectious Diseases Reference Laboratory, Doherty Institute Caly L., Seemann T., Sait, M., Schultz M., Druce J., Sherry, N.
EPI_ISL_427029 hCoV-19/Australia/VIC752/2020 Oceania / Australia / Victoria 2020-03-30 Victorian

Infectious Diseases Reference Laboratory (VIDRL)	Microbiological Diagnostic Unit Public Health Laboratory and Victorian Infectious Diseases Reference Laboratory, Doherty Institute	Caly L., Seemann T., Sait, M., Schultz M., Druce J., Sherry, N.
EPI_ISL_427031	hCoV-19/Australia/VIC754/2020	Oceania / Australia / Victoria	2020-03-30	Victorian Infectious Diseases Reference Laboratory (VIDRL)	Microbiological Diagnostic Unit Public Health Laboratory and Victorian Infectious Diseases Reference Laboratory, Doherty Institute	Caly L., Seemann T., Sait, M., Schultz M., Druce J., Sherry, N.
EPI_ISL_427034	hCoV-19/Australia/VIC757/2020	Oceania / Australia / Victoria	2020-03-31	Victorian Infectious Diseases Reference Laboratory (VIDRL)	Microbiological Diagnostic Unit Public Health Laboratory and Victorian Infectious Diseases Reference Laboratory, Doherty Institute	Caly L., Seemann T., Sait, M., Schultz M., Druce J., Sherry, N.
EPI_ISL_427038	hCoV-19/Australia/VIC762/2020	Oceania / Australia / Victoria	2020-03-29	Victorian Infectious Diseases Reference Laboratory (VIDRL)	Microbiological Diagnostic Unit Public Health Laboratory and Victorian Infectious Diseases Reference Laboratory, Doherty Institute	Caly L., Seemann T., Sait, M., Schultz M., Druce J., Sherry, N.
EPI_ISL_427040	hCoV-19/Australia/VIC764/2020	Oceania / Australia / Victoria	2020-03-29	Victorian Infectious Diseases Reference Laboratory (VIDRL)	Microbiological Diagnostic Unit Public Health Laboratory and Victorian Infectious Diseases Reference Laboratory, Doherty Institute	Caly L., Seemann T., Sait, M., Schultz M., Druce J., Sherry, N.
EPI_ISL_427041	hCoV-19/Australia/VIC765/2020	Oceania / Australia / Victoria	2020-03-29	Victorian Infectious Diseases Reference Laboratory (VIDRL)	Microbiological Diagnostic Unit Public Health Laboratory and Victorian Infectious Diseases Reference Laboratory, Doherty Institute	Caly L., Seemann T., Sait, M., Schultz M., Druce J., Sherry, N.
EPI_ISL_427043	hCoV-19/Greece/13/2020	Europe / Greece / Athens	2020-03-18	Laboratory of Microbiology, Medical School, National and Kapodistrian University of Athens	Laboratory of Biology, Department of Medicine, Democritus University of Thrace	Bampali,M., Dovrolis,N., Gatzidou,E., Froukala,E., Stavropoulou,A., Veletza,S., Tsakris,A., Spanakis,N. and Karakasiliotis,I.
EPI_ISL_427045	hCoV-19/Australia/VIC768/2020	Oceania / Australia / Victoria	2020-03-20	Victorian Infectious Diseases Reference Laboratory (VIDRL)	Microbiological Diagnostic Unit Public Health Laboratory and Victorian Infectious Diseases Reference Laboratory, Doherty Institute	Caly L., Seemann T., Sait, M., Schultz M., Druce J., Sherry, N.
EPI_ISL_427046	hCoV-19/Australia/VIC769/2020	Oceania / Australia / Victoria	2020-03-20	Victorian Infectious Diseases Reference Laboratory (VIDRL)	Microbiological Diagnostic Unit Public Health Laboratory and Victorian Infectious Diseases Reference Laboratory, Doherty Institute	Caly L., Seemann T., Sait, M., Schultz M., Druce J., Sherry, N.
EPI_ISL_427048	hCoV-19/Australia/VIC772/2020	Oceania / Australia / Victoria	2020-03-20	Victorian Infectious Diseases Reference Laboratory (VIDRL)	Microbiological Diagnostic Unit Public Health Laboratory and Victorian Infectious Diseases Reference Laboratory, Doherty Institute	Caly L., Seemann T., Sait, M., Schultz M., Druce J., Sherry, N.
EPI_ISL_427049	hCoV-19/Australia/VIC773/2020	Oceania / Australia / Victoria	2020-03-20	Victorian Infectious Diseases Reference Laboratory (VIDRL)	Microbiological Diagnostic Unit Public Health Laboratory and Victorian Infectious Diseases Reference Laboratory, Doherty Institute	Caly L., Seemann T., Sait, M., Schultz M., Druce J., Sherry, N.
EPI_ISL_427054	hCoV-19/Australia/VIC779/2020	Oceania / Australia / Victoria	2020-03-30	Microbiological Diagnostic Unit Public Health Laboratory	Microbiological Diagnostic Unit Public Health Laboratory	Seemann T., Schultz M., Sait, M., Sherry, N.
EPI_ISL_427055	hCoV-19/Australia/VIC780/2020	Oceania / Australia / Victoria	2020-03-30	Microbiological Diagnostic Unit Public Health Laboratory	Microbiological Diagnostic Unit Public Health Laboratory	Seemann T., Schultz M., Sait, M., Sherry, N.
EPI_ISL_427057	hCoV-19/Australia/VIC782/2020	Oceania / Australia / Victoria	2020-03-30	Microbiological Diagnostic Unit Public Health Laboratory	Microbiological Diagnostic Unit Public Health Laboratory	Seemann T., Schultz M., Sait, M., Sherry, N.
EPI_ISL_427058	hCoV-19/Australia/VIC783/2020	Oceania / Australia / Victoria	2020-03-30	Microbiological Diagnostic Unit Public Health Laboratory	Microbiological Diagnostic Unit Public Health Laboratory	Seemann T., Schultz M., Sait, M., Sherry, N.
EPI_ISL_427060	hCoV-19/Australia/VIC785/2020	Oceania / Australia / Victoria	2020-03-30	Microbiological Diagnostic Unit Public Health Laboratory	Microbiological Diagnostic Unit Public Health Laboratory	Seemann T., Schultz M., Sait, M., Sherry, N.
EPI_ISL_427061	hCoV-19/Australia/VIC786/2020	Oceania / Australia / Victoria	2020-03-30	Microbiological Diagnostic Unit Public Health Laboratory	Microbiological Diagnostic Unit Public Health Laboratory	Seemann T., Schultz M., Sait, M., Sherry, N.
EPI_ISL_427062	hCoV-19/Australia/VIC787/2020	Oceania / Australia / Victoria	2020-03-30	Microbiological Diagnostic Unit Public Health Laboratory	Microbiological Diagnostic Unit Public Health Laboratory	Seemann T., Schultz M., Sait, M., Sherry, N.
EPI_ISL_427063	hCoV-19/Australia/VIC788/2020	Oceania / Australia / Victoria	2020-03-31	Microbiological Diagnostic Unit Public Health Laboratory	Microbiological Diagnostic Unit Public Health Laboratory	Seemann T., Schultz M., Sait, M., Sherry, N.
EPI_ISL_427065	hCoV-19/Australia/VIC790/2020	Oceania / Australia / Victoria	2020-03-31	Microbiological Diagnostic Unit Public Health Laboratory	Microbiological Diagnostic Unit Public Health Laboratory	Seemann T., Schultz M., Sait, M., Sherry, N.

EPI_ISL_427068 hCoV-19/Australia/VIC793/2020 Oceania / Australia / Victoria 2020-03-31 Microbiological Diagnostic Unit Public Health Laboratory Microbiological Diagnostic Unit Public Health Laboratory Seemann T., Schultz M., Sait, M., Sherry, N.
EPI_ISL_427069 hCoV-19/Australia/VIC794/2020 Oceania / Australia / Victoria 2020-03-31 Microbiological Diagnostic Unit Public Health Laboratory Microbiological Diagnostic Unit Public Health Laboratory Seemann T., Schultz M., Sait, M., Sherry, N.
EPI_ISL_427070 hCoV-19/Australia/VIC796/2020 Oceania / Australia / Victoria 2020-03-31 Microbiological Diagnostic Unit Public Health Laboratory Microbiological Diagnostic Unit Public Health Laboratory Seemann T., Schultz M., Sait, M., Sherry, N.
EPI_ISL_427071 hCoV-19/Australia/VIC797/2020 Oceania / Australia / Victoria 2020-04-01 Microbiological Diagnostic Unit Public Health Laboratory Microbiological Diagnostic Unit Public Health Laboratory Seemann T., Schultz M., Sait, M., Sherry, N.
EPI_ISL_427072 hCoV-19/Australia/VIC798/2020 Oceania / Australia / Victoria 2020-04-01 Microbiological Diagnostic Unit Public Health Laboratory Microbiological Diagnostic Unit Public Health Laboratory Seemann T., Schultz M., Sait, M., Sherry, N.
EPI_ISL_427073 hCoV-19/Australia/VIC799/2020 Oceania / Australia / Victoria 2020-04-01 Microbiological Diagnostic Unit Public Health Laboratory Microbiological Diagnostic Unit Public Health Laboratory Seemann T., Schultz M., Sait, M., Sherry, N.
EPI_ISL_427074 hCoV-19/Australia/VIC800/2020 Oceania / Australia / Victoria 2020-04-01 Microbiological Diagnostic Unit Public Health Laboratory Microbiological Diagnostic Unit Public Health Laboratory Seemann T., Schultz M., Sait, M., Sherry, N.
EPI_ISL_427076 hCoV-19/Australia/VIC802/2020 Oceania / Australia / Victoria 2020-04-01 Microbiological Diagnostic Unit Public Health Laboratory Microbiological Diagnostic Unit Public Health Laboratory Seemann T., Schultz M., Sait, M., Sherry, N.
EPI_ISL_427079 hCoV-19/Australia/VIC805/2020 Oceania / Australia / Victoria 2020-03-16 Victorian Infectious Diseases Reference Laboratory (VIDRL) Microbiological Diagnostic Unit Public Health Laboratory and Victorian Infectious Diseases Reference Laboratory, Doherty Institute Caly L., Seemann T., Sait, M., Schultz M., Druce J., Sherry, N.
EPI_ISL_427080 hCoV-19/Australia/VIC806/2020 Oceania / Australia / Victoria 2020-03-23 Victorian Infectious Diseases Reference Laboratory (VIDRL) Microbiological Diagnostic Unit Public Health Laboratory and Victorian Infectious Diseases Reference Laboratory, Doherty Institute Caly L., Seemann T., Sait, M., Schultz M., Druce J., Sherry, N.
EPI_ISL_427081 hCoV-19/Australia/VIC807/2020 Oceania / Australia / Victoria 2020-03-24 Victorian Infectious Diseases Reference Laboratory (VIDRL) Microbiological Diagnostic Unit Public Health Laboratory and Victorian Infectious Diseases Reference Laboratory, Doherty Institute Caly L., Seemann T., Sait, M., Schultz M., Druce J., Sherry, N.
EPI_ISL_427082 hCoV-19/Australia/VIC808/2020 Oceania / Australia / Victoria 2020-03-28 Victorian Infectious Diseases Reference Laboratory (VIDRL) Microbiological Diagnostic Unit Public Health Laboratory and Victorian Infectious Diseases Reference Laboratory, Doherty Institute Caly L., Seemann T., Sait, M., Schultz M., Druce J., Sherry, N.
EPI_ISL_427085 hCoV-19/Australia/VIC812/2020 Oceania / Australia / Victoria 2020-04-02 Victorian Infectious Diseases Reference Laboratory (VIDRL) Microbiological Diagnostic Unit Public Health Laboratory and Victorian Infectious Diseases Reference Laboratory, Doherty Institute Caly L., Seemann T., Sait, M., Schultz M., Druce J., Sherry, N.
EPI_ISL_427092 hCoV-19/Australia/VIC820/2020 Oceania / Australia / Victoria 2020-04-03 Victorian Infectious Diseases Reference Laboratory (VIDRL) Microbiological Diagnostic Unit Public Health Laboratory and Victorian Infectious Diseases Reference Laboratory, Doherty Institute Caly L., Seemann T., Sait, M., Schultz M., Druce J., Sherry, N.
EPI_ISL_427093 hCoV-19/Australia/VIC821/2020 Oceania / Australia / Victoria 2020-04-03 Victorian Infectious Diseases Reference Laboratory (VIDRL) Microbiological Diagnostic Unit Public Health Laboratory and Victorian Infectious Diseases Reference Laboratory, Doherty Institute Caly L., Seemann T., Sait, M., Schultz M., Druce J., Sherry, N.
EPI_ISL_427094 hCoV-19/Australia/VIC823/2020 Oceania / Australia / Victoria 2020-04-03 Victorian Infectious Diseases Reference Laboratory (VIDRL) Microbiological Diagnostic Unit Public Health Laboratory and Victorian Infectious Diseases Reference Laboratory, Doherty Institute Caly L., Seemann T., Sait, M., Schultz M., Druce J., Sherry, N.
EPI_ISL_427095 hCoV-19/Australia/VIC824/2020 Oceania / Australia / Victoria 2020-04-03 Victorian Infectious Diseases Reference Laboratory (VIDRL) Microbiological Diagnostic Unit Public Health Laboratory and Victorian Infectious Diseases Reference Laboratory, Doherty Institute Caly L., Seemann T., Sait, M., Schultz M., Druce J., Sherry, N.
EPI_ISL_427096 hCoV-19/Australia/VIC828/2020 Oceania / Australia / Victoria 2020-04-03 Victorian Infectious Diseases Reference Laboratory (VIDRL) Microbiological Diagnostic Unit Public Health Laboratory and Victorian Infectious Diseases Reference Laboratory, Doherty Institute Caly L., Seemann T., Sait, M., Schultz M., Druce J., Sherry, N.
EPI_ISL_428300 hCoV-19/USA/WI-UW-204/2020 North America / USA / Wisconsin / Campbellsp 2020-03-25 University of Wisconsin-Madison AIDS Vaccine Research Laboratories University of Wisconsin-Madison AIDS Vaccine Research Laboratories Gage Moreno, Katarina Braun, et al. AIDS Vaccine Research Laboratories
EPI_ISL_428301 hCoV-19/USA/WI-UW-205/2020 North America / USA / Wisconsin / Milwaukee 2020-03-25 University of Wisconsin-Madison AIDS Vaccine Research Laboratories University of Wisconsin-Madison AIDS Vaccine Research Laboratories Gage Moreno, Katarina Braun, et al. AIDS Vaccine Research Laboratories

EPI_ISL_428302 hCoV-19/USA/WI-UW-206/2020 North America / USA / Wisconsin / Grafton 2020-03-25 University of Wisconsin-Madison AIDS Vaccine Research Laboratories University of Wisconsin-Madison AIDS Vaccine Research Laboratories Gage Moreno, Katarina Braun, et al. AIDS Vaccine Research Laboratories
EPI_ISL_428303 hCoV-19/USA/WI-UW-207/2020 North America / USA / Wisconsin / Milwaukee 2020-03-23 University of Wisconsin-Madison AIDS Vaccine Research Laboratories University of Wisconsin-Madison AIDS Vaccine Research Laboratories Gage Moreno, Katarina Braun, et al. AIDS Vaccine Research Laboratories
EPI_ISL_428304 hCoV-19/USA/WI-UW-208/2020 North America / USA / Wisconsin / Milwaukee 2020-03-25 University of Wisconsin-Madison AIDS Vaccine Research Laboratories University of Wisconsin-Madison AIDS Vaccine Research Laboratories Gage Moreno, Katarina Braun, et al. AIDS Vaccine Research Laboratories
EPI_ISL_428305 hCoV-19/USA/WI-UW-209/2020 North America / USA / Wisconsin / Milwaukee 2020-03-25 University of Wisconsin-Madison AIDS Vaccine Research Laboratories University of Wisconsin-Madison AIDS Vaccine Research Laboratories Gage Moreno, Katarina Braun, et al. AIDS Vaccine Research Laboratories
EPI_ISL_428306 hCoV-19/USA/WI-UW-210/2020 North America / USA / Wisconsin / Milwaukee 2020-03-24 University of Wisconsin-Madison AIDS Vaccine Research Laboratories University of Wisconsin-Madison AIDS Vaccine Research Laboratories Gage Moreno, Katarina Braun, et al. AIDS Vaccine Research Laboratories
EPI_ISL_428307 hCoV-19/USA/WI-UW-211/2020 North America / USA / Wisconsin / Milwaukee 2020-03-25 University of Wisconsin-Madison AIDS Vaccine Research Laboratories University of Wisconsin-Madison AIDS Vaccine Research Laboratories Gage Moreno, Katarina Braun, et al. AIDS Vaccine Research Laboratories
EPI_ISL_428308 hCoV-19/USA/WI-UW-212/2020 North America / USA / Wisconsin / Milwaukee 2020-03-25 University of Wisconsin-Madison AIDS Vaccine Research Laboratories University of Wisconsin-Madison AIDS Vaccine Research Laboratories Gage Moreno, Katarina Braun, et al. AIDS Vaccine Research Laboratories
EPI_ISL_428309 hCoV-19/USA/WI-UW-213/2020 North America / USA / Wisconsin / Milwaukee 2020-03-25 University of Wisconsin-Madison AIDS Vaccine Research Laboratories University of Wisconsin-Madison AIDS Vaccine Research Laboratories Gage Moreno, Katarina Braun, et al. AIDS Vaccine Research Laboratories
EPI_ISL_428310 hCoV-19/USA/WI-UW-214/2020 North America / USA / Wisconsin / Grafton 2020-03-25 University of Wisconsin-Madison AIDS Vaccine Research Laboratories University of Wisconsin-Madison AIDS Vaccine Research Laboratories Gage Moreno, Katarina Braun, et al. AIDS Vaccine Research Laboratories
EPI_ISL_428311 hCoV-19/USA/WI-UW-215/2020 North America / USA / Wisconsin / Milwaukee 2020-03-25 University of Wisconsin-Madison AIDS Vaccine Research Laboratories University of Wisconsin-Madison AIDS Vaccine Research Laboratories Gage Moreno, Katarina Braun, et al. AIDS Vaccine Research Laboratories
EPI_ISL_428312 hCoV-19/USA/WI-UW-216/2020 North America / USA / Wisconsin / Milwaukee 2020-03-25 University of Wisconsin-Madison AIDS Vaccine Research Laboratories University of Wisconsin-Madison AIDS Vaccine Research Laboratories Gage Moreno, Katarina Braun, et al. AIDS Vaccine Research Laboratories
EPI_ISL_428313 hCoV-19/USA/WI-UW-217/2020 North America / USA / Wisconsin / Milwaukee 2020-03-25 University of Wisconsin-Madison AIDS Vaccine Research Laboratories University of Wisconsin-Madison AIDS Vaccine Research Laboratories Gage Moreno, Katarina Braun, et al. AIDS Vaccine Research Laboratories
EPI_ISL_428314 hCoV-19/USA/WI-UW-218/2020 North America / USA / Wisconsin / Milwaukee 2020-03-25 University of Wisconsin-Madison AIDS Vaccine Research Laboratories University of Wisconsin-Madison AIDS Vaccine Research Laboratories Gage Moreno, Katarina Braun, et al. AIDS Vaccine Research Laboratories
EPI_ISL_428315 hCoV-19/USA/WI-UW-219/2020 North America / USA / Wisconsin / Milwaukee 2020-03-25 University of Wisconsin-Madison AIDS Vaccine Research Laboratories University of Wisconsin-Madison AIDS Vaccine Research Laboratories Gage Moreno, Katarina Braun, et al. AIDS Vaccine Research Laboratories
EPI_ISL_428316 hCoV-19/USA/WI-UW-220/2020 North America / USA / Wisconsin / Mequon 2020-03-25 University of Wisconsin-Madison AIDS Vaccine Research Laboratories University of Wisconsin-Madison AIDS Vaccine Research Laboratories Gage Moreno, Katarina Braun, et al. AIDS Vaccine Research Laboratories
EPI_ISL_428317 hCoV-19/USA/WI-UW-221/2020 North America / USA / Wisconsin / Milwaukee 2020-03-25 University of Wisconsin-Madison AIDS Vaccine Research Laboratories University of Wisconsin-Madison AIDS Vaccine Research Laboratories Gage Moreno, Katarina Braun, et al. AIDS Vaccine Research Laboratories
EPI_ISL_428318 hCoV-19/USA/WI-UW-222/2020 North America / USA / Wisconsin / Milwaukee 2020-03-25 University of Wisconsin-Madison AIDS Vaccine Research Laboratories University of Wisconsin-Madison AIDS Vaccine Research Laboratories Gage Moreno, Katarina Braun, et al. AIDS Vaccine Research Laboratories
EPI_ISL_428319 hCoV-19/USA/WI-UW-223/2020 North America / USA / Wisconsin / Milwaukee 2020-03-26 University of Wisconsin-Madison AIDS Vaccine Research Laboratories University of Wisconsin-Madison AIDS Vaccine Research Laboratories Gage Moreno, Katarina Braun, et al. AIDS Vaccine Research Laboratories
EPI_ISL_428320 hCoV-19/USA/WI-UW-224/2020 North America / USA / Wisconsin / Brookfield 2020-03-26 University of Wisconsin-Madison AIDS Vaccine Research Laboratories University of Wisconsin-Madison AIDS Vaccine Research Laboratories Gage Moreno, Katarina Braun, et al. AIDS Vaccine Research Laboratories
EPI_ISL_428321 hCoV-19/USA/WI-UW-225/2020 North America / USA / Wisconsin / Franklin 2020-03-26 University of Wisconsin-Madison AIDS Vaccine Research Laboratories University of Wisconsin-Madison AIDS Vaccine Research Laboratories Gage Moreno, Katarina Braun, et al. AIDS Vaccine Research Laboratories
EPI_ISL_428322 hCoV-19/USA/WI-UW-226/2020 North America / USA / Wisconsin / South Milwaukee 2020-03-26 University of Wisconsin-Madison AIDS Vaccine Research Laboratories University of Wisconsin-Madison AIDS Vaccine Research Laboratories Gage Moreno, Katarina Braun, et al. AIDS Vaccine Research Laboratories
EPI_ISL_428323 hCoV-19/USA/WI-UW-227/2020 North America / USA / Wisconsin / Milwaukee 2020-03-26 University of Wisconsin-Madison AIDS Vaccine Research Laboratories University of Wisconsin-Madison AIDS Vaccine Research Laboratories Gage Moreno, Katarina Braun, et al. AIDS Vaccine Research Laboratories
EPI_ISL_428324 hCoV-19/USA/WI-UW-228/2020 North America / USA / Wisconsin / Greenfield 2020-03-26 University of Wisconsin-Madison AIDS Vaccine Research Laboratories University of Wisconsin-Madison AIDS Vaccine Research Laboratories Gage Moreno, Katarina Braun, et al. AIDS Vaccine Research Laboratories
EPI_ISL_428325 hCoV-19/USA/WI-UW-229/2020 North America / USA / Wisconsin / Milwaukee 2020-03-26

University of Wisconsin-Madison AIDS Vaccine Research Laboratories	University of Wisconsin-Madison AIDS Vaccine Research Laboratories	Gage Moreno, Katarina Braun, et al. AIDS Vaccine Research Laboratories
EPI_ISL_428326	hCoV-19/USA/WI-UW-230/2020	North America / USA / Wisconsin / Milwaukee	2020-03-26	University of Wisconsin-Madison AIDS Vaccine Research Laboratories	University of Wisconsin-Madison AIDS Vaccine Research Laboratories	Gage Moreno, Katarina Braun, et al. AIDS Vaccine Research Laboratories
EPI_ISL_428327	hCoV-19/USA/WI-UW-231/2020	North America / USA / Wisconsin / Milwaukee	2020-03-26	University of Wisconsin-Madison AIDS Vaccine Research Laboratories	University of Wisconsin-Madison AIDS Vaccine Research Laboratories	Gage Moreno, Katarina Braun, et al. AIDS Vaccine Research Laboratories
EPI_ISL_428328	hCoV-19/USA/WI-UW-232/2020	North America / USA / Wisconsin / Glendale	2020-03-27	University of Wisconsin-Madison AIDS Vaccine Research Laboratories	University of Wisconsin-Madison AIDS Vaccine Research Laboratories	Gage Moreno, Katarina Braun, et al. AIDS Vaccine Research Laboratories
EPI_ISL_428329	hCoV-19/USA/WI-UW-233/2020	North America / USA / Wisconsin / Milwaukee	2020-03-27	University of Wisconsin-Madison AIDS Vaccine Research Laboratories	University of Wisconsin-Madison AIDS Vaccine Research Laboratories	Gage Moreno, Katarina Braun, et al. AIDS Vaccine Research Laboratories
EPI_ISL_428330	hCoV-19/USA/WI-UW-234/2020	North America / USA / Wisconsin / Milwaukee	2020-03-26	University of Wisconsin-Madison AIDS Vaccine Research Laboratories	University of Wisconsin-Madison AIDS Vaccine Research Laboratories	Gage Moreno, Katarina Braun, et al. AIDS Vaccine Research Laboratories
EPI_ISL_428331	hCoV-19/USA/WI-UW-235/2020	North America / USA / Wisconsin / Cudahy	2020-03-28	University of Wisconsin-Madison AIDS Vaccine Research Laboratories	University of Wisconsin-Madison AIDS Vaccine Research Laboratories	Gage Moreno, Katarina Braun, et al. AIDS Vaccine Research Laboratories
EPI_ISL_428332	hCoV-19/USA/WI-UW-236/2020	North America / USA / Wisconsin / Milwaukee	2020-03-27	University of Wisconsin-Madison AIDS Vaccine Research Laboratories	University of Wisconsin-Madison AIDS Vaccine Research Laboratories	Gage Moreno, Katarina Braun, et al. AIDS Vaccine Research Laboratories
EPI_ISL_428333	hCoV-19/USA/WI-UW-237/2020	North America / USA / Wisconsin / Whitefish	2020-03-27	University of Wisconsin-Madison AIDS Vaccine Research Laboratories	University of Wisconsin-Madison AIDS Vaccine Research Laboratories	Gage Moreno, Katarina Braun, et al. AIDS Vaccine Research Laboratories
EPI_ISL_428334	hCoV-19/USA/WI-UW-238/2020	North America / USA / Wisconsin / Milwaukee	2020-03-27	University of Wisconsin-Madison AIDS Vaccine Research Laboratories	University of Wisconsin-Madison AIDS Vaccine Research Laboratories	Gage Moreno, Katarina Braun, et al. AIDS Vaccine Research Laboratories
EPI_ISL_428335	hCoV-19/USA/WI-UW-239/2020	North America / USA / Wisconsin / Mequon	2020-03-27	University of Wisconsin-Madison AIDS Vaccine Research Laboratories	University of Wisconsin-Madison AIDS Vaccine Research Laboratories	Gage Moreno, Katarina Braun, et al. AIDS Vaccine Research Laboratories
EPI_ISL_428336	hCoV-19/USA/WI-UW-240/2020	North America / USA / Wisconsin / Milwaukee	2020-03-27	University of Wisconsin-Madison AIDS Vaccine Research Laboratories	University of Wisconsin-Madison AIDS Vaccine Research Laboratories	Gage Moreno, Katarina Braun, et al. AIDS Vaccine Research Laboratories
EPI_ISL_428337	hCoV-19/USA/WI-UW-241/2020	North America / USA / Wisconsin / Milwaukee	2020-03-27	University of Wisconsin-Madison AIDS Vaccine Research Laboratories	University of Wisconsin-Madison AIDS Vaccine Research Laboratories	Gage Moreno, Katarina Braun, et al. AIDS Vaccine Research Laboratories
EPI_ISL_428338	hCoV-19/USA/WI-UW-242/2020	North America / USA / Wisconsin / Milwaukee	2020-03-28	University of Wisconsin-Madison AIDS Vaccine Research Laboratories	University of Wisconsin-Madison AIDS Vaccine Research Laboratories	Gage Moreno, Katarina Braun, et al. AIDS Vaccine Research Laboratories
EPI_ISL_428339	hCoV-19/USA/WI-UW-243/2020	North America / USA / Wisconsin / Grafton	2020-03-28	University of Wisconsin-Madison AIDS Vaccine Research Laboratories	University of Wisconsin-Madison AIDS Vaccine Research Laboratories	Gage Moreno, Katarina Braun, et al. AIDS Vaccine Research Laboratories
EPI_ISL_428340	hCoV-19/USA/WI-UW-244/2020	North America / USA / Wisconsin / Thiensvill	2020-03-28	University of Wisconsin-Madison AIDS Vaccine Research Laboratories	University of Wisconsin-Madison AIDS Vaccine Research Laboratories	Gage Moreno, Katarina Braun, et al. AIDS Vaccine Research Laboratories
EPI_ISL_428341	hCoV-19/USA/WI-UW-245/2020	North America / USA / Wisconsin / Milwaukee	2020-03-28	University of Wisconsin-Madison AIDS Vaccine Research Laboratories	University of Wisconsin-Madison AIDS Vaccine Research Laboratories	Gage Moreno, Katarina Braun, et al. AIDS Vaccine Research Laboratories
EPI_ISL_428343	hCoV-19/USA/WI-UW-247/2020	North America / USA / Wisconsin / Port Washi	2020-03-28	University of Wisconsin-Madison AIDS Vaccine Research Laboratories	University of Wisconsin-Madison AIDS Vaccine Research Laboratories	Gage Moreno, Katarina Braun, et al. AIDS Vaccine Research Laboratories
EPI_ISL_428344	hCoV-19/USA/WI-UW-248/2020	North America / USA / Wisconsin / Milwaukee	2020-03-28	University of Wisconsin-Madison AIDS Vaccine Research Laboratories	University of Wisconsin-Madison AIDS Vaccine Research Laboratories	Gage Moreno, Katarina Braun, et al. AIDS Vaccine Research Laboratories
EPI_ISL_428345	hCoV-19/USA/WI-UW-249/2020	North America / USA / Wisconsin / Racine	2020-03-16	University of Wisconsin-Madison AIDS Vaccine Research Laboratories	University of Wisconsin-Madison AIDS Vaccine Research Laboratories	Gage Moreno, Katarina Braun, et al. AIDS Vaccine Research Laboratories
EPI_ISL_428346	hCoV-19/Turkey/GLAB-CoV012/2020 Europe / Turkey / Istanbul	2020-04-17	Genomic Laboratory (GLAB) (Conjoint lab of Health Directorate of Istanbul and Istanbul Technical University)	Genomic Laboratory (GLAB), Istanbul Technical University	Ilker Karacan, Tugba Kizilboga Akgun, Bugra Agaoglu, Gizem Alkurt, Jale Yildiz, Betsi Köse, Elifnaz Çelik, Arzu Irvem, Yasemin Kendir Demirkol, Ozlem Akgun Dogan, Mehtap Aydın, Levent Doganay, Gizem Dinler Doganay
EPI_ISL_428347	hCoV-19/France/IDF3384/2020	Europe / France / Ile De France / Longjumeau	2020-03-23	Service de Biologie Médicale - BP 125	National Reference Center for Viruses of Respiratory Infections, Institut Pasteur, Paris	Mélanie Albert, Marion Barbet, Sylvie Behillil, Méline Bizard, Angela Brisebarre, Flora Donati, Etienne Simon-Lorière, Vincent Enouf, Maud Vanpeene, Sylvie van der Werf
EPI_ISL_428348	hCoV-19/France/GE3372/2020	Europe / France / Grand-est / Gondrecourt-le-chateau	2020-03-20	Maison de Santé du Val d'Ormois	National Reference Center for Viruses of Respiratory Infections,

Institut Pasteur, Paris Mélanie Albert, Marion Barbet, Sylvie Behillil, Méline Bizard, Angela Brisebarre, Flora Donati, Etienne Simon-Lorière, Vincent Enouf, Maud Vanpeene, Sylvie van der Werf
EPI_ISL_428349 hCoV-19/France/IDF3386/2020 Europe / France / Ile De France / Longjumeau 2020-03-23 Service de Biologie Médicale - BP 125 National Reference Center for Viruses of Respiratory Infections, Institut Pasteur, Paris Mélanie Albert, Marion Barbet, Sylvie Behillil, Méline Bizard, Angela Brisebarre, Flora Donati, Etienne Simon-Lorière, Vincent Enouf, Maud Vanpeene, Sylvie van der Werf
EPI_ISL_428350 hCoV-19/France/HF3419/2020 Europe / France / Hauts De France / Chateau-thierry 2020-03-24 CH Jean de Navarre Laboratoire de Biologie National Reference Center for Viruses of Respiratory Infections, Institut Pasteur, Paris Mélanie Albert, Marion Barbet, Sylvie Behillil, Méline Bizard, Angela Brisebarre, Flora Donati, Etienne Simon-Lorière, Vincent Enouf, Maud Vanpeene, Sylvie van der Werf
EPI_ISL_428351 hCoV-19/France/IDF3509/2020 Europe / France / Ile De France / Orsay 2020-03-24 GH Nord Essonne Service de Biologie clinique National Reference Center for Viruses of Respiratory Infections, Institut Pasteur, Paris Mélanie Albert, Marion Barbet, Sylvie Behillil, Méline Bizard, Angela Brisebarre, Flora Donati, Etienne Simon-Lorière, Vincent Enouf, Maud Vanpeene, Sylvie van der Werf
EPI_ISL_428352 hCoV-19/France/IDF3518/2020 Europe / France / Ile De France / Orsay 2020-03-23 GH Nord Essonne Service de Biologie clinique National Reference Center for Viruses of Respiratory Infections, Institut Pasteur, Paris Mélanie Albert, Marion Barbet, Sylvie Behillil, Méline Bizard, Angela Brisebarre, Flora Donati, Etienne Simon-Lorière, Vincent Enouf, Maud Vanpeene, Sylvie van der Werf
EPI_ISL_428353 hCoV-19/France/HF3534/2020 Europe / France / Hauts De France / Compiegne 2020-03-24 CH Compiègne Laboratoire de Biologie National Reference Center for Viruses of Respiratory Infections, Institut Pasteur, Paris Mélanie Albert, Marion Barbet, Sylvie Behillil, Méline Bizard, Angela Brisebarre, Flora Donati, Etienne Simon-Lorière, Vincent Enouf, Maud Vanpeene, Sylvie van der Werf
EPI_ISL_428354 hCoV-19/France/IDF3577/2020 Europe / France / Ile De France / Longjumeau 2020-03-25 LABM GH nord Essonne de Longjumeau - BP 125 National Reference Center for Viruses of Respiratory Infections, Institut Pasteur, Paris Mélanie Albert, Marion Barbet, Sylvie Behillil, Méline Bizard, Angela Brisebarre, Flora Donati, Etienne Simon-Lorière, Vincent Enouf, Maud Vanpeene, Sylvie van der Werf
EPI_ISL_428355 hCoV-19/France/IDF2251/2020 Europe / France / Ile De France / Garches 2020-03-11 Institut Médico légal- Hop R. Poincaré National Reference Center for Viruses of Respiratory Infections, Institut Pasteur, Paris Mélanie Albert, Marion Barbet, Sylvie Behillil, Méline Bizard, Angela Brisebarre, Flora Donati, Etienne Simon-Lorière, Vincent Enouf, Maud Vanpeene, Sylvie van der Werf
EPI_ISL_428356 hCoV-19/France/IDF2252/2020 Europe / France / Ile De France / Garches 2020-03-11 Institut Médico légal- Hop R. Poincaré National Reference Center for Viruses of Respiratory Infections, Institut Pasteur, Paris Mélanie Albert, Marion Barbet, Sylvie Behillil, Méline Bizard, Angela Brisebarre, Flora Donati, Etienne Simon-Lorière, Vincent Enouf, Maud Vanpeene, Sylvie van der Werf
EPI_ISL_428357 hCoV-19/France/IDF2253/2020 Europe / France / Ile De France / Garches 2020-03-11 Institut Médico légal- Hop R. Poincaré National Reference Center for Viruses of Respiratory Infections, Institut Pasteur, Paris Mélanie Albert, Marion Barbet, Sylvie Behillil, Méline Bizard, Angela Brisebarre, Flora Donati, Etienne Simon-Lorière, Vincent Enouf, Maud Vanpeene, Sylvie van der Werf
EPI_ISL_428358 hCoV-19/France/HF3598/2020 Europe / France / Hauts De France / Chateau-thierry 2020-03-24 CH Jeanne de Navarre Laboratoire de Biologie National Reference Center for Viruses of Respiratory Infections, Institut Pasteur, Paris Mélanie Albert, Marion Barbet, Sylvie Behillil, Méline Bizard, Angela Brisebarre, Flora Donati, Etienne Simon-Lorière, Vincent Enouf, Maud Vanpeene, Sylvie van der Werf
EPI_ISL_428359 hCoV-19/France/HF3677/2020 Europe / France / Hauts De France / Compiegne 2020-03-23 CH Compiègne Laboratoire de Biologie National Reference Center for Viruses of Respiratory Infections, Institut Pasteur, Paris Mélanie Albert, Marion Barbet, Sylvie Behillil, Méline Bizard, Angela Brisebarre, Flora Donati, Etienne Simon-Lorière, Vincent Enouf, Maud Vanpeene, Sylvie van der Werf
EPI_ISL_428360 hCoV-19/France/HF3678/2020 Europe / France / Hauts De France / Compiegne 2020-03-25 CH Compiègne Laboratoire de Biologie National Reference Center for Viruses of Respiratory Infections, Institut Pasteur, Paris Mélanie Albert, Marion Barbet, Sylvie Behillil, Méline Bizard, Angela Brisebarre, Flora Donati, Etienne Simon-Lorière, Vincent Enouf, Maud Vanpeene, Sylvie van der Werf
EPI_ISL_428361 hCoV-19/France/IDF3703/2020 Europe / France / Ile De France / Longjumeau 2020-03-25 LABM GH nord Essonne de Longjumeau - BP 125 National Reference Center for Viruses of Respiratory Infections, Institut Pasteur, Paris Mélanie Albert, Marion Barbet, Sylvie Behillil, Méline Bizard, Angela Brisebarre, Flora Donati, Etienne Simon-Lorière, Vincent Enouf, Maud Vanpeene, Sylvie van der Werf
EPI_ISL_428363 hCoV-19/France/IDF3745/2020 Europe / France / Ile De France / Orsay 2020-03-26 GH Nord Essonne Service de Biologie clinique National Reference Center for Viruses of Respiratory Infections, Institut Pasteur, Paris Mélanie Albert, Marion Barbet, Sylvie Behillil, Méline Bizard, Angela Brisebarre, Flora Donati, Etienne Simon-Lorière, Vincent Enouf, Maud Vanpeene, Sylvie van der Werf
EPI_ISL_428365 hCoV-19/France/IDF3930/2020 Europe / France / Ile De France / Longjumeau 2020-03-26 LABM GH nord Essonne de Longjumeau - BP 125 National Reference Center for Viruses of Respiratory Infections, Institut Pasteur, Paris Mélanie Albert, Marion Barbet, Sylvie Behillil, Méline Bizard, Angela Brisebarre, Flora Donati, Etienne Simon-Lorière, Vincent Enouf, Maud Vanpeene, Sylvie van der Werf
EPI_ISL_428366 hCoV-19/France/HF4220/2020 Europe / France / Hauts De France / Chateau-thierry 2020-03-30 CH Jeanne de Navarre Laboratoire de Biologie National Reference Center for Viruses of Respiratory Infections, Institut Pasteur, Paris Mélanie Albert, Marion Barbet, Sylvie Behillil, Méline Bizard, Angela Brisebarre, Flora Donati, Etienne Simon-Lorière, Vincent Enouf, Maud Vanpeene, Sylvie van der Werf
EPI_ISL_428367 hCoV-19/France/N4427/2020 Europe / France / Normandie / Carentan 2020-03-23 Cabinet Médical National Reference Center for Viruses of Respiratory Infections, Institut Pasteur, Paris Mélanie Albert, Marion Barbet, Sylvie Behillil, Méline Bizard, Angela Brisebarre, Flora Donati, Etienne Simon-Lorière, Vincent Enouf, Maud Vanpeene, Sylvie van der Werf

EPI_ISL_428368 hCoV-19/Turkey/GLAB-CoV013/2020 Europe / Turkey / Istanbul 2020-04-16 Genomic Laboratory (GLAB) (Conjoint lab of Health Directorate of Istanbul and Istanbul Technical University) Genomic Laboratory (GLAB), Istanbul Technical University Ilker Karacan, Tugba Kizilboga Akgun, Bugra Agaoglu, Gizem Alkurt, Jale Yildiz, Betsi Köse, Elifnaz Çelik, Arzu Irvem, Yasemin Kendir Demirkol, Ozlem Akgun Dogan, Mehtap Aydın, Levent Doganay, Gizem Dinler Doganay
EPI_ISL_428385 hCoV-19/USA/CT-Yale-066/2020 North America / USA / Connecticut 2020-04-04 Yale COVID-19 Biorepository Grubaugh Lab - Yale School of Public Health Joseph Fauver, Tara Alpert, Anderson Brito, Anne Wyllie, Chantal Vogels, Mary Petrone, Chaney Kalinich, Isabel Ott, Arnau Casanovas, Catherine Muenker, Adam Moore, Alice Lu, Maria Tokuyama, Patrick Wong, Peiwen Lu, Saad Omer, Richard Martinello, Allison Nelson, Shelli Farhadian, Akiko Iwasaki, Charlese Dela Cruz, Albert Ko, Nathan Grubaugh
EPI_ISL_429350 hCoV-19/Denmark/ALAB-SSI121/2020 Europe / Denmark / Unknown 2020-03-08 Department of Virus and Microbiological Special Diagnostics, Statens Serum Institut, Copenhagen, Denmark, Artillerivej 5, 2300 Copenahgen S Albertsen lab, Department of Chemistry and Bioscience, Aalborg University, Denmark Rasmus Kirkegaard
EPI_ISL_429351 hCoV-19/Denmark/ALAB-SSI122/2020 Europe / Denmark / Unknown 2020-03-08 Department of Virus and Microbiological Special Diagnostics, Statens Serum Institut, Copenhagen, Denmark, Artillerivej 5, 2300 Copenahgen S Albertsen lab, Department of Chemistry and Bioscience, Aalborg University, Denmark Rasmus Kirkegaard
EPI_ISL_429352 hCoV-19/Denmark/ALAB-SSI123/2020 Europe / Denmark / Unknown 2020-03-08 Department of Virus and Microbiological Special Diagnostics, Statens Serum Institut, Copenhagen, Denmark, Artillerivej 5, 2300 Copenahgen S Albertsen lab, Department of Chemistry and Bioscience, Aalborg University, Denmark Rasmus Kirkegaard
EPI_ISL_429354 hCoV-19/Denmark/ALAB-SSI126/2020 Europe / Denmark / Unknown 2020-03-08 Department of Virus and Microbiological Special Diagnostics, Statens Serum Institut, Copenhagen, Denmark, Artillerivej 5, 2300 Copenahgen S Albertsen lab, Department of Chemistry and Bioscience, Aalborg University, Denmark Rasmus Kirkegaard
EPI_ISL_429370 hCoV-19/Denmark/ALAB-SSI154/2020 Europe / Denmark / Unknown 2020-03-09 Department of Virus and Microbiological Special Diagnostics, Statens Serum Institut, Copenhagen, Denmark, Artillerivej 5, 2300 Copenahgen S Albertsen lab, Department of Chemistry and Bioscience, Aalborg University, Denmark Rasmus Kirkegaard
EPI_ISL_429371 hCoV-19/Denmark/ALAB-SSI155/2020 Europe / Denmark / Unknown 2020-03-10 Department of Virus and Microbiological Special Diagnostics, Statens Serum Institut, Copenhagen, Denmark, Artillerivej 5, 2300 Copenahgen S Albertsen lab, Department of Chemistry and Bioscience, Aalborg University, Denmark Rasmus Kirkegaard
EPI_ISL_429372 hCoV-19/Denmark/ALAB-SSI156/2020 Europe / Denmark / Unknown 2020-03-10 Department of Virus and Microbiological Special Diagnostics, Statens Serum Institut, Copenhagen, Denmark, Artillerivej 5, 2300 Copenahgen S Albertsen lab, Department of Chemistry and Bioscience, Aalborg University, Denmark Rasmus Kirkegaard
EPI_ISL_429373 hCoV-19/Denmark/ALAB-SSI157/2020 Europe / Denmark / Unknown 2020-03-09 Department of Virus and Microbiological Special Diagnostics, Statens Serum Institut, Copenhagen, Denmark, Artillerivej 5, 2300 Copenahgen S Albertsen lab, Department of Chemistry and Bioscience, Aalborg University, Denmark Rasmus Kirkegaard
EPI_ISL_429374 hCoV-19/Denmark/ALAB-SSI158/2020 Europe / Denmark / Unknown 2020-03-09 Department of Virus and Microbiological Special Diagnostics, Statens Serum Institut, Copenhagen, Denmark, Artillerivej 5, 2300 Copenahgen S Albertsen lab, Department of Chemistry and Bioscience, Aalborg University, Denmark Rasmus Kirkegaard
EPI_ISL_429375 hCoV-19/Denmark/ALAB-SSI159/2020 Europe / Denmark / Unknown 2020-03-09 Department of Virus and Microbiological Special Diagnostics, Statens Serum Institut, Copenhagen, Denmark, Artillerivej 5, 2300 Copenahgen S Albertsen lab, Department of Chemistry and Bioscience, Aalborg University, Denmark Rasmus Kirkegaard
EPI_ISL_429376 hCoV-19/Denmark/ALAB-SSI160/2020 Europe / Denmark / Unknown 2020-03-09 Department of Virus and Microbiological Special Diagnostics, Statens Serum Institut, Copenhagen, Denmark, Artillerivej 5, 2300 Copenahgen S Albertsen lab, Department of Chemistry and Bioscience, Aalborg University, Denmark Rasmus Kirkegaard
EPI_ISL_429377 hCoV-19/Denmark/ALAB-SSI162/2020 Europe / Denmark / Unknown 2020-03-09 Department of Virus and Microbiological Special Diagnostics, Statens Serum Institut, Copenhagen, Denmark, Artillerivej 5, 2300 Copenahgen S Albertsen lab, Department of Chemistry and Bioscience, Aalborg University, Denmark Rasmus Kirkegaard
EPI_ISL_429378 hCoV-19/Denmark/ALAB-SSI163/2020 Europe / Denmark / Unknown 2020-03-09 Department of Virus and Microbiological Special Diagnostics, Statens Serum Institut, Copenhagen, Denmark, Artillerivej 5, 2300 Copenahgen S Albertsen lab, Department of Chemistry and Bioscience, Aalborg University, Denmark Rasmus Kirkegaard
EPI_ISL_429379 hCoV-19/Denmark/ALAB-SSI164/2020 Europe / Denmark / Unknown 2020-03-09 Department of Virus and Microbiological Special Diagnostics, Statens Serum Institut, Copenhagen, Denmark, Artillerivej 5, 2300 Copenahgen S Albertsen lab, Department of Chemistry and Bioscience, Aalborg University, Denmark Rasmus Kirkegaard
EPI_ISL_429381 hCoV-19/Denmark/ALAB-SSI167/2020 Europe / Denmark / Unknown 2020-03-09 Department of Virus and Microbiological Special Diagnostics, Statens Serum Institut, Copenhagen, Denmark, Artillerivej 5, 2300 Copenahgen S Albertsen lab, Department of Chemistry and Bioscience, Aalborg University, Denmark Rasmus Kirkegaard

EPI_ISL_429382  hCoV-19/Denmark/ALAB-SSI168/2020    Europe / Denmark / Unknown    2020-03-09
Department of Virus and Microbiological Special Diagnostics, Statens Serum Institut, Copenhagen, Denmark, Artillerivej 5, 2300 Copenahgen S    Albertsen lab, Department of Chemistry and Bioscience, Aalborg University, Denmark    Rasmus Kirkegaard
EPI_ISL_429383  hCoV-19/Denmark/ALAB-SSI169/2020    Europe / Denmark / Unknown    2020-03-09
Department of Virus and Microbiological Special Diagnostics, Statens Serum Institut, Copenhagen, Denmark, Artillerivej 5, 2300 Copenahgen S    Albertsen lab, Department of Chemistry and Bioscience, Aalborg University, Denmark    Rasmus Kirkegaard
EPI_ISL_429384  hCoV-19/Denmark/ALAB-SSI171/2020    Europe / Denmark / Unknown    2020-03-09
Department of Virus and Microbiological Special Diagnostics, Statens Serum Institut, Copenhagen, Denmark, Artillerivej 5, 2300 Copenahgen S    Albertsen lab, Department of Chemistry and Bioscience, Aalborg University, Denmark    Rasmus Kirkegaard
EPI_ISL_429385  hCoV-19/Denmark/ALAB-SSI172/2020    Europe / Denmark / Unknown    2020-03-09
Department of Virus and Microbiological Special Diagnostics, Statens Serum Institut, Copenhagen, Denmark, Artillerivej 5, 2300 Copenahgen S    Albertsen lab, Department of Chemistry and Bioscience, Aalborg University, Denmark    Rasmus Kirkegaard
EPI_ISL_429386  hCoV-19/Denmark/ALAB-SSI173/2020    Europe / Denmark / Unknown    2020-03-09
Department of Virus and Microbiological Special Diagnostics, Statens Serum Institut, Copenhagen, Denmark, Artillerivej 5, 2300 Copenahgen S    Albertsen lab, Department of Chemistry and Bioscience, Aalborg University, Denmark    Rasmus Kirkegaard
EPI_ISL_429387  hCoV-19/Denmark/ALAB-SSI174/2020    Europe / Denmark / Unknown    2020-03-09
Department of Virus and Microbiological Special Diagnostics, Statens Serum Institut, Copenhagen, Denmark, Artillerivej 5, 2300 Copenahgen S    Albertsen lab, Department of Chemistry and Bioscience, Aalborg University, Denmark    Rasmus Kirkegaard
EPI_ISL_429501  hCoV-19/Denmark/ALAB-SSI407/2020    Europe / Denmark / Unknown    2020-03-10
Department of Virus and Microbiological Special Diagnostics, Statens Serum Institut, Copenhagen, Denmark, Artillerivej 5, 2300 Copenahgen S    Albertsen lab, Department of Chemistry and Bioscience, Aalborg University, Denmark    Rasmus Kirkegaard
EPI_ISL_429502  hCoV-19/Denmark/ALAB-SSI408/2020    Europe / Denmark / Unknown    2020-03-10
Department of Virus and Microbiological Special Diagnostics, Statens Serum Institut, Copenhagen, Denmark, Artillerivej 5, 2300 Copenahgen S    Albertsen lab, Department of Chemistry and Bioscience, Aalborg University, Denmark    Rasmus Kirkegaard
EPI_ISL_429503  hCoV-19/Denmark/ALAB-SSI409/2020    Europe / Denmark / Unknown    2020-03-10
Department of Virus and Microbiological Special Diagnostics, Statens Serum Institut, Copenhagen, Denmark, Artillerivej 5, 2300 Copenahgen S    Albertsen lab, Department of Chemistry and Bioscience, Aalborg University, Denmark    Rasmus Kirkegaard
EPI_ISL_429505  hCoV-19/Denmark/ALAB-SSI411/2020    Europe / Denmark / Unknown    2020-03-10
Department of Virus and Microbiological Special Diagnostics, Statens Serum Institut, Copenhagen, Denmark, Artillerivej 5, 2300 Copenahgen S    Albertsen lab, Department of Chemistry and Bioscience, Aalborg University, Denmark    Rasmus Kirkegaard
EPI_ISL_429506  hCoV-19/Denmark/ALAB-SSI411A/2020    Europe / Denmark / Unknown    2020-03-23
Department of Virus and Microbiological Special Diagnostics, Statens Serum Institut, Copenhagen, Denmark, Artillerivej 5, 2300 Copenahgen S    Albertsen lab, Department of Chemistry and Bioscience, Aalborg University, Denmark    Rasmus Kirkegaard
EPI_ISL_429507  hCoV-19/Denmark/ALAB-SSI412/2020    Europe / Denmark / Unknown    2020-03-10
Department of Virus and Microbiological Special Diagnostics, Statens Serum Institut, Copenhagen, Denmark, Artillerivej 5, 2300 Copenahgen S    Albertsen lab, Department of Chemistry and Bioscience, Aalborg University, Denmark    Rasmus Kirkegaard
EPI_ISL_429600  hCoV-19/USA/WA-UW-4549/2020    North America / USA / Washington    2020-03-31    UW Virology Lab    UW Virology Lab Pavitra Roychoudhury, Hong Xie, Keith Jerome, Alexander Greninger
EPI_ISL_429601  hCoV-19/USA/ID-UW-4552/2020    North America / USA / Idaho    2020-03-29    UW Virology Lab    UW Virology Lab Pavitra Roychoudhury, Hong Xie, Keith Jerome, Alexander Greninger
EPI_ISL_429602  hCoV-19/USA/WA-UW-4564/2020    North America / USA / Washington    2020-03-30    UW Virology Lab    UW Virology Lab Pavitra Roychoudhury, Hong Xie, Keith Jerome, Alexander Greninger
EPI_ISL_429603  hCoV-19/USA/WA-UW-4565/2020    North America / USA / Washington    2020-03-30    UW Virology Lab    UW Virology Lab Pavitra Roychoudhury, Hong Xie, Keith Jerome, Alexander Greninger
EPI_ISL_429604  hCoV-19/USA/WA-UW-4568/2020    North America / USA / Washington    2020-03-29    UW Virology Lab    UW Virology Lab Pavitra Roychoudhury, Hong Xie, Keith Jerome, Alexander Greninger
EPI_ISL_429605  hCoV-19/USA/WA-UW-4578/2020    North America / USA / Washington    2020-03-31    UW Virology Lab    UW Virology Lab Pavitra Roychoudhury, Hong Xie, Keith Jerome, Alexander Greninger
EPI_ISL_429606  hCoV-19/USA/WA-UW-4579/2020    North America / USA / Washington    2020-03-31    UW Virology Lab    UW Virology Lab Pavitra Roychoudhury, Hong Xie, Keith Jerome, Alexander Greninger
EPI_ISL_429608  hCoV-19/USA/WA-UW-4593/2020    North America / USA / Washington    2020-03-31    UW Virology Lab    UW Virology Lab Pavitra Roychoudhury, Hong Xie, Keith Jerome, Alexander Greninger
EPI_ISL_429609  hCoV-19/USA/WA-UW-4594/2020    North America / USA / Washington    2020-03-31    UW Virology Lab    UW Virology Lab Pavitra Roychoudhury, Hong Xie, Keith Jerome, Alexander Greninger
EPI_ISL_429610  hCoV-19/USA/ID-UW-4598/2020    North America / USA / Idaho    2020-03-30    UW Virology Lab    UW Virology Lab Pavitra Roychoudhury, Hong Xie, Keith Jerome, Alexander Greninger
EPI_ISL_429611  hCoV-19/USA/WA-UW-4602/2020    North America / USA / Washington    2020-03-31    UW Virology Lab    UW Virology Lab Pavitra Roychoudhury, Hong Xie, Keith Jerome, Alexander Greninger

EPI_ISL_429612 hCoV-19/USA/WA-UW-4609/2020 North America / USA / Washington 2020-03-26 UW Virology Lab UW Virology Lab Pavitra Roychoudhury, Hong Xie, Keith Jerome, Alexander Greninger
EPI_ISL_429613 hCoV-19/USA/WA-UW-4611/2020 North America / USA / Washington 2020-03-26 UW Virology Lab UW Virology Lab Pavitra Roychoudhury, Hong Xie, Keith Jerome, Alexander Greninger
EPI_ISL_429614 hCoV-19/USA/WA-UW-4612/2020 North America / USA / Washington 2020-03-26 UW Virology Lab UW Virology Lab Pavitra Roychoudhury, Hong Xie, Keith Jerome, Alexander Greninger
EPI_ISL_429615 hCoV-19/USA/WA-UW-4703/2020 North America / USA / Washington 2020-03-29 UW Virology Lab UW Virology Lab Pavitra Roychoudhury, Hong Xie, Keith Jerome, Alexander Greninger
EPI_ISL_429616 hCoV-19/USA/WA-UW-4732/2020 North America / USA / Washington 2020-03-29 UW Virology Lab UW Virology Lab Pavitra Roychoudhury, Hong Xie, Keith Jerome, Alexander Greninger
EPI_ISL_429617 hCoV-19/USA/WA-UW-4734/2020 North America / USA / Washington 2020-03-28 UW Virology Lab UW Virology Lab Pavitra Roychoudhury, Hong Xie, Keith Jerome, Alexander Greninger
EPI_ISL_429618 hCoV-19/USA/WA-UW-4741/2020 North America / USA / Washington 2020-03-25 UW Virology Lab UW Virology Lab Pavitra Roychoudhury, Hong Xie, Keith Jerome, Alexander Greninger
EPI_ISL_429621 hCoV-19/USA/WA-UW-4760/2020 North America / USA / Washington 2020-03-28 UW Virology Lab UW Virology Lab Pavitra Roychoudhury, Hong Xie, Keith Jerome, Alexander Greninger
EPI_ISL_429622 hCoV-19/USA/WA-UW-4763/2020 North America / USA / Washington 2020-03-27 UW Virology Lab UW Virology Lab Pavitra Roychoudhury, Hong Xie, Keith Jerome, Alexander Greninger
EPI_ISL_429623 hCoV-19/USA/WA-UW-4766/2020 North America / USA / Washington 2020-03-28 UW Virology Lab UW Virology Lab Pavitra Roychoudhury, Hong Xie, Keith Jerome, Alexander Greninger
EPI_ISL_429624 hCoV-19/USA/WA-UW-4769/2020 North America / USA / Washington 2020-03-27 UW Virology Lab UW Virology Lab Pavitra Roychoudhury, Hong Xie, Keith Jerome, Alexander Greninger
EPI_ISL_429625 hCoV-19/USA/WA-UW-4770/2020 North America / USA / Washington 2020-03-27 UW Virology Lab UW Virology Lab Pavitra Roychoudhury, Hong Xie, Keith Jerome, Alexander Greninger
EPI_ISL_429626 hCoV-19/USA/WA-UW-4775/2020 North America / USA / Washington 2020-03-28 UW Virology Lab UW Virology Lab Pavitra Roychoudhury, Hong Xie, Keith Jerome, Alexander Greninger
EPI_ISL_429627 hCoV-19/USA/WA-UW-4777/2020 North America / USA / Washington 2020-03-27 UW Virology Lab UW Virology Lab Pavitra Roychoudhury, Hong Xie, Keith Jerome, Alexander Greninger
EPI_ISL_429628 hCoV-19/USA/UN-UW-4645/2020 North America / USA 2020-04-06 UW Virology Lab UW Virology Lab Pavitra Roychoudhury, Hong Xie, Keith Jerome, Alexander Greninger
EPI_ISL_429629 hCoV-19/USA/WA-UW-4649/2020 North America / USA / Washington 2020-04-06 UW Virology Lab UW Virology Lab Pavitra Roychoudhury, Hong Xie, Keith Jerome, Alexander Greninger
EPI_ISL_429630 hCoV-19/USA/WA-UW-4677/2020 North America / USA / Washington 2020-04-05 UW Virology Lab UW Virology Lab Pavitra Roychoudhury, Hong Xie, Keith Jerome, Alexander Greninger
EPI_ISL_429631 hCoV-19/USA/WA-UW-4695/2020 North America / USA / Washington 2020-04-06 UW Virology Lab UW Virology Lab Pavitra Roychoudhury, Hong Xie, Keith Jerome, Alexander Greninger
EPI_ISL_429632 hCoV-19/USA/WA-UW-4705/2020 North America / USA / Washington 2020-04-06 UW Virology Lab UW Virology Lab Pavitra Roychoudhury, Hong Xie, Keith Jerome, Alexander Greninger
EPI_ISL_429634 hCoV-19/USA/WA-UW-4892/2020 North America / USA / Washington 2020-04-07 UW Virology Lab UW Virology Lab Pavitra Roychoudhury, Hong Xie, Keith Jerome, Alexander Greninger
EPI_ISL_429635 hCoV-19/USA/WA-UW-4897/2020 North America / USA / Washington 2020-04-08 UW Virology Lab UW Virology Lab Pavitra Roychoudhury, Hong Xie, Keith Jerome, Alexander Greninger
EPI_ISL_429636 hCoV-19/USA/WA-UW-4921/2020 North America / USA / Washington 2020-04-07 UW Virology Lab UW Virology Lab Pavitra Roychoudhury, Hong Xie, Keith Jerome, Alexander Greninger
EPI_ISL_429637 hCoV-19/USA/WA-UW-4925/2020 North America / USA / Washington 2020-04-07 UW Virology Lab UW Virology Lab Pavitra Roychoudhury, Hong Xie, Keith Jerome, Alexander Greninger
EPI_ISL_429638 hCoV-19/USA/WA-UW-4999/2020 North America / USA / Washington 2020-04-07 UW Virology Lab UW Virology Lab Pavitra Roychoudhury, Hong Xie, Keith Jerome, Alexander Greninger
EPI_ISL_429639 hCoV-19/USA/WA-UW-5025/2020 North America / USA / Washington 2020-04-07 UW Virology Lab UW Virology Lab Pavitra Roychoudhury, Hong Xie, Keith Jerome, Alexander Greninger
EPI_ISL_429641 hCoV-19/USA/CT-UW-5038/2020 North America / USA / Connecticut 2020-04-06 UW Virology Lab UW Virology Lab Pavitra Roychoudhury, Hong Xie, Keith Jerome, Alexander Greninger
EPI_ISL_429642 hCoV-19/USA/CT-UW-5043/2020 North America / USA / Connecticut 2020-04-06 UW Virology Lab UW Virology Lab Pavitra Roychoudhury, Hong Xie, Keith Jerome, Alexander Greninger
EPI_ISL_429643 hCoV-19/USA/UN-UW-5044/2020 North America / USA 2020-04-08 UW Virology Lab UW Virology Lab Pavitra Roychoudhury, Hong Xie, Keith Jerome, Alexander Greninger
EPI_ISL_429644 hCoV-19/USA/WA-UW-5065/2020 North America / USA / Washington 2020-04-08 UW Virology Lab UW Virology Lab Pavitra Roychoudhury, Hong Xie, Keith Jerome, Alexander Greninger
EPI_ISL_429645 hCoV-19/USA/CT-UW-5073/2020 North America / USA / Connecticut 2020-04-06 UW Virology Lab UW Virology Lab Pavitra Roychoudhury, Hong Xie, Keith Jerome, Alexander Greninger
EPI_ISL_429646 hCoV-19/USA/CT-UW-5086/2020 North America / USA / Connecticut 2020-04-06 UW Virology Lab UW Virology Lab Pavitra Roychoudhury, Hong Xie, Keith Jerome, Alexander Greninger
EPI_ISL_429647 hCoV-19/USA/WA-UW-5095/2020 North America / USA / Washington 2020-04-08 UW Virology Lab UW Virology Lab Pavitra Roychoudhury, Hong Xie, Keith Jerome, Alexander Greninger
EPI_ISL_429649 hCoV-19/USA/WA-UW-5144/2020 North America / USA / Washington 2020-04-02 UW Virology Lab UW Virology Lab Pavitra Roychoudhury, Hong Xie, Keith Jerome, Alexander Greninger
EPI_ISL_429650 hCoV-19/USA/WA-UW-5152/2020 North America / USA / Washington 2020-04-02 UW Virology Lab UW Virology Lab Pavitra Roychoudhury, Hong Xie, Keith Jerome, Alexander Greninger
EPI_ISL_429651 hCoV-19/USA/WA-UW-5158/2020 North America / USA / Washington 2020-04-02 UW Virology Lab UW Virology Lab Pavitra Roychoudhury, Hong Xie, Keith Jerome, Alexander Greninger

EPI_ISL_429652 hCoV-19/USA/WA-UW-5164/2020 North America / USA / Washington 2020-04-02 UW Virology Lab UW Virology Lab Pavitra Roychoudhury, Hong Xie, Keith Jerome, Alexander Greninger
EPI_ISL_429653 hCoV-19/USA/WA-UW-5166/2020 North America / USA / Washington 2020-04-02 UW Virology Lab UW Virology Lab Pavitra Roychoudhury, Hong Xie, Keith Jerome, Alexander Greninger
EPI_ISL_429654 hCoV-19/USA/WA-UW-5169/2020 North America / USA / Washington 2020-04-02 UW Virology Lab UW Virology Lab Pavitra Roychoudhury, Hong Xie, Keith Jerome, Alexander Greninger
EPI_ISL_429655 hCoV-19/USA/WA-UW-5170/2020 North America / USA / Washington 2020-04-02 UW Virology Lab UW Virology Lab Pavitra Roychoudhury, Hong Xie, Keith Jerome, Alexander Greninger
EPI_ISL_429656 hCoV-19/USA/UN-UW-5172/2020 North America / USA 2020-04-02 UW Virology Lab UW Virology Lab Pavitra Roychoudhury, Hong Xie, Keith Jerome, Alexander Greninger
EPI_ISL_429667 hCoV-19/Brazil/CV4/2020 South America / Brazil / Minas gerais 2020-03-09 Central Public Health Laboratory/Octávio Magalhães Institute (IOM) from the Ezequiel Dias Foundation (FUNED) Instituto Octávio Magalhães / Fundação Ezequiel Dias (IOM/Funed) Talita Adelino, Joilson Xavier, Marta Giovanetti, Vagner Fonseca, Marcos Vinícius Silva, Luiz Carlos Junior Alcantara, Marluce Aparecida Assunção Oliveira
EPI_ISL_429669 hCoV-19/Brazil/CV6/2020 South America / Brazil / Minas gerais 2020-03-13 Central Public Health Laboratory/Octávio Magalhães Institute (IOM) from the Ezequiel Dias Foundation (FUNED) Instituto Octávio Magalhães / Fundação Ezequiel Dias (IOM/Funed) Talita Adelino, Joilson Xavier, Marta Giovanetti, Vagner Fonseca, Marcos Vinícius Silva, Luiz Carlos Junior Alcantara, Marluce Aparecida Assunção Oliveira
EPI_ISL_429676 hCoV-19/Brazil/CV16/2020 South America / Brazil / Minas gerais 2020-03-16 Central Public Health Laboratory/Octávio Magalhães Institute (IOM) from the Ezequiel Dias Foundation (FUNED) Instituto Octávio Magalhães / Fundação Ezequiel Dias (IOM/Funed) Talita Adelino, Joilson Xavier, Marta Giovanetti, Vagner Fonseca, Marcos Vinícius Silva, Luiz Carlos Junior Alcantara, Marluce Aparecida Assunção Oliveira
EPI_ISL_429681 hCoV-19/Brazil/CV21/2020 South America / Brazil / Minas gerais 2020-03-16 Central Public Health Laboratory/Octávio Magalhães Institute (IOM) from the Ezequiel Dias Foundation (FUNED) Instituto Octávio Magalhães / Fundação Ezequiel Dias (IOM/Funed) Talita Adelino, Joilson Xavier, Marta Giovanetti, Vagner Fonseca, Marcos Vinícius Silva, Luiz Carlos Junior Alcantara, Marluce Aparecida Assunção Oliveira
EPI_ISL_429687 hCoV-19/Brazil/CV31/2020 South America / Brazil / Minas gerais 2020-03-17 Central Public Health Laboratory/Octávio Magalhães Institute (IOM) from the Ezequiel Dias Foundation (FUNED) Instituto Octávio Magalhães / Fundação Ezequiel Dias (IOM/Funed) Talita Adelino, Joilson Xavier, Marta Giovanetti, Vagner Fonseca, Marcos Vinícius Silva, Luiz Carlos Junior Alcantara, Marluce Aparecida Assunção Oliveira
EPI_ISL_429688 hCoV-19/Brazil/CV32/2020 South America / Brazil / Minas gerais 2020-03-17 Central Public Health Laboratory/Octávio Magalhães Institute (IOM) from the Ezequiel Dias Foundation (FUNED) Instituto Octávio Magalhães / Fundação Ezequiel Dias (IOM/Funed) Talita Adelino, Joilson Xavier, Marta Giovanetti, Vagner Fonseca, Marcos Vinícius Silva, Luiz Carlos Junior Alcantara, Marluce Aparecida Assunção Oliveira
EPI_ISL_429695 hCoV-19/Brazil/CV42/2020 South America / Brazil / Minas gerais 2020-03-20 Central Public Health Laboratory/Octávio Magalhães Institute (IOM) from the Ezequiel Dias Foundation (FUNED) Instituto Octávio Magalhães / Fundação Ezequiel Dias (IOM/Funed) Talita Adelino, Joilson Xavier, Marta Giovanetti, Vagner Fonseca, Marcos Vinícius Silva, Luiz Carlos Junior Alcantara, Marluce Aparecida Assunção Oliveira
EPI_ISL_430333 hCoV-19/USA/NY-NYUMC306/2020 North America / USA / New York / Brooklyn 2020-03-30 NYU Langone Health Departments of Pathology and Medicine, New York University School of Medicine Maria Aguero-Rosenfeld, Brendan Belovarac, Margaret Black, Ludovic Boytard, John Cadley, Paolo Cotzia, John Chen, Dacia Dimartino, Xiaojun Feng, Tatyana Gindin, Emily Guzman, Adriana Heguy, Megan Hogan, Emily Huang, George Jour, Lawrence H. Lin, Raven Luther, Andrew Lytle, Christian Marier, Matthew T. Maurano, Mark J. Mulligan, Peter Meyn, Raquel Ordonez Ciriza, Iman Osman, Jared Pinnell, Vanessa Raabe, Sitharam Ramaswami, Amy Rapkiewicz, Andre M. Ribeiro-dos-Santos, Marie Samanovic-Golden, Antonio Serrano, Guomiao Shen, Matija Snuderl, Theodore Vougiouklakis, Nick Vulpescu, Gael Westby, Paul Zappile, Yutong Zhang
EPI_ISL_430334 hCoV-19/USA/NY-NYUMC307/2020 North America / USA / New York / Brooklyn 2020-03-30 NYU Langone Health Departments of Pathology and Medicine, New York University School of Medicine Maria Aguero-Rosenfeld, Brendan Belovarac, Margaret Black, Ludovic Boytard, John Cadley, Paolo Cotzia, John Chen, Dacia Dimartino, Xiaojun Feng, Tatyana Gindin, Emily Guzman, Adriana Heguy, Megan Hogan, Emily Huang, George Jour, Lawrence H. Lin, Raven Luther, Andrew Lytle, Christian Marier, Matthew T. Maurano, Mark J. Mulligan, Peter Meyn, Raquel Ordonez Ciriza, Iman Osman, Jared Pinnell, Vanessa Raabe, Sitharam Ramaswami, Amy Rapkiewicz, Andre M. Ribeiro-dos-Santos, Marie Samanovic-Golden, Antonio Serrano, Guomiao Shen, Matija Snuderl, Theodore Vougiouklakis, Nick Vulpescu, Gael Westby, Paul Zappile, Yutong Zhang
EPI_ISL_430335 hCoV-19/USA/NY-NYUMC308/2020 North America / USA / New York / Brooklyn 2020-03-30 NYU Langone Health Departments of Pathology and Medicine, New York University School of Medicine Maria Aguero-Rosenfeld, Brendan Belovarac, Margaret Black, Ludovic Boytard, John Cadley, Paolo Cotzia, John Chen, Dacia Dimartino, Xiaojun Feng, Tatyana Gindin, Emily Guzman, Adriana Heguy, Megan Hogan, Emily Huang, George Jour, Lawrence H. Lin, Raven Luther, Andrew Lytle, Christian Marier, Matthew T. Maurano, Mark J. Mulligan, Peter Meyn, Raquel Ordonez Ciriza, Iman Osman, Jared Pinnell, Vanessa Raabe, Sitharam Ramaswami, Amy Rapkiewicz, Andre M. Ribeiro-dos-Santos, Marie Samanovic-Golden, Antonio Serrano, Guomiao Shen, Matija Snuderl, Theodore Vougiouklakis, Nick Vulpescu, Gael Westby, Paul Zappile, Yutong Zhang
EPI_ISL_430336 hCoV-19/USA/NY-NYUMC309/2020 North America / USA / New York / Manhattan 2020-03-30 NYU Langone Health Departments of Pathology and Medicine, New York University School of Medicine Maria Aguero-Rosenfeld, Brendan Belovarac, Margaret Black, Ludovic Boytard, John Cadley, Paolo Cotzia, John Chen,

Dacia Dimartino, Xiaojun Feng, Tatyana Gindin, Emily Guzman, Adriana Heguy, Megan Hogan, Emily Huang, George Jour, Lawrence H. Lin, Raven Luther, Andrew Lytle, Christian Marier, Matthew T. Maurano, Mark J. Mulligan, Peter Meyn, Raquel Ordonez Ciriza, Iman Osman, Jared Pinnell, Vanessa Raabe, Sitharam Ramaswami, Amy Rapkiewicz, Andre M. Ribeiro-dos-Santos, Marie Samanovic-Golden, Antonio Serrano, Guomiao Shen, Matija Snuderl, Theodore Vougiouklakis, Nick Vulpescu, Gael Westby, Paul Zappile, Yutong Zhang
EPI_ISL_430337 hCoV-19/USA/NY-NYUMC310/2020 North America / USA / New York / Manhattan 2020-03-30 NYU Langone Health Departments of Pathology and Medicine, New York University School of Medicine Maria Aguero-Rosenfeld, Brendan Belovarac, Margaret Black, Ludovic Boytard, John Cadley, Paolo Cotzia, John Chen, Dacia Dimartino, Xiaojun Feng, Tatyana Gindin, Emily Guzman, Adriana Heguy, Megan Hogan, Emily Huang, George Jour, Lawrence H. Lin, Raven Luther, Andrew Lytle, Christian Marier, Matthew T. Maurano, Mark J. Mulligan, Peter Meyn, Raquel Ordonez Ciriza, Iman Osman, Jared Pinnell, Vanessa Raabe, Sitharam Ramaswami, Amy Rapkiewicz, Andre M. Ribeiro-dos-Santos, Marie Samanovic-Golden, Antonio Serrano, Guomiao Shen, Matija Snuderl, Theodore Vougiouklakis, Nick Vulpescu, Gael Westby, Paul Zappile, Yutong Zhang
EPI_ISL_430338 hCoV-19/USA/NY-NYUMC311/2020 North America / USA / New York / Manhattan 2020-03-30 NYU Langone Health Departments of Pathology and Medicine, New York University School of Medicine Maria Aguero-Rosenfeld, Brendan Belovarac, Margaret Black, Ludovic Boytard, John Cadley, Paolo Cotzia, John Chen, Dacia Dimartino, Xiaojun Feng, Tatyana Gindin, Emily Guzman, Adriana Heguy, Megan Hogan, Emily Huang, George Jour, Lawrence H. Lin, Raven Luther, Andrew Lytle, Christian Marier, Matthew T. Maurano, Mark J. Mulligan, Peter Meyn, Raquel Ordonez Ciriza, Iman Osman, Jared Pinnell, Vanessa Raabe, Sitharam Ramaswami, Amy Rapkiewicz, Andre M. Ribeiro-dos-Santos, Marie Samanovic-Golden, Antonio Serrano, Guomiao Shen, Matija Snuderl, Theodore Vougiouklakis, Nick Vulpescu, Gael Westby, Paul Zappile, Yutong Zhang
EPI_ISL_430339 hCoV-19/USA/NY-NYUMC312/2020 North America / USA / New York / Manhattan 2020-03-30 NYU Langone Health Departments of Pathology and Medicine, New York University School of Medicine Maria Aguero-Rosenfeld, Brendan Belovarac, Margaret Black, Ludovic Boytard, John Cadley, Paolo Cotzia, John Chen, Dacia Dimartino, Xiaojun Feng, Tatyana Gindin, Emily Guzman, Adriana Heguy, Megan Hogan, Emily Huang, George Jour, Lawrence H. Lin, Raven Luther, Andrew Lytle, Christian Marier, Matthew T. Maurano, Mark J. Mulligan, Peter Meyn, Raquel Ordonez Ciriza, Iman Osman, Jared Pinnell, Vanessa Raabe, Sitharam Ramaswami, Amy Rapkiewicz, Andre M. Ribeiro-dos-Santos, Marie Samanovic-Golden, Antonio Serrano, Guomiao Shen, Matija Snuderl, Theodore Vougiouklakis, Nick Vulpescu, Gael Westby, Paul Zappile, Yutong Zhang
EPI_ISL_430343 hCoV-19/USA/NY-NYUMC316/2020 North America / USA / New York / Manhattan 2020-03-30 NYU Langone Health Departments of Pathology and Medicine, New York University School of Medicine Maria Aguero-Rosenfeld, Brendan Belovarac, Margaret Black, Ludovic Boytard, John Cadley, Paolo Cotzia, John Chen, Dacia Dimartino, Xiaojun Feng, Tatyana Gindin, Emily Guzman, Adriana Heguy, Megan Hogan, Emily Huang, George Jour, Lawrence H. Lin, Raven Luther, Andrew Lytle, Christian Marier, Matthew T. Maurano, Mark J. Mulligan, Peter Meyn, Raquel Ordonez Ciriza, Iman Osman, Jared Pinnell, Vanessa Raabe, Sitharam Ramaswami, Amy Rapkiewicz, Andre M. Ribeiro-dos-Santos, Marie Samanovic-Golden, Antonio Serrano, Guomiao Shen, Matija Snuderl, Theodore Vougiouklakis, Nick Vulpescu, Gael Westby, Paul Zappile, Yutong Zhang
EPI_ISL_430355 hCoV-19/USA/NY-NYUMC328/2020 North America / USA / New York / Nassau County 2020-04-02 NYU Langone Health Departments of Pathology and Medicine, New York University School of Medicine Maria Aguero-Rosenfeld, Brendan Belovarac, Margaret Black, Ludovic Boytard, John Cadley, Paolo Cotzia, John Chen, Dacia Dimartino, Xiaojun Feng, Tatyana Gindin, Emily Guzman, Adriana Heguy, Megan Hogan, Emily Huang, George Jour, Lawrence H. Lin, Raven Luther, Andrew Lytle, Christian Marier, Matthew T. Maurano, Mark J. Mulligan, Peter Meyn, Raquel Ordonez Ciriza, Iman Osman, Jared Pinnell, Vanessa Raabe, Sitharam Ramaswami, Amy Rapkiewicz, Andre M. Ribeiro-dos-Santos, Marie Samanovic-Golden, Antonio Serrano, Guomiao Shen, Matija Snuderl, Theodore Vougiouklakis, Nick Vulpescu, Gael Westby, Paul Zappile, Yutong Zhang
EPI_ISL_430356 hCoV-19/USA/NJ-NYUMC329/2020 North America / USA / New Jersey / Bergen County 2020-04-02 NYU Langone Health Departments of Pathology and Medicine, New York University School of Medicine Maria Aguero-Rosenfeld, Brendan Belovarac, Margaret Black, Ludovic Boytard, John Cadley, Paolo Cotzia, John Chen, Dacia Dimartino, Xiaojun Feng, Tatyana Gindin, Emily Guzman, Adriana Heguy, Megan Hogan, Emily Huang, George Jour, Lawrence H. Lin, Raven Luther, Andrew Lytle, Christian Marier, Matthew T. Maurano, Mark J. Mulligan, Peter Meyn, Raquel Ordonez Ciriza, Iman Osman, Jared Pinnell, Vanessa Raabe, Sitharam Ramaswami, Amy Rapkiewicz, Andre M. Ribeiro-dos-Santos, Marie Samanovic-Golden, Antonio Serrano, Guomiao Shen, Matija Snuderl, Theodore Vougiouklakis, Nick Vulpescu, Gael Westby, Paul Zappile, Yutong Zhang
EPI_ISL_430357 hCoV-19/USA/NY-NYUMC330/2020 North America / USA / New York / Brooklyn 2020-04-03 NYU Langone Health Departments of Pathology and Medicine, New York University School of Medicine Maria Aguero-Rosenfeld, Brendan Belovarac, Margaret Black, Ludovic Boytard, John Cadley, Paolo Cotzia, John Chen, Dacia Dimartino, Xiaojun Feng, Tatyana Gindin, Emily Guzman, Adriana Heguy, Megan Hogan, Emily Huang, George Jour, Lawrence H. Lin, Raven Luther, Andrew Lytle, Christian Marier, Matthew T. Maurano, Mark J. Mulligan, Peter Meyn, Raquel Ordonez Ciriza, Iman Osman, Jared Pinnell, Vanessa Raabe, Sitharam Ramaswami, Amy Rapkiewicz, Andre M. Ribeiro-dos-Santos, Marie Samanovic-Golden, Antonio Serrano, Guomiao Shen, Matija Snuderl, Theodore Vougiouklakis, Nick Vulpescu, Gael Westby, Paul Zappile, Yutong Zhang
EPI_ISL_430358 hCoV-19/USA/NY-NYUMC331/2020 North America / USA / New York / Manhattan 2020-04-13 NYU Langone Health Departments of Pathology and Medicine, New York University School of Medicine Maria Aguero-Rosenfeld, Brendan Belovarac, Margaret Black, Ludovic Boytard, John Cadley, Paolo Cotzia, John Chen, Dacia Dimartino, Xiaojun Feng, Tatyana Gindin, Emily Guzman, Adriana Heguy, Megan Hogan, Emily Huang, George Jour, Lawrence H. Lin, Raven Luther, Andrew Lytle, Christian Marier, Matthew T. Maurano, Mark J. Mulligan, Peter Meyn, Raquel Ordonez Ciriza, Iman Osman, Jared Pinnell, Vanessa Raabe, Sitharam Ramaswami, Amy Rapkiewicz, Andre M. Ribeiro-dos-Santos, Marie Samanovic-Golden, Antonio Serrano, Guomiao Shen, Matija Snuderl, Theodore Vougiouklakis, Nick Vulpescu, Gael Westby, Paul Zappile, Yutong Zhang
EPI_ISL_430359 hCoV-19/USA/NY-NYUMC332/2020 North America / USA / New York / Brooklyn 2020-04-08

NYU Langone Health	Departments of Pathology and Medicine, New York University School of Medicine	Maria Aguero-Rosenfeld, Brendan Belovarac, Margaret Black, Ludovic Boytard, John Cadley, Paolo Cotzia, John Chen, Dacia Dimartino, Xiaojun Feng, Tatyana Gindin, Emily Guzman, Adriana Heguy, Megan Hogan, Emily Huang, George Jour, Lawrence H. Lin, Raven Luther, Andrew Lytle, Christian Marier, Matthew T. Maurano, Mark J. Mulligan, Peter Meyn, Raquel Ordonez Ciriza, Iman Osman, Jared Pinnell, Vanessa Raabe, Sitharam Ramaswami, Amy Rapkiewicz, Andre M. Ribeiro-dos-Santos, Marie Samanovic-Golden, Antonio Serrano, Guomiao Shen, Matija Snuderl, Theodore Vougiouklakis, Nick Vulpescu, Gael Westby, Paul Zappile, Yutong Zhang
EPI_ISL_430360	hCoV-19/USA/NY-NYUMC333/2020	North America / USA / New York / Queens	2020-04-07	NYU Langone Health	Departments of Pathology and Medicine, New York University School of Medicine	Maria Aguero-Rosenfeld, Brendan Belovarac, Margaret Black, Ludovic Boytard, John Cadley, Paolo Cotzia, John Chen, Dacia Dimartino, Xiaojun Feng, Tatyana Gindin, Emily Guzman, Adriana Heguy, Megan Hogan, Emily Huang, George Jour, Lawrence H. Lin, Raven Luther, Andrew Lytle, Christian Marier, Matthew T. Maurano, Mark J. Mulligan, Peter Meyn, Raquel Ordonez Ciriza, Iman Osman, Jared Pinnell, Vanessa Raabe, Sitharam Ramaswami, Amy Rapkiewicz, Andre M. Ribeiro-dos-Santos, Marie Samanovic-Golden, Antonio Serrano, Guomiao Shen, Matija Snuderl, Theodore Vougiouklakis, Nick Vulpescu, Gael Westby, Paul Zappile, Yutong Zhang
EPI_ISL_430361	hCoV-19/USA/NY-NYUMC334/2020	North America / USA / New York / Brooklyn	2020-04-07	NYU Langone Health	Departments of Pathology and Medicine, New York University School of Medicine	Maria Aguero-Rosenfeld, Brendan Belovarac, Margaret Black, Ludovic Boytard, John Cadley, Paolo Cotzia, John Chen, Dacia Dimartino, Xiaojun Feng, Tatyana Gindin, Emily Guzman, Adriana Heguy, Megan Hogan, Emily Huang, George Jour, Lawrence H. Lin, Raven Luther, Andrew Lytle, Christian Marier, Matthew T. Maurano, Mark J. Mulligan, Peter Meyn, Raquel Ordonez Ciriza, Iman Osman, Jared Pinnell, Vanessa Raabe, Sitharam Ramaswami, Amy Rapkiewicz, Andre M. Ribeiro-dos-Santos, Marie Samanovic-Golden, Antonio Serrano, Guomiao Shen, Matija Snuderl, Theodore Vougiouklakis, Nick Vulpescu, Gael Westby, Paul Zappile, Yutong Zhang
EPI_ISL_430362	hCoV-19/USA/NY-NYUMC335/2020	North America / USA / New York / Brooklyn	2020-04-08	NYU Langone Health	Departments of Pathology and Medicine, New York University School of Medicine	Maria Aguero-Rosenfeld, Brendan Belovarac, Margaret Black, Ludovic Boytard, John Cadley, Paolo Cotzia, John Chen, Dacia Dimartino, Xiaojun Feng, Tatyana Gindin, Emily Guzman, Adriana Heguy, Megan Hogan, Emily Huang, George Jour, Lawrence H. Lin, Raven Luther, Andrew Lytle, Christian Marier, Matthew T. Maurano, Mark J. Mulligan, Peter Meyn, Raquel Ordonez Ciriza, Iman Osman, Jared Pinnell, Vanessa Raabe, Sitharam Ramaswami, Amy Rapkiewicz, Andre M. Ribeiro-dos-Santos, Marie Samanovic-Golden, Antonio Serrano, Guomiao Shen, Matija Snuderl, Theodore Vougiouklakis, Nick Vulpescu, Gael Westby, Paul Zappile, Yutong Zhang
EPI_ISL_430363	hCoV-19/USA/NY-NYUMC336/2020	North America / USA / New York / Brooklyn	2020-04-08	NYU Langone Health	Departments of Pathology and Medicine, New York University School of Medicine	Maria Aguero-Rosenfeld, Brendan Belovarac, Margaret Black, Ludovic Boytard, John Cadley, Paolo Cotzia, John Chen, Dacia Dimartino, Xiaojun Feng, Tatyana Gindin, Emily Guzman, Adriana Heguy, Megan Hogan, Emily Huang, George Jour, Lawrence H. Lin, Raven Luther, Andrew Lytle, Christian Marier, Matthew T. Maurano, Mark J. Mulligan, Peter Meyn, Raquel Ordonez Ciriza, Iman Osman, Jared Pinnell, Vanessa Raabe, Sitharam Ramaswami, Amy Rapkiewicz, Andre M. Ribeiro-dos-Santos, Marie Samanovic-Golden, Antonio Serrano, Guomiao Shen, Matija Snuderl, Theodore Vougiouklakis, Nick Vulpescu, Gael Westby, Paul Zappile, Yutong Zhang
EPI_ISL_430364	hCoV-19/USA/NY-NYUMC337/2020	North America / USA / New York / Brooklyn	2020-04-08	NYU Langone Health	Departments of Pathology and Medicine, New York University School of Medicine	Maria Aguero-Rosenfeld, Brendan Belovarac, Margaret Black, Ludovic Boytard, John Cadley, Paolo Cotzia, John Chen, Dacia Dimartino, Xiaojun Feng, Tatyana Gindin, Emily Guzman, Adriana Heguy, Megan Hogan, Emily Huang, George Jour, Lawrence H. Lin, Raven Luther, Andrew Lytle, Christian Marier, Matthew T. Maurano, Mark J. Mulligan, Peter Meyn, Raquel Ordonez Ciriza, Iman Osman, Jared Pinnell, Vanessa Raabe, Sitharam Ramaswami, Amy Rapkiewicz, Andre M. Ribeiro-dos-Santos, Marie Samanovic-Golden, Antonio Serrano, Guomiao Shen, Matija Snuderl, Theodore Vougiouklakis, Nick Vulpescu, Gael Westby, Paul Zappile, Yutong Zhang
EPI_ISL_430365	hCoV-19/USA/NY-NYUMC338/2020	North America / USA / New York / Brooklyn	2020-04-08	NYU Langone Health	Departments of Pathology and Medicine, New York University School of Medicine	Maria Aguero-Rosenfeld, Brendan Belovarac, Margaret Black, Ludovic Boytard, John Cadley, Paolo Cotzia, John Chen, Dacia Dimartino, Xiaojun Feng, Tatyana Gindin, Emily Guzman, Adriana Heguy, Megan Hogan, Emily Huang, George Jour, Lawrence H. Lin, Raven Luther, Andrew Lytle, Christian Marier, Matthew T. Maurano, Mark J. Mulligan, Peter Meyn, Raquel Ordonez Ciriza, Iman Osman, Jared Pinnell, Vanessa Raabe, Sitharam Ramaswami, Amy Rapkiewicz, Andre M. Ribeiro-dos-Santos, Marie Samanovic-Golden, Antonio Serrano, Guomiao Shen, Matija Snuderl, Theodore Vougiouklakis, Nick Vulpescu, Gael Westby, Paul Zappile, Yutong Zhang
EPI_ISL_430366	hCoV-19/USA/NY-NYUMC339/2020	North America / USA / New York / Brooklyn	2020-04-08	NYU Langone Health	Departments of Pathology and Medicine, New York University School of Medicine	Maria Aguero-Rosenfeld, Brendan Belovarac, Margaret Black, Ludovic Boytard, John Cadley, Paolo Cotzia, John Chen, Dacia Dimartino, Xiaojun Feng, Tatyana Gindin, Emily Guzman, Adriana Heguy, Megan Hogan, Emily Huang, George Jour, Lawrence H. Lin, Raven Luther, Andrew Lytle, Christian Marier, Matthew T. Maurano, Mark J. Mulligan, Peter Meyn, Raquel Ordonez Ciriza, Iman Osman, Jared Pinnell, Vanessa Raabe, Sitharam Ramaswami, Amy Rapkiewicz, Andre M. Ribeiro-dos-Santos, Marie Samanovic-Golden, Antonio Serrano, Guomiao Shen, Matija Snuderl, Theodore Vougiouklakis, Nick Vulpescu, Gael Westby, Paul Zappile, Yutong Zhang
EPI_ISL_430367	hCoV-19/USA/NY-NYUMC340/2020	North America / USA / New York / Brooklyn	2020-04-07	NYU Langone Health	Departments of Pathology and Medicine, New York University School of Medicine	Maria Aguero-Rosenfeld, Brendan Belovarac, Margaret Black, Ludovic Boytard, John Cadley, Paolo Cotzia, John Chen, Dacia Dimartino, Xiaojun Feng, Tatyana Gindin, Emily Guzman, Adriana Heguy, Megan Hogan, Emily Huang, George Jour, Lawrence H. Lin, Raven Luther, Andrew Lytle, Christian Marier, Matthew T. Maurano, Mark J. Mulligan, Peter Meyn, Raquel Ordonez Ciriza, Iman Osman, Jared Pinnell, Vanessa Raabe, Sitharam Ramaswami, Amy Rapkiewicz, Andre M. Ribeiro-dos-Santos, Marie Samanovic-Golden, Antonio Serrano, Guomiao Shen, Matija Snuderl,

Theodore Vougiouklakis, Nick Vulpescu, Gael Westby, Paul Zappile, Yutong Zhang
EPI_ISL_430368 hCoV-19/USA/NY-NYUMC341/2020 North America / USA / New York / Brooklyn 2020-04-08 NYU Langone Health Departments of Pathology and Medicine, New York University School of Medicine Maria Aguero-Rosenfeld, Brendan Belovarac, Margaret Black, Ludovic Boytard, John Cadley, Paolo Cotzia, John Chen, Dacia Dimartino, Xiaojun Feng, Tatyana Gindin, Emily Guzman, Adriana Heguy, Megan Hogan, Emily Huang, George Jour, Lawrence H. Lin, Raven Luther, Andrew Lytle, Christian Marier, Matthew T. Maurano, Mark J. Mulligan, Peter Meyn, Raquel Ordonez Ciriza, Iman Osman, Jared Pinnell, Vanessa Raabe, Sitharam Ramaswami, Amy Rapkiewicz, Andre M. Ribeiro-dos-Santos, Marie Samanovic-Golden, Antonio Serrano, Guomiao Shen, Matija Snuderl, Theodore Vougiouklakis, Nick Vulpescu, Gael Westby, Paul Zappile, Yutong Zhang
EPI_ISL_430369 hCoV-19/USA/NY-NYUMC342/2020 North America / USA / New York / Manhattan 2020-04-08 NYU Langone Health Departments of Pathology and Medicine, New York University School of Medicine Maria Aguero-Rosenfeld, Brendan Belovarac, Margaret Black, Ludovic Boytard, John Cadley, Paolo Cotzia, John Chen, Dacia Dimartino, Xiaojun Feng, Tatyana Gindin, Emily Guzman, Adriana Heguy, Megan Hogan, Emily Huang, George Jour, Lawrence H. Lin, Raven Luther, Andrew Lytle, Christian Marier, Matthew T. Maurano, Mark J. Mulligan, Peter Meyn, Raquel Ordonez Ciriza, Iman Osman, Jared Pinnell, Vanessa Raabe, Sitharam Ramaswami, Amy Rapkiewicz, Andre M. Ribeiro-dos-Santos, Marie Samanovic-Golden, Antonio Serrano, Guomiao Shen, Matija Snuderl, Theodore Vougiouklakis, Nick Vulpescu, Gael Westby, Paul Zappile, Yutong Zhang
EPI_ISL_430370 hCoV-19/USA/NY-NYUMC343/2020 North America / USA / New York / Queens 2020-04-08 NYU Langone Health Departments of Pathology and Medicine, New York University School of Medicine Maria Aguero-Rosenfeld, Brendan Belovarac, Margaret Black, Ludovic Boytard, John Cadley, Paolo Cotzia, John Chen, Dacia Dimartino, Xiaojun Feng, Tatyana Gindin, Emily Guzman, Adriana Heguy, Megan Hogan, Emily Huang, George Jour, Lawrence H. Lin, Raven Luther, Andrew Lytle, Christian Marier, Matthew T. Maurano, Mark J. Mulligan, Peter Meyn, Raquel Ordonez Ciriza, Iman Osman, Jared Pinnell, Vanessa Raabe, Sitharam Ramaswami, Amy Rapkiewicz, Andre M. Ribeiro-dos-Santos, Marie Samanovic-Golden, Antonio Serrano, Guomiao Shen, Matija Snuderl, Theodore Vougiouklakis, Nick Vulpescu, Gael Westby, Paul Zappile, Yutong Zhang
EPI_ISL_430371 hCoV-19/USA/NJ-NYUMC344/2020 North America / USA / New Jersey / Hudson County 2020-04-08 NYU Langone Health Departments of Pathology and Medicine, New York University School of Medicine Maria Aguero-Rosenfeld, Brendan Belovarac, Margaret Black, Ludovic Boytard, John Cadley, Paolo Cotzia, John Chen, Dacia Dimartino, Xiaojun Feng, Tatyana Gindin, Emily Guzman, Adriana Heguy, Megan Hogan, Emily Huang, George Jour, Lawrence H. Lin, Raven Luther, Andrew Lytle, Christian Marier, Matthew T. Maurano, Mark J. Mulligan, Peter Meyn, Raquel Ordonez Ciriza, Iman Osman, Jared Pinnell, Vanessa Raabe, Sitharam Ramaswami, Amy Rapkiewicz, Andre M. Ribeiro-dos-Santos, Marie Samanovic-Golden, Antonio Serrano, Guomiao Shen, Matija Snuderl, Theodore Vougiouklakis, Nick Vulpescu, Gael Westby, Paul Zappile, Yutong Zhang
EPI_ISL_430372 hCoV-19/USA/NY-NYUMC345/2020 North America / USA / New York / Brooklyn 2020-04-07 NYU Langone Health Departments of Pathology and Medicine, New York University School of Medicine Maria Aguero-Rosenfeld, Brendan Belovarac, Margaret Black, Ludovic Boytard, John Cadley, Paolo Cotzia, John Chen, Dacia Dimartino, Xiaojun Feng, Tatyana Gindin, Emily Guzman, Adriana Heguy, Megan Hogan, Emily Huang, George Jour, Lawrence H. Lin, Raven Luther, Andrew Lytle, Christian Marier, Matthew T. Maurano, Mark J. Mulligan, Peter Meyn, Raquel Ordonez Ciriza, Iman Osman, Jared Pinnell, Vanessa Raabe, Sitharam Ramaswami, Amy Rapkiewicz, Andre M. Ribeiro-dos-Santos, Marie Samanovic-Golden, Antonio Serrano, Guomiao Shen, Matija Snuderl, Theodore Vougiouklakis, Nick Vulpescu, Gael Westby, Paul Zappile, Yutong Zhang
EPI_ISL_430373 hCoV-19/USA/NY-NYUMC346/2020 North America / USA / New York / Brooklyn 2020-04-08 NYU Langone Health Departments of Pathology and Medicine, New York University School of Medicine Maria Aguero-Rosenfeld, Brendan Belovarac, Margaret Black, Ludovic Boytard, John Cadley, Paolo Cotzia, John Chen, Dacia Dimartino, Xiaojun Feng, Tatyana Gindin, Emily Guzman, Adriana Heguy, Megan Hogan, Emily Huang, George Jour, Lawrence H. Lin, Raven Luther, Andrew Lytle, Christian Marier, Matthew T. Maurano, Mark J. Mulligan, Peter Meyn, Raquel Ordonez Ciriza, Iman Osman, Jared Pinnell, Vanessa Raabe, Sitharam Ramaswami, Amy Rapkiewicz, Andre M. Ribeiro-dos-Santos, Marie Samanovic-Golden, Antonio Serrano, Guomiao Shen, Matija Snuderl, Theodore Vougiouklakis, Nick Vulpescu, Gael Westby, Paul Zappile, Yutong Zhang
EPI_ISL_430374 hCoV-19/USA/NY-NYUMC347/2020 North America / USA / New York / Brooklyn 2020-04-08 NYU Langone Health Departments of Pathology and Medicine, New York University School of Medicine Maria Aguero-Rosenfeld, Brendan Belovarac, Margaret Black, Ludovic Boytard, John Cadley, Paolo Cotzia, John Chen, Dacia Dimartino, Xiaojun Feng, Tatyana Gindin, Emily Guzman, Adriana Heguy, Megan Hogan, Emily Huang, George Jour, Lawrence H. Lin, Raven Luther, Andrew Lytle, Christian Marier, Matthew T. Maurano, Mark J. Mulligan, Peter Meyn, Raquel Ordonez Ciriza, Iman Osman, Jared Pinnell, Vanessa Raabe, Sitharam Ramaswami, Amy Rapkiewicz, Andre M. Ribeiro-dos-Santos, Marie Samanovic-Golden, Antonio Serrano, Guomiao Shen, Matija Snuderl, Theodore Vougiouklakis, Nick Vulpescu, Gael Westby, Paul Zappile, Yutong Zhang
EPI_ISL_430375 hCoV-19/USA/NY-NYUMC348/2020 North America / USA / New York / Brooklyn 2020-04-08 NYU Langone Health Departments of Pathology and Medicine, New York University School of Medicine Maria Aguero-Rosenfeld, Brendan Belovarac, Margaret Black, Ludovic Boytard, John Cadley, Paolo Cotzia, John Chen, Dacia Dimartino, Xiaojun Feng, Tatyana Gindin, Emily Guzman, Adriana Heguy, Megan Hogan, Emily Huang, George Jour, Lawrence H. Lin, Raven Luther, Andrew Lytle, Christian Marier, Matthew T. Maurano, Mark J. Mulligan, Peter Meyn, Raquel Ordonez Ciriza, Iman Osman, Jared Pinnell, Vanessa Raabe, Sitharam Ramaswami, Amy Rapkiewicz, Andre M. Ribeiro-dos-Santos, Marie Samanovic-Golden, Antonio Serrano, Guomiao Shen, Matija Snuderl, Theodore Vougiouklakis, Nick Vulpescu, Gael Westby, Paul Zappile, Yutong Zhang
EPI_ISL_430376 hCoV-19/USA/NY-NYUMC349/2020 North America / USA / New York / Brooklyn 2020-04-08 NYU Langone Health Departments of Pathology and Medicine, New York University School of Medicine Maria Aguero-Rosenfeld, Brendan Belovarac, Margaret Black, Ludovic Boytard, John Cadley, Paolo Cotzia, John Chen, Dacia Dimartino, Xiaojun Feng, Tatyana Gindin, Emily Guzman, Adriana Heguy, Megan Hogan, Emily Huang, George Jour, Lawrence H. Lin, Raven Luther, Andrew Lytle, Christian Marier, Matthew T. Maurano, Mark J. Mulligan,

Peter Meyn, Raquel Ordonez Ciriza, Iman Osman, Jared Pinnell, Vanessa Raabe, Sitharam Ramaswami, Amy Rapkiewicz, Andre M. Ribeiro-dos-Santos, Marie Samanovic-Golden, Antonio Serrano, Guomiao Shen, Matija Snuderl, Theodore Vougiouklakis, Nick Vulpescu, Gael Westby, Paul Zappile, Yutong Zhang
EPI_ISL_430600 hCoV-19/Australia/VIC1134/2020 Oceania / Australia / Victoria 2020-04-08 Victorian Infectious Diseases Reference Laboratory (VIDRL) Microbiological Diagnostic Unit Public Health Laboratory and Victorian Infectious Diseases Reference Laboratory, The Peter Doherty Institute for Infection and Immunity Caly L., Seemann T., Sait, M., Schultz M., Druce J., Sherry, N.
EPI_ISL_430601 hCoV-19/Australia/VIC1135/2020 Oceania / Australia / Victoria 2020-04-08 Victorian Infectious Diseases Reference Laboratory (VIDRL) Microbiological Diagnostic Unit Public Health Laboratory and Victorian Infectious Diseases Reference Laboratory, The Peter Doherty Institute for Infection and Immunity Caly L., Seemann T., Sait, M., Schultz M., Druce J., Sherry, N.
EPI_ISL_430602 hCoV-19/Australia/VIC1158/2020 Oceania / Australia / Victoria 2020-04-09 Victorian Infectious Diseases Reference Laboratory (VIDRL) Microbiological Diagnostic Unit Public Health Laboratory and Victorian Infectious Diseases Reference Laboratory, The Peter Doherty Institute for Infection and Immunity Caly L., Seemann T., Sait, M., Schultz M., Druce J., Sherry, N.
EPI_ISL_430604 hCoV-19/Australia/VIC1159/2020 Oceania / Australia / Victoria 2020-04-09 Victorian Infectious Diseases Reference Laboratory (VIDRL) Microbiological Diagnostic Unit Public Health Laboratory and Victorian Infectious Diseases Reference Laboratory, The Peter Doherty Institute for Infection and Immunity Caly L., Seemann T., Sait, M., Schultz M., Druce J., Sherry, N.
EPI_ISL_430605 hCoV-19/Australia/VIC1160/2020 Oceania / Australia / Victoria 2020-04-09 Victorian Infectious Diseases Reference Laboratory (VIDRL) Microbiological Diagnostic Unit Public Health Laboratory and Victorian Infectious Diseases Reference Laboratory, The Peter Doherty Institute for Infection and Immunity Caly L., Seemann T., Sait, M., Schultz M., Druce J., Sherry, N.
EPI_ISL_430607 hCoV-19/Australia/VIC1137/2020 Oceania / Australia / Victoria 2020-04-08 Victorian Infectious Diseases Reference Laboratory (VIDRL) Microbiological Diagnostic Unit Public Health Laboratory and Victorian Infectious Diseases Reference Laboratory, The Peter Doherty Institute for Infection and Immunity Caly L., Seemann T., Sait, M., Schultz M., Druce J., Sherry, N.
EPI_ISL_430609 hCoV-19/Australia/VIC1138/2020 Oceania / Australia / Victoria 2020-04-08 Victorian Infectious Diseases Reference Laboratory (VIDRL) Microbiological Diagnostic Unit Public Health Laboratory and Victorian Infectious Diseases Reference Laboratory, The Peter Doherty Institute for Infection and Immunity Caly L., Seemann T., Sait, M., Schultz M., Druce J., Sherry, N.
EPI_ISL_430610 hCoV-19/Australia/VIC1139/2020 Oceania / Australia / Victoria 2020-04-08 Victorian Infectious Diseases Reference Laboratory (VIDRL) Microbiological Diagnostic Unit Public Health Laboratory and Victorian Infectious Diseases Reference Laboratory, The Peter Doherty Institute for Infection and Immunity Caly L., Seemann T., Sait, M., Schultz M., Druce J., Sherry, N.
EPI_ISL_430614 hCoV-19/Australia/VIC1143/2020 Oceania / Australia / Victoria 2020-04-08 Victorian Infectious Diseases Reference Laboratory (VIDRL) Microbiological Diagnostic Unit Public Health Laboratory and Victorian Infectious Diseases Reference Laboratory, The Peter Doherty Institute for Infection and Immunity Caly L., Seemann T., Sait, M., Schultz M., Druce J., Sherry, N.
EPI_ISL_430615 hCoV-19/Australia/VIC1161/2020 Oceania / Australia / Victoria 2020-04-09 Victorian Infectious Diseases Reference Laboratory (VIDRL) Microbiological Diagnostic Unit Public Health Laboratory and Victorian Infectious Diseases Reference Laboratory, The Peter Doherty Institute for Infection and Immunity Caly L., Seemann T., Sait, M., Schultz M., Druce J., Sherry, N.
EPI_ISL_430616 hCoV-19/Australia/VIC1162/2020 Oceania / Australia / Victoria 2020-04-09 Victorian Infectious Diseases Reference Laboratory (VIDRL) Microbiological Diagnostic Unit Public Health Laboratory and Victorian Infectious Diseases Reference Laboratory, The Peter Doherty Institute for Infection and Immunity Caly L., Seemann T., Sait, M., Schultz M., Druce J., Sherry, N.
EPI_ISL_430618 hCoV-19/Australia/VIC1109/2020 Oceania / Australia / Victoria 2020-04-07 Victorian Infectious Diseases Reference Laboratory (VIDRL) Microbiological Diagnostic Unit Public Health Laboratory and Victorian Infectious Diseases Reference Laboratory, The Peter Doherty Institute for Infection and Immunity Caly L., Seemann T., Sait, M., Schultz M., Druce J., Sherry, N.
EPI_ISL_430619 hCoV-19/Australia/VIC1144/2020 Oceania / Australia / Victoria 2020-04-08 Victorian Infectious Diseases Reference Laboratory (VIDRL) Microbiological Diagnostic Unit Public Health Laboratory and Victorian Infectious Diseases Reference Laboratory, The Peter Doherty Institute for Infection and Immunity Caly L., Seemann T., Sait, M., Schultz M., Druce J., Sherry, N.
EPI_ISL_430623 hCoV-19/Australia/VIC1174/2020 Oceania / Australia / Victoria 2020-04-10 Victorian Infectious Diseases Reference Laboratory (VIDRL) Microbiological Diagnostic Unit Public Health Laboratory and Victorian Infectious Diseases Reference Laboratory, The Peter Doherty Institute for Infection and Immunity Caly L., Seemann T., Sait, M., Schultz M., Druce J., Sherry, N.
EPI_ISL_430632 hCoV-19/Australia/NT29/2020 Oceania / Australia / Northern Territory 2020 Royal Darwin Hospital Pathology Microbiological Diagnostic Unit Public Health Laboratory and Victorian Infectious Diseases Reference Laboratory, The Peter Doherty Institute for Infection and Immunity Meumann, E., Caly L., Seemann T., Sait, M., Schultz M., Druce J., Sherry, N.
EPI_ISL_430633 hCoV-19/Australia/NT30/2020 Oceania / Australia / Northern Territory 2020 Royal Darwin Hospital Pathology Microbiological Diagnostic Unit Public Health Laboratory and Victorian Infectious Diseases Reference Laboratory, The Peter Doherty Institute for Infection and Immunity Meumann, E., Caly L., Seemann T., Sait, M., Schultz M., Druce J., Sherry, N.
EPI_ISL_430635 hCoV-19/Australia/NT31/2020 Oceania / Australia / Northern Territory 2020 Royal Darwin Hospital Pathology Microbiological Diagnostic Unit Public Health Laboratory and Victorian Infectious Diseases Reference Laboratory, The Peter Doherty Institute for Infection and Immunity

Meumann, E., Caly L., Seemann T., Sait, M., Schultz M., Druce J., Sherry, N.
EPI_ISL_430638 hCoV-19/Australia/VIC1355/2020 Oceania / Australia / Victoria 2020-04-09 Victorian Infectious Diseases Reference Laboratory (VIDRL) Microbiological Diagnostic Unit Public Health Laboratory and Victorian Infectious Diseases Reference Laboratory, The Peter Doherty Institute for Infection and Immunity Caly L., Seemann T., Sait, M., Schultz M., Druce J., Sherry, N.
EPI_ISL_430639 hCoV-19/Australia/VIC942/2020 Oceania / Australia / Victoria 2020-03-26 Microbiological Diagnostic Unit Public Health Laboratory Microbiological Diagnostic Unit Public Health Laboratory Seemann T., Schultz M., Sait, M., Sherry, N.
EPI_ISL_430640 hCoV-19/Australia/VIC946/2020 Oceania / Australia / Victoria 2020-03-26 Microbiological Diagnostic Unit Public Health Laboratory Microbiological Diagnostic Unit Public Health Laboratory Seemann T., Schultz M., Sait, M., Sherry, N.
EPI_ISL_430645 hCoV-19/Australia/VIC953/2020 Oceania / Australia / Victoria 2020-03-26 Microbiological Diagnostic Unit Public Health Laboratory Microbiological Diagnostic Unit Public Health Laboratory Seemann T., Schultz M., Sait, M., Sherry, N.
EPI_ISL_430646 hCoV-19/Australia/VIC951/2020 Oceania / Australia / Victoria 2020-03-26 Microbiological Diagnostic Unit Public Health Laboratory Microbiological Diagnostic Unit Public Health Laboratory Seemann T., Schultz M., Sait, M., Sherry, N.
EPI_ISL_430647 hCoV-19/Australia/VIC948/2020 Oceania / Australia / Victoria 2020-03-26 Microbiological Diagnostic Unit Public Health Laboratory Microbiological Diagnostic Unit Public Health Laboratory Seemann T., Schultz M., Sait, M., Sherry, N.
EPI_ISL_430649 hCoV-19/Australia/VIC957/2020 Oceania / Australia / Victoria 2020-03-27 Microbiological Diagnostic Unit Public Health Laboratory Microbiological Diagnostic Unit Public Health Laboratory Seemann T., Schultz M., Sait, M., Sherry, N.
EPI_ISL_430653 hCoV-19/Australia/VIC966/2020 Oceania / Australia / Victoria 2020-03-27 Microbiological Diagnostic Unit Public Health Laboratory Microbiological Diagnostic Unit Public Health Laboratory Seemann T., Schultz M., Sait, M., Sherry, N.
EPI_ISL_430654 hCoV-19/Australia/VIC961/2020 Oceania / Australia / Victoria 2020-03-27 Microbiological Diagnostic Unit Public Health Laboratory Microbiological Diagnostic Unit Public Health Laboratory Seemann T., Schultz M., Sait, M., Sherry, N.
EPI_ISL_430655 hCoV-19/Australia/VIC964/2020 Oceania / Australia / Victoria 2020-03-27 Microbiological Diagnostic Unit Public Health Laboratory Microbiological Diagnostic Unit Public Health Laboratory Seemann T., Schultz M., Sait, M., Sherry, N.
EPI_ISL_430658 hCoV-19/Australia/VIC975/2020 Oceania / Australia / Victoria 2020-03-28 Microbiological Diagnostic Unit Public Health Laboratory Microbiological Diagnostic Unit Public Health Laboratory Seemann T., Schultz M., Sait, M., Sherry, N.
EPI_ISL_430659 hCoV-19/Australia/VIC974/2020 Oceania / Australia / Victoria 2020-03-28 Microbiological Diagnostic Unit Public Health Laboratory Microbiological Diagnostic Unit Public Health Laboratory Seemann T., Schultz M., Sait, M., Sherry, N.
EPI_ISL_430661 hCoV-19/Australia/VIC971/2020 Oceania / Australia / Victoria 2020-03-27 Microbiological Diagnostic Unit Public Health Laboratory Microbiological Diagnostic Unit Public Health Laboratory Seemann T., Schultz M., Sait, M., Sherry, N.
EPI_ISL_430663 hCoV-19/Australia/VIC959/2020 Oceania / Australia / Victoria 2020-03-27 Microbiological Diagnostic Unit Public Health Laboratory Microbiological Diagnostic Unit Public Health Laboratory Seemann T., Schultz M., Sait, M., Sherry, N.
EPI_ISL_430669 hCoV-19/Australia/VIC1056/2020 Oceania / Australia / Victoria 2020-04-03 Microbiological Diagnostic Unit Public Health Laboratory Microbiological Diagnostic Unit Public Health Laboratory Seemann T., Schultz M., Sait, M., Sherry, N.
EPI_ISL_430670 hCoV-19/Australia/VIC1067/2020 Oceania / Australia / Victoria 2020-04-04 Microbiological Diagnostic Unit Public Health Laboratory Microbiological Diagnostic Unit Public Health Laboratory Seemann T., Schultz M., Sait, M., Sherry, N.
EPI_ISL_430674 hCoV-19/Australia/VIC1178/2020 Oceania / Australia / Victoria 2020-04-10 Microbiological Diagnostic Unit Public Health Laboratory Microbiological Diagnostic Unit Public Health Laboratory Seemann T., Schultz M., Sait, M., Sherry, N.
EPI_ISL_430676 hCoV-19/Australia/VIC1281/2020 Oceania / Australia / Victoria 2020-04-08 Microbiological Diagnostic Unit Public Health Laboratory Microbiological Diagnostic Unit Public Health Laboratory Seemann T., Schultz M., Sait, M., Sherry, N.
EPI_ISL_430677 hCoV-19/Australia/VIC1300/2020 Oceania / Australia / Victoria 2020-04-10 Microbiological Diagnostic Unit Public Health Laboratory Microbiological Diagnostic Unit Public Health Laboratory Seemann T., Schultz M., Sait, M., Sherry, N.
EPI_ISL_430678 hCoV-19/Australia/VIC1279/2020 Oceania / Australia / Victoria 2020-04-07 Microbiological Diagnostic Unit Public Health Laboratory Microbiological Diagnostic Unit Public Health Laboratory Seemann T., Schultz M., Sait, M., Sherry, N.
EPI_ISL_430679 hCoV-19/Australia/VIC1301/2020 Oceania / Australia / Victoria 2020-04-10 Microbiological Diagnostic Unit Public Health Laboratory Microbiological Diagnostic Unit Public Health Laboratory Seemann T., Schultz M., Sait, M., Sherry, N.
EPI_ISL_430680 hCoV-19/Australia/VIC1302/2020 Oceania / Australia / Victoria 2020-04-10 Microbiological Diagnostic Unit Public Health Laboratory Microbiological Diagnostic Unit Public Health Laboratory Seemann T., Schultz M., Sait, M., Sherry, N.
EPI_ISL_430681 hCoV-19/Australia/VIC1314/2020 Oceania / Australia / Victoria 2020-04-11 Microbiological Diagnostic Unit Public Health Laboratory Microbiological Diagnostic Unit Public Health Laboratory

Seemann T., Schultz M., Sait, M., Sherry, N.
EPI_ISL_430682 hCoV-19/Australia/VIC1334/2020 Oceania / Australia / Victoria 2020-04-13 Microbiological Diagnostic Unit Public Health Laboratory Microbiological Diagnostic Unit Public Health Laboratory Seemann T., Schultz M., Sait, M., Sherry, N.
EPI_ISL_430683 hCoV-19/Australia/VIC1342/2020 Oceania / Australia / Victoria 2020-04-14 Microbiological Diagnostic Unit Public Health Laboratory Microbiological Diagnostic Unit Public Health Laboratory Seemann T., Schultz M., Sait, M., Sherry, N.
EPI_ISL_430684 hCoV-19/Australia/VIC1343/2020 Oceania / Australia / Victoria 2020-04-14 Microbiological Diagnostic Unit Public Health Laboratory Microbiological Diagnostic Unit Public Health Laboratory Seemann T., Schultz M., Sait, M., Sherry, N.
EPI_ISL_430686 hCoV-19/Australia/VIC1340/2020 Oceania / Australia / Victoria 2020-04-13 Microbiological Diagnostic Unit Public Health Laboratory Microbiological Diagnostic Unit Public Health Laboratory Seemann T., Schultz M., Sait, M., Sherry, N.
EPI_ISL_430689 hCoV-19/Australia/VIC1315/2020 Oceania / Australia / Victoria 2020-04-11 Victorian Infectious Diseases Reference Laboratory (VIDRL) Microbiological Diagnostic Unit Public Health Laboratory and Victorian Infectious Diseases Reference Laboratory, The Peter Doherty Institute for Infection and Immunity Caly L., Seemann T., Sait, M., Schultz M., Druce J., Sherry, N.
EPI_ISL_430690 hCoV-19/Australia/VIC1316/2020 Oceania / Australia / Victoria 2020-04-11 Victorian Infectious Diseases Reference Laboratory (VIDRL) Microbiological Diagnostic Unit Public Health Laboratory and Victorian Infectious Diseases Reference Laboratory, The Peter Doherty Institute for Infection and Immunity Caly L., Seemann T., Sait, M., Schultz M., Druce J., Sherry, N.
EPI_ISL_430691 hCoV-19/Australia/VIC1317/2020 Oceania / Australia / Victoria 2020-04-11 Victorian Infectious Diseases Reference Laboratory (VIDRL) Microbiological Diagnostic Unit Public Health Laboratory and Victorian Infectious Diseases Reference Laboratory, The Peter Doherty Institute for Infection and Immunity Caly L., Seemann T., Sait, M., Schultz M., Druce J., Sherry, N.
EPI_ISL_430693 hCoV-19/Australia/VIC1318/2020 Oceania / Australia / Victoria 2020-04-11 Victorian Infectious Diseases Reference Laboratory (VIDRL) Microbiological Diagnostic Unit Public Health Laboratory and Victorian Infectious Diseases Reference Laboratory, The Peter Doherty Institute for Infection and Immunity Caly L., Seemann T., Sait, M., Schultz M., Druce J., Sherry, N.
EPI_ISL_430694 hCoV-19/Australia/VIC1303/2020 Oceania / Australia / Victoria 2020-04-10 Victorian Infectious Diseases Reference Laboratory (VIDRL) Microbiological Diagnostic Unit Public Health Laboratory and Victorian Infectious Diseases Reference Laboratory, The Peter Doherty Institute for Infection and Immunity Caly L., Seemann T., Sait, M., Schultz M., Druce J., Sherry, N.
EPI_ISL_430695 hCoV-19/Australia/VIC1328/2020 Oceania / Australia / Victoria 2020-04-12 Victorian Infectious Diseases Reference Laboratory (VIDRL) Microbiological Diagnostic Unit Public Health Laboratory and Victorian Infectious Diseases Reference Laboratory, The Peter Doherty Institute for Infection and Immunity Caly L., Seemann T., Sait, M., Schultz M., Druce J., Sherry, N.
EPI_ISL_430696 hCoV-19/Australia/VIC1329/2020 Oceania / Australia / Victoria 2020-04-12 Victorian Infectious Diseases Reference Laboratory (VIDRL) Microbiological Diagnostic Unit Public Health Laboratory and Victorian Infectious Diseases Reference Laboratory, The Peter Doherty Institute for Infection and Immunity Caly L., Seemann T., Sait, M., Schultz M., Druce J., Sherry, N.
EPI_ISL_430697 hCoV-19/Australia/VIC1344/2020 Oceania / Australia / Victoria 2020-04-14 Victorian Infectious Diseases Reference Laboratory (VIDRL) Microbiological Diagnostic Unit Public Health Laboratory and Victorian Infectious Diseases Reference Laboratory, The Peter Doherty Institute for Infection and Immunity Caly L., Seemann T., Sait, M., Schultz M., Druce J., Sherry, N.
EPI_ISL_430698 hCoV-19/Australia/VIC1336/2020 Oceania / Australia / Victoria 2020-04-13 Victorian Infectious Diseases Reference Laboratory (VIDRL) Microbiological Diagnostic Unit Public Health Laboratory and Victorian Infectious Diseases Reference Laboratory, The Peter Doherty Institute for Infection and Immunity Caly L., Seemann T., Sait, M., Schultz M., Druce J., Sherry, N.
EPI_ISL_406798 hCoV-19/Wuhan/WH01/2019 Asia / China / Hubei / Wuhan 2019-12-26 General Hospital of Central Theater Command of People's Liberation Army of China BGI & Institute of Microbiology, Chinese Academy of Sciences & Shandong First Medical University & Shandong Academy of Medical Sciences & General Hospital of Central Theater Command of People's Liberation Army of China Weijun Chen, Yuhai Bi, Weifeng Shi and Zhenhong Hu
EPI_ISL_407976 hCoV-19/Belgium/GHB-03021/2020 Europe / Belgium / Leuven 2020-02-03 KU Leuven, Clinical and Epidemiological Virology KU Leuven, Clinical and Epidemiological Virology Bert Vanmechelen, Elke Wollants, Annabel Rector, Els Keyaerts, Lies Laenen, Marc Van Ranst, and Piet Maes
EPI_ISL_407987 hCoV-19/Singapore/2/2020 Asia / Singapore 2020-01-25 Singapore General Hospital Programme in Emerging Infectious Diseases, Duke-NUS Medical School Danielle E Anderson, Martin Linster, Yan Zhuang, Jayanthi Jayakumar, Kian Sing Chan, Lynette LE Oon, Jenny GH Low, Yvonne CF Su, Linfa Wang, Gavin JD Smith
EPI_ISL_407988 hCoV-19/Singapore/3/2020 Asia / Singapore 2020-02-01 National Centre for Infectious Diseases Programme in Emerging Infectious Diseases, Duke-NUS Medical School Danielle E Anderson, Martin Linster, Yan Zhuang, Jayanthi Jayakumar, David CB Lye, Yee Sin Leo, Barnaby E Young, Yvonne CF Su, Linfa Wang, Gavin JD Smith
EPI_ISL_412459 hCoV-19/Jingzhou/HBCDC-HB-01/2020 Asia / China / Hubei / Jingzhou 2020-01-08 Jingzhou Center for Disease Control and Prevention Hubei Provincial Center for Disease Control and Prevention Bin Fang, Xiang Li, Xiao Yu, Linlin Liu, Bo Yang, Faxian Zhan, Guojun Ye, Xixiang Huo, Junqiang Xu, Bo Yu, Kun Cai, Jing Li, Maoyi Chen,Jie Hu, Chunlin Mao, Yongzhong Jiang.
EPI_ISL_413729 hCoV-19/China/WF0015/2020 Asia / China 2020-02 Weifang Center for Disease Control and

Prevention	Weifang Center for Disease Control and Prevention & BGI-Shenzhen	Qing Nie, Xingguang Li, Erik M Volz, Han Fu, Haowei Wang, Xiaoyue Xi, Wei Chen, Dehui Liu, Yingying Chen, Mengmeng Tian, Wei Tan, Junjie Zai, Wanying Sun, Jiandong Li, Junhua Li
EPI_ISL_413746	hCoV-19/China/WF0016/2020	Asia / China	2020-02 Weifang Center for Disease Control and Prevention	Weifang Center for Disease Control and Prevention & BGI-Shenzhen	Qing Nie, Xingguang Li, Erik M Volz, Han Fu, Haowei Wang, Xiaoyue Xi, Wei Chen, Dehui Liu, Yingying Chen, Mengmeng Tian, Wei Tan, Junjie Zai, Wanying Sun, Jiandong Li, Junhua Li
EPI_ISL_413748	hCoV-19/China/WF0018/2020	Asia / China	2020-02 Weifang Center for Disease Control and Prevention	Weifang Center for Disease Control and Prevention & BGI-Shenzhen	Qing Nie, Xingguang Li, Erik M Volz, Han Fu, Haowei Wang, Xiaoyue Xi, Wei Chen, Dehui Liu, Yingying Chen, Mengmeng Tian, Wei Tan, Junjie Zai, Wanying Sun, Jiandong Li, Junhua Li
EPI_ISL_413749	hCoV-19/China/WF0019/2020	Asia / China	2020-02 Weifang Center for Disease Control and Prevention	Weifang Center for Disease Control and Prevention & BGI-Shenzhen	Qing Nie, Xingguang Li, Erik M Volz, Han Fu, Haowei Wang, Xiaoyue Xi, Wei Chen, Dehui Liu, Yingying Chen, Mengmeng Tian, Wei Tan, Junjie Zai, Wanying Sun, Jiandong Li, Junhua Li
EPI_ISL_413750	hCoV-19/China/WF0020/2020	Asia / China	2020-02 Weifang Center for Disease Control and Prevention	Weifang Center for Disease Control and Prevention & BGI-Shenzhen	Qing Nie, Xingguang Li, Erik M Volz, Han Fu, Haowei Wang, Xiaoyue Xi, Wei Chen, Dehui Liu, Yingying Chen, Mengmeng Tian, Wei Tan, Junjie Zai, Wanying Sun, Jiandong Li, Junhua Li
EPI_ISL_413751	hCoV-19/China/WF0021/2020	Asia / China	2020-02 Weifang Center for Disease Control and Prevention	Weifang Center for Disease Control and Prevention & BGI-Shenzhen	Qing Nie, Xingguang Li, Erik M Volz, Han Fu, Haowei Wang, Xiaoyue Xi, Wei Chen, Dehui Liu, Yingying Chen, Mengmeng Tian, Wei Tan, Junjie Zai, Wanying Sun, Jiandong Li, Junhua Li
EPI_ISL_413753	hCoV-19/China/WF0024/2020	Asia / China	2020-02 Weifang Center for Disease Control and Prevention	Weifang Center for Disease Control and Prevention & BGI-Shenzhen	Qing Nie, Xingguang Li, Erik M Volz, Han Fu, Haowei Wang, Xiaoyue Xi, Wei Chen, Dehui Liu, Yingying Chen, Mengmeng Tian, Wei Tan, Junjie Zai, Wanying Sun, Jiandong Li, Junhua Li
EPI_ISL_413761	hCoV-19/China/WF0026/2020	Asia / China	2020-02 Weifang Center for Disease Control and Prevention	Weifang Center for Disease Control and Prevention & BGI-Shenzhen	Qing Nie, Xingguang Li, Erik M Volz, Han Fu, Haowei Wang, Xiaoyue Xi, Wei Chen, Dehui Liu, Yingying Chen, Mengmeng Tian, Wei Tan, Junjie Zai, Wanying Sun, Jiandong Li, Junhua Li
EPI_ISL_413791	hCoV-19/China/WF0028/2020	Asia / China	2020-02 Weifang Center for Disease Control and Prevention	Weifang Center for Disease Control and Prevention & BGI-Shenzhen	Qing Nie, Xingguang Li, Erik M Volz, Han Fu, Haowei Wang, Xiaoyue Xi, Wei Chen, Dehui Liu, Yingying Chen, Mengmeng Tian, Wei Tan, Junjie Zai, Wanying Sun, Jiandong Li, Junhua Li
EPI_ISL_413809	hCoV-19/China/WF0029/2020	Asia / China	2020-02 Weifang Center for Disease Control and Prevention	Weifang Center for Disease Control and Prevention & BGI-Shenzhen	Qing Nie, Xingguang Li, Erik M Volz, Han Fu, Haowei Wang, Xiaoyue Xi, Wei Chen, Dehui Liu, Yingying Chen, Mengmeng Tian, Wei Tan, Junjie Zai, Wanying Sun, Jiandong Li, Junhua Li
EPI_ISL_413924	hCoV-19/USA/CA-CDPH-UC6/2020	North America / USA / California / Grand Princess cruise ship	2020-03-05	California Department of Public Health	Chiu Laboratory, University of California, San Francisco	Xianding Deng, Scot Federman, Chao-Yang Pan, Hugo Guevara,Wei Gu, Debra A. Wadford, and Charles Y. Chiu
EPI_ISL_413925	hCoV-19/USA/CA-CDPH-UC7/2020	North America / USA / California / Grand Princess cruise ship	2020-03-05	California Department of Public Health	Chiu Laboratory, University of California, San Francisco	Xianding Deng, Scot Federman, Chao-Yang Pan, Hugo Guevara,Wei Gu, Debra A. Wadford, and Charles Y. Chiu
EPI_ISL_413928	hCoV-19/USA/CA-CDPH-UC9/2020	North America / USA / California / Grand Princess cruise ship	2020-03-05	California Department of Public Health	Chiu Laboratory, University of California, San Francisco	Xianding Deng, Scot Federman, Chao-Yang Pan, Hugo Guevara,Wei Gu, Debra A. Wadford, and Charles Y. Chiu
EPI_ISL_413931	hCoV-19/USA/CA-CDPH-UC11/2020	North America / USA / California / Grand Princess cruise ship	2020-03-05	California Department of Public Health	Chiu Laboratory, University of California, San Francisco	Xianding Deng, Scot Federman, Chao-Yang Pan, Hugo Guevara,Wei Gu, Debra A. Wadford, and Charles Y. Chiu
EPI_ISL_416024	hCoV-19/Wales/PHW35/2020	Europe / United Kingdom / Wales 2020-03-11	Wales Specialist Virology Centre	Public Health Wales Microbiology Cardiff	Catherine Moore, Joanne Watkins, Sally Corden, Tom Connor
EPI_ISL_416026	hCoV-19/Wales/PHW21/2020	Europe / United Kingdom / Wales 2020-03-11	Wales Specialist Virology Centre	Public Health Wales Microbiology Cardiff	Catherine Moore, Joanne Watkins, Sally Corden, Tom Connor
EPI_ISL_416028	hCoV-19/Brazil/SPBR-07/2020	South America / Brazil / Sao Paulo / Sao Paulo	2020-03-03	National Influenza Center - Instituto Adolfo Lutz	Instituto Adolfo Lutz, Interdiciplinary Procedures Center, Strategic Laboratory	Claudio Tavares Sacchi, Claudia Regina Gonçalves, Carlos Henrique Camargo, Fabiana Cristina Pereira dos Santos, Daniela Bernardes Borges da Silva, Simone Guadagnucci Morillo, Adriano Abbud, Adriana Bugno, Maria do Carmo Sampaio Tavares Timenetsky, Terezinha Maria de Paiva
EPI_ISL_416029	hCoV-19/Brazil/SPBR-08/2020	South America / Brazil / Sao Paulo / Sao Paulo	2020-03-04	Laboiratório Fleury	Instituto Adolfo Lutz, Interdiciplinary Procedures Center, Strategic Laboratory Claudio Tavares Sacchi, Claudia Regina Gonçalves, Carlos Henrique Camargo, Fabiana Cristina Pereira dos Santos, Daniela Bernardes Borges da Silva, Simone Guadagnucci Morillo, Adriano Abbud, Adriana Bugno, Maria do Carmo Sampaio

Tavares Timenetsky, Terezinha Maria de Paiva
EPI_ISL_416031 hCoV-19/Brazil/SPBR-09/2020 South America / Brazil / Sao Paulo / Sao Paulo 2020-03-04 National Influenza Center - Instituto Adolfo Lutz Instituto Adolfo Lutz, Interdiciplinary Procedures Center, Strategic Laboratory Claudio Tavares Sacchi, Claudia Regina Gonçalves, Carlos Henrique Camargo, Fabiana Cristina Pereira dos Santos, Daniela Bernardes Borges da Silva, Simone Guadagnucci Morillo, Adriano Abbud, Adriana Bugno, Maria do Carmo Sampaio Tavares Timenetsky, Terezinha Maria de Paiva
EPI_ISL_416032 hCoV-19/Brazil/SPBR-10/2020 South America / Brazil / Distrito Federal / Brasilia 2020-03-04 National Influenza Center - Instituto Adolfo Lutz Instituto Adolfo Lutz, Interdiciplinary Procedures Center, Strategic Laboratory Claudio Tavares Sacchi, Claudia Regina Gonçalves, Carlos Henrique Camargo, Fabiana Cristina Pereira dos Santos, Daniela Bernardes Borges da Silva, Simone Guadagnucci Morillo, Adriano Abbud, Adriana Bugno, Maria do Carmo Sampaio Tavares Timenetsky, Terezinha Maria de Paiva
EPI_ISL_416033 hCoV-19/Brazil/SPBR-11/2020 South America / Brazil / Sao Paulo / Sao Paulo 2020-03-03 Hospital Israelita Albert Einstein Instituto Adolfo Lutz, Interdiciplinary Procedures Center, Strategic Laboratory Claudio Tavares Sacchi, Claudia Regina Gonçalves, Carlos Henrique Camargo, Erica Valessa Ramos Gomes, Fabiana Cristina Pereira dos Santos, Daniela Bernardes Borges da Silva, Simone Guadagnucci Morillo, Adriano Abbud, Adriana Bugno, Maria do Carmo Sampaio Tavares Timenetsky, Terezinha Maria de Paiva
EPI_ISL_416034 hCoV-19/Brazil/SPBR-12/2020 South America / Brazil / Sao Paulo / Sao Paulo 2020-03-04 Hospital Israelita Albert Einstein Instituto Adolfo Lutz, Interdiciplinary Procedures Center, Strategic Laboratory Claudio Tavares Sacchi, Claudia Regina Gonçalves, Carlos Henrique Camargo, Erica Valessa Ramos Gomes, Fabiana Cristina Pereira dos Santos, Daniela Bernardes Borges da Silva, Simone Guadagnucci Morillo, Adriano Abbud, Adriana Bugno, Maria do Carmo Sampaio Tavares Timenetsky, Terezinha Maria de Paiva
EPI_ISL_416035 hCoV-19/Brazil/SPBR-13/2020 South America / Brazil / Sao Paulo / Sao Paulo 2020-03-05 National Influenza Center - Instituto Adolfo Lutz Instituto Adolfo Lutz, Interdiciplinary Procedures Center, Strategic Laboratory Claudio Tavares Sacchi, Claudia Regina Gonçalves, Carlos Henrique Camargo, Erica Valessa Ramos Gomes, Fabiana Cristina Pereira dos Santos, Daniela Bernardes Borges da Silva, Simone Guadagnucci Morillo, Adriano Abbud, Adriana Bugno, Maria do Carmo Sampaio Tavares Timenetsky, Terezinha Maria de Paiva
EPI_ISL_416036 hCoV-19/Brazil/SPBR-14/2020 South America / Brazil / Sao Paulo / Sao Paulo 2020-03-05 National Influenza Center - Instituto Adolfo Lutz Instituto Adolfo Lutz, Interdiciplinary Procedures Center, Strategic Laboratory Claudio Tavares Sacchi, Claudia Regina Gonçalves, Carlos Henrique Camargo, Erica Valessa Ramos Gomes, Fabiana Cristina Pereira dos Santos, Daniela Bernardes Borges da Silva, Simone Guadagnucci Morillo, Adriano Abbud, Adriana Bugno, Maria do Carmo Sampaio Tavares Timenetsky, Terezinha Maria de Paiva
EPI_ISL_416140 hCoV-19/Denmark/SSI-05/2020 Europe / Denmark / Copenhagen 2020-03-02 Department of Virus and Microbiological Special diagnostics, Statens Serum Institut, Copenhagen, Denmark. Statens Serum Institute Morten Rasmussen, Maiken Worsoe Rosenstierne , Anders Fomsgaard
EPI_ISL_416141 hCoV-19/Denmark/SSI-09/2020 Europe / Denmark / Copenhagen 2020-03-03 Department of Virus and Microbiological Special diagnostics, Statens Serum Institut, Copenhagen, Denmark. Statens Serum Institute Morten Rasmussen, Maiken Worsoe Rosenstierne , Anders Fomsgaard
EPI_ISL_416142 hCoV-19/Denmark/SSI-01/2020 Europe / Denmark / Copenhagen 2020-02-26 Department of Virus and Microbiological Special diagnostics, Statens Serum Institut, Copenhagen, Denmark. Statens Serum Institute Morten Rasmussen, Maiken Worsoe Rosenstierne , Anders Fomsgaard
EPI_ISL_416143 hCoV-19/Denmark/SSI-02/2020 Europe / Denmark / Copenhagen 2020-02-28 Department of Virus and Microbiological Special diagnostics, Statens Serum Institut, Copenhagen, Denmark. ViFU Morten Rasmussen, Maiken Worsoe Rosenstierne , Anders Fomsgaard
EPI_ISL_416144 hCoV-19/Denmark/SSI-03/2020 Europe / Denmark / Copenhagen 2020-03-01 Department of Virus and Microbiological Special diagnostics, Statens Serum Institut, Copenhagen, Denmark. ViFU Morten Rasmussen, Maiken Worsoe Rosenstierne , Anders Fomsgaard
EPI_ISL_416153 hCoV-19/Denmark/SSI-04/2020 Europe / Denmark / Copenhagen 2020-03-02 Department of Virus and Microbiological Special diagnostics, Statens Serum Institut, Copenhagen, Denmark. ViFU Morten Rasmussen, Maiken Worsoe Rosenstierne , Anders Fomsgaard
EPI_ISL_417301 hCoV-19/England/20110000606/2020 Europe / United Kingdom / England 2020-03-06 Respiratory Virus Unit, Microbiology Services Colindale, Public Health England Respiratory Virus Unit, Microbiology Services Colindale, Public Health England Monica Galiano, Shahjahan Miah, Angie Lackenby, Omolola Akinbami, Tiina Talts, Leena Bhaw, Richard Myers, Steven Platt, Kirstin Edwards, Jonathan Hubb, Joanna Ellis, Maria Zambon
EPI_ISL_417302 hCoV-19/England/20110000706/2020 Europe / United Kingdom / England 2020-03-05 Respiratory Virus Unit, Microbiology Services Colindale, Public Health England Respiratory Virus Unit, Microbiology Services Colindale, Public Health England Monica Galiano, Shahjahan Miah, Angie Lackenby, Omolola Akinbami, Tiina Talts, Leena Bhaw, Richard Myers, Steven Platt, Kirstin Edwards, Jonathan Hubb, Joanna Ellis, Maria Zambon
EPI_ISL_417307 hCoV-19/England/20110023706/2020 Europe / United Kingdom / England 2020-03-08 Respiratory Virus Unit, Microbiology Services Colindale, Public Health England Respiratory Virus Unit, Microbiology Services Colindale, Public Health England Monica Galiano, Shahjahan Miah, Angie Lackenby, Omolola Akinbami, Tiina Talts, Leena Bhaw, Richard Myers, Steven Platt, Kirstin Edwards, Jonathan Hubb, Joanna Ellis, Maria Zambon
EPI_ISL_417311 hCoV-19/England/20110058706/2020 Europe / United Kingdom / England 2020-03-09 Respiratory Virus Unit, Microbiology Services Colindale, Public Health England Respiratory Virus Unit, Microbiology Services Colindale, Public Health England Monica Galiano, Shahjahan Miah, Angie Lackenby, Omolola Akinbami, Tiina Talts, Leena Bhaw, Richard Myers, Steven Platt, Kirstin Edwards, Jonathan Hubb, Joanna Ellis,

Maria Zambon
EPI_ISL_417312 hCoV-19/England/20110059306/2020 Europe / United Kingdom / England 2020-03-08 Respiratory Virus Unit, Microbiology Services Colindale, Public Health England Respiratory Virus Unit, Microbiology Services Colindale, Public Health England Monica Galiano, Shahjahan Miah, Angie Lackenby, Omolola Akinbami, Tiina Talts, Leena Bhaw, Richard Myers, Steven Platt, Kirstin Edwards, Jonathan Hubb, Joanna Ellis, Maria Zambon
EPI_ISL_417313 hCoV-19/England/20110097506/2020 Europe / United Kingdom / England 2020-03-08 Respiratory Virus Unit, Microbiology Services Colindale, Public Health England Respiratory Virus Unit, Microbiology Services Colindale, Public Health England Monica Galiano, Shahjahan Miah, Angie Lackenby, Omolola Akinbami, Tiina Talts, Leena Bhaw, Richard Myers, Steven Platt, Kirstin Edwards, Jonathan Hubb, Joanna Ellis, Maria Zambon
EPI_ISL_417314 hCoV-19/England/20112112106/2020 Europe / United Kingdom / England 2020-03-10 Respiratory Virus Unit, Microbiology Services Colindale, Public Health England Respiratory Virus Unit, Microbiology Services Colindale, Public Health England Monica Galiano, Shahjahan Miah, Angie Lackenby, Omolola Akinbami, Tiina Talts, Leena Bhaw, Richard Myers, Steven Platt, Kirstin Edwards, Jonathan Hubb, Joanna Ellis, Maria Zambon
EPI_ISL_417315 hCoV-19/England/20118044606/2020 Europe / United Kingdom / England 2020-03-13 Respiratory Virus Unit, Microbiology Services Colindale, Public Health England Respiratory Virus Unit, Microbiology Services Colindale, Public Health England Monica Galiano, Shahjahan Miah, Angie Lackenby, Omolola Akinbami, Tiina Talts, Leena Bhaw, Richard Myers, Steven Platt, Kirstin Edwards, Jonathan Hubb, Joanna Ellis, Maria Zambon
EPI_ISL_417317 hCoV-19/USA/CA-SCCPHD-UC13/2020 North America / USA / California / Santa Clara County 2020-03-02 Santa Clara County Public Health Department Chiu Laboratory, University of California, San Francisco Xianding Deng, Scot Federman, Wei Gu, Elsa Villarino, Brandon Bonin, Debra A. Wadford, and Charles Y. Chiu
EPI_ISL_417318 hCoV-19/USA/CA-SCCPHD-UC14/2020 North America / USA / California / Santa Clara County 2020-02-29 Santa Clara County Public Health Department Chiu Laboratory, University of California, San Francisco Xianding Deng, Scot Federman, Wei Gu, Elsa Villarino, Brandon Bonin, Debra A. Wadford, and Charles Y. Chiu
EPI_ISL_417320 hCoV-19/USA/CA-SCCPHD-UC16/2020 North America / USA / California / Santa Clara County 2020-03-04 Santa Clara County Public Health Department Chiu Laboratory, University of California, San Francisco Xianding Deng, Scot Federman, Wei Gu, Elsa Villarino, Brandon Bonin, Debra A. Wadford, and Charles Y. Chiu
EPI_ISL_417330 hCoV-19/USA/CA-CDPH-UC26/2020 North America / USA / California / San Francisco County 2020-03-13 Chiu Laboratory, University of California, San Francisco Chiu Laboratory, University of California, San Francisco Xianding Deng, Scot Federman, Wei Gu, and Charles Y. Chiu
EPI_ISL_417333 hCoV-19/France/Lyon_06042/2020 Europe / France / ARA 2020-03-04 Institut des Agents Infectieux (IAI), Hospices Civils de Lyon CNR Virus des Infections Respiratoires - France SUD Antonin Bal, Gregory Destras, Gwendolyne Burfin, Solenne Brun, Carine Moustaud, Raphaelle Lamy, Alexandre Gaymard, Maude Bouscambert-Duchamp, Florence Morfin-Sherpa, Martine Valette, Laurence Josset, Bruno Lina
EPI_ISL_417334 hCoV-19/France/Lyon_06056/2020 Europe / France / ARA 2020-03-04 Institut des Agents Infectieux (IAI), Hospices Civils de Lyon CNR Virus des Infections Respiratoires - France SUD Antonin Bal, Gregory Destras, Gwendolyne Burfin, Solenne Brun, Carine Moustaud, Raphaelle Lamy, Alexandre Gaymard, Maude Bouscambert-Duchamp, Florence Morfin-Sherpa, Martine Valette, Laurence Josset, Bruno Lina
EPI_ISL_417335 hCoV-19/France/Lyon_06573/2020 Europe / France / ARA 2020-03-06 Institut des Agents Infectieux (IAI), Hospices Civils de Lyon CNR Virus des Infections Respiratoires - France SUD Antonin Bal, Gregory Destras, Gwendolyne Burfin, Solenne Brun, Carine Moustaud, Raphaelle Lamy, Alexandre Gaymard, Maude Bouscambert-Duchamp, Florence Morfin-Sherpa, Martine Valette, Laurence Josset, Bruno Lina
EPI_ISL_417336 hCoV-19/France/Lyon_0668/2020 Europe / France / ARA 2020-03-06 Institut des Agents Infectieux (IAI), Hospices Civils de Lyon CNR Virus des Infections Respiratoires - France SUD Antonin Bal, Gregory Destras, Gwendolyne Burfin, Solenne Brun, Carine Moustaud, Raphaelle Lamy, Alexandre Gaymard, Maude Bouscambert-Duchamp, Florence Morfin-Sherpa, Martine Valette, Laurence Josset, Bruno Lina
EPI_ISL_417337 hCoV-19/France/Lyon_06625/2020 Europe / France / ARA 2020-03-07 Institut des Agents Infectieux (IAI), Hospices Civils de Lyon CNR Virus des Infections Respiratoires - France SUD Antonin Bal, Gregory Destras, Gwendolyne Burfin, Solenne Brun, Carine Moustaud, Raphaelle Lamy, Alexandre Gaymard, Maude Bouscambert-Duchamp, Florence Morfin-Sherpa, Martine Valette, Laurence Josset, Bruno Lina
EPI_ISL_417338 hCoV-19/France/Macon_06756/2020 Europe / France / ARA 2020-03-07 Centre Hospitalier de Macon CNR Virus des Infections Respiratoires - France SUD Antonin Bal, Gregory Destras, Gwendolyne Burfin, Solenne Brun, Carine Moustaud, Raphaelle Lamy, Alexandre Gaymard, Maude Bouscambert-Duchamp, Florence Morfin-Sherpa, Martine Valette, Laurence Josset, Bruno Lina
EPI_ISL_417339 hCoV-19/France/Lyon_06820/2020 Europe / France / ARA 2020-03-08 Institut des Agents Infectieux (IAI), Hospices Civils de Lyon CNR Virus des Infections Respiratoires - France SUD Antonin Bal, Gregory Destras, Gwendolyne Burfin, Solenne Brun, Carine Moustaud, Raphaelle Lamy, Alexandre Gaymard, Maude Bouscambert-Duchamp, Florence Morfin-Sherpa, Martine Valette, Laurence Josset, Bruno Lina
EPI_ISL_417340 hCoV-19/France/Bourg-en-Bresse_06813/2020 Europe / France / ARA 2020-03-07 Centre Hospitalier de Bourg en Bresse CNR Virus des Infections Respiratoires - France SUD Antonin Bal, Gregory Destras, Gwendolyne Burfin, Solenne Brun, Carine Moustaud, Raphaelle Lamy, Alexandre Gaymard, Maude Bouscambert-Duchamp, Florence Morfin-Sherpa, Martine Valette, Laurence Josset, Bruno Lina
EPI_ISL_417341 hCoV-19/USA/WA-UW192/2020 North America / USA / Washington 2020-03-13 UW Virology Lab UW Virology Lab Pavitra Roychoudhury, Hong Xie, Keith Jerome, Alexander Greninger

EPI_ISL_417342 hCoV-19/USA/WA-UW193/2020 North America / USA / Washington 2020-03-13 UW Virology Lab UW Virology Lab Pavitra Roychoudhury, Hong Xie, Keith Jerome, Alexander Greninger
EPI_ISL_417343 hCoV-19/USA/WA-UW194/2020 North America / USA / Washington 2020-03-13 UW Virology Lab UW Virology Lab Pavitra Roychoudhury, Hong Xie, Keith Jerome, Alexander Greninger
EPI_ISL_417344 hCoV-19/USA/WA-UW195/2020 North America / USA / Washington 2020-03-13 UW Virology Lab UW Virology Lab Pavitra Roychoudhury, Hong Xie, Keith Jerome, Alexander Greninger
EPI_ISL_417345 hCoV-19/USA/WA-UW196/2020 North America / USA / Washington 2020-03-13 UW Virology Lab UW Virology Lab Pavitra Roychoudhury, Hong Xie, Keith Jerome, Alexander Greninger
EPI_ISL_417346 hCoV-19/USA/WA-UW197/2020 North America / USA / Washington 2020-03-12 UW Virology Lab UW Virology Lab Pavitra Roychoudhury, Hong Xie, Keith Jerome, Alexander Greninger
EPI_ISL_417347 hCoV-19/USA/WA-UW198/2020 North America / USA / Washington 2020-03-12 UW Virology Lab UW Virology Lab Pavitra Roychoudhury, Hong Xie, Keith Jerome, Alexander Greninger
EPI_ISL_417348 hCoV-19/USA/WA-UW199/2020 North America / USA / Washington 2020-03-13 UW Virology Lab UW Virology Lab Pavitra Roychoudhury, Hong Xie, Keith Jerome, Alexander Greninger
EPI_ISL_417349 hCoV-19/USA/WA-UW200/2020 North America / USA / Washington 2020-03-12 UW Virology Lab UW Virology Lab Pavitra Roychoudhury, Hong Xie, Keith Jerome, Alexander Greninger
EPI_ISL_417350 hCoV-19/USA/WA-UW201/2020 North America / USA / Washington 2020-03-12 UW Virology Lab UW Virology Lab Pavitra Roychoudhury, Hong Xie, Keith Jerome, Alexander Greninger
EPI_ISL_417351 hCoV-19/USA/WA-UW202/2020 North America / USA / Washington 2020-03-12 UW Virology Lab UW Virology Lab Pavitra Roychoudhury, Hong Xie, Keith Jerome, Alexander Greninger
EPI_ISL_417352 hCoV-19/USA/WA-UW203/2020 North America / USA / Washington 2020-03-12 UW Virology Lab UW Virology Lab Pavitra Roychoudhury, Hong Xie, Keith Jerome, Alexander Greninger
EPI_ISL_417353 hCoV-19/USA/WA-UW204/2020 North America / USA / Washington 2020-03-13 UW Virology Lab UW Virology Lab Pavitra Roychoudhury, Hong Xie, Keith Jerome, Alexander Greninger
EPI_ISL_417354 hCoV-19/USA/WA-UW205/2020 North America / USA / Washington 2020-03-13 UW Virology Lab UW Virology Lab Pavitra Roychoudhury, Hong Xie, Keith Jerome, Alexander Greninger
EPI_ISL_417355 hCoV-19/USA/WA-UW206/2020 North America / USA / Washington 2020-03-13 UW Virology Lab UW Virology Lab Pavitra Roychoudhury, Hong Xie, Keith Jerome, Alexander Greninger
EPI_ISL_417356 hCoV-19/USA/WA-UW207/2020 North America / USA / Washington 2020-03-13 UW Virology Lab UW Virology Lab Pavitra Roychoudhury, Hong Xie, Keith Jerome, Alexander Greninger
EPI_ISL_417358 hCoV-19/USA/WA-UW209/2020 North America / USA / Washington 2020-03-13 UW Virology Lab UW Virology Lab Pavitra Roychoudhury, Hong Xie, Keith Jerome, Alexander Greninger
EPI_ISL_417359 hCoV-19/USA/WA-UW210/2020 North America / USA / Washington 2020-03-14 UW Virology Lab UW Virology Lab Pavitra Roychoudhury, Hong Xie, Keith Jerome, Alexander Greninger
EPI_ISL_417360 hCoV-19/USA/WA-UW211/2020 North America / USA / Washington 2020-03-14 UW Virology Lab UW Virology Lab Pavitra Roychoudhury, Hong Xie, Keith Jerome, Alexander Greninger
EPI_ISL_417361 hCoV-19/USA/WA-UW212/2020 North America / USA / Washington 2020-03-14 UW Virology Lab UW Virology Lab Pavitra Roychoudhury, Hong Xie, Keith Jerome, Alexander Greninger
EPI_ISL_417362 hCoV-19/USA/CT-UW213/2020 North America / USA / Connecticut 2020-03-13 UW Virology Lab UW Virology Lab Pavitra Roychoudhury, Hong Xie, Keith Jerome, Alexander Greninger
EPI_ISL_417363 hCoV-19/USA/WA-UW214/2020 North America / USA / Washington 2020-03-14 UW Virology Lab UW Virology Lab Pavitra Roychoudhury, Hong Xie, Keith Jerome, Alexander Greninger
EPI_ISL_417364 hCoV-19/USA/WA-UW215/2020 North America / USA / Washington 2020-03-12 UW Virology Lab UW Virology Lab Pavitra Roychoudhury, Hong Xie, Keith Jerome, Alexander Greninger
EPI_ISL_417365 hCoV-19/USA/WA-UW216/2020 North America / USA / Washington 2020-03-14 UW Virology Lab UW Virology Lab Pavitra Roychoudhury, Hong Xie, Keith Jerome, Alexander Greninger
EPI_ISL_417366 hCoV-19/USA/WA-UW217/2020 North America / USA / Washington 2020-03-14 UW Virology Lab UW Virology Lab Pavitra Roychoudhury, Hong Xie, Keith Jerome, Alexander Greninger
EPI_ISL_417367 hCoV-19/USA/WA-UW218/2020 North America / USA / Washington 2020-03-13 UW Virology Lab UW Virology Lab Pavitra Roychoudhury, Hong Xie, Keith Jerome, Alexander Greninger
EPI_ISL_417368 hCoV-19/USA/UN-UW219/2020 North America / USA 2020-03-13 UW Virology Lab UW Virology Lab Pavitra Roychoudhury, Hong Xie, Keith Jerome, Alexander Greninger
EPI_ISL_417369 hCoV-19/USA/WA-UW220/2020 North America / USA / Washington 2020-03-14 UW Virology Lab UW Virology Lab Pavitra Roychoudhury, Hong Xie, Keith Jerome, Alexander Greninger
EPI_ISL_417390 hCoV-19/Australia/NSW27/2020 Oceania / Australia / New South Wales / Sydney 2020-03-08 Centre for Infectious Diseases and Microbiology Public Health NSW Health Pathology - Institute of Clinical Pathology and Medical Research; Westmead Hospital; University of Sydney Sadsad R, Carter I, Rahman H, Holmes EC, O'Sullivan MV, Sintchenko V, Chen SC, Maddocks S, Kok J, Dwyer DE, Rockett R, Eden J-S, Lam C, Gray K, Timms V, Gall M and Arnott A for the 2019-nCoV Study Group
EPI_ISL_417391 hCoV-19/Australia/NSW28/2020 Oceania / Australia / New South Wales / Sydney 2020-03-08 Centre for Infectious Diseases and Microbiology Public Health NSW Health Pathology - Institute of Clinical Pathology and Medical Research; Westmead Hospital; University of Sydney Carter I, Rahman H, Holmes EC, O'Sullivan MV, Sintchenko V, Chen SC, Maddocks S, Kok J, Dwyer DE, Rockett R, Eden J-S, Lam C, Gray K, Timms V, Gall M, Arnott A and Sadsad R for the 2019-nCoV Study Group
EPI_ISL_417392 hCoV-19/Australia/NSW31/2020 Oceania / Australia / New South Wales / Sydney 2020-03-10 Centre for Infectious Diseases and Microbiology Public Health NSW Health Pathology - Institute of Clinical Pathology and Medical Research; Westmead Hospital; University of Sydney Rahman H, Holmes EC, O'Sullivan MV, Sintchenko V, Chen SC, Maddocks S, Kok J, Dwyer DE, Rockett R, Eden J-S, Lam C, Gray K, Timms V, Gall M, Arnott A, Sadsad R and Carter I for the 2019-nCoV Study Group
EPI_ISL_417393 hCoV-19/Australia/NSW35/2020 Oceania / Australia / New South Wales / Sydney 2020-03-08

Centre for Infectious Diseases and Microbiology Public Health NSW Health Pathology - Institute of Clinical Pathology and Medical Research; Westmead Hospital; University of Sydney Holmes EC, O'Sullivan MV, Sintchenko V, Chen SC, Maddocks S, Kok J, Dwyer DE, Rockett R, Eden J-S, Lam C, Gray K, Timms V, Gall M, Arnott A, Sadsad R, Carter I and Rahman H for the 2019-nCoV Study Group
EPI_ISL_417394 hCoV-19/Australia/NSW36/2020 Oceania / Australia / New South Wales / Sydney 2020-03-10 Centre for Infectious Diseases and Microbiology Public Health NSW Health Pathology - Institute of Clinical Pathology and Medical Research; Westmead Hospital; University of Sydney O'Sullivan MV, Sintchenko V, Chen SC, Maddocks S, Kok J, Dwyer DE, Rockett R, Eden J-S, Lam C, Gray K, Timms V, Gall M, Arnott A, Sadsad R, Carter I, Rahman H and Holmes EC for the 2019-nCoV Study Group
EPI_ISL_417395 hCoV-19/Australia/NSW37/2020 Oceania / Australia / New South Wales / Sydney 2020-03-10 Centre for Infectious Diseases and Microbiology Public Health NSW Health Pathology - Institute of Clinical Pathology and Medical Research; Westmead Hospital; University of Sydney Sintchenko V, Chen SC, Maddocks S, Kok J, Dwyer DE, Rockett R, Eden J-S, Lam C, Gray K, Timms V, Gall M, Arnott A, Sadsad R, Carter I, Rahman H, Holmes EC and O'Sullivan MV for the 2019-nCoV Study Group
EPI_ISL_417396 hCoV-19/Australia/NSW38/2020 Oceania / Australia / New South Wales / Sydney 2020-03-10 Centre for Infectious Diseases and Microbiology Public Health NSW Health Pathology - Institute of Clinical Pathology and Medical Research; Westmead Hospital; University of Sydney Chen SC, Maddocks S, Kok J, Dwyer DE, Rockett R, Eden J-S, Lam C, Gray K, Timms V, Gall M, Arnott A, Sadsad R, Carter I, Rahman H, Holmes EC, O'Sullivan MV and Sintchenko V for the 2019-nCoV Study Group
EPI_ISL_417397 hCoV-19/Australia/NSW39/2020 Oceania / Australia / New South Wales / Sydney 2020-03-11 Centre for Infectious Diseases and Microbiology Public Health NSW Health Pathology - Institute of Clinical Pathology and Medical Research; Westmead Hospital; University of Sydney Maddocks S, Kok J, Dwyer DE, Rockett R, Eden J-S, Lam C, Gray K, Timms V, Gall M, Arnott A, Sadsad R, Carter I, Rahman H, Holmes EC, O'Sullivan MV, Sintchenko V and Chen SC for the 2019-nCoV Study Group
EPI_ISL_417398 hCoV-19/Australia/NSW40/2020 Oceania / Australia / New South Wales / Sydney 2020-03-11 Centre for Infectious Diseases and Microbiology Public Health NSW Health Pathology - Institute of Clinical Pathology and Medical Research; Westmead Hospital; University of Sydney Kok J, Dwyer DE, Rockett R, Eden J-S, Lam C, Gray K, Timms V, Gall M, Arnott A, Sadsad R, Carter I, Rahman H, Holmes EC, O'Sullivan MV, Sintchenko V, Chen SC and Maddocks S for the 2019-nCoV Study Group
EPI_ISL_417399 hCoV-19/Australia/NSW41/2020 Oceania / Australia / New South Wales / Sydney 2020-03-10 Centre for Infectious Diseases and Microbiology Public Health NSW Health Pathology - Institute of Clinical Pathology and Medical Research; Westmead Hospital; University of Sydney Dwyer DE, Rockett R, Eden J-S, Lam C, Gray K, Timms V, Gall M, Arnott A, Sadsad R, Carter I, Rahman H, Holmes EC, O'Sullivan MV, Sintchenko V, Chen SC, Maddocks S and Kok J for the 2019-nCoV Study Group
EPI_ISL_417408 hCoV-19/Australia/NSW56/2020 Oceania / Australia / New South Wales / Sydney 2020-03-17 Centre for Infectious Diseases and Microbiology Public Health NSW Health Pathology - Institute of Clinical Pathology and Medical Research; Westmead Hospital; University of Sydney Carter I, Rahman H, Holmes EC, O'Sullivan MV, Sintchenko V, Chen SC, Maddocks S, Kok J, Dwyer DE, Rockett R, Eden J-S, Lam C, Gray K, Timms V, Gall M, Arnott A and Sadsad R for the 2019-nCoV Study Group
EPI_ISL_417409 hCoV-19/Australia/NSW57/2020 Oceania / Australia / New South Wales / Sydney 2020-03-16 Centre for Infectious Diseases and Microbiology Public Health NSW Health Pathology - Institute of Clinical Pathology and Medical Research; Westmead Hospital; University of Sydney Rahman H, Holmes EC, O'Sullivan MV, Sintchenko V, Chen SC, Maddocks S, Kok J, Dwyer DE, Rockett R, Eden J-S, Lam C, Gray K, Timms V, Gall M, Arnott A, Sadsad R and Carter I for the 2019-nCoV Study Group
EPI_ISL_417410 hCoV-19/Australia/NSW58/2020 Oceania / Australia / New South Wales / Sydney 2020-03-16 Centre for Infectious Diseases and Microbiology Public Health NSW Health Pathology - Institute of Clinical Pathology and Medical Research; Westmead Hospital; University of Sydney Holmes EC, O'Sullivan MV, Sintchenko V, Chen SC, Maddocks S, Kok J, Dwyer DE, Rockett R, Eden J-S, Lam C, Gray K, Timms V, Gall M, Arnott A, Sadsad R, Carter I and Rahman H for the 2019-nCoV Study Group
EPI_ISL_417411 hCoV-19/Australia/NSW61/2020 Oceania / Australia / New South Wales / Sydney 2020-03-17 Centre for Infectious Diseases and Microbiology Public Health NSW Health Pathology - Institute of Clinical Pathology and Medical Research; Westmead Hospital; University of Sydney O'Sullivan MV, Sintchenko V, Chen SC, Maddocks S, Kok J, Dwyer DE, Rockett R, Eden J-S, Lam C, Gray K, Timms V, Gall M, Arnott A, Sadsad R, Carter I, Rahman H and Holmes EC for the 2019-nCoV Study Group
EPI_ISL_417412 hCoV-19/Australia/NSW62/2020 Oceania / Australia / New South Wales / Sydney 2020-03-17 Centre for Infectious Diseases and Microbiology Public Health NSW Health Pathology - Institute of Clinical Pathology and Medical Research; Westmead Hospital; University of Sydney Sintchenko V, Chen SC, Maddocks S, Kok J, Dwyer DE, Rockett R, Eden J-S, Lam C, Gray K, Timms V, Gall M, Arnott A, Sadsad R, Carter I, Rahman H, Holmes EC and O'Sullivan MV for the 2019-nCoV Study Group
EPI_ISL_417418 hCoV-19/Italy/FVG-ICGEB_S1/2020 Europe / Italy / Friuli Venezia Giulia 2020-03-01 Laboratory of Molecular Virology International Center fro Genetic Engineering and Biotechnology (ICGEB) ARGO Open Lab Platform for Genome sequencing Licastro D, Rajasekharan S, Dal Monego S, Segat L, D'Agaro P, Marcello A
EPI_ISL_417419 hCoV-19/Italy/FVG-ICGEB_S5/2020 Europe / Italy / Friuli Venezia Giulia 2020-03-01 Laboratory of Molecular Virology International Center for Genetic Engineering and Biotechnology (ICGEB) ARGO Open Lab Platform for Genome sequencing Licastro D, Rajasekharan S, Dal Monego S, Segat L, D'Agaro P, Marcello A
EPI_ISL_417420 hCoV-19/NanChang/JX216/2020 Asia / China / NanChang 2020-03-23 Jiangxi province Center for Disease Control and Prevention Jiangxi province Center for Disease Control and Prevention Li jian Xiong

EPI_ISL_417421 hCoV-19/Italy/FVG-ICGEB_S8/2020 Europe / Italy / Friuli Venezia Giulia 2020-03-01 Laboratory of Molecular Virology International Center for Genetic Engineering and Biotechnology (ICGEB) ARGO Open Lab Platform for Genome sequencing Licastro D, Rajasekharan S, Dal Monego S, Segat L, D'Agaro P, Marcello A
EPI_ISL_417422 hCoV-19/Belgium/DHWM-03041/2020 Europe / Belgium / Dilbeek 2020-03-04 KU Leuven, Clinical and Epidemiological Virology KU Leuven, Clinical and Epidemiological Virology Joan Marti-Carerras, Tony Wawina, Bert Vanmechelen, Piet Maes
EPI_ISL_417423 hCoV-19/Italy/FVG-ICGEB_S9/2020 Europe / Italy / Friuli Venezia Giulia 2020-03-01 Laboratory of Molecular Virology International Center for Genetic Engineering and Biotechnology (ICGEB) ARGO Open Lab Platform for Genome sequencing Licastro D, Rajasekharan, Dal Monego S, Segat L, D'Agaro P, Marcello A
EPI_ISL_417424 hCoV-19/Belgium/KA-03042/2020 Europe / Belgium / Kasterlee 2020-03-04 KU Leuven, Clinical and Epidemiological Virology KU Leuven, Clinical and Epidemiological Virology Joan Marti-Carerras, Tony Wawina, Bert Vanmechelen, Piet Maes
EPI_ISL_417425 hCoV-19/Belgium/VBK-03061/2020 Europe / Belgium / Schoten 2020-03-06 KU Leuven, Clinical and Epidemiological Virology KU Leuven, Clinical and Epidemiological Virology Joan Marti-Carerras, Tony Wawina, Bert Vanmechelen, Piet Maes
EPI_ISL_417426 hCoV-19/Belgium/SQ-03043/2020 Europe / Belgium / Leuven 2020-03-04 KU Leuven, Clinical and Epidemiological Virology KU Leuven, Clinical and Epidemiological Virology Joan Marti-Carerras, Tony Wawina, Bert Vanmechelen, Piet Maes
EPI_ISL_417427 hCoV-19/Belgium/VI-03027/2020 Europe / Belgium / Sint-Pieters-Woluwe 2020-03-02 KU Leuven, Clinical and Epidemiological Virology KU Leuven, Clinical and Epidemiological Virology Joan Marti-Carerras, Tony Wawina, Bert Vanmechelen, Piet Maes
EPI_ISL_417428 hCoV-19/Belgium/FR-03029/2020 Europe / Belgium / Rixensart 2020-03-02 KU Leuven, Clinical and Epidemiological Virology KU Leuven, Clinical and Epidemiological Virology Joan Marti-Carerras, Tony Wawina, Bert Vanmechelen, Piet Maes
EPI_ISL_417429 hCoV-19/Belgium/MMJ-03034/2020 Europe / Belgium / Holsbeek 2020-03-03 KU Leuven, Clinical and Epidemiological Virology KU Leuven, Clinical and Epidemiological Virology Joan Marti-Carerras, Tony Wawina, Bert Vanmechelen, Piet Maes
EPI_ISL_417430 hCoV-19/Belgium/RT-03035/2020 Europe / Belgium / Vorst 2020-03-03 KU Leuven, Clinical and Epidemiological Virology KU Leuven, Clinical and Epidemiological Virology Joan Marti-Carerras, Tony Wawina, Bert Vanmechelen, Piet Maes
EPI_ISL_417437 hCoV-19/DRC/KN-0054/2020 Africa / Democratic Republic of the Congo / Kinshasa 2020-03-17 Viral Respiratory Lab, National Institute for Biomedical Research (INRB) Pathogen Sequencing Lab, National Institute for Biomedical Research (INRB) Placide Mbala-Kingebeni, Edith Nkwembe, Eddy Kinganda-Lusamaki, Amuri Aziza, Catherine Pratt, Matthias Pauthner, Josh Quick, Allison Black, James Hadfield, Trevor Bedford, Ian Goodfellow, Nick Loman, Kristian Andersen, Michael Wiley, Steve Ahuka-Mundeke, Jean-Jacques Muyembe Tamfum
EPI_ISL_417438 hCoV-19/DRC/KN-0058/2020 Africa / Democratic Republic of the Congo / Kinshasa 2020-03-17 Viral Respiratory Lab, National Institute for Biomedical Research (INRB) Pathogen Sequencing Lab, National Institute for Biomedical Research (INRB) Placide Mbala-Kingebeni, Edith Nkwembe, Eddy Kinganda-Lusamaki, Amuri Aziza, Catherine Pratt, Matthias Pauthner, Josh Quick, Allison Black, James Hadfield, Trevor Bedford, Ian Goodfellow, Nick Loman, Kristian Andersen, Michael Wiley, Steve Ahuka-Mundeke, Jean-Jacques Muyembe Tamfum
EPI_ISL_417440 hCoV-19/DRC/KN-0060/2020 Africa / Democratic Republic of the Congo / Kinshasa 2020-03-17 Viral Respiratory Lab, National Institute for Biomedical Research (INRB) Pathogen Sequencing Lab, National Institute for Biomedical Research (INRB) Placide Mbala-Kingebeni, Edith Nkwembe, Eddy Kinganda-Lusamaki, Amuri Aziza, Catherine Pratt, Matthias Pauthner, Josh Quick, Allison Black, James Hadfield, Trevor Bedford, Ian Goodfellow, Nick Loman, Kristian Andersen, Michael Wiley, Steve Ahuka-Mundeke, Jean-Jacques Muyembe Tamfum
EPI_ISL_417442 hCoV-19/DRC/KN-0072/2020 Africa / Democratic Republic of the Congo / Kinshasa 2020-03-18 Viral Respiratory Lab, National Institute for Biomedical Research (INRB) Pathogen Sequencing Lab, National Institute for Biomedical Research (INRB) Placide Mbala-Kingebeni, Edith Nkwembe, Eddy Kinganda-Lusamaki, Amuri Aziza, Catherine Pratt, Matthias Pauthner, Josh Quick, Allison Black, James Hadfield, Trevor Bedford, Ian Goodfellow, Nick Loman, Kristian Andersen, Michael Wiley, Steve Ahuka-Mundeke, Jean-Jacques Muyembe Tamfum
EPI_ISL_417443 hCoV-19/Hong Kong/XM-PII-S4/2020 Asia / Hong Kong 2020-01-22 State Key Laboratory for Emerging Infectious Diseases Department of Microbiology Li Ka Shing Faculty of Medicine The University of Hong Kong State Key Laboratory for Emerging Infectious Diseases Department of Microbiology Li Ka Shing Faculty of Medicine The University of Hong Kong Pui Wang, Siu-Ying Lau, Shaofeng Deng, Bobo Wing-Yee Mok, Wenjun Song, Kwok-Yung Yuen, Honglin Chen
EPI_ISL_417444 hCoV-19/Pakistan/Gilgit1/2020 Asia / Pakistan / Gilgit 2020-03-04 unknown Department of Healthcare Biotechnology Javed,A., Niazi,S.K., Ghani,E., Saqib,M., Janjua,H.A., Corman,V.M. and Zohaib,A.
EPI_ISL_417445 hCoV-19/Italy/UniMI01/2020 Europe / Italy / Lombardy / Milan 2020-02-24 Laboratory of Infectious Diseases, Department of Biomedical and Clinical Sciences L. Sacco, University of Milan Laboratory of Infectious Diseases, Department of Biomedical and Clinical Sciences L. Sacco, University of Milan Gianguglielmo Zehender, Alessia Lai, Annalisa Bergna, Luca Meroni, Agostino Riva, Claudia Balotta, Maciej Tarkowski, Arianna Gabrieli, Dario Bernacchia, Stefano Rusconi, Giuliano Rizzardini, Spinello Antinori, Massimo Galli
EPI_ISL_417447 hCoV-19/Italy/UniMI03/2020 Europe / Italy / Lombardy / Milan 2020-02-24

Laboratory of Infectious Diseases, Department of Biomedical and Clinical Sciences L. Sacco, University of Milan
Laboratory of Infectious Diseases, Department of Biomedical and Clinical Sciences L. Sacco, University of Milan
Gianguglielmo Zehender, Alessia Lai, Annalisa Bergna, Luca Meroni, Agostino Riva, Claudia Balotta, Maciej Tarkowski, Arianna Gabrieli, Dario Bernacchia, Stefano Rusconi, Giuliano Rizzardini, Spinello Antinori, Massimo Galli
EPI_ISL_417448 hCoV-19/USA/WA-UW234/2020 North America / USA / Washington 2020-03-14 UW Virology Lab UW Virology Lab Pavitra Roychoudhury, Hong Xie, Keith Jerome, Alexander Greninger
EPI_ISL_417449 hCoV-19/USA/WA-UW235/2020 North America / USA / Washington 2020-03-14 UW Virology Lab UW Virology Lab Pavitra Roychoudhury, Hong Xie, Keith Jerome, Alexander Greninger
EPI_ISL_417450 hCoV-19/USA/WA-UW236/2020 North America / USA / Washington 2020-03-14 UW Virology Lab UW Virology Lab Pavitra Roychoudhury, Hong Xie, Keith Jerome, Alexander Greninger
EPI_ISL_417451 hCoV-19/USA/WA-UW237/2020 North America / USA / Washington 2020-03-14 UW Virology Lab UW Virology Lab Pavitra Roychoudhury, Hong Xie, Keith Jerome, Alexander Greninger
EPI_ISL_417452 hCoV-19/USA/CT-UW238/2020 North America / USA / Connecticut 2020-03-12 UW Virology Lab UW Virology Lab Pavitra Roychoudhury, Hong Xie, Keith Jerome, Alexander Greninger
EPI_ISL_417453 hCoV-19/USA/WA-UW239/2020 North America / USA / Washington 2020-03-14 UW Virology Lab UW Virology Lab Pavitra Roychoudhury, Hong Xie, Keith Jerome, Alexander Greninger
EPI_ISL_417454 hCoV-19/USA/WA-UW240/2020 North America / USA / Washington 2020-03-14 UW Virology Lab UW Virology Lab Pavitra Roychoudhury, Hong Xie, Keith Jerome, Alexander Greninger
EPI_ISL_417455 hCoV-19/USA/WA-UW241/2020 North America / USA / Washington 2020-03-14 UW Virology Lab UW Virology Lab Pavitra Roychoudhury, Hong Xie, Keith Jerome, Alexander Greninger
EPI_ISL_417456 hCoV-19/USA/WA-UW242/2020 North America / USA / Washington 2020-03-13 UW Virology Lab UW Virology Lab Pavitra Roychoudhury, Hong Xie, Keith Jerome, Alexander Greninger
EPI_ISL_417457 hCoV-19/Germany/NRW-12/2020 Europe / Germany / North Rhine Westphalia / Duesseldorf 2020-03-10 Center of Medical Microbiology, Virology, and Hospital Hygiene, University of Duesseldorf Center of Medical Microbiology, Virology, and Hospital Hygiene, University of Duesseldorf Ortwin Adams, Marcel Andree, Alexander Dilthey, Torsten Feldt, Sandra Hauka, Torsten Houwaart, Björn-Erik Jensen, Detlef Kindgen-Milles, Malte Kohns Vasconcelos, Klaus Pfeffer, Tina Senff, Daniel Strelow, Jörg Timm, Andreas Walker, Tobias Wienemann
EPI_ISL_417458 hCoV-19/Germany/NRW-13/2020 Europe / Germany / North Rhine Westphalia / Duesseldorf 2020-03-11 Center of Medical Microbiology, Virology, and Hospital Hygiene, University of Duesseldorf Center of Medical Microbiology, Virology, and Hospital Hygiene, University of Duesseldorf Ortwin Adams, Marcel Andree, Alexander Dilthey, Torsten Feldt, Sandra Hauka, Torsten Houwaart, Björn-Erik Jensen, Detlef Kindgen-Milles, Malte Kohns Vasconcelos, Klaus Pfeffer, Tina Senff, Daniel Strelow, Jörg Timm, Andreas Walker, Tobias Wienemann
EPI_ISL_417459 hCoV-19/Germany/NRW-14/2020 Europe / Germany / North Rhine Westphalia / Duesseldorf 2020-03-11 Center of Medical Microbiology, Virology, and Hospital Hygiene, University of Duesseldorf Center of Medical Microbiology, Virology, and Hospital Hygiene, University of Duesseldorf Ortwin Adams, Marcel Andree, Alexander Dilthey, Torsten Feldt, Sandra Hauka, Torsten Houwaart, Björn-Erik Jensen, Detlef Kindgen-Milles, Malte Kohns Vasconcelos, Klaus Pfeffer, Tina Senff, Daniel Strelow, Jörg Timm, Andreas Walker, Tobias Wienemann
EPI_ISL_417460 hCoV-19/Germany/NRW-15/2020 Europe / Germany / North Rhine Westphalia / Duesseldorf 2020-03-11 Center of Medical Microbiology, Virology, and Hospital Hygiene, University of Duesseldorf Center of Medical Microbiology, Virology, and Hospital Hygiene, University of Duesseldorf Ortwin Adams, Marcel Andree, Alexander Dilthey, Torsten Feldt, Sandra Hauka, Torsten Houwaart, Björn-Erik Jensen, Detlef Kindgen-Milles, Malte Kohns Vasconcelos, Klaus Pfeffer, Tina Senff, Daniel Strelow, Jörg Timm, Andreas Walker, Tobias Wienemann
EPI_ISL_417461 hCoV-19/Germany/NRW-16/2020 Europe / Germany / North Rhine Westphalia / Duesseldorf 2020-03-11 Center of Medical Microbiology, Virology, and Hospital Hygiene, University of Duesseldorf Center of Medical Microbiology, Virology, and Hospital Hygiene, University of Duesseldorf Ortwin Adams, Marcel Andree, Alexander Dilthey, Torsten Feldt, Sandra Hauka, Torsten Houwaart, Björn-Erik Jensen, Detlef Kindgen-Milles, Malte Kohns Vasconcelos, Klaus Pfeffer, Tina Senff, Daniel Strelow, Jörg Timm, Andreas Walker, Tobias Wienemann
EPI_ISL_417462 hCoV-19/Germany/NRW-17/2020 Europe / Germany / North Rhine Westphalia / Duesseldorf 2020-03-11 Center of Medical Microbiology, Virology, and Hospital Hygiene, University of Duesseldorf Center of Medical Microbiology, Virology, and Hospital Hygiene, University of Duesseldorf Ortwin Adams, Marcel Andree, Alexander Dilthey, Torsten Feldt, Sandra Hauka, Torsten Houwaart, Björn-Erik Jensen, Detlef Kindgen-Milles, Malte Kohns Vasconcelos, Klaus Pfeffer, Tina Senff, Daniel Strelow, Jörg Timm, Andreas Walker, Tobias Wienemann
EPI_ISL_417463 hCoV-19/Germany/NRW-18/2020 Europe / Germany / North Rhine Westphalia / Duesseldorf 2020-03-13 Center of Medical Microbiology, Virology, and Hospital Hygiene, University of Duesseldorf Center of Medical Microbiology, Virology, and Hospital Hygiene, University of Duesseldorf Ortwin Adams, Marcel Andree, Alexander Dilthey, Torsten Feldt, Sandra Hauka, Torsten Houwaart, Björn-Erik Jensen, Detlef Kindgen-Milles, Malte Kohns Vasconcelos, Klaus Pfeffer, Tina Senff, Daniel Strelow, Jörg Timm, Andreas Walker, Tobias Wienemann
EPI_ISL_417464 hCoV-19/Germany/NRW-19/2020 Europe / Germany / North Rhine Westphalia / Duesseldorf 2020-03-14 Center of Medical Microbiology, Virology, and Hospital Hygiene, University of Duesseldorf Center of Medical Microbiology, Virology, and Hospital Hygiene, University of Duesseldorf Ortwin Adams, Marcel Andree, Alexander Dilthey, Torsten Feldt, Sandra Hauka, Torsten Houwaart, Björn-Erik Jensen, Detlef Kindgen-Milles, Malte Kohns Vasconcelos, Klaus Pfeffer, Tina Senff, Daniel Strelow, Jörg Timm, Andreas Walker, Tobias

Wienemann
EPI_ISL_417465 hCoV-19/Germany/NRW-20/2020 Europe / Germany / North Rhine Westphalia / Duesseldorf 2020-03-14 Center of Medical Microbiology, Virology, and Hospital Hygiene, University of Duesseldorf Center of Medical Microbiology, Virology, and Hospital Hygiene, University of Duesseldorf Ortwin Adams, Marcel Andree, Alexander Dilthey, Torsten Feldt, Sandra Hauka, Torsten Houwaart, Björn-Erik Jensen, Detlef Kindgen-Milles, Malte Kohns Vasconcelos, Klaus Pfeffer, Tina Senff, Daniel Strelow, Jörg Timm, Andreas Walker, Tobias Wienemann
EPI_ISL_417466 hCoV-19/Germany/NRW-21/2020 Europe / Germany / North Rhine Westphalia / Duesseldorf 2020-03-14 Center of Medical Microbiology, Virology, and Hospital Hygiene, University of Duesseldorf Center of Medical Microbiology, Virology, and Hospital Hygiene, University of Duesseldorf Ortwin Adams, Marcel Andree, Alexander Dilthey, Torsten Feldt, Sandra Hauka, Torsten Houwaart, Björn-Erik Jensen, Detlef Kindgen-Milles, Malte Kohns Vasconcelos, Klaus Pfeffer, Tina Senff, Daniel Strelow, Jörg Timm, Andreas Walker, Tobias Wienemann
EPI_ISL_417467 hCoV-19/Germany/NRW-22/2020 Europe / Germany / North Rhine Westphalia / Duesseldorf 2020-03-15 Center of Medical Microbiology, Virology, and Hospital Hygiene, University of Duesseldorf Center of Medical Microbiology, Virology, and Hospital Hygiene, University of Duesseldorf Ortwin Adams, Marcel Andree, Alexander Dilthey, Torsten Feldt, Sandra Hauka, Torsten Houwaart, Björn-Erik Jensen, Detlef Kindgen-Milles, Malte Kohns Vasconcelos, Klaus Pfeffer, Tina Senff, Daniel Strelow, Jörg Timm, Andreas Walker, Tobias Wienemann
EPI_ISL_417468 hCoV-19/Germany/NRW-23/2020 Europe / Germany / North Rhine Westphalia / Duesseldorf 2020-03-16 Center of Medical Microbiology, Virology, and Hospital Hygiene, University of Duesseldorf Center of Medical Microbiology, Virology, and Hospital Hygiene, University of Duesseldorf Ortwin Adams, Marcel Andree, Alexander Dilthey, Torsten Feldt, Sandra Hauka, Torsten Houwaart, Björn-Erik Jensen, Detlef Kindgen-Milles, Malte Kohns Vasconcelos, Klaus Pfeffer, Tina Senff, Daniel Strelow, Jörg Timm, Andreas Walker, Tobias Wienemann
EPI_ISL_417469 hCoV-19/USA/MN31-MDH31/2020 North America / USA / Minnesota 2020-03-12 Minnesota Department of Health, Public Health Laboratory Minnesota Department of Health, Public Health Laboratory Matt Plumb, Jake Garfin and Xiong Wang
EPI_ISL_417470 hCoV-19/USA/MN32-MDH32/2020 North America / USA / Minnesota 2020-03-12 Minnesota Department of Health, Public Health Laboratory Minnesota Department of Health, Public Health Laboratory Matt Plumb, Jake Garfin and Xiong Wang
EPI_ISL_417471 hCoV-19/USA/MN34-MDH34/2020 North America / USA / Minnesota 2020-03-12 Minnesota Department of Health, Public Health Laboratory Minnesota Department of Health, Public Health Laboratory Matt Plumb, Jake Garfin and Xiong Wang
EPI_ISL_417472 hCoV-19/USA/MN35-MDH35/2020 North America / USA / Minnesota 2020-03-12 Minnesota Department of Health, Public Health Laboratory Minnesota Department of Health, Public Health Laboratory Matt Plumb, Jake Garfin and Xiong Wang
EPI_ISL_417473 hCoV-19/USA/MN36-MDH36/2020 North America / USA / Minnesota 2020-03-13 Minnesota Department of Health, Public Health Laboratory Minnesota Department of Health, Public Health Laboratory Matt Plumb, Jake Garfin and Xiong Wang
EPI_ISL_417474 hCoV-19/USA/MN38-MDH38/2020 North America / USA / Minnesota 2020-03-12 Minnesota Department of Health, Public Health Laboratory Minnesota Department of Health, Public Health Laboratory Matt Plumb, Jake Garfin and Xiong Wang
EPI_ISL_417475 hCoV-19/USA/MN39-MDH39/2020 North America / USA / Minnesota 2020-03-12 Minnesota Department of Health, Public Health Laboratory Minnesota Department of Health, Public Health Laboratory Matt Plumb, Jake Garfin and Xiong Wang
EPI_ISL_417476 hCoV-19/USA/MN40-MDH40/2020 North America / USA / Minnesota 2020-03-12 Minnesota Department of Health, Public Health Laboratory Minnesota Department of Health, Public Health Laboratory Matt Plumb, Jake Garfin and Xiong Wang
EPI_ISL_417477 hCoV-19/USA/MN41-MDH41/2020 North America / USA / Minnesota 2020-03-12 Minnesota Department of Health, Public Health Laboratory Minnesota Department of Health, Public Health Laboratory Matt Plumb, Jake Garfin and Xiong Wang
EPI_ISL_417478 hCoV-19/USA/MN42-MDH42/2020 North America / USA / Minnesota 2020-03-13 Minnesota Department of Health, Public Health Laboratory Minnesota Department of Health, Public Health Laboratory Matt Plumb, Jake Garfin and Xiong Wang
EPI_ISL_417479 hCoV-19/USA/MN43-MDH43/2020 North America / USA / Minnesota 2020-03-12 Minnesota Department of Health, Public Health Laboratory Minnesota Department of Health, Public Health Laboratory Matt Plumb, Jake Garfin and Xiong Wang
EPI_ISL_417480 hCoV-19/USA/MN44-MDH44/2020 North America / USA / Minnesota 2020-03-12 Minnesota Department of Health, Public Health Laboratory Minnesota Department of Health, Public Health Laboratory Matt Plumb, Jake Garfin and Xiong Wang
EPI_ISL_417481 hCoV-19/Iceland/LUH-01/2020 Europe / Iceland / Reykjavik 2020-03-13 deCODE genetics deCODE genetics Daniel F Gudbjartsson, Agnar Helgason, Hakon Jonsson, Olafur T Magnusson, Pall Melsted, Gudmundur L Norddahl, Jona Saemundsdottir, Asgeir Sigurdsson, Patrick Sulem, Arna B Agustsdottir, Berglind Eiriksdottir, Run Fridriksdottir, Elisabet E Gardarsdottir, Gudmundur Georgsson, Olafia S Gretarsdottir, Kjartan R Gudmundsson, Thora R Gunnarsdottir, Arnaldur Gylfason, Hilma Holm, Brynjar O Jensson, Aslaug Jonasdottir, Kamilla S Josefsdottir, Thordur Kristjansson, Droplaug N Magnusdottir, Louise le Roux, Gudrun Sigmundsdottir, Gardar Sveinbjornsson, Kristin E Sveinsdottir, Maney Sveinsdottir, Emil A Thorarensen, Bjarni Thorbjornsson, Gisli Masson, Ingileif Jonsdottir, Alma Moller, Thorolfur Gudnason, Karl G Kristinsson, Unnur Thorsteinsdottir, Kari Stefansson

EPI_ISL_417483  hCoV-19/Norway/1379/2020        Europe / Norway / Oslo  2020-02-27      Oslo University Hospital, Department of Medical Microbiology    Norwegian Institute of Public Health    Kathrine Stene-Johansen, Kamilla Heddeland Instefjord, Hilde Elshaug, Karoline Bragstad, Olav Hungnes
EPI_ISL_417484  hCoV-19/Norway/1380/2020        Europe / Norway / Oslo  2020-02-26      Oslo University Hospital, Department of Medical Microbiology    Norwegian Institute of Public Health, Department of Virology    Kathrine Stene-Johansen, Kamilla Heddeland Instefjord, Hilde Elshaug, Karoline Bragstad, Olav Hungnes
EPI_ISL_417485  hCoV-19/Norway/1526/2020        Europe / Norway / Oslo  2020-03-02      University Hospital of Northern Norway, Department for Microbiology and Infectious Disease Control     Norwegian Institute of Public Health, Department of Virology  Kathrine Stene-Johansen, Kamilla Heddeland Instefjord, Hilde Elshaug, Karoline Bragstad, Olav Hungnes
EPI_ISL_417486  hCoV-19/Norway/1538/2020        Europe / Norway / Oslo  2020-03-01      Hospital of Southern Norway - Kristiansand, Department of Medical Microbiology       Norwegian Institute of Public Health, Department of Virology  Kathrine Stene-Johansen, Kamilla Heddeland Instefjord, Hilde Elshaug, Karoline Bragstad, Olav Hungnes
EPI_ISL_417487  hCoV-19/Norway/1539/2020        Europe / Norway / Oslo  2020-03-01      Hospital of Southern Norway - Kristiansand, Department of Medical Microbiology       Norwegian Institute of Public Health, Department of Virology  Kathrine Stene-Johansen, Kamilla Heddeland Instefjord, Hilde Elshaug, Karoline Bragstad, Olav Hungnes
EPI_ISL_417488  hCoV-19/Norway/1493/2020        Europe / Norway / Oslo  2020-02-29      Oslo University Hospital, Department of Medical Microbiology    Norwegian Institute of Public Health, Department of Virology    Kathrine Stene-Johansen, Kamilla Heddeland Instefjord, Hilde Elshaug, Karoline Bragstad, Olav Hungnes
EPI_ISL_417491  hCoV-19/Italy/UnivPM1/2020      Europe / Italy / Marche 2020-03-03      Virology Laboratory, Department of Biomedical Sciences and Public Health, University Politecnica delle Marche        Virology and Legal Medicine Laboratories, Department of Biomedical Sciences and Public Health, University Politecnica delle Marche  Bagnarelli,P., Caucci,S., Di Sante,L., Menzo,S., Alessandrini,F., Onofri,V., Turchi,C., Tagliabracci,A.
EPI_ISL_417492  hCoV-19/USA/MN45-MDH45/2020     North America / USA / Minnesota 2020-03-13      Minnesota Department of Health, Public Health Laboratory  Minnesota Department of Health, Public Health Laboratory    Matt Plumb, Jake Garfin and Xiong Wang
EPI_ISL_417493  hCoV-19/USA/MN46-MDH46/2020     North America / USA / Minnesota 2020-03-13      Minnesota Department of Health, Public Health Laboratory  Minnesota Department of Health, Public Health Laboratory    Matt Plumb, Jake Garfin and Xiong Wang
EPI_ISL_417494  hCoV-19/USA/MN47-MDH47/2020     North America / USA / Minnesota 2020-03-13      Minnesota Department of Health, Public Health Laboratory  Minnesota Department of Health, Public Health Laboratory    Matt Plumb, Jake Garfin and Xiong Wang
EPI_ISL_417495  hCoV-19/USA/MN48-MDH48/2020     North America / USA / Minnesota 2020-03-13      Minnesota Department of Health, Public Health Laboratory  Minnesota Department of Health, Public Health Laboratory    Matt Plumb, Jake Garfin and Xiong Wang
EPI_ISL_417496  hCoV-19/USA/MN51-MDH51/2020     North America / USA / Minnesota 2020-03-13      Minnesota Department of Health, Public Health Laboratory  Minnesota Department of Health, Public Health Laboratory    Matt Plumb, Jake Garfin and Xiong Wang
EPI_ISL_417497  hCoV-19/USA/MN52-MDH52/2020     North America / USA / Minnesota 2020-03-13      Minnesota Department of Health, Public Health Laboratory  Minnesota Department of Health, Public Health Laboratory    Matt Plumb, Jake Garfin and Xiong Wang
EPI_ISL_417498  hCoV-19/USA/MN53-MDH53/2020     North America / USA / Minnesota 2020-03-13      Minnesota Department of Health, Public Health Laboratory  Minnesota Department of Health, Public Health Laboratory    Matt Plumb, Jake Garfin and Xiong Wang
EPI_ISL_417499  hCoV-19/USA/MN55-MDH55/2020     North America / USA / Minnesota 2020-03-13      Minnesota Department of Health, Public Health Laboratory  Minnesota Department of Health, Public Health Laboratory    Matt Plumb, Jake Garfin and Xiong Wang
EPI_ISL_418624  hCoV-19/Belgium/ULG-6972/2020   Europe / Belgium / Liège        2020-03-15      Department of Clinical Microbiology   GIGA Medical Genomics   Keith Durkin, Maria Artesi, Sébastien Bontems, Raphaël Boreux, Cécile Meex, Pierrette Melin, Marie-Pierre Hayette, Vincent Bours.
EPI_ISL_418625  hCoV-19/Belgium/ULG-6688/2020   Europe / Belgium / Liège        2020-03-14      Department of Clinical Microbiology   GIGA Medical Genomics   Keith Durkin, Maria Artesi, Sébastien Bontems, Raphaël Boreux, Cécile Meex, Pierrette Melin, Marie-Pierre Hayette, Vincent Bours.
EPI_ISL_418626  hCoV-19/Belgium/ULG-7242/2020   Europe / Belgium / Liège        2020-03-16      Department of Clinical Microbiology   GIGA Medical Genomics   Keith Durkin, Maria Artesi, Sébastien Bontems, Raphaël Boreux, Cécile Meex, Pierrette Melin, Marie-Pierre Hayette, Vincent Bours.
EPI_ISL_418627  hCoV-19/Belgium/ULG-6743/2020   Europe / Belgium / Liège        2020-03-14      Department of Clinical Microbiology   GIGA Medical Genomics   Keith Durkin, Maria Artesi, Sébastien Bontems, Raphaël Boreux, Cécile Meex, Pierrette Melin, Marie-Pierre Hayette, Vincent Bours.
EPI_ISL_418628  hCoV-19/Belgium/ULG-7134/2020   Europe / Belgium / Liège        2020-03-16      Department of Clinical Microbiology   GIGA Medical Genomics   Keith Durkin, Maria Artesi, Sébastien Bontems, Raphaël Boreux, Cécile Meex, Pierrette Melin, Marie-Pierre Hayette, Vincent Bours.
EPI_ISL_418629  hCoV-19/Belgium/ULG-7773/2020   Europe / Belgium / Liège        2020-03-17      Department of Clinical Microbiology   GIGA Medical Genomics   Keith Durkin, Maria Artesi, Sébastien Bontems, Raphaël Boreux, Cécile Meex, Pierrette Melin, Marie-Pierre Hayette, Vincent Bours.
EPI_ISL_418630  hCoV-19/Belgium/ULG-7729/2020   Europe / Belgium / Liège        2020-03-17      Department of Clinical Microbiology   GIGA Medical Genomics   Keith Durkin, Maria Artesi, Sébastien Bontems, Raphaël Boreux, Cécile Meex, Pierrette Melin, Marie-Pierre Hayette, Vincent Bours.

EPI_ISL_418631  hCoV-19/Belgium/ULG-6649/2020   Europe / Belgium / Liège        2020-03-14      Department of Clinical Microbiology   GIGA Medical Genomics   Keith Durkin, Maria Artesi, Sébastien Bontems, Raphaël Boreux, Cécile Meex, Pierrette Melin, Marie-Pierre Hayette, Vincent Bours.
EPI_ISL_418632  hCoV-19/Belgium/ULG-7500/2020   Europe / Belgium / Liège        2020-03-16      Department of Clinical Microbiology   GIGA Medical Genomics   Keith Durkin, Maria Artesi, Sébastien Bontems, Raphaël Boreux, Cécile Meex, Pierrette Melin, Marie-Pierre Hayette, Vincent Bours.
EPI_ISL_418633  hCoV-19/Belgium/ULG-7626/2020   Europe / Belgium / Liège        2020-03-17      Department of Clinical Microbiology   GIGA Medical Genomics   Keith Durkin, Maria Artesi, Sébastien Bontems, Raphaël Boreux, Cécile Meex, Pierrette Melin, Marie-Pierre Hayette, Vincent Bours.
EPI_ISL_418634  hCoV-19/Belgium/ULG-8595/2020   Europe / Belgium / Liège        2020-03-19      Department of Clinical Microbiology   GIGA Medical Genomics   Keith Durkin, Maria Artesi, Sébastien Bontems, Raphaël Boreux, Cécile Meex, Pierrette Melin, Marie-Pierre Hayette, Vincent Bours.
EPI_ISL_418635  hCoV-19/Belgium/ULG-8532/2020   Europe / Belgium / Liège        2020-03-19      Department of Clinical Microbiology   GIGA Medical Genomics   Keith Durkin, Maria Artesi, Sébastien Bontems, Raphaël Boreux, Cécile Meex, Pierrette Melin, Marie-Pierre Hayette, Vincent Bours.
EPI_ISL_418636  hCoV-19/Belgium/ULG-8541/2020   Europe / Belgium / Liège        2020-03-19      Department of Clinical Microbiology   GIGA Medical Genomics   Keith Durkin, Maria Artesi, Sébastien Bontems, Raphaël Boreux, Cécile Meex, Pierrette Melin, Marie-Pierre Hayette, Vincent Bours.
EPI_ISL_418638  hCoV-19/Belgium/ULG-8578/2020   Europe / Belgium / Liège        2020-03-19      Department of Clinical Microbiology   GIGA Medical Genomics   Keith Durkin, Maria Artesi, Sébastien Bontems, Raphaël Boreux, Cécile Meex, Pierrette Melin, Marie-Pierre Hayette, Vincent Bours.
EPI_ISL_418639  hCoV-19/Belgium/ULG-8617/2020   Europe / Belgium / Liège        2020-03-19      Department of Clinical Microbiology   GIGA Medical Genomics   Keith Durkin, Maria Artesi, Sébastien Bontems, Raphaël Boreux, Cécile Meex, Pierrette Melin, Marie-Pierre Hayette, Vincent Bours.
EPI_ISL_418640  hCoV-19/Belgium/ULG-8634/2020   Europe / Belgium / Liège        2020-03-19      Department of Clinical Microbiology   GIGA Medical Genomics   Keith Durkin, Maria Artesi, Sébastien Bontems, Raphaël Boreux, Cécile Meex, Pierrette Melin, Marie-Pierre Hayette, Vincent Bours.
EPI_ISL_418645  hCoV-19/Belgium/ULG-9558/2020   Europe / Belgium / Liège        2020-03-21      Department of Clinical Microbiology   GIGA Medical Genomics   Keith Durkin, Maria Artesi, Sébastien Bontems, Raphaël Boreux, Cécile Meex, Pierrette Melin, Marie-Pierre Hayette, Vincent Bours.
EPI_ISL_418646  hCoV-19/Belgium/ULG-9572/2020   Europe / Belgium / Liège        2020-03-21      Department of Clinical Microbiology   GIGA Medical Genomics   Keith Durkin, Maria Artesi, Sébastien Bontems, Raphaël Boreux, Cécile Meex, Pierrette Melin, Marie-Pierre Hayette, Vincent Bours.
EPI_ISL_418648  hCoV-19/Belgium/ULG-9618/2020   Europe / Belgium / Liège        2020-03-22      Department of Clinical Microbiology   GIGA Medical Genomics   Keith Durkin, Maria Artesi, Sébastien Bontems, Raphaël Boreux, Cécile Meex, Pierrette Melin, Marie-Pierre Hayette, Vincent Bours.
EPI_ISL_418649  hCoV-19/Belgium/ULG-9619/2020   Europe / Belgium / Liège        2020-03-22      Department of Clinical Microbiology   GIGA Medical Genomics   Keith Durkin, Maria Artesi, Sébastien Bontems, Raphaël Boreux, Cécile Meex, Pierrette Melin, Marie-Pierre Hayette, Vincent Bours.
EPI_ISL_418650  hCoV-19/Belgium/ULG-9620/2020   Europe / Belgium / Liège        2020-03-22      Department of Clinical Microbiology   GIGA Medical Genomics   Keith Durkin, Maria Artesi, Sébastien Bontems, Raphaël Boreux, Cécile Meex, Pierrette Melin, Marie-Pierre Hayette, Vincent Bours.
EPI_ISL_418651  hCoV-19/Belgium/ULG-9634/2020   Europe / Belgium / Liège        2020-03-22      Department of Clinical Microbiology   GIGA Medical Genomics   Keith Durkin, Maria Artesi, Sébastien Bontems, Raphaël Boreux, Cécile Meex, Pierrette Melin, Marie-Pierre Hayette, Vincent Bours.
EPI_ISL_418652  hCoV-19/Belgium/ULG-9641/2020   Europe / Belgium / Liège        2020-03-22      Department of Clinical Microbiology   GIGA Medical Genomics   Keith Durkin, Maria Artesi, Sébastien Bontems, Raphaël Boreux, Cécile Meex, Pierrette Melin, Marie-Pierre Hayette, Vincent Bours.
EPI_ISL_418653  hCoV-19/Belgium/ULG-9647/2020   Europe / Belgium / Liège        2020-03-22      Department of Clinical Microbiology   GIGA Medical Genomics   Keith Durkin, Maria Artesi, Sébastien Bontems, Raphaël Boreux, Cécile Meex, Pierrette Melin, Marie-Pierre Hayette, Vincent Bours.
EPI_ISL_418654  hCoV-19/Belgium/ULG-9694/2020   Europe / Belgium / Liège        2020-03-22      Department of Clinical Microbiology   GIGA Medical Genomics   Keith Durkin, Maria Artesi, Sébastien Bontems, Raphaël Boreux, Cécile Meex, Pierrette Melin, Marie-Pierre Hayette, Vincent Bours.
EPI_ISL_418655  hCoV-19/Belgium/ULG-9714/2020   Europe / Belgium / Liège        2020-03-22      Department of Clinical Microbiology   GIGA Medical Genomics   Keith Durkin, Maria Artesi, Sébastien Bontems, Raphaël Boreux, Cécile Meex, Pierrette Melin, Marie-Pierre Hayette, Vincent Bours.
EPI_ISL_418656  hCoV-19/Belgium/ULG-9715/2020   Europe / Belgium / Liège        2020-03-22      Department of Clinical Microbiology   GIGA Medical Genomics   Keith Durkin, Maria Artesi, Sébastien Bontems, Raphaël Boreux, Cécile Meex, Pierrette Melin, Marie-Pierre Hayette, Vincent Bours.
EPI_ISL_418657  hCoV-19/Belgium/ULG-9716/2020   Europe / Belgium / Liège        2020-03-22      Department of Clinical Microbiology   GIGA Medical Genomics   Keith Durkin, Maria Artesi, Sébastien Bontems, Raphaël Boreux, Cécile Meex, Pierrette Melin, Marie-Pierre Hayette, Vincent Bours.
EPI_ISL_418659  hCoV-19/Belgium/ULG-9725/2020   Europe / Belgium / Liège        2020-03-22      Department of Clinical Microbiology   GIGA Medical Genomics   Keith Durkin, Maria Artesi, Sébastien Bontems, Raphaël Boreux, Cécile Meex, Pierrette Melin, Marie-Pierre Hayette, Vincent Bours.
EPI_ISL_418660  hCoV-19/Belgium/ULG-9732/2020   Europe / Belgium / Liège        2020-03-22      Department of Clinical Microbiology   GIGA Medical Genomics   Keith Durkin, Maria Artesi, Sébastien Bontems, Raphaël Boreux, Cécile Meex, Pierrette Melin, Marie-Pierre Hayette, Vincent Bours.
EPI_ISL_418661  hCoV-19/Belgium/ULG-9735/2020   Europe / Belgium / Liège        2020-03-22      Department of

Clinical Microbiology   GIGA Medical Genomics   Keith Durkin, Maria Artesi, Sébastien Bontems, Raphaël Boreux, Cécile Meex, Pierrette Melin, Marie-Pierre Hayette, Vincent Bours.
EPI_ISL_418663  hCoV-19/Belgium/ULG-9739/2020   Europe / Belgium / Liège   2020-03-22   Department of Clinical Microbiology   GIGA Medical Genomics   Keith Durkin, Maria Artesi, Sébastien Bontems, Raphaël Boreux, Cécile Meex, Pierrette Melin, Marie-Pierre Hayette, Vincent Bours.
EPI_ISL_418664  hCoV-19/Belgium/ULG-9741/2020   Europe / Belgium / Liège   2020-03-22   Department of Clinical Microbiology   GIGA Medical Genomics   Keith Durkin, Maria Artesi, Sébastien Bontems, Raphaël Boreux, Cécile Meex, Pierrette Melin, Marie-Pierre Hayette, Vincent Bours.
EPI_ISL_418667  hCoV-19/England/20109047803/2020   Europe / United Kingdom / England   2020-03-07   Respiratory Virus Unit, Microbiology Services Colindale, Public Health England  Respiratory Virus Unit, Microbiology Services Colindale, Public Health England  Monica Galiano, Shahjahan Miah, Angie Lackenby, Omolola Akinbami, Tiina Talts, Leena Bhaw, Richard Myers, Steven Platt, Kirstin Edwards, Jonathan Hubb, Joanna Ellis, Maria Zambon
EPI_ISL_418668  hCoV-19/England/20110136302/2020   Europe / United Kingdom / England   2020-03-09   Respiratory Virus Unit, Microbiology Services Colindale, Public Health England  Respiratory Virus Unit, Microbiology Services Colindale, Public Health England  Monica Galiano, Shahjahan Miah, Angie Lackenby, Omolola Akinbami, Tiina Talts, Leena Bhaw, Richard Myers, Steven Platt, Kirstin Edwards, Jonathan Hubb, Joanna Ellis, Maria Zambon
EPI_ISL_418669  hCoV-19/England/20116022902/2020   Europe / United Kingdom / England   2020-03-11   Respiratory Virus Unit, Microbiology Services Colindale, Public Health England  Respiratory Virus Unit, Microbiology Services Colindale, Public Health England  Monica Galiano, Shahjahan Miah, Angie Lackenby, Omolola Akinbami, Tiina Talts, Leena Bhaw, Richard Myers, Steven Platt, Kirstin Edwards, Jonathan Hubb, Joanna Ellis, Maria Zambon
EPI_ISL_418670  hCoV-19/England/20116025603/2020   Europe / United Kingdom / England   2020-03-11   Respiratory Virus Unit, Microbiology Services Colindale, Public Health England  Respiratory Virus Unit, Microbiology Services Colindale, Public Health England  Monica Galiano, Shahjahan Miah, Angie Lackenby, Omolola Akinbami, Tiina Talts, Leena Bhaw, Richard Myers, Steven Platt, Kirstin Edwards, Jonathan Hubb, Joanna Ellis, Maria Zambon
EPI_ISL_418671  hCoV-19/England/20119026102/2020   Europe / United Kingdom / England   2020-03-13   Respiratory Virus Unit, Microbiology Services Colindale, Public Health England  Respiratory Virus Unit, Microbiology Services Colindale, Public Health England  Monica Galiano, Shahjahan Miah, Angie Lackenby, Omolola Akinbami, Tiina Talts, Leena Bhaw, Richard Myers, Steven Platt, Kirstin Edwards, Jonathan Hubb, Joanna Ellis, Maria Zambon
EPI_ISL_418672  hCoV-19/England/20120002102/2020   Europe / United Kingdom / England   2020-03-15   Respiratory Virus Unit, Microbiology Services Colindale, Public Health England  Respiratory Virus Unit, Microbiology Services Colindale, Public Health England  Monica Galiano, Shahjahan Miah, Angie Lackenby, Omolola Akinbami, Tiina Talts, Leena Bhaw, Richard Myers, Steven Platt, Kirstin Edwards, Jonathan Hubb, Joanna Ellis, Maria Zambon
EPI_ISL_418673  hCoV-19/England/20122032802/2020   Europe / United Kingdom / England   2020-03-16   Respiratory Virus Unit, Microbiology Services Colindale, Public Health England  Respiratory Virus Unit, Microbiology Services Colindale, Public Health England  Monica Galiano, Shahjahan Miah, Angie Lackenby, Omolola Akinbami, Tiina Talts, Leena Bhaw, Richard Myers, Steven Platt, Kirstin Edwards, Jonathan Hubb, Joanna Ellis, Maria Zambon
EPI_ISL_418675  hCoV-19/England/20122073102/2020   Europe / United Kingdom / England   2020-03-16   Respiratory Virus Unit, Microbiology Services Colindale, Public Health England  Respiratory Virus Unit, Microbiology Services Colindale, Public Health England  Monica Galiano, Shahjahan Miah, Angie Lackenby, Omolola Akinbami, Tiina Talts, Leena Bhaw, Richard Myers, Steven Platt, Kirstin Edwards, Jonathan Hubb, Joanna Ellis, Maria Zambon
EPI_ISL_418676  hCoV-19/England/20122074602/2020   Europe / United Kingdom / England   2020-03-10   Respiratory Virus Unit, Microbiology Services Colindale, Public Health England  Respiratory Virus Unit, Microbiology Services Colindale, Public Health England  Monica Galiano, Shahjahan Miah, Angie Lackenby, Omolola Akinbami, Tiina Talts, Leena Bhaw, Richard Myers, Steven Platt, Kirstin Edwards, Jonathan Hubb, Joanna Ellis, Maria Zambon
EPI_ISL_418677  hCoV-19/England/20122087702/2020   Europe / United Kingdom / England   2020-03-15   Respiratory Virus Unit, Microbiology Services Colindale, Public Health England  Respiratory Virus Unit, Microbiology Services Colindale, Public Health England  Monica Galiano, Shahjahan Miah, Angie Lackenby, Omolola Akinbami, Tiina Talts, Leena Bhaw, Richard Myers, Steven Platt, Kirstin Edwards, Jonathan Hubb, Joanna Ellis, Maria Zambon
EPI_ISL_418678  hCoV-19/England/20122088102/2020   Europe / United Kingdom / England   2020-03-16   Respiratory Virus Unit, Microbiology Services Colindale, Public Health England  Respiratory Virus Unit, Microbiology Services Colindale, Public Health England  Monica Galiano, Shahjahan Miah, Angie Lackenby, Omolola Akinbami, Tiina Talts, Leena Bhaw, Richard Myers, Steven Platt, Kirstin Edwards, Jonathan Hubb, Joanna Ellis, Maria Zambon
EPI_ISL_418679  hCoV-19/England/20122119202/2020   Europe / United Kingdom / England   2020-03-17   Respiratory Virus Unit, Microbiology Services Colindale, Public Health England  Respiratory Virus Unit, Microbiology Services Colindale, Public Health England  Monica Galiano, Shahjahan Miah, Angie Lackenby, Omolola Akinbami, Tiina Talts, Leena Bhaw, Richard Myers, Steven Platt, Kirstin Edwards, Jonathan Hubb, Joanna Ellis, Maria Zambon
EPI_ISL_418680  hCoV-19/England/20122119302/2020   Europe / United Kingdom / England   2020-03-17   Respiratory Virus Unit, Microbiology Services Colindale, Public Health England  Respiratory Virus Unit,

Microbiology Services Colindale, Public Health England  Monica Galiano, Shahjahan Miah, Angie Lackenby, Omolola Akinbami, Tiina Talts, Leena Bhaw, Richard Myers, Steven Platt, Kirstin Edwards, Jonathan Hubb, Joanna Ellis, Maria Zambon
EPI_ISL_418681  hCoV-19/England/20122119502/2020  Europe / United Kingdom / England  2020-03-17  Respiratory Virus Unit, Microbiology Services Colindale, Public Health England  Respiratory Virus Unit, Microbiology Services Colindale, Public Health England  Monica Galiano, Shahjahan Miah, Angie Lackenby, Omolola Akinbami, Tiina Talts, Leena Bhaw, Richard Myers, Steven Platt, Kirstin Edwards, Jonathan Hubb, Joanna Ellis, Maria Zambon
EPI_ISL_418684  hCoV-19/England/20124001302/2020  Europe / United Kingdom / England  2020-03-17  Respiratory Virus Unit, Microbiology Services Colindale, Public Health England  Respiratory Virus Unit, Microbiology Services Colindale, Public Health England  Monica Galiano, Shahjahan Miah, Angie Lackenby, Omolola Akinbami, Tiina Talts, Leena Bhaw, Richard Myers, Steven Platt, Kirstin Edwards, Jonathan Hubb, Joanna Ellis, Maria Zambon
EPI_ISL_418685  hCoV-19/England/20124001702/2020  Europe / United Kingdom / England  2020-03-17  Respiratory Virus Unit, Microbiology Services Colindale, Public Health England  Respiratory Virus Unit, Microbiology Services Colindale, Public Health England  Monica Galiano, Shahjahan Miah, Angie Lackenby, Omolola Akinbami, Tiina Talts, Leena Bhaw, Richard Myers, Steven Platt, Kirstin Edwards, Jonathan Hubb, Joanna Ellis, Maria Zambon
EPI_ISL_418686  hCoV-19/England/20124001902/2020  Europe / United Kingdom / England  2020-03-17  Respiratory Virus Unit, Microbiology Services Colindale, Public Health England  Respiratory Virus Unit, Microbiology Services Colindale, Public Health England  Monica Galiano, Shahjahan Miah, Angie Lackenby, Omolola Akinbami, Tiina Talts, Leena Bhaw, Richard Myers, Steven Platt, Kirstin Edwards, Jonathan Hubb, Joanna Ellis, Maria Zambon
EPI_ISL_418687  hCoV-19/England/20124002002/2020  Europe / United Kingdom / England  2020-03-17  Respiratory Virus Unit, Microbiology Services Colindale, Public Health England  Respiratory Virus Unit, Microbiology Services Colindale, Public Health England  Monica Galiano, Shahjahan Miah, Angie Lackenby, Omolola Akinbami, Tiina Talts, Leena Bhaw, Richard Myers, Steven Platt, Kirstin Edwards, Jonathan Hubb, Joanna Ellis, Maria Zambon
EPI_ISL_418688  hCoV-19/England/20124002102/2020  Europe / United Kingdom / England  2020-03-17  Respiratory Virus Unit, Microbiology Services Colindale, Public Health England  Respiratory Virus Unit, Microbiology Services Colindale, Public Health England  Monica Galiano, Shahjahan Miah, Angie Lackenby, Omolola Akinbami, Tiina Talts, Leena Bhaw, Richard Myers, Steven Platt, Kirstin Edwards, Jonathan Hubb, Joanna Ellis, Maria Zambon
EPI_ISL_418690  hCoV-19/England/20124003702/2020  Europe / United Kingdom / England  2020-03-17  Respiratory Virus Unit, Microbiology Services Colindale, Public Health England  Respiratory Virus Unit, Microbiology Services Colindale, Public Health England  Monica Galiano, Shahjahan Miah, Angie Lackenby, Omolola Akinbami, Tiina Talts, Leena Bhaw, Richard Myers, Steven Platt, Kirstin Edwards, Jonathan Hubb, Joanna Ellis, Maria Zambon
EPI_ISL_418691  hCoV-19/England/20124003902/2020  Europe / United Kingdom / England  2020-03-17  Respiratory Virus Unit, Microbiology Services Colindale, Public Health England  Respiratory Virus Unit, Microbiology Services Colindale, Public Health England  Monica Galiano, Shahjahan Miah, Angie Lackenby, Omolola Akinbami, Tiina Talts, Leena Bhaw, Richard Myers, Steven Platt, Kirstin Edwards, Jonathan Hubb, Joanna Ellis, Maria Zambon
EPI_ISL_418692  hCoV-19/England/20124004302/2020  Europe / United Kingdom / England  2020-03-17  Respiratory Virus Unit, Microbiology Services Colindale, Public Health England  Respiratory Virus Unit, Microbiology Services Colindale, Public Health England  Monica Galiano, Shahjahan Miah, Angie Lackenby, Omolola Akinbami, Tiina Talts, Leena Bhaw, Richard Myers, Steven Platt, Kirstin Edwards, Jonathan Hubb, Joanna Ellis, Maria Zambon
EPI_ISL_418694  hCoV-19/England/20124008902/2020  Europe / United Kingdom / England  2020-03-16  Respiratory Virus Unit, Microbiology Services Colindale, Public Health England  Respiratory Virus Unit, Microbiology Services Colindale, Public Health England  Monica Galiano, Shahjahan Miah, Angie Lackenby, Omolola Akinbami, Tiina Talts, Leena Bhaw, Richard Myers, Steven Platt, Kirstin Edwards, Jonathan Hubb, Joanna Ellis, Maria Zambon
EPI_ISL_418696  hCoV-19/England/20124009602/2020  Europe / United Kingdom / England  2020-03-18  Respiratory Virus Unit, Microbiology Services Colindale, Public Health England  Respiratory Virus Unit, Microbiology Services Colindale, Public Health England  Monica Galiano, Shahjahan Miah, Angie Lackenby, Omolola Akinbami, Tiina Talts, Leena Bhaw, Richard Myers, Steven Platt, Kirstin Edwards, Jonathan Hubb, Joanna Ellis, Maria Zambon
EPI_ISL_418698  hCoV-19/England/20124012702/2020  Europe / United Kingdom / England  2020-03-17  Respiratory Virus Unit, Microbiology Services Colindale, Public Health England  Respiratory Virus Unit, Microbiology Services Colindale, Public Health England  Monica Galiano, Shahjahan Miah, Angie Lackenby, Omolola Akinbami, Tiina Talts, Leena Bhaw, Richard Myers, Steven Platt, Kirstin Edwards, Jonathan Hubb, Joanna Ellis, Maria Zambon
EPI_ISL_418709  hCoV-19/England/20124034002/2020  Europe / United Kingdom / England  2020-03-16  Respiratory Virus Unit, Microbiology Services Colindale, Public Health England  Respiratory Virus Unit, Microbiology Services Colindale, Public Health England  Monica Galiano, Shahjahan Miah, Angie Lackenby, Omolola Akinbami, Tiina Talts, Leena Bhaw, Richard Myers, Steven Platt, Kirstin Edwards, Jonathan Hubb, Joanna Ellis, Maria Zambon
EPI_ISL_418711  hCoV-19/England/20124034702/2020  Europe / United Kingdom / England  2020-03-16  Respiratory Virus Unit, Microbiology Services Colindale, Public Health England  Respiratory Virus Unit,

Microbiology Services Colindale, Public Health England  Monica Galiano, Shahjahan Miah, Angie Lackenby, Omolola Akinbami, Tiina Talts, Leena Bhaw, Richard Myers, Steven Platt, Kirstin Edwards, Jonathan Hubb, Joanna Ellis, Maria Zambon
EPI_ISL_418715  hCoV-19/England/20124035802/2020  Europe / United Kingdom / England  2020-03-17  Respiratory Virus Unit, Microbiology Services Colindale, Public Health England  Respiratory Virus Unit, Microbiology Services Colindale, Public Health England  Monica Galiano, Shahjahan Miah, Angie Lackenby, Omolola Akinbami, Tiina Talts, Leena Bhaw, Richard Myers, Steven Platt, Kirstin Edwards, Jonathan Hubb, Joanna Ellis, Maria Zambon
EPI_ISL_418716  hCoV-19/England/20124036002/2020  Europe / United Kingdom / England  2020-03-17  Respiratory Virus Unit, Microbiology Services Colindale, Public Health England  Respiratory Virus Unit, Microbiology Services Colindale, Public Health England  Monica Galiano, Shahjahan Miah, Angie Lackenby, Omolola Akinbami, Tiina Talts, Leena Bhaw, Richard Myers, Steven Platt, Kirstin Edwards, Jonathan Hubb, Joanna Ellis, Maria Zambon
EPI_ISL_418739  hCoV-19/England/20124101102/2020  Europe / United Kingdom / England  2020-03-18  Respiratory Virus Unit, Microbiology Services Colindale, Public Health England  Respiratory Virus Unit, Microbiology Services Colindale, Public Health England  Monica Galiano, Shahjahan Miah, Angie Lackenby, Omolola Akinbami, Tiina Talts, Leena Bhaw, Richard Myers, Steven Platt, Kirstin Edwards, Jonathan Hubb, Joanna Ellis, Maria Zambon
EPI_ISL_418749  hCoV-19/England/20126006802/2020  Europe / United Kingdom / England  2020-03-18  Respiratory Virus Unit, Microbiology Services Colindale, Public Health England  Respiratory Virus Unit, Microbiology Services Colindale, Public Health England  Monica Galiano, Shahjahan Miah, Angie Lackenby, Omolola Akinbami, Tiina Talts, Leena Bhaw, Richard Myers, Steven Platt, Kirstin Edwards, Jonathan Hubb, Joanna Ellis, Maria Zambon
EPI_ISL_418751  hCoV-19/England/20126007102/2020  Europe / United Kingdom / England  2020-03-18  Respiratory Virus Unit, Microbiology Services Colindale, Public Health England  Respiratory Virus Unit, Microbiology Services Colindale, Public Health England  Monica Galiano, Shahjahan Miah, Angie Lackenby, Omolola Akinbami, Tiina Talts, Leena Bhaw, Richard Myers, Steven Platt, Kirstin Edwards, Jonathan Hubb, Joanna Ellis, Maria Zambon
EPI_ISL_418756  hCoV-19/England/20126012702/2020  Europe / United Kingdom / England  2020-03-17  Respiratory Virus Unit, Microbiology Services Colindale, Public Health England  Respiratory Virus Unit, Microbiology Services Colindale, Public Health England  Monica Galiano, Shahjahan Miah, Angie Lackenby, Omolola Akinbami, Tiina Talts, Leena Bhaw, Richard Myers, Steven Platt, Kirstin Edwards, Jonathan Hubb, Joanna Ellis, Maria Zambon
EPI_ISL_418764  hCoV-19/England/20126035502/2020  Europe / United Kingdom / England  2020-03-18  Respiratory Virus Unit, Microbiology Services Colindale, Public Health England  Respiratory Virus Unit, Microbiology Services Colindale, Public Health England  Monica Galiano, Shahjahan Miah, Angie Lackenby, Omolola Akinbami, Tiina Talts, Leena Bhaw, Richard Myers, Steven Platt, Kirstin Edwards, Jonathan Hubb, Joanna Ellis, Maria Zambon
EPI_ISL_418770  hCoV-19/England/20126096602/2020  Europe / United Kingdom / England  2020-03-18  Respiratory Virus Unit, Microbiology Services Colindale, Public Health England  Respiratory Virus Unit, Microbiology Services Colindale, Public Health England  Monica Galiano, Shahjahan Miah, Angie Lackenby, Omolola Akinbami, Tiina Talts, Leena Bhaw, Richard Myers, Steven Platt, Kirstin Edwards, Jonathan Hubb, Joanna Ellis, Maria Zambon
EPI_ISL_418771  hCoV-19/USA/WA-NH2/2020 North America / USA / Washington  2020-03-13  WA State Department of Health  Pathogen Discovery, Respiratory Viruses Branch, Division of Viral Diseases, Centers for Disease Control and Prevention  Jing Zhang, Ying Tao, Clinton R. Paden, Krista Queen, Anna Uehara, Yan Li, Haibin Wang, Jesica Jacobs, Denny Russell, Brian Hiatt, Jessica Gant, Suxiang Tong
EPI_ISL_418772  hCoV-19/USA/WA-NH3/2020 North America / USA / Washington  2020-03-13  WA State Department of Health  Pathogen Discovery, Respiratory Viruses Branch, Division of Viral Diseases, Centers for Disease Control and Prevention  Jing Zhang, Ying Tao, Clinton R. Paden, Krista Queen, Anna Uehara, Yan Li, Haibin Wang, Jesica Jacobs, Denny Russell, Brian Hiatt, Jessica Gant, Suxiang Tong
EPI_ISL_418773  hCoV-19/USA/WA-NH4/2020 North America / USA / Washington  2020-03-13  WA State Department of Health  Pathogen Discovery, Respiratory Viruses Branch, Division of Viral Diseases, Centers for Disease Control and Prevention  Jing Zhang, Ying Tao, Clinton R. Paden, Krista Queen, Anna Uehara, Yan Li, Haibin Wang, Jesica Jacobs, Denny Russell, Brian Hiatt, Jessica Gant, Suxiang Tong
EPI_ISL_418774  hCoV-19/USA/WA-NH5/2020 North America / USA / Washington  2020-03-13  WA State Department of Health  Pathogen Discovery, Respiratory Viruses Branch, Division of Viral Diseases, Centers for Disease Control and Prevention  Jing Zhang, Ying Tao, Clinton R. Paden, Krista Queen, Anna Uehara, Yan Li, Haibin Wang, Jesica Jacobs, Denny Russell, Brian Hiatt, Jessica Gant, Suxiang Tong
EPI_ISL_418775  hCoV-19/USA/WA-NH6/2020 North America / USA / Washington  2020-03-13  WA State Department of Health  Pathogen Discovery, Respiratory Viruses Branch, Division of Viral Diseases, Centers for Disease Control and Prevention  Jing Zhang, Ying Tao, Clinton R. Paden, Krista Queen, Anna Uehara, Yan Li, Haibin Wang, Jesica Jacobs, Denny Russell, Brian Hiatt, Jessica Gant, Suxiang Tong
EPI_ISL_418776  hCoV-19/USA/WA-NH7/2020 North America / USA / Washington  2020-03-13  WA State Department of Health  Pathogen Discovery, Respiratory Viruses Branch, Division of Viral Diseases, Centers for Disease Control and Prevention  Jing Zhang, Ying Tao, Clinton R. Paden, Krista Queen, Anna Uehara, Yan Li, Haibin Wang, Jesica Jacobs, Denny Russell, Brian Hiatt, Jessica Gant, Suxiang Tong
EPI_ISL_418777  hCoV-19/USA/WA-NH8/2020 North America / USA / Washington  2020-03-13  WA State Department of Health  Pathogen Discovery, Respiratory Viruses Branch, Division of Viral Diseases, Centers for Disease Control and Prevention  Jing Zhang, Ying Tao, Clinton R. Paden, Krista Queen, Anna Uehara, Yan Li,

Haibin Wang, Jesica Jacobs, Denny Russell, Brian Hiatt, Jessica Gant, Suxiang Tong
EPI_ISL_418778 hCoV-19/USA/WA-NH9/2020 North America / USA / Washington 2020-03-13 WA State Department of Health Pathogen Discovery, Respiratory Viruses Branch, Division of Viral Diseases, Centers for Disease Control and Prevention Ying Tao, Jing Zhang, Clinton R. Paden, Krista Queen, Anna Uehara, Yan Li, Haibin Wang, Jesica Jacobs, Denny Russell, Brian Hiatt, Jessica Gant, Suxiang Tong
EPI_ISL_418779 hCoV-19/USA/WA-NH10/2020 North America / USA / Washington 2020-03-13 WA State Department of Health Pathogen Discovery, Respiratory Viruses Branch, Division of Viral Diseases, Centers for Disease Control and Prevention Ying Tao, Jing Zhang, Clinton R. Paden, Krista Queen, Anna Uehara, Yan Li, Haibin Wang, Jesica Jacobs, Denny Russell, Brian Hiatt, Jessica Gant, Suxiang Tong
EPI_ISL_418780 hCoV-19/USA/WA-NH11/2020 North America / USA / Washington 2020-03-13 WA State Department of Health Pathogen Discovery, Respiratory Viruses Branch, Division of Viral Diseases, Centers for Disease Control and Prevention Ying Tao, Jing Zhang, Clinton R. Paden, Krista Queen, Anna Uehara, Yan Li, Haibin Wang, Jesica Jacobs, Denny Russell, Brian Hiatt, Jessica Gant, Suxiang Tong
EPI_ISL_418781 hCoV-19/USA/WA-NH12/2020 North America / USA / Washington 2020-03-13 WA State Department of Health Pathogen Discovery, Respiratory Viruses Branch, Division of Viral Diseases, Centers for Disease Control and Prevention Ying Tao, Jing Zhang, Clinton R. Paden, Krista Queen, Anna Uehara, Yan Li, Haibin Wang, Jesica Jacobs, Denny Russell, Brian Hiatt, Jessica Gant, Suxiang Tong
EPI_ISL_418782 hCoV-19/USA/WA-NH13/2020 North America / USA / Washington 2020-03-13 WA State Department of Health Pathogen Discovery, Respiratory Viruses Branch, Division of Viral Diseases, Centers for Disease Control and Prevention Ying Tao, Jing Zhang, Clinton R. Paden, Krista Queen, Anna Uehara, Yan Li, Haibin Wang, Jesica Jacobs, Denny Russell, Brian Hiatt, Jessica Gant, Suxiang Tong
EPI_ISL_418783 hCoV-19/USA/WA-NH14/2020 North America / USA / Washington 2020-03-13 WA State Department of Health Pathogen Discovery, Respiratory Viruses Branch, Division of Viral Diseases, Centers for Disease Control and Prevention Ying Tao, Jing Zhang, Clinton R. Paden, Krista Queen, Anna Uehara, Yan Li, Haibin Wang, Jesica Jacobs, Denny Russell, Brian Hiatt, Jessica Gant, Suxiang Tong
EPI_ISL_418784 hCoV-19/USA/WA-NH17/2020 North America / USA / Washington 2020-03-13 WA State Department of Health Pathogen Discovery, Respiratory Viruses Branch, Division of Viral Diseases, Centers for Disease Control and Prevention Ying Tao, Jing Zhang, Clinton R. Paden, Krista Queen, Anna Uehara, Yan Li, Haibin Wang, Jesica Jacobs, Denny Russell, Brian Hiatt, Jessica Gant, Suxiang Tong
EPI_ISL_418785 hCoV-19/USA/WA-NH18/2020 North America / USA / Washington 2020-03-13 WA State Department of Health Pathogen Discovery, Respiratory Viruses Branch, Division of Viral Diseases, Centers for Disease Control and Prevention Ying Tao, Jing Zhang, Clinton R. Paden, Krista Queen, Anna Uehara, Yan Li, Haibin Wang, Jesica Jacobs, Denny Russell, Brian Hiatt, Jessica Gant, Suxiang Tong
EPI_ISL_418786 hCoV-19/USA/WA-NH19/2020 North America / USA / Washington 2020-03-13 WA State Department of Health Pathogen Discovery, Respiratory Viruses Branch, Division of Viral Diseases, Centers for Disease Control and Prevention Ying Tao, Jing Zhang, Clinton R. Paden, Krista Queen, Anna Uehara, Yan Li, Haibin Wang, Jesica Jacobs, Denny Russell, Brian Hiatt, Jessica Gant, Suxiang Tong
EPI_ISL_418787 hCoV-19/USA/WA-NH20/2020 North America / USA / Washington 2020-03-13 WA State Department of Health Pathogen Discovery, Respiratory Viruses Branch, Division of Viral Diseases, Centers for Disease Control and Prevention Ying Tao, Jing Zhang, Clinton R. Paden, Krista Queen, Anna Uehara, Yan Li, Haibin Wang, Jesica Jacobs, Denny Russell, Brian Hiatt, Jessica Gant, Suxiang Tong
EPI_ISL_418788 hCoV-19/USA/WA-NH21/2020 North America / USA / Washington 2020-03-13 WA State Department of Health Pathogen Discovery, Respiratory Viruses Branch, Division of Viral Diseases, Centers for Disease Control and Prevention Ying Tao, Jing Zhang, Clinton R. Paden, Krista Queen, Anna Uehara, Yan Li, Haibin Wang, Jesica Jacobs, Denny Russell, Brian Hiatt, Jessica Gant, Suxiang Tong
EPI_ISL_418789 hCoV-19/USA/WA-NH22/2020 North America / USA / Washington 2020-03-13 WA State Department of Health Pathogen Discovery, Respiratory Viruses Branch, Division of Viral Diseases, Centers for Disease Control and Prevention Ying Tao, Jing Zhang, Clinton R. Paden, Krista Queen, Anna Uehara, Yan Li, Haibin Wang, Jesica Jacobs, Denny Russell, Brian Hiatt, Jessica Gant, Suxiang Tong
EPI_ISL_418790 hCoV-19/USA/WA-NH23/2020 North America / USA / Washington 2020-03-13 WA State Department of Health Pathogen Discovery, Respiratory Viruses Branch, Division of Viral Diseases, Centers for Disease Control and Prevention Ying Tao, Jing Zhang, Clinton R. Paden, Krista Queen, Anna Uehara, Yan Li, Haibin Wang, Jesica Jacobs, Denny Russell, Brian Hiatt, Jessica Gant, Suxiang Tong
EPI_ISL_418791 hCoV-19/USA/WA-NH24/2020 North America / USA / Washington 2020-03-13 WA State Department of Health Pathogen Discovery, Respiratory Viruses Branch, Division of Viral Diseases, Centers for Disease Control and Prevention Ying Tao, Jing Zhang, Clinton R. Paden, Krista Queen, Anna Uehara, Yan Li, Haibin Wang, Jesica Jacobs, Denny Russell, Brian Hiatt, Jessica Gant, Suxiang Tong
EPI_ISL_418792 hCoV-19/Belgium/GL-030546/2020 Europe / Belgium / Houthalen-Helchteren 2020-03-05 KU Leuven, Clinical and Epidemiological Virology KU Leuven, Clinical and Epidemiological Virology Bert Vanmechelen, Tony Wawina, Joan Marti-Carreras, Piet Maes
EPI_ISL_418793 hCoV-19/Belgium/SS-031047/2020 Europe / Belgium / Zonhoven 2020-03-10 KU Leuven, Clinical and Epidemiological Virology KU Leuven, Clinical and Epidemiological Virology Bert Vanmechelen, Tony Wawina, Joan Marti-Carreras, Piet Maes
EPI_ISL_418794 hCoV-19/Belgium/VHRA-030644/2020 Europe / Belgium / Nevele 2020-03-06 KU Leuven, Clinical and Epidemiological Virology KU Leuven, Clinical and Epidemiological Virology Bert Vanmechelen, Joan Marti-Carreras, Tony Wawina, Piet Maes
EPI_ISL_418795 hCoV-19/Belgium/NKR-030645/2020 Europe / Belgium / Ganshoren 2020-03-06 KU Leuven, Clinical and Epidemiological Virology KU Leuven, Clinical and Epidemiological Virology Bert Vanmechelen, Joan Marti-Carreras, Tony Wawina, Piet Maes
EPI_ISL_418796 hCoV-19/Belgium/VDLH-030548/2020 Europe / Belgium / Kalmthout 2020-03-05 KU

Leuven, Clinical and Epidemiological Virology KU Leuven, Clinical and Epidemiological Virology Bert Vanmechelen, Joan Marti-Carreras, Tony Wawina, Piet Maes
EPI_ISL_418798 hCoV-19/Belgium/GS-030549/2020 Europe / Belgium / Chaineux 2020-03-05 KU Leuven, Clinical and Epidemiological Virology KU Leuven, Clinical and Epidemiological Virology Bert Vanmechelen, Joan Marti-Carreras, Tony Wawina, Piet Maes
EPI_ISL_418799 hCoV-19/Australia/QLDID922/2020 Oceania / Australia / Queensland / Brisbane 2020-02-28 Mater Pathology Public Health Virology Laboratory Bixing Huang, Alyssa Pyke, Amanda De Jong, Andrew Van Den Hurk, Carmel Taylor, David Warrilow, Doris Genge, Elisabeth Gamez, Glen Hewitson, Ian Maxwell Mackay, Inga Sultana, Jamie McMahon, Jean Barcelon, Judy Northill, Mitchell Finger, Natalie Simpson, Neelima Nair, Peter Burtonclay, Peter Moore, Sarah Wheatley, Sean Moody, Sonja Hall-Mendelin, Timothy Gardam, and Frederick Moore
EPI_ISL_419900 hCoV-19/Australia/VIC205/2020 Oceania / Australia / Victoria 2020-03-19 Victorian Infectious Diseases Reference Laboratory (VIDRL) Victorian Infectious Diseases Reference Laboratory and Microbiological Diagnostic Unit Public Health Laboratory, Doherty Institute Caly L., Seemann T., Sait, M., Schultz M., Druce J., Sherry, N.
EPI_ISL_419901 hCoV-19/Australia/VIC206/2020 Oceania / Australia / Victoria 2020-03-19 Victorian Infectious Diseases Reference Laboratory (VIDRL) Victorian Infectious Diseases Reference Laboratory and Microbiological Diagnostic Unit Public Health Laboratory, Doherty Institute Caly L., Seemann T., Sait, M., Schultz M., Druce J., Sherry, N.
EPI_ISL_419902 hCoV-19/Australia/VIC207/2020 Oceania / Australia / Victoria 2020-03-19 Victorian Infectious Diseases Reference Laboratory (VIDRL) Victorian Infectious Diseases Reference Laboratory and Microbiological Diagnostic Unit Public Health Laboratory, Doherty Institute Caly L., Seemann T., Sait, M., Schultz M., Druce J., Sherry, N.
EPI_ISL_419903 hCoV-19/Australia/VIC208/2020 Oceania / Australia / Victoria 2020-03-20 Victorian Infectious Diseases Reference Laboratory (VIDRL) Victorian Infectious Diseases Reference Laboratory and Microbiological Diagnostic Unit Public Health Laboratory, Doherty Institute Caly L., Seemann T., Sait, M., Schultz M., Druce J., Sherry, N.
EPI_ISL_419904 hCoV-19/Australia/VIC209/2020 Oceania / Australia / Victoria 2020-03-20 Victorian Infectious Diseases Reference Laboratory (VIDRL) Victorian Infectious Diseases Reference Laboratory and Microbiological Diagnostic Unit Public Health Laboratory, Doherty Institute Caly L., Seemann T., Sait, M., Schultz M., Druce J., Sherry, N.
EPI_ISL_419905 hCoV-19/Australia/VIC210/2020 Oceania / Australia / Victoria 2020-03-20 Victorian Infectious Diseases Reference Laboratory (VIDRL) Victorian Infectious Diseases Reference Laboratory and Microbiological Diagnostic Unit Public Health Laboratory, Doherty Institute Caly L., Seemann T., Sait, M., Schultz M., Druce J., Sherry, N.
EPI_ISL_419906 hCoV-19/Australia/VIC211/2020 Oceania / Australia / Victoria 2020-03-20 Victorian Infectious Diseases Reference Laboratory (VIDRL) Victorian Infectious Diseases Reference Laboratory and Microbiological Diagnostic Unit Public Health Laboratory, Doherty Institute Caly L., Seemann T., Sait, M., Schultz M., Druce J., Sherry, N.
EPI_ISL_419907 hCoV-19/Australia/VIC212/2020 Oceania / Australia / Victoria 2020-03-20 Victorian Infectious Diseases Reference Laboratory (VIDRL) Victorian Infectious Diseases Reference Laboratory and Microbiological Diagnostic Unit Public Health Laboratory, Doherty Institute Caly L., Seemann T., Sait, M., Schultz M., Druce J., Sherry, N.
EPI_ISL_419908 hCoV-19/Australia/VIC213/2020 Oceania / Australia / Victoria 2020-03-20 Victorian Infectious Diseases Reference Laboratory (VIDRL) Victorian Infectious Diseases Reference Laboratory and Microbiological Diagnostic Unit Public Health Laboratory, Doherty Institute Caly L., Seemann T., Sait, M., Schultz M., Druce J., Sherry, N.
EPI_ISL_419909 hCoV-19/Australia/VIC214/2020 Oceania / Australia / Victoria 2020-03-20 Victorian Infectious Diseases Reference Laboratory (VIDRL) Victorian Infectious Diseases Reference Laboratory and Microbiological Diagnostic Unit Public Health Laboratory, Doherty Institute Caly L., Seemann T., Sait, M., Schultz M., Druce J., Sherry, N.
EPI_ISL_419910 hCoV-19/Australia/VIC215/2020 Oceania / Australia / Victoria 2020-03-20 Victorian Infectious Diseases Reference Laboratory (VIDRL) Victorian Infectious Diseases Reference Laboratory and Microbiological Diagnostic Unit Public Health Laboratory, Doherty Institute Caly L., Seemann T., Sait, M., Schultz M., Druce J., Sherry, N.
EPI_ISL_419911 hCoV-19/Australia/VIC216/2020 Oceania / Australia / Victoria 2020-03-20 Victorian Infectious Diseases Reference Laboratory (VIDRL) Victorian Infectious Diseases Reference Laboratory and Microbiological Diagnostic Unit Public Health Laboratory, Doherty Institute Caly L., Seemann T., Sait, M., Schultz M., Druce J., Sherry, N.
EPI_ISL_419912 hCoV-19/Australia/VIC217/2020 Oceania / Australia / Victoria 2020-03-20 Victorian Infectious Diseases Reference Laboratory (VIDRL) Victorian Infectious Diseases Reference Laboratory and Microbiological Diagnostic Unit Public Health Laboratory, Doherty Institute Caly L., Seemann T., Sait, M., Schultz M., Druce J., Sherry, N.
EPI_ISL_419913 hCoV-19/Australia/VIC218/2020 Oceania / Australia / Victoria 2020-03-20 Victorian Infectious Diseases Reference Laboratory (VIDRL) Victorian Infectious Diseases Reference Laboratory and Microbiological Diagnostic Unit Public Health Laboratory, Doherty Institute Caly L., Seemann T., Sait, M., Schultz M., Druce J., Sherry, N.
EPI_ISL_419914 hCoV-19/Australia/VIC219/2020 Oceania / Australia / Victoria 2020-03-20 Victorian Infectious Diseases Reference Laboratory (VIDRL) Victorian Infectious Diseases Reference Laboratory and Microbiological Diagnostic Unit Public Health Laboratory, Doherty Institute Caly L., Seemann T., Sait, M., Schultz M., Druce J., Sherry, N.

EPI_ISL_419915 hCoV-19/Australia/VIC220/2020 Oceania / Australia / Victoria 2020-03-20 Victorian Infectious Diseases Reference Laboratory (VIDRL) Victorian Infectious Diseases Reference Laboratory and Microbiological Diagnostic Unit Public Health Laboratory, Doherty Institute Caly L., Seemann T., Sait, M., Schultz M., Druce J., Sherry, N.
EPI_ISL_419916 hCoV-19/Australia/VIC221/2020 Oceania / Australia / Victoria 2020-03-20 Victorian Infectious Diseases Reference Laboratory (VIDRL) Victorian Infectious Diseases Reference Laboratory and Microbiological Diagnostic Unit Public Health Laboratory, Doherty Institute Caly L., Seemann T., Sait, M., Schultz M., Druce J., Sherry, N.
EPI_ISL_419917 hCoV-19/Australia/VIC222/2020 Oceania / Australia / Victoria 2020-03-20 Victorian Infectious Diseases Reference Laboratory (VIDRL) Victorian Infectious Diseases Reference Laboratory and Microbiological Diagnostic Unit Public Health Laboratory, Doherty Institute Caly L., Seemann T., Sait, M., Schultz M., Druce J., Sherry, N.
EPI_ISL_419918 hCoV-19/Australia/VIC223/2020 Oceania / Australia / Victoria 2020-03-20 Victorian Infectious Diseases Reference Laboratory (VIDRL) Victorian Infectious Diseases Reference Laboratory and Microbiological Diagnostic Unit Public Health Laboratory, Doherty Institute Caly L., Seemann T., Sait, M., Schultz M., Druce J., Sherry, N.
EPI_ISL_419919 hCoV-19/Australia/VIC224/2020 Oceania / Australia / Victoria 2020-03-20 Victorian Infectious Diseases Reference Laboratory (VIDRL) Victorian Infectious Diseases Reference Laboratory and Microbiological Diagnostic Unit Public Health Laboratory, Doherty Institute Caly L., Seemann T., Sait, M., Schultz M., Druce J., Sherry, N.
EPI_ISL_419920 hCoV-19/Australia/VIC225/2020 Oceania / Australia / Victoria 2020-03-20 Victorian Infectious Diseases Reference Laboratory (VIDRL) Victorian Infectious Diseases Reference Laboratory and Microbiological Diagnostic Unit Public Health Laboratory, Doherty Institute Caly L., Seemann T., Sait, M., Schultz M., Druce J., Sherry, N.
EPI_ISL_419921 hCoV-19/Australia/VIC226/2020 Oceania / Australia / Victoria 2020-03-20 Victorian Infectious Diseases Reference Laboratory (VIDRL) Victorian Infectious Diseases Reference Laboratory and Microbiological Diagnostic Unit Public Health Laboratory, Doherty Institute Caly L., Seemann T., Sait, M., Schultz M., Druce J., Sherry, N.
EPI_ISL_419922 hCoV-19/Australia/VIC227/2020 Oceania / Australia / Victoria 2020-03-20 Victorian Infectious Diseases Reference Laboratory (VIDRL) Victorian Infectious Diseases Reference Laboratory and Microbiological Diagnostic Unit Public Health Laboratory, Doherty Institute Caly L., Seemann T., Sait, M., Schultz M., Druce J., Sherry, N.
EPI_ISL_419923 hCoV-19/Australia/VIC228/2020 Oceania / Australia / Victoria 2020-03-20 Victorian Infectious Diseases Reference Laboratory (VIDRL) Victorian Infectious Diseases Reference Laboratory and Microbiological Diagnostic Unit Public Health Laboratory, Doherty Institute Caly L., Seemann T., Sait, M., Schultz M., Druce J., Sherry, N.
EPI_ISL_419924 hCoV-19/Australia/VIC229/2020 Oceania / Australia / Victoria 2020-03-21 Victorian Infectious Diseases Reference Laboratory (VIDRL) Victorian Infectious Diseases Reference Laboratory and Microbiological Diagnostic Unit Public Health Laboratory, Doherty Institute Caly L., Seemann T., Sait, M., Schultz M., Druce J., Sherry, N.
EPI_ISL_419925 hCoV-19/Australia/VIC230/2020 Oceania / Australia / Victoria 2020-03-21 Victorian Infectious Diseases Reference Laboratory (VIDRL) Victorian Infectious Diseases Reference Laboratory and Microbiological Diagnostic Unit Public Health Laboratory, Doherty Institute Caly L., Seemann T., Sait, M., Schultz M., Druce J., Sherry, N.
EPI_ISL_419926 hCoV-19/Australia/VIC231/2020 Oceania / Australia / Victoria 2020-03-21 Victorian Infectious Diseases Reference Laboratory (VIDRL) Victorian Infectious Diseases Reference Laboratory and Microbiological Diagnostic Unit Public Health Laboratory, Doherty Institute Caly L., Seemann T., Sait, M., Schultz M., Druce J., Sherry, N.
EPI_ISL_419927 hCoV-19/Australia/VIC232/2020 Oceania / Australia / Victoria 2020-03-21 Victorian Infectious Diseases Reference Laboratory (VIDRL) Victorian Infectious Diseases Reference Laboratory and Microbiological Diagnostic Unit Public Health Laboratory, Doherty Institute Caly L., Seemann T., Sait, M., Schultz M., Druce J., Sherry, N.
EPI_ISL_419928 hCoV-19/Australia/VIC233/2020 Oceania / Australia / Victoria 2020-03-21 Victorian Infectious Diseases Reference Laboratory (VIDRL) Victorian Infectious Diseases Reference Laboratory and Microbiological Diagnostic Unit Public Health Laboratory, Doherty Institute Caly L., Seemann T., Sait, M., Schultz M., Druce J., Sherry, N.
EPI_ISL_419929 hCoV-19/Australia/VIC234/2020 Oceania / Australia / Victoria 2020-03-21 Victorian Infectious Diseases Reference Laboratory (VIDRL) Victorian Infectious Diseases Reference Laboratory and Microbiological Diagnostic Unit Public Health Laboratory, Doherty Institute Caly L., Seemann T., Sait, M., Schultz M., Druce J., Sherry, N.
EPI_ISL_419931 hCoV-19/Australia/VIC236/2020 Oceania / Australia / Victoria 2020-03-21 Victorian Infectious Diseases Reference Laboratory (VIDRL) Victorian Infectious Diseases Reference Laboratory and Microbiological Diagnostic Unit Public Health Laboratory, Doherty Institute Caly L., Seemann T., Sait, M., Schultz M., Druce J., Sherry, N.
EPI_ISL_419932 hCoV-19/Australia/VIC237/2020 Oceania / Australia / Victoria 2020-03-21 Victorian Infectious Diseases Reference Laboratory (VIDRL) Victorian Infectious Diseases Reference Laboratory and Microbiological Diagnostic Unit Public Health Laboratory, Doherty Institute Caly L., Seemann T., Sait, M., Schultz M., Druce J., Sherry, N.
EPI_ISL_419933 hCoV-19/Australia/VIC238/2020 Oceania / Australia / Victoria 2020-03-21 Victorian Infectious Diseases Reference Laboratory (VIDRL) Victorian Infectious Diseases Reference Laboratory and

Microbiological Diagnostic Unit Public Health Laboratory, Doherty Institute	Caly L., Seemann T., Sait, M., Schultz M., Druce J., Sherry, N.
EPI_ISL_419934	hCoV-19/Australia/VIC239/2020	Oceania / Australia / Victoria	2020-03-21	Victorian Infectious Diseases Reference Laboratory (VIDRL)	Victorian Infectious Diseases Reference Laboratory and Microbiological Diagnostic Unit Public Health Laboratory, Doherty Institute	Caly L., Seemann T., Sait, M., Schultz M., Druce J., Sherry, N.
EPI_ISL_419935	hCoV-19/Australia/VIC240/2020	Oceania / Australia / Victoria	2020-03-21	Victorian Infectious Diseases Reference Laboratory (VIDRL)	Victorian Infectious Diseases Reference Laboratory and Microbiological Diagnostic Unit Public Health Laboratory, Doherty Institute	Caly L., Seemann T., Sait, M., Schultz M., Druce J., Sherry, N.
EPI_ISL_419936	hCoV-19/Australia/VIC241/2020	Oceania / Australia / Victoria	2020-03-21	Victorian Infectious Diseases Reference Laboratory (VIDRL)	Victorian Infectious Diseases Reference Laboratory and Microbiological Diagnostic Unit Public Health Laboratory, Doherty Institute	Caly L., Seemann T., Sait, M., Schultz M., Druce J., Sherry, N.
EPI_ISL_419937	hCoV-19/Australia/VIC242/2020	Oceania / Australia / Victoria	2020-03-21	Victorian Infectious Diseases Reference Laboratory (VIDRL)	Victorian Infectious Diseases Reference Laboratory and Microbiological Diagnostic Unit Public Health Laboratory, Doherty Institute	Caly L., Seemann T., Sait, M., Schultz M., Druce J., Sherry, N.
EPI_ISL_419938	hCoV-19/Australia/VIC243/2020	Oceania / Australia / Victoria	2020-03-21	Victorian Infectious Diseases Reference Laboratory (VIDRL)	Victorian Infectious Diseases Reference Laboratory and Microbiological Diagnostic Unit Public Health Laboratory, Doherty Institute	Caly L., Seemann T., Sait, M., Schultz M., Druce J., Sherry, N.
EPI_ISL_419939	hCoV-19/Australia/VIC244/2020	Oceania / Australia / Victoria	2020-03-21	Victorian Infectious Diseases Reference Laboratory (VIDRL)	Victorian Infectious Diseases Reference Laboratory and Microbiological Diagnostic Unit Public Health Laboratory, Doherty Institute	Caly L., Seemann T., Sait, M., Schultz M., Druce J., Sherry, N.
EPI_ISL_419940	hCoV-19/Australia/VIC245/2020	Oceania / Australia / Victoria	2020-03-21	Victorian Infectious Diseases Reference Laboratory (VIDRL)	Victorian Infectious Diseases Reference Laboratory and Microbiological Diagnostic Unit Public Health Laboratory, Doherty Institute	Caly L., Seemann T., Sait, M., Schultz M., Druce J., Sherry, N.
EPI_ISL_419941	hCoV-19/Australia/VIC246/2020	Oceania / Australia / Victoria	2020-03-21	Victorian Infectious Diseases Reference Laboratory (VIDRL)	Victorian Infectious Diseases Reference Laboratory and Microbiological Diagnostic Unit Public Health Laboratory, Doherty Institute	Caly L., Seemann T., Sait, M., Schultz M., Druce J., Sherry, N.
EPI_ISL_419942	hCoV-19/Australia/VIC247/2020	Oceania / Australia / Victoria	2020-03-21	Victorian Infectious Diseases Reference Laboratory (VIDRL)	Victorian Infectious Diseases Reference Laboratory and Microbiological Diagnostic Unit Public Health Laboratory, Doherty Institute	Caly L., Seemann T., Sait, M., Schultz M., Druce J., Sherry, N.
EPI_ISL_419943	hCoV-19/Australia/VIC248/2020	Oceania / Australia / Victoria	2020-03-21	Victorian Infectious Diseases Reference Laboratory (VIDRL)	Victorian Infectious Diseases Reference Laboratory and Microbiological Diagnostic Unit Public Health Laboratory, Doherty Institute	Caly L., Seemann T., Sait, M., Schultz M., Druce J., Sherry, N.
EPI_ISL_419944	hCoV-19/Australia/VIC249/2020	Oceania / Australia / Victoria	2020-03-21	Victorian Infectious Diseases Reference Laboratory (VIDRL)	Victorian Infectious Diseases Reference Laboratory and Microbiological Diagnostic Unit Public Health Laboratory, Doherty Institute	Caly L., Seemann T., Sait, M., Schultz M., Druce J., Sherry, N.
EPI_ISL_419945	hCoV-19/Australia/VIC250/2020	Oceania / Australia / Victoria	2020-03-21	Victorian Infectious Diseases Reference Laboratory (VIDRL)	Victorian Infectious Diseases Reference Laboratory and Microbiological Diagnostic Unit Public Health Laboratory, Doherty Institute	Caly L., Seemann T., Sait, M., Schultz M., Druce J., Sherry, N.
EPI_ISL_419946	hCoV-19/Australia/VIC251/2020	Oceania / Australia / Victoria	2020-03-21	Victorian Infectious Diseases Reference Laboratory (VIDRL)	Victorian Infectious Diseases Reference Laboratory and Microbiological Diagnostic Unit Public Health Laboratory, Doherty Institute	Caly L., Seemann T., Sait, M., Schultz M., Druce J., Sherry, N.
EPI_ISL_419947	hCoV-19/Australia/VIC252/2020	Oceania / Australia / Victoria	2020-03-21	Victorian Infectious Diseases Reference Laboratory (VIDRL)	Victorian Infectious Diseases Reference Laboratory and Microbiological Diagnostic Unit Public Health Laboratory, Doherty Institute	Caly L., Seemann T., Sait, M., Schultz M., Druce J., Sherry, N.
EPI_ISL_419948	hCoV-19/Australia/VIC253/2020	Oceania / Australia / Victoria	2020-03-21	Victorian Infectious Diseases Reference Laboratory (VIDRL)	Victorian Infectious Diseases Reference Laboratory and Microbiological Diagnostic Unit Public Health Laboratory, Doherty Institute	Caly L., Seemann T., Sait, M., Schultz M., Druce J., Sherry, N.
EPI_ISL_419949	hCoV-19/Australia/VIC254/2020	Oceania / Australia / Victoria	2020-03-21	Victorian Infectious Diseases Reference Laboratory (VIDRL)	Victorian Infectious Diseases Reference Laboratory and Microbiological Diagnostic Unit Public Health Laboratory, Doherty Institute	Caly L., Seemann T., Sait, M., Schultz M., Druce J., Sherry, N.
EPI_ISL_419950	hCoV-19/Australia/VIC257/2020	Oceania / Australia / Victoria	2020-03-21	Victorian Infectious Diseases Reference Laboratory (VIDRL)	Victorian Infectious Diseases Reference Laboratory and Microbiological Diagnostic Unit Public Health Laboratory, Doherty Institute	Caly L., Seemann T., Sait, M., Schultz M., Druce J., Sherry, N.

EPI_ISL_419951  hCoV-19/Australia/VIC258/2020   Oceania / Australia / Victoria  2020-03-21      Victorian Infectious Diseases Reference Laboratory (VIDRL)        Victorian Infectious Diseases Reference Laboratory and Microbiological Diagnostic Unit Public Health Laboratory, Doherty Institute     Caly L., Seemann T., Sait, M., Schultz M., Druce J., Sherry, N.
EPI_ISL_419952  hCoV-19/Australia/VIC259/2020   Oceania / Australia / Victoria  2020-03-21      Victorian Infectious Diseases Reference Laboratory (VIDRL)        Victorian Infectious Diseases Reference Laboratory and Microbiological Diagnostic Unit Public Health Laboratory, Doherty Institute     Caly L., Seemann T., Sait, M., Schultz M., Druce J., Sherry, N.
EPI_ISL_419953  hCoV-19/Australia/VIC260/2020   Oceania / Australia / Victoria  2020-03-21      Victorian Infectious Diseases Reference Laboratory (VIDRL)        Victorian Infectious Diseases Reference Laboratory and Microbiological Diagnostic Unit Public Health Laboratory, Doherty Institute     Caly L., Seemann T., Sait, M., Schultz M., Druce J., Sherry, N.
EPI_ISL_419954  hCoV-19/Australia/VIC261/2020   Oceania / Australia / Victoria  2020-03-21      Victorian Infectious Diseases Reference Laboratory (VIDRL)        Victorian Infectious Diseases Reference Laboratory and Microbiological Diagnostic Unit Public Health Laboratory, Doherty Institute     Caly L., Seemann T., Sait, M., Schultz M., Druce J., Sherry, N.
EPI_ISL_419955  hCoV-19/Australia/VIC262/2020   Oceania / Australia / Victoria  2020-03-21      Victorian Infectious Diseases Reference Laboratory (VIDRL)        Victorian Infectious Diseases Reference Laboratory and Microbiological Diagnostic Unit Public Health Laboratory, Doherty Institute     Caly L., Seemann T., Sait, M., Schultz M., Druce J., Sherry, N.
EPI_ISL_419956  hCoV-19/Australia/VIC263/2020   Oceania / Australia / Victoria  2020-03-21      Victorian Infectious Diseases Reference Laboratory (VIDRL)        Victorian Infectious Diseases Reference Laboratory and Microbiological Diagnostic Unit Public Health Laboratory, Doherty Institute     Caly L., Seemann T., Sait, M., Schultz M., Druce J., Sherry, N.
EPI_ISL_419957  hCoV-19/Australia/VIC264/2020   Oceania / Australia / Victoria  2020-03-22      Victorian Infectious Diseases Reference Laboratory (VIDRL)        Victorian Infectious Diseases Reference Laboratory and Microbiological Diagnostic Unit Public Health Laboratory, Doherty Institute     Caly L., Seemann T., Sait, M., Schultz M., Druce J., Sherry, N.
EPI_ISL_419958  hCoV-19/Australia/VIC265/2020   Oceania / Australia / Victoria  2020-03-22      Victorian Infectious Diseases Reference Laboratory (VIDRL)        Victorian Infectious Diseases Reference Laboratory and Microbiological Diagnostic Unit Public Health Laboratory, Doherty Institute     Caly L., Seemann T., Sait, M., Schultz M., Druce J., Sherry, N.
EPI_ISL_419959  hCoV-19/Australia/VIC266/2020   Oceania / Australia / Victoria  2020-03-22      Victorian Infectious Diseases Reference Laboratory (VIDRL)        Victorian Infectious Diseases Reference Laboratory and Microbiological Diagnostic Unit Public Health Laboratory, Doherty Institute     Caly L., Seemann T., Sait, M., Schultz M., Druce J., Sherry, N.
EPI_ISL_419960  hCoV-19/Australia/VIC267/2020   Oceania / Australia / Victoria  2020-03-22      Victorian Infectious Diseases Reference Laboratory (VIDRL)        Victorian Infectious Diseases Reference Laboratory and Microbiological Diagnostic Unit Public Health Laboratory, Doherty Institute     Caly L., Seemann T., Sait, M., Schultz M., Druce J., Sherry, N.
EPI_ISL_419961  hCoV-19/Australia/VIC268/2020   Oceania / Australia / Victoria  2020-03-22      Victorian Infectious Diseases Reference Laboratory (VIDRL)        Victorian Infectious Diseases Reference Laboratory and Microbiological Diagnostic Unit Public Health Laboratory, Doherty Institute     Caly L., Seemann T., Sait, M., Schultz M., Druce J., Sherry, N.
EPI_ISL_419962  hCoV-19/Australia/VIC269/2020   Oceania / Australia / Victoria  2020-03-22      Victorian Infectious Diseases Reference Laboratory (VIDRL)        Victorian Infectious Diseases Reference Laboratory and Microbiological Diagnostic Unit Public Health Laboratory, Doherty Institute     Caly L., Seemann T., Sait, M., Schultz M., Druce J., Sherry, N.
EPI_ISL_419963  hCoV-19/Australia/VIC270/2020   Oceania / Australia / Victoria  2020-03-22      Victorian Infectious Diseases Reference Laboratory (VIDRL)        Victorian Infectious Diseases Reference Laboratory and Microbiological Diagnostic Unit Public Health Laboratory, Doherty Institute     Caly L., Seemann T., Sait, M., Schultz M., Druce J., Sherry, N.
EPI_ISL_419964  hCoV-19/Australia/VIC271/2020   Oceania / Australia / Victoria  2020-03-22      Victorian Infectious Diseases Reference Laboratory (VIDRL)        Victorian Infectious Diseases Reference Laboratory and Microbiological Diagnostic Unit Public Health Laboratory, Doherty Institute     Caly L., Seemann T., Sait, M., Schultz M., Druce J., Sherry, N.
EPI_ISL_419965  hCoV-19/Australia/VIC272/2020   Oceania / Australia / Victoria  2020-03-22      Victorian Infectious Diseases Reference Laboratory (VIDRL)        Victorian Infectious Diseases Reference Laboratory and Microbiological Diagnostic Unit Public Health Laboratory, Doherty Institute     Caly L., Seemann T., Sait, M., Schultz M., Druce J., Sherry, N.
EPI_ISL_419966  hCoV-19/Australia/VIC273/2020   Oceania / Australia / Victoria  2020-03-22      Victorian Infectious Diseases Reference Laboratory (VIDRL)        Victorian Infectious Diseases Reference Laboratory and Microbiological Diagnostic Unit Public Health Laboratory, Doherty Institute     Caly L., Seemann T., Sait, M., Schultz M., Druce J., Sherry, N.
EPI_ISL_419967  hCoV-19/Australia/VIC274/2020   Oceania / Australia / Victoria  2020-03-22      Victorian Infectious Diseases Reference Laboratory (VIDRL)        Victorian Infectious Diseases Reference Laboratory and Microbiological Diagnostic Unit Public Health Laboratory, Doherty Institute     Caly L., Seemann T., Sait, M., Schultz M., Druce J., Sherry, N.
EPI_ISL_419968  hCoV-19/Australia/VIC275/2020   Oceania / Australia / Victoria  2020-03-22      Victorian Infectious Diseases Reference Laboratory (VIDRL)        Victorian Infectious Diseases Reference Laboratory and

Microbiological Diagnostic Unit Public Health Laboratory, Doherty Institute	Caly L., Seemann T., Sait, M., Schultz M., Druce J., Sherry, N.
EPI_ISL_419969	hCoV-19/Australia/VIC276/2020	Oceania / Australia / Victoria	2020-03-22	Victorian Infectious Diseases Reference Laboratory (VIDRL)	Victorian Infectious Diseases Reference Laboratory and Microbiological Diagnostic Unit Public Health Laboratory, Doherty Institute	Caly L., Seemann T., Sait, M., Schultz M., Druce J., Sherry, N.
EPI_ISL_419970	hCoV-19/Australia/VIC277/2020	Oceania / Australia / Victoria	2020-03-22	Victorian Infectious Diseases Reference Laboratory (VIDRL)	Victorian Infectious Diseases Reference Laboratory and Microbiological Diagnostic Unit Public Health Laboratory, Doherty Institute	Caly L., Seemann T., Sait, M., Schultz M., Druce J., Sherry, N.
EPI_ISL_419971	hCoV-19/Australia/VIC278/2020	Oceania / Australia / Victoria	2020-03-22	Victorian Infectious Diseases Reference Laboratory (VIDRL)	Victorian Infectious Diseases Reference Laboratory and Microbiological Diagnostic Unit Public Health Laboratory, Doherty Institute	Caly L., Seemann T., Sait, M., Schultz M., Druce J., Sherry, N.
EPI_ISL_419972	hCoV-19/Australia/VIC279/2020	Oceania / Australia / Victoria	2020-03-22	Victorian Infectious Diseases Reference Laboratory (VIDRL)	Victorian Infectious Diseases Reference Laboratory and Microbiological Diagnostic Unit Public Health Laboratory, Doherty Institute	Caly L., Seemann T., Sait, M., Schultz M., Druce J., Sherry, N.
EPI_ISL_419973	hCoV-19/Australia/VIC280/2020	Oceania / Australia / Victoria	2020-03-22	Victorian Infectious Diseases Reference Laboratory (VIDRL)	Victorian Infectious Diseases Reference Laboratory and Microbiological Diagnostic Unit Public Health Laboratory, Doherty Institute	Caly L., Seemann T., Sait, M., Schultz M., Druce J., Sherry, N.
EPI_ISL_419974	hCoV-19/Australia/VIC283/2020	Oceania / Australia / Victoria	2020-03-22	Victorian Infectious Diseases Reference Laboratory (VIDRL)	Victorian Infectious Diseases Reference Laboratory and Microbiological Diagnostic Unit Public Health Laboratory, Doherty Institute	Caly L., Seemann T., Sait, M., Schultz M., Druce J., Sherry, N.
EPI_ISL_419975	hCoV-19/Australia/VIC284/2020	Oceania / Australia / Victoria	2020-03-22	Victorian Infectious Diseases Reference Laboratory (VIDRL)	Victorian Infectious Diseases Reference Laboratory and Microbiological Diagnostic Unit Public Health Laboratory, Doherty Institute	Caly L., Seemann T., Sait, M., Schultz M., Druce J., Sherry, N.
EPI_ISL_419976	hCoV-19/Australia/VIC285/2020	Oceania / Australia / Victoria	2020-03-22	Victorian Infectious Diseases Reference Laboratory (VIDRL)	Victorian Infectious Diseases Reference Laboratory and Microbiological Diagnostic Unit Public Health Laboratory, Doherty Institute	Caly L., Seemann T., Sait, M., Schultz M., Druce J., Sherry, N.
EPI_ISL_419977	hCoV-19/Australia/VIC286/2020	Oceania / Australia / Victoria	2020-03-22	Victorian Infectious Diseases Reference Laboratory (VIDRL)	Victorian Infectious Diseases Reference Laboratory and Microbiological Diagnostic Unit Public Health Laboratory, Doherty Institute	Caly L., Seemann T., Sait, M., Schultz M., Druce J., Sherry, N.
EPI_ISL_419978	hCoV-19/Australia/VIC287/2020	Oceania / Australia / Victoria	2020-03-22	Victorian Infectious Diseases Reference Laboratory (VIDRL)	Victorian Infectious Diseases Reference Laboratory and Microbiological Diagnostic Unit Public Health Laboratory, Doherty Institute	Caly L., Seemann T., Sait, M., Schultz M., Druce J., Sherry, N.
EPI_ISL_419979	hCoV-19/Australia/VIC288/2020	Oceania / Australia / Victoria	2020-03-22	Victorian Infectious Diseases Reference Laboratory (VIDRL)	Victorian Infectious Diseases Reference Laboratory and Microbiological Diagnostic Unit Public Health Laboratory, Doherty Institute	Caly L., Seemann T., Sait, M., Schultz M., Druce J., Sherry, N.
EPI_ISL_419980	hCoV-19/Australia/VIC289/2020	Oceania / Australia / Victoria	2020-03-22	Microbiological Diagnostic Unit Public Health Laboratory	Microbiological Diagnostic Unit Public Health Laboratory	Seemann T., Schultz M., Sait, M., Sherry, N.
EPI_ISL_419982	hCoV-19/Australia/VIC291/2020	Oceania / Australia / Victoria	2020-03-23	Victorian Infectious Diseases Reference Laboratory (VIDRL)	Victorian Infectious Diseases Reference Laboratory and Microbiological Diagnostic Unit Public Health Laboratory, Doherty Institute	Caly L., Seemann T., Sait, M., Schultz M., Druce J., Sherry, N.
EPI_ISL_419983	hCoV-19/Australia/VIC292/2020	Oceania / Australia / Victoria	2020-03-23	Victorian Infectious Diseases Reference Laboratory (VIDRL)	Victorian Infectious Diseases Reference Laboratory and Microbiological Diagnostic Unit Public Health Laboratory, Doherty Institute	Caly L., Seemann T., Sait, M., Schultz M., Druce J., Sherry, N.
EPI_ISL_419984	hCoV-19/Australia/VIC293/2020	Oceania / Australia / Victoria	2020-03-23	Victorian Infectious Diseases Reference Laboratory (VIDRL)	Victorian Infectious Diseases Reference Laboratory and Microbiological Diagnostic Unit Public Health Laboratory, Doherty Institute	Caly L., Seemann T., Sait, M., Schultz M., Druce J., Sherry, N.
EPI_ISL_419985	hCoV-19/Australia/VIC294/2020	Oceania / Australia / Victoria	2020-03-23	Victorian Infectious Diseases Reference Laboratory (VIDRL)	Victorian Infectious Diseases Reference Laboratory and Microbiological Diagnostic Unit Public Health Laboratory, Doherty Institute	Caly L., Seemann T., Sait, M., Schultz M., Druce J., Sherry, N.
EPI_ISL_419986	hCoV-19/Australia/VIC295/2020	Oceania / Australia / Victoria	2020-03-23	Victorian Infectious Diseases Reference Laboratory (VIDRL)	Victorian Infectious Diseases Reference Laboratory and Microbiological Diagnostic Unit Public Health Laboratory, Doherty Institute	Caly L., Seemann T., Sait, M., Schultz M., Druce J., Sherry, N.
EPI_ISL_419987	hCoV-19/Australia/VIC296/2020	Oceania / Australia / Victoria	2020-03-23	Victorian

Infectious Diseases Reference Laboratory (VIDRL)	Victorian Infectious Diseases Reference Laboratory and Microbiological Diagnostic Unit Public Health Laboratory, Doherty Institute	Caly L., Seemann T., Sait, M., Schultz M., Druce J., Sherry, N.
EPI_ISL_419988	hCoV-19/Australia/VIC297/2020	Oceania / Australia / Victoria	2020-03-23	Victorian Infectious Diseases Reference Laboratory (VIDRL)	Victorian Infectious Diseases Reference Laboratory and Microbiological Diagnostic Unit Public Health Laboratory, Doherty Institute	Caly L., Seemann T., Sait, M., Schultz M., Druce J., Sherry, N.
EPI_ISL_419989	hCoV-19/Australia/VIC298/2020	Oceania / Australia / Victoria	2020-03-23	Victorian Infectious Diseases Reference Laboratory (VIDRL)	Victorian Infectious Diseases Reference Laboratory and Microbiological Diagnostic Unit Public Health Laboratory, Doherty Institute	Caly L., Seemann T., Sait, M., Schultz M., Druce J., Sherry, N.
EPI_ISL_419990	hCoV-19/Australia/VIC299/2020	Oceania / Australia / Victoria	2020-03-23	Victorian Infectious Diseases Reference Laboratory (VIDRL)	Victorian Infectious Diseases Reference Laboratory and Microbiological Diagnostic Unit Public Health Laboratory, Doherty Institute	Caly L., Seemann T., Sait, M., Schultz M., Druce J., Sherry, N.
EPI_ISL_419991	hCoV-19/Australia/VIC300/2020	Oceania / Australia / Victoria	2020-03-23	Victorian Infectious Diseases Reference Laboratory (VIDRL)	Victorian Infectious Diseases Reference Laboratory and Microbiological Diagnostic Unit Public Health Laboratory, Doherty Institute	Caly L., Seemann T., Sait, M., Schultz M., Druce J., Sherry, N.
EPI_ISL_419992	hCoV-19/Australia/VIC301/2020	Oceania / Australia / Victoria	2020-03-23	Victorian Infectious Diseases Reference Laboratory (VIDRL)	Victorian Infectious Diseases Reference Laboratory and Microbiological Diagnostic Unit Public Health Laboratory, Doherty Institute	Caly L., Seemann T., Sait, M., Schultz M., Druce J., Sherry, N.
EPI_ISL_419993	hCoV-19/Australia/VIC302/2020	Oceania / Australia / Victoria	2020-03-23	Victorian Infectious Diseases Reference Laboratory (VIDRL)	Victorian Infectious Diseases Reference Laboratory and Microbiological Diagnostic Unit Public Health Laboratory, Doherty Institute	Caly L., Seemann T., Sait, M., Schultz M., Druce J., Sherry, N.
EPI_ISL_419994	hCoV-19/Australia/VIC303/2020	Oceania / Australia / Victoria	2020-03-23	Victorian Infectious Diseases Reference Laboratory (VIDRL)	Victorian Infectious Diseases Reference Laboratory and Microbiological Diagnostic Unit Public Health Laboratory, Doherty Institute	Caly L., Seemann T., Sait, M., Schultz M., Druce J., Sherry, N.
EPI_ISL_419995	hCoV-19/Australia/VIC304/2020	Oceania / Australia / Victoria	2020-03-23	Victorian Infectious Diseases Reference Laboratory (VIDRL)	Victorian Infectious Diseases Reference Laboratory and Microbiological Diagnostic Unit Public Health Laboratory, Doherty Institute	Caly L., Seemann T., Sait, M., Schultz M., Druce J., Sherry, N.
EPI_ISL_419996	hCoV-19/Australia/VIC305/2020	Oceania / Australia / Victoria	2020-03-23	Victorian Infectious Diseases Reference Laboratory (VIDRL)	Victorian Infectious Diseases Reference Laboratory and Microbiological Diagnostic Unit Public Health Laboratory, Doherty Institute	Caly L., Seemann T., Sait, M., Schultz M., Druce J., Sherry, N.
EPI_ISL_419997	hCoV-19/Australia/VIC306/2020	Oceania / Australia / Victoria	2020-03-23	Victorian Infectious Diseases Reference Laboratory (VIDRL)	Victorian Infectious Diseases Reference Laboratory and Microbiological Diagnostic Unit Public Health Laboratory, Doherty Institute	Caly L., Seemann T., Sait, M., Schultz M., Druce J., Sherry, N.
EPI_ISL_419998	hCoV-19/Australia/VIC307/2020	Oceania / Australia / Victoria	2020-03-23	Victorian Infectious Diseases Reference Laboratory (VIDRL)	Victorian Infectious Diseases Reference Laboratory and Microbiological Diagnostic Unit Public Health Laboratory, Doherty Institute	Caly L., Seemann T., Sait, M., Schultz M., Druce J., Sherry, N.
EPI_ISL_419999	hCoV-19/Australia/VIC308/2020	Oceania / Australia / Victoria	2020-03-23	Microbiological Diagnostic Unit Public Health Laboratory	Microbiological Diagnostic Unit Public Health Laboratory	Seemann T., Schultz M., Sait, M., Sherry, N.
EPI_ISL_420901	hCoV-19/Germany/BAV-V2010840/2020	Europe / Germany / Munich	2020-03-12	Max von Pettenkofer Institute, Virology, National Reference Center for Retroviruses, LMU Munich	Laboratory for Functional Genome Analysis, Dept. Genomics, Gene Center of the LMU Munich	Max Muenchhoff, Stefan Krebs, Alexander Graf, Ashok Varadharajan, Oliver Keppler, Helmut Blum
EPI_ISL_420902	hCoV-19/Germany/BAV-V2011376/2020	Europe / Germany / Munich	2020-03-13	Max von Pettenkofer Institute, Virology, National Reference Center for Retroviruses, LMU Munich	Laboratory for Functional Genome Analysis, Dept. Genomics, Gene Center of the LMU Munich	Max Muenchhoff, Stefan Krebs, Alexander Graf, Ashok Varadharajan, Oliver Keppler, Helmut Blum
EPI_ISL_420903	hCoV-19/Germany/BAV-V2011395/2020	Europe / Germany / Munich	2020-03-14	Max von Pettenkofer Institute, Virology, National Reference Center for Retroviruses, LMU Munich	Laboratory for Functional Genome Analysis, Dept. Genomics, Gene Center of the LMU Munich	Max Muenchhoff, Stefan Krebs, Alexander Graf, Ashok Varadharajan, Oliver Keppler, Helmut Blum
EPI_ISL_420906	hCoV-19/Germany/BAV-V2011673/2020	Europe / Germany / Munich	2020-03-16	Max von Pettenkofer Institute, Virology, National Reference Center for Retroviruses, LMU Munich	Laboratory for Functional Genome Analysis, Dept. Genomics, Gene Center of the LMU Munich	Max Muenchhoff, Stefan Krebs, Alexander Graf, Ashok Varadharajan, Oliver Keppler, Helmut Blum
EPI_ISL_420907	hCoV-19/Germany/BAV-V2011675/2020	Europe / Germany / Munich	2020-03-16	Max von Pettenkofer Institute, Virology, National Reference Center for Retroviruses, LMU Munich	Laboratory for Functional Genome Analysis, Dept. Genomics, Gene Center of the LMU Munich	Max Muenchhoff, Stefan Krebs, Alexander Graf, Ashok Varadharajan, Oliver Keppler, Helmut Blum

EPI_ISL_420910 hCoV-19/England/20136087804/2020 Europe / United Kingdom / England 2020-03-25 Respiratory Virus Unit, Microbiology Services Colindale, Public Health England Respiratory Virus Unit, Microbiology Services Colindale, Public Health England Monica Galiano, Shahjahan Miah, Angie Lackenby, Omolola Akinbami, Tiina Talts, Leena Bhaw, Richard Myers, Steven Platt, Kirstin Edwards, Jonathan Hubb, Joanna Ellis, Maria Zambon
EPI_ISL_420912 hCoV-19/Germany/BAV-V2012856/2020 Europe / Germany / Munich 2020-03-21 Max von Pettenkofer Institute, Virology, National Reference Center for Retroviruses, LMU Munich Laboratory for Functional Genome Analysis, Dept. Genomics, Gene Center of the LMU Munich Max Muenchhoff, Stefan Krebs, Alexander Graf, Ashok Varadharajan, Oliver Keppler, Helmut Blum
EPI_ISL_420929 hCoV-19/Wales/PHWC-25005/2020 Europe / United Kingdom / Wales 2020-03-20 Wales Specialist Virology Centre Public Health Wales Microbiology Cardiff Catherine Moore, Joanne Watkins, Sally Corden, Malorie Perry, Simon Cottrell Sara Rey, Matt Bull, Tom Connor
EPI_ISL_420937 hCoV-19/Wales/PHWC-24CB7/2020 Europe / United Kingdom / Wales 2020-03-21 Wales Specialist Virology Centre Public Health Wales Microbiology Cardiff Catherine Moore, Joanne Watkins, Sally Corden, Malorie Perry, Simon Cottrell Sara Rey, Matt Bull, Tom Connor
EPI_ISL_420940 hCoV-19/Wales/PHWC-24DD2/2020 Europe / United Kingdom / Wales 2020-03-21 Wales Specialist Virology Centre Public Health Wales Microbiology Cardiff Catherine Moore, Joanne Watkins, Sally Corden, Malorie Perry, Simon Cottrell Sara Rey, Matt Bull, Tom Connor
EPI_ISL_420943 hCoV-19/Wales/PHWC-24F36/2020 Europe / United Kingdom / Wales 2020-03-21 Wales Specialist Virology Centre Public Health Wales Microbiology Cardiff Catherine Moore, Joanne Watkins, Sally Corden, Malorie Perry, Simon Cottrell Sara Rey, Matt Bull, Tom Connor
EPI_ISL_420944 hCoV-19/Wales/PHWC-24FFA/2020 Europe / United Kingdom / Wales 2020-03-21 Wales Specialist Virology Centre Public Health Wales Microbiology Cardiff Catherine Moore, Joanne Watkins, Sally Corden, Malorie Perry, Simon Cottrell Sara Rey, Matt Bull, Tom Connor
EPI_ISL_420949 hCoV-19/Wales/PHWC-2522D/2020 Europe / United Kingdom / Wales 2020-03-21 Wales Specialist Virology Centre Public Health Wales Microbiology Cardiff Catherine Moore, Joanne Watkins, Sally Corden, Malorie Perry, Simon Cottrell Sara Rey, Matt Bull, Tom Connor
EPI_ISL_420951 hCoV-19/Wales/PHWC-24CF3/2020 Europe / United Kingdom / Wales 2020-03-22 Wales Specialist Virology Centre Public Health Wales Microbiology Cardiff Catherine Moore, Joanne Watkins, Sally Corden, Malorie Perry, Simon Cottrell Sara Rey, Matt Bull, Tom Connor
EPI_ISL_420960 hCoV-19/Wales/PHWC-24F72/2020 Europe / United Kingdom / Wales 2020-03-22 Wales Specialist Virology Centre Public Health Wales Microbiology Cardiff Catherine Moore, Joanne Watkins, Sally Corden, Malorie Perry, Simon Cottrell Sara Rey, Matt Bull, Tom Connor
EPI_ISL_420961 hCoV-19/Wales/PHWC-24F81/2020 Europe / United Kingdom / Wales 2020-03-22 Wales Specialist Virology Centre Public Health Wales Microbiology Cardiff Catherine Moore, Joanne Watkins, Sally Corden, Malorie Perry, Simon Cottrell Sara Rey, Matt Bull, Tom Connor
EPI_ISL_420969 hCoV-19/Wales/PHWC-25111/2020 Europe / United Kingdom / Wales 2020-03-22 Wales Specialist Virology Centre Public Health Wales Microbiology Cardiff Catherine Moore, Joanne Watkins, Sally Corden, Malorie Perry, Simon Cottrell Sara Rey, Matt Bull, Tom Connor
EPI_ISL_420970 hCoV-19/Wales/PHWC-25120/2020 Europe / United Kingdom / Wales 2020-03-22 Wales Specialist Virology Centre Public Health Wales Microbiology Cardiff Catherine Moore, Joanne Watkins, Sally Corden, Malorie Perry, Simon Cottrell Sara Rey, Matt Bull, Tom Connor
EPI_ISL_420974 hCoV-19/Wales/PHWC-2516C/2020 Europe / United Kingdom / Wales 2020-03-22 Wales Specialist Virology Centre Public Health Wales Microbiology Cardiff Catherine Moore, Joanne Watkins, Sally Corden, Malorie Perry, Simon Cottrell Sara Rey, Matt Bull, Tom Connor
EPI_ISL_420976 hCoV-19/Wales/PHWC-251B7/2020 Europe / United Kingdom / Wales 2020-03-22 Wales Specialist Virology Centre Public Health Wales Microbiology Cardiff Catherine Moore, Joanne Watkins, Sally Corden, Malorie Perry, Simon Cottrell Sara Rey, Matt Bull, Tom Connor
EPI_ISL_420977 hCoV-19/Wales/PHWC-2523C/2020 Europe / United Kingdom / Wales 2020-03-22 Wales Specialist Virology Centre Public Health Wales Microbiology Cardiff Catherine Moore, Joanne Watkins, Sally Corden, Malorie Perry, Simon Cottrell Sara Rey, Matt Bull, Tom Connor
EPI_ISL_420978 hCoV-19/Wales/PHWC-253B1/2020 Europe / United Kingdom / Wales 2020-03-22 Wales Specialist Virology Centre Public Health Wales Microbiology Cardiff Catherine Moore, Joanne Watkins, Sally Corden, Malorie Perry, Simon Cottrell Sara Rey, Matt Bull, Tom Connor
EPI_ISL_420982 hCoV-19/Wales/PHWC-251D5/2020 Europe / United Kingdom / Wales 2020-03-23 Wales Specialist Virology Centre Public Health Wales Microbiology Cardiff Catherine Moore, Joanne Watkins, Sally Corden, Malorie Perry, Simon Cottrell Sara Rey, Matt Bull, Tom Connor
EPI_ISL_420985 hCoV-19/Wales/PHWC-252B4/2020 Europe / United Kingdom / Wales 2020-03-23 Wales Specialist Virology Centre Public Health Wales Microbiology Cardiff Catherine Moore, Joanne Watkins, Sally Corden, Malorie Perry, Simon Cottrell Sara Rey, Matt Bull, Tom Connor
EPI_ISL_420988 hCoV-19/Wales/PHWC-253DF/2020 Europe / United Kingdom / Wales 2020-03-23 Wales Specialist Virology Centre Public Health Wales Microbiology Cardiff Catherine Moore, Joanne Watkins, Sally Corden, Malorie Perry, Simon Cottrell Sara Rey, Matt Bull, Tom Connor
EPI_ISL_420990 hCoV-19/Wales/PHWC-25418/2020 Europe / United Kingdom / Wales 2020-03-23 Wales Specialist Virology Centre Public Health Wales Microbiology Cardiff Catherine Moore, Joanne Watkins, Sally Corden, Malorie Perry, Simon Cottrell Sara Rey, Matt Bull, Tom Connor
EPI_ISL_420994 hCoV-19/Wales/PHWC-25463/2020 Europe / United Kingdom / Wales 2020-03-23 Wales Specialist Virology Centre Public Health Wales Microbiology Cardiff Catherine Moore, Joanne Watkins, Sally Corden, Malorie Perry, Simon Cottrell Sara Rey, Matt Bull, Tom Connor
EPI_ISL_420995 hCoV-19/Wales/PHWC-25472/2020 Europe / United Kingdom / Wales 2020-03-23 Wales

Specialist Virology Centre	Public Health Wales Microbiology Cardiff	Catherine Moore, Joanne Watkins, Sally Corden, Malorie Perry, Simon Cottrell Sara Rey, Matt Bull, Tom Connor
EPI_ISL_420998	hCoV-19/Wales/PHWC-254AF/2020	Europe / United Kingdom / Wales 2020-03-23	Wales Specialist Virology Centre	Public Health Wales Microbiology Cardiff	Catherine Moore, Joanne Watkins, Sally Corden, Malorie Perry, Simon Cottrell Sara Rey, Matt Bull, Tom Connor
EPI_ISL_423100	hCoV-19/England/20134018902/2020	Europe / United Kingdom / England	2020-03-24	Respiratory Virus Unit, Microbiology Services Colindale, Public Health England	Respiratory Virus Unit, Microbiology Services Colindale, Public Health England	Monica Galiano, Shahjahan Miah, Angie Lackenby, Omolola Akinbami, Tiina Talts, Leena Bhaw, Richard Myers, Steven Platt, Kirstin Edwards, Jonathan Hubb, Joanna Ellis, Maria Zambon
EPI_ISL_423101	hCoV-19/England/20134019102/2020	Europe / United Kingdom / England	2020-03-24	Respiratory Virus Unit, Microbiology Services Colindale, Public Health England	Respiratory Virus Unit, Microbiology Services Colindale, Public Health England	Monica Galiano, Shahjahan Miah, Angie Lackenby, Omolola Akinbami, Tiina Talts, Leena Bhaw, Richard Myers, Steven Platt, Kirstin Edwards, Jonathan Hubb, Joanna Ellis, Maria Zambon
EPI_ISL_423102	hCoV-19/England/20134019202/2020	Europe / United Kingdom / England	2020-03-24	Respiratory Virus Unit, Microbiology Services Colindale, Public Health England	Respiratory Virus Unit, Microbiology Services Colindale, Public Health England	Monica Galiano, Shahjahan Miah, Angie Lackenby, Omolola Akinbami, Tiina Talts, Leena Bhaw, Richard Myers, Steven Platt, Kirstin Edwards, Jonathan Hubb, Joanna Ellis, Maria Zambon
EPI_ISL_423104	hCoV-19/England/20134019402/2020	Europe / United Kingdom / England	2020-03-23	Respiratory Virus Unit, Microbiology Services Colindale, Public Health England	Respiratory Virus Unit, Microbiology Services Colindale, Public Health England	Monica Galiano, Shahjahan Miah, Angie Lackenby, Omolola Akinbami, Tiina Talts, Leena Bhaw, Richard Myers, Steven Platt, Kirstin Edwards, Jonathan Hubb, Joanna Ellis, Maria Zambon
EPI_ISL_423105	hCoV-19/England/20134019502/2020	Europe / United Kingdom / England	2020-03-23	Respiratory Virus Unit, Microbiology Services Colindale, Public Health England	Respiratory Virus Unit, Microbiology Services Colindale, Public Health England	Monica Galiano, Shahjahan Miah, Angie Lackenby, Omolola Akinbami, Tiina Talts, Leena Bhaw, Richard Myers, Steven Platt, Kirstin Edwards, Jonathan Hubb, Joanna Ellis, Maria Zambon
EPI_ISL_423107	hCoV-19/England/20134019904/2020	Europe / United Kingdom / England	2020-03-23	Respiratory Virus Unit, Microbiology Services Colindale, Public Health England	Respiratory Virus Unit, Microbiology Services Colindale, Public Health England	Monica Galiano, Shahjahan Miah, Angie Lackenby, Omolola Akinbami, Tiina Talts, Leena Bhaw, Richard Myers, Steven Platt, Kirstin Edwards, Jonathan Hubb, Joanna Ellis, Maria Zambon
EPI_ISL_423109	hCoV-19/England/20134020104/2020	Europe / United Kingdom / England	2020-03-23	Respiratory Virus Unit, Microbiology Services Colindale, Public Health England	Respiratory Virus Unit, Microbiology Services Colindale, Public Health England	Monica Galiano, Shahjahan Miah, Angie Lackenby, Omolola Akinbami, Tiina Talts, Leena Bhaw, Richard Myers, Steven Platt, Kirstin Edwards, Jonathan Hubb, Joanna Ellis, Maria Zambon
EPI_ISL_423110	hCoV-19/England/20134020504/2020	Europe / United Kingdom / England	2020-03-24	Respiratory Virus Unit, Microbiology Services Colindale, Public Health England	Respiratory Virus Unit, Microbiology Services Colindale, Public Health England	Monica Galiano, Shahjahan Miah, Angie Lackenby, Omolola Akinbami, Tiina Talts, Leena Bhaw, Richard Myers, Steven Platt, Kirstin Edwards, Jonathan Hubb, Joanna Ellis, Maria Zambon
EPI_ISL_423111	hCoV-19/England/20134020904/2020	Europe / United Kingdom / England	2020-03-24	Respiratory Virus Unit, Microbiology Services Colindale, Public Health England	Respiratory Virus Unit, Microbiology Services Colindale, Public Health England	Monica Galiano, Shahjahan Miah, Angie Lackenby, Omolola Akinbami, Tiina Talts, Leena Bhaw, Richard Myers, Steven Platt, Kirstin Edwards, Jonathan Hubb, Joanna Ellis, Maria Zambon
EPI_ISL_423112	hCoV-19/England/20134021004/2020	Europe / United Kingdom / England	2020-03-24	Respiratory Virus Unit, Microbiology Services Colindale, Public Health England	Respiratory Virus Unit, Microbiology Services Colindale, Public Health England	Monica Galiano, Shahjahan Miah, Angie Lackenby, Omolola Akinbami, Tiina Talts, Leena Bhaw, Richard Myers, Steven Platt, Kirstin Edwards, Jonathan Hubb, Joanna Ellis, Maria Zambon
EPI_ISL_423113	hCoV-19/England/20134021104/2020	Europe / United Kingdom / England	2020-03-24	Respiratory Virus Unit, Microbiology Services Colindale, Public Health England	Respiratory Virus Unit, Microbiology Services Colindale, Public Health England	Monica Galiano, Shahjahan Miah, Angie Lackenby, Omolola Akinbami, Tiina Talts, Leena Bhaw, Richard Myers, Steven Platt, Kirstin Edwards, Jonathan Hubb, Joanna Ellis, Maria Zambon
EPI_ISL_423117	hCoV-19/England/20134021804/2020	Europe / United Kingdom / England	2020-03-24	Respiratory Virus Unit, Microbiology Services Colindale, Public Health England	Respiratory Virus Unit, Microbiology Services Colindale, Public Health England	Monica Galiano, Shahjahan Miah, Angie Lackenby, Omolola Akinbami, Tiina Talts, Leena Bhaw, Richard Myers, Steven Platt, Kirstin Edwards, Jonathan Hubb, Joanna Ellis, Maria Zambon
EPI_ISL_423118	hCoV-19/England/20134021904/2020	Europe / United Kingdom / England	2020-03-23	Respiratory Virus Unit, Microbiology Services Colindale, Public Health England	Respiratory Virus Unit, Microbiology Services Colindale, Public Health England	Monica Galiano, Shahjahan Miah, Angie Lackenby, Omolola Akinbami, Tiina Talts, Leena Bhaw, Richard Myers, Steven Platt, Kirstin Edwards, Jonathan Hubb, Joanna Ellis, Maria Zambon

EPI_ISL_423119  hCoV-19/England/20134022104/2020        Europe / United Kingdom / England       2020-03-25
Respiratory Virus Unit, Microbiology Services Colindale, Public Health England  Respiratory Virus Unit, Microbiology Services Colindale, Public Health England  Monica Galiano, Shahjahan Miah, Angie Lackenby, Omolola Akinbami, Tiina Talts, Leena Bhaw, Richard Myers, Steven Platt, Kirstin Edwards, Jonathan Hubb, Joanna Ellis, Maria Zambon
EPI_ISL_423121  hCoV-19/England/20134023004/2020        Europe / United Kingdom / England       2020-03-24
Respiratory Virus Unit, Microbiology Services Colindale, Public Health England  Respiratory Virus Unit, Microbiology Services Colindale, Public Health England  Monica Galiano, Shahjahan Miah, Angie Lackenby, Omolola Akinbami, Tiina Talts, Leena Bhaw, Richard Myers, Steven Platt, Kirstin Edwards, Jonathan Hubb, Joanna Ellis, Maria Zambon
EPI_ISL_423122  hCoV-19/England/20134023104/2020        Europe / United Kingdom / England       2020-03-24
Respiratory Virus Unit, Microbiology Services Colindale, Public Health England  Respiratory Virus Unit, Microbiology Services Colindale, Public Health England  Monica Galiano, Shahjahan Miah, Angie Lackenby, Omolola Akinbami, Tiina Talts, Leena Bhaw, Richard Myers, Steven Platt, Kirstin Edwards, Jonathan Hubb, Joanna Ellis, Maria Zambon
EPI_ISL_423123  hCoV-19/England/20134023204/2020        Europe / United Kingdom / England       2020-03-24
Respiratory Virus Unit, Microbiology Services Colindale, Public Health England  Respiratory Virus Unit, Microbiology Services Colindale, Public Health England  Monica Galiano, Shahjahan Miah, Angie Lackenby, Omolola Akinbami, Tiina Talts, Leena Bhaw, Richard Myers, Steven Platt, Kirstin Edwards, Jonathan Hubb, Joanna Ellis, Maria Zambon
EPI_ISL_423124  hCoV-19/England/20134023704/2020        Europe / United Kingdom / England       2020-03-24
Respiratory Virus Unit, Microbiology Services Colindale, Public Health England  Respiratory Virus Unit, Microbiology Services Colindale, Public Health England  Monica Galiano, Shahjahan Miah, Angie Lackenby, Omolola Akinbami, Tiina Talts, Leena Bhaw, Richard Myers, Steven Platt, Kirstin Edwards, Jonathan Hubb, Joanna Ellis, Maria Zambon
EPI_ISL_423126  hCoV-19/England/20134024104/2020        Europe / United Kingdom / England       2020-03-24
Respiratory Virus Unit, Microbiology Services Colindale, Public Health England  Respiratory Virus Unit, Microbiology Services Colindale, Public Health England  Monica Galiano, Shahjahan Miah, Angie Lackenby, Omolola Akinbami, Tiina Talts, Leena Bhaw, Richard Myers, Steven Platt, Kirstin Edwards, Jonathan Hubb, Joanna Ellis, Maria Zambon
EPI_ISL_423128  hCoV-19/England/20134024504/2020        Europe / United Kingdom / England       2020-03-23
Respiratory Virus Unit, Microbiology Services Colindale, Public Health England  Respiratory Virus Unit, Microbiology Services Colindale, Public Health England  Monica Galiano, Shahjahan Miah, Angie Lackenby, Omolola Akinbami, Tiina Talts, Leena Bhaw, Richard Myers, Steven Platt, Kirstin Edwards, Jonathan Hubb, Joanna Ellis, Maria Zambon
EPI_ISL_423129  hCoV-19/England/20134025204/2020        Europe / United Kingdom / England       2020-03-23
Respiratory Virus Unit, Microbiology Services Colindale, Public Health England  Respiratory Virus Unit, Microbiology Services Colindale, Public Health England  Monica Galiano, Shahjahan Miah, Angie Lackenby, Omolola Akinbami, Tiina Talts, Leena Bhaw, Richard Myers, Steven Platt, Kirstin Edwards, Jonathan Hubb, Joanna Ellis, Maria Zambon
EPI_ISL_423130  hCoV-19/England/20134025504/2020        Europe / United Kingdom / England       2020-03-24
Respiratory Virus Unit, Microbiology Services Colindale, Public Health England  Respiratory Virus Unit, Microbiology Services Colindale, Public Health England  Monica Galiano, Shahjahan Miah, Angie Lackenby, Omolola Akinbami, Tiina Talts, Leena Bhaw, Richard Myers, Steven Platt, Kirstin Edwards, Jonathan Hubb, Joanna Ellis, Maria Zambon
EPI_ISL_423131  hCoV-19/England/20134025704/2020        Europe / United Kingdom / England       2020-03-24
Respiratory Virus Unit, Microbiology Services Colindale, Public Health England  Respiratory Virus Unit, Microbiology Services Colindale, Public Health England  Monica Galiano, Shahjahan Miah, Angie Lackenby, Omolola Akinbami, Tiina Talts, Leena Bhaw, Richard Myers, Steven Platt, Kirstin Edwards, Jonathan Hubb, Joanna Ellis, Maria Zambon
EPI_ISL_423132  hCoV-19/England/20134025904/2020        Europe / United Kingdom / England       2020-03-24
Respiratory Virus Unit, Microbiology Services Colindale, Public Health England  Respiratory Virus Unit, Microbiology Services Colindale, Public Health England  Monica Galiano, Shahjahan Miah, Angie Lackenby, Omolola Akinbami, Tiina Talts, Leena Bhaw, Richard Myers, Steven Platt, Kirstin Edwards, Jonathan Hubb, Joanna Ellis, Maria Zambon
EPI_ISL_423134  hCoV-19/England/20134026904/2020        Europe / United Kingdom / England       2020-03-23
Respiratory Virus Unit, Microbiology Services Colindale, Public Health England  Respiratory Virus Unit, Microbiology Services Colindale, Public Health England  Monica Galiano, Shahjahan Miah, Angie Lackenby, Omolola Akinbami, Tiina Talts, Leena Bhaw, Richard Myers, Steven Platt, Kirstin Edwards, Jonathan Hubb, Joanna Ellis, Maria Zambon
EPI_ISL_423136  hCoV-19/England/20134027504/2020        Europe / United Kingdom / England       2020-03-23
Respiratory Virus Unit, Microbiology Services Colindale, Public Health England  Respiratory Virus Unit, Microbiology Services Colindale, Public Health England  Monica Galiano, Shahjahan Miah, Angie Lackenby, Omolola Akinbami, Tiina Talts, Leena Bhaw, Richard Myers, Steven Platt, Kirstin Edwards, Jonathan Hubb, Joanna Ellis, Maria Zambon
EPI_ISL_423142  hCoV-19/England/20134047404/2020        Europe / United Kingdom / England       2020-03-24
Respiratory Virus Unit, Microbiology Services Colindale, Public Health England  Respiratory Virus Unit, Microbiology Services Colindale, Public Health England  Monica Galiano, Shahjahan Miah, Angie Lackenby, Omolola Akinbami, Tiina Talts, Leena Bhaw, Richard Myers, Steven Platt, Kirstin Edwards, Jonathan Hubb, Joanna Ellis, Maria Zambon

EPI_ISL_423144  hCoV-19/England/20134059604/2020        Europe / United Kingdom / England       2020-03-24
Respiratory Virus Unit, Microbiology Services Colindale, Public Health England  Respiratory Virus Unit, Microbiology Services Colindale, Public Health England  Monica Galiano, Shahjahan Miah, Angie Lackenby, Omolola Akinbami, Tiina Talts, Leena Bhaw, Richard Myers, Steven Platt, Kirstin Edwards, Jonathan Hubb, Joanna Ellis, Maria Zambon
EPI_ISL_423145  hCoV-19/England/20134059704/2020        Europe / United Kingdom / England       2020-03-24
Respiratory Virus Unit, Microbiology Services Colindale, Public Health England  Respiratory Virus Unit, Microbiology Services Colindale, Public Health England  Monica Galiano, Shahjahan Miah, Angie Lackenby, Omolola Akinbami, Tiina Talts, Leena Bhaw, Richard Myers, Steven Platt, Kirstin Edwards, Jonathan Hubb, Joanna Ellis, Maria Zambon
EPI_ISL_423147  hCoV-19/England/20134060504/2020        Europe / United Kingdom / England       2020-03-23
Respiratory Virus Unit, Microbiology Services Colindale, Public Health England  Respiratory Virus Unit, Microbiology Services Colindale, Public Health England  Monica Galiano, Shahjahan Miah, Angie Lackenby, Omolola Akinbami, Tiina Talts, Leena Bhaw, Richard Myers, Steven Platt, Kirstin Edwards, Jonathan Hubb, Joanna Ellis, Maria Zambon
EPI_ISL_423148  hCoV-19/England/20134060804/2020        Europe / United Kingdom / England       2020-03-24
Respiratory Virus Unit, Microbiology Services Colindale, Public Health England  Respiratory Virus Unit, Microbiology Services Colindale, Public Health England  Monica Galiano, Shahjahan Miah, Angie Lackenby, Omolola Akinbami, Tiina Talts, Leena Bhaw, Richard Myers, Steven Platt, Kirstin Edwards, Jonathan Hubb, Joanna Ellis, Maria Zambon
EPI_ISL_423149  hCoV-19/England/20134060904/2020        Europe / United Kingdom / England       2020-03-21
Respiratory Virus Unit, Microbiology Services Colindale, Public Health England  Respiratory Virus Unit, Microbiology Services Colindale, Public Health England  Monica Galiano, Shahjahan Miah, Angie Lackenby, Omolola Akinbami, Tiina Talts, Leena Bhaw, Richard Myers, Steven Platt, Kirstin Edwards, Jonathan Hubb, Joanna Ellis, Maria Zambon
EPI_ISL_423150  hCoV-19/England/20134064004/2020        Europe / United Kingdom / England       2020-03-23
Respiratory Virus Unit, Microbiology Services Colindale, Public Health England  Respiratory Virus Unit, Microbiology Services Colindale, Public Health England  Monica Galiano, Shahjahan Miah, Angie Lackenby, Omolola Akinbami, Tiina Talts, Leena Bhaw, Richard Myers, Steven Platt, Kirstin Edwards, Jonathan Hubb, Joanna Ellis, Maria Zambon
EPI_ISL_423151  hCoV-19/England/20134064204/2020        Europe / United Kingdom / England       2020-03-24
Respiratory Virus Unit, Microbiology Services Colindale, Public Health England  Respiratory Virus Unit, Microbiology Services Colindale, Public Health England  Monica Galiano, Shahjahan Miah, Angie Lackenby, Omolola Akinbami, Tiina Talts, Leena Bhaw, Richard Myers, Steven Platt, Kirstin Edwards, Jonathan Hubb, Joanna Ellis, Maria Zambon
EPI_ISL_423152  hCoV-19/England/20134064904/2020        Europe / United Kingdom / England       2020-03-24
Respiratory Virus Unit, Microbiology Services Colindale, Public Health England  Respiratory Virus Unit, Microbiology Services Colindale, Public Health England  Monica Galiano, Shahjahan Miah, Angie Lackenby, Omolola Akinbami, Tiina Talts, Leena Bhaw, Richard Myers, Steven Platt, Kirstin Edwards, Jonathan Hubb, Joanna Ellis, Maria Zambon
EPI_ISL_423154  hCoV-19/England/20134067304/2020        Europe / United Kingdom / England       2020-03-24
Respiratory Virus Unit, Microbiology Services Colindale, Public Health England  Respiratory Virus Unit, Microbiology Services Colindale, Public Health England  Monica Galiano, Shahjahan Miah, Angie Lackenby, Omolola Akinbami, Tiina Talts, Leena Bhaw, Richard Myers, Steven Platt, Kirstin Edwards, Jonathan Hubb, Joanna Ellis, Maria Zambon
EPI_ISL_423156  hCoV-19/England/20134067904/2020        Europe / United Kingdom / England       2020-03-24
Respiratory Virus Unit, Microbiology Services Colindale, Public Health England  Respiratory Virus Unit, Microbiology Services Colindale, Public Health England  Monica Galiano, Shahjahan Miah, Angie Lackenby, Omolola Akinbami, Tiina Talts, Leena Bhaw, Richard Myers, Steven Platt, Kirstin Edwards, Jonathan Hubb, Joanna Ellis, Maria Zambon
EPI_ISL_423157  hCoV-19/England/20134068504/2020        Europe / United Kingdom / England       2020-03-24
Respiratory Virus Unit, Microbiology Services Colindale, Public Health England  Respiratory Virus Unit, Microbiology Services Colindale, Public Health England  Monica Galiano, Shahjahan Miah, Angie Lackenby, Omolola Akinbami, Tiina Talts, Leena Bhaw, Richard Myers, Steven Platt, Kirstin Edwards, Jonathan Hubb, Joanna Ellis, Maria Zambon
EPI_ISL_423158  hCoV-19/England/20134068604/2020        Europe / United Kingdom / England       2020-03-24
Respiratory Virus Unit, Microbiology Services Colindale, Public Health England  Respiratory Virus Unit, Microbiology Services Colindale, Public Health England  Monica Galiano, Shahjahan Miah, Angie Lackenby, Omolola Akinbami, Tiina Talts, Leena Bhaw, Richard Myers, Steven Platt, Kirstin Edwards, Jonathan Hubb, Joanna Ellis, Maria Zambon
EPI_ISL_423159  hCoV-19/England/20134068704/2020        Europe / United Kingdom / England       2020-03-24
Respiratory Virus Unit, Microbiology Services Colindale, Public Health England  Respiratory Virus Unit, Microbiology Services Colindale, Public Health England  Monica Galiano, Shahjahan Miah, Angie Lackenby, Omolola Akinbami, Tiina Talts, Leena Bhaw, Richard Myers, Steven Platt, Kirstin Edwards, Jonathan Hubb, Joanna Ellis, Maria Zambon
EPI_ISL_423160  hCoV-19/England/20134068904/2020        Europe / United Kingdom / England       2020-03-24
Respiratory Virus Unit, Microbiology Services Colindale, Public Health England  Respiratory Virus Unit, Microbiology Services Colindale, Public Health England  Monica Galiano, Shahjahan Miah, Angie Lackenby, Omolola Akinbami, Tiina Talts, Leena Bhaw, Richard Myers, Steven Platt, Kirstin Edwards, Jonathan Hubb, Joanna Ellis, Maria Zambon

EPI_ISL_423161  hCoV-19/England/20134069004/2020        Europe / United Kingdom / England       2020-03-24
Respiratory Virus Unit, Microbiology Services Colindale, Public Health England  Respiratory Virus Unit, Microbiology Services Colindale, Public Health England  Monica Galiano, Shahjahan Miah, Angie Lackenby, Omolola Akinbami, Tiina Talts, Leena Bhaw, Richard Myers, Steven Platt, Kirstin Edwards, Jonathan Hubb, Joanna Ellis, Maria Zambon
EPI_ISL_423162  hCoV-19/England/20134071504/2020        Europe / United Kingdom / England       2020-03-24
Respiratory Virus Unit, Microbiology Services Colindale, Public Health England  Respiratory Virus Unit, Microbiology Services Colindale, Public Health England  Monica Galiano, Shahjahan Miah, Angie Lackenby, Omolola Akinbami, Tiina Talts, Leena Bhaw, Richard Myers, Steven Platt, Kirstin Edwards, Jonathan Hubb, Joanna Ellis, Maria Zambon
EPI_ISL_423163  hCoV-19/England/20134071804/2020        Europe / United Kingdom / England       2020-03-23
Respiratory Virus Unit, Microbiology Services Colindale, Public Health England  Respiratory Virus Unit, Microbiology Services Colindale, Public Health England  Monica Galiano, Shahjahan Miah, Angie Lackenby, Omolola Akinbami, Tiina Talts, Leena Bhaw, Richard Myers, Steven Platt, Kirstin Edwards, Jonathan Hubb, Joanna Ellis, Maria Zambon
EPI_ISL_423170  hCoV-19/England/20134077504/2020        Europe / United Kingdom / England       2020-03-24
Respiratory Virus Unit, Microbiology Services Colindale, Public Health England  Respiratory Virus Unit, Microbiology Services Colindale, Public Health England  Monica Galiano, Shahjahan Miah, Angie Lackenby, Omolola Akinbami, Tiina Talts, Leena Bhaw, Richard Myers, Steven Platt, Kirstin Edwards, Jonathan Hubb, Joanna Ellis, Maria Zambon
EPI_ISL_423172  hCoV-19/England/20134078004/2020        Europe / United Kingdom / England       2020-03-24
Respiratory Virus Unit, Microbiology Services Colindale, Public Health England  Respiratory Virus Unit, Microbiology Services Colindale, Public Health England  Monica Galiano, Shahjahan Miah, Angie Lackenby, Omolola Akinbami, Tiina Talts, Leena Bhaw, Richard Myers, Steven Platt, Kirstin Edwards, Jonathan Hubb, Joanna Ellis, Maria Zambon
EPI_ISL_423173  hCoV-19/England/20134078204/2020        Europe / United Kingdom / England       2020-03-24
Respiratory Virus Unit, Microbiology Services Colindale, Public Health England  Respiratory Virus Unit, Microbiology Services Colindale, Public Health England  Monica Galiano, Shahjahan Miah, Angie Lackenby, Omolola Akinbami, Tiina Talts, Leena Bhaw, Richard Myers, Steven Platt, Kirstin Edwards, Jonathan Hubb, Joanna Ellis, Maria Zambon
EPI_ISL_423174  hCoV-19/England/20134078404/2020        Europe / United Kingdom / England       2020-03-24
Respiratory Virus Unit, Microbiology Services Colindale, Public Health England  Respiratory Virus Unit, Microbiology Services Colindale, Public Health England  Monica Galiano, Shahjahan Miah, Angie Lackenby, Omolola Akinbami, Tiina Talts, Leena Bhaw, Richard Myers, Steven Platt, Kirstin Edwards, Jonathan Hubb, Joanna Ellis, Maria Zambon
EPI_ISL_423175  hCoV-19/England/20134078604/2020        Europe / United Kingdom / England       2020-03-24
Respiratory Virus Unit, Microbiology Services Colindale, Public Health England  Respiratory Virus Unit, Microbiology Services Colindale, Public Health England  Monica Galiano, Shahjahan Miah, Angie Lackenby, Omolola Akinbami, Tiina Talts, Leena Bhaw, Richard Myers, Steven Platt, Kirstin Edwards, Jonathan Hubb, Joanna Ellis, Maria Zambon
EPI_ISL_423176  hCoV-19/England/20134078804/2020        Europe / United Kingdom / England       2020-03-24
Respiratory Virus Unit, Microbiology Services Colindale, Public Health England  Respiratory Virus Unit, Microbiology Services Colindale, Public Health England  Monica Galiano, Shahjahan Miah, Angie Lackenby, Omolola Akinbami, Tiina Talts, Leena Bhaw, Richard Myers, Steven Platt, Kirstin Edwards, Jonathan Hubb, Joanna Ellis, Maria Zambon
EPI_ISL_423177  hCoV-19/England/20134078904/2020        Europe / United Kingdom / England       2020-03-24
Respiratory Virus Unit, Microbiology Services Colindale, Public Health England  Respiratory Virus Unit, Microbiology Services Colindale, Public Health England  Monica Galiano, Shahjahan Miah, Angie Lackenby, Omolola Akinbami, Tiina Talts, Leena Bhaw, Richard Myers, Steven Platt, Kirstin Edwards, Jonathan Hubb, Joanna Ellis, Maria Zambon
EPI_ISL_423178  hCoV-19/England/20134079504/2020        Europe / United Kingdom / England       2020-03-24
Respiratory Virus Unit, Microbiology Services Colindale, Public Health England  Respiratory Virus Unit, Microbiology Services Colindale, Public Health England  Monica Galiano, Shahjahan Miah, Angie Lackenby, Omolola Akinbami, Tiina Talts, Leena Bhaw, Richard Myers, Steven Platt, Kirstin Edwards, Jonathan Hubb, Joanna Ellis, Maria Zambon
EPI_ISL_423181  hCoV-19/England/20134081904/2020        Europe / United Kingdom / England       2020-03-20
Respiratory Virus Unit, Microbiology Services Colindale, Public Health England  Respiratory Virus Unit, Microbiology Services Colindale, Public Health England  Monica Galiano, Shahjahan Miah, Angie Lackenby, Omolola Akinbami, Tiina Talts, Leena Bhaw, Richard Myers, Steven Platt, Kirstin Edwards, Jonathan Hubb, Joanna Ellis, Maria Zambon
EPI_ISL_423193  hCoV-19/England/201340875/2020  Europe / United Kingdom / England       2020-03-24
Respiratory Virus Unit, Microbiology Services Colindale, Public Health England  Respiratory Virus Unit, Microbiology Services Colindale, Public Health England  Monica Galiano, Shahjahan Miah, Angie Lackenby, Omolola Akinbami, Tiina Talts, Leena Bhaw, Richard Myers, Steven Platt, Kirstin Edwards, Jonathan Hubb, Joanna Ellis, Maria Zambon
EPI_ISL_423194  hCoV-19/England/201360001/2020  Europe / United Kingdom / England       2020-03-25
Respiratory Virus Unit, Microbiology Services Colindale, Public Health England  Respiratory Virus Unit, Microbiology Services Colindale, Public Health England  Monica Galiano, Shahjahan Miah, Angie Lackenby, Omolola Akinbami, Tiina Talts, Leena Bhaw, Richard Myers, Steven Platt, Kirstin Edwards, Jonathan Hubb, Joanna Ellis, Maria Zambon

EPI_ISL_423196 hCoV-19/England/201360015/2020 Europe / United Kingdom / England 2020-03-24
Respiratory Virus Unit, Microbiology Services Colindale, Public Health England Respiratory Virus Unit, Microbiology Services Colindale, Public Health England Monica Galiano, Shahjahan Miah, Angie Lackenby, Omolola Akinbami, Tiina Talts, Leena Bhaw, Richard Myers, Steven Platt, Kirstin Edwards, Jonathan Hubb, Joanna Ellis, Maria Zambon
EPI_ISL_423199 hCoV-19/England/201360037/2020 Europe / United Kingdom / England 2020-03-24
Respiratory Virus Unit, Microbiology Services Colindale, Public Health England Respiratory Virus Unit, Microbiology Services Colindale, Public Health England Monica Galiano, Shahjahan Miah, Angie Lackenby, Omolola Akinbami, Tiina Talts, Leena Bhaw, Richard Myers, Steven Platt, Kirstin Edwards, Jonathan Hubb, Joanna Ellis, Maria Zambon
EPI_ISL_423250 hCoV-19/England/201360220/2020 Europe / United Kingdom / England 2020-03-25
Respiratory Virus Unit, Microbiology Services Colindale, Public Health England Respiratory Virus Unit, Microbiology Services Colindale, Public Health England Monica Galiano, Shahjahan Miah, Angie Lackenby, Omolola Akinbami, Tiina Talts, Leena Bhaw, Richard Myers, Steven Platt, Kirstin Edwards, Jonathan Hubb, Joanna Ellis, Maria Zambon
EPI_ISL_423251 hCoV-19/England/201360221/2020 Europe / United Kingdom / England 2020-03-25
Respiratory Virus Unit, Microbiology Services Colindale, Public Health England Respiratory Virus Unit, Microbiology Services Colindale, Public Health England Monica Galiano, Shahjahan Miah, Angie Lackenby, Omolola Akinbami, Tiina Talts, Leena Bhaw, Richard Myers, Steven Platt, Kirstin Edwards, Jonathan Hubb, Joanna Ellis, Maria Zambon
EPI_ISL_423253 hCoV-19/England/201360223/2020 Europe / United Kingdom / England 2020-03-25
Respiratory Virus Unit, Microbiology Services Colindale, Public Health England Respiratory Virus Unit, Microbiology Services Colindale, Public Health England Monica Galiano, Shahjahan Miah, Angie Lackenby, Omolola Akinbami, Tiina Talts, Leena Bhaw, Richard Myers, Steven Platt, Kirstin Edwards, Jonathan Hubb, Joanna Ellis, Maria Zambon
EPI_ISL_423254 hCoV-19/England/201360224/2020 Europe / United Kingdom / England 2020-03-25
Respiratory Virus Unit, Microbiology Services Colindale, Public Health England Respiratory Virus Unit, Microbiology Services Colindale, Public Health England Monica Galiano, Shahjahan Miah, Angie Lackenby, Omolola Akinbami, Tiina Talts, Leena Bhaw, Richard Myers, Steven Platt, Kirstin Edwards, Jonathan Hubb, Joanna Ellis, Maria Zambon
EPI_ISL_423255 hCoV-19/England/201360229/2020 Europe / United Kingdom / England 2020-03-25
Respiratory Virus Unit, Microbiology Services Colindale, Public Health England Respiratory Virus Unit, Microbiology Services Colindale, Public Health England Monica Galiano, Shahjahan Miah, Angie Lackenby, Omolola Akinbami, Tiina Talts, Leena Bhaw, Richard Myers, Steven Platt, Kirstin Edwards, Jonathan Hubb, Joanna Ellis, Maria Zambon
EPI_ISL_423256 hCoV-19/England/201360236/2020 Europe / United Kingdom / England 2020-03-25
Respiratory Virus Unit, Microbiology Services Colindale, Public Health England Respiratory Virus Unit, Microbiology Services Colindale, Public Health England Monica Galiano, Shahjahan Miah, Angie Lackenby, Omolola Akinbami, Tiina Talts, Leena Bhaw, Richard Myers, Steven Platt, Kirstin Edwards, Jonathan Hubb, Joanna Ellis, Maria Zambon
EPI_ISL_423258 hCoV-19/England/201360249/2020 Europe / United Kingdom / England 2020-03-23
Respiratory Virus Unit, Microbiology Services Colindale, Public Health England Respiratory Virus Unit, Microbiology Services Colindale, Public Health England Monica Galiano, Shahjahan Miah, Angie Lackenby, Omolola Akinbami, Tiina Talts, Leena Bhaw, Richard Myers, Steven Platt, Kirstin Edwards, Jonathan Hubb, Joanna Ellis, Maria Zambon
EPI_ISL_423259 hCoV-19/England/201360251/2020 Europe / United Kingdom / England 2020-03-24
Respiratory Virus Unit, Microbiology Services Colindale, Public Health England Respiratory Virus Unit, Microbiology Services Colindale, Public Health England Monica Galiano, Shahjahan Miah, Angie Lackenby, Omolola Akinbami, Tiina Talts, Leena Bhaw, Richard Myers, Steven Platt, Kirstin Edwards, Jonathan Hubb, Joanna Ellis, Maria Zambon
EPI_ISL_423261 hCoV-19/England/201360276/2020 Europe / United Kingdom / England 2020-03-25
Respiratory Virus Unit, Microbiology Services Colindale, Public Health England Respiratory Virus Unit, Microbiology Services Colindale, Public Health England Monica Galiano, Shahjahan Miah, Angie Lackenby, Omolola Akinbami, Tiina Talts, Leena Bhaw, Richard Myers, Steven Platt, Kirstin Edwards, Jonathan Hubb, Joanna Ellis, Maria Zambon
EPI_ISL_423262 hCoV-19/England/201360277/2020 Europe / United Kingdom / England 2020-03-25
Respiratory Virus Unit, Microbiology Services Colindale, Public Health England Respiratory Virus Unit, Microbiology Services Colindale, Public Health England Monica Galiano, Shahjahan Miah, Angie Lackenby, Omolola Akinbami, Tiina Talts, Leena Bhaw, Richard Myers, Steven Platt, Kirstin Edwards, Jonathan Hubb, Joanna Ellis, Maria Zambon
EPI_ISL_423263 hCoV-19/England/201360281/2020 Europe / United Kingdom / England 2020-03-26
Respiratory Virus Unit, Microbiology Services Colindale, Public Health England Respiratory Virus Unit, Microbiology Services Colindale, Public Health England Monica Galiano, Shahjahan Miah, Angie Lackenby, Omolola Akinbami, Tiina Talts, Leena Bhaw, Richard Myers, Steven Platt, Kirstin Edwards, Jonathan Hubb, Joanna Ellis, Maria Zambon
EPI_ISL_423264 hCoV-19/England/201360317/2020 Europe / United Kingdom / England 2020-03-25
Respiratory Virus Unit, Microbiology Services Colindale, Public Health England Respiratory Virus Unit, Microbiology Services Colindale, Public Health England Monica Galiano, Shahjahan Miah, Angie Lackenby, Omolola Akinbami, Tiina Talts, Leena Bhaw, Richard Myers, Steven Platt, Kirstin Edwards, Jonathan Hubb, Joanna Ellis, Maria Zambon

EPI_ISL_423265  hCoV-19/England/201360318/2020  Europe / United Kingdom / England       2020-03-25
Respiratory Virus Unit, Microbiology Services Colindale, Public Health England  Respiratory Virus Unit, Microbiology Services Colindale, Public Health England  Monica Galiano, Shahjahan Miah, Angie Lackenby, Omolola Akinbami, Tiina Talts, Leena Bhaw, Richard Myers, Steven Platt, Kirstin Edwards, Jonathan Hubb, Joanna Ellis, Maria Zambon
EPI_ISL_423266  hCoV-19/England/201360538/2020  Europe / United Kingdom / England       2020-03-25
Respiratory Virus Unit, Microbiology Services Colindale, Public Health England  Respiratory Virus Unit, Microbiology Services Colindale, Public Health England  Monica Galiano, Shahjahan Miah, Angie Lackenby, Omolola Akinbami, Tiina Talts, Leena Bhaw, Richard Myers, Steven Platt, Kirstin Edwards, Jonathan Hubb, Joanna Ellis, Maria Zambon
EPI_ISL_423269  hCoV-19/England/20138007104/2020        Europe / United Kingdom / England       2020-03-27
Respiratory Virus Unit, Microbiology Services Colindale, Public Health England  Respiratory Virus Unit, Microbiology Services Colindale, Public Health England  Monica Galiano, Shahjahan Miah, Angie Lackenby, Omolola Akinbami, Tiina Talts, Leena Bhaw, Richard Myers, Steven Platt, Kirstin Edwards, Jonathan Hubb, Joanna Ellis, Maria Zambon
EPI_ISL_423288  hCoV-19/England/20138017804/2020        Europe / United Kingdom / England       2020-03-26
Respiratory Virus Unit, Microbiology Services Colindale, Public Health England  Respiratory Virus Unit, Microbiology Services Colindale, Public Health England  Monica Galiano, Shahjahan Miah, Angie Lackenby, Omolola Akinbami, Tiina Talts, Leena Bhaw, Richard Myers, Steven Platt, Kirstin Edwards, Jonathan Hubb, Joanna Ellis, Maria Zambon
EPI_ISL_424400  hCoV-19/Iceland/376/2020        Europe / Iceland / Reykjavik    2020-03-19      The National University Hospital of Iceland  deCODE genetics Daniel F Gudbjartsson; Agnar Helgason; Hakon Jonsson; Olafur T Magnusson; Pall Melsted; Gudmundur L Norddahl; Jona Saemundsdottir; Asgeir Sigurdsson; Patrick Sulem; Arna B Agustsdottir; Berglind Eiriksdottir; Run Fridriksdottir; Elisabet E Gardarsdottir; Gudmundur Georgsson; Olafia S Gretarsdottir; Kjartan R Gudmundsson; Thora R Gunnarsdottir; Arnaldur Gylfason; Hilma Holm; Brynjar O Jensson; Aslaug Jonasdottir; Kamilla S Josefsdottir; Thordur Kristjansson; Droplaug N Magnusdottir; Louise le Roux; Gudrun Sigmundsdottir; Gardar Sveinbjornsson; Kristin E Sveinsdottir; Maney Sveinsdottir; Emil A Thorarensen; Bjarni Thorbjornsson; Gisli Masson; Ingileif Jonsdottir; Alma Moller; Thorolfur Gudnason; Karl G Kristinsson; Unnur Thorsteinsdottir; Kari Stefansson
EPI_ISL_424401  hCoV-19/Iceland/377/2020        Europe / Iceland / Reykjavik    2020-03-19      The National University Hospital of Iceland  deCODE genetics Daniel F Gudbjartsson; Agnar Helgason; Hakon Jonsson; Olafur T Magnusson; Pall Melsted; Gudmundur L Norddahl; Jona Saemundsdottir; Asgeir Sigurdsson; Patrick Sulem; Arna B Agustsdottir; Berglind Eiriksdottir; Run Fridriksdottir; Elisabet E Gardarsdottir; Gudmundur Georgsson; Olafia S Gretarsdottir; Kjartan R Gudmundsson; Thora R Gunnarsdottir; Arnaldur Gylfason; Hilma Holm; Brynjar O Jensson; Aslaug Jonasdottir; Kamilla S Josefsdottir; Thordur Kristjansson; Droplaug N Magnusdottir; Louise le Roux; Gudrun Sigmundsdottir; Gardar Sveinbjornsson; Kristin E Sveinsdottir; Maney Sveinsdottir; Emil A Thorarensen; Bjarni Thorbjornsson; Gisli Masson; Ingileif Jonsdottir; Alma Moller; Thorolfur Gudnason; Karl G Kristinsson; Unnur Thorsteinsdottir; Kari Stefansson
EPI_ISL_424402  hCoV-19/Iceland/378/2020        Europe / Iceland / Reykjavik    2020-03-19      The National University Hospital of Iceland  deCODE genetics Daniel F Gudbjartsson; Agnar Helgason; Hakon Jonsson; Olafur T Magnusson; Pall Melsted; Gudmundur L Norddahl; Jona Saemundsdottir; Asgeir Sigurdsson; Patrick Sulem; Arna B Agustsdottir; Berglind Eiriksdottir; Run Fridriksdottir; Elisabet E Gardarsdottir; Gudmundur Georgsson; Olafia S Gretarsdottir; Kjartan R Gudmundsson; Thora R Gunnarsdottir; Arnaldur Gylfason; Hilma Holm; Brynjar O Jensson; Aslaug Jonasdottir; Kamilla S Josefsdottir; Thordur Kristjansson; Droplaug N Magnusdottir; Louise le Roux; Gudrun Sigmundsdottir; Gardar Sveinbjornsson; Kristin E Sveinsdottir; Maney Sveinsdottir; Emil A Thorarensen; Bjarni Thorbjornsson; Gisli Masson; Ingileif Jonsdottir; Alma Moller; Thorolfur Gudnason; Karl G Kristinsson; Unnur Thorsteinsdottir; Kari Stefansson
EPI_ISL_424403  hCoV-19/Iceland/379/2020        Europe / Iceland / Reykjavik    2020-03-18      The National University Hospital of Iceland  deCODE genetics Daniel F Gudbjartsson; Agnar Helgason; Hakon Jonsson; Olafur T Magnusson; Pall Melsted; Gudmundur L Norddahl; Jona Saemundsdottir; Asgeir Sigurdsson; Patrick Sulem; Arna B Agustsdottir; Berglind Eiriksdottir; Run Fridriksdottir; Elisabet E Gardarsdottir; Gudmundur Georgsson; Olafia S Gretarsdottir; Kjartan R Gudmundsson; Thora R Gunnarsdottir; Arnaldur Gylfason; Hilma Holm; Brynjar O Jensson; Aslaug Jonasdottir; Kamilla S Josefsdottir; Thordur Kristjansson; Droplaug N Magnusdottir; Louise le Roux; Gudrun Sigmundsdottir; Gardar Sveinbjornsson; Kristin E Sveinsdottir; Maney Sveinsdottir; Emil A Thorarensen; Bjarni Thorbjornsson; Gisli Masson; Ingileif Jonsdottir; Alma Moller; Thorolfur Gudnason; Karl G Kristinsson; Unnur Thorsteinsdottir; Kari Stefansson
EPI_ISL_424404  hCoV-19/Iceland/380/2020        Europe / Iceland / Reykjavik    2020-03-19      The National University Hospital of Iceland  deCODE genetics Daniel F Gudbjartsson; Agnar Helgason; Hakon Jonsson; Olafur T Magnusson; Pall Melsted; Gudmundur L Norddahl; Jona Saemundsdottir; Asgeir Sigurdsson; Patrick Sulem; Arna B Agustsdottir; Berglind Eiriksdottir; Run Fridriksdottir; Elisabet E Gardarsdottir; Gudmundur Georgsson; Olafia S Gretarsdottir; Kjartan R Gudmundsson; Thora R Gunnarsdottir; Arnaldur Gylfason; Hilma Holm; Brynjar O Jensson; Aslaug Jonasdottir; Kamilla S Josefsdottir; Thordur Kristjansson; Droplaug N Magnusdottir; Louise le Roux; Gudrun Sigmundsdottir; Gardar Sveinbjornsson; Kristin E Sveinsdottir; Maney Sveinsdottir; Emil A Thorarensen; Bjarni Thorbjornsson; Gisli Masson; Ingileif Jonsdottir; Alma Moller; Thorolfur Gudnason; Karl G Kristinsson; Unnur Thorsteinsdottir; Kari Stefansson
EPI_ISL_424405  hCoV-19/Iceland/381/2020        Europe / Iceland / Reykjavik    2020-03-19      The National University Hospital of Iceland  deCODE genetics Daniel F Gudbjartsson; Agnar Helgason; Hakon Jonsson; Olafur T Magnusson; Pall Melsted; Gudmundur L Norddahl; Jona Saemundsdottir; Asgeir Sigurdsson; Patrick Sulem; Arna B Agustsdottir; Berglind Eiriksdottir; Run Fridriksdottir; Elisabet E Gardarsdottir; Gudmundur Georgsson; Olafia S Gretarsdottir; Kjartan R Gudmundsson; Thora R Gunnarsdottir; Arnaldur Gylfason; Hilma Holm; Brynjar O

Jensson; Aslaug Jonasdottir; Kamilla S Josefsdottir; Thordur Kristjansson; Droplaug N Magnusdottir; Louise le Roux; Gudrun Sigmundsdottir; Gardar Sveinbjornsson; Kristin E Sveinsdottir; Maney Sveinsdottir; Emil A Thorarensen; Bjarni Thorbjornsson; Gisli Masson; Ingileif Jonsdottir; Alma Moller; Thorolfur Gudnason; Karl G Kristinsson; Unnur Thorsteinsdottir; Kari Stefansson
EPI_ISL_424406 hCoV-19/Iceland/382/2020 Europe / Iceland / Reykjavik 2020-03-19 The National University Hospital of Iceland deCODE genetics Daniel F Gudbjartsson; Agnar Helgason; Hakon Jonsson; Olafur T Magnusson; Pall Melsted; Gudmundur L Norddahl; Jona Saemundsdottir; Asgeir Sigurdsson; Patrick Sulem; Arna B Agustsdottir; Berglind Eiriksdottir; Run Fridriksdottir; Elisabet E Gardarsdottir; Gudmundur Georgsson; Olafia S Gretarsdottir; Kjartan R Gudmundsson; Thora R Gunnarsdottir; Arnaldur Gylfason; Hilma Holm; Brynjar O Jensson; Aslaug Jonasdottir; Kamilla S Josefsdottir; Thordur Kristjansson; Droplaug N Magnusdottir; Louise le Roux; Gudrun Sigmundsdottir; Gardar Sveinbjornsson; Kristin E Sveinsdottir; Maney Sveinsdottir; Emil A Thorarensen; Bjarni Thorbjornsson; Gisli Masson; Ingileif Jonsdottir; Alma Moller; Thorolfur Gudnason; Karl G Kristinsson; Unnur Thorsteinsdottir; Kari Stefansson
EPI_ISL_424407 hCoV-19/Iceland/383/2020 Europe / Iceland / Reykjavik 2020-03-19 The National University Hospital of Iceland deCODE genetics Daniel F Gudbjartsson; Agnar Helgason; Hakon Jonsson; Olafur T Magnusson; Pall Melsted; Gudmundur L Norddahl; Jona Saemundsdottir; Asgeir Sigurdsson; Patrick Sulem; Arna B Agustsdottir; Berglind Eiriksdottir; Run Fridriksdottir; Elisabet E Gardarsdottir; Gudmundur Georgsson; Olafia S Gretarsdottir; Kjartan R Gudmundsson; Thora R Gunnarsdottir; Arnaldur Gylfason; Hilma Holm; Brynjar O Jensson; Aslaug Jonasdottir; Kamilla S Josefsdottir; Thordur Kristjansson; Droplaug N Magnusdottir; Louise le Roux; Gudrun Sigmundsdottir; Gardar Sveinbjornsson; Kristin E Sveinsdottir; Maney Sveinsdottir; Emil A Thorarensen; Bjarni Thorbjornsson; Gisli Masson; Ingileif Jonsdottir; Alma Moller; Thorolfur Gudnason; Karl G Kristinsson; Unnur Thorsteinsdottir; Kari Stefansson
EPI_ISL_424410 hCoV-19/Iceland/386/2020 Europe / Iceland / Reykjavik 2020-03-19 The National University Hospital of Iceland deCODE genetics Daniel F Gudbjartsson; Agnar Helgason; Hakon Jonsson; Olafur T Magnusson; Pall Melsted; Gudmundur L Norddahl; Jona Saemundsdottir; Asgeir Sigurdsson; Patrick Sulem; Arna B Agustsdottir; Berglind Eiriksdottir; Run Fridriksdottir; Elisabet E Gardarsdottir; Gudmundur Georgsson; Olafia S Gretarsdottir; Kjartan R Gudmundsson; Thora R Gunnarsdottir; Arnaldur Gylfason; Hilma Holm; Brynjar O Jensson; Aslaug Jonasdottir; Kamilla S Josefsdottir; Thordur Kristjansson; Droplaug N Magnusdottir; Louise le Roux; Gudrun Sigmundsdottir; Gardar Sveinbjornsson; Kristin E Sveinsdottir; Maney Sveinsdottir; Emil A Thorarensen; Bjarni Thorbjornsson; Gisli Masson; Ingileif Jonsdottir; Alma Moller; Thorolfur Gudnason; Karl G Kristinsson; Unnur Thorsteinsdottir; Kari Stefansson
EPI_ISL_424411 hCoV-19/Iceland/387/2020 Europe / Iceland / Reykjavik 2020-03-19 The National University Hospital of Iceland deCODE genetics Daniel F Gudbjartsson; Agnar Helgason; Hakon Jonsson; Olafur T Magnusson; Pall Melsted; Gudmundur L Norddahl; Jona Saemundsdottir; Asgeir Sigurdsson; Patrick Sulem; Arna B Agustsdottir; Berglind Eiriksdottir; Run Fridriksdottir; Elisabet E Gardarsdottir; Gudmundur Georgsson; Olafia S Gretarsdottir; Kjartan R Gudmundsson; Thora R Gunnarsdottir; Arnaldur Gylfason; Hilma Holm; Brynjar O Jensson; Aslaug Jonasdottir; Kamilla S Josefsdottir; Thordur Kristjansson; Droplaug N Magnusdottir; Louise le Roux; Gudrun Sigmundsdottir; Gardar Sveinbjornsson; Kristin E Sveinsdottir; Maney Sveinsdottir; Emil A Thorarensen; Bjarni Thorbjornsson; Gisli Masson; Ingileif Jonsdottir; Alma Moller; Thorolfur Gudnason; Karl G Kristinsson; Unnur Thorsteinsdottir; Kari Stefansson
EPI_ISL_424412 hCoV-19/Iceland/388/2020 Europe / Iceland / Reykjavik 2020-03-19 The National University Hospital of Iceland deCODE genetics Daniel F Gudbjartsson; Agnar Helgason; Hakon Jonsson; Olafur T Magnusson; Pall Melsted; Gudmundur L Norddahl; Jona Saemundsdottir; Asgeir Sigurdsson; Patrick Sulem; Arna B Agustsdottir; Berglind Eiriksdottir; Run Fridriksdottir; Elisabet E Gardarsdottir; Gudmundur Georgsson; Olafia S Gretarsdottir; Kjartan R Gudmundsson; Thora R Gunnarsdottir; Arnaldur Gylfason; Hilma Holm; Brynjar O Jensson; Aslaug Jonasdottir; Kamilla S Josefsdottir; Thordur Kristjansson; Droplaug N Magnusdottir; Louise le Roux; Gudrun Sigmundsdottir; Gardar Sveinbjornsson; Kristin E Sveinsdottir; Maney Sveinsdottir; Emil A Thorarensen; Bjarni Thorbjornsson; Gisli Masson; Ingileif Jonsdottir; Alma Moller; Thorolfur Gudnason; Karl G Kristinsson; Unnur Thorsteinsdottir; Kari Stefansson
EPI_ISL_424413 hCoV-19/Iceland/389/2020 Europe / Iceland / Reykjavik 2020-03-19 The National University Hospital of Iceland deCODE genetics Daniel F Gudbjartsson; Agnar Helgason; Hakon Jonsson; Olafur T Magnusson; Pall Melsted; Gudmundur L Norddahl; Jona Saemundsdottir; Asgeir Sigurdsson; Patrick Sulem; Arna B Agustsdottir; Berglind Eiriksdottir; Run Fridriksdottir; Elisabet E Gardarsdottir; Gudmundur Georgsson; Olafia S Gretarsdottir; Kjartan R Gudmundsson; Thora R Gunnarsdottir; Arnaldur Gylfason; Hilma Holm; Brynjar O Jensson; Aslaug Jonasdottir; Kamilla S Josefsdottir; Thordur Kristjansson; Droplaug N Magnusdottir; Louise le Roux; Gudrun Sigmundsdottir; Gardar Sveinbjornsson; Kristin E Sveinsdottir; Maney Sveinsdottir; Emil A Thorarensen; Bjarni Thorbjornsson; Gisli Masson; Ingileif Jonsdottir; Alma Moller; Thorolfur Gudnason; Karl G Kristinsson; Unnur Thorsteinsdottir; Kari Stefansson
EPI_ISL_424414 hCoV-19/Iceland/390/2020 Europe / Iceland / Reykjavik 2020-03-19 The National University Hospital of Iceland deCODE genetics Daniel F Gudbjartsson; Agnar Helgason; Hakon Jonsson; Olafur T Magnusson; Pall Melsted; Gudmundur L Norddahl; Jona Saemundsdottir; Asgeir Sigurdsson; Patrick Sulem; Arna B Agustsdottir; Berglind Eiriksdottir; Run Fridriksdottir; Elisabet E Gardarsdottir; Gudmundur Georgsson; Olafia S Gretarsdottir; Kjartan R Gudmundsson; Thora R Gunnarsdottir; Arnaldur Gylfason; Hilma Holm; Brynjar O Jensson; Aslaug Jonasdottir; Kamilla S Josefsdottir; Thordur Kristjansson; Droplaug N Magnusdottir; Louise le Roux; Gudrun Sigmundsdottir; Gardar Sveinbjornsson; Kristin E Sveinsdottir; Maney Sveinsdottir; Emil A Thorarensen; Bjarni Thorbjornsson; Gisli Masson; Ingileif Jonsdottir; Alma Moller; Thorolfur Gudnason; Karl G Kristinsson; Unnur Thorsteinsdottir; Kari Stefansson
EPI_ISL_424415 hCoV-19/Iceland/391/2020 Europe / Iceland / Reykjavik 2020-03-19 The National University Hospital of Iceland deCODE genetics Daniel F Gudbjartsson; Agnar Helgason; Hakon Jonsson; Olafur T Magnusson; Pall Melsted; Gudmundur L Norddahl; Jona Saemundsdottir; Asgeir Sigurdsson; Patrick Sulem; Arna B

Agustsdottir; Berglind Eiriksdottir; Run Fridriksdottir; Elisabet E Gardarsdottir; Gudmundur Georgsson; Olafia S Gretarsdottir; Kjartan R Gudmundsson; Thora R Gunnarsdottir; Arnaldur Gylfason; Hilma Holm; Brynjar O Jensson; Aslaug Jonasdottir; Kamilla S Josefsdottir; Thordur Kristjansson; Droplaug N Magnusdottir; Louise le Roux; Gudrun Sigmundsdottir; Gardar Sveinbjornsson; Kristin E Sveinsdottir; Maney Sveinsdottir; Emil A Thorarensen; Bjarni Thorbjornsson; Gisli Masson; Ingileif Jonsdottir; Alma Moller; Thorolfur Gudnason; Karl G Kristinsson; Unnur Thorsteinsdottir; Kari Stefansson
EPI_ISL_424418 hCoV-19/Iceland/394/2020 Europe / Iceland / Reykjavik 2020-03-19 The National University Hospital of Iceland deCODE genetics Daniel F Gudbjartsson; Agnar Helgason; Hakon Jonsson; Olafur T Magnusson; Pall Melsted; Gudmundur L Norddahl; Jona Saemundsdottir; Asgeir Sigurdsson; Patrick Sulem; Arna B Agustsdottir; Berglind Eiriksdottir; Run Fridriksdottir; Elisabet E Gardarsdottir; Gudmundur Georgsson; Olafia S Gretarsdottir; Kjartan R Gudmundsson; Thora R Gunnarsdottir; Arnaldur Gylfason; Hilma Holm; Brynjar O Jensson; Aslaug Jonasdottir; Kamilla S Josefsdottir; Thordur Kristjansson; Droplaug N Magnusdottir; Louise le Roux; Gudrun Sigmundsdottir; Gardar Sveinbjornsson; Kristin E Sveinsdottir; Maney Sveinsdottir; Emil A Thorarensen; Bjarni Thorbjornsson; Gisli Masson; Ingileif Jonsdottir; Alma Moller; Thorolfur Gudnason; Karl G Kristinsson; Unnur Thorsteinsdottir; Kari Stefansson
EPI_ISL_424419 hCoV-19/Iceland/395/2020 Europe / Iceland / Reykjavik 2020-03-19 The National University Hospital of Iceland deCODE genetics Daniel F Gudbjartsson; Agnar Helgason; Hakon Jonsson; Olafur T Magnusson; Pall Melsted; Gudmundur L Norddahl; Jona Saemundsdottir; Asgeir Sigurdsson; Patrick Sulem; Arna B Agustsdottir; Berglind Eiriksdottir; Run Fridriksdottir; Elisabet E Gardarsdottir; Gudmundur Georgsson; Olafia S Gretarsdottir; Kjartan R Gudmundsson; Thora R Gunnarsdottir; Arnaldur Gylfason; Hilma Holm; Brynjar O Jensson; Aslaug Jonasdottir; Kamilla S Josefsdottir; Thordur Kristjansson; Droplaug N Magnusdottir; Louise le Roux; Gudrun Sigmundsdottir; Gardar Sveinbjornsson; Kristin E Sveinsdottir; Maney Sveinsdottir; Emil A Thorarensen; Bjarni Thorbjornsson; Gisli Masson; Ingileif Jonsdottir; Alma Moller; Thorolfur Gudnason; Karl G Kristinsson; Unnur Thorsteinsdottir; Kari Stefansson
EPI_ISL_424422 hCoV-19/Iceland/398/2020 Europe / Iceland / Reykjavik 2020-03-19 The National University Hospital of Iceland deCODE genetics Daniel F Gudbjartsson; Agnar Helgason; Hakon Jonsson; Olafur T Magnusson; Pall Melsted; Gudmundur L Norddahl; Jona Saemundsdottir; Asgeir Sigurdsson; Patrick Sulem; Arna B Agustsdottir; Berglind Eiriksdottir; Run Fridriksdottir; Elisabet E Gardarsdottir; Gudmundur Georgsson; Olafia S Gretarsdottir; Kjartan R Gudmundsson; Thora R Gunnarsdottir; Arnaldur Gylfason; Hilma Holm; Brynjar O Jensson; Aslaug Jonasdottir; Kamilla S Josefsdottir; Thordur Kristjansson; Droplaug N Magnusdottir; Louise le Roux; Gudrun Sigmundsdottir; Gardar Sveinbjornsson; Kristin E Sveinsdottir; Maney Sveinsdottir; Emil A Thorarensen; Bjarni Thorbjornsson; Gisli Masson; Ingileif Jonsdottir; Alma Moller; Thorolfur Gudnason; Karl G Kristinsson; Unnur Thorsteinsdottir; Kari Stefansson
EPI_ISL_424423 hCoV-19/Iceland/399/2020 Europe / Iceland / Reykjavik 2020-03-19 The National University Hospital of Iceland deCODE genetics Daniel F Gudbjartsson; Agnar Helgason; Hakon Jonsson; Olafur T Magnusson; Pall Melsted; Gudmundur L Norddahl; Jona Saemundsdottir; Asgeir Sigurdsson; Patrick Sulem; Arna B Agustsdottir; Berglind Eiriksdottir; Run Fridriksdottir; Elisabet E Gardarsdottir; Gudmundur Georgsson; Olafia S Gretarsdottir; Kjartan R Gudmundsson; Thora R Gunnarsdottir; Arnaldur Gylfason; Hilma Holm; Brynjar O Jensson; Aslaug Jonasdottir; Kamilla S Josefsdottir; Thordur Kristjansson; Droplaug N Magnusdottir; Louise le Roux; Gudrun Sigmundsdottir; Gardar Sveinbjornsson; Kristin E Sveinsdottir; Maney Sveinsdottir; Emil A Thorarensen; Bjarni Thorbjornsson; Gisli Masson; Ingileif Jonsdottir; Alma Moller; Thorolfur Gudnason; Karl G Kristinsson; Unnur Thorsteinsdottir; Kari Stefansson
EPI_ISL_424424 hCoV-19/Iceland/400/2020 Europe / Iceland / Reykjavik 2020-03-19 The National University Hospital of Iceland deCODE genetics Daniel F Gudbjartsson; Agnar Helgason; Hakon Jonsson; Olafur T Magnusson; Pall Melsted; Gudmundur L Norddahl; Jona Saemundsdottir; Asgeir Sigurdsson; Patrick Sulem; Arna B Agustsdottir; Berglind Eiriksdottir; Run Fridriksdottir; Elisabet E Gardarsdottir; Gudmundur Georgsson; Olafia S Gretarsdottir; Kjartan R Gudmundsson; Thora R Gunnarsdottir; Arnaldur Gylfason; Hilma Holm; Brynjar O Jensson; Aslaug Jonasdottir; Kamilla S Josefsdottir; Thordur Kristjansson; Droplaug N Magnusdottir; Louise le Roux; Gudrun Sigmundsdottir; Gardar Sveinbjornsson; Kristin E Sveinsdottir; Maney Sveinsdottir; Emil A Thorarensen; Bjarni Thorbjornsson; Gisli Masson; Ingileif Jonsdottir; Alma Moller; Thorolfur Gudnason; Karl G Kristinsson; Unnur Thorsteinsdottir; Kari Stefansson
EPI_ISL_424425 hCoV-19/Iceland/401/2020 Europe / Iceland / Reykjavik 2020-03-19 The National University Hospital of Iceland deCODE genetics Daniel F Gudbjartsson; Agnar Helgason; Hakon Jonsson; Olafur T Magnusson; Pall Melsted; Gudmundur L Norddahl; Jona Saemundsdottir; Asgeir Sigurdsson; Patrick Sulem; Arna B Agustsdottir; Berglind Eiriksdottir; Run Fridriksdottir; Elisabet E Gardarsdottir; Gudmundur Georgsson; Olafia S Gretarsdottir; Kjartan R Gudmundsson; Thora R Gunnarsdottir; Arnaldur Gylfason; Hilma Holm; Brynjar O Jensson; Aslaug Jonasdottir; Kamilla S Josefsdottir; Thordur Kristjansson; Droplaug N Magnusdottir; Louise le Roux; Gudrun Sigmundsdottir; Gardar Sveinbjornsson; Kristin E Sveinsdottir; Maney Sveinsdottir; Emil A Thorarensen; Bjarni Thorbjornsson; Gisli Masson; Ingileif Jonsdottir; Alma Moller; Thorolfur Gudnason; Karl G Kristinsson; Unnur Thorsteinsdottir; Kari Stefansson
EPI_ISL_424426 hCoV-19/Iceland/402/2020 Europe / Iceland / Reykjavik 2020-03-19 The National University Hospital of Iceland deCODE genetics Daniel F Gudbjartsson; Agnar Helgason; Hakon Jonsson; Olafur T Magnusson; Pall Melsted; Gudmundur L Norddahl; Jona Saemundsdottir; Asgeir Sigurdsson; Patrick Sulem; Arna B Agustsdottir; Berglind Eiriksdottir; Run Fridriksdottir; Elisabet E Gardarsdottir; Gudmundur Georgsson; Olafia S Gretarsdottir; Kjartan R Gudmundsson; Thora R Gunnarsdottir; Arnaldur Gylfason; Hilma Holm; Brynjar O Jensson; Aslaug Jonasdottir; Kamilla S Josefsdottir; Thordur Kristjansson; Droplaug N Magnusdottir; Louise le Roux; Gudrun Sigmundsdottir; Gardar Sveinbjornsson; Kristin E Sveinsdottir; Maney Sveinsdottir; Emil A Thorarensen; Bjarni Thorbjornsson; Gisli Masson; Ingileif Jonsdottir; Alma Moller; Thorolfur Gudnason; Karl G Kristinsson; Unnur Thorsteinsdottir; Kari Stefansson
EPI_ISL_424427 hCoV-19/Iceland/403/2020 Europe / Iceland / Reykjavik 2020-03-19 The National

University Hospital of Iceland deCODE genetics Daniel F Gudbjartsson; Agnar Helgason; Hakon Jonsson; Olafur T Magnusson; Pall Melsted; Gudmundur L Norddahl; Jona Saemundsdottir; Asgeir Sigurdsson; Patrick Sulem; Arna B Agustsdottir; Berglind Eiriksdottir; Run Fridriksdottir; Elisabet E Gardarsdottir; Gudmundur Georgsson; Olafia S Gretarsdottir; Kjartan R Gudmundsson; Thora R Gunnarsdottir; Arnaldur Gylfason; Hilma Holm; Brynjar O Jensson; Aslaug Jonasdottir; Kamilla S Josefsdottir; Thordur Kristjansson; Droplaug N Magnusdottir; Louise le Roux; Gudrun Sigmundsdottir; Gardar Sveinbjornsson; Kristin E Sveinsdottir; Maney Sveinsdottir; Emil A Thorarensen; Bjarni Thorbjornsson; Gisli Masson; Ingileif Jonsdottir; Alma Moller; Thorolfur Gudnason; Karl G Kristinsson; Unnur Thorsteinsdottir; Kari Stefansson
EPI_ISL_424429 hCoV-19/Iceland/405/2020 Europe / Iceland / Reykjavik 2020-03-19 The National University Hospital of Iceland deCODE genetics Daniel F Gudbjartsson; Agnar Helgason; Hakon Jonsson; Olafur T Magnusson; Pall Melsted; Gudmundur L Norddahl; Jona Saemundsdottir; Asgeir Sigurdsson; Patrick Sulem; Arna B Agustsdottir; Berglind Eiriksdottir; Run Fridriksdottir; Elisabet E Gardarsdottir; Gudmundur Georgsson; Olafia S Gretarsdottir; Kjartan R Gudmundsson; Thora R Gunnarsdottir; Arnaldur Gylfason; Hilma Holm; Brynjar O Jensson; Aslaug Jonasdottir; Kamilla S Josefsdottir; Thordur Kristjansson; Droplaug N Magnusdottir; Louise le Roux; Gudrun Sigmundsdottir; Gardar Sveinbjornsson; Kristin E Sveinsdottir; Maney Sveinsdottir; Emil A Thorarensen; Bjarni Thorbjornsson; Gisli Masson; Ingileif Jonsdottir; Alma Moller; Thorolfur Gudnason; Karl G Kristinsson; Unnur Thorsteinsdottir; Kari Stefansson
EPI_ISL_424433 hCoV-19/Iceland/409/2020 Europe / Iceland / Reykjavik 2020-03-19 The National University Hospital of Iceland deCODE genetics Daniel F Gudbjartsson; Agnar Helgason; Hakon Jonsson; Olafur T Magnusson; Pall Melsted; Gudmundur L Norddahl; Jona Saemundsdottir; Asgeir Sigurdsson; Patrick Sulem; Arna B Agustsdottir; Berglind Eiriksdottir; Run Fridriksdottir; Elisabet E Gardarsdottir; Gudmundur Georgsson; Olafia S Gretarsdottir; Kjartan R Gudmundsson; Thora R Gunnarsdottir; Arnaldur Gylfason; Hilma Holm; Brynjar O Jensson; Aslaug Jonasdottir; Kamilla S Josefsdottir; Thordur Kristjansson; Droplaug N Magnusdottir; Louise le Roux; Gudrun Sigmundsdottir; Gardar Sveinbjornsson; Kristin E Sveinsdottir; Maney Sveinsdottir; Emil A Thorarensen; Bjarni Thorbjornsson; Gisli Masson; Ingileif Jonsdottir; Alma Moller; Thorolfur Gudnason; Karl G Kristinsson; Unnur Thorsteinsdottir; Kari Stefansson
EPI_ISL_424434 hCoV-19/Iceland/410/2020 Europe / Iceland / Reykjavik 2020-03-19 The National University Hospital of Iceland deCODE genetics Daniel F Gudbjartsson; Agnar Helgason; Hakon Jonsson; Olafur T Magnusson; Pall Melsted; Gudmundur L Norddahl; Jona Saemundsdottir; Asgeir Sigurdsson; Patrick Sulem; Arna B Agustsdottir; Berglind Eiriksdottir; Run Fridriksdottir; Elisabet E Gardarsdottir; Gudmundur Georgsson; Olafia S Gretarsdottir; Kjartan R Gudmundsson; Thora R Gunnarsdottir; Arnaldur Gylfason; Hilma Holm; Brynjar O Jensson; Aslaug Jonasdottir; Kamilla S Josefsdottir; Thordur Kristjansson; Droplaug N Magnusdottir; Louise le Roux; Gudrun Sigmundsdottir; Gardar Sveinbjornsson; Kristin E Sveinsdottir; Maney Sveinsdottir; Emil A Thorarensen; Bjarni Thorbjornsson; Gisli Masson; Ingileif Jonsdottir; Alma Moller; Thorolfur Gudnason; Karl G Kristinsson; Unnur Thorsteinsdottir; Kari Stefansson
EPI_ISL_424435 hCoV-19/Iceland/411/2020 Europe / Iceland / Reykjavik 2020-03-19 The National University Hospital of Iceland deCODE genetics Daniel F Gudbjartsson; Agnar Helgason; Hakon Jonsson; Olafur T Magnusson; Pall Melsted; Gudmundur L Norddahl; Jona Saemundsdottir; Asgeir Sigurdsson; Patrick Sulem; Arna B Agustsdottir; Berglind Eiriksdottir; Run Fridriksdottir; Elisabet E Gardarsdottir; Gudmundur Georgsson; Olafia S Gretarsdottir; Kjartan R Gudmundsson; Thora R Gunnarsdottir; Arnaldur Gylfason; Hilma Holm; Brynjar O Jensson; Aslaug Jonasdottir; Kamilla S Josefsdottir; Thordur Kristjansson; Droplaug N Magnusdottir; Louise le Roux; Gudrun Sigmundsdottir; Gardar Sveinbjornsson; Kristin E Sveinsdottir; Maney Sveinsdottir; Emil A Thorarensen; Bjarni Thorbjornsson; Gisli Masson; Ingileif Jonsdottir; Alma Moller; Thorolfur Gudnason; Karl G Kristinsson; Unnur Thorsteinsdottir; Kari Stefansson
EPI_ISL_424436 hCoV-19/Iceland/412/2020 Europe / Iceland / Reykjavik 2020-03-19 The National University Hospital of Iceland deCODE genetics Daniel F Gudbjartsson; Agnar Helgason; Hakon Jonsson; Olafur T Magnusson; Pall Melsted; Gudmundur L Norddahl; Jona Saemundsdottir; Asgeir Sigurdsson; Patrick Sulem; Arna B Agustsdottir; Berglind Eiriksdottir; Run Fridriksdottir; Elisabet E Gardarsdottir; Gudmundur Georgsson; Olafia S Gretarsdottir; Kjartan R Gudmundsson; Thora R Gunnarsdottir; Arnaldur Gylfason; Hilma Holm; Brynjar O Jensson; Aslaug Jonasdottir; Kamilla S Josefsdottir; Thordur Kristjansson; Droplaug N Magnusdottir; Louise le Roux; Gudrun Sigmundsdottir; Gardar Sveinbjornsson; Kristin E Sveinsdottir; Maney Sveinsdottir; Emil A Thorarensen; Bjarni Thorbjornsson; Gisli Masson; Ingileif Jonsdottir; Alma Moller; Thorolfur Gudnason; Karl G Kristinsson; Unnur Thorsteinsdottir; Kari Stefansson
EPI_ISL_424437 hCoV-19/Iceland/413/2020 Europe / Iceland / Reykjavik 2020-03-19 The National University Hospital of Iceland deCODE genetics Daniel F Gudbjartsson; Agnar Helgason; Hakon Jonsson; Olafur T Magnusson; Pall Melsted; Gudmundur L Norddahl; Jona Saemundsdottir; Asgeir Sigurdsson; Patrick Sulem; Arna B Agustsdottir; Berglind Eiriksdottir; Run Fridriksdottir; Elisabet E Gardarsdottir; Gudmundur Georgsson; Olafia S Gretarsdottir; Kjartan R Gudmundsson; Thora R Gunnarsdottir; Arnaldur Gylfason; Hilma Holm; Brynjar O Jensson; Aslaug Jonasdottir; Kamilla S Josefsdottir; Thordur Kristjansson; Droplaug N Magnusdottir; Louise le Roux; Gudrun Sigmundsdottir; Gardar Sveinbjornsson; Kristin E Sveinsdottir; Maney Sveinsdottir; Emil A Thorarensen; Bjarni Thorbjornsson; Gisli Masson; Ingileif Jonsdottir; Alma Moller; Thorolfur Gudnason; Karl G Kristinsson; Unnur Thorsteinsdottir; Kari Stefansson
EPI_ISL_424438 hCoV-19/Iceland/414/2020 Europe / Iceland / Reykjavik 2020-03-19 The National University Hospital of Iceland deCODE genetics Daniel F Gudbjartsson; Agnar Helgason; Hakon Jonsson; Olafur T Magnusson; Pall Melsted; Gudmundur L Norddahl; Jona Saemundsdottir; Asgeir Sigurdsson; Patrick Sulem; Arna B Agustsdottir; Berglind Eiriksdottir; Run Fridriksdottir; Elisabet E Gardarsdottir; Gudmundur Georgsson; Olafia S Gretarsdottir; Kjartan R Gudmundsson; Thora R Gunnarsdottir; Arnaldur Gylfason; Hilma Holm; Brynjar O Jensson; Aslaug Jonasdottir; Kamilla S Josefsdottir; Thordur Kristjansson; Droplaug N Magnusdottir; Louise le Roux; Gudrun Sigmundsdottir; Gardar Sveinbjornsson; Kristin E Sveinsdottir; Maney Sveinsdottir; Emil A Thorarensen; Bjarni Thorbjornsson; Gisli Masson; Ingileif Jonsdottir; Alma Moller; Thorolfur Gudnason; Karl G

Kristinsson; Unnur Thorsteinsdottir; Kari Stefansson
EPI_ISL_424439 hCoV-19/Iceland/415/2020 Europe / Iceland / Reykjavik 2020-03-19 The National University Hospital of Iceland deCODE genetics Daniel F Gudbjartsson; Agnar Helgason; Hakon Jonsson; Olafur T Magnusson; Pall Melsted; Gudmundur L Norddahl; Jona Saemundsdottir; Asgeir Sigurdsson; Patrick Sulem; Arna B Agustsdottir; Berglind Eiriksdottir; Run Fridriksdottir; Elisabet E Gardarsdottir; Gudmundur Georgsson; Olafia S Gretarsdottir; Kjartan R Gudmundsson; Thora R Gunnarsdottir; Arnaldur Gylfason; Hilma Holm; Brynjar O Jensson; Aslaug Jonasdottir; Kamilla S Josefsdottir; Thordur Kristjansson; Droplaug N Magnusdottir; Louise le Roux; Gudrun Sigmundsdottir; Gardar Sveinbjornsson; Kristin E Sveinsdottir; Maney Sveinsdottir; Emil A Thorarensen; Bjarni Thorbjornsson; Gisli Masson; Ingileif Jonsdottir; Alma Moller; Thorolfur Gudnason; Karl G Kristinsson; Unnur Thorsteinsdottir; Kari Stefansson
EPI_ISL_424440 hCoV-19/Iceland/416/2020 Europe / Iceland / Reykjavik 2020-03-19 The National University Hospital of Iceland deCODE genetics Daniel F Gudbjartsson; Agnar Helgason; Hakon Jonsson; Olafur T Magnusson; Pall Melsted; Gudmundur L Norddahl; Jona Saemundsdottir; Asgeir Sigurdsson; Patrick Sulem; Arna B Agustsdottir; Berglind Eiriksdottir; Run Fridriksdottir; Elisabet E Gardarsdottir; Gudmundur Georgsson; Olafia S Gretarsdottir; Kjartan R Gudmundsson; Thora R Gunnarsdottir; Arnaldur Gylfason; Hilma Holm; Brynjar O Jensson; Aslaug Jonasdottir; Kamilla S Josefsdottir; Thordur Kristjansson; Droplaug N Magnusdottir; Louise le Roux; Gudrun Sigmundsdottir; Gardar Sveinbjornsson; Kristin E Sveinsdottir; Maney Sveinsdottir; Emil A Thorarensen; Bjarni Thorbjornsson; Gisli Masson; Ingileif Jonsdottir; Alma Moller; Thorolfur Gudnason; Karl G Kristinsson; Unnur Thorsteinsdottir; Kari Stefansson
EPI_ISL_424441 hCoV-19/Iceland/417/2020 Europe / Iceland / Reykjavik 2020-03-19 The National University Hospital of Iceland deCODE genetics Daniel F Gudbjartsson; Agnar Helgason; Hakon Jonsson; Olafur T Magnusson; Pall Melsted; Gudmundur L Norddahl; Jona Saemundsdottir; Asgeir Sigurdsson; Patrick Sulem; Arna B Agustsdottir; Berglind Eiriksdottir; Run Fridriksdottir; Elisabet E Gardarsdottir; Gudmundur Georgsson; Olafia S Gretarsdottir; Kjartan R Gudmundsson; Thora R Gunnarsdottir; Arnaldur Gylfason; Hilma Holm; Brynjar O Jensson; Aslaug Jonasdottir; Kamilla S Josefsdottir; Thordur Kristjansson; Droplaug N Magnusdottir; Louise le Roux; Gudrun Sigmundsdottir; Gardar Sveinbjornsson; Kristin E Sveinsdottir; Maney Sveinsdottir; Emil A Thorarensen; Bjarni Thorbjornsson; Gisli Masson; Ingileif Jonsdottir; Alma Moller; Thorolfur Gudnason; Karl G Kristinsson; Unnur Thorsteinsdottir; Kari Stefansson
EPI_ISL_424442 hCoV-19/Iceland/418/2020 Europe / Iceland / Reykjavik 2020-03-19 The National University Hospital of Iceland deCODE genetics Daniel F Gudbjartsson; Agnar Helgason; Hakon Jonsson; Olafur T Magnusson; Pall Melsted; Gudmundur L Norddahl; Jona Saemundsdottir; Asgeir Sigurdsson; Patrick Sulem; Arna B Agustsdottir; Berglind Eiriksdottir; Run Fridriksdottir; Elisabet E Gardarsdottir; Gudmundur Georgsson; Olafia S Gretarsdottir; Kjartan R Gudmundsson; Thora R Gunnarsdottir; Arnaldur Gylfason; Hilma Holm; Brynjar O Jensson; Aslaug Jonasdottir; Kamilla S Josefsdottir; Thordur Kristjansson; Droplaug N Magnusdottir; Louise le Roux; Gudrun Sigmundsdottir; Gardar Sveinbjornsson; Kristin E Sveinsdottir; Maney Sveinsdottir; Emil A Thorarensen; Bjarni Thorbjornsson; Gisli Masson; Ingileif Jonsdottir; Alma Moller; Thorolfur Gudnason; Karl G Kristinsson; Unnur Thorsteinsdottir; Kari Stefansson
EPI_ISL_424443 hCoV-19/Iceland/419/2020 Europe / Iceland / Reykjavik 2020-03-19 The National University Hospital of Iceland deCODE genetics Daniel F Gudbjartsson; Agnar Helgason; Hakon Jonsson; Olafur T Magnusson; Pall Melsted; Gudmundur L Norddahl; Jona Saemundsdottir; Asgeir Sigurdsson; Patrick Sulem; Arna B Agustsdottir; Berglind Eiriksdottir; Run Fridriksdottir; Elisabet E Gardarsdottir; Gudmundur Georgsson; Olafia S Gretarsdottir; Kjartan R Gudmundsson; Thora R Gunnarsdottir; Arnaldur Gylfason; Hilma Holm; Brynjar O Jensson; Aslaug Jonasdottir; Kamilla S Josefsdottir; Thordur Kristjansson; Droplaug N Magnusdottir; Louise le Roux; Gudrun Sigmundsdottir; Gardar Sveinbjornsson; Kristin E Sveinsdottir; Maney Sveinsdottir; Emil A Thorarensen; Bjarni Thorbjornsson; Gisli Masson; Ingileif Jonsdottir; Alma Moller; Thorolfur Gudnason; Karl G Kristinsson; Unnur Thorsteinsdottir; Kari Stefansson
EPI_ISL_424445 hCoV-19/Iceland/421/2020 Europe / Iceland / Reykjavik 2020-03-19 The National University Hospital of Iceland deCODE genetics Daniel F Gudbjartsson; Agnar Helgason; Hakon Jonsson; Olafur T Magnusson; Pall Melsted; Gudmundur L Norddahl; Jona Saemundsdottir; Asgeir Sigurdsson; Patrick Sulem; Arna B Agustsdottir; Berglind Eiriksdottir; Run Fridriksdottir; Elisabet E Gardarsdottir; Gudmundur Georgsson; Olafia S Gretarsdottir; Kjartan R Gudmundsson; Thora R Gunnarsdottir; Arnaldur Gylfason; Hilma Holm; Brynjar O Jensson; Aslaug Jonasdottir; Kamilla S Josefsdottir; Thordur Kristjansson; Droplaug N Magnusdottir; Louise le Roux; Gudrun Sigmundsdottir; Gardar Sveinbjornsson; Kristin E Sveinsdottir; Maney Sveinsdottir; Emil A Thorarensen; Bjarni Thorbjornsson; Gisli Masson; Ingileif Jonsdottir; Alma Moller; Thorolfur Gudnason; Karl G Kristinsson; Unnur Thorsteinsdottir; Kari Stefansson
EPI_ISL_424446 hCoV-19/Iceland/422/2020 Europe / Iceland / Reykjavik 2020-03-19 The National University Hospital of Iceland deCODE genetics Daniel F Gudbjartsson; Agnar Helgason; Hakon Jonsson; Olafur T Magnusson; Pall Melsted; Gudmundur L Norddahl; Jona Saemundsdottir; Asgeir Sigurdsson; Patrick Sulem; Arna B Agustsdottir; Berglind Eiriksdottir; Run Fridriksdottir; Elisabet E Gardarsdottir; Gudmundur Georgsson; Olafia S Gretarsdottir; Kjartan R Gudmundsson; Thora R Gunnarsdottir; Arnaldur Gylfason; Hilma Holm; Brynjar O Jensson; Aslaug Jonasdottir; Kamilla S Josefsdottir; Thordur Kristjansson; Droplaug N Magnusdottir; Louise le Roux; Gudrun Sigmundsdottir; Gardar Sveinbjornsson; Kristin E Sveinsdottir; Maney Sveinsdottir; Emil A Thorarensen; Bjarni Thorbjornsson; Gisli Masson; Ingileif Jonsdottir; Alma Moller; Thorolfur Gudnason; Karl G Kristinsson; Unnur Thorsteinsdottir; Kari Stefansson
EPI_ISL_424447 hCoV-19/Iceland/423/2020 Europe / Iceland / Reykjavik 2020-03-19 The National University Hospital of Iceland deCODE genetics Daniel F Gudbjartsson; Agnar Helgason; Hakon Jonsson; Olafur T Magnusson; Pall Melsted; Gudmundur L Norddahl; Jona Saemundsdottir; Asgeir Sigurdsson; Patrick Sulem; Arna B Agustsdottir; Berglind Eiriksdottir; Run Fridriksdottir; Elisabet E Gardarsdottir; Gudmundur Georgsson; Olafia S Gretarsdottir; Kjartan R Gudmundsson; Thora R Gunnarsdottir; Arnaldur Gylfason; Hilma Holm; Brynjar O Jensson; Aslaug Jonasdottir; Kamilla S Josefsdottir; Thordur Kristjansson; Droplaug N Magnusdottir; Louise le

Roux; Gudrun Sigmundsdottir; Gardar Sveinbjornsson; Kristin E Sveinsdottir; Maney Sveinsdottir; Emil A Thorarensen; Bjarni Thorbjornsson; Gisli Masson; Ingileif Jonsdottir; Alma Moller; Thorolfur Gudnason; Karl G Kristinsson; Unnur Thorsteinsdottir; Kari Stefansson
EPI_ISL_424448 hCoV-19/Iceland/424/2020 Europe / Iceland / Reykjavik 2020-03-20 The National University Hospital of Iceland deCODE genetics Daniel F Gudbjartsson; Agnar Helgason; Hakon Jonsson; Olafur T Magnusson; Pall Melsted; Gudmundur L Norddahl; Jona Saemundsdottir; Asgeir Sigurdsson; Patrick Sulem; Arna B Agustsdottir; Berglind Eiriksdottir; Run Fridriksdottir; Elisabet E Gardarsdottir; Gudmundur Georgsson; Olafia S Gretarsdottir; Kjartan R Gudmundsson; Thora R Gunnarsdottir; Arnaldur Gylfason; Hilma Holm; Brynjar O Jensson; Aslaug Jonasdottir; Kamilla S Josefsdottir; Thordur Kristjansson; Droplaug N Magnusdottir; Louise le Roux; Gudrun Sigmundsdottir; Gardar Sveinbjornsson; Kristin E Sveinsdottir; Maney Sveinsdottir; Emil A Thorarensen; Bjarni Thorbjornsson; Gisli Masson; Ingileif Jonsdottir; Alma Moller; Thorolfur Gudnason; Karl G Kristinsson; Unnur Thorsteinsdottir; Kari Stefansson
EPI_ISL_424449 hCoV-19/Iceland/425/2020 Europe / Iceland / Reykjavik 2020-03-20 The National University Hospital of Iceland deCODE genetics Daniel F Gudbjartsson; Agnar Helgason; Hakon Jonsson; Olafur T Magnusson; Pall Melsted; Gudmundur L Norddahl; Jona Saemundsdottir; Asgeir Sigurdsson; Patrick Sulem; Arna B Agustsdottir; Berglind Eiriksdottir; Run Fridriksdottir; Elisabet E Gardarsdottir; Gudmundur Georgsson; Olafia S Gretarsdottir; Kjartan R Gudmundsson; Thora R Gunnarsdottir; Arnaldur Gylfason; Hilma Holm; Brynjar O Jensson; Aslaug Jonasdottir; Kamilla S Josefsdottir; Thordur Kristjansson; Droplaug N Magnusdottir; Louise le Roux; Gudrun Sigmundsdottir; Gardar Sveinbjornsson; Kristin E Sveinsdottir; Maney Sveinsdottir; Emil A Thorarensen; Bjarni Thorbjornsson; Gisli Masson; Ingileif Jonsdottir; Alma Moller; Thorolfur Gudnason; Karl G Kristinsson; Unnur Thorsteinsdottir; Kari Stefansson
EPI_ISL_424450 hCoV-19/Iceland/426/2020 Europe / Iceland / Reykjavik 2020-03-20 The National University Hospital of Iceland deCODE genetics Daniel F Gudbjartsson; Agnar Helgason; Hakon Jonsson; Olafur T Magnusson; Pall Melsted; Gudmundur L Norddahl; Jona Saemundsdottir; Asgeir Sigurdsson; Patrick Sulem; Arna B Agustsdottir; Berglind Eiriksdottir; Run Fridriksdottir; Elisabet E Gardarsdottir; Gudmundur Georgsson; Olafia S Gretarsdottir; Kjartan R Gudmundsson; Thora R Gunnarsdottir; Arnaldur Gylfason; Hilma Holm; Brynjar O Jensson; Aslaug Jonasdottir; Kamilla S Josefsdottir; Thordur Kristjansson; Droplaug N Magnusdottir; Louise le Roux; Gudrun Sigmundsdottir; Gardar Sveinbjornsson; Kristin E Sveinsdottir; Maney Sveinsdottir; Emil A Thorarensen; Bjarni Thorbjornsson; Gisli Masson; Ingileif Jonsdottir; Alma Moller; Thorolfur Gudnason; Karl G Kristinsson; Unnur Thorsteinsdottir; Kari Stefansson
EPI_ISL_424451 hCoV-19/Iceland/427/2020 Europe / Iceland / Reykjavik 2020-03-20 The National University Hospital of Iceland deCODE genetics Daniel F Gudbjartsson; Agnar Helgason; Hakon Jonsson; Olafur T Magnusson; Pall Melsted; Gudmundur L Norddahl; Jona Saemundsdottir; Asgeir Sigurdsson; Patrick Sulem; Arna B Agustsdottir; Berglind Eiriksdottir; Run Fridriksdottir; Elisabet E Gardarsdottir; Gudmundur Georgsson; Olafia S Gretarsdottir; Kjartan R Gudmundsson; Thora R Gunnarsdottir; Arnaldur Gylfason; Hilma Holm; Brynjar O Jensson; Aslaug Jonasdottir; Kamilla S Josefsdottir; Thordur Kristjansson; Droplaug N Magnusdottir; Louise le Roux; Gudrun Sigmundsdottir; Gardar Sveinbjornsson; Kristin E Sveinsdottir; Maney Sveinsdottir; Emil A Thorarensen; Bjarni Thorbjornsson; Gisli Masson; Ingileif Jonsdottir; Alma Moller; Thorolfur Gudnason; Karl G Kristinsson; Unnur Thorsteinsdottir; Kari Stefansson
EPI_ISL_424452 hCoV-19/Iceland/428/2020 Europe / Iceland / Reykjavik 2020-03-20 The National University Hospital of Iceland deCODE genetics Daniel F Gudbjartsson; Agnar Helgason; Hakon Jonsson; Olafur T Magnusson; Pall Melsted; Gudmundur L Norddahl; Jona Saemundsdottir; Asgeir Sigurdsson; Patrick Sulem; Arna B Agustsdottir; Berglind Eiriksdottir; Run Fridriksdottir; Elisabet E Gardarsdottir; Gudmundur Georgsson; Olafia S Gretarsdottir; Kjartan R Gudmundsson; Thora R Gunnarsdottir; Arnaldur Gylfason; Hilma Holm; Brynjar O Jensson; Aslaug Jonasdottir; Kamilla S Josefsdottir; Thordur Kristjansson; Droplaug N Magnusdottir; Louise le Roux; Gudrun Sigmundsdottir; Gardar Sveinbjornsson; Kristin E Sveinsdottir; Maney Sveinsdottir; Emil A Thorarensen; Bjarni Thorbjornsson; Gisli Masson; Ingileif Jonsdottir; Alma Moller; Thorolfur Gudnason; Karl G Kristinsson; Unnur Thorsteinsdottir; Kari Stefansson
EPI_ISL_424454 hCoV-19/Iceland/430/2020 Europe / Iceland / Reykjavik 2020-03-20 The National University Hospital of Iceland deCODE genetics Daniel F Gudbjartsson; Agnar Helgason; Hakon Jonsson; Olafur T Magnusson; Pall Melsted; Gudmundur L Norddahl; Jona Saemundsdottir; Asgeir Sigurdsson; Patrick Sulem; Arna B Agustsdottir; Berglind Eiriksdottir; Run Fridriksdottir; Elisabet E Gardarsdottir; Gudmundur Georgsson; Olafia S Gretarsdottir; Kjartan R Gudmundsson; Thora R Gunnarsdottir; Arnaldur Gylfason; Hilma Holm; Brynjar O Jensson; Aslaug Jonasdottir; Kamilla S Josefsdottir; Thordur Kristjansson; Droplaug N Magnusdottir; Louise le Roux; Gudrun Sigmundsdottir; Gardar Sveinbjornsson; Kristin E Sveinsdottir; Maney Sveinsdottir; Emil A Thorarensen; Bjarni Thorbjornsson; Gisli Masson; Ingileif Jonsdottir; Alma Moller; Thorolfur Gudnason; Karl G Kristinsson; Unnur Thorsteinsdottir; Kari Stefansson
EPI_ISL_424455 hCoV-19/Iceland/431/2020 Europe / Iceland / Reykjavik 2020-03-20 The National University Hospital of Iceland deCODE genetics Daniel F Gudbjartsson; Agnar Helgason; Hakon Jonsson; Olafur T Magnusson; Pall Melsted; Gudmundur L Norddahl; Jona Saemundsdottir; Asgeir Sigurdsson; Patrick Sulem; Arna B Agustsdottir; Berglind Eiriksdottir; Run Fridriksdottir; Elisabet E Gardarsdottir; Gudmundur Georgsson; Olafia S Gretarsdottir; Kjartan R Gudmundsson; Thora R Gunnarsdottir; Arnaldur Gylfason; Hilma Holm; Brynjar O Jensson; Aslaug Jonasdottir; Kamilla S Josefsdottir; Thordur Kristjansson; Droplaug N Magnusdottir; Louise le Roux; Gudrun Sigmundsdottir; Gardar Sveinbjornsson; Kristin E Sveinsdottir; Maney Sveinsdottir; Emil A Thorarensen; Bjarni Thorbjornsson; Gisli Masson; Ingileif Jonsdottir; Alma Moller; Thorolfur Gudnason; Karl G Kristinsson; Unnur Thorsteinsdottir; Kari Stefansson
EPI_ISL_424456 hCoV-19/Iceland/432/2020 Europe / Iceland / Reykjavik 2020-03-20 The National University Hospital of Iceland deCODE genetics Daniel F Gudbjartsson; Agnar Helgason; Hakon Jonsson; Olafur T Magnusson; Pall Melsted; Gudmundur L Norddahl; Jona Saemundsdottir; Asgeir Sigurdsson; Patrick Sulem; Arna B Agustsdottir; Berglind Eiriksdottir; Run Fridriksdottir; Elisabet E Gardarsdottir; Gudmundur Georgsson; Olafia

S Gretarsdottir; Kjartan R Gudmundsson; Thora R Gunnarsdottir; Arnaldur Gylfason; Hilma Holm; Brynjar O Jensson; Aslaug Jonasdottir; Kamilla S Josefsdottir; Thordur Kristjansson; Droplaug N Magnusdottir; Louise le Roux; Gudrun Sigmundsdottir; Gardar Sveinbjornsson; Kristin E Sveinsdottir; Maney Sveinsdottir; Emil A Thorarensen; Bjarni Thorbjornsson; Gisli Masson; Ingileif Jonsdottir; Alma Moller; Thorolfur Gudnason; Karl G Kristinsson; Unnur Thorsteinsdottir; Kari Stefansson

EPI_ISL_424457 hCoV-19/Iceland/433/2020 Europe / Iceland / Reykjavik 2020-03-20 The National University Hospital of Iceland deCODE genetics Daniel F Gudbjartsson; Agnar Helgason; Hakon Jonsson; Olafur T Magnusson; Pall Melsted; Gudmundur L Norddahl; Jona Saemundsdottir; Asgeir Sigurdsson; Patrick Sulem; Arna B Agustsdottir; Berglind Eiriksdottir; Run Fridriksdottir; Elisabet E Gardarsdottir; Gudmundur Georgsson; Olafia S Gretarsdottir; Kjartan R Gudmundsson; Thora R Gunnarsdottir; Arnaldur Gylfason; Hilma Holm; Brynjar O Jensson; Aslaug Jonasdottir; Kamilla S Josefsdottir; Thordur Kristjansson; Droplaug N Magnusdottir; Louise le Roux; Gudrun Sigmundsdottir; Gardar Sveinbjornsson; Kristin E Sveinsdottir; Maney Sveinsdottir; Emil A Thorarensen; Bjarni Thorbjornsson; Gisli Masson; Ingileif Jonsdottir; Alma Moller; Thorolfur Gudnason; Karl G Kristinsson; Unnur Thorsteinsdottir; Kari Stefansson

EPI_ISL_424459 hCoV-19/Iceland/435/2020 Europe / Iceland / Reykjavik 2020-03-20 The National University Hospital of Iceland deCODE genetics Daniel F Gudbjartsson; Agnar Helgason; Hakon Jonsson; Olafur T Magnusson; Pall Melsted; Gudmundur L Norddahl; Jona Saemundsdottir; Asgeir Sigurdsson; Patrick Sulem; Arna B Agustsdottir; Berglind Eiriksdottir; Run Fridriksdottir; Elisabet E Gardarsdottir; Gudmundur Georgsson; Olafia S Gretarsdottir; Kjartan R Gudmundsson; Thora R Gunnarsdottir; Arnaldur Gylfason; Hilma Holm; Brynjar O Jensson; Aslaug Jonasdottir; Kamilla S Josefsdottir; Thordur Kristjansson; Droplaug N Magnusdottir; Louise le Roux; Gudrun Sigmundsdottir; Gardar Sveinbjornsson; Kristin E Sveinsdottir; Maney Sveinsdottir; Emil A Thorarensen; Bjarni Thorbjornsson; Gisli Masson; Ingileif Jonsdottir; Alma Moller; Thorolfur Gudnason; Karl G Kristinsson; Unnur Thorsteinsdottir; Kari Stefansson

EPI_ISL_424460 hCoV-19/Iceland/436/2020 Europe / Iceland / Reykjavik 2020-03-20 The National University Hospital of Iceland deCODE genetics Daniel F Gudbjartsson; Agnar Helgason; Hakon Jonsson; Olafur T Magnusson; Pall Melsted; Gudmundur L Norddahl; Jona Saemundsdottir; Asgeir Sigurdsson; Patrick Sulem; Arna B Agustsdottir; Berglind Eiriksdottir; Run Fridriksdottir; Elisabet E Gardarsdottir; Gudmundur Georgsson; Olafia S Gretarsdottir; Kjartan R Gudmundsson; Thora R Gunnarsdottir; Arnaldur Gylfason; Hilma Holm; Brynjar O Jensson; Aslaug Jonasdottir; Kamilla S Josefsdottir; Thordur Kristjansson; Droplaug N Magnusdottir; Louise le Roux; Gudrun Sigmundsdottir; Gardar Sveinbjornsson; Kristin E Sveinsdottir; Maney Sveinsdottir; Emil A Thorarensen; Bjarni Thorbjornsson; Gisli Masson; Ingileif Jonsdottir; Alma Moller; Thorolfur Gudnason; Karl G Kristinsson; Unnur Thorsteinsdottir; Kari Stefansson

EPI_ISL_424461 hCoV-19/Iceland/437/2020 Europe / Iceland / Reykjavik 2020-03-20 The National University Hospital of Iceland deCODE genetics Daniel F Gudbjartsson; Agnar Helgason; Hakon Jonsson; Olafur T Magnusson; Pall Melsted; Gudmundur L Norddahl; Jona Saemundsdottir; Asgeir Sigurdsson; Patrick Sulem; Arna B Agustsdottir; Berglind Eiriksdottir; Run Fridriksdottir; Elisabet E Gardarsdottir; Gudmundur Georgsson; Olafia S Gretarsdottir; Kjartan R Gudmundsson; Thora R Gunnarsdottir; Arnaldur Gylfason; Hilma Holm; Brynjar O Jensson; Aslaug Jonasdottir; Kamilla S Josefsdottir; Thordur Kristjansson; Droplaug N Magnusdottir; Louise le Roux; Gudrun Sigmundsdottir; Gardar Sveinbjornsson; Kristin E Sveinsdottir; Maney Sveinsdottir; Emil A Thorarensen; Bjarni Thorbjornsson; Gisli Masson; Ingileif Jonsdottir; Alma Moller; Thorolfur Gudnason; Karl G Kristinsson; Unnur Thorsteinsdottir; Kari Stefansson

EPI_ISL_424462 hCoV-19/Iceland/438/2020 Europe / Iceland / Reykjavik 2020-03-20 The National University Hospital of Iceland deCODE genetics Daniel F Gudbjartsson; Agnar Helgason; Hakon Jonsson; Olafur T Magnusson; Pall Melsted; Gudmundur L Norddahl; Jona Saemundsdottir; Asgeir Sigurdsson; Patrick Sulem; Arna B Agustsdottir; Berglind Eiriksdottir; Run Fridriksdottir; Elisabet E Gardarsdottir; Gudmundur Georgsson; Olafia S Gretarsdottir; Kjartan R Gudmundsson; Thora R Gunnarsdottir; Arnaldur Gylfason; Hilma Holm; Brynjar O Jensson; Aslaug Jonasdottir; Kamilla S Josefsdottir; Thordur Kristjansson; Droplaug N Magnusdottir; Louise le Roux; Gudrun Sigmundsdottir; Gardar Sveinbjornsson; Kristin E Sveinsdottir; Maney Sveinsdottir; Emil A Thorarensen; Bjarni Thorbjornsson; Gisli Masson; Ingileif Jonsdottir; Alma Moller; Thorolfur Gudnason; Karl G Kristinsson; Unnur Thorsteinsdottir; Kari Stefansson

EPI_ISL_424463 hCoV-19/Iceland/439/2020 Europe / Iceland / Reykjavik 2020-03-20 The National University Hospital of Iceland deCODE genetics Daniel F Gudbjartsson; Agnar Helgason; Hakon Jonsson; Olafur T Magnusson; Pall Melsted; Gudmundur L Norddahl; Jona Saemundsdottir; Asgeir Sigurdsson; Patrick Sulem; Arna B Agustsdottir; Berglind Eiriksdottir; Run Fridriksdottir; Elisabet E Gardarsdottir; Gudmundur Georgsson; Olafia S Gretarsdottir; Kjartan R Gudmundsson; Thora R Gunnarsdottir; Arnaldur Gylfason; Hilma Holm; Brynjar O Jensson; Aslaug Jonasdottir; Kamilla S Josefsdottir; Thordur Kristjansson; Droplaug N Magnusdottir; Louise le Roux; Gudrun Sigmundsdottir; Gardar Sveinbjornsson; Kristin E Sveinsdottir; Maney Sveinsdottir; Emil A Thorarensen; Bjarni Thorbjornsson; Gisli Masson; Ingileif Jonsdottir; Alma Moller; Thorolfur Gudnason; Karl G Kristinsson; Unnur Thorsteinsdottir; Kari Stefansson

EPI_ISL_424465 hCoV-19/Iceland/441/2020 Europe / Iceland / Reykjavik 2020-03-20 The National University Hospital of Iceland deCODE genetics Daniel F Gudbjartsson; Agnar Helgason; Hakon Jonsson; Olafur T Magnusson; Pall Melsted; Gudmundur L Norddahl; Jona Saemundsdottir; Asgeir Sigurdsson; Patrick Sulem; Arna B Agustsdottir; Berglind Eiriksdottir; Run Fridriksdottir; Elisabet E Gardarsdottir; Gudmundur Georgsson; Olafia S Gretarsdottir; Kjartan R Gudmundsson; Thora R Gunnarsdottir; Arnaldur Gylfason; Hilma Holm; Brynjar O Jensson; Aslaug Jonasdottir; Kamilla S Josefsdottir; Thordur Kristjansson; Droplaug N Magnusdottir; Louise le Roux; Gudrun Sigmundsdottir; Gardar Sveinbjornsson; Kristin E Sveinsdottir; Maney Sveinsdottir; Emil A Thorarensen; Bjarni Thorbjornsson; Gisli Masson; Ingileif Jonsdottir; Alma Moller; Thorolfur Gudnason; Karl G Kristinsson; Unnur Thorsteinsdottir; Kari Stefansson

EPI_ISL_424466 hCoV-19/Iceland/442/2020 Europe / Iceland / Reykjavik 2020-03-20 The National University Hospital of Iceland deCODE genetics Daniel F Gudbjartsson; Agnar Helgason; Hakon Jonsson; Olafur T

Magnusson; Pall Melsted; Gudmundur L Norddahl; Jona Saemundsdottir; Asgeir Sigurdsson; Patrick Sulem; Arna B Agustsdottir; Berglind Eiriksdottir; Run Fridriksdottir; Elisabet E Gardarsdottir; Gudmundur Georgsson; Olafia S Gretarsdottir; Kjartan R Gudmundsson; Thora R Gunnarsdottir; Arnaldur Gylfason; Hilma Holm; Brynjar O Jensson; Aslaug Jonasdottir; Kamilla S Josefsdottir; Thordur Kristjansson; Droplaug N Magnusdottir; Louise le Roux; Gudrun Sigmundsdottir; Gardar Sveinbjornsson; Kristin E Sveinsdottir; Maney Sveinsdottir; Emil A Thorarensen; Bjarni Thorbjornsson; Gisli Masson; Ingileif Jonsdottir; Alma Moller; Thorolfur Gudnason; Karl G Kristinsson; Unnur Thorsteinsdottir; Kari Stefansson
EPI_ISL_424467 hCoV-19/Iceland/443/2020 Europe / Iceland / Reykjavik 2020-03-20 The National University Hospital of Iceland deCODE genetics Daniel F Gudbjartsson; Agnar Helgason; Hakon Jonsson; Olafur T Magnusson; Pall Melsted; Gudmundur L Norddahl; Jona Saemundsdottir; Asgeir Sigurdsson; Patrick Sulem; Arna B Agustsdottir; Berglind Eiriksdottir; Run Fridriksdottir; Elisabet E Gardarsdottir; Gudmundur Georgsson; Olafia S Gretarsdottir; Kjartan R Gudmundsson; Thora R Gunnarsdottir; Arnaldur Gylfason; Hilma Holm; Brynjar O Jensson; Aslaug Jonasdottir; Kamilla S Josefsdottir; Thordur Kristjansson; Droplaug N Magnusdottir; Louise le Roux; Gudrun Sigmundsdottir; Gardar Sveinbjornsson; Kristin E Sveinsdottir; Maney Sveinsdottir; Emil A Thorarensen; Bjarni Thorbjornsson; Gisli Masson; Ingileif Jonsdottir; Alma Moller; Thorolfur Gudnason; Karl G Kristinsson; Unnur Thorsteinsdottir; Kari Stefansson
EPI_ISL_424468 hCoV-19/Iceland/444/2020 Europe / Iceland / Reykjavik 2020-03-20 The National University Hospital of Iceland deCODE genetics Daniel F Gudbjartsson; Agnar Helgason; Hakon Jonsson; Olafur T Magnusson; Pall Melsted; Gudmundur L Norddahl; Jona Saemundsdottir; Asgeir Sigurdsson; Patrick Sulem; Arna B Agustsdottir; Berglind Eiriksdottir; Run Fridriksdottir; Elisabet E Gardarsdottir; Gudmundur Georgsson; Olafia S Gretarsdottir; Kjartan R Gudmundsson; Thora R Gunnarsdottir; Arnaldur Gylfason; Hilma Holm; Brynjar O Jensson; Aslaug Jonasdottir; Kamilla S Josefsdottir; Thordur Kristjansson; Droplaug N Magnusdottir; Louise le Roux; Gudrun Sigmundsdottir; Gardar Sveinbjornsson; Kristin E Sveinsdottir; Maney Sveinsdottir; Emil A Thorarensen; Bjarni Thorbjornsson; Gisli Masson; Ingileif Jonsdottir; Alma Moller; Thorolfur Gudnason; Karl G Kristinsson; Unnur Thorsteinsdottir; Kari Stefansson
EPI_ISL_424469 hCoV-19/Iceland/445/2020 Europe / Iceland / Reykjavik 2020-03-20 The National University Hospital of Iceland deCODE genetics Daniel F Gudbjartsson; Agnar Helgason; Hakon Jonsson; Olafur T Magnusson; Pall Melsted; Gudmundur L Norddahl; Jona Saemundsdottir; Asgeir Sigurdsson; Patrick Sulem; Arna B Agustsdottir; Berglind Eiriksdottir; Run Fridriksdottir; Elisabet E Gardarsdottir; Gudmundur Georgsson; Olafia S Gretarsdottir; Kjartan R Gudmundsson; Thora R Gunnarsdottir; Arnaldur Gylfason; Hilma Holm; Brynjar O Jensson; Aslaug Jonasdottir; Kamilla S Josefsdottir; Thordur Kristjansson; Droplaug N Magnusdottir; Louise le Roux; Gudrun Sigmundsdottir; Gardar Sveinbjornsson; Kristin E Sveinsdottir; Maney Sveinsdottir; Emil A Thorarensen; Bjarni Thorbjornsson; Gisli Masson; Ingileif Jonsdottir; Alma Moller; Thorolfur Gudnason; Karl G Kristinsson; Unnur Thorsteinsdottir; Kari Stefansson
EPI_ISL_424470 hCoV-19/Iceland/446/2020 Europe / Iceland / Reykjavik 2020-03-20 The National University Hospital of Iceland deCODE genetics Daniel F Gudbjartsson; Agnar Helgason; Hakon Jonsson; Olafur T Magnusson; Pall Melsted; Gudmundur L Norddahl; Jona Saemundsdottir; Asgeir Sigurdsson; Patrick Sulem; Arna B Agustsdottir; Berglind Eiriksdottir; Run Fridriksdottir; Elisabet E Gardarsdottir; Gudmundur Georgsson; Olafia S Gretarsdottir; Kjartan R Gudmundsson; Thora R Gunnarsdottir; Arnaldur Gylfason; Hilma Holm; Brynjar O Jensson; Aslaug Jonasdottir; Kamilla S Josefsdottir; Thordur Kristjansson; Droplaug N Magnusdottir; Louise le Roux; Gudrun Sigmundsdottir; Gardar Sveinbjornsson; Kristin E Sveinsdottir; Maney Sveinsdottir; Emil A Thorarensen; Bjarni Thorbjornsson; Gisli Masson; Ingileif Jonsdottir; Alma Moller; Thorolfur Gudnason; Karl G Kristinsson; Unnur Thorsteinsdottir; Kari Stefansson
EPI_ISL_424471 hCoV-19/Iceland/447/2020 Europe / Iceland / Reykjavik 2020-03-20 The National University Hospital of Iceland deCODE genetics Daniel F Gudbjartsson; Agnar Helgason; Hakon Jonsson; Olafur T Magnusson; Pall Melsted; Gudmundur L Norddahl; Jona Saemundsdottir; Asgeir Sigurdsson; Patrick Sulem; Arna B Agustsdottir; Berglind Eiriksdottir; Run Fridriksdottir; Elisabet E Gardarsdottir; Gudmundur Georgsson; Olafia S Gretarsdottir; Kjartan R Gudmundsson; Thora R Gunnarsdottir; Arnaldur Gylfason; Hilma Holm; Brynjar O Jensson; Aslaug Jonasdottir; Kamilla S Josefsdottir; Thordur Kristjansson; Droplaug N Magnusdottir; Louise le Roux; Gudrun Sigmundsdottir; Gardar Sveinbjornsson; Kristin E Sveinsdottir; Maney Sveinsdottir; Emil A Thorarensen; Bjarni Thorbjornsson; Gisli Masson; Ingileif Jonsdottir; Alma Moller; Thorolfur Gudnason; Karl G Kristinsson; Unnur Thorsteinsdottir; Kari Stefansson
EPI_ISL_424472 hCoV-19/Iceland/448/2020 Europe / Iceland / Reykjavik 2020-03-20 The National University Hospital of Iceland deCODE genetics Daniel F Gudbjartsson; Agnar Helgason; Hakon Jonsson; Olafur T Magnusson; Pall Melsted; Gudmundur L Norddahl; Jona Saemundsdottir; Asgeir Sigurdsson; Patrick Sulem; Arna B Agustsdottir; Berglind Eiriksdottir; Run Fridriksdottir; Elisabet E Gardarsdottir; Gudmundur Georgsson; Olafia S Gretarsdottir; Kjartan R Gudmundsson; Thora R Gunnarsdottir; Arnaldur Gylfason; Hilma Holm; Brynjar O Jensson; Aslaug Jonasdottir; Kamilla S Josefsdottir; Thordur Kristjansson; Droplaug N Magnusdottir; Louise le Roux; Gudrun Sigmundsdottir; Gardar Sveinbjornsson; Kristin E Sveinsdottir; Maney Sveinsdottir; Emil A Thorarensen; Bjarni Thorbjornsson; Gisli Masson; Ingileif Jonsdottir; Alma Moller; Thorolfur Gudnason; Karl G Kristinsson; Unnur Thorsteinsdottir; Kari Stefansson
EPI_ISL_424473 hCoV-19/Iceland/449/2020 Europe / Iceland / Reykjavik 2020-03-20 The National University Hospital of Iceland deCODE genetics Daniel F Gudbjartsson; Agnar Helgason; Hakon Jonsson; Olafur T Magnusson; Pall Melsted; Gudmundur L Norddahl; Jona Saemundsdottir; Asgeir Sigurdsson; Patrick Sulem; Arna B Agustsdottir; Berglind Eiriksdottir; Run Fridriksdottir; Elisabet E Gardarsdottir; Gudmundur Georgsson; Olafia S Gretarsdottir; Kjartan R Gudmundsson; Thora R Gunnarsdottir; Arnaldur Gylfason; Hilma Holm; Brynjar O Jensson; Aslaug Jonasdottir; Kamilla S Josefsdottir; Thordur Kristjansson; Droplaug N Magnusdottir; Louise le Roux; Gudrun Sigmundsdottir; Gardar Sveinbjornsson; Kristin E Sveinsdottir; Maney Sveinsdottir; Emil A Thorarensen; Bjarni Thorbjornsson; Gisli Masson; Ingileif Jonsdottir; Alma Moller; Thorolfur Gudnason; Karl G Kristinsson; Unnur Thorsteinsdottir; Kari Stefansson

EPI_ISL_424474 hCoV-19/Iceland/450/2020 Europe / Iceland / Reykjavik 2020-03-20 The National University Hospital of Iceland deCODE genetics Daniel F Gudbjartsson; Agnar Helgason; Hakon Jonsson; Olafur T Magnusson; Pall Melsted; Gudmundur L Norddahl; Jona Saemundsdottir; Asgeir Sigurdsson; Patrick Sulem; Arna B Agustsdottir; Berglind Eiriksdottir; Run Fridriksdottir; Elisabet E Gardarsdottir; Gudmundur Georgsson; Olafia S Gretarsdottir; Kjartan R Gudmundsson; Thora R Gunnarsdottir; Arnaldur Gylfason; Hilma Holm; Brynjar O Jensson; Aslaug Jonasdottir; Kamilla S Josefsdottir; Thordur Kristjansson; Droplaug N Magnusdottir; Louise le Roux; Gudrun Sigmundsdottir; Gardar Sveinbjornsson; Kristin E Sveinsdottir; Maney Sveinsdottir; Emil A Thorarensen; Bjarni Thorbjornsson; Gisli Masson; Ingileif Jonsdottir; Alma Moller; Thorolfur Gudnason; Karl G Kristinsson; Unnur Thorsteinsdottir; Kari Stefansson
EPI_ISL_424475 hCoV-19/Iceland/451/2020 Europe / Iceland / Reykjavik 2020-03-20 The National University Hospital of Iceland deCODE genetics Daniel F Gudbjartsson; Agnar Helgason; Hakon Jonsson; Olafur T Magnusson; Pall Melsted; Gudmundur L Norddahl; Jona Saemundsdottir; Asgeir Sigurdsson; Patrick Sulem; Arna B Agustsdottir; Berglind Eiriksdottir; Run Fridriksdottir; Elisabet E Gardarsdottir; Gudmundur Georgsson; Olafia S Gretarsdottir; Kjartan R Gudmundsson; Thora R Gunnarsdottir; Arnaldur Gylfason; Hilma Holm; Brynjar O Jensson; Aslaug Jonasdottir; Kamilla S Josefsdottir; Thordur Kristjansson; Droplaug N Magnusdottir; Louise le Roux; Gudrun Sigmundsdottir; Gardar Sveinbjornsson; Kristin E Sveinsdottir; Maney Sveinsdottir; Emil A Thorarensen; Bjarni Thorbjornsson; Gisli Masson; Ingileif Jonsdottir; Alma Moller; Thorolfur Gudnason; Karl G Kristinsson; Unnur Thorsteinsdottir; Kari Stefansson
EPI_ISL_424476 hCoV-19/Iceland/452/2020 Europe / Iceland / Reykjavik 2020-03-19 The National University Hospital of Iceland deCODE genetics Daniel F Gudbjartsson; Agnar Helgason; Hakon Jonsson; Olafur T Magnusson; Pall Melsted; Gudmundur L Norddahl; Jona Saemundsdottir; Asgeir Sigurdsson; Patrick Sulem; Arna B Agustsdottir; Berglind Eiriksdottir; Run Fridriksdottir; Elisabet E Gardarsdottir; Gudmundur Georgsson; Olafia S Gretarsdottir; Kjartan R Gudmundsson; Thora R Gunnarsdottir; Arnaldur Gylfason; Hilma Holm; Brynjar O Jensson; Aslaug Jonasdottir; Kamilla S Josefsdottir; Thordur Kristjansson; Droplaug N Magnusdottir; Louise le Roux; Gudrun Sigmundsdottir; Gardar Sveinbjornsson; Kristin E Sveinsdottir; Maney Sveinsdottir; Emil A Thorarensen; Bjarni Thorbjornsson; Gisli Masson; Ingileif Jonsdottir; Alma Moller; Thorolfur Gudnason; Karl G Kristinsson; Unnur Thorsteinsdottir; Kari Stefansson
EPI_ISL_424477 hCoV-19/Iceland/454/2020 Europe / Iceland / Reykjavik 2020-03-20 The National University Hospital of Iceland deCODE genetics Daniel F Gudbjartsson; Agnar Helgason; Hakon Jonsson; Olafur T Magnusson; Pall Melsted; Gudmundur L Norddahl; Jona Saemundsdottir; Asgeir Sigurdsson; Patrick Sulem; Arna B Agustsdottir; Berglind Eiriksdottir; Run Fridriksdottir; Elisabet E Gardarsdottir; Gudmundur Georgsson; Olafia S Gretarsdottir; Kjartan R Gudmundsson; Thora R Gunnarsdottir; Arnaldur Gylfason; Hilma Holm; Brynjar O Jensson; Aslaug Jonasdottir; Kamilla S Josefsdottir; Thordur Kristjansson; Droplaug N Magnusdottir; Louise le Roux; Gudrun Sigmundsdottir; Gardar Sveinbjornsson; Kristin E Sveinsdottir; Maney Sveinsdottir; Emil A Thorarensen; Bjarni Thorbjornsson; Gisli Masson; Ingileif Jonsdottir; Alma Moller; Thorolfur Gudnason; Karl G Kristinsson; Unnur Thorsteinsdottir; Kari Stefansson
EPI_ISL_424478 hCoV-19/Iceland/455/2020 Europe / Iceland / Reykjavik 2020-03-20 The National University Hospital of Iceland deCODE genetics Daniel F Gudbjartsson; Agnar Helgason; Hakon Jonsson; Olafur T Magnusson; Pall Melsted; Gudmundur L Norddahl; Jona Saemundsdottir; Asgeir Sigurdsson; Patrick Sulem; Arna B Agustsdottir; Berglind Eiriksdottir; Run Fridriksdottir; Elisabet E Gardarsdottir; Gudmundur Georgsson; Olafia S Gretarsdottir; Kjartan R Gudmundsson; Thora R Gunnarsdottir; Arnaldur Gylfason; Hilma Holm; Brynjar O Jensson; Aslaug Jonasdottir; Kamilla S Josefsdottir; Thordur Kristjansson; Droplaug N Magnusdottir; Louise le Roux; Gudrun Sigmundsdottir; Gardar Sveinbjornsson; Kristin E Sveinsdottir; Maney Sveinsdottir; Emil A Thorarensen; Bjarni Thorbjornsson; Gisli Masson; Ingileif Jonsdottir; Alma Moller; Thorolfur Gudnason; Karl G Kristinsson; Unnur Thorsteinsdottir; Kari Stefansson
EPI_ISL_424479 hCoV-19/Iceland/456/2020 Europe / Iceland / Reykjavik 2020-03-20 The National University Hospital of Iceland deCODE genetics Daniel F Gudbjartsson; Agnar Helgason; Hakon Jonsson; Olafur T Magnusson; Pall Melsted; Gudmundur L Norddahl; Jona Saemundsdottir; Asgeir Sigurdsson; Patrick Sulem; Arna B Agustsdottir; Berglind Eiriksdottir; Run Fridriksdottir; Elisabet E Gardarsdottir; Gudmundur Georgsson; Olafia S Gretarsdottir; Kjartan R Gudmundsson; Thora R Gunnarsdottir; Arnaldur Gylfason; Hilma Holm; Brynjar O Jensson; Aslaug Jonasdottir; Kamilla S Josefsdottir; Thordur Kristjansson; Droplaug N Magnusdottir; Louise le Roux; Gudrun Sigmundsdottir; Gardar Sveinbjornsson; Kristin E Sveinsdottir; Maney Sveinsdottir; Emil A Thorarensen; Bjarni Thorbjornsson; Gisli Masson; Ingileif Jonsdottir; Alma Moller; Thorolfur Gudnason; Karl G Kristinsson; Unnur Thorsteinsdottir; Kari Stefansson
EPI_ISL_424480 hCoV-19/Iceland/457/2020 Europe / Iceland / Reykjavik 2020-03-20 The National University Hospital of Iceland deCODE genetics Daniel F Gudbjartsson; Agnar Helgason; Hakon Jonsson; Olafur T Magnusson; Pall Melsted; Gudmundur L Norddahl; Jona Saemundsdottir; Asgeir Sigurdsson; Patrick Sulem; Arna B Agustsdottir; Berglind Eiriksdottir; Run Fridriksdottir; Elisabet E Gardarsdottir; Gudmundur Georgsson; Olafia S Gretarsdottir; Kjartan R Gudmundsson; Thora R Gunnarsdottir; Arnaldur Gylfason; Hilma Holm; Brynjar O Jensson; Aslaug Jonasdottir; Kamilla S Josefsdottir; Thordur Kristjansson; Droplaug N Magnusdottir; Louise le Roux; Gudrun Sigmundsdottir; Gardar Sveinbjornsson; Kristin E Sveinsdottir; Maney Sveinsdottir; Emil A Thorarensen; Bjarni Thorbjornsson; Gisli Masson; Ingileif Jonsdottir; Alma Moller; Thorolfur Gudnason; Karl G Kristinsson; Unnur Thorsteinsdottir; Kari Stefansson
EPI_ISL_424482 hCoV-19/Iceland/459/2020 Europe / Iceland / Reykjavik 2020-03-20 The National University Hospital of Iceland deCODE genetics Daniel F Gudbjartsson; Agnar Helgason; Hakon Jonsson; Olafur T Magnusson; Pall Melsted; Gudmundur L Norddahl; Jona Saemundsdottir; Asgeir Sigurdsson; Patrick Sulem; Arna B Agustsdottir; Berglind Eiriksdottir; Run Fridriksdottir; Elisabet E Gardarsdottir; Gudmundur Georgsson; Olafia S Gretarsdottir; Kjartan R Gudmundsson; Thora R Gunnarsdottir; Arnaldur Gylfason; Hilma Holm; Brynjar O Jensson; Aslaug Jonasdottir; Kamilla S Josefsdottir; Thordur Kristjansson; Droplaug N Magnusdottir; Louise le Roux; Gudrun Sigmundsdottir; Gardar Sveinbjornsson; Kristin E Sveinsdottir; Maney Sveinsdottir; Emil A

Thorarensen; Bjarni Thorbjornsson; Gisli Masson; Ingileif Jonsdottir; Alma Moller; Thorolfur Gudnason; Karl G Kristinsson; Unnur Thorsteinsdottir; Kari Stefansson
EPI_ISL_424483 hCoV-19/Iceland/460/2020 Europe / Iceland / Reykjavik 2020-03-20 The National University Hospital of Iceland deCODE genetics Daniel F Gudbjartsson; Agnar Helgason; Hakon Jonsson; Olafur T Magnusson; Pall Melsted; Gudmundur L Norddahl; Jona Saemundsdottir; Asgeir Sigurdsson; Patrick Sulem; Arna B Agustsdottir; Berglind Eiriksdottir; Run Fridriksdottir; Elisabet E Gardarsdottir; Gudmundur Georgsson; Olafia S Gretarsdottir; Kjartan R Gudmundsson; Thora R Gunnarsdottir; Arnaldur Gylfason; Hilma Holm; Brynjar O Jensson; Aslaug Jonasdottir; Kamilla S Josefsdottir; Thordur Kristjansson; Droplaug N Magnusdottir; Louise le Roux; Gudrun Sigmundsdottir; Gardar Sveinbjornsson; Kristin E Sveinsdottir; Maney Sveinsdottir; Emil A Thorarensen; Bjarni Thorbjornsson; Gisli Masson; Ingileif Jonsdottir; Alma Moller; Thorolfur Gudnason; Karl G Kristinsson; Unnur Thorsteinsdottir; Kari Stefansson
EPI_ISL_424484 hCoV-19/Iceland/461/2020 Europe / Iceland / Reykjavik 2020-03-20 The National University Hospital of Iceland deCODE genetics Daniel F Gudbjartsson; Agnar Helgason; Hakon Jonsson; Olafur T Magnusson; Pall Melsted; Gudmundur L Norddahl; Jona Saemundsdottir; Asgeir Sigurdsson; Patrick Sulem; Arna B Agustsdottir; Berglind Eiriksdottir; Run Fridriksdottir; Elisabet E Gardarsdottir; Gudmundur Georgsson; Olafia S Gretarsdottir; Kjartan R Gudmundsson; Thora R Gunnarsdottir; Arnaldur Gylfason; Hilma Holm; Brynjar O Jensson; Aslaug Jonasdottir; Kamilla S Josefsdottir; Thordur Kristjansson; Droplaug N Magnusdottir; Louise le Roux; Gudrun Sigmundsdottir; Gardar Sveinbjornsson; Kristin E Sveinsdottir; Maney Sveinsdottir; Emil A Thorarensen; Bjarni Thorbjornsson; Gisli Masson; Ingileif Jonsdottir; Alma Moller; Thorolfur Gudnason; Karl G Kristinsson; Unnur Thorsteinsdottir; Kari Stefansson
EPI_ISL_424485 hCoV-19/Iceland/462/2020 Europe / Iceland / Reykjavik 2020-03-20 The National University Hospital of Iceland deCODE genetics Daniel F Gudbjartsson; Agnar Helgason; Hakon Jonsson; Olafur T Magnusson; Pall Melsted; Gudmundur L Norddahl; Jona Saemundsdottir; Asgeir Sigurdsson; Patrick Sulem; Arna B Agustsdottir; Berglind Eiriksdottir; Run Fridriksdottir; Elisabet E Gardarsdottir; Gudmundur Georgsson; Olafia S Gretarsdottir; Kjartan R Gudmundsson; Thora R Gunnarsdottir; Arnaldur Gylfason; Hilma Holm; Brynjar O Jensson; Aslaug Jonasdottir; Kamilla S Josefsdottir; Thordur Kristjansson; Droplaug N Magnusdottir; Louise le Roux; Gudrun Sigmundsdottir; Gardar Sveinbjornsson; Kristin E Sveinsdottir; Maney Sveinsdottir; Emil A Thorarensen; Bjarni Thorbjornsson; Gisli Masson; Ingileif Jonsdottir; Alma Moller; Thorolfur Gudnason; Karl G Kristinsson; Unnur Thorsteinsdottir; Kari Stefansson
EPI_ISL_424486 hCoV-19/Iceland/463/2020 Europe / Iceland / Reykjavik 2020-03-20 The National University Hospital of Iceland deCODE genetics Daniel F Gudbjartsson; Agnar Helgason; Hakon Jonsson; Olafur T Magnusson; Pall Melsted; Gudmundur L Norddahl; Jona Saemundsdottir; Asgeir Sigurdsson; Patrick Sulem; Arna B Agustsdottir; Berglind Eiriksdottir; Run Fridriksdottir; Elisabet E Gardarsdottir; Gudmundur Georgsson; Olafia S Gretarsdottir; Kjartan R Gudmundsson; Thora R Gunnarsdottir; Arnaldur Gylfason; Hilma Holm; Brynjar O Jensson; Aslaug Jonasdottir; Kamilla S Josefsdottir; Thordur Kristjansson; Droplaug N Magnusdottir; Louise le Roux; Gudrun Sigmundsdottir; Gardar Sveinbjornsson; Kristin E Sveinsdottir; Maney Sveinsdottir; Emil A Thorarensen; Bjarni Thorbjornsson; Gisli Masson; Ingileif Jonsdottir; Alma Moller; Thorolfur Gudnason; Karl G Kristinsson; Unnur Thorsteinsdottir; Kari Stefansson
EPI_ISL_424487 hCoV-19/Iceland/464/2020 Europe / Iceland / Reykjavik 2020-03-20 The National University Hospital of Iceland deCODE genetics Daniel F Gudbjartsson; Agnar Helgason; Hakon Jonsson; Olafur T Magnusson; Pall Melsted; Gudmundur L Norddahl; Jona Saemundsdottir; Asgeir Sigurdsson; Patrick Sulem; Arna B Agustsdottir; Berglind Eiriksdottir; Run Fridriksdottir; Elisabet E Gardarsdottir; Gudmundur Georgsson; Olafia S Gretarsdottir; Kjartan R Gudmundsson; Thora R Gunnarsdottir; Arnaldur Gylfason; Hilma Holm; Brynjar O Jensson; Aslaug Jonasdottir; Kamilla S Josefsdottir; Thordur Kristjansson; Droplaug N Magnusdottir; Louise le Roux; Gudrun Sigmundsdottir; Gardar Sveinbjornsson; Kristin E Sveinsdottir; Maney Sveinsdottir; Emil A Thorarensen; Bjarni Thorbjornsson; Gisli Masson; Ingileif Jonsdottir; Alma Moller; Thorolfur Gudnason; Karl G Kristinsson; Unnur Thorsteinsdottir; Kari Stefansson
EPI_ISL_424489 hCoV-19/Iceland/466/2020 Europe / Iceland / Reykjavik 2020-03-20 The National University Hospital of Iceland deCODE genetics Daniel F Gudbjartsson; Agnar Helgason; Hakon Jonsson; Olafur T Magnusson; Pall Melsted; Gudmundur L Norddahl; Jona Saemundsdottir; Asgeir Sigurdsson; Patrick Sulem; Arna B Agustsdottir; Berglind Eiriksdottir; Run Fridriksdottir; Elisabet E Gardarsdottir; Gudmundur Georgsson; Olafia S Gretarsdottir; Kjartan R Gudmundsson; Thora R Gunnarsdottir; Arnaldur Gylfason; Hilma Holm; Brynjar O Jensson; Aslaug Jonasdottir; Kamilla S Josefsdottir; Thordur Kristjansson; Droplaug N Magnusdottir; Louise le Roux; Gudrun Sigmundsdottir; Gardar Sveinbjornsson; Kristin E Sveinsdottir; Maney Sveinsdottir; Emil A Thorarensen; Bjarni Thorbjornsson; Gisli Masson; Ingileif Jonsdottir; Alma Moller; Thorolfur Gudnason; Karl G Kristinsson; Unnur Thorsteinsdottir; Kari Stefansson
EPI_ISL_424490 hCoV-19/Iceland/467/2020 Europe / Iceland / Reykjavik 2020-03-20 The National University Hospital of Iceland deCODE genetics Daniel F Gudbjartsson; Agnar Helgason; Hakon Jonsson; Olafur T Magnusson; Pall Melsted; Gudmundur L Norddahl; Jona Saemundsdottir; Asgeir Sigurdsson; Patrick Sulem; Arna B Agustsdottir; Berglind Eiriksdottir; Run Fridriksdottir; Elisabet E Gardarsdottir; Gudmundur Georgsson; Olafia S Gretarsdottir; Kjartan R Gudmundsson; Thora R Gunnarsdottir; Arnaldur Gylfason; Hilma Holm; Brynjar O Jensson; Aslaug Jonasdottir; Kamilla S Josefsdottir; Thordur Kristjansson; Droplaug N Magnusdottir; Louise le Roux; Gudrun Sigmundsdottir; Gardar Sveinbjornsson; Kristin E Sveinsdottir; Maney Sveinsdottir; Emil A Thorarensen; Bjarni Thorbjornsson; Gisli Masson; Ingileif Jonsdottir; Alma Moller; Thorolfur Gudnason; Karl G Kristinsson; Unnur Thorsteinsdottir; Kari Stefansson
EPI_ISL_424491 hCoV-19/Iceland/468/2020 Europe / Iceland / Reykjavik 2020-03-20 The National University Hospital of Iceland deCODE genetics Daniel F Gudbjartsson; Agnar Helgason; Hakon Jonsson; Olafur T Magnusson; Pall Melsted; Gudmundur L Norddahl; Jona Saemundsdottir; Asgeir Sigurdsson; Patrick Sulem; Arna B Agustsdottir; Berglind Eiriksdottir; Run Fridriksdottir; Elisabet E Gardarsdottir; Gudmundur Georgsson; Olafia S Gretarsdottir; Kjartan R Gudmundsson; Thora R Gunnarsdottir; Arnaldur Gylfason; Hilma Holm; Brynjar O

Jensson; Aslaug Jonasdottir; Kamilla S Josefsdottir; Thordur Kristjansson; Droplaug N Magnusdottir; Louise le Roux; Gudrun Sigmundsdottir; Gardar Sveinbjornsson; Kristin E Sveinsdottir; Maney Sveinsdottir; Emil A Thorarensen; Bjarni Thorbjornsson; Gisli Masson; Ingileif Jonsdottir; Alma Moller; Thorolfur Gudnason; Karl G Kristinsson; Unnur Thorsteinsdottir; Kari Stefansson
EPI_ISL_424492 hCoV-19/Iceland/469/2020 Europe / Iceland / Reykjavik 2020-03-21 The National University Hospital of Iceland deCODE genetics Daniel F Gudbjartsson; Agnar Helgason; Hakon Jonsson; Olafur T Magnusson; Pall Melsted; Gudmundur L Norddahl; Jona Saemundsdottir; Asgeir Sigurdsson; Patrick Sulem; Arna B Agustsdottir; Berglind Eiriksdottir; Run Fridriksdottir; Elisabet E Gardarsdottir; Gudmundur Georgsson; Olafia S Gretarsdottir; Kjartan R Gudmundsson; Thora R Gunnarsdottir; Arnaldur Gylfason; Hilma Holm; Brynjar O Jensson; Aslaug Jonasdottir; Kamilla S Josefsdottir; Thordur Kristjansson; Droplaug N Magnusdottir; Louise le Roux; Gudrun Sigmundsdottir; Gardar Sveinbjornsson; Kristin E Sveinsdottir; Maney Sveinsdottir; Emil A Thorarensen; Bjarni Thorbjornsson; Gisli Masson; Ingileif Jonsdottir; Alma Moller; Thorolfur Gudnason; Karl G Kristinsson; Unnur Thorsteinsdottir; Kari Stefansson
EPI_ISL_424493 hCoV-19/Iceland/470/2020 Europe / Iceland / Reykjavik 2020-03-21 The National University Hospital of Iceland deCODE genetics Daniel F Gudbjartsson; Agnar Helgason; Hakon Jonsson; Olafur T Magnusson; Pall Melsted; Gudmundur L Norddahl; Jona Saemundsdottir; Asgeir Sigurdsson; Patrick Sulem; Arna B Agustsdottir; Berglind Eiriksdottir; Run Fridriksdottir; Elisabet E Gardarsdottir; Gudmundur Georgsson; Olafia S Gretarsdottir; Kjartan R Gudmundsson; Thora R Gunnarsdottir; Arnaldur Gylfason; Hilma Holm; Brynjar O Jensson; Aslaug Jonasdottir; Kamilla S Josefsdottir; Thordur Kristjansson; Droplaug N Magnusdottir; Louise le Roux; Gudrun Sigmundsdottir; Gardar Sveinbjornsson; Kristin E Sveinsdottir; Maney Sveinsdottir; Emil A Thorarensen; Bjarni Thorbjornsson; Gisli Masson; Ingileif Jonsdottir; Alma Moller; Thorolfur Gudnason; Karl G Kristinsson; Unnur Thorsteinsdottir; Kari Stefansson
EPI_ISL_424494 hCoV-19/Iceland/471/2020 Europe / Iceland / Reykjavik 2020-03-20 The National University Hospital of Iceland deCODE genetics Daniel F Gudbjartsson; Agnar Helgason; Hakon Jonsson; Olafur T Magnusson; Pall Melsted; Gudmundur L Norddahl; Jona Saemundsdottir; Asgeir Sigurdsson; Patrick Sulem; Arna B Agustsdottir; Berglind Eiriksdottir; Run Fridriksdottir; Elisabet E Gardarsdottir; Gudmundur Georgsson; Olafia S Gretarsdottir; Kjartan R Gudmundsson; Thora R Gunnarsdottir; Arnaldur Gylfason; Hilma Holm; Brynjar O Jensson; Aslaug Jonasdottir; Kamilla S Josefsdottir; Thordur Kristjansson; Droplaug N Magnusdottir; Louise le Roux; Gudrun Sigmundsdottir; Gardar Sveinbjornsson; Kristin E Sveinsdottir; Maney Sveinsdottir; Emil A Thorarensen; Bjarni Thorbjornsson; Gisli Masson; Ingileif Jonsdottir; Alma Moller; Thorolfur Gudnason; Karl G Kristinsson; Unnur Thorsteinsdottir; Kari Stefansson
EPI_ISL_424495 hCoV-19/Iceland/472/2020 Europe / Iceland / Reykjavik 2020-03-20 The National University Hospital of Iceland deCODE genetics Daniel F Gudbjartsson; Agnar Helgason; Hakon Jonsson; Olafur T Magnusson; Pall Melsted; Gudmundur L Norddahl; Jona Saemundsdottir; Asgeir Sigurdsson; Patrick Sulem; Arna B Agustsdottir; Berglind Eiriksdottir; Run Fridriksdottir; Elisabet E Gardarsdottir; Gudmundur Georgsson; Olafia S Gretarsdottir; Kjartan R Gudmundsson; Thora R Gunnarsdottir; Arnaldur Gylfason; Hilma Holm; Brynjar O Jensson; Aslaug Jonasdottir; Kamilla S Josefsdottir; Thordur Kristjansson; Droplaug N Magnusdottir; Louise le Roux; Gudrun Sigmundsdottir; Gardar Sveinbjornsson; Kristin E Sveinsdottir; Maney Sveinsdottir; Emil A Thorarensen; Bjarni Thorbjornsson; Gisli Masson; Ingileif Jonsdottir; Alma Moller; Thorolfur Gudnason; Karl G Kristinsson; Unnur Thorsteinsdottir; Kari Stefansson
EPI_ISL_424496 hCoV-19/Iceland/473/2020 Europe / Iceland / Reykjavik 2020-03-20 The National University Hospital of Iceland deCODE genetics Daniel F Gudbjartsson; Agnar Helgason; Hakon Jonsson; Olafur T Magnusson; Pall Melsted; Gudmundur L Norddahl; Jona Saemundsdottir; Asgeir Sigurdsson; Patrick Sulem; Arna B Agustsdottir; Berglind Eiriksdottir; Run Fridriksdottir; Elisabet E Gardarsdottir; Gudmundur Georgsson; Olafia S Gretarsdottir; Kjartan R Gudmundsson; Thora R Gunnarsdottir; Arnaldur Gylfason; Hilma Holm; Brynjar O Jensson; Aslaug Jonasdottir; Kamilla S Josefsdottir; Thordur Kristjansson; Droplaug N Magnusdottir; Louise le Roux; Gudrun Sigmundsdottir; Gardar Sveinbjornsson; Kristin E Sveinsdottir; Maney Sveinsdottir; Emil A Thorarensen; Bjarni Thorbjornsson; Gisli Masson; Ingileif Jonsdottir; Alma Moller; Thorolfur Gudnason; Karl G Kristinsson; Unnur Thorsteinsdottir; Kari Stefansson
EPI_ISL_424498 hCoV-19/Iceland/475/2020 Europe / Iceland / Reykjavik 2020-03-20 The National University Hospital of Iceland deCODE genetics Daniel F Gudbjartsson; Agnar Helgason; Hakon Jonsson; Olafur T Magnusson; Pall Melsted; Gudmundur L Norddahl; Jona Saemundsdottir; Asgeir Sigurdsson; Patrick Sulem; Arna B Agustsdottir; Berglind Eiriksdottir; Run Fridriksdottir; Elisabet E Gardarsdottir; Gudmundur Georgsson; Olafia S Gretarsdottir; Kjartan R Gudmundsson; Thora R Gunnarsdottir; Arnaldur Gylfason; Hilma Holm; Brynjar O Jensson; Aslaug Jonasdottir; Kamilla S Josefsdottir; Thordur Kristjansson; Droplaug N Magnusdottir; Louise le Roux; Gudrun Sigmundsdottir; Gardar Sveinbjornsson; Kristin E Sveinsdottir; Maney Sveinsdottir; Emil A Thorarensen; Bjarni Thorbjornsson; Gisli Masson; Ingileif Jonsdottir; Alma Moller; Thorolfur Gudnason; Karl G Kristinsson; Unnur Thorsteinsdottir; Kari Stefansson
EPI_ISL_424499 hCoV-19/Iceland/476/2020 Europe / Iceland / Reykjavik 2020-03-20 The National University Hospital of Iceland deCODE genetics Daniel F Gudbjartsson; Agnar Helgason; Hakon Jonsson; Olafur T Magnusson; Pall Melsted; Gudmundur L Norddahl; Jona Saemundsdottir; Asgeir Sigurdsson; Patrick Sulem; Arna B Agustsdottir; Berglind Eiriksdottir; Run Fridriksdottir; Elisabet E Gardarsdottir; Gudmundur Georgsson; Olafia S Gretarsdottir; Kjartan R Gudmundsson; Thora R Gunnarsdottir; Arnaldur Gylfason; Hilma Holm; Brynjar O Jensson; Aslaug Jonasdottir; Kamilla S Josefsdottir; Thordur Kristjansson; Droplaug N Magnusdottir; Louise le Roux; Gudrun Sigmundsdottir; Gardar Sveinbjornsson; Kristin E Sveinsdottir; Maney Sveinsdottir; Emil A Thorarensen; Bjarni Thorbjornsson; Gisli Masson; Ingileif Jonsdottir; Alma Moller; Thorolfur Gudnason; Karl G Kristinsson; Unnur Thorsteinsdottir; Kari Stefansson
EPI_ISL_424580 hCoV-19/Iceland/560/2020 Europe / Iceland / Reykjavik 2020-03-27 The National University Hospital of Iceland deCODE genetics Daniel F Gudbjartsson; Agnar Helgason; Hakon Jonsson; Olafur T Magnusson; Pall Melsted; Gudmundur L Norddahl; Jona Saemundsdottir; Asgeir Sigurdsson; Patrick Sulem; Arna B

Agustsdottir; Berglind Eiriksdottir; Run Fridriksdottir; Elisabet E Gardarsdottir; Gudmundur Georgsson; Olafia S Gretarsdottir; Kjartan R Gudmundsson; Thora R Gunnarsdottir; Arnaldur Gylfason; Hilma Holm; Brynjar O Jensson; Aslaug Jonasdottir; Kamilla S Josefsdottir; Thordur Kristjansson; Droplaug N Magnusdottir; Louise le Roux; Gudrun Sigmundsdottir; Gardar Sveinbjornsson; Kristin E Sveinsdottir; Maney Sveinsdottir; Emil A Thorarensen; Bjarni Thorbjornsson; Gisli Masson; Ingileif Jonsdottir; Alma Moller; Thorolfur Gudnason; Karl G Kristinsson; Unnur Thorsteinsdottir; Kari Stefansson
EPI_ISL_424581 hCoV-19/Iceland/561/2020 Europe / Iceland / Reykjavik 2020-03-28 The National University Hospital of Iceland deCODE genetics Daniel F Gudbjartsson; Agnar Helgason; Hakon Jonsson; Olafur T Magnusson; Pall Melsted; Gudmundur L Norddahl; Jona Saemundsdottir; Asgeir Sigurdsson; Patrick Sulem; Arna B Agustsdottir; Berglind Eiriksdottir; Run Fridriksdottir; Elisabet E Gardarsdottir; Gudmundur Georgsson; Olafia S Gretarsdottir; Kjartan R Gudmundsson; Thora R Gunnarsdottir; Arnaldur Gylfason; Hilma Holm; Brynjar O Jensson; Aslaug Jonasdottir; Kamilla S Josefsdottir; Thordur Kristjansson; Droplaug N Magnusdottir; Louise le Roux; Gudrun Sigmundsdottir; Gardar Sveinbjornsson; Kristin E Sveinsdottir; Maney Sveinsdottir; Emil A Thorarensen; Bjarni Thorbjornsson; Gisli Masson; Ingileif Jonsdottir; Alma Moller; Thorolfur Gudnason; Karl G Kristinsson; Unnur Thorsteinsdottir; Kari Stefansson
EPI_ISL_424582 hCoV-19/Iceland/562/2020 Europe / Iceland / Reykjavik 2020-03-27 The National University Hospital of Iceland deCODE genetics Daniel F Gudbjartsson; Agnar Helgason; Hakon Jonsson; Olafur T Magnusson; Pall Melsted; Gudmundur L Norddahl; Jona Saemundsdottir; Asgeir Sigurdsson; Patrick Sulem; Arna B Agustsdottir; Berglind Eiriksdottir; Run Fridriksdottir; Elisabet E Gardarsdottir; Gudmundur Georgsson; Olafia S Gretarsdottir; Kjartan R Gudmundsson; Thora R Gunnarsdottir; Arnaldur Gylfason; Hilma Holm; Brynjar O Jensson; Aslaug Jonasdottir; Kamilla S Josefsdottir; Thordur Kristjansson; Droplaug N Magnusdottir; Louise le Roux; Gudrun Sigmundsdottir; Gardar Sveinbjornsson; Kristin E Sveinsdottir; Maney Sveinsdottir; Emil A Thorarensen; Bjarni Thorbjornsson; Gisli Masson; Ingileif Jonsdottir; Alma Moller; Thorolfur Gudnason; Karl G Kristinsson; Unnur Thorsteinsdottir; Kari Stefansson
EPI_ISL_424584 hCoV-19/Iceland/564/2020 Europe / Iceland / Reykjavik 2020-03-28 The National University Hospital of Iceland deCODE genetics Daniel F Gudbjartsson; Agnar Helgason; Hakon Jonsson; Olafur T Magnusson; Pall Melsted; Gudmundur L Norddahl; Jona Saemundsdottir; Asgeir Sigurdsson; Patrick Sulem; Arna B Agustsdottir; Berglind Eiriksdottir; Run Fridriksdottir; Elisabet E Gardarsdottir; Gudmundur Georgsson; Olafia S Gretarsdottir; Kjartan R Gudmundsson; Thora R Gunnarsdottir; Arnaldur Gylfason; Hilma Holm; Brynjar O Jensson; Aslaug Jonasdottir; Kamilla S Josefsdottir; Thordur Kristjansson; Droplaug N Magnusdottir; Louise le Roux; Gudrun Sigmundsdottir; Gardar Sveinbjornsson; Kristin E Sveinsdottir; Maney Sveinsdottir; Emil A Thorarensen; Bjarni Thorbjornsson; Gisli Masson; Ingileif Jonsdottir; Alma Moller; Thorolfur Gudnason; Karl G Kristinsson; Unnur Thorsteinsdottir; Kari Stefansson
EPI_ISL_424585 hCoV-19/Iceland/565/2020 Europe / Iceland / Reykjavik 2020-03-28 The National University Hospital of Iceland deCODE genetics Daniel F Gudbjartsson; Agnar Helgason; Hakon Jonsson; Olafur T Magnusson; Pall Melsted; Gudmundur L Norddahl; Jona Saemundsdottir; Asgeir Sigurdsson; Patrick Sulem; Arna B Agustsdottir; Berglind Eiriksdottir; Run Fridriksdottir; Elisabet E Gardarsdottir; Gudmundur Georgsson; Olafia S Gretarsdottir; Kjartan R Gudmundsson; Thora R Gunnarsdottir; Arnaldur Gylfason; Hilma Holm; Brynjar O Jensson; Aslaug Jonasdottir; Kamilla S Josefsdottir; Thordur Kristjansson; Droplaug N Magnusdottir; Louise le Roux; Gudrun Sigmundsdottir; Gardar Sveinbjornsson; Kristin E Sveinsdottir; Maney Sveinsdottir; Emil A Thorarensen; Bjarni Thorbjornsson; Gisli Masson; Ingileif Jonsdottir; Alma Moller; Thorolfur Gudnason; Karl G Kristinsson; Unnur Thorsteinsdottir; Kari Stefansson
EPI_ISL_424588 hCoV-19/Iceland/568/2020 Europe / Iceland / Reykjavik 2020-03-28 The National University Hospital of Iceland deCODE genetics Daniel F Gudbjartsson; Agnar Helgason; Hakon Jonsson; Olafur T Magnusson; Pall Melsted; Gudmundur L Norddahl; Jona Saemundsdottir; Asgeir Sigurdsson; Patrick Sulem; Arna B Agustsdottir; Berglind Eiriksdottir; Run Fridriksdottir; Elisabet E Gardarsdottir; Gudmundur Georgsson; Olafia S Gretarsdottir; Kjartan R Gudmundsson; Thora R Gunnarsdottir; Arnaldur Gylfason; Hilma Holm; Brynjar O Jensson; Aslaug Jonasdottir; Kamilla S Josefsdottir; Thordur Kristjansson; Droplaug N Magnusdottir; Louise le Roux; Gudrun Sigmundsdottir; Gardar Sveinbjornsson; Kristin E Sveinsdottir; Maney Sveinsdottir; Emil A Thorarensen; Bjarni Thorbjornsson; Gisli Masson; Ingileif Jonsdottir; Alma Moller; Thorolfur Gudnason; Karl G Kristinsson; Unnur Thorsteinsdottir; Kari Stefansson
EPI_ISL_424589 hCoV-19/Iceland/569/2020 Europe / Iceland / Reykjavik 2020-03-28 The National University Hospital of Iceland deCODE genetics Daniel F Gudbjartsson; Agnar Helgason; Hakon Jonsson; Olafur T Magnusson; Pall Melsted; Gudmundur L Norddahl; Jona Saemundsdottir; Asgeir Sigurdsson; Patrick Sulem; Arna B Agustsdottir; Berglind Eiriksdottir; Run Fridriksdottir; Elisabet E Gardarsdottir; Gudmundur Georgsson; Olafia S Gretarsdottir; Kjartan R Gudmundsson; Thora R Gunnarsdottir; Arnaldur Gylfason; Hilma Holm; Brynjar O Jensson; Aslaug Jonasdottir; Kamilla S Josefsdottir; Thordur Kristjansson; Droplaug N Magnusdottir; Louise le Roux; Gudrun Sigmundsdottir; Gardar Sveinbjornsson; Kristin E Sveinsdottir; Maney Sveinsdottir; Emil A Thorarensen; Bjarni Thorbjornsson; Gisli Masson; Ingileif Jonsdottir; Alma Moller; Thorolfur Gudnason; Karl G Kristinsson; Unnur Thorsteinsdottir; Kari Stefansson
EPI_ISL_424590 hCoV-19/Iceland/570/2020 Europe / Iceland / Reykjavik 2020-03-28 The National University Hospital of Iceland deCODE genetics Daniel F Gudbjartsson; Agnar Helgason; Hakon Jonsson; Olafur T Magnusson; Pall Melsted; Gudmundur L Norddahl; Jona Saemundsdottir; Asgeir Sigurdsson; Patrick Sulem; Arna B Agustsdottir; Berglind Eiriksdottir; Run Fridriksdottir; Elisabet E Gardarsdottir; Gudmundur Georgsson; Olafia S Gretarsdottir; Kjartan R Gudmundsson; Thora R Gunnarsdottir; Arnaldur Gylfason; Hilma Holm; Brynjar O Jensson; Aslaug Jonasdottir; Kamilla S Josefsdottir; Thordur Kristjansson; Droplaug N Magnusdottir; Louise le Roux; Gudrun Sigmundsdottir; Gardar Sveinbjornsson; Kristin E Sveinsdottir; Maney Sveinsdottir; Emil A Thorarensen; Bjarni Thorbjornsson; Gisli Masson; Ingileif Jonsdottir; Alma Moller; Thorolfur Gudnason; Karl G Kristinsson; Unnur Thorsteinsdottir; Kari Stefansson
EPI_ISL_424591 hCoV-19/Iceland/571/2020 Europe / Iceland / Reykjavik 2020-03-28 The National

University Hospital of Iceland  deCODE genetics Daniel F Gudbjartsson; Agnar Helgason; Hakon Jonsson; Olafur T Magnusson; Pall Melsted; Gudmundur L Norddahl; Jona Saemundsdottir; Asgeir Sigurdsson; Patrick Sulem; Arna B Agustsdottir; Berglind Eiriksdottir; Run Fridriksdottir; Elisabet E Gardarsdottir; Gudmundur Georgsson; Olafia S Gretarsdottir; Kjartan R Gudmundsson; Thora R Gunnarsdottir; Arnaldur Gylfason; Hilma Holm; Brynjar O Jensson; Aslaug Jonasdottir; Kamilla S Josefsdottir; Thordur Kristjansson; Droplaug N Magnusdottir; Louise le Roux; Gudrun Sigmundsdottir; Gardar Sveinbjornsson; Kristin E Sveinsdottir; Maney Sveinsdottir; Emil A Thorarensen; Bjarni Thorbjornsson; Gisli Masson; Ingileif Jonsdottir; Alma Moller; Thorolfur Gudnason; Karl G Kristinsson; Unnur Thorsteinsdottir; Kari Stefansson
EPI_ISL_424592  hCoV-19/Iceland/572/2020  Europe / Iceland / Reykjavik  2020-03-28  The National University Hospital of Iceland  deCODE genetics Daniel F Gudbjartsson; Agnar Helgason; Hakon Jonsson; Olafur T Magnusson; Pall Melsted; Gudmundur L Norddahl; Jona Saemundsdottir; Asgeir Sigurdsson; Patrick Sulem; Arna B Agustsdottir; Berglind Eiriksdottir; Run Fridriksdottir; Elisabet E Gardarsdottir; Gudmundur Georgsson; Olafia S Gretarsdottir; Kjartan R Gudmundsson; Thora R Gunnarsdottir; Arnaldur Gylfason; Hilma Holm; Brynjar O Jensson; Aslaug Jonasdottir; Kamilla S Josefsdottir; Thordur Kristjansson; Droplaug N Magnusdottir; Louise le Roux; Gudrun Sigmundsdottir; Gardar Sveinbjornsson; Kristin E Sveinsdottir; Maney Sveinsdottir; Emil A Thorarensen; Bjarni Thorbjornsson; Gisli Masson; Ingileif Jonsdottir; Alma Moller; Thorolfur Gudnason; Karl G Kristinsson; Unnur Thorsteinsdottir; Kari Stefansson
EPI_ISL_424593  hCoV-19/Iceland/573/2020  Europe / Iceland / Reykjavik  2020-03-28  The National University Hospital of Iceland  deCODE genetics Daniel F Gudbjartsson; Agnar Helgason; Hakon Jonsson; Olafur T Magnusson; Pall Melsted; Gudmundur L Norddahl; Jona Saemundsdottir; Asgeir Sigurdsson; Patrick Sulem; Arna B Agustsdottir; Berglind Eiriksdottir; Run Fridriksdottir; Elisabet E Gardarsdottir; Gudmundur Georgsson; Olafia S Gretarsdottir; Kjartan R Gudmundsson; Thora R Gunnarsdottir; Arnaldur Gylfason; Hilma Holm; Brynjar O Jensson; Aslaug Jonasdottir; Kamilla S Josefsdottir; Thordur Kristjansson; Droplaug N Magnusdottir; Louise le Roux; Gudrun Sigmundsdottir; Gardar Sveinbjornsson; Kristin E Sveinsdottir; Maney Sveinsdottir; Emil A Thorarensen; Bjarni Thorbjornsson; Gisli Masson; Ingileif Jonsdottir; Alma Moller; Thorolfur Gudnason; Karl G Kristinsson; Unnur Thorsteinsdottir; Kari Stefansson
EPI_ISL_424595  hCoV-19/Iceland/575/2020  Europe / Iceland / Reykjavik  2020-03-28  The National University Hospital of Iceland  deCODE genetics Daniel F Gudbjartsson; Agnar Helgason; Hakon Jonsson; Olafur T Magnusson; Pall Melsted; Gudmundur L Norddahl; Jona Saemundsdottir; Asgeir Sigurdsson; Patrick Sulem; Arna B Agustsdottir; Berglind Eiriksdottir; Run Fridriksdottir; Elisabet E Gardarsdottir; Gudmundur Georgsson; Olafia S Gretarsdottir; Kjartan R Gudmundsson; Thora R Gunnarsdottir; Arnaldur Gylfason; Hilma Holm; Brynjar O Jensson; Aslaug Jonasdottir; Kamilla S Josefsdottir; Thordur Kristjansson; Droplaug N Magnusdottir; Louise le Roux; Gudrun Sigmundsdottir; Gardar Sveinbjornsson; Kristin E Sveinsdottir; Maney Sveinsdottir; Emil A Thorarensen; Bjarni Thorbjornsson; Gisli Masson; Ingileif Jonsdottir; Alma Moller; Thorolfur Gudnason; Karl G Kristinsson; Unnur Thorsteinsdottir; Kari Stefansson
EPI_ISL_424596  hCoV-19/Iceland/576/2020  Europe / Iceland / Reykjavik  2020-03-28  The National University Hospital of Iceland  deCODE genetics Daniel F Gudbjartsson; Agnar Helgason; Hakon Jonsson; Olafur T Magnusson; Pall Melsted; Gudmundur L Norddahl; Jona Saemundsdottir; Asgeir Sigurdsson; Patrick Sulem; Arna B Agustsdottir; Berglind Eiriksdottir; Run Fridriksdottir; Elisabet E Gardarsdottir; Gudmundur Georgsson; Olafia S Gretarsdottir; Kjartan R Gudmundsson; Thora R Gunnarsdottir; Arnaldur Gylfason; Hilma Holm; Brynjar O Jensson; Aslaug Jonasdottir; Kamilla S Josefsdottir; Thordur Kristjansson; Droplaug N Magnusdottir; Louise le Roux; Gudrun Sigmundsdottir; Gardar Sveinbjornsson; Kristin E Sveinsdottir; Maney Sveinsdottir; Emil A Thorarensen; Bjarni Thorbjornsson; Gisli Masson; Ingileif Jonsdottir; Alma Moller; Thorolfur Gudnason; Karl G Kristinsson; Unnur Thorsteinsdottir; Kari Stefansson
EPI_ISL_424597  hCoV-19/Iceland/577/2020  Europe / Iceland / Reykjavik  2020-03-27  The National University Hospital of Iceland  deCODE genetics Daniel F Gudbjartsson; Agnar Helgason; Hakon Jonsson; Olafur T Magnusson; Pall Melsted; Gudmundur L Norddahl; Jona Saemundsdottir; Asgeir Sigurdsson; Patrick Sulem; Arna B Agustsdottir; Berglind Eiriksdottir; Run Fridriksdottir; Elisabet E Gardarsdottir; Gudmundur Georgsson; Olafia S Gretarsdottir; Kjartan R Gudmundsson; Thora R Gunnarsdottir; Arnaldur Gylfason; Hilma Holm; Brynjar O Jensson; Aslaug Jonasdottir; Kamilla S Josefsdottir; Thordur Kristjansson; Droplaug N Magnusdottir; Louise le Roux; Gudrun Sigmundsdottir; Gardar Sveinbjornsson; Kristin E Sveinsdottir; Maney Sveinsdottir; Emil A Thorarensen; Bjarni Thorbjornsson; Gisli Masson; Ingileif Jonsdottir; Alma Moller; Thorolfur Gudnason; Karl G Kristinsson; Unnur Thorsteinsdottir; Kari Stefansson
EPI_ISL_424598  hCoV-19/Iceland/578/2020  Europe / Iceland / Reykjavik  2020-03-27  The National University Hospital of Iceland  deCODE genetics Daniel F Gudbjartsson; Agnar Helgason; Hakon Jonsson; Olafur T Magnusson; Pall Melsted; Gudmundur L Norddahl; Jona Saemundsdottir; Asgeir Sigurdsson; Patrick Sulem; Arna B Agustsdottir; Berglind Eiriksdottir; Run Fridriksdottir; Elisabet E Gardarsdottir; Gudmundur Georgsson; Olafia S Gretarsdottir; Kjartan R Gudmundsson; Thora R Gunnarsdottir; Arnaldur Gylfason; Hilma Holm; Brynjar O Jensson; Aslaug Jonasdottir; Kamilla S Josefsdottir; Thordur Kristjansson; Droplaug N Magnusdottir; Louise le Roux; Gudrun Sigmundsdottir; Gardar Sveinbjornsson; Kristin E Sveinsdottir; Maney Sveinsdottir; Emil A Thorarensen; Bjarni Thorbjornsson; Gisli Masson; Ingileif Jonsdottir; Alma Moller; Thorolfur Gudnason; Karl G Kristinsson; Unnur Thorsteinsdottir; Kari Stefansson
EPI_ISL_424599  hCoV-19/Iceland/579/2020  Europe / Iceland / Reykjavik  2020-03-27  The National University Hospital of Iceland  deCODE genetics Daniel F Gudbjartsson; Agnar Helgason; Hakon Jonsson; Olafur T Magnusson; Pall Melsted; Gudmundur L Norddahl; Jona Saemundsdottir; Asgeir Sigurdsson; Patrick Sulem; Arna B Agustsdottir; Berglind Eiriksdottir; Run Fridriksdottir; Elisabet E Gardarsdottir; Gudmundur Georgsson; Olafia S Gretarsdottir; Kjartan R Gudmundsson; Thora R Gunnarsdottir; Arnaldur Gylfason; Hilma Holm; Brynjar O Jensson; Aslaug Jonasdottir; Kamilla S Josefsdottir; Thordur Kristjansson; Droplaug N Magnusdottir; Louise le Roux; Gudrun Sigmundsdottir; Gardar Sveinbjornsson; Kristin E Sveinsdottir; Maney Sveinsdottir; Emil A Thorarensen; Bjarni Thorbjornsson; Gisli Masson; Ingileif Jonsdottir; Alma Moller; Thorolfur Gudnason; Karl G

Kristinsson; Unnur Thorsteinsdottir; Kari Stefansson
EPI_ISL_424601 hCoV-19/Iceland/581/2020 Europe / Iceland / Reykjavik 2020-03-27 The National University Hospital of Iceland deCODE genetics Daniel F Gudbjartsson; Agnar Helgason; Hakon Jonsson; Olafur T Magnusson; Pall Melsted; Gudmundur L Norddahl; Jona Saemundsdottir; Asgeir Sigurdsson; Patrick Sulem; Arna B Agustsdottir; Berglind Eiriksdottir; Run Fridriksdottir; Elisabet E Gardarsdottir; Gudmundur Georgsson; Olafia S Gretarsdottir; Kjartan R Gudmundsson; Thora R Gunnarsdottir; Arnaldur Gylfason; Hilma Holm; Brynjar O Jensson; Aslaug Jonasdottir; Kamilla S Josefsdottir; Thordur Kristjansson; Droplaug N Magnusdottir; Louise le Roux; Gudrun Sigmundsdottir; Gardar Sveinbjornsson; Kristin E Sveinsdottir; Maney Sveinsdottir; Emil A Thorarensen; Bjarni Thorbjornsson; Gisli Masson; Ingileif Jonsdottir; Alma Moller; Thorolfur Gudnason; Karl G Kristinsson; Unnur Thorsteinsdottir; Kari Stefansson
EPI_ISL_424602 hCoV-19/Iceland/582/2020 Europe / Iceland / Reykjavik 2020-03-27 The National University Hospital of Iceland deCODE genetics Daniel F Gudbjartsson; Agnar Helgason; Hakon Jonsson; Olafur T Magnusson; Pall Melsted; Gudmundur L Norddahl; Jona Saemundsdottir; Asgeir Sigurdsson; Patrick Sulem; Arna B Agustsdottir; Berglind Eiriksdottir; Run Fridriksdottir; Elisabet E Gardarsdottir; Gudmundur Georgsson; Olafia S Gretarsdottir; Kjartan R Gudmundsson; Thora R Gunnarsdottir; Arnaldur Gylfason; Hilma Holm; Brynjar O Jensson; Aslaug Jonasdottir; Kamilla S Josefsdottir; Thordur Kristjansson; Droplaug N Magnusdottir; Louise le Roux; Gudrun Sigmundsdottir; Gardar Sveinbjornsson; Kristin E Sveinsdottir; Maney Sveinsdottir; Emil A Thorarensen; Bjarni Thorbjornsson; Gisli Masson; Ingileif Jonsdottir; Alma Moller; Thorolfur Gudnason; Karl G Kristinsson; Unnur Thorsteinsdottir; Kari Stefansson
EPI_ISL_424603 hCoV-19/Iceland/583/2020 Europe / Iceland / Reykjavik 2020-03-27 The National University Hospital of Iceland deCODE genetics Daniel F Gudbjartsson; Agnar Helgason; Hakon Jonsson; Olafur T Magnusson; Pall Melsted; Gudmundur L Norddahl; Jona Saemundsdottir; Asgeir Sigurdsson; Patrick Sulem; Arna B Agustsdottir; Berglind Eiriksdottir; Run Fridriksdottir; Elisabet E Gardarsdottir; Gudmundur Georgsson; Olafia S Gretarsdottir; Kjartan R Gudmundsson; Thora R Gunnarsdottir; Arnaldur Gylfason; Hilma Holm; Brynjar O Jensson; Aslaug Jonasdottir; Kamilla S Josefsdottir; Thordur Kristjansson; Droplaug N Magnusdottir; Louise le Roux; Gudrun Sigmundsdottir; Gardar Sveinbjornsson; Kristin E Sveinsdottir; Maney Sveinsdottir; Emil A Thorarensen; Bjarni Thorbjornsson; Gisli Masson; Ingileif Jonsdottir; Alma Moller; Thorolfur Gudnason; Karl G Kristinsson; Unnur Thorsteinsdottir; Kari Stefansson
EPI_ISL_424604 hCoV-19/Iceland/584/2020 Europe / Iceland / Reykjavik 2020-03-27 The National University Hospital of Iceland deCODE genetics Daniel F Gudbjartsson; Agnar Helgason; Hakon Jonsson; Olafur T Magnusson; Pall Melsted; Gudmundur L Norddahl; Jona Saemundsdottir; Asgeir Sigurdsson; Patrick Sulem; Arna B Agustsdottir; Berglind Eiriksdottir; Run Fridriksdottir; Elisabet E Gardarsdottir; Gudmundur Georgsson; Olafia S Gretarsdottir; Kjartan R Gudmundsson; Thora R Gunnarsdottir; Arnaldur Gylfason; Hilma Holm; Brynjar O Jensson; Aslaug Jonasdottir; Kamilla S Josefsdottir; Thordur Kristjansson; Droplaug N Magnusdottir; Louise le Roux; Gudrun Sigmundsdottir; Gardar Sveinbjornsson; Kristin E Sveinsdottir; Maney Sveinsdottir; Emil A Thorarensen; Bjarni Thorbjornsson; Gisli Masson; Ingileif Jonsdottir; Alma Moller; Thorolfur Gudnason; Karl G Kristinsson; Unnur Thorsteinsdottir; Kari Stefansson
EPI_ISL_424605 hCoV-19/Iceland/585/2020 Europe / Iceland / Reykjavik 2020-03-27 The National University Hospital of Iceland deCODE genetics Daniel F Gudbjartsson; Agnar Helgason; Hakon Jonsson; Olafur T Magnusson; Pall Melsted; Gudmundur L Norddahl; Jona Saemundsdottir; Asgeir Sigurdsson; Patrick Sulem; Arna B Agustsdottir; Berglind Eiriksdottir; Run Fridriksdottir; Elisabet E Gardarsdottir; Gudmundur Georgsson; Olafia S Gretarsdottir; Kjartan R Gudmundsson; Thora R Gunnarsdottir; Arnaldur Gylfason; Hilma Holm; Brynjar O Jensson; Aslaug Jonasdottir; Kamilla S Josefsdottir; Thordur Kristjansson; Droplaug N Magnusdottir; Louise le Roux; Gudrun Sigmundsdottir; Gardar Sveinbjornsson; Kristin E Sveinsdottir; Maney Sveinsdottir; Emil A Thorarensen; Bjarni Thorbjornsson; Gisli Masson; Ingileif Jonsdottir; Alma Moller; Thorolfur Gudnason; Karl G Kristinsson; Unnur Thorsteinsdottir; Kari Stefansson
EPI_ISL_424606 hCoV-19/Iceland/586/2020 Europe / Iceland / Reykjavik 2020-03-28 The National University Hospital of Iceland deCODE genetics Daniel F Gudbjartsson; Agnar Helgason; Hakon Jonsson; Olafur T Magnusson; Pall Melsted; Gudmundur L Norddahl; Jona Saemundsdottir; Asgeir Sigurdsson; Patrick Sulem; Arna B Agustsdottir; Berglind Eiriksdottir; Run Fridriksdottir; Elisabet E Gardarsdottir; Gudmundur Georgsson; Olafia S Gretarsdottir; Kjartan R Gudmundsson; Thora R Gunnarsdottir; Arnaldur Gylfason; Hilma Holm; Brynjar O Jensson; Aslaug Jonasdottir; Kamilla S Josefsdottir; Thordur Kristjansson; Droplaug N Magnusdottir; Louise le Roux; Gudrun Sigmundsdottir; Gardar Sveinbjornsson; Kristin E Sveinsdottir; Maney Sveinsdottir; Emil A Thorarensen; Bjarni Thorbjornsson; Gisli Masson; Ingileif Jonsdottir; Alma Moller; Thorolfur Gudnason; Karl G Kristinsson; Unnur Thorsteinsdottir; Kari Stefansson
EPI_ISL_424609 hCoV-19/Iceland/589/2020 Europe / Iceland / Reykjavik 2020-03-29 deCODE genetics deCODE genetics Daniel F Gudbjartsson; Agnar Helgason; Hakon Jonsson; Olafur T Magnusson; Pall Melsted; Gudmundur L Norddahl; Jona Saemundsdottir; Asgeir Sigurdsson; Patrick Sulem; Arna B Agustsdottir; Berglind Eiriksdottir; Run Fridriksdottir; Elisabet E Gardarsdottir; Gudmundur Georgsson; Olafia S Gretarsdottir; Kjartan R Gudmundsson; Thora R Gunnarsdottir; Arnaldur Gylfason; Hilma Holm; Brynjar O Jensson; Aslaug Jonasdottir; Kamilla S Josefsdottir; Thordur Kristjansson; Droplaug N Magnusdottir; Louise le Roux; Gudrun Sigmundsdottir; Gardar Sveinbjornsson; Kristin E Sveinsdottir; Maney Sveinsdottir; Emil A Thorarensen; Bjarni Thorbjornsson; Gisli Masson; Ingileif Jonsdottir; Alma Moller; Thorolfur Gudnason; Karl G Kristinsson; Unnur Thorsteinsdottir; Kari Stefansson
EPI_ISL_424610 hCoV-19/Iceland/590/2020 Europe / Iceland / Reykjavik 2020-03-26 The National University Hospital of Iceland deCODE genetics Daniel F Gudbjartsson; Agnar Helgason; Hakon Jonsson; Olafur T Magnusson; Pall Melsted; Gudmundur L Norddahl; Jona Saemundsdottir; Asgeir Sigurdsson; Patrick Sulem; Arna B Agustsdottir; Berglind Eiriksdottir; Run Fridriksdottir; Elisabet E Gardarsdottir; Gudmundur Georgsson; Olafia S Gretarsdottir; Kjartan R Gudmundsson; Thora R Gunnarsdottir; Arnaldur Gylfason; Hilma Holm; Brynjar O Jensson; Aslaug Jonasdottir; Kamilla S Josefsdottir; Thordur Kristjansson; Droplaug N Magnusdottir; Louise le

Roux; Gudrun Sigmundsdottir; Gardar Sveinbjornsson; Kristin E Sveinsdottir; Maney Sveinsdottir; Emil A Thorarensen; Bjarni Thorbjornsson; Gisli Masson; Ingileif Jonsdottir; Alma Moller; Thorolfur Gudnason; Karl G Kristinsson; Unnur Thorsteinsdottir; Kari Stefansson
EPI_ISL_424612 hCoV-19/Iceland/592/2020 Europe / Iceland / Reykjavik 2020-03-27 The National University Hospital of Iceland deCODE genetics Daniel F Gudbjartsson; Agnar Helgason; Hakon Jonsson; Olafur T Magnusson; Pall Melsted; Gudmundur L Norddahl; Jona Saemundsdottir; Asgeir Sigurdsson; Patrick Sulem; Arna B Agustsdottir; Berglind Eiriksdottir; Run Fridriksdottir; Elisabet E Gardarsdottir; Gudmundur Georgsson; Olafia S Gretarsdottir; Kjartan R Gudmundsson; Thora R Gunnarsdottir; Arnaldur Gylfason; Hilma Holm; Brynjar O Jensson; Aslaug Jonasdottir; Kamilla S Josefsdottir; Thordur Kristjansson; Droplaug N Magnusdottir; Louise le Roux; Gudrun Sigmundsdottir; Gardar Sveinbjornsson; Kristin E Sveinsdottir; Maney Sveinsdottir; Emil A Thorarensen; Bjarni Thorbjornsson; Gisli Masson; Ingileif Jonsdottir; Alma Moller; Thorolfur Gudnason; Karl G Kristinsson; Unnur Thorsteinsdottir; Kari Stefansson
EPI_ISL_425702 hCoV-19/Scotland/CVR163/2020 Europe / United Kingdom / Scotland 2020-03-17 West of Scotland Specialist Virology Centre, NHSGGC / MRC-University of Glasgow Centre for Virus Research COVID-19 Genomics UK (COG-UK) Consortium Ana da Silva Filipe, Kathy Smollett, Stephen Carmichael, Natasha Johnson, Daniel Mair, Lily Tong, Jenna Nichols; Sarah McDonald; Richard Orton, Joseph Hughes, Sreenu Vattipally, David L Robertson; Kathy Li, Natasha Jesudason, Rajiv Shah, James Shepherd, Antonia Ho, Emma Thomson; Alasdair MacLean, Rory Gunson.
EPI_ISL_425703 hCoV-19/Scotland/CVR164/2020 Europe / United Kingdom / Scotland 2020-03-17 West of Scotland Specialist Virology Centre, NHSGGC / MRC-University of Glasgow Centre for Virus Research COVID-19 Genomics UK (COG-UK) Consortium Ana da Silva Filipe, Kathy Smollett, Stephen Carmichael, Natasha Johnson, Daniel Mair, Lily Tong, Jenna Nichols; Sarah McDonald; Richard Orton, Joseph Hughes, Sreenu Vattipally, David L Robertson; Kathy Li, Natasha Jesudason, Rajiv Shah, James Shepherd, Antonia Ho, Emma Thomson; Alasdair MacLean, Rory Gunson.
EPI_ISL_425704 hCoV-19/Scotland/CVR165/2020 Europe / United Kingdom / Scotland 2020-03-17 West of Scotland Specialist Virology Centre, NHSGGC / MRC-University of Glasgow Centre for Virus Research COVID-19 Genomics UK (COG-UK) Consortium Ana da Silva Filipe, Kathy Smollett, Stephen Carmichael, Natasha Johnson, Daniel Mair, Lily Tong, Jenna Nichols; Sarah McDonald; Richard Orton, Joseph Hughes, Sreenu Vattipally, David L Robertson; Kathy Li, Natasha Jesudason, Rajiv Shah, James Shepherd, Antonia Ho, Emma Thomson; Alasdair MacLean, Rory Gunson.
EPI_ISL_425705 hCoV-19/Scotland/CVR166/2020 Europe / United Kingdom / Scotland 2020-03-17 West of Scotland Specialist Virology Centre, NHSGGC / MRC-University of Glasgow Centre for Virus Research COVID-19 Genomics UK (COG-UK) Consortium Ana da Silva Filipe, Kathy Smollett, Stephen Carmichael, Natasha Johnson, Daniel Mair, Lily Tong, Jenna Nichols; Sarah McDonald; Richard Orton, Joseph Hughes, Sreenu Vattipally, David L Robertson; Kathy Li, Natasha Jesudason, Rajiv Shah, James Shepherd, Antonia Ho, Emma Thomson; Alasdair MacLean, Rory Gunson.
EPI_ISL_425707 hCoV-19/Scotland/CVR168/2020 Europe / United Kingdom / Scotland 2020-03-17 West of Scotland Specialist Virology Centre, NHSGGC / MRC-University of Glasgow Centre for Virus Research COVID-19 Genomics UK (COG-UK) Consortium Ana da Silva Filipe, Kathy Smollett, Stephen Carmichael, Natasha Johnson, Daniel Mair, Lily Tong, Jenna Nichols; Sarah McDonald; Richard Orton, Joseph Hughes, Sreenu Vattipally, David L Robertson; Kathy Li, Natasha Jesudason, Rajiv Shah, James Shepherd, Antonia Ho, Emma Thomson; Alasdair MacLean, Rory Gunson.
EPI_ISL_425708 hCoV-19/Scotland/CVR169/2020 Europe / United Kingdom / Scotland 2020-03-17 West of Scotland Specialist Virology Centre, NHSGGC / MRC-University of Glasgow Centre for Virus Research COVID-19 Genomics UK (COG-UK) Consortium Ana da Silva Filipe, Kathy Smollett, Stephen Carmichael, Natasha Johnson, Daniel Mair, Lily Tong, Jenna Nichols; Sarah McDonald; Richard Orton, Joseph Hughes, Sreenu Vattipally, David L Robertson; Kathy Li, Natasha Jesudason, Rajiv Shah, James Shepherd, Antonia Ho, Emma Thomson; Alasdair MacLean, Rory Gunson.
EPI_ISL_425709 hCoV-19/Scotland/CVR17/2020 Europe / United Kingdom / Scotland 2020-03-09 West of Scotland Specialist Virology Centre, NHSGGC / MRC-University of Glasgow Centre for Virus Research COVID-19 Genomics UK (COG-UK) Consortium Ana da Silva Filipe, Kathy Smollett, Stephen Carmichael, Natasha Johnson, Daniel Mair, Lily Tong, Jenna Nichols; Sarah McDonald; Richard Orton, Joseph Hughes, Sreenu Vattipally, David L Robertson; Kathy Li, Natasha Jesudason, Rajiv Shah, James Shepherd, Antonia Ho, Emma Thomson; Alasdair MacLean, Rory Gunson.
EPI_ISL_425711 hCoV-19/Scotland/CVR171/2020 Europe / United Kingdom / Scotland 2020-03-17 West of Scotland Specialist Virology Centre, NHSGGC / MRC-University of Glasgow Centre for Virus Research COVID-19 Genomics UK (COG-UK) Consortium Ana da Silva Filipe, Kathy Smollett, Stephen Carmichael, Natasha Johnson, Daniel Mair, Lily Tong, Jenna Nichols; Sarah McDonald; Richard Orton, Joseph Hughes, Sreenu Vattipally, David L Robertson; Kathy Li, Natasha Jesudason, Rajiv Shah, James Shepherd, Antonia Ho, Emma Thomson; Alasdair MacLean, Rory Gunson.
EPI_ISL_425713 hCoV-19/Scotland/CVR173/2020 Europe / United Kingdom / Scotland 2020-03-17 West of Scotland Specialist Virology Centre, NHSGGC / MRC-University of Glasgow Centre for Virus Research COVID-19 Genomics UK (COG-UK) Consortium Ana da Silva Filipe, Kathy Smollett, Stephen Carmichael, Natasha Johnson, Daniel Mair, Lily Tong, Jenna Nichols; Sarah McDonald; Richard Orton, Joseph Hughes, Sreenu Vattipally, David L Robertson; Kathy Li, Natasha Jesudason, Rajiv Shah, James Shepherd, Antonia Ho, Emma Thomson; Alasdair MacLean, Rory Gunson.
EPI_ISL_425715 hCoV-19/Scotland/CVR175/2020 Europe / United Kingdom / Scotland 2020-03-17 West of Scotland Specialist Virology Centre, NHSGGC / MRC-University of Glasgow Centre for Virus Research COVID-19 Genomics UK (COG-UK) Consortium Ana da Silva Filipe, Kathy Smollett, Stephen Carmichael, Natasha Johnson, Daniel Mair, Lily Tong, Jenna Nichols; Sarah McDonald; Richard Orton, Joseph Hughes, Sreenu

Vattipally, David L Robertson; Kathy Li, Natasha Jesudason, Rajiv Shah, James Shepherd, Antonia Ho, Emma Thomson; Alasdair MacLean, Rory Gunson.
EPI_ISL_425716 hCoV-19/Scotland/CVR176/2020 Europe / United Kingdom / Scotland 2020-03-17 West of Scotland Specialist Virology Centre, NHSGGC / MRC-University of Glasgow Centre for Virus Research COVID-19 Genomics UK (COG-UK) Consortium Ana da Silva Filipe, Kathy Smollett, Stephen Carmichael, Natasha Johnson, Daniel Mair, Lily Tong, Jenna Nichols; Sarah McDonald; Richard Orton, Joseph Hughes, Sreenu Vattipally, David L Robertson; Kathy Li, Natasha Jesudason, Rajiv Shah, James Shepherd, Antonia Ho, Emma Thomson; Alasdair MacLean, Rory Gunson.
EPI_ISL_425717 hCoV-19/Scotland/CVR177/2020 Europe / United Kingdom / Scotland 2020-03-17 West of Scotland Specialist Virology Centre, NHSGGC / MRC-University of Glasgow Centre for Virus Research COVID-19 Genomics UK (COG-UK) Consortium Ana da Silva Filipe, Kathy Smollett, Stephen Carmichael, Natasha Johnson, Daniel Mair, Lily Tong, Jenna Nichols; Sarah McDonald; Richard Orton, Joseph Hughes, Sreenu Vattipally, David L Robertson; Kathy Li, Natasha Jesudason, Rajiv Shah, James Shepherd, Antonia Ho, Emma Thomson; Alasdair MacLean, Rory Gunson.
EPI_ISL_425720 hCoV-19/Scotland/CVR18/2020 Europe / United Kingdom / Scotland 2020-03-12 West of Scotland Specialist Virology Centre, NHSGGC / MRC-University of Glasgow Centre for Virus Research COVID-19 Genomics UK (COG-UK) Consortium Ana da Silva Filipe, Kathy Smollett, Stephen Carmichael, Natasha Johnson, Daniel Mair, Lily Tong, Jenna Nichols; Sarah McDonald; Richard Orton, Joseph Hughes, Sreenu Vattipally, David L Robertson; Kathy Li, Natasha Jesudason, Rajiv Shah, James Shepherd, Antonia Ho, Emma Thomson; Alasdair MacLean, Rory Gunson.
EPI_ISL_425721 hCoV-19/Scotland/CVR180/2020 Europe / United Kingdom / Scotland 2020-03-17 West of Scotland Specialist Virology Centre, NHSGGC / MRC-University of Glasgow Centre for Virus Research COVID-19 Genomics UK (COG-UK) Consortium Ana da Silva Filipe, Kathy Smollett, Stephen Carmichael, Natasha Johnson, Daniel Mair, Lily Tong, Jenna Nichols; Sarah McDonald; Richard Orton, Joseph Hughes, Sreenu Vattipally, David L Robertson; Kathy Li, Natasha Jesudason, Rajiv Shah, James Shepherd, Antonia Ho, Emma Thomson; Alasdair MacLean, Rory Gunson.
EPI_ISL_425722 hCoV-19/Scotland/CVR183/2020 Europe / United Kingdom / Scotland 2020-03-18 West of Scotland Specialist Virology Centre, NHSGGC / MRC-University of Glasgow Centre for Virus Research COVID-19 Genomics UK (COG-UK) Consortium Ana da Silva Filipe, Kathy Smollett, Stephen Carmichael, Natasha Johnson, Daniel Mair, Lily Tong, Jenna Nichols; Sarah McDonald; Richard Orton, Joseph Hughes, Sreenu Vattipally, David L Robertson; Kathy Li, Natasha Jesudason, Rajiv Shah, James Shepherd, Antonia Ho, Emma Thomson; Alasdair MacLean, Rory Gunson.
EPI_ISL_425724 hCoV-19/Scotland/CVR19/2020 Europe / United Kingdom / Scotland 2020-03-12 West of Scotland Specialist Virology Centre, NHSGGC / MRC-University of Glasgow Centre for Virus Research COVID-19 Genomics UK (COG-UK) Consortium Ana da Silva Filipe, Kathy Smollett, Stephen Carmichael, Natasha Johnson, Daniel Mair, Lily Tong, Jenna Nichols; Sarah McDonald; Richard Orton, Joseph Hughes, Sreenu Vattipally, David L Robertson; Kathy Li, Natasha Jesudason, Rajiv Shah, James Shepherd, Antonia Ho, Emma Thomson; Alasdair MacLean, Rory Gunson.
EPI_ISL_425725 hCoV-19/Scotland/CVR190/2020 Europe / United Kingdom / Scotland 2020-03-18 West of Scotland Specialist Virology Centre, NHSGGC / MRC-University of Glasgow Centre for Virus Research COVID-19 Genomics UK (COG-UK) Consortium Ana da Silva Filipe, Kathy Smollett, Stephen Carmichael, Natasha Johnson, Daniel Mair, Lily Tong, Jenna Nichols; Sarah McDonald; Richard Orton, Joseph Hughes, Sreenu Vattipally, David L Robertson; Kathy Li, Natasha Jesudason, Rajiv Shah, James Shepherd, Antonia Ho, Emma Thomson; Alasdair MacLean, Rory Gunson.
EPI_ISL_425726 hCoV-19/Scotland/CVR191/2020 Europe / United Kingdom / Scotland 2020-03-18 West of Scotland Specialist Virology Centre, NHSGGC / MRC-University of Glasgow Centre for Virus Research COVID-19 Genomics UK (COG-UK) Consortium Ana da Silva Filipe, Kathy Smollett, Stephen Carmichael, Natasha Johnson, Daniel Mair, Lily Tong, Jenna Nichols; Sarah McDonald; Richard Orton, Joseph Hughes, Sreenu Vattipally, David L Robertson; Kathy Li, Natasha Jesudason, Rajiv Shah, James Shepherd, Antonia Ho, Emma Thomson; Alasdair MacLean, Rory Gunson.
EPI_ISL_425727 hCoV-19/Scotland/CVR195/2020 Europe / United Kingdom / Scotland 2020-03-18 West of Scotland Specialist Virology Centre, NHSGGC / MRC-University of Glasgow Centre for Virus Research COVID-19 Genomics UK (COG-UK) Consortium Ana da Silva Filipe, Kathy Smollett, Stephen Carmichael, Natasha Johnson, Daniel Mair, Lily Tong, Jenna Nichols; Sarah McDonald; Richard Orton, Joseph Hughes, Sreenu Vattipally, David L Robertson; Kathy Li, Natasha Jesudason, Rajiv Shah, James Shepherd, Antonia Ho, Emma Thomson; Alasdair MacLean, Rory Gunson.
EPI_ISL_425728 hCoV-19/Scotland/CVR196/2020 Europe / United Kingdom / Scotland 2020-03-19 West of Scotland Specialist Virology Centre, NHSGGC / MRC-University of Glasgow Centre for Virus Research COVID-19 Genomics UK (COG-UK) Consortium Ana da Silva Filipe, Kathy Smollett, Stephen Carmichael, Natasha Johnson, Daniel Mair, Lily Tong, Jenna Nichols; Sarah McDonald; Richard Orton, Joseph Hughes, Sreenu Vattipally, David L Robertson; Kathy Li, Natasha Jesudason, Rajiv Shah, James Shepherd, Antonia Ho, Emma Thomson; Alasdair MacLean, Rory Gunson.
EPI_ISL_425729 hCoV-19/Scotland/CVR199/2020 Europe / United Kingdom / Scotland 2020-03-18 West of Scotland Specialist Virology Centre, NHSGGC / MRC-University of Glasgow Centre for Virus Research COVID-19 Genomics UK (COG-UK) Consortium Ana da Silva Filipe, Kathy Smollett, Stephen Carmichael, Natasha Johnson, Daniel Mair, Lily Tong, Jenna Nichols; Sarah McDonald; Richard Orton, Joseph Hughes, Sreenu Vattipally, David L Robertson; Kathy Li, Natasha Jesudason, Rajiv Shah, James Shepherd, Antonia Ho, Emma Thomson; Alasdair MacLean, Rory Gunson.
EPI_ISL_425730 hCoV-19/Scotland/CVR20/2020 Europe / United Kingdom / Scotland 2020-03-13 West of Scotland Specialist Virology Centre, NHSGGC / MRC-University of Glasgow Centre for Virus Research COVID-

19 Genomics UK (COG-UK) Consortium	Ana da Silva Filipe, Kathy Smollett, Stephen Carmichael, Natasha Johnson, Daniel Mair, Lily Tong, Jenna Nichols; Sarah McDonald; Richard Orton, Joseph Hughes, Sreenu Vattipally, David L Robertson; Kathy Li, Natasha Jesudason, Rajiv Shah, James Shepherd, Antonia Ho, Emma Thomson; Alasdair MacLean, Rory Gunson.
EPI_ISL_425731	hCoV-19/Scotland/CVR200/2020	Europe / United Kingdom / Scotland	2020-03-17	West of Scotland Specialist Virology Centre, NHSGGC / MRC-University of Glasgow Centre for Virus Research	COVID-19 Genomics UK (COG-UK) Consortium	Ana da Silva Filipe, Kathy Smollett, Stephen Carmichael, Natasha Johnson, Daniel Mair, Lily Tong, Jenna Nichols; Sarah McDonald; Richard Orton, Joseph Hughes, Sreenu Vattipally, David L Robertson; Kathy Li, Natasha Jesudason, Rajiv Shah, James Shepherd, Antonia Ho, Emma Thomson; Alasdair MacLean, Rory Gunson.
EPI_ISL_425732	hCoV-19/Scotland/CVR201/2020	Europe / United Kingdom / Scotland	2020-03-18	West of Scotland Specialist Virology Centre, NHSGGC / MRC-University of Glasgow Centre for Virus Research	COVID-19 Genomics UK (COG-UK) Consortium	Ana da Silva Filipe, Kathy Smollett, Stephen Carmichael, Natasha Johnson, Daniel Mair, Lily Tong, Jenna Nichols; Sarah McDonald; Richard Orton, Joseph Hughes, Sreenu Vattipally, David L Robertson; Kathy Li, Natasha Jesudason, Rajiv Shah, James Shepherd, Antonia Ho, Emma Thomson; Alasdair MacLean, Rory Gunson.
EPI_ISL_425733	hCoV-19/Scotland/CVR202/2020	Europe / United Kingdom / Scotland	2020-03-19	West of Scotland Specialist Virology Centre, NHSGGC / MRC-University of Glasgow Centre for Virus Research	COVID-19 Genomics UK (COG-UK) Consortium	Ana da Silva Filipe, Kathy Smollett, Stephen Carmichael, Natasha Johnson, Daniel Mair, Lily Tong, Jenna Nichols; Sarah McDonald; Richard Orton, Joseph Hughes, Sreenu Vattipally, David L Robertson; Kathy Li, Natasha Jesudason, Rajiv Shah, James Shepherd, Antonia Ho, Emma Thomson; Alasdair MacLean, Rory Gunson.
EPI_ISL_425734	hCoV-19/Scotland/CVR203/2020	Europe / United Kingdom / Scotland	2020-03-18	West of Scotland Specialist Virology Centre, NHSGGC / MRC-University of Glasgow Centre for Virus Research	COVID-19 Genomics UK (COG-UK) Consortium	Ana da Silva Filipe, Kathy Smollett, Stephen Carmichael, Natasha Johnson, Daniel Mair, Lily Tong, Jenna Nichols; Sarah McDonald; Richard Orton, Joseph Hughes, Sreenu Vattipally, David L Robertson; Kathy Li, Natasha Jesudason, Rajiv Shah, James Shepherd, Antonia Ho, Emma Thomson; Alasdair MacLean, Rory Gunson.
EPI_ISL_425735	hCoV-19/Scotland/CVR205/2020	Europe / United Kingdom / Scotland	2020-03-19	West of Scotland Specialist Virology Centre, NHSGGC / MRC-University of Glasgow Centre for Virus Research	COVID-19 Genomics UK (COG-UK) Consortium	Ana da Silva Filipe, Kathy Smollett, Stephen Carmichael, Natasha Johnson, Daniel Mair, Lily Tong, Jenna Nichols; Sarah McDonald; Richard Orton, Joseph Hughes, Sreenu Vattipally, David L Robertson; Kathy Li, Natasha Jesudason, Rajiv Shah, James Shepherd, Antonia Ho, Emma Thomson; Alasdair MacLean, Rory Gunson.
EPI_ISL_425736	hCoV-19/Scotland/CVR206/2020	Europe / United Kingdom / Scotland	2020-03-16	West of Scotland Specialist Virology Centre, NHSGGC / MRC-University of Glasgow Centre for Virus Research	COVID-19 Genomics UK (COG-UK) Consortium	Ana da Silva Filipe, Kathy Smollett, Stephen Carmichael, Natasha Johnson, Daniel Mair, Lily Tong, Jenna Nichols; Sarah McDonald; Richard Orton, Joseph Hughes, Sreenu Vattipally, David L Robertson; Kathy Li, Natasha Jesudason, Rajiv Shah, James Shepherd, Antonia Ho, Emma Thomson; Alasdair MacLean, Rory Gunson.
EPI_ISL_425737	hCoV-19/Scotland/CVR207/2020	Europe / United Kingdom / Scotland	2020-03-18	West of Scotland Specialist Virology Centre, NHSGGC / MRC-University of Glasgow Centre for Virus Research	COVID-19 Genomics UK (COG-UK) Consortium	Ana da Silva Filipe, Kathy Smollett, Stephen Carmichael, Natasha Johnson, Daniel Mair, Lily Tong, Jenna Nichols; Sarah McDonald; Richard Orton, Joseph Hughes, Sreenu Vattipally, David L Robertson; Kathy Li, Natasha Jesudason, Rajiv Shah, James Shepherd, Antonia Ho, Emma Thomson; Alasdair MacLean, Rory Gunson.
EPI_ISL_425739	hCoV-19/Scotland/CVR21/2020	Europe / United Kingdom / Scotland	2020-03-13	West of Scotland Specialist Virology Centre, NHSGGC / MRC-University of Glasgow Centre for Virus Research	COVID-19 Genomics UK (COG-UK) Consortium	Ana da Silva Filipe, Kathy Smollett, Stephen Carmichael, Natasha Johnson, Daniel Mair, Lily Tong, Jenna Nichols; Sarah McDonald; Richard Orton, Joseph Hughes, Sreenu Vattipally, David L Robertson; Kathy Li, Natasha Jesudason, Rajiv Shah, James Shepherd, Antonia Ho, Emma Thomson; Alasdair MacLean, Rory Gunson.
EPI_ISL_425741	hCoV-19/Scotland/CVR212/2020	Europe / United Kingdom / Scotland	2020-03-18	West of Scotland Specialist Virology Centre, NHSGGC / MRC-University of Glasgow Centre for Virus Research	COVID-19 Genomics UK (COG-UK) Consortium	Ana da Silva Filipe, Kathy Smollett, Stephen Carmichael, Natasha Johnson, Daniel Mair, Lily Tong, Jenna Nichols; Sarah McDonald; Richard Orton, Joseph Hughes, Sreenu Vattipally, David L Robertson; Kathy Li, Natasha Jesudason, Rajiv Shah, James Shepherd, Antonia Ho, Emma Thomson; Alasdair MacLean, Rory Gunson.
EPI_ISL_425742	hCoV-19/Scotland/CVR213/2020	Europe / United Kingdom / Scotland	2020-03-19	West of Scotland Specialist Virology Centre, NHSGGC / MRC-University of Glasgow Centre for Virus Research	COVID-19 Genomics UK (COG-UK) Consortium	Ana da Silva Filipe, Kathy Smollett, Stephen Carmichael, Natasha Johnson, Daniel Mair, Lily Tong, Jenna Nichols; Sarah McDonald; Richard Orton, Joseph Hughes, Sreenu Vattipally, David L Robertson; Kathy Li, Natasha Jesudason, Rajiv Shah, James Shepherd, Antonia Ho, Emma Thomson; Alasdair MacLean, Rory Gunson.
EPI_ISL_425743	hCoV-19/Scotland/CVR215/2020	Europe / United Kingdom / Scotland	2020-03-18	West of Scotland Specialist Virology Centre, NHSGGC / MRC-University of Glasgow Centre for Virus Research	COVID-19 Genomics UK (COG-UK) Consortium	Ana da Silva Filipe, Kathy Smollett, Stephen Carmichael, Natasha Johnson, Daniel Mair, Lily Tong, Jenna Nichols; Sarah McDonald; Richard Orton, Joseph Hughes, Sreenu Vattipally, David L Robertson; Kathy Li, Natasha Jesudason, Rajiv Shah, James Shepherd, Antonia Ho, Emma Thomson; Alasdair MacLean, Rory Gunson.

EPI_ISL_425744 hCoV-19/Scotland/CVR216/2020 Europe / United Kingdom / Scotland 2020-03-19 West of Scotland Specialist Virology Centre, NHSGGC / MRC-University of Glasgow Centre for Virus Research COVID-19 Genomics UK (COG-UK) Consortium Ana da Silva Filipe, Kathy Smollett, Stephen Carmichael, Natasha Johnson, Daniel Mair, Lily Tong, Jenna Nichols; Sarah McDonald; Richard Orton, Joseph Hughes, Sreenu Vattipally, David L Robertson; Kathy Li, Natasha Jesudason, Rajiv Shah, James Shepherd, Antonia Ho, Emma Thomson; Alasdair MacLean, Rory Gunson.
EPI_ISL_425745 hCoV-19/Scotland/CVR218/2020 Europe / United Kingdom / Scotland 2020-03-19 West of Scotland Specialist Virology Centre, NHSGGC / MRC-University of Glasgow Centre for Virus Research COVID-19 Genomics UK (COG-UK) Consortium Ana da Silva Filipe, Kathy Smollett, Stephen Carmichael, Natasha Johnson, Daniel Mair, Lily Tong, Jenna Nichols; Sarah McDonald; Richard Orton, Joseph Hughes, Sreenu Vattipally, David L Robertson; Kathy Li, Natasha Jesudason, Rajiv Shah, James Shepherd, Antonia Ho, Emma Thomson; Alasdair MacLean, Rory Gunson.
EPI_ISL_425746 hCoV-19/Scotland/CVR219/2020 Europe / United Kingdom / Scotland 2020-03-19 West of Scotland Specialist Virology Centre, NHSGGC / MRC-University of Glasgow Centre for Virus Research COVID-19 Genomics UK (COG-UK) Consortium Ana da Silva Filipe, Kathy Smollett, Stephen Carmichael, Natasha Johnson, Daniel Mair, Lily Tong, Jenna Nichols; Sarah McDonald; Richard Orton, Joseph Hughes, Sreenu Vattipally, David L Robertson; Kathy Li, Natasha Jesudason, Rajiv Shah, James Shepherd, Antonia Ho, Emma Thomson; Alasdair MacLean, Rory Gunson.
EPI_ISL_425747 hCoV-19/Scotland/CVR220/2020 Europe / United Kingdom / Scotland 2020-03-18 West of Scotland Specialist Virology Centre, NHSGGC / MRC-University of Glasgow Centre for Virus Research COVID-19 Genomics UK (COG-UK) Consortium Ana da Silva Filipe, Kathy Smollett, Stephen Carmichael, Natasha Johnson, Daniel Mair, Lily Tong, Jenna Nichols; Sarah McDonald; Richard Orton, Joseph Hughes, Sreenu Vattipally, David L Robertson; Kathy Li, Natasha Jesudason, Rajiv Shah, James Shepherd, Antonia Ho, Emma Thomson; Alasdair MacLean, Rory Gunson.
EPI_ISL_425749 hCoV-19/Scotland/CVR23/2020 Europe / United Kingdom / Scotland 2020-03-12 West of Scotland Specialist Virology Centre, NHSGGC / MRC-University of Glasgow Centre for Virus Research COVID-19 Genomics UK (COG-UK) Consortium Ana da Silva Filipe, Kathy Smollett, Stephen Carmichael, Natasha Johnson, Daniel Mair, Lily Tong, Jenna Nichols; Sarah McDonald; Richard Orton, Joseph Hughes, Sreenu Vattipally, David L Robertson; Kathy Li, Natasha Jesudason, Rajiv Shah, James Shepherd, Antonia Ho, Emma Thomson; Alasdair MacLean, Rory Gunson.
EPI_ISL_425750 hCoV-19/Scotland/CVR235/2020 Europe / United Kingdom / Scotland 2020-03-19 West of Scotland Specialist Virology Centre, NHSGGC / MRC-University of Glasgow Centre for Virus Research COVID-19 Genomics UK (COG-UK) Consortium Ana da Silva Filipe, Kathy Smollett, Stephen Carmichael, Natasha Johnson, Daniel Mair, Lily Tong, Jenna Nichols; Sarah McDonald; Richard Orton, Joseph Hughes, Sreenu Vattipally, David L Robertson; Kathy Li, Natasha Jesudason, Rajiv Shah, James Shepherd, Antonia Ho, Emma Thomson; Alasdair MacLean, Rory Gunson.
EPI_ISL_425751 hCoV-19/Scotland/CVR236/2020 Europe / United Kingdom / Scotland 2020-03-19 West of Scotland Specialist Virology Centre, NHSGGC / MRC-University of Glasgow Centre for Virus Research COVID-19 Genomics UK (COG-UK) Consortium Ana da Silva Filipe, Kathy Smollett, Stephen Carmichael, Natasha Johnson, Daniel Mair, Lily Tong, Jenna Nichols; Sarah McDonald; Richard Orton, Joseph Hughes, Sreenu Vattipally, David L Robertson; Kathy Li, Natasha Jesudason, Rajiv Shah, James Shepherd, Antonia Ho, Emma Thomson; Alasdair MacLean, Rory Gunson.
EPI_ISL_425752 hCoV-19/Scotland/CVR24/2020 Europe / United Kingdom / Scotland 2020-03-12 West of Scotland Specialist Virology Centre, NHSGGC / MRC-University of Glasgow Centre for Virus Research COVID-19 Genomics UK (COG-UK) Consortium Ana da Silva Filipe, Kathy Smollett, Stephen Carmichael, Natasha Johnson, Daniel Mair, Lily Tong, Jenna Nichols; Sarah McDonald; Richard Orton, Joseph Hughes, Sreenu Vattipally, David L Robertson; Kathy Li, Natasha Jesudason, Rajiv Shah, James Shepherd, Antonia Ho, Emma Thomson; Alasdair MacLean, Rory Gunson.
EPI_ISL_425754 hCoV-19/Scotland/CVR264/2020 Europe / United Kingdom / Scotland 2020-03-20 West of Scotland Specialist Virology Centre, NHSGGC / MRC-University of Glasgow Centre for Virus Research COVID-19 Genomics UK (COG-UK) Consortium Ana da Silva Filipe, Kathy Smollett, Stephen Carmichael, Natasha Johnson, Daniel Mair, Lily Tong, Jenna Nichols; Sarah McDonald; Richard Orton, Joseph Hughes, Sreenu Vattipally, David L Robertson; Kathy Li, Natasha Jesudason, Rajiv Shah, James Shepherd, Antonia Ho, Emma Thomson; Alasdair MacLean, Rory Gunson.
EPI_ISL_425755 hCoV-19/Scotland/CVR268/2020 Europe / United Kingdom / Scotland 2020-03-19 West of Scotland Specialist Virology Centre, NHSGGC / MRC-University of Glasgow Centre for Virus Research COVID-19 Genomics UK (COG-UK) Consortium Ana da Silva Filipe, Kathy Smollett, Stephen Carmichael, Natasha Johnson, Daniel Mair, Lily Tong, Jenna Nichols; Sarah McDonald; Richard Orton, Joseph Hughes, Sreenu Vattipally, David L Robertson; Kathy Li, Natasha Jesudason, Rajiv Shah, James Shepherd, Antonia Ho, Emma Thomson; Alasdair MacLean, Rory Gunson.
EPI_ISL_425756 hCoV-19/Scotland/CVR27/2020 Europe / United Kingdom / Scotland 2020-03-12 West of Scotland Specialist Virology Centre, NHSGGC / MRC-University of Glasgow Centre for Virus Research COVID-19 Genomics UK (COG-UK) Consortium Ana da Silva Filipe, Kathy Smollett, Stephen Carmichael, Natasha Johnson, Daniel Mair, Lily Tong, Jenna Nichols; Sarah McDonald; Richard Orton, Joseph Hughes, Sreenu Vattipally, David L Robertson; Kathy Li, Natasha Jesudason, Rajiv Shah, James Shepherd, Antonia Ho, Emma Thomson; Alasdair MacLean, Rory Gunson.
EPI_ISL_425757 hCoV-19/Scotland/CVR28/2020 Europe / United Kingdom / Scotland 2020-03-12 West of Scotland Specialist Virology Centre, NHSGGC / MRC-University of Glasgow Centre for Virus Research COVID-19 Genomics UK (COG-UK) Consortium Ana da Silva Filipe, Kathy Smollett, Stephen Carmichael, Natasha Johnson, Daniel Mair, Lily Tong, Jenna Nichols; Sarah McDonald; Richard Orton, Joseph Hughes, Sreenu

Vattipally, David L Robertson; Kathy Li, Natasha Jesudason, Rajiv Shah, James Shepherd, Antonia Ho, Emma Thomson; Alasdair MacLean, Rory Gunson.
EPI_ISL_425758 hCoV-19/Scotland/CVR29/2020 Europe / United Kingdom / Scotland 2020-03-11 West of Scotland Specialist Virology Centre, NHSGGC / MRC-University of Glasgow Centre for Virus Research COVID-19 Genomics UK (COG-UK) Consortium Ana da Silva Filipe, Kathy Smollett, Stephen Carmichael, Natasha Johnson, Daniel Mair, Lily Tong, Jenna Nichols; Sarah McDonald; Richard Orton, Joseph Hughes, Sreenu Vattipally, David L Robertson; Kathy Li, Natasha Jesudason, Rajiv Shah, James Shepherd, Antonia Ho, Emma Thomson; Alasdair MacLean, Rory Gunson.
EPI_ISL_425759 hCoV-19/Scotland/CVR30/2020 Europe / United Kingdom / Scotland 2020-03-13 West of Scotland Specialist Virology Centre, NHSGGC / MRC-University of Glasgow Centre for Virus Research COVID-19 Genomics UK (COG-UK) Consortium Ana da Silva Filipe, Kathy Smollett, Stephen Carmichael, Natasha Johnson, Daniel Mair, Lily Tong, Jenna Nichols; Sarah McDonald; Richard Orton, Joseph Hughes, Sreenu Vattipally, David L Robertson; Kathy Li, Natasha Jesudason, Rajiv Shah, James Shepherd, Antonia Ho, Emma Thomson; Alasdair MacLean, Rory Gunson.
EPI_ISL_425760 hCoV-19/Scotland/CVR31/2020 Europe / United Kingdom / Scotland 2020-03-13 West of Scotland Specialist Virology Centre, NHSGGC / MRC-University of Glasgow Centre for Virus Research COVID-19 Genomics UK (COG-UK) Consortium Ana da Silva Filipe, Kathy Smollett, Stephen Carmichael, Natasha Johnson, Daniel Mair, Lily Tong, Jenna Nichols; Sarah McDonald; Richard Orton, Joseph Hughes, Sreenu Vattipally, David L Robertson; Kathy Li, Natasha Jesudason, Rajiv Shah, James Shepherd, Antonia Ho, Emma Thomson; Alasdair MacLean, Rory Gunson.
EPI_ISL_425761 hCoV-19/Scotland/CVR32/2020 Europe / United Kingdom / Scotland 2020-03-13 West of Scotland Specialist Virology Centre, NHSGGC / MRC-University of Glasgow Centre for Virus Research COVID-19 Genomics UK (COG-UK) Consortium Ana da Silva Filipe, Kathy Smollett, Stephen Carmichael, Natasha Johnson, Daniel Mair, Lily Tong, Jenna Nichols; Sarah McDonald; Richard Orton, Joseph Hughes, Sreenu Vattipally, David L Robertson; Kathy Li, Natasha Jesudason, Rajiv Shah, James Shepherd, Antonia Ho, Emma Thomson; Alasdair MacLean, Rory Gunson.
EPI_ISL_425762 hCoV-19/Scotland/CVR33/2020 Europe / United Kingdom / Scotland 2020-03-13 West of Scotland Specialist Virology Centre, NHSGGC / MRC-University of Glasgow Centre for Virus Research COVID-19 Genomics UK (COG-UK) Consortium Ana da Silva Filipe, Kathy Smollett, Stephen Carmichael, Natasha Johnson, Daniel Mair, Lily Tong, Jenna Nichols; Sarah McDonald; Richard Orton, Joseph Hughes, Sreenu Vattipally, David L Robertson; Kathy Li, Natasha Jesudason, Rajiv Shah, James Shepherd, Antonia Ho, Emma Thomson; Alasdair MacLean, Rory Gunson.
EPI_ISL_425763 hCoV-19/Scotland/CVR34/2020 Europe / United Kingdom / Scotland 2020-03-12 West of Scotland Specialist Virology Centre, NHSGGC / MRC-University of Glasgow Centre for Virus Research COVID-19 Genomics UK (COG-UK) Consortium Ana da Silva Filipe, Kathy Smollett, Stephen Carmichael, Natasha Johnson, Daniel Mair, Lily Tong, Jenna Nichols; Sarah McDonald; Richard Orton, Joseph Hughes, Sreenu Vattipally, David L Robertson; Kathy Li, Natasha Jesudason, Rajiv Shah, James Shepherd, Antonia Ho, Emma Thomson; Alasdair MacLean, Rory Gunson.
EPI_ISL_425764 hCoV-19/Scotland/CVR35/2020 Europe / United Kingdom / Scotland 2020-03-13 West of Scotland Specialist Virology Centre, NHSGGC / MRC-University of Glasgow Centre for Virus Research COVID-19 Genomics UK (COG-UK) Consortium Ana da Silva Filipe, Kathy Smollett, Stephen Carmichael, Natasha Johnson, Daniel Mair, Lily Tong, Jenna Nichols; Sarah McDonald; Richard Orton, Joseph Hughes, Sreenu Vattipally, David L Robertson; Kathy Li, Natasha Jesudason, Rajiv Shah, James Shepherd, Antonia Ho, Emma Thomson; Alasdair MacLean, Rory Gunson.
EPI_ISL_425765 hCoV-19/Scotland/CVR36/2020 Europe / United Kingdom / Scotland 2020-03-13 West of Scotland Specialist Virology Centre, NHSGGC / MRC-University of Glasgow Centre for Virus Research COVID-19 Genomics UK (COG-UK) Consortium Ana da Silva Filipe, Kathy Smollett, Stephen Carmichael, Natasha Johnson, Daniel Mair, Lily Tong, Jenna Nichols; Sarah McDonald; Richard Orton, Joseph Hughes, Sreenu Vattipally, David L Robertson; Kathy Li, Natasha Jesudason, Rajiv Shah, James Shepherd, Antonia Ho, Emma Thomson; Alasdair MacLean, Rory Gunson.
EPI_ISL_425766 hCoV-19/Scotland/CVR37/2020 Europe / United Kingdom / Scotland 2020-03-13 West of Scotland Specialist Virology Centre, NHSGGC / MRC-University of Glasgow Centre for Virus Research COVID-19 Genomics UK (COG-UK) Consortium Ana da Silva Filipe, Kathy Smollett, Stephen Carmichael, Natasha Johnson, Daniel Mair, Lily Tong, Jenna Nichols; Sarah McDonald; Richard Orton, Joseph Hughes, Sreenu Vattipally, David L Robertson; Kathy Li, Natasha Jesudason, Rajiv Shah, James Shepherd, Antonia Ho, Emma Thomson; Alasdair MacLean, Rory Gunson.
EPI_ISL_425767 hCoV-19/Scotland/CVR38/2020 Europe / United Kingdom / Scotland 2020-03-13 West of Scotland Specialist Virology Centre, NHSGGC / MRC-University of Glasgow Centre for Virus Research COVID-19 Genomics UK (COG-UK) Consortium Ana da Silva Filipe, Kathy Smollett, Stephen Carmichael, Natasha Johnson, Daniel Mair, Lily Tong, Jenna Nichols; Sarah McDonald; Richard Orton, Joseph Hughes, Sreenu Vattipally, David L Robertson; Kathy Li, Natasha Jesudason, Rajiv Shah, James Shepherd, Antonia Ho, Emma Thomson; Alasdair MacLean, Rory Gunson.
EPI_ISL_425768 hCoV-19/Scotland/CVR40/2020 Europe / United Kingdom / Scotland 2020-03-13 West of Scotland Specialist Virology Centre, NHSGGC / MRC-University of Glasgow Centre for Virus Research COVID-19 Genomics UK (COG-UK) Consortium Ana da Silva Filipe, Kathy Smollett, Stephen Carmichael, Natasha Johnson, Daniel Mair, Lily Tong, Jenna Nichols; Sarah McDonald; Richard Orton, Joseph Hughes, Sreenu Vattipally, David L Robertson; Kathy Li, Natasha Jesudason, Rajiv Shah, James Shepherd, Antonia Ho, Emma Thomson; Alasdair MacLean, Rory Gunson.
EPI_ISL_425769 hCoV-19/Scotland/CVR41/2020 Europe / United Kingdom / Scotland 2020-03-13 West of Scotland Specialist Virology Centre, NHSGGC / MRC-University of Glasgow Centre for Virus Research COVID-

19 Genomics UK (COG-UK) Consortium	Ana da Silva Filipe, Kathy Smollett, Stephen Carmichael, Natasha Johnson, Daniel Mair, Lily Tong, Jenna Nichols; Sarah McDonald; Richard Orton, Joseph Hughes, Sreenu Vattipally, David L Robertson; Kathy Li, Natasha Jesudason, Rajiv Shah, James Shepherd, Antonia Ho, Emma Thomson; Alasdair MacLean, Rory Gunson.
EPI_ISL_425770	hCoV-19/Scotland/CVR42/2020	Europe / United Kingdom / Scotland	2020-03-13	West of Scotland Specialist Virology Centre, NHSGGC / MRC-University of Glasgow Centre for Virus Research	COVID-19 Genomics UK (COG-UK) Consortium	Ana da Silva Filipe, Kathy Smollett, Stephen Carmichael, Natasha Johnson, Daniel Mair, Lily Tong, Jenna Nichols; Sarah McDonald; Richard Orton, Joseph Hughes, Sreenu Vattipally, David L Robertson; Kathy Li, Natasha Jesudason, Rajiv Shah, James Shepherd, Antonia Ho, Emma Thomson; Alasdair MacLean, Rory Gunson.
EPI_ISL_425772	hCoV-19/Scotland/CVR44/2020	Europe / United Kingdom / Scotland	2020-03-13	West of Scotland Specialist Virology Centre, NHSGGC / MRC-University of Glasgow Centre for Virus Research	COVID-19 Genomics UK (COG-UK) Consortium	Ana da Silva Filipe, Kathy Smollett, Stephen Carmichael, Natasha Johnson, Daniel Mair, Lily Tong, Jenna Nichols; Sarah McDonald; Richard Orton, Joseph Hughes, Sreenu Vattipally, David L Robertson; Kathy Li, Natasha Jesudason, Rajiv Shah, James Shepherd, Antonia Ho, Emma Thomson; Alasdair MacLean, Rory Gunson.
EPI_ISL_425773	hCoV-19/Scotland/CVR45/2020	Europe / United Kingdom / Scotland	2020-03-13	West of Scotland Specialist Virology Centre, NHSGGC / MRC-University of Glasgow Centre for Virus Research	COVID-19 Genomics UK (COG-UK) Consortium	Ana da Silva Filipe, Kathy Smollett, Stephen Carmichael, Natasha Johnson, Daniel Mair, Lily Tong, Jenna Nichols; Sarah McDonald; Richard Orton, Joseph Hughes, Sreenu Vattipally, David L Robertson; Kathy Li, Natasha Jesudason, Rajiv Shah, James Shepherd, Antonia Ho, Emma Thomson; Alasdair MacLean, Rory Gunson.
EPI_ISL_425774	hCoV-19/Scotland/CVR46/2020	Europe / United Kingdom / Scotland	2020-03-13	West of Scotland Specialist Virology Centre, NHSGGC / MRC-University of Glasgow Centre for Virus Research	COVID-19 Genomics UK (COG-UK) Consortium	Ana da Silva Filipe, Kathy Smollett, Stephen Carmichael, Natasha Johnson, Daniel Mair, Lily Tong, Jenna Nichols; Sarah McDonald; Richard Orton, Joseph Hughes, Sreenu Vattipally, David L Robertson; Kathy Li, Natasha Jesudason, Rajiv Shah, James Shepherd, Antonia Ho, Emma Thomson; Alasdair MacLean, Rory Gunson.
EPI_ISL_425775	hCoV-19/Scotland/CVR462/2020	Europe / United Kingdom / Scotland	2020-03-23	West of Scotland Specialist Virology Centre, NHSGGC / MRC-University of Glasgow Centre for Virus Research	COVID-19 Genomics UK (COG-UK) Consortium	Ana da Silva Filipe, Kathy Smollett, Stephen Carmichael, Natasha Johnson, Daniel Mair, Lily Tong, Jenna Nichols; Sarah McDonald; Richard Orton, Joseph Hughes, Sreenu Vattipally, David L Robertson; Kathy Li, Natasha Jesudason, Rajiv Shah, James Shepherd, Antonia Ho, Emma Thomson; Alasdair MacLean, Rory Gunson.
EPI_ISL_425776	hCoV-19/Scotland/CVR47/2020	Europe / United Kingdom / Scotland	2020-03-13	West of Scotland Specialist Virology Centre, NHSGGC / MRC-University of Glasgow Centre for Virus Research	COVID-19 Genomics UK (COG-UK) Consortium	Ana da Silva Filipe, Kathy Smollett, Stephen Carmichael, Natasha Johnson, Daniel Mair, Lily Tong, Jenna Nichols; Sarah McDonald; Richard Orton, Joseph Hughes, Sreenu Vattipally, David L Robertson; Kathy Li, Natasha Jesudason, Rajiv Shah, James Shepherd, Antonia Ho, Emma Thomson; Alasdair MacLean, Rory Gunson.
EPI_ISL_425777	hCoV-19/Scotland/CVR49/2020	Europe / United Kingdom / Scotland	2020-03-13	West of Scotland Specialist Virology Centre, NHSGGC / MRC-University of Glasgow Centre for Virus Research	COVID-19 Genomics UK (COG-UK) Consortium	Ana da Silva Filipe, Kathy Smollett, Stephen Carmichael, Natasha Johnson, Daniel Mair, Lily Tong, Jenna Nichols; Sarah McDonald; Richard Orton, Joseph Hughes, Sreenu Vattipally, David L Robertson; Kathy Li, Natasha Jesudason, Rajiv Shah, James Shepherd, Antonia Ho, Emma Thomson; Alasdair MacLean, Rory Gunson.
EPI_ISL_425778	hCoV-19/Scotland/CVR50/2020	Europe / United Kingdom / Scotland	2020-03-13	West of Scotland Specialist Virology Centre, NHSGGC / MRC-University of Glasgow Centre for Virus Research	COVID-19 Genomics UK (COG-UK) Consortium	Ana da Silva Filipe, Kathy Smollett, Stephen Carmichael, Natasha Johnson, Daniel Mair, Lily Tong, Jenna Nichols; Sarah McDonald; Richard Orton, Joseph Hughes, Sreenu Vattipally, David L Robertson; Kathy Li, Natasha Jesudason, Rajiv Shah, James Shepherd, Antonia Ho, Emma Thomson; Alasdair MacLean, Rory Gunson.
EPI_ISL_425779	hCoV-19/Scotland/CVR51/2020	Europe / United Kingdom / Scotland	2020-03-13	West of Scotland Specialist Virology Centre, NHSGGC / MRC-University of Glasgow Centre for Virus Research	COVID-19 Genomics UK (COG-UK) Consortium	Ana da Silva Filipe, Kathy Smollett, Stephen Carmichael, Natasha Johnson, Daniel Mair, Lily Tong, Jenna Nichols; Sarah McDonald; Richard Orton, Joseph Hughes, Sreenu Vattipally, David L Robertson; Kathy Li, Natasha Jesudason, Rajiv Shah, James Shepherd, Antonia Ho, Emma Thomson; Alasdair MacLean, Rory Gunson.
EPI_ISL_425780	hCoV-19/Scotland/CVR52/2020	Europe / United Kingdom / Scotland	2020-03-14	West of Scotland Specialist Virology Centre, NHSGGC / MRC-University of Glasgow Centre for Virus Research	COVID-19 Genomics UK (COG-UK) Consortium	Ana da Silva Filipe, Kathy Smollett, Stephen Carmichael, Natasha Johnson, Daniel Mair, Lily Tong, Jenna Nichols; Sarah McDonald; Richard Orton, Joseph Hughes, Sreenu Vattipally, David L Robertson; Kathy Li, Natasha Jesudason, Rajiv Shah, James Shepherd, Antonia Ho, Emma Thomson; Alasdair MacLean, Rory Gunson.
EPI_ISL_425782	hCoV-19/Scotland/CVR53/2020	Europe / United Kingdom / Scotland	2020-03-14	West of Scotland Specialist Virology Centre, NHSGGC / MRC-University of Glasgow Centre for Virus Research	COVID-19 Genomics UK (COG-UK) Consortium	Ana da Silva Filipe, Kathy Smollett, Stephen Carmichael, Natasha Johnson, Daniel Mair, Lily Tong, Jenna Nichols; Sarah McDonald; Richard Orton, Joseph Hughes, Sreenu Vattipally, David L Robertson; Kathy Li, Natasha Jesudason, Rajiv Shah, James Shepherd, Antonia Ho, Emma Thomson; Alasdair MacLean, Rory Gunson.

EPI_ISL_425783	hCoV-19/Scotland/CVR54/2020	Europe / United Kingdom / Scotland	2020-03-13	West of Scotland Specialist Virology Centre, NHSGGC / MRC-University of Glasgow Centre for Virus Research	COVID-19 Genomics UK (COG-UK) Consortium	Ana da Silva Filipe, Kathy Smollett, Stephen Carmichael, Natasha Johnson, Daniel Mair, Lily Tong, Jenna Nichols; Sarah McDonald; Richard Orton, Joseph Hughes, Sreenu Vattipally, David L Robertson; Kathy Li, Natasha Jesudason, Rajiv Shah, James Shepherd, Antonia Ho, Emma Thomson; Alasdair MacLean, Rory Gunson.
EPI_ISL_425784	hCoV-19/Scotland/CVR55/2020	Europe / United Kingdom / Scotland	2020-03-13	West of Scotland Specialist Virology Centre, NHSGGC / MRC-University of Glasgow Centre for Virus Research	COVID-19 Genomics UK (COG-UK) Consortium	Ana da Silva Filipe, Kathy Smollett, Stephen Carmichael, Natasha Johnson, Daniel Mair, Lily Tong, Jenna Nichols; Sarah McDonald; Richard Orton, Joseph Hughes, Sreenu Vattipally, David L Robertson; Kathy Li, Natasha Jesudason, Rajiv Shah, James Shepherd, Antonia Ho, Emma Thomson; Alasdair MacLean, Rory Gunson.
EPI_ISL_425785	hCoV-19/Scotland/CVR56/2020	Europe / United Kingdom / Scotland	2020-03-12	West of Scotland Specialist Virology Centre, NHSGGC / MRC-University of Glasgow Centre for Virus Research	COVID-19 Genomics UK (COG-UK) Consortium	Ana da Silva Filipe, Kathy Smollett, Stephen Carmichael, Natasha Johnson, Daniel Mair, Lily Tong, Jenna Nichols; Sarah McDonald; Richard Orton, Joseph Hughes, Sreenu Vattipally, David L Robertson; Kathy Li, Natasha Jesudason, Rajiv Shah, James Shepherd, Antonia Ho, Emma Thomson; Alasdair MacLean, Rory Gunson.
EPI_ISL_425786	hCoV-19/Scotland/CVR562/2020	Europe / United Kingdom / Scotland	2020-03-23	West of Scotland Specialist Virology Centre, NHSGGC / MRC-University of Glasgow Centre for Virus Research	COVID-19 Genomics UK (COG-UK) Consortium	Ana da Silva Filipe, Kathy Smollett, Stephen Carmichael, Natasha Johnson, Daniel Mair, Lily Tong, Jenna Nichols; Sarah McDonald; Richard Orton, Joseph Hughes, Sreenu Vattipally, David L Robertson; Kathy Li, Natasha Jesudason, Rajiv Shah, James Shepherd, Antonia Ho, Emma Thomson; Alasdair MacLean, Rory Gunson.
EPI_ISL_425787	hCoV-19/Scotland/CVR57/2020	Europe / United Kingdom / Scotland	2020-03-12	West of Scotland Specialist Virology Centre, NHSGGC / MRC-University of Glasgow Centre for Virus Research	COVID-19 Genomics UK (COG-UK) Consortium	Ana da Silva Filipe, Kathy Smollett, Stephen Carmichael, Natasha Johnson, Daniel Mair, Lily Tong, Jenna Nichols; Sarah McDonald; Richard Orton, Joseph Hughes, Sreenu Vattipally, David L Robertson; Kathy Li, Natasha Jesudason, Rajiv Shah, James Shepherd, Antonia Ho, Emma Thomson; Alasdair MacLean, Rory Gunson.
EPI_ISL_425788	hCoV-19/Scotland/CVR58/2020	Europe / United Kingdom / Scotland	2020-03-12	West of Scotland Specialist Virology Centre, NHSGGC / MRC-University of Glasgow Centre for Virus Research	COVID-19 Genomics UK (COG-UK) Consortium	Ana da Silva Filipe, Kathy Smollett, Stephen Carmichael, Natasha Johnson, Daniel Mair, Lily Tong, Jenna Nichols; Sarah McDonald; Richard Orton, Joseph Hughes, Sreenu Vattipally, David L Robertson; Kathy Li, Natasha Jesudason, Rajiv Shah, James Shepherd, Antonia Ho, Emma Thomson; Alasdair MacLean, Rory Gunson.
EPI_ISL_425789	hCoV-19/Scotland/CVR60/2020	Europe / United Kingdom / Scotland	2020-03-12	West of Scotland Specialist Virology Centre, NHSGGC / MRC-University of Glasgow Centre for Virus Research	COVID-19 Genomics UK (COG-UK) Consortium	Ana da Silva Filipe, Kathy Smollett, Stephen Carmichael, Natasha Johnson, Daniel Mair, Lily Tong, Jenna Nichols; Sarah McDonald; Richard Orton, Joseph Hughes, Sreenu Vattipally, David L Robertson; Kathy Li, Natasha Jesudason, Rajiv Shah, James Shepherd, Antonia Ho, Emma Thomson; Alasdair MacLean, Rory Gunson.
EPI_ISL_425790	hCoV-19/Scotland/CVR62/2020	Europe / United Kingdom / Scotland	2020-03-12	West of Scotland Specialist Virology Centre, NHSGGC / MRC-University of Glasgow Centre for Virus Research	COVID-19 Genomics UK (COG-UK) Consortium	Ana da Silva Filipe, Kathy Smollett, Stephen Carmichael, Natasha Johnson, Daniel Mair, Lily Tong, Jenna Nichols; Sarah McDonald; Richard Orton, Joseph Hughes, Sreenu Vattipally, David L Robertson; Kathy Li, Natasha Jesudason, Rajiv Shah, James Shepherd, Antonia Ho, Emma Thomson; Alasdair MacLean, Rory Gunson.
EPI_ISL_425791	hCoV-19/Scotland/CVR63/2020	Europe / United Kingdom / Scotland	2020-03-13	West of Scotland Specialist Virology Centre, NHSGGC / MRC-University of Glasgow Centre for Virus Research	COVID-19 Genomics UK (COG-UK) Consortium	Ana da Silva Filipe, Kathy Smollett, Stephen Carmichael, Natasha Johnson, Daniel Mair, Lily Tong, Jenna Nichols; Sarah McDonald; Richard Orton, Joseph Hughes, Sreenu Vattipally, David L Robertson; Kathy Li, Natasha Jesudason, Rajiv Shah, James Shepherd, Antonia Ho, Emma Thomson; Alasdair MacLean, Rory Gunson.
EPI_ISL_425792	hCoV-19/Scotland/CVR632/2020	Europe / United Kingdom / Scotland	2020-03-25	West of Scotland Specialist Virology Centre, NHSGGC / MRC-University of Glasgow Centre for Virus Research	COVID-19 Genomics UK (COG-UK) Consortium	Ana da Silva Filipe, Kathy Smollett, Stephen Carmichael, Natasha Johnson, Daniel Mair, Lily Tong, Jenna Nichols; Sarah McDonald; Richard Orton, Joseph Hughes, Sreenu Vattipally, David L Robertson; Kathy Li, Natasha Jesudason, Rajiv Shah, James Shepherd, Antonia Ho, Emma Thomson; Alasdair MacLean, Rory Gunson.
EPI_ISL_425793	hCoV-19/Scotland/CVR634/2020	Europe / United Kingdom / Scotland	2020-03-25	West of Scotland Specialist Virology Centre, NHSGGC / MRC-University of Glasgow Centre for Virus Research	COVID-19 Genomics UK (COG-UK) Consortium	Ana da Silva Filipe, Kathy Smollett, Stephen Carmichael, Natasha Johnson, Daniel Mair, Lily Tong, Jenna Nichols; Sarah McDonald; Richard Orton, Joseph Hughes, Sreenu Vattipally, David L Robertson; Kathy Li, Natasha Jesudason, Rajiv Shah, James Shepherd, Antonia Ho, Emma Thomson; Alasdair MacLean, Rory Gunson.
EPI_ISL_425794	hCoV-19/Scotland/CVR635/2020	Europe / United Kingdom / Scotland	2020-03-25	West of Scotland Specialist Virology Centre, NHSGGC / MRC-University of Glasgow Centre for Virus Research	COVID-19 Genomics UK (COG-UK) Consortium	Ana da Silva Filipe, Kathy Smollett, Stephen Carmichael, Natasha Johnson, Daniel Mair, Lily Tong, Jenna Nichols; Sarah McDonald; Richard Orton, Joseph Hughes, Sreenu

Vattipally, David L Robertson; Kathy Li, Natasha Jesudason, Rajiv Shah, James Shepherd, Antonia Ho, Emma Thomson; Alasdair MacLean, Rory Gunson.
EPI_ISL_425795 hCoV-19/Scotland/CVR64/2020 Europe / United Kingdom / Scotland 2020-03-03 West of Scotland Specialist Virology Centre, NHSGGC / MRC-University of Glasgow Centre for Virus Research COVID-19 Genomics UK (COG-UK) Consortium Ana da Silva Filipe, Kathy Smollett, Stephen Carmichael, Natasha Johnson, Daniel Mair, Lily Tong, Jenna Nichols; Sarah McDonald; Richard Orton, Joseph Hughes, Sreenu Vattipally, David L Robertson; Kathy Li, Natasha Jesudason, Rajiv Shah, James Shepherd, Antonia Ho, Emma Thomson; Alasdair MacLean, Rory Gunson.
EPI_ISL_425796 hCoV-19/Scotland/CVR65/2020 Europe / United Kingdom / Scotland 2020-03-06 West of Scotland Specialist Virology Centre, NHSGGC / MRC-University of Glasgow Centre for Virus Research COVID-19 Genomics UK (COG-UK) Consortium Ana da Silva Filipe, Kathy Smollett, Stephen Carmichael, Natasha Johnson, Daniel Mair, Lily Tong, Jenna Nichols; Sarah McDonald; Richard Orton, Joseph Hughes, Sreenu Vattipally, David L Robertson; Kathy Li, Natasha Jesudason, Rajiv Shah, James Shepherd, Antonia Ho, Emma Thomson; Alasdair MacLean, Rory Gunson.
EPI_ISL_425797 hCoV-19/Scotland/CVR66/2020 Europe / United Kingdom / Scotland 2020-03-06 West of Scotland Specialist Virology Centre, NHSGGC / MRC-University of Glasgow Centre for Virus Research COVID-19 Genomics UK (COG-UK) Consortium Ana da Silva Filipe, Kathy Smollett, Stephen Carmichael, Natasha Johnson, Daniel Mair, Lily Tong, Jenna Nichols; Sarah McDonald; Richard Orton, Joseph Hughes, Sreenu Vattipally, David L Robertson; Kathy Li, Natasha Jesudason, Rajiv Shah, James Shepherd, Antonia Ho, Emma Thomson; Alasdair MacLean, Rory Gunson.
EPI_ISL_425798 hCoV-19/Scotland/CVR67/2020 Europe / United Kingdom / Scotland 2020-03-07 West of Scotland Specialist Virology Centre, NHSGGC / MRC-University of Glasgow Centre for Virus Research COVID-19 Genomics UK (COG-UK) Consortium Ana da Silva Filipe, Kathy Smollett, Stephen Carmichael, Natasha Johnson, Daniel Mair, Lily Tong, Jenna Nichols; Sarah McDonald; Richard Orton, Joseph Hughes, Sreenu Vattipally, David L Robertson; Kathy Li, Natasha Jesudason, Rajiv Shah, James Shepherd, Antonia Ho, Emma Thomson; Alasdair MacLean, Rory Gunson.
EPI_ISL_425799 hCoV-19/Scotland/CVR68/2020 Europe / United Kingdom / Scotland 2020-03-09 West of Scotland Specialist Virology Centre, NHSGGC / MRC-University of Glasgow Centre for Virus Research COVID-19 Genomics UK (COG-UK) Consortium Ana da Silva Filipe, Kathy Smollett, Stephen Carmichael, Natasha Johnson, Daniel Mair, Lily Tong, Jenna Nichols; Sarah McDonald; Richard Orton, Joseph Hughes, Sreenu Vattipally, David L Robertson; Kathy Li, Natasha Jesudason, Rajiv Shah, James Shepherd, Antonia Ho, Emma Thomson; Alasdair MacLean, Rory Gunson.
EPI_ISL_425800 hCoV-19/Scotland/CVR69/2020 Europe / United Kingdom / Scotland 2020-03-10 West of Scotland Specialist Virology Centre, NHSGGC / MRC-University of Glasgow Centre for Virus Research COVID-19 Genomics UK (COG-UK) Consortium Ana da Silva Filipe, Kathy Smollett, Stephen Carmichael, Natasha Johnson, Daniel Mair, Lily Tong, Jenna Nichols; Sarah McDonald; Richard Orton, Joseph Hughes, Sreenu Vattipally, David L Robertson; Kathy Li, Natasha Jesudason, Rajiv Shah, James Shepherd, Antonia Ho, Emma Thomson; Alasdair MacLean, Rory Gunson.
EPI_ISL_425801 hCoV-19/Scotland/CVR70/2020 Europe / United Kingdom / Scotland 2020-03-09 West of Scotland Specialist Virology Centre, NHSGGC / MRC-University of Glasgow Centre for Virus Research COVID-19 Genomics UK (COG-UK) Consortium Ana da Silva Filipe, Kathy Smollett, Stephen Carmichael, Natasha Johnson, Daniel Mair, Lily Tong, Jenna Nichols; Sarah McDonald; Richard Orton, Joseph Hughes, Sreenu Vattipally, David L Robertson; Kathy Li, Natasha Jesudason, Rajiv Shah, James Shepherd, Antonia Ho, Emma Thomson; Alasdair MacLean, Rory Gunson.
EPI_ISL_425802 hCoV-19/Scotland/CVR71/2020 Europe / United Kingdom / Scotland 2020-03-06 West of Scotland Specialist Virology Centre, NHSGGC / MRC-University of Glasgow Centre for Virus Research COVID-19 Genomics UK (COG-UK) Consortium Ana da Silva Filipe, Kathy Smollett, Stephen Carmichael, Natasha Johnson, Daniel Mair, Lily Tong, Jenna Nichols; Sarah McDonald; Richard Orton, Joseph Hughes, Sreenu Vattipally, David L Robertson; Kathy Li, Natasha Jesudason, Rajiv Shah, James Shepherd, Antonia Ho, Emma Thomson; Alasdair MacLean, Rory Gunson.
EPI_ISL_425803 hCoV-19/Scotland/CVR72/2020 Europe / United Kingdom / Scotland 2020-03-04 West of Scotland Specialist Virology Centre, NHSGGC / MRC-University of Glasgow Centre for Virus Research COVID-19 Genomics UK (COG-UK) Consortium Ana da Silva Filipe, Kathy Smollett, Stephen Carmichael, Natasha Johnson, Daniel Mair, Lily Tong, Jenna Nichols; Sarah McDonald; Richard Orton, Joseph Hughes, Sreenu Vattipally, David L Robertson; Kathy Li, Natasha Jesudason, Rajiv Shah, James Shepherd, Antonia Ho, Emma Thomson; Alasdair MacLean, Rory Gunson.
EPI_ISL_425804 hCoV-19/Scotland/CVR74/2020 Europe / United Kingdom / Scotland 2020-03-07 West of Scotland Specialist Virology Centre, NHSGGC / MRC-University of Glasgow Centre for Virus Research COVID-19 Genomics UK (COG-UK) Consortium Ana da Silva Filipe, Kathy Smollett, Stephen Carmichael, Natasha Johnson, Daniel Mair, Lily Tong, Jenna Nichols; Sarah McDonald; Richard Orton, Joseph Hughes, Sreenu Vattipally, David L Robertson; Kathy Li, Natasha Jesudason, Rajiv Shah, James Shepherd, Antonia Ho, Emma Thomson; Alasdair MacLean, Rory Gunson.
EPI_ISL_425805 hCoV-19/Scotland/CVR75/2020 Europe / United Kingdom / Scotland 2020-03-06 West of Scotland Specialist Virology Centre, NHSGGC / MRC-University of Glasgow Centre for Virus Research COVID-19 Genomics UK (COG-UK) Consortium Ana da Silva Filipe, Kathy Smollett, Stephen Carmichael, Natasha Johnson, Daniel Mair, Lily Tong, Jenna Nichols; Sarah McDonald; Richard Orton, Joseph Hughes, Sreenu Vattipally, David L Robertson; Kathy Li, Natasha Jesudason, Rajiv Shah, James Shepherd, Antonia Ho, Emma Thomson; Alasdair MacLean, Rory Gunson.
EPI_ISL_425806 hCoV-19/Scotland/CVR76/2020 Europe / United Kingdom / Scotland 2020-03-05 West of Scotland Specialist Virology Centre, NHSGGC / MRC-University of Glasgow Centre for Virus Research COVID-

19 Genomics UK (COG-UK) Consortium	Ana da Silva Filipe, Kathy Smollett, Stephen Carmichael, Natasha Johnson, Daniel Mair, Lily Tong, Jenna Nichols; Sarah McDonald; Richard Orton, Joseph Hughes, Sreenu Vattipally, David L Robertson; Kathy Li, Natasha Jesudason, Rajiv Shah, James Shepherd, Antonia Ho, Emma Thomson; Alasdair MacLean, Rory Gunson.
EPI_ISL_425807	hCoV-19/Scotland/CVR77/2020	Europe / United Kingdom / Scotland	2020-03-03	West of Scotland Specialist Virology Centre, NHSGGC / MRC-University of Glasgow Centre for Virus Research	COVID-19 Genomics UK (COG-UK) Consortium	Ana da Silva Filipe, Kathy Smollett, Stephen Carmichael, Natasha Johnson, Daniel Mair, Lily Tong, Jenna Nichols; Sarah McDonald; Richard Orton, Joseph Hughes, Sreenu Vattipally, David L Robertson; Kathy Li, Natasha Jesudason, Rajiv Shah, James Shepherd, Antonia Ho, Emma Thomson; Alasdair MacLean, Rory Gunson.
EPI_ISL_425808	hCoV-19/Scotland/CVR78/2020	Europe / United Kingdom / Scotland	2020-03-06	West of Scotland Specialist Virology Centre, NHSGGC / MRC-University of Glasgow Centre for Virus Research	COVID-19 Genomics UK (COG-UK) Consortium	Ana da Silva Filipe, Kathy Smollett, Stephen Carmichael, Natasha Johnson, Daniel Mair, Lily Tong, Jenna Nichols; Sarah McDonald; Richard Orton, Joseph Hughes, Sreenu Vattipally, David L Robertson; Kathy Li, Natasha Jesudason, Rajiv Shah, James Shepherd, Antonia Ho, Emma Thomson; Alasdair MacLean, Rory Gunson.
EPI_ISL_425809	hCoV-19/Scotland/CVR79/2020	Europe / United Kingdom / Scotland	2020-03-09	West of Scotland Specialist Virology Centre, NHSGGC / MRC-University of Glasgow Centre for Virus Research	COVID-19 Genomics UK (COG-UK) Consortium	Ana da Silva Filipe, Kathy Smollett, Stephen Carmichael, Natasha Johnson, Daniel Mair, Lily Tong, Jenna Nichols; Sarah McDonald; Richard Orton, Joseph Hughes, Sreenu Vattipally, David L Robertson; Kathy Li, Natasha Jesudason, Rajiv Shah, James Shepherd, Antonia Ho, Emma Thomson; Alasdair MacLean, Rory Gunson.
EPI_ISL_425810	hCoV-19/Scotland/CVR80/2020	Europe / United Kingdom / Scotland	2020-03-05	West of Scotland Specialist Virology Centre, NHSGGC / MRC-University of Glasgow Centre for Virus Research	COVID-19 Genomics UK (COG-UK) Consortium	Ana da Silva Filipe, Kathy Smollett, Stephen Carmichael, Natasha Johnson, Daniel Mair, Lily Tong, Jenna Nichols; Sarah McDonald; Richard Orton, Joseph Hughes, Sreenu Vattipally, David L Robertson; Kathy Li, Natasha Jesudason, Rajiv Shah, James Shepherd, Antonia Ho, Emma Thomson; Alasdair MacLean, Rory Gunson.
EPI_ISL_425811	hCoV-19/Scotland/CVR83/2020	Europe / United Kingdom / Scotland	2020-03-09	West of Scotland Specialist Virology Centre, NHSGGC / MRC-University of Glasgow Centre for Virus Research	COVID-19 Genomics UK (COG-UK) Consortium	Ana da Silva Filipe, Kathy Smollett, Stephen Carmichael, Natasha Johnson, Daniel Mair, Lily Tong, Jenna Nichols; Sarah McDonald; Richard Orton, Joseph Hughes, Sreenu Vattipally, David L Robertson; Kathy Li, Natasha Jesudason, Rajiv Shah, James Shepherd, Antonia Ho, Emma Thomson; Alasdair MacLean, Rory Gunson.
EPI_ISL_425812	hCoV-19/Scotland/CVR85/2020	Europe / United Kingdom / Scotland	2020-03-09	West of Scotland Specialist Virology Centre, NHSGGC / MRC-University of Glasgow Centre for Virus Research	COVID-19 Genomics UK (COG-UK) Consortium	Ana da Silva Filipe, Kathy Smollett, Stephen Carmichael, Natasha Johnson, Daniel Mair, Lily Tong, Jenna Nichols; Sarah McDonald; Richard Orton, Joseph Hughes, Sreenu Vattipally, David L Robertson; Kathy Li, Natasha Jesudason, Rajiv Shah, James Shepherd, Antonia Ho, Emma Thomson; Alasdair MacLean, Rory Gunson.
EPI_ISL_425813	hCoV-19/Scotland/CVR89/2020	Europe / United Kingdom / Scotland	2020-03-12	West of Scotland Specialist Virology Centre, NHSGGC / MRC-University of Glasgow Centre for Virus Research	COVID-19 Genomics UK (COG-UK) Consortium	Ana da Silva Filipe, Kathy Smollett, Stephen Carmichael, Natasha Johnson, Daniel Mair, Lily Tong, Jenna Nichols; Sarah McDonald; Richard Orton, Joseph Hughes, Sreenu Vattipally, David L Robertson; Kathy Li, Natasha Jesudason, Rajiv Shah, James Shepherd, Antonia Ho, Emma Thomson; Alasdair MacLean, Rory Gunson.
EPI_ISL_425815	hCoV-19/Scotland/CVR91/2020	Europe / United Kingdom / Scotland	2020-03-12	West of Scotland Specialist Virology Centre, NHSGGC / MRC-University of Glasgow Centre for Virus Research	COVID-19 Genomics UK (COG-UK) Consortium	Ana da Silva Filipe, Kathy Smollett, Stephen Carmichael, Natasha Johnson, Daniel Mair, Lily Tong, Jenna Nichols; Sarah McDonald; Richard Orton, Joseph Hughes, Sreenu Vattipally, David L Robertson; Kathy Li, Natasha Jesudason, Rajiv Shah, James Shepherd, Antonia Ho, Emma Thomson; Alasdair MacLean, Rory Gunson.
EPI_ISL_425816	hCoV-19/Scotland/CVR96/2020	Europe / United Kingdom / Scotland	2020-03-14	West of Scotland Specialist Virology Centre, NHSGGC / MRC-University of Glasgow Centre for Virus Research	COVID-19 Genomics UK (COG-UK) Consortium	Ana da Silva Filipe, Kathy Smollett, Stephen Carmichael, Natasha Johnson, Daniel Mair, Lily Tong, Jenna Nichols; Sarah McDonald; Richard Orton, Joseph Hughes, Sreenu Vattipally, David L Robertson; Kathy Li, Natasha Jesudason, Rajiv Shah, James Shepherd, Antonia Ho, Emma Thomson; Alasdair MacLean, Rory Gunson.
EPI_ISL_425817	hCoV-19/Scotland/CVR97/2020	Europe / United Kingdom / Scotland	2020-03-13	West of Scotland Specialist Virology Centre, NHSGGC / MRC-University of Glasgow Centre for Virus Research	COVID-19 Genomics UK (COG-UK) Consortium	Ana da Silva Filipe, Kathy Smollett, Stephen Carmichael, Natasha Johnson, Daniel Mair, Lily Tong, Jenna Nichols; Sarah McDonald; Richard Orton, Joseph Hughes, Sreenu Vattipally, David L Robertson; Kathy Li, Natasha Jesudason, Rajiv Shah, James Shepherd, Antonia Ho, Emma Thomson; Alasdair MacLean, Rory Gunson.
EPI_ISL_425818	hCoV-19/Scotland/CVR99/2020	Europe / United Kingdom / Scotland	2020-03-14	West of Scotland Specialist Virology Centre, NHSGGC / MRC-University of Glasgow Centre for Virus Research	COVID-19 Genomics UK (COG-UK) Consortium	Ana da Silva Filipe, Kathy Smollett, Stephen Carmichael, Natasha Johnson, Daniel Mair, Lily Tong, Jenna Nichols; Sarah McDonald; Richard Orton, Joseph Hughes, Sreenu Vattipally, David L Robertson; Kathy Li, Natasha Jesudason, Rajiv Shah, James Shepherd, Antonia Ho, Emma Thomson; Alasdair MacLean, Rory Gunson.

EPI_ISL_425819 hCoV-19/Scotland/EDB002/2020 Europe / United Kingdom / Scotland 2020-03-05
Virology Department, Royal Infirmary of Edinburgh, NHS Lothian / School of Biological Sciences, University of Edinburgh / Institute of Genetics and Molecular Medicine, University of Edinburgh COVID-19 Genomics UK (COG-UK) Consortium McHugh M, Dewar R, Rooke S, Gallagher M, Balcaza C, O'Toole A, Hill V, McCrone JT, Colquhoun R, Yu X, Jackson B, Scher E, Rambaut A, Williams TC, Templeton K
EPI_ISL_425820 hCoV-19/Scotland/EDB014/2020 Europe / United Kingdom / Scotland 2020-03-11
Virology Department, Royal Infirmary of Edinburgh, NHS Lothian / School of Biological Sciences, University of Edinburgh / Institute of Genetics and Molecular Medicine, University of Edinburgh COVID-19 Genomics UK (COG-UK) Consortium McHugh M, Dewar R, Rooke S, Gallagher M, Balcaza C, O'Toole A, Hill V, McCrone JT, Colquhoun R, Yu X, Jackson B, Scher E, Rambaut A, Williams TC, Templeton K
EPI_ISL_425821 hCoV-19/Scotland/EDB015/2020 Europe / United Kingdom / Scotland 2020-03-11
Virology Department, Royal Infirmary of Edinburgh, NHS Lothian / School of Biological Sciences, University of Edinburgh / Institute of Genetics and Molecular Medicine, University of Edinburgh COVID-19 Genomics UK (COG-UK) Consortium McHugh M, Dewar R, Rooke S, Gallagher M, Balcaza C, O'Toole A, Hill V, McCrone JT, Colquhoun R, Yu X, Jackson B, Scher E, Rambaut A, Williams TC, Templeton K
EPI_ISL_425822 hCoV-19/Scotland/EDB016/2020 Europe / United Kingdom / Scotland 2020-03-11
Virology Department, Royal Infirmary of Edinburgh, NHS Lothian / School of Biological Sciences, University of Edinburgh / Institute of Genetics and Molecular Medicine, University of Edinburgh COVID-19 Genomics UK (COG-UK) Consortium McHugh M, Dewar R, Rooke S, Gallagher M, Balcaza C, O'Toole A, Hill V, McCrone JT, Colquhoun R, Yu X, Jackson B, Scher E, Rambaut A, Williams TC, Templeton K
EPI_ISL_425900 hCoV-19/Scotland/EDB113/2020 Europe / United Kingdom / Scotland 2020-03-22
Virology Department, Royal Infirmary of Edinburgh, NHS Lothian / School of Biological Sciences, University of Edinburgh / Institute of Genetics and Molecular Medicine, University of Edinburgh COVID-19 Genomics UK (COG-UK) Consortium McHugh M, Dewar R, Rooke S, Gallagher M, Balcaza C, O'Toole A, Hill V, McCrone JT, Colquhoun R, Yu X, Jackson B, Scher E, Rambaut A, Williams TC, Templeton K
EPI_ISL_425902 hCoV-19/Scotland/EDB118/2020 Europe / United Kingdom / Scotland 2020-03-23
Virology Department, Royal Infirmary of Edinburgh, NHS Lothian / School of Biological Sciences, University of Edinburgh / Institute of Genetics and Molecular Medicine, University of Edinburgh COVID-19 Genomics UK (COG-UK) Consortium McHugh M, Dewar R, Rooke S, Gallagher M, Balcaza C, O'Toole A, Hill V, McCrone JT, Colquhoun R, Yu X, Jackson B, Scher E, Rambaut A, Williams TC, Templeton K
EPI_ISL_425904 hCoV-19/Scotland/EDB120/2020 Europe / United Kingdom / Scotland 2020-03-22
Virology Department, Royal Infirmary of Edinburgh, NHS Lothian / School of Biological Sciences, University of Edinburgh / Institute of Genetics and Molecular Medicine, University of Edinburgh COVID-19 Genomics UK (COG-UK) Consortium McHugh M, Dewar R, Rooke S, Gallagher M, Balcaza C, O'Toole A, Hill V, McCrone JT, Colquhoun R, Yu X, Jackson B, Scher E, Rambaut A, Williams TC, Templeton K
EPI_ISL_425906 hCoV-19/Scotland/EDB124/2020 Europe / United Kingdom / Scotland 2020-03-18
Virology Department, Royal Infirmary of Edinburgh, NHS Lothian / School of Biological Sciences, University of Edinburgh / Institute of Genetics and Molecular Medicine, University of Edinburgh COVID-19 Genomics UK (COG-UK) Consortium McHugh M, Dewar R, Rooke S, Gallagher M, Balcaza C, O'Toole A, Hill V, McCrone JT, Colquhoun R, Yu X, Jackson B, Scher E, Rambaut A, Williams TC, Templeton K
EPI_ISL_425907 hCoV-19/Scotland/EDB125/2020 Europe / United Kingdom / Scotland 2020-03-23
Virology Department, Royal Infirmary of Edinburgh, NHS Lothian / School of Biological Sciences, University of Edinburgh / Institute of Genetics and Molecular Medicine, University of Edinburgh COVID-19 Genomics UK (COG-UK) Consortium McHugh M, Dewar R, Rooke S, Gallagher M, Balcaza C, O'Toole A, Hill V, McCrone JT, Colquhoun R, Yu X, Jackson B, Scher E, Rambaut A, Williams TC, Templeton K
EPI_ISL_425911 hCoV-19/Scotland/EDB132/2020 Europe / United Kingdom / Scotland 2020-03-23
Virology Department, Royal Infirmary of Edinburgh, NHS Lothian / School of Biological Sciences, University of Edinburgh / Institute of Genetics and Molecular Medicine, University of Edinburgh COVID-19 Genomics UK (COG-UK) Consortium McHugh M, Dewar R, Rooke S, Gallagher M, Balcaza C, O'Toole A, Hill V, McCrone JT, Colquhoun R, Yu X, Jackson B, Scher E, Rambaut A, Williams TC, Templeton K
EPI_ISL_425912 hCoV-19/Scotland/EDB142/2020 Europe / United Kingdom / Scotland 2020-03-23
Virology Department, Royal Infirmary of Edinburgh, NHS Lothian / School of Biological Sciences, University of Edinburgh / Institute of Genetics and Molecular Medicine, University of Edinburgh COVID-19 Genomics UK (COG-UK) Consortium McHugh M, Dewar R, Rooke S, Gallagher M, Balcaza C, O'Toole A, Hill V, McCrone JT, Colquhoun R, Yu X, Jackson B, Scher E, Rambaut A, Williams TC, Templeton K
EPI_ISL_425916 hCoV-19/Scotland/EDB148/2020 Europe / United Kingdom / Scotland 2020-03-23
Virology Department, Royal Infirmary of Edinburgh, NHS Lothian / School of Biological Sciences, University of Edinburgh / Institute of Genetics and Molecular Medicine, University of Edinburgh COVID-19 Genomics UK (COG-UK) Consortium McHugh M, Dewar R, Rooke S, Gallagher M, Balcaza C, O'Toole A, Hill V, McCrone JT, Colquhoun R, Yu X, Jackson B, Scher E, Rambaut A, Williams TC, Templeton K
EPI_ISL_425917 hCoV-19/Scotland/EDB149/2020 Europe / United Kingdom / Scotland 2020-03-22
Virology Department, Royal Infirmary of Edinburgh, NHS Lothian / School of Biological Sciences, University of Edinburgh / Institute of Genetics and Molecular Medicine, University of Edinburgh COVID-19 Genomics UK (COG-UK) Consortium McHugh M, Dewar R, Rooke S, Gallagher M, Balcaza C, O'Toole A, Hill V, McCrone JT, Colquhoun R, Yu X, Jackson B, Scher E, Rambaut A, Williams TC, Templeton K
EPI_ISL_425918 hCoV-19/Scotland/EDB150/2020 Europe / United Kingdom / Scotland 2020-03-24
Virology Department, Royal Infirmary of Edinburgh, NHS Lothian / School of Biological Sciences, University of Edinburgh / Institute of Genetics and Molecular Medicine, University of Edinburgh COVID-19 Genomics UK (COG-UK) Consortium McHugh M, Dewar R, Rooke S, Gallagher M, Balcaza C, O'Toole A, Hill V, McCrone JT, Colquhoun R, Yu X, Jackson B, Scher E, Rambaut A, Williams TC, Templeton K

EPI_ISL_425919  hCoV-19/Scotland/EDB151/2020    Europe / United Kingdom / Scotland      2020-03-24
Virology Department, Royal Infirmary of Edinburgh, NHS Lothian / School of Biological Sciences, University of Edinburgh / Institute of Genetics and Molecular Medicine, University of Edinburgh       COVID-19 Genomics UK (COG-UK) Consortium     McHugh M, Dewar R, Rooke S, Gallagher M, Balcaza C, O'Toole A, Hill V, McCrone JT, Colquhoun R, Yu X, Jackson B, Scher E, Rambaut A, Williams TC, Templeton K
EPI_ISL_425921  hCoV-19/Scotland/EDB156/2020    Europe / United Kingdom / Scotland      2020-03-23
Virology Department, Royal Infirmary of Edinburgh, NHS Lothian / School of Biological Sciences, University of Edinburgh / Institute of Genetics and Molecular Medicine, University of Edinburgh       COVID-19 Genomics UK (COG-UK) Consortium     McHugh M, Dewar R, Rooke S, Gallagher M, Balcaza C, O'Toole A, Hill V, McCrone JT, Colquhoun R, Yu X, Jackson B, Scher E, Rambaut A, Williams TC, Templeton K
EPI_ISL_425922  hCoV-19/Scotland/EDB157/2020    Europe / United Kingdom / Scotland      2020-03-23
Virology Department, Royal Infirmary of Edinburgh, NHS Lothian / School of Biological Sciences, University of Edinburgh / Institute of Genetics and Molecular Medicine, University of Edinburgh       COVID-19 Genomics UK (COG-UK) Consortium     McHugh M, Dewar R, Rooke S, Gallagher M, Balcaza C, O'Toole A, Hill V, McCrone JT, Colquhoun R, Yu X, Jackson B, Scher E, Rambaut A, Williams TC, Templeton K
EPI_ISL_425924  hCoV-19/Scotland/EDB162/2020    Europe / United Kingdom / Scotland      2020-03-24
Virology Department, Royal Infirmary of Edinburgh, NHS Lothian / School of Biological Sciences, University of Edinburgh / Institute of Genetics and Molecular Medicine, University of Edinburgh       COVID-19 Genomics UK (COG-UK) Consortium     McHugh M, Dewar R, Rooke S, Gallagher M, Balcaza C, O'Toole A, Hill V, McCrone JT, Colquhoun R, Yu X, Jackson B, Scher E, Rambaut A, Williams TC, Templeton K
EPI_ISL_425925  hCoV-19/Scotland/EDB168/2020    Europe / United Kingdom / Scotland      2020-03-24
Virology Department, Royal Infirmary of Edinburgh, NHS Lothian / School of Biological Sciences, University of Edinburgh / Institute of Genetics and Molecular Medicine, University of Edinburgh       COVID-19 Genomics UK (COG-UK) Consortium     McHugh M, Dewar R, Rooke S, Gallagher M, Balcaza C, O'Toole A, Hill V, McCrone JT, Colquhoun R, Yu X, Jackson B, Scher E, Rambaut A, Williams TC, Templeton K
EPI_ISL_425927  hCoV-19/Scotland/EDB181/2020    Europe / United Kingdom / Scotland      2020-03-24
Virology Department, Royal Infirmary of Edinburgh, NHS Lothian / School of Biological Sciences, University of Edinburgh / Institute of Genetics and Molecular Medicine, University of Edinburgh       COVID-19 Genomics UK (COG-UK) Consortium     McHugh M, Dewar R, Rooke S, Gallagher M, Balcaza C, O'Toole A, Hill V, McCrone JT, Colquhoun R, Yu X, Jackson B, Scher E, Rambaut A, Williams TC, Templeton K
EPI_ISL_425933  hCoV-19/Scotland/EDB189/2020    Europe / United Kingdom / Scotland      2020-03-25
Virology Department, Royal Infirmary of Edinburgh, NHS Lothian / School of Biological Sciences, University of Edinburgh / Institute of Genetics and Molecular Medicine, University of Edinburgh       COVID-19 Genomics UK (COG-UK) Consortium     McHugh M, Dewar R, Rooke S, Gallagher M, Balcaza C, O'Toole A, Hill V, McCrone JT, Colquhoun R, Yu X, Jackson B, Scher E, Rambaut A, Williams TC, Templeton K
EPI_ISL_425937  hCoV-19/Scotland/EDB194/2020    Europe / United Kingdom / Scotland      2020-03-25
Virology Department, Royal Infirmary of Edinburgh, NHS Lothian / School of Biological Sciences, University of Edinburgh / Institute of Genetics and Molecular Medicine, University of Edinburgh       COVID-19 Genomics UK (COG-UK) Consortium     McHugh M, Dewar R, Rooke S, Gallagher M, Balcaza C, O'Toole A, Hill V, McCrone JT, Colquhoun R, Yu X, Jackson B, Scher E, Rambaut A, Williams TC, Templeton K
EPI_ISL_425939  hCoV-19/Scotland/EDB198/2020    Europe / United Kingdom / Scotland      2020-03-25
Virology Department, Royal Infirmary of Edinburgh, NHS Lothian / School of Biological Sciences, University of Edinburgh / Institute of Genetics and Molecular Medicine, University of Edinburgh       COVID-19 Genomics UK (COG-UK) Consortium     McHugh M, Dewar R, Rooke S, Gallagher M, Balcaza C, O'Toole A, Hill V, McCrone JT, Colquhoun R, Yu X, Jackson B, Scher E, Rambaut A, Williams TC, Templeton K
EPI_ISL_425942  hCoV-19/Scotland/EDB204/2020    Europe / United Kingdom / Scotland      2020-03-26
Virology Department, Royal Infirmary of Edinburgh, NHS Lothian / School of Biological Sciences, University of Edinburgh / Institute of Genetics and Molecular Medicine, University of Edinburgh       COVID-19 Genomics UK (COG-UK) Consortium     McHugh M, Dewar R, Rooke S, Gallagher M, Balcaza C, O'Toole A, Hill V, McCrone JT, Colquhoun R, Yu X, Jackson B, Scher E, Rambaut A, Williams TC, Templeton K
EPI_ISL_425943  hCoV-19/Scotland/EDB212/2020    Europe / United Kingdom / Scotland      2020-03-26
Virology Department, Royal Infirmary of Edinburgh, NHS Lothian / School of Biological Sciences, University of Edinburgh / Institute of Genetics and Molecular Medicine, University of Edinburgh       COVID-19 Genomics UK (COG-UK) Consortium     McHugh M, Dewar R, Rooke S, Gallagher M, Balcaza C, O'Toole A, Hill V, McCrone JT, Colquhoun R, Yu X, Jackson B, Scher E, Rambaut A, Williams TC, Templeton K
EPI_ISL_428148  hCoV-19/Sweden/RV-FOI-4/2020    Europe / Sweden / Västerbotten  2020-03-18      Klinisk mikrobiologi, Region Västerbotten       Unit for Biological Agents, Department for CBRN Defence and Security, Swedish Defence Research Agency FOI bioinformatics team
EPI_ISL_429300  hCoV-19/Denmark/ALAB-HH46/2020  Europe / Denmark / Unknown      2020-03-13      Department of Clinical Microbiology, Copenhagen University Hospital, Hvidovre, Kettegaard Alle 30, 2650 Hvidovre.
Albertsen lab, Department of Chemistry and Bioscience, Aalborg University, Denmark      Rasmus Kirkegaard
EPI_ISL_429301  hCoV-19/Denmark/ALAB-HH49/2020  Europe / Denmark / Unknown      2020-03-16      Department of Clinical Microbiology, Copenhagen University Hospital, Hvidovre, Kettegaard Alle 30, 2650 Hvidovre.
Albertsen lab, Department of Chemistry and Bioscience, Aalborg University, Denmark      Rasmus Kirkegaard
EPI_ISL_429302  hCoV-19/Denmark/ALAB-HH50/2020  Europe / Denmark / Unknown      2020-03-16      Department of Clinical Microbiology, Copenhagen University Hospital, Hvidovre, Kettegaard Alle 30, 2650 Hvidovre.
Albertsen lab, Department of Chemistry and Bioscience, Aalborg University, Denmark      Rasmus Kirkegaard
EPI_ISL_429303  hCoV-19/Denmark/ALAB-HH51/2020  Europe / Denmark / Unknown      2020-03-16      Department of Clinical Microbiology, Copenhagen University Hospital, Hvidovre, Kettegaard Alle 30, 2650 Hvidovre.
Albertsen lab, Department of Chemistry and Bioscience, Aalborg University, Denmark      Rasmus Kirkegaard

EPI_ISL_429304 hCoV-19/Denmark/ALAB-HH56/2020 Europe / Denmark / Unknown 2020-03-17 Department of Clinical Microbiology, Copenhagen University Hospital, Hvidovre, Kettegaard Alle 30, 2650 Hvidovre. Albertsen lab, Department of Chemistry and Bioscience, Aalborg University, Denmark Rasmus Kirkegaard
EPI_ISL_429305 hCoV-19/Denmark/ALAB-HH57/2020 Europe / Denmark / Unknown 2020-03-17 Department of Clinical Microbiology, Copenhagen University Hospital, Hvidovre, Kettegaard Alle 30, 2650 Hvidovre. Albertsen lab, Department of Chemistry and Bioscience, Aalborg University, Denmark Rasmus Kirkegaard
EPI_ISL_429306 hCoV-19/Denmark/ALAB-HH60/2020 Europe / Denmark / Unknown 2020-03-17 Department of Clinical Microbiology, Copenhagen University Hospital, Hvidovre, Kettegaard Alle 30, 2650 Hvidovre. Albertsen lab, Department of Chemistry and Bioscience, Aalborg University, Denmark Rasmus Kirkegaard
EPI_ISL_429307 hCoV-19/Denmark/ALAB-HH61/2020 Europe / Denmark / Unknown 2020-03-17 Department of Clinical Microbiology, Copenhagen University Hospital, Hvidovre, Kettegaard Alle 30, 2650 Hvidovre. Albertsen lab, Department of Chemistry and Bioscience, Aalborg University, Denmark Rasmus Kirkegaard
EPI_ISL_429308 hCoV-19/Denmark/ALAB-HH62/2020 Europe / Denmark / Unknown 2020-03-17 Department of Clinical Microbiology, Copenhagen University Hospital, Hvidovre, Kettegaard Alle 30, 2650 Hvidovre. Albertsen lab, Department of Chemistry and Bioscience, Aalborg University, Denmark Rasmus Kirkegaard
EPI_ISL_429309 hCoV-19/Denmark/ALAB-HH63/2020 Europe / Denmark / Unknown 2020-03-18 Department of Clinical Microbiology, Copenhagen University Hospital, Hvidovre, Kettegaard Alle 30, 2650 Hvidovre. Albertsen lab, Department of Chemistry and Bioscience, Aalborg University, Denmark Rasmus Kirkegaard
EPI_ISL_429310 hCoV-19/Denmark/ALAB-HH64/2020 Europe / Denmark / Unknown 2020-03-17 Department of Clinical Microbiology, Copenhagen University Hospital, Hvidovre, Kettegaard Alle 30, 2650 Hvidovre. Albertsen lab, Department of Chemistry and Bioscience, Aalborg University, Denmark Rasmus Kirkegaard
EPI_ISL_429311 hCoV-19/Denmark/ALAB-HH65/2020 Europe / Denmark / Unknown 2020-03-17 Department of Clinical Microbiology, Copenhagen University Hospital, Hvidovre, Kettegaard Alle 30, 2650 Hvidovre. Albertsen lab, Department of Chemistry and Bioscience, Aalborg University, Denmark Rasmus Kirkegaard
EPI_ISL_429312 hCoV-19/Denmark/ALAB-HH66/2020 Europe / Denmark / Unknown 2020-03-18 Department of Clinical Microbiology, Copenhagen University Hospital, Hvidovre, Kettegaard Alle 30, 2650 Hvidovre. Albertsen lab, Department of Chemistry and Bioscience, Aalborg University, Denmark Rasmus Kirkegaard
EPI_ISL_429315 hCoV-19/Denmark/ALAB-HH74/2020 Europe / Denmark / Unknown 2020-03-18 Department of Clinical Microbiology, Copenhagen University Hospital, Hvidovre, Kettegaard Alle 30, 2650 Hvidovre. Albertsen lab, Department of Chemistry and Bioscience, Aalborg University, Denmark Rasmus Kirkegaard
EPI_ISL_429316 hCoV-19/Denmark/ALAB-HH75/2020 Europe / Denmark / Unknown 2020-03-18 Department of Clinical Microbiology, Copenhagen University Hospital, Hvidovre, Kettegaard Alle 30, 2650 Hvidovre. Albertsen lab, Department of Chemistry and Bioscience, Aalborg University, Denmark Rasmus Kirkegaard
EPI_ISL_429318 hCoV-19/Denmark/ALAB-HH77/2020 Europe / Denmark / Unknown 2020-03-16 Department of Clinical Microbiology, Copenhagen University Hospital, Hvidovre, Kettegaard Alle 30, 2650 Hvidovre. Albertsen lab, Department of Chemistry and Bioscience, Aalborg University, Denmark Rasmus Kirkegaard
EPI_ISL_429321 hCoV-19/Denmark/ALAB-HH80/2020 Europe / Denmark / Unknown 2020-03-16 Department of Clinical Microbiology, Copenhagen University Hospital, Hvidovre, Kettegaard Alle 30, 2650 Hvidovre. Albertsen lab, Department of Chemistry and Bioscience, Aalborg University, Denmark Rasmus Kirkegaard
EPI_ISL_429322 hCoV-19/Denmark/ALAB-HH81/2020 Europe / Denmark / Unknown 2020-03-16 Department of Clinical Microbiology, Copenhagen University Hospital, Hvidovre, Kettegaard Alle 30, 2650 Hvidovre. Albertsen lab, Department of Chemistry and Bioscience, Aalborg University, Denmark Rasmus Kirkegaard
EPI_ISL_429323 hCoV-19/Denmark/ALAB-HH82/2020 Europe / Denmark / Unknown 2020-03-15 Department of Clinical Microbiology, Copenhagen University Hospital, Hvidovre, Kettegaard Alle 30, 2650 Hvidovre. Albertsen lab, Department of Chemistry and Bioscience, Aalborg University, Denmark Rasmus Kirkegaard
EPI_ISL_429324 hCoV-19/Denmark/ALAB-HH83/2020 Europe / Denmark / Unknown 2020-03-16 Department of Clinical Microbiology, Copenhagen University Hospital, Hvidovre, Kettegaard Alle 30, 2650 Hvidovre. Albertsen lab, Department of Chemistry and Bioscience, Aalborg University, Denmark Rasmus Kirkegaard
EPI_ISL_429325 hCoV-19/Denmark/ALAB-HH84/2020 Europe / Denmark / Unknown 2020-03-16 Department of Clinical Microbiology, Copenhagen University Hospital, Hvidovre, Kettegaard Alle 30, 2650 Hvidovre. Albertsen lab, Department of Chemistry and Bioscience, Aalborg University, Denmark Rasmus Kirkegaard
EPI_ISL_429327 hCoV-19/Denmark/ALAB-HH87/2020 Europe / Denmark / Unknown 2020-03-15 Department of Clinical Microbiology, Copenhagen University Hospital, Hvidovre, Kettegaard Alle 30, 2650 Hvidovre. Albertsen lab, Department of Chemistry and Bioscience, Aalborg University, Denmark Rasmus Kirkegaard
EPI_ISL_429328 hCoV-19/Denmark/ALAB-HH88/2020 Europe / Denmark / Unknown 2020-03-15 Department of Clinical Microbiology, Copenhagen University Hospital, Hvidovre, Kettegaard Alle 30, 2650 Hvidovre. Albertsen lab, Department of Chemistry and Bioscience, Aalborg University, Denmark Rasmus Kirkegaard
EPI_ISL_429329 hCoV-19/Denmark/ALAB-HH89/2020 Europe / Denmark / Unknown 2020-03-15 Department of Clinical Microbiology, Copenhagen University Hospital, Hvidovre, Kettegaard Alle 30, 2650 Hvidovre. Albertsen lab, Department of Chemistry and Bioscience, Aalborg University, Denmark Rasmus Kirkegaard
EPI_ISL_429331 hCoV-19/Denmark/ALAB-HH92/2020 Europe / Denmark / Unknown 2020-03-15 Department of Clinical Microbiology, Copenhagen University Hospital, Hvidovre, Kettegaard Alle 30, 2650 Hvidovre. Albertsen lab, Department of Chemistry and Bioscience, Aalborg University, Denmark Rasmus Kirkegaard
EPI_ISL_429335 hCoV-19/Denmark/ALAB-SSI103/2020 Europe / Denmark / Unknown 2020-03-02 Department of Virus and Microbiological Special Diagnostics, Statens Serum Institut, Copenhagen, Denmark, Artillerivej 5, 2300 Copenahgen S Albertsen lab, Department of Chemistry and Bioscience, Aalborg University, Denmark Rasmus Kirkegaard
EPI_ISL_429337 hCoV-19/Denmark/ALAB-SSI105/2020 Europe / Denmark / Unknown 2020-03-08 Department of Virus and Microbiological Special Diagnostics, Statens Serum Institut, Copenhagen, Denmark, Artillerivej 5, 2300 Copenahgen S Albertsen lab, Department of Chemistry and Bioscience, Aalborg

University, Denmark	Rasmus Kirkegaard
EPI_ISL_429339	hCoV-19/Denmark/ALAB-SSI107/2020	Europe / Denmark / Unknown	2020-03-07
Department of Virus and Microbiological Special Diagnostics, Statens Serum Institut, Copenhagen, Denmark, Artillerivej 5, 2300 Copenahgen S	Albertsen lab, Department of Chemistry and Bioscience, Aalborg University, Denmark	Rasmus Kirkegaard
EPI_ISL_429340	hCoV-19/Denmark/ALAB-SSI108/2020	Europe / Denmark / Unknown	2020-03-07
Department of Virus and Microbiological Special Diagnostics, Statens Serum Institut, Copenhagen, Denmark, Artillerivej 5, 2300 Copenahgen S	Albertsen lab, Department of Chemistry and Bioscience, Aalborg University, Denmark	Rasmus Kirkegaard
EPI_ISL_429341	hCoV-19/Denmark/ALAB-SSI109/2020	Europe / Denmark / Unknown	2020-03-04
Department of Virus and Microbiological Special Diagnostics, Statens Serum Institut, Copenhagen, Denmark, Artillerivej 5, 2300 Copenahgen S	Albertsen lab, Department of Chemistry and Bioscience, Aalborg University, Denmark	Rasmus Kirkegaard
EPI_ISL_429342	hCoV-19/Denmark/ALAB-SSI110/2020	Europe / Denmark / Unknown	2020-03-08
Department of Virus and Microbiological Special Diagnostics, Statens Serum Institut, Copenhagen, Denmark, Artillerivej 5, 2300 Copenahgen S	Albertsen lab, Department of Chemistry and Bioscience, Aalborg University, Denmark	Rasmus Kirkegaard
EPI_ISL_429343	hCoV-19/Denmark/ALAB-SSI111/2020	Europe / Denmark / Unknown	2020-03-09
Department of Virus and Microbiological Special Diagnostics, Statens Serum Institut, Copenhagen, Denmark, Artillerivej 5, 2300 Copenahgen S	Albertsen lab, Department of Chemistry and Bioscience, Aalborg University, Denmark	Rasmus Kirkegaard
EPI_ISL_429344	hCoV-19/Denmark/ALAB-SSI113/2020	Europe / Denmark / Unknown	2020-03-08
Department of Virus and Microbiological Special Diagnostics, Statens Serum Institut, Copenhagen, Denmark, Artillerivej 5, 2300 Copenahgen S	Albertsen lab, Department of Chemistry and Bioscience, Aalborg University, Denmark	Rasmus Kirkegaard
EPI_ISL_429345	hCoV-19/Denmark/ALAB-SSI114/2020	Europe / Denmark / Unknown	2020-03-09
Department of Virus and Microbiological Special Diagnostics, Statens Serum Institut, Copenhagen, Denmark, Artillerivej 5, 2300 Copenahgen S	Albertsen lab, Department of Chemistry and Bioscience, Aalborg University, Denmark	Rasmus Kirkegaard
EPI_ISL_429346	hCoV-19/Denmark/ALAB-SSI117/2020	Europe / Denmark / Unknown	2020-03-06
Department of Virus and Microbiological Special Diagnostics, Statens Serum Institut, Copenhagen, Denmark, Artillerivej 5, 2300 Copenahgen S	Albertsen lab, Department of Chemistry and Bioscience, Aalborg University, Denmark	Rasmus Kirkegaard
EPI_ISL_429348	hCoV-19/Denmark/ALAB-SSI119/2020	Europe / Denmark / Unknown	2020-03-08
Department of Virus and Microbiological Special Diagnostics, Statens Serum Institut, Copenhagen, Denmark, Artillerivej 5, 2300 Copenahgen S	Albertsen lab, Department of Chemistry and Bioscience, Aalborg University, Denmark	Rasmus Kirkegaard
EPI_ISL_429349	hCoV-19/Denmark/ALAB-SSI120/2020	Europe / Denmark / Unknown	2020-03-08
Department of Virus and Microbiological Special Diagnostics, Statens Serum Institut, Copenhagen, Denmark, Artillerivej 5, 2300 Copenahgen S	Albertsen lab, Department of Chemistry and Bioscience, Aalborg University, Denmark	Rasmus Kirkegaard
EPI_ISL_429353	hCoV-19/Denmark/ALAB-SSI125/2020	Europe / Denmark / Unknown	2020-03-08
Department of Virus and Microbiological Special Diagnostics, Statens Serum Institut, Copenhagen, Denmark, Artillerivej 5, 2300 Copenahgen S	Albertsen lab, Department of Chemistry and Bioscience, Aalborg University, Denmark	Rasmus Kirkegaard
EPI_ISL_429355	hCoV-19/Denmark/ALAB-SSI128/2020	Europe / Denmark / Unknown	2020-03-08
Department of Virus and Microbiological Special Diagnostics, Statens Serum Institut, Copenhagen, Denmark, Artillerivej 5, 2300 Copenahgen S	Albertsen lab, Department of Chemistry and Bioscience, Aalborg University, Denmark	Rasmus Kirkegaard
EPI_ISL_429356	hCoV-19/Denmark/ALAB-SSI129/2020	Europe / Denmark / Unknown	2020-03-08
Department of Virus and Microbiological Special Diagnostics, Statens Serum Institut, Copenhagen, Denmark, Artillerivej 5, 2300 Copenahgen S	Albertsen lab, Department of Chemistry and Bioscience, Aalborg University, Denmark	Rasmus Kirkegaard
EPI_ISL_429357	hCoV-19/Denmark/ALAB-SSI132/2020	Europe / Denmark / Unknown	2020-03-08
Department of Virus and Microbiological Special Diagnostics, Statens Serum Institut, Copenhagen, Denmark, Artillerivej 5, 2300 Copenahgen S	Albertsen lab, Department of Chemistry and Bioscience, Aalborg University, Denmark	Rasmus Kirkegaard
EPI_ISL_429358	hCoV-19/Denmark/ALAB-SSI139/2020	Europe / Denmark / Unknown	2020-03-09
Department of Virus and Microbiological Special Diagnostics, Statens Serum Institut, Copenhagen, Denmark, Artillerivej 5, 2300 Copenahgen S	Albertsen lab, Department of Chemistry and Bioscience, Aalborg University, Denmark	Rasmus Kirkegaard
EPI_ISL_429360	hCoV-19/Denmark/ALAB-SSI142/2020	Europe / Denmark / Unknown	2020-03-09
Department of Virus and Microbiological Special Diagnostics, Statens Serum Institut, Copenhagen, Denmark, Artillerivej 5, 2300 Copenahgen S	Albertsen lab, Department of Chemistry and Bioscience, Aalborg University, Denmark	Rasmus Kirkegaard
EPI_ISL_429361	hCoV-19/Denmark/ALAB-SSI143/2020	Europe / Denmark / Unknown	2020-03-09
Department of Virus and Microbiological Special Diagnostics, Statens Serum Institut, Copenhagen, Denmark, Artillerivej 5, 2300 Copenahgen S	Albertsen lab, Department of Chemistry and Bioscience, Aalborg University, Denmark	Rasmus Kirkegaard
EPI_ISL_429364	hCoV-19/Denmark/ALAB-SSI146/2020	Europe / Denmark / Unknown	2020-03-09

Department of Virus and Microbiological Special Diagnostics, Statens Serum Institut, Copenhagen, Denmark, Artillerivej 5, 2300 Copenahgen S	Albertsen lab, Department of Chemistry and Bioscience, Aalborg University, Denmark	Rasmus Kirkegaard
EPI_ISL_429368	hCoV-19/Denmark/ALAB-SSI150/2020	Europe / Denmark / Unknown	2020-03-09	Department of Virus and Microbiological Special Diagnostics, Statens Serum Institut, Copenhagen, Denmark, Artillerivej 5, 2300 Copenahgen S	Albertsen lab, Department of Chemistry and Bioscience, Aalborg University, Denmark	Rasmus Kirkegaard
EPI_ISL_429369	hCoV-19/Denmark/ALAB-SSI151/2020	Europe / Denmark / Unknown	2020-03-09	Department of Virus and Microbiological Special Diagnostics, Statens Serum Institut, Copenhagen, Denmark, Artillerivej 5, 2300 Copenahgen S	Albertsen lab, Department of Chemistry and Bioscience, Aalborg University, Denmark	Rasmus Kirkegaard
EPI_ISL_429395	hCoV-19/Denmark/ALAB-SSI194/2020	Europe / Denmark / Unknown	2020-03-09	Department of Virus and Microbiological Special Diagnostics, Statens Serum Institut, Copenhagen, Denmark, Artillerivej 5, 2300 Copenahgen S	Albertsen lab, Department of Chemistry and Bioscience, Aalborg University, Denmark	Rasmus Kirkegaard
EPI_ISL_429396	hCoV-19/Denmark/ALAB-SSI195/2020	Europe / Denmark / Unknown	2020-03-09	Department of Virus and Microbiological Special Diagnostics, Statens Serum Institut, Copenhagen, Denmark, Artillerivej 5, 2300 Copenahgen S	Albertsen lab, Department of Chemistry and Bioscience, Aalborg University, Denmark	Rasmus Kirkegaard
EPI_ISL_429399	hCoV-19/Denmark/ALAB-SSI201/2020	Europe / Denmark / Unknown	2020-03-09	Department of Virus and Microbiological Special Diagnostics, Statens Serum Institut, Copenhagen, Denmark, Artillerivej 5, 2300 Copenahgen S	Albertsen lab, Department of Chemistry and Bioscience, Aalborg University, Denmark	Rasmus Kirkegaard
EPI_ISL_429402	hCoV-19/Denmark/ALAB-SSI204/2020	Europe / Denmark / Unknown	2020-03-09	Department of Virus and Microbiological Special Diagnostics, Statens Serum Institut, Copenhagen, Denmark, Artillerivej 5, 2300 Copenahgen S	Albertsen lab, Department of Chemistry and Bioscience, Aalborg University, Denmark	Rasmus Kirkegaard
EPI_ISL_429404	hCoV-19/Denmark/ALAB-SSI207/2020	Europe / Denmark / Unknown	2020-03-10	Department of Virus and Microbiological Special Diagnostics, Statens Serum Institut, Copenhagen, Denmark, Artillerivej 5, 2300 Copenahgen S	Albertsen lab, Department of Chemistry and Bioscience, Aalborg University, Denmark	Rasmus Kirkegaard
EPI_ISL_429405	hCoV-19/Denmark/ALAB-SSI209/2020	Europe / Denmark / Unknown	2020-03-10	Department of Virus and Microbiological Special Diagnostics, Statens Serum Institut, Copenhagen, Denmark, Artillerivej 5, 2300 Copenahgen S	Albertsen lab, Department of Chemistry and Bioscience, Aalborg University, Denmark	Rasmus Kirkegaard
EPI_ISL_429406	hCoV-19/Denmark/ALAB-SSI213/2020	Europe / Denmark / Unknown	2020-03-11	Department of Virus and Microbiological Special Diagnostics, Statens Serum Institut, Copenhagen, Denmark, Artillerivej 5, 2300 Copenahgen S	Albertsen lab, Department of Chemistry and Bioscience, Aalborg University, Denmark	Rasmus Kirkegaard
EPI_ISL_429407	hCoV-19/Denmark/ALAB-SSI215/2020	Europe / Denmark / Unknown	2020-03-11	Department of Virus and Microbiological Special Diagnostics, Statens Serum Institut, Copenhagen, Denmark, Artillerivej 5, 2300 Copenahgen S	Albertsen lab, Department of Chemistry and Bioscience, Aalborg University, Denmark	Rasmus Kirkegaard
EPI_ISL_429408	hCoV-19/Denmark/ALAB-SSI216/2020	Europe / Denmark / Unknown	2020-03-10	Department of Virus and Microbiological Special Diagnostics, Statens Serum Institut, Copenhagen, Denmark, Artillerivej 5, 2300 Copenahgen S	Albertsen lab, Department of Chemistry and Bioscience, Aalborg University, Denmark	Rasmus Kirkegaard
EPI_ISL_429409	hCoV-19/Denmark/ALAB-SSI219/2020	Europe / Denmark / Unknown	2020-03-10	Department of Virus and Microbiological Special Diagnostics, Statens Serum Institut, Copenhagen, Denmark, Artillerivej 5, 2300 Copenahgen S	Albertsen lab, Department of Chemistry and Bioscience, Aalborg University, Denmark	Rasmus Kirkegaard
EPI_ISL_429410	hCoV-19/Denmark/ALAB-SSI221/2020	Europe / Denmark / Unknown	2020-03-10	Department of Virus and Microbiological Special Diagnostics, Statens Serum Institut, Copenhagen, Denmark, Artillerivej 5, 2300 Copenahgen S	Albertsen lab, Department of Chemistry and Bioscience, Aalborg University, Denmark	Rasmus Kirkegaard
EPI_ISL_429414	hCoV-19/Denmark/ALAB-SSI235/2020	Europe / Denmark / Unknown	2020-03-09	Department of Virus and Microbiological Special Diagnostics, Statens Serum Institut, Copenhagen, Denmark, Artillerivej 5, 2300 Copenahgen S	Albertsen lab, Department of Chemistry and Bioscience, Aalborg University, Denmark	Rasmus Kirkegaard
EPI_ISL_429420	hCoV-19/Denmark/ALAB-SSI258/2020	Europe / Denmark / Unknown	2020-03-10	Department of Virus and Microbiological Special Diagnostics, Statens Serum Institut, Copenhagen, Denmark, Artillerivej 5, 2300 Copenahgen S	Albertsen lab, Department of Chemistry and Bioscience, Aalborg University, Denmark	Rasmus Kirkegaard
EPI_ISL_429421	hCoV-19/Denmark/ALAB-SSI259/2020	Europe / Denmark / Unknown	2020-03-10	Department of Virus and Microbiological Special Diagnostics, Statens Serum Institut, Copenhagen, Denmark, Artillerivej 5, 2300 Copenahgen S	Albertsen lab, Department of Chemistry and Bioscience, Aalborg University, Denmark	Rasmus Kirkegaard
EPI_ISL_429424	hCoV-19/Denmark/ALAB-SSI262/2020	Europe / Denmark / Unknown	2020-03-10	Department of Virus and Microbiological Special Diagnostics, Statens Serum Institut, Copenhagen, Denmark, Artillerivej 5, 2300 Copenahgen S	Albertsen lab, Department of Chemistry and Bioscience, Aalborg

University, Denmark	Rasmus Kirkegaard
EPI_ISL_429425	hCoV-19/Denmark/ALAB-SSI263/2020	Europe / Denmark / Unknown	2020-03-10	Department of Virus and Microbiological Special Diagnostics, Statens Serum Institut, Copenhagen, Denmark, Artillerivej 5, 2300 Copenahgen S	Albertsen lab, Department of Chemistry and Bioscience, Aalborg University, Denmark	Rasmus Kirkegaard
EPI_ISL_429426	hCoV-19/Denmark/ALAB-SSI264/2020	Europe / Denmark / Unknown	2020-03-10	Department of Virus and Microbiological Special Diagnostics, Statens Serum Institut, Copenhagen, Denmark, Artillerivej 5, 2300 Copenahgen S	Albertsen lab, Department of Chemistry and Bioscience, Aalborg University, Denmark	Rasmus Kirkegaard
EPI_ISL_429427	hCoV-19/Denmark/ALAB-SSI265/2020	Europe / Denmark / Unknown	2020-03-10	Department of Virus and Microbiological Special Diagnostics, Statens Serum Institut, Copenhagen, Denmark, Artillerivej 5, 2300 Copenahgen S	Albertsen lab, Department of Chemistry and Bioscience, Aalborg University, Denmark	Rasmus Kirkegaard
EPI_ISL_429428	hCoV-19/Denmark/ALAB-SSI268/2020	Europe / Denmark / Unknown	2020-03-10	Department of Virus and Microbiological Special Diagnostics, Statens Serum Institut, Copenhagen, Denmark, Artillerivej 5, 2300 Copenahgen S	Albertsen lab, Department of Chemistry and Bioscience, Aalborg University, Denmark	Rasmus Kirkegaard
EPI_ISL_429431	hCoV-19/Denmark/ALAB-SSI271/2020	Europe / Denmark / Unknown	2020-03-10	Department of Virus and Microbiological Special Diagnostics, Statens Serum Institut, Copenhagen, Denmark, Artillerivej 5, 2300 Copenahgen S	Albertsen lab, Department of Chemistry and Bioscience, Aalborg University, Denmark	Rasmus Kirkegaard
EPI_ISL_429433	hCoV-19/Denmark/ALAB-SSI272/2020	Europe / Denmark / Unknown	2020-03-10	Department of Virus and Microbiological Special Diagnostics, Statens Serum Institut, Copenhagen, Denmark, Artillerivej 5, 2300 Copenahgen S	Albertsen lab, Department of Chemistry and Bioscience, Aalborg University, Denmark	Rasmus Kirkegaard
EPI_ISL_429434	hCoV-19/Denmark/ALAB-SSI273/2020	Europe / Denmark / Unknown	2020-03-10	Department of Virus and Microbiological Special Diagnostics, Statens Serum Institut, Copenhagen, Denmark, Artillerivej 5, 2300 Copenahgen S	Albertsen lab, Department of Chemistry and Bioscience, Aalborg University, Denmark	Rasmus Kirkegaard
EPI_ISL_429439	hCoV-19/Denmark/ALAB-SSI279/2020	Europe / Denmark / Unknown	2020-03-10	Department of Virus and Microbiological Special Diagnostics, Statens Serum Institut, Copenhagen, Denmark, Artillerivej 5, 2300 Copenahgen S	Albertsen lab, Department of Chemistry and Bioscience, Aalborg University, Denmark	Rasmus Kirkegaard
EPI_ISL_429440	hCoV-19/Denmark/ALAB-SSI280/2020	Europe / Denmark / Unknown	2020-03-10	Department of Virus and Microbiological Special Diagnostics, Statens Serum Institut, Copenhagen, Denmark, Artillerivej 5, 2300 Copenahgen S	Albertsen lab, Department of Chemistry and Bioscience, Aalborg University, Denmark	Rasmus Kirkegaard
EPI_ISL_429447	hCoV-19/Denmark/ALAB-SSI29/2020 Europe / Denmark / Unknown	2020-03-03	Department of Virus and Microbiological Special Diagnostics, Statens Serum Institut, Copenhagen, Denmark, Artillerivej 5, 2300 Copenahgen S	Albertsen lab, Department of Chemistry and Bioscience, Aalborg University, Denmark	Rasmus Kirkegaard
EPI_ISL_429448	hCoV-19/Denmark/ALAB-SSI293/2020	Europe / Denmark / Unknown	2020-03-10	Department of Virus and Microbiological Special Diagnostics, Statens Serum Institut, Copenhagen, Denmark, Artillerivej 5, 2300 Copenahgen S	Albertsen lab, Department of Chemistry and Bioscience, Aalborg University, Denmark	Rasmus Kirkegaard
EPI_ISL_429449	hCoV-19/Denmark/ALAB-SSI298/2020	Europe / Denmark / Unknown	2020-03-10	Department of Virus and Microbiological Special Diagnostics, Statens Serum Institut, Copenhagen, Denmark, Artillerivej 5, 2300 Copenahgen S	Albertsen lab, Department of Chemistry and Bioscience, Aalborg University, Denmark	Rasmus Kirkegaard
EPI_ISL_429450	hCoV-19/Denmark/ALAB-SSI299/2020	Europe / Denmark / Unknown	2020-03-10	Department of Virus and Microbiological Special Diagnostics, Statens Serum Institut, Copenhagen, Denmark, Artillerivej 5, 2300 Copenahgen S	Albertsen lab, Department of Chemistry and Bioscience, Aalborg University, Denmark	Rasmus Kirkegaard
EPI_ISL_429451	hCoV-19/Denmark/ALAB-SSI300/2020	Europe / Denmark / Unknown	2020-03-10	Department of Virus and Microbiological Special Diagnostics, Statens Serum Institut, Copenhagen, Denmark, Artillerivej 5, 2300 Copenahgen S	Albertsen lab, Department of Chemistry and Bioscience, Aalborg University, Denmark	Rasmus Kirkegaard
EPI_ISL_429452	hCoV-19/Denmark/ALAB-SSI301/2020	Europe / Denmark / Unknown	2020-03-10	Department of Virus and Microbiological Special Diagnostics, Statens Serum Institut, Copenhagen, Denmark, Artillerivej 5, 2300 Copenahgen S	Albertsen lab, Department of Chemistry and Bioscience, Aalborg University, Denmark	Rasmus Kirkegaard
EPI_ISL_429453	hCoV-19/Denmark/ALAB-SSI302/2020	Europe / Denmark / Unknown	2020-03-10	Department of Virus and Microbiological Special Diagnostics, Statens Serum Institut, Copenhagen, Denmark, Artillerivej 5, 2300 Copenahgen S	Albertsen lab, Department of Chemistry and Bioscience, Aalborg University, Denmark	Rasmus Kirkegaard
EPI_ISL_429460	hCoV-19/Denmark/ALAB-SSI346/2020	Europe / Denmark / Unknown	2020-03-23	Department of Virus and Microbiological Special Diagnostics, Statens Serum Institut, Copenhagen, Denmark, Artillerivej 5, 2300 Copenahgen S	Albertsen lab, Department of Chemistry and Bioscience, Aalborg University, Denmark	Rasmus Kirkegaard
EPI_ISL_429461	hCoV-19/Denmark/ALAB-SSI347/2020	Europe / Denmark / Unknown	2020-03-23

Department of Virus and Microbiological Special Diagnostics, Statens Serum Institut, Copenhagen, Denmark, Artillerivej 5, 2300 Copenahgen S	Albertsen lab, Department of Chemistry and Bioscience, Aalborg University, Denmark	Rasmus Kirkegaard
EPI_ISL_429463	hCoV-19/Denmark/ALAB-SSI349/2020	Europe / Denmark / Unknown	2020-03-23
Department of Virus and Microbiological Special Diagnostics, Statens Serum Institut, Copenhagen, Denmark, Artillerivej 5, 2300 Copenahgen S	Albertsen lab, Department of Chemistry and Bioscience, Aalborg University, Denmark	Rasmus Kirkegaard
EPI_ISL_429464	hCoV-19/Denmark/ALAB-SSI351/2020	Europe / Denmark / Unknown	2020-03-23
Department of Virus and Microbiological Special Diagnostics, Statens Serum Institut, Copenhagen, Denmark, Artillerivej 5, 2300 Copenahgen S	Albertsen lab, Department of Chemistry and Bioscience, Aalborg University, Denmark	Rasmus Kirkegaard
EPI_ISL_429465	hCoV-19/Denmark/ALAB-SSI352/2020	Europe / Denmark / Unknown	2020-03-23
Department of Virus and Microbiological Special Diagnostics, Statens Serum Institut, Copenhagen, Denmark, Artillerivej 5, 2300 Copenahgen S	Albertsen lab, Department of Chemistry and Bioscience, Aalborg University, Denmark	Rasmus Kirkegaard
EPI_ISL_429466	hCoV-19/Denmark/ALAB-SSI354/2020	Europe / Denmark / Unknown	2020-03-23
Department of Virus and Microbiological Special Diagnostics, Statens Serum Institut, Copenhagen, Denmark, Artillerivej 5, 2300 Copenahgen S	Albertsen lab, Department of Chemistry and Bioscience, Aalborg University, Denmark	Rasmus Kirkegaard
EPI_ISL_429468	hCoV-19/Denmark/ALAB-SSI357/2020	Europe / Denmark / Unknown	2020-03-23
Department of Virus and Microbiological Special Diagnostics, Statens Serum Institut, Copenhagen, Denmark, Artillerivej 5, 2300 Copenahgen S	Albertsen lab, Department of Chemistry and Bioscience, Aalborg University, Denmark	Rasmus Kirkegaard
EPI_ISL_429469	hCoV-19/Denmark/ALAB-SSI358/2020	Europe / Denmark / Unknown	2020-03-24
Department of Virus and Microbiological Special Diagnostics, Statens Serum Institut, Copenhagen, Denmark, Artillerivej 5, 2300 Copenahgen S	Albertsen lab, Department of Chemistry and Bioscience, Aalborg University, Denmark	Rasmus Kirkegaard
EPI_ISL_429471	hCoV-19/Denmark/ALAB-SSI361/2020	Europe / Denmark / Unknown	2020-03-23
Department of Virus and Microbiological Special Diagnostics, Statens Serum Institut, Copenhagen, Denmark, Artillerivej 5, 2300 Copenahgen S	Albertsen lab, Department of Chemistry and Bioscience, Aalborg University, Denmark	Rasmus Kirkegaard
EPI_ISL_429472	hCoV-19/Denmark/ALAB-SSI364/2020	Europe / Denmark / Unknown	2020-03-23
Department of Virus and Microbiological Special Diagnostics, Statens Serum Institut, Copenhagen, Denmark, Artillerivej 5, 2300 Copenahgen S	Albertsen lab, Department of Chemistry and Bioscience, Aalborg University, Denmark	Rasmus Kirkegaard
EPI_ISL_429473	hCoV-19/Denmark/ALAB-SSI365/2020	Europe / Denmark / Unknown	2020-03-23
Department of Virus and Microbiological Special Diagnostics, Statens Serum Institut, Copenhagen, Denmark, Artillerivej 5, 2300 Copenahgen S	Albertsen lab, Department of Chemistry and Bioscience, Aalborg University, Denmark	Rasmus Kirkegaard
EPI_ISL_429474	hCoV-19/Denmark/ALAB-SSI366/2020	Europe / Denmark / Unknown	2020-03-23
Department of Virus and Microbiological Special Diagnostics, Statens Serum Institut, Copenhagen, Denmark, Artillerivej 5, 2300 Copenahgen S	Albertsen lab, Department of Chemistry and Bioscience, Aalborg University, Denmark	Rasmus Kirkegaard
EPI_ISL_429476	hCoV-19/Denmark/ALAB-SSI370/2020	Europe / Denmark / Unknown	2020-03-23
Department of Virus and Microbiological Special Diagnostics, Statens Serum Institut, Copenhagen, Denmark, Artillerivej 5, 2300 Copenahgen S	Albertsen lab, Department of Chemistry and Bioscience, Aalborg University, Denmark	Rasmus Kirkegaard
EPI_ISL_429477	hCoV-19/Denmark/ALAB-SSI371/2020	Europe / Denmark / Unknown	2020-03-23
Department of Virus and Microbiological Special Diagnostics, Statens Serum Institut, Copenhagen, Denmark, Artillerivej 5, 2300 Copenahgen S	Albertsen lab, Department of Chemistry and Bioscience, Aalborg University, Denmark	Rasmus Kirkegaard
EPI_ISL_429478	hCoV-19/Denmark/ALAB-SSI372/2020	Europe / Denmark / Unknown	2020-03-23
Department of Virus and Microbiological Special Diagnostics, Statens Serum Institut, Copenhagen, Denmark, Artillerivej 5, 2300 Copenahgen S	Albertsen lab, Department of Chemistry and Bioscience, Aalborg University, Denmark	Rasmus Kirkegaard
EPI_ISL_429479	hCoV-19/Denmark/ALAB-SSI374/2020	Europe / Denmark / Unknown	2020-03-22
Department of Virus and Microbiological Special Diagnostics, Statens Serum Institut, Copenhagen, Denmark, Artillerivej 5, 2300 Copenahgen S	Albertsen lab, Department of Chemistry and Bioscience, Aalborg University, Denmark	Rasmus Kirkegaard
EPI_ISL_429480	hCoV-19/Denmark/ALAB-SSI375/2020	Europe / Denmark / Unknown	2020-03-22
Department of Virus and Microbiological Special Diagnostics, Statens Serum Institut, Copenhagen, Denmark, Artillerivej 5, 2300 Copenahgen S	Albertsen lab, Department of Chemistry and Bioscience, Aalborg University, Denmark	Rasmus Kirkegaard
EPI_ISL_429481	hCoV-19/Denmark/ALAB-SSI376/2020	Europe / Denmark / Unknown	2020-03-22
Department of Virus and Microbiological Special Diagnostics, Statens Serum Institut, Copenhagen, Denmark, Artillerivej 5, 2300 Copenahgen S	Albertsen lab, Department of Chemistry and Bioscience, Aalborg University, Denmark	Rasmus Kirkegaard
EPI_ISL_429482	hCoV-19/Denmark/ALAB-SSI377/2020	Europe / Denmark / Unknown	2020-03-22
Department of Virus and Microbiological Special Diagnostics, Statens Serum Institut, Copenhagen, Denmark, Artillerivej 5, 2300 Copenahgen S	Albertsen lab, Department of Chemistry and Bioscience, Aalborg

University, Denmark	Rasmus Kirkegaard
EPI_ISL_429483	hCoV-19/Denmark/ALAB-SSI383/2020	Europe / Denmark / Unknown	2020-03-24	Department of Virus and Microbiological Special Diagnostics, Statens Serum Institut, Copenhagen, Denmark, Artillerivej 5, 2300 Copenahgen S	Albertsen lab, Department of Chemistry and Bioscience, Aalborg University, Denmark	Rasmus Kirkegaard
EPI_ISL_429484	hCoV-19/Denmark/ALAB-SSI384/2020	Europe / Denmark / Unknown	2020-03-24	Department of Virus and Microbiological Special Diagnostics, Statens Serum Institut, Copenhagen, Denmark, Artillerivej 5, 2300 Copenahgen S	Albertsen lab, Department of Chemistry and Bioscience, Aalborg University, Denmark	Rasmus Kirkegaard
EPI_ISL_429485	hCoV-19/Denmark/ALAB-SSI385/2020	Europe / Denmark / Unknown	2020-03-24	Department of Virus and Microbiological Special Diagnostics, Statens Serum Institut, Copenhagen, Denmark, Artillerivej 5, 2300 Copenahgen S	Albertsen lab, Department of Chemistry and Bioscience, Aalborg University, Denmark	Rasmus Kirkegaard
EPI_ISL_429486	hCoV-19/Denmark/ALAB-SSI390/2020	Europe / Denmark / Unknown	2020-03-24	Department of Virus and Microbiological Special Diagnostics, Statens Serum Institut, Copenhagen, Denmark, Artillerivej 5, 2300 Copenahgen S	Albertsen lab, Department of Chemistry and Bioscience, Aalborg University, Denmark	Rasmus Kirkegaard
EPI_ISL_429488	hCoV-19/Denmark/ALAB-SSI393/2020	Europe / Denmark / Unknown	2020-03-24	Department of Virus and Microbiological Special Diagnostics, Statens Serum Institut, Copenhagen, Denmark, Artillerivej 5, 2300 Copenahgen S	Albertsen lab, Department of Chemistry and Bioscience, Aalborg University, Denmark	Rasmus Kirkegaard
EPI_ISL_429489	hCoV-19/Denmark/ALAB-SSI394/2020	Europe / Denmark / Unknown	2020-03-24	Department of Virus and Microbiological Special Diagnostics, Statens Serum Institut, Copenhagen, Denmark, Artillerivej 5, 2300 Copenahgen S	Albertsen lab, Department of Chemistry and Bioscience, Aalborg University, Denmark	Rasmus Kirkegaard
EPI_ISL_429490	hCoV-19/Denmark/ALAB-SSI395/2020	Europe / Denmark / Unknown	2020-03-23	Department of Virus and Microbiological Special Diagnostics, Statens Serum Institut, Copenhagen, Denmark, Artillerivej 5, 2300 Copenahgen S	Albertsen lab, Department of Chemistry and Bioscience, Aalborg University, Denmark	Rasmus Kirkegaard
EPI_ISL_429492	hCoV-19/Denmark/ALAB-SSI399/2020	Europe / Denmark / Unknown	2020-03-23	Department of Virus and Microbiological Special Diagnostics, Statens Serum Institut, Copenhagen, Denmark, Artillerivej 5, 2300 Copenahgen S	Albertsen lab, Department of Chemistry and Bioscience, Aalborg University, Denmark	Rasmus Kirkegaard
EPI_ISL_429493	hCoV-19/Denmark/ALAB-SSI400/2020	Europe / Denmark / Unknown	2020-03-23	Department of Virus and Microbiological Special Diagnostics, Statens Serum Institut, Copenhagen, Denmark, Artillerivej 5, 2300 Copenahgen S	Albertsen lab, Department of Chemistry and Bioscience, Aalborg University, Denmark	Rasmus Kirkegaard
EPI_ISL_429495	hCoV-19/Denmark/ALAB-SSI402/2020	Europe / Denmark / Unknown	2020-03-09	Department of Virus and Microbiological Special Diagnostics, Statens Serum Institut, Copenhagen, Denmark, Artillerivej 5, 2300 Copenahgen S	Albertsen lab, Department of Chemistry and Bioscience, Aalborg University, Denmark	Rasmus Kirkegaard
EPI_ISL_429496	hCoV-19/Denmark/ALAB-SSI403A/2020	Europe / Denmark / Unknown	2020-03-23	Department of Virus and Microbiological Special Diagnostics, Statens Serum Institut, Copenhagen, Denmark, Artillerivej 5, 2300 Copenahgen S	Albertsen lab, Department of Chemistry and Bioscience, Aalborg University, Denmark	Rasmus Kirkegaard
EPI_ISL_429497	hCoV-19/Denmark/ALAB-SSI404/2020	Europe / Denmark / Unknown	2020-03-08	Department of Virus and Microbiological Special Diagnostics, Statens Serum Institut, Copenhagen, Denmark, Artillerivej 5, 2300 Copenahgen S	Albertsen lab, Department of Chemistry and Bioscience, Aalborg University, Denmark	Rasmus Kirkegaard
EPI_ISL_429499	hCoV-19/Denmark/ALAB-SSI405/2020	Europe / Denmark / Unknown	2020-03-08	Department of Virus and Microbiological Special Diagnostics, Statens Serum Institut, Copenhagen, Denmark, Artillerivej 5, 2300 Copenahgen S	Albertsen lab, Department of Chemistry and Bioscience, Aalborg University, Denmark	Rasmus Kirkegaard
EPI_ISL_430323	hCoV-19/USA/NY-NYUMC296/2020	North America / USA / New York / Queens 2020-04-10	NYU Langone Health	Departments of Pathology and Medicine, New York University School of Medicine	Maria Aguero-Rosenfeld, Brendan Belovarac, Margaret Black, Ludovic Boytard, John Cadley, Paolo Cotzia, John Chen, Dacia Dimartino, Xiaojun Feng, Tatyana Gindin, Emily Guzman, Adriana Heguy, Megan Hogan, Emily Huang, George Jour, Lawrence H. Lin, Raven Luther, Andrew Lytle, Christian Marier, Matthew T. Maurano, Mark J. Mulligan, Peter Meyn, Raquel Ordonez Ciriza, Iman Osman, Jared Pinnell, Vanessa Raabe, Sitharam Ramaswami, Amy Rapkiewicz, Andre M. Ribeiro-dos-Santos, Marie Samanovic-Golden, Antonio Serrano, Guomiao Shen, Matija Snuderl, Theodore Vougiouklakis, Nick Vulpescu, Gael Westby, Paul Zappile, Yutong Zhang
EPI_ISL_430324	hCoV-19/USA/NY-NYUMC297/2020	North America / USA / New York / Westchester County	2020-04-10	NYU Langone Health	Departments of Pathology and Medicine, New York University School of Medicine	Maria Aguero-Rosenfeld, Brendan Belovarac, Margaret Black, Ludovic Boytard, John Cadley, Paolo Cotzia, John Chen, Dacia Dimartino, Xiaojun Feng, Tatyana Gindin, Emily Guzman, Adriana Heguy, Megan Hogan, Emily Huang, George Jour, Lawrence H. Lin, Raven Luther, Andrew Lytle, Christian Marier, Matthew T. Maurano, Mark J. Mulligan, Peter Meyn, Raquel Ordonez Ciriza, Iman Osman, Jared Pinnell, Vanessa Raabe, Sitharam Ramaswami, Amy Rapkiewicz, Andre M. Ribeiro-dos-Santos, Marie Samanovic-Golden, Antonio Serrano, Guomiao Shen, Matija Snuderl, Theodore Vougiouklakis, Nick Vulpescu, Gael Westby, Paul Zappile, Yutong Zhang
EPI_ISL_430325	hCoV-19/USA/NY-NYUMC298/2020	North America / USA / New York / Brooklyn	2020-04-08

NYU Langone Health	Departments of Pathology and Medicine, New York University School of Medicine	Maria Aguero-Rosenfeld, Brendan Belovarac, Margaret Black, Ludovic Boytard, John Cadley, Paolo Cotzia, John Chen, Dacia Dimartino, Xiaojun Feng, Tatyana Gindin, Emily Guzman, Adriana Heguy, Megan Hogan, Emily Huang, George Jour, Lawrence H. Lin, Raven Luther, Andrew Lytle, Christian Marier, Matthew T. Maurano, Mark J. Mulligan, Peter Meyn, Raquel Ordonez Ciriza, Iman Osman, Jared Pinnell, Vanessa Raabe, Sitharam Ramaswami, Amy Rapkiewicz, Andre M. Ribeiro-dos-Santos, Marie Samanovic-Golden, Antonio Serrano, Guomiao Shen, Matija Snuderl, Theodore Vougiouklakis, Nick Vulpescu, Gael Westby, Paul Zappile, Yutong Zhang
EPI_ISL_430326	hCoV-19/USA/NY-NYUMC299/2020	North America / USA / New York / Brooklyn	2020-03-30	NYU Langone Health	Departments of Pathology and Medicine, New York University School of Medicine	Maria Aguero-Rosenfeld, Brendan Belovarac, Margaret Black, Ludovic Boytard, John Cadley, Paolo Cotzia, John Chen, Dacia Dimartino, Xiaojun Feng, Tatyana Gindin, Emily Guzman, Adriana Heguy, Megan Hogan, Emily Huang, George Jour, Lawrence H. Lin, Raven Luther, Andrew Lytle, Christian Marier, Matthew T. Maurano, Mark J. Mulligan, Peter Meyn, Raquel Ordonez Ciriza, Iman Osman, Jared Pinnell, Vanessa Raabe, Sitharam Ramaswami, Amy Rapkiewicz, Andre M. Ribeiro-dos-Santos, Marie Samanovic-Golden, Antonio Serrano, Guomiao Shen, Matija Snuderl, Theodore Vougiouklakis, Nick Vulpescu, Gael Westby, Paul Zappile, Yutong Zhang
EPI_ISL_430327	hCoV-19/USA/NY-NYUMC300/2020	North America / USA / New York / Manhattan	2020-03-30	NYU Langone Health	Departments of Pathology and Medicine, New York University School of Medicine	Maria Aguero-Rosenfeld, Brendan Belovarac, Margaret Black, Ludovic Boytard, John Cadley, Paolo Cotzia, John Chen, Dacia Dimartino, Xiaojun Feng, Tatyana Gindin, Emily Guzman, Adriana Heguy, Megan Hogan, Emily Huang, George Jour, Lawrence H. Lin, Raven Luther, Andrew Lytle, Christian Marier, Matthew T. Maurano, Mark J. Mulligan, Peter Meyn, Raquel Ordonez Ciriza, Iman Osman, Jared Pinnell, Vanessa Raabe, Sitharam Ramaswami, Amy Rapkiewicz, Andre M. Ribeiro-dos-Santos, Marie Samanovic-Golden, Antonio Serrano, Guomiao Shen, Matija Snuderl, Theodore Vougiouklakis, Nick Vulpescu, Gael Westby, Paul Zappile, Yutong Zhang
EPI_ISL_430328	hCoV-19/USA/NY-NYUMC301/2020	North America / USA / New York / Brooklyn	2020-03-30	NYU Langone Health	Departments of Pathology and Medicine, New York University School of Medicine	Maria Aguero-Rosenfeld, Brendan Belovarac, Margaret Black, Ludovic Boytard, John Cadley, Paolo Cotzia, John Chen, Dacia Dimartino, Xiaojun Feng, Tatyana Gindin, Emily Guzman, Adriana Heguy, Megan Hogan, Emily Huang, George Jour, Lawrence H. Lin, Raven Luther, Andrew Lytle, Christian Marier, Matthew T. Maurano, Mark J. Mulligan, Peter Meyn, Raquel Ordonez Ciriza, Iman Osman, Jared Pinnell, Vanessa Raabe, Sitharam Ramaswami, Amy Rapkiewicz, Andre M. Ribeiro-dos-Santos, Marie Samanovic-Golden, Antonio Serrano, Guomiao Shen, Matija Snuderl, Theodore Vougiouklakis, Nick Vulpescu, Gael Westby, Paul Zappile, Yutong Zhang
EPI_ISL_430329	hCoV-19/USA/NY-NYUMC302/2020	North America / USA / New York / Staten Island	2020-03-30	NYU Langone Health	Departments of Pathology and Medicine, New York University School of Medicine	Maria Aguero-Rosenfeld, Brendan Belovarac, Margaret Black, Ludovic Boytard, John Cadley, Paolo Cotzia, John Chen, Dacia Dimartino, Xiaojun Feng, Tatyana Gindin, Emily Guzman, Adriana Heguy, Megan Hogan, Emily Huang, George Jour, Lawrence H. Lin, Raven Luther, Andrew Lytle, Christian Marier, Matthew T. Maurano, Mark J. Mulligan, Peter Meyn, Raquel Ordonez Ciriza, Iman Osman, Jared Pinnell, Vanessa Raabe, Sitharam Ramaswami, Amy Rapkiewicz, Andre M. Ribeiro-dos-Santos, Marie Samanovic-Golden, Antonio Serrano, Guomiao Shen, Matija Snuderl, Theodore Vougiouklakis, Nick Vulpescu, Gael Westby, Paul Zappile, Yutong Zhang
EPI_ISL_430330	hCoV-19/USA/NY-NYUMC303/2020	North America / USA / New York / Nassau County	2020-03-30	NYU Langone Health	Departments of Pathology and Medicine, New York University School of Medicine	Maria Aguero-Rosenfeld, Brendan Belovarac, Margaret Black, Ludovic Boytard, John Cadley, Paolo Cotzia, John Chen, Dacia Dimartino, Xiaojun Feng, Tatyana Gindin, Emily Guzman, Adriana Heguy, Megan Hogan, Emily Huang, George Jour, Lawrence H. Lin, Raven Luther, Andrew Lytle, Christian Marier, Matthew T. Maurano, Mark J. Mulligan, Peter Meyn, Raquel Ordonez Ciriza, Iman Osman, Jared Pinnell, Vanessa Raabe, Sitharam Ramaswami, Amy Rapkiewicz, Andre M. Ribeiro-dos-Santos, Marie Samanovic-Golden, Antonio Serrano, Guomiao Shen, Matija Snuderl, Theodore Vougiouklakis, Nick Vulpescu, Gael Westby, Paul Zappile, Yutong Zhang
EPI_ISL_430331	hCoV-19/USA/NY-NYUMC304/2020	North America / USA / New York / Manhattan	2020-03-30	NYU Langone Health	Departments of Pathology and Medicine, New York University School of Medicine	Maria Aguero-Rosenfeld, Brendan Belovarac, Margaret Black, Ludovic Boytard, John Cadley, Paolo Cotzia, John Chen, Dacia Dimartino, Xiaojun Feng, Tatyana Gindin, Emily Guzman, Adriana Heguy, Megan Hogan, Emily Huang, George Jour, Lawrence H. Lin, Raven Luther, Andrew Lytle, Christian Marier, Matthew T. Maurano, Mark J. Mulligan, Peter Meyn, Raquel Ordonez Ciriza, Iman Osman, Jared Pinnell, Vanessa Raabe, Sitharam Ramaswami, Amy Rapkiewicz, Andre M. Ribeiro-dos-Santos, Marie Samanovic-Golden, Antonio Serrano, Guomiao Shen, Matija Snuderl, Theodore Vougiouklakis, Nick Vulpescu, Gael Westby, Paul Zappile, Yutong Zhang
EPI_ISL_430332	hCoV-19/USA/NY-NYUMC305/2020	North America / USA / New York / Brooklyn	2020-03-30	NYU Langone Health	Departments of Pathology and Medicine, New York University School of Medicine	Maria Aguero-Rosenfeld, Brendan Belovarac, Margaret Black, Ludovic Boytard, John Cadley, Paolo Cotzia, John Chen, Dacia Dimartino, Xiaojun Feng, Tatyana Gindin, Emily Guzman, Adriana Heguy, Megan Hogan, Emily Huang, George Jour, Lawrence H. Lin, Raven Luther, Andrew Lytle, Christian Marier, Matthew T. Maurano, Mark J. Mulligan, Peter Meyn, Raquel Ordonez Ciriza, Iman Osman, Jared Pinnell, Vanessa Raabe, Sitharam Ramaswami, Amy Rapkiewicz, Andre M. Ribeiro-dos-Santos, Marie Samanovic-Golden, Antonio Serrano, Guomiao Shen, Matija Snuderl, Theodore Vougiouklakis, Nick Vulpescu, Gael Westby, Paul Zappile, Yutong Zhang
EPI_ISL_430340	hCoV-19/USA/NY-NYUMC313/2020	North America / USA / New York / Westchester County	2020-03-30	NYU Langone Health	Departments of Pathology and Medicine, New York University School of Medicine	Maria Aguero-Rosenfeld, Brendan Belovarac, Margaret Black, Ludovic Boytard, John Cadley, Paolo Cotzia, John Chen, Dacia Dimartino, Xiaojun Feng, Tatyana Gindin, Emily Guzman, Adriana Heguy, Megan Hogan, Emily Huang, George Jour, Lawrence H. Lin, Raven Luther, Andrew Lytle, Christian Marier, Matthew T. Maurano, Mark J. Mulligan, Peter Meyn, Raquel Ordonez Ciriza, Iman Osman, Jared Pinnell, Vanessa Raabe, Sitharam Ramaswami, Amy Rapkiewicz, Andre M. Ribeiro-dos-Santos, Marie Samanovic-Golden, Antonio Serrano, Guomiao Shen, Matija Snuderl,

Theodore Vougiouklakis, Nick Vulpescu, Gael Westby, Paul Zappile, Yutong Zhang
EPI_ISL_430341 hCoV-19/USA/NY-NYUMC314/2020 North America / USA / New York / Brooklyn 2020-03-30 NYU Langone Health Departments of Pathology and Medicine, New York University School of Medicine Maria Aguero-Rosenfeld, Brendan Belovarac, Margaret Black, Ludovic Boytard, John Cadley, Paolo Cotzia, John Chen, Dacia Dimartino, Xiaojun Feng, Tatyana Gindin, Emily Guzman, Adriana Heguy, Megan Hogan, Emily Huang, George Jour, Lawrence H. Lin, Raven Luther, Andrew Lytle, Christian Marier, Matthew T. Maurano, Mark J. Mulligan, Peter Meyn, Raquel Ordonez Ciriza, Iman Osman, Jared Pinnell, Vanessa Raabe, Sitharam Ramaswami, Amy Rapkiewicz, Andre M. Ribeiro-dos-Santos, Marie Samanovic-Golden, Antonio Serrano, Guomiao Shen, Matija Snuderl, Theodore Vougiouklakis, Nick Vulpescu, Gael Westby, Paul Zappile, Yutong Zhang
EPI_ISL_430342 hCoV-19/USA/NY-NYUMC315/2020 North America / USA / New York / Queens 2020-03-30 NYU Langone Health Departments of Pathology and Medicine, New York University School of Medicine Maria Aguero-Rosenfeld, Brendan Belovarac, Margaret Black, Ludovic Boytard, John Cadley, Paolo Cotzia, John Chen, Dacia Dimartino, Xiaojun Feng, Tatyana Gindin, Emily Guzman, Adriana Heguy, Megan Hogan, Emily Huang, George Jour, Lawrence H. Lin, Raven Luther, Andrew Lytle, Christian Marier, Matthew T. Maurano, Mark J. Mulligan, Peter Meyn, Raquel Ordonez Ciriza, Iman Osman, Jared Pinnell, Vanessa Raabe, Sitharam Ramaswami, Amy Rapkiewicz, Andre M. Ribeiro-dos-Santos, Marie Samanovic-Golden, Antonio Serrano, Guomiao Shen, Matija Snuderl, Theodore Vougiouklakis, Nick Vulpescu, Gael Westby, Paul Zappile, Yutong Zhang
EPI_ISL_430344 hCoV-19/USA/NY-NYUMC317/2020 North America / USA / New York / Brooklyn 2020-03-30 NYU Langone Health Departments of Pathology and Medicine, New York University School of Medicine Maria Aguero-Rosenfeld, Brendan Belovarac, Margaret Black, Ludovic Boytard, John Cadley, Paolo Cotzia, John Chen, Dacia Dimartino, Xiaojun Feng, Tatyana Gindin, Emily Guzman, Adriana Heguy, Megan Hogan, Emily Huang, George Jour, Lawrence H. Lin, Raven Luther, Andrew Lytle, Christian Marier, Matthew T. Maurano, Mark J. Mulligan, Peter Meyn, Raquel Ordonez Ciriza, Iman Osman, Jared Pinnell, Vanessa Raabe, Sitharam Ramaswami, Amy Rapkiewicz, Andre M. Ribeiro-dos-Santos, Marie Samanovic-Golden, Antonio Serrano, Guomiao Shen, Matija Snuderl, Theodore Vougiouklakis, Nick Vulpescu, Gael Westby, Paul Zappile, Yutong Zhang
EPI_ISL_430345 hCoV-19/USA/NY-NYUMC318/2020 North America / USA / New York / Manhattan 2020-03-30 NYU Langone Health Departments of Pathology and Medicine, New York University School of Medicine Maria Aguero-Rosenfeld, Brendan Belovarac, Margaret Black, Ludovic Boytard, John Cadley, Paolo Cotzia, John Chen, Dacia Dimartino, Xiaojun Feng, Tatyana Gindin, Emily Guzman, Adriana Heguy, Megan Hogan, Emily Huang, George Jour, Lawrence H. Lin, Raven Luther, Andrew Lytle, Christian Marier, Matthew T. Maurano, Mark J. Mulligan, Peter Meyn, Raquel Ordonez Ciriza, Iman Osman, Jared Pinnell, Vanessa Raabe, Sitharam Ramaswami, Amy Rapkiewicz, Andre M. Ribeiro-dos-Santos, Marie Samanovic-Golden, Antonio Serrano, Guomiao Shen, Matija Snuderl, Theodore Vougiouklakis, Nick Vulpescu, Gael Westby, Paul Zappile, Yutong Zhang
EPI_ISL_430346 hCoV-19/USA/NY-NYUMC319/2020 North America / USA / New York / Brooklyn 2020-03-30 NYU Langone Health Departments of Pathology and Medicine, New York University School of Medicine Maria Aguero-Rosenfeld, Brendan Belovarac, Margaret Black, Ludovic Boytard, John Cadley, Paolo Cotzia, John Chen, Dacia Dimartino, Xiaojun Feng, Tatyana Gindin, Emily Guzman, Adriana Heguy, Megan Hogan, Emily Huang, George Jour, Lawrence H. Lin, Raven Luther, Andrew Lytle, Christian Marier, Matthew T. Maurano, Mark J. Mulligan, Peter Meyn, Raquel Ordonez Ciriza, Iman Osman, Jared Pinnell, Vanessa Raabe, Sitharam Ramaswami, Amy Rapkiewicz, Andre M. Ribeiro-dos-Santos, Marie Samanovic-Golden, Antonio Serrano, Guomiao Shen, Matija Snuderl, Theodore Vougiouklakis, Nick Vulpescu, Gael Westby, Paul Zappile, Yutong Zhang
EPI_ISL_430347 hCoV-19/USA/NY-NYUMC320/2020 North America / USA / New York / Manhattan 2020-03-30 NYU Langone Health Departments of Pathology and Medicine, New York University School of Medicine Maria Aguero-Rosenfeld, Brendan Belovarac, Margaret Black, Ludovic Boytard, John Cadley, Paolo Cotzia, John Chen, Dacia Dimartino, Xiaojun Feng, Tatyana Gindin, Emily Guzman, Adriana Heguy, Megan Hogan, Emily Huang, George Jour, Lawrence H. Lin, Raven Luther, Andrew Lytle, Christian Marier, Matthew T. Maurano, Mark J. Mulligan, Peter Meyn, Raquel Ordonez Ciriza, Iman Osman, Jared Pinnell, Vanessa Raabe, Sitharam Ramaswami, Amy Rapkiewicz, Andre M. Ribeiro-dos-Santos, Marie Samanovic-Golden, Antonio Serrano, Guomiao Shen, Matija Snuderl, Theodore Vougiouklakis, Nick Vulpescu, Gael Westby, Paul Zappile, Yutong Zhang
EPI_ISL_430348 hCoV-19/USA/NY-NYUMC321/2020 North America / USA / New York / Manhattan 2020-03-30 NYU Langone Health Departments of Pathology and Medicine, New York University School of Medicine Maria Aguero-Rosenfeld, Brendan Belovarac, Margaret Black, Ludovic Boytard, John Cadley, Paolo Cotzia, John Chen, Dacia Dimartino, Xiaojun Feng, Tatyana Gindin, Emily Guzman, Adriana Heguy, Megan Hogan, Emily Huang, George Jour, Lawrence H. Lin, Raven Luther, Andrew Lytle, Christian Marier, Matthew T. Maurano, Mark J. Mulligan, Peter Meyn, Raquel Ordonez Ciriza, Iman Osman, Jared Pinnell, Vanessa Raabe, Sitharam Ramaswami, Amy Rapkiewicz, Andre M. Ribeiro-dos-Santos, Marie Samanovic-Golden, Antonio Serrano, Guomiao Shen, Matija Snuderl, Theodore Vougiouklakis, Nick Vulpescu, Gael Westby, Paul Zappile, Yutong Zhang
EPI_ISL_430349 hCoV-19/USA/NY-NYUMC322/2020 North America / USA / New York / Staten Island 2020-03-30 NYU Langone Health Departments of Pathology and Medicine, New York University School of Medicine Maria Aguero-Rosenfeld, Brendan Belovarac, Margaret Black, Ludovic Boytard, John Cadley, Paolo Cotzia, John Chen, Dacia Dimartino, Xiaojun Feng, Tatyana Gindin, Emily Guzman, Adriana Heguy, Megan Hogan, Emily Huang, George Jour, Lawrence H. Lin, Raven Luther, Andrew Lytle, Christian Marier, Matthew T. Maurano, Mark J. Mulligan, Peter Meyn, Raquel Ordonez Ciriza, Iman Osman, Jared Pinnell, Vanessa Raabe, Sitharam Ramaswami, Amy Rapkiewicz, Andre M. Ribeiro-dos-Santos, Marie Samanovic-Golden, Antonio Serrano, Guomiao Shen, Matija Snuderl, Theodore Vougiouklakis, Nick Vulpescu, Gael Westby, Paul Zappile, Yutong Zhang
EPI_ISL_430350 hCoV-19/USA/NY-NYUMC323/2020 North America / USA / New York / Queens 2020-04-02 NYU Langone Health Departments of Pathology and Medicine, New York University School of Medicine Maria Aguero-Rosenfeld, Brendan Belovarac, Margaret Black, Ludovic Boytard, John Cadley, Paolo Cotzia, John Chen, Dacia Dimartino, Xiaojun Feng, Tatyana Gindin, Emily Guzman, Adriana Heguy, Megan Hogan, Emily Huang, George Jour, Lawrence H. Lin, Raven Luther, Andrew Lytle, Christian Marier, Matthew T. Maurano, Mark J. Mulligan, Peter

Meyn, Raquel Ordonez Ciriza, Iman Osman, Jared Pinnell, Vanessa Raabe, Sitharam Ramaswami, Amy Rapkiewicz, Andre M. Ribeiro-dos-Santos, Marie Samanovic-Golden, Antonio Serrano, Guomiao Shen, Matija Snuderl, Theodore Vougiouklakis, Nick Vulpescu, Gael Westby, Paul Zappile, Yutong Zhang
EPI_ISL_430351 hCoV-19/USA/NY-NYUMC324/2020 North America / USA / New York / Brooklyn 2020-03-31 NYU Langone Health Departments of Pathology and Medicine, New York University School of Medicine Maria Aguero-Rosenfeld, Brendan Belovarac, Margaret Black, Ludovic Boytard, John Cadley, Paolo Cotzia, John Chen, Dacia Dimartino, Xiaojun Feng, Tatyana Gindin, Emily Guzman, Adriana Heguy, Megan Hogan, Emily Huang, George Jour, Lawrence H. Lin, Raven Luther, Andrew Lytle, Christian Marier, Matthew T. Maurano, Mark J. Mulligan, Peter Meyn, Raquel Ordonez Ciriza, Iman Osman, Jared Pinnell, Vanessa Raabe, Sitharam Ramaswami, Amy Rapkiewicz, Andre M. Ribeiro-dos-Santos, Marie Samanovic-Golden, Antonio Serrano, Guomiao Shen, Matija Snuderl, Theodore Vougiouklakis, Nick Vulpescu, Gael Westby, Paul Zappile, Yutong Zhang
EPI_ISL_430352 hCoV-19/USA/NY-NYUMC325/2020 North America / USA / New York / Suffolk County 2020-04-01 NYU Langone Health Departments of Pathology and Medicine, New York University School of Medicine Maria Aguero-Rosenfeld, Brendan Belovarac, Margaret Black, Ludovic Boytard, John Cadley, Paolo Cotzia, John Chen, Dacia Dimartino, Xiaojun Feng, Tatyana Gindin, Emily Guzman, Adriana Heguy, Megan Hogan, Emily Huang, George Jour, Lawrence H. Lin, Raven Luther, Andrew Lytle, Christian Marier, Matthew T. Maurano, Mark J. Mulligan, Peter Meyn, Raquel Ordonez Ciriza, Iman Osman, Jared Pinnell, Vanessa Raabe, Sitharam Ramaswami, Amy Rapkiewicz, Andre M. Ribeiro-dos-Santos, Marie Samanovic-Golden, Antonio Serrano, Guomiao Shen, Matija Snuderl, Theodore Vougiouklakis, Nick Vulpescu, Gael Westby, Paul Zappile, Yutong Zhang
EPI_ISL_430353 hCoV-19/USA/NY-NYUMC326/2020 North America / USA / New York / Brooklyn 2020-04-01 NYU Langone Health Departments of Pathology and Medicine, New York University School of Medicine Maria Aguero-Rosenfeld, Brendan Belovarac, Margaret Black, Ludovic Boytard, John Cadley, Paolo Cotzia, John Chen, Dacia Dimartino, Xiaojun Feng, Tatyana Gindin, Emily Guzman, Adriana Heguy, Megan Hogan, Emily Huang, George Jour, Lawrence H. Lin, Raven Luther, Andrew Lytle, Christian Marier, Matthew T. Maurano, Mark J. Mulligan, Peter Meyn, Raquel Ordonez Ciriza, Iman Osman, Jared Pinnell, Vanessa Raabe, Sitharam Ramaswami, Amy Rapkiewicz, Andre M. Ribeiro-dos-Santos, Marie Samanovic-Golden, Antonio Serrano, Guomiao Shen, Matija Snuderl, Theodore Vougiouklakis, Nick Vulpescu, Gael Westby, Paul Zappile, Yutong Zhang
EPI_ISL_430354 hCoV-19/USA/NY-NYUMC327/2020 North America / USA / New York / Nassau County 2020-04-03 NYU Langone Health Departments of Pathology and Medicine, New York University School of Medicine Maria Aguero-Rosenfeld, Brendan Belovarac, Margaret Black, Ludovic Boytard, John Cadley, Paolo Cotzia, John Chen, Dacia Dimartino, Xiaojun Feng, Tatyana Gindin, Emily Guzman, Adriana Heguy, Megan Hogan, Emily Huang, George Jour, Lawrence H. Lin, Raven Luther, Andrew Lytle, Christian Marier, Matthew T. Maurano, Mark J. Mulligan, Peter Meyn, Raquel Ordonez Ciriza, Iman Osman, Jared Pinnell, Vanessa Raabe, Sitharam Ramaswami, Amy Rapkiewicz, Andre M. Ribeiro-dos-Santos, Marie Samanovic-Golden, Antonio Serrano, Guomiao Shen, Matija Snuderl, Theodore Vougiouklakis, Nick Vulpescu, Gael Westby, Paul Zappile, Yutong Zhang
EPI_ISL_430377 hCoV-19/USA/NY-NYUMC350/2020 North America / USA / New York / Brooklyn 2020-04-08 NYU Langone Health Departments of Pathology and Medicine, New York University School of Medicine Maria Aguero-Rosenfeld, Brendan Belovarac, Margaret Black, Ludovic Boytard, John Cadley, Paolo Cotzia, John Chen, Dacia Dimartino, Xiaojun Feng, Tatyana Gindin, Emily Guzman, Adriana Heguy, Megan Hogan, Emily Huang, George Jour, Lawrence H. Lin, Raven Luther, Andrew Lytle, Christian Marier, Matthew T. Maurano, Mark J. Mulligan, Peter Meyn, Raquel Ordonez Ciriza, Iman Osman, Jared Pinnell, Vanessa Raabe, Sitharam Ramaswami, Amy Rapkiewicz, Andre M. Ribeiro-dos-Santos, Marie Samanovic-Golden, Antonio Serrano, Guomiao Shen, Matija Snuderl, Theodore Vougiouklakis, Nick Vulpescu, Gael Westby, Paul Zappile, Yutong Zhang
EPI_ISL_430378 hCoV-19/USA/NY-NYUMC351/2020 North America / USA / New York / Brooklyn 2020-04-07 NYU Langone Health Departments of Pathology and Medicine, New York University School of Medicine Maria Aguero-Rosenfeld, Brendan Belovarac, Margaret Black, Ludovic Boytard, John Cadley, Paolo Cotzia, John Chen, Dacia Dimartino, Xiaojun Feng, Tatyana Gindin, Emily Guzman, Adriana Heguy, Megan Hogan, Emily Huang, George Jour, Lawrence H. Lin, Raven Luther, Andrew Lytle, Christian Marier, Matthew T. Maurano, Mark J. Mulligan, Peter Meyn, Raquel Ordonez Ciriza, Iman Osman, Jared Pinnell, Vanessa Raabe, Sitharam Ramaswami, Amy Rapkiewicz, Andre M. Ribeiro-dos-Santos, Marie Samanovic-Golden, Antonio Serrano, Guomiao Shen, Matija Snuderl, Theodore Vougiouklakis, Nick Vulpescu, Gael Westby, Paul Zappile, Yutong Zhang
EPI_ISL_430379 hCoV-19/USA/NY-NYUMC352/2020 North America / USA / New York / Brooklyn 2020-04-07 NYU Langone Health Departments of Pathology and Medicine, New York University School of Medicine Maria Aguero-Rosenfeld, Brendan Belovarac, Margaret Black, Ludovic Boytard, John Cadley, Paolo Cotzia, John Chen, Dacia Dimartino, Xiaojun Feng, Tatyana Gindin, Emily Guzman, Adriana Heguy, Megan Hogan, Emily Huang, George Jour, Lawrence H. Lin, Raven Luther, Andrew Lytle, Christian Marier, Matthew T. Maurano, Mark J. Mulligan, Peter Meyn, Raquel Ordonez Ciriza, Iman Osman, Jared Pinnell, Vanessa Raabe, Sitharam Ramaswami, Amy Rapkiewicz, Andre M. Ribeiro-dos-Santos, Marie Samanovic-Golden, Antonio Serrano, Guomiao Shen, Matija Snuderl, Theodore Vougiouklakis, Nick Vulpescu, Gael Westby, Paul Zappile, Yutong Zhang
EPI_ISL_430380 hCoV-19/USA/NY-NYUMC353/2020 North America / USA / New York / Queens 2020-04-08 NYU Langone Health Departments of Pathology and Medicine, New York University School of Medicine Maria Aguero-Rosenfeld, Brendan Belovarac, Margaret Black, Ludovic Boytard, John Cadley, Paolo Cotzia, John Chen, Dacia Dimartino, Xiaojun Feng, Tatyana Gindin, Emily Guzman, Adriana Heguy, Megan Hogan, Emily Huang, George Jour, Lawrence H. Lin, Raven Luther, Andrew Lytle, Christian Marier, Matthew T. Maurano, Mark J. Mulligan, Peter Meyn, Raquel Ordonez Ciriza, Iman Osman, Jared Pinnell, Vanessa Raabe, Sitharam Ramaswami, Amy Rapkiewicz, Andre M. Ribeiro-dos-Santos, Marie Samanovic-Golden, Antonio Serrano, Guomiao Shen, Matija Snuderl, Theodore Vougiouklakis, Nick Vulpescu, Gael Westby, Paul Zappile, Yutong Zhang
EPI_ISL_430381 hCoV-19/USA/NY-NYUMC354/2020 North America / USA / New York / Brooklyn 2020-04-07 NYU Langone Health Departments of Pathology and Medicine, New York University School of Medicine Maria Aguero-Rosenfeld, Brendan Belovarac, Margaret Black, Ludovic Boytard, John Cadley, Paolo Cotzia, John Chen,

Dacia Dimartino, Xiaojun Feng, Tatyana Gindin, Emily Guzman, Adriana Heguy, Megan Hogan, Emily Huang, George Jour, Lawrence H. Lin, Raven Luther, Andrew Lytle, Christian Marier, Matthew T. Maurano, Mark J. Mulligan, Peter Meyn, Raquel Ordonez Ciriza, Iman Osman, Jared Pinnell, Vanessa Raabe, Sitharam Ramaswami, Amy Rapkiewicz, Andre M. Ribeiro-dos-Santos, Marie Samanovic-Golden, Antonio Serrano, Guomiao Shen, Matija Snuderl, Theodore Vougiouklakis, Nick Vulpescu, Gael Westby, Paul Zappile, Yutong Zhang
EPI_ISL_430382 hCoV-19/USA/NY-NYUMC355/2020 North America / USA / New York / Brooklyn 2020-04-09 NYU Langone Health Departments of Pathology and Medicine, New York University School of Medicine Maria Aguero-Rosenfeld, Brendan Belovarac, Margaret Black, Ludovic Boytard, John Cadley, Paolo Cotzia, John Chen, Dacia Dimartino, Xiaojun Feng, Tatyana Gindin, Emily Guzman, Adriana Heguy, Megan Hogan, Emily Huang, George Jour, Lawrence H. Lin, Raven Luther, Andrew Lytle, Christian Marier, Matthew T. Maurano, Mark J. Mulligan, Peter Meyn, Raquel Ordonez Ciriza, Iman Osman, Jared Pinnell, Vanessa Raabe, Sitharam Ramaswami, Amy Rapkiewicz, Andre M. Ribeiro-dos-Santos, Marie Samanovic-Golden, Antonio Serrano, Guomiao Shen, Matija Snuderl, Theodore Vougiouklakis, Nick Vulpescu, Gael Westby, Paul Zappile, Yutong Zhang
EPI_ISL_430383 hCoV-19/USA/NY-NYUMC356/2020 North America / USA / New York / Brooklyn 2020-04-08 NYU Langone Health Departments of Pathology and Medicine, New York University School of Medicine Maria Aguero-Rosenfeld, Brendan Belovarac, Margaret Black, Ludovic Boytard, John Cadley, Paolo Cotzia, John Chen, Dacia Dimartino, Xiaojun Feng, Tatyana Gindin, Emily Guzman, Adriana Heguy, Megan Hogan, Emily Huang, George Jour, Lawrence H. Lin, Raven Luther, Andrew Lytle, Christian Marier, Matthew T. Maurano, Mark J. Mulligan, Peter Meyn, Raquel Ordonez Ciriza, Iman Osman, Jared Pinnell, Vanessa Raabe, Sitharam Ramaswami, Amy Rapkiewicz, Andre M. Ribeiro-dos-Santos, Marie Samanovic-Golden, Antonio Serrano, Guomiao Shen, Matija Snuderl, Theodore Vougiouklakis, Nick Vulpescu, Gael Westby, Paul Zappile, Yutong Zhang
EPI_ISL_430384 hCoV-19/USA/NY-NYUMC357/2020 North America / USA / New York / Brooklyn 2020-04-09 NYU Langone Health Departments of Pathology and Medicine, New York University School of Medicine Maria Aguero-Rosenfeld, Brendan Belovarac, Margaret Black, Ludovic Boytard, John Cadley, Paolo Cotzia, John Chen, Dacia Dimartino, Xiaojun Feng, Tatyana Gindin, Emily Guzman, Adriana Heguy, Megan Hogan, Emily Huang, George Jour, Lawrence H. Lin, Raven Luther, Andrew Lytle, Christian Marier, Matthew T. Maurano, Mark J. Mulligan, Peter Meyn, Raquel Ordonez Ciriza, Iman Osman, Jared Pinnell, Vanessa Raabe, Sitharam Ramaswami, Amy Rapkiewicz, Andre M. Ribeiro-dos-Santos, Marie Samanovic-Golden, Antonio Serrano, Guomiao Shen, Matija Snuderl, Theodore Vougiouklakis, Nick Vulpescu, Gael Westby, Paul Zappile, Yutong Zhang
EPI_ISL_430385 hCoV-19/USA/NY-NYUMC358/2020 North America / USA / New York / Brooklyn 2020-04-08 NYU Langone Health Departments of Pathology and Medicine, New York University School of Medicine Maria Aguero-Rosenfeld, Brendan Belovarac, Margaret Black, Ludovic Boytard, John Cadley, Paolo Cotzia, John Chen, Dacia Dimartino, Xiaojun Feng, Tatyana Gindin, Emily Guzman, Adriana Heguy, Megan Hogan, Emily Huang, George Jour, Lawrence H. Lin, Raven Luther, Andrew Lytle, Christian Marier, Matthew T. Maurano, Mark J. Mulligan, Peter Meyn, Raquel Ordonez Ciriza, Iman Osman, Jared Pinnell, Vanessa Raabe, Sitharam Ramaswami, Amy Rapkiewicz, Andre M. Ribeiro-dos-Santos, Marie Samanovic-Golden, Antonio Serrano, Guomiao Shen, Matija Snuderl, Theodore Vougiouklakis, Nick Vulpescu, Gael Westby, Paul Zappile, Yutong Zhang
EPI_ISL_430386 hCoV-19/USA/NY-NYUMC359/2020 North America / USA / New York / Brooklyn 2020-04-08 NYU Langone Health Departments of Pathology and Medicine, New York University School of Medicine Maria Aguero-Rosenfeld, Brendan Belovarac, Margaret Black, Ludovic Boytard, John Cadley, Paolo Cotzia, John Chen, Dacia Dimartino, Xiaojun Feng, Tatyana Gindin, Emily Guzman, Adriana Heguy, Megan Hogan, Emily Huang, George Jour, Lawrence H. Lin, Raven Luther, Andrew Lytle, Christian Marier, Matthew T. Maurano, Mark J. Mulligan, Peter Meyn, Raquel Ordonez Ciriza, Iman Osman, Jared Pinnell, Vanessa Raabe, Sitharam Ramaswami, Amy Rapkiewicz, Andre M. Ribeiro-dos-Santos, Marie Samanovic-Golden, Antonio Serrano, Guomiao Shen, Matija Snuderl, Theodore Vougiouklakis, Nick Vulpescu, Gael Westby, Paul Zappile, Yutong Zhang
EPI_ISL_430387 hCoV-19/USA/NY-NYUMC360/2020 North America / USA / New York / Brooklyn 2020-04-08 NYU Langone Health Departments of Pathology and Medicine, New York University School of Medicine Maria Aguero-Rosenfeld, Brendan Belovarac, Margaret Black, Ludovic Boytard, John Cadley, Paolo Cotzia, John Chen, Dacia Dimartino, Xiaojun Feng, Tatyana Gindin, Emily Guzman, Adriana Heguy, Megan Hogan, Emily Huang, George Jour, Lawrence H. Lin, Raven Luther, Andrew Lytle, Christian Marier, Matthew T. Maurano, Mark J. Mulligan, Peter Meyn, Raquel Ordonez Ciriza, Iman Osman, Jared Pinnell, Vanessa Raabe, Sitharam Ramaswami, Amy Rapkiewicz, Andre M. Ribeiro-dos-Santos, Marie Samanovic-Golden, Antonio Serrano, Guomiao Shen, Matija Snuderl, Theodore Vougiouklakis, Nick Vulpescu, Gael Westby, Paul Zappile, Yutong Zhang
EPI_ISL_430388 hCoV-19/USA/NY-NYUMC361/2020 North America / USA / New York / Queens 2020-04-09 NYU Langone Health Departments of Pathology and Medicine, New York University School of Medicine Maria Aguero-Rosenfeld, Brendan Belovarac, Margaret Black, Ludovic Boytard, John Cadley, Paolo Cotzia, John Chen, Dacia Dimartino, Xiaojun Feng, Tatyana Gindin, Emily Guzman, Adriana Heguy, Megan Hogan, Emily Huang, George Jour, Lawrence H. Lin, Raven Luther, Andrew Lytle, Christian Marier, Matthew T. Maurano, Mark J. Mulligan, Peter Meyn, Raquel Ordonez Ciriza, Iman Osman, Jared Pinnell, Vanessa Raabe, Sitharam Ramaswami, Amy Rapkiewicz, Andre M. Ribeiro-dos-Santos, Marie Samanovic-Golden, Antonio Serrano, Guomiao Shen, Matija Snuderl, Theodore Vougiouklakis, Nick Vulpescu, Gael Westby, Paul Zappile, Yutong Zhang
EPI_ISL_430389 hCoV-19/USA/NY-NYUMC362/2020 North America / USA / New York / Brooklyn 2020-04-08 NYU Langone Health Departments of Pathology and Medicine, New York University School of Medicine Maria Aguero-Rosenfeld, Brendan Belovarac, Margaret Black, Ludovic Boytard, John Cadley, Paolo Cotzia, John Chen, Dacia Dimartino, Xiaojun Feng, Tatyana Gindin, Emily Guzman, Adriana Heguy, Megan Hogan, Emily Huang, George Jour, Lawrence H. Lin, Raven Luther, Andrew Lytle, Christian Marier, Matthew T. Maurano, Mark J. Mulligan, Peter Meyn, Raquel Ordonez Ciriza, Iman Osman, Jared Pinnell, Vanessa Raabe, Sitharam Ramaswami, Amy Rapkiewicz, Andre M. Ribeiro-dos-Santos, Marie Samanovic-Golden, Antonio Serrano, Guomiao Shen, Matija Snuderl, Theodore Vougiouklakis, Nick Vulpescu, Gael Westby, Paul Zappile, Yutong Zhang
EPI_ISL_430390 hCoV-19/USA/NY-NYUMC363/2020 North America / USA / New York / Brooklyn 2020-04-08

NYU Langone Health	Departments of Pathology and Medicine, New York University School of Medicine	Maria Aguero-Rosenfeld, Brendan Belovarac, Margaret Black, Ludovic Boytard, John Cadley, Paolo Cotzia, John Chen, Dacia Dimartino, Xiaojun Feng, Tatyana Gindin, Emily Guzman, Adriana Heguy, Megan Hogan, Emily Huang, George Jour, Lawrence H. Lin, Raven Luther, Andrew Lytle, Christian Marier, Matthew T. Maurano, Mark J. Mulligan, Peter Meyn, Raquel Ordonez Ciriza, Iman Osman, Jared Pinnell, Vanessa Raabe, Sitharam Ramaswami, Amy Rapkiewicz, Andre M. Ribeiro-dos-Santos, Marie Samanovic-Golden, Antonio Serrano, Guomiao Shen, Matija Snuderl, Theodore Vougiouklakis, Nick Vulpescu, Gael Westby, Paul Zappile, Yutong Zhang
EPI_ISL_430391	hCoV-19/USA/NY-NYUMC364/2020	North America / USA / New York / Queens 2020-04-09	NYU Langone Health	Departments of Pathology and Medicine, New York University School of Medicine	Maria Aguero-Rosenfeld, Brendan Belovarac, Margaret Black, Ludovic Boytard, John Cadley, Paolo Cotzia, John Chen, Dacia Dimartino, Xiaojun Feng, Tatyana Gindin, Emily Guzman, Adriana Heguy, Megan Hogan, Emily Huang, George Jour, Lawrence H. Lin, Raven Luther, Andrew Lytle, Christian Marier, Matthew T. Maurano, Mark J. Mulligan, Peter Meyn, Raquel Ordonez Ciriza, Iman Osman, Jared Pinnell, Vanessa Raabe, Sitharam Ramaswami, Amy Rapkiewicz, Andre M. Ribeiro-dos-Santos, Marie Samanovic-Golden, Antonio Serrano, Guomiao Shen, Matija Snuderl, Theodore Vougiouklakis, Nick Vulpescu, Gael Westby, Paul Zappile, Yutong Zhang
EPI_ISL_430392	hCoV-19/USA/NY-NYUMC365/2020	North America / USA / New York / Brooklyn	2020-04-08	NYU Langone Health	Departments of Pathology and Medicine, New York University School of Medicine	Maria Aguero-Rosenfeld, Brendan Belovarac, Margaret Black, Ludovic Boytard, John Cadley, Paolo Cotzia, John Chen, Dacia Dimartino, Xiaojun Feng, Tatyana Gindin, Emily Guzman, Adriana Heguy, Megan Hogan, Emily Huang, George Jour, Lawrence H. Lin, Raven Luther, Andrew Lytle, Christian Marier, Matthew T. Maurano, Mark J. Mulligan, Peter Meyn, Raquel Ordonez Ciriza, Iman Osman, Jared Pinnell, Vanessa Raabe, Sitharam Ramaswami, Amy Rapkiewicz, Andre M. Ribeiro-dos-Santos, Marie Samanovic-Golden, Antonio Serrano, Guomiao Shen, Matija Snuderl, Theodore Vougiouklakis, Nick Vulpescu, Gael Westby, Paul Zappile, Yutong Zhang
EPI_ISL_430393	hCoV-19/USA/NY-NYUMC366/2020	North America / USA / New York / Manhattan	2020-04-09	NYU Langone Health	Departments of Pathology and Medicine, New York University School of Medicine	Maria Aguero-Rosenfeld, Brendan Belovarac, Margaret Black, Ludovic Boytard, John Cadley, Paolo Cotzia, John Chen, Dacia Dimartino, Xiaojun Feng, Tatyana Gindin, Emily Guzman, Adriana Heguy, Megan Hogan, Emily Huang, George Jour, Lawrence H. Lin, Raven Luther, Andrew Lytle, Christian Marier, Matthew T. Maurano, Mark J. Mulligan, Peter Meyn, Raquel Ordonez Ciriza, Iman Osman, Jared Pinnell, Vanessa Raabe, Sitharam Ramaswami, Amy Rapkiewicz, Andre M. Ribeiro-dos-Santos, Marie Samanovic-Golden, Antonio Serrano, Guomiao Shen, Matija Snuderl, Theodore Vougiouklakis, Nick Vulpescu, Gael Westby, Paul Zappile, Yutong Zhang
EPI_ISL_430394	hCoV-19/USA/NJ-NYUMC367/2020	North America / USA / New Jersey / Hudson County	2020-04-09	NYU Langone Health	Departments of Pathology and Medicine, New York University School of Medicine	Maria Aguero-Rosenfeld, Brendan Belovarac, Margaret Black, Ludovic Boytard, John Cadley, Paolo Cotzia, John Chen, Dacia Dimartino, Xiaojun Feng, Tatyana Gindin, Emily Guzman, Adriana Heguy, Megan Hogan, Emily Huang, George Jour, Lawrence H. Lin, Raven Luther, Andrew Lytle, Christian Marier, Matthew T. Maurano, Mark J. Mulligan, Peter Meyn, Raquel Ordonez Ciriza, Iman Osman, Jared Pinnell, Vanessa Raabe, Sitharam Ramaswami, Amy Rapkiewicz, Andre M. Ribeiro-dos-Santos, Marie Samanovic-Golden, Antonio Serrano, Guomiao Shen, Matija Snuderl, Theodore Vougiouklakis, Nick Vulpescu, Gael Westby, Paul Zappile, Yutong Zhang
EPI_ISL_430395	hCoV-19/USA/NY-NYUMC368/2020	North America / USA / New York / Brooklyn	2020-04-09	NYU Langone Health	Departments of Pathology and Medicine, New York University School of Medicine	Maria Aguero-Rosenfeld, Brendan Belovarac, Margaret Black, Ludovic Boytard, John Cadley, Paolo Cotzia, John Chen, Dacia Dimartino, Xiaojun Feng, Tatyana Gindin, Emily Guzman, Adriana Heguy, Megan Hogan, Emily Huang, George Jour, Lawrence H. Lin, Raven Luther, Andrew Lytle, Christian Marier, Matthew T. Maurano, Mark J. Mulligan, Peter Meyn, Raquel Ordonez Ciriza, Iman Osman, Jared Pinnell, Vanessa Raabe, Sitharam Ramaswami, Amy Rapkiewicz, Andre M. Ribeiro-dos-Santos, Marie Samanovic-Golden, Antonio Serrano, Guomiao Shen, Matija Snuderl, Theodore Vougiouklakis, Nick Vulpescu, Gael Westby, Paul Zappile, Yutong Zhang
EPI_ISL_430396	hCoV-19/USA/NY-NYUMC369/2020	North America / USA / New York / Nassau County	2020-04-10	NYU Langone Health	Departments of Pathology and Medicine, New York University School of Medicine	Maria Aguero-Rosenfeld, Brendan Belovarac, Margaret Black, Ludovic Boytard, John Cadley, Paolo Cotzia, John Chen, Dacia Dimartino, Xiaojun Feng, Tatyana Gindin, Emily Guzman, Adriana Heguy, Megan Hogan, Emily Huang, George Jour, Lawrence H. Lin, Raven Luther, Andrew Lytle, Christian Marier, Matthew T. Maurano, Mark J. Mulligan, Peter Meyn, Raquel Ordonez Ciriza, Iman Osman, Jared Pinnell, Vanessa Raabe, Sitharam Ramaswami, Amy Rapkiewicz, Andre M. Ribeiro-dos-Santos, Marie Samanovic-Golden, Antonio Serrano, Guomiao Shen, Matija Snuderl, Theodore Vougiouklakis, Nick Vulpescu, Gael Westby, Paul Zappile, Yutong Zhang
EPI_ISL_430397	hCoV-19/USA/NY-NYUMC370/2020	North America / USA / New York / Nassau County	2020-04-09	NYU Langone Health	Departments of Pathology and Medicine, New York University School of Medicine	Maria Aguero-Rosenfeld, Brendan Belovarac, Margaret Black, Ludovic Boytard, John Cadley, Paolo Cotzia, John Chen, Dacia Dimartino, Xiaojun Feng, Tatyana Gindin, Emily Guzman, Adriana Heguy, Megan Hogan, Emily Huang, George Jour, Lawrence H. Lin, Raven Luther, Andrew Lytle, Christian Marier, Matthew T. Maurano, Mark J. Mulligan, Peter Meyn, Raquel Ordonez Ciriza, Iman Osman, Jared Pinnell, Vanessa Raabe, Sitharam Ramaswami, Amy Rapkiewicz, Andre M. Ribeiro-dos-Santos, Marie Samanovic-Golden, Antonio Serrano, Guomiao Shen, Matija Snuderl, Theodore Vougiouklakis, Nick Vulpescu, Gael Westby, Paul Zappile, Yutong Zhang
EPI_ISL_430398	hCoV-19/USA/NY-NYUMC371/2020	North America / USA / New York / Nassau County	2020-04-10	NYU Langone Health	Departments of Pathology and Medicine, New York University School of Medicine	Maria Aguero-Rosenfeld, Brendan Belovarac, Margaret Black, Ludovic Boytard, John Cadley, Paolo Cotzia, John Chen, Dacia Dimartino, Xiaojun Feng, Tatyana Gindin, Emily Guzman, Adriana Heguy, Megan Hogan, Emily Huang, George Jour, Lawrence H. Lin, Raven Luther, Andrew Lytle, Christian Marier, Matthew T. Maurano, Mark J. Mulligan, Peter Meyn, Raquel Ordonez Ciriza, Iman Osman, Jared Pinnell, Vanessa Raabe, Sitharam Ramaswami, Amy Rapkiewicz, Andre M. Ribeiro-dos-Santos, Marie Samanovic-Golden, Antonio Serrano, Guomiao Shen, Matija Snuderl,

Theodore Vougiouklakis, Nick Vulpescu, Gael Westby, Paul Zappile, Yutong Zhang
EPI_ISL_430399 hCoV-19/USA/NY-NYUMC372/2020 North America / USA / New York / Nassau County 2020-04-10 NYU Langone Health Departments of Pathology and Medicine, New York University School of Medicine Maria Aguero-Rosenfeld, Brendan Belovarac, Margaret Black, Ludovic Boytard, John Cadley, Paolo Cotzia, John Chen, Dacia Dimartino, Xiaojun Feng, Tatyana Gindin, Emily Guzman, Adriana Heguy, Megan Hogan, Emily Huang, George Jour, Lawrence H. Lin, Raven Luther, Andrew Lytle, Christian Marier, Matthew T. Maurano, Mark J. Mulligan, Peter Meyn, Raquel Ordonez Ciriza, Iman Osman, Jared Pinnell, Vanessa Raabe, Sitharam Ramaswami, Amy Rapkiewicz, Andre M. Ribeiro-dos-Santos, Marie Samanovic-Golden, Antonio Serrano, Guomiao Shen, Matija Snuderl, Theodore Vougiouklakis, Nick Vulpescu, Gael Westby, Paul Zappile, Yutong Zhang
EPI_ISL_430400 hCoV-19/USA/NY-NYUMC373/2020 North America / USA / New York / Brooklyn 2020-04-10 NYU Langone Health Departments of Pathology and Medicine, New York University School of Medicine Maria Aguero-Rosenfeld, Brendan Belovarac, Margaret Black, Ludovic Boytard, John Cadley, Paolo Cotzia, John Chen, Dacia Dimartino, Xiaojun Feng, Tatyana Gindin, Emily Guzman, Adriana Heguy, Megan Hogan, Emily Huang, George Jour, Lawrence H. Lin, Raven Luther, Andrew Lytle, Christian Marier, Matthew T. Maurano, Mark J. Mulligan, Peter Meyn, Raquel Ordonez Ciriza, Iman Osman, Jared Pinnell, Vanessa Raabe, Sitharam Ramaswami, Amy Rapkiewicz, Andre M. Ribeiro-dos-Santos, Marie Samanovic-Golden, Antonio Serrano, Guomiao Shen, Matija Snuderl, Theodore Vougiouklakis, Nick Vulpescu, Gael Westby, Paul Zappile, Yutong Zhang
EPI_ISL_430401 hCoV-19/USA/CT-NYUMC374/2020 North America / USA / Connecticut / Fairfield County 2020-04-10 NYU Langone Health Departments of Pathology and Medicine, New York University School of Medicine Maria Aguero-Rosenfeld, Brendan Belovarac, Margaret Black, Ludovic Boytard, John Cadley, Paolo Cotzia, John Chen, Dacia Dimartino, Xiaojun Feng, Tatyana Gindin, Emily Guzman, Adriana Heguy, Megan Hogan, Emily Huang, George Jour, Lawrence H. Lin, Raven Luther, Andrew Lytle, Christian Marier, Matthew T. Maurano, Mark J. Mulligan, Peter Meyn, Raquel Ordonez Ciriza, Iman Osman, Jared Pinnell, Vanessa Raabe, Sitharam Ramaswami, Amy Rapkiewicz, Andre M. Ribeiro-dos-Santos, Marie Samanovic-Golden, Antonio Serrano, Guomiao Shen, Matija Snuderl, Theodore Vougiouklakis, Nick Vulpescu, Gael Westby, Paul Zappile, Yutong Zhang
EPI_ISL_430402 hCoV-19/USA/NY-NYUMC375/2020 North America / USA / New York / Brooklyn 2020-04-10 NYU Langone Health Departments of Pathology and Medicine, New York University School of Medicine Maria Aguero-Rosenfeld, Brendan Belovarac, Margaret Black, Ludovic Boytard, John Cadley, Paolo Cotzia, John Chen, Dacia Dimartino, Xiaojun Feng, Tatyana Gindin, Emily Guzman, Adriana Heguy, Megan Hogan, Emily Huang, George Jour, Lawrence H. Lin, Raven Luther, Andrew Lytle, Christian Marier, Matthew T. Maurano, Mark J. Mulligan, Peter Meyn, Raquel Ordonez Ciriza, Iman Osman, Jared Pinnell, Vanessa Raabe, Sitharam Ramaswami, Amy Rapkiewicz, Andre M. Ribeiro-dos-Santos, Marie Samanovic-Golden, Antonio Serrano, Guomiao Shen, Matija Snuderl, Theodore Vougiouklakis, Nick Vulpescu, Gael Westby, Paul Zappile, Yutong Zhang
EPI_ISL_430403 hCoV-19/USA/NJ-NYUMC376/2020 North America / USA / New York 2020-04-10 NYU Langone Health Departments of Pathology and Medicine, New York University School of Medicine Maria Aguero-Rosenfeld, Brendan Belovarac, Margaret Black, Ludovic Boytard, John Cadley, Paolo Cotzia, John Chen, Dacia Dimartino, Xiaojun Feng, Tatyana Gindin, Emily Guzman, Adriana Heguy, Megan Hogan, Emily Huang, George Jour, Lawrence H. Lin, Raven Luther, Andrew Lytle, Christian Marier, Matthew T. Maurano, Mark J. Mulligan, Peter Meyn, Raquel Ordonez Ciriza, Iman Osman, Jared Pinnell, Vanessa Raabe, Sitharam Ramaswami, Amy Rapkiewicz, Andre M. Ribeiro-dos-Santos, Marie Samanovic-Golden, Antonio Serrano, Guomiao Shen, Matija Snuderl, Theodore Vougiouklakis, Nick Vulpescu, Gael Westby, Paul Zappile, Yutong Zhang
EPI_ISL_430404 hCoV-19/USA/NY-NYUMC377/2020 North America / USA / New York / Brooklyn 2020-04-10 NYU Langone Health Departments of Pathology and Medicine, New York University School of Medicine Maria Aguero-Rosenfeld, Brendan Belovarac, Margaret Black, Ludovic Boytard, John Cadley, Paolo Cotzia, John Chen, Dacia Dimartino, Xiaojun Feng, Tatyana Gindin, Emily Guzman, Adriana Heguy, Megan Hogan, Emily Huang, George Jour, Lawrence H. Lin, Raven Luther, Andrew Lytle, Christian Marier, Matthew T. Maurano, Mark J. Mulligan, Peter Meyn, Raquel Ordonez Ciriza, Iman Osman, Jared Pinnell, Vanessa Raabe, Sitharam Ramaswami, Amy Rapkiewicz, Andre M. Ribeiro-dos-Santos, Marie Samanovic-Golden, Antonio Serrano, Guomiao Shen, Matija Snuderl, Theodore Vougiouklakis, Nick Vulpescu, Gael Westby, Paul Zappile, Yutong Zhang
EPI_ISL_430405 hCoV-19/USA/NJ-NYUMC378/2020 North America / USA / New Jersey / Hudson County 2020-04-10 NYU Langone Health Departments of Pathology and Medicine, New York University School of Medicine Maria Aguero-Rosenfeld, Brendan Belovarac, Margaret Black, Ludovic Boytard, John Cadley, Paolo Cotzia, John Chen, Dacia Dimartino, Xiaojun Feng, Tatyana Gindin, Emily Guzman, Adriana Heguy, Megan Hogan, Emily Huang, George Jour, Lawrence H. Lin, Raven Luther, Andrew Lytle, Christian Marier, Matthew T. Maurano, Mark J. Mulligan, Peter Meyn, Raquel Ordonez Ciriza, Iman Osman, Jared Pinnell, Vanessa Raabe, Sitharam Ramaswami, Amy Rapkiewicz, Andre M. Ribeiro-dos-Santos, Marie Samanovic-Golden, Antonio Serrano, Guomiao Shen, Matija Snuderl, Theodore Vougiouklakis, Nick Vulpescu, Gael Westby, Paul Zappile, Yutong Zhang
EPI_ISL_430406 hCoV-19/USA/NY-NYUMC379/2020 North America / USA / New York / Manhattan 2020-04-10 NYU Langone Health Departments of Pathology and Medicine, New York University School of Medicine Maria Aguero-Rosenfeld, Brendan Belovarac, Margaret Black, Ludovic Boytard, John Cadley, Paolo Cotzia, John Chen, Dacia Dimartino, Xiaojun Feng, Tatyana Gindin, Emily Guzman, Adriana Heguy, Megan Hogan, Emily Huang, George Jour, Lawrence H. Lin, Raven Luther, Andrew Lytle, Christian Marier, Matthew T. Maurano, Mark J. Mulligan, Peter Meyn, Raquel Ordonez Ciriza, Iman Osman, Jared Pinnell, Vanessa Raabe, Sitharam Ramaswami, Amy Rapkiewicz, Andre M. Ribeiro-dos-Santos, Marie Samanovic-Golden, Antonio Serrano, Guomiao Shen, Matija Snuderl, Theodore Vougiouklakis, Nick Vulpescu, Gael Westby, Paul Zappile, Yutong Zhang
EPI_ISL_430407 hCoV-19/USA/NY-NYUMC380/2020 North America / USA / New York / Brooklyn 2020-04-10 NYU Langone Health Departments of Pathology and Medicine, New York University School of Medicine Maria Aguero-Rosenfeld, Brendan Belovarac, Margaret Black, Ludovic Boytard, John Cadley, Paolo Cotzia, John Chen, Dacia Dimartino, Xiaojun Feng, Tatyana Gindin, Emily Guzman, Adriana Heguy, Megan Hogan, Emily Huang, George Jour, Lawrence H. Lin, Raven Luther, Andrew Lytle, Christian Marier, Matthew T. Maurano, Mark J. Mulligan,

Peter Meyn, Raquel Ordonez Ciriza, Iman Osman, Jared Pinnell, Vanessa Raabe, Sitharam Ramaswami, Amy Rapkiewicz, Andre M. Ribeiro-dos-Santos, Marie Samanovic-Golden, Antonio Serrano, Guomiao Shen, Matija Snuderl, Theodore Vougiouklakis, Nick Vulpescu, Gael Westby, Paul Zappile, Yutong Zhang
EPI_ISL_430408 hCoV-19/USA/NY-NYUMC381/2020 North America / USA / New York / Nassau County 2020-04-09 NYU Langone Health Departments of Pathology and Medicine, New York University School of Medicine Maria Aguero-Rosenfeld, Brendan Belovarac, Margaret Black, Ludovic Boytard, John Cadley, Paolo Cotzia, John Chen, Dacia Dimartino, Xiaojun Feng, Tatyana Gindin, Emily Guzman, Adriana Heguy, Megan Hogan, Emily Huang, George Jour, Lawrence H. Lin, Raven Luther, Andrew Lytle, Christian Marier, Matthew T. Maurano, Mark J. Mulligan, Peter Meyn, Raquel Ordonez Ciriza, Iman Osman, Jared Pinnell, Vanessa Raabe, Sitharam Ramaswami, Amy Rapkiewicz, Andre M. Ribeiro-dos-Santos, Marie Samanovic-Golden, Antonio Serrano, Guomiao Shen, Matija Snuderl, Theodore Vougiouklakis, Nick Vulpescu, Gael Westby, Paul Zappile, Yutong Zhang
EPI_ISL_430409 hCoV-19/USA/NY-NYUMC382/2020 North America / USA / New York / Brooklyn 2020-04-10 NYU Langone Health Departments of Pathology and Medicine, New York University School of Medicine Maria Aguero-Rosenfeld, Brendan Belovarac, Margaret Black, Ludovic Boytard, John Cadley, Paolo Cotzia, John Chen, Dacia Dimartino, Xiaojun Feng, Tatyana Gindin, Emily Guzman, Adriana Heguy, Megan Hogan, Emily Huang, George Jour, Lawrence H. Lin, Raven Luther, Andrew Lytle, Christian Marier, Matthew T. Maurano, Mark J. Mulligan, Peter Meyn, Raquel Ordonez Ciriza, Iman Osman, Jared Pinnell, Vanessa Raabe, Sitharam Ramaswami, Amy Rapkiewicz, Andre M. Ribeiro-dos-Santos, Marie Samanovic-Golden, Antonio Serrano, Guomiao Shen, Matija Snuderl, Theodore Vougiouklakis, Nick Vulpescu, Gael Westby, Paul Zappile, Yutong Zhang
EPI_ISL_430410 hCoV-19/USA/NY-NYUMC383/2020 North America / USA / New York / Brooklyn 2020-04-10 NYU Langone Health Departments of Pathology and Medicine, New York University School of Medicine Maria Aguero-Rosenfeld, Brendan Belovarac, Margaret Black, Ludovic Boytard, John Cadley, Paolo Cotzia, John Chen, Dacia Dimartino, Xiaojun Feng, Tatyana Gindin, Emily Guzman, Adriana Heguy, Megan Hogan, Emily Huang, George Jour, Lawrence H. Lin, Raven Luther, Andrew Lytle, Christian Marier, Matthew T. Maurano, Mark J. Mulligan, Peter Meyn, Raquel Ordonez Ciriza, Iman Osman, Jared Pinnell, Vanessa Raabe, Sitharam Ramaswami, Amy Rapkiewicz, Andre M. Ribeiro-dos-Santos, Marie Samanovic-Golden, Antonio Serrano, Guomiao Shen, Matija Snuderl, Theodore Vougiouklakis, Nick Vulpescu, Gael Westby, Paul Zappile, Yutong Zhang
EPI_ISL_430411 hCoV-19/USA/NY-NYUMC384/2020 North America / USA / New York / Brooklyn 2020-04-09 NYU Langone Health Departments of Pathology and Medicine, New York University School of Medicine Maria Aguero-Rosenfeld, Brendan Belovarac, Margaret Black, Ludovic Boytard, John Cadley, Paolo Cotzia, John Chen, Dacia Dimartino, Xiaojun Feng, Tatyana Gindin, Emily Guzman, Adriana Heguy, Megan Hogan, Emily Huang, George Jour, Lawrence H. Lin, Raven Luther, Andrew Lytle, Christian Marier, Matthew T. Maurano, Mark J. Mulligan, Peter Meyn, Raquel Ordonez Ciriza, Iman Osman, Jared Pinnell, Vanessa Raabe, Sitharam Ramaswami, Amy Rapkiewicz, Andre M. Ribeiro-dos-Santos, Marie Samanovic-Golden, Antonio Serrano, Guomiao Shen, Matija Snuderl, Theodore Vougiouklakis, Nick Vulpescu, Gael Westby, Paul Zappile, Yutong Zhang
EPI_ISL_430412 hCoV-19/USA/NY-NYUMC385/2020 North America / USA / New York / Staten Island 2020-04-10 NYU Langone Health Departments of Pathology and Medicine, New York University School of Medicine Maria Aguero-Rosenfeld, Brendan Belovarac, Margaret Black, Ludovic Boytard, John Cadley, Paolo Cotzia, John Chen, Dacia Dimartino, Xiaojun Feng, Tatyana Gindin, Emily Guzman, Adriana Heguy, Megan Hogan, Emily Huang, George Jour, Lawrence H. Lin, Raven Luther, Andrew Lytle, Christian Marier, Matthew T. Maurano, Mark J. Mulligan, Peter Meyn, Raquel Ordonez Ciriza, Iman Osman, Jared Pinnell, Vanessa Raabe, Sitharam Ramaswami, Amy Rapkiewicz, Andre M. Ribeiro-dos-Santos, Marie Samanovic-Golden, Antonio Serrano, Guomiao Shen, Matija Snuderl, Theodore Vougiouklakis, Nick Vulpescu, Gael Westby, Paul Zappile, Yutong Zhang
EPI_ISL_430413 hCoV-19/USA/NY-NYUMC386/2020 North America / USA / New York / Nassau County 2020-04-09 NYU Langone Health Departments of Pathology and Medicine, New York University School of Medicine Maria Aguero-Rosenfeld, Brendan Belovarac, Margaret Black, Ludovic Boytard, John Cadley, Paolo Cotzia, John Chen, Dacia Dimartino, Xiaojun Feng, Tatyana Gindin, Emily Guzman, Adriana Heguy, Megan Hogan, Emily Huang, George Jour, Lawrence H. Lin, Raven Luther, Andrew Lytle, Christian Marier, Matthew T. Maurano, Mark J. Mulligan, Peter Meyn, Raquel Ordonez Ciriza, Iman Osman, Jared Pinnell, Vanessa Raabe, Sitharam Ramaswami, Amy Rapkiewicz, Andre M. Ribeiro-dos-Santos, Marie Samanovic-Golden, Antonio Serrano, Guomiao Shen, Matija Snuderl, Theodore Vougiouklakis, Nick Vulpescu, Gael Westby, Paul Zappile, Yutong Zhang
EPI_ISL_430414 hCoV-19/USA/NY-NYUMC387/2020 North America / USA / New York / Rockland County 2020-04-10 NYU Langone Health Departments of Pathology and Medicine, New York University School of Medicine Maria Aguero-Rosenfeld, Brendan Belovarac, Margaret Black, Ludovic Boytard, John Cadley, Paolo Cotzia, John Chen, Dacia Dimartino, Xiaojun Feng, Tatyana Gindin, Emily Guzman, Adriana Heguy, Megan Hogan, Emily Huang, George Jour, Lawrence H. Lin, Raven Luther, Andrew Lytle, Christian Marier, Matthew T. Maurano, Mark J. Mulligan, Peter Meyn, Raquel Ordonez Ciriza, Iman Osman, Jared Pinnell, Vanessa Raabe, Sitharam Ramaswami, Amy Rapkiewicz, Andre M. Ribeiro-dos-Santos, Marie Samanovic-Golden, Antonio Serrano, Guomiao Shen, Matija Snuderl, Theodore Vougiouklakis, Nick Vulpescu, Gael Westby, Paul Zappile, Yutong Zhang
EPI_ISL_430415 hCoV-19/USA/NY-NYUMC388/2020 North America / USA / New York / Manhattan 2020-04-10 NYU Langone Health Departments of Pathology and Medicine, New York University School of Medicine Maria Aguero-Rosenfeld, Brendan Belovarac, Margaret Black, Ludovic Boytard, John Cadley, Paolo Cotzia, John Chen, Dacia Dimartino, Xiaojun Feng, Tatyana Gindin, Emily Guzman, Adriana Heguy, Megan Hogan, Emily Huang, George Jour, Lawrence H. Lin, Raven Luther, Andrew Lytle, Christian Marier, Matthew T. Maurano, Mark J. Mulligan, Peter Meyn, Raquel Ordonez Ciriza, Iman Osman, Jared Pinnell, Vanessa Raabe, Sitharam Ramaswami, Amy Rapkiewicz, Andre M. Ribeiro-dos-Santos, Marie Samanovic-Golden, Antonio Serrano, Guomiao Shen, Matija Snuderl, Theodore Vougiouklakis, Nick Vulpescu, Gael Westby, Paul Zappile, Yutong Zhang
EPI_ISL_430416 hCoV-19/USA/NY-NYUMC389/2020 North America / USA / New York / Manhattan 2020-04-13 NYU Langone Health Departments of Pathology and Medicine, New York University School of Medicine Maria Aguero-Rosenfeld, Brendan Belovarac, Margaret Black, Ludovic Boytard, John Cadley, Paolo Cotzia, John Chen,

Dacia Dimartino, Xiaojun Feng, Tatyana Gindin, Emily Guzman, Adriana Heguy, Megan Hogan, Emily Huang, George Jour, Lawrence H. Lin, Raven Luther, Andrew Lytle, Christian Marier, Matthew T. Maurano, Mark J. Mulligan, Peter Meyn, Raquel Ordonez Ciriza, Iman Osman, Jared Pinnell, Vanessa Raabe, Sitharam Ramaswami, Amy Rapkiewicz, Andre M. Ribeiro-dos-Santos, Marie Samanovic-Golden, Antonio Serrano, Guomiao Shen, Matija Snuderl, Theodore Vougiouklakis, Nick Vulpescu, Gael Westby, Paul Zappile, Yutong Zhang
EPI_ISL_430417 hCoV-19/USA/NY-NYUMC390/2020 North America / USA / New York / Nassau County 2020-04-13 NYU Langone Health Departments of Pathology and Medicine, New York University School of Medicine Maria Aguero-Rosenfeld, Brendan Belovarac, Margaret Black, Ludovic Boytard, John Cadley, Paolo Cotzia, John Chen, Dacia Dimartino, Xiaojun Feng, Tatyana Gindin, Emily Guzman, Adriana Heguy, Megan Hogan, Emily Huang, George Jour, Lawrence H. Lin, Raven Luther, Andrew Lytle, Christian Marier, Matthew T. Maurano, Mark J. Mulligan, Peter Meyn, Raquel Ordonez Ciriza, Iman Osman, Jared Pinnell, Vanessa Raabe, Sitharam Ramaswami, Amy Rapkiewicz, Andre M. Ribeiro-dos-Santos, Marie Samanovic-Golden, Antonio Serrano, Guomiao Shen, Matija Snuderl, Theodore Vougiouklakis, Nick Vulpescu, Gael Westby, Paul Zappile, Yutong Zhang
EPI_ISL_430418 hCoV-19/USA/NY-NYUMC391/2020 North America / USA / New York / Nassau County 2020-04-13 NYU Langone Health Departments of Pathology and Medicine, New York University School of Medicine Maria Aguero-Rosenfeld, Brendan Belovarac, Margaret Black, Ludovic Boytard, John Cadley, Paolo Cotzia, John Chen, Dacia Dimartino, Xiaojun Feng, Tatyana Gindin, Emily Guzman, Adriana Heguy, Megan Hogan, Emily Huang, George Jour, Lawrence H. Lin, Raven Luther, Andrew Lytle, Christian Marier, Matthew T. Maurano, Mark J. Mulligan, Peter Meyn, Raquel Ordonez Ciriza, Iman Osman, Jared Pinnell, Vanessa Raabe, Sitharam Ramaswami, Amy Rapkiewicz, Andre M. Ribeiro-dos-Santos, Marie Samanovic-Golden, Antonio Serrano, Guomiao Shen, Matija Snuderl, Theodore Vougiouklakis, Nick Vulpescu, Gael Westby, Paul Zappile, Yutong Zhang
EPI_ISL_430419 hCoV-19/USA/NY-NYUMC392/2020 North America / USA / New York / Nassau County 2020-04-13 NYU Langone Health Departments of Pathology and Medicine, New York University School of Medicine Maria Aguero-Rosenfeld, Brendan Belovarac, Margaret Black, Ludovic Boytard, John Cadley, Paolo Cotzia, John Chen, Dacia Dimartino, Xiaojun Feng, Tatyana Gindin, Emily Guzman, Adriana Heguy, Megan Hogan, Emily Huang, George Jour, Lawrence H. Lin, Raven Luther, Andrew Lytle, Christian Marier, Matthew T. Maurano, Mark J. Mulligan, Peter Meyn, Raquel Ordonez Ciriza, Iman Osman, Jared Pinnell, Vanessa Raabe, Sitharam Ramaswami, Amy Rapkiewicz, Andre M. Ribeiro-dos-Santos, Marie Samanovic-Golden, Antonio Serrano, Guomiao Shen, Matija Snuderl, Theodore Vougiouklakis, Nick Vulpescu, Gael Westby, Paul Zappile, Yutong Zhang
EPI_ISL_430420 hCoV-19/USA/NY-NYUMC393/2020 North America / USA / New York / Staten Island 2020-04-13 NYU Langone Health Departments of Pathology and Medicine, New York University School of Medicine Maria Aguero-Rosenfeld, Brendan Belovarac, Margaret Black, Ludovic Boytard, John Cadley, Paolo Cotzia, John Chen, Dacia Dimartino, Xiaojun Feng, Tatyana Gindin, Emily Guzman, Adriana Heguy, Megan Hogan, Emily Huang, George Jour, Lawrence H. Lin, Raven Luther, Andrew Lytle, Christian Marier, Matthew T. Maurano, Mark J. Mulligan, Peter Meyn, Raquel Ordonez Ciriza, Iman Osman, Jared Pinnell, Vanessa Raabe, Sitharam Ramaswami, Amy Rapkiewicz, Andre M. Ribeiro-dos-Santos, Marie Samanovic-Golden, Antonio Serrano, Guomiao Shen, Matija Snuderl, Theodore Vougiouklakis, Nick Vulpescu, Gael Westby, Paul Zappile, Yutong Zhang
EPI_ISL_430421 hCoV-19/USA/NY-NYUMC394/2020 North America / USA / New York / Brooklyn 2020-04-13 NYU Langone Health Departments of Pathology and Medicine, New York University School of Medicine Maria Aguero-Rosenfeld, Brendan Belovarac, Margaret Black, Ludovic Boytard, John Cadley, Paolo Cotzia, John Chen, Dacia Dimartino, Xiaojun Feng, Tatyana Gindin, Emily Guzman, Adriana Heguy, Megan Hogan, Emily Huang, George Jour, Lawrence H. Lin, Raven Luther, Andrew Lytle, Christian Marier, Matthew T. Maurano, Mark J. Mulligan, Peter Meyn, Raquel Ordonez Ciriza, Iman Osman, Jared Pinnell, Vanessa Raabe, Sitharam Ramaswami, Amy Rapkiewicz, Andre M. Ribeiro-dos-Santos, Marie Samanovic-Golden, Antonio Serrano, Guomiao Shen, Matija Snuderl, Theodore Vougiouklakis, Nick Vulpescu, Gael Westby, Paul Zappile, Yutong Zhang
EPI_ISL_430422 hCoV-19/USA/NY-NYUMC395/2020 North America / USA / New York / Queens 2020-04-13 NYU Langone Health Departments of Pathology and Medicine, New York University School of Medicine Maria Aguero-Rosenfeld, Brendan Belovarac, Margaret Black, Ludovic Boytard, John Cadley, Paolo Cotzia, John Chen, Dacia Dimartino, Xiaojun Feng, Tatyana Gindin, Emily Guzman, Adriana Heguy, Megan Hogan, Emily Huang, George Jour, Lawrence H. Lin, Raven Luther, Andrew Lytle, Christian Marier, Matthew T. Maurano, Mark J. Mulligan, Peter Meyn, Raquel Ordonez Ciriza, Iman Osman, Jared Pinnell, Vanessa Raabe, Sitharam Ramaswami, Amy Rapkiewicz, Andre M. Ribeiro-dos-Santos, Marie Samanovic-Golden, Antonio Serrano, Guomiao Shen, Matija Snuderl, Theodore Vougiouklakis, Nick Vulpescu, Gael Westby, Paul Zappile, Yutong Zhang
EPI_ISL_430423 hCoV-19/USA/NY-NYUMC396/2020 North America / USA / New York / Queens 2020-04-11 NYU Langone Health Departments of Pathology and Medicine, New York University School of Medicine Maria Aguero-Rosenfeld, Brendan Belovarac, Margaret Black, Ludovic Boytard, John Cadley, Paolo Cotzia, John Chen, Dacia Dimartino, Xiaojun Feng, Tatyana Gindin, Emily Guzman, Adriana Heguy, Megan Hogan, Emily Huang, George Jour, Lawrence H. Lin, Raven Luther, Andrew Lytle, Christian Marier, Matthew T. Maurano, Mark J. Mulligan, Peter Meyn, Raquel Ordonez Ciriza, Iman Osman, Jared Pinnell, Vanessa Raabe, Sitharam Ramaswami, Amy Rapkiewicz, Andre M. Ribeiro-dos-Santos, Marie Samanovic-Golden, Antonio Serrano, Guomiao Shen, Matija Snuderl, Theodore Vougiouklakis, Nick Vulpescu, Gael Westby, Paul Zappile, Yutong Zhang
EPI_ISL_430424 hCoV-19/USA/NY-NYUMC397/2020 North America / USA / New York / Brooklyn 2020-04-13 NYU Langone Health Departments of Pathology and Medicine, New York University School of Medicine Maria Aguero-Rosenfeld, Brendan Belovarac, Margaret Black, Ludovic Boytard, John Cadley, Paolo Cotzia, John Chen, Dacia Dimartino, Xiaojun Feng, Tatyana Gindin, Emily Guzman, Adriana Heguy, Megan Hogan, Emily Huang, George Jour, Lawrence H. Lin, Raven Luther, Andrew Lytle, Christian Marier, Matthew T. Maurano, Mark J. Mulligan, Peter Meyn, Raquel Ordonez Ciriza, Iman Osman, Jared Pinnell, Vanessa Raabe, Sitharam Ramaswami, Amy Rapkiewicz, Andre M. Ribeiro-dos-Santos, Marie Samanovic-Golden, Antonio Serrano, Guomiao Shen, Matija Snuderl, Theodore Vougiouklakis, Nick Vulpescu, Gael Westby, Paul Zappile, Yutong Zhang
EPI_ISL_430425 hCoV-19/USA/NY-NYUMC398/2020 North America / USA / New York / Manhattan 2020-04-13

NYU Langone Health	Departments of Pathology and Medicine, New York University School of Medicine	Maria Aguero-Rosenfeld, Brendan Belovarac, Margaret Black, Ludovic Boytard, John Cadley, Paolo Cotzia, John Chen, Dacia Dimartino, Xiaojun Feng, Tatyana Gindin, Emily Guzman, Adriana Heguy, Megan Hogan, Emily Huang, George Jour, Lawrence H. Lin, Raven Luther, Andrew Lytle, Christian Marier, Matthew T. Maurano, Mark J. Mulligan, Peter Meyn, Raquel Ordonez Ciriza, Iman Osman, Jared Pinnell, Vanessa Raabe, Sitharam Ramaswami, Amy Rapkiewicz, Andre M. Ribeiro-dos-Santos, Marie Samanovic-Golden, Antonio Serrano, Guomiao Shen, Matija Snuderl, Theodore Vougiouklakis, Nick Vulpescu, Gael Westby, Paul Zappile, Yutong Zhang
EPI_ISL_430426	hCoV-19/USA/NY-NYUMC399/2020	North America / USA / New York / Manhattan	2020-04-13	NYU Langone Health	Departments of Pathology and Medicine, New York University School of Medicine	Maria Aguero-Rosenfeld, Brendan Belovarac, Margaret Black, Ludovic Boytard, John Cadley, Paolo Cotzia, John Chen, Dacia Dimartino, Xiaojun Feng, Tatyana Gindin, Emily Guzman, Adriana Heguy, Megan Hogan, Emily Huang, George Jour, Lawrence H. Lin, Raven Luther, Andrew Lytle, Christian Marier, Matthew T. Maurano, Mark J. Mulligan, Peter Meyn, Raquel Ordonez Ciriza, Iman Osman, Jared Pinnell, Vanessa Raabe, Sitharam Ramaswami, Amy Rapkiewicz, Andre M. Ribeiro-dos-Santos, Marie Samanovic-Golden, Antonio Serrano, Guomiao Shen, Matija Snuderl, Theodore Vougiouklakis, Nick Vulpescu, Gael Westby, Paul Zappile, Yutong Zhang
EPI_ISL_430427	hCoV-19/USA/NY-NYUMC400/2020	North America / USA / New York / Nassau County	2020-04-13	NYU Langone Health	Departments of Pathology and Medicine, New York University School of Medicine	Maria Aguero-Rosenfeld, Brendan Belovarac, Margaret Black, Ludovic Boytard, John Cadley, Paolo Cotzia, John Chen, Dacia Dimartino, Xiaojun Feng, Tatyana Gindin, Emily Guzman, Adriana Heguy, Megan Hogan, Emily Huang, George Jour, Lawrence H. Lin, Raven Luther, Andrew Lytle, Christian Marier, Matthew T. Maurano, Mark J. Mulligan, Peter Meyn, Raquel Ordonez Ciriza, Iman Osman, Jared Pinnell, Vanessa Raabe, Sitharam Ramaswami, Amy Rapkiewicz, Andre M. Ribeiro-dos-Santos, Marie Samanovic-Golden, Antonio Serrano, Guomiao Shen, Matija Snuderl, Theodore Vougiouklakis, Nick Vulpescu, Gael Westby, Paul Zappile, Yutong Zhang
EPI_ISL_430428	hCoV-19/USA/NJ-NYUMC401/2020	North America / USA / New Jersey / Hudson County	2020-04-13	NYU Langone Health	Departments of Pathology and Medicine, New York University School of Medicine	Maria Aguero-Rosenfeld, Brendan Belovarac, Margaret Black, Ludovic Boytard, John Cadley, Paolo Cotzia, John Chen, Dacia Dimartino, Xiaojun Feng, Tatyana Gindin, Emily Guzman, Adriana Heguy, Megan Hogan, Emily Huang, George Jour, Lawrence H. Lin, Raven Luther, Andrew Lytle, Christian Marier, Matthew T. Maurano, Mark J. Mulligan, Peter Meyn, Raquel Ordonez Ciriza, Iman Osman, Jared Pinnell, Vanessa Raabe, Sitharam Ramaswami, Amy Rapkiewicz, Andre M. Ribeiro-dos-Santos, Marie Samanovic-Golden, Antonio Serrano, Guomiao Shen, Matija Snuderl, Theodore Vougiouklakis, Nick Vulpescu, Gael Westby, Paul Zappile, Yutong Zhang
EPI_ISL_430429	hCoV-19/USA/NY-NYUMC402/2020	North America / USA / New York / Brooklyn	2020-04-13	NYU Langone Health	Departments of Pathology and Medicine, New York University School of Medicine	Maria Aguero-Rosenfeld, Brendan Belovarac, Margaret Black, Ludovic Boytard, John Cadley, Paolo Cotzia, John Chen, Dacia Dimartino, Xiaojun Feng, Tatyana Gindin, Emily Guzman, Adriana Heguy, Megan Hogan, Emily Huang, George Jour, Lawrence H. Lin, Raven Luther, Andrew Lytle, Christian Marier, Matthew T. Maurano, Mark J. Mulligan, Peter Meyn, Raquel Ordonez Ciriza, Iman Osman, Jared Pinnell, Vanessa Raabe, Sitharam Ramaswami, Amy Rapkiewicz, Andre M. Ribeiro-dos-Santos, Marie Samanovic-Golden, Antonio Serrano, Guomiao Shen, Matija Snuderl, Theodore Vougiouklakis, Nick Vulpescu, Gael Westby, Paul Zappile, Yutong Zhang
EPI_ISL_430430	hCoV-19/USA/NY-NYUMC403/2020	North America / USA / New York / Bronx	2020-04-13	NYU Langone Health	Departments of Pathology and Medicine, New York University School of Medicine	Maria Aguero-Rosenfeld, Brendan Belovarac, Margaret Black, Ludovic Boytard, John Cadley, Paolo Cotzia, John Chen, Dacia Dimartino, Xiaojun Feng, Tatyana Gindin, Emily Guzman, Adriana Heguy, Megan Hogan, Emily Huang, George Jour, Lawrence H. Lin, Raven Luther, Andrew Lytle, Christian Marier, Matthew T. Maurano, Mark J. Mulligan, Peter Meyn, Raquel Ordonez Ciriza, Iman Osman, Jared Pinnell, Vanessa Raabe, Sitharam Ramaswami, Amy Rapkiewicz, Andre M. Ribeiro-dos-Santos, Marie Samanovic-Golden, Antonio Serrano, Guomiao Shen, Matija Snuderl, Theodore Vougiouklakis, Nick Vulpescu, Gael Westby, Paul Zappile, Yutong Zhang
EPI_ISL_430431	hCoV-19/USA/NY-NYUMC404/2020	North America / USA / New York / Nassau County	2020-04-13	NYU Langone Health	Departments of Pathology and Medicine, New York University School of Medicine	Maria Aguero-Rosenfeld, Brendan Belovarac, Margaret Black, Ludovic Boytard, John Cadley, Paolo Cotzia, John Chen, Dacia Dimartino, Xiaojun Feng, Tatyana Gindin, Emily Guzman, Adriana Heguy, Megan Hogan, Emily Huang, George Jour, Lawrence H. Lin, Raven Luther, Andrew Lytle, Christian Marier, Matthew T. Maurano, Mark J. Mulligan, Peter Meyn, Raquel Ordonez Ciriza, Iman Osman, Jared Pinnell, Vanessa Raabe, Sitharam Ramaswami, Amy Rapkiewicz, Andre M. Ribeiro-dos-Santos, Marie Samanovic-Golden, Antonio Serrano, Guomiao Shen, Matija Snuderl, Theodore Vougiouklakis, Nick Vulpescu, Gael Westby, Paul Zappile, Yutong Zhang
EPI_ISL_430432	hCoV-19/USA/NY-NYUMC405/2020	North America / USA / New York / Staten Island	2020-04-13	NYU Langone Health	Departments of Pathology and Medicine, New York University School of Medicine	Maria Aguero-Rosenfeld, Brendan Belovarac, Margaret Black, Ludovic Boytard, John Cadley, Paolo Cotzia, John Chen, Dacia Dimartino, Xiaojun Feng, Tatyana Gindin, Emily Guzman, Adriana Heguy, Megan Hogan, Emily Huang, George Jour, Lawrence H. Lin, Raven Luther, Andrew Lytle, Christian Marier, Matthew T. Maurano, Mark J. Mulligan, Peter Meyn, Raquel Ordonez Ciriza, Iman Osman, Jared Pinnell, Vanessa Raabe, Sitharam Ramaswami, Amy Rapkiewicz, Andre M. Ribeiro-dos-Santos, Marie Samanovic-Golden, Antonio Serrano, Guomiao Shen, Matija Snuderl, Theodore Vougiouklakis, Nick Vulpescu, Gael Westby, Paul Zappile, Yutong Zhang
EPI_ISL_430433	hCoV-19/USA/NY-NYUMC406/2020	North America / USA / New York / Queens	2020-04-13	NYU Langone Health	Departments of Pathology and Medicine, New York University School of Medicine	Maria Aguero-Rosenfeld, Brendan Belovarac, Margaret Black, Ludovic Boytard, John Cadley, Paolo Cotzia, John Chen, Dacia Dimartino, Xiaojun Feng, Tatyana Gindin, Emily Guzman, Adriana Heguy, Megan Hogan, Emily Huang, George Jour, Lawrence H. Lin, Raven Luther, Andrew Lytle, Christian Marier, Matthew T. Maurano, Mark J. Mulligan, Peter Meyn, Raquel Ordonez Ciriza, Iman Osman, Jared Pinnell, Vanessa Raabe, Sitharam Ramaswami, Amy Rapkiewicz, Andre M. Ribeiro-dos-Santos, Marie Samanovic-Golden, Antonio Serrano, Guomiao Shen, Matija Snuderl, Theodore

Vougiouklakis, Nick Vulpescu, Gael Westby, Paul Zappile, Yutong Zhang
EPI_ISL_430434 hCoV-19/USA/NY-NYUMC407/2020 North America / USA / New York / Queens 2020-04-12 NYU Langone Health Departments of Pathology and Medicine, New York University School of Medicine Maria Aguero-Rosenfeld, Brendan Belovarac, Margaret Black, Ludovic Boytard, John Cadley, Paolo Cotzia, John Chen, Dacia Dimartino, Xiaojun Feng, Tatyana Gindin, Emily Guzman, Adriana Heguy, Megan Hogan, Emily Huang, George Jour, Lawrence H. Lin, Raven Luther, Andrew Lytle, Christian Marier, Matthew T. Maurano, Mark J. Mulligan, Peter Meyn, Raquel Ordonez Ciriza, Iman Osman, Jared Pinnell, Vanessa Raabe, Sitharam Ramaswami, Amy Rapkiewicz, Andre M. Ribeiro-dos-Santos, Marie Samanovic-Golden, Antonio Serrano, Guomiao Shen, Matija Snuderl, Theodore Vougiouklakis, Nick Vulpescu, Gael Westby, Paul Zappile, Yutong Zhang
EPI_ISL_430439 hCoV-19/Malaysia/IMR_WC1177/2020 Asia / Malaysia 2020-03-05 Institute for Medical Research, Infectious Disease Research Centre, National Institutes of Health, Ministry of Health Malaysia Institute for Medical Research Infectious Disease Research Centre, National Institutes of Health, Ministry of Health Malaysia Suppiah.J, Mohd-Zawawi.Z, Kalyanasundram.J, Azizan.M-A, Mat-Sharani.S, Hisham.H-A, Tan.L-P, Abdul-Wahid.M-Z, Mohd-Zain.R, Ahmad.N, Thayan.R
EPI_ISL_430440 hCoV-19/Malaysia/IMR_WC1170/2020 Asia / Malaysia 2020-03-05 Institute for Medical Research, Infectious Disease Research Centre, National Institutes of Health, Ministry of Health Malaysia Institute for Medical Research, Infectious Disease Research Centre, National Institutes of Health, Ministry of Health Malaysia Suppiah.J, Mohd-Zawawi.Z, Kalyanasundram.J, Azizan.M-A, Mat-Sharani.S, Hisham.H-A, Tan.L-P, Abdul-Wahid.M-Z, Tengku-Abd-Rashid.T-R, Mohd-Zain.R, Ahmad.N, Thayan.R
EPI_ISL_430441 hCoV-19/Malaysia/IMR_WC1097/2020 Asia / Malaysia 2020-02-29 Institute for Medical Research, Infectious Disease Research Centre, National Institutes of Health, Ministry of Health Malaysia Institute for Medical Research, Infectious Disease Research Centre, National Institutes of Health, Ministry of Health Malaysia Suppiah.J, Mohd-Zawawi.Z, Kalyanasundram.J, Azizan.M-A, Mat-Sharani.S, Hisham.H-A, Tan.L-P, Abdul-Wahid.M-Z, Tengku-Rogayah.TAR, Mohd-Zain.R, Ahmad.N, Thayan.R
EPI_ISL_430442 hCoV-19/Malaysia/IMR_WC1098/2020 Asia / Malaysia 2020-02-29 Institute for Medical Research, Infectious Disease Research Centre, National Institutes of Health, Ministry of Health Malaysia Institute for Medical Research, Infectious Disease Research Centre, National Institutes of Health, Ministry of Health Malaysia Suppiah.J, Mohd-Zawawi.Z, Kalyanasundram.J, Azizan.M-A, Mat-Sharani.S, Hisham.H-A, Tan.L-P, Abdul-Wahid.M-Z, Tengku-Rogayah.TAR, Mohd-Zain.R, Ahmad.N, Thayan.R
EPI_ISL_430443 hCoV-19/Malaysia/IMR_WC085/2020 Asia / Malaysia 2020-01-28 Institute for Medical Research, Infectious Disease Research Centre, National Institutes of Health, Ministry of Health Malaysia Institute for Medical Research, Infectious Disease Research Centre, National Institutes of Health, Ministry of Health Malaysia Suppiah.J, Mohd-Zawawi.Z, Kalyanasundram.J, Azizan.M-A, Mat-Sharani.S, Hisham.H-A, Tan.L-P, Abdul-Wahid.M-Z, Tengku-Rogayah.TAR, Mohd-Zain.R, Ahmad.N, Thayan.R
EPI_ISL_430444 hCoV-19/Malaysia/IMR_WC627/2020 Asia / Malaysia 2020-02-12 Institute for Medical Research, Infectious Disease Research Centre, National Institutes of Health, Ministry of Health Malaysia Institute for Medical Research, Infectious Disease Research Centre, National Institutes of Health, Ministry of Health Malaysia Suppiah.J, Mohd-Zawawi.Z, Kalyanasundram.J, Azizan.M-A, Mat-Sharani.S, Hisham.H-A, Tan.L-P, Abdul-Wahid.M-Z, Tengku-Rogayah.TAR, Mohd-Zain.R, Ahmad.N, Thayan.R
EPI_ISL_430464 hCoV-19/India/S3/2020 Asia / India / West Bengal / Kolkata 2020-03-21 ICMR-National Institute of Cholera and Enteric Diseases National Institute of Biomedical Genomics Arindam Maitra, Mamta Chawla Sarkar, Sreedhar Chinnaswamy, Hasina Banu, Ananya Chatterjee, Shanta Dutta, Saumitra Das
EPI_ISL_430465 hCoV-19/India/S5/2020 Asia / India / West Bengal / Darjeeling 2020-03-28 ICMR-National Institute of Cholera and Enteric Diseases National Institute of Biomedical Genomics Arindam Maitra, Mamta Chawla Sarkar, Sreedhar Chinnaswamy, Hasina Banu, Ananya Chatterjee, Shanta Dutta, Saumitra Das
EPI_ISL_430466 hCoV-19/India/S6/2020 Asia / India / West Bengal / Tehatta 2020-03-26 ICMR-National Institute of Cholera and Enteric Diseases National Institute of Biomedical Genomics Arindam Maitra, Mamta Chawla Sarkar, Sreedhar Chinnaswamy, Hasina Banu, Ananya Chatterjee, Shanta Dutta, Saumitra Das
EPI_ISL_430467 hCoV-19/India/S11/2020 Asia / India / West Bengal / East Medinipur 2020-04-03 ICMR-National Institute of Cholera and Enteric Diseases National Institute of Biomedical Genomics Arindam Maitra, Mamta Chawla Sarkar, Sreedhar Chinnaswamy, Hasina Banu, Ananya Chatterjee, Shanta Dutta, Saumitra Das
EPI_ISL_430468 hCoV-19/India/S2/2020 Asia / India / West_Bengal / Kolkata 2020-03-21 ICMR-National Institute of Cholera and Enteric Diseases National Institute of Biomedical Genomics Arindam Maitra, Mamta Chawla Sarkar, Sreedhar Chinnaswamy, Hasina Banu, Ananya Chatterjee, Shanta Dutta, Saumitra Das
EPI_ISL_430469 hCoV-19/Greece/127_HPI/2020 Europe / Greece / Athens 2020-02-29 Hellenic Pasteur Institute, Public Health Laboratories Hellenic Pasteur Institute, Public Health Laboratories, Unit of Bioinformatics and Applied Genomics Vasiliki Pogka, Timokratis Karamitros, Athanasios Kossyvakis, Antonios Kalliaropoulos, Horefti Elina, Evangelidou Maria, Androniki Voulgari-Kokota, Aspasia Kontou, Andreas Mentis
EPI_ISL_430471 hCoV-19/Australia/VIC935/2020 Oceania / Australia / Victoria 2020-03-25 Microbiological Diagnostic Unit Public Health Laboratory Microbiological Diagnostic Unit Public Health Laboratory Seemann T., Schultz M., Sait, M., Sherry, N.
EPI_ISL_430472 hCoV-19/Australia/VIC938/2020 Oceania / Australia / Victoria 2020-03-25 Microbiological Diagnostic Unit Public Health Laboratory Microbiological Diagnostic Unit Public Health Laboratory Seemann T., Schultz M., Sait, M., Sherry, N.
EPI_ISL_430473 hCoV-19/Australia/VIC1008/2020 Oceania / Australia / Victoria 2020-03-31 Victorian Infectious Diseases Reference Laboratory (VIDRL) Microbiological Diagnostic Unit Public Health Laboratory and Victorian Infectious Diseases Reference Laboratory, The Peter Doherty Institute for Infection and Immunity Caly L., Seemann T., Sait, M., Schultz M., Druce J., Sherry, N.
EPI_ISL_430476 hCoV-19/Australia/VIC1016/2020 Oceania / Australia / Victoria 2020-04-01 Victorian Infectious Diseases Reference Laboratory (VIDRL) Microbiological Diagnostic Unit Public Health

Laboratory and Victorian Infectious Diseases Reference Laboratory, The Peter Doherty Institute for Infection and Immunity    Caly L., Seemann T., Sait, M., Schultz M., Druce J., Sherry, N.
EPI_ISL_430478  hCoV-19/Australia/VIC994/2020   Oceania / Australia / Victoria  2020-03-31      Victorian Infectious Diseases Reference Laboratory (VIDRL)        Microbiological Diagnostic Unit Public Health Laboratory and Victorian Infectious Diseases Reference Laboratory, The Peter Doherty Institute for Infection and Immunity    Caly L., Seemann T., Sait, M., Schultz M., Druce J., Sherry, N.
EPI_ISL_430479  hCoV-19/Australia/VIC995/2020   Oceania / Australia / Victoria  2020-03-31      Victorian Infectious Diseases Reference Laboratory (VIDRL)        Microbiological Diagnostic Unit Public Health Laboratory and Victorian Infectious Diseases Reference Laboratory, The Peter Doherty Institute for Infection and Immunity    Caly L., Seemann T., Sait, M., Schultz M., Druce J., Sherry, N.
EPI_ISL_430481  hCoV-19/Australia/VIC997/2020   Oceania / Australia / Victoria  2020-03-31      Victorian Infectious Diseases Reference Laboratory (VIDRL)        Microbiological Diagnostic Unit Public Health Laboratory and Victorian Infectious Diseases Reference Laboratory, The Peter Doherty Institute for Infection and Immunity    Caly L., Seemann T., Sait, M., Schultz M., Druce J., Sherry, N.
EPI_ISL_430482  hCoV-19/Australia/VIC998/2020   Oceania / Australia / Victoria  2020-03-31      Victorian Infectious Diseases Reference Laboratory (VIDRL)        Microbiological Diagnostic Unit Public Health Laboratory and Victorian Infectious Diseases Reference Laboratory, The Peter Doherty Institute for Infection and Immunity    Caly L., Seemann T., Sait, M., Schultz M., Druce J., Sherry, N.
EPI_ISL_430484  hCoV-19/Australia/VIC1000/2020  Oceania / Australia / Victoria  2020-03-31      Victorian Infectious Diseases Reference Laboratory (VIDRL)        Microbiological Diagnostic Unit Public Health Laboratory and Victorian Infectious Diseases Reference Laboratory, The Peter Doherty Institute for Infection and Immunity    Caly L., Seemann T., Sait, M., Schultz M., Druce J., Sherry, N.
EPI_ISL_430486  hCoV-19/Australia/VIC1002/2020  Oceania / Australia / Victoria  2020-03-31      Victorian Infectious Diseases Reference Laboratory (VIDRL)        Microbiological Diagnostic Unit Public Health Laboratory and Victorian Infectious Diseases Reference Laboratory, The Peter Doherty Institute for Infection and Immunity    Caly L., Seemann T., Sait, M., Schultz M., Druce J., Sherry, N.
EPI_ISL_430490  hCoV-19/Australia/VIC1018/2020  Oceania / Australia / Victoria  2020-04-01      Victorian Infectious Diseases Reference Laboratory (VIDRL)        Microbiological Diagnostic Unit Public Health Laboratory and Victorian Infectious Diseases Reference Laboratory, The Peter Doherty Institute for Infection and Immunity    Caly L., Seemann T., Sait, M., Schultz M., Druce J., Sherry, N.
EPI_ISL_430498  hCoV-19/Australia/VIC1208/2020  Oceania / Australia / Victoria  2020-04-01      Victorian Infectious Diseases Reference Laboratory (VIDRL)        Microbiological Diagnostic Unit Public Health Laboratory and Victorian Infectious Diseases Reference Laboratory, The Peter Doherty Institute for Infection and Immunity    Caly L., Seemann T., Sait, M., Schultz M., Druce J., Sherry, N.
EPI_ISL_406716  hCoV-19/Wuhan/WHU01/2020        Asia / China / Hubei / Wuhan    2020-01-02      unknown State Key Laboratory of Virology, Wuhan University    Chen,L., Liu,W., Zhang,Q., Xu,K., Ye,G., Wu,W., Sun,Z., Liu,F., Wu,K., Mei,Y., Zhang,W., Chen,Y., Li,Y., Shi,M., Lan,K. and Liu,Y.
EPI_ISL_406717  hCoV-19/Wuhan/WHU02/2020        Asia / China / Hubei / Wuhan    2020-01-02      unknown State Key Laboratory of Virology, Wuhan University    Chen,L., Liu,W., Zhang,Q., Xu,K., Ye,G., Wu,W., Sun,Z., Liu,F., Wu,K., Mei,Y., Zhang,W., Chen,Y., Li,Y., Shi,M., Lan,K. and Liu,Y.
EPI_ISL_410486  hCoV-19/France/RA739/2020       Europe / France / Rhone-Alpes / Contamines      2020-02-08      CNR Virus des Infections Respiratoires - France SUD     CNR Virus des Infections Respiratoires - France SUD     Bal, Antonin; Destras, Gregory; Gaymard, Alexandre; Bouscambert-Duchamp, Maude; Cheynet, Valérie; Brengel-Pesce, Karen; Morfin-Sherpa, Florence; Valette, Martine; Josset, Laurence; Lina, Bruno.
EPI_ISL_410531  hCoV-19/Japan/NA-20-05-1/2020   Asia / Japan / Nara     2020-01-25      Dept. of Pathology, National Institute of Infectious Diseases       Pathogen Genomics Center, National Institute of Infectious Diseases       Tsuyoshi Sekizuka, Harutaka Katano, Shutoku Matsuyama, Naganori Nao, Kazuya Shirato, Motoi Suzuki, Hideki Hasegawa, Takaji Wakita, Makoto Takeda, Tadaki Suzuki, Makoto Kuroda
EPI_ISL_410532  hCoV-19/Japan/OS-20-07-1/2020   Asia / Japan / Osaka    2020-01-23      Dept. of Pathology, National Institute of Infectious Diseases       Pathogen Genomics Center, National Institute of Infectious Diseases       Tsuyoshi Sekizuka, Harutaka Katano, Shutoku Matsuyama, Naganori Nao, Kazuya Shirato, Motoi Suzuki, Hideki Hasegawa, Takaji Wakita, Makoto Takeda, Tadaki Suzuki, Makoto Kuroda
EPI_ISL_410535  hCoV-19/Singapore/4/2020        Asia / Singapore        2020-02-03      National Centre for Infectious Diseases     Programme in Emerging Infectious Diseases, Duke-NUS Medical School      Danielle E Anderson, Martin Linster, Yan Zhuang, Jayanthi Jayakumar, David CB Lye, Yee Sin Leo, Barnaby E Young, Yvonne CF Su, Gavin JD Smith
EPI_ISL_410536  hCoV-19/Singapore/5/2020        Asia / Singapore        2020-02-06      Singapore General Hospital, Molecular Laboratory, Division of Pathology   Programme in Emerging Infectious Diseases, Duke-NUS Medical School  Danielle E Anderson, Martin Linster, Yan Zhuang, Jayanthi Jayakumar, Kian Sing Chan, Lynette LE Oon, Shirin Kalimuddin, Jenny GH Low, Yvonne CF Su, Gavin JD Smith
EPI_ISL_410537  hCoV-19/Singapore/6/2020        Asia / Singapore        2020-02-09      Singapore General Hospital, Molecular Laboratory, Division of Pathology   Programme in Emerging Infectious Diseases, Duke-NUS Medical School  Danielle E Anderson, Martin Linster, Yan Zhuang, Jayanthi Jayakumar, Kian Sing Chan, Lynette LE Oon, Shirin Kalimuddin, Jenny GH Low, Yvonne CF Su, Gavin JD Smith
EPI_ISL_410545  hCoV-19/Italy/INMI1-isl/2020    Europe / Italy / Rome   2020-01-29      INMI Lazzaro Spallanzani IRCCS       Laboratory of Virology, INMI Lazzaro Spallanzani IRCCS  Maria R. Capobianchi, Cesare E. M. Gruber, Martina Rueca, Barbara Bartolini, Francesco Messina, Emanuela Giombini, Francesca Colavita, Concetta Castilletti, Eleonora Lalle, Fabrizio Carletti, Emanuele Nicastri, Giuseppe Ippolito.
EPI_ISL_410546  hCoV-19/Italy/INMI1-cs/2020     Europe / Italy / Rome   2020-01-31      INMI Lazzaro Spallanzani IRCCS       Laboratory of Virology, INMI Lazzaro Spallanzani IRCCS  Maria R. Capobianchi, Cesare E.

M. Gruber, Martina Rueca, Fabrizio Carletti, Barbara Bartolini, Francesco Messina, Emanuela Giombini, Francesca Colavita, Concetta Castilletti, Eleonora Lalle, Emanuele Nicastri, Giuseppe Ippolito.
EPI_ISL_411218 hCoV-19/France/IDF0571/2020 Europe / France / Ile-de-France / Paris 2020-02-02 Department of Infectious and Tropical Diseases, Bichat Claude Bernard Hospital, Paris Laboratoire Virpath, CIRI U111, UCBL1, INSERM, CNRS, ENS Lyon Olivier Terrier, Aurélien Traversier, Julien Fouret, Yazdan Yazdanpanah, Xavier Lescure, Catherine Legras-Lachuer, Alexandre Gaymard, Bruno Lina, Manuel Rosa-Calatrava
EPI_ISL_411219 hCoV-19/France/IDF0386-islP1/2020 Europe / France / Ile-de-France / Paris 2020-01-28 Department of Infectious and Tropical Diseases, Bichat Claude Bernard Hospital, Paris Laboratoire Virpath, CIRI U111, UCBL1, INSERM, CNRS, ENS Lyon Olivier Terrier, Aurélien Traversier, Julien Fouret, Yazdan Yazdanpanah, Xavier Lescure, Alexandre Gaymard, Bruno Lina, Manuel Rosa-Calatrava
EPI_ISL_411220 hCoV-19/France/IDF0386-islP3/2020 Europe / France / Ile-de-France / Paris 2020-01-28 Department of Infectious and Tropical Diseases, Bichat Claude Bernard Hospital, Paris Laboratoire Virpath, CIRI U111, UCBL1, INSERM, CNRS, ENS Lyon Olivier Terrier, Aurélien Traversier, Julien Fouret, Yazdan Yazdanpanah, Xavier Lescure, Alexandre Gaymard, Bruno Lina, Manuel Rosa-Calatrava
EPI_ISL_411950 hCoV-19/Jiangsu/JS01/2020 Asia / China / Jiangsu 2020-01-23 NHC Key laboratory of Enteric Pathogenic Microbiology, Institute of Pathogenic Microbiology Jiangsu Provincial Center for Disease Control & Prevention Lunbiao Cui,Kangchen Zhao,Xiaojuan Zhu,Yiyue Ge,Tao Wu,Bin Wu,Yin Chen,Fengcai Zhu,Baoli Zhu,Ming Wu
EPI_ISL_411951 hCoV-19/Sweden/01/2020 Europe / Sweden 2020-02-07 unknown Unit for Laboratory Development and Technology Transfer, Public Health Agency of Sweden Bengner,M., Palmerus,M., Lindsjo,O., Lind Karlberg,M., Monteil,V., Appelberg,S., Brave,A., Muradrasoli,S. and Tegmark-Wisell,K.
EPI_ISL_411952 hCoV-19/Jiangsu/JS02/2020 Asia / China / Jiangsu 2020-01-24 NHC Key laboratory of Enteric Pathogenic Microbiology, Institute of Pathogenic Microbiology Jiangsu Provincial Center for Disease Control & Prevention Kangchen Zhao, Xiaojuan Zhu, Lunbiao Cui, Tao Wu, Yiyue Ge, Bin Wu, Yin Chen, Fengcai Zhu, Baoli Zhu, Ming Wu
EPI_ISL_411953 hCoV-19/Jiangsu/JS03/2020 Asia / China / Jiangsu 2020-01-24 NHC Key laboratory of Enteric Pathogenic Microbiology, Institute of Pathogenic Microbiology Jiangsu Provincial Center for Disease Control & Prevention Kangchen Zhao, Xiaojuan Zhu, Lunbiao Cui, Tao Wu, Yiyue Ge, Bin Wu, Yin Chen, Fengcai Zhu, Baoli Zhu, Ming Wu
EPI_ISL_411954 hCoV-19/USA/CA7/2020 North America / USA / California 2020-02-06 California Department of Public Health Pathogen Discovery, Respiratory Viruses Branch, Division of Viral Diseases, Centers for Dieases Control and Prevention Krista Queen, Anna Uehara, Jing Zhang, Yan Li, Ying Tao, Clinton R. Paden, Haibin Wang, Shifaq Kamili, Xiaoyan Lu, Brian Lynch, Senthil Kumar K. Sakthivel, Brett L. Whitaker, Lijuan Wang, Janna' R. Murray, Susan I. Gerber, Stephen Lindstrom, Suxiang Tong
EPI_ISL_411955 hCoV-19/USA/CA8/2020 North America / USA / California 2020-02-10 California Department of Public Health Pathogen Discovery, Respiratory Viruses Branch, Division of Viral Diseases, Centers for Dieases Control and Prevention Krista Queen, Anna Uehara, Jing Zhang, Yan Li, Ying Tao, Clinton R. Paden, Haibin Wang, Shifaq Kamili, Xiaoyan Lu, Brian Lynch, Senthil Kumar K. Sakthivel, Brett L. Whitaker, Lijuan Wang, Janna' R. Murray, Susan I. Gerber, Stephen Lindstrom, Suxiang Tong
EPI_ISL_411956 hCoV-19/USA/TX1/2020 North America / USA / Texas 2020-02-11 Texas Department of State Health Services Pathogen Discovery, Respiratory Viruses Branch, Division of Viral Diseases, Centers for Dieases Control and Prevention Krista Queen, Anna Uehara, Jing Zhang, Yan Li, Ying Tao, Clinton R. Paden, Haibin Wang, Shifaq Kamili, Xiaoyan Lu, Brian Lynch, Senthil Kumar K. Sakthivel, Brett L. Whitaker, Lijuan Wang, Janna' R. Murray, Susan I. Gerber, Stephen Lindstrom, Suxiang Tong
EPI_ISL_411957 hCoV-19/China/WH-09/2020 Asia / China 2020-01-08 unknown Key Laboratory of Human Diseases, Comparative Medicine, Institute of Laboratory Animal Science Linlin,B., Lili,R., Shuran,G., Jiangning,L., Feifei,Q., Qi,L., Fengdi,L., Jing,X., Wei,D., Pin,Y., Yanfeng,X., Yajin,Q., Hong,G., Qiang,W., Mingya,L., Guanpeng,W., Shunyi,W., Zhiqi,S., Li,G., Lan,C., Conghui,W., Ying,W., Xinming,W., Yan,X., Qi,J. and Chuan,Q.
EPI_ISL_413851 hCoV-19/Guangdong/2020XN4373-P0039/2020 Asia / China / Guangdong 2020-01-30 Guangdong Provincial Institution of Public Health, Guangdong Provinical Center for Disease Control and Prevention Guangdong Provincial Institution of Public Health Jing Lu, Louis du Plessis, Liu Zhe, Jiufeng Sun, Sarah François, Huifang Lin, Moritz Kraemer, Jingju Peng, Qianlin Xiong, Runyu Yuan, Lilian Zeng, Pingping Zhou, Chuming Liang, Tao Liu, Wei Li, Juan Su, Huanying Zheng, Kang Min, Song Tie, Bo Peng, Shisong Fang, Wenzhe Su, Kuibiao Li, Ruilin Sun, Ru bai, Xi Tang, Minfeng Liang, Nuno Faria, Josh Quick, Andrew Rambaut, Verity Hill, Wenjun Ma, Nick Loman, Oliver Pybus, Changwen Ke
EPI_ISL_413852 hCoV-19/Guangdong/2020XN4433-P0040/2020 Asia / China / Guangdong 2020-01-30 Guangdong Provincial Institution of Public Health, Guangdong Provinical Center for Disease Control and Prevention Guangdong Provincial Institution of Public Health Jing Lu, Louis du Plessis, Liu Zhe, Jiufeng Sun, Sarah François, Huifang Lin, Moritz Kraemer, Jingju Peng, Qianlin Xiong, Runyu Yuan, Lilian Zeng, Pingping Zhou, Chuming Liang, Tao Liu, Wei Li, Juan Su, Huanying Zheng, Kang Min, Song Tie, Bo Peng, Shisong Fang, Wenzhe Su, Kuibiao Li, Ruilin Sun, Ru bai, Xi Tang, Minfeng Liang, Nuno Faria, Josh Quick, Andrew Rambaut, Verity Hill, Wenjun Ma, Nick Loman, Oliver Pybus, Changwen Ke
EPI_ISL_413853 hCoV-19/Guangdong/2020XN4243-P0035/2020 Asia / China / Guangdong 2020-01-30 Guangdong Provincial Institution of Public Health, Guangdong Provinical Center for Disease Control and Prevention Guangdong Provincial Institution of Public Health Jing Lu, Louis du Plessis, Liu Zhe, Jiufeng Sun, Sarah François, Huifang Lin, Moritz Kraemer, Jingju Peng, Qianlin Xiong, Runyu Yuan, Lilian Zeng, Pingping Zhou, Chuming Liang, Tao Liu, Wei Li, Juan Su, Huanying Zheng, Kang Min, Song Tie, Bo Peng, Shisong Fang, Wenzhe Su, Kuibiao Li, Ruilin Sun, Ru bai, Xi Tang, Minfeng Liang, Nuno Faria, Josh Quick, Andrew Rambaut, Verity Hill, Wenjun Ma, Nick Loman, Oliver Pybus, Changwen Ke

EPI_ISL_413854	hCoV-19/Guangdong/2020XN4475-P0042/2020	Asia / China / Guangdong	2020-01-30	Guangdong Provincial Institution of Public Health, Guangdong Provinical Center for Disease Control and Prevention	Guangdong Provincial Institution of Public Health	Jing Lu, Louis du Plessis, Liu Zhe, Jiufeng Sun, Sarah François, Huifang Lin, Moritz Kraemer, Jingju Peng, Qianlin Xiong, Runyu Yuan, Lilian Zeng, Pingping Zhou, Chuming Liang, Tao Liu, Wei Li, Juan Su, Huanying Zheng, Kang Min, Song Tie, Bo Peng, Shisong Fang, Wenzhe Su, Kuibiao Li, Ruilin Sun, Ru bai, Xi Tang, Minfeng Liang, Nuno Faria, Josh Quick, Andrew Rambaut, Verity Hill, Wenjun Ma, Nick Loman, Oliver Pybus, Changwen Ke
EPI_ISL_413855	hCoV-19/Guangdong/GD2020234-P0023/2020	Asia / China / Guangdong	2020-02-07	Guangdong Provincial Institution of Public Health, Guangdong Provinical Center for Disease Control and Prevention	Guangdong Provincial Institution of Public Health	Jing Lu, Louis du Plessis, Liu Zhe, Jiufeng Sun, Sarah François, Huifang Lin, Moritz Kraemer, Jingju Peng, Qianlin Xiong, Runyu Yuan, Lilian Zeng, Pingping Zhou, Chuming Liang, Tao Liu, Wei Li, Juan Su, Huanying Zheng, Kang Min, Song Tie, Bo Peng, Shisong Fang, Wenzhe Su, Kuibiao Li, Ruilin Sun, Ru bai, Xi Tang, Minfeng Liang, Nuno Faria, Josh Quick, Andrew Rambaut, Verity Hill, Wenjun Ma, Nick Loman, Oliver Pybus, Changwen Ke
EPI_ISL_413856	hCoV-19/Guangdong/GD2020233-P0027/2020	Asia / China / Guangdong	2020-02-07	Guangdong Provincial Institution of Public Health, Guangdong Provinical Center for Disease Control and Prevention	Guangdong Provincial Institution of Public Health	Jing Lu, Louis du Plessis, Liu Zhe, Jiufeng Sun, Sarah François, Huifang Lin, Moritz Kraemer, Jingju Peng, Qianlin Xiong, Runyu Yuan, Lilian Zeng, Pingping Zhou, Chuming Liang, Tao Liu, Wei Li, Juan Su, Huanying Zheng, Kang Min, Song Tie, Bo Peng, Shisong Fang, Wenzhe Su, Kuibiao Li, Ruilin Sun, Ru bai, Xi Tang, Minfeng Liang, Nuno Faria, Josh Quick, Andrew Rambaut, Verity Hill, Wenjun Ma, Nick Loman, Oliver Pybus, Changwen Ke
EPI_ISL_413857	hCoV-19/Guangdong/2020XN4448-P0002/2020	Asia / China / Guangdong	2020-01-31	Guangdong Provincial Institution of Public Health, Guangdong Provinical Center for Disease Control and Prevention	Guangdong Provincial Institution of Public Health	Jing Lu, Louis du Plessis, Liu Zhe, Jiufeng Sun, Sarah François, Huifang Lin, Moritz Kraemer, Jingju Peng, Qianlin Xiong, Runyu Yuan, Lilian Zeng, Pingping Zhou, Chuming Liang, Tao Liu, Wei Li, Juan Su, Huanying Zheng, Kang Min, Song Tie, Bo Peng, Shisong Fang, Wenzhe Su, Kuibiao Li, Ruilin Sun, Ru bai, Xi Tang, Minfeng Liang, Nuno Faria, Josh Quick, Andrew Rambaut, Verity Hill, Wenjun Ma, Nick Loman, Oliver Pybus, Changwen Ke
EPI_ISL_413858	hCoV-19/Guangdong/2020XN4459-P0041/2020	Asia / China / Guangdong	2020-01-30	Guangdong Provincial Institution of Public Health, Guangdong Provinical Center for Disease Control and Prevention	Guangdong Provincial Institution of Public Health	Jing Lu, Louis du Plessis, Liu Zhe, Jiufeng Sun, Sarah François, Huifang Lin, Moritz Kraemer, Jingju Peng, Qianlin Xiong, Runyu Yuan, Lilian Zeng, Pingping Zhou, Chuming Liang, Tao Liu, Wei Li, Juan Su, Huanying Zheng, Kang Min, Song Tie, Bo Peng, Shisong Fang, Wenzhe Su, Kuibiao Li, Ruilin Sun, Ru bai, Xi Tang, Minfeng Liang, Nuno Faria, Josh Quick, Andrew Rambaut, Verity Hill, Wenjun Ma, Nick Loman, Oliver Pybus, Changwen Ke
EPI_ISL_413860	hCoV-19/Guangdong/2020XN4273-P0036/2020	Asia / China / Guangdong	2020-01-30	Guangdong Provincial Institution of Public Health, Guangdong Provinical Center for Disease Control and Prevention	Guangdong Provincial Institution of Public Health	Jing Lu, Louis du Plessis, Liu Zhe, Jiufeng Sun, Sarah François, Huifang Lin, Moritz Kraemer, Jingju Peng, Qianlin Xiong, Runyu Yuan, Lilian Zeng, Pingping Zhou, Chuming Liang, Tao Liu, Wei Li, Juan Su, Huanying Zheng, Kang Min, Song Tie, Bo Peng, Shisong Fang, Wenzhe Su, Kuibiao Li, Ruilin Sun, Ru bai, Xi Tang, Minfeng Liang, Nuno Faria, Josh Quick, Andrew Rambaut, Verity Hill, Wenjun Ma, Nick Loman, Oliver Pybus, Changwen Ke
EPI_ISL_413861	hCoV-19/Guangdong/GD2020080-P0010/2020	Asia / China / Guangdong	2020-02-01	Guangdong Provincial Institution of Public Health, Guangdong Provinical Center for Disease Control and Prevention	Guangdong Provincial Institution of Public Health	Jing Lu, Louis du Plessis, Liu Zhe, Jiufeng Sun, Sarah François, Huifang Lin, Moritz Kraemer, Jingju Peng, Qianlin Xiong, Runyu Yuan, Lilian Zeng, Pingping Zhou, Chuming Liang, Tao Liu, Wei Li, Juan Su, Huanying Zheng, Kang Min, Song Tie, Bo Peng, Shisong Fang, Wenzhe Su, Kuibiao Li, Ruilin Sun, Ru bai, Xi Tang, Minfeng Liang, Nuno Faria, Josh Quick, Andrew Rambaut, Verity Hill, Wenjun Ma, Nick Loman, Oliver Pybus, Changwen Ke
EPI_ISL_413862	hCoV-19/Guangdong/GD2020086-P0021/2020	Asia / China / Guangdong	2020-02-01	Guangdong Provincial Institution of Public Health, Guangdong Provinical Center for Disease Control and Prevention	Guangdong Provincial Institution of Public Health	Jing Lu, Louis du Plessis, Liu Zhe, Jiufeng Sun, Sarah François, Huifang Lin, Moritz Kraemer, Jingju Peng, Qianlin Xiong, Runyu Yuan, Lilian Zeng, Pingping Zhou, Chuming Liang, Tao Liu, Wei Li, Juan Su, Huanying Zheng, Kang Min, Song Tie, Bo Peng, Shisong Fang, Wenzhe Su, Kuibiao Li, Ruilin Sun, Ru bai, Xi Tang, Minfeng Liang, Nuno Faria, Josh Quick, Andrew Rambaut, Verity Hill, Wenjun Ma, Nick Loman, Oliver Pybus, Changwen Ke
EPI_ISL_413863	hCoV-19/Guangdong/GD2020087-P0008/2020	Asia / China / Guangdong	2020-02-01	Guangdong Provincial Institution of Public Health, Guangdong Provinical Center for Disease Control and Prevention	Guangdong Provincial Institution of Public Health	Jing Lu, Louis du Plessis, Liu Zhe, Jiufeng Sun, Sarah François, Huifang Lin, Moritz Kraemer, Jingju Peng, Qianlin Xiong, Runyu Yuan, Lilian Zeng, Pingping Zhou, Chuming Liang, Tao Liu, Wei Li, Juan Su, Huanying Zheng, Kang Min, Song Tie, Bo Peng, Shisong Fang, Wenzhe Su, Kuibiao Li, Ruilin Sun, Ru bai, Xi Tang, Minfeng Liang, Nuno Faria, Josh Quick, Andrew Rambaut, Verity Hill, Wenjun Ma, Nick Loman, Oliver Pybus, Changwen Ke
EPI_ISL_413864	hCoV-19/Guangdong/GD2020246-P0028/2020	Asia / China / Guangdong	2020-02-09	Guangdong Provincial Institution of Public Health, Guangdong Provinical Center for Disease Control and Prevention	Guangdong Provincial Institution of Public Health	Jing Lu, Louis du Plessis, Liu Zhe, Jiufeng Sun, Sarah François, Huifang Lin, Moritz Kraemer, Jingju Peng, Qianlin Xiong, Runyu Yuan, Lilian Zeng, Pingping Zhou, Chuming Liang, Tao Liu, Wei Li, Juan Su, Huanying Zheng, Kang Min, Song Tie, Bo Peng, Shisong Fang, Wenzhe Su, Kuibiao Li, Ruilin Sun, Ru bai, Xi Tang, Minfeng Liang, Nuno Faria, Josh Quick, Andrew Rambaut, Verity Hill, Wenjun Ma, Nick Loman, Oliver Pybus, Changwen Ke

EPI_ISL_413867  hCoV-19/Guangdong/GDSZ202015-P0019/2020 Asia / China / Guangdong        2020-02-05
Guangdong Provincial Institution of Public Health, Guangdong Provinical Center for Disease Control and Prevention      Guangdong Provincial Institution of Public Health       Jing Lu, Louis du Plessis, Liu Zhe, Jiufeng Sun, Sarah François, Huifang Lin, Moritz Kraemer, Jingju Peng, Qianlin Xiong, Runyu Yuan, Lilian Zeng, Pingping Zhou, Chuming Liang, Tao Liu, Wei Li, Juan Su, Huanying Zheng, Kang Min, Song Tie, Bo Peng, Shisong Fang, Wenzhe Su, Kuibiao Li, Ruilin Sun, Ru bai, Xi Tang, Minfeng Liang, Nuno Faria, Josh Quick, Andrew Rambaut, Verity Hill, Wenjun Ma, Nick Loman, Oliver Pybus, Changwen Ke
EPI_ISL_414005  hCoV-19/England/200940527/2020  Europe / United Kingdom / England       2020-02-25
Respiratory Virus Unit, Microbiology Services Colindale, Public Health England  Respiratory Virus Unit, Microbiology Services Colindale, Public Health England  Monica Galiano, Shahjahan Miah, Angie Lackenby, Omolola Akinbami, Tiina Talts, Leena Bhaw, Richard Myers, Steven Platt, Kirstin Edwards, Jonathan Hubb, Joanna Ellis, Maria Zambon
EPI_ISL_414006  hCoV-19/England/200990724/2020  Europe / United Kingdom / England       2020-02-28
Respiratory Virus Unit, Microbiology Services Colindale, Public Health England  Respiratory Virus Unit, Microbiology Services Colindale, Public Health England  Monica Galiano, Shahjahan Miah, Angie Lackenby, Omolola Akinbami, Tiina Talts, Leena Bhaw, Richard Myers, Steven Platt, Kirstin Edwards, Jonathan Hubb, Joanna Ellis, Maria Zambon
EPI_ISL_414007  hCoV-19/England/200990725/2020  Europe / United Kingdom / England       2020-02-28
Respiratory Virus Unit, Microbiology Services Colindale, Public Health England  Respiratory Virus Unit, Microbiology Services Colindale, Public Health England  Monica Galiano, Shahjahan Miah, Angie Lackenby, Omolola Akinbami, Tiina Talts, Leena Bhaw, Richard Myers, Steven Platt, Kirstin Edwards, Jonathan Hubb, Joanna Ellis, Maria Zambon
EPI_ISL_414008  hCoV-19/England/200960041/2020  Europe / United Kingdom / England       2020-02-27
Respiratory Virus Unit, Microbiology Services Colindale, Public Health England  Respiratory Virus Unit, Microbiology Services Colindale, Public Health England  Monica Galiano, Shahjahan Miah, Angie Lackenby, Omolola Akinbami, Tiina Talts, Leena Bhaw, Richard Myers, Steven Platt, Kirstin Edwards, Jonathan Hubb, Joanna Ellis, Maria Zambon
EPI_ISL_414009  hCoV-19/England/200960515/2020  Europe / United Kingdom / England       2020-02-25
Respiratory Virus Unit, Microbiology Services Colindale, Public Health England  Respiratory Virus Unit, Microbiology Services Colindale, Public Health England  Monica Galiano, Shahjahan Miah, Angie Lackenby, Omolola Akinbami, Tiina Talts, Leena Bhaw, Richard Myers, Steven Platt, Kirstin Edwards, Jonathan Hubb, Joanna Ellis, Maria Zambon
EPI_ISL_414010  hCoV-19/England/200981386/2020  Europe / United Kingdom / England       2020-02-26
Respiratory Virus Unit, Microbiology Services Colindale, Public Health England  Respiratory Virus Unit, Microbiology Services Colindale, Public Health England  Monica Galiano, Shahjahan Miah, Angie Lackenby, Omolola Akinbami, Tiina Talts, Leena Bhaw, Richard Myers, Steven Platt, Kirstin Edwards, Jonathan Hubb, Joanna Ellis, Maria Zambon
EPI_ISL_414011  hCoV-19/England/200990006/2020  Europe / United Kingdom / England       2020-02-26
Respiratory Virus Unit, Microbiology Services Colindale, Public Health England  Respiratory Virus Unit, Microbiology Services Colindale, Public Health England  Monica Galiano, Shahjahan Miah, Angie Lackenby, Omolola Akinbami, Tiina Talts, Leena Bhaw, Richard Myers, Steven Platt, Kirstin Edwards, Jonathan Hubb, Joanna Ellis, Maria Zambon
EPI_ISL_414012  hCoV-19/England/200990723/2020  Europe / United Kingdom / England       2020-02-27
Respiratory Virus Unit, Microbiology Services Colindale, Public Health England  Respiratory Virus Unit, Microbiology Services Colindale, Public Health England  Monica Galiano, Shahjahan Miah, Angie Lackenby, Omolola Akinbami, Tiina Talts, Leena Bhaw, Richard Myers, Steven Platt, Kirstin Edwards, Jonathan Hubb, Joanna Ellis, Maria Zambon
EPI_ISL_414013  hCoV-19/England/201000003/2020  Europe / United Kingdom / England       2020-03-01
Respiratory Virus Unit, Microbiology Services Colindale, Public Health England  Respiratory Virus Unit, Microbiology Services Colindale, Public Health England  Monica Galiano, Shahjahan Miah, Angie Lackenby, Omolola Akinbami, Tiina Talts, Leena Bhaw, Richard Myers, Steven Platt, Kirstin Edwards, Jonathan Hubb, Joanna Ellis, Maria Zambon
EPI_ISL_414014  hCoV-19/Brazil/SPBR-03/2020     South America / Brazil / Sao Paulo      2020-03-02
Hospital Israelita Albert Einstein      Instituto Adolfo Lutz, Interdiciplinary Procedures Center, Strategic Laboratory      Claudio Tavares Sacchi, Claudia Regina Gonçalves, Katia Correia dos Santos, Carlos Henrique Camargo, Maria do Carmo Sampaito Tavares Timenetsky, Terezinha Maria de Paiva, Ester Cerdeira Sabino
EPI_ISL_414015  hCoV-19/Brazil/SPBR-06/2020     South America / Brazil / Sao Paulo / Sao Paulo  2020-02-29
Hospital São Joaquim Beneficencia Portuguesa    Instituto Adolfo Lutz, Interdiciplinary Procedures Center, Strategic Laboratory    Claudio Tavares Sacchi, Claudia Regina Gonçalves, SimoneGuadagnucci Morillo, Carlos Henrique Camargo, Maria do Carmo Sampaito Tavares Timenetsky, Fabiana Cristina Pereira dos Santos Terezinha Maria de Paiva, Ester Cerdeira Sabino
EPI_ISL_414016  hCoV-19/Brazil/SPBR-05/2020     South America / Brazil / Sao Paulo / Sao Paulo  2020-02-29
Hospital São Joaquim Beneficencia Portuguesa    Instituto Adolfo Lutz, Interdiciplinary Procedures Center, Strategic Laboratory    Claudio Tavares Sacchi, Claudia Regina Gonçalves, Audrey Cilli, Carlos Henrique Camargo, Maria do Carmo Sampaito Tavares Timenetsky, Daniela Bernardes Borges da Silva, Terezinha Maria de Paiva, Ester Cerdeira Sabino
EPI_ISL_414017  hCoV-19/Brazil/SPBR-04/2020     South America / Brazil / Sao Paulo      2020-03-04
Hospital São Joaquim Beneficencia Portuguesa    Instituto Adolfo Lutz, Interdiciplinary Procedures Center, Strategic Laboratory    Claudio Tavares Sacchi, Claudia Regina Gonçalves, Fabiana Cristina Pereira dos Santos, Carlos Henrique Camargo, Maria do Carmo Sampaito Tavares Timenetsky, Daniela Bernardes Borges da Silva,

Terezinha Maria de Paiva, Ester Cerdeira Sabino
EPI_ISL_414019 hCoV-19/Switzerland/GE3121/2020 Europe / Switzerland / Geneva 2020-02-27 Laboratoire de Virologie, HUG Swiss National Reference Centre for Influenza LAUBSCHER Florian et al.
EPI_ISL_414020 hCoV-19/Switzerland/GE5373/2020 Europe / Switzerland / Geneva 2020-02-27 Laboratoire de Virologie, HUG Swiss National Reference Centre for Influenza LAUBSCHER Florian et al.
EPI_ISL_414021 hCoV-19/Switzerland/BL0902/2020 Europe / Switzerland / Basel 2020-02-27 Laboratoire de Virologie, HUG Swiss National Reference Centre for Influenza LAUBSCHER Florian et al.
EPI_ISL_414022 hCoV-19/Switzerland/GE9586/2020 Europe / Switzerland / Geneva 2020-02-27 Laboratoire de Virologie, HUG Swiss National Reference Centre for Influenza LAUBSCHER Florian et al.
EPI_ISL_414023 hCoV-19/Switzerland/VD5615/2020 Europe / Switzerland / Vaud 2020-03-01 Laboratoire de Virologie, HUG Swiss National Reference Centre for Influenza LAUBSCHER Florian et al.
EPI_ISL_414027 hCoV-19/Scotland/CVR05/2020 Europe / United Kingdom / Scotland 2020-03-04 West of Scotland Specialist Virology Centre, NHSGGC MRC-University of Glasgow Centre for Virus Research Emma Thomson, Antonia Ho; Kathy Smollett, Daniel Mair, Stephen Carmichael, Ana da Silva Filipe; Richard Orton, David L Robertson; Alasdair MacLean, Rory Gunson.
EPI_ISL_414040 hCoV-19/England/200641094/2020 Europe / United Kingdom / England 2020-02-05 Respiratory Virus Unit, Microbiology Services Colindale, Public Health England Respiratory Virus Unit, Microbiology Services Colindale, Public Health England Monica Galiano, Shahjahan Miah, Angie Lackenby, Omolola Akinbami, Tiina Talts, Leena Bhaw, Richard Myers, Steven Platt, Kirstin Edwards, Jonathan Hubb, Joanna Ellis, Maria Zambon
EPI_ISL_414041 hCoV-19/England/200690245/2020 Europe / United Kingdom / England 2020-02-08 Respiratory Virus Unit, Microbiology Services Colindale, Public Health England Respiratory Virus Unit, Microbiology Services Colindale, Public Health England Monica Galiano, Shahjahan Miah, Angie Lackenby, Omolola Akinbami, Tiina Talts, Leena Bhaw, Richard Myers, Steven Platt, Kirstin Edwards, Jonathan Hubb, Joanna Ellis, Maria Zambon
EPI_ISL_414042 hCoV-19/England/200690300/2020 Europe / United Kingdom / England 2020-02-08 Respiratory Virus Unit, Microbiology Services Colindale, Public Health England Respiratory Virus Unit, Microbiology Services Colindale, Public Health England Monica Galiano, Shahjahan Miah, Angie Lackenby, Omolola Akinbami, Tiina Talts, Leena Bhaw, Richard Myers, Steven Platt, Kirstin Edwards, Jonathan Hubb, Joanna Ellis, Maria Zambon
EPI_ISL_414043 hCoV-19/England/200690306/2020 Europe / United Kingdom / England 2020-02-07 Respiratory Virus Unit, Microbiology Services Colindale, Public Health England Respiratory Virus Unit, Microbiology Services Colindale, Public Health England Monica Galiano, Shahjahan Miah, Angie Lackenby, Omolola Akinbami, Tiina Talts, Leena Bhaw, Richard Myers, Steven Platt, Kirstin Edwards, Jonathan Hubb, Joanna Ellis, Maria Zambon
EPI_ISL_414044 hCoV-19/England/200690756/2020 Europe / United Kingdom / England 2020-02-08 Respiratory Virus Unit, Microbiology Services Colindale, Public Health England Respiratory Virus Unit, Microbiology Services Colindale, Public Health England Monica Galiano, Shahjahan Miah, Angie Lackenby, Omolola Akinbami, Tiina Talts, Leena Bhaw, Richard Myers, Steven Platt, Kirstin Edwards, Jonathan Hubb, Joanna Ellis, Maria Zambon
EPI_ISL_414045 hCoV-19/Brazil/RJ-314/2020 South America / Brazil / Rio de Janeiro / Rio de Janeiro 2020-03-04 LACEN RJ - Laboratório Central de Saúde Pública Noel Nutels Instituto Oswaldo Cruz FIOCRUZ - Laboratory of Respiratory Viruses and Measles (LVRS) Paola Resende, Alisson Fabri, Joilson Xavier, Sunando Roy, Fernando Motta, Aline Mattos, Milene Miranda, Cristiana Garcia, Braulia Caetano, Maria Ogrzewalska, Jonathan Lopes, Luciana Appolinario, Maria Nóbrega, Marilda Siqueira
EPI_ISL_416600 hCoV-19/Japan/DP0438/2020 Asia / Japan / unknown 2020-02-16 Japanese Quarantine Stations Pathogen Genomics Center, National Institute of Infectious Diseases Tsuyoshi Sekizuka, Kentaro Itokawa, Rina Tanaka, Masanori Hashino, Tsutomu Kageyama, Shinji Saito, Ikuyo Takayama, Hideki Hasegawa, Takuri Takahashi, Hajime Kamiya, Takuya Yamagishi, Motoi Suzuki, Takaji Wakita, Makoto Kuroda
EPI_ISL_416601 hCoV-19/Japan/DP0457/2020 Asia / Japan / unknown 2020-02-16 Japanese Quarantine Stations Pathogen Genomics Center, National Institute of Infectious Diseases Tsuyoshi Sekizuka, Kentaro Itokawa, Rina Tanaka, Masanori Hashino, Tsutomu Kageyama, Shinji Saito, Ikuyo Takayama, Hideki Hasegawa, Takuri Takahashi, Hajime Kamiya, Takuya Yamagishi, Motoi Suzuki, Takaji Wakita, Makoto Kuroda
EPI_ISL_416602 hCoV-19/Japan/DP0462/2020 Asia / Japan / unknown 2020-02-16 Japanese Quarantine Stations Pathogen Genomics Center, National Institute of Infectious Diseases Tsuyoshi Sekizuka, Kentaro Itokawa, Rina Tanaka, Masanori Hashino, Tsutomu Kageyama, Shinji Saito, Ikuyo Takayama, Hideki Hasegawa, Takuri Takahashi, Hajime Kamiya, Takuya Yamagishi, Motoi Suzuki, Takaji Wakita, Makoto Kuroda
EPI_ISL_416603 hCoV-19/Japan/DP0464/2020 Asia / Japan / unknown 2020-02-16 Japanese Quarantine Stations Pathogen Genomics Center, National Institute of Infectious Diseases Tsuyoshi Sekizuka, Kentaro Itokawa, Rina Tanaka, Masanori Hashino, Tsutomu Kageyama, Shinji Saito, Ikuyo Takayama, Hideki Hasegawa, Takuri Takahashi, Hajime Kamiya, Takuya Yamagishi, Motoi Suzuki, Takaji Wakita, Makoto Kuroda
EPI_ISL_416604 hCoV-19/Japan/DP0476/2020 Asia / Japan / unknown 2020-02-16 Japanese Quarantine Stations Pathogen Genomics Center, National Institute of Infectious Diseases Tsuyoshi Sekizuka, Kentaro Itokawa, Rina Tanaka, Masanori Hashino, Tsutomu Kageyama, Shinji Saito, Ikuyo Takayama, Hideki Hasegawa, Takuri Takahashi, Hajime Kamiya, Takuya Yamagishi, Motoi Suzuki, Takaji Wakita, Makoto Kuroda
EPI_ISL_416605 hCoV-19/Japan/DP0481/2020 Asia / Japan / unknown 2020-02-16 Japanese Quarantine Stations Pathogen Genomics Center, National Institute of Infectious Diseases Tsuyoshi Sekizuka, Kentaro Itokawa, Rina Tanaka, Masanori Hashino, Tsutomu Kageyama, Shinji Saito, Ikuyo Takayama, Hideki Hasegawa, Takuri Takahashi, Hajime Kamiya, Takuya Yamagishi, Motoi Suzuki, Takaji Wakita, Makoto Kuroda
EPI_ISL_416606 hCoV-19/Japan/DP0482/2020 Asia / Japan / unknown 2020-02-16 Japanese Quarantine

Stations	Pathogen Genomics Center, National Institute of Infectious Diseases	Tsuyoshi Sekizuka, Kentaro Itokawa, Rina Tanaka, Masanori Hashino, Tsutomu Kageyama, Shinji Saito, Ikuyo Takayama, Hideki Hasegawa, Takuri Takahashi, Hajime Kamiya, Takuya Yamagishi, Motoi Suzuki, Takaji Wakita, Makoto Kuroda
EPI_ISL_416607	hCoV-19/Japan/DP0543/2020	Asia / Japan / unknown	2020-02-17	Japanese Quarantine Stations	Pathogen Genomics Center, National Institute of Infectious Diseases	Tsuyoshi Sekizuka, Kentaro Itokawa, Rina Tanaka, Masanori Hashino, Tsutomu Kageyama, Shinji Saito, Ikuyo Takayama, Hideki Hasegawa, Takuri Takahashi, Hajime Kamiya, Takuya Yamagishi, Motoi Suzuki, Takaji Wakita, Makoto Kuroda
EPI_ISL_416608	hCoV-19/Japan/DP0544/2020	Asia / Japan / unknown	2020-02-17	Japanese Quarantine Stations	Pathogen Genomics Center, National Institute of Infectious Diseases	Tsuyoshi Sekizuka, Kentaro Itokawa, Rina Tanaka, Masanori Hashino, Tsutomu Kageyama, Shinji Saito, Ikuyo Takayama, Hideki Hasegawa, Takuri Takahashi, Hajime Kamiya, Takuya Yamagishi, Motoi Suzuki, Takaji Wakita, Makoto Kuroda
EPI_ISL_416609	hCoV-19/Japan/DP0568/2020	Asia / Japan / unknown	2020-02-17	Japanese Quarantine Stations	Pathogen Genomics Center, National Institute of Infectious Diseases	Tsuyoshi Sekizuka, Kentaro Itokawa, Rina Tanaka, Masanori Hashino, Tsutomu Kageyama, Shinji Saito, Ikuyo Takayama, Hideki Hasegawa, Takuri Takahashi, Hajime Kamiya, Takuya Yamagishi, Motoi Suzuki, Takaji Wakita, Makoto Kuroda
EPI_ISL_416610	hCoV-19/Japan/DP0588/2020	Asia / Japan / unknown	2020-02-17	Japanese Quarantine Stations	Pathogen Genomics Center, National Institute of Infectious Diseases	Tsuyoshi Sekizuka, Kentaro Itokawa, Rina Tanaka, Masanori Hashino, Tsutomu Kageyama, Shinji Saito, Ikuyo Takayama, Hideki Hasegawa, Takuri Takahashi, Hajime Kamiya, Takuya Yamagishi, Motoi Suzuki, Takaji Wakita, Makoto Kuroda
EPI_ISL_416611	hCoV-19/Japan/DP0644/2020	Asia / Japan / unknown	2020-02-17	Japanese Quarantine Stations	Pathogen Genomics Center, National Institute of Infectious Diseases	Tsuyoshi Sekizuka, Kentaro Itokawa, Rina Tanaka, Masanori Hashino, Tsutomu Kageyama, Shinji Saito, Ikuyo Takayama, Hideki Hasegawa, Takuri Takahashi, Hajime Kamiya, Takuya Yamagishi, Motoi Suzuki, Takaji Wakita, Makoto Kuroda
EPI_ISL_416612	hCoV-19/Japan/DP0645/2020	Asia / Japan / unknown	2020-02-17	Japanese Quarantine Stations	Pathogen Genomics Center, National Institute of Infectious Diseases	Tsuyoshi Sekizuka, Kentaro Itokawa, Rina Tanaka, Masanori Hashino, Tsutomu Kageyama, Shinji Saito, Ikuyo Takayama, Hideki Hasegawa, Takuri Takahashi, Hajime Kamiya, Takuya Yamagishi, Motoi Suzuki, Takaji Wakita, Makoto Kuroda
EPI_ISL_416613	hCoV-19/Japan/DP0654/2020	Asia / Japan / unknown	2020-02-17	Japanese Quarantine Stations	Pathogen Genomics Center, National Institute of Infectious Diseases	Tsuyoshi Sekizuka, Kentaro Itokawa, Rina Tanaka, Masanori Hashino, Tsutomu Kageyama, Shinji Saito, Ikuyo Takayama, Hideki Hasegawa, Takuri Takahashi, Hajime Kamiya, Takuya Yamagishi, Motoi Suzuki, Takaji Wakita, Makoto Kuroda
EPI_ISL_416614	hCoV-19/Japan/DP0687/2020	Asia / Japan / unknown	2020-02-17	Japanese Quarantine Stations	Pathogen Genomics Center, National Institute of Infectious Diseases	Tsuyoshi Sekizuka, Kentaro Itokawa, Rina Tanaka, Masanori Hashino, Tsutomu Kageyama, Shinji Saito, Ikuyo Takayama, Hideki Hasegawa, Takuri Takahashi, Hajime Kamiya, Takuya Yamagishi, Motoi Suzuki, Takaji Wakita, Makoto Kuroda
EPI_ISL_416615	hCoV-19/Japan/DP0690/2020	Asia / Japan / unknown	2020-02-17	Japanese Quarantine Stations	Pathogen Genomics Center, National Institute of Infectious Diseases	Tsuyoshi Sekizuka, Kentaro Itokawa, Rina Tanaka, Masanori Hashino, Tsutomu Kageyama, Shinji Saito, Ikuyo Takayama, Hideki Hasegawa, Takuri Takahashi, Hajime Kamiya, Takuya Yamagishi, Motoi Suzuki, Takaji Wakita, Makoto Kuroda
EPI_ISL_416617	hCoV-19/Japan/DP0699/2020	Asia / Japan / unknown	2020-02-17	Japanese Quarantine Stations	Pathogen Genomics Center, National Institute of Infectious Diseases	Tsuyoshi Sekizuka, Kentaro Itokawa, Rina Tanaka, Masanori Hashino, Tsutomu Kageyama, Shinji Saito, Ikuyo Takayama, Hideki Hasegawa, Takuri Takahashi, Hajime Kamiya, Takuya Yamagishi, Motoi Suzuki, Takaji Wakita, Makoto Kuroda
EPI_ISL_416618	hCoV-19/Japan/DP0700/2020	Asia / Japan / unknown	2020-02-17	Japanese Quarantine Stations	Pathogen Genomics Center, National Institute of Infectious Diseases	Tsuyoshi Sekizuka, Kentaro Itokawa, Rina Tanaka, Masanori Hashino, Tsutomu Kageyama, Shinji Saito, Ikuyo Takayama, Hideki Hasegawa, Takuri Takahashi, Hajime Kamiya, Takuya Yamagishi, Motoi Suzuki, Takaji Wakita, Makoto Kuroda
EPI_ISL_416619	hCoV-19/Japan/DP0703/2020	Asia / Japan / unknown	2020-02-17	Japanese Quarantine Stations	Pathogen Genomics Center, National Institute of Infectious Diseases	Tsuyoshi Sekizuka, Kentaro Itokawa, Rina Tanaka, Masanori Hashino, Tsutomu Kageyama, Shinji Saito, Ikuyo Takayama, Hideki Hasegawa, Takuri Takahashi, Hajime Kamiya, Takuya Yamagishi, Motoi Suzuki, Takaji Wakita, Makoto Kuroda
EPI_ISL_416620	hCoV-19/Japan/DP0724/2020	Asia / Japan / unknown	2020-02-17	Japanese Quarantine Stations	Pathogen Genomics Center, National Institute of Infectious Diseases	Tsuyoshi Sekizuka, Kentaro Itokawa, Rina Tanaka, Masanori Hashino, Tsutomu Kageyama, Shinji Saito, Ikuyo Takayama, Hideki Hasegawa, Takuri Takahashi, Hajime Kamiya, Takuya Yamagishi, Motoi Suzuki, Takaji Wakita, Makoto Kuroda
EPI_ISL_416621	hCoV-19/Japan/DP0743/2020	Asia / Japan / unknown	2020-02-17	Japanese Quarantine Stations	Pathogen Genomics Center, National Institute of Infectious Diseases	Tsuyoshi Sekizuka, Kentaro Itokawa, Rina Tanaka, Masanori Hashino, Tsutomu Kageyama, Shinji Saito, Ikuyo Takayama, Hideki Hasegawa, Takuri Takahashi, Hajime Kamiya, Takuya Yamagishi, Motoi Suzuki, Takaji Wakita, Makoto Kuroda
EPI_ISL_416622	hCoV-19/Japan/DP0752/2020	Asia / Japan / unknown	2020-02-17	Japanese Quarantine Stations	Pathogen Genomics Center, National Institute of Infectious Diseases	Tsuyoshi Sekizuka, Kentaro Itokawa, Rina Tanaka, Masanori Hashino, Tsutomu Kageyama, Shinji Saito, Ikuyo Takayama, Hideki Hasegawa, Takuri Takahashi, Hajime Kamiya, Takuya Yamagishi, Motoi Suzuki, Takaji Wakita, Makoto Kuroda
EPI_ISL_416624	hCoV-19/Japan/DP0764/2020	Asia / Japan / unknown	2020-02-17	Japanese Quarantine Stations	Pathogen Genomics Center, National Institute of Infectious Diseases	Tsuyoshi Sekizuka, Kentaro Itokawa, Rina Tanaka, Masanori Hashino, Tsutomu Kageyama, Shinji Saito, Ikuyo Takayama, Hideki Hasegawa, Takuri Takahashi, Hajime Kamiya, Takuya Yamagishi, Motoi Suzuki, Takaji Wakita, Makoto Kuroda
EPI_ISL_416625	hCoV-19/Japan/DP0765/2020	Asia / Japan / unknown	2020-02-17	Japanese Quarantine Stations	Pathogen Genomics Center, National Institute of Infectious Diseases	Tsuyoshi Sekizuka, Kentaro Itokawa, Rina Tanaka, Masanori Hashino, Tsutomu Kageyama, Shinji Saito, Ikuyo Takayama, Hideki Hasegawa

Hasegawa, Takuri Takahashi, Hajime Kamiya, Takuya Yamagishi, Motoi Suzuki, Takaji Wakita, Makoto Kuroda
EPI_ISL_416626 hCoV-19/Japan/DP0779/2020 Asia / Japan / unknown 2020-02-17 Japanese Quarantine Stations Pathogen Genomics Center, National Institute of Infectious Diseases Tsuyoshi Sekizuka, Kentaro Itokawa, Rina Tanaka, Masanori Hashino, Tsutomu Kageyama, Shinji Saito, Ikuyo Takayama, Hideki Hasegawa, Takuri Takahashi, Hajime Kamiya, Takuya Yamagishi, Motoi Suzuki, Takaji Wakita, Makoto Kuroda
EPI_ISL_416627 hCoV-19/Japan/DP0785/2020 Asia / Japan / unknown 2020-02-17 Japanese Quarantine Stations Pathogen Genomics Center, National Institute of Infectious Diseases Tsuyoshi Sekizuka, Kentaro Itokawa, Rina Tanaka, Masanori Hashino, Tsutomu Kageyama, Shinji Saito, Ikuyo Takayama, Hideki Hasegawa, Takuri Takahashi, Hajime Kamiya, Takuya Yamagishi, Motoi Suzuki, Takaji Wakita, Makoto Kuroda
EPI_ISL_416628 hCoV-19/Japan/DP0786/2020 Asia / Japan / unknown 2020-02-17 Japanese Quarantine Stations Pathogen Genomics Center, National Institute of Infectious Diseases Tsuyoshi Sekizuka, Kentaro Itokawa, Rina Tanaka, Masanori Hashino, Tsutomu Kageyama, Shinji Saito, Ikuyo Takayama, Hideki Hasegawa, Takuri Takahashi, Hajime Kamiya, Takuya Yamagishi, Motoi Suzuki, Takaji Wakita, Makoto Kuroda
EPI_ISL_416629 hCoV-19/Japan/DP0802/2020 Asia / Japan / unknown 2020-02-17 Japanese Quarantine Stations Pathogen Genomics Center, National Institute of Infectious Diseases Tsuyoshi Sekizuka, Kentaro Itokawa, Rina Tanaka, Masanori Hashino, Tsutomu Kageyama, Shinji Saito, Ikuyo Takayama, Hideki Hasegawa, Takuri Takahashi, Hajime Kamiya, Takuya Yamagishi, Motoi Suzuki, Takaji Wakita, Makoto Kuroda
EPI_ISL_416630 hCoV-19/Japan/DP0803/2020 Asia / Japan / unknown 2020-02-17 Japanese Quarantine Stations Pathogen Genomics Center, National Institute of Infectious Diseases Tsuyoshi Sekizuka, Kentaro Itokawa, Rina Tanaka, Masanori Hashino, Tsutomu Kageyama, Shinji Saito, Ikuyo Takayama, Hideki Hasegawa, Takuri Takahashi, Hajime Kamiya, Takuya Yamagishi, Motoi Suzuki, Takaji Wakita, Makoto Kuroda
EPI_ISL_416631 hCoV-19/Japan/DP0804/2020 Asia / Japan / unknown 2020-02-17 Japanese Quarantine Stations Pathogen Genomics Center, National Institute of Infectious Diseases Tsuyoshi Sekizuka, Kentaro Itokawa, Rina Tanaka, Masanori Hashino, Tsutomu Kageyama, Shinji Saito, Ikuyo Takayama, Hideki Hasegawa, Takuri Takahashi, Hajime Kamiya, Takuya Yamagishi, Motoi Suzuki, Takaji Wakita, Makoto Kuroda
EPI_ISL_416632 hCoV-19/Japan/DP0827/2020 Asia / Japan / unknown 2020-02-17 Japanese Quarantine Stations Pathogen Genomics Center, National Institute of Infectious Diseases Tsuyoshi Sekizuka, Kentaro Itokawa, Rina Tanaka, Masanori Hashino, Tsutomu Kageyama, Shinji Saito, Ikuyo Takayama, Hideki Hasegawa, Takuri Takahashi, Hajime Kamiya, Takuya Yamagishi, Motoi Suzuki, Takaji Wakita, Makoto Kuroda
EPI_ISL_416633 hCoV-19/Japan/DP0880/2020 Asia / Japan / unknown 2020-02-17 Japanese Quarantine Stations Pathogen Genomics Center, National Institute of Infectious Diseases Tsuyoshi Sekizuka, Kentaro Itokawa, Rina Tanaka, Masanori Hashino, Tsutomu Kageyama, Shinji Saito, Ikuyo Takayama, Hideki Hasegawa, Takuri Takahashi, Hajime Kamiya, Takuya Yamagishi, Motoi Suzuki, Takaji Wakita, Makoto Kuroda
EPI_ISL_416634 hCoV-19/Japan/DP0890/2020 Asia / Japan / unknown 2020-02-17 Japanese Quarantine Stations Pathogen Genomics Center, National Institute of Infectious Diseases Tsuyoshi Sekizuka, Kentaro Itokawa, Rina Tanaka, Masanori Hashino, Tsutomu Kageyama, Shinji Saito, Ikuyo Takayama, Hideki Hasegawa, Takuri Takahashi, Hajime Kamiya, Takuya Yamagishi, Motoi Suzuki, Takaji Wakita, Makoto Kuroda
EPI_ISL_416635 hCoV-19/USA/WA-UW97/2020 North America / USA / Washington 2020-03-12 UW Virology Lab UW Virology Lab Pavitra Roychoudhury, Hong Xie, Keith Jerome, Alexander Greninger
EPI_ISL_416636 hCoV-19/USA/WA-UW98/2020 North America / USA / Washington 2020-03-12 UW Virology Lab UW Virology Lab Pavitra Roychoudhury, Hong Xie, Keith Jerome, Alexander Greninger
EPI_ISL_416637 hCoV-19/USA/WA-UW99/2020 North America / USA / Washington 2020-03-12 UW Virology Lab UW Virology Lab Pavitra Roychoudhury, Hong Xie, Keith Jerome, Alexander Greninger
EPI_ISL_416638 hCoV-19/USA/WA-UW100/2020 North America / USA / Washington 2020-03-12 UW Virology Lab UW Virology Lab Pavitra Roychoudhury, Hong Xie, Keith Jerome, Alexander Greninger
EPI_ISL_416639 hCoV-19/USA/WA-UW101/2020 North America / USA / Washington 2020-03-10 UW Virology Lab UW Virology Lab Pavitra Roychoudhury, Hong Xie, Keith Jerome, Alexander Greninger
EPI_ISL_416641 hCoV-19/USA/WA-UW103/2020 North America / USA / Washington 2020-03-11 UW Virology Lab UW Virology Lab Pavitra Roychoudhury, Hong Xie, Keith Jerome, Alexander Greninger
EPI_ISL_416642 hCoV-19/USA/WA-UW104/2020 North America / USA / Washington 2020-03-11 UW Virology Lab UW Virology Lab Pavitra Roychoudhury, Hong Xie, Keith Jerome, Alexander Greninger
EPI_ISL_416643 hCoV-19/USA/WA-UW105/2020 North America / USA / Washington 2020-03-11 UW Virology Lab UW Virology Lab Pavitra Roychoudhury, Hong Xie, Keith Jerome, Alexander Greninger
EPI_ISL_416644 hCoV-19/USA/WA-UW106/2020 North America / USA / Washington 2020-03-11 UW Virology Lab UW Virology Lab Pavitra Roychoudhury, Hong Xie, Keith Jerome, Alexander Greninger
EPI_ISL_416646 hCoV-19/USA/WA-UW108/2020 North America / USA / Washington 2020-03-11 UW Virology Lab UW Virology Lab Pavitra Roychoudhury, Hong Xie, Keith Jerome, Alexander Greninger
EPI_ISL_416647 hCoV-19/USA/WA-UW109/2020 North America / USA / Washington 2020-03-11 UW Virology Lab UW Virology Lab Pavitra Roychoudhury, Hong Xie, Keith Jerome, Alexander Greninger
EPI_ISL_416648 hCoV-19/USA/WA-UW110/2020 North America / USA / Washington 2020-03-11 UW Virology Lab UW Virology Lab Pavitra Roychoudhury, Hong Xie, Keith Jerome, Alexander Greninger
EPI_ISL_416649 hCoV-19/USA/WA-UW111/2020 North America / USA / Washington 2020-03-11 UW Virology Lab UW Virology Lab Pavitra Roychoudhury, Hong Xie, Keith Jerome, Alexander Greninger
EPI_ISL_416650 hCoV-19/USA/WA-UW112/2020 North America / USA 2020-03-10 UW Virology Lab UW Virology Lab Pavitra Roychoudhury, Hong Xie, Keith Jerome, Alexander Greninger
EPI_ISL_416651 hCoV-19/USA/WA-UW113/2020 North America / USA 2020-03-11 UW Virology Lab UW Virology Lab Pavitra Roychoudhury, Hong Xie, Keith Jerome, Alexander Greninger
EPI_ISL_416652 hCoV-19/USA/WA-UW114/2020 North America / USA / Washington 2020-03-11 UW Virology Lab UW Virology Lab Pavitra Roychoudhury, Hong Xie, Keith Jerome, Alexander Greninger
EPI_ISL_416653 hCoV-19/USA/WA-UW115/2020 North America / USA / Washington 2020-03-11 UW

Virology Lab	UW Virology Lab Pavitra Roychoudhury, Hong Xie, Keith Jerome, Alexander Greninger
EPI_ISL_416654	hCoV-19/USA/WA-UW116/2020	North America / USA / Washington	2020-03-11	UW Virology Lab	UW Virology Lab Pavitra Roychoudhury, Hong Xie, Keith Jerome, Alexander Greninger
EPI_ISL_416655	hCoV-19/USA/WA-UW117/2020	North America / USA / Washington	2020-03-11	UW Virology Lab	UW Virology Lab Pavitra Roychoudhury, Hong Xie, Keith Jerome, Alexander Greninger
EPI_ISL_416656	hCoV-19/USA/WA-UW118/2020	North America / USA / Washington	2020-03-11	UW Virology Lab	UW Virology Lab Pavitra Roychoudhury, Hong Xie, Keith Jerome, Alexander Greninger
EPI_ISL_416657	hCoV-19/USA/WA-UW119/2020	North America / USA / Washington	2020-03-11	UW Virology Lab	UW Virology Lab Pavitra Roychoudhury, Hong Xie, Keith Jerome, Alexander Greninger
EPI_ISL_416658	hCoV-19/USA/WA-UW120/2020	North America / USA / Washington	2020-03-11	UW Virology Lab	UW Virology Lab Pavitra Roychoudhury, Hong Xie, Keith Jerome, Alexander Greninger
EPI_ISL_416659	hCoV-19/USA/WA-UW121/2020	North America / USA / Washington	2020-03-11	UW Virology Lab	UW Virology Lab Pavitra Roychoudhury, Hong Xie, Keith Jerome, Alexander Greninger
EPI_ISL_416660	hCoV-19/USA/WA-UW122/2020	North America / USA	2020-03-10	UW Virology Lab UW Virology Lab	Pavitra Roychoudhury, Hong Xie, Keith Jerome, Alexander Greninger
EPI_ISL_416661	hCoV-19/USA/WA-UW123/2020	North America / USA / Washington	2020-03-12	UW Virology Lab	UW Virology Lab Pavitra Roychoudhury, Hong Xie, Keith Jerome, Alexander Greninger
EPI_ISL_416662	hCoV-19/USA/WA-UW124/2020	North America / USA / Washington	2020-03-12	UW Virology Lab	UW Virology Lab Pavitra Roychoudhury, Hong Xie, Keith Jerome, Alexander Greninger
EPI_ISL_416663	hCoV-19/USA/WA-UW125/2020	North America / USA / Washington	2020-03-12	UW Virology Lab	UW Virology Lab Pavitra Roychoudhury, Hong Xie, Keith Jerome, Alexander Greninger
EPI_ISL_416664	hCoV-19/USA/WA-UW126/2020	North America / USA / Washington	2020-03-12	UW Virology Lab	UW Virology Lab Pavitra Roychoudhury, Hong Xie, Keith Jerome, Alexander Greninger
EPI_ISL_416665	hCoV-19/USA/WA-UW127/2020	North America / USA / Washington	2020-03-12	UW Virology Lab	UW Virology Lab Pavitra Roychoudhury, Hong Xie, Keith Jerome, Alexander Greninger
EPI_ISL_416666	hCoV-19/USA/WA-UW128/2020	North America / USA / Washington	2020-03-12	UW Virology Lab	UW Virology Lab Pavitra Roychoudhury, Hong Xie, Keith Jerome, Alexander Greninger
EPI_ISL_416667	hCoV-19/USA/WA-UW129/2020	North America / USA / Washington	2020-03-12	UW Virology Lab	UW Virology Lab Pavitra Roychoudhury, Hong Xie, Keith Jerome, Alexander Greninger
EPI_ISL_416668	hCoV-19/USA/WA-UW130/2020	North America / USA / Washington	2020-03-11	UW Virology Lab	UW Virology Lab Pavitra Roychoudhury, Hong Xie, Keith Jerome, Alexander Greninger
EPI_ISL_416669	hCoV-19/USA/WA-UW131/2020	North America / USA / Washington	2020-03-11	UW Virology Lab	UW Virology Lab Pavitra Roychoudhury, Hong Xie, Keith Jerome, Alexander Greninger
EPI_ISL_416671	hCoV-19/USA/WA-UW133/2020	North America / USA / Washington	2020-03-12	UW Virology Lab	UW Virology Lab Pavitra Roychoudhury, Hong Xie, Keith Jerome, Alexander Greninger
EPI_ISL_416672	hCoV-19/USA/WA-UW134/2020	North America / USA	2020-03-10	UW Virology Lab UW Virology Lab	Pavitra Roychoudhury, Hong Xie, Keith Jerome, Alexander Greninger
EPI_ISL_416673	hCoV-19/USA/OR-UW135/2020	North America / USA / Oregon	2020-03-11	UW Virology Lab UW Virology Lab Pavitra Roychoudhury, Hong Xie, Keith Jerome, Alexander Greninger
EPI_ISL_416674	hCoV-19/USA/WA-UW136/2020	North America / USA	2020-03-11	UW Virology Lab UW Virology Lab	Pavitra Roychoudhury, Hong Xie, Keith Jerome, Alexander Greninger
EPI_ISL_416675	hCoV-19/USA/WA-UW137/2020	North America / USA	2020-03-11	UW Virology Lab UW Virology Lab	Pavitra Roychoudhury, Hong Xie, Keith Jerome, Alexander Greninger
EPI_ISL_416676	hCoV-19/USA/WA-UW138/2020	North America / USA	2020-03-11	UW Virology Lab UW Virology Lab	Pavitra Roychoudhury, Hong Xie, Keith Jerome, Alexander Greninger
EPI_ISL_416677	hCoV-19/USA/WA-UW139/2020	North America / USA / Washington	2020-03-12	UW Virology Lab	UW Virology Lab Pavitra Roychoudhury, Hong Xie, Keith Jerome, Alexander Greninger
EPI_ISL_416678	hCoV-19/USA/WA-UW140/2020	North America / USA / Washington	2020-03-11	UW Virology Lab	UW Virology Lab Pavitra Roychoudhury, Hong Xie, Keith Jerome, Alexander Greninger
EPI_ISL_416679	hCoV-19/USA/WA-UW141/2020	North America / USA / Washington	2020-03-11	UW Virology Lab	UW Virology Lab Pavitra Roychoudhury, Hong Xie, Keith Jerome, Alexander Greninger
EPI_ISL_416680	hCoV-19/USA/WA-UW142/2020	North America / USA / Washington	2020-03-11	UW Virology Lab	UW Virology Lab Pavitra Roychoudhury, Hong Xie, Keith Jerome, Alexander Greninger
EPI_ISL_416681	hCoV-19/USA/WA-UW143/2020	North America / USA / Washington	2020-03-11	UW Virology Lab	UW Virology Lab Pavitra Roychoudhury, Hong Xie, Keith Jerome, Alexander Greninger
EPI_ISL_416682	hCoV-19/USA/WA-UW144/2020	North America / USA	2020-03-09	UW Virology Lab UW Virology Lab	Pavitra Roychoudhury, Hong Xie, Keith Jerome, Alexander Greninger
EPI_ISL_416683	hCoV-19/USA/WA-UW145/2020	North America / USA / Washington	2020-03-15	UW Virology Lab	UW Virology Lab Pavitra Roychoudhury, Hong Xie, Keith Jerome, Alexander Greninger
EPI_ISL_416684	hCoV-19/USA/WA-UW146/2020	North America / USA / Washington	2020-03-14	UW Virology Lab	UW Virology Lab Pavitra Roychoudhury, Hong Xie, Keith Jerome, Alexander Greninger
EPI_ISL_416685	hCoV-19/USA/WA-UW147/2020	North America / USA / Washington	2020-03-15	UW Virology Lab	UW Virology Lab Pavitra Roychoudhury, Hong Xie, Keith Jerome, Alexander Greninger
EPI_ISL_416686	hCoV-19/USA/WA-UW148/2020	North America / USA / Washington	2020-03-14	UW Virology Lab	UW Virology Lab Pavitra Roychoudhury, Hong Xie, Keith Jerome, Alexander Greninger
EPI_ISL_416687	hCoV-19/USA/WA-UW149/2020	North America / USA / Washington	2020-03-14	UW Virology Lab	UW Virology Lab Pavitra Roychoudhury, Hong Xie, Keith Jerome, Alexander Greninger
EPI_ISL_416688	hCoV-19/USA/WA-UW150/2020	North America / USA / Washington	2020-03-14	UW Virology Lab	UW Virology Lab Pavitra Roychoudhury, Hong Xie, Keith Jerome, Alexander Greninger
EPI_ISL_416689	hCoV-19/USA/WA-UW151/2020	North America / USA / Washington	2020-03-14	UW

Virology Lab	UW Virology Lab Pavitra Roychoudhury, Hong Xie, Keith Jerome, Alexander Greninger
EPI_ISL_416690	hCoV-19/USA/WA-UW152/2020	North America / USA / Washington	2020-03-13	UW Virology Lab	UW Virology Lab Pavitra Roychoudhury, Hong Xie, Keith Jerome, Alexander Greninger
EPI_ISL_416691	hCoV-19/USA/WA-UW153/2020	North America / USA / Washington	2020-03-13	UW Virology Lab	UW Virology Lab Pavitra Roychoudhury, Hong Xie, Keith Jerome, Alexander Greninger
EPI_ISL_416692	hCoV-19/USA/WA-UW154/2020	North America / USA / Washington	2020-03-14	UW Virology Lab	UW Virology Lab Pavitra Roychoudhury, Hong Xie, Keith Jerome, Alexander Greninger
EPI_ISL_416693	hCoV-19/USA/WA-UW155/2020	North America / USA / Washington	2020-03-12	UW Virology Lab	UW Virology Lab Pavitra Roychoudhury, Hong Xie, Keith Jerome, Alexander Greninger
EPI_ISL_416694	hCoV-19/USA/WA-UW156/2020	North America / USA / Washington	2020-03-13	UW Virology Lab	UW Virology Lab Pavitra Roychoudhury, Hong Xie, Keith Jerome, Alexander Greninger
EPI_ISL_416696	hCoV-19/USA/CT-UW158/2020	North America / USA / Connecticut	2020-03-13	UW Virology Lab	UW Virology Lab Pavitra Roychoudhury, Hong Xie, Keith Jerome, Alexander Greninger
EPI_ISL_416697	hCoV-19/USA/CT-UW159/2020	North America / USA / Connecticut	2020-03-13	UW Virology Lab	UW Virology Lab Pavitra Roychoudhury, Hong Xie, Keith Jerome, Alexander Greninger
EPI_ISL_416698	hCoV-19/USA/WA-UW160/2020	North America / USA / Washington	2020-03-13	UW Virology Lab	UW Virology Lab Pavitra Roychoudhury, Hong Xie, Keith Jerome, Alexander Greninger
EPI_ISL_416699	hCoV-19/USA/WA-UW161/2020	North America / USA / Washington	2020-03-13	UW Virology Lab	UW Virology Lab Pavitra Roychoudhury, Hong Xie, Keith Jerome, Alexander Greninger
EPI_ISL_417400	hCoV-19/Australia/NSW44/2020	Oceania / Australia / New South Wales / Sydney	2020-03-10	Centre for Infectious Diseases and Microbiology Public Health	NSW Health Pathology - Institute of Clinical Pathology and Medical Research; Westmead Hospital; University of Sydney Rockett R, Eden J-S, Lam C, Gray K, Timms V, Gall M, Arnott A, Sadsad R, Carter I, Rahman H, Holmes EC, O’Sullivan MV, Sintchenko V, Chen SC, Maddocks S, Kok J and Dwyer DE for the 2019-nCoV Study Group
EPI_ISL_417401	hCoV-19/Australia/NSW45/2020	Oceania / Australia / New South Wales / Sydney	2020-03-13	Centre for Infectious Diseases and Microbiology Public Health	NSW Health Pathology - Institute of Clinical Pathology and Medical Research; Westmead Hospital; University of Sydney Eden J-S, Lam C, Gray K, Timms V, Gall M, Arnott A, Sadsad R, Carter I, Rahman H, Holmes EC, O’Sullivan MV, Sintchenko V, Chen SC, Maddocks S, Kok J, Dwyer DE and Rockett R for the 2019-nCoV Study Group
EPI_ISL_417402	hCoV-19/Australia/NSW47/2020	Oceania / Australia / New South Wales / Sydney	2020-03-13	Centre for Infectious Diseases and Microbiology Public Health	NSW Health Pathology - Institute of Clinical Pathology and Medical Research; Westmead Hospital; University of Sydney Lam C, Gray K, Timms V, Gall M, Arnott A, Sadsad R, Carter I, Rahman H, Holmes EC, O’Sullivan MV, Sintchenko V, Chen SC, Maddocks S, Kok J, Dwyer DE, Rockett R and Eden J-S for the 2019-nCoV Study Group
EPI_ISL_417403	hCoV-19/Australia/NSW48/2020	Oceania / Australia / New South Wales / Sydney	2020-03-14	Centre for Infectious Diseases and Microbiology Public Health	NSW Health Pathology - Institute of Clinical Pathology and Medical Research; Westmead Hospital; University of Sydney Gray K, Timms V, Gall M, Arnott A, Sadsad R, Carter I, Rahman H, Holmes EC, O’Sullivan MV, Sintchenko V, Chen SC, Maddocks S, Kok J, Dwyer DE, Rockett R, Eden J-S and Lam C for the 2019-nCoV Study Group
EPI_ISL_417404	hCoV-19/Australia/NSW50/2020	Oceania / Australia / New South Wales / Sydney	2020-03-12	Centre for Infectious Diseases and Microbiology Public Health	NSW Health Pathology - Institute of Clinical Pathology and Medical Research; Westmead Hospital; University of Sydney Timms V, Gall M, Arnott A, Sadsad R, Carter I, Rahman H, Holmes EC, O’Sullivan MV, Sintchenko V, Chen SC, Maddocks S, Kok J, Dwyer DE, Rockett R, Eden J-S, Lam C and Gray K for the 2019-nCoV Study Group
EPI_ISL_417405	hCoV-19/Australia/NSW52/2020	Oceania / Australia / New South Wales / Sydney	2020-03-11	Centre for Infectious Diseases and Microbiology Public Health	NSW Health Pathology - Institute of Clinical Pathology and Medical Research; Westmead Hospital; University of Sydney Gall M, Arnott A, Sadsad R, Carter I, Rahman H, Holmes EC, O’Sullivan MV, Sintchenko V, Chen SC, Maddocks S, Kok J, Dwyer DE, Rockett R, Eden J-S, Lam C, Gray K and Timms V for the 2019-nCoV Study Group
EPI_ISL_417406	hCoV-19/Australia/NSW53/2020	Oceania / Australia / New South Wales / Sydney	2020-03-11	Centre for Infectious Diseases and Microbiology Public Health	NSW Health Pathology - Institute of Clinical Pathology and Medical Research; Westmead Hospital; University of Sydney Arnott A, Sadsad R, Carter I, Rahman H, Holmes EC, O’Sullivan MV, Sintchenko V, Chen SC, Maddocks S, Kok J, Dwyer DE, Rockett R, Eden J-S, Lam C, Gray K, Timms V and Gall M for the 2019-nCoV Study Group
EPI_ISL_417407	hCoV-19/Australia/NSW54/2020	Oceania / Australia / New South Wales / Sydney	2020-03-14	Centre for Infectious Diseases and Microbiology Public Health	NSW Health Pathology - Institute of Clinical Pathology and Medical Research; Westmead Hospital; University of Sydney Sadsad R, Carter I, Rahman H, Holmes EC, O’Sullivan MV, Sintchenko V, Chen SC, Maddocks S, Kok J, Dwyer DE, Rockett R, Eden J-S, Lam C, Gray K, Timms V, Gall M and Arnott A for the 2019-nCoV Study Group
EPI_ISL_417917	hCoV-19/Malaysia/189332/2020	Asia / Malaysia / Kuala Lumpur	2020-03-20	Department of Medical Microbiology, University Malaya Medical Centre	Department of Medical Microbiology	Yoong Min CHONG, Sasheela PONNAMPALAVANAR, Sharifah Faridah SYED OMAR, Adeeba KAMARULZAMAN,Vijayan MUNUSAMY, Chee Kuan WONG, Cindy Shuan Ju TEH, I-Ching SAM, Yoke Fun Chan, University Malaya Medical Centre COVID Team
EPI_ISL_417918	hCoV-19/Malaysia/188407/2020	Asia / Malaysia / Kuala Lumpur	2020-03-18	Department of Medical Microbiology, University Malaya Medical Centre	Department of Medical Microbiology, Faculty of Medicine, University of Malaya	Yoong Min CHONG, Sasheela PONNAMPALAVANAR, Sharifah Faridah SYED OMAR, Adeeba KAMARULZAMAN,Vijayan MUNUSAMY, Chee Kuan WONG, Cindy Shuan Ju TEH, I-Ching SAM, Yoke Fun Chan, University Malaya Medical Centre COVID Team
EPI_ISL_417921	hCoV-19/Italy/INMI3/2020	Europe / Italy / Rome	2020-03-01	INMI Lazzaro Spallanzani IRCCS	Laboratory of Virology, INMI Lazzaro Spallanzani IRCCS	Martina Rueca, Barbara

Bartolini, Francesco Messina, Cesare E. M. Gruber, Emanuela Giombini, Maria R. Capobianchi, Fabrizio Carletti, Francesca Colavita, Concetta Castilletti, Eleonora Lalle, Daniele Lapa, Giuseppe Ippolito.
EPI_ISL_417922 hCoV-19/Italy/INMI4/2020 Europe / Italy / Rome 2020-02-28 INMI Lazzaro Spallanzani IRCCS Laboratory of Virology, INMI Lazzaro Spallanzani IRCCS Cesare E. M. Gruber, Martina Rueca, Barbara Bartolini, Francesco Messina, Emanuela Giombini, Maria R. Capobianchi, Fabrizio Carletti, Francesca Colavita, Concetta Castilletti, Eleonora Lalle, Daniele Lapa, Giuseppe Ippolito.
EPI_ISL_417923 hCoV-19/Italy/INMI5/2020 Europe / Italy / Rome 2020-03-04 INMI Lazzaro Spallanzani IRCCS Laboratory of Virology, INMI Lazzaro Spallanzani IRCCS Francesco Messina, Barbara Bartolini, Martina Rueca, Cesare E. M. Gruber, Emanuela Giombini, Maria R. Capobianchi, Fabrizio Carletti, Francesca Colavita, Concetta Castilletti, Eleonora Lalle, Daniele Lapa, Giuseppe Ippolito.
EPI_ISL_417924 hCoV-19/Colombia/INS-79256/2020 South America / Colombia / Antioquia 2020-03-11 Secretaría de Salud Medellín Instituto Nacional de Salud, Universidad Cooperativa de Colombia, Instituto Alexander von Humboldt, Imperial College-London, London School of Hygiene & Tropical Medicine Marcela Mercado-Reyes, Katherine Laiton-Donato, Diego A. Álvarez-Díaz, Carlos Franco-Muñoz, Jose A. Usme-Ciro, Gloria Puerto, Nicolás D. Franco-Sierra, Mailyn A. Gonzalez, Zulma M. Cucunubá, Christian Julian Villabona-Arenas, Liz Villabona-Arenas, Sussy Echeverria-Londoño, Astrid C. Flórez, Sergio Gomez Rangel, Luz Dary Rodriguez, Juliana Barbosa, Erika Ospitia, Diana Marcela Walteros-Acero, Martha Lucia Ospina Martinez
EPI_ISL_417931 hCoV-19/USA/CA-CZB05/2020 North America / USA / California / San Francisco 2020-03-18 UCSF Clinical Microbiology Laboratory Chan-Zuckerberg Biohub Shaun Arevalo, Josh Batson, Olga Botvinnik, Gloria Castaneda, Angela Detweiler, David Dynerman, Samantha Hao, Jack Kamm, Amy Kistler, G. Renuka Kumar, Chaz Langelier, Lucy Li, Steve Miller, Lusajo Mwakibete, Norma Neff, Angela Pisco, Maira Phelps, Michelle Tan, Chunyu Zhao
EPI_ISL_417932 hCoV-19/USA/CA-CZB06/2020 North America / USA / California / San Francisco 2020-03-18 UCSF Clinical Microbiology Laboratory Chan-Zuckerberg Biohub Shaun Arevalo, Josh Batson, Olga Botvinnik, Gloria Castaneda, Angela Detweiler, David Dynerman, Samantha Hao, Jack Kamm, Amy Kistler, G. Renuka Kumar, Chaz Langelier, Lucy Li, Steve Miller, Lusajo Mwakibete, Norma Neff, Angela Pisco, Maira Phelps, Michelle Tan, Chunyu Zhao
EPI_ISL_417933 hCoV-19/USA/CA-CZB07/2020 North America / USA / California / San Francisco 2020-03-18 UCSF Clinical Microbiology Laboratory Chan-Zuckerberg Biohub Shaun Arevalo, Josh Batson, Olga Botvinnik, Gloria Castaneda, Angela Detweiler, David Dynerman, Samantha Hao, Jack Kamm, Amy Kistler, G. Renuka Kumar, Chaz Langelier, Lucy Li, Steve Miller, Lusajo Mwakibete, Norma Neff, Angela Pisco, Maira Phelps, Michelle Tan, Chunyu Zhao
EPI_ISL_417935 hCoV-19/USA/CA-CZB011a/2020 North America / USA / California / San Francisco 2020-03-18 UCSF Clinical Microbiology Laboratory Chan-Zuckerberg Biohub Shaun Arevalo, Josh Batson, Olga Botvinnik, Gloria Castaneda, Angela Detweiler, David Dynerman, Samantha Hao, Jack Kamm, Amy Kistler, G. Renuka Kumar, Chaz Langelier, Lucy Li, Steve Miller, Lusajo Mwakibete, Norma Neff, Angela Pisco, Maira Phelps, Michelle Tan, Chunyu Zhao
EPI_ISL_417937 hCoV-19/USA/CA-CZB013/2020 North America / USA / California / San Francisco 2020-03-18 UCSF Clinical Microbiology Laboratory Chan-Zuckerberg Biohub Shaun Arevalo, Josh Batson, Olga Botvinnik, Gloria Castaneda, Angela Detweiler, David Dynerman, Samantha Hao, Jack Kamm, Amy Kistler, G. Renuka Kumar, Chaz Langelier, Lucy Li, Steve Miller, Lusajo Mwakibete, Norma Neff, Angela Pisco, Maira Phelps, Michelle Tan, Chunyu Zhao
EPI_ISL_417938 hCoV-19/USA/CA-CZB014/2020 North America / USA / California / San Francisco 2020-03-18 UCSF Clinical Microbiology Laboratory Chan-Zuckerberg Biohub Shaun Arevalo, Josh Batson, Olga Botvinnik, Gloria Castaneda, Angela Detweiler, David Dynerman, Samantha Hao, Jack Kamm, Amy Kistler, G. Renuka Kumar, Chaz Langelier, Lucy Li, Steve Miller, Lusajo Mwakibete, Norma Neff, Angela Pisco, Maira Phelps, Michelle Tan, Chunyu Zhao
EPI_ISL_417939 hCoV-19/USA/CA-CZB015a/2020 North America / USA / California / San Francisco 2020-03-18 UCSF Clinical Microbiology Laboratory Chan-Zuckerberg Biohub Shaun Arevalo, Josh Batson, Olga Botvinnik, Gloria Castaneda, Angela Detweiler, David Dynerman, Samantha Hao, Jack Kamm, Amy Kistler, G. Renuka Kumar, Chaz Langelier, Lucy Li, Steve Miller, Lusajo Mwakibete, Norma Neff, Angela Pisco, Maira Phelps, Michelle Tan, Chunyu Zhao
EPI_ISL_417941 hCoV-19/DRC/73/2020 Africa / Democratic Republic of the Congo / Kinshasa 2020-03-18 Viral Respiratory Lab, National Institute for Biomedical Research (INRB) Pathogen Sequencing Lab, National Institute for Biomedical Research (INRB) Placide Mbala-Kingebeni, Edith Nkwembe, Eddy Kinganda-Lusamaki, Amuri Aziza, Catherine Pratt, Matthias Pauthner, Josh Quick, Allison Black, James Hadfield, Trevor Bedford, Ian Goodfellow, Nick Loman, Kristian Andersen, Michael Wiley, Steve Ahuka-Mundeke, Jean-Jacques Muyembe Tamfum
EPI_ISL_417942 hCoV-19/DRC/80/2020 Africa / Democratic Republic of the Congo / Kinshasa 2020-03-18 Viral Respiratory Lab, National Institute for Biomedical Research (INRB) Pathogen Sequencing Lab, National Institute for Biomedical Research (INRB) Placide Mbala-Kingebeni, Edith Nkwembe, Eddy Kinganda-Lusamaki, Amuri Aziza, Catherine Pratt, Matthias Pauthner, Josh Quick, Allison Black, James Hadfield, Trevor Bedford, Ian Goodfellow, Nick Loman, Kristian Andersen, Michael Wiley, Steve Ahuka-Mundeke, Jean-Jacques Muyembe Tamfum
EPI_ISL_417944 hCoV-19/DRC/81/2020 Africa / Democratic Republic of the Congo / Kinshasa 2020-03-18 Viral Respiratory Lab, National Institute for Biomedical Research (INRB) Pathogen Sequencing Lab, National Institute for Biomedical Research (INRB) Placide Mbala-Kingebeni, Edith Nkwembe, Eddy Kinganda-Lusamaki, Amuri Aziza, Catherine Pratt, Matthias Pauthner, Josh Quick, Allison Black, James Hadfield, Trevor Bedford, Ian Goodfellow, Nick Loman, Kristian Andersen, Michael Wiley, Steve Ahuka-Mundeke, Jean-Jacques Muyembe Tamfum

EPI_ISL_417946 hCoV-19/DRC/82/2020 Africa / Democratic Republic of the Congo / Kinshasa 2020-03-18 Viral Respiratory Lab, National Institute for Biomedical Research (INRB) Pathogen Sequencing Lab, National Institute for Biomedical Research (INRB) Placide Mbala-Kingebeni, Edith Nkwembe, Eddy Kinganda-Lusamaki, Amuri Aziza, Catherine Pratt, Matthias Pauthner, Josh Quick, Allison Black, James Hadfield, Trevor Bedford, Ian Goodfellow, Nick Loman, Kristian Andersen, Michael Wiley, Steve Ahuka-Mundeke, Jean-Jacques Muyembe Tamfum
EPI_ISL_417947 hCoV-19/DRC/94/2020 Africa / Democratic Republic of the Congo / Kinshasa 2020-03-19 Viral Respiratory Lab, National Institute for Biomedical Research (INRB) Pathogen Sequencing Lab, National Institute for Biomedical Research (INRB) Placide Mbala-Kingebeni, Edith Nkwembe, Eddy Kinganda-Lusamaki, Amuri Aziza, Catherine Pratt, Matthias Pauthner, Josh Quick, Allison Black, James Hadfield, Trevor Bedford, Ian Goodfellow, Nick Loman, Kristian Andersen, Michael Wiley, Steve Ahuka-Mundeke, Jean-Jacques Muyembe Tamfum
EPI_ISL_417948 hCoV-19/DRC/108/2020 Africa / Democratic Republic of the Congo / Kinshasa 2020-03-19 Viral Respiratory Lab, National Institute for Biomedical Research (INRB) Pathogen Sequencing Lab, National Institute for Biomedical Research (INRB) Placide Mbala-Kingebeni, Edith Nkwembe, Eddy Kinganda-Lusamaki, Amuri Aziza, Catherine Pratt, Matthias Pauthner, Josh Quick, Allison Black, James Hadfield, Trevor Bedford, Ian Goodfellow, Nick Loman, Kristian Andersen, Michael Wiley, Steve Ahuka-Mundeke, Jean-Jacques Muyembe Tamfum
EPI_ISL_417950 hCoV-19/DRC/158/2020 Africa / Democratic Republic of the Congo / Kinshasa 2020-03-20 Viral Respiratory Lab, National Institute for Biomedical Research (INRB) Pathogen Sequencing Lab, National Institute for Biomedical Research (INRB) Placide Mbala-Kingebeni, Edith Nkwembe, Eddy Kinganda-Lusamaki, Amuri Aziza, Catherine Pratt, Matthias Pauthner, Josh Quick, Allison Black, James Hadfield, Trevor Bedford, Ian Goodfellow, Nick Loman, Kristian Andersen, Michael Wiley, Steve Ahuka-Mundeke, Jean-Jacques Muyembe Tamfum
EPI_ISL_417954 hCoV-19/Spain/Madrid_H3_10/2020 Europe / Spain / Madrid 2020-03-12 Hospital Universitario 12 de Octubre Hospital Universitario La Paz Elias Dahdouh, Sara González, Fernando Lázaro, Esther Viedma, Natalia Stella, Julio García, Juan Carlos Galán, Rafael Cantón, Mª Dolores Folgueira, Rafael Delgado, Jesús Mingorance
EPI_ISL_417955 hCoV-19/DRC/191/2020 Africa / Democratic Republic of the Congo / Kinshasa 2020-03-21 Viral Respiratory Lab, National Institute for Biomedical Research (INRB) Pathogen Sequencing Lab, National Institute for Biomedical Research (INRB) Placide Mbala-Kingebeni, Edith Nkwembe, Eddy Kinganda-Lusamaki, Amuri Aziza, Catherine Pratt, Matthias Pauthner, Josh Quick, Allison Black, James Hadfield, Trevor Bedford, Ian Goodfellow, Nick Loman, Kristian Andersen, Michael Wiley, Steve Ahuka-Mundeke, Jean-Jacques Muyembe Tamfum
EPI_ISL_417956 hCoV-19/Spain/Madrid_H5_34/2020 Europe / Spain / Madrid 2020-03-11 Hospital Universitario 12 de Octubre Hospital Universitario La Paz Elias Dahdouh, Sara González, Fernando Lázaro, Esther Viedma, Natalia Stella, Julio García, Juan Carlos Galán, Rafael Cantón, Mª Dolores Folgueira, Rafael Delgado, Jesús Mingorance
EPI_ISL_417957 hCoV-19/Spain/Madrid_H7_36/2020 Europe / Spain / Madrid 2020-03-12 Hospital Universitario 12 de Octubre Hospital Universitario La Paz Elias Dahdouh, Sara González, Fernando Lázaro, Esther Viedma, Natalia Stella, Julio García, Juan Carlos Galán, Rafael Cantón, Mª Dolores Folgueira, Rafael Delgado, Jesús Mingorance
EPI_ISL_417958 hCoV-19/USA/UT-010/2020 North America / USA / Utah 2020-03-10 Utah Public Health Laboratory Utah Public Health Laboratory Erin Young, Kelly Oakeson
EPI_ISL_417960 hCoV-19/USA/UT-012/2020 North America / USA / Utah 2020-03-12 Utah Public Health Laboratory Utah Public Health Laboratory Erin Young, Kelly Oakeson
EPI_ISL_417961 hCoV-19/Spain/Madrid_H9_38/2020 Europe / Spain / Madrid 2020-03-12 Hospital Universitario 12 de Octubre Hospital Universitario La Paz Elias Dahdouh, Sara González, Fernando Lázaro, Esther Viedma, Natalia Stella, Julio García, Juan Carlos Galán, Rafael Cantón, Mª Dolores Folgueira, Rafael Delgado, Jesús Mingorance
EPI_ISL_417963 hCoV-19/Spain/Madrid_H10_39/2020 Europe / Spain / Madrid 2020-03-12 Hospital Universitario 12 de Octubre Hospital Universitario La Paz Elias Dahdouh, Sara González, Fernando Lázaro, Esther Viedma, Natalia Stella, Julio García, Juan Carlos Galán, Rafael Cantón, Mª Dolores Folgueira, Rafael Delgado, Jesús Mingorance
EPI_ISL_417964 hCoV-19/USA/UT-014/2020 North America / USA / Utah 2020-03-13 Utah Public Health Laboratory Utah Public Health Laboratory Erin Young, Kelly Oakeson
EPI_ISL_417966 hCoV-19/USA/UT-016/2020 North America / USA / Utah 2020-03-13 Utah Public Health Laboratory Utah Public Health Laboratory Erin Young, Kelly Oakeson
EPI_ISL_417967 hCoV-19/Spain/Madrid_H11_40/2020 Europe / Spain / Madrid 2020-03-12 Hospital Universitario 12 de Octubre Hospital Universitario La Paz Elias Dahdouh, Sara González, Fernando Lázaro, Esther Viedma, Natalia Stella, Julio García, Juan Carlos Galán, Rafael Cantón, Mª Dolores Folgueira, Rafael Delgado, Jesús Mingorance
EPI_ISL_417969 hCoV-19/Spain/Madrid_LP10_12/2020 Europe / Spain / Madrid 2020-03-09 Hospital Universitario La Paz Hospital Universitario La Paz Elias Dahdouh, Sara González, Fernando Lázaro, Esther Viedma, Natalia Stella, Julio García, Juan Carlos Galán, Rafael Cantón, Mª Dolores Folgueira, Rafael Delgado, Jesús Mingorance
EPI_ISL_417971 hCoV-19/USA/UT-028/2020 North America / USA / Utah 2020-03-19 Utah Public Health Laboratory Utah Public Health Laboratory Erin Young, Kelly Oakeson
EPI_ISL_417972 hCoV-19/Spain/Madrid_LP12_21/2020 Europe / Spain / Madrid 2020-03-09 Hospital Universitario La Paz Hospital Universitario La Paz Elias Dahdouh, Sara González, Fernando Lázaro, Esther

Viedma, Natalia Stella, Julio García, Juan Carlos Galán, Rafael Cantón, Mª Dolores Folgueira, Rafael Delgado, Jesús Mingorance
EPI_ISL_417973 hCoV-19/USA/UT-031/2020 North America / USA / Utah 2020-03-19 Utah Public Health Laboratory Utah Public Health Laboratory Erin Young, Kelly Oakeson
EPI_ISL_417974 hCoV-19/USA/UT-032/2020 North America / USA / Utah 2020-03-19 Utah Public Health Laboratory Utah Public Health Laboratory Erin Young, Kelly Oakeson
EPI_ISL_417975 hCoV-19/Spain/Madrid_LP14_3/2020 Europe / Spain / Madrid 2020-03-09 Hospital Universitario La Paz Hospital Universitario La Paz Elias Dahdouh, Sara González, Fernando Lázaro, Esther Viedma, Natalia Stella, Julio García, Juan Carlos Galán, Rafael Cantón, Mª Dolores Folgueira, Rafael Delgado, Jesús Mingorance
EPI_ISL_417976 hCoV-19/USA/UT-033/2020 North America / USA / Utah 2020-03-19 Utah Public Health Laboratory Utah Public Health Laboratory Erin Young, Kelly Oakeson
EPI_ISL_417977 hCoV-19/USA/UT-034/2020 North America / USA / Utah 2020-03-20 Utah Public Health Laboratory Utah Public Health Laboratory Erin Young, Kelly Oakeson
EPI_ISL_417978 hCoV-19/Spain/Madrid_LP15_4/2020 Europe / Spain / Madrid 2020-03-09 Hospital Universitario La Paz Hospital Universitario La Paz Elias Dahdouh, Sara González, Fernando Lázaro, Esther Viedma, Natalia Stella, Julio García, Juan Carlos Galán, Rafael Cantón, Mª Dolores Folgueira, Rafael Delgado, Jesús Mingorance
EPI_ISL_417979 hCoV-19/Spain/Madrid_R2_15/2020 Europe / Spain / Madrid 2020-03-03 Hospital Universitario Ramón y Cajal Hospital Universitario La Paz Elias Dahdouh, Sara González, Fernando Lázaro, Esther Viedma, Natalia Stella, Julio García, Juan Carlos Galán, Rafael Cantón, Mª Dolores Folgueira, Rafael Delgado, Jesús Mingorance
EPI_ISL_417980 hCoV-19/Spain/Madrid_R5_8/2020 Europe / Spain / Madrid 2020-03-03 Hospital Universitario Ramón y Cajal Hospital Universitario La Paz Elias Dahdouh, Sara González, Fernando Lázaro, Esther Viedma, Natalia Stella, Julio García, Juan Carlos Galán, Rafael Cantón, Mª Dolores Folgueira, Rafael Delgado, Jesús Mingorance
EPI_ISL_417981 hCoV-19/Spain/Madrid_R10_33/2020 Europe / Spain / Madrid 2020-03-02 Hospital Universitario Ramón y Cajal Hospital Universitario La Paz Elias Dahdouh, Sara González, Fernando Lázaro, Esther Viedma, Natalia Stella, Julio García, Juan Carlos Galán, Rafael Cantón, Mª Dolores Folgueira, Rafael Delgado, Jesús Mingorance
EPI_ISL_417986 hCoV-19/Portugal/PT0001b/2020 Europe / Portugal 2020-03-03 Centro Hospitalar e Universitario de Sao Joao, Porto Instituto Nacional de Saude (INSA) Guiomar et al
EPI_ISL_417987 hCoV-19/Portugal/PT0003/2020 Europe / Portugal 2020-03-03 Centro Hospitalar e Universitario de Sao Joao, Porto Instituto Nacional de Saude (INSA) Guiomar et al
EPI_ISL_417988 hCoV-19/Portugal/PT0004/2020 Europe / Portugal 2020-03-05 CHULC - H Curry Cabral Instituto Nacional de Saude (INSA) Guiomar et al
EPI_ISL_417989 hCoV-19/Portugal/PT0005/2020 Europe / Portugal 2020-03-04 Centro Hospitalar e Universitario de Sao Joao, Porto Instituto Nacional de Saude (INSA) Guiomar et al
EPI_ISL_417990 hCoV-19/Portugal/PT0006a/2020 Europe / Portugal 2020-03-06 CHULC - H Curry Cabral Instituto Nacional de Saude (INSA) Guiomar et al
EPI_ISL_417991 hCoV-19/Portugal/PT0006b/2020 Europe / Portugal 2020-03-08 CHULC - H Curry Cabral Instituto Nacional de Saude (INSA) Guiomar et al
EPI_ISL_417992 hCoV-19/Portugal/PT0007/2020 Europe / Portugal 2020-03-07 CHULC - H D Estefania Instituto Nacional de Saude (INSA) Guiomar et al
EPI_ISL_417993 hCoV-19/Portugal/PT0008/2020 Europe / Portugal 2020-03-08 CHULC - H D Estefania Instituto Nacional de Saude (INSA) Guiomar et al
EPI_ISL_417994 hCoV-19/Portugal/PT0009/2020 Europe / Portugal 2020-03-08 CHULC - H Curry Cabral Instituto Nacional de Saude (INSA) Guiomar et al
EPI_ISL_417995 hCoV-19/Portugal/PT0010/2020 Europe / Portugal 2020-03-08 CHULC - H Curry Cabral Instituto Nacional de Saude (INSA) Guiomar et al
EPI_ISL_417996 hCoV-19/Portugal/PT0011/2020 Europe / Portugal 2020-03-08 CHULC - H Curry Cabral Instituto Nacional de Saude (INSA) Guiomar et al
EPI_ISL_417997 hCoV-19/Portugal/PT0012/2020 Europe / Portugal 2020-03-07 Centro Hospital do Porto, E.P.E. - H. Geral de Santo Antonio Instituto Nacional de Saude (INSA) Guiomar et al
EPI_ISL_417998 hCoV-19/Portugal/PT0013/2020 Europe / Portugal 2020-03-08 Centro Hospital do Porto, E.P.E. - H. Geral de Santo Antonio Instituto Nacional de Saude (INSA) Guiomar et al
EPI_ISL_417999 hCoV-19/Portugal/PT0014/2020 Europe / Portugal 2020-03-07 Centro Hospital do Porto, E.P.E. - H. Geral de Santo Antonio Instituto Nacional de Saude (INSA) Guiomar et al
EPI_ISL_418700 hCoV-19/England/20124013202/2020 Europe / United Kingdom / England 2020-03-17 Respiratory Virus Unit, Microbiology Services Colindale, Public Health England Respiratory Virus Unit, Microbiology Services Colindale, Public Health England Monica Galiano, Shahjahan Miah, Angie Lackenby, Omolola Akinbami, Tiina Talts, Leena Bhaw, Richard Myers, Steven Platt, Kirstin Edwards, Jonathan Hubb, Joanna Ellis, Maria Zambon
EPI_ISL_418701 hCoV-19/England/20124020402/2020 Europe / United Kingdom / England 2020-03-17 Respiratory Virus Unit, Microbiology Services Colindale, Public Health England Respiratory Virus Unit, Microbiology Services Colindale, Public Health England Monica Galiano, Shahjahan Miah, Angie Lackenby, Omolola Akinbami, Tiina Talts, Leena Bhaw, Richard Myers, Steven Platt, Kirstin Edwards, Jonathan Hubb, Joanna Ellis, Maria Zambon
EPI_ISL_418702 hCoV-19/England/20124020802/2020 Europe / United Kingdom / England 2020-03-16 Respiratory Virus Unit, Microbiology Services Colindale, Public Health England Respiratory Virus Unit,

Microbiology Services Colindale, Public Health England Monica Galiano, Shahjahan Miah, Angie Lackenby, Omolola Akinbami, Tiina Talts, Leena Bhaw, Richard Myers, Steven Platt, Kirstin Edwards, Jonathan Hubb, Joanna Ellis, Maria Zambon
EPI_ISL_418704 hCoV-19/England/20124021402/2020 Europe / United Kingdom / England 2020-03-17 Respiratory Virus Unit, Microbiology Services Colindale, Public Health England Respiratory Virus Unit, Microbiology Services Colindale, Public Health England Monica Galiano, Shahjahan Miah, Angie Lackenby, Omolola Akinbami, Tiina Talts, Leena Bhaw, Richard Myers, Steven Platt, Kirstin Edwards, Jonathan Hubb, Joanna Ellis, Maria Zambon
EPI_ISL_418706 hCoV-19/England/20124029802/2020 Europe / United Kingdom / England 2020-03-17 Respiratory Virus Unit, Microbiology Services Colindale, Public Health England Respiratory Virus Unit, Microbiology Services Colindale, Public Health England Monica Galiano, Shahjahan Miah, Angie Lackenby, Omolola Akinbami, Tiina Talts, Leena Bhaw, Richard Myers, Steven Platt, Kirstin Edwards, Jonathan Hubb, Joanna Ellis, Maria Zambon
EPI_ISL_418707 hCoV-19/England/20124030003/2020 Europe / United Kingdom / England 2020-03-17 Respiratory Virus Unit, Microbiology Services Colindale, Public Health England Respiratory Virus Unit, Microbiology Services Colindale, Public Health England Monica Galiano, Shahjahan Miah, Angie Lackenby, Omolola Akinbami, Tiina Talts, Leena Bhaw, Richard Myers, Steven Platt, Kirstin Edwards, Jonathan Hubb, Joanna Ellis, Maria Zambon
EPI_ISL_418708 hCoV-19/England/20124030802/2020 Europe / United Kingdom / England 2020-03-17 Respiratory Virus Unit, Microbiology Services Colindale, Public Health England Respiratory Virus Unit, Microbiology Services Colindale, Public Health England Monica Galiano, Shahjahan Miah, Angie Lackenby, Omolola Akinbami, Tiina Talts, Leena Bhaw, Richard Myers, Steven Platt, Kirstin Edwards, Jonathan Hubb, Joanna Ellis, Maria Zambon
EPI_ISL_418718 hCoV-19/England/20124036902/2020 Europe / United Kingdom / England 2020-03-18 Respiratory Virus Unit, Microbiology Services Colindale, Public Health England Respiratory Virus Unit, Microbiology Services Colindale, Public Health England Monica Galiano, Shahjahan Miah, Angie Lackenby, Omolola Akinbami, Tiina Talts, Leena Bhaw, Richard Myers, Steven Platt, Kirstin Edwards, Jonathan Hubb, Joanna Ellis, Maria Zambon
EPI_ISL_418720 hCoV-19/England/20124037302/2020 Europe / United Kingdom / England 2020-03-17 Respiratory Virus Unit, Microbiology Services Colindale, Public Health England Respiratory Virus Unit, Microbiology Services Colindale, Public Health England Monica Galiano, Shahjahan Miah, Angie Lackenby, Omolola Akinbami, Tiina Talts, Leena Bhaw, Richard Myers, Steven Platt, Kirstin Edwards, Jonathan Hubb, Joanna Ellis, Maria Zambon
EPI_ISL_418722 hCoV-19/England/20124048202/2020 Europe / United Kingdom / England 2020-03-17 Respiratory Virus Unit, Microbiology Services Colindale, Public Health England Respiratory Virus Unit, Microbiology Services Colindale, Public Health England Monica Galiano, Shahjahan Miah, Angie Lackenby, Omolola Akinbami, Tiina Talts, Leena Bhaw, Richard Myers, Steven Platt, Kirstin Edwards, Jonathan Hubb, Joanna Ellis, Maria Zambon
EPI_ISL_418723 hCoV-19/England/20124049402/2020 Europe / United Kingdom / England 2020-03-16 Respiratory Virus Unit, Microbiology Services Colindale, Public Health England Respiratory Virus Unit, Microbiology Services Colindale, Public Health England Monica Galiano, Shahjahan Miah, Angie Lackenby, Omolola Akinbami, Tiina Talts, Leena Bhaw, Richard Myers, Steven Platt, Kirstin Edwards, Jonathan Hubb, Joanna Ellis, Maria Zambon
EPI_ISL_418729 hCoV-19/England/20124095502/2020 Europe / United Kingdom / England 2020-03-18 Respiratory Virus Unit, Microbiology Services Colindale, Public Health England Respiratory Virus Unit, Microbiology Services Colindale, Public Health England Monica Galiano, Shahjahan Miah, Angie Lackenby, Omolola Akinbami, Tiina Talts, Leena Bhaw, Richard Myers, Steven Platt, Kirstin Edwards, Jonathan Hubb, Joanna Ellis, Maria Zambon
EPI_ISL_418733 hCoV-19/England/20124096802/2020 Europe / United Kingdom / England 2020-03-18 Respiratory Virus Unit, Microbiology Services Colindale, Public Health England Respiratory Virus Unit, Microbiology Services Colindale, Public Health England Monica Galiano, Shahjahan Miah, Angie Lackenby, Omolola Akinbami, Tiina Talts, Leena Bhaw, Richard Myers, Steven Platt, Kirstin Edwards, Jonathan Hubb, Joanna Ellis, Maria Zambon
EPI_ISL_418734 hCoV-19/England/20124097102/2020 Europe / United Kingdom / England 2020-03-13 Respiratory Virus Unit, Microbiology Services Colindale, Public Health England Respiratory Virus Unit, Microbiology Services Colindale, Public Health England Monica Galiano, Shahjahan Miah, Angie Lackenby, Omolola Akinbami, Tiina Talts, Leena Bhaw, Richard Myers, Steven Platt, Kirstin Edwards, Jonathan Hubb, Joanna Ellis, Maria Zambon
EPI_ISL_418737 hCoV-19/England/20124100002/2020 Europe / United Kingdom / England 2020-03-17 Respiratory Virus Unit, Microbiology Services Colindale, Public Health England Respiratory Virus Unit, Microbiology Services Colindale, Public Health England Monica Galiano, Shahjahan Miah, Angie Lackenby, Omolola Akinbami, Tiina Talts, Leena Bhaw, Richard Myers, Steven Platt, Kirstin Edwards, Jonathan Hubb, Joanna Ellis, Maria Zambon
EPI_ISL_419000 hCoV-19/Singapore/21/2020 Asia / Singapore 2020-02-13 National Public Health Laboratory, National Centre for Infectious Diseases National Public Health Laboratory, National Centre for Infectious Diseases Mak TM, Octavia S, Cui L, Lin RTP
EPI_ISL_419001 hCoV-19/Singapore/19/2020 Asia / Singapore 2020-03-02 National Public Health Laboratory, National Centre for Infectious Diseases National Public Health Laboratory, National Centre for Infectious Diseases Mak TM, Octavia S, Cui L, Lin RTP
EPI_ISL_421171 hCoV-19/Spain/Madrid_H12_1301/2020 Europe / Spain/ Madrid 2020-03-05 Hospital

Universitario 12 de Octubre	Hospital Universitario 12 de Octubre	Esther Viedma, Sara González, Elias Dahdouh, Raúl Recio, Fernando Lázaro, Julio García, Mª Dolores Folgueira, Jesús Mingorance, Rafael Delgado
EPI_ISL_421172	hCoV-19/Spain/Madrid_H12_1502/2020	Europe / Spain/ Madrid	2020-03-09	Hospital Universitario 12 de Octubre	Hospital Universitario 12 de Octubre	Esther Viedma, Sara González, Elias Dahdouh, Raúl Recio, Fernando Lázaro, Julio García, Mª Dolores Folgueira, Jesús Mingorance, Rafael Delgado
EPI_ISL_421173	hCoV-19/Spain/Madrid_H12_1703/2020	Europe / Spain / Madrid	2020-03-08	Hospital Universitario 12 de Octubre	Hospital Universitario 12 de Octubre	Esther Viedma, Sara González, Elias Dahdouh, Raúl Recio, Fernando Lázaro, Julio García, Mª Dolores Folgueira, Jesús Mingorance, Rafael Delgado
EPI_ISL_421174	hCoV-19/Spain/Madrid_H12_1804/2021	Europe / Spain / Madrid	2020-03-29	Hospital Universitario 12 de Octubre	Hospital Universitario 12 de Octubre	Esther Viedma, Sara González, Elias Dahdouh, Raúl Recio, Fernando Lázaro, Julio García, Mª Dolores Folgueira, Jesús Mingorance, Rafael Delgado
EPI_ISL_421175	hCoV-19/Spain/Madrid_H12_1905/2020	Europe / Spain / Madrid	2020-03-29	Hospital Universitario 12 de Octubre	Hospital Universitario 12 de Octubre	Esther Viedma, Sara González, Elias Dahdouh, Raúl Recio, Fernando Lázaro, Julio García, Mª Dolores Folgueira, Jesús Mingorance, Rafael Delgado
EPI_ISL_421176	hCoV-19/Spain/Madrid_H12_2006/2020	Europe / Spain / Madrid	2020-03-27	Hospital Universitario 12 de Octubre	Hospital Universitario 12 de Octubre	Esther Viedma, Sara González, Elias Dahdouh, Raúl Recio, Fernando Lázaro, Julio García, Mª Dolores Folgueira, Jesús Mingorance, Rafael Delgado
EPI_ISL_421177	hCoV-19/Spain/Madrid_H12_2107/2020	Europe / Spain / Madrid	2020-03-27	Hospital Universitario 12 de Octubre	Hospital Universitario 12 de Octubre	Esther Viedma, Sara González, Elias Dahdouh, Raúl Recio, Fernando Lázaro, Julio García, Mª Dolores Folgueira, Jesús Mingorance, Rafael Delgado
EPI_ISL_421178	hCoV-19/Spain/Madrid_H12_2208/2020	Europe / Spain / Madrid	2020-03-27	Hospital Universitario 12 de Octubre	Hospital Universitario 12 de Octubre	Esther Viedma, Sara González, Elias Dahdouh, Raúl Recio, Fernando Lázaro, Julio García, Mª Dolores Folgueira, Jesús Mingorance, Rafael Delgado
EPI_ISL_421179	hCoV-19/Spain/Madrid_H12_2309/2020	Europe / Spain / Madrid	2020-03-27	Hospital Universitario 12 de Octubre	Hospital Universitario 12 de Octubre	Esther Viedma, Sara González, Elias Dahdouh, Raúl Recio, Fernando Lázaro, Julio García, Mª Dolores Folgueira, Jesús Mingorance, Rafael Delgado
EPI_ISL_421180	hCoV-19/Spain/Madrid_H12_2410/2020	Europe / Spain / Madrid	2020-03-27	Hospital Universitario 12 de Octubre	Hospital Universitario 12 de Octubre	Esther Viedma, Sara González, Elias Dahdouh, Raúl Recio, Fernando Lázaro, Julio García, Mª Dolores Folgueira, Jesús Mingorance, Rafael Delgado
EPI_ISL_421182	hCoV-19/Belgium/ULG-10001/2020	Europe / Belgium / Liège	2020-03-30	Department of Clinical Microbiology	GIGA Medical Genomics	Keith Durkin, Maria Artesi, Sébastien Bontems, Raphaël Boreux, Cécile Meex, Pierrette Melin, Marie-Pierre Hayette, Vincent Bours.
EPI_ISL_421183	hCoV-19/Belgium/ULG-10003/2020	Europe / Belgium / Liège	2020-03-30	Department of Clinical Microbiology	GIGA Medical Genomics	Keith Durkin, Maria Artesi, Sébastien Bontems, Raphaël Boreux, Cécile Meex, Pierrette Melin, Marie-Pierre Hayette, Vincent Bours.
EPI_ISL_421184	hCoV-19/Belgium/ULG-10004/2020	Europe / Belgium / Liège	2020-03-31	Department of Clinical Microbiology	GIGA Medical Genomics	Keith Durkin, Maria Artesi, Sébastien Bontems, Raphaël Boreux, Cécile Meex, Pierrette Melin, Marie-Pierre Hayette, Vincent Bours.
EPI_ISL_421185	hCoV-19/Belgium/ULG-10006/2020	Europe / Belgium / Liège	2020-03-30	Department of Clinical Microbiology	GIGA Medical Genomics	Keith Durkin, Maria Artesi, Sébastien Bontems, Raphaël Boreux, Cécile Meex, Pierrette Melin, Marie-Pierre Hayette, Vincent Bours.
EPI_ISL_421186	hCoV-19/Belgium/ULG-10007/2020	Europe / Belgium / Liège	2020-03-30	Department of Clinical Microbiology	GIGA Medical Genomics	Keith Durkin, Maria Artesi, Sébastien Bontems, Raphaël Boreux, Cécile Meex, Pierrette Melin, Marie-Pierre Hayette, Vincent Bours.
EPI_ISL_421187	hCoV-19/Belgium/ULG-10008/2020	Europe / Belgium / Liège	2020-03-30	Department of Clinical Microbiology	GIGA Medical Genomics	Keith Durkin, Maria Artesi, Sébastien Bontems, Raphaël Boreux, Cécile Meex, Pierrette Melin, Marie-Pierre Hayette, Vincent Bours.
EPI_ISL_421188	hCoV-19/Belgium/ULG-10009/2020	Europe / Belgium / Liège	2020-03-30	Department of Clinical Microbiology	GIGA Medical Genomics	Keith Durkin, Maria Artesi, Sébastien Bontems, Raphaël Boreux, Cécile Meex, Pierrette Melin, Marie-Pierre Hayette, Vincent Bours.
EPI_ISL_421189	hCoV-19/Belgium/ULG-10011/2020	Europe / Belgium / Liège	2020-03-30	Department of Clinical Microbiology	GIGA Medical Genomics	Keith Durkin, Maria Artesi, Sébastien Bontems, Raphaël Boreux, Cécile Meex, Pierrette Melin, Marie-Pierre Hayette, Vincent Bours.
EPI_ISL_421190	hCoV-19/Belgium/ULG-10012/2020	Europe / Belgium / Liège	2020-03-31	Department of Clinical Microbiology	GIGA Medical Genomics	Keith Durkin, Maria Artesi, Sébastien Bontems, Raphaël Boreux, Cécile Meex, Pierrette Melin, Marie-Pierre Hayette, Vincent Bours.
EPI_ISL_421191	hCoV-19/Belgium/ULG-10013/2020	Europe / Belgium / Liège	2020-03-30	Department of Clinical Microbiology	GIGA Medical Genomics	Keith Durkin, Maria Artesi, Sébastien Bontems, Raphaël Boreux, Cécile Meex, Pierrette Melin, Marie-Pierre Hayette, Vincent Bours.
EPI_ISL_421192	hCoV-19/Belgium/ULG-10014/2020	Europe / Belgium / Liège	2020-03-30	Department of Clinical Microbiology	GIGA Medical Genomics	Keith Durkin, Maria Artesi, Sébastien Bontems, Raphaël Boreux, Cécile Meex, Pierrette Melin, Marie-Pierre Hayette, Vincent Bours.
EPI_ISL_421193	hCoV-19/Belgium/ULG-10015/2020	Europe / Belgium / Liège	2020-03-31	Department of Clinical Microbiology	GIGA Medical Genomics	Keith Durkin, Maria Artesi, Sébastien Bontems, Raphaël Boreux, Cécile Meex, Pierrette Melin, Marie-Pierre Hayette, Vincent Bours.
EPI_ISL_421194	hCoV-19/Belgium/ULG-10016/2020	Europe / Belgium / Liège	2020-03-30	Department of Clinical Microbiology	GIGA Medical Genomics	Keith Durkin, Maria Artesi, Sébastien Bontems, Raphaël Boreux, Cécile Meex, Pierrette Melin, Marie-Pierre Hayette, Vincent Bours.
EPI_ISL_421195	hCoV-19/Belgium/ULG-10017/2020	Europe / Belgium / Liège	2020-03-30	Department of Clinical Microbiology	GIGA Medical Genomics	Keith Durkin, Maria Artesi, Sébastien Bontems, Raphaël Boreux,

Cécile Meex, Pierrette Melin, Marie-Pierre Hayette, Vincent Bours.
EPI_ISL_421196 hCoV-19/Belgium/ULG-10018/2020 Europe / Belgium / Liège 2020-03-30 Department of Clinical Microbiology GIGA Medical Genomics Keith Durkin, Maria Artesi, Sébastien Bontems, Raphaël Boreux, Cécile Meex, Pierrette Melin, Marie-Pierre Hayette, Vincent Bours.
EPI_ISL_421198 hCoV-19/Belgium/ULG-10024/2020 Europe / Belgium / Liège 2020-03-30 Department of Clinical Microbiology GIGA Medical Genomics Keith Durkin, Maria Artesi, Sébastien Bontems, Raphaël Boreux, Cécile Meex, Pierrette Melin, Marie-Pierre Hayette, Vincent Bours.
EPI_ISL_421200 hCoV-19/Belgium/ULG-10026/2020 Europe / Belgium / Liège 2020-03-31 Department of Clinical Microbiology GIGA Medical Genomics Keith Durkin, Maria Artesi, Sébastien Bontems, Raphaël Boreux, Cécile Meex, Pierrette Melin, Marie-Pierre Hayette, Vincent Bours.
EPI_ISL_421201 hCoV-19/Belgium/ULG-10027/2020 Europe / Belgium / Liège 2020-03-31 Department of Clinical Microbiology GIGA Medical Genomics Keith Durkin, Maria Artesi, Sébastien Bontems, Raphaël Boreux, Cécile Meex, Pierrette Melin, Marie-Pierre Hayette, Vincent Bours.
EPI_ISL_421202 hCoV-19/Belgium/ULG-10028/2020 Europe / Belgium / Liège 2020-03-30 Department of Clinical Microbiology GIGA Medical Genomics Keith Durkin, Maria Artesi, Sébastien Bontems, Raphaël Boreux, Cécile Meex, Pierrette Melin, Marie-Pierre Hayette, Vincent Bours.
EPI_ISL_421203 hCoV-19/Belgium/ULG-10029/2020 Europe / Belgium / Liège 2020-03-30 Department of Clinical Microbiology GIGA Medical Genomics Keith Durkin, Maria Artesi, Sébastien Bontems, Raphaël Boreux, Cécile Meex, Pierrette Melin, Marie-Pierre Hayette, Vincent Bours.
EPI_ISL_421204 hCoV-19/Belgium/ULG-10030/2020 Europe / Belgium / Liège 2020-03-31 Department of Clinical Microbiology GIGA Medical Genomics Keith Durkin, Maria Artesi, Sébastien Bontems, Raphaël Boreux, Cécile Meex, Pierrette Melin, Marie-Pierre Hayette, Vincent Bours.
EPI_ISL_421205 hCoV-19/Belgium/ULG-10031/2020 Europe / Belgium / Liège 2020-03-31 Department of Clinical Microbiology GIGA Medical Genomics Keith Durkin, Maria Artesi, Sébastien Bontems, Raphaël Boreux, Cécile Meex, Pierrette Melin, Marie-Pierre Hayette, Vincent Bours.
EPI_ISL_421206 hCoV-19/Belgium/ULG-10032/2020 Europe / Belgium / Liège 2020-03-31 Department of Clinical Microbiology GIGA Medical Genomics Keith Durkin, Maria Artesi, Sébastien Bontems, Raphaël Boreux, Cécile Meex, Pierrette Melin, Marie-Pierre Hayette, Vincent Bours.
EPI_ISL_421207 hCoV-19/Belgium/ULG-10033/2020 Europe / Belgium / Liège 2020-03-31 Department of Clinical Microbiology GIGA Medical Genomics Keith Durkin, Maria Artesi, Sébastien Bontems, Raphaël Boreux, Cécile Meex, Pierrette Melin, Marie-Pierre Hayette, Vincent Bours.
EPI_ISL_421210 hCoV-19/Belgium/ULG-10043/2020 Europe / Belgium / Liège 2020-03-31 Department of Clinical Microbiology GIGA Medical Genomics Keith Durkin, Maria Artesi, Sébastien Bontems, Raphaël Boreux, Cécile Meex, Pierrette Melin, Marie-Pierre Hayette, Vincent Bours.
EPI_ISL_421212 hCoV-19/Belgium/ULG-10045/2020 Europe / Belgium / Liège 2020-03-31 Department of Clinical Microbiology GIGA Medical Genomics Keith Durkin, Maria Artesi, Sébastien Bontems, Raphaël Boreux, Cécile Meex, Pierrette Melin, Marie-Pierre Hayette, Vincent Bours.
EPI_ISL_421213 hCoV-19/Belgium/ULG-10046/2020 Europe / Belgium / Liège 2020-03-31 Department of Clinical Microbiology GIGA Medical Genomics Keith Durkin, Maria Artesi, Sébastien Bontems, Raphaël Boreux, Cécile Meex, Pierrette Melin, Marie-Pierre Hayette, Vincent Bours.
EPI_ISL_421214 hCoV-19/Belgium/ULG-10047/2020 Europe / Belgium / Liège 2020-03-30 Department of Clinical Microbiology GIGA Medical Genomics Keith Durkin, Maria Artesi, Sébastien Bontems, Raphaël Boreux, Cécile Meex, Pierrette Melin, Marie-Pierre Hayette, Vincent Bours.
EPI_ISL_421221 hCoV-19/Hangzhou/HZCDC6789/2020 Asia / China / Hangzhou 2020-03-15 Hangzhou Center for Diseases Control and Prevention Hangzhou Center for Diseases Control and Prevention Jun Li, Haoqiu Wang, Lingfeng Mao, Hua Yu, Xinfen Yu, Zhou Sun, Xin Qian, Shuchang Chen, Junfang Chen, Xuchu Wang
EPI_ISL_421222 hCoV-19/Hangzhou/HZCDC6706/2020 Asia / China / Hangzhou 2020-03-14 Hangzhou Center for Diseases Control and Prevention Hangzhou Center for Diseases Control and Prevention Jun Li, Haoqiu Wang, Lingfeng Mao, Hua Yu, Xinfen Yu, Zhou Sun, Xin Qian, Shuchang Chen, Junfang Chen, Xuchu Wang
EPI_ISL_421224 hCoV-19/Hangzhou/HZCDC0162/2020 Asia / China / Hangzhou 2020-01-23 Hangzhou Center for Diseases Control and Prevention Hangzhou Center for Diseases Control and Prevention Jun Li, Haoqiu Wang, Lingfeng Mao, Hua Yu, Xinfen Yu, Zhou Sun, Xin Qian, Shuchang Chen, Junfang Chen, Xuchu Wang
EPI_ISL_421300 hCoV-19/USA/WI-UW-42/2020 North America / USA / Wisconsin / Madison 2020-03-18 University of Wisconsin-Madison AIDS Vaccine Research Laboratories University of Wisconsin-Madison AIDS Vaccine Research Laboratories Gage Moreno, Katarina Braun, et al. AIDS Vaccine Research Laboratories
EPI_ISL_421301 hCoV-19/USA/WI-UW-43/2020 North America / USA / Wisconsin / Waunakee 2020-03-19 University of Wisconsin-Madison AIDS Vaccine Research Laboratories University of Wisconsin-Madison AIDS Vaccine Research Laboratories Gage Moreno, Katarina Braun, et al. AIDS Vaccine Research Laboratories
EPI_ISL_421302 hCoV-19/USA/WI-UW-44/2020 North America / USA / Wisconsin / Madison 2020-03-17 University of Wisconsin-Madison AIDS Vaccine Research Laboratories University of Wisconsin-Madison AIDS Vaccine Research Laboratories Gage Moreno, Katarina Braun, et al. AIDS Vaccine Research Laboratories
EPI_ISL_421304 hCoV-19/USA/WI-UW-46/2020 North America / USA / Wisconsin / Belleville 2020-03-22 University of Wisconsin-Madison AIDS Vaccine Research Laboratories University of Wisconsin-Madison AIDS Vaccine Research Laboratories Gage Moreno, Katarina Braun, et al. AIDS Vaccine Research Laboratories
EPI_ISL_421305 hCoV-19/USA/WI-UW-47/2020 North America / USA / Wisconsin / Chippewa Falls 2020-03-19 University of Wisconsin-Madison AIDS Vaccine Research Laboratories University of Wisconsin-Madison AIDS Vaccine Research Laboratories Gage Moreno, Katarina Braun, et al. AIDS Vaccine Research Laboratories
EPI_ISL_421306 hCoV-19/USA/WI-UW-48/2020 North America / USA / Wisconsin / DeForest 2020-03-25 University of Wisconsin-Madison AIDS Vaccine Research Laboratories University of Wisconsin-Madison AIDS Vaccine Research Laboratories Gage Moreno, Katarina Braun, et al. AIDS Vaccine Research Laboratories

EPI_ISL_421307 hCoV-19/USA/WI-UW-49/2020 North America / USA / Wisconsin / Sun Prarie 2020-03-25 University of Wisconsin-Madison AIDS Vaccine Research Laboratories University of Wisconsin-Madison AIDS Vaccine Research Laboratories Gage Moreno, Katarina Braun, et al. AIDS Vaccine Research Laboratories
EPI_ISL_421308 hCoV-19/USA/WI-UW-50/2020 North America / USA / Wisconsin / Brooklyn 2020-03-25 University of Wisconsin-Madison AIDS Vaccine Research Laboratories University of Wisconsin-Madison AIDS Vaccine Research Laboratories Gage Moreno, Katarina Braun, et al. AIDS Vaccine Research Laboratories
EPI_ISL_421309 hCoV-19/USA/WI-UW-51/2020 North America / USA / Wisconsin / Madison 2020-03-20 University of Wisconsin-Madison AIDS Vaccine Research Laboratories University of Wisconsin-Madison AIDS Vaccine Research Laboratories Gage Moreno, Katarina Braun, et al. AIDS Vaccine Research Laboratories
EPI_ISL_421310 hCoV-19/USA/WI-UW-52/2020 North America / USA / Wisconsin / Madison 2020-03-18 University of Wisconsin-Madison AIDS Vaccine Research Laboratories University of Wisconsin-Madison AIDS Vaccine Research Laboratories Gage Moreno, Katarina Braun, et al. AIDS Vaccine Research Laboratories
EPI_ISL_421311 hCoV-19/USA/WI-UW-53/2020 North America / USA / Wisconsin / Madison 2020-03-18 University of Wisconsin-Madison AIDS Vaccine Research Laboratories University of Wisconsin-Madison AIDS Vaccine Research Laboratories Gage Moreno, Katarina Braun, et al. AIDS Vaccine Research Laboratories
EPI_ISL_421312 hCoV-19/USA/WI-UW-54/2020 North America / USA / Wisconsin / Monona 2020-03-20 University of Wisconsin-Madison AIDS Vaccine Research Laboratories University of Wisconsin-Madison AIDS Vaccine Research Laboratories Gage Moreno, Katarina Braun, et al. AIDS Vaccine Research Laboratories
EPI_ISL_421313 hCoV-19/USA/WI-UW-55/2020 North America / USA / Wisconsin / Madison 2020-03-23 University of Wisconsin-Madison AIDS Vaccine Research Laboratories University of Wisconsin-Madison AIDS Vaccine Research Laboratories Gage Moreno, Katarina Braun, et al. AIDS Vaccine Research Laboratories
EPI_ISL_421314 hCoV-19/USA/WI-UW-56/2020 North America / USA / Wisconsin / Waunakee 2020-03-18 University of Wisconsin-Madison AIDS Vaccine Research Laboratories University of Wisconsin-Madison AIDS Vaccine Research Laboratories Gage Moreno, Katarina Braun, et al. AIDS Vaccine Research Laboratories
EPI_ISL_421315 hCoV-19/USA/WI-UW-57/2020 North America / USA / Wisconsin / Blanchardville 2020-03-19 University of Wisconsin-Madison AIDS Vaccine Research Laboratories University of Wisconsin-Madison AIDS Vaccine Research Laboratories Gage Moreno, Katarina Braun, et al. AIDS Vaccine Research Laboratories
EPI_ISL_421316 hCoV-19/USA/WI-UW-58/2020 North America / USA / Wisconsin / Madison 2020-03-23 University of Wisconsin-Madison AIDS Vaccine Research Laboratories University of Wisconsin-Madison AIDS Vaccine Research Laboratories Gage Moreno, Katarina Braun, et al. AIDS Vaccine Research Laboratories
EPI_ISL_421317 hCoV-19/USA/WI-UW-59/2020 North America / USA / Wisconsin / DeForest 2020-03-24 University of Wisconsin-Madison AIDS Vaccine Research Laboratories University of Wisconsin-Madison AIDS Vaccine Research Laboratories Gage Moreno, Katarina Braun, et al. AIDS Vaccine Research Laboratories
EPI_ISL_421318 hCoV-19/USA/WI-UW-60/2020 North America / USA / Wisconsin / Madison 2020-03-19 University of Wisconsin-Madison AIDS Vaccine Research Laboratories University of Wisconsin-Madison AIDS Vaccine Research Laboratories Gage Moreno, Katarina Braun, et al. AIDS Vaccine Research Laboratories
EPI_ISL_421319 hCoV-19/USA/WI-UW-61/2020 North America / USA / Wisconsin / Stoughton 2020-03-23 University of Wisconsin-Madison AIDS Vaccine Research Laboratories University of Wisconsin-Madison AIDS Vaccine Research Laboratories Gage Moreno, Katarina Braun, et al. AIDS Vaccine Research Laboratories
EPI_ISL_421320 hCoV-19/USA/WI-UW-62/2020 North America / USA / Wisconsin / Madison 2020-03-25 University of Wisconsin-Madison AIDS Vaccine Research Laboratories University of Wisconsin-Madison AIDS Vaccine Research Laboratories Gage Moreno, Katarina Braun, et al. AIDS Vaccine Research Laboratories
EPI_ISL_421321 hCoV-19/USA/WI-UW-63/2020 North America / USA / Wisconsin / Sun Prarie 2020-03-24 University of Wisconsin-Madison AIDS Vaccine Research Laboratories University of Wisconsin-Madison AIDS Vaccine Research Laboratories Gage Moreno, Katarina Braun, et al. AIDS Vaccine Research Laboratories
EPI_ISL_421322 hCoV-19/USA/WI-UW-64/2020 North America / USA / Wisconsin / Verona 2020-03-24 University of Wisconsin-Madison AIDS Vaccine Research Laboratories University of Wisconsin-Madison AIDS Vaccine Research Laboratories Gage Moreno, Katarina Braun, et al. AIDS Vaccine Research Laboratories
EPI_ISL_421323 hCoV-19/USA/WI-UW-65/2020 North America / USA / Wisconsin / Madison 2020-03-22 University of Wisconsin-Madison AIDS Vaccine Research Laboratories University of Wisconsin-Madison AIDS Vaccine Research Laboratories Gage Moreno, Katarina Braun, et al. AIDS Vaccine Research Laboratories
EPI_ISL_422435 hCoV-19/Singapore/42/2020 Asia / Singapore 2020-03-31 National Public Health Laboratory, National Centre for Infectious Diseases National Public Health Laboratory, National Centre for Infectious Diseases Mak TM, Octavia S, Cui L, Lin RTP
EPI_ISL_422437 hCoV-19/Italy/VR_20COV21-26/2020 Europe / Italy / Veneto / Verona 2020-03-25 ULSS9 Distretto di Bussolengo Istituto Zooprofilattico Sperimentale delle Venezie Adelaide Milani, Alessia Schivo, Annalisa Salviato, Erika Giorgia Quaranta, Gianpiero Zamperin, Ambra Pastori, Bianca Zecchin, Alice Fusaro, Calogero Terregino, Antonia Ricci
EPI_ISL_422438 hCoV-19/Italy/VR_20COV21-37/2020 Europe / Italy / Veneto / Verona 2020-03-25 ULSS9 Distretto di Bussolengo Istituto Zooprofilattico Sperimentale delle Venezie Adelaide Milani, Alessia Schivo, Annalisa Salviato, Erika Giorgia Quaranta, Gianpiero Zamperin, Ambra Pastori, Bianca Zecchin, Alice Fusaro, Calogero Terregino, Antonia Ricci
EPI_ISL_422453 hCoV-19/USA/WI-GMF-00498/2020 North America / USA / Wisconsin / Adams County 2020-03-31 Gundersen Molecular Diagnostics Laboratory Kabara Cancer Research Institute Craig S. Richmond & Paraic A. Kenny
EPI_ISL_422459 hCoV-19/USA/WI-GMF-00557/2020 North America / USA / Wisconsin / Monroe County 2020-04-02 Gundersen Molecular Diagnostics Laboratory Kabara Cancer Research Institute Craig S. Richmond & Paraic A. Kenny
EPI_ISL_422461 hCoV-19/USA/WI-GMF-00707/2020 North America / USA / Iowa / Allamakee County 2020-04-06 Gundersen Molecular Diagnostics Laboratory Kabara Cancer Research Institute Craig S. Richmond,

Paraic A. Kenny
EPI_ISL_422462 hCoV-19/USA/WI-GMF-00744/2020 North America / USA / Iowa / Winneshiek County 2020-04-03 Gundersen Molecular Diagnostics Laboratory Kabara Cancer Research Institute Craig S. Richmond, Paraic A. Kenny
EPI_ISL_422463 hCoV-19/USA/WI-GMF-00588/2020 North America / USA / Minnesota / Winona County 2020-04-06 Gundersen Molecular Diagnostics Laboratory Kabara Cancer Research Institute Craig S. Richmond, Paraic A. Kenny
EPI_ISL_422465 hCoV-19/USA/WI-GMF-00441/2020 North America / USA / Wisconsin / Jackson County 2020-03-28 Gundersen Molecular Diagnostics Laboratory Kabara Cancer Research Institute Craig S. Richmond, Paraic A. Kenny
EPI_ISL_422488 hCoV-19/USA/NY-PV09045/2020 North America / USA / New York / Manhattan 2020-03-21 MSHS Clinical Microbiology Laboratories MSHS Pathogen Surveillance Program Ana S. Gonzalez-Reiche, Mitchell Sullivan, Ajay Obla, Gopi Patel, Emilia Sordillo, Melissa Gitman, Alberto Paniz-mondolfi, Matthew Hernandez, Shelcie Fabre, Jose Polanco, Zenab Khan, Bremy Albuquerque, Jayeeta Dutta, Juan Soto, Shwetha Sridhar Hara, Ying-Chih Wang, Melissa Smith, Robert Sebra, Lisa Miorin, Wen-chun Liu, Randy Albrecht, Judith Aberg, Florian Krammer, Adolfo Garcia-Sarstre, Viviana Simon, Harm van Bakel
EPI_ISL_422489 hCoV-19/USA/NY-PV09057/2020 North America / USA / New York / Brooklyn 2020-03-21 MSHS Clinical Microbiology Laboratories MSHS Pathogen Surveillance Program Ana S. Gonzalez-Reiche, Mitchell Sullivan, Ajay Obla, Gopi Patel, Emilia Sordillo, Melissa Gitman, Alberto Paniz-mondolfi, Matthew Hernandez, Shelcie Fabre, Jose Polanco, Zenab Khan, Bremy Albuquerque, Jayeeta Dutta, Juan Soto, Shwetha Sridhar Hara, Ying-Chih Wang, Melissa Smith, Robert Sebra, Lisa Miorin, Wen-chun Liu, Randy Albrecht, Judith Aberg, Florian Krammer, Adolfo Garcia-Sarstre, Viviana Simon, Harm van Bakel
EPI_ISL_422490 hCoV-19/USA/NY-PV09069/2020 North America / USA / New York / Brooklyn 2020-03-20 MSHS Clinical Microbiology Laboratories MSHS Pathogen Surveillance Program Ana S. Gonzalez-Reiche, Mitchell Sullivan, Ajay Obla, Gopi Patel, Emilia Sordillo, Melissa Gitman, Alberto Paniz-mondolfi, Matthew Hernandez, Shelcie Fabre, Jose Polanco, Zenab Khan, Bremy Albuquerque, Jayeeta Dutta, Juan Soto, Shwetha Sridhar Hara, Ying-Chih Wang, Melissa Smith, Robert Sebra, Lisa Miorin, Wen-chun Liu, Randy Albrecht, Judith Aberg, Florian Krammer, Adolfo Garcia-Sarstre, Viviana Simon, Harm van Bakel
EPI_ISL_422491 hCoV-19/USA/NY-PV09115/2020 North America / USA / New York / Nassau County 2020-03-19 MSHS Clinical Microbiology Laboratories MSHS Pathogen Surveillance Program Ana S. Gonzalez-Reiche, Mitchell Sullivan, Ajay Obla, Gopi Patel, Emilia Sordillo, Melissa Gitman, Alberto Paniz-mondolfi, Matthew Hernandez, Shelcie Fabre, Jose Polanco, Zenab Khan, Bremy Albuquerque, Jayeeta Dutta, Juan Soto, Shwetha Sridhar Hara, Ying-Chih Wang, Melissa Smith, Robert Sebra, Lisa Miorin, Wen-chun Liu, Randy Albrecht, Judith Aberg, Florian Krammer, Adolfo Garcia-Sarstre, Viviana Simon, Harm van Bakel
EPI_ISL_422492 hCoV-19/USA/NY-PV09116/2020 North America / USA / New York / Nassau County 2020-03-19 MSHS Clinical Microbiology Laboratories MSHS Pathogen Surveillance Program Ana S. Gonzalez-Reiche, Mitchell Sullivan, Ajay Obla, Gopi Patel, Emilia Sordillo, Melissa Gitman, Alberto Paniz-mondolfi, Matthew Hernandez, Shelcie Fabre, Jose Polanco, Zenab Khan, Bremy Albuquerque, Jayeeta Dutta, Juan Soto, Shwetha Sridhar Hara, Ying-Chih Wang, Melissa Smith, Robert Sebra, Lisa Miorin, Wen-chun Liu, Randy Albrecht, Judith Aberg, Florian Krammer, Adolfo Garcia-Sarstre, Viviana Simon, Harm van Bakel
EPI_ISL_422494 hCoV-19/USA/NY-PV09119/2020 North America / USA / New York / Manhattan 2020-03-19 MSHS Clinical Microbiology Laboratories MSHS Pathogen Surveillance Program Ana S. Gonzalez-Reiche, Mitchell Sullivan, Ajay Obla, Gopi Patel, Emilia Sordillo, Melissa Gitman, Alberto Paniz-mondolfi, Matthew Hernandez, Shelcie Fabre, Jose Polanco, Zenab Khan, Bremy Albuquerque, Jayeeta Dutta, Juan Soto, Shwetha Sridhar Hara, Ying-Chih Wang, Melissa Smith, Robert Sebra, Lisa Miorin, Wen-chun Liu, Randy Albrecht, Judith Aberg, Florian Krammer, Adolfo Garcia-Sarstre, Viviana Simon, Harm van Bakel
EPI_ISL_422495 hCoV-19/USA/NY-PV09120/2020 North America / USA / New York / Brooklyn 2020-03-19 MSHS Clinical Microbiology Laboratories MSHS Pathogen Surveillance Program Ana S. Gonzalez-Reiche, Mitchell Sullivan, Ajay Obla, Gopi Patel, Emilia Sordillo, Melissa Gitman, Alberto Paniz-mondolfi, Matthew Hernandez, Shelcie Fabre, Jose Polanco, Zenab Khan, Bremy Albuquerque, Jayeeta Dutta, Juan Soto, Shwetha Sridhar Hara, Ying-Chih Wang, Melissa Smith, Robert Sebra, Lisa Miorin, Wen-chun Liu, Randy Albrecht, Judith Aberg, Florian Krammer, Adolfo Garcia-Sarstre, Viviana Simon, Harm van Bakel
EPI_ISL_422496 hCoV-19/USA/NY-PV09121/2020 North America / USA / New York / Nassau County 2020-03-19 MSHS Clinical Microbiology Laboratories MSHS Pathogen Surveillance Program Ana S. Gonzalez-Reiche, Mitchell Sullivan, Ajay Obla, Gopi Patel, Emilia Sordillo, Melissa Gitman, Alberto Paniz-mondolfi, Matthew Hernandez, Shelcie Fabre, Jose Polanco, Zenab Khan, Bremy Albuquerque, Jayeeta Dutta, Juan Soto, Shwetha Sridhar Hara, Ying-Chih Wang, Melissa Smith, Robert Sebra, Lisa Miorin, Wen-chun Liu, Randy Albrecht, Judith Aberg, Florian Krammer, Adolfo Garcia-Sarstre, Viviana Simon, Harm van Bakel
EPI_ISL_422497 hCoV-19/USA/NY-PV09122/2020 North America / USA / New York / Nassau County 2020-03-19 MSHS Clinical Microbiology Laboratories MSHS Pathogen Surveillance Program Ana S. Gonzalez-Reiche, Mitchell Sullivan, Ajay Obla, Gopi Patel, Emilia Sordillo, Melissa Gitman, Alberto Paniz-mondolfi, Matthew Hernandez, Shelcie Fabre, Jose Polanco, Zenab Khan, Bremy Albuquerque, Jayeeta Dutta, Juan Soto, Shwetha Sridhar Hara, Ying-Chih Wang, Melissa Smith, Robert Sebra, Lisa Miorin, Wen-chun Liu, Randy Albrecht, Judith Aberg, Florian Krammer, Adolfo Garcia-Sarstre, Viviana Simon, Harm van Bakel
EPI_ISL_422498 hCoV-19/USA/NY-PV09123/2020 North America / USA / New York / Brooklyn 2020-03-19 MSHS Clinical Microbiology Laboratories MSHS Pathogen Surveillance Program Ana S. Gonzalez-Reiche, Mitchell Sullivan, Ajay Obla, Gopi Patel, Emilia Sordillo, Melissa Gitman, Alberto Paniz-mondolfi, Matthew Hernandez, Shelcie Fabre, Jose Polanco, Zenab Khan, Bremy Albuquerque, Jayeeta Dutta, Juan Soto, Shwetha Sridhar Hara, Ying-Chih Wang, Melissa Smith, Robert Sebra, Lisa Miorin, Wen-chun Liu, Randy Albrecht, Judith Aberg, Florian Krammer, Adolfo Garcia-Sarstre, Viviana Simon, Harm van Bakel

EPI_ISL_422499 hCoV-19/USA/NY-PV09125/2020 North America / USA / New York / Queens 2020-03-19 MSHS Clinical Microbiology Laboratories MSHS Pathogen Surveillance Program Ana S. Gonzalez-Reiche, Mitchell Sullivan, Ajay Obla, Gopi Patel, Emilia Sordillo, Melissa Gitman, Alberto Paniz-mondolfi, Matthew Hernandez, Shelcie Fabre, Jose Polanco, Zenab Khan, Bremy Albuquerque, Jayeeta Dutta, Juan Soto, Shwetha Sridhar Hara, Ying-Chih Wang, Melissa Smith, Robert Sebra, Lisa Miorin, Wen-chun Liu, Randy Albrecht, Judith Aberg, Florian Krammer, Adolfo Garcia-Sarstre, Viviana Simon, Harm van Bakel
EPI_ISL_422500 hCoV-19/USA/NY-PV09126/2020 North America / USA / New York / Nassau County 2020-03-19 MSHS Clinical Microbiology Laboratories MSHS Pathogen Surveillance Program Ana S. Gonzalez-Reiche, Mitchell Sullivan, Ajay Obla, Gopi Patel, Emilia Sordillo, Melissa Gitman, Alberto Paniz-mondolfi, Matthew Hernandez, Shelcie Fabre, Jose Polanco, Zenab Khan, Bremy Albuquerque, Jayeeta Dutta, Juan Soto, Shwetha Sridhar Hara, Ying-Chih Wang, Melissa Smith, Robert Sebra, Lisa Miorin, Wen-chun Liu, Randy Albrecht, Judith Aberg, Florian Krammer, Adolfo Garcia-Sarstre, Viviana Simon, Harm van Bakel
EPI_ISL_422501 hCoV-19/USA/NY-PV09127/2020 North America / USA / New York / Brooklyn 2020-03-19 MSHS Clinical Microbiology Laboratories MSHS Pathogen Surveillance Program Ana S. Gonzalez-Reiche, Mitchell Sullivan, Ajay Obla, Gopi Patel, Emilia Sordillo, Melissa Gitman, Alberto Paniz-mondolfi, Matthew Hernandez, Shelcie Fabre, Jose Polanco, Zenab Khan, Bremy Albuquerque, Jayeeta Dutta, Juan Soto, Shwetha Sridhar Hara, Ying-Chih Wang, Melissa Smith, Robert Sebra, Lisa Miorin, Wen-chun Liu, Randy Albrecht, Judith Aberg, Florian Krammer, Adolfo Garcia-Sarstre, Viviana Simon, Harm van Bakel
EPI_ISL_422502 hCoV-19/USA/NY-PV09128/2020 North America / USA / New York / Manhattan 2020-03-19 MSHS Clinical Microbiology Laboratories MSHS Pathogen Surveillance Program Ana S. Gonzalez-Reiche, Mitchell Sullivan, Ajay Obla, Gopi Patel, Emilia Sordillo, Melissa Gitman, Alberto Paniz-mondolfi, Matthew Hernandez, Shelcie Fabre, Jose Polanco, Zenab Khan, Bremy Albuquerque, Jayeeta Dutta, Juan Soto, Shwetha Sridhar Hara, Ying-Chih Wang, Melissa Smith, Robert Sebra, Lisa Miorin, Wen-chun Liu, Randy Albrecht, Judith Aberg, Florian Krammer, Adolfo Garcia-Sarstre, Viviana Simon, Harm van Bakel
EPI_ISL_422503 hCoV-19/USA/NY-PV09129/2020 North America / USA / New York / Nassau County 2020-03-19 MSHS Clinical Microbiology Laboratories MSHS Pathogen Surveillance Program Ana S. Gonzalez-Reiche, Mitchell Sullivan, Ajay Obla, Gopi Patel, Emilia Sordillo, Melissa Gitman, Alberto Paniz-mondolfi, Matthew Hernandez, Shelcie Fabre, Jose Polanco, Zenab Khan, Bremy Albuquerque, Jayeeta Dutta, Juan Soto, Shwetha Sridhar Hara, Ying-Chih Wang, Melissa Smith, Robert Sebra, Lisa Miorin, Wen-chun Liu, Randy Albrecht, Judith Aberg, Florian Krammer, Adolfo Garcia-Sarstre, Viviana Simon, Harm van Bakel
EPI_ISL_422504 hCoV-19/USA/NY-PV09130/2020 North America / USA / New York / Manhattan 2020-03-19 MSHS Clinical Microbiology Laboratories MSHS Pathogen Surveillance Program Ana S. Gonzalez-Reiche, Mitchell Sullivan, Ajay Obla, Gopi Patel, Emilia Sordillo, Melissa Gitman, Alberto Paniz-mondolfi, Matthew Hernandez, Shelcie Fabre, Jose Polanco, Zenab Khan, Bremy Albuquerque, Jayeeta Dutta, Juan Soto, Shwetha Sridhar Hara, Ying-Chih Wang, Melissa Smith, Robert Sebra, Lisa Miorin, Wen-chun Liu, Randy Albrecht, Judith Aberg, Florian Krammer, Adolfo Garcia-Sarstre, Viviana Simon, Harm van Bakel
EPI_ISL_422505 hCoV-19/USA/NY-PV09132/2020 North America / USA / New York / Manhattan 2020-03-19 MSHS Clinical Microbiology Laboratories MSHS Pathogen Surveillance Program Ana S. Gonzalez-Reiche, Mitchell Sullivan, Ajay Obla, Gopi Patel, Emilia Sordillo, Melissa Gitman, Alberto Paniz-mondolfi, Matthew Hernandez, Shelcie Fabre, Jose Polanco, Zenab Khan, Bremy Albuquerque, Jayeeta Dutta, Juan Soto, Shwetha Sridhar Hara, Ying-Chih Wang, Melissa Smith, Robert Sebra, Lisa Miorin, Wen-chun Liu, Randy Albrecht, Judith Aberg, Florian Krammer, Adolfo Garcia-Sarstre, Viviana Simon, Harm van Bakel
EPI_ISL_422506 hCoV-19/USA/NY-PV09134/2020 North America / USA / New York / Manhattan 2020-03-19 MSHS Clinical Microbiology Laboratories MSHS Pathogen Surveillance Program Ana S. Gonzalez-Reiche, Mitchell Sullivan, Ajay Obla, Gopi Patel, Emilia Sordillo, Melissa Gitman, Alberto Paniz-mondolfi, Matthew Hernandez, Shelcie Fabre, Jose Polanco, Zenab Khan, Bremy Albuquerque, Jayeeta Dutta, Juan Soto, Shwetha Sridhar Hara, Ying-Chih Wang, Melissa Smith, Robert Sebra, Lisa Miorin, Wen-chun Liu, Randy Albrecht, Judith Aberg, Florian Krammer, Adolfo Garcia-Sarstre, Viviana Simon, Harm van Bakel
EPI_ISL_422507 hCoV-19/USA/NY-PV09135/2020 North America / USA / New York / Nassau County 2020-03-20 MSHS Clinical Microbiology Laboratories MSHS Pathogen Surveillance Program Ana S. Gonzalez-Reiche, Mitchell Sullivan, Ajay Obla, Gopi Patel, Emilia Sordillo, Melissa Gitman, Alberto Paniz-mondolfi, Matthew Hernandez, Shelcie Fabre, Jose Polanco, Zenab Khan, Bremy Albuquerque, Jayeeta Dutta, Juan Soto, Shwetha Sridhar Hara, Ying-Chih Wang, Melissa Smith, Robert Sebra, Lisa Miorin, Wen-chun Liu, Randy Albrecht, Judith Aberg, Florian Krammer, Adolfo Garcia-Sarstre, Viviana Simon, Harm van Bakel
EPI_ISL_422508 hCoV-19/USA/NY-PV09137/2020 North America / USA / New York / Nassau County 2020-03-20 MSHS Clinical Microbiology Laboratories MSHS Pathogen Surveillance Program Ana S. Gonzalez-Reiche, Mitchell Sullivan, Ajay Obla, Gopi Patel, Emilia Sordillo, Melissa Gitman, Alberto Paniz-mondolfi, Matthew Hernandez, Shelcie Fabre, Jose Polanco, Zenab Khan, Bremy Albuquerque, Jayeeta Dutta, Juan Soto, Shwetha Sridhar Hara, Ying-Chih Wang, Melissa Smith, Robert Sebra, Lisa Miorin, Wen-chun Liu, Randy Albrecht, Judith Aberg, Florian Krammer, Adolfo Garcia-Sarstre, Viviana Simon, Harm van Bakel
EPI_ISL_422509 hCoV-19/USA/NY-PV09138/2020 North America / USA / New York / Manhattan 2020-03-20 MSHS Clinical Microbiology Laboratories MSHS Pathogen Surveillance Program Ana S. Gonzalez-Reiche, Mitchell Sullivan, Ajay Obla, Gopi Patel, Emilia Sordillo, Melissa Gitman, Alberto Paniz-mondolfi, Matthew Hernandez, Shelcie Fabre, Jose Polanco, Zenab Khan, Bremy Albuquerque, Jayeeta Dutta, Juan Soto, Shwetha Sridhar Hara, Ying-Chih Wang, Melissa Smith, Robert Sebra, Lisa Miorin, Wen-chun Liu, Randy Albrecht, Judith Aberg, Florian Krammer, Adolfo Garcia-Sarstre, Viviana Simon, Harm van Bakel
EPI_ISL_422510 hCoV-19/USA/NY-PV09139/2020 North America / USA / New York / Queens 2020-03-20 MSHS Clinical Microbiology Laboratories MSHS Pathogen Surveillance Program Ana S. Gonzalez-Reiche, Mitchell Sullivan, Ajay Obla, Gopi Patel, Emilia Sordillo, Melissa Gitman, Alberto Paniz-mondolfi, Matthew Hernandez, Shelcie Fabre, Jose Polanco, Zenab Khan, Bremy Albuquerque, Jayeeta Dutta, Juan Soto, Shwetha

Sridhar Hara, Ying-Chih Wang, Melissa Smith, Robert Sebra, Lisa Miorin, Wen-chun Liu, Randy Albrecht, Judith Aberg, Florian Krammer, Adolfo Garcia-Sarstre, Viviana Simon, Harm van Bakel
EPI_ISL_422511 hCoV-19/USA/NY-PV09140/2020 North America / USA / New York / Manhattan 2020-03-20 MSHS Clinical Microbiology Laboratories MSHS Pathogen Surveillance Program Ana S. Gonzalez-Reiche, Mitchell Sullivan, Ajay Obla, Gopi Patel, Emilia Sordillo, Melissa Gitman, Alberto Paniz-mondolfi, Matthew Hernandez, Shelcie Fabre, Jose Polanco, Zenab Khan, Bremy Albuquerque, Jayeeta Dutta, Juan Soto, Shwetha Sridhar Hara, Ying-Chih Wang, Melissa Smith, Robert Sebra, Lisa Miorin, Wen-chun Liu, Randy Albrecht, Judith Aberg, Florian Krammer, Adolfo Garcia-Sarstre, Viviana Simon, Harm van Bakel
EPI_ISL_422513 hCoV-19/USA/NY-PV09142/2020 North America / USA / New York / Manhattan 2020-03-20 MSHS Clinical Microbiology Laboratories MSHS Pathogen Surveillance Program Ana S. Gonzalez-Reiche, Mitchell Sullivan, Ajay Obla, Gopi Patel, Emilia Sordillo, Melissa Gitman, Alberto Paniz-mondolfi, Matthew Hernandez, Shelcie Fabre, Jose Polanco, Zenab Khan, Bremy Albuquerque, Jayeeta Dutta, Juan Soto, Shwetha Sridhar Hara, Ying-Chih Wang, Melissa Smith, Robert Sebra, Lisa Miorin, Wen-chun Liu, Randy Albrecht, Judith Aberg, Florian Krammer, Adolfo Garcia-Sarstre, Viviana Simon, Harm van Bakel
EPI_ISL_422514 hCoV-19/USA/NY-PV09143/2020 North America / USA / New York / Manhattan 2020-03-20 MSHS Clinical Microbiology Laboratories MSHS Pathogen Surveillance Program Ana S. Gonzalez-Reiche, Mitchell Sullivan, Ajay Obla, Gopi Patel, Emilia Sordillo, Melissa Gitman, Alberto Paniz-mondolfi, Matthew Hernandez, Shelcie Fabre, Jose Polanco, Zenab Khan, Bremy Albuquerque, Jayeeta Dutta, Juan Soto, Shwetha Sridhar Hara, Ying-Chih Wang, Melissa Smith, Robert Sebra, Lisa Miorin, Wen-chun Liu, Randy Albrecht, Judith Aberg, Florian Krammer, Adolfo Garcia-Sarstre, Viviana Simon, Harm van Bakel
EPI_ISL_422515 hCoV-19/USA/NY-PV09144/2020 North America / USA / New York / Manhattan 2020-03-20 MSHS Clinical Microbiology Laboratories MSHS Pathogen Surveillance Program Ana S. Gonzalez-Reiche, Mitchell Sullivan, Ajay Obla, Gopi Patel, Emilia Sordillo, Melissa Gitman, Alberto Paniz-mondolfi, Matthew Hernandez, Shelcie Fabre, Jose Polanco, Zenab Khan, Bremy Albuquerque, Jayeeta Dutta, Juan Soto, Shwetha Sridhar Hara, Ying-Chih Wang, Melissa Smith, Robert Sebra, Lisa Miorin, Wen-chun Liu, Randy Albrecht, Judith Aberg, Florian Krammer, Adolfo Garcia-Sarstre, Viviana Simon, Harm van Bakel
EPI_ISL_422516 hCoV-19/USA/NY-PV09145/2020 North America / USA / New York / Queens 2020-03-20 MSHS Clinical Microbiology Laboratories MSHS Pathogen Surveillance Program Ana S. Gonzalez-Reiche, Mitchell Sullivan, Ajay Obla, Gopi Patel, Emilia Sordillo, Melissa Gitman, Alberto Paniz-mondolfi, Matthew Hernandez, Shelcie Fabre, Jose Polanco, Zenab Khan, Bremy Albuquerque, Jayeeta Dutta, Juan Soto, Shwetha Sridhar Hara, Ying-Chih Wang, Melissa Smith, Robert Sebra, Lisa Miorin, Wen-chun Liu, Randy Albrecht, Judith Aberg, Florian Krammer, Adolfo Garcia-Sarstre, Viviana Simon, Harm van Bakel
EPI_ISL_422517 hCoV-19/USA/NY-PV09147/2020 North America / USA / New York / Manhattan 2020-03-20 MSHS Clinical Microbiology Laboratories MSHS Pathogen Surveillance Program Ana S. Gonzalez-Reiche, Mitchell Sullivan, Ajay Obla, Gopi Patel, Emilia Sordillo, Melissa Gitman, Alberto Paniz-mondolfi, Matthew Hernandez, Shelcie Fabre, Jose Polanco, Zenab Khan, Bremy Albuquerque, Jayeeta Dutta, Juan Soto, Shwetha Sridhar Hara, Ying-Chih Wang, Melissa Smith, Robert Sebra, Lisa Miorin, Wen-chun Liu, Randy Albrecht, Judith Aberg, Florian Krammer, Adolfo Garcia-Sarstre, Viviana Simon, Harm van Bakel
EPI_ISL_422519 hCoV-19/USA/NY-PV09151/2020 North America / USA / New York / Brooklyn 2020-03-22 MSHS Clinical Microbiology Laboratories MSHS Pathogen Surveillance Program Ana S. Gonzalez-Reiche, Mitchell Sullivan, Ajay Obla, Gopi Patel, Emilia Sordillo, Melissa Gitman, Alberto Paniz-mondolfi, Matthew Hernandez, Shelcie Fabre, Jose Polanco, Zenab Khan, Bremy Albuquerque, Jayeeta Dutta, Juan Soto, Shwetha Sridhar Hara, Ying-Chih Wang, Melissa Smith, Robert Sebra, Lisa Miorin, Wen-chun Liu, Randy Albrecht, Judith Aberg, Florian Krammer, Adolfo Garcia-Sarstre, Viviana Simon, Harm van Bakel
EPI_ISL_422520 hCoV-19/USA/NY-PV09152/2020 North America / USA / New York / Brooklyn 2020-03-21 MSHS Clinical Microbiology Laboratories MSHS Pathogen Surveillance Program Ana S. Gonzalez-Reiche, Mitchell Sullivan, Ajay Obla, Gopi Patel, Emilia Sordillo, Melissa Gitman, Alberto Paniz-mondolfi, Matthew Hernandez, Shelcie Fabre, Jose Polanco, Zenab Khan, Bremy Albuquerque, Jayeeta Dutta, Juan Soto, Shwetha Sridhar Hara, Ying-Chih Wang, Melissa Smith, Robert Sebra, Lisa Miorin, Wen-chun Liu, Randy Albrecht, Judith Aberg, Florian Krammer, Adolfo Garcia-Sarstre, Viviana Simon, Harm van Bakel
EPI_ISL_422521 hCoV-19/USA/NY-PV09153/2020 North America / USA / New York / Brooklyn 2020-03-22 MSHS Clinical Microbiology Laboratories MSHS Pathogen Surveillance Program Ana S. Gonzalez-Reiche, Mitchell Sullivan, Ajay Obla, Gopi Patel, Emilia Sordillo, Melissa Gitman, Alberto Paniz-mondolfi, Matthew Hernandez, Shelcie Fabre, Jose Polanco, Zenab Khan, Bremy Albuquerque, Jayeeta Dutta, Juan Soto, Shwetha Sridhar Hara, Ying-Chih Wang, Melissa Smith, Robert Sebra, Lisa Miorin, Wen-chun Liu, Randy Albrecht, Judith Aberg, Florian Krammer, Adolfo Garcia-Sarstre, Viviana Simon, Harm van Bakel
EPI_ISL_422522 hCoV-19/USA/NY-PV09154/2020 North America / USA / New York / Manhattan 2020-03-22 MSHS Clinical Microbiology Laboratories MSHS Pathogen Surveillance Program Ana S. Gonzalez-Reiche, Mitchell Sullivan, Ajay Obla, Gopi Patel, Emilia Sordillo, Melissa Gitman, Alberto Paniz-mondolfi, Matthew Hernandez, Shelcie Fabre, Jose Polanco, Zenab Khan, Bremy Albuquerque, Jayeeta Dutta, Juan Soto, Shwetha Sridhar Hara, Ying-Chih Wang, Melissa Smith, Robert Sebra, Lisa Miorin, Wen-chun Liu, Randy Albrecht, Judith Aberg, Florian Krammer, Adolfo Garcia-Sarstre, Viviana Simon, Harm van Bakel
EPI_ISL_422523 hCoV-19/USA/NY-PV09156/2020 North America / USA / New York / Bronx 2020-03-21 MSHS Clinical Microbiology Laboratories MSHS Pathogen Surveillance Program Ana S. Gonzalez-Reiche, Mitchell Sullivan, Ajay Obla, Gopi Patel, Emilia Sordillo, Melissa Gitman, Alberto Paniz-mondolfi, Matthew Hernandez, Shelcie Fabre, Jose Polanco, Zenab Khan, Bremy Albuquerque, Jayeeta Dutta, Juan Soto, Shwetha Sridhar Hara, Ying-Chih Wang, Melissa Smith, Robert Sebra, Lisa Miorin, Wen-chun Liu, Randy Albrecht, Judith Aberg, Florian Krammer, Adolfo Garcia-Sarstre, Viviana Simon, Harm van Bakel
EPI_ISL_422524 hCoV-19/USA/NY-PV09157/2020 North America / USA / New York / Manhattan 2020-03-22 MSHS Clinical Microbiology Laboratories MSHS Pathogen Surveillance Program Ana S. Gonzalez-Reiche,

Mitchell Sullivan, Ajay Obla, Gopi Patel, Emilia Sordillo, Melissa Gitman, Alberto Paniz-mondolfi, Matthew Hernandez, Shelcie Fabre, Jose Polanco, Zenab Khan, Bremy Albuquerque, Jayeeta Dutta, Juan Soto, Shwetha Sridhar Hara, Ying-Chih Wang, Melissa Smith, Robert Sebra, Lisa Miorin, Wen-chun Liu, Randy Albrecht, Judith Aberg, Florian Krammer, Adolfo Garcia-Sarstre, Viviana Simon, Harm van Bakel
EPI_ISL_422525 hCoV-19/USA/NY-PV09158/2020 North America / USA / New York / Brooklyn 2020-03-22 MSHS Clinical Microbiology Laboratories MSHS Pathogen Surveillance Program Ana S. Gonzalez-Reiche, Mitchell Sullivan, Ajay Obla, Gopi Patel, Emilia Sordillo, Melissa Gitman, Alberto Paniz-mondolfi, Matthew Hernandez, Shelcie Fabre, Jose Polanco, Zenab Khan, Bremy Albuquerque, Jayeeta Dutta, Juan Soto, Shwetha Sridhar Hara, Ying-Chih Wang, Melissa Smith, Robert Sebra, Lisa Miorin, Wen-chun Liu, Randy Albrecht, Judith Aberg, Florian Krammer, Adolfo Garcia-Sarstre, Viviana Simon, Harm van Bakel
EPI_ISL_422527 hCoV-19/USA/NY-PV09163/2020 North America / USA / New York / Brooklyn 2020-03-21 MSHS Clinical Microbiology Laboratories MSHS Pathogen Surveillance Program Ana S. Gonzalez-Reiche, Mitchell Sullivan, Ajay Obla, Gopi Patel, Emilia Sordillo, Melissa Gitman, Alberto Paniz-mondolfi, Matthew Hernandez, Shelcie Fabre, Jose Polanco, Zenab Khan, Bremy Albuquerque, Jayeeta Dutta, Juan Soto, Shwetha Sridhar Hara, Ying-Chih Wang, Melissa Smith, Robert Sebra, Lisa Miorin, Wen-chun Liu, Randy Albrecht, Judith Aberg, Florian Krammer, Adolfo Garcia-Sarstre, Viviana Simon, Harm van Bakel
EPI_ISL_422528 hCoV-19/USA/NY-PV09164/2020 North America / USA / New York / Brooklyn 2020-03-21 MSHS Clinical Microbiology Laboratories MSHS Pathogen Surveillance Program Ana S. Gonzalez-Reiche, Mitchell Sullivan, Ajay Obla, Gopi Patel, Emilia Sordillo, Melissa Gitman, Alberto Paniz-mondolfi, Matthew Hernandez, Shelcie Fabre, Jose Polanco, Zenab Khan, Bremy Albuquerque, Jayeeta Dutta, Juan Soto, Shwetha Sridhar Hara, Ying-Chih Wang, Melissa Smith, Robert Sebra, Lisa Miorin, Wen-chun Liu, Randy Albrecht, Judith Aberg, Florian Krammer, Adolfo Garcia-Sarstre, Viviana Simon, Harm van Bakel
EPI_ISL_422529 hCoV-19/USA/NY-PV09165/2020 North America / USA / New York / Brooklyn 2020-03-22 MSHS Clinical Microbiology Laboratories MSHS Pathogen Surveillance Program Ana S. Gonzalez-Reiche, Mitchell Sullivan, Ajay Obla, Gopi Patel, Emilia Sordillo, Melissa Gitman, Alberto Paniz-mondolfi, Matthew Hernandez, Shelcie Fabre, Jose Polanco, Zenab Khan, Bremy Albuquerque, Jayeeta Dutta, Juan Soto, Shwetha Sridhar Hara, Ying-Chih Wang, Melissa Smith, Robert Sebra, Lisa Miorin, Wen-chun Liu, Randy Albrecht, Judith Aberg, Florian Krammer, Adolfo Garcia-Sarstre, Viviana Simon, Harm van Bakel
EPI_ISL_422530 hCoV-19/USA/NY-PV09168/2020 North America / USA / New York / Manhattan 2020-03-22 MSHS Clinical Microbiology Laboratories MSHS Pathogen Surveillance Program Ana S. Gonzalez-Reiche, Mitchell Sullivan, Ajay Obla, Gopi Patel, Emilia Sordillo, Melissa Gitman, Alberto Paniz-mondolfi, Matthew Hernandez, Shelcie Fabre, Jose Polanco, Zenab Khan, Bremy Albuquerque, Jayeeta Dutta, Juan Soto, Shwetha Sridhar Hara, Ying-Chih Wang, Melissa Smith, Robert Sebra, Lisa Miorin, Wen-chun Liu, Randy Albrecht, Judith Aberg, Florian Krammer, Adolfo Garcia-Sarstre, Viviana Simon, Harm van Bakel
EPI_ISL_422531 hCoV-19/USA/NY-PV09169/2020 North America / USA / New York / Manhattan 2020-03-19 MSHS Clinical Microbiology Laboratories MSHS Pathogen Surveillance Program Ana S. Gonzalez-Reiche, Mitchell Sullivan, Ajay Obla, Gopi Patel, Emilia Sordillo, Melissa Gitman, Alberto Paniz-mondolfi, Matthew Hernandez, Shelcie Fabre, Jose Polanco, Zenab Khan, Bremy Albuquerque, Jayeeta Dutta, Juan Soto, Shwetha Sridhar Hara, Ying-Chih Wang, Melissa Smith, Robert Sebra, Lisa Miorin, Wen-chun Liu, Randy Albrecht, Judith Aberg, Florian Krammer, Adolfo Garcia-Sarstre, Viviana Simon, Harm van Bakel
EPI_ISL_422532 hCoV-19/USA/NY-PV09170/2020 North America / USA / New York / Brooklyn 2020-03-19 MSHS Clinical Microbiology Laboratories MSHS Pathogen Surveillance Program Ana S. Gonzalez-Reiche, Mitchell Sullivan, Ajay Obla, Gopi Patel, Emilia Sordillo, Melissa Gitman, Alberto Paniz-mondolfi, Matthew Hernandez, Shelcie Fabre, Jose Polanco, Zenab Khan, Bremy Albuquerque, Jayeeta Dutta, Juan Soto, Shwetha Sridhar Hara, Ying-Chih Wang, Melissa Smith, Robert Sebra, Lisa Miorin, Wen-chun Liu, Randy Albrecht, Judith Aberg, Florian Krammer, Adolfo Garcia-Sarstre, Viviana Simon, Harm van Bakel
EPI_ISL_422533 hCoV-19/USA/NY-PV09171/2020 North America / USA / New York / Manhattan 2020-03-19 MSHS Clinical Microbiology Laboratories MSHS Pathogen Surveillance Program Ana S. Gonzalez-Reiche, Mitchell Sullivan, Ajay Obla, Gopi Patel, Emilia Sordillo, Melissa Gitman, Alberto Paniz-mondolfi, Matthew Hernandez, Shelcie Fabre, Jose Polanco, Zenab Khan, Bremy Albuquerque, Jayeeta Dutta, Juan Soto, Shwetha Sridhar Hara, Ying-Chih Wang, Melissa Smith, Robert Sebra, Lisa Miorin, Wen-chun Liu, Randy Albrecht, Judith Aberg, Florian Krammer, Adolfo Garcia-Sarstre, Viviana Simon, Harm van Bakel
EPI_ISL_422534 hCoV-19/USA/NY-PV09172/2020 North America / USA / New York / Queens 2020-03-19 MSHS Clinical Microbiology Laboratories MSHS Pathogen Surveillance Program Ana S. Gonzalez-Reiche, Mitchell Sullivan, Ajay Obla, Gopi Patel, Emilia Sordillo, Melissa Gitman, Alberto Paniz-mondolfi, Matthew Hernandez, Shelcie Fabre, Jose Polanco, Zenab Khan, Bremy Albuquerque, Jayeeta Dutta, Juan Soto, Shwetha Sridhar Hara, Ying-Chih Wang, Melissa Smith, Robert Sebra, Lisa Miorin, Wen-chun Liu, Randy Albrecht, Judith Aberg, Florian Krammer, Adolfo Garcia-Sarstre, Viviana Simon, Harm van Bakel
EPI_ISL_422535 hCoV-19/USA/NY-PV09176/2020 North America / USA / New York / Brooklyn 2020-03-19 MSHS Clinical Microbiology Laboratories MSHS Pathogen Surveillance Program Ana S. Gonzalez-Reiche, Mitchell Sullivan, Ajay Obla, Gopi Patel, Emilia Sordillo, Melissa Gitman, Alberto Paniz-mondolfi, Matthew Hernandez, Shelcie Fabre, Jose Polanco, Zenab Khan, Bremy Albuquerque, Jayeeta Dutta, Juan Soto, Shwetha Sridhar Hara, Ying-Chih Wang, Melissa Smith, Robert Sebra, Lisa Miorin, Wen-chun Liu, Randy Albrecht, Judith Aberg, Florian Krammer, Adolfo Garcia-Sarstre, Viviana Simon, Harm van Bakel
EPI_ISL_422536 hCoV-19/USA/NY-PV09177/2020 North America / USA / New York / Brooklyn 2020-03-19 MSHS Clinical Microbiology Laboratories MSHS Pathogen Surveillance Program Ana S. Gonzalez-Reiche, Mitchell Sullivan, Ajay Obla, Gopi Patel, Emilia Sordillo, Melissa Gitman, Alberto Paniz-mondolfi, Matthew Hernandez, Shelcie Fabre, Jose Polanco, Zenab Khan, Bremy Albuquerque, Jayeeta Dutta, Juan Soto, Shwetha Sridhar Hara, Ying-Chih Wang, Melissa Smith, Robert Sebra, Lisa Miorin, Wen-chun Liu, Randy Albrecht, Judith Aberg, Florian Krammer, Adolfo Garcia-Sarstre, Viviana Simon, Harm van Bakel

EPI_ISL_422537 hCoV-19/USA/NY-PV09178/2020 North America / USA / New York / Manhattan 2020-03-19 MSHS Clinical Microbiology Laboratories MSHS Pathogen Surveillance Program Ana S. Gonzalez-Reiche, Mitchell Sullivan, Ajay Obla, Gopi Patel, Emilia Sordillo, Melissa Gitman, Alberto Paniz-mondolfi, Matthew Hernandez, Shelcie Fabre, Jose Polanco, Zenab Khan, Bremy Albuquerque, Jayeeta Dutta, Juan Soto, Shwetha Sridhar Hara, Ying-Chih Wang, Melissa Smith, Robert Sebra, Lisa Miorin, Wen-chun Liu, Randy Albrecht, Judith Aberg, Florian Krammer, Adolfo Garcia-Sarstre, Viviana Simon, Harm van Bakel
EPI_ISL_422539 hCoV-19/USA/NY-PV09180/2020 North America / USA / New York / Bronx 2020-03-19 MSHS Clinical Microbiology Laboratories MSHS Pathogen Surveillance Program Ana S. Gonzalez-Reiche, Mitchell Sullivan, Ajay Obla, Gopi Patel, Emilia Sordillo, Melissa Gitman, Alberto Paniz-mondolfi, Matthew Hernandez, Shelcie Fabre, Jose Polanco, Zenab Khan, Bremy Albuquerque, Jayeeta Dutta, Juan Soto, Shwetha Sridhar Hara, Ying-Chih Wang, Melissa Smith, Robert Sebra, Lisa Miorin, Wen-chun Liu, Randy Albrecht, Judith Aberg, Florian Krammer, Adolfo Garcia-Sarstre, Viviana Simon, Harm van Bakel
EPI_ISL_422540 hCoV-19/USA/NY-PV09181/2020 North America / USA / New York / Manhattan 2020-03-19 MSHS Clinical Microbiology Laboratories MSHS Pathogen Surveillance Program Ana S. Gonzalez-Reiche, Mitchell Sullivan, Ajay Obla, Gopi Patel, Emilia Sordillo, Melissa Gitman, Alberto Paniz-mondolfi, Matthew Hernandez, Shelcie Fabre, Jose Polanco, Zenab Khan, Bremy Albuquerque, Jayeeta Dutta, Juan Soto, Shwetha Sridhar Hara, Ying-Chih Wang, Melissa Smith, Robert Sebra, Lisa Miorin, Wen-chun Liu, Randy Albrecht, Judith Aberg, Florian Krammer, Adolfo Garcia-Sarstre, Viviana Simon, Harm van Bakel
EPI_ISL_422541 hCoV-19/USA/NY-PV09182/2020 North America / USA / New York / Manhattan 2020-03-19 MSHS Clinical Microbiology Laboratories MSHS Pathogen Surveillance Program Ana S. Gonzalez-Reiche, Mitchell Sullivan, Ajay Obla, Gopi Patel, Emilia Sordillo, Melissa Gitman, Alberto Paniz-mondolfi, Matthew Hernandez, Shelcie Fabre, Jose Polanco, Zenab Khan, Bremy Albuquerque, Jayeeta Dutta, Juan Soto, Shwetha Sridhar Hara, Ying-Chih Wang, Melissa Smith, Robert Sebra, Lisa Miorin, Wen-chun Liu, Randy Albrecht, Judith Aberg, Florian Krammer, Adolfo Garcia-Sarstre, Viviana Simon, Harm van Bakel
EPI_ISL_422542 hCoV-19/USA/NY-PV09183/2020 North America / USA / New York / Bronx 2020-03-19 MSHS Clinical Microbiology Laboratories MSHS Pathogen Surveillance Program Ana S. Gonzalez-Reiche, Mitchell Sullivan, Ajay Obla, Gopi Patel, Emilia Sordillo, Melissa Gitman, Alberto Paniz-mondolfi, Matthew Hernandez, Shelcie Fabre, Jose Polanco, Zenab Khan, Bremy Albuquerque, Jayeeta Dutta, Juan Soto, Shwetha Sridhar Hara, Ying-Chih Wang, Melissa Smith, Robert Sebra, Lisa Miorin, Wen-chun Liu, Randy Albrecht, Judith Aberg, Florian Krammer, Adolfo Garcia-Sarstre, Viviana Simon, Harm van Bakel
EPI_ISL_422543 hCoV-19/USA/NY-PV09186/2020 North America / USA / New York / Bronx 2020-03-20 MSHS Clinical Microbiology Laboratories MSHS Pathogen Surveillance Program Ana S. Gonzalez-Reiche, Mitchell Sullivan, Ajay Obla, Gopi Patel, Emilia Sordillo, Melissa Gitman, Alberto Paniz-mondolfi, Matthew Hernandez, Shelcie Fabre, Jose Polanco, Zenab Khan, Bremy Albuquerque, Jayeeta Dutta, Juan Soto, Shwetha Sridhar Hara, Ying-Chih Wang, Melissa Smith, Robert Sebra, Lisa Miorin, Wen-chun Liu, Randy Albrecht, Judith Aberg, Florian Krammer, Adolfo Garcia-Sarstre, Viviana Simon, Harm van Bakel
EPI_ISL_422544 hCoV-19/USA/NY-PV09187/2020 North America / USA / New York / Westchester County 2020-03-20 MSHS Clinical Microbiology Laboratories MSHS Pathogen Surveillance Program Ana S. Gonzalez-Reiche, Mitchell Sullivan, Ajay Obla, Gopi Patel, Emilia Sordillo, Melissa Gitman, Alberto Paniz-mondolfi, Matthew Hernandez, Shelcie Fabre, Jose Polanco, Zenab Khan, Bremy Albuquerque, Jayeeta Dutta, Juan Soto, Shwetha Sridhar Hara, Ying-Chih Wang, Melissa Smith, Robert Sebra, Lisa Miorin, Wen-chun Liu, Randy Albrecht, Judith Aberg, Florian Krammer, Adolfo Garcia-Sarstre, Viviana Simon, Harm van Bakel
EPI_ISL_422546 hCoV-19/USA/NY-PV09189/2020 North America / USA / New York / Queens 2020-03-19 MSHS Clinical Microbiology Laboratories MSHS Pathogen Surveillance Program Ana S. Gonzalez-Reiche, Mitchell Sullivan, Ajay Obla, Gopi Patel, Emilia Sordillo, Melissa Gitman, Alberto Paniz-mondolfi, Matthew Hernandez, Shelcie Fabre, Jose Polanco, Zenab Khan, Bremy Albuquerque, Jayeeta Dutta, Juan Soto, Shwetha Sridhar Hara, Ying-Chih Wang, Melissa Smith, Robert Sebra, Lisa Miorin, Wen-chun Liu, Randy Albrecht, Judith Aberg, Florian Krammer, Adolfo Garcia-Sarstre, Viviana Simon, Harm van Bakel
EPI_ISL_422547 hCoV-19/USA/NY-PV09190/2020 North America / USA / New York / Manhattan 2020-03-20 MSHS Clinical Microbiology Laboratories MSHS Pathogen Surveillance Program Ana S. Gonzalez-Reiche, Mitchell Sullivan, Ajay Obla, Gopi Patel, Emilia Sordillo, Melissa Gitman, Alberto Paniz-mondolfi, Matthew Hernandez, Shelcie Fabre, Jose Polanco, Zenab Khan, Bremy Albuquerque, Jayeeta Dutta, Juan Soto, Shwetha Sridhar Hara, Ying-Chih Wang, Melissa Smith, Robert Sebra, Lisa Miorin, Wen-chun Liu, Randy Albrecht, Judith Aberg, Florian Krammer, Adolfo Garcia-Sarstre, Viviana Simon, Harm van Bakel
EPI_ISL_422548 hCoV-19/USA/NY-PV09192/2020 North America / USA / New York / Bronx 2020-03-20 MSHS Clinical Microbiology Laboratories MSHS Pathogen Surveillance Program Ana S. Gonzalez-Reiche, Mitchell Sullivan, Ajay Obla, Gopi Patel, Emilia Sordillo, Melissa Gitman, Alberto Paniz-mondolfi, Matthew Hernandez, Shelcie Fabre, Jose Polanco, Zenab Khan, Bremy Albuquerque, Jayeeta Dutta, Juan Soto, Shwetha Sridhar Hara, Ying-Chih Wang, Melissa Smith, Robert Sebra, Lisa Miorin, Wen-chun Liu, Randy Albrecht, Judith Aberg, Florian Krammer, Adolfo Garcia-Sarstre, Viviana Simon, Harm van Bakel
EPI_ISL_422549 hCoV-19/USA/NY-PV09193/2020 North America / USA / New York / Manhattan 2020-03-20 MSHS Clinical Microbiology Laboratories MSHS Pathogen Surveillance Program Ana S. Gonzalez-Reiche, Mitchell Sullivan, Ajay Obla, Gopi Patel, Emilia Sordillo, Melissa Gitman, Alberto Paniz-mondolfi, Matthew Hernandez, Shelcie Fabre, Jose Polanco, Zenab Khan, Bremy Albuquerque, Jayeeta Dutta, Juan Soto, Shwetha Sridhar Hara, Ying-Chih Wang, Melissa Smith, Robert Sebra, Lisa Miorin, Wen-chun Liu, Randy Albrecht, Judith Aberg, Florian Krammer, Adolfo Garcia-Sarstre, Viviana Simon, Harm van Bakel
EPI_ISL_422550 hCoV-19/USA/NY-PV09194/2020 North America / USA / New York / Bronx 2020-03-20 MSHS Clinical Microbiology Laboratories MSHS Pathogen Surveillance Program Ana S. Gonzalez-Reiche, Mitchell Sullivan, Ajay Obla, Gopi Patel, Emilia Sordillo, Melissa Gitman, Alberto Paniz-mondolfi, Matthew Hernandez, Shelcie Fabre, Jose Polanco, Zenab Khan, Bremy Albuquerque, Jayeeta Dutta, Juan Soto, Shwetha

Sridhar Hara, Ying-Chih Wang, Melissa Smith, Robert Sebra, Lisa Miorin, Wen-chun Liu, Randy Albrecht, Judith Aberg, Florian Krammer, Adolfo Garcia-Sarstre, Viviana Simon, Harm van Bakel
EPI_ISL_422551 hCoV-19/USA/NY-PV09195/2020 North America / USA / New York / Bronx 2020-03-20 MSHS Clinical Microbiology Laboratories MSHS Pathogen Surveillance Program Ana S. Gonzalez-Reiche, Mitchell Sullivan, Ajay Obla, Gopi Patel, Emilia Sordillo, Melissa Gitman, Alberto Paniz-mondolfi, Matthew Hernandez, Shelcie Fabre, Jose Polanco, Zenab Khan, Bremy Albuquerque, Jayeeta Dutta, Juan Soto, Shwetha Sridhar Hara, Ying-Chih Wang, Melissa Smith, Robert Sebra, Lisa Miorin, Wen-chun Liu, Randy Albrecht, Judith Aberg, Florian Krammer, Adolfo Garcia-Sarstre, Viviana Simon, Harm van Bakel
EPI_ISL_422552 hCoV-19/USA/NY-PV09197/2020 North America / USA / New York / Manhattan 2020-03-20 MSHS Clinical Microbiology Laboratories MSHS Pathogen Surveillance Program Ana S. Gonzalez-Reiche, Mitchell Sullivan, Ajay Obla, Gopi Patel, Emilia Sordillo, Melissa Gitman, Alberto Paniz-mondolfi, Matthew Hernandez, Shelcie Fabre, Jose Polanco, Zenab Khan, Bremy Albuquerque, Jayeeta Dutta, Juan Soto, Shwetha Sridhar Hara, Ying-Chih Wang, Melissa Smith, Robert Sebra, Lisa Miorin, Wen-chun Liu, Randy Albrecht, Judith Aberg, Florian Krammer, Adolfo Garcia-Sarstre, Viviana Simon, Harm van Bakel
EPI_ISL_422553 hCoV-19/USA/NY-PV09198/2020 North America / USA / New York / Manhattan 2020-03-20 MSHS Clinical Microbiology Laboratories MSHS Pathogen Surveillance Program Ana S. Gonzalez-Reiche, Mitchell Sullivan, Ajay Obla, Gopi Patel, Emilia Sordillo, Melissa Gitman, Alberto Paniz-mondolfi, Matthew Hernandez, Shelcie Fabre, Jose Polanco, Zenab Khan, Bremy Albuquerque, Jayeeta Dutta, Juan Soto, Shwetha Sridhar Hara, Ying-Chih Wang, Melissa Smith, Robert Sebra, Lisa Miorin, Wen-chun Liu, Randy Albrecht, Judith Aberg, Florian Krammer, Adolfo Garcia-Sarstre, Viviana Simon, Harm van Bakel
EPI_ISL_422554 hCoV-19/USA/NY-PV09199/2020 North America / USA / New York / Brooklyn 2020-03-21 MSHS Clinical Microbiology Laboratories MSHS Pathogen Surveillance Program Ana S. Gonzalez-Reiche, Mitchell Sullivan, Ajay Obla, Gopi Patel, Emilia Sordillo, Melissa Gitman, Alberto Paniz-mondolfi, Matthew Hernandez, Shelcie Fabre, Jose Polanco, Zenab Khan, Bremy Albuquerque, Jayeeta Dutta, Juan Soto, Shwetha Sridhar Hara, Ying-Chih Wang, Melissa Smith, Robert Sebra, Lisa Miorin, Wen-chun Liu, Randy Albrecht, Judith Aberg, Florian Krammer, Adolfo Garcia-Sarstre, Viviana Simon, Harm van Bakel
EPI_ISL_422555 hCoV-19/USA/NY-PV09200/2020 North America / USA / New York / Manhattan 2020-03-22 MSHS Clinical Microbiology Laboratories MSHS Pathogen Surveillance Program Ana S. Gonzalez-Reiche, Mitchell Sullivan, Ajay Obla, Gopi Patel, Emilia Sordillo, Melissa Gitman, Alberto Paniz-mondolfi, Matthew Hernandez, Shelcie Fabre, Jose Polanco, Zenab Khan, Bremy Albuquerque, Jayeeta Dutta, Juan Soto, Shwetha Sridhar Hara, Ying-Chih Wang, Melissa Smith, Robert Sebra, Lisa Miorin, Wen-chun Liu, Randy Albrecht, Judith Aberg, Florian Krammer, Adolfo Garcia-Sarstre, Viviana Simon, Harm van Bakel
EPI_ISL_422600 hCoV-19/Netherlands/NA_291/2020 Europe / Netherlands 2020-04-01 Dutch COVID-19 response team Erasmus Medical Center Bas Oude Munnink, David Nieuwenhuijse, Reina Sikkema, Claudia Schapendonk, Irina Chestakova, Anne van der Linden, Theo Bestebroer, Stefan van Nieuwkoop, Mark Pronk, Pascal Lexmond, Corien Swaan, Manon Haverkate, Madelief Mollers, Mart Stein, Sandra Kengne Kamga Mobou, Jeroen van Kampen, Jolanda Voermans, Aura Timen, Corine GeurtsvanKessel, Annemiek van der Eijk, Richard Molenkamp, Marion Koopmans, on behalf of the Dutch national COVID-19 response team.
EPI_ISL_422601 hCoV-19/Netherlands/NA_292/2020 Europe / Netherlands 2020-04-01 Dutch COVID-19 response team Erasmus Medical Center Bas Oude Munnink, David Nieuwenhuijse, Reina Sikkema, Claudia Schapendonk, Irina Chestakova, Anne van der Linden, Theo Bestebroer, Stefan van Nieuwkoop, Mark Pronk, Pascal Lexmond, Corien Swaan, Manon Haverkate, Madelief Mollers, Mart Stein, Sandra Kengne Kamga Mobou, Jeroen van Kampen, Jolanda Voermans, Aura Timen, Corine GeurtsvanKessel, Annemiek van der Eijk, Richard Molenkamp, Marion Koopmans, on behalf of the Dutch national COVID-19 response team.
EPI_ISL_422602 hCoV-19/Netherlands/NA_293/2020 Europe / Netherlands 2020-04-01 Dutch COVID-19 response team Erasmus Medical Center Bas Oude Munnink, David Nieuwenhuijse, Reina Sikkema, Claudia Schapendonk, Irina Chestakova, Anne van der Linden, Theo Bestebroer, Stefan van Nieuwkoop, Mark Pronk, Pascal Lexmond, Corien Swaan, Manon Haverkate, Madelief Mollers, Mart Stein, Sandra Kengne Kamga Mobou, Jeroen van Kampen, Jolanda Voermans, Aura Timen, Corine GeurtsvanKessel, Annemiek van der Eijk, Richard Molenkamp, Marion Koopmans, on behalf of the Dutch national COVID-19 response team.
EPI_ISL_422603 hCoV-19/Netherlands/NA_294/2020 Europe / Netherlands 2020-04-01 Dutch COVID-19 response team Erasmus Medical Center Bas Oude Munnink, David Nieuwenhuijse, Reina Sikkema, Claudia Schapendonk, Irina Chestakova, Anne van der Linden, Theo Bestebroer, Stefan van Nieuwkoop, Mark Pronk, Pascal Lexmond, Corien Swaan, Manon Haverkate, Madelief Mollers, Mart Stein, Sandra Kengne Kamga Mobou, Jeroen van Kampen, Jolanda Voermans, Aura Timen, Corine GeurtsvanKessel, Annemiek van der Eijk, Richard Molenkamp, Marion Koopmans, on behalf of the Dutch national COVID-19 response team.
EPI_ISL_422604 hCoV-19/Netherlands/NA_295/2020 Europe / Netherlands 2020-04-01 Dutch COVID-19 response team Erasmus Medical Center Bas Oude Munnink, David Nieuwenhuijse, Reina Sikkema, Claudia Schapendonk, Irina Chestakova, Anne van der Linden, Theo Bestebroer, Stefan van Nieuwkoop, Mark Pronk, Pascal Lexmond, Corien Swaan, Manon Haverkate, Madelief Mollers, Mart Stein, Sandra Kengne Kamga Mobou, Jeroen van Kampen, Jolanda Voermans, Aura Timen, Corine GeurtsvanKessel, Annemiek van der Eijk, Richard Molenkamp, Marion Koopmans, on behalf of the Dutch national COVID-19 response team.
EPI_ISL_422605 hCoV-19/Netherlands/NA_296/2020 Europe / Netherlands 2020-04-01 Dutch COVID-19 response team Erasmus Medical Center Bas Oude Munnink, David Nieuwenhuijse, Reina Sikkema, Claudia Schapendonk, Irina Chestakova, Anne van der Linden, Theo Bestebroer, Stefan van Nieuwkoop, Mark Pronk, Pascal Lexmond, Corien Swaan, Manon Haverkate, Madelief Mollers, Mart Stein, Sandra Kengne Kamga Mobou, Jeroen van Kampen, Jolanda Voermans, Aura Timen, Corine GeurtsvanKessel, Annemiek van der Eijk, Richard Molenkamp, Marion Koopmans, on behalf of the Dutch national COVID-19 response team.
EPI_ISL_422606 hCoV-19/Netherlands/NA_297/2020 Europe / Netherlands 2020-04-01 Dutch COVID-19 response team Erasmus Medical Center Bas Oude Munnink, David Nieuwenhuijse, Reina Sikkema, Claudia Schapendonk,

Irina Chestakova, Anne van der Linden, Theo Bestebroer, Stefan van Nieuwkoop, Mark Pronk, Pascal Lexmond, Corien Swaan, Manon Haverkate, Madelief Mollers, Mart Stein, Sandra Kengne Kamga Mobou, Jeroen van Kampen, Jolanda Voermans, Aura Timen, Corine GeurtsvanKessel, Annemiek van der Eijk, Richard Molenkamp, Marion Koopmans, on behalf of the Dutch national COVID-19 response team.
EPI_ISL_422607 hCoV-19/Netherlands/NA_298/2020 Europe / Netherlands 2020-04-01 Dutch COVID-19 response team Erasmus Medical Center Bas Oude Munnink, David Nieuwenhuijse, Reina Sikkema, Claudia Schapendonk, Irina Chestakova, Anne van der Linden, Theo Bestebroer, Stefan van Nieuwkoop, Mark Pronk, Pascal Lexmond, Corien Swaan, Manon Haverkate, Madelief Mollers, Mart Stein, Sandra Kengne Kamga Mobou, Jeroen van Kampen, Jolanda Voermans, Aura Timen, Corine GeurtsvanKessel, Annemiek van der Eijk, Richard Molenkamp, Marion Koopmans, on behalf of the Dutch national COVID-19 response team.
EPI_ISL_422608 hCoV-19/Netherlands/NA_299/2020 Europe / Netherlands 2020-04-01 Dutch COVID-19 response team Erasmus Medical Center Bas Oude Munnink, David Nieuwenhuijse, Reina Sikkema, Claudia Schapendonk, Irina Chestakova, Anne van der Linden, Theo Bestebroer, Stefan van Nieuwkoop, Mark Pronk, Pascal Lexmond, Corien Swaan, Manon Haverkate, Madelief Mollers, Mart Stein, Sandra Kengne Kamga Mobou, Jeroen van Kampen, Jolanda Voermans, Aura Timen, Corine GeurtsvanKessel, Annemiek van der Eijk, Richard Molenkamp, Marion Koopmans, on behalf of the Dutch national COVID-19 response team.
EPI_ISL_422609 hCoV-19/Netherlands/NA_300/2020 Europe / Netherlands 2020-04-01 Dutch COVID-19 response team Erasmus Medical Center Bas Oude Munnink, David Nieuwenhuijse, Reina Sikkema, Claudia Schapendonk, Irina Chestakova, Anne van der Linden, Theo Bestebroer, Stefan van Nieuwkoop, Mark Pronk, Pascal Lexmond, Corien Swaan, Manon Haverkate, Madelief Mollers, Mart Stein, Sandra Kengne Kamga Mobou, Jeroen van Kampen, Jolanda Voermans, Aura Timen, Corine GeurtsvanKessel, Annemiek van der Eijk, Richard Molenkamp, Marion Koopmans, on behalf of the Dutch national COVID-19 response team.
EPI_ISL_422610 hCoV-19/Netherlands/NA_301/2020 Europe / Netherlands 2020-04-01 Dutch COVID-19 response team Erasmus Medical Center Bas Oude Munnink, David Nieuwenhuijse, Reina Sikkema, Claudia Schapendonk, Irina Chestakova, Anne van der Linden, Theo Bestebroer, Stefan van Nieuwkoop, Mark Pronk, Pascal Lexmond, Corien Swaan, Manon Haverkate, Madelief Mollers, Mart Stein, Sandra Kengne Kamga Mobou, Jeroen van Kampen, Jolanda Voermans, Aura Timen, Corine GeurtsvanKessel, Annemiek van der Eijk, Richard Molenkamp, Marion Koopmans, on behalf of the Dutch national COVID-19 response team.
EPI_ISL_422611 hCoV-19/Netherlands/NA_302/2020 Europe / Netherlands 2020-04-01 Dutch COVID-19 response team Erasmus Medical Center Bas Oude Munnink, David Nieuwenhuijse, Reina Sikkema, Claudia Schapendonk, Irina Chestakova, Anne van der Linden, Theo Bestebroer, Stefan van Nieuwkoop, Mark Pronk, Pascal Lexmond, Corien Swaan, Manon Haverkate, Madelief Mollers, Mart Stein, Sandra Kengne Kamga Mobou, Jeroen van Kampen, Jolanda Voermans, Aura Timen, Corine GeurtsvanKessel, Annemiek van der Eijk, Richard Molenkamp, Marion Koopmans, on behalf of the Dutch national COVID-19 response team.
EPI_ISL_422612 hCoV-19/Netherlands/NA_303/2020 Europe / Netherlands 2020-04-01 Dutch COVID-19 response team Erasmus Medical Center Bas Oude Munnink, David Nieuwenhuijse, Reina Sikkema, Claudia Schapendonk, Irina Chestakova, Anne van der Linden, Theo Bestebroer, Stefan van Nieuwkoop, Mark Pronk, Pascal Lexmond, Corien Swaan, Manon Haverkate, Madelief Mollers, Mart Stein, Sandra Kengne Kamga Mobou, Jeroen van Kampen, Jolanda Voermans, Aura Timen, Corine GeurtsvanKessel, Annemiek van der Eijk, Richard Molenkamp, Marion Koopmans, on behalf of the Dutch national COVID-19 response team.
EPI_ISL_422615 hCoV-19/Netherlands/NA_306/2020 Europe / Netherlands 2020-04-01 Dutch COVID-19 response team Erasmus Medical Center Bas Oude Munnink, David Nieuwenhuijse, Reina Sikkema, Claudia Schapendonk, Irina Chestakova, Anne van der Linden, Theo Bestebroer, Stefan van Nieuwkoop, Mark Pronk, Pascal Lexmond, Corien Swaan, Manon Haverkate, Madelief Mollers, Mart Stein, Sandra Kengne Kamga Mobou, Jeroen van Kampen, Jolanda Voermans, Aura Timen, Corine GeurtsvanKessel, Annemiek van der Eijk, Richard Molenkamp, Marion Koopmans, on behalf of the Dutch national COVID-19 response team.
EPI_ISL_422616 hCoV-19/Netherlands/NA_307/2020 Europe / Netherlands 2020-04-02 Dutch COVID-19 response team Erasmus Medical Center Bas Oude Munnink, David Nieuwenhuijse, Reina Sikkema, Claudia Schapendonk, Irina Chestakova, Anne van der Linden, Theo Bestebroer, Stefan van Nieuwkoop, Mark Pronk, Pascal Lexmond, Corien Swaan, Manon Haverkate, Madelief Mollers, Mart Stein, Sandra Kengne Kamga Mobou, Jeroen van Kampen, Jolanda Voermans, Aura Timen, Corine GeurtsvanKessel, Annemiek van der Eijk, Richard Molenkamp, Marion Koopmans, on behalf of the Dutch national COVID-19 response team.
EPI_ISL_422619 hCoV-19/Netherlands/NA_310/2020 Europe / Netherlands 2020-04-02 Dutch COVID-19 response team Erasmus Medical Center Bas Oude Munnink, David Nieuwenhuijse, Reina Sikkema, Claudia Schapendonk, Irina Chestakova, Anne van der Linden, Theo Bestebroer, Stefan van Nieuwkoop, Mark Pronk, Pascal Lexmond, Corien Swaan, Manon Haverkate, Madelief Mollers, Mart Stein, Sandra Kengne Kamga Mobou, Jeroen van Kampen, Jolanda Voermans, Aura Timen, Corine GeurtsvanKessel, Annemiek van der Eijk, Richard Molenkamp, Marion Koopmans, on behalf of the Dutch national COVID-19 response team.
EPI_ISL_422620 hCoV-19/Netherlands/NA_311/2020 Europe / Netherlands 2020-04-02 Dutch COVID-19 response team Erasmus Medical Center Bas Oude Munnink, David Nieuwenhuijse, Reina Sikkema, Claudia Schapendonk, Irina Chestakova, Anne van der Linden, Theo Bestebroer, Stefan van Nieuwkoop, Mark Pronk, Pascal Lexmond, Corien Swaan, Manon Haverkate, Madelief Mollers, Mart Stein, Sandra Kengne Kamga Mobou, Jeroen van Kampen, Jolanda Voermans, Aura Timen, Corine GeurtsvanKessel, Annemiek van der Eijk, Richard Molenkamp, Marion Koopmans, on behalf of the Dutch national COVID-19 response team.
EPI_ISL_422621 hCoV-19/Netherlands/NA_312/2020 Europe / Netherlands 2020-04-02 Dutch COVID-19 response team Erasmus Medical Center Bas Oude Munnink, David Nieuwenhuijse, Reina Sikkema, Claudia Schapendonk, Irina Chestakova, Anne van der Linden, Theo Bestebroer, Stefan van Nieuwkoop, Mark Pronk, Pascal Lexmond, Corien Swaan, Manon Haverkate, Madelief Mollers, Mart Stein, Sandra Kengne Kamga Mobou, Jeroen van Kampen, Jolanda Voermans, Aura Timen, Corine GeurtsvanKessel, Annemiek van der Eijk, Richard Molenkamp, Marion Koopmans, on behalf of the Dutch national COVID-19 response team.

EPI_ISL_422622 hCoV-19/Netherlands/NA_313/2020 Europe / Netherlands 2020-04-02 Dutch COVID-19 response team Erasmus Medical Center Bas Oude Munnink, David Nieuwenhuijse, Reina Sikkema, Claudia Schapendonk, Irina Chestakova, Anne van der Linden, Theo Bestebroer, Stefan van Nieuwkoop, Mark Pronk, Pascal Lexmond, Corien Swaan, Manon Haverkate, Madelief Mollers, Mart Stein, Sandra Kengne Kamga Mobou, Jeroen van Kampen, Jolanda Voermans, Aura Timen, Corine GeurtsvanKessel, Annemiek van der Eijk, Richard Molenkamp, Marion Koopmans, on behalf of the Dutch national COVID-19 response team.
EPI_ISL_422623 hCoV-19/Netherlands/NA_314/2020 Europe / Netherlands 2020-04-02 Dutch COVID-19 response team Erasmus Medical Center Bas Oude Munnink, David Nieuwenhuijse, Reina Sikkema, Claudia Schapendonk, Irina Chestakova, Anne van der Linden, Theo Bestebroer, Stefan van Nieuwkoop, Mark Pronk, Pascal Lexmond, Corien Swaan, Manon Haverkate, Madelief Mollers, Mart Stein, Sandra Kengne Kamga Mobou, Jeroen van Kampen, Jolanda Voermans, Aura Timen, Corine GeurtsvanKessel, Annemiek van der Eijk, Richard Molenkamp, Marion Koopmans, on behalf of the Dutch national COVID-19 response team.
EPI_ISL_422625 hCoV-19/Netherlands/NA_316/2020 Europe / Netherlands 2020-04-02 Dutch COVID-19 response team Erasmus Medical Center Bas Oude Munnink, David Nieuwenhuijse, Reina Sikkema, Claudia Schapendonk, Irina Chestakova, Anne van der Linden, Theo Bestebroer, Stefan van Nieuwkoop, Mark Pronk, Pascal Lexmond, Corien Swaan, Manon Haverkate, Madelief Mollers, Mart Stein, Sandra Kengne Kamga Mobou, Jeroen van Kampen, Jolanda Voermans, Aura Timen, Corine GeurtsvanKessel, Annemiek van der Eijk, Richard Molenkamp, Marion Koopmans, on behalf of the Dutch national COVID-19 response team.
EPI_ISL_422626 hCoV-19/Netherlands/NA_317/2020 Europe / Netherlands 2020-04-02 Dutch COVID-19 response team Erasmus Medical Center Bas Oude Munnink, David Nieuwenhuijse, Reina Sikkema, Claudia Schapendonk, Irina Chestakova, Anne van der Linden, Theo Bestebroer, Stefan van Nieuwkoop, Mark Pronk, Pascal Lexmond, Corien Swaan, Manon Haverkate, Madelief Mollers, Mart Stein, Sandra Kengne Kamga Mobou, Jeroen van Kampen, Jolanda Voermans, Aura Timen, Corine GeurtsvanKessel, Annemiek van der Eijk, Richard Molenkamp, Marion Koopmans, on behalf of the Dutch national COVID-19 response team.
EPI_ISL_422628 hCoV-19/Netherlands/ZuidHolland_100/2020 Europe / Netherlands / Zuid Holland 2020-03-31 Dutch COVID-19 response team Erasmus Medical Center Bas Oude Munnink, David Nieuwenhuijse, Reina Sikkema, Claudia Schapendonk, Irina Chestakova, Anne van der Linden, Theo Bestebroer, Stefan van Nieuwkoop, Mark Pronk, Pascal Lexmond, Corien Swaan, Manon Haverkate, Madelief Mollers, Mart Stein, Sandra Kengne Kamga Mobou, Jeroen van Kampen, Jolanda Voermans, Aura Timen, Corine GeurtsvanKessel, Annemiek van der Eijk, Richard Molenkamp, Marion Koopmans, on behalf of the Dutch national COVID-19 response team.
EPI_ISL_422629 hCoV-19/Netherlands/ZuidHolland_101/2020 Europe / Netherlands / Zuid Holland 2020-03-31 Dutch COVID-19 response team Erasmus Medical Center Bas Oude Munnink, David Nieuwenhuijse, Reina Sikkema, Claudia Schapendonk, Irina Chestakova, Anne van der Linden, Theo Bestebroer, Stefan van Nieuwkoop, Mark Pronk, Pascal Lexmond, Corien Swaan, Manon Haverkate, Madelief Mollers, Mart Stein, Sandra Kengne Kamga Mobou, Jeroen van Kampen, Jolanda Voermans, Aura Timen, Corine GeurtsvanKessel, Annemiek van der Eijk, Richard Molenkamp, Marion Koopmans, on behalf of the Dutch national COVID-19 response team.
EPI_ISL_422630 hCoV-19/Netherlands/ZuidHolland_94/2020 Europe / Netherlands / Zuid Holland 2020-03-31 Dutch COVID-19 response team Erasmus Medical Center Bas Oude Munnink, David Nieuwenhuijse, Reina Sikkema, Claudia Schapendonk, Irina Chestakova, Anne van der Linden, Theo Bestebroer, Stefan van Nieuwkoop, Mark Pronk, Pascal Lexmond, Corien Swaan, Manon Haverkate, Madelief Mollers, Mart Stein, Sandra Kengne Kamga Mobou, Jeroen van Kampen, Jolanda Voermans, Aura Timen, Corine GeurtsvanKessel, Annemiek van der Eijk, Richard Molenkamp, Marion Koopmans, on behalf of the Dutch national COVID-19 response team.
EPI_ISL_422631 hCoV-19/Netherlands/ZuidHolland_95/2020 Europe / Netherlands / Zuid Holland 2020-03-31 Dutch COVID-19 response team Erasmus Medical Center Bas Oude Munnink, David Nieuwenhuijse, Reina Sikkema, Claudia Schapendonk, Irina Chestakova, Anne van der Linden, Theo Bestebroer, Stefan van Nieuwkoop, Mark Pronk, Pascal Lexmond, Corien Swaan, Manon Haverkate, Madelief Mollers, Mart Stein, Sandra Kengne Kamga Mobou, Jeroen van Kampen, Jolanda Voermans, Aura Timen, Corine GeurtsvanKessel, Annemiek van der Eijk, Richard Molenkamp, Marion Koopmans, on behalf of the Dutch national COVID-19 response team.
EPI_ISL_422632 hCoV-19/Netherlands/ZuidHolland_96/2020 Europe / Netherlands / Zuid Holland 2020-03-30 Dutch COVID-19 response team Erasmus Medical Center Bas Oude Munnink, David Nieuwenhuijse, Reina Sikkema, Claudia Schapendonk, Irina Chestakova, Anne van der Linden, Theo Bestebroer, Stefan van Nieuwkoop, Mark Pronk, Pascal Lexmond, Corien Swaan, Manon Haverkate, Madelief Mollers, Mart Stein, Sandra Kengne Kamga Mobou, Jeroen van Kampen, Jolanda Voermans, Aura Timen, Corine GeurtsvanKessel, Annemiek van der Eijk, Richard Molenkamp, Marion Koopmans, on behalf of the Dutch national COVID-19 response team.
EPI_ISL_422633 hCoV-19/Netherlands/ZuidHolland_97/2020 Europe / Netherlands / Zuid Holland 2020-03-31 Dutch COVID-19 response team Erasmus Medical Center Bas Oude Munnink, David Nieuwenhuijse, Reina Sikkema, Claudia Schapendonk, Irina Chestakova, Anne van der Linden, Theo Bestebroer, Stefan van Nieuwkoop, Mark Pronk, Pascal Lexmond, Corien Swaan, Manon Haverkate, Madelief Mollers, Mart Stein, Sandra Kengne Kamga Mobou, Jeroen van Kampen, Jolanda Voermans, Aura Timen, Corine GeurtsvanKessel, Annemiek van der Eijk, Richard Molenkamp, Marion Koopmans, on behalf of the Dutch national COVID-19 response team.
EPI_ISL_422635 hCoV-19/Netherlands/ZuidHolland_99/2020 Europe / Netherlands / Zuid Holland 2020-03-31 Dutch COVID-19 response team Erasmus Medical Center Bas Oude Munnink, David Nieuwenhuijse, Reina Sikkema, Claudia Schapendonk, Irina Chestakova, Anne van der Linden, Theo Bestebroer, Stefan van Nieuwkoop, Mark Pronk, Pascal Lexmond, Corien Swaan, Manon Haverkate, Madelief Mollers, Mart Stein, Sandra Kengne Kamga Mobou, Jeroen van Kampen, Jolanda Voermans, Aura Timen, Corine GeurtsvanKessel, Annemiek van der Eijk, Richard Molenkamp, Marion Koopmans, on behalf of the Dutch national COVID-19 response team.
EPI_ISL_422636 hCoV-19/Czech Republic/2741/2020 Europe / Czech Republic / Prague 2020-03-08 The National Institute of Public Health Center for Epidemiology and Microbiology State Veterinary Institute Prague Alexander Nagy, Helena Jirincova, Klara Labska, Ludmila Novakova, Olga Storkanova, Dusan Trnka, Jaromira Vecerova

EPI_ISL_422638 hCoV-19/Netherlands/Gelderland_10/2020 Europe / Netherlands / Gelderland 2020-03-20 Dutch COVID-19 response team Erasmus Medical Center Bas Oude Munnink, David Nieuwenhuijse, Reina Sikkema, Claudia Schapendonk, Irina Chestakova, Anne van der Linden, Theo Bestebroer, Stefan van Nieuwkoop, Mark Pronk, Pascal Lexmond, Corien Swaan, Manon Haverkate, Madelief Mollers, Mart Stein, Sandra Kengne Kamga Mobou, Jeroen van Kampen, Jolanda Voermans, Aura Timen, Corine GeurtsvanKessel, Annemiek van der Eijk, Richard Molenkamp, Marion Koopmans, on behalf of the Dutch national COVID-19 response team.
EPI_ISL_422639 hCoV-19/Netherlands/Gelderland_11/2020 Europe / Netherlands / Gelderland 2020-03-21 Dutch COVID-19 response team Erasmus Medical Center Bas Oude Munnink, David Nieuwenhuijse, Reina Sikkema, Claudia Schapendonk, Irina Chestakova, Anne van der Linden, Theo Bestebroer, Stefan van Nieuwkoop, Mark Pronk, Pascal Lexmond, Corien Swaan, Manon Haverkate, Madelief Mollers, Mart Stein, Sandra Kengne Kamga Mobou, Jeroen van Kampen, Jolanda Voermans, Aura Timen, Corine GeurtsvanKessel, Annemiek van der Eijk, Richard Molenkamp, Marion Koopmans, on behalf of the Dutch national COVID-19 response team.
EPI_ISL_422640 hCoV-19/Netherlands/Gelderland_12/2020 Europe / Netherlands / Gelderland 2020-03-20 Dutch COVID-19 response team Erasmus Medical Center Bas Oude Munnink, David Nieuwenhuijse, Reina Sikkema, Claudia Schapendonk, Irina Chestakova, Anne van der Linden, Theo Bestebroer, Stefan van Nieuwkoop, Mark Pronk, Pascal Lexmond, Corien Swaan, Manon Haverkate, Madelief Mollers, Mart Stein, Sandra Kengne Kamga Mobou, Jeroen van Kampen, Jolanda Voermans, Aura Timen, Corine GeurtsvanKessel, Annemiek van der Eijk, Richard Molenkamp, Marion Koopmans, on behalf of the Dutch national COVID-19 response team.
EPI_ISL_422641 hCoV-19/Netherlands/Gelderland_5/2020 Europe / Netherlands / Gelderland 2020-03-10 Dutch COVID-19 response team Erasmus Medical Center Bas Oude Munnink, David Nieuwenhuijse, Reina Sikkema, Claudia Schapendonk, Irina Chestakova, Anne van der Linden, Theo Bestebroer, Stefan van Nieuwkoop, Mark Pronk, Pascal Lexmond, Corien Swaan, Manon Haverkate, Madelief Mollers, Mart Stein, Sandra Kengne Kamga Mobou, Jeroen van Kampen, Jolanda Voermans, Aura Timen, Corine GeurtsvanKessel, Annemiek van der Eijk, Richard Molenkamp, Marion Koopmans, on behalf of the Dutch national COVID-19 response team.
EPI_ISL_422642 hCoV-19/Netherlands/Gelderland_6/2020 Europe / Netherlands / Gelderland 2020-03-09 Dutch COVID-19 response team Erasmus Medical Center Bas Oude Munnink, David Nieuwenhuijse, Reina Sikkema, Claudia Schapendonk, Irina Chestakova, Anne van der Linden, Theo Bestebroer, Stefan van Nieuwkoop, Mark Pronk, Pascal Lexmond, Corien Swaan, Manon Haverkate, Madelief Mollers, Mart Stein, Sandra Kengne Kamga Mobou, Jeroen van Kampen, Jolanda Voermans, Aura Timen, Corine GeurtsvanKessel, Annemiek van der Eijk, Richard Molenkamp, Marion Koopmans, on behalf of the Dutch national COVID-19 response team.
EPI_ISL_422643 hCoV-19/Netherlands/Gelderland_7/2020 Europe / Netherlands / Gelderland 2020-03-11 Dutch COVID-19 response team Erasmus Medical Center Bas Oude Munnink, David Nieuwenhuijse, Reina Sikkema, Claudia Schapendonk, Irina Chestakova, Anne van der Linden, Theo Bestebroer, Stefan van Nieuwkoop, Mark Pronk, Pascal Lexmond, Corien Swaan, Manon Haverkate, Madelief Mollers, Mart Stein, Sandra Kengne Kamga Mobou, Jeroen van Kampen, Jolanda Voermans, Aura Timen, Corine GeurtsvanKessel, Annemiek van der Eijk, Richard Molenkamp, Marion Koopmans, on behalf of the Dutch national COVID-19 response team.
EPI_ISL_422644 hCoV-19/Netherlands/Gelderland_8/2020 Europe / Netherlands / Gelderland 2020-03-13 Dutch COVID-19 response team Erasmus Medical Center Bas Oude Munnink, David Nieuwenhuijse, Reina Sikkema, Claudia Schapendonk, Irina Chestakova, Anne van der Linden, Theo Bestebroer, Stefan van Nieuwkoop, Mark Pronk, Pascal Lexmond, Corien Swaan, Manon Haverkate, Madelief Mollers, Mart Stein, Sandra Kengne Kamga Mobou, Jeroen van Kampen, Jolanda Voermans, Aura Timen, Corine GeurtsvanKessel, Annemiek van der Eijk, Richard Molenkamp, Marion Koopmans, on behalf of the Dutch national COVID-19 response team.
EPI_ISL_422645 hCoV-19/Netherlands/Gelderland_9/2020 Europe / Netherlands / Gelderland 2020-03-18 Dutch COVID-19 response team Erasmus Medical Center Bas Oude Munnink, David Nieuwenhuijse, Reina Sikkema, Claudia Schapendonk, Irina Chestakova, Anne van der Linden, Theo Bestebroer, Stefan van Nieuwkoop, Mark Pronk, Pascal Lexmond, Corien Swaan, Manon Haverkate, Madelief Mollers, Mart Stein, Sandra Kengne Kamga Mobou, Jeroen van Kampen, Jolanda Voermans, Aura Timen, Corine GeurtsvanKessel, Annemiek van der Eijk, Richard Molenkamp, Marion Koopmans, on behalf of the Dutch national COVID-19 response team.
EPI_ISL_422646 hCoV-19/Netherlands/Limburg_10/2020 Europe / Netherlands / Limburg 2020-03-20 Dutch COVID-19 response team Erasmus Medical Center Bas Oude Munnink, David Nieuwenhuijse, Reina Sikkema, Claudia Schapendonk, Irina Chestakova, Anne van der Linden, Theo Bestebroer, Stefan van Nieuwkoop, Mark Pronk, Pascal Lexmond, Corien Swaan, Manon Haverkate, Madelief Mollers, Mart Stein, Sandra Kengne Kamga Mobou, Jeroen van Kampen, Jolanda Voermans, Aura Timen, Corine GeurtsvanKessel, Annemiek van der Eijk, Richard Molenkamp, Marion Koopmans, on behalf of the Dutch national COVID-19 response team.
EPI_ISL_422647 hCoV-19/Netherlands/Limburg_8/2020 Europe / Netherlands / Limburg 2020-03-19 Dutch COVID-19 response team Erasmus Medical Center Bas Oude Munnink, David Nieuwenhuijse, Reina Sikkema, Claudia Schapendonk, Irina Chestakova, Anne van der Linden, Theo Bestebroer, Stefan van Nieuwkoop, Mark Pronk, Pascal Lexmond, Corien Swaan, Manon Haverkate, Madelief Mollers, Mart Stein, Sandra Kengne Kamga Mobou, Jeroen van Kampen, Jolanda Voermans, Aura Timen, Corine GeurtsvanKessel, Annemiek van der Eijk, Richard Molenkamp, Marion Koopmans, on behalf of the Dutch national COVID-19 response team.
EPI_ISL_422648 hCoV-19/Netherlands/Limburg_9/2020 Europe / Netherlands / Limburg 2020-03-20 Dutch COVID-19 response team Erasmus Medical Center Bas Oude Munnink, David Nieuwenhuijse, Reina Sikkema, Claudia Schapendonk, Irina Chestakova, Anne van der Linden, Theo Bestebroer, Stefan van Nieuwkoop, Mark Pronk, Pascal Lexmond, Corien Swaan, Manon Haverkate, Madelief Mollers, Mart Stein, Sandra Kengne Kamga Mobou, Jeroen van Kampen, Jolanda Voermans, Aura Timen, Corine GeurtsvanKessel, Annemiek van der Eijk, Richard Molenkamp, Marion Koopmans, on behalf of the Dutch national COVID-19 response team.
EPI_ISL_422649 hCoV-19/Netherlands/NA_100/2020 Europe / Netherlands 2020-03-18 Dutch COVID-19 response team Erasmus Medical Center Bas Oude Munnink, David Nieuwenhuijse, Reina Sikkema, Claudia Schapendonk, Irina Chestakova, Anne van der Linden, Theo Bestebroer, Stefan van Nieuwkoop, Mark Pronk, Pascal Lexmond, Corien Swaan, Manon Haverkate, Madelief Mollers, Mart Stein, Sandra Kengne Kamga Mobou, Jeroen van Kampen,

Jolanda Voermans, Aura Timen, Corine GeurtsvanKessel, Annemiek van der Eijk, Richard Molenkamp, Marion Koopmans, on behalf of the Dutch national COVID-19 response team.
EPI_ISL_422650 hCoV-19/Netherlands/NA_101/2020 Europe / Netherlands 2020-03-18 Dutch COVID-19 response team Erasmus Medical Center Bas Oude Munnink, David Nieuwenhuijse, Reina Sikkema, Claudia Schapendonk, Irina Chestakova, Anne van der Linden, Theo Bestebroer, Stefan van Nieuwkoop, Mark Pronk, Pascal Lexmond, Corien Swaan, Manon Haverkate, Madelief Mollers, Mart Stein, Sandra Kengne Kamga Mobou, Jeroen van Kampen, Jolanda Voermans, Aura Timen, Corine GeurtsvanKessel, Annemiek van der Eijk, Richard Molenkamp, Marion Koopmans, on behalf of the Dutch national COVID-19 response team.
EPI_ISL_422651 hCoV-19/Netherlands/NA_102/2020 Europe / Netherlands 2020-03-18 Dutch COVID-19 response team Erasmus Medical Center Bas Oude Munnink, David Nieuwenhuijse, Reina Sikkema, Claudia Schapendonk, Irina Chestakova, Anne van der Linden, Theo Bestebroer, Stefan van Nieuwkoop, Mark Pronk, Pascal Lexmond, Corien Swaan, Manon Haverkate, Madelief Mollers, Mart Stein, Sandra Kengne Kamga Mobou, Jeroen van Kampen, Jolanda Voermans, Aura Timen, Corine GeurtsvanKessel, Annemiek van der Eijk, Richard Molenkamp, Marion Koopmans, on behalf of the Dutch national COVID-19 response team.
EPI_ISL_422652 hCoV-19/Netherlands/NA_103/2020 Europe / Netherlands 2020-03-18 Dutch COVID-19 response team Erasmus Medical Center Bas Oude Munnink, David Nieuwenhuijse, Reina Sikkema, Claudia Schapendonk, Irina Chestakova, Anne van der Linden, Theo Bestebroer, Stefan van Nieuwkoop, Mark Pronk, Pascal Lexmond, Corien Swaan, Manon Haverkate, Madelief Mollers, Mart Stein, Sandra Kengne Kamga Mobou, Jeroen van Kampen, Jolanda Voermans, Aura Timen, Corine GeurtsvanKessel, Annemiek van der Eijk, Richard Molenkamp, Marion Koopmans, on behalf of the Dutch national COVID-19 response team.
EPI_ISL_422653 hCoV-19/Netherlands/NA_104/2020 Europe / Netherlands 2020-03-18 Dutch COVID-19 response team Erasmus Medical Center Bas Oude Munnink, David Nieuwenhuijse, Reina Sikkema, Claudia Schapendonk, Irina Chestakova, Anne van der Linden, Theo Bestebroer, Stefan van Nieuwkoop, Mark Pronk, Pascal Lexmond, Corien Swaan, Manon Haverkate, Madelief Mollers, Mart Stein, Sandra Kengne Kamga Mobou, Jeroen van Kampen, Jolanda Voermans, Aura Timen, Corine GeurtsvanKessel, Annemiek van der Eijk, Richard Molenkamp, Marion Koopmans, on behalf of the Dutch national COVID-19 response team.
EPI_ISL_423202 hCoV-19/England/201360052/2020 Europe / United Kingdom / England 2020-03-24 Respiratory Virus Unit, Microbiology Services Colindale, Public Health England Respiratory Virus Unit, Microbiology Services Colindale, Public Health England Monica Galiano, Shahjahan Miah, Angie Lackenby, Omolola Akinbami, Tiina Talts, Leena Bhaw, Richard Myers, Steven Platt, Kirstin Edwards, Jonathan Hubb, Joanna Ellis, Maria Zambon
EPI_ISL_423203 hCoV-19/England/201360053/2020 Europe / United Kingdom / England 2020-03-24 Respiratory Virus Unit, Microbiology Services Colindale, Public Health England Respiratory Virus Unit, Microbiology Services Colindale, Public Health England Monica Galiano, Shahjahan Miah, Angie Lackenby, Omolola Akinbami, Tiina Talts, Leena Bhaw, Richard Myers, Steven Platt, Kirstin Edwards, Jonathan Hubb, Joanna Ellis, Maria Zambon
EPI_ISL_423204 hCoV-19/England/201360055/2020 Europe / United Kingdom / England 2020-03-26 Respiratory Virus Unit, Microbiology Services Colindale, Public Health England Respiratory Virus Unit, Microbiology Services Colindale, Public Health England Monica Galiano, Shahjahan Miah, Angie Lackenby, Omolola Akinbami, Tiina Talts, Leena Bhaw, Richard Myers, Steven Platt, Kirstin Edwards, Jonathan Hubb, Joanna Ellis, Maria Zambon
EPI_ISL_423205 hCoV-19/England/201360057/2020 Europe / United Kingdom / England 2020-03-25 Respiratory Virus Unit, Microbiology Services Colindale, Public Health England Respiratory Virus Unit, Microbiology Services Colindale, Public Health England Monica Galiano, Shahjahan Miah, Angie Lackenby, Omolola Akinbami, Tiina Talts, Leena Bhaw, Richard Myers, Steven Platt, Kirstin Edwards, Jonathan Hubb, Joanna Ellis, Maria Zambon
EPI_ISL_423208 hCoV-19/England/201360069/2020 Europe / United Kingdom / England 2020-03-24 Respiratory Virus Unit, Microbiology Services Colindale, Public Health England Respiratory Virus Unit, Microbiology Services Colindale, Public Health England Monica Galiano, Shahjahan Miah, Angie Lackenby, Omolola Akinbami, Tiina Talts, Leena Bhaw, Richard Myers, Steven Platt, Kirstin Edwards, Jonathan Hubb, Joanna Ellis, Maria Zambon
EPI_ISL_423209 hCoV-19/England/201360071/2020 Europe / United Kingdom / England 2020-03-24 Respiratory Virus Unit, Microbiology Services Colindale, Public Health England Respiratory Virus Unit, Microbiology Services Colindale, Public Health England Monica Galiano, Shahjahan Miah, Angie Lackenby, Omolola Akinbami, Tiina Talts, Leena Bhaw, Richard Myers, Steven Platt, Kirstin Edwards, Jonathan Hubb, Joanna Ellis, Maria Zambon
EPI_ISL_423210 hCoV-19/England/201360075/2020 Europe / United Kingdom / England 2020-03-27 Respiratory Virus Unit, Microbiology Services Colindale, Public Health England Respiratory Virus Unit, Microbiology Services Colindale, Public Health England Monica Galiano, Shahjahan Miah, Angie Lackenby, Omolola Akinbami, Tiina Talts, Leena Bhaw, Richard Myers, Steven Platt, Kirstin Edwards, Jonathan Hubb, Joanna Ellis, Maria Zambon
EPI_ISL_423214 hCoV-19/England/201360119/2020 Europe / United Kingdom / England 2020-03-25 Respiratory Virus Unit, Microbiology Services Colindale, Public Health England Respiratory Virus Unit, Microbiology Services Colindale, Public Health England Monica Galiano, Shahjahan Miah, Angie Lackenby, Omolola Akinbami, Tiina Talts, Leena Bhaw, Richard Myers, Steven Platt, Kirstin Edwards, Jonathan Hubb, Joanna Ellis, Maria Zambon
EPI_ISL_423215 hCoV-19/England/20136012004/2020 Europe / United Kingdom / England 2020-03-25 Respiratory Virus Unit, Microbiology Services Colindale, Public Health England Respiratory Virus Unit, Microbiology Services Colindale, Public Health England Monica Galiano, Shahjahan Miah, Angie Lackenby, Omolola Akinbami, Tiina Talts, Leena Bhaw, Richard Myers, Steven Platt, Kirstin Edwards, Jonathan Hubb, Joanna Ellis,

Maria Zambon
EPI_ISL_423216 hCoV-19/England/201360121/2020 Europe / United Kingdom / England 2020-03-25 Respiratory Virus Unit, Microbiology Services Colindale, Public Health England Respiratory Virus Unit, Microbiology Services Colindale, Public Health England Monica Galiano, Shahjahan Miah, Angie Lackenby, Omolola Akinbami, Tiina Talts, Leena Bhaw, Richard Myers, Steven Platt, Kirstin Edwards, Jonathan Hubb, Joanna Ellis, Maria Zambon
EPI_ISL_423218 hCoV-19/England/201360126/2020 Europe / United Kingdom / England 2020-03-25 Respiratory Virus Unit, Microbiology Services Colindale, Public Health England Respiratory Virus Unit, Microbiology Services Colindale, Public Health England Monica Galiano, Shahjahan Miah, Angie Lackenby, Omolola Akinbami, Tiina Talts, Leena Bhaw, Richard Myers, Steven Platt, Kirstin Edwards, Jonathan Hubb, Joanna Ellis, Maria Zambon
EPI_ISL_423219 hCoV-19/England/201360127/2020 Europe / United Kingdom / England 2020-03-25 Respiratory Virus Unit, Microbiology Services Colindale, Public Health England Respiratory Virus Unit, Microbiology Services Colindale, Public Health England Monica Galiano, Shahjahan Miah, Angie Lackenby, Omolola Akinbami, Tiina Talts, Leena Bhaw, Richard Myers, Steven Platt, Kirstin Edwards, Jonathan Hubb, Joanna Ellis, Maria Zambon
EPI_ISL_423221 hCoV-19/England/201360131/2020 Europe / United Kingdom / England 2020-03-24 Respiratory Virus Unit, Microbiology Services Colindale, Public Health England Respiratory Virus Unit, Microbiology Services Colindale, Public Health England Monica Galiano, Shahjahan Miah, Angie Lackenby, Omolola Akinbami, Tiina Talts, Leena Bhaw, Richard Myers, Steven Platt, Kirstin Edwards, Jonathan Hubb, Joanna Ellis, Maria Zambon
EPI_ISL_423223 hCoV-19/England/201360136/2020 Europe / United Kingdom / England 2020-03-25 Respiratory Virus Unit, Microbiology Services Colindale, Public Health England Respiratory Virus Unit, Microbiology Services Colindale, Public Health England Monica Galiano, Shahjahan Miah, Angie Lackenby, Omolola Akinbami, Tiina Talts, Leena Bhaw, Richard Myers, Steven Platt, Kirstin Edwards, Jonathan Hubb, Joanna Ellis, Maria Zambon
EPI_ISL_423224 hCoV-19/England/201360137/2020 Europe / United Kingdom / England 2020-03-26 Respiratory Virus Unit, Microbiology Services Colindale, Public Health England Respiratory Virus Unit, Microbiology Services Colindale, Public Health England Monica Galiano, Shahjahan Miah, Angie Lackenby, Omolola Akinbami, Tiina Talts, Leena Bhaw, Richard Myers, Steven Platt, Kirstin Edwards, Jonathan Hubb, Joanna Ellis, Maria Zambon
EPI_ISL_423227 hCoV-19/England/201360143/2020 Europe / United Kingdom / England 2020-03-24 Respiratory Virus Unit, Microbiology Services Colindale, Public Health England Respiratory Virus Unit, Microbiology Services Colindale, Public Health England Monica Galiano, Shahjahan Miah, Angie Lackenby, Omolola Akinbami, Tiina Talts, Leena Bhaw, Richard Myers, Steven Platt, Kirstin Edwards, Jonathan Hubb, Joanna Ellis, Maria Zambon
EPI_ISL_423228 hCoV-19/England/201360171/2020 Europe / United Kingdom / England 2020-03-25 Respiratory Virus Unit, Microbiology Services Colindale, Public Health England Respiratory Virus Unit, Microbiology Services Colindale, Public Health England Monica Galiano, Shahjahan Miah, Angie Lackenby, Omolola Akinbami, Tiina Talts, Leena Bhaw, Richard Myers, Steven Platt, Kirstin Edwards, Jonathan Hubb, Joanna Ellis, Maria Zambon
EPI_ISL_423231 hCoV-19/England/201360178/2020 Europe / United Kingdom / England 2020-03-24 Respiratory Virus Unit, Microbiology Services Colindale, Public Health England Respiratory Virus Unit, Microbiology Services Colindale, Public Health England Monica Galiano, Shahjahan Miah, Angie Lackenby, Omolola Akinbami, Tiina Talts, Leena Bhaw, Richard Myers, Steven Platt, Kirstin Edwards, Jonathan Hubb, Joanna Ellis, Maria Zambon
EPI_ISL_423232 hCoV-19/England/201360181/2020 Europe / United Kingdom / England 2020-03-26 Respiratory Virus Unit, Microbiology Services Colindale, Public Health England Respiratory Virus Unit, Microbiology Services Colindale, Public Health England Monica Galiano, Shahjahan Miah, Angie Lackenby, Omolola Akinbami, Tiina Talts, Leena Bhaw, Richard Myers, Steven Platt, Kirstin Edwards, Jonathan Hubb, Joanna Ellis, Maria Zambon
EPI_ISL_423233 hCoV-19/England/201360188/2020 Europe / United Kingdom / England 2020-03-24 Respiratory Virus Unit, Microbiology Services Colindale, Public Health England Respiratory Virus Unit, Microbiology Services Colindale, Public Health England Monica Galiano, Shahjahan Miah, Angie Lackenby, Omolola Akinbami, Tiina Talts, Leena Bhaw, Richard Myers, Steven Platt, Kirstin Edwards, Jonathan Hubb, Joanna Ellis, Maria Zambon
EPI_ISL_423235 hCoV-19/England/201360191/2020 Europe / United Kingdom / England 2020-03-24 Respiratory Virus Unit, Microbiology Services Colindale, Public Health England Respiratory Virus Unit, Microbiology Services Colindale, Public Health England Monica Galiano, Shahjahan Miah, Angie Lackenby, Omolola Akinbami, Tiina Talts, Leena Bhaw, Richard Myers, Steven Platt, Kirstin Edwards, Jonathan Hubb, Joanna Ellis, Maria Zambon
EPI_ISL_423236 hCoV-19/England/201360198/2020 Europe / United Kingdom / England 2020-03-24 Respiratory Virus Unit, Microbiology Services Colindale, Public Health England Respiratory Virus Unit, Microbiology Services Colindale, Public Health England Monica Galiano, Shahjahan Miah, Angie Lackenby, Omolola Akinbami, Tiina Talts, Leena Bhaw, Richard Myers, Steven Platt, Kirstin Edwards, Jonathan Hubb, Joanna Ellis, Maria Zambon
EPI_ISL_423237 hCoV-19/England/201360200/2020 Europe / United Kingdom / England 2020-03-24 Respiratory Virus Unit, Microbiology Services Colindale, Public Health England Respiratory Virus Unit, Microbiology Services Colindale, Public Health England Monica Galiano, Shahjahan Miah, Angie Lackenby, Omolola Akinbami, Tiina Talts, Leena Bhaw, Richard Myers, Steven Platt, Kirstin Edwards, Jonathan Hubb, Joanna Ellis,

Maria Zambon
EPI_ISL_423238 hCoV-19/England/201360201/2020 Europe / United Kingdom / England 2020-03-24 Respiratory Virus Unit, Microbiology Services Colindale, Public Health England Respiratory Virus Unit, Microbiology Services Colindale, Public Health England Monica Galiano, Shahjahan Miah, Angie Lackenby, Omolola Akinbami, Tiina Talts, Leena Bhaw, Richard Myers, Steven Platt, Kirstin Edwards, Jonathan Hubb, Joanna Ellis, Maria Zambon
EPI_ISL_423241 hCoV-19/England/201360206/2020 Europe / United Kingdom / England 2020-03-24 Respiratory Virus Unit, Microbiology Services Colindale, Public Health England Respiratory Virus Unit, Microbiology Services Colindale, Public Health England Monica Galiano, Shahjahan Miah, Angie Lackenby, Omolola Akinbami, Tiina Talts, Leena Bhaw, Richard Myers, Steven Platt, Kirstin Edwards, Jonathan Hubb, Joanna Ellis, Maria Zambon
EPI_ISL_423242 hCoV-19/England/201360210/2020 Europe / United Kingdom / England 2020-03-25 Respiratory Virus Unit, Microbiology Services Colindale, Public Health England Respiratory Virus Unit, Microbiology Services Colindale, Public Health England Monica Galiano, Shahjahan Miah, Angie Lackenby, Omolola Akinbami, Tiina Talts, Leena Bhaw, Richard Myers, Steven Platt, Kirstin Edwards, Jonathan Hubb, Joanna Ellis, Maria Zambon
EPI_ISL_423243 hCoV-19/England/201360211/2020 Europe / United Kingdom / England 2020-03-25 Respiratory Virus Unit, Microbiology Services Colindale, Public Health England Respiratory Virus Unit, Microbiology Services Colindale, Public Health England Monica Galiano, Shahjahan Miah, Angie Lackenby, Omolola Akinbami, Tiina Talts, Leena Bhaw, Richard Myers, Steven Platt, Kirstin Edwards, Jonathan Hubb, Joanna Ellis, Maria Zambon
EPI_ISL_423244 hCoV-19/England/201360213/2020 Europe / United Kingdom / England 2020-03-25 Respiratory Virus Unit, Microbiology Services Colindale, Public Health England Respiratory Virus Unit, Microbiology Services Colindale, Public Health England Monica Galiano, Shahjahan Miah, Angie Lackenby, Omolola Akinbami, Tiina Talts, Leena Bhaw, Richard Myers, Steven Platt, Kirstin Edwards, Jonathan Hubb, Joanna Ellis, Maria Zambon
EPI_ISL_423246 hCoV-19/England/201360215/2020 Europe / United Kingdom / England 2020-03-25 Respiratory Virus Unit, Microbiology Services Colindale, Public Health England Respiratory Virus Unit, Microbiology Services Colindale, Public Health England Monica Galiano, Shahjahan Miah, Angie Lackenby, Omolola Akinbami, Tiina Talts, Leena Bhaw, Richard Myers, Steven Platt, Kirstin Edwards, Jonathan Hubb, Joanna Ellis, Maria Zambon
EPI_ISL_423248 hCoV-19/England/201360217/2020 Europe / United Kingdom / England 2020-03-25 Respiratory Virus Unit, Microbiology Services Colindale, Public Health England Respiratory Virus Unit, Microbiology Services Colindale, Public Health England Monica Galiano, Shahjahan Miah, Angie Lackenby, Omolola Akinbami, Tiina Talts, Leena Bhaw, Richard Myers, Steven Platt, Kirstin Edwards, Jonathan Hubb, Joanna Ellis, Maria Zambon
EPI_ISL_423769 hCoV-19/England/20132031404/2020 Europe / United Kingdom / England 2020-03-22 Respiratory Virus Unit, Microbiology Services Colindale, Public Health England Respiratory Virus Unit, Microbiology Services Colindale, Public Health England Monica Galiano, Shahjahan Miah, Angie Lackenby, Omolola Akinbami, Tiina Talts, Leena Bhaw, Richard Myers, Steven Platt, Kirstin Edwards, Jonathan Hubb, Joanna Ellis, Maria Zambon
EPI_ISL_423771 hCoV-19/England/20132032504/2020 Europe / United Kingdom / England 2020-03-24 Respiratory Virus Unit, Microbiology Services Colindale, Public Health England Respiratory Virus Unit, Microbiology Services Colindale, Public Health England Monica Galiano, Shahjahan Miah, Angie Lackenby, Omolola Akinbami, Tiina Talts, Leena Bhaw, Richard Myers, Steven Platt, Kirstin Edwards, Jonathan Hubb, Joanna Ellis, Maria Zambon
EPI_ISL_423772 hCoV-19/England/20132032604/2020 Europe / United Kingdom / England 2020-03-22 Respiratory Virus Unit, Microbiology Services Colindale, Public Health England Respiratory Virus Unit, Microbiology Services Colindale, Public Health England Monica Galiano, Shahjahan Miah, Angie Lackenby, Omolola Akinbami, Tiina Talts, Leena Bhaw, Richard Myers, Steven Platt, Kirstin Edwards, Jonathan Hubb, Joanna Ellis, Maria Zambon
EPI_ISL_423773 hCoV-19/England/20132032704/2020 Europe / United Kingdom / England 2020-03-24 Respiratory Virus Unit, Microbiology Services Colindale, Public Health England Respiratory Virus Unit, Microbiology Services Colindale, Public Health England Monica Galiano, Shahjahan Miah, Angie Lackenby, Omolola Akinbami, Tiina Talts, Leena Bhaw, Richard Myers, Steven Platt, Kirstin Edwards, Jonathan Hubb, Joanna Ellis, Maria Zambon
EPI_ISL_423774 hCoV-19/England/20132032804/2020 Europe / United Kingdom / England 2020-03-22 Respiratory Virus Unit, Microbiology Services Colindale, Public Health England Respiratory Virus Unit, Microbiology Services Colindale, Public Health England Monica Galiano, Shahjahan Miah, Angie Lackenby, Omolola Akinbami, Tiina Talts, Leena Bhaw, Richard Myers, Steven Platt, Kirstin Edwards, Jonathan Hubb, Joanna Ellis, Maria Zambon
EPI_ISL_423775 hCoV-19/England/20132032904/2020 Europe / United Kingdom / England 2020-03-22 Respiratory Virus Unit, Microbiology Services Colindale, Public Health England Respiratory Virus Unit, Microbiology Services Colindale, Public Health England Monica Galiano, Shahjahan Miah, Angie Lackenby, Omolola Akinbami, Tiina Talts, Leena Bhaw, Richard Myers, Steven Platt, Kirstin Edwards, Jonathan Hubb, Joanna Ellis, Maria Zambon
EPI_ISL_423778 hCoV-19/England/20132035704/2020 Europe / United Kingdom / England 2020-03-24 Respiratory Virus Unit, Microbiology Services Colindale, Public Health England Respiratory Virus Unit, Microbiology Services Colindale, Public Health England Monica Galiano, Shahjahan Miah, Angie Lackenby, Omolola Akinbami, Tiina Talts, Leena Bhaw, Richard Myers, Steven Platt, Kirstin Edwards, Jonathan Hubb, Joanna Ellis,

Maria Zambon
EPI_ISL_423779 hCoV-19/England/20132035904/2020 Europe / United Kingdom / England 2020-03-23 Respiratory Virus Unit, Microbiology Services Colindale, Public Health England Respiratory Virus Unit, Microbiology Services Colindale, Public Health England Monica Galiano, Shahjahan Miah, Angie Lackenby, Omolola Akinbami, Tiina Talts, Leena Bhaw, Richard Myers, Steven Platt, Kirstin Edwards, Jonathan Hubb, Joanna Ellis, Maria Zambon
EPI_ISL_423782 hCoV-19/England/20132047304/2020 Europe / United Kingdom / England 2020-03-23 Respiratory Virus Unit, Microbiology Services Colindale, Public Health England Respiratory Virus Unit, Microbiology Services Colindale, Public Health England Monica Galiano, Shahjahan Miah, Angie Lackenby, Omolola Akinbami, Tiina Talts, Leena Bhaw, Richard Myers, Steven Platt, Kirstin Edwards, Jonathan Hubb, Joanna Ellis, Maria Zambon
EPI_ISL_423783 hCoV-19/England/20132047404/2020 Europe / United Kingdom / England 2020-03-24 Respiratory Virus Unit, Microbiology Services Colindale, Public Health England Respiratory Virus Unit, Microbiology Services Colindale, Public Health England Monica Galiano, Shahjahan Miah, Angie Lackenby, Omolola Akinbami, Tiina Talts, Leena Bhaw, Richard Myers, Steven Platt, Kirstin Edwards, Jonathan Hubb, Joanna Ellis, Maria Zambon
EPI_ISL_423786 hCoV-19/England/20132054604/2020 Europe / United Kingdom / England 2020-03-23 Respiratory Virus Unit, Microbiology Services Colindale, Public Health England Respiratory Virus Unit, Microbiology Services Colindale, Public Health England Monica Galiano, Shahjahan Miah, Angie Lackenby, Omolola Akinbami, Tiina Talts, Leena Bhaw, Richard Myers, Steven Platt, Kirstin Edwards, Jonathan Hubb, Joanna Ellis, Maria Zambon
EPI_ISL_423787 hCoV-19/England/20132054704/2020 Europe / United Kingdom / England 2020-03-23 Respiratory Virus Unit, Microbiology Services Colindale, Public Health England Respiratory Virus Unit, Microbiology Services Colindale, Public Health England Monica Galiano, Shahjahan Miah, Angie Lackenby, Omolola Akinbami, Tiina Talts, Leena Bhaw, Richard Myers, Steven Platt, Kirstin Edwards, Jonathan Hubb, Joanna Ellis, Maria Zambon
EPI_ISL_423791 hCoV-19/England/20132066804/2020 Europe / United Kingdom / England 2020-03-23 Respiratory Virus Unit, Microbiology Services Colindale, Public Health England Respiratory Virus Unit, Microbiology Services Colindale, Public Health England Monica Galiano, Shahjahan Miah, Angie Lackenby, Omolola Akinbami, Tiina Talts, Leena Bhaw, Richard Myers, Steven Platt, Kirstin Edwards, Jonathan Hubb, Joanna Ellis, Maria Zambon
EPI_ISL_423796 hCoV-19/England/20132068204/2020 Europe / United Kingdom / England 2020-03-23 Respiratory Virus Unit, Microbiology Services Colindale, Public Health England Respiratory Virus Unit, Microbiology Services Colindale, Public Health England Monica Galiano, Shahjahan Miah, Angie Lackenby, Omolola Akinbami, Tiina Talts, Leena Bhaw, Richard Myers, Steven Platt, Kirstin Edwards, Jonathan Hubb, Joanna Ellis, Maria Zambon
EPI_ISL_423797 hCoV-19/England/20132068304/2020 Europe / United Kingdom / England 2020-03-23 Respiratory Virus Unit, Microbiology Services Colindale, Public Health England Respiratory Virus Unit, Microbiology Services Colindale, Public Health England Monica Galiano, Shahjahan Miah, Angie Lackenby, Omolola Akinbami, Tiina Talts, Leena Bhaw, Richard Myers, Steven Platt, Kirstin Edwards, Jonathan Hubb, Joanna Ellis, Maria Zambon
EPI_ISL_423798 hCoV-19/England/20132069004/2020 Europe / United Kingdom / England 2020-03-23 Respiratory Virus Unit, Microbiology Services Colindale, Public Health England Respiratory Virus Unit, Microbiology Services Colindale, Public Health England Monica Galiano, Shahjahan Miah, Angie Lackenby, Omolola Akinbami, Tiina Talts, Leena Bhaw, Richard Myers, Steven Platt, Kirstin Edwards, Jonathan Hubb, Joanna Ellis, Maria Zambon
EPI_ISL_423799 hCoV-19/England/20132069104/2020 Europe / United Kingdom / England 2020-03-23 Respiratory Virus Unit, Microbiology Services Colindale, Public Health England Respiratory Virus Unit, Microbiology Services Colindale, Public Health England Monica Galiano, Shahjahan Miah, Angie Lackenby, Omolola Akinbami, Tiina Talts, Leena Bhaw, Richard Myers, Steven Platt, Kirstin Edwards, Jonathan Hubb, Joanna Ellis, Maria Zambon
EPI_ISL_423803 hCoV-19/England/20132080004/2020 Europe / United Kingdom / England 2020-03-24 Respiratory Virus Unit, Microbiology Services Colindale, Public Health England Respiratory Virus Unit, Microbiology Services Colindale, Public Health England Monica Galiano, Shahjahan Miah, Angie Lackenby, Omolola Akinbami, Tiina Talts, Leena Bhaw, Richard Myers, Steven Platt, Kirstin Edwards, Jonathan Hubb, Joanna Ellis, Maria Zambon
EPI_ISL_423808 hCoV-19/England/20132080804/2020 Europe / United Kingdom / England 2020-03-23 Respiratory Virus Unit, Microbiology Services Colindale, Public Health England Respiratory Virus Unit, Microbiology Services Colindale, Public Health England Monica Galiano, Shahjahan Miah, Angie Lackenby, Omolola Akinbami, Tiina Talts, Leena Bhaw, Richard Myers, Steven Platt, Kirstin Edwards, Jonathan Hubb, Joanna Ellis, Maria Zambon
EPI_ISL_423809 hCoV-19/England/20132081104/2020 Europe / United Kingdom / England 2020-03-24 Respiratory Virus Unit, Microbiology Services Colindale, Public Health England Respiratory Virus Unit, Microbiology Services Colindale, Public Health England Monica Galiano, Shahjahan Miah, Angie Lackenby, Omolola Akinbami, Tiina Talts, Leena Bhaw, Richard Myers, Steven Platt, Kirstin Edwards, Jonathan Hubb, Joanna Ellis, Maria Zambon
EPI_ISL_423810 hCoV-19/England/20132081304/2020 Europe / United Kingdom / England 2020-03-23 Respiratory Virus Unit, Microbiology Services Colindale, Public Health England Respiratory Virus Unit, Microbiology Services Colindale, Public Health England Monica Galiano, Shahjahan Miah, Angie Lackenby, Omolola Akinbami, Tiina Talts, Leena Bhaw, Richard Myers, Steven Platt, Kirstin Edwards, Jonathan Hubb, Joanna Ellis,

Maria Zambon
EPI_ISL_423811 hCoV-19/England/20132081404/2020 Europe / United Kingdom / England 2020-03-23 Respiratory Virus Unit, Microbiology Services Colindale, Public Health England Respiratory Virus Unit, Microbiology Services Colindale, Public Health England Monica Galiano, Shahjahan Miah, Angie Lackenby, Omolola Akinbami, Tiina Talts, Leena Bhaw, Richard Myers, Steven Platt, Kirstin Edwards, Jonathan Hubb, Joanna Ellis, Maria Zambon
EPI_ISL_423812 hCoV-19/England/20132081804/2020 Europe / United Kingdom / England 2020-03-23 Respiratory Virus Unit, Microbiology Services Colindale, Public Health England Respiratory Virus Unit, Microbiology Services Colindale, Public Health England Monica Galiano, Shahjahan Miah, Angie Lackenby, Omolola Akinbami, Tiina Talts, Leena Bhaw, Richard Myers, Steven Platt, Kirstin Edwards, Jonathan Hubb, Joanna Ellis, Maria Zambon
EPI_ISL_423813 hCoV-19/England/20132082004/2020 Europe / United Kingdom / England 2020-03-23 Respiratory Virus Unit, Microbiology Services Colindale, Public Health England Respiratory Virus Unit, Microbiology Services Colindale, Public Health England Monica Galiano, Shahjahan Miah, Angie Lackenby, Omolola Akinbami, Tiina Talts, Leena Bhaw, Richard Myers, Steven Platt, Kirstin Edwards, Jonathan Hubb, Joanna Ellis, Maria Zambon
EPI_ISL_423817 hCoV-19/England/20132082504/2020 Europe / United Kingdom / England 2020-03-23 Respiratory Virus Unit, Microbiology Services Colindale, Public Health England Respiratory Virus Unit, Microbiology Services Colindale, Public Health England Monica Galiano, Shahjahan Miah, Angie Lackenby, Omolola Akinbami, Tiina Talts, Leena Bhaw, Richard Myers, Steven Platt, Kirstin Edwards, Jonathan Hubb, Joanna Ellis, Maria Zambon
EPI_ISL_423818 hCoV-19/England/20132090404/2020 Europe / United Kingdom / England 2020-03-22 Respiratory Virus Unit, Microbiology Services Colindale, Public Health England Respiratory Virus Unit, Microbiology Services Colindale, Public Health England Monica Galiano, Shahjahan Miah, Angie Lackenby, Omolola Akinbami, Tiina Talts, Leena Bhaw, Richard Myers, Steven Platt, Kirstin Edwards, Jonathan Hubb, Joanna Ellis, Maria Zambon
EPI_ISL_423819 hCoV-19/England/20132090504/2020 Europe / United Kingdom / England 2020-03-23 Respiratory Virus Unit, Microbiology Services Colindale, Public Health England Respiratory Virus Unit, Microbiology Services Colindale, Public Health England Monica Galiano, Shahjahan Miah, Angie Lackenby, Omolola Akinbami, Tiina Talts, Leena Bhaw, Richard Myers, Steven Platt, Kirstin Edwards, Jonathan Hubb, Joanna Ellis, Maria Zambon
EPI_ISL_423822 hCoV-19/England/20132091504/2020 Europe / United Kingdom / England 2020-03-22 Respiratory Virus Unit, Microbiology Services Colindale, Public Health England Respiratory Virus Unit, Microbiology Services Colindale, Public Health England Monica Galiano, Shahjahan Miah, Angie Lackenby, Omolola Akinbami, Tiina Talts, Leena Bhaw, Richard Myers, Steven Platt, Kirstin Edwards, Jonathan Hubb, Joanna Ellis, Maria Zambon
EPI_ISL_423823 hCoV-19/England/20132091704/2020 Europe / United Kingdom / England 2020-03-22 Respiratory Virus Unit, Microbiology Services Colindale, Public Health England Respiratory Virus Unit, Microbiology Services Colindale, Public Health England Monica Galiano, Shahjahan Miah, Angie Lackenby, Omolola Akinbami, Tiina Talts, Leena Bhaw, Richard Myers, Steven Platt, Kirstin Edwards, Jonathan Hubb, Joanna Ellis, Maria Zambon
EPI_ISL_423824 hCoV-19/England/20132091804/2020 Europe / United Kingdom / England 2020-03-22 Respiratory Virus Unit, Microbiology Services Colindale, Public Health England Respiratory Virus Unit, Microbiology Services Colindale, Public Health England Monica Galiano, Shahjahan Miah, Angie Lackenby, Omolola Akinbami, Tiina Talts, Leena Bhaw, Richard Myers, Steven Platt, Kirstin Edwards, Jonathan Hubb, Joanna Ellis, Maria Zambon
EPI_ISL_423825 hCoV-19/England/20132091904/2020 Europe / United Kingdom / England 2020-03-22 Respiratory Virus Unit, Microbiology Services Colindale, Public Health England Respiratory Virus Unit, Microbiology Services Colindale, Public Health England Monica Galiano, Shahjahan Miah, Angie Lackenby, Omolola Akinbami, Tiina Talts, Leena Bhaw, Richard Myers, Steven Platt, Kirstin Edwards, Jonathan Hubb, Joanna Ellis, Maria Zambon
EPI_ISL_423828 hCoV-19/England/20132095704/2020 Europe / United Kingdom / England 2020-03-23 Respiratory Virus Unit, Microbiology Services Colindale, Public Health England Respiratory Virus Unit, Microbiology Services Colindale, Public Health England Monica Galiano, Shahjahan Miah, Angie Lackenby, Omolola Akinbami, Tiina Talts, Leena Bhaw, Richard Myers, Steven Platt, Kirstin Edwards, Jonathan Hubb, Joanna Ellis, Maria Zambon
EPI_ISL_423829 hCoV-19/England/20132096004/2020 Europe / United Kingdom / England 2020-03-23 Respiratory Virus Unit, Microbiology Services Colindale, Public Health England Respiratory Virus Unit, Microbiology Services Colindale, Public Health England Monica Galiano, Shahjahan Miah, Angie Lackenby, Omolola Akinbami, Tiina Talts, Leena Bhaw, Richard Myers, Steven Platt, Kirstin Edwards, Jonathan Hubb, Joanna Ellis, Maria Zambon
EPI_ISL_423830 hCoV-19/England/20132096304/2020 Europe / United Kingdom / England 2020-03-23 Respiratory Virus Unit, Microbiology Services Colindale, Public Health England Respiratory Virus Unit, Microbiology Services Colindale, Public Health England Monica Galiano, Shahjahan Miah, Angie Lackenby, Omolola Akinbami, Tiina Talts, Leena Bhaw, Richard Myers, Steven Platt, Kirstin Edwards, Jonathan Hubb, Joanna Ellis, Maria Zambon
EPI_ISL_423832 hCoV-19/England/20132099304/2020 Europe / United Kingdom / England 2020-03-23 Respiratory Virus Unit, Microbiology Services Colindale, Public Health England Respiratory Virus Unit, Microbiology Services Colindale, Public Health England Monica Galiano, Shahjahan Miah, Angie Lackenby, Omolola Akinbami, Tiina Talts, Leena Bhaw, Richard Myers, Steven Platt, Kirstin Edwards, Jonathan Hubb, Joanna Ellis,

Maria Zambon
EPI_ISL_423834 hCoV-19/England/20132100404/2020 Europe / United Kingdom / England 2020-03-23 Respiratory Virus Unit, Microbiology Services Colindale, Public Health England Respiratory Virus Unit, Microbiology Services Colindale, Public Health England Monica Galiano, Shahjahan Miah, Angie Lackenby, Omolola Akinbami, Tiina Talts, Leena Bhaw, Richard Myers, Steven Platt, Kirstin Edwards, Jonathan Hubb, Joanna Ellis, Maria Zambon
EPI_ISL_423835 hCoV-19/England/20132100704/2020 Europe / United Kingdom / England 2020-03-23 Respiratory Virus Unit, Microbiology Services Colindale, Public Health England Respiratory Virus Unit, Microbiology Services Colindale, Public Health England Monica Galiano, Shahjahan Miah, Angie Lackenby, Omolola Akinbami, Tiina Talts, Leena Bhaw, Richard Myers, Steven Platt, Kirstin Edwards, Jonathan Hubb, Joanna Ellis, Maria Zambon
EPI_ISL_423837 hCoV-19/England/20132104104/2020 Europe / United Kingdom / England 2020-03-24 Respiratory Virus Unit, Microbiology Services Colindale, Public Health England Respiratory Virus Unit, Microbiology Services Colindale, Public Health England Monica Galiano, Shahjahan Miah, Angie Lackenby, Omolola Akinbami, Tiina Talts, Leena Bhaw, Richard Myers, Steven Platt, Kirstin Edwards, Jonathan Hubb, Joanna Ellis, Maria Zambon
EPI_ISL_423838 hCoV-19/England/20132104404/2020 Europe / United Kingdom / England 2020-03-24 Respiratory Virus Unit, Microbiology Services Colindale, Public Health England Respiratory Virus Unit, Microbiology Services Colindale, Public Health England Monica Galiano, Shahjahan Miah, Angie Lackenby, Omolola Akinbami, Tiina Talts, Leena Bhaw, Richard Myers, Steven Platt, Kirstin Edwards, Jonathan Hubb, Joanna Ellis, Maria Zambon
EPI_ISL_423839 hCoV-19/England/20132105704/2020 Europe / United Kingdom / England 2020-03-23 Respiratory Virus Unit, Microbiology Services Colindale, Public Health England Respiratory Virus Unit, Microbiology Services Colindale, Public Health England Monica Galiano, Shahjahan Miah, Angie Lackenby, Omolola Akinbami, Tiina Talts, Leena Bhaw, Richard Myers, Steven Platt, Kirstin Edwards, Jonathan Hubb, Joanna Ellis, Maria Zambon
EPI_ISL_423841 hCoV-19/England/20132106104/2020 Europe / United Kingdom / England 2020-03-28 Respiratory Virus Unit, Microbiology Services Colindale, Public Health England Respiratory Virus Unit, Microbiology Services Colindale, Public Health England Monica Galiano, Shahjahan Miah, Angie Lackenby, Omolola Akinbami, Tiina Talts, Leena Bhaw, Richard Myers, Steven Platt, Kirstin Edwards, Jonathan Hubb, Joanna Ellis, Maria Zambon
EPI_ISL_423844 hCoV-19/England/20132107304/2020 Europe / United Kingdom / England 2020-03-23 Respiratory Virus Unit, Microbiology Services Colindale, Public Health England Respiratory Virus Unit, Microbiology Services Colindale, Public Health England Monica Galiano, Shahjahan Miah, Angie Lackenby, Omolola Akinbami, Tiina Talts, Leena Bhaw, Richard Myers, Steven Platt, Kirstin Edwards, Jonathan Hubb, Joanna Ellis, Maria Zambon
EPI_ISL_423845 hCoV-19/England/20132107704/2020 Europe / United Kingdom / England 2020-03-24 Respiratory Virus Unit, Microbiology Services Colindale, Public Health England Respiratory Virus Unit, Microbiology Services Colindale, Public Health England Monica Galiano, Shahjahan Miah, Angie Lackenby, Omolola Akinbami, Tiina Talts, Leena Bhaw, Richard Myers, Steven Platt, Kirstin Edwards, Jonathan Hubb, Joanna Ellis, Maria Zambon
EPI_ISL_423847 hCoV-19/England/20132108004/2020 Europe / United Kingdom / England 2020-03-23 Respiratory Virus Unit, Microbiology Services Colindale, Public Health England Respiratory Virus Unit, Microbiology Services Colindale, Public Health England Monica Galiano, Shahjahan Miah, Angie Lackenby, Omolola Akinbami, Tiina Talts, Leena Bhaw, Richard Myers, Steven Platt, Kirstin Edwards, Jonathan Hubb, Joanna Ellis, Maria Zambon
EPI_ISL_423852 hCoV-19/England/20132108704/2020 Europe / United Kingdom / England 2020-03-23 Respiratory Virus Unit, Microbiology Services Colindale, Public Health England Respiratory Virus Unit, Microbiology Services Colindale, Public Health England Monica Galiano, Shahjahan Miah, Angie Lackenby, Omolola Akinbami, Tiina Talts, Leena Bhaw, Richard Myers, Steven Platt, Kirstin Edwards, Jonathan Hubb, Joanna Ellis, Maria Zambon
EPI_ISL_423857 hCoV-19/England/20124036306/2020 Europe / United Kingdom / England 2020-03-17 Respiratory Virus Unit, Microbiology Services Colindale, Public Health England Respiratory Virus Unit, Microbiology Services Colindale, Public Health England Monica Galiano, Shahjahan Miah, Angie Lackenby, Omolola Akinbami, Tiina Talts, Leena Bhaw, Richard Myers, Steven Platt, Kirstin Edwards, Jonathan Hubb, Joanna Ellis, Maria Zambon
EPI_ISL_423859 hCoV-19/England/20126000104/2020 Europe / United Kingdom / England 2020-03-17 Respiratory Virus Unit, Microbiology Services Colindale, Public Health England Respiratory Virus Unit, Microbiology Services Colindale, Public Health England Monica Galiano, Shahjahan Miah, Angie Lackenby, Omolola Akinbami, Tiina Talts, Leena Bhaw, Richard Myers, Steven Platt, Kirstin Edwards, Jonathan Hubb, Joanna Ellis, Maria Zambon
EPI_ISL_423867 hCoV-19/England/20126001904/2020 Europe / United Kingdom / England 2020-03-18 Respiratory Virus Unit, Microbiology Services Colindale, Public Health England Respiratory Virus Unit, Microbiology Services Colindale, Public Health England Monica Galiano, Shahjahan Miah, Angie Lackenby, Omolola Akinbami, Tiina Talts, Leena Bhaw, Richard Myers, Steven Platt, Kirstin Edwards, Jonathan Hubb, Joanna Ellis, Maria Zambon
EPI_ISL_423877 hCoV-19/England/20126003406/2020 Europe / United Kingdom / England 2020-03-17 Respiratory Virus Unit, Microbiology Services Colindale, Public Health England Respiratory Virus Unit, Microbiology Services Colindale, Public Health England Monica Galiano, Shahjahan Miah, Angie Lackenby, Omolola Akinbami, Tiina Talts, Leena Bhaw, Richard Myers, Steven Platt, Kirstin Edwards, Jonathan Hubb, Joanna Ellis,

Maria Zambon
EPI_ISL_423883  hCoV-19/England/20126009204/2020        Europe / United Kingdom / England       2020-03-18
Respiratory Virus Unit, Microbiology Services Colindale, Public Health England  Respiratory Virus Unit, Microbiology Services Colindale, Public Health England  Monica Galiano, Shahjahan Miah, Angie Lackenby, Omolola Akinbami, Tiina Talts, Leena Bhaw, Richard Myers, Steven Platt, Kirstin Edwards, Jonathan Hubb, Joanna Ellis, Maria Zambon
EPI_ISL_423923  hCoV-19/England/20128002204/2020        Europe / United Kingdom / England       2020-03-19
Respiratory Virus Unit, Microbiology Services Colindale, Public Health England  Respiratory Virus Unit, Microbiology Services Colindale, Public Health England  Monica Galiano, Shahjahan Miah, Angie Lackenby, Omolola Akinbami, Tiina Talts, Leena Bhaw, Richard Myers, Steven Platt, Kirstin Edwards, Jonathan Hubb, Joanna Ellis, Maria Zambon
EPI_ISL_423926  hCoV-19/England/20128003506/2020        Europe / United Kingdom / England       2020-03-18
Respiratory Virus Unit, Microbiology Services Colindale, Public Health England  Respiratory Virus Unit, Microbiology Services Colindale, Public Health England  Monica Galiano, Shahjahan Miah, Angie Lackenby, Omolola Akinbami, Tiina Talts, Leena Bhaw, Richard Myers, Steven Platt, Kirstin Edwards, Jonathan Hubb, Joanna Ellis, Maria Zambon
EPI_ISL_423928  hCoV-19/England/20128009404/2020        Europe / United Kingdom / England       2020-03-19
Respiratory Virus Unit, Microbiology Services Colindale, Public Health England  Respiratory Virus Unit, Microbiology Services Colindale, Public Health England  Monica Galiano, Shahjahan Miah, Angie Lackenby, Omolola Akinbami, Tiina Talts, Leena Bhaw, Richard Myers, Steven Platt, Kirstin Edwards, Jonathan Hubb, Joanna Ellis, Maria Zambon
EPI_ISL_423935  hCoV-19/England/20128012704/2020        Europe / United Kingdom / England       2020-03-18
Respiratory Virus Unit, Microbiology Services Colindale, Public Health England  Respiratory Virus Unit, Microbiology Services Colindale, Public Health England  Monica Galiano, Shahjahan Miah, Angie Lackenby, Omolola Akinbami, Tiina Talts, Leena Bhaw, Richard Myers, Steven Platt, Kirstin Edwards, Jonathan Hubb, Joanna Ellis, Maria Zambon
EPI_ISL_423938  hCoV-19/England/20128014404/2020        Europe / United Kingdom / England       2020-03-19
Respiratory Virus Unit, Microbiology Services Colindale, Public Health England  Respiratory Virus Unit, Microbiology Services Colindale, Public Health England  Monica Galiano, Shahjahan Miah, Angie Lackenby, Omolola Akinbami, Tiina Talts, Leena Bhaw, Richard Myers, Steven Platt, Kirstin Edwards, Jonathan Hubb, Joanna Ellis, Maria Zambon
EPI_ISL_423940  hCoV-19/England/20128014806/2020        Europe / United Kingdom / England       2020-03-18
Respiratory Virus Unit, Microbiology Services Colindale, Public Health England  Respiratory Virus Unit, Microbiology Services Colindale, Public Health England  Monica Galiano, Shahjahan Miah, Angie Lackenby, Omolola Akinbami, Tiina Talts, Leena Bhaw, Richard Myers, Steven Platt, Kirstin Edwards, Jonathan Hubb, Joanna Ellis, Maria Zambon
EPI_ISL_423942  hCoV-19/England/20128015104/2020        Europe / United Kingdom / England       2020-03-19
Respiratory Virus Unit, Microbiology Services Colindale, Public Health England  Respiratory Virus Unit, Microbiology Services Colindale, Public Health England  Monica Galiano, Shahjahan Miah, Angie Lackenby, Omolola Akinbami, Tiina Talts, Leena Bhaw, Richard Myers, Steven Platt, Kirstin Edwards, Jonathan Hubb, Joanna Ellis, Maria Zambon
EPI_ISL_423943  hCoV-19/England/20128015604/2020        Europe / United Kingdom / England       2020-03-19
Respiratory Virus Unit, Microbiology Services Colindale, Public Health England  Respiratory Virus Unit, Microbiology Services Colindale, Public Health England  Monica Galiano, Shahjahan Miah, Angie Lackenby, Omolola Akinbami, Tiina Talts, Leena Bhaw, Richard Myers, Steven Platt, Kirstin Edwards, Jonathan Hubb, Joanna Ellis, Maria Zambon
EPI_ISL_423948  hCoV-19/England/20128018804/2020        Europe / United Kingdom / England       2020-03-18
Respiratory Virus Unit, Microbiology Services Colindale, Public Health England  Respiratory Virus Unit, Microbiology Services Colindale, Public Health England  Monica Galiano, Shahjahan Miah, Angie Lackenby, Omolola Akinbami, Tiina Talts, Leena Bhaw, Richard Myers, Steven Platt, Kirstin Edwards, Jonathan Hubb, Joanna Ellis, Maria Zambon
EPI_ISL_423951  hCoV-19/England/20128035406/2020        Europe / United Kingdom / England       2020-03-19
Respiratory Virus Unit, Microbiology Services Colindale, Public Health England  Respiratory Virus Unit, Microbiology Services Colindale, Public Health England  Monica Galiano, Shahjahan Miah, Angie Lackenby, Omolola Akinbami, Tiina Talts, Leena Bhaw, Richard Myers, Steven Platt, Kirstin Edwards, Jonathan Hubb, Joanna Ellis, Maria Zambon
EPI_ISL_423952  hCoV-19/England/20128036004/2020        Europe / United Kingdom / England       2020-03-19
Respiratory Virus Unit, Microbiology Services Colindale, Public Health England  Respiratory Virus Unit, Microbiology Services Colindale, Public Health England  Monica Galiano, Shahjahan Miah, Angie Lackenby, Omolola Akinbami, Tiina Talts, Leena Bhaw, Richard Myers, Steven Platt, Kirstin Edwards, Jonathan Hubb, Joanna Ellis, Maria Zambon
EPI_ISL_423953  hCoV-19/England/20128036304/2020        Europe / United Kingdom / England       2020-03-18
Respiratory Virus Unit, Microbiology Services Colindale, Public Health England  Respiratory Virus Unit, Microbiology Services Colindale, Public Health England  Monica Galiano, Shahjahan Miah, Angie Lackenby, Omolola Akinbami, Tiina Talts, Leena Bhaw, Richard Myers, Steven Platt, Kirstin Edwards, Jonathan Hubb, Joanna Ellis, Maria Zambon
EPI_ISL_423956  hCoV-19/England/20128037406/2020        Europe / United Kingdom / England       2020-03-18
Respiratory Virus Unit, Microbiology Services Colindale, Public Health England  Respiratory Virus Unit, Microbiology Services Colindale, Public Health England  Monica Galiano, Shahjahan Miah, Angie Lackenby, Omolola Akinbami, Tiina Talts, Leena Bhaw, Richard Myers, Steven Platt, Kirstin Edwards, Jonathan Hubb, Joanna Ellis,

Maria Zambon
EPI_ISL_423973 hCoV-19/England/20129000404/2020 Europe / United Kingdom / England 2020-03-20 Respiratory Virus Unit, Microbiology Services Colindale, Public Health England Respiratory Virus Unit, Microbiology Services Colindale, Public Health England Monica Galiano, Shahjahan Miah, Angie Lackenby, Omolola Akinbami, Tiina Talts, Leena Bhaw, Richard Myers, Steven Platt, Kirstin Edwards, Jonathan Hubb, Joanna Ellis, Maria Zambon
EPI_ISL_423979 hCoV-19/England/20129009304/2020 Europe / United Kingdom / England 2020-03-20 Respiratory Virus Unit, Microbiology Services Colindale, Public Health England Respiratory Virus Unit, Microbiology Services Colindale, Public Health England Monica Galiano, Shahjahan Miah, Angie Lackenby, Omolola Akinbami, Tiina Talts, Leena Bhaw, Richard Myers, Steven Platt, Kirstin Edwards, Jonathan Hubb, Joanna Ellis, Maria Zambon
EPI_ISL_424500 hCoV-19/Iceland/477/2020 Europe / Iceland / Reykjavik 2020-03-20 The National University Hospital of Iceland deCODE genetics Daniel F Gudbjartsson; Agnar Helgason; Hakon Jonsson; Olafur T Magnusson; Pall Melsted; Gudmundur L Norddahl; Jona Saemundsdottir; Asgeir Sigurdsson; Patrick Sulem; Arna B Agustsdottir; Berglind Eiriksdottir; Run Fridriksdottir; Elisabet E Gardarsdottir; Gudmundur Georgsson; Olafia S Gretarsdottir; Kjartan R Gudmundsson; Thora R Gunnarsdottir; Arnaldur Gylfason; Hilma Holm; Brynjar O Jensson; Aslaug Jonasdottir; Kamilla S Josefsdottir; Thordur Kristjansson; Droplaug N Magnusdottir; Louise le Roux; Gudrun Sigmundsdottir; Gardar Sveinbjornsson; Kristin E Sveinsdottir; Maney Sveinsdottir; Emil A Thorarensen; Bjarni Thorbjornsson; Gisli Masson; Ingileif Jonsdottir; Alma Moller; Thorolfur Gudnason; Karl G Kristinsson; Unnur Thorsteinsdottir; Kari Stefansson
EPI_ISL_424502 hCoV-19/Iceland/479/2020 Europe / Iceland / Reykjavik 2020-03-20 The National University Hospital of Iceland deCODE genetics Daniel F Gudbjartsson; Agnar Helgason; Hakon Jonsson; Olafur T Magnusson; Pall Melsted; Gudmundur L Norddahl; Jona Saemundsdottir; Asgeir Sigurdsson; Patrick Sulem; Arna B Agustsdottir; Berglind Eiriksdottir; Run Fridriksdottir; Elisabet E Gardarsdottir; Gudmundur Georgsson; Olafia S Gretarsdottir; Kjartan R Gudmundsson; Thora R Gunnarsdottir; Arnaldur Gylfason; Hilma Holm; Brynjar O Jensson; Aslaug Jonasdottir; Kamilla S Josefsdottir; Thordur Kristjansson; Droplaug N Magnusdottir; Louise le Roux; Gudrun Sigmundsdottir; Gardar Sveinbjornsson; Kristin E Sveinsdottir; Maney Sveinsdottir; Emil A Thorarensen; Bjarni Thorbjornsson; Gisli Masson; Ingileif Jonsdottir; Alma Moller; Thorolfur Gudnason; Karl G Kristinsson; Unnur Thorsteinsdottir; Kari Stefansson
EPI_ISL_424503 hCoV-19/Iceland/480/2020 Europe / Iceland / Reykjavik 2020-03-20 The National University Hospital of Iceland deCODE genetics Daniel F Gudbjartsson; Agnar Helgason; Hakon Jonsson; Olafur T Magnusson; Pall Melsted; Gudmundur L Norddahl; Jona Saemundsdottir; Asgeir Sigurdsson; Patrick Sulem; Arna B Agustsdottir; Berglind Eiriksdottir; Run Fridriksdottir; Elisabet E Gardarsdottir; Gudmundur Georgsson; Olafia S Gretarsdottir; Kjartan R Gudmundsson; Thora R Gunnarsdottir; Arnaldur Gylfason; Hilma Holm; Brynjar O Jensson; Aslaug Jonasdottir; Kamilla S Josefsdottir; Thordur Kristjansson; Droplaug N Magnusdottir; Louise le Roux; Gudrun Sigmundsdottir; Gardar Sveinbjornsson; Kristin E Sveinsdottir; Maney Sveinsdottir; Emil A Thorarensen; Bjarni Thorbjornsson; Gisli Masson; Ingileif Jonsdottir; Alma Moller; Thorolfur Gudnason; Karl G Kristinsson; Unnur Thorsteinsdottir; Kari Stefansson
EPI_ISL_424506 hCoV-19/Iceland/483/2020 Europe / Iceland / Reykjavik 2020-03-20 The National University Hospital of Iceland deCODE genetics Daniel F Gudbjartsson; Agnar Helgason; Hakon Jonsson; Olafur T Magnusson; Pall Melsted; Gudmundur L Norddahl; Jona Saemundsdottir; Asgeir Sigurdsson; Patrick Sulem; Arna B Agustsdottir; Berglind Eiriksdottir; Run Fridriksdottir; Elisabet E Gardarsdottir; Gudmundur Georgsson; Olafia S Gretarsdottir; Kjartan R Gudmundsson; Thora R Gunnarsdottir; Arnaldur Gylfason; Hilma Holm; Brynjar O Jensson; Aslaug Jonasdottir; Kamilla S Josefsdottir; Thordur Kristjansson; Droplaug N Magnusdottir; Louise le Roux; Gudrun Sigmundsdottir; Gardar Sveinbjornsson; Kristin E Sveinsdottir; Maney Sveinsdottir; Emil A Thorarensen; Bjarni Thorbjornsson; Gisli Masson; Ingileif Jonsdottir; Alma Moller; Thorolfur Gudnason; Karl G Kristinsson; Unnur Thorsteinsdottir; Kari Stefansson
EPI_ISL_424507 hCoV-19/Iceland/484/2020 Europe / Iceland / Reykjavik 2020-03-20 The National University Hospital of Iceland deCODE genetics Daniel F Gudbjartsson; Agnar Helgason; Hakon Jonsson; Olafur T Magnusson; Pall Melsted; Gudmundur L Norddahl; Jona Saemundsdottir; Asgeir Sigurdsson; Patrick Sulem; Arna B Agustsdottir; Berglind Eiriksdottir; Run Fridriksdottir; Elisabet E Gardarsdottir; Gudmundur Georgsson; Olafia S Gretarsdottir; Kjartan R Gudmundsson; Thora R Gunnarsdottir; Arnaldur Gylfason; Hilma Holm; Brynjar O Jensson; Aslaug Jonasdottir; Kamilla S Josefsdottir; Thordur Kristjansson; Droplaug N Magnusdottir; Louise le Roux; Gudrun Sigmundsdottir; Gardar Sveinbjornsson; Kristin E Sveinsdottir; Maney Sveinsdottir; Emil A Thorarensen; Bjarni Thorbjornsson; Gisli Masson; Ingileif Jonsdottir; Alma Moller; Thorolfur Gudnason; Karl G Kristinsson; Unnur Thorsteinsdottir; Kari Stefansson
EPI_ISL_424508 hCoV-19/Iceland/485/2020 Europe / Iceland / Reykjavik 2020-03-20 The National University Hospital of Iceland deCODE genetics Daniel F Gudbjartsson; Agnar Helgason; Hakon Jonsson; Olafur T Magnusson; Pall Melsted; Gudmundur L Norddahl; Jona Saemundsdottir; Asgeir Sigurdsson; Patrick Sulem; Arna B Agustsdottir; Berglind Eiriksdottir; Run Fridriksdottir; Elisabet E Gardarsdottir; Gudmundur Georgsson; Olafia S Gretarsdottir; Kjartan R Gudmundsson; Thora R Gunnarsdottir; Arnaldur Gylfason; Hilma Holm; Brynjar O Jensson; Aslaug Jonasdottir; Kamilla S Josefsdottir; Thordur Kristjansson; Droplaug N Magnusdottir; Louise le Roux; Gudrun Sigmundsdottir; Gardar Sveinbjornsson; Kristin E Sveinsdottir; Maney Sveinsdottir; Emil A Thorarensen; Bjarni Thorbjornsson; Gisli Masson; Ingileif Jonsdottir; Alma Moller; Thorolfur Gudnason; Karl G Kristinsson; Unnur Thorsteinsdottir; Kari Stefansson
EPI_ISL_424509 hCoV-19/Iceland/487/2020 Europe / Iceland / Reykjavik 2020-03-20 The National University Hospital of Iceland deCODE genetics Daniel F Gudbjartsson; Agnar Helgason; Hakon Jonsson; Olafur T Magnusson; Pall Melsted; Gudmundur L Norddahl; Jona Saemundsdottir; Asgeir Sigurdsson; Patrick Sulem; Arna B Agustsdottir; Berglind Eiriksdottir; Run Fridriksdottir; Elisabet E Gardarsdottir; Gudmundur Georgsson; Olafia S Gretarsdottir; Kjartan R Gudmundsson; Thora R Gunnarsdottir; Arnaldur Gylfason; Hilma Holm; Brynjar O

Jensson; Aslaug Jonasdottir; Kamilla S Josefsdottir; Thordur Kristjansson; Droplaug N Magnusdottir; Louise le Roux; Gudrun Sigmundsdottir; Gardar Sveinbjornsson; Kristin E Sveinsdottir; Maney Sveinsdottir; Emil A Thorarensen; Bjarni Thorbjornsson; Gisli Masson; Ingileif Jonsdottir; Alma Moller; Thorolfur Gudnason; Karl G Kristinsson; Unnur Thorsteinsdottir; Kari Stefansson
EPI_ISL_424510 hCoV-19/Iceland/488/2020 Europe / Iceland / Reykjavik 2020-03-20 The National University Hospital of Iceland deCODE genetics Daniel F Gudbjartsson; Agnar Helgason; Hakon Jonsson; Olafur T Magnusson; Pall Melsted; Gudmundur L Norddahl; Jona Saemundsdottir; Asgeir Sigurdsson; Patrick Sulem; Arna B Agustsdottir; Berglind Eiriksdottir; Run Fridriksdottir; Elisabet E Gardarsdottir; Gudmundur Georgsson; Olafia S Gretarsdottir; Kjartan R Gudmundsson; Thora R Gunnarsdottir; Arnaldur Gylfason; Hilma Holm; Brynjar O Jensson; Aslaug Jonasdottir; Kamilla S Josefsdottir; Thordur Kristjansson; Droplaug N Magnusdottir; Louise le Roux; Gudrun Sigmundsdottir; Gardar Sveinbjornsson; Kristin E Sveinsdottir; Maney Sveinsdottir; Emil A Thorarensen; Bjarni Thorbjornsson; Gisli Masson; Ingileif Jonsdottir; Alma Moller; Thorolfur Gudnason; Karl G Kristinsson; Unnur Thorsteinsdottir; Kari Stefansson
EPI_ISL_424511 hCoV-19/Iceland/489/2020 Europe / Iceland / Reykjavik 2020-03-20 The National University Hospital of Iceland deCODE genetics Daniel F Gudbjartsson; Agnar Helgason; Hakon Jonsson; Olafur T Magnusson; Pall Melsted; Gudmundur L Norddahl; Jona Saemundsdottir; Asgeir Sigurdsson; Patrick Sulem; Arna B Agustsdottir; Berglind Eiriksdottir; Run Fridriksdottir; Elisabet E Gardarsdottir; Gudmundur Georgsson; Olafia S Gretarsdottir; Kjartan R Gudmundsson; Thora R Gunnarsdottir; Arnaldur Gylfason; Hilma Holm; Brynjar O Jensson; Aslaug Jonasdottir; Kamilla S Josefsdottir; Thordur Kristjansson; Droplaug N Magnusdottir; Louise le Roux; Gudrun Sigmundsdottir; Gardar Sveinbjornsson; Kristin E Sveinsdottir; Maney Sveinsdottir; Emil A Thorarensen; Bjarni Thorbjornsson; Gisli Masson; Ingileif Jonsdottir; Alma Moller; Thorolfur Gudnason; Karl G Kristinsson; Unnur Thorsteinsdottir; Kari Stefansson
EPI_ISL_424512 hCoV-19/Iceland/490/2020 Europe / Iceland / Reykjavik 2020-03-20 The National University Hospital of Iceland deCODE genetics Daniel F Gudbjartsson; Agnar Helgason; Hakon Jonsson; Olafur T Magnusson; Pall Melsted; Gudmundur L Norddahl; Jona Saemundsdottir; Asgeir Sigurdsson; Patrick Sulem; Arna B Agustsdottir; Berglind Eiriksdottir; Run Fridriksdottir; Elisabet E Gardarsdottir; Gudmundur Georgsson; Olafia S Gretarsdottir; Kjartan R Gudmundsson; Thora R Gunnarsdottir; Arnaldur Gylfason; Hilma Holm; Brynjar O Jensson; Aslaug Jonasdottir; Kamilla S Josefsdottir; Thordur Kristjansson; Droplaug N Magnusdottir; Louise le Roux; Gudrun Sigmundsdottir; Gardar Sveinbjornsson; Kristin E Sveinsdottir; Maney Sveinsdottir; Emil A Thorarensen; Bjarni Thorbjornsson; Gisli Masson; Ingileif Jonsdottir; Alma Moller; Thorolfur Gudnason; Karl G Kristinsson; Unnur Thorsteinsdottir; Kari Stefansson
EPI_ISL_424514 hCoV-19/Iceland/492/2020 Europe / Iceland / Reykjavik 2020-03-20 The National University Hospital of Iceland deCODE genetics Daniel F Gudbjartsson; Agnar Helgason; Hakon Jonsson; Olafur T Magnusson; Pall Melsted; Gudmundur L Norddahl; Jona Saemundsdottir; Asgeir Sigurdsson; Patrick Sulem; Arna B Agustsdottir; Berglind Eiriksdottir; Run Fridriksdottir; Elisabet E Gardarsdottir; Gudmundur Georgsson; Olafia S Gretarsdottir; Kjartan R Gudmundsson; Thora R Gunnarsdottir; Arnaldur Gylfason; Hilma Holm; Brynjar O Jensson; Aslaug Jonasdottir; Kamilla S Josefsdottir; Thordur Kristjansson; Droplaug N Magnusdottir; Louise le Roux; Gudrun Sigmundsdottir; Gardar Sveinbjornsson; Kristin E Sveinsdottir; Maney Sveinsdottir; Emil A Thorarensen; Bjarni Thorbjornsson; Gisli Masson; Ingileif Jonsdottir; Alma Moller; Thorolfur Gudnason; Karl G Kristinsson; Unnur Thorsteinsdottir; Kari Stefansson
EPI_ISL_424515 hCoV-19/Iceland/493/2020 Europe / Iceland / Reykjavik 2020-03-20 The National University Hospital of Iceland deCODE genetics Daniel F Gudbjartsson; Agnar Helgason; Hakon Jonsson; Olafur T Magnusson; Pall Melsted; Gudmundur L Norddahl; Jona Saemundsdottir; Asgeir Sigurdsson; Patrick Sulem; Arna B Agustsdottir; Berglind Eiriksdottir; Run Fridriksdottir; Elisabet E Gardarsdottir; Gudmundur Georgsson; Olafia S Gretarsdottir; Kjartan R Gudmundsson; Thora R Gunnarsdottir; Arnaldur Gylfason; Hilma Holm; Brynjar O Jensson; Aslaug Jonasdottir; Kamilla S Josefsdottir; Thordur Kristjansson; Droplaug N Magnusdottir; Louise le Roux; Gudrun Sigmundsdottir; Gardar Sveinbjornsson; Kristin E Sveinsdottir; Maney Sveinsdottir; Emil A Thorarensen; Bjarni Thorbjornsson; Gisli Masson; Ingileif Jonsdottir; Alma Moller; Thorolfur Gudnason; Karl G Kristinsson; Unnur Thorsteinsdottir; Kari Stefansson
EPI_ISL_424516 hCoV-19/Iceland/494/2020 Europe / Iceland / Reykjavik 2020-03-20 The National University Hospital of Iceland deCODE genetics Daniel F Gudbjartsson; Agnar Helgason; Hakon Jonsson; Olafur T Magnusson; Pall Melsted; Gudmundur L Norddahl; Jona Saemundsdottir; Asgeir Sigurdsson; Patrick Sulem; Arna B Agustsdottir; Berglind Eiriksdottir; Run Fridriksdottir; Elisabet E Gardarsdottir; Gudmundur Georgsson; Olafia S Gretarsdottir; Kjartan R Gudmundsson; Thora R Gunnarsdottir; Arnaldur Gylfason; Hilma Holm; Brynjar O Jensson; Aslaug Jonasdottir; Kamilla S Josefsdottir; Thordur Kristjansson; Droplaug N Magnusdottir; Louise le Roux; Gudrun Sigmundsdottir; Gardar Sveinbjornsson; Kristin E Sveinsdottir; Maney Sveinsdottir; Emil A Thorarensen; Bjarni Thorbjornsson; Gisli Masson; Ingileif Jonsdottir; Alma Moller; Thorolfur Gudnason; Karl G Kristinsson; Unnur Thorsteinsdottir; Kari Stefansson
EPI_ISL_424517 hCoV-19/Iceland/495/2020 Europe / Iceland / Reykjavik 2020-03-21 The National University Hospital of Iceland deCODE genetics Daniel F Gudbjartsson; Agnar Helgason; Hakon Jonsson; Olafur T Magnusson; Pall Melsted; Gudmundur L Norddahl; Jona Saemundsdottir; Asgeir Sigurdsson; Patrick Sulem; Arna B Agustsdottir; Berglind Eiriksdottir; Run Fridriksdottir; Elisabet E Gardarsdottir; Gudmundur Georgsson; Olafia S Gretarsdottir; Kjartan R Gudmundsson; Thora R Gunnarsdottir; Arnaldur Gylfason; Hilma Holm; Brynjar O Jensson; Aslaug Jonasdottir; Kamilla S Josefsdottir; Thordur Kristjansson; Droplaug N Magnusdottir; Louise le Roux; Gudrun Sigmundsdottir; Gardar Sveinbjornsson; Kristin E Sveinsdottir; Maney Sveinsdottir; Emil A Thorarensen; Bjarni Thorbjornsson; Gisli Masson; Ingileif Jonsdottir; Alma Moller; Thorolfur Gudnason; Karl G Kristinsson; Unnur Thorsteinsdottir; Kari Stefansson
EPI_ISL_424518 hCoV-19/Iceland/496/2020 Europe / Iceland / Reykjavik 2020-03-21 The National University Hospital of Iceland deCODE genetics Daniel F Gudbjartsson; Agnar Helgason; Hakon Jonsson; Olafur T Magnusson; Pall Melsted; Gudmundur L Norddahl; Jona Saemundsdottir; Asgeir Sigurdsson; Patrick Sulem; Arna B

Agustsdottir; Berglind Eiriksdottir; Run Fridriksdottir; Elisabet E Gardarsdottir; Gudmundur Georgsson; Olafia S Gretarsdottir; Kjartan R Gudmundsson; Thora R Gunnarsdottir; Arnaldur Gylfason; Hilma Holm; Brynjar O Jensson; Aslaug Jonasdottir; Kamilla S Josefsdottir; Thordur Kristjansson; Droplaug N Magnusdottir; Louise le Roux; Gudrun Sigmundsdottir; Gardar Sveinbjornsson; Kristin E Sveinsdottir; Maney Sveinsdottir; Emil A Thorarensen; Bjarni Thorbjornsson; Gisli Masson; Ingileif Jonsdottir; Alma Moller; Thorolfur Gudnason; Karl G Kristinsson; Unnur Thorsteinsdottir; Kari Stefansson
EPI_ISL_424519 hCoV-19/Iceland/497/2020 Europe / Iceland / Reykjavik 2020-03-21 The National University Hospital of Iceland deCODE genetics Daniel F Gudbjartsson; Agnar Helgason; Hakon Jonsson; Olafur T Magnusson; Pall Melsted; Gudmundur L Norddahl; Jona Saemundsdottir; Asgeir Sigurdsson; Patrick Sulem; Arna B Agustsdottir; Berglind Eiriksdottir; Run Fridriksdottir; Elisabet E Gardarsdottir; Gudmundur Georgsson; Olafia S Gretarsdottir; Kjartan R Gudmundsson; Thora R Gunnarsdottir; Arnaldur Gylfason; Hilma Holm; Brynjar O Jensson; Aslaug Jonasdottir; Kamilla S Josefsdottir; Thordur Kristjansson; Droplaug N Magnusdottir; Louise le Roux; Gudrun Sigmundsdottir; Gardar Sveinbjornsson; Kristin E Sveinsdottir; Maney Sveinsdottir; Emil A Thorarensen; Bjarni Thorbjornsson; Gisli Masson; Ingileif Jonsdottir; Alma Moller; Thorolfur Gudnason; Karl G Kristinsson; Unnur Thorsteinsdottir; Kari Stefansson
EPI_ISL_424520 hCoV-19/Iceland/498/2020 Europe / Iceland / Reykjavik 2020-03-21 The National University Hospital of Iceland deCODE genetics Daniel F Gudbjartsson; Agnar Helgason; Hakon Jonsson; Olafur T Magnusson; Pall Melsted; Gudmundur L Norddahl; Jona Saemundsdottir; Asgeir Sigurdsson; Patrick Sulem; Arna B Agustsdottir; Berglind Eiriksdottir; Run Fridriksdottir; Elisabet E Gardarsdottir; Gudmundur Georgsson; Olafia S Gretarsdottir; Kjartan R Gudmundsson; Thora R Gunnarsdottir; Arnaldur Gylfason; Hilma Holm; Brynjar O Jensson; Aslaug Jonasdottir; Kamilla S Josefsdottir; Thordur Kristjansson; Droplaug N Magnusdottir; Louise le Roux; Gudrun Sigmundsdottir; Gardar Sveinbjornsson; Kristin E Sveinsdottir; Maney Sveinsdottir; Emil A Thorarensen; Bjarni Thorbjornsson; Gisli Masson; Ingileif Jonsdottir; Alma Moller; Thorolfur Gudnason; Karl G Kristinsson; Unnur Thorsteinsdottir; Kari Stefansson
EPI_ISL_424521 hCoV-19/Iceland/499/2020 Europe / Iceland / Reykjavik 2020-03-21 The National University Hospital of Iceland deCODE genetics Daniel F Gudbjartsson; Agnar Helgason; Hakon Jonsson; Olafur T Magnusson; Pall Melsted; Gudmundur L Norddahl; Jona Saemundsdottir; Asgeir Sigurdsson; Patrick Sulem; Arna B Agustsdottir; Berglind Eiriksdottir; Run Fridriksdottir; Elisabet E Gardarsdottir; Gudmundur Georgsson; Olafia S Gretarsdottir; Kjartan R Gudmundsson; Thora R Gunnarsdottir; Arnaldur Gylfason; Hilma Holm; Brynjar O Jensson; Aslaug Jonasdottir; Kamilla S Josefsdottir; Thordur Kristjansson; Droplaug N Magnusdottir; Louise le Roux; Gudrun Sigmundsdottir; Gardar Sveinbjornsson; Kristin E Sveinsdottir; Maney Sveinsdottir; Emil A Thorarensen; Bjarni Thorbjornsson; Gisli Masson; Ingileif Jonsdottir; Alma Moller; Thorolfur Gudnason; Karl G Kristinsson; Unnur Thorsteinsdottir; Kari Stefansson
EPI_ISL_424522 hCoV-19/Iceland/500/2020 Europe / Iceland / Reykjavik 2020-03-21 The National University Hospital of Iceland deCODE genetics Daniel F Gudbjartsson; Agnar Helgason; Hakon Jonsson; Olafur T Magnusson; Pall Melsted; Gudmundur L Norddahl; Jona Saemundsdottir; Asgeir Sigurdsson; Patrick Sulem; Arna B Agustsdottir; Berglind Eiriksdottir; Run Fridriksdottir; Elisabet E Gardarsdottir; Gudmundur Georgsson; Olafia S Gretarsdottir; Kjartan R Gudmundsson; Thora R Gunnarsdottir; Arnaldur Gylfason; Hilma Holm; Brynjar O Jensson; Aslaug Jonasdottir; Kamilla S Josefsdottir; Thordur Kristjansson; Droplaug N Magnusdottir; Louise le Roux; Gudrun Sigmundsdottir; Gardar Sveinbjornsson; Kristin E Sveinsdottir; Maney Sveinsdottir; Emil A Thorarensen; Bjarni Thorbjornsson; Gisli Masson; Ingileif Jonsdottir; Alma Moller; Thorolfur Gudnason; Karl G Kristinsson; Unnur Thorsteinsdottir; Kari Stefansson
EPI_ISL_424523 hCoV-19/Iceland/501/2020 Europe / Iceland / Reykjavik 2020-03-21 The National University Hospital of Iceland deCODE genetics Daniel F Gudbjartsson; Agnar Helgason; Hakon Jonsson; Olafur T Magnusson; Pall Melsted; Gudmundur L Norddahl; Jona Saemundsdottir; Asgeir Sigurdsson; Patrick Sulem; Arna B Agustsdottir; Berglind Eiriksdottir; Run Fridriksdottir; Elisabet E Gardarsdottir; Gudmundur Georgsson; Olafia S Gretarsdottir; Kjartan R Gudmundsson; Thora R Gunnarsdottir; Arnaldur Gylfason; Hilma Holm; Brynjar O Jensson; Aslaug Jonasdottir; Kamilla S Josefsdottir; Thordur Kristjansson; Droplaug N Magnusdottir; Louise le Roux; Gudrun Sigmundsdottir; Gardar Sveinbjornsson; Kristin E Sveinsdottir; Maney Sveinsdottir; Emil A Thorarensen; Bjarni Thorbjornsson; Gisli Masson; Ingileif Jonsdottir; Alma Moller; Thorolfur Gudnason; Karl G Kristinsson; Unnur Thorsteinsdottir; Kari Stefansson
EPI_ISL_424524 hCoV-19/Iceland/502/2020 Europe / Iceland / Reykjavik 2020-03-21 The National University Hospital of Iceland deCODE genetics Daniel F Gudbjartsson; Agnar Helgason; Hakon Jonsson; Olafur T Magnusson; Pall Melsted; Gudmundur L Norddahl; Jona Saemundsdottir; Asgeir Sigurdsson; Patrick Sulem; Arna B Agustsdottir; Berglind Eiriksdottir; Run Fridriksdottir; Elisabet E Gardarsdottir; Gudmundur Georgsson; Olafia S Gretarsdottir; Kjartan R Gudmundsson; Thora R Gunnarsdottir; Arnaldur Gylfason; Hilma Holm; Brynjar O Jensson; Aslaug Jonasdottir; Kamilla S Josefsdottir; Thordur Kristjansson; Droplaug N Magnusdottir; Louise le Roux; Gudrun Sigmundsdottir; Gardar Sveinbjornsson; Kristin E Sveinsdottir; Maney Sveinsdottir; Emil A Thorarensen; Bjarni Thorbjornsson; Gisli Masson; Ingileif Jonsdottir; Alma Moller; Thorolfur Gudnason; Karl G Kristinsson; Unnur Thorsteinsdottir; Kari Stefansson
EPI_ISL_424525 hCoV-19/Iceland/503/2020 Europe / Iceland / Reykjavik 2020-03-21 The National University Hospital of Iceland deCODE genetics Daniel F Gudbjartsson; Agnar Helgason; Hakon Jonsson; Olafur T Magnusson; Pall Melsted; Gudmundur L Norddahl; Jona Saemundsdottir; Asgeir Sigurdsson; Patrick Sulem; Arna B Agustsdottir; Berglind Eiriksdottir; Run Fridriksdottir; Elisabet E Gardarsdottir; Gudmundur Georgsson; Olafia S Gretarsdottir; Kjartan R Gudmundsson; Thora R Gunnarsdottir; Arnaldur Gylfason; Hilma Holm; Brynjar O Jensson; Aslaug Jonasdottir; Kamilla S Josefsdottir; Thordur Kristjansson; Droplaug N Magnusdottir; Louise le Roux; Gudrun Sigmundsdottir; Gardar Sveinbjornsson; Kristin E Sveinsdottir; Maney Sveinsdottir; Emil A Thorarensen; Bjarni Thorbjornsson; Gisli Masson; Ingileif Jonsdottir; Alma Moller; Thorolfur Gudnason; Karl G Kristinsson; Unnur Thorsteinsdottir; Kari Stefansson
EPI_ISL_424526 hCoV-19/Iceland/504/2020 Europe / Iceland / Reykjavik 2020-03-21 The National

University Hospital of Iceland  deCODE genetics Daniel F Gudbjartsson; Agnar Helgason; Hakon Jonsson; Olafur T Magnusson; Pall Melsted; Gudmundur L Norddahl; Jona Saemundsdottir; Asgeir Sigurdsson; Patrick Sulem; Arna B Agustsdottir; Berglind Eiriksdottir; Run Fridriksdottir; Elisabet E Gardarsdottir; Gudmundur Georgsson; Olafia S Gretarsdottir; Kjartan R Gudmundsson; Thora R Gunnarsdottir; Arnaldur Gylfason; Hilma Holm; Brynjar O Jensson; Aslaug Jonasdottir; Kamilla S Josefsdottir; Thordur Kristjansson; Droplaug N Magnusdottir; Louise le Roux; Gudrun Sigmundsdottir; Gardar Sveinbjornsson; Kristin E Sveinsdottir; Maney Sveinsdottir; Emil A Thorarensen; Bjarni Thorbjornsson; Gisli Masson; Ingileif Jonsdottir; Alma Moller; Thorolfur Gudnason; Karl G Kristinsson; Unnur Thorsteinsdottir; Kari Stefansson
EPI_ISL_424527  hCoV-19/Iceland/505/2020  Europe / Iceland / Reykjavik  2020-03-21  The National University Hospital of Iceland  deCODE genetics Daniel F Gudbjartsson; Agnar Helgason; Hakon Jonsson; Olafur T Magnusson; Pall Melsted; Gudmundur L Norddahl; Jona Saemundsdottir; Asgeir Sigurdsson; Patrick Sulem; Arna B Agustsdottir; Berglind Eiriksdottir; Run Fridriksdottir; Elisabet E Gardarsdottir; Gudmundur Georgsson; Olafia S Gretarsdottir; Kjartan R Gudmundsson; Thora R Gunnarsdottir; Arnaldur Gylfason; Hilma Holm; Brynjar O Jensson; Aslaug Jonasdottir; Kamilla S Josefsdottir; Thordur Kristjansson; Droplaug N Magnusdottir; Louise le Roux; Gudrun Sigmundsdottir; Gardar Sveinbjornsson; Kristin E Sveinsdottir; Maney Sveinsdottir; Emil A Thorarensen; Bjarni Thorbjornsson; Gisli Masson; Ingileif Jonsdottir; Alma Moller; Thorolfur Gudnason; Karl G Kristinsson; Unnur Thorsteinsdottir; Kari Stefansson
EPI_ISL_424528  hCoV-19/Iceland/506/2020  Europe / Iceland / Reykjavik  2020-03-21  The National University Hospital of Iceland  deCODE genetics Daniel F Gudbjartsson; Agnar Helgason; Hakon Jonsson; Olafur T Magnusson; Pall Melsted; Gudmundur L Norddahl; Jona Saemundsdottir; Asgeir Sigurdsson; Patrick Sulem; Arna B Agustsdottir; Berglind Eiriksdottir; Run Fridriksdottir; Elisabet E Gardarsdottir; Gudmundur Georgsson; Olafia S Gretarsdottir; Kjartan R Gudmundsson; Thora R Gunnarsdottir; Arnaldur Gylfason; Hilma Holm; Brynjar O Jensson; Aslaug Jonasdottir; Kamilla S Josefsdottir; Thordur Kristjansson; Droplaug N Magnusdottir; Louise le Roux; Gudrun Sigmundsdottir; Gardar Sveinbjornsson; Kristin E Sveinsdottir; Maney Sveinsdottir; Emil A Thorarensen; Bjarni Thorbjornsson; Gisli Masson; Ingileif Jonsdottir; Alma Moller; Thorolfur Gudnason; Karl G Kristinsson; Unnur Thorsteinsdottir; Kari Stefansson
EPI_ISL_424529  hCoV-19/Iceland/507/2020  Europe / Iceland / Reykjavik  2020-03-20  The National University Hospital of Iceland  deCODE genetics Daniel F Gudbjartsson; Agnar Helgason; Hakon Jonsson; Olafur T Magnusson; Pall Melsted; Gudmundur L Norddahl; Jona Saemundsdottir; Asgeir Sigurdsson; Patrick Sulem; Arna B Agustsdottir; Berglind Eiriksdottir; Run Fridriksdottir; Elisabet E Gardarsdottir; Gudmundur Georgsson; Olafia S Gretarsdottir; Kjartan R Gudmundsson; Thora R Gunnarsdottir; Arnaldur Gylfason; Hilma Holm; Brynjar O Jensson; Aslaug Jonasdottir; Kamilla S Josefsdottir; Thordur Kristjansson; Droplaug N Magnusdottir; Louise le Roux; Gudrun Sigmundsdottir; Gardar Sveinbjornsson; Kristin E Sveinsdottir; Maney Sveinsdottir; Emil A Thorarensen; Bjarni Thorbjornsson; Gisli Masson; Ingileif Jonsdottir; Alma Moller; Thorolfur Gudnason; Karl G Kristinsson; Unnur Thorsteinsdottir; Kari Stefansson
EPI_ISL_424530  hCoV-19/Iceland/508/2020  Europe / Iceland / Reykjavik  2020-03-20  The National University Hospital of Iceland  deCODE genetics Daniel F Gudbjartsson; Agnar Helgason; Hakon Jonsson; Olafur T Magnusson; Pall Melsted; Gudmundur L Norddahl; Jona Saemundsdottir; Asgeir Sigurdsson; Patrick Sulem; Arna B Agustsdottir; Berglind Eiriksdottir; Run Fridriksdottir; Elisabet E Gardarsdottir; Gudmundur Georgsson; Olafia S Gretarsdottir; Kjartan R Gudmundsson; Thora R Gunnarsdottir; Arnaldur Gylfason; Hilma Holm; Brynjar O Jensson; Aslaug Jonasdottir; Kamilla S Josefsdottir; Thordur Kristjansson; Droplaug N Magnusdottir; Louise le Roux; Gudrun Sigmundsdottir; Gardar Sveinbjornsson; Kristin E Sveinsdottir; Maney Sveinsdottir; Emil A Thorarensen; Bjarni Thorbjornsson; Gisli Masson; Ingileif Jonsdottir; Alma Moller; Thorolfur Gudnason; Karl G Kristinsson; Unnur Thorsteinsdottir; Kari Stefansson
EPI_ISL_424531  hCoV-19/Iceland/509/2020  Europe / Iceland / Reykjavik  2020-03-20  The National University Hospital of Iceland  deCODE genetics Daniel F Gudbjartsson; Agnar Helgason; Hakon Jonsson; Olafur T Magnusson; Pall Melsted; Gudmundur L Norddahl; Jona Saemundsdottir; Asgeir Sigurdsson; Patrick Sulem; Arna B Agustsdottir; Berglind Eiriksdottir; Run Fridriksdottir; Elisabet E Gardarsdottir; Gudmundur Georgsson; Olafia S Gretarsdottir; Kjartan R Gudmundsson; Thora R Gunnarsdottir; Arnaldur Gylfason; Hilma Holm; Brynjar O Jensson; Aslaug Jonasdottir; Kamilla S Josefsdottir; Thordur Kristjansson; Droplaug N Magnusdottir; Louise le Roux; Gudrun Sigmundsdottir; Gardar Sveinbjornsson; Kristin E Sveinsdottir; Maney Sveinsdottir; Emil A Thorarensen; Bjarni Thorbjornsson; Gisli Masson; Ingileif Jonsdottir; Alma Moller; Thorolfur Gudnason; Karl G Kristinsson; Unnur Thorsteinsdottir; Kari Stefansson
EPI_ISL_424532  hCoV-19/Iceland/510/2020  Europe / Iceland / Reykjavik  2020-03-20  The National University Hospital of Iceland  deCODE genetics Daniel F Gudbjartsson; Agnar Helgason; Hakon Jonsson; Olafur T Magnusson; Pall Melsted; Gudmundur L Norddahl; Jona Saemundsdottir; Asgeir Sigurdsson; Patrick Sulem; Arna B Agustsdottir; Berglind Eiriksdottir; Run Fridriksdottir; Elisabet E Gardarsdottir; Gudmundur Georgsson; Olafia S Gretarsdottir; Kjartan R Gudmundsson; Thora R Gunnarsdottir; Arnaldur Gylfason; Hilma Holm; Brynjar O Jensson; Aslaug Jonasdottir; Kamilla S Josefsdottir; Thordur Kristjansson; Droplaug N Magnusdottir; Louise le Roux; Gudrun Sigmundsdottir; Gardar Sveinbjornsson; Kristin E Sveinsdottir; Maney Sveinsdottir; Emil A Thorarensen; Bjarni Thorbjornsson; Gisli Masson; Ingileif Jonsdottir; Alma Moller; Thorolfur Gudnason; Karl G Kristinsson; Unnur Thorsteinsdottir; Kari Stefansson
EPI_ISL_424533  hCoV-19/Iceland/511/2020  Europe / Iceland / Reykjavik  2020-03-20  deCODE genetics  deCODE genetics Daniel F Gudbjartsson; Agnar Helgason; Hakon Jonsson; Olafur T Magnusson; Pall Melsted; Gudmundur L Norddahl; Jona Saemundsdottir; Asgeir Sigurdsson; Patrick Sulem; Arna B Agustsdottir; Berglind Eiriksdottir; Run Fridriksdottir; Elisabet E Gardarsdottir; Gudmundur Georgsson; Olafia S Gretarsdottir; Kjartan R Gudmundsson; Thora R Gunnarsdottir; Arnaldur Gylfason; Hilma Holm; Brynjar O Jensson; Aslaug Jonasdottir; Kamilla S Josefsdottir; Thordur Kristjansson; Droplaug N Magnusdottir; Louise le Roux; Gudrun Sigmundsdottir; Gardar Sveinbjornsson; Kristin E Sveinsdottir; Maney Sveinsdottir; Emil A Thorarensen; Bjarni Thorbjornsson; Gisli Masson; Ingileif Jonsdottir; Alma Moller; Thorolfur Gudnason; Karl G Kristinsson; Unnur

Thorsteinsdottir; Kari Stefansson
EPI_ISL_424534 hCoV-19/Iceland/512/2020 Europe / Iceland / Reykjavik 2020-03 deCODE genetics deCODE genetics Daniel F Gudbjartsson; Agnar Helgason; Hakon Jonsson; Olafur T Magnusson; Pall Melsted; Gudmundur L Norddahl; Jona Saemundsdottir; Asgeir Sigurdsson; Patrick Sulem; Arna B Agustsdottir; Berglind Eiriksdottir; Run Fridriksdottir; Elisabet E Gardarsdottir; Gudmundur Georgsson; Olafia S Gretarsdottir; Kjartan R Gudmundsson; Thora R Gunnarsdottir; Arnaldur Gylfason; Hilma Holm; Brynjar O Jensson; Aslaug Jonasdottir; Kamilla S Josefsdottir; Thordur Kristjansson; Droplaug N Magnusdottir; Louise le Roux; Gudrun Sigmundsdottir; Gardar Sveinbjornsson; Kristin E Sveinsdottir; Maney Sveinsdottir; Emil A Thorarensen; Bjarni Thorbjornsson; Gisli Masson; Ingileif Jonsdottir; Alma Moller; Thorolfur Gudnason; Karl G Kristinsson; Unnur Thorsteinsdottir; Kari Stefansson
EPI_ISL_424535 hCoV-19/Iceland/513/2020 Europe / Iceland / Reykjavik 2020-03-21 deCODE genetics deCODE genetics Daniel F Gudbjartsson; Agnar Helgason; Hakon Jonsson; Olafur T Magnusson; Pall Melsted; Gudmundur L Norddahl; Jona Saemundsdottir; Asgeir Sigurdsson; Patrick Sulem; Arna B Agustsdottir; Berglind Eiriksdottir; Run Fridriksdottir; Elisabet E Gardarsdottir; Gudmundur Georgsson; Olafia S Gretarsdottir; Kjartan R Gudmundsson; Thora R Gunnarsdottir; Arnaldur Gylfason; Hilma Holm; Brynjar O Jensson; Aslaug Jonasdottir; Kamilla S Josefsdottir; Thordur Kristjansson; Droplaug N Magnusdottir; Louise le Roux; Gudrun Sigmundsdottir; Gardar Sveinbjornsson; Kristin E Sveinsdottir; Maney Sveinsdottir; Emil A Thorarensen; Bjarni Thorbjornsson; Gisli Masson; Ingileif Jonsdottir; Alma Moller; Thorolfur Gudnason; Karl G Kristinsson; Unnur Thorsteinsdottir; Kari Stefansson
EPI_ISL_424536 hCoV-19/Iceland/514/2020 Europe / Iceland / Reykjavik 2020-03-21 deCODE genetics deCODE genetics Daniel F Gudbjartsson; Agnar Helgason; Hakon Jonsson; Olafur T Magnusson; Pall Melsted; Gudmundur L Norddahl; Jona Saemundsdottir; Asgeir Sigurdsson; Patrick Sulem; Arna B Agustsdottir; Berglind Eiriksdottir; Run Fridriksdottir; Elisabet E Gardarsdottir; Gudmundur Georgsson; Olafia S Gretarsdottir; Kjartan R Gudmundsson; Thora R Gunnarsdottir; Arnaldur Gylfason; Hilma Holm; Brynjar O Jensson; Aslaug Jonasdottir; Kamilla S Josefsdottir; Thordur Kristjansson; Droplaug N Magnusdottir; Louise le Roux; Gudrun Sigmundsdottir; Gardar Sveinbjornsson; Kristin E Sveinsdottir; Maney Sveinsdottir; Emil A Thorarensen; Bjarni Thorbjornsson; Gisli Masson; Ingileif Jonsdottir; Alma Moller; Thorolfur Gudnason; Karl G Kristinsson; Unnur Thorsteinsdottir; Kari Stefansson
EPI_ISL_424537 hCoV-19/Iceland/515/2020 Europe / Iceland / Reykjavik 2020-03 deCODE genetics deCODE genetics Daniel F Gudbjartsson; Agnar Helgason; Hakon Jonsson; Olafur T Magnusson; Pall Melsted; Gudmundur L Norddahl; Jona Saemundsdottir; Asgeir Sigurdsson; Patrick Sulem; Arna B Agustsdottir; Berglind Eiriksdottir; Run Fridriksdottir; Elisabet E Gardarsdottir; Gudmundur Georgsson; Olafia S Gretarsdottir; Kjartan R Gudmundsson; Thora R Gunnarsdottir; Arnaldur Gylfason; Hilma Holm; Brynjar O Jensson; Aslaug Jonasdottir; Kamilla S Josefsdottir; Thordur Kristjansson; Droplaug N Magnusdottir; Louise le Roux; Gudrun Sigmundsdottir; Gardar Sveinbjornsson; Kristin E Sveinsdottir; Maney Sveinsdottir; Emil A Thorarensen; Bjarni Thorbjornsson; Gisli Masson; Ingileif Jonsdottir; Alma Moller; Thorolfur Gudnason; Karl G Kristinsson; Unnur Thorsteinsdottir; Kari Stefansson
EPI_ISL_424538 hCoV-19/Iceland/516/2020 Europe / Iceland / Reykjavik 2020-03-24 deCODE genetics deCODE genetics Daniel F Gudbjartsson; Agnar Helgason; Hakon Jonsson; Olafur T Magnusson; Pall Melsted; Gudmundur L Norddahl; Jona Saemundsdottir; Asgeir Sigurdsson; Patrick Sulem; Arna B Agustsdottir; Berglind Eiriksdottir; Run Fridriksdottir; Elisabet E Gardarsdottir; Gudmundur Georgsson; Olafia S Gretarsdottir; Kjartan R Gudmundsson; Thora R Gunnarsdottir; Arnaldur Gylfason; Hilma Holm; Brynjar O Jensson; Aslaug Jonasdottir; Kamilla S Josefsdottir; Thordur Kristjansson; Droplaug N Magnusdottir; Louise le Roux; Gudrun Sigmundsdottir; Gardar Sveinbjornsson; Kristin E Sveinsdottir; Maney Sveinsdottir; Emil A Thorarensen; Bjarni Thorbjornsson; Gisli Masson; Ingileif Jonsdottir; Alma Moller; Thorolfur Gudnason; Karl G Kristinsson; Unnur Thorsteinsdottir; Kari Stefansson
EPI_ISL_424540 hCoV-19/Iceland/519/2020 Europe / Iceland / Reykjavik 2020-03 deCODE genetics deCODE genetics Daniel F Gudbjartsson; Agnar Helgason; Hakon Jonsson; Olafur T Magnusson; Pall Melsted; Gudmundur L Norddahl; Jona Saemundsdottir; Asgeir Sigurdsson; Patrick Sulem; Arna B Agustsdottir; Berglind Eiriksdottir; Run Fridriksdottir; Elisabet E Gardarsdottir; Gudmundur Georgsson; Olafia S Gretarsdottir; Kjartan R Gudmundsson; Thora R Gunnarsdottir; Arnaldur Gylfason; Hilma Holm; Brynjar O Jensson; Aslaug Jonasdottir; Kamilla S Josefsdottir; Thordur Kristjansson; Droplaug N Magnusdottir; Louise le Roux; Gudrun Sigmundsdottir; Gardar Sveinbjornsson; Kristin E Sveinsdottir; Maney Sveinsdottir; Emil A Thorarensen; Bjarni Thorbjornsson; Gisli Masson; Ingileif Jonsdottir; Alma Moller; Thorolfur Gudnason; Karl G Kristinsson; Unnur Thorsteinsdottir; Kari Stefansson
EPI_ISL_424541 hCoV-19/Iceland/520/2020 Europe / Iceland / Reykjavik 2020-03 deCODE genetics deCODE genetics Daniel F Gudbjartsson; Agnar Helgason; Hakon Jonsson; Olafur T Magnusson; Pall Melsted; Gudmundur L Norddahl; Jona Saemundsdottir; Asgeir Sigurdsson; Patrick Sulem; Arna B Agustsdottir; Berglind Eiriksdottir; Run Fridriksdottir; Elisabet E Gardarsdottir; Gudmundur Georgsson; Olafia S Gretarsdottir; Kjartan R Gudmundsson; Thora R Gunnarsdottir; Arnaldur Gylfason; Hilma Holm; Brynjar O Jensson; Aslaug Jonasdottir; Kamilla S Josefsdottir; Thordur Kristjansson; Droplaug N Magnusdottir; Louise le Roux; Gudrun Sigmundsdottir; Gardar Sveinbjornsson; Kristin E Sveinsdottir; Maney Sveinsdottir; Emil A Thorarensen; Bjarni Thorbjornsson; Gisli Masson; Ingileif Jonsdottir; Alma Moller; Thorolfur Gudnason; Karl G Kristinsson; Unnur Thorsteinsdottir; Kari Stefansson
EPI_ISL_424542 hCoV-19/Iceland/521/2020 Europe / Iceland / Reykjavik 2020-03 deCODE genetics deCODE genetics Daniel F Gudbjartsson; Agnar Helgason; Hakon Jonsson; Olafur T Magnusson; Pall Melsted; Gudmundur L Norddahl; Jona Saemundsdottir; Asgeir Sigurdsson; Patrick Sulem; Arna B Agustsdottir; Berglind Eiriksdottir; Run Fridriksdottir; Elisabet E Gardarsdottir; Gudmundur Georgsson; Olafia S Gretarsdottir; Kjartan R Gudmundsson; Thora R Gunnarsdottir; Arnaldur Gylfason; Hilma Holm; Brynjar O Jensson; Aslaug Jonasdottir; Kamilla S Josefsdottir; Thordur Kristjansson; Droplaug N Magnusdottir; Louise le Roux; Gudrun

Sigmundsdottir; Gardar Sveinbjornsson; Kristin E Sveinsdottir; Maney Sveinsdottir; Emil A Thorarensen; Bjarni Thorbjornsson; Gisli Masson; Ingileif Jonsdottir; Alma Moller; Thorolfur Gudnason; Karl G Kristinsson; Unnur Thorsteinsdottir; Kari Stefansson
EPI_ISL_424545 hCoV-19/Iceland/524/2020 Europe / Iceland / Reykjavik 2020-03-25 deCODE genetics deCODE genetics Daniel F Gudbjartsson; Agnar Helgason; Hakon Jonsson; Olafur T Magnusson; Pall Melsted; Gudmundur L Norddahl; Jona Saemundsdottir; Asgeir Sigurdsson; Patrick Sulem; Arna B Agustsdottir; Berglind Eiriksdottir; Run Fridriksdottir; Elisabet E Gardarsdottir; Gudmundur Georgsson; Olafia S Gretarsdottir; Kjartan R Gudmundsson; Thora R Gunnarsdottir; Arnaldur Gylfason; Hilma Holm; Brynjar O Jensson; Aslaug Jonasdottir; Kamilla S Josefsdottir; Thordur Kristjansson; Droplaug N Magnusdottir; Louise le Roux; Gudrun Sigmundsdottir; Gardar Sveinbjornsson; Kristin E Sveinsdottir; Maney Sveinsdottir; Emil A Thorarensen; Bjarni Thorbjornsson; Gisli Masson; Ingileif Jonsdottir; Alma Moller; Thorolfur Gudnason; Karl G Kristinsson; Unnur Thorsteinsdottir; Kari Stefansson
EPI_ISL_424548 hCoV-19/Iceland/527/2020 Europe / Iceland / Reykjavik 2020-03 deCODE genetics deCODE genetics Daniel F Gudbjartsson; Agnar Helgason; Hakon Jonsson; Olafur T Magnusson; Pall Melsted; Gudmundur L Norddahl; Jona Saemundsdottir; Asgeir Sigurdsson; Patrick Sulem; Arna B Agustsdottir; Berglind Eiriksdottir; Run Fridriksdottir; Elisabet E Gardarsdottir; Gudmundur Georgsson; Olafia S Gretarsdottir; Kjartan R Gudmundsson; Thora R Gunnarsdottir; Arnaldur Gylfason; Hilma Holm; Brynjar O Jensson; Aslaug Jonasdottir; Kamilla S Josefsdottir; Thordur Kristjansson; Droplaug N Magnusdottir; Louise le Roux; Gudrun Sigmundsdottir; Gardar Sveinbjornsson; Kristin E Sveinsdottir; Maney Sveinsdottir; Emil A Thorarensen; Bjarni Thorbjornsson; Gisli Masson; Ingileif Jonsdottir; Alma Moller; Thorolfur Gudnason; Karl G Kristinsson; Unnur Thorsteinsdottir; Kari Stefansson
EPI_ISL_424549 hCoV-19/Iceland/528/2020 Europe / Iceland / Reykjavik 2020-03 deCODE genetics deCODE genetics Daniel F Gudbjartsson; Agnar Helgason; Hakon Jonsson; Olafur T Magnusson; Pall Melsted; Gudmundur L Norddahl; Jona Saemundsdottir; Asgeir Sigurdsson; Patrick Sulem; Arna B Agustsdottir; Berglind Eiriksdottir; Run Fridriksdottir; Elisabet E Gardarsdottir; Gudmundur Georgsson; Olafia S Gretarsdottir; Kjartan R Gudmundsson; Thora R Gunnarsdottir; Arnaldur Gylfason; Hilma Holm; Brynjar O Jensson; Aslaug Jonasdottir; Kamilla S Josefsdottir; Thordur Kristjansson; Droplaug N Magnusdottir; Louise le Roux; Gudrun Sigmundsdottir; Gardar Sveinbjornsson; Kristin E Sveinsdottir; Maney Sveinsdottir; Emil A Thorarensen; Bjarni Thorbjornsson; Gisli Masson; Ingileif Jonsdottir; Alma Moller; Thorolfur Gudnason; Karl G Kristinsson; Unnur Thorsteinsdottir; Kari Stefansson
EPI_ISL_424550 hCoV-19/Iceland/529/2020 Europe / Iceland / Reykjavik 2020-03 deCODE genetics deCODE genetics Daniel F Gudbjartsson; Agnar Helgason; Hakon Jonsson; Olafur T Magnusson; Pall Melsted; Gudmundur L Norddahl; Jona Saemundsdottir; Asgeir Sigurdsson; Patrick Sulem; Arna B Agustsdottir; Berglind Eiriksdottir; Run Fridriksdottir; Elisabet E Gardarsdottir; Gudmundur Georgsson; Olafia S Gretarsdottir; Kjartan R Gudmundsson; Thora R Gunnarsdottir; Arnaldur Gylfason; Hilma Holm; Brynjar O Jensson; Aslaug Jonasdottir; Kamilla S Josefsdottir; Thordur Kristjansson; Droplaug N Magnusdottir; Louise le Roux; Gudrun Sigmundsdottir; Gardar Sveinbjornsson; Kristin E Sveinsdottir; Maney Sveinsdottir; Emil A Thorarensen; Bjarni Thorbjornsson; Gisli Masson; Ingileif Jonsdottir; Alma Moller; Thorolfur Gudnason; Karl G Kristinsson; Unnur Thorsteinsdottir; Kari Stefansson
EPI_ISL_424551 hCoV-19/Iceland/530/2020 Europe / Iceland / Reykjavik 2020-03-25 deCODE genetics deCODE genetics Daniel F Gudbjartsson; Agnar Helgason; Hakon Jonsson; Olafur T Magnusson; Pall Melsted; Gudmundur L Norddahl; Jona Saemundsdottir; Asgeir Sigurdsson; Patrick Sulem; Arna B Agustsdottir; Berglind Eiriksdottir; Run Fridriksdottir; Elisabet E Gardarsdottir; Gudmundur Georgsson; Olafia S Gretarsdottir; Kjartan R Gudmundsson; Thora R Gunnarsdottir; Arnaldur Gylfason; Hilma Holm; Brynjar O Jensson; Aslaug Jonasdottir; Kamilla S Josefsdottir; Thordur Kristjansson; Droplaug N Magnusdottir; Louise le Roux; Gudrun Sigmundsdottir; Gardar Sveinbjornsson; Kristin E Sveinsdottir; Maney Sveinsdottir; Emil A Thorarensen; Bjarni Thorbjornsson; Gisli Masson; Ingileif Jonsdottir; Alma Moller; Thorolfur Gudnason; Karl G Kristinsson; Unnur Thorsteinsdottir; Kari Stefansson
EPI_ISL_424555 hCoV-19/Iceland/535/2020 Europe / Iceland / Reykjavik 2020-03-29 The National University Hospital of Iceland deCODE genetics Daniel F Gudbjartsson; Agnar Helgason; Hakon Jonsson; Olafur T Magnusson; Pall Melsted; Gudmundur L Norddahl; Jona Saemundsdottir; Asgeir Sigurdsson; Patrick Sulem; Arna B Agustsdottir; Berglind Eiriksdottir; Run Fridriksdottir; Elisabet E Gardarsdottir; Gudmundur Georgsson; Olafia S Gretarsdottir; Kjartan R Gudmundsson; Thora R Gunnarsdottir; Arnaldur Gylfason; Hilma Holm; Brynjar O Jensson; Aslaug Jonasdottir; Kamilla S Josefsdottir; Thordur Kristjansson; Droplaug N Magnusdottir; Louise le Roux; Gudrun Sigmundsdottir; Gardar Sveinbjornsson; Kristin E Sveinsdottir; Maney Sveinsdottir; Emil A Thorarensen; Bjarni Thorbjornsson; Gisli Masson; Ingileif Jonsdottir; Alma Moller; Thorolfur Gudnason; Karl G Kristinsson; Unnur Thorsteinsdottir; Kari Stefansson
EPI_ISL_424556 hCoV-19/Iceland/536/2020 Europe / Iceland / Reykjavik 2020-03-29 The National University Hospital of Iceland deCODE genetics Daniel F Gudbjartsson; Agnar Helgason; Hakon Jonsson; Olafur T Magnusson; Pall Melsted; Gudmundur L Norddahl; Jona Saemundsdottir; Asgeir Sigurdsson; Patrick Sulem; Arna B Agustsdottir; Berglind Eiriksdottir; Run Fridriksdottir; Elisabet E Gardarsdottir; Gudmundur Georgsson; Olafia S Gretarsdottir; Kjartan R Gudmundsson; Thora R Gunnarsdottir; Arnaldur Gylfason; Hilma Holm; Brynjar O Jensson; Aslaug Jonasdottir; Kamilla S Josefsdottir; Thordur Kristjansson; Droplaug N Magnusdottir; Louise le Roux; Gudrun Sigmundsdottir; Gardar Sveinbjornsson; Kristin E Sveinsdottir; Maney Sveinsdottir; Emil A Thorarensen; Bjarni Thorbjornsson; Gisli Masson; Ingileif Jonsdottir; Alma Moller; Thorolfur Gudnason; Karl G Kristinsson; Unnur Thorsteinsdottir; Kari Stefansson
EPI_ISL_424557 hCoV-19/Iceland/537/2020 Europe / Iceland / Reykjavik 2020-03-27 The National University Hospital of Iceland deCODE genetics Daniel F Gudbjartsson; Agnar Helgason; Hakon Jonsson; Olafur T Magnusson; Pall Melsted; Gudmundur L Norddahl; Jona Saemundsdottir; Asgeir Sigurdsson; Patrick Sulem; Arna B Agustsdottir; Berglind Eiriksdottir; Run Fridriksdottir; Elisabet E Gardarsdottir; Gudmundur Georgsson; Olafia

S Gretarsdottir; Kjartan R Gudmundsson; Thora R Gunnarsdottir; Arnaldur Gylfason; Hilma Holm; Brynjar O Jensson; Aslaug Jonasdottir; Kamilla S Josefsdottir; Thordur Kristjansson; Droplaug N Magnusdottir; Louise le Roux; Gudrun Sigmundsdottir; Gardar Sveinbjornsson; Kristin E Sveinsdottir; Maney Sveinsdottir; Emil A Thorarensen; Bjarni Thorbjornsson; Gisli Masson; Ingileif Jonsdottir; Alma Moller; Thorolfur Gudnason; Karl G Kristinsson; Unnur Thorsteinsdottir; Kari Stefansson
EPI_ISL_424558 hCoV-19/Iceland/538/2020 Europe / Iceland / Reykjavik 2020-03-27 The National University Hospital of Iceland deCODE genetics Daniel F Gudbjartsson; Agnar Helgason; Hakon Jonsson; Olafur T Magnusson; Pall Melsted; Gudmundur L Norddahl; Jona Saemundsdottir; Asgeir Sigurdsson; Patrick Sulem; Arna B Agustsdottir; Berglind Eiriksdottir; Run Fridriksdottir; Elisabet E Gardarsdottir; Gudmundur Georgsson; Olafia S Gretarsdottir; Kjartan R Gudmundsson; Thora R Gunnarsdottir; Arnaldur Gylfason; Hilma Holm; Brynjar O Jensson; Aslaug Jonasdottir; Kamilla S Josefsdottir; Thordur Kristjansson; Droplaug N Magnusdottir; Louise le Roux; Gudrun Sigmundsdottir; Gardar Sveinbjornsson; Kristin E Sveinsdottir; Maney Sveinsdottir; Emil A Thorarensen; Bjarni Thorbjornsson; Gisli Masson; Ingileif Jonsdottir; Alma Moller; Thorolfur Gudnason; Karl G Kristinsson; Unnur Thorsteinsdottir; Kari Stefansson
EPI_ISL_424559 hCoV-19/Iceland/539/2020 Europe / Iceland / Reykjavik 2020-03-27 The National University Hospital of Iceland deCODE genetics Daniel F Gudbjartsson; Agnar Helgason; Hakon Jonsson; Olafur T Magnusson; Pall Melsted; Gudmundur L Norddahl; Jona Saemundsdottir; Asgeir Sigurdsson; Patrick Sulem; Arna B Agustsdottir; Berglind Eiriksdottir; Run Fridriksdottir; Elisabet E Gardarsdottir; Gudmundur Georgsson; Olafia S Gretarsdottir; Kjartan R Gudmundsson; Thora R Gunnarsdottir; Arnaldur Gylfason; Hilma Holm; Brynjar O Jensson; Aslaug Jonasdottir; Kamilla S Josefsdottir; Thordur Kristjansson; Droplaug N Magnusdottir; Louise le Roux; Gudrun Sigmundsdottir; Gardar Sveinbjornsson; Kristin E Sveinsdottir; Maney Sveinsdottir; Emil A Thorarensen; Bjarni Thorbjornsson; Gisli Masson; Ingileif Jonsdottir; Alma Moller; Thorolfur Gudnason; Karl G Kristinsson; Unnur Thorsteinsdottir; Kari Stefansson
EPI_ISL_424560 hCoV-19/Iceland/540/2020 Europe / Iceland / Reykjavik 2020-03-27 deCODE genetics deCODE genetics Daniel F Gudbjartsson; Agnar Helgason; Hakon Jonsson; Olafur T Magnusson; Pall Melsted; Gudmundur L Norddahl; Jona Saemundsdottir; Asgeir Sigurdsson; Patrick Sulem; Arna B Agustsdottir; Berglind Eiriksdottir; Run Fridriksdottir; Elisabet E Gardarsdottir; Gudmundur Georgsson; Olafia S Gretarsdottir; Kjartan R Gudmundsson; Thora R Gunnarsdottir; Arnaldur Gylfason; Hilma Holm; Brynjar O Jensson; Aslaug Jonasdottir; Kamilla S Josefsdottir; Thordur Kristjansson; Droplaug N Magnusdottir; Louise le Roux; Gudrun Sigmundsdottir; Gardar Sveinbjornsson; Kristin E Sveinsdottir; Maney Sveinsdottir; Emil A Thorarensen; Bjarni Thorbjornsson; Gisli Masson; Ingileif Jonsdottir; Alma Moller; Thorolfur Gudnason; Karl G Kristinsson; Unnur Thorsteinsdottir; Kari Stefansson
EPI_ISL_424562 hCoV-19/Iceland/542/2020 Europe / Iceland / Reykjavik 2020-03-26 deCODE genetics deCODE genetics Daniel F Gudbjartsson; Agnar Helgason; Hakon Jonsson; Olafur T Magnusson; Pall Melsted; Gudmundur L Norddahl; Jona Saemundsdottir; Asgeir Sigurdsson; Patrick Sulem; Arna B Agustsdottir; Berglind Eiriksdottir; Run Fridriksdottir; Elisabet E Gardarsdottir; Gudmundur Georgsson; Olafia S Gretarsdottir; Kjartan R Gudmundsson; Thora R Gunnarsdottir; Arnaldur Gylfason; Hilma Holm; Brynjar O Jensson; Aslaug Jonasdottir; Kamilla S Josefsdottir; Thordur Kristjansson; Droplaug N Magnusdottir; Louise le Roux; Gudrun Sigmundsdottir; Gardar Sveinbjornsson; Kristin E Sveinsdottir; Maney Sveinsdottir; Emil A Thorarensen; Bjarni Thorbjornsson; Gisli Masson; Ingileif Jonsdottir; Alma Moller; Thorolfur Gudnason; Karl G Kristinsson; Unnur Thorsteinsdottir; Kari Stefansson
EPI_ISL_424563 hCoV-19/Iceland/543/2020 Europe / Iceland / Reykjavik 2020-03-27 The National University Hospital of Iceland deCODE genetics Daniel F Gudbjartsson; Agnar Helgason; Hakon Jonsson; Olafur T Magnusson; Pall Melsted; Gudmundur L Norddahl; Jona Saemundsdottir; Asgeir Sigurdsson; Patrick Sulem; Arna B Agustsdottir; Berglind Eiriksdottir; Run Fridriksdottir; Elisabet E Gardarsdottir; Gudmundur Georgsson; Olafia S Gretarsdottir; Kjartan R Gudmundsson; Thora R Gunnarsdottir; Arnaldur Gylfason; Hilma Holm; Brynjar O Jensson; Aslaug Jonasdottir; Kamilla S Josefsdottir; Thordur Kristjansson; Droplaug N Magnusdottir; Louise le Roux; Gudrun Sigmundsdottir; Gardar Sveinbjornsson; Kristin E Sveinsdottir; Maney Sveinsdottir; Emil A Thorarensen; Bjarni Thorbjornsson; Gisli Masson; Ingileif Jonsdottir; Alma Moller; Thorolfur Gudnason; Karl G Kristinsson; Unnur Thorsteinsdottir; Kari Stefansson
EPI_ISL_424564 hCoV-19/Iceland/544/2020 Europe / Iceland / Reykjavik 2020-03-27 The National University Hospital of Iceland deCODE genetics Daniel F Gudbjartsson; Agnar Helgason; Hakon Jonsson; Olafur T Magnusson; Pall Melsted; Gudmundur L Norddahl; Jona Saemundsdottir; Asgeir Sigurdsson; Patrick Sulem; Arna B Agustsdottir; Berglind Eiriksdottir; Run Fridriksdottir; Elisabet E Gardarsdottir; Gudmundur Georgsson; Olafia S Gretarsdottir; Kjartan R Gudmundsson; Thora R Gunnarsdottir; Arnaldur Gylfason; Hilma Holm; Brynjar O Jensson; Aslaug Jonasdottir; Kamilla S Josefsdottir; Thordur Kristjansson; Droplaug N Magnusdottir; Louise le Roux; Gudrun Sigmundsdottir; Gardar Sveinbjornsson; Kristin E Sveinsdottir; Maney Sveinsdottir; Emil A Thorarensen; Bjarni Thorbjornsson; Gisli Masson; Ingileif Jonsdottir; Alma Moller; Thorolfur Gudnason; Karl G Kristinsson; Unnur Thorsteinsdottir; Kari Stefansson
EPI_ISL_424567 hCoV-19/Iceland/547/2020 Europe / Iceland / Reykjavik 2020-03-28 deCODE genetics deCODE genetics Daniel F Gudbjartsson; Agnar Helgason; Hakon Jonsson; Olafur T Magnusson; Pall Melsted; Gudmundur L Norddahl; Jona Saemundsdottir; Asgeir Sigurdsson; Patrick Sulem; Arna B Agustsdottir; Berglind Eiriksdottir; Run Fridriksdottir; Elisabet E Gardarsdottir; Gudmundur Georgsson; Olafia S Gretarsdottir; Kjartan R Gudmundsson; Thora R Gunnarsdottir; Arnaldur Gylfason; Hilma Holm; Brynjar O Jensson; Aslaug Jonasdottir; Kamilla S Josefsdottir; Thordur Kristjansson; Droplaug N Magnusdottir; Louise le Roux; Gudrun Sigmundsdottir; Gardar Sveinbjornsson; Kristin E Sveinsdottir; Maney Sveinsdottir; Emil A Thorarensen; Bjarni Thorbjornsson; Gisli Masson; Ingileif Jonsdottir; Alma Moller; Thorolfur Gudnason; Karl G Kristinsson; Unnur Thorsteinsdottir; Kari Stefansson
EPI_ISL_424568 hCoV-19/Iceland/548/2020 Europe / Iceland / Reykjavik 2020-03-28 deCODE genetics deCODE genetics Daniel F Gudbjartsson; Agnar Helgason; Hakon Jonsson; Olafur T Magnusson; Pall Melsted;

Gudmundur L Norddahl; Jona Saemundsdottir; Asgeir Sigurdsson; Patrick Sulem; Arna B Agustsdottir; Berglind Eiriksdottir; Run Fridriksdottir; Elisabet E Gardarsdottir; Gudmundur Georgsson; Olafia S Gretarsdottir; Kjartan R Gudmundsson; Thora R Gunnarsdottir; Arnaldur Gylfason; Hilma Holm; Brynjar O Jensson; Aslaug Jonasdottir; Kamilla S Josefsdottir; Thordur Kristjansson; Droplaug N Magnusdottir; Louise le Roux; Gudrun Sigmundsdottir; Gardar Sveinbjornsson; Kristin E Sveinsdottir; Maney Sveinsdottir; Emil A Thorarensen; Bjarni Thorbjornsson; Gisli Masson; Ingileif Jonsdottir; Alma Moller; Thorolfur Gudnason; Karl G Kristinsson; Unnur Thorsteinsdottir; Kari Stefansson
EPI_ISL_424570 hCoV-19/Iceland/550/2020 Europe / Iceland / Reykjavik 2020-03-28 deCODE genetics deCODE genetics Daniel F Gudbjartsson; Agnar Helgason; Hakon Jonsson; Olafur T Magnusson; Pall Melsted; Gudmundur L Norddahl; Jona Saemundsdottir; Asgeir Sigurdsson; Patrick Sulem; Arna B Agustsdottir; Berglind Eiriksdottir; Run Fridriksdottir; Elisabet E Gardarsdottir; Gudmundur Georgsson; Olafia S Gretarsdottir; Kjartan R Gudmundsson; Thora R Gunnarsdottir; Arnaldur Gylfason; Hilma Holm; Brynjar O Jensson; Aslaug Jonasdottir; Kamilla S Josefsdottir; Thordur Kristjansson; Droplaug N Magnusdottir; Louise le Roux; Gudrun Sigmundsdottir; Gardar Sveinbjornsson; Kristin E Sveinsdottir; Maney Sveinsdottir; Emil A Thorarensen; Bjarni Thorbjornsson; Gisli Masson; Ingileif Jonsdottir; Alma Moller; Thorolfur Gudnason; Karl G Kristinsson; Unnur Thorsteinsdottir; Kari Stefansson
EPI_ISL_424571 hCoV-19/Iceland/551/2020 Europe / Iceland / Reykjavik 2020-03-27 The National University Hospital of Iceland deCODE genetics Daniel F Gudbjartsson; Agnar Helgason; Hakon Jonsson; Olafur T Magnusson; Pall Melsted; Gudmundur L Norddahl; Jona Saemundsdottir; Asgeir Sigurdsson; Patrick Sulem; Arna B Agustsdottir; Berglind Eiriksdottir; Run Fridriksdottir; Elisabet E Gardarsdottir; Gudmundur Georgsson; Olafia S Gretarsdottir; Kjartan R Gudmundsson; Thora R Gunnarsdottir; Arnaldur Gylfason; Hilma Holm; Brynjar O Jensson; Aslaug Jonasdottir; Kamilla S Josefsdottir; Thordur Kristjansson; Droplaug N Magnusdottir; Louise le Roux; Gudrun Sigmundsdottir; Gardar Sveinbjornsson; Kristin E Sveinsdottir; Maney Sveinsdottir; Emil A Thorarensen; Bjarni Thorbjornsson; Gisli Masson; Ingileif Jonsdottir; Alma Moller; Thorolfur Gudnason; Karl G Kristinsson; Unnur Thorsteinsdottir; Kari Stefansson
EPI_ISL_424572 hCoV-19/Iceland/552/2020 Europe / Iceland / Reykjavik 2020-03-26 The National University Hospital of Iceland deCODE genetics Daniel F Gudbjartsson; Agnar Helgason; Hakon Jonsson; Olafur T Magnusson; Pall Melsted; Gudmundur L Norddahl; Jona Saemundsdottir; Asgeir Sigurdsson; Patrick Sulem; Arna B Agustsdottir; Berglind Eiriksdottir; Run Fridriksdottir; Elisabet E Gardarsdottir; Gudmundur Georgsson; Olafia S Gretarsdottir; Kjartan R Gudmundsson; Thora R Gunnarsdottir; Arnaldur Gylfason; Hilma Holm; Brynjar O Jensson; Aslaug Jonasdottir; Kamilla S Josefsdottir; Thordur Kristjansson; Droplaug N Magnusdottir; Louise le Roux; Gudrun Sigmundsdottir; Gardar Sveinbjornsson; Kristin E Sveinsdottir; Maney Sveinsdottir; Emil A Thorarensen; Bjarni Thorbjornsson; Gisli Masson; Ingileif Jonsdottir; Alma Moller; Thorolfur Gudnason; Karl G Kristinsson; Unnur Thorsteinsdottir; Kari Stefansson
EPI_ISL_424573 hCoV-19/Iceland/553/2020 Europe / Iceland / Reykjavik 2020-03-27 The National University Hospital of Iceland deCODE genetics Daniel F Gudbjartsson; Agnar Helgason; Hakon Jonsson; Olafur T Magnusson; Pall Melsted; Gudmundur L Norddahl; Jona Saemundsdottir; Asgeir Sigurdsson; Patrick Sulem; Arna B Agustsdottir; Berglind Eiriksdottir; Run Fridriksdottir; Elisabet E Gardarsdottir; Gudmundur Georgsson; Olafia S Gretarsdottir; Kjartan R Gudmundsson; Thora R Gunnarsdottir; Arnaldur Gylfason; Hilma Holm; Brynjar O Jensson; Aslaug Jonasdottir; Kamilla S Josefsdottir; Thordur Kristjansson; Droplaug N Magnusdottir; Louise le Roux; Gudrun Sigmundsdottir; Gardar Sveinbjornsson; Kristin E Sveinsdottir; Maney Sveinsdottir; Emil A Thorarensen; Bjarni Thorbjornsson; Gisli Masson; Ingileif Jonsdottir; Alma Moller; Thorolfur Gudnason; Karl G Kristinsson; Unnur Thorsteinsdottir; Kari Stefansson
EPI_ISL_424574 hCoV-19/Iceland/554/2020 Europe / Iceland / Reykjavik 2020-03-27 The National University Hospital of Iceland deCODE genetics Daniel F Gudbjartsson; Agnar Helgason; Hakon Jonsson; Olafur T Magnusson; Pall Melsted; Gudmundur L Norddahl; Jona Saemundsdottir; Asgeir Sigurdsson; Patrick Sulem; Arna B Agustsdottir; Berglind Eiriksdottir; Run Fridriksdottir; Elisabet E Gardarsdottir; Gudmundur Georgsson; Olafia S Gretarsdottir; Kjartan R Gudmundsson; Thora R Gunnarsdottir; Arnaldur Gylfason; Hilma Holm; Brynjar O Jensson; Aslaug Jonasdottir; Kamilla S Josefsdottir; Thordur Kristjansson; Droplaug N Magnusdottir; Louise le Roux; Gudrun Sigmundsdottir; Gardar Sveinbjornsson; Kristin E Sveinsdottir; Maney Sveinsdottir; Emil A Thorarensen; Bjarni Thorbjornsson; Gisli Masson; Ingileif Jonsdottir; Alma Moller; Thorolfur Gudnason; Karl G Kristinsson; Unnur Thorsteinsdottir; Kari Stefansson
EPI_ISL_424575 hCoV-19/Iceland/555/2020 Europe / Iceland / Reykjavik 2020-03-28 The National University Hospital of Iceland deCODE genetics Daniel F Gudbjartsson; Agnar Helgason; Hakon Jonsson; Olafur T Magnusson; Pall Melsted; Gudmundur L Norddahl; Jona Saemundsdottir; Asgeir Sigurdsson; Patrick Sulem; Arna B Agustsdottir; Berglind Eiriksdottir; Run Fridriksdottir; Elisabet E Gardarsdottir; Gudmundur Georgsson; Olafia S Gretarsdottir; Kjartan R Gudmundsson; Thora R Gunnarsdottir; Arnaldur Gylfason; Hilma Holm; Brynjar O Jensson; Aslaug Jonasdottir; Kamilla S Josefsdottir; Thordur Kristjansson; Droplaug N Magnusdottir; Louise le Roux; Gudrun Sigmundsdottir; Gardar Sveinbjornsson; Kristin E Sveinsdottir; Maney Sveinsdottir; Emil A Thorarensen; Bjarni Thorbjornsson; Gisli Masson; Ingileif Jonsdottir; Alma Moller; Thorolfur Gudnason; Karl G Kristinsson; Unnur Thorsteinsdottir; Kari Stefansson
EPI_ISL_424576 hCoV-19/Iceland/556/2020 Europe / Iceland / Reykjavik 2020-03-27 The National University Hospital of Iceland deCODE genetics Daniel F Gudbjartsson; Agnar Helgason; Hakon Jonsson; Olafur T Magnusson; Pall Melsted; Gudmundur L Norddahl; Jona Saemundsdottir; Asgeir Sigurdsson; Patrick Sulem; Arna B Agustsdottir; Berglind Eiriksdottir; Run Fridriksdottir; Elisabet E Gardarsdottir; Gudmundur Georgsson; Olafia S Gretarsdottir; Kjartan R Gudmundsson; Thora R Gunnarsdottir; Arnaldur Gylfason; Hilma Holm; Brynjar O Jensson; Aslaug Jonasdottir; Kamilla S Josefsdottir; Thordur Kristjansson; Droplaug N Magnusdottir; Louise le Roux; Gudrun Sigmundsdottir; Gardar Sveinbjornsson; Kristin E Sveinsdottir; Maney Sveinsdottir; Emil A Thorarensen; Bjarni Thorbjornsson; Gisli Masson; Ingileif Jonsdottir; Alma Moller; Thorolfur Gudnason; Karl G Kristinsson; Unnur Thorsteinsdottir; Kari Stefansson

EPI_ISL_424577 hCoV-19/Iceland/557/2020 Europe / Iceland / Reykjavik 2020-03-27 The National University Hospital of Iceland deCODE genetics Daniel F Gudbjartsson; Agnar Helgason; Hakon Jonsson; Olafur T Magnusson; Pall Melsted; Gudmundur L Norddahl; Jona Saemundsdottir; Asgeir Sigurdsson; Patrick Sulem; Arna B Agustsdottir; Berglind Eiriksdottir; Run Fridriksdottir; Elisabet E Gardarsdottir; Gudmundur Georgsson; Olafia S Gretarsdottir; Kjartan R Gudmundsson; Thora R Gunnarsdottir; Arnaldur Gylfason; Hilma Holm; Brynjar O Jensson; Aslaug Jonasdottir; Kamilla S Josefsdottir; Thordur Kristjansson; Droplaug N Magnusdottir; Louise le Roux; Gudrun Sigmundsdottir; Gardar Sveinbjornsson; Kristin E Sveinsdottir; Maney Sveinsdottir; Emil A Thorarensen; Bjarni Thorbjornsson; Gisli Masson; Ingileif Jonsdottir; Alma Moller; Thorolfur Gudnason; Karl G Kristinsson; Unnur Thorsteinsdottir; Kari Stefansson
EPI_ISL_424578 hCoV-19/Iceland/558/2020 Europe / Iceland / Reykjavik 2020-03-27 The National University Hospital of Iceland deCODE genetics Daniel F Gudbjartsson; Agnar Helgason; Hakon Jonsson; Olafur T Magnusson; Pall Melsted; Gudmundur L Norddahl; Jona Saemundsdottir; Asgeir Sigurdsson; Patrick Sulem; Arna B Agustsdottir; Berglind Eiriksdottir; Run Fridriksdottir; Elisabet E Gardarsdottir; Gudmundur Georgsson; Olafia S Gretarsdottir; Kjartan R Gudmundsson; Thora R Gunnarsdottir; Arnaldur Gylfason; Hilma Holm; Brynjar O Jensson; Aslaug Jonasdottir; Kamilla S Josefsdottir; Thordur Kristjansson; Droplaug N Magnusdottir; Louise le Roux; Gudrun Sigmundsdottir; Gardar Sveinbjornsson; Kristin E Sveinsdottir; Maney Sveinsdottir; Emil A Thorarensen; Bjarni Thorbjornsson; Gisli Masson; Ingileif Jonsdottir; Alma Moller; Thorolfur Gudnason; Karl G Kristinsson; Unnur Thorsteinsdottir; Kari Stefansson
EPI_ISL_425823 hCoV-19/Scotland/EDB017/2020 Europe / United Kingdom / Scotland 2020-03-14 Virology Department, Royal Infirmary of Edinburgh, NHS Lothian / School of Biological Sciences, University of Edinburgh / Institute of Genetics and Molecular Medicine, University of Edinburgh COVID-19 Genomics UK (COG-UK) Consortium McHugh M, Dewar R, Rooke S, Gallagher M, Balcaza C, O'Toole A, Hill V, McCrone JT, Colquhoun R, Yu X, Jackson B, Scher E, Rambaut A, Williams TC, Templeton K
EPI_ISL_425824 hCoV-19/Scotland/EDB018/2020 Europe / United Kingdom / Scotland 2020-03-12 Virology Department, Royal Infirmary of Edinburgh, NHS Lothian / School of Biological Sciences, University of Edinburgh / Institute of Genetics and Molecular Medicine, University of Edinburgh COVID-19 Genomics UK (COG-UK) Consortium McHugh M, Dewar R, Rooke S, Gallagher M, Balcaza C, O'Toole A, Hill V, McCrone JT, Colquhoun R, Yu X, Jackson B, Scher E, Rambaut A, Williams TC, Templeton K
EPI_ISL_425825 hCoV-19/Scotland/EDB019/2020 Europe / United Kingdom / Scotland 2020-03-13 Virology Department, Royal Infirmary of Edinburgh, NHS Lothian / School of Biological Sciences, University of Edinburgh / Institute of Genetics and Molecular Medicine, University of Edinburgh COVID-19 Genomics UK (COG-UK) Consortium McHugh M, Dewar R, Rooke S, Gallagher M, Balcaza C, O'Toole A, Hill V, McCrone JT, Colquhoun R, Yu X, Jackson B, Scher E, Rambaut A, Williams TC, Templeton K
EPI_ISL_425826 hCoV-19/Scotland/EDB020/2020 Europe / United Kingdom / Scotland 2020-03-14 Virology Department, Royal Infirmary of Edinburgh, NHS Lothian / School of Biological Sciences, University of Edinburgh / Institute of Genetics and Molecular Medicine, University of Edinburgh COVID-19 Genomics UK (COG-UK) Consortium McHugh M, Dewar R, Rooke S, Gallagher M, Balcaza C, O'Toole A, Hill V, McCrone JT, Colquhoun R, Yu X, Jackson B, Scher E, Rambaut A, Williams TC, Templeton K
EPI_ISL_425827 hCoV-19/Scotland/EDB021/2020 Europe / United Kingdom / Scotland 2020-03-13 Virology Department, Royal Infirmary of Edinburgh, NHS Lothian / School of Biological Sciences, University of Edinburgh / Institute of Genetics and Molecular Medicine, University of Edinburgh COVID-19 Genomics UK (COG-UK) Consortium McHugh M, Dewar R, Rooke S, Gallagher M, Balcaza C, O'Toole A, Hill V, McCrone JT, Colquhoun R, Yu X, Jackson B, Scher E, Rambaut A, Williams TC, Templeton K
EPI_ISL_425828 hCoV-19/Scotland/EDB022/2020 Europe / United Kingdom / Scotland 2020-03-12 Virology Department, Royal Infirmary of Edinburgh, NHS Lothian / School of Biological Sciences, University of Edinburgh / Institute of Genetics and Molecular Medicine, University of Edinburgh COVID-19 Genomics UK (COG-UK) Consortium McHugh M, Dewar R, Rooke S, Gallagher M, Balcaza C, O'Toole A, Hill V, McCrone JT, Colquhoun R, Yu X, Jackson B, Scher E, Rambaut A, Williams TC, Templeton K
EPI_ISL_425829 hCoV-19/Scotland/EDB023/2020 Europe / United Kingdom / Scotland 2020-03-12 Virology Department, Royal Infirmary of Edinburgh, NHS Lothian / School of Biological Sciences, University of Edinburgh / Institute of Genetics and Molecular Medicine, University of Edinburgh COVID-19 Genomics UK (COG-UK) Consortium McHugh M, Dewar R, Rooke S, Gallagher M, Balcaza C, O'Toole A, Hill V, McCrone JT, Colquhoun R, Yu X, Jackson B, Scher E, Rambaut A, Williams TC, Templeton K
EPI_ISL_425830 hCoV-19/Scotland/EDB024/2020 Europe / United Kingdom / Scotland 2020-03-12 Virology Department, Royal Infirmary of Edinburgh, NHS Lothian / School of Biological Sciences, University of Edinburgh / Institute of Genetics and Molecular Medicine, University of Edinburgh COVID-19 Genomics UK (COG-UK) Consortium McHugh M, Dewar R, Rooke S, Gallagher M, Balcaza C, O'Toole A, Hill V, McCrone JT, Colquhoun R, Yu X, Jackson B, Scher E, Rambaut A, Williams TC, Templeton K
EPI_ISL_425836 hCoV-19/Scotland/EDB030/2020 Europe / United Kingdom / Scotland 2020-03-11 Virology Department, Royal Infirmary of Edinburgh, NHS Lothian / School of Biological Sciences, University of Edinburgh / Institute of Genetics and Molecular Medicine, University of Edinburgh COVID-19 Genomics UK (COG-UK) Consortium McHugh M, Dewar R, Rooke S, Gallagher M, Balcaza C, O'Toole A, Hill V, McCrone JT, Colquhoun R, Yu X, Jackson B, Scher E, Rambaut A, Williams TC, Templeton K
EPI_ISL_425839 hCoV-19/Scotland/EDB033/2020 Europe / United Kingdom / Scotland 2020-03-13 Virology Department, Royal Infirmary of Edinburgh, NHS Lothian / School of Biological Sciences, University of Edinburgh / Institute of Genetics and Molecular Medicine, University of Edinburgh COVID-19 Genomics UK (COG-UK) Consortium McHugh M, Dewar R, Rooke S, Gallagher M, Balcaza C, O'Toole A, Hill V, McCrone JT, Colquhoun R, Yu X, Jackson B, Scher E, Rambaut A, Williams TC, Templeton K
EPI_ISL_425842 hCoV-19/Scotland/EDB036/2020 Europe / United Kingdom / Scotland 2020-03-11 Virology Department, Royal Infirmary of Edinburgh, NHS Lothian / School of Biological Sciences, University of

Edinburgh / Institute of Genetics and Molecular Medicine, University of Edinburgh COVID-19 Genomics UK (COG-UK) Consortium McHugh M, Dewar R, Rooke S, Gallagher M, Balcaza C, O'Toole A, Hill V, McCrone JT, Colquhoun R, Yu X, Jackson B, Scher E, Rambaut A, Williams TC, Templeton K
EPI_ISL_425843 hCoV-19/Scotland/EDB037/2020 Europe / United Kingdom / Scotland 2020-03-12
Virology Department, Royal Infirmary of Edinburgh, NHS Lothian / School of Biological Sciences, University of Edinburgh / Institute of Genetics and Molecular Medicine, University of Edinburgh COVID-19 Genomics UK (COG-UK) Consortium McHugh M, Dewar R, Rooke S, Gallagher M, Balcaza C, O'Toole A, Hill V, McCrone JT, Colquhoun R, Yu X, Jackson B, Scher E, Rambaut A, Williams TC, Templeton K
EPI_ISL_425844 hCoV-19/Scotland/EDB038/2020 Europe / United Kingdom / Scotland 2020-03-11
Virology Department, Royal Infirmary of Edinburgh, NHS Lothian / School of Biological Sciences, University of Edinburgh / Institute of Genetics and Molecular Medicine, University of Edinburgh COVID-19 Genomics UK (COG-UK) Consortium McHugh M, Dewar R, Rooke S, Gallagher M, Balcaza C, O'Toole A, Hill V, McCrone JT, Colquhoun R, Yu X, Jackson B, Scher E, Rambaut A, Williams TC, Templeton K
EPI_ISL_425845 hCoV-19/Scotland/EDB039/2020 Europe / United Kingdom / Scotland 2020-03-09
Virology Department, Royal Infirmary of Edinburgh, NHS Lothian / School of Biological Sciences, University of Edinburgh / Institute of Genetics and Molecular Medicine, University of Edinburgh COVID-19 Genomics UK (COG-UK) Consortium McHugh M, Dewar R, Rooke S, Gallagher M, Balcaza C, O'Toole A, Hill V, McCrone JT, Colquhoun R, Yu X, Jackson B, Scher E, Rambaut A, Williams TC, Templeton K
EPI_ISL_425847 hCoV-19/Scotland/EDB042/2020 Europe / United Kingdom / Scotland 2020-03-11
Virology Department, Royal Infirmary of Edinburgh, NHS Lothian / School of Biological Sciences, University of Edinburgh / Institute of Genetics and Molecular Medicine, University of Edinburgh COVID-19 Genomics UK (COG-UK) Consortium McHugh M, Dewar R, Rooke S, Gallagher M, Balcaza C, O'Toole A, Hill V, McCrone JT, Colquhoun R, Yu X, Jackson B, Scher E, Rambaut A, Williams TC, Templeton K
EPI_ISL_425848 hCoV-19/Scotland/EDB044/2020 Europe / United Kingdom / Scotland 2020-03-12
Virology Department, Royal Infirmary of Edinburgh, NHS Lothian / School of Biological Sciences, University of Edinburgh / Institute of Genetics and Molecular Medicine, University of Edinburgh COVID-19 Genomics UK (COG-UK) Consortium McHugh M, Dewar R, Rooke S, Gallagher M, Balcaza C, O'Toole A, Hill V, McCrone JT, Colquhoun R, Yu X, Jackson B, Scher E, Rambaut A, Williams TC, Templeton K
EPI_ISL_425849 hCoV-19/Scotland/EDB045/2020 Europe / United Kingdom / Scotland 2020-03-14
Virology Department, Royal Infirmary of Edinburgh, NHS Lothian / School of Biological Sciences, University of Edinburgh / Institute of Genetics and Molecular Medicine, University of Edinburgh COVID-19 Genomics UK (COG-UK) Consortium McHugh M, Dewar R, Rooke S, Gallagher M, Balcaza C, O'Toole A, Hill V, McCrone JT, Colquhoun R, Yu X, Jackson B, Scher E, Rambaut A, Williams TC, Templeton K
EPI_ISL_425850 hCoV-19/Scotland/EDB046/2020 Europe / United Kingdom / Scotland 2020-03-15
Virology Department, Royal Infirmary of Edinburgh, NHS Lothian / School of Biological Sciences, University of Edinburgh / Institute of Genetics and Molecular Medicine, University of Edinburgh COVID-19 Genomics UK (COG-UK) Consortium McHugh M, Dewar R, Rooke S, Gallagher M, Balcaza C, O'Toole A, Hill V, McCrone JT, Colquhoun R, Yu X, Jackson B, Scher E, Rambaut A, Williams TC, Templeton K
EPI_ISL_425851 hCoV-19/Scotland/EDB047/2020 Europe / United Kingdom / Scotland 2020-03-17
Virology Department, Royal Infirmary of Edinburgh, NHS Lothian / School of Biological Sciences, University of Edinburgh / Institute of Genetics and Molecular Medicine, University of Edinburgh COVID-19 Genomics UK (COG-UK) Consortium McHugh M, Dewar R, Rooke S, Gallagher M, Balcaza C, O'Toole A, Hill V, McCrone JT, Colquhoun R, Yu X, Jackson B, Scher E, Rambaut A, Williams TC, Templeton K
EPI_ISL_425852 hCoV-19/Scotland/EDB048/2020 Europe / United Kingdom / Scotland 2020-03-17
Virology Department, Royal Infirmary of Edinburgh, NHS Lothian / School of Biological Sciences, University of Edinburgh / Institute of Genetics and Molecular Medicine, University of Edinburgh COVID-19 Genomics UK (COG-UK) Consortium McHugh M, Dewar R, Rooke S, Gallagher M, Balcaza C, O'Toole A, Hill V, McCrone JT, Colquhoun R, Yu X, Jackson B, Scher E, Rambaut A, Williams TC, Templeton K
EPI_ISL_425856 hCoV-19/Scotland/EDB052/2020 Europe / United Kingdom / Scotland 2020-03-18
Virology Department, Royal Infirmary of Edinburgh, NHS Lothian / School of Biological Sciences, University of Edinburgh / Institute of Genetics and Molecular Medicine, University of Edinburgh COVID-19 Genomics UK (COG-UK) Consortium McHugh M, Dewar R, Rooke S, Gallagher M, Balcaza C, O'Toole A, Hill V, McCrone JT, Colquhoun R, Yu X, Jackson B, Scher E, Rambaut A, Williams TC, Templeton K
EPI_ISL_425861 hCoV-19/Scotland/EDB057/2020 Europe / United Kingdom / Scotland 2020-03-18
Virology Department, Royal Infirmary of Edinburgh, NHS Lothian / School of Biological Sciences, University of Edinburgh / Institute of Genetics and Molecular Medicine, University of Edinburgh COVID-19 Genomics UK (COG-UK) Consortium McHugh M, Dewar R, Rooke S, Gallagher M, Balcaza C, O'Toole A, Hill V, McCrone JT, Colquhoun R, Yu X, Jackson B, Scher E, Rambaut A, Williams TC, Templeton K
EPI_ISL_425862 hCoV-19/Scotland/EDB058/2020 Europe / United Kingdom / Scotland 2020-03-18
Virology Department, Royal Infirmary of Edinburgh, NHS Lothian / School of Biological Sciences, University of Edinburgh / Institute of Genetics and Molecular Medicine, University of Edinburgh COVID-19 Genomics UK (COG-UK) Consortium McHugh M, Dewar R, Rooke S, Gallagher M, Balcaza C, O'Toole A, Hill V, McCrone JT, Colquhoun R, Yu X, Jackson B, Scher E, Rambaut A, Williams TC, Templeton K
EPI_ISL_425863 hCoV-19/Scotland/EDB063/2020 Europe / United Kingdom / Scotland 2020-03-19
Virology Department, Royal Infirmary of Edinburgh, NHS Lothian / School of Biological Sciences, University of Edinburgh / Institute of Genetics and Molecular Medicine, University of Edinburgh COVID-19 Genomics UK (COG-UK) Consortium McHugh M, Dewar R, Rooke S, Gallagher M, Balcaza C, O'Toole A, Hill V, McCrone JT, Colquhoun R, Yu X, Jackson B, Scher E, Rambaut A, Williams TC, Templeton K
EPI_ISL_425864 hCoV-19/Scotland/EDB065/2020 Europe / United Kingdom / Scotland 2020-03-20
Virology Department, Royal Infirmary of Edinburgh, NHS Lothian / School of Biological Sciences, University of

Edinburgh / Institute of Genetics and Molecular Medicine, University of Edinburgh	COVID-19 Genomics UK (COG-UK) Consortium	McHugh M, Dewar R, Rooke S, Gallagher M, Balcaza C, O'Toole A, Hill V, McCrone JT, Colquhoun R, Yu X, Jackson B, Scher E, Rambaut A, Williams TC, Templeton K
EPI_ISL_425865	hCoV-19/Scotland/EDB066/2020	Europe / United Kingdom / Scotland	2020-03-18
Virology Department, Royal Infirmary of Edinburgh, NHS Lothian / School of Biological Sciences, University of Edinburgh / Institute of Genetics and Molecular Medicine, University of Edinburgh	COVID-19 Genomics UK (COG-UK) Consortium	McHugh M, Dewar R, Rooke S, Gallagher M, Balcaza C, O'Toole A, Hill V, McCrone JT, Colquhoun R, Yu X, Jackson B, Scher E, Rambaut A, Williams TC, Templeton K
EPI_ISL_425866	hCoV-19/Scotland/EDB067/2020	Europe / United Kingdom / Scotland	2020-03-19
Virology Department, Royal Infirmary of Edinburgh, NHS Lothian / School of Biological Sciences, University of Edinburgh / Institute of Genetics and Molecular Medicine, University of Edinburgh	COVID-19 Genomics UK (COG-UK) Consortium	McHugh M, Dewar R, Rooke S, Gallagher M, Balcaza C, O'Toole A, Hill V, McCrone JT, Colquhoun R, Yu X, Jackson B, Scher E, Rambaut A, Williams TC, Templeton K
EPI_ISL_425867	hCoV-19/Scotland/EDB068/2020	Europe / United Kingdom / Scotland	2020-03-20
Virology Department, Royal Infirmary of Edinburgh, NHS Lothian / School of Biological Sciences, University of Edinburgh / Institute of Genetics and Molecular Medicine, University of Edinburgh	COVID-19 Genomics UK (COG-UK) Consortium	McHugh M, Dewar R, Rooke S, Gallagher M, Balcaza C, O'Toole A, Hill V, McCrone JT, Colquhoun R, Yu X, Jackson B, Scher E, Rambaut A, Williams TC, Templeton K
EPI_ISL_425868	hCoV-19/Scotland/EDB070/2020	Europe / United Kingdom / Scotland	2020-03-22
Virology Department, Royal Infirmary of Edinburgh, NHS Lothian / School of Biological Sciences, University of Edinburgh / Institute of Genetics and Molecular Medicine, University of Edinburgh	COVID-19 Genomics UK (COG-UK) Consortium	McHugh M, Dewar R, Rooke S, Gallagher M, Balcaza C, O'Toole A, Hill V, McCrone JT, Colquhoun R, Yu X, Jackson B, Scher E, Rambaut A, Williams TC, Templeton K
EPI_ISL_425869	hCoV-19/Scotland/EDB071/2020	Europe / United Kingdom / Scotland	2020-03-21
Virology Department, Royal Infirmary of Edinburgh, NHS Lothian / School of Biological Sciences, University of Edinburgh / Institute of Genetics and Molecular Medicine, University of Edinburgh	COVID-19 Genomics UK (COG-UK) Consortium	McHugh M, Dewar R, Rooke S, Gallagher M, Balcaza C, O'Toole A, Hill V, McCrone JT, Colquhoun R, Yu X, Jackson B, Scher E, Rambaut A, Williams TC, Templeton K
EPI_ISL_425870	hCoV-19/Scotland/EDB073/2020	Europe / United Kingdom / Scotland	2020-03-20
Virology Department, Royal Infirmary of Edinburgh, NHS Lothian / School of Biological Sciences, University of Edinburgh / Institute of Genetics and Molecular Medicine, University of Edinburgh	COVID-19 Genomics UK (COG-UK) Consortium	McHugh M, Dewar R, Rooke S, Gallagher M, Balcaza C, O'Toole A, Hill V, McCrone JT, Colquhoun R, Yu X, Jackson B, Scher E, Rambaut A, Williams TC, Templeton K
EPI_ISL_425872	hCoV-19/Scotland/EDB075/2020	Europe / United Kingdom / Scotland	2020-03-20
Virology Department, Royal Infirmary of Edinburgh, NHS Lothian / School of Biological Sciences, University of Edinburgh / Institute of Genetics and Molecular Medicine, University of Edinburgh	COVID-19 Genomics UK (COG-UK) Consortium	McHugh M, Dewar R, Rooke S, Gallagher M, Balcaza C, O'Toole A, Hill V, McCrone JT, Colquhoun R, Yu X, Jackson B, Scher E, Rambaut A, Williams TC, Templeton K
EPI_ISL_425873	hCoV-19/Scotland/EDB081/2020	Europe / United Kingdom / Scotland	2020-03-20
Virology Department, Royal Infirmary of Edinburgh, NHS Lothian / School of Biological Sciences, University of Edinburgh / Institute of Genetics and Molecular Medicine, University of Edinburgh	COVID-19 Genomics UK (COG-UK) Consortium	McHugh M, Dewar R, Rooke S, Gallagher M, Balcaza C, O'Toole A, Hill V, McCrone JT, Colquhoun R, Yu X, Jackson B, Scher E, Rambaut A, Williams TC, Templeton K
EPI_ISL_425875	hCoV-19/Scotland/EDB083/2020	Europe / United Kingdom / Scotland	2020-03-21
Virology Department, Royal Infirmary of Edinburgh, NHS Lothian / School of Biological Sciences, University of Edinburgh / Institute of Genetics and Molecular Medicine, University of Edinburgh	COVID-19 Genomics UK (COG-UK) Consortium	McHugh M, Dewar R, Rooke S, Gallagher M, Balcaza C, O'Toole A, Hill V, McCrone JT, Colquhoun R, Yu X, Jackson B, Scher E, Rambaut A, Williams TC, Templeton K
EPI_ISL_425876	hCoV-19/Scotland/EDB084/2020	Europe / United Kingdom / Scotland	2020-03-21
Virology Department, Royal Infirmary of Edinburgh, NHS Lothian / School of Biological Sciences, University of Edinburgh / Institute of Genetics and Molecular Medicine, University of Edinburgh	COVID-19 Genomics UK (COG-UK) Consortium	McHugh M, Dewar R, Rooke S, Gallagher M, Balcaza C, O'Toole A, Hill V, McCrone JT, Colquhoun R, Yu X, Jackson B, Scher E, Rambaut A, Williams TC, Templeton K
EPI_ISL_425877	hCoV-19/Scotland/EDB085/2020	Europe / United Kingdom / Scotland	2020-03-21
Virology Department, Royal Infirmary of Edinburgh, NHS Lothian / School of Biological Sciences, University of Edinburgh / Institute of Genetics and Molecular Medicine, University of Edinburgh	COVID-19 Genomics UK (COG-UK) Consortium	McHugh M, Dewar R, Rooke S, Gallagher M, Balcaza C, O'Toole A, Hill V, McCrone JT, Colquhoun R, Yu X, Jackson B, Scher E, Rambaut A, Williams TC, Templeton K
EPI_ISL_425878	hCoV-19/Scotland/EDB086/2020	Europe / United Kingdom / Scotland	2020-03-21
Virology Department, Royal Infirmary of Edinburgh, NHS Lothian / School of Biological Sciences, University of Edinburgh / Institute of Genetics and Molecular Medicine, University of Edinburgh	COVID-19 Genomics UK (COG-UK) Consortium	McHugh M, Dewar R, Rooke S, Gallagher M, Balcaza C, O'Toole A, Hill V, McCrone JT, Colquhoun R, Yu X, Jackson B, Scher E, Rambaut A, Williams TC, Templeton K
EPI_ISL_425879	hCoV-19/Scotland/EDB087/2020	Europe / United Kingdom / Scotland	2020-03-21
Virology Department, Royal Infirmary of Edinburgh, NHS Lothian / School of Biological Sciences, University of Edinburgh / Institute of Genetics and Molecular Medicine, University of Edinburgh	COVID-19 Genomics UK (COG-UK) Consortium	McHugh M, Dewar R, Rooke S, Gallagher M, Balcaza C, O'Toole A, Hill V, McCrone JT, Colquhoun R, Yu X, Jackson B, Scher E, Rambaut A, Williams TC, Templeton K
EPI_ISL_425880	hCoV-19/Scotland/EDB089/2020	Europe / United Kingdom / Scotland	2020-03-21
Virology Department, Royal Infirmary of Edinburgh, NHS Lothian / School of Biological Sciences, University of

Edinburgh / Institute of Genetics and Molecular Medicine, University of Edinburgh	COVID-19 Genomics UK (COG-UK) Consortium	McHugh M, Dewar R, Rooke S, Gallagher M, Balcaza C, O'Toole A, Hill V, McCrone JT, Colquhoun R, Yu X, Jackson B, Scher E, Rambaut A, Williams TC, Templeton K
EPI_ISL_425881	hCoV-19/Scotland/EDB090/2020	Europe / United Kingdom / Scotland	2020-03-21	Virology Department, Royal Infirmary of Edinburgh, NHS Lothian / School of Biological Sciences, University of Edinburgh / Institute of Genetics and Molecular Medicine, University of Edinburgh	COVID-19 Genomics UK (COG-UK) Consortium	McHugh M, Dewar R, Rooke S, Gallagher M, Balcaza C, O'Toole A, Hill V, McCrone JT, Colquhoun R, Yu X, Jackson B, Scher E, Rambaut A, Williams TC, Templeton K
EPI_ISL_425882	hCoV-19/Scotland/EDB091/2020	Europe / United Kingdom / Scotland	2020-03-21	Virology Department, Royal Infirmary of Edinburgh, NHS Lothian / School of Biological Sciences, University of Edinburgh / Institute of Genetics and Molecular Medicine, University of Edinburgh	COVID-19 Genomics UK (COG-UK) Consortium	McHugh M, Dewar R, Rooke S, Gallagher M, Balcaza C, O'Toole A, Hill V, McCrone JT, Colquhoun R, Yu X, Jackson B, Scher E, Rambaut A, Williams TC, Templeton K
EPI_ISL_425885	hCoV-19/Scotland/EDB095/2020	Europe / United Kingdom / Scotland	2020-03-21	Virology Department, Royal Infirmary of Edinburgh, NHS Lothian / School of Biological Sciences, University of Edinburgh / Institute of Genetics and Molecular Medicine, University of Edinburgh	COVID-19 Genomics UK (COG-UK) Consortium	McHugh M, Dewar R, Rooke S, Gallagher M, Balcaza C, O'Toole A, Hill V, McCrone JT, Colquhoun R, Yu X, Jackson B, Scher E, Rambaut A, Williams TC, Templeton K
EPI_ISL_425887	hCoV-19/Scotland/EDB097/2020	Europe / United Kingdom / Scotland	2020-03-20	Virology Department, Royal Infirmary of Edinburgh, NHS Lothian / School of Biological Sciences, University of Edinburgh / Institute of Genetics and Molecular Medicine, University of Edinburgh	COVID-19 Genomics UK (COG-UK) Consortium	McHugh M, Dewar R, Rooke S, Gallagher M, Balcaza C, O'Toole A, Hill V, McCrone JT, Colquhoun R, Yu X, Jackson B, Scher E, Rambaut A, Williams TC, Templeton K
EPI_ISL_425888	hCoV-19/Scotland/EDB098/2020	Europe / United Kingdom / Scotland	2020-03-20	Virology Department, Royal Infirmary of Edinburgh, NHS Lothian / School of Biological Sciences, University of Edinburgh / Institute of Genetics and Molecular Medicine, University of Edinburgh	COVID-19 Genomics UK (COG-UK) Consortium	McHugh M, Dewar R, Rooke S, Gallagher M, Balcaza C, O'Toole A, Hill V, McCrone JT, Colquhoun R, Yu X, Jackson B, Scher E, Rambaut A, Williams TC, Templeton K
EPI_ISL_425889	hCoV-19/Scotland/EDB099/2020	Europe / United Kingdom / Scotland	2020-03-20	Virology Department, Royal Infirmary of Edinburgh, NHS Lothian / School of Biological Sciences, University of Edinburgh / Institute of Genetics and Molecular Medicine, University of Edinburgh	COVID-19 Genomics UK (COG-UK) Consortium	McHugh M, Dewar R, Rooke S, Gallagher M, Balcaza C, O'Toole A, Hill V, McCrone JT, Colquhoun R, Yu X, Jackson B, Scher E, Rambaut A, Williams TC, Templeton K
EPI_ISL_425891	hCoV-19/Scotland/EDB101/2020	Europe / United Kingdom / Scotland	2020-03-21	Virology Department, Royal Infirmary of Edinburgh, NHS Lothian / School of Biological Sciences, University of Edinburgh / Institute of Genetics and Molecular Medicine, University of Edinburgh	COVID-19 Genomics UK (COG-UK) Consortium	McHugh M, Dewar R, Rooke S, Gallagher M, Balcaza C, O'Toole A, Hill V, McCrone JT, Colquhoun R, Yu X, Jackson B, Scher E, Rambaut A, Williams TC, Templeton K
EPI_ISL_425892	hCoV-19/Scotland/EDB102/2020	Europe / United Kingdom / Scotland	2020-03-22	Virology Department, Royal Infirmary of Edinburgh, NHS Lothian / School of Biological Sciences, University of Edinburgh / Institute of Genetics and Molecular Medicine, University of Edinburgh	COVID-19 Genomics UK (COG-UK) Consortium	McHugh M, Dewar R, Rooke S, Gallagher M, Balcaza C, O'Toole A, Hill V, McCrone JT, Colquhoun R, Yu X, Jackson B, Scher E, Rambaut A, Williams TC, Templeton K
EPI_ISL_425893	hCoV-19/Scotland/EDB103/2020	Europe / United Kingdom / Scotland	2020-03-22	Virology Department, Royal Infirmary of Edinburgh, NHS Lothian / School of Biological Sciences, University of Edinburgh / Institute of Genetics and Molecular Medicine, University of Edinburgh	COVID-19 Genomics UK (COG-UK) Consortium	McHugh M, Dewar R, Rooke S, Gallagher M, Balcaza C, O'Toole A, Hill V, McCrone JT, Colquhoun R, Yu X, Jackson B, Scher E, Rambaut A, Williams TC, Templeton K
EPI_ISL_425894	hCoV-19/Scotland/EDB106/2020	Europe / United Kingdom / Scotland	2020-03-22	Virology Department, Royal Infirmary of Edinburgh, NHS Lothian / School of Biological Sciences, University of Edinburgh / Institute of Genetics and Molecular Medicine, University of Edinburgh	COVID-19 Genomics UK (COG-UK) Consortium	McHugh M, Dewar R, Rooke S, Gallagher M, Balcaza C, O'Toole A, Hill V, McCrone JT, Colquhoun R, Yu X, Jackson B, Scher E, Rambaut A, Williams TC, Templeton K
EPI_ISL_425896	hCoV-19/Scotland/EDB108/2020	Europe / United Kingdom / Scotland	2020-03-22	Virology Department, Royal Infirmary of Edinburgh, NHS Lothian / School of Biological Sciences, University of Edinburgh / Institute of Genetics and Molecular Medicine, University of Edinburgh	COVID-19 Genomics UK (COG-UK) Consortium	McHugh M, Dewar R, Rooke S, Gallagher M, Balcaza C, O'Toole A, Hill V, McCrone JT, Colquhoun R, Yu X, Jackson B, Scher E, Rambaut A, Williams TC, Templeton K
EPI_ISL_425897	hCoV-19/Scotland/EDB109/2020	Europe / United Kingdom / Scotland	2020-03-22	Virology Department, Royal Infirmary of Edinburgh, NHS Lothian / School of Biological Sciences, University of Edinburgh / Institute of Genetics and Molecular Medicine, University of Edinburgh	COVID-19 Genomics UK (COG-UK) Consortium	McHugh M, Dewar R, Rooke S, Gallagher M, Balcaza C, O'Toole A, Hill V, McCrone JT, Colquhoun R, Yu X, Jackson B, Scher E, Rambaut A, Williams TC, Templeton K
EPI_ISL_425899	hCoV-19/Scotland/EDB111/2020	Europe / United Kingdom / Scotland	2020-03-22	Virology Department, Royal Infirmary of Edinburgh, NHS Lothian / School of Biological Sciences, University of Edinburgh / Institute of Genetics and Molecular Medicine, University of Edinburgh	COVID-19 Genomics UK (COG-UK) Consortium	McHugh M, Dewar R, Rooke S, Gallagher M, Balcaza C, O'Toole A, Hill V, McCrone JT, Colquhoun R, Yu X, Jackson B, Scher E, Rambaut A, Williams TC, Templeton K
EPI_ISL_426000	hCoV-19/Scotland/EDB407/2020	Europe / United Kingdom / Scotland	2020-03-30	Virology Department, Royal Infirmary of Edinburgh, NHS Lothian / School of Biological Sciences, University of

Edinburgh / Institute of Genetics and Molecular Medicine, University of Edinburgh	COVID-19 Genomics UK (COG-UK) Consortium	McHugh M, Dewar R, Rooke S, Gallagher M, Balcaza C, O'Toole A, Hill V, McCrone JT, Colquhoun R, Yu X, Jackson B, Scher E, Rambaut A, Williams TC, Templeton K
EPI_ISL_426001	hCoV-19/Scotland/EDB409/2020	Europe / United Kingdom / Scotland	2020-03-30	Virology Department, Royal Infirmary of Edinburgh, NHS Lothian / School of Biological Sciences, University of Edinburgh / Institute of Genetics and Molecular Medicine, University of Edinburgh	COVID-19 Genomics UK (COG-UK) Consortium	McHugh M, Dewar R, Rooke S, Gallagher M, Balcaza C, O'Toole A, Hill V, McCrone JT, Colquhoun R, Yu X, Jackson B, Scher E, Rambaut A, Williams TC, Templeton K
EPI_ISL_426002	hCoV-19/Scotland/EDB410/2020	Europe / United Kingdom / Scotland	2020-03-30	Virology Department, Royal Infirmary of Edinburgh, NHS Lothian / School of Biological Sciences, University of Edinburgh / Institute of Genetics and Molecular Medicine, University of Edinburgh	COVID-19 Genomics UK (COG-UK) Consortium	McHugh M, Dewar R, Rooke S, Gallagher M, Balcaza C, O'Toole A, Hill V, McCrone JT, Colquhoun R, Yu X, Jackson B, Scher E, Rambaut A, Williams TC, Templeton K
EPI_ISL_426003	hCoV-19/Scotland/EDB412/2020	Europe / United Kingdom / Scotland	2020-03-30	Virology Department, Royal Infirmary of Edinburgh, NHS Lothian / School of Biological Sciences, University of Edinburgh / Institute of Genetics and Molecular Medicine, University of Edinburgh	COVID-19 Genomics UK (COG-UK) Consortium	McHugh M, Dewar R, Rooke S, Gallagher M, Balcaza C, O'Toole A, Hill V, McCrone JT, Colquhoun R, Yu X, Jackson B, Scher E, Rambaut A, Williams TC, Templeton K
EPI_ISL_426004	hCoV-19/Scotland/EDB413/2020	Europe / United Kingdom / Scotland	2020-03-29	Virology Department, Royal Infirmary of Edinburgh, NHS Lothian / School of Biological Sciences, University of Edinburgh / Institute of Genetics and Molecular Medicine, University of Edinburgh	COVID-19 Genomics UK (COG-UK) Consortium	McHugh M, Dewar R, Rooke S, Gallagher M, Balcaza C, O'Toole A, Hill V, McCrone JT, Colquhoun R, Yu X, Jackson B, Scher E, Rambaut A, Williams TC, Templeton K
EPI_ISL_426005	hCoV-19/Scotland/EDB420/2020	Europe / United Kingdom / Scotland	2020-03-30	Virology Department, Royal Infirmary of Edinburgh, NHS Lothian / School of Biological Sciences, University of Edinburgh / Institute of Genetics and Molecular Medicine, University of Edinburgh	COVID-19 Genomics UK (COG-UK) Consortium	McHugh M, Dewar R, Rooke S, Gallagher M, Balcaza C, O'Toole A, Hill V, McCrone JT, Colquhoun R, Yu X, Jackson B, Scher E, Rambaut A, Williams TC, Templeton K
EPI_ISL_426006	hCoV-19/Scotland/EDB421/2020	Europe / United Kingdom / Scotland	2020-03-30	Virology Department, Royal Infirmary of Edinburgh, NHS Lothian / School of Biological Sciences, University of Edinburgh / Institute of Genetics and Molecular Medicine, University of Edinburgh	COVID-19 Genomics UK (COG-UK) Consortium	McHugh M, Dewar R, Rooke S, Gallagher M, Balcaza C, O'Toole A, Hill V, McCrone JT, Colquhoun R, Yu X, Jackson B, Scher E, Rambaut A, Williams TC, Templeton K
EPI_ISL_426007	hCoV-19/Scotland/EDB442/2020	Europe / United Kingdom / Scotland	2020-03-31	Virology Department, Royal Infirmary of Edinburgh, NHS Lothian / School of Biological Sciences, University of Edinburgh / Institute of Genetics and Molecular Medicine, University of Edinburgh	COVID-19 Genomics UK (COG-UK) Consortium	McHugh M, Dewar R, Rooke S, Gallagher M, Balcaza C, O'Toole A, Hill V, McCrone JT, Colquhoun R, Yu X, Jackson B, Scher E, Rambaut A, Williams TC, Templeton K
EPI_ISL_426008	hCoV-19/Scotland/EDB501/2020	Europe / United Kingdom / Scotland	2020-03-31	Virology Department, Royal Infirmary of Edinburgh, NHS Lothian / School of Biological Sciences, University of Edinburgh / Institute of Genetics and Molecular Medicine, University of Edinburgh	COVID-19 Genomics UK (COG-UK) Consortium	McHugh M, Dewar R, Rooke S, Gallagher M, Balcaza C, O'Toole A, Hill V, McCrone JT, Colquhoun R, Yu X, Jackson B, Scher E, Rambaut A, Williams TC, Templeton K
EPI_ISL_426009	hCoV-19/Scotland/EDB502/2020	Europe / United Kingdom / Scotland	2020-03-31	Virology Department, Royal Infirmary of Edinburgh, NHS Lothian / School of Biological Sciences, University of Edinburgh / Institute of Genetics and Molecular Medicine, University of Edinburgh	COVID-19 Genomics UK (COG-UK) Consortium	McHugh M, Dewar R, Rooke S, Gallagher M, Balcaza C, O'Toole A, Hill V, McCrone JT, Colquhoun R, Yu X, Jackson B, Scher E, Rambaut A, Williams TC, Templeton K
EPI_ISL_426011	hCoV-19/Scotland/EDB513/2020	Europe / United Kingdom / Scotland	2020-03-31	Virology Department, Royal Infirmary of Edinburgh, NHS Lothian / School of Biological Sciences, University of Edinburgh / Institute of Genetics and Molecular Medicine, University of Edinburgh	COVID-19 Genomics UK (COG-UK) Consortium	McHugh M, Dewar R, Rooke S, Gallagher M, Balcaza C, O'Toole A, Hill V, McCrone JT, Colquhoun R, Yu X, Jackson B, Scher E, Rambaut A, Williams TC, Templeton K
EPI_ISL_426013	hCoV-19/Scotland/EDB530/2020	Europe / United Kingdom / Scotland	2020-04-01	Virology Department, Royal Infirmary of Edinburgh, NHS Lothian / School of Biological Sciences, University of Edinburgh / Institute of Genetics and Molecular Medicine, University of Edinburgh	COVID-19 Genomics UK (COG-UK) Consortium	McHugh M, Dewar R, Rooke S, Gallagher M, Balcaza C, O'Toole A, Hill V, McCrone JT, Colquhoun R, Yu X, Jackson B, Scher E, Rambaut A, Williams TC, Templeton K
EPI_ISL_426014	hCoV-19/Scotland/EDB533/2020	Europe / United Kingdom / Scotland	2020-04-01	Virology Department, Royal Infirmary of Edinburgh, NHS Lothian / School of Biological Sciences, University of Edinburgh / Institute of Genetics and Molecular Medicine, University of Edinburgh	COVID-19 Genomics UK (COG-UK) Consortium	McHugh M, Dewar R, Rooke S, Gallagher M, Balcaza C, O'Toole A, Hill V, McCrone JT, Colquhoun R, Yu X, Jackson B, Scher E, Rambaut A, Williams TC, Templeton K
EPI_ISL_426015	hCoV-19/Scotland/EDB551/2020	Europe / United Kingdom / Scotland	2020-03-31	Virology Department, Royal Infirmary of Edinburgh, NHS Lothian / School of Biological Sciences, University of Edinburgh / Institute of Genetics and Molecular Medicine, University of Edinburgh	COVID-19 Genomics UK (COG-UK) Consortium	McHugh M, Dewar R, Rooke S, Gallagher M, Balcaza C, O'Toole A, Hill V, McCrone JT, Colquhoun R, Yu X, Jackson B, Scher E, Rambaut A, Williams TC, Templeton K
EPI_ISL_426016	hCoV-19/Scotland/EDB555/2020	Europe / United Kingdom / Scotland	2020-03-31	Virology Department, Royal Infirmary of Edinburgh, NHS Lothian / School of Biological Sciences, University of

Edinburgh / Institute of Genetics and Molecular Medicine, University of Edinburgh	COVID-19 Genomics UK (COG-UK) Consortium	McHugh M, Dewar R, Rooke S, Gallagher M, Balcaza C, O'Toole A, Hill V, McCrone JT, Colquhoun R, Yu X, Jackson B, Scher E, Rambaut A, Williams TC, Templeton K
EPI_ISL_426020	hCoV-19/Scotland/EDB561/2020	Europe / United Kingdom / Scotland	2020-04-01	Virology Department, Royal Infirmary of Edinburgh, NHS Lothian / School of Biological Sciences, University of Edinburgh / Institute of Genetics and Molecular Medicine, University of Edinburgh	COVID-19 Genomics UK (COG-UK) Consortium	McHugh M, Dewar R, Rooke S, Gallagher M, Balcaza C, O'Toole A, Hill V, McCrone JT, Colquhoun R, Yu X, Jackson B, Scher E, Rambaut A, Williams TC, Templeton K
EPI_ISL_426021	hCoV-19/Scotland/EDB562/2020	Europe / United Kingdom / Scotland	2020-04-01	Virology Department, Royal Infirmary of Edinburgh, NHS Lothian / School of Biological Sciences, University of Edinburgh / Institute of Genetics and Molecular Medicine, University of Edinburgh	COVID-19 Genomics UK (COG-UK) Consortium	McHugh M, Dewar R, Rooke S, Gallagher M, Balcaza C, O'Toole A, Hill V, McCrone JT, Colquhoun R, Yu X, Jackson B, Scher E, Rambaut A, Williams TC, Templeton K
EPI_ISL_426023	hCoV-19/Scotland/EDB565/2020	Europe / United Kingdom / Scotland	2020-03-31	Virology Department, Royal Infirmary of Edinburgh, NHS Lothian / School of Biological Sciences, University of Edinburgh / Institute of Genetics and Molecular Medicine, University of Edinburgh	COVID-19 Genomics UK (COG-UK) Consortium	McHugh M, Dewar R, Rooke S, Gallagher M, Balcaza C, O'Toole A, Hill V, McCrone JT, Colquhoun R, Yu X, Jackson B, Scher E, Rambaut A, Williams TC, Templeton K
EPI_ISL_426024	hCoV-19/Scotland/EDB566/2020	Europe / United Kingdom / Scotland	2020-03-31	Virology Department, Royal Infirmary of Edinburgh, NHS Lothian / School of Biological Sciences, University of Edinburgh / Institute of Genetics and Molecular Medicine, University of Edinburgh	COVID-19 Genomics UK (COG-UK) Consortium	McHugh M, Dewar R, Rooke S, Gallagher M, Balcaza C, O'Toole A, Hill V, McCrone JT, Colquhoun R, Yu X, Jackson B, Scher E, Rambaut A, Williams TC, Templeton K
EPI_ISL_426025	hCoV-19/USA/NY-Wadsworth-10496-01/2020	North America / USA / New York	2020-03-03	Wadsworth Center, New York State Department.of Health	Wadsworth Center, New York State Department.of Health	Kirsten St. George, Daryl M. Lamson, Sara Griesemer, Jonathan Plitnick, Navjot Singh, Matthew D. Shudt, Erica Lasek-Nesselquist
EPI_ISL_426026	hCoV-19/USA/NY-Wadsworth-10528-01/2020	North America / USA / New York	2020-03-03	Wadsworth Center, New York State Department.of Health	Wadsworth Center, New York State Department.of Health	Kirsten St. George, Daryl M. Lamson, Sara Griesemer, Jonathan Plitnick, Navjot Singh, Matthew D. Shudt, Erica Lasek-Nesselquist
EPI_ISL_426027	hCoV-19/USA/NY-Wadsworth-10683-01/2020	North America / USA / New York	2020-03-04	Wadsworth Center, New York State Department.of Health	Wadsworth Center, New York State Department.of Health	Kirsten St. George, Daryl M. Lamson, Sara Griesemer, Jonathan Plitnick, Navjot Singh, Matthew D. Shudt, Erica Lasek-Nesselquist
EPI_ISL_426028	hCoV-19/USA/NY-Wadsworth-10690-01/2020	North America / USA / New York	2020-03-04	Wadsworth Center, New York State Department.of Health	Wadsworth Center, New York State Department.of Health	Kirsten St. George, Daryl M. Lamson, Sara Griesemer, Jonathan Plitnick, Navjot Singh, Matthew D. Shudt, Erica Lasek-Nesselquist
EPI_ISL_426029	hCoV-19/USA/NY-Wadsworth-10695-01/2020	North America / USA / New York	2020-03-04	Wadsworth Center, New York State Department.of Health	Wadsworth Center, New York State Department.of Health	Kirsten St. George, Daryl M. Lamson, Sara Griesemer, Jonathan Plitnick, Navjot Singh, Matthew D. Shudt, Erica Lasek-Nesselquist
EPI_ISL_426030	hCoV-19/USA/NY-Wadsworth-10703-01/2020	North America / USA / New York	2020-03-04	Wadsworth Center, New York State Department.of Health	Wadsworth Center, New York State Department.of Health	Kirsten St. George, Daryl M. Lamson, Sara Griesemer, Jonathan Plitnick, Navjot Singh, Matthew D. Shudt, Erica Lasek-Nesselquist
EPI_ISL_426031	hCoV-19/USA/NY-Wadsworth-10704-01/2020	North America / USA / New York	2020-03-04	Wadsworth Center, New York State Department.of Health	Wadsworth Center, New York State Department.of Health	Kirsten St. George, Daryl M. Lamson, Sara Griesemer, Jonathan Plitnick, Navjot Singh, Matthew D. Shudt, Erica Lasek-Nesselquist
EPI_ISL_426032	hCoV-19/USA/NY-Wadsworth-10707-01/2020	North America / USA / New York	2020-03-04	Wadsworth Center, New York State Department.of Health	Wadsworth Center, New York State Department.of Health	Kirsten St. George, Daryl M. Lamson, Sara Griesemer, Jonathan Plitnick, Navjot Singh, Matthew D. Shudt, Erica Lasek-Nesselquist
EPI_ISL_426033	hCoV-19/USA/NY-Wadsworth-10935-01/2020	North America / USA / New York	2020-03-04	Wadsworth Center, New York State Department.of Health	Wadsworth Center, New York State Department.of Health	Kirsten St. George, Daryl M. Lamson, Sara Griesemer, Jonathan Plitnick, Navjot Singh, Matthew D. Shudt, Erica Lasek-Nesselquist
EPI_ISL_426034	hCoV-19/USA/NY-Wadsworth-10955-01/2020	North America / USA / New York	2020-03-04	Wadsworth Center, New York State Department.of Health	Wadsworth Center, New York State Department.of Health	Kirsten St. George, Daryl M. Lamson, Sara Griesemer, Jonathan Plitnick, Navjot Singh, Matthew D. Shudt, Erica Lasek-Nesselquist
EPI_ISL_426035	hCoV-19/USA/NY-Wadsworth-10957-02/2020	North America / USA / New York	2020-03-04	Wadsworth Center, New York State Department.of Health	Wadsworth Center, New York State Department.of Health	Kirsten St. George, Daryl M. Lamson, Sara Griesemer, Jonathan Plitnick, Navjot Singh, Matthew D. Shudt, Erica Lasek-Nesselquist
EPI_ISL_426036	hCoV-19/USA/NY-Wadsworth-10958-01/2020	North America / USA / New York	2020-03-04	Wadsworth Center, New York State Department.of Health	Wadsworth Center, New York State Department.of Health	Kirsten St. George, Daryl M. Lamson, Sara Griesemer, Jonathan Plitnick, Navjot Singh, Matthew D. Shudt, Erica

Lasek-Nesselquist
EPI_ISL_426037 hCoV-19/USA/NY-Wadsworth-10999-01/2020 North America / USA / New York 2020-03-05
Wadsworth Center, New York State Department.of Health Wadsworth Center, New York State Department.of Health
Kirsten St. George, Daryl M. Lamson, Sara Griesemer, Jonathan Plitnick, Navjot Singh, Matthew D. Shudt, Erica
Lasek-Nesselquist
EPI_ISL_426038 hCoV-19/USA/NY-Wadsworth-11000-01/2020 North America / USA / New York 2020-03-05
Wadsworth Center, New York State Department.of Health Wadsworth Center, New York State Department.of Health
Kirsten St. George, Daryl M. Lamson, Sara Griesemer, Jonathan Plitnick, Navjot Singh, Matthew D. Shudt, Erica
Lasek-Nesselquist
EPI_ISL_426039 hCoV-19/USA/NY-Wadsworth-11003-01/2020 North America / USA / New York 2020-03-05
Wadsworth Center, New York State Department.of Health Wadsworth Center, New York State Department.of Health
Kirsten St. George, Daryl M. Lamson, Sara Griesemer, Jonathan Plitnick, Navjot Singh, Matthew D. Shudt, Erica
Lasek-Nesselquist
EPI_ISL_426040 hCoV-19/USA/NY-Wadsworth-11017-01/2020 North America / USA / New York 2020-03-05
Wadsworth Center, New York State Department.of Health Wadsworth Center, New York State Department.of Health
Kirsten St. George, Daryl M. Lamson, Sara Griesemer, Jonathan Plitnick, Navjot Singh, Matthew D. Shudt, Erica
Lasek-Nesselquist
EPI_ISL_426041 hCoV-19/USA/NY-Wadsworth-11180-01/2020 North America / USA / New York 2020-03-06
Wadsworth Center, New York State Department.of Health Wadsworth Center, New York State Department.of Health
Kirsten St. George, Daryl M. Lamson, Sara Griesemer, Jonathan Plitnick, Navjot Singh, Matthew D. Shudt, Erica
Lasek-Nesselquist
EPI_ISL_426042 hCoV-19/USA/NY-Wadsworth-11202-01/2020 North America / USA / New York 2020-03-06
Wadsworth Center, New York State Department.of Health Wadsworth Center, New York State Department.of Health
Kirsten St. George, Daryl M. Lamson, Sara Griesemer, Jonathan Plitnick, Navjot Singh, Matthew D. Shudt, Erica
Lasek-Nesselquist
EPI_ISL_426043 hCoV-19/USA/NY-Wadsworth-11284-01/2020 North America / USA / New York 2020-03-06
Wadsworth Center, New York State Department.of Health Wadsworth Center, New York State Department.of Health
Kirsten St. George, Daryl M. Lamson, Sara Griesemer, Jonathan Plitnick, Navjot Singh, Matthew D. Shudt, Erica
Lasek-Nesselquist
EPI_ISL_426044 hCoV-19/USA/NY-Wadsworth-11291-01/2020 North America / USA / New York 2020-03-06
Wadsworth Center, New York State Department.of Health Wadsworth Center, New York State Department.of Health
Kirsten St. George, Daryl M. Lamson, Sara Griesemer, Jonathan Plitnick, Navjot Singh, Matthew D. Shudt, Erica
Lasek-Nesselquist
EPI_ISL_426045 hCoV-19/USA/NY-Wadsworth-11344-01/2020 North America / USA / New York 2020-03-06
Wadsworth Center, New York State Department.of Health Wadsworth Center, New York State Department.of Health
Kirsten St. George, Daryl M. Lamson, Sara Griesemer, Jonathan Plitnick, Navjot Singh, Matthew D. Shudt, Erica
Lasek-Nesselquist
EPI_ISL_426046 hCoV-19/USA/NY-Wadsworth-11353-01/2020 North America / USA / New York 2020-03-06
Wadsworth Center, New York State Department.of Health Wadsworth Center, New York State Department.of Health
Kirsten St. George, Daryl M. Lamson, Sara Griesemer, Jonathan Plitnick, Navjot Singh, Matthew D. Shudt, Erica
Lasek-Nesselquist
EPI_ISL_426047 hCoV-19/USA/NY-Wadsworth-11354-01/2020 North America / USA / New York 2020-03-06
Wadsworth Center, New York State Department.of Health Wadsworth Center, New York State Department.of Health
Kirsten St. George, Daryl M. Lamson, Sara Griesemer, Jonathan Plitnick, Navjot Singh, Matthew D. Shudt, Erica
Lasek-Nesselquist
EPI_ISL_426048 hCoV-19/USA/NY-Wadsworth-11379-01/2020 North America / USA / New York 2020-03-06
Wadsworth Center, New York State Department.of Health Wadsworth Center, New York State Department.of Health
Kirsten St. George, Daryl M. Lamson, Sara Griesemer, Jonathan Plitnick, Navjot Singh, Matthew D. Shudt, Erica
Lasek-Nesselquist
EPI_ISL_426049 hCoV-19/USA/NY-Wadsworth-11380-01/2020 North America / USA / New York 2020-03-06
Wadsworth Center, New York State Department.of Health Wadsworth Center, New York State Department.of Health
Kirsten St. George, Daryl M. Lamson, Sara Griesemer, Jonathan Plitnick, Navjot Singh, Matthew D. Shudt, Erica
Lasek-Nesselquist
EPI_ISL_426050 hCoV-19/USA/NY-Wadsworth-11666-01/2020 North America / USA / New York 2020-03-08
Wadsworth Center, New York State Department.of Health Wadsworth Center, New York State Department.of Health
Kirsten St. George, Daryl M. Lamson, Sara Griesemer, Jonathan Plitnick, Navjot Singh, Matthew D. Shudt, Erica
Lasek-Nesselquist
EPI_ISL_426051 hCoV-19/Czech Republic/2282/2020 Europe / Czech Republic / Prague 2020-03-20
Laboratory of Molecular Genetics, 2nd Faculty of Medicine, Charles University in Prague, Prague, Czech Republic
Laboratory of Molecular Genetics, 2nd Faculty of Medicine, Charles University in Prague, Prague, Czech Republic
Lenka Kramná, Kateřina Poláčková, Ondřej Cinek
EPI_ISL_426052 hCoV-19/USA/CT-UW-4344/2020 North America / USA / Connecticut 2020-03-30 UW
Virology Lab UW Virology Lab Pavitra Roychoudhury, Hong Xie, Keith Jerome, Alexander Greninger
EPI_ISL_426053 hCoV-19/USA/CT-UW-4346/2020 North America / USA / Connecticut 2020-03-30 UW
Virology Lab UW Virology Lab Pavitra Roychoudhury, Hong Xie, Keith Jerome, Alexander Greninger
EPI_ISL_426054 hCoV-19/USA/CT-UW-4347/2020 North America / USA / Connecticut 2020-03-30 UW
Virology Lab UW Virology Lab Pavitra Roychoudhury, Hong Xie, Keith Jerome, Alexander Greninger
EPI_ISL_426055 hCoV-19/USA/CT-UW-4366/2020 North America / USA / Connecticut 2020-03-30 UW
Virology Lab UW Virology Lab Pavitra Roychoudhury, Hong Xie, Keith Jerome, Alexander Greninger
EPI_ISL_426056 hCoV-19/USA/CT-UW-4372/2020 North America / USA / Connecticut 2020-03-30 UW

Virology Lab	UW Virology Lab Pavitra Roychoudhury, Hong Xie, Keith Jerome, Alexander Greninger
EPI_ISL_426057	hCoV-19/USA/ID-UW-2127/2020	North America / USA / Idaho	2020-03-21	UW Virology Lab
UW Virology Lab Pavitra Roychoudhury, Hong Xie, Keith Jerome, Alexander Greninger
EPI_ISL_426058	hCoV-19/USA/ID-UW-2134/2020	North America / USA / Idaho	2020-03-24	UW Virology Lab
UW Virology Lab Pavitra Roychoudhury, Hong Xie, Keith Jerome, Alexander Greninger
EPI_ISL_426059	hCoV-19/USA/ID-UW-2159/2020	North America / USA / Idaho	2020-03-23	UW Virology Lab
UW Virology Lab Pavitra Roychoudhury, Hong Xie, Keith Jerome, Alexander Greninger
EPI_ISL_426060	hCoV-19/USA/ID-UW-2160/2020	North America / USA / Idaho	2020-03-23	UW Virology Lab
UW Virology Lab Pavitra Roychoudhury, Hong Xie, Keith Jerome, Alexander Greninger
EPI_ISL_426061	hCoV-19/USA/ID-UW-2174/2020	North America / USA / Idaho	2020-03-23	UW Virology Lab
UW Virology Lab Pavitra Roychoudhury, Hong Xie, Keith Jerome, Alexander Greninger
EPI_ISL_426062	hCoV-19/USA/ID-UW-2198/2020	North America / USA / Idaho	2020-03-24	UW Virology Lab
UW Virology Lab Pavitra Roychoudhury, Hong Xie, Keith Jerome, Alexander Greninger
EPI_ISL_426063	hCoV-19/USA/ID-UW-2255/2020	North America / USA / Idaho	2020-03-23	UW Virology Lab
UW Virology Lab Pavitra Roychoudhury, Hong Xie, Keith Jerome, Alexander Greninger
EPI_ISL_426064	hCoV-19/USA/ID-UW-4355/2020	North America / USA / Idaho	2020-03-27	UW Virology Lab
UW Virology Lab Pavitra Roychoudhury, Hong Xie, Keith Jerome, Alexander Greninger
EPI_ISL_426065	hCoV-19/USA/ID-UW-4357/2020	North America / USA / Idaho	2020-03-27	UW Virology Lab
UW Virology Lab Pavitra Roychoudhury, Hong Xie, Keith Jerome, Alexander Greninger
EPI_ISL_426066	hCoV-19/USA/ID-UW-4375/2020	North America / USA / Idaho	2020-03-28	UW Virology Lab
UW Virology Lab Pavitra Roychoudhury, Hong Xie, Keith Jerome, Alexander Greninger
EPI_ISL_426067	hCoV-19/USA/ID-UW-4378/2020	North America / USA / Idaho	2020-03-28	UW Virology Lab
UW Virology Lab Pavitra Roychoudhury, Hong Xie, Keith Jerome, Alexander Greninger
EPI_ISL_426068	hCoV-19/USA/ID-UW-4380/2020	North America / USA / Idaho	2020-03-27	UW Virology Lab
UW Virology Lab Pavitra Roychoudhury, Hong Xie, Keith Jerome, Alexander Greninger
EPI_ISL_426069	hCoV-19/USA/ID-UW-4396/2020	North America / USA / Idaho	2020-03-28	UW Virology Lab
UW Virology Lab Pavitra Roychoudhury, Hong Xie, Keith Jerome, Alexander Greninger
EPI_ISL_426070	hCoV-19/USA/ID-UW-4400/2020	North America / USA / Idaho	2020-03-28	UW Virology Lab
UW Virology Lab Pavitra Roychoudhury, Hong Xie, Keith Jerome, Alexander Greninger
EPI_ISL_426072	hCoV-19/USA/ID-UW-4427/2020	North America / USA / Idaho	2020-03-30	UW Virology Lab
UW Virology Lab Pavitra Roychoudhury, Hong Xie, Keith Jerome, Alexander Greninger
EPI_ISL_426073	hCoV-19/USA/ID-UW-4433/2020	North America / USA / Idaho	2020-03-27	UW Virology Lab
UW Virology Lab Pavitra Roychoudhury, Hong Xie, Keith Jerome, Alexander Greninger
EPI_ISL_426074	hCoV-19/USA/ID-UW-4448/2020	North America / USA / Idaho	2020-03-29	UW Virology Lab
UW Virology Lab Pavitra Roychoudhury, Hong Xie, Keith Jerome, Alexander Greninger
EPI_ISL_426075	hCoV-19/USA/ID-UW-4461/2020	North America / USA / Idaho	2020-03-29	UW Virology Lab
UW Virology Lab Pavitra Roychoudhury, Hong Xie, Keith Jerome, Alexander Greninger
EPI_ISL_426076	hCoV-19/USA/OR-UW-4388/2020	North America / USA / Oregon	2020-03-29	UW Virology Lab
UW Virology Lab Pavitra Roychoudhury, Hong Xie, Keith Jerome, Alexander Greninger
EPI_ISL_426077	hCoV-19/USA/UN-UW-2069/2020	North America / USA	2020-03-21	UW Virology Lab UW
Virology Lab	Pavitra Roychoudhury, Hong Xie, Keith Jerome, Alexander Greninger
EPI_ISL_426078	hCoV-19/USA/UN-UW-2139/2020	North America / USA	2020-03-23	UW Virology Lab UW
Virology Lab	Pavitra Roychoudhury, Hong Xie, Keith Jerome, Alexander Greninger
EPI_ISL_426079	hCoV-19/USA/WA-UW-2072/2020	North America / USA / Washington	2020-03-24	UW
Virology Lab	UW Virology Lab Pavitra Roychoudhury, Hong Xie, Keith Jerome, Alexander Greninger
EPI_ISL_426080	hCoV-19/USA/WA-UW-2074/2020	North America / USA / Washington	2020-03-25	UW
Virology Lab	UW Virology Lab Pavitra Roychoudhury, Hong Xie, Keith Jerome, Alexander Greninger
EPI_ISL_426081	hCoV-19/USA/WA-UW-2078/2020	North America / USA / Washington	2020-03-23	UW
Virology Lab	UW Virology Lab Pavitra Roychoudhury, Hong Xie, Keith Jerome, Alexander Greninger
EPI_ISL_426082	hCoV-19/USA/WA-UW-2082/2020	North America / USA / Washington	2020-03-25	UW
Virology Lab	UW Virology Lab Pavitra Roychoudhury, Hong Xie, Keith Jerome, Alexander Greninger
EPI_ISL_426083	hCoV-19/USA/WA-UW-2087/2020	North America / USA / Washington	2020-03-25	UW
Virology Lab	UW Virology Lab Pavitra Roychoudhury, Hong Xie, Keith Jerome, Alexander Greninger
EPI_ISL_426084	hCoV-19/USA/WA-UW-2091/2020	North America / USA / Washington	2020-03-24	UW
Virology Lab	UW Virology Lab Pavitra Roychoudhury, Hong Xie, Keith Jerome, Alexander Greninger
EPI_ISL_426085	hCoV-19/USA/WA-UW-2094/2020	North America / USA / Washington	2020-03-25	UW
Virology Lab	UW Virology Lab Pavitra Roychoudhury, Hong Xie, Keith Jerome, Alexander Greninger
EPI_ISL_426086	hCoV-19/USA/WA-UW-2100/2020	North America / USA / Washington	2020-03-25	UW
Virology Lab	UW Virology Lab Pavitra Roychoudhury, Hong Xie, Keith Jerome, Alexander Greninger
EPI_ISL_426087	hCoV-19/USA/WA-UW-2104/2020	North America / USA / Washington	2020-03-24	UW
Virology Lab	UW Virology Lab Pavitra Roychoudhury, Hong Xie, Keith Jerome, Alexander Greninger
EPI_ISL_426089	hCoV-19/USA/WA-UW-2108/2020	North America / USA / Washington	2020-03-24	UW
Virology Lab	UW Virology Lab Pavitra Roychoudhury, Hong Xie, Keith Jerome, Alexander Greninger
EPI_ISL_426090	hCoV-19/USA/WA-UW-2112/2020	North America / USA / Washington	2020-03-25	UW
Virology Lab	UW Virology Lab Pavitra Roychoudhury, Hong Xie, Keith Jerome, Alexander Greninger
EPI_ISL_426091	hCoV-19/USA/WA-UW-2119/2020	North America / USA / Washington	2020-03-25	UW
Virology Lab	UW Virology Lab Pavitra Roychoudhury, Hong Xie, Keith Jerome, Alexander Greninger
EPI_ISL_426092	hCoV-19/USA/WA-UW-2131/2020	North America / USA / Washington	2020-03-25	UW
Virology Lab	UW Virology Lab Pavitra Roychoudhury, Hong Xie, Keith Jerome, Alexander Greninger
EPI_ISL_426093	hCoV-19/USA/WA-UW-2132/2020	North America / USA / Washington	2020-03-25	UW

Virology Lab	UW Virology Lab Pavitra Roychoudhury, Hong Xie, Keith Jerome, Alexander Greninger
EPI_ISL_426094	hCoV-19/USA/WA-UW-2137/2020	North America / USA / Washington	2020-03-24	UW Virology Lab	UW Virology Lab Pavitra Roychoudhury, Hong Xie, Keith Jerome, Alexander Greninger
EPI_ISL_426095	hCoV-19/USA/WA-UW-2140/2020	North America / USA / Washington	2020-03-25	UW Virology Lab	UW Virology Lab Pavitra Roychoudhury, Hong Xie, Keith Jerome, Alexander Greninger
EPI_ISL_426096	hCoV-19/USA/WA-UW-2141/2020	North America / USA / Washington	2020-03-25	UW Virology Lab	UW Virology Lab Pavitra Roychoudhury, Hong Xie, Keith Jerome, Alexander Greninger
EPI_ISL_426097	hCoV-19/USA/WA-UW-2142/2020	North America / USA / Washington	2020-03-25	UW Virology Lab	UW Virology Lab Pavitra Roychoudhury, Hong Xie, Keith Jerome, Alexander Greninger
EPI_ISL_426098	hCoV-19/USA/WA-UW-2144/2020	North America / USA / Washington	2020-03-25	UW Virology Lab	UW Virology Lab Pavitra Roychoudhury, Hong Xie, Keith Jerome, Alexander Greninger
EPI_ISL_426099	hCoV-19/USA/WA-UW-2145/2020	North America / USA / Washington	2020-03-25	UW Virology Lab	UW Virology Lab Pavitra Roychoudhury, Hong Xie, Keith Jerome, Alexander Greninger
EPI_ISL_427300	hCoV-19/Brazil/RJ-477/2020	South America / Brazil / Rio de Janeiro / Rio de Janeiro	2020-03-11	Instituto Oswaldo Cruz FIOCRUZ - Laboratory of Respiratory Viruses and Measles (LVRS)	Instituto Oswaldo Cruz FIOCRUZ - Laboratory of Respiratory Viruses and Measles (LVRS)	Paola Resende, Fernando Motta, Luciana Appolinario, Sunando Roy, Aline Mattos, Milene Miranda, Cristiana Garcia, Braulia Caetano, Maria Ogrzewalska, Priscila Born, Jonathan Lopes, Marilda Siqueira
EPI_ISL_427301	hCoV-19/Brazil/RJ-477i/2020	South America / Brazil / Rio de Janeiro / Rio de Janeiro	2020-03-11	Instituto Oswaldo Cruz FIOCRUZ - Laboratory of Respiratory Viruses and Measles (LVRS)	Instituto Oswaldo Cruz FIOCRUZ - Laboratory of Respiratory Viruses and Measles (LVRS)	Paola Resende, Fernando Motta, Luciana Appolinario, Sunando Roy, Aline Mattos, Milene Miranda, Cristiana Garcia, Braulia Caetano, Maria Ogrzewalska, Priscila Born, Jonathan Lopes, Marilda Siqueira
EPI_ISL_427302	hCoV-19/Brazil/RJ-763/2020	South America / Brazil / Rio de Janeiro / Rio de Janeiro	2020-03-20	Instituto Oswaldo Cruz FIOCRUZ - Laboratory of Respiratory Viruses and Measles (LVRS)	Instituto Oswaldo Cruz FIOCRUZ - Laboratory of Respiratory Viruses and Measles (LVRS)	Paola Resende, Fernando Motta, Luciana Appolinario, Sunando Roy, Aline Mattos, Milene Miranda, Cristiana Garcia, Braulia Caetano, Maria Ogrzewalska, Priscila Born, Jonathan Lopes, Marilda Siqueira
EPI_ISL_427303	hCoV-19/Brazil/RJ-818/2020	South America / Brazil / Rio de Janeiro / Rio de Janeiro	2020-03-25	Instituto Oswaldo Cruz FIOCRUZ - Laboratory of Respiratory Viruses and Measles (LVRS)	Instituto Oswaldo Cruz FIOCRUZ - Laboratory of Respiratory Viruses and Measles (LVRS)	Paola Resende, Fernando Motta, Luciana Appolinario, Sunando Roy, Aline Mattos, Milene Miranda, Cristiana Garcia, Braulia Caetano, Maria Ogrzewalska, Priscila Born, Jonathan Lopes, Marilda Siqueira
EPI_ISL_427304	hCoV-19/Brazil/RJ-872/2020	South America / Brazil / Rio de Janeiro / Rio de Janeiro	2020-03-26	Instituto Oswaldo Cruz FIOCRUZ - Laboratory of Respiratory Viruses and Measles (LVRS)	Instituto Oswaldo Cruz FIOCRUZ - Laboratory of Respiratory Viruses and Measles (LVRS)	Paola Resende, Fernando Motta, Luciana Appolinario, Sunando Roy, Aline Mattos, Milene Miranda, Cristiana Garcia, Braulia Caetano, Maria Ogrzewalska, Priscila Born, Jonathan Lopes, Marilda Siqueira
EPI_ISL_427305	hCoV-19/Brazil/SC-766/2020	South America / Brazil / Santa Catarina / Joinville	2020-03-10	LACEN-SC - Laboratorio Central de Santa Catarina	Instituto Oswaldo Cruz FIOCRUZ - Laboratory of Respiratory Viruses and Measles (LVRS)	Paola Resende, Fernando Motta, Luciana Appolinario, Sunando Roy, Aline Mattos, Milene Miranda, Cristiana Garcia, Braulia Caetano, Maria Ogrzewalska, Priscila Born, Jonathan Lopes, Marilda Siqueira
EPI_ISL_427306	hCoV-19/Brazil/SC-769/2020	South America / Brazil / Santa Catarina / Florianopolis	2020-03-10	LACEN-SC - Laboratorio Central de Santa Catarina	Instituto Oswaldo Cruz FIOCRUZ - Laboratory of Respiratory Viruses and Measles (LVRS)	Paola Resende, Fernando Motta, Luciana Appolinario, Sunando Roy, Aline Mattos, Milene Miranda, Cristiana Garcia, Braulia Caetano, Maria Ogrzewalska, Priscila Born, Jonathan Lopes, Marilda Siqueira
EPI_ISL_427307	hCoV-19/Russia/StPetersburg-RII4144S/2020	Europe / Russia / Saint-Petersburg	2020-03-20	WHO National Influenza Centre Russian Federation	WHO National Influenza Centre Russian Federation	Andrey Komissarov, Artem Fadeev, Mariia Sergeeva, Anna Ivanova, Daria Danilenko
EPI_ISL_427308	hCoV-19/Russia/StPetersburg-RII4307V/2020	Europe / Russia / Saint-Petersburg	2020-03-24	WHO National Influenza Centre Russian Federation	WHO National Influenza Centre Russian Federation	Andrey Komissarov, Artem Fadeev, Mariia Sergeeva, Anna Ivanova, Daria Danilenko
EPI_ISL_427309	hCoV-19/Russia/StPetersburg-RII4308V/2020	Europe / Russia / Saint-Petersburg	2020-03-24	WHO National Influenza Centre Russian Federation	WHO National Influenza Centre Russian Federation	Andrey Komissarov, Artem Fadeev, Mariia Sergeeva, Anna Ivanova, Daria Danilenko
EPI_ISL_427310	hCoV-19/Russia/StPetersburg-RII4382V/2020	Europe / Russia / Saint-Petersburg	2020-03-29	WHO National Influenza Centre Russian Federation	WHO National Influenza Centre Russian Federation	Andrey Komissarov, Artem Fadeev, Mariia Sergeeva, Anna Ivanova, Daria Danilenko
EPI_ISL_427311	hCoV-19/Russia/StPetersburg-RII4384V/2020	Europe / Russia / Saint-Petersburg	2020-03-29	WHO National Influenza Centre Russian Federation	WHO National Influenza Centre Russian Federation	Andrey Komissarov, Artem Fadeev, Mariia Sergeeva, Anna Ivanova, Daria Danilenko
EPI_ISL_427312	hCoV-19/Russia/StPetersburg-RII4386V/2020	Europe / Russia / Saint-Petersburg	2020-03-29	WHO National Influenza Centre Russian Federation	WHO National Influenza Centre Russian Federation	Andrey Komissarov, Artem Fadeev, Mariia Sergeeva, Anna Ivanova, Daria Danilenko
EPI_ISL_427313	hCoV-19/Russia/StPetersburg-RII4532V/2020	Europe / Russia / Saint-Petersburg	2020-04-03	WHO National Influenza Centre Russian Federation	WHO National Influenza Centre Russian Federation	Andrey Komissarov, Artem Fadeev, Mariia Sergeeva, Anna Ivanova, Daria Danilenko
EPI_ISL_427314	hCoV-19/Russia/StPetersburg-RII4546V/2020	Europe / Russia / Saint-Petersburg	2020-

04-02	WHO National Influenza Centre Russian Federation	WHO National Influenza Centre Russian Federation	Andrey Komissarov, Artem Fadeev, Mariia Sergeeva, Anna Ivanova, Daria Danilenko
EPI_ISL_427316	hCoV-19/Russia/StPetersburg-RII4584S/2020	Europe / Russia / Saint-Petersburg	2020-04-03	WHO National Influenza Centre Russian Federation	WHO National Influenza Centre Russian Federation	Andrey Komissarov, Artem Fadeev, Mariia Sergeeva, Anna Ivanova, Daria Danilenko
EPI_ISL_427317	hCoV-19/Russia/StPetersburg-RII4585S/2020	Europe / Russia / Saint-Petersburg	2020-04-03	WHO National Influenza Centre Russian Federation	WHO National Influenza Centre Russian Federation	Andrey Komissarov, Artem Fadeev, Mariia Sergeeva, Anna Ivanova, Daria Danilenko
EPI_ISL_427318	hCoV-19/Russia/StPetersburg-RII4591S/2020	Europe / Russia / Saint-Petersburg	2020-04-03	WHO National Influenza Centre Russian Federation	WHO National Influenza Centre Russian Federation	Andrey Komissarov, Artem Fadeev, Mariia Sergeeva, Anna Ivanova, Daria Danilenko
EPI_ISL_427319	hCoV-19/Russia/StPetersburg-RII4597S/2020	Europe / Russia / Saint-Petersburg	2020-04-04	WHO National Influenza Centre Russian Federation	WHO National Influenza Centre Russian Federation	Andrey Komissarov, Artem Fadeev, Mariia Sergeeva, Anna Ivanova, Daria Danilenko
EPI_ISL_427320	hCoV-19/Russia/StPetersburg-RII4637S/2020	Europe / Russia / Saint-Petersburg	2020-04-05	WHO National Influenza Centre Russian Federation	WHO National Influenza Centre Russian Federation	Andrey Komissarov, Artem Fadeev, Mariia Sergeeva, Anna Ivanova, Daria Danilenko
EPI_ISL_427321	hCoV-19/Russia/StPetersburg-RII4649S/2020	Europe / Russia / Saint-Petersburg	2020-04-05	WHO National Influenza Centre Russian Federation	WHO National Influenza Centre Russian Federation	Andrey Komissarov, Artem Fadeev, Mariia Sergeeva, Anna Ivanova, Daria Danilenko
EPI_ISL_427322	hCoV-19/Russia/StPetersburg-RII4655S/2020	Europe / Russia / Saint-Petersburg	2020-04-05	WHO National Influenza Centre Russian Federation	WHO National Influenza Centre Russian Federation	Andrey Komissarov, Artem Fadeev, Mariia Sergeeva, Anna Ivanova, Daria Danilenko
EPI_ISL_427323	hCoV-19/Russia/StPetersburg-RII4678S/2020	Europe / Russia / Saint-Petersburg	2020-04-06	WHO National Influenza Centre Russian Federation	WHO National Influenza Centre Russian Federation	Andrey Komissarov, Artem Fadeev, Mariia Sergeeva, Anna Ivanova, Daria Danilenko
EPI_ISL_427324	hCoV-19/Russia/StPetersburg-RII4686S/2020	Europe / Russia / Saint-Petersburg	2020-04-07	WHO National Influenza Centre Russian Federation	WHO National Influenza Centre Russian Federation	Andrey Komissarov, Artem Fadeev, Mariia Sergeeva, Anna Ivanova, Daria Danilenko
EPI_ISL_427325	hCoV-19/Russia/StPetersburg-RII4693S/2020	Europe / Russia / Saint-Petersburg	2020-04-07	WHO National Influenza Centre Russian Federation	WHO National Influenza Centre Russian Federation	Andrey Komissarov, Artem Fadeev, Mariia Sergeeva, Anna Ivanova, Daria Danilenko
EPI_ISL_427326	hCoV-19/Russia/StPetersburg-RII4694S/2020	Europe / Russia / Saint-Petersburg	2020-04-07	WHO National Influenza Centre Russian Federation	WHO National Influenza Centre Russian Federation	Andrey Komissarov, Artem Fadeev, Mariia Sergeeva, Anna Ivanova, Daria Danilenko
EPI_ISL_427327	hCoV-19/Russia/StPetersburg-RII4696S/2020	Europe / Russia / Saint-Petersburg	2020-04-07	WHO National Influenza Centre Russian Federation	WHO National Influenza Centre Russian Federation	Andrey Komissarov, Artem Fadeev, Mariia Sergeeva, Anna Ivanova, Daria Danilenko
EPI_ISL_427328	hCoV-19/Russia/StPetersburg-RII4697S/2020	Europe / Russia / Saint-Petersburg	2020-04-07	WHO National Influenza Centre Russian Federation	WHO National Influenza Centre Russian Federation	Andrey Komissarov, Artem Fadeev, Mariia Sergeeva, Anna Ivanova, Daria Danilenko
EPI_ISL_427329	hCoV-19/Russia/StPetersburg-RII4707S/2020	Europe / Russia / Saint-Petersburg	2020-04-07	WHO National Influenza Centre Russian Federation	WHO National Influenza Centre Russian Federation	Andrey Komissarov, Artem Fadeev, Mariia Sergeeva, Anna Ivanova, Daria Danilenko
EPI_ISL_427330	hCoV-19/Russia/StPetersburg-RII4711S/2020	Europe / Russia / Saint-Petersburg	2020-04-07	WHO National Influenza Centre Russian Federation	WHO National Influenza Centre Russian Federation	Andrey Komissarov, Artem Fadeev, Mariia Sergeeva, Anna Ivanova, Daria Danilenko
EPI_ISL_427331	hCoV-19/Russia/StPetersburg-RII4713S/2020	Europe / Russia / Saint-Petersburg	2020-04-07	WHO National Influenza Centre Russian Federation	WHO National Influenza Centre Russian Federation	Andrey Komissarov, Artem Fadeev, Mariia Sergeeva, Anna Ivanova, Daria Danilenko
EPI_ISL_427332	hCoV-19/Russia/StPetersburg-RII4723S/2020	Europe / Russia / Saint-Petersburg	2020-04-07	WHO National Influenza Centre Russian Federation	WHO National Influenza Centre Russian Federation	Andrey Komissarov, Artem Fadeev, Mariia Sergeeva, Anna Ivanova, Daria Danilenko
EPI_ISL_427333	hCoV-19/Russia/StPetersburg-RII4724S/2020	Europe / Russia / Saint-Petersburg	2020-04-07	WHO National Influenza Centre Russian Federation	WHO National Influenza Centre Russian Federation	Andrey Komissarov, Artem Fadeev, Mariia Sergeeva, Anna Ivanova, Daria Danilenko
EPI_ISL_427334	hCoV-19/Russia/StPetersburg-RII4726S/2020	Europe / Russia / Saint-Petersburg	2020-04-07	WHO National Influenza Centre Russian Federation	WHO National Influenza Centre Russian Federation	Andrey Komissarov, Artem Fadeev, Mariia Sergeeva, Anna Ivanova, Daria Danilenko
EPI_ISL_427335	hCoV-19/Russia/StPetersburg-RII4728S/2020	Europe / Russia / Saint-Petersburg	2020-04-07	WHO National Influenza Centre Russian Federation	WHO National Influenza Centre Russian Federation	Andrey Komissarov, Artem Fadeev, Mariia Sergeeva, Anna Ivanova, Daria Danilenko
EPI_ISL_427336	hCoV-19/Russia/StPetersburg-RII4781S/2020	Europe / Russia / Saint-Petersburg	2020-04-07	WHO National Influenza Centre Russian Federation	WHO National Influenza Centre Russian Federation	Andrey Komissarov, Artem Fadeev, Mariia Sergeeva, Anna Ivanova, Daria Danilenko
EPI_ISL_427337	hCoV-19/Russia/StPetersburg-RII4326S/2020	Europe / Russia / Saint-Petersburg	2020-03-25	WHO National Influenza Centre Russian Federation	WHO National Influenza Centre Russian Federation	Andrey Komissarov, Artem Fadeev, Mariia Sergeeva, Anna Ivanova, Daria Danilenko
EPI_ISL_427338	hCoV-19/Russia/StPetersburg-RII4328S/2020	Europe / Russia / Saint-Petersburg	2020-03-25	WHO National Influenza Centre Russian Federation	WHO National Influenza Centre Russian

Federation	Andrey Komissarov, Artem Fadeev, Mariia Sergeeva, Anna Ivanova, Daria Danilenko
EPI_ISL_427339	hCoV-19/Russia/StPetersburg-RII4332S/2020	Europe / Russia / Saint-Petersburg	2020-03-25	WHO National Influenza Centre Russian Federation	WHO National Influenza Centre Russian Federation	Andrey Komissarov, Artem Fadeev, Mariia Sergeeva, Anna Ivanova, Daria Danilenko
EPI_ISL_427349	hCoV-19/Belgium/ULG-10098/2020	Europe / Belgium / Liege	2020-04-06	Department of Clinical Microbiology	GIGA Medical Genomics	Keith Durkin, Maria Artesi, Sébastien Bontems, Raphaël Boreux, Cécile Meex, Pierrette Melin, Marie-Pierre Hayette, Vincent Bours.
EPI_ISL_427350	hCoV-19/Belgium/ULG-10099/2020	Europe / Belgium / Liege	2020-04-06	Department of Clinical Microbiology	GIGA Medical Genomics	Keith Durkin, Maria Artesi, Sébastien Bontems, Raphaël Boreux, Cécile Meex, Pierrette Melin, Marie-Pierre Hayette, Vincent Bours.
EPI_ISL_427351	hCoV-19/Belgium/ULG-10100/2020	Europe / Belgium / Liege	2020-04-07	Department of Clinical Microbiology	GIGA Medical Genomics	Keith Durkin, Maria Artesi, Sébastien Bontems, Raphaël Boreux, Cécile Meex, Pierrette Melin, Marie-Pierre Hayette, Vincent Bours.
EPI_ISL_427352	hCoV-19/Belgium/ULG-10101/2020	Europe / Belgium / Liege	2020-04-06	Department of Clinical Microbiology	GIGA Medical Genomics	Keith Durkin, Maria Artesi, Sébastien Bontems, Raphaël Boreux, Cécile Meex, Pierrette Melin, Marie-Pierre Hayette, Vincent Bours.
EPI_ISL_427354	hCoV-19/Belgium/ULG-10103/2020	Europe / Belgium / Liege	2020-04-06	Department of Clinical Microbiology	GIGA Medical Genomics	Keith Durkin, Maria Artesi, Sébastien Bontems, Raphaël Boreux, Cécile Meex, Pierrette Melin, Marie-Pierre Hayette, Vincent Bours.
EPI_ISL_427357	hCoV-19/Belgium/ULG-10106/2020	Europe / Belgium / Liege	2020-04-07	Department of Clinical Microbiology	GIGA Medical Genomics	Keith Durkin, Maria Artesi, Sébastien Bontems, Raphaël Boreux, Cécile Meex, Pierrette Melin, Marie-Pierre Hayette, Vincent Bours.
EPI_ISL_427358	hCoV-19/Belgium/ULG-10107/2020	Europe / Belgium / Liege	2020-04-06	Department of Clinical Microbiology	GIGA Medical Genomics	Keith Durkin, Maria Artesi, Sébastien Bontems, Raphaël Boreux, Cécile Meex, Pierrette Melin, Marie-Pierre Hayette, Vincent Bours.
EPI_ISL_427359	hCoV-19/Belgium/ULG-10108/2020	Europe / Belgium / Liege	2020-04-06	Department of Clinical Microbiology	GIGA Medical Genomics	Keith Durkin, Maria Artesi, Sébastien Bontems, Raphaël Boreux, Cécile Meex, Pierrette Melin, Marie-Pierre Hayette, Vincent Bours.
EPI_ISL_427364	hCoV-19/Belgium/ULG-10115/2020	Europe / Belgium / Liege	2020-04-07	Department of Clinical Microbiology	GIGA Medical Genomics	Keith Durkin, Maria Artesi, Sébastien Bontems, Raphaël Boreux, Cécile Meex, Pierrette Melin, Marie-Pierre Hayette, Vincent Bours.
EPI_ISL_427366	hCoV-19/Belgium/ULG-10118/2020	Europe / Belgium / Liege	2020-04-07	Department of Clinical Microbiology	GIGA Medical Genomics	Keith Durkin, Maria Artesi, Sébastien Bontems, Raphaël Boreux, Cécile Meex, Pierrette Melin, Marie-Pierre Hayette, Vincent Bours.
EPI_ISL_427370	hCoV-19/Belgium/ULG-10122/2020	Europe / Belgium / Liege	2020-04-07	Department of Clinical Microbiology	GIGA Medical Genomics	Keith Durkin, Maria Artesi, Sébastien Bontems, Raphaël Boreux, Cécile Meex, Pierrette Melin, Marie-Pierre Hayette, Vincent Bours.
EPI_ISL_427371	hCoV-19/Belgium/ULG-10124/2020	Europe / Belgium / Liege	2020-04-07	Department of Clinical Microbiology	GIGA Medical Genomics	Keith Durkin, Maria Artesi, Sébastien Bontems, Raphaël Boreux, Cécile Meex, Pierrette Melin, Marie-Pierre Hayette, Vincent Bours.
EPI_ISL_427372	hCoV-19/Belgium/ULG-10126/2020	Europe / Belgium / Liege	2020-04-07	Department of Clinical Microbiology	GIGA Medical Genomics	Keith Durkin, Maria Artesi, Sébastien Bontems, Raphaël Boreux, Cécile Meex, Pierrette Melin, Marie-Pierre Hayette, Vincent Bours.
EPI_ISL_427379	hCoV-19/Belgium/ULG-10134/2020	Europe / Belgium / Liege	2020-04-08	Department of Clinical Microbiology	GIGA Medical Genomics	Keith Durkin, Maria Artesi, Sébastien Bontems, Raphaël Boreux, Cécile Meex, Pierrette Melin, Marie-Pierre Hayette, Vincent Bours.
EPI_ISL_427385	hCoV-19/Belgium/ULG-10142/2020	Europe / Belgium / Liege	2020-04-08	Department of Clinical Microbiology	GIGA Medical Genomics	Keith Durkin, Maria Artesi, Sébastien Bontems, Raphaël Boreux, Cécile Meex, Pierrette Melin, Marie-Pierre Hayette, Vincent Bours.
EPI_ISL_427388	hCoV-19/Belgium/ULG-10144/2020	Europe / Belgium / Liege	2020-04-07	Department of Clinical Microbiology	GIGA Medical Genomics	Keith Durkin, Maria Artesi, Sébastien Bontems, Raphaël Boreux, Cécile Meex, Pierrette Melin, Marie-Pierre Hayette, Vincent Bours.
EPI_ISL_427390	hCoV-19/Belgium/ULG-10125/2020	Europe / Belgium / Liege	2020-04-07	Department of Clinical Microbiology	GIGA Medical Genomics	Keith Durkin, Maria Artesi, Sébastien Bontems, Raphaël Boreux, Cécile Meex, Pierrette Melin, Marie-Pierre Hayette, Vincent Bours.
EPI_ISL_427391	hCoV-19/Turkey/GLAB-CoV008/2020	Europe / Turkey / Istanbul	2020-04-13	Genomic Laboratory (GLAB) (Conjoint lab of Health Directorate of Istanbul and Istanbul Technical University)	Genomic Laboratory (GLAB), Istanbul Technical University	Ilker Karacan, Tugba Kizilboga Akgun, Bugra Agaoglu, Gizem Alkurt, Jale Yildiz, Betsi Köse, Elifnaz Çelik, Mehtap Aydın, Levent Doganay, Gizem Dinler Doganay
EPI_ISL_427392	hCoV-19/Taiwan/TSGH-05/2020	Asia / Taiwan / New Taipei City	2020-03-14	TSGH-CP molecular lab	TSGH-CP molecular lab	Cherng-Lih Perng, Ming-Jr Jian, Chih-Kai Chang, Jung-Chung Lin, Kuo-Ming Yeh, Chien-Wen Chen, Sheng-Kang Chiu, Hsing-Yi Chung, Shih-Hung Tsai, Kuo-Sheng Hung, Tien-Yao Chang, Feng-Yee Chang, Hung-Sheng Shang
EPI_ISL_427393	hCoV-19/Taiwan/TSGH-06/2020	Asia / Taiwan / Taipei	2020-03-15	TSGH-CP molecular lab	TSGH-CP molecular lab	Cherng-Lih Perng, Ming-Jr Jian, Chih-Kai Chang, Jung-Chung Lin, Kuo-Ming Yeh, Chien-Wen Chen, Sheng-Kang Chiu, Hsing-Yi Chung, Shih-Hung Tsai, Kuo-Sheng Hung, Tien-Yao Chang, Feng-Yee Chang, Hung-Sheng Shang
EPI_ISL_427394	hCoV-19/Taiwan/TSGH-07/2020	Asia / Taiwan / New Taipei City	2020-03-22	TSGH-CP molecular lab	TSGH-CP molecular lab	Cherng-Lih Perng, Ming-Jr Jian, Chih-Kai Chang, Jung-Chung Lin, Kuo-Ming Yeh, Chien-Wen Chen, Sheng-Kang Chiu, Hsing-Yi Chung, Shih-Hung Tsai, Kuo-Sheng Hung, Tien-Yao Chang,

Feng-Yee Chang, Hung-Sheng Shang
EPI_ISL_427395 hCoV-19/Taiwan/TSGH-08/2020 Asia / Taiwan / Keelung 2020-03-23 TSGH-CP molecular lab TSGH-CP molecular lab Cherng-Lih Perng, Ming-Jr Jian, Chih-Kai Chang, Jung-Chung Lin, Kuo-Ming Yeh, Chien-Wen Chen, Sheng-Kang Chiu, Hsing-Yi Chung, Shih-Hung Tsai, Kuo-Sheng Hung, Tien-Yao Chang, Feng-Yee Chang, Hung-Sheng Shang
EPI_ISL_427396 hCoV-19/Taiwan/TSGH-09/2020 Asia / Taiwan / New Taipei City 2020-03-23 TSGH-CP molecular lab TSGH-CP molecular lab Cherng-Lih Perng, Ming-Jr Jian, Chih-Kai Chang, Jung-Chung Lin, Kuo-Ming Yeh, Chien-Wen Chen, Sheng-Kang Chiu, Hsing-Yi Chung, Shih-Hung Tsai, Kuo-Sheng Hung, Tien-Yao Chang, Feng-Yee Chang, Hung-Sheng Shang
EPI_ISL_427397 hCoV-19/Taiwan/TSGH-10/2020 Asia / Taiwan / Taipei 2020-03-24 TSGH-CP molecular lab TSGH-CP molecular lab Cherng-Lih Perng, Ming-Jr Jian, Chih-Kai Chang, Jung-Chung Lin, Kuo-Ming Yeh, Chien-Wen Chen, Sheng-Kang Chiu, Hsing-Yi Chung, Shih-Hung Tsai, Kuo-Sheng Hung, Tien-Yao Chang, Feng-Yee Chang, Hung-Sheng Shang
EPI_ISL_427398 hCoV-19/Taiwan/TSGH-11/2020 Asia / Taiwan / New Taipei City 2020-03-25 TSGH-CP molecular lab TSGH-CP molecular lab Cherng-Lih Perng, Ming-Jr Jian, Chih-Kai Chang, Jung-Chung Lin, Kuo-Ming Yeh, Chien-Wen Chen, Sheng-Kang Chiu, Hsing-Yi Chung, Shih-Hung Tsai, Kuo-Sheng Hung, Tien-Yao Chang, Feng-Yee Chang, Hung-Sheng Shang
EPI_ISL_428670 hCoV-19/Sri Lanka/COV38/2020 Asia / Sri Lanka 2020-03-16 Centre for Dengue Research Centre for Dengue Research Chandima Jeewandara, Dinuka Ariyaratne, Laksiri Gomes, Deshni Jayathilaka, Ananda Wijewickrama, Eranga Narangoda, Damayanthi Idampitiya, Neelika Malaige
EPI_ISL_428671 hCoV-19/Sri Lanka/COV53/2020 Asia / Sri Lanka 2020-03-10 Centre for Dengue Research Centre for Dengue Research Chandima Jeewandara, Dinuka Ariyatane, Laksiri Gomes, Deshni Jayathilaka, Diyanath Ranasinghe, Ananda Wijewickrama, Eranga Narangoda, Damayanthi Tdampitiya, Neelika Malavige
EPI_ISL_428672 hCoV-19/Sri Lanka/COV91/2020 Asia / Sri Lanka 2020-03-19 Centre for Dengue Research Centre for Dengue Research Chandima Jeewandara, Dinuka Ariyaratne, Laksiri Gomes, Deshni Jayathilaka, Diyanath Ranasinghe, Ananda Wijewickrama, Eranga Narangoda, Damayanthi Idampitiya, Neelika Malavige
EPI_ISL_428673 hCoV-19/Sri Lanka/COV486/2020 Asia / Sri Lanka 2020-03-31 Centre for Dengue Research Centre for Dengue Research Chandima Jeewandara, Dinuka Ariyaratne, Laksiri Gomes, Deshni Jayathilaka, Diyanath Ranasinghe, Ananda Wijewickrama, Eranga Narangoda, Damayanthi Idampitiya, Neelika Malavige
EPI_ISL_428674 hCoV-19/Spain/Madrid_LP11_2271/2020 Europe / Spain / Madrid 2020-03-09 Hospital Universitario La Paz Hospital Universitario 12 de Octubre Elias Dahdouh, Sara González, Raúl Recio, Fernando Lázaro, Esther Viedma, Natalia Stella, Julio García, Juan Carlos Galán, Rafael Cantón, Mª Dolores Folgueira, Rafael Delgado, Jesús Mingorance
EPI_ISL_428675 hCoV-19/Spain/Madrid_LP16_6193/2020 Europe / Spain / Madrid 2020-03-09 Hospital Universitario La Paz Hospital Universitario 12 de Octubre Elias Dahdouh, Sara González, Raúl Recio, Fernando Lázaro, Esther Viedma, Natalia Stella, Julio García, Juan Carlos Galán, Rafael Cantón, Mª Dolores Folgueira, Rafael Delgado, Jesús Mingorance
EPI_ISL_428676 hCoV-19/Spain/Madrid_LP19_4952/2020 Europe / Spain / Madrid 2020-03-07 Hospital Universitario La Paz Hospital Universitario 12 de Octubre Elias Dahdouh, Sara González, Raúl Recio, Fernando Lázaro, Esther Viedma, Natalia Stella, Julio García, Juan Carlos Galán, Rafael Cantón, Mª Dolores Folgueira, Rafael Delgado, Jesús Mingorance
EPI_ISL_428677 hCoV-19/Spain/Madrid_LP20_2327/2020 Europe / Spain / Madrid 2020-03-11 Hospital Universitario La Paz Hospital Universitario 12 de Octubre Elias Dahdouh, Sara González, Raúl Recio, Fernando Lázaro, Esther Viedma, Natalia Stella, Julio García, Juan Carlos Galán, Rafael Cantón, Mª Dolores Folgueira, Rafael Delgado, Jesús Mingorance
EPI_ISL_428678 hCoV-19/Spain/Madrid_LP22_5885/2020 Europe / Spain / Madrid 2020-03-08 Hospital Universitario La Paz Hospital Universitario 12 de Octubre Elias Dahdouh, Sara González, Raúl Recio, Fernando Lázaro, Esther Viedma, Natalia Stella, Julio García, Juan Carlos Galán, Rafael Cantón, Mª Dolores Folgueira, Rafael Delgado, Jesús Mingorance
EPI_ISL_428679 hCoV-19/Spain/Madrid_LP23_5852/2020 Europe / Spain / Madrid 2020-03-08 Hospital Universitario La Paz Hospital Universitario 12 de Octubre Elias Dahdouh, Sara González, Raúl Recio, Fernando Lázaro, Esther Viedma, Natalia Stella, Julio García, Juan Carlos Galán, Rafael Cantón, Mª Dolores Folgueira, Rafael Delgado, Jesús Mingorance
EPI_ISL_428680 hCoV-19/Spain/Madrid_LP24_5999/2020 Europe / Spain / Madrid 2020-03-09 Hospital Universitario La Paz Hospital Universitario 12 de Octubre Elias Dahdouh, Sara González, Raúl Recio, Fernando Lázaro, Esther Viedma, Natalia Stella, Julio García, Juan Carlos Galán, Rafael Cantón, Mª Dolores Folgueira, Rafael Delgado, Jesús Mingorance
EPI_ISL_428681 hCoV-19/Spain/Madrid_LP27_2548/2020 Europe / Spain / Madrid 2020-03-11 Hospital Universitario La Paz Hospital Universitario 12 de Octubre Elias Dahdouh, Sara González, Raúl Recio, Fernando Lázaro, Esther Viedma, Natalia Stella, Julio García, Juan Carlos Galán, Rafael Cantón, Mª Dolores Folgueira, Rafael Delgado, Jesús Mingorance
EPI_ISL_428682 hCoV-19/Spain/Madrid_LP30_2226/2020 Europe / Spain / Madrid 2020-03-11 Hospital Universitario La Paz Hospital Universitario 12 de Octubre Elias Dahdouh, Sara González, Raúl Recio, Fernando Lázaro, Esther Viedma, Natalia Stella, Julio García, Juan Carlos Galán, Rafael Cantón, Mª Dolores Folgueira, Rafael Delgado, Jesús Mingorance
EPI_ISL_428683 hCoV-19/Spain/Madrid_H12_13/2020 Europe / Spain / Madrid 2020-03-03 Hospital Universitario 12 de Octubre Hospital Universitario 12 de Octubre Sara González, Raúl Recio,Elias

Dahdouh, Fernando Lázaro, Esther Viedma, Natalia Stella, Julio García, Juan Carlos Galán, Rafael Cantón, Mª Dolores Folgueira, Rafael Delgado, Jesús Mingorance
EPI_ISL_428684 hCoV-19/Spain/Madrid_H12_20/2020 Europe / Spain / Madrid 2020-03-27 Hospital Universitario 12 de Octubre Hospital Universitario 12 de Octubre Sara González, Raúl Recio,Elias Dahdouh, Fernando Lázaro, Esther Viedma, Natalia Stella, Julio García, Juan Carlos Galán, Rafael Cantón, Mª Dolores Folgueira, Rafael Delgado, Jesús Mingorance
EPI_ISL_428686 hCoV-19/Spain/Madrid_H12_23/2020 Europe / Spain / Madrid 2020-03-22 Hospital Universitario 12 de Octubre Hospital Universitario 12 de Octubre Sara González, Raúl Recio,Elias Dahdouh, Fernando Lázaro, Esther Viedma, Natalia Stella, Julio García, Juan Carlos Galán, Rafael Cantón, Mª Dolores Folgueira, Rafael Delgado, Jesús Mingorance
EPI_ISL_428687 hCoV-19/Spain/Madrid_H12_24/2020 Europe / Spain / Madrid 2020-03-22 Hospital Universitario 12 de Octubre Hospital Universitario 12 de Octubre Sara González, Raúl Recio,Elias Dahdouh, Fernando Lázaro, Esther Viedma, Natalia Stella, Julio García, Juan Carlos Galán, Rafael Cantón, Mª Dolores Folgueira, Rafael Delgado, Jesús Mingorance
EPI_ISL_428688 hCoV-19/Spain/Madrid_H12_25/2020 Europe / Spain / Madrid 2020-03-22 Hospital Universitario 12 de Octubre Hospital Universitario 12 de Octubre Sara González, Raúl Recio,Elias Dahdouh, Fernando Lázaro, Esther Viedma, Natalia Stella, Julio García, Juan Carlos Galán, Rafael Cantón, Mª Dolores Folgueira, Rafael Delgado, Jesús Mingorance
EPI_ISL_428689 hCoV-19/Spain/Madrid_H12_26/2020 Europe / Spain / Madrid 2020-03-26 Hospital Universitario 12 de Octubre Hospital Universitario 12 de Octubre Sara González, Raúl Recio,Elias Dahdouh, Fernando Lázaro, Esther Viedma, Natalia Stella, Julio García, Juan Carlos Galán, Rafael Cantón, Mª Dolores Folgueira, Rafael Delgado, Jesús Mingorance
EPI_ISL_428690 hCoV-19/Spain/Madrid_H12_27/2020 Europe / Spain / Madrid 2020-03-21 Hospital Universitario 12 de Octubre Hospital Universitario 12 de Octubre Sara González, Raúl Recio,Elias Dahdouh, Fernando Lázaro, Esther Viedma, Natalia Stella, Julio García, Juan Carlos Galán, Rafael Cantón, Mª Dolores Folgueira, Rafael Delgado, Jesús Mingorance
EPI_ISL_428691 hCoV-19/Spain/Madrid_H12_28/2020 Europe / Spain / Madrid 2020-03-21 Hospital Universitario 12 de Octubre Hospital Universitario 12 de Octubre Sara González, Raúl Recio,Elias Dahdouh, Fernando Lázaro, Esther Viedma, Natalia Stella, Julio García, Juan Carlos Galán, Rafael Cantón, Mª Dolores Folgueira, Rafael Delgado, Jesús Mingorance
EPI_ISL_428692 hCoV-19/Spain/Madrid_H12_29/2020 Europe / Spain / Madrid 2020-03-12 Hospital Universitario 12 de Octubre Hospital Universitario 12 de Octubre Sara González, Raúl Recio,Elias Dahdouh, Fernando Lázaro, Esther Viedma, Natalia Stella, Julio García, Juan Carlos Galán, Rafael Cantón, Mª Dolores Folgueira, Rafael Delgado, Jesús Mingorance
EPI_ISL_428693 hCoV-19/Spain/Madrid_H12_30/2020 Europe / Spain / Madrid 2020-03-12 Hospital Universitario 12 de Octubre Hospital Universitario 12 de Octubre Sara González, Raúl Recio,Elias Dahdouh, Fernando Lázaro, Esther Viedma, Natalia Stella, Julio García, Juan Carlos Galán, Rafael Cantón, Mª Dolores Folgueira, Rafael Delgado, Jesús Mingorance
EPI_ISL_428694 hCoV-19/Spain/Madrid_H12_31/2020 Europe / Spain / Madrid 2020-03-12 Hospital Universitario 12 de Octubre Hospital Universitario 12 de Octubre Sara González, Raúl Recio,Elias Dahdouh, Fernando Lázaro, Esther Viedma, Natalia Stella, Julio García, Juan Carlos Galán, Rafael Cantón, Mª Dolores Folgueira, Rafael Delgado, Jesús Mingorance
EPI_ISL_428695 hCoV-19/Spain/Madrid_H12_32/2020 Europe / Spain / Madrid 2020-03-12 Hospital Universitario 12 de Octubre Hospital Universitario 12 de Octubre Raúl Recio, Sara González, Elias Dahdouh, Fernando Lázaro, Esther Viedma, Natalia Stella, Julio García, Juan Carlos Galán, Rafael Cantón, Mª Dolores Folgueira, Rafael Delgado, Jesús Mingorance
EPI_ISL_428696 hCoV-19/Spain/Madrid_H12_33/2020 Europe / Spain / Madrid 2020-03-12 Hospital Universitario 12 de Octubre Hospital Universitario 12 de Octubre Raúl Recio, Sara González, Elias Dahdouh, Fernando Lázaro, Esther Viedma, Natalia Stella, Julio García, Juan Carlos Galán, Rafael Cantón, Mª Dolores Folgueira, Rafael Delgado, Jesús Mingorance
EPI_ISL_428697 hCoV-19/Spain/Madrid_H12_34/2020 Europe / Spain / Madrid 2020-03-12 Hospital Universitario 12 de Octubre Hospital Universitario 12 de Octubre Raúl Recio, Sara González, Elias Dahdouh, Fernando Lázaro, Esther Viedma, Natalia Stella, Julio García, Juan Carlos Galán, Rafael Cantón, Mª Dolores Folgueira, Rafael Delgado, Jesús Mingorance
EPI_ISL_428698 hCoV-19/Spain/Madrid_H12_35/2020 Europe / Spain / Madrid 2020-03-12 Hospital Universitario 12 de Octubre Hospital Universitario 12 de Octubre Raúl Recio, Sara González, Elias Dahdouh, Fernando Lázaro, Esther Viedma, Natalia Stella, Julio García, Juan Carlos Galán, Rafael Cantón, Mª Dolores Folgueira, Rafael Delgado, Jesús Mingorance
EPI_ISL_428699 hCoV-19/Spain/Madrid_H12_36/2020 Europe / Spain / Madrid 2020-03-09 Hospital Universitario 12 de Octubre Hospital Universitario 12 de Octubre Raúl Recio, Sara González, Elias Dahdouh, Fernando Lázaro, Esther Viedma, Natalia Stella, Julio García, Juan Carlos Galán, Rafael Cantón, Mª Dolores Folgueira, Rafael Delgado, Jesús Mingorance
EPI_ISL_429968 hCoV-19/France/HF1463/2020 Europe / France / Hauts de France / Compiegne 2020-02-21 Centre Hospitalier Compiègne Laboratoire de Biologie National Reference Center for Viruses of Respiratory Infections, Institut Pasteur, Paris Mélanie Albert, Marion Barbet, Sylvie Behillil, Méline Bizard, Angela Brisebarre, Flora Donati, Fabiana Gambaro, Etienne Simon-Lorière, Vincent Enouf, Maud Vanpeene, Sylvie van der Werf, Raulin Olivia
EPI_ISL_429969 hCoV-19/USA/VA-DCLS-0069/2020 North America / USA / Virginia 2020-03-26 Virginia DCLS Virginia DCLS Virginia DCLS
EPI_ISL_429970 hCoV-19/USA/VA-DCLS-0070/2020 North America / USA / Virginia 2020-03-28 Virginia DCLS

Virginia DCLS	Virginia DCLS
EPI_ISL_429971	hCoV-19/USA/VA-DCLS-0071/2020	North America / USA / Virginia	2020-03-30	Virginia DCLS
Virginia DCLS	Virginia DCLS
EPI_ISL_429972	hCoV-19/USA/VA-DCLS-0072/2020	North America / USA / Virginia	2020-03-28	Virginia DCLS
Virginia DCLS	Virginia DCLS
EPI_ISL_429973	hCoV-19/USA/VA-DCLS-0073/2020	North America / USA / Virginia	2020-03-28	Virginia DCLS
Virginia DCLS	Virginia DCLS
EPI_ISL_429974	hCoV-19/USA/VA-DCLS-0075/2020	North America / USA / Virginia	2020-03-29	Virginia DCLS
Virginia DCLS	Virginia DCLS
EPI_ISL_429975	hCoV-19/USA/VA-DCLS-0076/2020	North America / USA / Virginia	2020-03-27	Virginia DCLS
Virginia DCLS	Virginia DCLS
EPI_ISL_429976	hCoV-19/USA/VA-DCLS-0077/2020	North America / USA / Virginia	2020-04-04	Virginia DCLS
Virginia DCLS	Virginia DCLS
EPI_ISL_429977	hCoV-19/USA/VA-DCLS-0078/2020	North America / USA / Virginia	2020-03-27	Virginia DCLS
Virginia DCLS	Virginia DCLS
EPI_ISL_429978	hCoV-19/USA/VA-DCLS-0079/2020	North America / USA / Virginia	2020-03-30	Virginia DCLS
Virginia DCLS	Virginia DCLS
EPI_ISL_429979	hCoV-19/USA/VA-DCLS-0080/2020	North America / USA / Virginia	2020-03-26	Virginia DCLS
Virginia DCLS	Virginia DCLS
EPI_ISL_429980	hCoV-19/USA/VA-DCLS-0081/2020	North America / USA / Virginia	2020-04-03	Virginia DCLS
Virginia DCLS	Virginia DCLS
EPI_ISL_429981	hCoV-19/USA/VA-DCLS-0083/2020	North America / USA / Virginia	2020-04-03	Virginia DCLS
Virginia DCLS	Virginia DCLS
EPI_ISL_429982	hCoV-19/USA/VA-DCLS-0084/2020	North America / USA / Virginia	2020-04-06	Virginia DCLS
Virginia DCLS	Virginia DCLS
EPI_ISL_429983	hCoV-19/USA/VA-DCLS-0086/2020	North America / USA / Virginia	2020-03-30	Virginia DCLS
Virginia DCLS	Virginia DCLS
EPI_ISL_429984	hCoV-19/USA/VA-DCLS-0087/2020	North America / USA / Virginia	2020-04-02	Virginia DCLS
Virginia DCLS	Virginia DCLS
EPI_ISL_429985	hCoV-19/USA/VA-DCLS-0088/2020	North America / USA / Virginia	2020-04-02	Virginia DCLS
Virginia DCLS	Virginia DCLS
EPI_ISL_429986	hCoV-19/USA/VA-DCLS-0089/2020	North America / USA / Virginia	2020-04-02	Virginia DCLS
Virginia DCLS	Virginia DCLS
EPI_ISL_429987	hCoV-19/USA/VA-DCLS-0090/2020	North America / USA / Virginia	2020-04-02	Virginia DCLS
Virginia DCLS	Virginia DCLS
EPI_ISL_429988	hCoV-19/USA/VA-DCLS-0091/2020	North America / USA / Virginia	2020-04-02	Virginia DCLS
Virginia DCLS	Virginia DCLS
EPI_ISL_429989	hCoV-19/USA/VA-DCLS-0092/2020	North America / USA / Virginia	2020-04-01	Virginia DCLS
Virginia DCLS	Virginia DCLS
EPI_ISL_429990	hCoV-19/USA/CA-SR007/2020	North America / USA / California / San Diego	2020-03-21
Rady's Childrens Hospital	Andersen lab at Scripps Research	SEARCH Alliance San Diego with
Christina Clarke, Michelle Vanderpool, Teresa Mueller, Denise Malicki
EPI_ISL_429991	hCoV-19/USA/CA-SR017/2020	North America / USA / California / San Diego	2020-03-24
Andersen lab at Scripps Research	Andersen lab at Scripps Research	SEARCH Alliance San Diego
EPI_ISL_429992	hCoV-19/Jordan/SR-032/2020	Asia / Jordan / Amman	2020-03-22	Biolab Diagnostic
Laboratories	Andersen lab at Scripps Research	Issa Abu-Dayyeh, Ahmad Tibi, Lama Hussein, Lina
Mohammad, Zein Naber, Amid Abdelnour with SEARCH Alliance San Diego
EPI_ISL_429993	hCoV-19/Jordan/SR-033/2020	Asia / Jordan / Amman	2020-03-16	Biolab Diagnostic
Laboratories	Andersen lab at Scripps Research	Issa Abu-Dayyeh, Ahmad Tibi, Lama Hussein, Lina
Mohammad, Zein Naber, Amid Abdelnour with SEARCH Alliance San Diego
EPI_ISL_429994	hCoV-19/Jordan/SR-034/2020	Asia / Jordan / Amman	2020-03-17	Biolab Diagnostic
Laboratories	Andersen lab at Scripps Research	Issa Abu-Dayyeh, Ahmad Tibi, Lama Hussein, Lina
Mohammad, Zein Naber, Amid Abdelnour with SEARCH Alliance San Diego
EPI_ISL_429995	hCoV-19/Jordan/SR-035/2020	Asia / Jordan / Amman	2020-03-30	Biolab Diagnostic
Laboratories	Andersen lab at Scripps Research	Issa Abu-Dayyeh, Ahmad Tibi, Lama Hussein, Lina
Mohammad, Zein Naber, Amid Abdelnour with SEARCH Alliance San Diego
EPI_ISL_429996	hCoV-19/Jordan/SR-036/2020	Asia / Jordan / Amman	2020-03-23	Biolab Diagnostic
Laboratories	Andersen lab at Scripps Research	Issa Abu-Dayyeh, Ahmad Tibi, Lama Hussein, Lina
Mohammad, Zein Naber, Amid Abdelnour with SEARCH Alliance San Diego
EPI_ISL_429997	hCoV-19/Jordan/SR-037/2020	Asia / Jordan / Amman	2020-03-16	Biolab Diagnostic
Laboratories	Andersen lab at Scripps Research	Issa Abu-Dayyeh, Ahmad Tibi, Lama Hussein, Lina
Mohammad, Zein Naber, Amid Abdelnour with SEARCH Alliance San Diego
EPI_ISL_429998	hCoV-19/Jordan/SR-039/2020	Asia / Jordan / Amman	2020-03-28	Biolab Diagnostic
Laboratories	Andersen lab at Scripps Research	Issa Abu-Dayyeh, Ahmad Tibi, Lama Hussein, Lina
Mohammad, Zein Naber, Amid Abdelnour with SEARCH Alliance San Diego
EPI_ISL_429999	hCoV-19/Jordan/SR-041/2020	Asia / Jordan / Amman	2020-03-17	Biolab Diagnostic
Laboratories	Andersen lab at Scripps Research	Issa Abu-Dayyeh, Ahmad Tibi, Lama Hussein, Lina
Mohammad, Zein Naber, Amid Abdelnour with SEARCH Alliance San Diego
EPI_ISL_402119	hCoV-19/Wuhan/IVDC-HB-01/2019	Asia / China / Hubei / Wuhan	2019-12-30	National
Institute for Viral Disease Control and Prevention, China CDC	National Institute for Viral Disease Control

and Prevention, China CDC	Wenjie Tan, Xiang Zhao, Wenling Wang, Xuejun Ma, Yongzhong Jiang, Roujian Lu, Ji Wang, Weimin Zhou, Peihua Niu, Peipei Liu, Faxian Zhan, Weifeng Shi, Baoying Huang, Jun Liu, Li Zhao, Yao Meng, Xiaozhou He, Fei Ye, Na Zhu, Yang Li, Jing Chen, Wenbo Xu, George F. Gao, Guizhen Wu
EPI_ISL_402120	hCoV-19/Wuhan/IVDC-HB-04/2020	Asia / China / Hubei / Wuhan	2020-01-01	National Institute for Viral Disease Control and Prevention, China CDC	National Institute for Viral Disease Control and Prevention, China CDC	Wenjie Tan, Xiang Zhao, Wenling Wang, Xuejun Ma, Yongzhong Jiang, Roujian Lu, Ji Wang, Weimin Zhou, Peihua Niu, Peipei Liu, Faxian Zhan, Weifeng Shi, Baoying Huang, Jun Liu, Li Zhao, Yao Meng, Xiaozhou He, Fei Ye, Na Zhu, Yang Li, Jing Chen, Wenbo Xu, George F. Gao, Guizhen Wu
EPI_ISL_402121	hCoV-19/Wuhan/IVDC-HB-05/2019	Asia / China / Hubei / Wuhan	2019-12-30	National Institute for Viral Disease Control and Prevention, China CDC	National Institute for Viral Disease Control and Prevention, China CDC	Wenjie Tan, Xuejun Ma, Xiang Zhao, Wenling Wang, Yongzhong Jiang, Roujian Lu, Ji Wang, Peihua Niu, Weimin Zhou, Faxian Zhan, Weifeng Shi, Baoying Huang, Jun Liu, Li Zhao, Yao Meng, Fei Ye, Na Zhu, Xiaozhou He, Peipei Liu, Yang Li, Jing Chen, Wenbo Xu, George F. Gao, Guizhen Wu
EPI_ISL_402123	hCoV-19/Wuhan/IPBCAMS-WH-01/2019	Asia / China / Hubei / Wuhan	2019-12-24	Institute of Pathogen Biology, Chinese Academy of Medical Sciences & Peking Union Medical College	Institute of Pathogen Biology, Chinese Academy of Medical Sciences & Peking Union Medical College	Lili Ren, Jianwei Wang, Qi Jin, Zichun Xiang, Zhiqiang Wu, Chao Wu, Yiwei Liu
EPI_ISL_402124	hCoV-19/Wuhan/WIV04/2019	Asia / China / Hubei / Wuhan	2019-12-30	Wuhan Jinyintan Hospital	Wuhan Institute of Virology, Chinese Academy of Sciences	Peng Zhou, Xing-Lou Yang, Ding-Yu Zhang, Lei Zhang, Yan Zhu, Hao-Rui Si, Zhengli Shi
EPI_ISL_402127	hCoV-19/Wuhan/WIV02/2019	Asia / China / Hubei / Wuhan	2019-12-30	Wuhan Jinyintan Hospital	Wuhan Institute of Virology, Chinese Academy of Sciences	Peng Zhou, Xing-Lou Yang, Ding-Yu Zhang, Lei Zhang, Yan Zhu, Hao-Rui Si, Zhengli Shi
EPI_ISL_402128	hCoV-19/Wuhan/WIV05/2019	Asia / China / Hubei / Wuhan	2019-12-30	Wuhan Jinyintan Hospital	Wuhan Institute of Virology, Chinese Academy of Sciences	Peng Zhou, Xing-Lou Yang, Ding-Yu Zhang, Lei Zhang, Yan Zhu, Hao-Rui Si, Zhengli Shi
EPI_ISL_402129	hCoV-19/Wuhan/WIV06/2019	Asia / China / Hubei / Wuhan	2019-12-30	Wuhan Jinyintan Hospital	Wuhan Institute of Virology, Chinese Academy of Sciences	Peng Zhou, Xing-Lou Yang, Ding-Yu Zhang, Lei Zhang, Yan Zhu, Hao-Rui Si, Zhengli Shi
EPI_ISL_402130	hCoV-19/Wuhan/WIV07/2019	Asia / China / Hubei / Wuhan	2019-12-30	Wuhan Jinyintan Hospital	Wuhan Institute of Virology, Chinese Academy of Sciences	Peng Zhou, Xing-Lou Yang, Ding-Yu Zhang, Lei Zhang, Yan Zhu, Hao-Rui Si, Zhengli Shi
EPI_ISL_402132	hCoV-19/Wuhan/HBCDC-HB-01/2019	Asia / China / Hubei / Wuhan	2019-12-30	Wuhan Jinyintan Hospital	Hubei Provincial Center for Disease Control and Prevention	Bin Fang, Xiang Li, Xiao Yu, Linlin Liu, Bo Yang, Faxian Zhan, Guojun Ye, Xixiang Huo, Junqiang Xu, Bo Yu, Kun Cai, Jing Li, Yongzhong Jiang.
EPI_ISL_408008	hCoV-19/USA/CA3/2020	North America / USA / California	2020-01-29	California Department of Health	Pathogen Discovery, Respiratory Viruses Branch, Division of Viral Diseases, Centers for Disease Control and Prevention	Krista Queen, Jing Zhang, Yan Li, Ying Tao, Anna Uehara, Clinton Paden, Xiaoyan Lu, Brian Lynch, Senthil Kumar K. Sakthivel, Brett L. Whitaker, Shifaq Kamili, Lijuan Wang, Janna' R. Murray, Susan I. Gerber, Stephen Lindstrom, Suxiang Tong
EPI_ISL_408009	hCoV-19/USA/CA4/2020	North America / USA / California	2020-01-29	California Department of Health	Pathogen Discovery, Respiratory Viruses Branch, Division of Viral Diseases, Centers for Dieases Control and Prevention	Krista Queen, Jing Zhang, Yan Li, Ying Tao, Anna Uehara, Clinton Paden, Xiaoyan Lu, Brian Lynch, Senthil Kumar K. Sakthivel, Brett L. Whitaker, Shifaq Kamili, Lijuan Wang, Janna' R. Murray, Susan I. Gerber, Stephen Lindstrom, Suxiang Tong
EPI_ISL_408010	hCoV-19/USA/CA5/2020	North America / USA / California	2020-01-29	California Department of Health	Pathogen Discovery, Respiratory Viruses Branch, Division of Viral Diseases, Centers for Dieases Control and Prevention	Ying Tao, Krista Queen, Jing Zhang, Yan Li, Anna Uehara, Clinton Paden, Xiaoyan Lu, Brian Lynch, Senthil Kumar K. Sakthivel, Brett L. Whitaker, Shifaq Kamili, Lijuan Wang, Janna' R. Murray, Susan I. Gerber, Stephen Lindstrom, Suxiang Tong
EPI_ISL_410301	hCoV-19/Nepal/61/2020	Asia / Nepal / Kathmandu	2020-01-13	National Influenza Centre, National Public Health Laboratory, Kathmandu, Nepal	The University of Hong Kong	Ranjit Sah , Runa Jha, Daniel Chu, Haogao Gu, Malik Peiris, Anup Bastola, Alfonso J. Rodriguez-Morales, Bibek Kumar Lal, Basu Dev Pandey, Leo Poon
EPI_ISL_411902	hCoV-19/Cambodia/0012/2020	Asia / Cambodia / Sihanoukville 2020-01-27	Virology Unit, Institut Pasteur du Cambodge.	Virology Unit, Institut Pasteur du Cambodge (Sequencing done by: Jessica E Manning/Jennifer A Bohl at Malaria and Vector Research Research Laboratory, National Institute of Allergy and Infectious Diseases and Vida Ahyong from Chan-Zuckerberg Biohub)	Erik A Karlsson, Jennifer A Bohl, Vida Ahyong, Veasna Duong, Philippe Dussart, Jessica E Manning.
EPI_ISL_411915	hCoV-19/Taiwan/CGMH-CGU-01/2020 Asia / Taiwan / Taoyuan 2020-01-25	Laboratory Medicine Department of Laboratory Medicine, Lin-Kou Chang Gung Memorial Hospital, Taoyuan, Taiwan.	Kuo-Chien Tsao, Yu-Nong Gong, Shu-Li Yang, Yi-Chun Li, Chung-Guei Huang, Yhu-Chering Huang, Shin-Ru Shih
EPI_ISL_411926	hCoV-19/Taiwan/3/2020	Asia / Taiwan / Taipei	2020-01-24	Taiwan Centers for Disease Control Taiwan Centers for Disease Control	Ji-Rong Yang, Yu-Chi-Lin, Jung-Jung Mu, Ming-Tsan-Liu
EPI_ISL_411927	hCoV-19/Taiwan/4/2020	Asia / Taiwan / Taipei	2020-01-28	Taiwan Centers for Disease Control Taiwan Centers for Disease Control	Ji-Rong Yang, Yu-Chi-Lin, Jung-Jung Mu, Ming-Tsan-Liu
EPI_ISL_411929	hCoV-19/South Korea/SNU01/2020	Asia / South Korea	2020-01 unknown Department of Clinical Diagnostics	Park,W.B., Kwon,N.-J., Choi,S.-J., Kang,C.K., Choe,P.G., Kim,J.Y., Yun,J., Lee,G.-W., Seong,M.-

W., Kim,N., Seo,J.-S. and Oh,M.-D.
EPI_ISL_412912 hCoV-19/Germany/Baden-Wuerttemberg-1/2020 Europe / Germany / Baden-Wuerttemberg 2020-02-25 State Health Office Baden-Wuerttemberg Charité Universitätsmedizin Berlin, Institute of Virology Victor M Corman, Julia Schneider, Barbara Mühlemann, Talitha Veith, Jörn Beheim-Schwarzbach, Terry Jones, Rainer Oehme, Silke Fischer, Christian Drosten
EPI_ISL_412964 hCoV-19/Brazil/SPBR-01/2020 South America / Brazil / Sao Paulo / Sao Paulo 2020-02-25 Hospital Israelita Albert Einstein Instituto Adolfo Lutz Interdisciplinary Procedures Center Strategic Laboratory Jaqueline Goes de Jesus, Claudio Tavares Sacchi, Daniela Bernardes Borges da Silva, Ingra Morales Claro, Flávia Cristina da Silva Sales, Claudia Regina Gonçalves, Joshua Quick, Maria do Carmo, Sampaio Tavares Timenetsky, Nicholas James Loman, Andrew Rambaut, Ester Cerdeira Sabino, Nuno Rodrigues Faria
EPI_ISL_412966 hCoV-19/China/IQTC01/2020 Asia / China / Guangdong / Guangzhou 2020-02-05 unknown Technology Centre, Guangzhou Customs Shi,Y., Sun,J., Zheng,K., Huang,J. and Zhao,J.
EPI_ISL_412967 hCoV-19/China/IQTC02/2020 Asia / China / Guangzhou 2020-01-29 unknown Technology Centre, Guangzhou Customs Shi,Y., Zheng,K., Sun,J., Huang,J., Zhu,A., Zhuang,Z., Dai,J., Chen,Z., Sun,F., Zhang,Z., Li,X. and Wang,Y.
EPI_ISL_412968 hCoV-19/Japan/Hu_DP_Kng_19-020/2020 Asia / Japan 2020-02-10 unknown Takayuki Hishiki Kanagawa Prefectural Institute of Public Health, Department of Microbiology Hishiki,T., Suzuki,R., Sakuragi,J., Usui,K., Tanaka,Y., Kawai,J., Kogo,Y., Matsuki,Y., An,T., Hayashizaki,Y. and Takasaki,T.
EPI_ISL_412969 hCoV-19/Japan/Hu_DP_Kng_19-027/2020 Asia / Japan 2020-02-10 unknown Takayuki Hishiki Kanagawa Prefectural Institute of Public Health, Department of Microbiology Hishiki,T., Suzuki,R., Sakuragi,J., Usui,K., Tanaka,Y., Kawai,J., Kogo,Y., Matsuki,Y., An,T., Hayashizaki,Y. and Takasaki,T.
EPI_ISL_412970 hCoV-19/USA/WA2/2020 North America / USA / Washington / Snohomish County 2020-02-24 Washington State Department of Health Seattle Flu Study Helen Chu, Michael Boeckh, Janet Englund, Michael Famulare, Barry Lutz, Deborah Nickerson, Mark Rieder, Lea Starita, Matthew Thompson, Jay Shendure, and Trevor Bedford
EPI_ISL_412972 hCoV-19/Mexico/CDMX-InDRE_01/2020 North America / Mexico / Mexico City 2020-02-27 Instituto Nacional de Enfermedades Respiratorias Instituto de Diagnostico y Referencia Epidemiologicos (INDRE) Ramirez-Gonzalez Ernesto, Garces-Ayala Fabiola, Araiza-Rodriguez Adnan, Mendieta-Condado Edgar, Rodriguez-Maldonado Abril, Wong-Arambula Claudia, Vazquez-Perez Joel, Martinez Arturo, Boukadida Celia, Munoz-Medina Esteban, Sanchez Alejandro, Isa Pavel, Taboada Blanca, Lopez Susana, Arias Carlos, Barrera-Badillo Gisela, Hernandez-Rivas Lucia, Lopez-Martinez Irma
EPI_ISL_412973 hCoV-19/Italy/CDG1/2020 Europe / Italy / Lombardy 2020-02-20 Department of Infectious Diseases, Istituto Superiore di Sanità, Roma , Italy Virology Laboratory, Scientific Department, Army Medical Center Paola Stefanelli, Stefano Fiore, Antonella Marchi, Eleonora Benedetti, Concetta Fabiani, Giovanni Faggioni, Antonella Fortunato, Riccardo De Santis, Silvia Fillo, Anna Anselmo, Andrea Ciammaruconi, Stefano Palomba, Florigio Lista
EPI_ISL_412974 hCoV-19/Italy/SPL1/2020 Europe / Italy / Rome 2020-01-29 Department of Infectious Diseases, Istituto Superiore di Sanità, Rome, Italy Virology Laboratory, Scientific Department, Army Medical Center Paola Stefanelli, Stefano Fiore, Antonella Marchi, Eleonora Benedetti, Concetta Fabiani, Giovanni Faggioni, Antonella Fortunato, Silvia Fillo, Riccardo De Santis, Andrea Ciammaruconi, Giancarlo Petralito, Filippo Molinari, Florigio Lista
EPI_ISL_412975 hCoV-19/Australia/NSW05/2020 Oceania / Australia / New South Wales / Sydney 2020-02-28 Centre for Infectious Diseases and Microbiology Laboratory Services NSW Health Pathology - Institute of Clinical Pathology and Medical Research; Westmead Hospital; University of Sydney Eden J-S, Carter I, Rahman H, Holmes EC, Rockett R, O’Sullivan MV, Sintchenko V, Chen SC, Maddocks S, Kok J and Dwyer DE for the 2019-nCoV Study Group
EPI_ISL_412978 hCoV-19/Wuhan/HBCDC-HB-02/2020 Asia / China / Hubei / Wuhan 2020-01-17 The Central Hospital Of Wuhan Hubei Provincial Center for Disease Control and Prevention Bin Fang, Xiang Li, Xiao Yu, Linlin Liu, Bo Yang, Faxian Zhan, Guojun Ye, Xixiang Huo, Junqiang Xu, Bo Yu, Kun Cai, Jing Li, Yongzhong Jiang.
EPI_ISL_412979 hCoV-19/Wuhan/HBCDC-HB-03/2020 Asia / China / Hubei / Wuhan 2020-01-18 Union Hospital of Tongji Medical College, Huazhong University of Science and Technology Hubei Provincial Center for Disease Control and Prevention Bin Fang, Xiang Li, Xiao Yu, Linlin Liu, Bo Yang, Faxian Zhan, Guojun Ye, Xixiang Huo, Junqiang Xu, Bo Yu, Kun Cai, Jing Li, Yongzhong Jiang.
EPI_ISL_412980 hCoV-19/Wuhan/HBCDC-HB-04/2020 Asia / China / Hubei / Wuhan 2020-01-18 Union Hospital of Tongji Medical College, Huazhong University of Science and Technology Hubei Provincial Center for Disease Control and Prevention Bin Fang, Xiang Li, Xiao Yu, Linlin Liu, Bo Yang, Faxian Zhan, Guojun Ye, Xixiang Huo, Junqiang Xu, Bo Yu, Kun Cai, Jing Li, Yongzhong Jiang.
EPI_ISL_412981 hCoV-19/Wuhan/HBCDC-HB-05/2020 Asia / China / Hubei / Wuhan 2020-01-18 CR&WISCO GENERAL HOSPITAL Hubei Provincial Center for Disease Control and Prevention Bin Fang, Xiang Li, Xiao Yu, Linlin Liu, Bo Yang, Faxian Zhan, Guojun Ye, Xixiang Huo, Junqiang Xu, Bo Yu, Kun Cai, Jing Li, Yongzhong Jiang.
EPI_ISL_412982 hCoV-19/Wuhan/HBCDC-HB-06/2020 Asia / China / Hubei / Wuhan 2020-02-07 Wuhan Lung Hospital Hubei Provincial Center for Disease Control and Prevention Bin Fang, Xiang Li, Xiao Yu, Linlin Liu, Bo Yang, Faxian Zhan, Guojun Ye, Xixiang Huo, Junqiang Xu, Bo Yu, Kun Cai, Jing Li, Yongzhong Jiang.
EPI_ISL_412983 hCoV-19/Tianmen/HBCDC-HB-07/2020 Asia / China / Hubei / Tianmen 2020-02-08 Tianmen Center for Disease Control and Prevention Hubei Provincial Center for Disease Control and Prevention Bin Fang, Xiang Li, Xiao Yu, Linlin Liu, Bo Yang, Faxian Zhan, Guojun Ye, Xixiang Huo, Junqiang Xu, Bo Yu, Kun Cai, Jing Li, YiFa Zhu, Yangyang Tao,Xierong Li,Yongzhong Jiang.

EPI_ISL_415105 hCoV-19/Brazil/BA-312/2020 South America / Brazil / Bahia / Feira de Santana 2020-03-04 Laboratório Central de Saúde Pública Professor Gonçalo Moniz – LACEN/BA Instituto Oswaldo Cruz FIOCRUZ - Laboratory of Respiratory Viruses and Measles (LVRS) Paola Resende, Allison Fabri, Joilson Xavier, Sunando Roy, Fernando Motta, Aline Mattos, Milene Miranda, Cristiana Garcia, Braulia Caetano, Maria Ogrzewalska, Jonathan Lopes, Luciana Appolinario, Maria Nóbrega, Marilda Siqueira
EPI_ISL_415128 hCoV-19/Brazil/ES-225/2020 South America / Brazil / Espirito Santo / Vila Velha 2020-02-29 LACEN/ES - Laboratório Central de Saúde Pública do Espírito Santo Instituto Oswaldo Cruz FIOCRUZ - Laboratory of Respiratory Viruses and Measles (LVRS) Paola Resende, Allison Fabri, Joilson Xavier, Sunando Roy, Fernando Motta, Aline Mattos, Milene Miranda, Cristiana Garcia, Braulia Caetano, Maria Ogrzewalska, Jonathan Lopes, Luciana Appolinario, Maria Nóbrega, Marilda Siqueira
EPI_ISL_415129 hCoV-19/England/20099038206/2020 Europe / United Kingdom / England 2020-02-29 Respiratory Virus Unit, Microbiology Services Colindale, Public Health England Respiratory Virus Unit, Microbiology Services Colindale, Public Health England Monica Galiano, Shahjahan Miah, Angie Lackenby, Omolola Akinbami, Tiina Talts, Leena Bhaw, Richard Myers, Steven Platt, Kirstin Edwards, Jonathan Hubb, Joanna Ellis, Maria Zambon
EPI_ISL_415134 hCoV-19/England/20100004806/2020 Europe / United Kingdom / England 2020-02-29 Respiratory Virus Unit, Microbiology Services Colindale, Public Health England Respiratory Virus Unit, Microbiology Services Colindale, Public Health England Monica Galiano, Shahjahan Miah, Angie Lackenby, Omolola Akinbami, Tiina Talts, Leena Bhaw, Richard Myers, Steven Platt, Kirstin Edwards, Jonathan Hubb, Joanna Ellis, Maria Zambon
EPI_ISL_415136 hCoV-19/England/20100022706/2020 Europe / United Kingdom / England 2020-02-29 Respiratory Virus Unit, Microbiology Services Colindale, Public Health England Respiratory Virus Unit, Microbiology Services Colindale, Public Health England Monica Galiano, Shahjahan Miah, Angie Lackenby, Omolola Akinbami, Tiina Talts, Leena Bhaw, Richard Myers, Steven Platt, Kirstin Edwards, Jonathan Hubb, Joanna Ellis, Maria Zambon
EPI_ISL_415141 hCoV-19/England/20100121007/2020 Europe / United Kingdom / England 2020-02-29 Respiratory Virus Unit, Microbiology Services Colindale, Public Health England Respiratory Virus Unit, Microbiology Services Colindale, Public Health England Monica Galiano, Shahjahan Miah, Angie Lackenby, Omolola Akinbami, Tiina Talts, Leena Bhaw, Richard Myers, Steven Platt, Kirstin Edwards, Jonathan Hubb, Joanna Ellis, Maria Zambon
EPI_ISL_415142 hCoV-19/England/20100122106/2020 Europe / United Kingdom / England 2020-03-02 Respiratory Virus Unit, Microbiology Services Colindale, Public Health England Respiratory Virus Unit, Microbiology Services Colindale, Public Health England Monica Galiano, Shahjahan Miah, Angie Lackenby, Omolola Akinbami, Tiina Talts, Leena Bhaw, Richard Myers, Steven Platt, Kirstin Edwards, Jonathan Hubb, Joanna Ellis, Maria Zambon
EPI_ISL_415147 hCoV-19/England/20102000506/2020 Europe / United Kingdom / England 2020-03-01 Respiratory Virus Unit, Microbiology Services Colindale, Public Health England Respiratory Virus Unit, Microbiology Services Colindale, Public Health England Monica Galiano, Shahjahan Miah, Angie Lackenby, Omolola Akinbami, Tiina Talts, Leena Bhaw, Richard Myers, Steven Platt, Kirstin Edwards, Jonathan Hubb, Joanna Ellis, Maria Zambon
EPI_ISL_415148 hCoV-19/England/20102000906/2020 Europe / United Kingdom / England 2020-03-03 Respiratory Virus Unit, Microbiology Services Colindale, Public Health England Respiratory Virus Unit, Microbiology Services Colindale, Public Health England Monica Galiano, Shahjahan Miah, Angie Lackenby, Omolola Akinbami, Tiina Talts, Leena Bhaw, Richard Myers, Steven Platt, Kirstin Edwards, Jonathan Hubb, Joanna Ellis, Maria Zambon
EPI_ISL_415150 hCoV-19/England/20110003506/2020 Europe / United Kingdom / England 2020-03-09 Respiratory Virus Unit, Microbiology Services Colindale, Public Health England Respiratory Virus Unit, Microbiology Services Colindale, Public Health England Monica Galiano, Shahjahan Miah, Angie Lackenby, Omolola Akinbami, Tiina Talts, Leena Bhaw, Richard Myers, Steven Platt, Kirstin Edwards, Jonathan Hubb, Joanna Ellis, Maria Zambon
EPI_ISL_415151 hCoV-19/USA/NY2-PV08100/2020 North America / USA / New York 2020-03-04 MSHS Clinical Microbiology Laboratories MSHS Pathogen Surveillance Program Gopi Patel, Emilia Sordillo, Melissa Gitman, Alberto Paniz-mondolfi, Matthew Hernandez, Shelcie Fabre, Jose Polanco, Ana Silvia Gonzalez-Reiche, Zenab Khan, Nancy Francoeur, Melissa Smith, Robert Sebra, Lisa Miorin, Wen-chun Liu, Randy Albrecht, Judith Aberg, Florian Krammer, Adolfo Garcia-Sarstre, Viviana Simon, Harm van Bakel
EPI_ISL_415152 hCoV-19/Panama/328677/2020 Central America / Panama / Panama City 2020-03-06 Gorgas Memorial Institute for Health Studies Gorgas Memorial Institute for Health Studies Danilo Franco, Sandra Lopez-Verges, Elimelec Valdespino, Claudia Gonzalez, Oris Chavarria, Ambar Moreno, Yamilka Diaz, Leyda Abrego, Juan M. Pascale, Alexander A. Martinez.
EPI_ISL_415153 hCoV-19/Belgium/VLM-03011/2020 Europe / Belgium / Huldenberg 2020-03-03 KU Leuven, Clinical and Epidemiological Virology KU Leuven, Clinical and Epidemiological Virology Bert Vanmechelen, Joan Marti-Carreras, Tony Wawina, Marc Van Ranst, Piet Maes
EPI_ISL_415154 hCoV-19/Belgium/BM-03012/2020 Europe / Belgium / Kraainem 2020-03-01 KU Leuven, Clincal and Epidemiological Virology KU Leuven, Clincal and Epidemiological Virology Bert Vanmechelen, Joan Marti-Careras, Tony Wawina, Marc Van Ranst, Piet Maes.
EPI_ISL_415155 hCoV-19/Belgium/VAG-03013/2020 Europe / Belgium / Huldenberg 2020-03-01 KU Leuven, Clinical and Epidemiological Virology KU Leuven, Clinical and Epidemiological Virology Bert Vanmechelen, Joan Marti-Carreras, Tony Wawina, Marc Van Ranst, Piet Maes
EPI_ISL_415156 hCoV-19/Belgium/SH-03014/2020 Europe / Belgium / Huldenberg 2020-03-01 KU Leuven, Clinical and Epidemiological Virology KU Leuven, Clinical and Epidemiological Virology Bert

Vanmechelen, Joan Marti-Carreras, Tony Wawina, Piet Maes
EPI_ISL_415157 hCoV-19/Belgium/BC-03016/2020 Europe / Belgium / Sint-Niklaas 2020-03-01 KU Leuven, Clinical and Epidemiological Virology KU Leuven, Clinical and Epidemiological Virology Bert Vanmechelen, Joan Marti-Carreras, Tony Wawina, Piet Maes
EPI_ISL_415158 hCoV-19/Belgium/QKJ-03015/2020 Europe / Belgium / Brussels 2020-03-01 KU Leuven, Clinical and Epidemiological Virology KU Leuven, Clinical and Epidemiological Virology Bert Vanmechelen, Joan Marti-Carreras, Tony Wawina, Piet Maes
EPI_ISL_415159 hCoV-19/Belgium/BA-02291/2020 Europe / Belgium / Leuven 2020-02-29 KU Leuven, Clinical and Epidemiological Virology KU Leuven, Clinical and Epidemiological Virology Bert Vanmechelen, Joan Marti-Carreras, Tony Wawina, Piet Maes
EPI_ISL_415435 hCoV-19/Wales/PHW06/2020 Europe / United Kingdom / Wales 2020-03-06 Wales Specialist Virology Centre Public Health Wales Microbiology Cardiff Catherine Moore, Joanne Watkins, Sally Corden, Tom Connor
EPI_ISL_415454 hCoV-19/Switzerland/GE1422/2020 Europe / Switzerland 2020-02-28 Hôpitaux universitaires de Genève Laboratoire de Virologie Hôpitaux universitaires de Genève Laboratoire de Virologie Laubscher F.
EPI_ISL_415455 hCoV-19/Switzerland/GE0199/2020 Europe / Switzerland 2020-02-28 Hôpitaux universitaires de Genève Laboratoire de Virologie Hôpitaux universitaires de Genève Laboratoire de Virologie Laubscher F.
EPI_ISL_415456 hCoV-19/Switzerland/BE6651/2020 Europe / Switzerland 2020-02-29 Hôpitaux universitaires de Genève Laboratoire de Virologie Hôpitaux universitaires de Genève Laboratoire de Virologie Laubscher F.
EPI_ISL_415457 hCoV-19/Switzerland/AG7120/2020 Europe / Switzerland 2020-02-29 Hôpitaux universitaires de Genève Laboratoire de Virologie Hôpitaux universitaires de Genève Laboratoire de Virologie Laubscher F.
EPI_ISL_415458 hCoV-19/Switzerland/GE8102/2020 Europe / Switzerland 2020-03-01 Hôpitaux universitaires de Genève Laboratoire de Virologie Hôpitaux universitaires de Genève Laboratoire de Virologie Laubscher F.
EPI_ISL_415459 hCoV-19/Switzerland/VD0503/2020 Europe / Switzerland / Genève 2020-02-29 Hôpitaux universitaires de Genève Laboratoire de Virologie Hôpitaux universitaires de Genève Laboratoire de Virologie Laubscher F.
EPI_ISL_415460 hCoV-19/Netherlands/Flevoland_1/2020 Europe / Netherlands / Flevoland 2020-03-09 Dutch COVID-19 response team Erasmus Medical Center David Nieuwenhuijse, Bas Oude Munnink, Reina Sikkema, Claudia Schapendonk, Irina Chestakova, Anne van der Linden, Mark Pronk, Pascal Lexmond, Corien Swaan, Manon Haverkate, Madelief Mollers, Mart Stein, Sandra Kengne Kamga Mobou, Jeroen van Kampen, Jolanda Voermans, Aura Timen, Corine GeurtsvanKessel, Annemiek van der Eijk, Richard Molenkamp, Marion Koopmans, on behalf of the Dutch national COVID-19 response team.
EPI_ISL_415461 hCoV-19/Netherlands/Gelderland_1-2/2020 Europe / Netherlands / Gelderland 2020-03-10 Dutch COVID-19 response team Erasmus Medical Center David Nieuwenhuijse, Bas Oude Munnink, Reina Sikkema, Claudia Schapendonk, Irina Chestakova, Anne van der Linden, Mark Pronk, Pascal Lexmond, Corien Swaan, Manon Haverkate, Madelief Mollers, Mart Stein, Sandra Kengne Kamga Mobou, Jeroen van Kampen, Jolanda Voermans, Aura Timen, Corine GeurtsvanKessel, Annemiek van der Eijk, Richard Molenkamp, Marion Koopmans, on behalf of the Dutch national COVID-19 response team.
EPI_ISL_415462 hCoV-19/Netherlands/Gelderland_2/2020 Europe / Netherlands / Gelderland 2020-03-09 Dutch COVID-19 response team Erasmus Medical Center David Nieuwenhuijse, Bas Oude Munnink, Reina Sikkema, Claudia Schapendonk, Irina Chestakova, Anne van der Linden, Mark Pronk, Pascal Lexmond, Corien Swaan, Manon Haverkate, Madelief Mollers, Mart Stein, Sandra Kengne Kamga Mobou, Jeroen van Kampen, Jolanda Voermans, Aura Timen, Corine GeurtsvanKessel, Annemiek van der Eijk, Richard Molenkamp, Marion Koopmans, on behalf of the Dutch national COVID-19 response team.
EPI_ISL_415463 hCoV-19/Netherlands/Gelderland_3/2020 Europe / Netherlands / Gelderland 2020-03-09 Dutch COVID-19 response team Erasmus Medical Center David Nieuwenhuijse, Bas Oude Munnink, Reina Sikkema, Claudia Schapendonk, Irina Chestakova, Anne van der Linden, Mark Pronk, Pascal Lexmond, Corien Swaan, Manon Haverkate, Madelief Mollers, Mart Stein, Sandra Kengne Kamga Mobou, Jeroen van Kampen, Jolanda Voermans, Aura Timen, Corine GeurtsvanKessel, Annemiek van der Eijk, Richard Molenkamp, Marion Koopmans, on behalf of the Dutch national COVID-19 response team.
EPI_ISL_415464 hCoV-19/Netherlands/Limburg_7/2020 Europe / Netherlands / Limburg 2020 Dutch COVID-19 response team Erasmus Medical Center David Nieuwenhuijse, Bas Oude Munnink, Reina Sikkema, Claudia Schapendonk, Irina Chestakova, Anne van der Linden, Mark Pronk, Pascal Lexmond, Corien Swaan, Manon Haverkate, Madelief Mollers, Mart Stein, Sandra Kengne Kamga Mobou, Jeroen van Kampen, Jolanda Voermans, Aura Timen, Corine GeurtsvanKessel, Annemiek van der Eijk, Richard Molenkamp, Marion Koopmans, on behalf of the Dutch national COVID-19 response team.
EPI_ISL_415465 hCoV-19/Netherlands/NA_1/2020 Europe / Netherlands 2020-03-10 Dutch COVID-19 response team Erasmus Medical Center David Nieuwenhuijse, Bas Oude Munnink, Reina Sikkema, Claudia Schapendonk, Irina Chestakova, Anne van der Linden, Mark Pronk, Pascal Lexmond, Corien Swaan, Manon Haverkate, Madelief Mollers, Mart Stein, Sandra Kengne Kamga Mobou, Jeroen van Kampen, Jolanda Voermans, Aura Timen, Corine GeurtsvanKessel, Annemiek van der Eijk, Richard Molenkamp, Marion Koopmans, on behalf of the Dutch national COVID-19 response team.
EPI_ISL_415466 hCoV-19/Netherlands/NA_10/2020 Europe / Netherlands 2020-03-09 Dutch COVID-19 response team Erasmus Medical Center David Nieuwenhuijse, Bas Oude Munnink, Reina Sikkema, Claudia Schapendonk, Irina Chestakova, Anne van der Linden, Mark Pronk, Pascal Lexmond, Corien Swaan, Manon Haverkate, Madelief

Mollers, Mart Stein, Sandra Kengne Kamga Mobou, Jeroen van Kampen, Jolanda Voermans, Aura Timen, Corine GeurtsvanKessel, Annemiek van der Eijk, Richard Molenkamp, Marion Koopmans, on behalf of the Dutch national COVID-19 response team.
EPI_ISL_415467 hCoV-19/Netherlands/NA_11/2020 Europe / Netherlands 2020-03-10 Dutch COVID-19 response team Erasmus Medical Center David Nieuwenhuijse, Bas Oude Munnink, Reina Sikkema, Claudia Schapendonk, Irina Chestakova, Anne van der Linden, Mark Pronk, Pascal Lexmond, Corien Swaan, Manon Haverkate, Madelief Mollers, Mart Stein, Sandra Kengne Kamga Mobou, Jeroen van Kampen, Jolanda Voermans, Aura Timen, Corine GeurtsvanKessel, Annemiek van der Eijk, Richard Molenkamp, Marion Koopmans, on behalf of the Dutch national COVID-19 response team.
EPI_ISL_415468 hCoV-19/Netherlands/NA_12/2020 Europe / Netherlands 2020-03-10 Dutch COVID-19 response team Erasmus Medical Center David Nieuwenhuijse, Bas Oude Munnink, Reina Sikkema, Claudia Schapendonk, Irina Chestakova, Anne van der Linden, Mark Pronk, Pascal Lexmond, Corien Swaan, Manon Haverkate, Madelief Mollers, Mart Stein, Sandra Kengne Kamga Mobou, Jeroen van Kampen, Jolanda Voermans, Aura Timen, Corine GeurtsvanKessel, Annemiek van der Eijk, Richard Molenkamp, Marion Koopmans, on behalf of the Dutch national COVID-19 response team.
EPI_ISL_415469 hCoV-19/Netherlands/NA_13/2020 Europe / Netherlands 2020-03-10 Dutch COVID-19 response team Erasmus Medical Center David Nieuwenhuijse, Bas Oude Munnink, Reina Sikkema, Claudia Schapendonk, Irina Chestakova, Anne van der Linden, Mark Pronk, Pascal Lexmond, Corien Swaan, Manon Haverkate, Madelief Mollers, Mart Stein, Sandra Kengne Kamga Mobou, Jeroen van Kampen, Jolanda Voermans, Aura Timen, Corine GeurtsvanKessel, Annemiek van der Eijk, Richard Molenkamp, Marion Koopmans, on behalf of the Dutch national COVID-19 response team.
EPI_ISL_415470 hCoV-19/Netherlands/NA_14/2020 Europe / Netherlands 2020-03-10 Dutch COVID-19 response team Erasmus Medical Center David Nieuwenhuijse, Bas Oude Munnink, Reina Sikkema, Claudia Schapendonk, Irina Chestakova, Anne van der Linden, Mark Pronk, Pascal Lexmond, Corien Swaan, Manon Haverkate, Madelief Mollers, Mart Stein, Sandra Kengne Kamga Mobou, Jeroen van Kampen, Jolanda Voermans, Aura Timen, Corine GeurtsvanKessel, Annemiek van der Eijk, Richard Molenkamp, Marion Koopmans, on behalf of the Dutch national COVID-19 response team.
EPI_ISL_415471 hCoV-19/Netherlands/NA_15/2020 Europe / Netherlands 2020-03-11 Dutch COVID-19 response team Erasmus Medical Center David Nieuwenhuijse, Bas Oude Munnink, Reina Sikkema, Claudia Schapendonk, Irina Chestakova, Anne van der Linden, Mark Pronk, Pascal Lexmond, Corien Swaan, Manon Haverkate, Madelief Mollers, Mart Stein, Sandra Kengne Kamga Mobou, Jeroen van Kampen, Jolanda Voermans, Aura Timen, Corine GeurtsvanKessel, Annemiek van der Eijk, Richard Molenkamp, Marion Koopmans, on behalf of the Dutch national COVID-19 response team.
EPI_ISL_415472 hCoV-19/Netherlands/NA_16/2020 Europe / Netherlands 2020-03-11 Dutch COVID-19 response team Erasmus Medical Center David Nieuwenhuijse, Bas Oude Munnink, Reina Sikkema, Claudia Schapendonk, Irina Chestakova, Anne van der Linden, Mark Pronk, Pascal Lexmond, Corien Swaan, Manon Haverkate, Madelief Mollers, Mart Stein, Sandra Kengne Kamga Mobou, Jeroen van Kampen, Jolanda Voermans, Aura Timen, Corine GeurtsvanKessel, Annemiek van der Eijk, Richard Molenkamp, Marion Koopmans, on behalf of the Dutch national COVID-19 response team.
EPI_ISL_415473 hCoV-19/Netherlands/NA_17/2020 Europe / Netherlands 2020-03-09 Dutch COVID-19 response team Erasmus Medical Center David Nieuwenhuijse, Bas Oude Munnink, Reina Sikkema, Claudia Schapendonk, Irina Chestakova, Anne van der Linden, Mark Pronk, Pascal Lexmond, Corien Swaan, Manon Haverkate, Madelief Mollers, Mart Stein, Sandra Kengne Kamga Mobou, Jeroen van Kampen, Jolanda Voermans, Aura Timen, Corine GeurtsvanKessel, Annemiek van der Eijk, Richard Molenkamp, Marion Koopmans, on behalf of the Dutch national COVID-19 response team.
EPI_ISL_415474 hCoV-19/Netherlands/NA_18/2020 Europe / Netherlands 2020-03-09 Dutch COVID-19 response team Erasmus Medical Center David Nieuwenhuijse, Bas Oude Munnink, Reina Sikkema, Claudia Schapendonk, Irina Chestakova, Anne van der Linden, Mark Pronk, Pascal Lexmond, Corien Swaan, Manon Haverkate, Madelief Mollers, Mart Stein, Sandra Kengne Kamga Mobou, Jeroen van Kampen, Jolanda Voermans, Aura Timen, Corine GeurtsvanKessel, Annemiek van der Eijk, Richard Molenkamp, Marion Koopmans, on behalf of the Dutch national COVID-19 response team.
EPI_ISL_415475 hCoV-19/Netherlands/NA_19/2020 Europe / Netherlands 2020-03-12 Dutch COVID-19 response team Erasmus Medical Center David Nieuwenhuijse, Bas Oude Munnink, Reina Sikkema, Claudia Schapendonk, Irina Chestakova, Anne van der Linden, Mark Pronk, Pascal Lexmond, Corien Swaan, Manon Haverkate, Madelief Mollers, Mart Stein, Sandra Kengne Kamga Mobou, Jeroen van Kampen, Jolanda Voermans, Aura Timen, Corine GeurtsvanKessel, Annemiek van der Eijk, Richard Molenkamp, Marion Koopmans, on behalf of the Dutch national COVID-19 response team.
EPI_ISL_415476 hCoV-19/Netherlands/NA_2/2020 Europe / Netherlands 2020-03-10 Dutch COVID-19 response team Erasmus Medical Center David Nieuwenhuijse, Bas Oude Munnink, Reina Sikkema, Claudia Schapendonk, Irina Chestakova, Anne van der Linden, Mark Pronk, Pascal Lexmond, Corien Swaan, Manon Haverkate, Madelief Mollers, Mart Stein, Sandra Kengne Kamga Mobou, Jeroen van Kampen, Jolanda Voermans, Aura Timen, Corine GeurtsvanKessel, Annemiek van der Eijk, Richard Molenkamp, Marion Koopmans, on behalf of the Dutch national COVID-19 response team.
EPI_ISL_415478 hCoV-19/Netherlands/NA_21/2020 Europe / Netherlands 2020-03-08 Dutch COVID-19 response team Erasmus Medical Center David Nieuwenhuijse, Bas Oude Munnink, Reina Sikkema, Claudia Schapendonk, Irina Chestakova, Anne van der Linden, Mark Pronk, Pascal Lexmond, Corien Swaan, Manon Haverkate, Madelief Mollers, Mart Stein, Sandra Kengne Kamga Mobou, Jeroen van Kampen, Jolanda Voermans, Aura Timen, Corine GeurtsvanKessel, Annemiek van der Eijk, Richard Molenkamp, Marion Koopmans, on behalf of the Dutch national COVID-19 response team.
EPI_ISL_415479 hCoV-19/Netherlands/NA_22/2020 Europe / Netherlands 2020-03-08 Dutch COVID-19 response

team    Erasmus Medical Center  David Nieuwenhuijse, Bas Oude Munnink, Reina Sikkema, Claudia Schapendonk, Irina Chestakova, Anne van der Linden, Mark Pronk, Pascal Lexmond, Corien Swaan, Manon Haverkate, Madelief Mollers, Mart Stein, Sandra Kengne Kamga Mobou, Jeroen van Kampen, Jolanda Voermans, Aura Timen, Corine GeurtsvanKessel, Annemiek van der Eijk, Richard Molenkamp, Marion Koopmans, on behalf of the Dutch national COVID-19 response team.
EPI_ISL_415480  hCoV-19/Netherlands/NA_23/2020  Europe / Netherlands    2020-03-09      Dutch COVID-19 response team    Erasmus Medical Center  David Nieuwenhuijse, Bas Oude Munnink, Reina Sikkema, Claudia Schapendonk, Irina Chestakova, Anne van der Linden, Mark Pronk, Pascal Lexmond, Corien Swaan, Manon Haverkate, Madelief Mollers, Mart Stein, Sandra Kengne Kamga Mobou, Jeroen van Kampen, Jolanda Voermans, Aura Timen, Corine GeurtsvanKessel, Annemiek van der Eijk, Richard Molenkamp, Marion Koopmans, on behalf of the Dutch national COVID-19 response team.
EPI_ISL_415481  hCoV-19/Netherlands/NA_24/2020  Europe / Netherlands    2020-03-08      Dutch COVID-19 response team    Erasmus Medical Center  David Nieuwenhuijse, Bas Oude Munnink, Reina Sikkema, Claudia Schapendonk, Irina Chestakova, Anne van der Linden, Mark Pronk, Pascal Lexmond, Corien Swaan, Manon Haverkate, Madelief Mollers, Mart Stein, Sandra Kengne Kamga Mobou, Jeroen van Kampen, Jolanda Voermans, Aura Timen, Corine GeurtsvanKessel, Annemiek van der Eijk, Richard Molenkamp, Marion Koopmans, on behalf of the Dutch national COVID-19 response team.
EPI_ISL_415482  hCoV-19/Netherlands/NA_25/2020  Europe / Netherlands    2020-03-09      Dutch COVID-19 response team    Erasmus Medical Center  David Nieuwenhuijse, Bas Oude Munnink, Reina Sikkema, Claudia Schapendonk, Irina Chestakova, Anne van der Linden, Mark Pronk, Pascal Lexmond, Corien Swaan, Manon Haverkate, Madelief Mollers, Mart Stein, Sandra Kengne Kamga Mobou, Jeroen van Kampen, Jolanda Voermans, Aura Timen, Corine GeurtsvanKessel, Annemiek van der Eijk, Richard Molenkamp, Marion Koopmans, on behalf of the Dutch national COVID-19 response team.
EPI_ISL_415483  hCoV-19/Netherlands/NA_26/2020  Europe / Netherlands    2020-03-09      Dutch COVID-19 response team    Erasmus Medical Center  David Nieuwenhuijse, Bas Oude Munnink, Reina Sikkema, Claudia Schapendonk, Irina Chestakova, Anne van der Linden, Mark Pronk, Pascal Lexmond, Corien Swaan, Manon Haverkate, Madelief Mollers, Mart Stein, Sandra Kengne Kamga Mobou, Jeroen van Kampen, Jolanda Voermans, Aura Timen, Corine GeurtsvanKessel, Annemiek van der Eijk, Richard Molenkamp, Marion Koopmans, on behalf of the Dutch national COVID-19 response team.
EPI_ISL_415484  hCoV-19/Netherlands/NA_27/2020  Europe / Netherlands    2020-03-13      Dutch COVID-19 response team    Erasmus Medical Center  David Nieuwenhuijse, Bas Oude Munnink, Reina Sikkema, Claudia Schapendonk, Irina Chestakova, Anne van der Linden, Mark Pronk, Pascal Lexmond, Corien Swaan, Manon Haverkate, Madelief Mollers, Mart Stein, Sandra Kengne Kamga Mobou, Jeroen van Kampen, Jolanda Voermans, Aura Timen, Corine GeurtsvanKessel, Annemiek van der Eijk, Richard Molenkamp, Marion Koopmans, on behalf of the Dutch national COVID-19 response team.
EPI_ISL_415485  hCoV-19/Netherlands/NA_28/2020  Europe / Netherlands    2020-03-12      Dutch COVID-19 response team    Erasmus Medical Center  David Nieuwenhuijse, Bas Oude Munnink, Reina Sikkema, Claudia Schapendonk, Irina Chestakova, Anne van der Linden, Mark Pronk, Pascal Lexmond, Corien Swaan, Manon Haverkate, Madelief Mollers, Mart Stein, Sandra Kengne Kamga Mobou, Jeroen van Kampen, Jolanda Voermans, Aura Timen, Corine GeurtsvanKessel, Annemiek van der Eijk, Richard Molenkamp, Marion Koopmans, on behalf of the Dutch national COVID-19 response team.
EPI_ISL_415486  hCoV-19/Netherlands/NA_29/2020  Europe / Netherlands    2020-03-13      Dutch COVID-19 response team    Erasmus Medical Center  David Nieuwenhuijse, Bas Oude Munnink, Reina Sikkema, Claudia Schapendonk, Irina Chestakova, Anne van der Linden, Mark Pronk, Pascal Lexmond, Corien Swaan, Manon Haverkate, Madelief Mollers, Mart Stein, Sandra Kengne Kamga Mobou, Jeroen van Kampen, Jolanda Voermans, Aura Timen, Corine GeurtsvanKessel, Annemiek van der Eijk, Richard Molenkamp, Marion Koopmans, on behalf of the Dutch national COVID-19 response team.
EPI_ISL_415487  hCoV-19/Netherlands/NA_30/2020  Europe / Netherlands    2020-03-13      Dutch COVID-19 response team    Erasmus Medical Center  David Nieuwenhuijse, Bas Oude Munnink, Reina Sikkema, Claudia Schapendonk, Irina Chestakova, Anne van der Linden, Mark Pronk, Pascal Lexmond, Corien Swaan, Manon Haverkate, Madelief Mollers, Mart Stein, Sandra Kengne Kamga Mobou, Jeroen van Kampen, Jolanda Voermans, Aura Timen, Corine GeurtsvanKessel, Annemiek van der Eijk, Richard Molenkamp, Marion Koopmans, on behalf of the Dutch national COVID-19 response team.
EPI_ISL_415488  hCoV-19/Netherlands/NA_31/2020  Europe / Netherlands    2020-03-13      Dutch COVID-19 response team    Erasmus Medical Center  David Nieuwenhuijse, Bas Oude Munnink, Reina Sikkema, Claudia Schapendonk, Irina Chestakova, Anne van der Linden, Mark Pronk, Pascal Lexmond, Corien Swaan, Manon Haverkate, Madelief Mollers, Mart Stein, Sandra Kengne Kamga Mobou, Jeroen van Kampen, Jolanda Voermans, Aura Timen, Corine GeurtsvanKessel, Annemiek van der Eijk, Richard Molenkamp, Marion Koopmans, on behalf of the Dutch national COVID-19 response team.
EPI_ISL_415489  hCoV-19/Netherlands/NA_32/2020  Europe / Netherlands    2020-03-13      Dutch COVID-19 response team    Erasmus Medical Center  David Nieuwenhuijse, Bas Oude Munnink, Reina Sikkema, Claudia Schapendonk, Irina Chestakova, Anne van der Linden, Mark Pronk, Pascal Lexmond, Corien Swaan, Manon Haverkate, Madelief Mollers, Mart Stein, Sandra Kengne Kamga Mobou, Jeroen van Kampen, Jolanda Voermans, Aura Timen, Corine GeurtsvanKessel, Annemiek van der Eijk, Richard Molenkamp, Marion Koopmans, on behalf of the Dutch national COVID-19 response team.
EPI_ISL_415491  hCoV-19/Netherlands/NA_34/2020  Europe / Netherlands    2020-03-07      Dutch COVID-19 response team    Erasmus Medical Center  David Nieuwenhuijse, Bas Oude Munnink, Reina Sikkema, Claudia Schapendonk, Irina Chestakova, Anne van der Linden, Mark Pronk, Pascal Lexmond, Corien Swaan, Manon Haverkate, Madelief Mollers, Mart Stein, Sandra Kengne Kamga Mobou, Jeroen van Kampen, Jolanda Voermans, Aura Timen, Corine GeurtsvanKessel, Annemiek van der Eijk, Richard Molenkamp, Marion Koopmans, on behalf of the Dutch national

COVID-19 response team.
EPI_ISL_415492 hCoV-19/Netherlands/NA_35/2020 Europe / Netherlands 2020-03-10 Dutch COVID-19 response team Erasmus Medical Center David Nieuwenhuijse, Bas Oude Munnink, Reina Sikkema, Claudia Schapendonk, Irina Chestakova, Anne van der Linden, Mark Pronk, Pascal Lexmond, Corien Swaan, Manon Haverkate, Madelief Mollers, Mart Stein, Sandra Kengne Kamga Mobou, Jeroen van Kampen, Jolanda Voermans, Aura Timen, Corine GeurtsvanKessel, Annemiek van der Eijk, Richard Molenkamp, Marion Koopmans, on behalf of the Dutch national COVID-19 response team.
EPI_ISL_415493 hCoV-19/Netherlands/NA_4/2020 Europe / Netherlands 2020 Dutch COVID-19 response team Erasmus Medical Center David Nieuwenhuijse, Bas Oude Munnink, Reina Sikkema, Claudia Schapendonk, Irina Chestakova, Anne van der Linden, Mark Pronk, Pascal Lexmond, Corien Swaan, Manon Haverkate, Madelief Mollers, Mart Stein, Sandra Kengne Kamga Mobou, Jeroen van Kampen, Jolanda Voermans, Aura Timen, Corine GeurtsvanKessel, Annemiek van der Eijk, Richard Molenkamp, Marion Koopmans, on behalf of the Dutch national COVID-19 response team.
EPI_ISL_415494 hCoV-19/Netherlands/NA_5/2020 Europe / Netherlands 2020 Dutch COVID-19 response team Erasmus Medical Center David Nieuwenhuijse, Bas Oude Munnink, Reina Sikkema, Claudia Schapendonk, Irina Chestakova, Anne van der Linden, Mark Pronk, Pascal Lexmond, Corien Swaan, Manon Haverkate, Madelief Mollers, Mart Stein, Sandra Kengne Kamga Mobou, Jeroen van Kampen, Jolanda Voermans, Aura Timen, Corine GeurtsvanKessel, Annemiek van der Eijk, Richard Molenkamp, Marion Koopmans, on behalf of the Dutch national COVID-19 response team.
EPI_ISL_415495 hCoV-19/Netherlands/NA_6/2020 Europe / Netherlands 2020-03-10 Dutch COVID-19 response team Erasmus Medical Center David Nieuwenhuijse, Bas Oude Munnink, Reina Sikkema, Claudia Schapendonk, Irina Chestakova, Anne van der Linden, Mark Pronk, Pascal Lexmond, Corien Swaan, Manon Haverkate, Madelief Mollers, Mart Stein, Sandra Kengne Kamga Mobou, Jeroen van Kampen, Jolanda Voermans, Aura Timen, Corine GeurtsvanKessel, Annemiek van der Eijk, Richard Molenkamp, Marion Koopmans, on behalf of the Dutch national COVID-19 response team.
EPI_ISL_415496 hCoV-19/Netherlands/NA_7/2020 Europe / Netherlands 2020-03-09 Dutch COVID-19 response team Erasmus Medical Center David Nieuwenhuijse, Bas Oude Munnink, Reina Sikkema, Claudia Schapendonk, Irina Chestakova, Anne van der Linden, Mark Pronk, Pascal Lexmond, Corien Swaan, Manon Haverkate, Madelief Mollers, Mart Stein, Sandra Kengne Kamga Mobou, Jeroen van Kampen, Jolanda Voermans, Aura Timen, Corine GeurtsvanKessel, Annemiek van der Eijk, Richard Molenkamp, Marion Koopmans, on behalf of the Dutch national COVID-19 response team.
EPI_ISL_415497 hCoV-19/Netherlands/NA_8/2020 Europe / Netherlands 2020-03-09 Dutch COVID-19 response team Erasmus Medical Center David Nieuwenhuijse, Bas Oude Munnink, Reina Sikkema, Claudia Schapendonk, Irina Chestakova, Anne van der Linden, Mark Pronk, Pascal Lexmond, Corien Swaan, Manon Haverkate, Madelief Mollers, Mart Stein, Sandra Kengne Kamga Mobou, Jeroen van Kampen, Jolanda Voermans, Aura Timen, Corine GeurtsvanKessel, Annemiek van der Eijk, Richard Molenkamp, Marion Koopmans, on behalf of the Dutch national COVID-19 response team.
EPI_ISL_415498 hCoV-19/Netherlands/NA_9/2020 Europe / Netherlands 2020-03-09 Dutch COVID-19 response team Erasmus Medical Center David Nieuwenhuijse, Bas Oude Munnink, Reina Sikkema, Claudia Schapendonk, Irina Chestakova, Anne van der Linden, Mark Pronk, Pascal Lexmond, Corien Swaan, Manon Haverkate, Madelief Mollers, Mart Stein, Sandra Kengne Kamga Mobou, Jeroen van Kampen, Jolanda Voermans, Aura Timen, Corine GeurtsvanKessel, Annemiek van der Eijk, Richard Molenkamp, Marion Koopmans, on behalf of the Dutch national COVID-19 response team.
EPI_ISL_415499 hCoV-19/Netherlands/NoordBrabant_41/2020 Europe / Netherlands / Noord Brabant 2020 Dutch COVID-19 response team Erasmus Medical Center David Nieuwenhuijse, Bas Oude Munnink, Reina Sikkema, Claudia Schapendonk, Irina Chestakova, Anne van der Linden, Mark Pronk, Pascal Lexmond, Corien Swaan, Manon Haverkate, Madelief Mollers, Mart Stein, Sandra Kengne Kamga Mobou, Jeroen van Kampen, Jolanda Voermans, Aura Timen, Corine GeurtsvanKessel, Annemiek van der Eijk, Richard Molenkamp, Marion Koopmans, on behalf of the Dutch national COVID-19 response team.
EPI_ISL_416400 hCoV-19/Shanghai/SH0112/2020 Asia / China / Shanghai 2020-02-02 Shanghai Public Health Clinical Center, Shanghai Medical College, Fudan University National Research Center for Translational Medicine (Shanghai), Ruijin Hospital affiliated to Shanghai Jiao Tong University School of Medicine & Shanghai Public Health Clinical Center Shengyue Wang, Xiaonan Zhang, Gang Lu, Yun Tan, Yun Ling, Hongzhou Lu, Saijuan Chen
EPI_ISL_416401 hCoV-19/Shanghai/SH0114/2020 Asia / China / Shanghai 2020-02-02 Shanghai Public Health Clinical Center, Shanghai Medical College, Fudan University National Research Center for Translational Medicine (Shanghai), Ruijin Hospital affiliated to Shanghai Jiao Tong University School of Medicine & Shanghai Public Health Clinical Center Shengyue Wang, Xiaonan Zhang, Gang Lu, Yun Tan, Yun Ling, Hongzhou Lu, Saijuan Chen
EPI_ISL_416402 hCoV-19/Shanghai/SH0115/2020 Asia / China / Shanghai 2020-02-11 Shanghai Public Health Clinical Center, Shanghai Medical College, Fudan University National Research Center for Translational Medicine (Shanghai), Ruijin Hospital affiliated to Shanghai Jiao Tong University School of Medicine & Shanghai Public Health Clinical Center Shengyue Wang, Xiaonan Zhang, Gang Lu, Yun Tan, Yun Ling, Hongzhou Lu, Saijuan Chen
EPI_ISL_416403 hCoV-19/Shanghai/SH0117/2020 Asia / China / Shanghai 2020-02-02 Shanghai Public Health Clinical Center, Shanghai Medical College, Fudan University National Research Center for Translational Medicine (Shanghai), Ruijin Hospital affiliated to Shanghai Jiao Tong University School of Medicine & Shanghai Public Health Clinical Center Shengyue Wang, Xiaonan Zhang, Gang Lu, Yun Tan, Yun Ling, Hongzhou Lu, Saijuan Chen
EPI_ISL_416404 hCoV-19/Shanghai/SH0119/2020 Asia / China / Shanghai 2020-02-09 Shanghai Public Health

Clinical Center, Shanghai Medical College, Fudan University National Research Center for Translational Medicine (Shanghai), Ruijin Hospital affiliated to Shanghai Jiao Tong University School of Medicine & Shanghai Public Health Clinical Center Shengyue Wang, Xiaonan Zhang, Gang Lu, Yun Tan, Yun Ling, Hongzhou Lu, Saijuan Chen
EPI_ISL_416405 hCoV-19/Shanghai/SH0121/2020 Asia / China / Shanghai 2020-02-02 Shanghai Public Health Clinical Center, Shanghai Medical College, Fudan University National Research Center for Translational Medicine (Shanghai), Ruijin Hospital affiliated to Shanghai Jiao Tong University School of Medicine & Shanghai Public Health Clinical Center Shengyue Wang, Xiaonan Zhang, Gang Lu, Yun Tan, Yun Ling, Hongzhou Lu, Saijuan Chen
EPI_ISL_416406 hCoV-19/Shanghai/SH0125/2020 Asia / China / Shanghai 2020-02-15 Shanghai Public Health Clinical Center, Shanghai Medical College, Fudan University National Research Center for Translational Medicine (Shanghai), Ruijin Hospital affiliated to Shanghai Jiao Tong University School of Medicine & Shanghai Public Health Clinical Center Shengyue Wang, Xiaonan Zhang, Gang Lu, Yun Tan, Yun Ling, Hongzhou Lu, Saijuan Chen
EPI_ISL_416407 hCoV-19/Shanghai/SH0126/2020 Asia / China / Shanghai 2020-02-15 Shanghai Public Health Clinical Center, Shanghai Medical College, Fudan University National Research Center for Translational Medicine (Shanghai), Ruijin Hospital affiliated to Shanghai Jiao Tong University School of Medicine & Shanghai Public Health Clinical Center Shengyue Wang, Xiaonan Zhang, Gang Lu, Yun Tan, Yun Ling, Hongzhou Lu, Saijuan Chen
EPI_ISL_416409 hCoV-19/Shanghai/SH0128/2020 Asia / China / Shanghai 2020-02-02 Shanghai Public Health Clinical Center, Shanghai Medical College, Fudan University National Research Center for Translational Medicine (Shanghai), Ruijin Hospital affiliated to Shanghai Jiao Tong University School of Medicine & Shanghai Public Health Clinical Center Shengyue Wang, Xiaonan Zhang, Gang Lu, Yun Tan, Yun Ling, Hongzhou Lu, Saijuan Chen
EPI_ISL_416410 hCoV-19/Australia/VIC02/2020 Oceania / Australia / Victoria / Melbourne 2020-01-24 Victorian Infectious Diseases Reference Laboratory (VIDRL) Victorian Infectious Diseases Reference Laboratory and Microbiological Diagnostic Unit Public Health Laboratory, Doherty Institute Caly L., Seemann T., Schultz M., Druce J., Taiaroa, G.
EPI_ISL_416411 hCoV-19/Australia/VIC03/2020 Oceania / Australia / Victoria / Melbourne 2020-01-25 Victorian Infectious Diseases Reference Laboratory (VIDRL) Victorian Infectious Diseases Reference Laboratory and Microbiological Diagnostic Unit Public Health Laboratory, Doherty Institute Caly L., Seemann T., Schultz M., Druce J., Taiaroa, G.
EPI_ISL_416412 hCoV-19/Australia/VIC04/2020 Oceania / Australia / Victoria / Melbourne 2020-03-02 Victorian Infectious Diseases Reference Laboratory (VIDRL) Victorian Infectious Diseases Reference Laboratory and Microbiological Diagnostic Unit Public Health Laboratory, Doherty Institute Caly L., Seemann T., Schultz M., Druce J., Taiaroa, G.
EPI_ISL_416413 hCoV-19/Australia/VIC05/2020 Oceania / Australia / Victoria / Melbourne 2020-03-05 Victorian Infectious Diseases Reference Laboratory (VIDRL) Victorian Infectious Diseases Reference Laboratory and Microbiological Diagnostic Unit Public Health Laboratory, Doherty Institute Caly L., Seemann T., Schultz M., Druce J., Taiaroa, G.
EPI_ISL_416415 hCoV-19/Australia/VIC07/2020 Oceania / Australia / Victoria / Melbourne 2020-02-08 Victorian Infectious Diseases Reference Laboratory (VIDRL) Victorian Infectious Diseases Reference Laboratory and Microbiological Diagnostic Unit Public Health Laboratory, Doherty Institute Caly L., Seemann T., Schultz M., Druce J., Taiaroa, G.
EPI_ISL_416430 hCoV-19/Vietnam/CM295/2020 Asia / Vietnam / Hanoi 2020-03-06 National Influenza Center, National Institute of Hygiene and Epidemiology (NIHE) National Influenza Center, National Institute of Hygiene and Epidemiology (NIHE) Le Quynh Mai, Taichiro Takemura, Meng Ling Moi, Takeshi Nabeshima, Nguyen Le Khanh Hang, Hoang Vu Mai Phuong, Ung Thi Hong Trang, Le Thi Thanh, Nguyen Vu Son, Vuong Duc Cuong, Pham Thi Hien, Tran Thu Huong, Nguyen Phuong Anh, Pham Hong Quynh Anh, Kouichi Morita, Futoshi Hasebe, Dang Duc Anh
EPI_ISL_416431 hCoV-19/Vietnam/CM296/2020 Asia / Vietnam / Hanoi 2020-03-06 National Influenza Center, National Institute of Hygiene and Epidemiology (NIHE) National Influenza Center, National Institute of Hygiene and Epidemiology (NIHE) Le Quynh Mai, Taichiro Takemura, Meng Ling Moi, Takeshi Nabeshima, Nguyen Le Khanh Hang, Hoang Vu Mai Phuong, Ung Thi Hong Trang, Le Thi Thanh, Nguyen Vu Son, Vuong Duc Cuong, Pham Thi Hien, Tran Thu Huong, Nguyen Phuong Anh, Pham Hong Quynh Anh, Kouichi Morita, Futoshi Hasebe, Dang Duc Anh
EPI_ISL_416432 hCoV-19/Saudi Arabia/KAIMRC-Alghoribi/2020 Asia / Saudi Arabia / Riyadh 2020-03-07 Clinical Microbiology Lab Infectious Disease Research Department, King Abdullah International Medical Research Center (KAIMRC) Majed Alghoribi, Sadeem Alhayli, Abdulrahman Alswaji, Liliane Okdah, Sameera Al Johani, Michel Doumith
EPI_ISL_416433 hCoV-19/USA/WA-UW77/2020 North America / USA / Washington 2020-03-10 UW Virology Lab UW Virology Lab Pavitra Roychoudhury, Hong Xie, Keith Jerome, Alexander Greninger
EPI_ISL_416434 hCoV-19/USA/WA-UW78/2020 North America / USA / Washington 2020-03-10 UW Virology Lab UW Virology Lab Pavitra Roychoudhury, Hong Xie, Keith Jerome, Alexander Greninger
EPI_ISL_416435 hCoV-19/USA/WA-UW79/2020 North America / USA / Washington 2020-03-10 UW Virology Lab UW Virology Lab Pavitra Roychoudhury, Hong Xie, Keith Jerome, Alexander Greninger
EPI_ISL_416436 hCoV-19/USA/WA-UW80/2020 North America / USA / Washington 2020-03-10 UW Virology Lab UW Virology Lab Pavitra Roychoudhury, Hong Xie, Keith Jerome, Alexander Greninger
EPI_ISL_416438 hCoV-19/USA/WA-UW82/2020 North America / USA / Washington 2020-03-10 UW Virology Lab UW Virology Lab Pavitra Roychoudhury, Hong Xie, Keith Jerome, Alexander Greninger

EPI_ISL_416439 hCoV-19/USA/WA-UW83/2020 North America / USA / Washington 2020-03-10 UW Virology Lab UW Virology Lab Pavitra Roychoudhury, Hong Xie, Keith Jerome, Alexander Greninger
EPI_ISL_416440 hCoV-19/USA/WA-UW84/2020 North America / USA / Washington 2020-03-10 UW Virology Lab UW Virology Lab Pavitra Roychoudhury, Hong Xie, Keith Jerome, Alexander Greninger
EPI_ISL_416441 hCoV-19/USA/WA-UW85/2020 North America / USA / Washington 2020-03-10 UW Virology Lab UW Virology Lab Pavitra Roychoudhury, Hong Xie, Keith Jerome, Alexander Greninger
EPI_ISL_416442 hCoV-19/USA/WA-UW86/2020 North America / USA / Washington 2020-03-10 UW Virology Lab UW Virology Lab Pavitra Roychoudhury, Hong Xie, Keith Jerome, Alexander Greninger
EPI_ISL_416443 hCoV-19/USA/WA-UW87/2020 North America / USA / Washington 2020-03-10 UW Virology Lab UW Virology Lab Pavitra Roychoudhury, Hong Xie, Keith Jerome, Alexander Greninger
EPI_ISL_416444 hCoV-19/USA/WA-UW88/2020 North America / USA / Washington 2020-03-10 UW Virology Lab UW Virology Lab Pavitra Roychoudhury, Hong Xie, Keith Jerome, Alexander Greninger
EPI_ISL_416445 hCoV-19/USA/WA-UW89/2020 North America / USA / Washington 2020-03-10 UW Virology Lab UW Virology Lab Pavitra Roychoudhury, Hong Xie, Keith Jerome, Alexander Greninger
EPI_ISL_416446 hCoV-19/USA/WA-UW90/2020 North America / USA / Washington 2020-03-10 UW Virology Lab UW Virology Lab Pavitra Roychoudhury, Hong Xie, Keith Jerome, Alexander Greninger
EPI_ISL_416447 hCoV-19/USA/WA-UW91/2020 North America / USA / Washington 2020-03-10 UW Virology Lab UW Virology Lab Pavitra Roychoudhury, Hong Xie, Keith Jerome, Alexander Greninger
EPI_ISL_416448 hCoV-19/USA/WA-UW92/2020 North America / USA / Washington 2020-03-11 UW Virology Lab UW Virology Lab Pavitra Roychoudhury, Hong Xie, Keith Jerome, Alexander Greninger
EPI_ISL_416449 hCoV-19/USA/WA-UW93/2020 North America / USA / Washington 2020-03-11 UW Virology Lab UW Virology Lab Pavitra Roychoudhury, Hong Xie, Keith Jerome, Alexander Greninger
EPI_ISL_416450 hCoV-19/USA/WA-UW94/2020 North America / USA / Washington 2020-03-11 UW Virology Lab UW Virology Lab Pavitra Roychoudhury, Hong Xie, Keith Jerome, Alexander Greninger
EPI_ISL_416451 hCoV-19/USA/WA-UW95/2020 North America / USA / Washington 2020-03-10 UW Virology Lab UW Virology Lab Pavitra Roychoudhury, Hong Xie, Keith Jerome, Alexander Greninger
EPI_ISL_416452 hCoV-19/USA/WA-UW96/2020 North America / USA / Washington 2020-03-10 UW Virology Lab UW Virology Lab Pavitra Roychoudhury, Hong Xie, Keith Jerome, Alexander Greninger
EPI_ISL_416453 hCoV-19/USA/WA-UW36/2020 North America / USA / Washington 2020-03-07 UW Virology Lab UW Virology Lab Pavitra Roychoudhury, Hong Xie, Keith Jerome, Alexander Greninger
EPI_ISL_416454 hCoV-19/USA/WA-UW37/2020 North America / USA / Washington 2020-03-06 UW Virology Lab UW Virology Lab Pavitra Roychoudhury, Hong Xie, Keith Jerome, Alexander Greninger
EPI_ISL_416455 hCoV-19/USA/WA-UW38/2020 North America / USA / Washington 2020-03-07 UW Virology Lab UW Virology Lab Pavitra Roychoudhury, Hong Xie, Keith Jerome, Alexander Greninger
EPI_ISL_416456 hCoV-19/USA/WA-UW39/2020 North America / USA / Washington 2020-03-06 UW Virology Lab UW Virology Lab Pavitra Roychoudhury, Hong Xie, Keith Jerome, Alexander Greninger
EPI_ISL_416457 hCoV-19/USA/CA-MG0987/2020 North America / USA / California / San Diego County 2020-03-18 Andersen Lab, The Scripps Research Institute Andersen Lab, The Scripps Research Institute Mark Zeller, Catie Anderson, Emily Spender, Sarah Topol, Raphaelle Klitting, Refugio Robles-Sikisaka, Karthik Gangavarapu, Laura Nicholson, Kristian Andersen
EPI_ISL_416458 hCoV-19/Kuwait/KU12/2020 Asia / Kuwait / Hawali 2020-03-02 Virology laboratory Ministry of Health Kuwait sequenced at Dasman Diabetes Institute Dasman Diabetes Institute Fahd Al-Mulla, Sumi John, Sara Alqabandi, Rasheeba iqbal, Motasem Melhem, Ebaa alOzairi, Qais Al-Duwairi
EPI_ISL_416460 hCoV-19/USA/WA-S5/2020 North America / USA / Washington / King County 2020-02-29 Seattle Flu Study Seattle Flu Study Chu et al
EPI_ISL_416461 hCoV-19/USA/WA-S6/2020 North America / USA / Washington / King County 2020-02-29 Seattle Flu Study Seattle Flu Study Chu et al
EPI_ISL_416462 hCoV-19/USA/WA-S7/2020 North America / USA / Washington 2020-02-24 Seattle Flu Study Seattle Flu Study Chu et al
EPI_ISL_416465 hCoV-19/USA/WA-S10/2020 North America / USA / Washington / King County 2020-02-29 Seattle Flu Study Seattle Flu Study Chu et al
EPI_ISL_416466 hCoV-19/USA/WA-S11/2020 North America / USA / Washington / King County 2020-03-03 Seattle Flu Study Seattle Flu Study Chu et al
EPI_ISL_416467 hCoV-19/Belgium/MTR-03021/2020 Europe / Belgium / Holsbeek 2020-03-02 KU Leuven, Clinical and Epidemiological Virology KU Leuven, Clinical and Epidemiological Virology Bert Vanmechelen, Tony Wawina, Joan Marti-Carreras, Piet Maes
EPI_ISL_416468 hCoV-19/Belgium/GMH-03022/2020 Europe / Belgium / Holsbeek 2020-03-02 KU Leuven, Clinical and Epidemiological Virology KU Leuven, Clinical and Epidemiological Virology Bert Vanmechelen, Tony Wawina, Joan Marti-Carreras, Piet Maes
EPI_ISL_416469 hCoV-19/Belgium/SN-03031/2020 Europe / Belgium / Kessel-Lo 2020-03-03 KU Leuven, Clinical and Epidemiological Virology KU Leuven, Clinical and Epidemiological Virology Bert Vanmechelen, Tony Wawina, Joan Marti-Carreras, Piet Maes
EPI_ISL_416470 hCoV-19/Belgium/DB-03023/2020 Europe / Belgium / Couthuin 2020-03-02 KU Leuven, Clinical and Epidemiological Virology KU Leuven, Clinical and Epidemiological Virology Bert Vanmechelen, Tony Wawina, Joan Marti-Carreras, Piet Maes
EPI_ISL_416471 hCoV-19/Belgium/DBD-03024/2020 Europe / Belgium / Kessel-Lo 2020-03-02 KU Leuven, Clinical and Epidemiological Virology KU Leuven, Clinical and Epidemiological Virology Bert Vanmechelen, Tony Wawina, Joan Marti-Carreras, Piet Maes
EPI_ISL_416472 hCoV-19/Belgium/UMF-03025/2020 Europe / Belgium / Kessel-Lo 2020-03-02 KU Leuven, Clinical and Epidemiological Virology KU Leuven, Clinical and Epidemiological Virology Bert

Vanmechelen, Tony Wawina, Joan Marti-Carreras, Piet Maes
EPI_ISL_416473 hCoV-19/Hangzhou/ZJU-08/2020 Asia / China / Hangzhou 2020-01-26 State Key Laboratory for Diagnosis and Treatment of Infectious Diseases, National Clinical Research Center for Infectious Diseases, First Affiliated Hospital, Zhejiang University School of Medicine, Hangzhou, China 310003 State Key Laboratory for Diagnosis and Treatment of Infectious Diseases, National Clinical Research Center for Infectious Diseases, First Affiliated Hospital, Zhejiang University School of Medicine, Hangzhou, China 310003 Hangping Yao, Nanping Wu, Chao Jiang, Xiangyun Lu, Linfang Cheng, Fumin Liu, Zhigang Wu, Haibo Wu, Changzhong Jin, Min Zheng, Lanjuan Li
EPI_ISL_416474 hCoV-19/Hangzhou/ZJU-09/2020 Asia / China / Hangzhou 2020-01-28 State Key Laboratory for Diagnosis and Treatment of Infectious Diseases, National Clinical Research Center for Infectious Diseases, First Affiliated Hospital, Zhejiang University School of Medicine, Hangzhou, China 310003 State Key Laboratory for Diagnosis and Treatment of Infectious Diseases, National Clinical Research Center for Infectious Diseases, First Affiliated Hospital, Zhejiang University School of Medicine, Hangzhou, China 310003 Hangping Yao, Nanping Wu, Chao Jiang, Xiangyun Lu, Linfang Cheng, Fumin Liu, Zhigang Wu, Haibo Wu, Changzhong Jin, Min Zheng, Lanjuan Li
EPI_ISL_416475 hCoV-19/Belgium/DBA-03032/2020 Europe / Belgium / Leuven 2020-03-03 KU Leuven, Clinical and Epidemiological Virology KU Leuven, Clinical and Epidemiological Virology Bert Vanmechelen, Tony Wawina, Joan Marti-Carreras, Piet Maes
EPI_ISL_416476 hCoV-19/Belgium/MTR-03026/2020 Europe / Belgium / Holsbeek 2020-03-02 KU Leuven, Clinical and Epidemiological Virology KU Leuven, Clinical and Epidemiological Virology Bert Vanmechelen, Tony Wawina, Joan Marti-Carreras, Piet Maes
EPI_ISL_416477 hCoV-19/Georgia/Tb-390/2020 Asia / Georgia / Tbilisi 2020-03-08 R. G. Lugar Center for Public Health Research, National Center for Disease Control and Public Health (NCDC) of Georgia. R. G. Lugar Center for Public Health Research, National Center for Disease Control and Public Health (NCDC) of Georgia. Marine Murtskhvaladze, Nato Kotaria, Ann Machablishvili, Lela Sabadze, Mari Gavashelidze, Ana Papkiauri, Meri Pantsulaia, Gvantsa Brachveli, Tata Imnadze, Tamar Jashiashvili, Tea Tevdoradze, Ketevan Sidamonidze, Ekaterine Khmaladze, Ekaterine Zhghenti, Roena Sukhiashvili, Mariam Zakalashvili, Lela Urushadze, Magda Dgebuadze, Giorgi Tomashvili, Davit Tsaguria, Ekaterine Zangaladze, Nino Berishvili, Gvantsa Chanturia, Adam Kotorashvili, Maia Alkhazashvili, Irma Burjanadze, Anna Kasradze, Khatuna Zakhashvili, Paata Imnadze, Amiran Gamkrelidze.
EPI_ISL_416478 hCoV-19/Georgia/Tb-673/2020 Asia / Georgia / Tbilisi 2020-03-14 R. G. Lugar Center for Public Health Research, National Center for Disease Control and Public Health (NCDC) of Georgia. R. G. Lugar Center for Public Health Research, National Center for Disease Control and Public Health (NCDC) of Georgia. Marine Murtskhvaladze, Nato Kotaria, Ann Machablishvili, Lela Sabadze, Mari Gavashelidze, Ana Papkiauri, Meri Pantsulaia, Gvantsa Brachveli, Tata Imnadze, Tamar Jashiashvili, Tea Tevdoradze, Ketevan Sidamonidze, Ekaterine Khmaladze, Ekaterine Zhghenti, Roena Sukhiashvili, Mariam Zakalashvili, Lela Urushadze, Magda Dgebuadze, Giorgi Tomashvili, Davit Tsaguria, Ekaterine Zangaladze, Nino Berishvili, Gvantsa Chanturia, Adam Kotorashvili, Maia Alkhazashvili, Irma Burjanadze, Anna Kasradze, Khatuna Zakhashvili, Paata Imnadze, Amiran Gamkrelidze.
EPI_ISL_416479 hCoV-19/Georgia/Tb-273/2020 Asia / Georgia / Tbilisi 2020-03-05 R. G. Lugar Center for Public Health Research, National Center for Disease Control and Public Health (NCDC) of Georgia. R. G. Lugar Center for Public Health Research, National Center for Disease Control and Public Health (NCDC) of Georgia. Marine Murtskhvaladze, Nato Kotaria, Ann Machablishvili, Lela Sabadze, Mari Gavashelidze, Ana Papkiauri, Meri Pantsulaia, Gvantsa Brachveli, Tata Imnadze, Tamar Jashiashvili, Tea Tevdoradze, Ketevan Sidamonidze, Ekaterine Khmaladze, Ekaterine Zhghenti, Roena Sukhiashvili, Mariam Zakalashvili, Lela Urushadze, Magda Dgebuadze, Giorgi Tomashvili, Davit Tsaguria, Ekaterine Zangaladze, Nino Berishvili, Gvantsa Chanturia, Adam Kotorashvili, Maia Alkhazashvili, Irma Burjanadze, Anna Kasradze, Khatuna Zakhashvili, Paata Imnadze, Amiran Gamkrelidze.
EPI_ISL_416480 hCoV-19/Georgia/Tb-537/2020 Asia / Georgia / Tbilisi 2020-03-11 R. G. Lugar Center for Public Health Research, National Center for Disease Control and Public Health (NCDC) of Georgia. R. G. Lugar Center for Public Health Research, National Center for Disease Control and Public Health (NCDC) of Georgia. Ann Machablishvili, Nato Kotaria, Marine Murtskhvaladze, Lela Sabadze, Mari Gavashelidze, Ana Papkiauri, Meri Pantsulaia, Gvantsa Brachveli, Tata Imnadze, Tamar Jashiashvili, Tea Tevdoradze, Ketevan Sidamonidze, Ekaterine Khmaladze, Ekaterine Zhghenti, Roena Sukhiashvili, Mariam Zakalashvili, Lela Urushadze, Magda Dgebuadze, Giorgi Tomashvili, Davit Tsaguria, Ekaterine Zangaladze, Nino Berishvili, Gvantsa Chanturia, Adam Kotorashvili, Maia Alkhazashvili, Irma Burjanadze, Anna Kasradze, Khatuna Zakhashvili, Paata Imnadze, Amiran Gamkrelidze.
EPI_ISL_416481 hCoV-19/Georgia/Tb-712/2020 Asia / Georgia / Tbilisi 2020-03-16 R. G. Lugar Center for Public Health Research, National Center for Disease Control and Public Health (NCDC) of Georgia. R. G. Lugar Center for Public Health Research, National Center for Disease Control and Public Health (NCDC) of Georgia. Gvantsa Chanturia, Marine Murtskhvaladze, Nato Kotaria, Ann Machablishvili, Lela Sabadze, Mari Gavashelidze, Ana Papkiauri, Meri Pantsulaia, Gvantsa Brachveli, Tata Imnadze, Tamar Jashiashvili, Tea Tevdoradze, Ketevan Sidamonidze, Ekaterine Khmaladze, Ekaterine Zhghenti, Roena Sukhiashvili, Mariam Zakalashvili, Lela Urushadze, Magda Dgebuadze, Giorgi Tomashvili, Davit Tsaguria, Ekaterine Zangaladze, Nino Berishvili, Adam Kotorashvili, Maia Alkhazashvili, Irma Burjanadze, Anna Kasradze, Khatuna Zakhashvili, Paata Imnadze, Amiran Gamkrelidze.
EPI_ISL_416482 hCoV-19/Georgia/Tb/2020 Asia / Georgia / Tbilisi 2020-03-13 R. G. Lugar Center for Public Health Research, National Center for Disease Control and Public Health (NCDC) of Georgia. R. G. Lugar Center for Public Health Research, National Center for Disease Control and Public Health (NCDC) of Georgia. Adam Kotorashvili, Marine Murtskhvaladze, Nato Kotaria, Ann Machablishvili, Lela Sabadze, Mari

Gavashelidze, Ana Papkiauri, Meri Pantsulaia, Gvantsa Brachveli, Tata Imnadze, Tamar Jashiashvili, Tea Tevdoradze, Ketevan Sidamonidze, Ekaterine Khmaladze, Ekaterine Zhghenti, Roena Sukhiashvili, Mariam Zakalashvili, Lela Urushadze, Magda Dgebuadze, Giorgi Tomashvili, Davit Tsaguria, Ekaterine Zangaladze, Nino Berishvili, Gvantsa Chanturia, Maia Alkhazashvili, Irma Burjanadze, Anna Kasradze, Khatuna Zakhashvili, Paata Imnadze, Amiran Gamkrelidze.
EPI_ISL_416484 hCoV-19/Spain/Valencia5/2020 Europe / Spain / Comunitat Valenciana / Valencia 2020-02-27 Servicio de Microbiología. Consorcio Hospital General Universitario de Valencia Sequencing and Bioinformatics Service and Molecular Epidemiology Research Group. FISABIO-Public Health Maria Dolores Ocete, Concepcion Gimeno, Giuseppe D'Auria, Griselda De Marco, Neris Garcia-Gonzalez, Maria Alma Bracho, Fernando Gonzalez-Candelas
EPI_ISL_416485 hCoV-19/Spain/Valencia6/2020 Europe / Spain / Comunitat Valenciana / Valencia 2020-02-27 Servicio de Microbiologia. Consorcio Hospital General Universitario de Valencia Sequencing and Bioinformatics Service and Molecular Epidemiology Research Group. FISABIO-Public Health Griselda De Marco, Neris Garcia-Gonzalez, Maria Alma Bracho, Maria Dolores Ocete, Concepcion Gimeno, Giuseppe D'Auria, Fernando Gonzalez-Candelas
EPI_ISL_416486 hCoV-19/Spain/Valencia7/2020 Europe / Spain / Comunitat Valenciana / Valencia 2020-03-02 Servicio de Microbiología. Consorcio Hospital General Universitario de Valencia Sequencing and Bioinformatics Service and Molecular Epidemiology Research Group. FISABIO-Public Health Neris Garcia-Gonzalez, Maria Alma Bracho, Maria Dolores Ocete, Concepcion Gimeno, Giuseppe D'Auria, Griselda De Marco, Fernando Gonzalez-Candelas
EPI_ISL_416487 hCoV-19/Spain/Valencia8/2020 Europe / Spain / Comunitat Valenciana / Valencia 2020-03-04 Servicio de Microbiología. Consorcio Hospital General Universitario de Valencia Sequencing and Bioinformatics Service and Molecular Epidemiology Research Group. FISABIO-Public Health Giuseppe D'Auria, Griselda De Marco, Neris Garcia-Gonzalez, Maria Alma Bracho, Maria Dolores Ocete, Concepcion Gimeno, Fernando Gonzalez-Candelas
EPI_ISL_416488 hCoV-19/Poland/PL_P1/2020 Europe / Poland / Zielonogorskie 2020-03-03 ViroGenetics - BSL3 Laboratory of Virology; Human Genome Variation Research Group & Genomics Centre MCB; Bioinformatics Research Group Department of Virology ViroGenetics - BSL3 Laboratory of Virology; Human Genome Variation Research Group & Genomics Centre MCB; Bioinformatics Research Group Department of Virology Aleksandra Milewska, Ewelina Pośpiech, Agata Jarosz, Adrianna Klajmon, Kamila Marszałek, Katarzyna Pancer, Magdalena Rzeczkowska, Tomasz Wołkowicz, Katarzyna Zacharczuk, Agnieszka Kołakowska-Kulesza, Natalia Wolaniuk, Ewelina Hallman-Szelińska, Paweł P Łabaj, Wojciech Branicki, Krzysztof Pyrć
EPI_ISL_416489 hCoV-19/USA/WI-UW-02/2020 North America / USA / Wisconsin / Madison 2020-03-15 University of Wisconsin-Madison AIDS Vaccine Research Laboratories University of Wisconsin-Madison AIDS Vaccine Research Laboratories Gage Moreno, Katarina Braun, et al. AIDS Vaccine Research Laboratories
EPI_ISL_416491 hCoV-19/USA/WI-UW-04/2020 North America / USA / Wisconsin / Waunakee 2020-03-15 University of Wisconsin-Madison AIDS Vaccine Research Laboratories University of Wisconsin-Madison AIDS Vaccine Research Laboratories Gage Moreno, Katarina Braun, et al. AIDS Vaccine Research Laboratories
EPI_ISL_416492 hCoV-19/USA/WI-UW-05/2020 North America / USA / Wisconsin / Fitchburg 2020-03-15 University of Wisconsin-Madison AIDS Vaccine Research Laboratories University of Wisconsin-Madison AIDS Vaccine Research Laboratories Gage Moreno, Katarina Braun, et al. AIDS Vaccine Research Laboratories
EPI_ISL_416493 hCoV-19/France/HF2196/2020 Europe / France / Hauts de France / Château-Thierry 2020-03-08 CH Jean de Navarre Laboratoire de Biologie National Reference Center for Viruses of Respiratory Infections, Institut Pasteur, Paris Mélnie Albert, Marion Barbet, Sylvie Behillil, Méline Bizard, Angela Brisebarre, Flora Donati, Etienne Simon-Lorière, Vincent Enouf, Maud Vanpeene, Sylvie van der Werf
EPI_ISL_416494 hCoV-19/France/N2223/2020 Europe / France / Normandie / Rouen 2020-03-04 Centre Hositalier Universitaire de Rouen Laboratoire de Virologie National Reference Center for Viruses of Respiratory Infections, Institut Pasteur, Paris Mélnie Albert, Marion Barbet, Sylvie Behillil, Méline Bizard, Angela Brisebarre, Flora Donati, Etienne Simon-Lorière, Vincent Enouf, Maud Vanpeene, Sylvie van der Werf, Jean-Christophe Plantier
EPI_ISL_416495 hCoV-19/France/HF2234/2020 Europe / France / Hauts de France / Compiègne 2020-03-10 Centre Hospitalier Compiègne Laboratoire de Biologie National Reference Center for Viruses of Respiratory Infections, Institut Pasteur, Paris Mélnie Albert, Marion Barbet, Sylvie Behillil, Méline Bizard, Angela Brisebarre, Flora Donati, Etienne Simon-Lorière, Vincent Enouf, Maud Vanpeene, Sylvie van der Werf, Raulin Olivia
EPI_ISL_416496 hCoV-19/France/HF2237/2020 Europe / France / Hauts de France / Compiègne 2020-03-10 Centre Hospitalier Compiègne Laboratoire de Biologie National Reference Center for Viruses of Respiratory Infections, Institut Pasteur, Paris Mélnie Albert, Marion Barbet, Sylvie Behillil, Méline Bizard, Angela Brisebarre, Flora Donati, Etienne Simon-Lorière, Vincent Enouf, Maud Vanpeene, Sylvie van der Werf, Raulin Olivia
EPI_ISL_416497 hCoV-19/France/HF2239/2020 Europe / France / Hauts de France / Compiègne 2020-03-10 Centre Hospitalier Compiègne Laboratoire de Biologie National Reference Center for Viruses of Respiratory Infections, Institut Pasteur, Paris Mélnie Albert, Marion Barbet, Sylvie Behillil, Méline Bizard, Angela Brisebarre, Flora Donati, Etienne Simon-Lorière, Vincent Enouf, Maud Vanpeene, Sylvie van der Werf, Raulin Olivia
EPI_ISL_416498 hCoV-19/France/IDF2256/2020 Europe / France / Ile de France / Garches 2020-03-11 Institut Médico légal- Hop R. Poincaré National Reference Center for Viruses of Respiratory Infections, Institut Pasteur, Paris Mélnie Albert, Marion Barbet, Sylvie Behillil, Méline Bizard, Angela Brisebarre, Flora Donati, Etienne Simon-Lorière, Vincent Enouf, Maud Vanpeene, Sylvie van der Werf
EPI_ISL_416499 hCoV-19/France/IDF2278/2020 Europe / France / Ile de France / Longjumeau 2020-03-11

LABM GH nord Essonne	National Reference Center for Viruses of Respiratory Infections, Institut Pasteur, Paris	Mélnie Albert, Marion Barbet, Sylvie Behillil, Méline Bizard, Angela Brisebarre, Flora Donati, Etienne Simon-Lorière, Vincent Enouf, Maud Vanpeene, Sylvie van der Werf
EPI_ISL_416700	hCoV-19/USA/WA-UW162/2020	North America / USA / Washington	2020-03-13	UW Virology Lab	UW Virology Lab	Pavitra Roychoudhury, Hong Xie, Keith Jerome, Alexander Greninger
EPI_ISL_416701	hCoV-19/USA/WA-UW163/2020	North America / USA / Washington	2020-03-13	UW Virology Lab	UW Virology Lab	Pavitra Roychoudhury, Hong Xie, Keith Jerome, Alexander Greninger
EPI_ISL_416702	hCoV-19/USA/WA-UW164/2020	North America / USA / Washington	2020-03-13	UW Virology Lab	UW Virology Lab	Pavitra Roychoudhury, Hong Xie, Keith Jerome, Alexander Greninger
EPI_ISL_416703	hCoV-19/USA/WA-UW165/2020	North America / USA / Washington	2020-03-13	UW Virology Lab	UW Virology Lab	Pavitra Roychoudhury, Hong Xie, Keith Jerome, Alexander Greninger
EPI_ISL_416705	hCoV-19/USA/WA-UW167/2020	North America / USA / Washington	2020-03-13	UW Virology Lab	UW Virology Lab	Pavitra Roychoudhury, Hong Xie, Keith Jerome, Alexander Greninger
EPI_ISL_416706	hCoV-19/USA/WA-UW168/2020	North America / USA / Washington	2020-03-13	UW Virology Lab	UW Virology Lab	Pavitra Roychoudhury, Hong Xie, Keith Jerome, Alexander Greninger
EPI_ISL_416707	hCoV-19/USA/WA-UW169/2020	North America / USA / Washington	2020-03-14	UW Virology Lab	UW Virology Lab	Pavitra Roychoudhury, Hong Xie, Keith Jerome, Alexander Greninger
EPI_ISL_416708	hCoV-19/USA/WA-UW170/2020	North America / USA / Washington	2020-03-14	UW Virology Lab	UW Virology Lab	Pavitra Roychoudhury, Hong Xie, Keith Jerome, Alexander Greninger
EPI_ISL_416709	hCoV-19/USA/WA-UW171/2020	North America / USA / Washington	2020-03-13	UW Virology Lab	UW Virology Lab	Pavitra Roychoudhury, Hong Xie, Keith Jerome, Alexander Greninger
EPI_ISL_416710	hCoV-19/USA/WA-UW172/2020	North America / USA / Washington	2020-03-13	UW Virology Lab	UW Virology Lab	Pavitra Roychoudhury, Hong Xie, Keith Jerome, Alexander Greninger
EPI_ISL_416711	hCoV-19/USA/WA-UW173/2020	North America / USA / Washington	2020-03-15	UW Virology Lab	UW Virology Lab	Pavitra Roychoudhury, Hong Xie, Keith Jerome, Alexander Greninger
EPI_ISL_416712	hCoV-19/USA/WA-UW174/2020	North America / USA / Washington	2020-03-14	UW Virology Lab	UW Virology Lab	Pavitra Roychoudhury, Hong Xie, Keith Jerome, Alexander Greninger
EPI_ISL_416713	hCoV-19/USA/WA-UW175/2020	North America / USA / Washington	2020-03-15	UW Virology Lab	UW Virology Lab	Pavitra Roychoudhury, Hong Xie, Keith Jerome, Alexander Greninger
EPI_ISL_416714	hCoV-19/USA/WA-UW176/2020	North America / USA / Washington	2020-03-14	UW Virology Lab	UW Virology Lab	Pavitra Roychoudhury, Hong Xie, Keith Jerome, Alexander Greninger
EPI_ISL_416715	hCoV-19/USA/WA-UW177/2020	North America / USA / Washington	2020-03-15	UW Virology Lab	UW Virology Lab	Pavitra Roychoudhury, Hong Xie, Keith Jerome, Alexander Greninger
EPI_ISL_416716	hCoV-19/USA/WA-UW178/2020	North America / USA / Washington	2020-03-13	UW Virology Lab	UW Virology Lab	Pavitra Roychoudhury, Hong Xie, Keith Jerome, Alexander Greninger
EPI_ISL_416717	hCoV-19/USA/WA-UW179/2020	North America / USA / Washington	2020-03-15	UW Virology Lab	UW Virology Lab	Pavitra Roychoudhury, Hong Xie, Keith Jerome, Alexander Greninger
EPI_ISL_416718	hCoV-19/USA/WA-UW180/2020	North America / USA / Washington	2020-03-14	UW Virology Lab	UW Virology Lab	Pavitra Roychoudhury, Hong Xie, Keith Jerome, Alexander Greninger
EPI_ISL_416719	hCoV-19/USA/WA-UW181/2020	North America / USA / Washington	2020-03-14	UW Virology Lab	UW Virology Lab	Pavitra Roychoudhury, Hong Xie, Keith Jerome, Alexander Greninger
EPI_ISL_416720	hCoV-19/USA/WA-UW182/2020	North America / USA / Washington	2020-03-13	UW Virology Lab	UW Virology Lab	Pavitra Roychoudhury, Hong Xie, Keith Jerome, Alexander Greninger
EPI_ISL_416721	hCoV-19/USA/WA-UW183/2020	North America / USA / Washington	2020-03-13	UW Virology Lab	UW Virology Lab	Pavitra Roychoudhury, Hong Xie, Keith Jerome, Alexander Greninger
EPI_ISL_416722	hCoV-19/USA/WA-UW184/2020	North America / USA / Washington	2020-03-12	UW Virology Lab	UW Virology Lab	Pavitra Roychoudhury, Hong Xie, Keith Jerome, Alexander Greninger
EPI_ISL_416723	hCoV-19/USA/WA-UW185/2020	North America / USA / Washington	2020-03-14	UW Virology Lab	UW Virology Lab	Pavitra Roychoudhury, Hong Xie, Keith Jerome, Alexander Greninger
EPI_ISL_416724	hCoV-19/USA/WA-UW186/2020	North America / USA / Washington	2020-03-13	UW Virology Lab	UW Virology Lab	Pavitra Roychoudhury, Hong Xie, Keith Jerome, Alexander Greninger
EPI_ISL_416725	hCoV-19/USA/WA-UW187/2020	North America / USA / Washington	2020-03-13	UW Virology Lab	UW Virology Lab	Pavitra Roychoudhury, Hong Xie, Keith Jerome, Alexander Greninger
EPI_ISL_416726	hCoV-19/USA/WA-UW188/2020	North America / USA / Washington	2020-03-13	UW Virology Lab	UW Virology Lab	Pavitra Roychoudhury, Hong Xie, Keith Jerome, Alexander Greninger
EPI_ISL_416727	hCoV-19/USA/WA-UW189/2020	North America / USA / Washington	2020-03-13	UW Virology Lab	UW Virology Lab	Pavitra Roychoudhury, Hong Xie, Keith Jerome, Alexander Greninger
EPI_ISL_416728	hCoV-19/USA/WA-UW190/2020	North America / USA / Washington	2020-03-13	UW Virology Lab	UW Virology Lab	Pavitra Roychoudhury, Hong Xie, Keith Jerome, Alexander Greninger
EPI_ISL_416729	hCoV-19/USA/WA-UW191/2020	North America / USA / Washington	2020-03-13	UW Virology Lab	UW Virology Lab	Pavitra Roychoudhury, Hong Xie, Keith Jerome, Alexander Greninger
EPI_ISL_416730	hCoV-19/England/SHEF-BFCB1/2020	Europe / United Kingdom / England / South Yorkshire	2020-03-03	Virology Department, Sheffield Teaching Hospitals NHS Foundation Trust	Department of Infection, Immunity and Cardiovascular Disease, The Florey Institute, The Medical School, University of Sheffield	Thushan de Silva, Matthew Parker, Adri Angyal, Rebecca Brown, Matthew Wyles, Mehmet Yavuz, Mohammad Raza, Cariad Evans
EPI_ISL_416731	hCoV-19/England/SHEF-BFCC0/2020	Europe / United Kingdom / England / Northamtonshire	2020-03-03	Virology Department, Sheffield Teaching Hospitals NHS Foundation Trust	Department of Infection, Immunity and Cardiovascular Disease, The Florey Institute, The Medical School, University of Sheffield	Thushan de Silva, Matthew Parker, Adri Angyal, Rebecca Brown, Matthew Wyles, Mehmet Yavuz, Mohammad Raza, Cariad Evans
EPI_ISL_416732	hCoV-19/England/SHEF-BFCDF/2020	Europe / United Kingdom / England / London	2020-03-03

Virology Department, Sheffield Teaching Hospitals NHS Foundation Trust  Department of Infection, Immunity and Cardiovascular Disease, The Florey Institute, The Medical School, University of Sheffield  Thushan de Silva, Matthew Parker, Adri Angyal, Rebecca Brown, Matthew Wyles, Mehmet Yavuz, Mohammad Raza, Cariad Evans
EPI_ISL_416733  hCoV-19/England/SHEF-BFCEE/2020 Europe / United Kingdom / England / South Yorkshire  2020-03-07  Virology Department, Sheffield Teaching Hospitals NHS Foundation Trust  Department of Infection, Immunity and Cardiovascular Disease, The Florey Institute, The Medical School, University of Sheffield  Thushan de Silva, Matthew Parker, Adri Angyal, Rebecca Brown, Matthew Wyles, Mehmet Yavuz, Mohammad Raza, Cariad Evans
EPI_ISL_416734  hCoV-19/England/SHEF-BFCFD/2020 Europe / United Kingdom / England / South Yorkshire  2020-03-09  Virology Department, Sheffield Teaching Hospitals NHS Foundation Trust  Department of Infection, Immunity and Cardiovascular Disease, The Florey Institute, The Medical School, University of Sheffield  Thushan de Silva, Matthew Parker, Adri Angyal, Rebecca Brown, Matthew Wyles, Mehmet Yavuz, Mohammad Raza, Cariad Evans
EPI_ISL_416739  hCoV-19/England/SHEF-BFD45/2020 Europe / United Kingdom / England / South Yorkshire  2020-03-09  Virology Department, Sheffield Teaching Hospitals NHS Foundation Trust  Department of Infection, Immunity and Cardiovascular Disease, The Florey Institute, The Medical School, University of Sheffield  Thushan de Silva, Matthew Parker, Adri Angyal, Rebecca Brown, Matthew Wyles, Mehmet Yavuz, Mohammad Raza, Cariad Evans
EPI_ISL_416740  hCoV-19/England/SHEF-BFD54/2020 Europe / United Kingdom / England / South Yorkshire  2020-03-03  Virology Department, Sheffield Teaching Hospitals NHS Foundation Trust  Department of Infection, Immunity and Cardiovascular Disease, The Florey Institute, The Medical School, University of Sheffield  Thushan de Silva, Matthew Parker, Adri Angyal, Rebecca Brown, Matthew Wyles, Mehmet Yavuz, Mohammad Raza, Cariad Evans
EPI_ISL_416741  hCoV-19/Lithuania/ChVir1632/2020  Europe / Lithuania / Vilnius  2020-02 National Public Health Surveillance Laboratory, Vilnius, Lithuania  Charite Universitaetsmedizin Berlin, Institute of Virology  Victor M Corman, Julia Schneider, Jörn Beheim-Schwarzbach, Talitha Veith, Barbara Muehlemann, Terry Jones, Ana Steponkiene, Christian Drosten
EPI_ISL_416742  hCoV-19/Czech Republic/ChVir1630/2020  Europe / Czech Republic / Prague  2020-02-29  NRL for Influenza, Centrum Epidemiology and Microbiology of National Institute of Public Health, Czech Republic  Charite Universitaetsmedizin Berlin, Institute of Virology  Victor M Corman, Julia Schneider, Jörn Beheim-Schwarzbach, Talitha Veith, Barbara Muehlemann, Terry Jones, Akexander Nagy, Jaromira Vecerova, Dusan Trnka, Ludmila Novakova, Helena Jirincova, Christian Drosten
EPI_ISL_416743  hCoV-19/Czech Republic/ChVir1912/2020  Europe / Czech Republic / Prague  2020-03-05  NRL for Influenza, Centrum Epidemiology and Microbiology of National Institute of Public Health, Czech Republic  Charite Universitaetsmedizin Berlin, Institute of Virology  Victor M Corman, Julia Schneider, Jörn Beheim-Schwarzbach, Talitha Veith, Barbara Muehlemann, Terry Jones, Akexander Nagy, Jaromira Vecerova, Dusan Trnka, Ludmila Novakova, Helena Jirincova, Christian Drosten
EPI_ISL_416744  hCoV-19/Hungary/mbl49/2020  Europe / Hungary / Baranya  2020-03-20  Virological Research Group, Szentágothai Research Centre  Bioinformatics Research Group, Szentágothai Research Centre  Péter Urbán, Endre Gábor Tóth, Gábor Kemenesi, Róbert Herczeg, Attila Gyenesei, Ferenc Jakab
EPI_ISL_416745  hCoV-19/France/Pollionay_1733/2020  Europe / France / ARA  2020-03-10  CNR Virus des Infections Respiratoires - France SUD  CNR Virus des Infections Respiratoires - France SUD  Bal, Antonin; Destras, Gregory; Gaymard, Alexandre; Bouscambert-Duchamp, Maude; Cheynet, Valérie; Brengel-Pesce, Karen; Morfin-Sherpa, Florence; Valette, Martine; Josset, Laurence; Lina, Bruno.
EPI_ISL_416746  hCoV-19/France/Valence_425/2020 Europe / France / ARA  2020-03-03  CNR Virus des Infections Respiratoires - France SUD  CNR Virus des Infections Respiratoires - France SUD  Bal, Antonin; Destras, Gregory; Gaymard, Alexandre; Bouscambert-Duchamp, Maude; Cheynet, Valérie; Brengel-Pesce, Karen; Morfin-Sherpa, Florence; Valette, Martine; Josset, Laurence; Lina, Bruno.
EPI_ISL_416747  hCoV-19/France/Lyon_487/2020  Europe / France / ARA  2020-03-04  Institut des Agents Infectieux (IAI) Hospices Civils de Lyon  CNR Virus des Infections Respiratoires - France SUD  Bal, Antonin; Destras, Gregory; Gaymard, Alexandre; Bouscambert-Duchamp, Maude; Cheynet, Valérie; Brengel-Pesce, Karen; Morfin-Sherpa, Florence; Valette, Martine; Josset, Laurence; Lina, Bruno.
EPI_ISL_416748  hCoV-19/France/Lyon_508/2020  Europe / France / ARA  2020-03-04  Institut des Agents Infectieux (IAI) Hospices Civils de Lyon  CNR Virus des Infections Respiratoires - France SUD  Bal, Antonin; Destras, Gregory; Gaymard, Alexandre; Bouscambert-Duchamp, Maude; Cheynet, Valérie; Brengel-Pesce, Karen; Morfin-Sherpa, Florence; Valette, Martine; Josset, Laurence; Lina, Bruno.
EPI_ISL_416749  hCoV-19/France/Valence_532/2020 Europe / France / ARA  2020-03-04  Centre Hospitalier de Valence CNR Virus des Infections Respiratoires - France SUD  Bal, Antonin; Destras, Gregory; Gaymard, Alexandre; Bouscambert-Duchamp, Maude; Cheynet, Valérie; Brengel-Pesce, Karen; Morfin-Sherpa, Florence; Valette, Martine; Josset, Laurence; Lina, Bruno.
EPI_ISL_416750  hCoV-19/France/Lyon_683/2020  Europe / France / ARA  2020-03-06  Institut des Agents Infectieux (IAI) Hospices Civils de Lyon  CNR Virus des Infections Respiratoires - France SUD  Bal, Antonin; Destras, Gregory; Gaymard, Alexandre; Bouscambert-Duchamp, Maude; Cheynet, Valérie; Brengel-Pesce, Karen; Morfin-Sherpa, Florence; Valette, Martine; Josset, Laurence; Lina, Bruno.
EPI_ISL_416751  hCoV-19/France/Clermont-Ferrand_651/2020  Europe / France / ARA  2020-03-05  CHU Gabriel Montpied  CNR Virus des Infections Respiratoires - France SUD  Bal, Antonin; Destras, Gregory; Gaymard, Alexandre; Bouscambert-Duchamp, Maude; Cheynet, Valérie; Brengel-Pesce, Karen; Morfin-Sherpa, Florence; Valette, Martine; Josset, Laurence; Lina, Bruno.
EPI_ISL_416752  hCoV-19/France/Clermont-Ferrand_650/2020  Europe / France / ARA  2020-03-04  CHU Gabriel Montpied  CNR Virus des Infections Respiratoires - France SUD  Bal, Antonin; Destras, Gregory; Gaymard, Alexandre; Bouscambert-Duchamp, Maude; Cheynet, Valérie; Brengel-Pesce, Karen; Morfin-Sherpa, Florence; Valette, Martine; Josset, Laurence; Lina, Bruno.
EPI_ISL_416753  hCoV-19/France/Lyon_06464 /2020 Europe / France / ARA  2020-03-06  Institut des Agents Infectieux (IAI) Hospices Civils de Lyon  CNR Virus des Infections Respiratoires - France SUD  Bal,

Antonin; Destras, Gregory; Gaymard, Alexandre; Bouscambert-Duchamp, Maude; Cheynet, Valérie; Brengel-Pesce, Karen; Morfin-Sherpa, Florence; Valette, Martine; Josset, Laurence; Lina, Bruno.
EPI_ISL_416754 hCoV-19/France/Lyon_06487/2020 Europe / France / ARA 2020-03-06 Institut des Agents Infectieux (IAI) Hospices Civils de Lyon CNR Virus des Infections Respiratoires - France SUD Bal, Antonin; Destras, Gregory; Gaymard, Alexandre; Bouscambert-Duchamp, Maude; Cheynet, Valérie; Brengel-Pesce, Karen; Morfin-Sherpa, Florence; Valette, Martine; Josset, Laurence; Lina, Bruno.
EPI_ISL_416756 hCoV-19/France/Lyon_06531/2020 Europe / France / ARA 2020-03-06 Institut des Agents Infectieux (IAI) Hospices Civils de Lyon CNR Virus des Infections Respiratoires - France SUD Bal, Antonin; Destras, Gregory; Gaymard, Alexandre; Bouscambert-Duchamp, Maude; Cheynet, Valérie; Brengel-Pesce, Karen; Morfin-Sherpa, Florence; Valette, Martine; Josset, Laurence; Lina, Bruno.
EPI_ISL_416757 hCoV-19/France/Bourg-en-Bresse_06678/2020 Europe / France / ARA 2020-03-07 Centre Hospitalier de Bourg en Bresse CNR Virus des Infections Respiratoires - France SUD Bal, Antonin; Destras, Gregory; Gaymard, Alexandre; Bouscambert-Duchamp, Maude; Cheynet, Valérie; Brengel-Pesce, Karen; Morfin-Sherpa, Florence; Valette, Martine; Josset, Laurence; Lina, Bruno.
EPI_ISL_416758 hCoV-19/France/Lyon_0693/2020 Europe / France / ARA 2020-03-08 Institut des Agents Infectieux (IAI) Hospices Civils de Lyon CNR Virus des Infections Respiratoires - France SUD Bal, Antonin; Destras, Gregory; Gaymard, Alexandre; Bouscambert-Duchamp, Maude; Cheynet, Valérie; Brengel-Pesce, Karen; Morfin-Sherpa, Florence; Valette, Martine; Josset, Laurence; Lina, Bruno.
EPI_ISL_417700 hCoV-19/Iceland/173/2020 Europe / Iceland / Reykjavik 2020-03-15 The National University Hospital of Iceland deCODE genetics Daniel F Gudbjartsson; Agnar Helgason; Hakon Jonsson; Olafur T Magnusson; Pall Melsted; Gudmundur L Norddahl; Jona Saemundsdottir; Asgeir Sigurdsson; Patrick Sulem; Arna B Agustsdottir; Berglind Eiriksdottir; Run Fridriksdottir; Elisabet E Gardarsdottir; Gudmundur Georgsson; Olafia S Gretarsdottir; Kjartan R Gudmundsson; Thora R Gunnarsdottir; Arnaldur Gylfason; Hilma Holm; Brynjar O Jensson; Aslaug Jonasdottir; Kamilla S Josefsdottir; Thordur Kristjansson; Droplaug N Magnusdottir; Louise le Roux; Gudrun Sigmundsdottir; Gardar Sveinbjornsson; Kristin E Sveinsdottir; Maney Sveinsdottir; Emil A Thorarensen; Bjarni Thorbjornsson; Gisli Masson; Ingileif Jonsdottir; Alma Moller; Thorolfur Gudnason; Karl G Kristinsson; Unnur Thorsteinsdottir; Kari Stefansson
EPI_ISL_417701 hCoV-19/Iceland/74/2020 Europe / Iceland / Reykjavik 2020-03-09 The National University Hospital of Iceland deCODE genetics Daniel F Gudbjartsson; Agnar Helgason; Hakon Jonsson; Olafur T Magnusson; Pall Melsted; Gudmundur L Norddahl; Jona Saemundsdottir; Asgeir Sigurdsson; Patrick Sulem; Arna B Agustsdottir; Berglind Eiriksdottir; Run Fridriksdottir; Elisabet E Gardarsdottir; Gudmundur Georgsson; Olafia S Gretarsdottir; Kjartan R Gudmundsson; Thora R Gunnarsdottir; Arnaldur Gylfason; Hilma Holm; Brynjar O Jensson; Aslaug Jonasdottir; Kamilla S Josefsdottir; Thordur Kristjansson; Droplaug N Magnusdottir; Louise le Roux; Gudrun Sigmundsdottir; Gardar Sveinbjornsson; Kristin E Sveinsdottir; Maney Sveinsdottir; Emil A Thorarensen; Bjarni Thorbjornsson; Gisli Masson; Ingileif Jonsdottir; Alma Moller; Thorolfur Gudnason; Karl G Kristinsson; Unnur Thorsteinsdottir; Kari Stefansson
EPI_ISL_417702 hCoV-19/Iceland/33/2020 Europe / Iceland / Reykjavik 2020-03-03 The National University Hospital of Iceland deCODE genetics Daniel F Gudbjartsson; Agnar Helgason; Hakon Jonsson; Olafur T Magnusson; Pall Melsted; Gudmundur L Norddahl; Jona Saemundsdottir; Asgeir Sigurdsson; Patrick Sulem; Arna B Agustsdottir; Berglind Eiriksdottir; Run Fridriksdottir; Elisabet E Gardarsdottir; Gudmundur Georgsson; Olafia S Gretarsdottir; Kjartan R Gudmundsson; Thora R Gunnarsdottir; Arnaldur Gylfason; Hilma Holm; Brynjar O Jensson; Aslaug Jonasdottir; Kamilla S Josefsdottir; Thordur Kristjansson; Droplaug N Magnusdottir; Louise le Roux; Gudrun Sigmundsdottir; Gardar Sveinbjornsson; Kristin E Sveinsdottir; Maney Sveinsdottir; Emil A Thorarensen; Bjarni Thorbjornsson; Gisli Masson; Ingileif Jonsdottir; Alma Moller; Thorolfur Gudnason; Karl G Kristinsson; Unnur Thorsteinsdottir; Kari Stefansson
EPI_ISL_417703 hCoV-19/Iceland/179/2020 Europe / Iceland / Reykjavik 2020-03-16 The National University Hospital of Iceland deCODE genetics Daniel F Gudbjartsson; Agnar Helgason; Hakon Jonsson; Olafur T Magnusson; Pall Melsted; Gudmundur L Norddahl; Jona Saemundsdottir; Asgeir Sigurdsson; Patrick Sulem; Arna B Agustsdottir; Berglind Eiriksdottir; Run Fridriksdottir; Elisabet E Gardarsdottir; Gudmundur Georgsson; Olafia S Gretarsdottir; Kjartan R Gudmundsson; Thora R Gunnarsdottir; Arnaldur Gylfason; Hilma Holm; Brynjar O Jensson; Aslaug Jonasdottir; Kamilla S Josefsdottir; Thordur Kristjansson; Droplaug N Magnusdottir; Louise le Roux; Gudrun Sigmundsdottir; Gardar Sveinbjornsson; Kristin E Sveinsdottir; Maney Sveinsdottir; Emil A Thorarensen; Bjarni Thorbjornsson; Gisli Masson; Ingileif Jonsdottir; Alma Moller; Thorolfur Gudnason; Karl G Kristinsson; Unnur Thorsteinsdottir; Kari Stefansson
EPI_ISL_417705 hCoV-19/Iceland/65/2020 Europe / Iceland / Reykjavik 2020-03-07 The National University Hospital of Iceland deCODE genetics Daniel F Gudbjartsson; Agnar Helgason; Hakon Jonsson; Olafur T Magnusson; Pall Melsted; Gudmundur L Norddahl; Jona Saemundsdottir; Asgeir Sigurdsson; Patrick Sulem; Arna B Agustsdottir; Berglind Eiriksdottir; Run Fridriksdottir; Elisabet E Gardarsdottir; Gudmundur Georgsson; Olafia S Gretarsdottir; Kjartan R Gudmundsson; Thora R Gunnarsdottir; Arnaldur Gylfason; Hilma Holm; Brynjar O Jensson; Aslaug Jonasdottir; Kamilla S Josefsdottir; Thordur Kristjansson; Droplaug N Magnusdottir; Louise le Roux; Gudrun Sigmundsdottir; Gardar Sveinbjornsson; Kristin E Sveinsdottir; Maney Sveinsdottir; Emil A Thorarensen; Bjarni Thorbjornsson; Gisli Masson; Ingileif Jonsdottir; Alma Moller; Thorolfur Gudnason; Karl G Kristinsson; Unnur Thorsteinsdottir; Kari Stefansson
EPI_ISL_417706 hCoV-19/Iceland/199/2020 Europe / Iceland / Reykjavik 2020-03-16 The National University Hospital of Iceland deCODE genetics Daniel F Gudbjartsson; Agnar Helgason; Hakon Jonsson; Olafur T Magnusson; Pall Melsted; Gudmundur L Norddahl; Jona Saemundsdottir; Asgeir Sigurdsson; Patrick Sulem; Arna B Agustsdottir; Berglind Eiriksdottir; Run Fridriksdottir; Elisabet E Gardarsdottir; Gudmundur Georgsson; Olafia S Gretarsdottir; Kjartan R Gudmundsson; Thora R Gunnarsdottir; Arnaldur Gylfason; Hilma Holm; Brynjar O Jensson; Aslaug Jonasdottir; Kamilla S Josefsdottir; Thordur Kristjansson; Droplaug N Magnusdottir; Louise le Roux; Gudrun Sigmundsdottir; Gardar Sveinbjornsson; Kristin E Sveinsdottir; Maney Sveinsdottir; Emil A

Thorarensen; Bjarni Thorbjornsson; Gisli Masson; Ingileif Jonsdottir; Alma Moller; Thorolfur Gudnason; Karl G Kristinsson; Unnur Thorsteinsdottir; Kari Stefansson
EPI_ISL_417709 hCoV-19/Iceland/189/2020 Europe / Iceland / Reykjavik 2020-03-16 The National University Hospital of Iceland deCODE genetics Daniel F Gudbjartsson; Agnar Helgason; Hakon Jonsson; Olafur T Magnusson; Pall Melsted; Gudmundur L Norddahl; Jona Saemundsdottir; Asgeir Sigurdsson; Patrick Sulem; Arna B Agustsdottir; Berglind Eiriksdottir; Run Fridriksdottir; Elisabet E Gardarsdottir; Gudmundur Georgsson; Olafia S Gretarsdottir; Kjartan R Gudmundsson; Thora R Gunnarsdottir; Arnaldur Gylfason; Hilma Holm; Brynjar O Jensson; Aslaug Jonasdottir; Kamilla S Josefsdottir; Thordur Kristjansson; Droplaug N Magnusdottir; Louise le Roux; Gudrun Sigmundsdottir; Gardar Sveinbjornsson; Kristin E Sveinsdottir; Maney Sveinsdottir; Emil A Thorarensen; Bjarni Thorbjornsson; Gisli Masson; Ingileif Jonsdottir; Alma Moller; Thorolfur Gudnason; Karl G Kristinsson; Unnur Thorsteinsdottir; Kari Stefansson
EPI_ISL_417711 hCoV-19/Iceland/150/2020 Europe / Iceland / Reykjavik 2020-03-13 The National University Hospital of Iceland deCODE genetics Daniel F Gudbjartsson; Agnar Helgason; Hakon Jonsson; Olafur T Magnusson; Pall Melsted; Gudmundur L Norddahl; Jona Saemundsdottir; Asgeir Sigurdsson; Patrick Sulem; Arna B Agustsdottir; Berglind Eiriksdottir; Run Fridriksdottir; Elisabet E Gardarsdottir; Gudmundur Georgsson; Olafia S Gretarsdottir; Kjartan R Gudmundsson; Thora R Gunnarsdottir; Arnaldur Gylfason; Hilma Holm; Brynjar O Jensson; Aslaug Jonasdottir; Kamilla S Josefsdottir; Thordur Kristjansson; Droplaug N Magnusdottir; Louise le Roux; Gudrun Sigmundsdottir; Gardar Sveinbjornsson; Kristin E Sveinsdottir; Maney Sveinsdottir; Emil A Thorarensen; Bjarni Thorbjornsson; Gisli Masson; Ingileif Jonsdottir; Alma Moller; Thorolfur Gudnason; Karl G Kristinsson; Unnur Thorsteinsdottir; Kari Stefansson
EPI_ISL_417712 hCoV-19/Iceland/172/2020 Europe / Iceland / Reykjavik 2020-03-15 The National University Hospital of Iceland deCODE genetics Daniel F Gudbjartsson; Agnar Helgason; Hakon Jonsson; Olafur T Magnusson; Pall Melsted; Gudmundur L Norddahl; Jona Saemundsdottir; Asgeir Sigurdsson; Patrick Sulem; Arna B Agustsdottir; Berglind Eiriksdottir; Run Fridriksdottir; Elisabet E Gardarsdottir; Gudmundur Georgsson; Olafia S Gretarsdottir; Kjartan R Gudmundsson; Thora R Gunnarsdottir; Arnaldur Gylfason; Hilma Holm; Brynjar O Jensson; Aslaug Jonasdottir; Kamilla S Josefsdottir; Thordur Kristjansson; Droplaug N Magnusdottir; Louise le Roux; Gudrun Sigmundsdottir; Gardar Sveinbjornsson; Kristin E Sveinsdottir; Maney Sveinsdottir; Emil A Thorarensen; Bjarni Thorbjornsson; Gisli Masson; Ingileif Jonsdottir; Alma Moller; Thorolfur Gudnason; Karl G Kristinsson; Unnur Thorsteinsdottir; Kari Stefansson
EPI_ISL_417713 hCoV-19/Iceland/130/2020 Europe / Iceland / Reykjavik 2020-03-12 The National University Hospital of Iceland deCODE genetics Daniel F Gudbjartsson; Agnar Helgason; Hakon Jonsson; Olafur T Magnusson; Pall Melsted; Gudmundur L Norddahl; Jona Saemundsdottir; Asgeir Sigurdsson; Patrick Sulem; Arna B Agustsdottir; Berglind Eiriksdottir; Run Fridriksdottir; Elisabet E Gardarsdottir; Gudmundur Georgsson; Olafia S Gretarsdottir; Kjartan R Gudmundsson; Thora R Gunnarsdottir; Arnaldur Gylfason; Hilma Holm; Brynjar O Jensson; Aslaug Jonasdottir; Kamilla S Josefsdottir; Thordur Kristjansson; Droplaug N Magnusdottir; Louise le Roux; Gudrun Sigmundsdottir; Gardar Sveinbjornsson; Kristin E Sveinsdottir; Maney Sveinsdottir; Emil A Thorarensen; Bjarni Thorbjornsson; Gisli Masson; Ingileif Jonsdottir; Alma Moller; Thorolfur Gudnason; Karl G Kristinsson; Unnur Thorsteinsdottir; Kari Stefansson
EPI_ISL_417714 hCoV-19/Iceland/129/2020 Europe / Iceland / Reykjavik 2020-03-12 The National University Hospital of Iceland deCODE genetics Daniel F Gudbjartsson; Agnar Helgason; Hakon Jonsson; Olafur T Magnusson; Pall Melsted; Gudmundur L Norddahl; Jona Saemundsdottir; Asgeir Sigurdsson; Patrick Sulem; Arna B Agustsdottir; Berglind Eiriksdottir; Run Fridriksdottir; Elisabet E Gardarsdottir; Gudmundur Georgsson; Olafia S Gretarsdottir; Kjartan R Gudmundsson; Thora R Gunnarsdottir; Arnaldur Gylfason; Hilma Holm; Brynjar O Jensson; Aslaug Jonasdottir; Kamilla S Josefsdottir; Thordur Kristjansson; Droplaug N Magnusdottir; Louise le Roux; Gudrun Sigmundsdottir; Gardar Sveinbjornsson; Kristin E Sveinsdottir; Maney Sveinsdottir; Emil A Thorarensen; Bjarni Thorbjornsson; Gisli Masson; Ingileif Jonsdottir; Alma Moller; Thorolfur Gudnason; Karl G Kristinsson; Unnur Thorsteinsdottir; Kari Stefansson
EPI_ISL_417715 hCoV-19/Iceland/45/2020 Europe / Iceland / Reykjavik 2020-03-04 The National University Hospital of Iceland deCODE genetics Daniel F Gudbjartsson; Agnar Helgason; Hakon Jonsson; Olafur T Magnusson; Pall Melsted; Gudmundur L Norddahl; Jona Saemundsdottir; Asgeir Sigurdsson; Patrick Sulem; Arna B Agustsdottir; Berglind Eiriksdottir; Run Fridriksdottir; Elisabet E Gardarsdottir; Gudmundur Georgsson; Olafia S Gretarsdottir; Kjartan R Gudmundsson; Thora R Gunnarsdottir; Arnaldur Gylfason; Hilma Holm; Brynjar O Jensson; Aslaug Jonasdottir; Kamilla S Josefsdottir; Thordur Kristjansson; Droplaug N Magnusdottir; Louise le Roux; Gudrun Sigmundsdottir; Gardar Sveinbjornsson; Kristin E Sveinsdottir; Maney Sveinsdottir; Emil A Thorarensen; Bjarni Thorbjornsson; Gisli Masson; Ingileif Jonsdottir; Alma Moller; Thorolfur Gudnason; Karl G Kristinsson; Unnur Thorsteinsdottir; Kari Stefansson
EPI_ISL_417716 hCoV-19/Iceland/174/2020 Europe / Iceland / Reykjavik 2020-03-16 The National University Hospital of Iceland deCODE genetics Daniel F Gudbjartsson; Agnar Helgason; Hakon Jonsson; Olafur T Magnusson; Pall Melsted; Gudmundur L Norddahl; Jona Saemundsdottir; Asgeir Sigurdsson; Patrick Sulem; Arna B Agustsdottir; Berglind Eiriksdottir; Run Fridriksdottir; Elisabet E Gardarsdottir; Gudmundur Georgsson; Olafia S Gretarsdottir; Kjartan R Gudmundsson; Thora R Gunnarsdottir; Arnaldur Gylfason; Hilma Holm; Brynjar O Jensson; Aslaug Jonasdottir; Kamilla S Josefsdottir; Thordur Kristjansson; Droplaug N Magnusdottir; Louise le Roux; Gudrun Sigmundsdottir; Gardar Sveinbjornsson; Kristin E Sveinsdottir; Maney Sveinsdottir; Emil A Thorarensen; Bjarni Thorbjornsson; Gisli Masson; Ingileif Jonsdottir; Alma Moller; Thorolfur Gudnason; Karl G Kristinsson; Unnur Thorsteinsdottir; Kari Stefansson
EPI_ISL_417717 hCoV-19/Iceland/195/2020 Europe / Iceland / Reykjavik 2020-03-16 The National University Hospital of Iceland deCODE genetics Daniel F Gudbjartsson; Agnar Helgason; Hakon Jonsson; Olafur T Magnusson; Pall Melsted; Gudmundur L Norddahl; Jona Saemundsdottir; Asgeir Sigurdsson; Patrick Sulem; Arna B Agustsdottir; Berglind Eiriksdottir; Run Fridriksdottir; Elisabet E Gardarsdottir; Gudmundur Georgsson; Olafia S Gretarsdottir; Kjartan R Gudmundsson; Thora R Gunnarsdottir; Arnaldur Gylfason; Hilma Holm; Brynjar O

Jensson; Aslaug Jonasdottir; Kamilla S Josefsdottir; Thordur Kristjansson; Droplaug N Magnusdottir; Louise le Roux; Gudrun Sigmundsdottir; Gardar Sveinbjornsson; Kristin E Sveinsdottir; Maney Sveinsdottir; Emil A Thorarensen; Bjarni Thorbjornsson; Gisli Masson; Ingileif Jonsdottir; Alma Moller; Thorolfur Gudnason; Karl G Kristinsson; Unnur Thorsteinsdottir; Kari Stefansson
EPI_ISL_417720 hCoV-19/Iceland/138/2020 Europe / Iceland / Reykjavik 2020-03-12 The National University Hospital of Iceland deCODE genetics Daniel F Gudbjartsson; Agnar Helgason; Hakon Jonsson; Olafur T Magnusson; Pall Melsted; Gudmundur L Norddahl; Jona Saemundsdottir; Asgeir Sigurdsson; Patrick Sulem; Arna B Agustsdottir; Berglind Eiriksdottir; Run Fridriksdottir; Elisabet E Gardarsdottir; Gudmundur Georgsson; Olafia S Gretarsdottir; Kjartan R Gudmundsson; Thora R Gunnarsdottir; Arnaldur Gylfason; Hilma Holm; Brynjar O Jensson; Aslaug Jonasdottir; Kamilla S Josefsdottir; Thordur Kristjansson; Droplaug N Magnusdottir; Louise le Roux; Gudrun Sigmundsdottir; Gardar Sveinbjornsson; Kristin E Sveinsdottir; Maney Sveinsdottir; Emil A Thorarensen; Bjarni Thorbjornsson; Gisli Masson; Ingileif Jonsdottir; Alma Moller; Thorolfur Gudnason; Karl G Kristinsson; Unnur Thorsteinsdottir; Kari Stefansson
EPI_ISL_417721 hCoV-19/Iceland/27/2020 Europe / Iceland / Reykjavik 2020-03-03 The National University Hospital of Iceland deCODE genetics Daniel F Gudbjartsson; Agnar Helgason; Hakon Jonsson; Olafur T Magnusson; Pall Melsted; Gudmundur L Norddahl; Jona Saemundsdottir; Asgeir Sigurdsson; Patrick Sulem; Arna B Agustsdottir; Berglind Eiriksdottir; Run Fridriksdottir; Elisabet E Gardarsdottir; Gudmundur Georgsson; Olafia S Gretarsdottir; Kjartan R Gudmundsson; Thora R Gunnarsdottir; Arnaldur Gylfason; Hilma Holm; Brynjar O Jensson; Aslaug Jonasdottir; Kamilla S Josefsdottir; Thordur Kristjansson; Droplaug N Magnusdottir; Louise le Roux; Gudrun Sigmundsdottir; Gardar Sveinbjornsson; Kristin E Sveinsdottir; Maney Sveinsdottir; Emil A Thorarensen; Bjarni Thorbjornsson; Gisli Masson; Ingileif Jonsdottir; Alma Moller; Thorolfur Gudnason; Karl G Kristinsson; Unnur Thorsteinsdottir; Kari Stefansson
EPI_ISL_417722 hCoV-19/Iceland/110/2020 Europe / Iceland / Reykjavik 2020-03-11 The National University Hospital of Iceland deCODE genetics Daniel F Gudbjartsson; Agnar Helgason; Hakon Jonsson; Olafur T Magnusson; Pall Melsted; Gudmundur L Norddahl; Jona Saemundsdottir; Asgeir Sigurdsson; Patrick Sulem; Arna B Agustsdottir; Berglind Eiriksdottir; Run Fridriksdottir; Elisabet E Gardarsdottir; Gudmundur Georgsson; Olafia S Gretarsdottir; Kjartan R Gudmundsson; Thora R Gunnarsdottir; Arnaldur Gylfason; Hilma Holm; Brynjar O Jensson; Aslaug Jonasdottir; Kamilla S Josefsdottir; Thordur Kristjansson; Droplaug N Magnusdottir; Louise le Roux; Gudrun Sigmundsdottir; Gardar Sveinbjornsson; Kristin E Sveinsdottir; Maney Sveinsdottir; Emil A Thorarensen; Bjarni Thorbjornsson; Gisli Masson; Ingileif Jonsdottir; Alma Moller; Thorolfur Gudnason; Karl G Kristinsson; Unnur Thorsteinsdottir; Kari Stefansson
EPI_ISL_417724 hCoV-19/Iceland/182/2020 Europe / Iceland / Reykjavik 2020-03-16 The National University Hospital of Iceland deCODE genetics Daniel F Gudbjartsson; Agnar Helgason; Hakon Jonsson; Olafur T Magnusson; Pall Melsted; Gudmundur L Norddahl; Jona Saemundsdottir; Asgeir Sigurdsson; Patrick Sulem; Arna B Agustsdottir; Berglind Eiriksdottir; Run Fridriksdottir; Elisabet E Gardarsdottir; Gudmundur Georgsson; Olafia S Gretarsdottir; Kjartan R Gudmundsson; Thora R Gunnarsdottir; Arnaldur Gylfason; Hilma Holm; Brynjar O Jensson; Aslaug Jonasdottir; Kamilla S Josefsdottir; Thordur Kristjansson; Droplaug N Magnusdottir; Louise le Roux; Gudrun Sigmundsdottir; Gardar Sveinbjornsson; Kristin E Sveinsdottir; Maney Sveinsdottir; Emil A Thorarensen; Bjarni Thorbjornsson; Gisli Masson; Ingileif Jonsdottir; Alma Moller; Thorolfur Gudnason; Karl G Kristinsson; Unnur Thorsteinsdottir; Kari Stefansson
EPI_ISL_417725 hCoV-19/Iceland/161/2020 Europe / Iceland / Reykjavik 2020-03-14 The National University Hospital of Iceland deCODE genetics Daniel F Gudbjartsson; Agnar Helgason; Hakon Jonsson; Olafur T Magnusson; Pall Melsted; Gudmundur L Norddahl; Jona Saemundsdottir; Asgeir Sigurdsson; Patrick Sulem; Arna B Agustsdottir; Berglind Eiriksdottir; Run Fridriksdottir; Elisabet E Gardarsdottir; Gudmundur Georgsson; Olafia S Gretarsdottir; Kjartan R Gudmundsson; Thora R Gunnarsdottir; Arnaldur Gylfason; Hilma Holm; Brynjar O Jensson; Aslaug Jonasdottir; Kamilla S Josefsdottir; Thordur Kristjansson; Droplaug N Magnusdottir; Louise le Roux; Gudrun Sigmundsdottir; Gardar Sveinbjornsson; Kristin E Sveinsdottir; Maney Sveinsdottir; Emil A Thorarensen; Bjarni Thorbjornsson; Gisli Masson; Ingileif Jonsdottir; Alma Moller; Thorolfur Gudnason; Karl G Kristinsson; Unnur Thorsteinsdottir; Kari Stefansson
EPI_ISL_417727 hCoV-19/Iceland/127/2020 Europe / Iceland / Reykjavik 2020-03-12 The National University Hospital of Iceland deCODE genetics Daniel F Gudbjartsson; Agnar Helgason; Hakon Jonsson; Olafur T Magnusson; Pall Melsted; Gudmundur L Norddahl; Jona Saemundsdottir; Asgeir Sigurdsson; Patrick Sulem; Arna B Agustsdottir; Berglind Eiriksdottir; Run Fridriksdottir; Elisabet E Gardarsdottir; Gudmundur Georgsson; Olafia S Gretarsdottir; Kjartan R Gudmundsson; Thora R Gunnarsdottir; Arnaldur Gylfason; Hilma Holm; Brynjar O Jensson; Aslaug Jonasdottir; Kamilla S Josefsdottir; Thordur Kristjansson; Droplaug N Magnusdottir; Louise le Roux; Gudrun Sigmundsdottir; Gardar Sveinbjornsson; Kristin E Sveinsdottir; Maney Sveinsdottir; Emil A Thorarensen; Bjarni Thorbjornsson; Gisli Masson; Ingileif Jonsdottir; Alma Moller; Thorolfur Gudnason; Karl G Kristinsson; Unnur Thorsteinsdottir; Kari Stefansson
EPI_ISL_417728 hCoV-19/Iceland/70/2020 Europe / Iceland / Reykjavik 2020-03-08 The National University Hospital of Iceland deCODE genetics Daniel F Gudbjartsson; Agnar Helgason; Hakon Jonsson; Olafur T Magnusson; Pall Melsted; Gudmundur L Norddahl; Jona Saemundsdottir; Asgeir Sigurdsson; Patrick Sulem; Arna B Agustsdottir; Berglind Eiriksdottir; Run Fridriksdottir; Elisabet E Gardarsdottir; Gudmundur Georgsson; Olafia S Gretarsdottir; Kjartan R Gudmundsson; Thora R Gunnarsdottir; Arnaldur Gylfason; Hilma Holm; Brynjar O Jensson; Aslaug Jonasdottir; Kamilla S Josefsdottir; Thordur Kristjansson; Droplaug N Magnusdottir; Louise le Roux; Gudrun Sigmundsdottir; Gardar Sveinbjornsson; Kristin E Sveinsdottir; Maney Sveinsdottir; Emil A Thorarensen; Bjarni Thorbjornsson; Gisli Masson; Ingileif Jonsdottir; Alma Moller; Thorolfur Gudnason; Karl G Kristinsson; Unnur Thorsteinsdottir; Kari Stefansson
EPI_ISL_417730 hCoV-19/Iceland/93/2020 Europe / Iceland / Reykjavik 2020-03-10 The National University Hospital of Iceland deCODE genetics Daniel F Gudbjartsson; Agnar Helgason; Hakon Jonsson; Olafur T Magnusson; Pall Melsted; Gudmundur L Norddahl; Jona Saemundsdottir; Asgeir Sigurdsson; Patrick Sulem; Arna B

Agustsdottir; Berglind Eiriksdottir; Run Fridriksdottir; Elisabet E Gardarsdottir; Gudmundur Georgsson; Olafia S Gretarsdottir; Kjartan R Gudmundsson; Thora R Gunnarsdottir; Arnaldur Gylfason; Hilma Holm; Brynjar O Jensson; Aslaug Jonasdottir; Kamilla S Josefsdottir; Thordur Kristjansson; Droplaug N Magnusdottir; Louise le Roux; Gudrun Sigmundsdottir; Gardar Sveinbjornsson; Kristin E Sveinsdottir; Maney Sveinsdottir; Emil A Thorarensen; Bjarni Thorbjornsson; Gisli Masson; Ingileif Jonsdottir; Alma Moller; Thorolfur Gudnason; Karl G Kristinsson; Unnur Thorsteinsdottir; Kari Stefansson
EPI_ISL_417731 hCoV-19/Iceland/61/2020 Europe / Iceland / Reykjavik 2020-03-06 The National University Hospital of Iceland deCODE genetics Daniel F Gudbjartsson; Agnar Helgason; Hakon Jonsson; Olafur T Magnusson; Pall Melsted; Gudmundur L Norddahl; Jona Saemundsdottir; Asgeir Sigurdsson; Patrick Sulem; Arna B Agustsdottir; Berglind Eiriksdottir; Run Fridriksdottir; Elisabet E Gardarsdottir; Gudmundur Georgsson; Olafia S Gretarsdottir; Kjartan R Gudmundsson; Thora R Gunnarsdottir; Arnaldur Gylfason; Hilma Holm; Brynjar O Jensson; Aslaug Jonasdottir; Kamilla S Josefsdottir; Thordur Kristjansson; Droplaug N Magnusdottir; Louise le Roux; Gudrun Sigmundsdottir; Gardar Sveinbjornsson; Kristin E Sveinsdottir; Maney Sveinsdottir; Emil A Thorarensen; Bjarni Thorbjornsson; Gisli Masson; Ingileif Jonsdottir; Alma Moller; Thorolfur Gudnason; Karl G Kristinsson; Unnur Thorsteinsdottir; Kari Stefansson
EPI_ISL_417732 hCoV-19/Iceland/94/2020 Europe / Iceland / Reykjavik 2020-03-10 The National University Hospital of Iceland deCODE genetics Daniel F Gudbjartsson; Agnar Helgason; Hakon Jonsson; Olafur T Magnusson; Pall Melsted; Gudmundur L Norddahl; Jona Saemundsdottir; Asgeir Sigurdsson; Patrick Sulem; Arna B Agustsdottir; Berglind Eiriksdottir; Run Fridriksdottir; Elisabet E Gardarsdottir; Gudmundur Georgsson; Olafia S Gretarsdottir; Kjartan R Gudmundsson; Thora R Gunnarsdottir; Arnaldur Gylfason; Hilma Holm; Brynjar O Jensson; Aslaug Jonasdottir; Kamilla S Josefsdottir; Thordur Kristjansson; Droplaug N Magnusdottir; Louise le Roux; Gudrun Sigmundsdottir; Gardar Sveinbjornsson; Kristin E Sveinsdottir; Maney Sveinsdottir; Emil A Thorarensen; Bjarni Thorbjornsson; Gisli Masson; Ingileif Jonsdottir; Alma Moller; Thorolfur Gudnason; Karl G Kristinsson; Unnur Thorsteinsdottir; Kari Stefansson
EPI_ISL_417733 hCoV-19/Iceland/207/2020 Europe / Iceland / Reykjavik 2020-03-16 The National University Hospital of Iceland deCODE genetics Daniel F Gudbjartsson; Agnar Helgason; Hakon Jonsson; Olafur T Magnusson; Pall Melsted; Gudmundur L Norddahl; Jona Saemundsdottir; Asgeir Sigurdsson; Patrick Sulem; Arna B Agustsdottir; Berglind Eiriksdottir; Run Fridriksdottir; Elisabet E Gardarsdottir; Gudmundur Georgsson; Olafia S Gretarsdottir; Kjartan R Gudmundsson; Thora R Gunnarsdottir; Arnaldur Gylfason; Hilma Holm; Brynjar O Jensson; Aslaug Jonasdottir; Kamilla S Josefsdottir; Thordur Kristjansson; Droplaug N Magnusdottir; Louise le Roux; Gudrun Sigmundsdottir; Gardar Sveinbjornsson; Kristin E Sveinsdottir; Maney Sveinsdottir; Emil A Thorarensen; Bjarni Thorbjornsson; Gisli Masson; Ingileif Jonsdottir; Alma Moller; Thorolfur Gudnason; Karl G Kristinsson; Unnur Thorsteinsdottir; Kari Stefansson
EPI_ISL_417734 hCoV-19/Iceland/22/2020 Europe / Iceland / Reykjavik 2020-03-02 The National University Hospital of Iceland deCODE genetics Daniel F Gudbjartsson; Agnar Helgason; Hakon Jonsson; Olafur T Magnusson; Pall Melsted; Gudmundur L Norddahl; Jona Saemundsdottir; Asgeir Sigurdsson; Patrick Sulem; Arna B Agustsdottir; Berglind Eiriksdottir; Run Fridriksdottir; Elisabet E Gardarsdottir; Gudmundur Georgsson; Olafia S Gretarsdottir; Kjartan R Gudmundsson; Thora R Gunnarsdottir; Arnaldur Gylfason; Hilma Holm; Brynjar O Jensson; Aslaug Jonasdottir; Kamilla S Josefsdottir; Thordur Kristjansson; Droplaug N Magnusdottir; Louise le Roux; Gudrun Sigmundsdottir; Gardar Sveinbjornsson; Kristin E Sveinsdottir; Maney Sveinsdottir; Emil A Thorarensen; Bjarni Thorbjornsson; Gisli Masson; Ingileif Jonsdottir; Alma Moller; Thorolfur Gudnason; Karl G Kristinsson; Unnur Thorsteinsdottir; Kari Stefansson
EPI_ISL_417735 hCoV-19/Iceland/134/2020 Europe / Iceland / Reykjavik 2020-03-12 The National University Hospital of Iceland deCODE genetics Daniel F Gudbjartsson; Agnar Helgason; Hakon Jonsson; Olafur T Magnusson; Pall Melsted; Gudmundur L Norddahl; Jona Saemundsdottir; Asgeir Sigurdsson; Patrick Sulem; Arna B Agustsdottir; Berglind Eiriksdottir; Run Fridriksdottir; Elisabet E Gardarsdottir; Gudmundur Georgsson; Olafia S Gretarsdottir; Kjartan R Gudmundsson; Thora R Gunnarsdottir; Arnaldur Gylfason; Hilma Holm; Brynjar O Jensson; Aslaug Jonasdottir; Kamilla S Josefsdottir; Thordur Kristjansson; Droplaug N Magnusdottir; Louise le Roux; Gudrun Sigmundsdottir; Gardar Sveinbjornsson; Kristin E Sveinsdottir; Maney Sveinsdottir; Emil A Thorarensen; Bjarni Thorbjornsson; Gisli Masson; Ingileif Jonsdottir; Alma Moller; Thorolfur Gudnason; Karl G Kristinsson; Unnur Thorsteinsdottir; Kari Stefansson
EPI_ISL_417736 hCoV-19/Iceland/14/2020 Europe / Iceland / Reykjavik 2020-03-01 The National University Hospital of Iceland deCODE genetics Daniel F Gudbjartsson; Agnar Helgason; Hakon Jonsson; Olafur T Magnusson; Pall Melsted; Gudmundur L Norddahl; Jona Saemundsdottir; Asgeir Sigurdsson; Patrick Sulem; Arna B Agustsdottir; Berglind Eiriksdottir; Run Fridriksdottir; Elisabet E Gardarsdottir; Gudmundur Georgsson; Olafia S Gretarsdottir; Kjartan R Gudmundsson; Thora R Gunnarsdottir; Arnaldur Gylfason; Hilma Holm; Brynjar O Jensson; Aslaug Jonasdottir; Kamilla S Josefsdottir; Thordur Kristjansson; Droplaug N Magnusdottir; Louise le Roux; Gudrun Sigmundsdottir; Gardar Sveinbjornsson; Kristin E Sveinsdottir; Maney Sveinsdottir; Emil A Thorarensen; Bjarni Thorbjornsson; Gisli Masson; Ingileif Jonsdottir; Alma Moller; Thorolfur Gudnason; Karl G Kristinsson; Unnur Thorsteinsdottir; Kari Stefansson
EPI_ISL_417737 hCoV-19/Iceland/155/2020 Europe / Iceland / Reykjavik 2020-03-15 The National University Hospital of Iceland deCODE genetics Daniel F Gudbjartsson; Agnar Helgason; Hakon Jonsson; Olafur T Magnusson; Pall Melsted; Gudmundur L Norddahl; Jona Saemundsdottir; Asgeir Sigurdsson; Patrick Sulem; Arna B Agustsdottir; Berglind Eiriksdottir; Run Fridriksdottir; Elisabet E Gardarsdottir; Gudmundur Georgsson; Olafia S Gretarsdottir; Kjartan R Gudmundsson; Thora R Gunnarsdottir; Arnaldur Gylfason; Hilma Holm; Brynjar O Jensson; Aslaug Jonasdottir; Kamilla S Josefsdottir; Thordur Kristjansson; Droplaug N Magnusdottir; Louise le Roux; Gudrun Sigmundsdottir; Gardar Sveinbjornsson; Kristin E Sveinsdottir; Maney Sveinsdottir; Emil A Thorarensen; Bjarni Thorbjornsson; Gisli Masson; Ingileif Jonsdottir; Alma Moller; Thorolfur Gudnason; Karl G Kristinsson; Unnur Thorsteinsdottir; Kari Stefansson
EPI_ISL_417738 hCoV-19/Iceland/89/2020 Europe / Iceland / Reykjavik 2020-03-10 The National University

Hospital of Iceland	deCODE genetics Daniel F Gudbjartsson; Agnar Helgason; Hakon Jonsson; Olafur T Magnusson; Pall Melsted; Gudmundur L Norddahl; Jona Saemundsdottir; Asgeir Sigurdsson; Patrick Sulem; Arna B Agustsdottir; Berglind Eiriksdottir; Run Fridriksdottir; Elisabet E Gardarsdottir; Gudmundur Georgsson; Olafia S Gretarsdottir; Kjartan R Gudmundsson; Thora R Gunnarsdottir; Arnaldur Gylfason; Hilma Holm; Brynjar O Jensson; Aslaug Jonasdottir; Kamilla S Josefsdottir; Thordur Kristjansson; Droplaug N Magnusdottir; Louise le Roux; Gudrun Sigmundsdottir; Gardar Sveinbjornsson; Kristin E Sveinsdottir; Maney Sveinsdottir; Emil A Thorarensen; Bjarni Thorbjornsson; Gisli Masson; Ingileif Jonsdottir; Alma Moller; Thorolfur Gudnason; Karl G Kristinsson; Unnur Thorsteinsdottir; Kari Stefansson
EPI_ISL_417739	hCoV-19/Iceland/5/2020	Europe / Iceland / Reykjavik	2020-03-13	The National University Hospital of Iceland	deCODE genetics Daniel F Gudbjartsson; Agnar Helgason; Hakon Jonsson; Olafur T Magnusson; Pall Melsted; Gudmundur L Norddahl; Jona Saemundsdottir; Asgeir Sigurdsson; Patrick Sulem; Arna B Agustsdottir; Berglind Eiriksdottir; Run Fridriksdottir; Elisabet E Gardarsdottir; Gudmundur Georgsson; Olafia S Gretarsdottir; Kjartan R Gudmundsson; Thora R Gunnarsdottir; Arnaldur Gylfason; Hilma Holm; Brynjar O Jensson; Aslaug Jonasdottir; Kamilla S Josefsdottir; Thordur Kristjansson; Droplaug N Magnusdottir; Louise le Roux; Gudrun Sigmundsdottir; Gardar Sveinbjornsson; Kristin E Sveinsdottir; Maney Sveinsdottir; Emil A Thorarensen; Bjarni Thorbjornsson; Gisli Masson; Ingileif Jonsdottir; Alma Moller; Thorolfur Gudnason; Karl G Kristinsson; Unnur Thorsteinsdottir; Kari Stefansson
EPI_ISL_417740	hCoV-19/Iceland/7/2020	Europe / Iceland / Reykjavik	2020-03-15	The National University Hospital of Iceland	deCODE genetics Daniel F Gudbjartsson; Agnar Helgason; Hakon Jonsson; Olafur T Magnusson; Pall Melsted; Gudmundur L Norddahl; Jona Saemundsdottir; Asgeir Sigurdsson; Patrick Sulem; Arna B Agustsdottir; Berglind Eiriksdottir; Run Fridriksdottir; Elisabet E Gardarsdottir; Gudmundur Georgsson; Olafia S Gretarsdottir; Kjartan R Gudmundsson; Thora R Gunnarsdottir; Arnaldur Gylfason; Hilma Holm; Brynjar O Jensson; Aslaug Jonasdottir; Kamilla S Josefsdottir; Thordur Kristjansson; Droplaug N Magnusdottir; Louise le Roux; Gudrun Sigmundsdottir; Gardar Sveinbjornsson; Kristin E Sveinsdottir; Maney Sveinsdottir; Emil A Thorarensen; Bjarni Thorbjornsson; Gisli Masson; Ingileif Jonsdottir; Alma Moller; Thorolfur Gudnason; Karl G Kristinsson; Unnur Thorsteinsdottir; Kari Stefansson
EPI_ISL_417741	hCoV-19/Iceland/145/2020	Europe / Iceland / Reykjavik	2020-03-13	The National University Hospital of Iceland	deCODE genetics Daniel F Gudbjartsson; Agnar Helgason; Hakon Jonsson; Olafur T Magnusson; Pall Melsted; Gudmundur L Norddahl; Jona Saemundsdottir; Asgeir Sigurdsson; Patrick Sulem; Arna B Agustsdottir; Berglind Eiriksdottir; Run Fridriksdottir; Elisabet E Gardarsdottir; Gudmundur Georgsson; Olafia S Gretarsdottir; Kjartan R Gudmundsson; Thora R Gunnarsdottir; Arnaldur Gylfason; Hilma Holm; Brynjar O Jensson; Aslaug Jonasdottir; Kamilla S Josefsdottir; Thordur Kristjansson; Droplaug N Magnusdottir; Louise le Roux; Gudrun Sigmundsdottir; Gardar Sveinbjornsson; Kristin E Sveinsdottir; Maney Sveinsdottir; Emil A Thorarensen; Bjarni Thorbjornsson; Gisli Masson; Ingileif Jonsdottir; Alma Moller; Thorolfur Gudnason; Karl G Kristinsson; Unnur Thorsteinsdottir; Kari Stefansson
EPI_ISL_417742	hCoV-19/Iceland/197/2020	Europe / Iceland / Reykjavik	2020-03-16	The National University Hospital of Iceland	deCODE genetics Daniel F Gudbjartsson; Agnar Helgason; Hakon Jonsson; Olafur T Magnusson; Pall Melsted; Gudmundur L Norddahl; Jona Saemundsdottir; Asgeir Sigurdsson; Patrick Sulem; Arna B Agustsdottir; Berglind Eiriksdottir; Run Fridriksdottir; Elisabet E Gardarsdottir; Gudmundur Georgsson; Olafia S Gretarsdottir; Kjartan R Gudmundsson; Thora R Gunnarsdottir; Arnaldur Gylfason; Hilma Holm; Brynjar O Jensson; Aslaug Jonasdottir; Kamilla S Josefsdottir; Thordur Kristjansson; Droplaug N Magnusdottir; Louise le Roux; Gudrun Sigmundsdottir; Gardar Sveinbjornsson; Kristin E Sveinsdottir; Maney Sveinsdottir; Emil A Thorarensen; Bjarni Thorbjornsson; Gisli Masson; Ingileif Jonsdottir; Alma Moller; Thorolfur Gudnason; Karl G Kristinsson; Unnur Thorsteinsdottir; Kari Stefansson
EPI_ISL_417743	hCoV-19/Iceland/200/2020	Europe / Iceland / Reykjavik	2020-03-16	The National University Hospital of Iceland	deCODE genetics Daniel F Gudbjartsson; Agnar Helgason; Hakon Jonsson; Olafur T Magnusson; Pall Melsted; Gudmundur L Norddahl; Jona Saemundsdottir; Asgeir Sigurdsson; Patrick Sulem; Arna B Agustsdottir; Berglind Eiriksdottir; Run Fridriksdottir; Elisabet E Gardarsdottir; Gudmundur Georgsson; Olafia S Gretarsdottir; Kjartan R Gudmundsson; Thora R Gunnarsdottir; Arnaldur Gylfason; Hilma Holm; Brynjar O Jensson; Aslaug Jonasdottir; Kamilla S Josefsdottir; Thordur Kristjansson; Droplaug N Magnusdottir; Louise le Roux; Gudrun Sigmundsdottir; Gardar Sveinbjornsson; Kristin E Sveinsdottir; Maney Sveinsdottir; Emil A Thorarensen; Bjarni Thorbjornsson; Gisli Masson; Ingileif Jonsdottir; Alma Moller; Thorolfur Gudnason; Karl G Kristinsson; Unnur Thorsteinsdottir; Kari Stefansson
EPI_ISL_417744	hCoV-19/Iceland/137/2020	Europe / Iceland / Reykjavik	2020-03-13	The National University Hospital of Iceland	deCODE genetics Daniel F Gudbjartsson; Agnar Helgason; Hakon Jonsson; Olafur T Magnusson; Pall Melsted; Gudmundur L Norddahl; Jona Saemundsdottir; Asgeir Sigurdsson; Patrick Sulem; Arna B Agustsdottir; Berglind Eiriksdottir; Run Fridriksdottir; Elisabet E Gardarsdottir; Gudmundur Georgsson; Olafia S Gretarsdottir; Kjartan R Gudmundsson; Thora R Gunnarsdottir; Arnaldur Gylfason; Hilma Holm; Brynjar O Jensson; Aslaug Jonasdottir; Kamilla S Josefsdottir; Thordur Kristjansson; Droplaug N Magnusdottir; Louise le Roux; Gudrun Sigmundsdottir; Gardar Sveinbjornsson; Kristin E Sveinsdottir; Maney Sveinsdottir; Emil A Thorarensen; Bjarni Thorbjornsson; Gisli Masson; Ingileif Jonsdottir; Alma Moller; Thorolfur Gudnason; Karl G Kristinsson; Unnur Thorsteinsdottir; Kari Stefansson
EPI_ISL_417745	hCoV-19/Iceland/152/2020	Europe / Iceland / Reykjavik	2020-03-13	The National University Hospital of Iceland	deCODE genetics Daniel F Gudbjartsson; Agnar Helgason; Hakon Jonsson; Olafur T Magnusson; Pall Melsted; Gudmundur L Norddahl; Jona Saemundsdottir; Asgeir Sigurdsson; Patrick Sulem; Arna B Agustsdottir; Berglind Eiriksdottir; Run Fridriksdottir; Elisabet E Gardarsdottir; Gudmundur Georgsson; Olafia S Gretarsdottir; Kjartan R Gudmundsson; Thora R Gunnarsdottir; Arnaldur Gylfason; Hilma Holm; Brynjar O Jensson; Aslaug Jonasdottir; Kamilla S Josefsdottir; Thordur Kristjansson; Droplaug N Magnusdottir; Louise le Roux; Gudrun Sigmundsdottir; Gardar Sveinbjornsson; Kristin E Sveinsdottir; Maney Sveinsdottir; Emil A Thorarensen; Bjarni Thorbjornsson; Gisli Masson; Ingileif Jonsdottir; Alma Moller; Thorolfur Gudnason; Karl G

Kristinsson; Unnur Thorsteinsdottir; Kari Stefansson
EPI_ISL_417746 hCoV-19/Iceland/183/2020 Europe / Iceland / Reykjavik 2020-03-16 The National University Hospital of Iceland deCODE genetics Daniel F Gudbjartsson; Agnar Helgason; Hakon Jonsson; Olafur T Magnusson; Pall Melsted; Gudmundur L Norddahl; Jona Saemundsdottir; Asgeir Sigurdsson; Patrick Sulem; Arna B Agustsdottir; Berglind Eiriksdottir; Run Fridriksdottir; Elisabet E Gardarsdottir; Gudmundur Georgsson; Olafia S Gretarsdottir; Kjartan R Gudmundsson; Thora R Gunnarsdottir; Arnaldur Gylfason; Hilma Holm; Brynjar O Jensson; Aslaug Jonasdottir; Kamilla S Josefsdottir; Thordur Kristjansson; Droplaug N Magnusdottir; Louise le Roux; Gudrun Sigmundsdottir; Gardar Sveinbjornsson; Kristin E Sveinsdottir; Maney Sveinsdottir; Emil A Thorarensen; Bjarni Thorbjornsson; Gisli Masson; Ingileif Jonsdottir; Alma Moller; Thorolfur Gudnason; Karl G Kristinsson; Unnur Thorsteinsdottir; Kari Stefansson
EPI_ISL_417747 hCoV-19/Iceland/24/2020 Europe / Iceland / Reykjavik 2020-03-11 The National University Hospital of Iceland deCODE genetics Daniel F Gudbjartsson; Agnar Helgason; Hakon Jonsson; Olafur T Magnusson; Pall Melsted; Gudmundur L Norddahl; Jona Saemundsdottir; Asgeir Sigurdsson; Patrick Sulem; Arna B Agustsdottir; Berglind Eiriksdottir; Run Fridriksdottir; Elisabet E Gardarsdottir; Gudmundur Georgsson; Olafia S Gretarsdottir; Kjartan R Gudmundsson; Thora R Gunnarsdottir; Arnaldur Gylfason; Hilma Holm; Brynjar O Jensson; Aslaug Jonasdottir; Kamilla S Josefsdottir; Thordur Kristjansson; Droplaug N Magnusdottir; Louise le Roux; Gudrun Sigmundsdottir; Gardar Sveinbjornsson; Kristin E Sveinsdottir; Maney Sveinsdottir; Emil A Thorarensen; Bjarni Thorbjornsson; Gisli Masson; Ingileif Jonsdottir; Alma Moller; Thorolfur Gudnason; Karl G Kristinsson; Unnur Thorsteinsdottir; Kari Stefansson
EPI_ISL_417748 hCoV-19/Iceland/109/2020 Europe / Iceland / Reykjavik 2020-03-11 The National University Hospital of Iceland deCODE genetics Daniel F Gudbjartsson; Agnar Helgason; Hakon Jonsson; Olafur T Magnusson; Pall Melsted; Gudmundur L Norddahl; Jona Saemundsdottir; Asgeir Sigurdsson; Patrick Sulem; Arna B Agustsdottir; Berglind Eiriksdottir; Run Fridriksdottir; Elisabet E Gardarsdottir; Gudmundur Georgsson; Olafia S Gretarsdottir; Kjartan R Gudmundsson; Thora R Gunnarsdottir; Arnaldur Gylfason; Hilma Holm; Brynjar O Jensson; Aslaug Jonasdottir; Kamilla S Josefsdottir; Thordur Kristjansson; Droplaug N Magnusdottir; Louise le Roux; Gudrun Sigmundsdottir; Gardar Sveinbjornsson; Kristin E Sveinsdottir; Maney Sveinsdottir; Emil A Thorarensen; Bjarni Thorbjornsson; Gisli Masson; Ingileif Jonsdottir; Alma Moller; Thorolfur Gudnason; Karl G Kristinsson; Unnur Thorsteinsdottir; Kari Stefansson
EPI_ISL_417761 hCoV-19/Iceland/84/2020 Europe / Iceland / Reykjavik 2020-03-10 The National University Hospital of Iceland deCODE genetics Daniel F Gudbjartsson; Agnar Helgason; Hakon Jonsson; Olafur T Magnusson; Pall Melsted; Gudmundur L Norddahl; Jona Saemundsdottir; Asgeir Sigurdsson; Patrick Sulem; Arna B Agustsdottir; Berglind Eiriksdottir; Run Fridriksdottir; Elisabet E Gardarsdottir; Gudmundur Georgsson; Olafia S Gretarsdottir; Kjartan R Gudmundsson; Thora R Gunnarsdottir; Arnaldur Gylfason; Hilma Holm; Brynjar O Jensson; Aslaug Jonasdottir; Kamilla S Josefsdottir; Thordur Kristjansson; Droplaug N Magnusdottir; Louise le Roux; Gudrun Sigmundsdottir; Gardar Sveinbjornsson; Kristin E Sveinsdottir; Maney Sveinsdottir; Emil A Thorarensen; Bjarni Thorbjornsson; Gisli Masson; Ingileif Jonsdottir; Alma Moller; Thorolfur Gudnason; Karl G Kristinsson; Unnur Thorsteinsdottir; Kari Stefansson
EPI_ISL_417762 hCoV-19/Iceland/191/2020 Europe / Iceland / Reykjavik 2020-03-16 The National University Hospital of Iceland deCODE genetics Daniel F Gudbjartsson; Agnar Helgason; Hakon Jonsson; Olafur T Magnusson; Pall Melsted; Gudmundur L Norddahl; Jona Saemundsdottir; Asgeir Sigurdsson; Patrick Sulem; Arna B Agustsdottir; Berglind Eiriksdottir; Run Fridriksdottir; Elisabet E Gardarsdottir; Gudmundur Georgsson; Olafia S Gretarsdottir; Kjartan R Gudmundsson; Thora R Gunnarsdottir; Arnaldur Gylfason; Hilma Holm; Brynjar O Jensson; Aslaug Jonasdottir; Kamilla S Josefsdottir; Thordur Kristjansson; Droplaug N Magnusdottir; Louise le Roux; Gudrun Sigmundsdottir; Gardar Sveinbjornsson; Kristin E Sveinsdottir; Maney Sveinsdottir; Emil A Thorarensen; Bjarni Thorbjornsson; Gisli Masson; Ingileif Jonsdottir; Alma Moller; Thorolfur Gudnason; Karl G Kristinsson; Unnur Thorsteinsdottir; Kari Stefansson
EPI_ISL_417763 hCoV-19/Iceland/211/2020 Europe / Iceland / Reykjavik 2020-03-16 The National University Hospital of Iceland deCODE genetics Daniel F Gudbjartsson; Agnar Helgason; Hakon Jonsson; Olafur T Magnusson; Pall Melsted; Gudmundur L Norddahl; Jona Saemundsdottir; Asgeir Sigurdsson; Patrick Sulem; Arna B Agustsdottir; Berglind Eiriksdottir; Run Fridriksdottir; Elisabet E Gardarsdottir; Gudmundur Georgsson; Olafia S Gretarsdottir; Kjartan R Gudmundsson; Thora R Gunnarsdottir; Arnaldur Gylfason; Hilma Holm; Brynjar O Jensson; Aslaug Jonasdottir; Kamilla S Josefsdottir; Thordur Kristjansson; Droplaug N Magnusdottir; Louise le Roux; Gudrun Sigmundsdottir; Gardar Sveinbjornsson; Kristin E Sveinsdottir; Maney Sveinsdottir; Emil A Thorarensen; Bjarni Thorbjornsson; Gisli Masson; Ingileif Jonsdottir; Alma Moller; Thorolfur Gudnason; Karl G Kristinsson; Unnur Thorsteinsdottir; Kari Stefansson
EPI_ISL_417764 hCoV-19/Iceland/209/2020 Europe / Iceland / Reykjavik 2020-03-16 The National University Hospital of Iceland deCODE genetics Daniel F Gudbjartsson; Agnar Helgason; Hakon Jonsson; Olafur T Magnusson; Pall Melsted; Gudmundur L Norddahl; Jona Saemundsdottir; Asgeir Sigurdsson; Patrick Sulem; Arna B Agustsdottir; Berglind Eiriksdottir; Run Fridriksdottir; Elisabet E Gardarsdottir; Gudmundur Georgsson; Olafia S Gretarsdottir; Kjartan R Gudmundsson; Thora R Gunnarsdottir; Arnaldur Gylfason; Hilma Holm; Brynjar O Jensson; Aslaug Jonasdottir; Kamilla S Josefsdottir; Thordur Kristjansson; Droplaug N Magnusdottir; Louise le Roux; Gudrun Sigmundsdottir; Gardar Sveinbjornsson; Kristin E Sveinsdottir; Maney Sveinsdottir; Emil A Thorarensen; Bjarni Thorbjornsson; Gisli Masson; Ingileif Jonsdottir; Alma Moller; Thorolfur Gudnason; Karl G Kristinsson; Unnur Thorsteinsdottir; Kari Stefansson
EPI_ISL_417766 hCoV-19/Iceland/208/2020 Europe / Iceland / Reykjavik 2020-03-16 The National University Hospital of Iceland deCODE genetics Daniel F Gudbjartsson; Agnar Helgason; Hakon Jonsson; Olafur T Magnusson; Pall Melsted; Gudmundur L Norddahl; Jona Saemundsdottir; Asgeir Sigurdsson; Patrick Sulem; Arna B Agustsdottir; Berglind Eiriksdottir; Run Fridriksdottir; Elisabet E Gardarsdottir; Gudmundur Georgsson; Olafia S Gretarsdottir; Kjartan R Gudmundsson; Thora R Gunnarsdottir; Arnaldur Gylfason; Hilma Holm; Brynjar O Jensson; Aslaug Jonasdottir; Kamilla S Josefsdottir; Thordur Kristjansson; Droplaug N Magnusdottir; Louise le

Roux; Gudrun Sigmundsdottir; Gardar Sveinbjornsson; Kristin E Sveinsdottir; Maney Sveinsdottir; Emil A Thorarensen; Bjarni Thorbjornsson; Gisli Masson; Ingileif Jonsdottir; Alma Moller; Thorolfur Gudnason; Karl G Kristinsson; Unnur Thorsteinsdottir; Kari Stefansson
EPI_ISL_417769 hCoV-19/Iceland/50/2020 Europe / Iceland / Reykjavik 2020-03-05 The National University Hospital of Iceland deCODE genetics Daniel F Gudbjartsson; Agnar Helgason; Hakon Jonsson; Olafur T Magnusson; Pall Melsted; Gudmundur L Norddahl; Jona Saemundsdottir; Asgeir Sigurdsson; Patrick Sulem; Arna B Agustsdottir; Berglind Eiriksdottir; Run Fridriksdottir; Elisabet E Gardarsdottir; Gudmundur Georgsson; Olafia S Gretarsdottir; Kjartan R Gudmundsson; Thora R Gunnarsdottir; Arnaldur Gylfason; Hilma Holm; Brynjar O Jensson; Aslaug Jonasdottir; Kamilla S Josefsdottir; Thordur Kristjansson; Droplaug N Magnusdottir; Louise le Roux; Gudrun Sigmundsdottir; Gardar Sveinbjornsson; Kristin E Sveinsdottir; Maney Sveinsdottir; Emil A Thorarensen; Bjarni Thorbjornsson; Gisli Masson; Ingileif Jonsdottir; Alma Moller; Thorolfur Gudnason; Karl G Kristinsson; Unnur Thorsteinsdottir; Kari Stefansson
EPI_ISL_417770 hCoV-19/Iceland/112/2020 Europe / Iceland / Reykjavik 2020-03-11 The National University Hospital of Iceland deCODE genetics Daniel F Gudbjartsson; Agnar Helgason; Hakon Jonsson; Olafur T Magnusson; Pall Melsted; Gudmundur L Norddahl; Jona Saemundsdottir; Asgeir Sigurdsson; Patrick Sulem; Arna B Agustsdottir; Berglind Eiriksdottir; Run Fridriksdottir; Elisabet E Gardarsdottir; Gudmundur Georgsson; Olafia S Gretarsdottir; Kjartan R Gudmundsson; Thora R Gunnarsdottir; Arnaldur Gylfason; Hilma Holm; Brynjar O Jensson; Aslaug Jonasdottir; Kamilla S Josefsdottir; Thordur Kristjansson; Droplaug N Magnusdottir; Louise le Roux; Gudrun Sigmundsdottir; Gardar Sveinbjornsson; Kristin E Sveinsdottir; Maney Sveinsdottir; Emil A Thorarensen; Bjarni Thorbjornsson; Gisli Masson; Ingileif Jonsdottir; Alma Moller; Thorolfur Gudnason; Karl G Kristinsson; Unnur Thorsteinsdottir; Kari Stefansson
EPI_ISL_417771 hCoV-19/Iceland/131/2020 Europe / Iceland / Reykjavik 2020-03-12 The National University Hospital of Iceland deCODE genetics Daniel F Gudbjartsson; Agnar Helgason; Hakon Jonsson; Olafur T Magnusson; Pall Melsted; Gudmundur L Norddahl; Jona Saemundsdottir; Asgeir Sigurdsson; Patrick Sulem; Arna B Agustsdottir; Berglind Eiriksdottir; Run Fridriksdottir; Elisabet E Gardarsdottir; Gudmundur Georgsson; Olafia S Gretarsdottir; Kjartan R Gudmundsson; Thora R Gunnarsdottir; Arnaldur Gylfason; Hilma Holm; Brynjar O Jensson; Aslaug Jonasdottir; Kamilla S Josefsdottir; Thordur Kristjansson; Droplaug N Magnusdottir; Louise le Roux; Gudrun Sigmundsdottir; Gardar Sveinbjornsson; Kristin E Sveinsdottir; Maney Sveinsdottir; Emil A Thorarensen; Bjarni Thorbjornsson; Gisli Masson; Ingileif Jonsdottir; Alma Moller; Thorolfur Gudnason; Karl G Kristinsson; Unnur Thorsteinsdottir; Kari Stefansson
EPI_ISL_417772 hCoV-19/Iceland/162/2020 Europe / Iceland / Reykjavik 2020-03-14 The National University Hospital of Iceland deCODE genetics Daniel F Gudbjartsson; Agnar Helgason; Hakon Jonsson; Olafur T Magnusson; Pall Melsted; Gudmundur L Norddahl; Jona Saemundsdottir; Asgeir Sigurdsson; Patrick Sulem; Arna B Agustsdottir; Berglind Eiriksdottir; Run Fridriksdottir; Elisabet E Gardarsdottir; Gudmundur Georgsson; Olafia S Gretarsdottir; Kjartan R Gudmundsson; Thora R Gunnarsdottir; Arnaldur Gylfason; Hilma Holm; Brynjar O Jensson; Aslaug Jonasdottir; Kamilla S Josefsdottir; Thordur Kristjansson; Droplaug N Magnusdottir; Louise le Roux; Gudrun Sigmundsdottir; Gardar Sveinbjornsson; Kristin E Sveinsdottir; Maney Sveinsdottir; Emil A Thorarensen; Bjarni Thorbjornsson; Gisli Masson; Ingileif Jonsdottir; Alma Moller; Thorolfur Gudnason; Karl G Kristinsson; Unnur Thorsteinsdottir; Kari Stefansson
EPI_ISL_417773 hCoV-19/Iceland/30/2020 Europe / Iceland / Reykjavik 2020-03-03 The National University Hospital of Iceland deCODE genetics Daniel F Gudbjartsson; Agnar Helgason; Hakon Jonsson; Olafur T Magnusson; Pall Melsted; Gudmundur L Norddahl; Jona Saemundsdottir; Asgeir Sigurdsson; Patrick Sulem; Arna B Agustsdottir; Berglind Eiriksdottir; Run Fridriksdottir; Elisabet E Gardarsdottir; Gudmundur Georgsson; Olafia S Gretarsdottir; Kjartan R Gudmundsson; Thora R Gunnarsdottir; Arnaldur Gylfason; Hilma Holm; Brynjar O Jensson; Aslaug Jonasdottir; Kamilla S Josefsdottir; Thordur Kristjansson; Droplaug N Magnusdottir; Louise le Roux; Gudrun Sigmundsdottir; Gardar Sveinbjornsson; Kristin E Sveinsdottir; Maney Sveinsdottir; Emil A Thorarensen; Bjarni Thorbjornsson; Gisli Masson; Ingileif Jonsdottir; Alma Moller; Thorolfur Gudnason; Karl G Kristinsson; Unnur Thorsteinsdottir; Kari Stefansson
EPI_ISL_417774 hCoV-19/Iceland/175/2020 Europe / Iceland / Reykjavik 2020-03-16 The National University Hospital of Iceland deCODE genetics Daniel F Gudbjartsson; Agnar Helgason; Hakon Jonsson; Olafur T Magnusson; Pall Melsted; Gudmundur L Norddahl; Jona Saemundsdottir; Asgeir Sigurdsson; Patrick Sulem; Arna B Agustsdottir; Berglind Eiriksdottir; Run Fridriksdottir; Elisabet E Gardarsdottir; Gudmundur Georgsson; Olafia S Gretarsdottir; Kjartan R Gudmundsson; Thora R Gunnarsdottir; Arnaldur Gylfason; Hilma Holm; Brynjar O Jensson; Aslaug Jonasdottir; Kamilla S Josefsdottir; Thordur Kristjansson; Droplaug N Magnusdottir; Louise le Roux; Gudrun Sigmundsdottir; Gardar Sveinbjornsson; Kristin E Sveinsdottir; Maney Sveinsdottir; Emil A Thorarensen; Bjarni Thorbjornsson; Gisli Masson; Ingileif Jonsdottir; Alma Moller; Thorolfur Gudnason; Karl G Kristinsson; Unnur Thorsteinsdottir; Kari Stefansson
EPI_ISL_417775 hCoV-19/Iceland/163/2020 Europe / Iceland / Reykjavik 2020-03-14 The National University Hospital of Iceland deCODE genetics Daniel F Gudbjartsson; Agnar Helgason; Hakon Jonsson; Olafur T Magnusson; Pall Melsted; Gudmundur L Norddahl; Jona Saemundsdottir; Asgeir Sigurdsson; Patrick Sulem; Arna B Agustsdottir; Berglind Eiriksdottir; Run Fridriksdottir; Elisabet E Gardarsdottir; Gudmundur Georgsson; Olafia S Gretarsdottir; Kjartan R Gudmundsson; Thora R Gunnarsdottir; Arnaldur Gylfason; Hilma Holm; Brynjar O Jensson; Aslaug Jonasdottir; Kamilla S Josefsdottir; Thordur Kristjansson; Droplaug N Magnusdottir; Louise le Roux; Gudrun Sigmundsdottir; Gardar Sveinbjornsson; Kristin E Sveinsdottir; Maney Sveinsdottir; Emil A Thorarensen; Bjarni Thorbjornsson; Gisli Masson; Ingileif Jonsdottir; Alma Moller; Thorolfur Gudnason; Karl G Kristinsson; Unnur Thorsteinsdottir; Kari Stefansson
EPI_ISL_417777 hCoV-19/Iceland/102/2020 Europe / Iceland / Reykjavik 2020-03-10 The National University Hospital of Iceland deCODE genetics Daniel F Gudbjartsson; Agnar Helgason; Hakon Jonsson; Olafur T Magnusson; Pall Melsted; Gudmundur L Norddahl; Jona Saemundsdottir; Asgeir Sigurdsson; Patrick Sulem; Arna B Agustsdottir; Berglind Eiriksdottir; Run Fridriksdottir; Elisabet E Gardarsdottir; Gudmundur Georgsson; Olafia

S Gretarsdottir; Kjartan R Gudmundsson; Thora R Gunnarsdottir; Arnaldur Gylfason; Hilma Holm; Brynjar O Jensson; Aslaug Jonasdottir; Kamilla S Josefsdottir; Thordur Kristjansson; Droplaug N Magnusdottir; Louise le Roux; Gudrun Sigmundsdottir; Gardar Sveinbjornsson; Kristin E Sveinsdottir; Maney Sveinsdottir; Emil A Thorarensen; Bjarni Thorbjornsson; Gisli Masson; Ingileif Jonsdottir; Alma Moller; Thorolfur Gudnason; Karl G Kristinsson; Unnur Thorsteinsdottir; Kari Stefansson
EPI_ISL_417779 hCoV-19/Iceland/106/2020 Europe / Iceland / Reykjavik 2020-03-11 The National University Hospital of Iceland deCODE genetics Daniel F Gudbjartsson; Agnar Helgason; Hakon Jonsson; Olafur T Magnusson; Pall Melsted; Gudmundur L Norddahl; Jona Saemundsdottir; Asgeir Sigurdsson; Patrick Sulem; Arna B Agustsdottir; Berglind Eiriksdottir; Run Fridriksdottir; Elisabet E Gardarsdottir; Gudmundur Georgsson; Olafia S Gretarsdottir; Kjartan R Gudmundsson; Thora R Gunnarsdottir; Arnaldur Gylfason; Hilma Holm; Brynjar O Jensson; Aslaug Jonasdottir; Kamilla S Josefsdottir; Thordur Kristjansson; Droplaug N Magnusdottir; Louise le Roux; Gudrun Sigmundsdottir; Gardar Sveinbjornsson; Kristin E Sveinsdottir; Maney Sveinsdottir; Emil A Thorarensen; Bjarni Thorbjornsson; Gisli Masson; Ingileif Jonsdottir; Alma Moller; Thorolfur Gudnason; Karl G Kristinsson; Unnur Thorsteinsdottir; Kari Stefansson
EPI_ISL_417781 hCoV-19/Iceland/113/2020 Europe / Iceland / Reykjavik 2020-03-11 The National University Hospital of Iceland deCODE genetics Daniel F Gudbjartsson; Agnar Helgason; Hakon Jonsson; Olafur T Magnusson; Pall Melsted; Gudmundur L Norddahl; Jona Saemundsdottir; Asgeir Sigurdsson; Patrick Sulem; Arna B Agustsdottir; Berglind Eiriksdottir; Run Fridriksdottir; Elisabet E Gardarsdottir; Gudmundur Georgsson; Olafia S Gretarsdottir; Kjartan R Gudmundsson; Thora R Gunnarsdottir; Arnaldur Gylfason; Hilma Holm; Brynjar O Jensson; Aslaug Jonasdottir; Kamilla S Josefsdottir; Thordur Kristjansson; Droplaug N Magnusdottir; Louise le Roux; Gudrun Sigmundsdottir; Gardar Sveinbjornsson; Kristin E Sveinsdottir; Maney Sveinsdottir; Emil A Thorarensen; Bjarni Thorbjornsson; Gisli Masson; Ingileif Jonsdottir; Alma Moller; Thorolfur Gudnason; Karl G Kristinsson; Unnur Thorsteinsdottir; Kari Stefansson
EPI_ISL_417783 hCoV-19/Iceland/123/2020 Europe / Iceland / Reykjavik 2020-03-11 The National University Hospital of Iceland deCODE genetics Daniel F Gudbjartsson; Agnar Helgason; Hakon Jonsson; Olafur T Magnusson; Pall Melsted; Gudmundur L Norddahl; Jona Saemundsdottir; Asgeir Sigurdsson; Patrick Sulem; Arna B Agustsdottir; Berglind Eiriksdottir; Run Fridriksdottir; Elisabet E Gardarsdottir; Gudmundur Georgsson; Olafia S Gretarsdottir; Kjartan R Gudmundsson; Thora R Gunnarsdottir; Arnaldur Gylfason; Hilma Holm; Brynjar O Jensson; Aslaug Jonasdottir; Kamilla S Josefsdottir; Thordur Kristjansson; Droplaug N Magnusdottir; Louise le Roux; Gudrun Sigmundsdottir; Gardar Sveinbjornsson; Kristin E Sveinsdottir; Maney Sveinsdottir; Emil A Thorarensen; Bjarni Thorbjornsson; Gisli Masson; Ingileif Jonsdottir; Alma Moller; Thorolfur Gudnason; Karl G Kristinsson; Unnur Thorsteinsdottir; Kari Stefansson
EPI_ISL_417784 hCoV-19/Iceland/124/2020 Europe / Iceland / Reykjavik 2020-03-12 The National University Hospital of Iceland deCODE genetics Daniel F Gudbjartsson; Agnar Helgason; Hakon Jonsson; Olafur T Magnusson; Pall Melsted; Gudmundur L Norddahl; Jona Saemundsdottir; Asgeir Sigurdsson; Patrick Sulem; Arna B Agustsdottir; Berglind Eiriksdottir; Run Fridriksdottir; Elisabet E Gardarsdottir; Gudmundur Georgsson; Olafia S Gretarsdottir; Kjartan R Gudmundsson; Thora R Gunnarsdottir; Arnaldur Gylfason; Hilma Holm; Brynjar O Jensson; Aslaug Jonasdottir; Kamilla S Josefsdottir; Thordur Kristjansson; Droplaug N Magnusdottir; Louise le Roux; Gudrun Sigmundsdottir; Gardar Sveinbjornsson; Kristin E Sveinsdottir; Maney Sveinsdottir; Emil A Thorarensen; Bjarni Thorbjornsson; Gisli Masson; Ingileif Jonsdottir; Alma Moller; Thorolfur Gudnason; Karl G Kristinsson; Unnur Thorsteinsdottir; Kari Stefansson
EPI_ISL_417785 hCoV-19/Iceland/126/2020 Europe / Iceland / Reykjavik 2020-03-12 The National University Hospital of Iceland deCODE genetics Daniel F Gudbjartsson; Agnar Helgason; Hakon Jonsson; Olafur T Magnusson; Pall Melsted; Gudmundur L Norddahl; Jona Saemundsdottir; Asgeir Sigurdsson; Patrick Sulem; Arna B Agustsdottir; Berglind Eiriksdottir; Run Fridriksdottir; Elisabet E Gardarsdottir; Gudmundur Georgsson; Olafia S Gretarsdottir; Kjartan R Gudmundsson; Thora R Gunnarsdottir; Arnaldur Gylfason; Hilma Holm; Brynjar O Jensson; Aslaug Jonasdottir; Kamilla S Josefsdottir; Thordur Kristjansson; Droplaug N Magnusdottir; Louise le Roux; Gudrun Sigmundsdottir; Gardar Sveinbjornsson; Kristin E Sveinsdottir; Maney Sveinsdottir; Emil A Thorarensen; Bjarni Thorbjornsson; Gisli Masson; Ingileif Jonsdottir; Alma Moller; Thorolfur Gudnason; Karl G Kristinsson; Unnur Thorsteinsdottir; Kari Stefansson
EPI_ISL_417786 hCoV-19/Iceland/128/2020 Europe / Iceland / Reykjavik 2020-03-12 The National University Hospital of Iceland deCODE genetics Daniel F Gudbjartsson; Agnar Helgason; Hakon Jonsson; Olafur T Magnusson; Pall Melsted; Gudmundur L Norddahl; Jona Saemundsdottir; Asgeir Sigurdsson; Patrick Sulem; Arna B Agustsdottir; Berglind Eiriksdottir; Run Fridriksdottir; Elisabet E Gardarsdottir; Gudmundur Georgsson; Olafia S Gretarsdottir; Kjartan R Gudmundsson; Thora R Gunnarsdottir; Arnaldur Gylfason; Hilma Holm; Brynjar O Jensson; Aslaug Jonasdottir; Kamilla S Josefsdottir; Thordur Kristjansson; Droplaug N Magnusdottir; Louise le Roux; Gudrun Sigmundsdottir; Gardar Sveinbjornsson; Kristin E Sveinsdottir; Maney Sveinsdottir; Emil A Thorarensen; Bjarni Thorbjornsson; Gisli Masson; Ingileif Jonsdottir; Alma Moller; Thorolfur Gudnason; Karl G Kristinsson; Unnur Thorsteinsdottir; Kari Stefansson
EPI_ISL_417787 hCoV-19/Iceland/132/2020 Europe / Iceland / Reykjavik 2020-03-12 The National University Hospital of Iceland deCODE genetics Daniel F Gudbjartsson; Agnar Helgason; Hakon Jonsson; Olafur T Magnusson; Pall Melsted; Gudmundur L Norddahl; Jona Saemundsdottir; Asgeir Sigurdsson; Patrick Sulem; Arna B Agustsdottir; Berglind Eiriksdottir; Run Fridriksdottir; Elisabet E Gardarsdottir; Gudmundur Georgsson; Olafia S Gretarsdottir; Kjartan R Gudmundsson; Thora R Gunnarsdottir; Arnaldur Gylfason; Hilma Holm; Brynjar O Jensson; Aslaug Jonasdottir; Kamilla S Josefsdottir; Thordur Kristjansson; Droplaug N Magnusdottir; Louise le Roux; Gudrun Sigmundsdottir; Gardar Sveinbjornsson; Kristin E Sveinsdottir; Maney Sveinsdottir; Emil A Thorarensen; Bjarni Thorbjornsson; Gisli Masson; Ingileif Jonsdottir; Alma Moller; Thorolfur Gudnason; Karl G Kristinsson; Unnur Thorsteinsdottir; Kari Stefansson
EPI_ISL_417788 hCoV-19/Iceland/133/2020 Europe / Iceland / Reykjavik 2020-03-12 The National University Hospital of Iceland deCODE genetics Daniel F Gudbjartsson; Agnar Helgason; Hakon Jonsson; Olafur T

Magnusson; Pall Melsted; Gudmundur L Norddahl; Jona Saemundsdottir; Asgeir Sigurdsson; Patrick Sulem; Arna B Agustsdottir; Berglind Eiriksdottir; Run Fridriksdottir; Elisabet E Gardarsdottir; Gudmundur Georgsson; Olafia S Gretarsdottir; Kjartan R Gudmundsson; Thora R Gunnarsdottir; Arnaldur Gylfason; Hilma Holm; Brynjar O Jensson; Aslaug Jonasdottir; Kamilla S Josefsdottir; Thordur Kristjansson; Droplaug N Magnusdottir; Louise le Roux; Gudrun Sigmundsdottir; Gardar Sveinbjornsson; Kristin E Sveinsdottir; Maney Sveinsdottir; Emil A Thorarensen; Bjarni Thorbjornsson; Gisli Masson; Ingileif Jonsdottir; Alma Moller; Thorolfur Gudnason; Karl G Kristinsson; Unnur Thorsteinsdottir; Kari Stefansson
EPI_ISL_417789 hCoV-19/Iceland/136/2020 Europe / Iceland / Reykjavik 2020-03-12 The National University Hospital of Iceland deCODE genetics Daniel F Gudbjartsson; Agnar Helgason; Hakon Jonsson; Olafur T Magnusson; Pall Melsted; Gudmundur L Norddahl; Jona Saemundsdottir; Asgeir Sigurdsson; Patrick Sulem; Arna B Agustsdottir; Berglind Eiriksdottir; Run Fridriksdottir; Elisabet E Gardarsdottir; Gudmundur Georgsson; Olafia S Gretarsdottir; Kjartan R Gudmundsson; Thora R Gunnarsdottir; Arnaldur Gylfason; Hilma Holm; Brynjar O Jensson; Aslaug Jonasdottir; Kamilla S Josefsdottir; Thordur Kristjansson; Droplaug N Magnusdottir; Louise le Roux; Gudrun Sigmundsdottir; Gardar Sveinbjornsson; Kristin E Sveinsdottir; Maney Sveinsdottir; Emil A Thorarensen; Bjarni Thorbjornsson; Gisli Masson; Ingileif Jonsdottir; Alma Moller; Thorolfur Gudnason; Karl G Kristinsson; Unnur Thorsteinsdottir; Kari Stefansson
EPI_ISL_417790 hCoV-19/Iceland/139/2020 Europe / Iceland / Reykjavik 2020-03-12 The National University Hospital of Iceland deCODE genetics Daniel F Gudbjartsson; Agnar Helgason; Hakon Jonsson; Olafur T Magnusson; Pall Melsted; Gudmundur L Norddahl; Jona Saemundsdottir; Asgeir Sigurdsson; Patrick Sulem; Arna B Agustsdottir; Berglind Eiriksdottir; Run Fridriksdottir; Elisabet E Gardarsdottir; Gudmundur Georgsson; Olafia S Gretarsdottir; Kjartan R Gudmundsson; Thora R Gunnarsdottir; Arnaldur Gylfason; Hilma Holm; Brynjar O Jensson; Aslaug Jonasdottir; Kamilla S Josefsdottir; Thordur Kristjansson; Droplaug N Magnusdottir; Louise le Roux; Gudrun Sigmundsdottir; Gardar Sveinbjornsson; Kristin E Sveinsdottir; Maney Sveinsdottir; Emil A Thorarensen; Bjarni Thorbjornsson; Gisli Masson; Ingileif Jonsdottir; Alma Moller; Thorolfur Gudnason; Karl G Kristinsson; Unnur Thorsteinsdottir; Kari Stefansson
EPI_ISL_417791 hCoV-19/Iceland/140/2020 Europe / Iceland / Reykjavik 2020-03-12 The National University Hospital of Iceland deCODE genetics Daniel F Gudbjartsson; Agnar Helgason; Hakon Jonsson; Olafur T Magnusson; Pall Melsted; Gudmundur L Norddahl; Jona Saemundsdottir; Asgeir Sigurdsson; Patrick Sulem; Arna B Agustsdottir; Berglind Eiriksdottir; Run Fridriksdottir; Elisabet E Gardarsdottir; Gudmundur Georgsson; Olafia S Gretarsdottir; Kjartan R Gudmundsson; Thora R Gunnarsdottir; Arnaldur Gylfason; Hilma Holm; Brynjar O Jensson; Aslaug Jonasdottir; Kamilla S Josefsdottir; Thordur Kristjansson; Droplaug N Magnusdottir; Louise le Roux; Gudrun Sigmundsdottir; Gardar Sveinbjornsson; Kristin E Sveinsdottir; Maney Sveinsdottir; Emil A Thorarensen; Bjarni Thorbjornsson; Gisli Masson; Ingileif Jonsdottir; Alma Moller; Thorolfur Gudnason; Karl G Kristinsson; Unnur Thorsteinsdottir; Kari Stefansson
EPI_ISL_417792 hCoV-19/Iceland/141/2020 Europe / Iceland / Reykjavik 2020-03-13 The National University Hospital of Iceland deCODE genetics Daniel F Gudbjartsson; Agnar Helgason; Hakon Jonsson; Olafur T Magnusson; Pall Melsted; Gudmundur L Norddahl; Jona Saemundsdottir; Asgeir Sigurdsson; Patrick Sulem; Arna B Agustsdottir; Berglind Eiriksdottir; Run Fridriksdottir; Elisabet E Gardarsdottir; Gudmundur Georgsson; Olafia S Gretarsdottir; Kjartan R Gudmundsson; Thora R Gunnarsdottir; Arnaldur Gylfason; Hilma Holm; Brynjar O Jensson; Aslaug Jonasdottir; Kamilla S Josefsdottir; Thordur Kristjansson; Droplaug N Magnusdottir; Louise le Roux; Gudrun Sigmundsdottir; Gardar Sveinbjornsson; Kristin E Sveinsdottir; Maney Sveinsdottir; Emil A Thorarensen; Bjarni Thorbjornsson; Gisli Masson; Ingileif Jonsdottir; Alma Moller; Thorolfur Gudnason; Karl G Kristinsson; Unnur Thorsteinsdottir; Kari Stefansson
EPI_ISL_417793 hCoV-19/Iceland/142/2020 Europe / Iceland / Reykjavik 2020-03-13 The National University Hospital of Iceland deCODE genetics Daniel F Gudbjartsson; Agnar Helgason; Hakon Jonsson; Olafur T Magnusson; Pall Melsted; Gudmundur L Norddahl; Jona Saemundsdottir; Asgeir Sigurdsson; Patrick Sulem; Arna B Agustsdottir; Berglind Eiriksdottir; Run Fridriksdottir; Elisabet E Gardarsdottir; Gudmundur Georgsson; Olafia S Gretarsdottir; Kjartan R Gudmundsson; Thora R Gunnarsdottir; Arnaldur Gylfason; Hilma Holm; Brynjar O Jensson; Aslaug Jonasdottir; Kamilla S Josefsdottir; Thordur Kristjansson; Droplaug N Magnusdottir; Louise le Roux; Gudrun Sigmundsdottir; Gardar Sveinbjornsson; Kristin E Sveinsdottir; Maney Sveinsdottir; Emil A Thorarensen; Bjarni Thorbjornsson; Gisli Masson; Ingileif Jonsdottir; Alma Moller; Thorolfur Gudnason; Karl G Kristinsson; Unnur Thorsteinsdottir; Kari Stefansson
EPI_ISL_417795 hCoV-19/Iceland/144/2020 Europe / Iceland / Reykjavik 2020-03-13 The National University Hospital of Iceland deCODE genetics Daniel F Gudbjartsson; Agnar Helgason; Hakon Jonsson; Olafur T Magnusson; Pall Melsted; Gudmundur L Norddahl; Jona Saemundsdottir; Asgeir Sigurdsson; Patrick Sulem; Arna B Agustsdottir; Berglind Eiriksdottir; Run Fridriksdottir; Elisabet E Gardarsdottir; Gudmundur Georgsson; Olafia S Gretarsdottir; Kjartan R Gudmundsson; Thora R Gunnarsdottir; Arnaldur Gylfason; Hilma Holm; Brynjar O Jensson; Aslaug Jonasdottir; Kamilla S Josefsdottir; Thordur Kristjansson; Droplaug N Magnusdottir; Louise le Roux; Gudrun Sigmundsdottir; Gardar Sveinbjornsson; Kristin E Sveinsdottir; Maney Sveinsdottir; Emil A Thorarensen; Bjarni Thorbjornsson; Gisli Masson; Ingileif Jonsdottir; Alma Moller; Thorolfur Gudnason; Karl G Kristinsson; Unnur Thorsteinsdottir; Kari Stefansson
EPI_ISL_417796 hCoV-19/Iceland/146/2020 Europe / Iceland / Reykjavik 2020-03-13 The National University Hospital of Iceland deCODE genetics Daniel F Gudbjartsson; Agnar Helgason; Hakon Jonsson; Olafur T Magnusson; Pall Melsted; Gudmundur L Norddahl; Jona Saemundsdottir; Asgeir Sigurdsson; Patrick Sulem; Arna B Agustsdottir; Berglind Eiriksdottir; Run Fridriksdottir; Elisabet E Gardarsdottir; Gudmundur Georgsson; Olafia S Gretarsdottir; Kjartan R Gudmundsson; Thora R Gunnarsdottir; Arnaldur Gylfason; Hilma Holm; Brynjar O Jensson; Aslaug Jonasdottir; Kamilla S Josefsdottir; Thordur Kristjansson; Droplaug N Magnusdottir; Louise le Roux; Gudrun Sigmundsdottir; Gardar Sveinbjornsson; Kristin E Sveinsdottir; Maney Sveinsdottir; Emil A Thorarensen; Bjarni Thorbjornsson; Gisli Masson; Ingileif Jonsdottir; Alma Moller; Thorolfur Gudnason; Karl G Kristinsson; Unnur Thorsteinsdottir; Kari Stefansson

EPI_ISL_417797  hCoV-19/Iceland/147/2020        Europe / Iceland / Reykjavik    2020-03-13      The National University Hospital of Iceland  deCODE genetics Daniel F Gudbjartsson; Agnar Helgason; Hakon Jonsson; Olafur T Magnusson; Pall Melsted; Gudmundur L Norddahl; Jona Saemundsdottir; Asgeir Sigurdsson; Patrick Sulem; Arna B Agustsdottir; Berglind Eiriksdottir; Run Fridriksdottir; Elisabet E Gardarsdottir; Gudmundur Georgsson; Olafia S Gretarsdottir; Kjartan R Gudmundsson; Thora R Gunnarsdottir; Arnaldur Gylfason; Hilma Holm; Brynjar O Jensson; Aslaug Jonasdottir; Kamilla S Josefsdottir; Thordur Kristjansson; Droplaug N Magnusdottir; Louise le Roux; Gudrun Sigmundsdottir; Gardar Sveinbjornsson; Kristin E Sveinsdottir; Maney Sveinsdottir; Emil A Thorarensen; Bjarni Thorbjornsson; Gisli Masson; Ingileif Jonsdottir; Alma Moller; Thorolfur Gudnason; Karl G Kristinsson; Unnur Thorsteinsdottir; Kari Stefansson
EPI_ISL_417798  hCoV-19/Iceland/148/2020        Europe / Iceland / Reykjavik    2020-03-13      The National University Hospital of Iceland  deCODE genetics Daniel F Gudbjartsson; Agnar Helgason; Hakon Jonsson; Olafur T Magnusson; Pall Melsted; Gudmundur L Norddahl; Jona Saemundsdottir; Asgeir Sigurdsson; Patrick Sulem; Arna B Agustsdottir; Berglind Eiriksdottir; Run Fridriksdottir; Elisabet E Gardarsdottir; Gudmundur Georgsson; Olafia S Gretarsdottir; Kjartan R Gudmundsson; Thora R Gunnarsdottir; Arnaldur Gylfason; Hilma Holm; Brynjar O Jensson; Aslaug Jonasdottir; Kamilla S Josefsdottir; Thordur Kristjansson; Droplaug N Magnusdottir; Louise le Roux; Gudrun Sigmundsdottir; Gardar Sveinbjornsson; Kristin E Sveinsdottir; Maney Sveinsdottir; Emil A Thorarensen; Bjarni Thorbjornsson; Gisli Masson; Ingileif Jonsdottir; Alma Moller; Thorolfur Gudnason; Karl G Kristinsson; Unnur Thorsteinsdottir; Kari Stefansson
EPI_ISL_417799  hCoV-19/Iceland/149/2020        Europe / Iceland / Reykjavik    2020-03-13      The National University Hospital of Iceland  deCODE genetics Daniel F Gudbjartsson; Agnar Helgason; Hakon Jonsson; Olafur T Magnusson; Pall Melsted; Gudmundur L Norddahl; Jona Saemundsdottir; Asgeir Sigurdsson; Patrick Sulem; Arna B Agustsdottir; Berglind Eiriksdottir; Run Fridriksdottir; Elisabet E Gardarsdottir; Gudmundur Georgsson; Olafia S Gretarsdottir; Kjartan R Gudmundsson; Thora R Gunnarsdottir; Arnaldur Gylfason; Hilma Holm; Brynjar O Jensson; Aslaug Jonasdottir; Kamilla S Josefsdottir; Thordur Kristjansson; Droplaug N Magnusdottir; Louise le Roux; Gudrun Sigmundsdottir; Gardar Sveinbjornsson; Kristin E Sveinsdottir; Maney Sveinsdottir; Emil A Thorarensen; Bjarni Thorbjornsson; Gisli Masson; Ingileif Jonsdottir; Alma Moller; Thorolfur Gudnason; Karl G Kristinsson; Unnur Thorsteinsdottir; Kari Stefansson
EPI_ISL_420000  hCoV-19/Australia/VIC309/2020   Oceania / Australia / Victoria  2020-03-23      Microbiological Diagnostic Unit Public Health Laboratory        Microbiological Diagnostic Unit Public Health Laboratory Seemann T., Schultz M., Sait, M., Sherry, N.
EPI_ISL_420001  hCoV-19/Australia/VIC310/2020   Oceania / Australia / Victoria  2020-03-23      Microbiological Diagnostic Unit Public Health Laboratory        Microbiological Diagnostic Unit Public Health Laboratory Seemann T., Schultz M., Sait, M., Sherry, N.
EPI_ISL_420002  hCoV-19/Australia/VIC311/2020   Oceania / Australia / Victoria  2020-03-23      Microbiological Diagnostic Unit Public Health Laboratory        Microbiological Diagnostic Unit Public Health Laboratory Seemann T., Schultz M., Sait, M., Sherry, N.
EPI_ISL_420003  hCoV-19/Australia/VIC312/2020   Oceania / Australia / Victoria  2020-03-23      Microbiological Diagnostic Unit Public Health Laboratory        Microbiological Diagnostic Unit Public Health Laboratory Seemann T., Schultz M., Sait, M., Sherry, N.
EPI_ISL_420004  hCoV-19/Australia/VIC313/2020   Oceania / Australia / Victoria  2020-03-24      Victorian Infectious Diseases Reference Laboratory (VIDRL)        Victorian Infectious Diseases Reference Laboratory and Microbiological Diagnostic Unit Public Health Laboratory, Doherty Institute     Caly L., Seemann T., Sait, M., Schultz M., Druce J., Sherry, N.
EPI_ISL_420005  hCoV-19/Australia/VIC314/2020   Oceania / Australia / Victoria  2020-03-24      Victorian Infectious Diseases Reference Laboratory (VIDRL)        Victorian Infectious Diseases Reference Laboratory and Microbiological Diagnostic Unit Public Health Laboratory, Doherty Institute     Caly L., Seemann T., Sait, M., Schultz M., Druce J., Sherry, N.
EPI_ISL_420006  hCoV-19/Australia/VIC315/2020   Oceania / Australia / Victoria  2020-03-24      Microbiological Diagnostic Unit Public Health Laboratory        Microbiological Diagnostic Unit Public Health Laboratory Seemann T., Schultz M., Sait, M., Sherry, N.
EPI_ISL_420007  hCoV-19/Australia/VIC316/2020   Oceania / Australia / Victoria  2020-03-24      Microbiological Diagnostic Unit Public Health Laboratory        Microbiological Diagnostic Unit Public Health Laboratory Seemann T., Schultz M., Sait, M., Sherry, N.
EPI_ISL_420008  hCoV-19/Australia/VIC317/2020   Oceania / Australia / Victoria  2020-03-24      Microbiological Diagnostic Unit Public Health Laboratory        Microbiological Diagnostic Unit Public Health Laboratory Seemann T., Schultz M., Sait, M., Sherry, N.
EPI_ISL_420009  hCoV-19/Australia/VIC318/2020   Oceania / Australia / Victoria  2020-03-24      Microbiological Diagnostic Unit Public Health Laboratory        Microbiological Diagnostic Unit Public Health Laboratory Seemann T., Schultz M., Sait, M., Sherry, N.
EPI_ISL_420010  hCoV-19/Australia/VIC319/2020   Oceania / Australia / Victoria  2020-03-24      Microbiological Diagnostic Unit Public Health Laboratory        Microbiological Diagnostic Unit Public Health Laboratory Seemann T., Schultz M., Sait, M., Sherry, N.
EPI_ISL_420011  hCoV-19/Australia/VIC320/2020   Oceania / Australia / Victoria  2020-03-24      Microbiological Diagnostic Unit Public Health Laboratory        Microbiological Diagnostic Unit Public Health Laboratory Seemann T., Schultz M., Sait, M., Sherry, N.
EPI_ISL_420012  hCoV-19/Australia/VIC321/2020   Oceania / Australia / Victoria  2020-03-24      Microbiological Diagnostic Unit Public Health Laboratory        Microbiological Diagnostic Unit Public Health Laboratory Seemann T., Schultz M., Sait, M., Sherry, N.
EPI_ISL_420013  hCoV-19/Australia/VIC322/2020   Oceania / Australia / Victoria  2020-03-24      Microbiological Diagnostic Unit Public Health Laboratory        Microbiological Diagnostic Unit Public Health Laboratory

Seemann T., Schultz M., Sait, M., Sherry, N.
EPI_ISL_420014 hCoV-19/Australia/VIC323/2020 Oceania / Australia / Victoria 2020-03-24 Microbiological Diagnostic Unit Public Health Laboratory Microbiological Diagnostic Unit Public Health Laboratory Seemann T., Schultz M., Sait, M., Sherry, N.
EPI_ISL_421225 hCoV-19/Hangzhou/HZCDC0135/2020 Asia / China / Hangzhou 2020-01-22 Hangzhou Center for Diseases Control and Prevention Hangzhou Center for Diseases Control and Prevention Jun Li, Haoqiu Wang, Lingfeng Mao, Hua Yu, Xinfen Yu, Zhou Sun, Xin Qian, Shuchang Chen, Junfang Chen, Xuchu Wang
EPI_ISL_421226 hCoV-19/Hangzhou/HZCDC0119/2020 Asia / China / Hangzhou 2020-01-22 Hangzhou Center for Diseases Control and Prevention Hangzhou Center for Diseases Control and Prevention Jun Li, Haoqiu Wang, Lingfeng Mao, Hua Yu, Xinfen Yu, Zhou Sun, Xin Qian, Shuchang Chen, Junfang Chen, Xuchu Wang
EPI_ISL_421227 hCoV-19/Hangzhou/HZCDC0091L/2020 Asia / China / Hangzhou 2020-01-21 Hangzhou Center for Diseases Control and Prevention Hangzhou Center for Diseases Control and Prevention Jun Li, Haoqiu Wang, Lingfeng Mao, Hua Yu, Xinfen Yu, Zhou Sun, Xin Qian, Shuchang Chen, Junfang Chen, Xuchu Wang
EPI_ISL_421228 hCoV-19/Hangzhou/HZCDC0091/2020 Asia / China / Hangzhou 2020-01-21 Hangzhou Center for Diseases Control and Prevention Hangzhou Center for Diseases Control and Prevention Jun Li, Haoqiu Wang, Lingfeng Mao, Hua Yu, Xinfen Yu, Zhou Sun, Xin Qian, Shuchang Chen, Junfang Chen, Xuchu Wang
EPI_ISL_421229 hCoV-19/Hangzhou/HZCDC0090L/2020 Asia / China / Hangzhou 2020-01-21 Hangzhou Center for Diseases Control and Prevention Hangzhou Center for Diseases Control and Prevention Jun Li, Haoqiu Wang, Lingfeng Mao, Hua Yu, Xinfen Yu, Zhou Sun, Xin Qian, Shuchang Chen, Junfang Chen, Xuchu Wang
EPI_ISL_421230 hCoV-19/Hangzhou/HZCDC0090/2020 Asia / China / Hangzhou 2020-01-21 Hangzhou Center for Diseases Control and Prevention Hangzhou Center for Diseases Control and Prevention Jun Li, Haoqiu Wang, Lingfeng Mao, Hua Yu, Xinfen Yu, Zhou Sun, Xin Qian, Shuchang Chen, Junfang Chen, Xuchu Wang
EPI_ISL_421231 hCoV-19/Hangzhou/HZCDC0049L/2020 Asia / China / Hangzhou 2020-01-21 Hangzhou Center for Diseases Control and Prevention Hangzhou Center for Diseases Control and Prevention Jun Li, Haoqiu Wang, Lingfeng Mao, Hua Yu, Xinfen Yu, Zhou Sun, Xin Qian, Shuchang Chen, Junfang Chen, Xuchu Wang
EPI_ISL_421232 hCoV-19/Hangzhou/HZCDC0048L/2020 Asia / China / Hangzhou 2020-01-21 Hangzhou Center for Diseases Control and Prevention Hangzhou Center for Diseases Control and Prevention Jun Li, Haoqiu Wang, Lingfeng Mao, Hua Yu, Xinfen Yu, Zhou Sun, Xin Qian, Shuchang Chen, Junfang Chen, Xuchu Wang
EPI_ISL_421233 hCoV-19/Hangzhou/HZCDC0048/2020 Asia / China / Hangzhou 2020-01-21 Hangzhou Center for Diseases Control and Prevention Hangzhou Center for Diseases Control and Prevention Jun Li, Haoqiu Wang, Lingfeng Mao, Hua Yu, Xinfen Yu, Zhou Sun, Xin Qian, Shuchang Chen, Junfang Chen, Xuchu Wang
EPI_ISL_421234 hCoV-19/Hangzhou/HZCDC0025/2020 Asia / China / Hangzhou 2020-01-21 Hangzhou Center for Diseases Control and Prevention Hangzhou Center for Diseases Control and Prevention Jun Li, Haoqiu Wang, Lingfeng Mao, Hua Yu, Xinfen Yu, Zhou Sun, Xin Qian, Shuchang Chen, Junfang Chen, Xuchu Wang
EPI_ISL_421235 hCoV-19/Hangzhou/HZCDC0013/2020 Asia / China / Hangzhou 2020-01-20 Hangzhou Center for Diseases Control and Prevention Hangzhou Center for Diseases Control and Prevention Jun Li, Haoqiu Wang, Lingfeng Mao, Hua Yu, Xinfen Yu, Zhou Sun, Xin Qian, Shuchang Chen, Junfang Chen, Xuchu Wang
EPI_ISL_421236 hCoV-19/Hangzhou/HZCDC0012/2020 Asia / China / Hangzhou 2020-01-20 Hangzhou Center for Diseases Control and Prevention Hangzhou Center for Diseases Control and Prevention Jun Li, Haoqiu Wang, Lingfeng Mao, Hua Yu, Xinfen Yu, Zhou Sun, Xin Qian, Shuchang Chen, Junfang Chen, Xuchu Wang
EPI_ISL_421237 hCoV-19/Jiujiang/JX22/2020 Asia / China / Jiangxi / Jiujiang 2020-01-22 Jiangxi Province Center for Disease Control and Prevention Jiangxi Province Center for Disease Control and Prevention JianXiong Li,Ying Xiong,Tian Gong,Yong Shi,Jun Zhou,Fang Xiao,ShiWen Liu,XiaoQing Liu,Gang Xu,DaJin Xiao,Xin Ran,YanNi Zhang
EPI_ISL_421238 hCoV-19/Nanchang/JX174/2020 Asia / China / Jiangxi / Nanchang 2020-01-29 Jiangxi Province Center for Disease Control and Prevention Jiangxi Province Center for Disease Control and Prevention JianXiong Li,Ying Xiong,Tian Gong,Yong Shi,Jun Zhou,Fang Xiao,ShiWen Liu,XiaoQing Liu,Gang Xu,DaJin Xiao,Xin Ran,YanNi Zhang
EPI_ISL_421239 hCoV-19/Shangrao/JX105/2020 Asia / China / Jiangxi / Shangrao 2020-01-26 Jiangxi Province Center for Disease Control and Prevention Jiangxi Province Center for Disease Control and Prevention JianXiong Li,Ying Xiong,Tian Gong,Yong Shi,Jun Zhou,Fang Xiao,ShiWen Liu,XiaoQing Liu,Gang Xu,DaJin Xiao,Xin Ran,YanNi Zhang
EPI_ISL_421240 hCoV-19/Shangrao/JX1948/2020 Asia / China / Jiangxi / Shangrao 2020-02-05 Jiangxi Province Center for Disease Control and Prevention Jiangxi Province Center for Disease Control and Prevention JianXiong Li,Ying Xiong,Tian Gong,Yong Shi,Jun Zhou,Fang Xiao,ShiWen Liu,XiaoQing Liu,Gang Xu,DaJin Xiao,Xin Ran,YanNi Zhang
EPI_ISL_421241 hCoV-19/Nanchang/JX177/2020 Asia / China / Jiangxi / Nanchang 2020-01-30 Jiangxi Province Center for Disease Control and Prevention Jiangxi Province Center for Disease Control and Prevention JianXiong Li,Ying Xiong,Tian Gong,Yong Shi,Jun Zhou,Fang Xiao,ShiWen Liu,XiaoQing Liu,Gang Xu,DaJin Xiao,Xin Ran,YanNi Zhang
EPI_ISL_421242 hCoV-19/Ganzhou/JX81/2020 Asia / China / Jiangxi / Ganzhou 2020-01-25 Jiangxi Province Center for Disease Control and Prevention Jiangxi Province Center for Disease Control and Prevention JianXiong Li,Ying Xiong,Tian Gong,Yong Shi,Jun Zhou,Fang Xiao,ShiWen Liu,XiaoQing Liu,Gang Xu,DaJin Xiao,Xin Ran,YanNi Zhang
EPI_ISL_421243 hCoV-19/Nanchang/JX14/2020 Asia / China / Jiangxi / Nanchang 2020-01-21 Jiangxi Province Center for Disease Control and Prevention Jiangxi Province Center for Disease Control and Prevention JianXiong Li,Ying Xiong,Tian Gong,Yong Shi,Jun Zhou,Fang Xiao,ShiWen Liu,XiaoQing Liu,Gang Xu,DaJin Xiao,Xin Ran,YanNi Zhang
EPI_ISL_421244 hCoV-19/Shangrao/JX29/2020 Asia / China / Jiangxi / Shangrao 2020-01-22 Jiangxi Province Center for Disease Control and Prevention Jiangxi Province Center for Disease Control and

Prevention	JianXiong Li,Ying Xiong,Tian Gong,Yong Shi,Jun Zhou,Fang Xiao,ShiWen Liu,XiaoQing Liu,Gang Xu,DaJin Xiao,Xin Ran,YanNi Zhang
EPI_ISL_421245	hCoV-19/Nanchang/JX90/2020	Asia / China / Jiangxi / Nanchang	2020-01-25	Jiangxi Province Center for Disease Control and Prevention	Jiangxi Province Center for Disease Control and Prevention	JianXiong Li,Ying Xiong,Tian Gong,Yong Shi,Jun Zhou,Fang Xiao,ShiWen Liu,XiaoQing Liu,Gang Xu,DaJin Xiao,Xin Ran,YanNi Zhang
EPI_ISL_421246	hCoV-19/Nanchang/JXN3T4/2020	Asia / China / Jiangxi / Nanchang	2020-02-26	Jiangxi Province Center for Disease Control and Prevention	Jiangxi Province Center for Disease Control and Prevention	JianXiong Li,Ying Xiong,Tian Gong,Yong Shi,Jun Zhou,Fang Xiao,ShiWen Liu,XiaoQing Liu,Gang Xu,DaJin Xiao,Xin Ran,YanNi Zhang
EPI_ISL_421247	hCoV-19/Nanchang/JX149/2020	Asia / China / Jiangxi / Nanchang	2020-01-27	Jiangxi Province Center for Disease Control and Prevention	Jiangxi Province Center for Disease Control and Prevention	JianXiong Li,Ying Xiong,Tian Gong,Yong Shi,Jun Zhou,Fang Xiao,ShiWen Liu,XiaoQing Liu,Gang Xu,DaJin Xiao,Xin Ran,YanNi Zhang
EPI_ISL_421248	hCoV-19/Nanchang/JX39/2020	Asia / China / Jiangxi / Nanchang	2020-01-23	Jiangxi Province Center for Disease Control and Prevention	Jiangxi Province Center for Disease Control and Prevention	JianXiong Li,Ying Xiong,Tian Gong,Yong Shi,Jun Zhou,Fang Xiao,ShiWen Liu,XiaoQing Liu,Gang Xu,DaJin Xiao,Xin Ran,YanNi Zhang
EPI_ISL_421249	hCoV-19/Xinyu/JX122/2020	Asia / China / Jiangxi / Xinyu	2020-01-25	Jiangxi Province Center for Disease Control and Prevention	Jiangxi Province Center for Disease Control and Prevention	JianXiong Li,Ying Xiong,Tian Gong,Yong Shi,Jun Zhou,Fang Xiao,ShiWen Liu,XiaoQing Liu,Gang Xu,DaJin Xiao,Xin Ran,YanNi Zhang
EPI_ISL_421250	hCoV-19/Shangrao/JX1177/2020	Asia / China / Jiangxi / Shangrao	2020-02-03	Jiangxi Province Center for Disease Control and Prevention	Jiangxi Province Center for Disease Control and Prevention	JianXiong Li,Ying Xiong,Tian Gong,Yong Shi,Jun Zhou,Fang Xiao,ShiWen Liu,XiaoQing Liu,Gang Xu,DaJin Xiao,Xin Ran,YanNi Zhang
EPI_ISL_421251	hCoV-19/Shangrao/JX1176/2020	Asia / China / Jiangxi / Shangrao	2020-02-03	Jiangxi Province Center for Disease Control and Prevention	Jiangxi Province Center for Disease Control and Prevention	JianXiong Li,Ying Xiong,Tian Gong,Yong Shi,Jun Zhou,Fang Xiao,ShiWen Liu,XiaoQing Liu,Gang Xu,DaJin Xiao,Xin Ran,YanNi Zhang
EPI_ISL_421252	hCoV-19/Pingxiang/JX5/2020	Asia / China / Jiangxi / Pingxiang	2020-01-11	Jiangxi Province Center for Disease Control and Prevention	Jiangxi Province Center for Disease Control and Prevention	JianXiong Li,Ying Xiong,Tian Gong,Yong Shi,Jun Zhou,Fang Xiao,ShiWen Liu,XiaoQing Liu,Gang Xu,DaJin Xiao,Xin Ran,YanNi Zhang
EPI_ISL_421253	hCoV-19/Jian/JX129/2020	Asia / China / Jiangxi / Jian	2020-01-26	Jiangxi Province Center for Disease Control and Prevention	Jiangxi Province Center for Disease Control and Prevention	JianXiong Li,Ying Xiong,Tian Gong,Yong Shi,Jun Zhou,Fang Xiao,ShiWen Liu,XiaoQing Liu,Gang Xu,DaJin Xiao,Xin Ran,YanNi Zhang
EPI_ISL_421254	hCoV-19/Shangrao/JX1178/2020	Asia / China / Jiangxi / Shangrao	2020-02-03	Jiangxi Province Center for Disease Control and Prevention	Jiangxi Province Center for Disease Control and Prevention	JianXiong Li,Ying Xiong,Tian Gong,Yong Shi,Jun Zhou,Fang Xiao,ShiWen Liu,XiaoQing Liu,Gang Xu,DaJin Xiao,Xin Ran,YanNi Zhang
EPI_ISL_421256	hCoV-19/Jian/JX169/2020	Asia / China / Jiangxi / Jian	2020-01-27	Jiangxi Province Center for Disease Control and Prevention	Jiangxi Province Center for Disease Control and Prevention	JianXiong Li,Ying Xiong,Tian Gong,Yong Shi,Jun Zhou,Fang Xiao,ShiWen Liu,XiaoQing Liu,Gang Xu,DaJin Xiao,Xin Ran,YanNi Zhang
EPI_ISL_421257	hCoV-19/Shangrao/JX1215/2020	Asia / China / Jiangxi / Shangrao	2020-02-02	Jiangxi Province Center for Disease Control and Prevention	Jiangxi Province Center for Disease Control and Prevention	JianXiong Li,Ying Xiong,Tian Gong,Yong Shi,Jun Zhou,Fang Xiao,ShiWen Liu,XiaoQing Liu,Gang Xu,DaJin Xiao,Xin Ran,YanNi Zhang
EPI_ISL_421258	hCoV-19/Shangrao/JX1974/2020	Asia / China / Jiangxi / Shangrao	2020-02-08	Jiangxi Province Center for Disease Control and Prevention	Jiangxi Province Center for Disease Control and Prevention	JianXiong Li,Ying Xiong,Tian Gong,Yong Shi,Jun Zhou,Fang Xiao,ShiWen Liu,XiaoQing Liu,Gang Xu,DaJin Xiao,Xin Ran,YanNi Zhang
EPI_ISL_421259	hCoV-19/Pingxiang/JX151/2020	Asia / China / Jiangxi / Pingxiang	2020-01-27	Jiangxi Province Center for Disease Control and Prevention	Jiangxi Province Center for Disease Control and Prevention	JianXiong Li,Ying Xiong,Tian Gong,Yong Shi,Jun Zhou,Fang Xiao,ShiWen Liu,XiaoQing Liu,Gang Xu,DaJin Xiao,Xin Ran,YanNi Zhang
EPI_ISL_421260	hCoV-19/Xinyu/JX124/2020	Asia / China / Jiangxi / Xinyu	2020-01-26	Jiangxi Province Center for Disease Control and Prevention	Jiangxi Province Center for Disease Control and Prevention	JianXiong Li,Ying Xiong,Tian Gong,Yong Shi,Jun Zhou,Fang Xiao,ShiWen Liu,XiaoQing Liu,Gang Xu,DaJin Xiao,Xin Ran,YanNi Zhang
EPI_ISL_421261	hCoV-19/Nanchang/JX176/2020	Asia / China / Jiangxi / Nanchang	2020-01-29	Jiangxi Province Center for Disease Control and Prevention	Jiangxi Province Center for Disease Control and Prevention	JianXiong Li,Ying Xiong,Tian Gong,Yong Shi,Jun Zhou,Fang Xiao,ShiWen Liu,XiaoQing Liu,Gang Xu,DaJin Xiao,Xin Ran,YanNi Zhang
EPI_ISL_421262	hCoV-19/Nanchang/JX155/2020	Asia / China / Jiangxi / Nanchang	2020-01-29	Jiangxi Province Center for Disease Control and Prevention	Jiangxi Province Center for Disease Control and Prevention	JianXiong Li,Ying Xiong,Tian Gong,Yong Shi,Jun Zhou,Fang Xiao,ShiWen Liu,XiaoQing Liu,Gang Xu,DaJin Xiao,Xin Ran,YanNi Zhang

EPI_ISL_421272 hCoV-19/USA/WY-WYPHL001/2020 North America / USA / Wyoming 2020-03-10 Wyoming Public Health Laboratory Center for Global Health, University of New Mexico Health Sciences Center Daryl Domman, Kurt Schwalm, Rob Christensen, Wanda Manley, Cari Sloma, Noah Hull, Darrell Dinwiddie
EPI_ISL_421275 hCoV-19/Russia/Moscow_PMVL-1/2020 Europe / Russia / Moscow 2020-03-18 Russian State Collection of Viruses Pathogenic Microorganisms Variability Laboratory Alexey Shchetinin, Maria Nikiforova, Nadezhda Kuznetsova, Ekaterina Aksenova, Marina Kunda, Natalia Ryzhova, Olga Voronina, Inna Dolzhikova, Daria Grousova, Andrey Botikov, Denis Logunov, Alexander Gintsburg, Vladimir Gushchin
EPI_ISL_421279 hCoV-19/USA/IN-Lilly-IPB0170-4bc20/2020 North America / USA / Indiana / Indianapolis 2020-03-19 Clinical Diagnostics Laboratory, Diagnostic & Experimental Pathology, Lilly Research Laboratories Clinical Diagnostics Laboratory, Diagnostic & Experimental Pathology, Lilly Research Laboratories Tim Holzer, Mayuri Vaidya, Angie Fulford, Sam McNeely, Rachael Redmond, Phil Ebert, John Calley, Leslie O’Neill Reising, Pat Finnegan, Erin Wray, John McElwee, Jeff Fill, Joe Oakley, Andrew Schade
EPI_ISL_421281 hCoV-19/USA/IN-Lilly-IPB0170-4bc21/2020 North America / USA / Indiana / Indianapolis 2020-03-19 Clinical Diagnostics Laboratory, Diagnostic & Experimental Pathology, Lilly Research Laboratories Clinical Diagnostics Laboratory, Diagnostic & Experimental Pathology, Lilly Research Laboratories Tim Holzer, Mayuri Vaidya, Angie Fulford, Sam McNeely, Rachael Redmond, Phil Ebert, John Calley, Leslie O’Neill Reising, Pat Finnegan, Erin Wray, John McElwee, Jeff Fill, Joe Oakley, Andrew Schade
EPI_ISL_421283 hCoV-19/USA/WI-UW-25/2020 North America / USA / Wisconsin / Madison 2020-03-23 University of Wisconsin-Madison AIDS Vaccine Research Laboratories University of Wisconsin-Madison AIDS Vaccine Research Laboratories Gage Moreno, Katarina Braun, et al. AIDS Vaccine Research Laboratories
EPI_ISL_421284 hCoV-19/USA/WI-UW-26/2020 North America / USA / Wisconsin / Verona 2020-03-24 University of Wisconsin-Madison AIDS Vaccine Research Laboratories University of Wisconsin-Madison AIDS Vaccine Research Laboratories Gage Moreno, Katarina Braun, et al. AIDS Vaccine Research Laboratories
EPI_ISL_421285 hCoV-19/USA/WI-UW-27/2020 North America / USA / Wisconsin / Mount Horeb 2020-03-25 University of Wisconsin-Madison AIDS Vaccine Research Laboratories University of Wisconsin-Madison AIDS Vaccine Research Laboratories Gage Moreno, Katarina Braun, et al. AIDS Vaccine Research Laboratories
EPI_ISL_421286 hCoV-19/USA/WI-UW-28/2020 North America / USA / Wisconsin / Madison 2020-03-25 University of Wisconsin-Madison AIDS Vaccine Research Laboratories University of Wisconsin-Madison AIDS Vaccine Research Laboratories Gage Moreno, Katarina Braun, et al. AIDS Vaccine Research Laboratories
EPI_ISL_421287 hCoV-19/USA/WI-UW-29/2020 North America / USA / Wisconsin / Fitchburg 2020-03-24 University of Wisconsin-Madison AIDS Vaccine Research Laboratories University of Wisconsin-Madison AIDS Vaccine Research Laboratories Gage Moreno, Katarina Braun, et al. AIDS Vaccine Research Laboratories
EPI_ISL_421288 hCoV-19/USA/WI-UW-30/2020 North America / USA / Wisconsin / Columbus 2020-03-26 University of Wisconsin-Madison AIDS Vaccine Research Laboratories University of Wisconsin-Madison AIDS Vaccine Research Laboratories Gage Moreno, Katarina Braun, et al. AIDS Vaccine Research Laboratories
EPI_ISL_421289 hCoV-19/USA/WI-UW-31/2020 North America / USA / Wisconsin / Madison 2020-03-25 University of Wisconsin-Madison AIDS Vaccine Research Laboratories University of Wisconsin-Madison AIDS Vaccine Research Laboratories Gage Moreno, Katarina Braun, et al. AIDS Vaccine Research Laboratories
EPI_ISL_421290 hCoV-19/USA/WI-UW-32/2020 North America / USA / Wisconsin / Madison 2020-03-24 University of Wisconsin-Madison AIDS Vaccine Research Laboratories University of Wisconsin-Madison AIDS Vaccine Research Laboratories Gage Moreno, Katarina Braun, et al. AIDS Vaccine Research Laboratories
EPI_ISL_421291 hCoV-19/USA/WI-UW-33/2020 North America / USA / Wisconsin / Madison 2020-03-23 University of Wisconsin-Madison AIDS Vaccine Research Laboratories University of Wisconsin-Madison AIDS Vaccine Research Laboratories Gage Moreno, Katarina Braun, et al. AIDS Vaccine Research Laboratories
EPI_ISL_421292 hCoV-19/USA/WI-UW-34/2020 North America / USA / Wisconsin / Madison 2020-03-26 University of Wisconsin-Madison AIDS Vaccine Research Laboratories University of Wisconsin-Madison AIDS Vaccine Research Laboratories Gage Moreno, Katarina Braun, et al. AIDS Vaccine Research Laboratories
EPI_ISL_421293 hCoV-19/USA/WI-UW-35/2020 North America / USA / Wisconsin / Stoughton 2020-03-24 University of Wisconsin-Madison AIDS Vaccine Research Laboratories University of Wisconsin-Madison AIDS Vaccine Research Laboratories Gage Moreno, Katarina Braun, et al. AIDS Vaccine Research Laboratories
EPI_ISL_421294 hCoV-19/USA/WI-UW-36/2020 North America / USA / Wisconsin / Janesville 2020-03-25 University of Wisconsin-Madison AIDS Vaccine Research Laboratories University of Wisconsin-Madison AIDS Vaccine Research Laboratories Gage Moreno, Katarina Braun, et al. AIDS Vaccine Research Laboratories
EPI_ISL_421295 hCoV-19/USA/WI-UW-37/2020 North America / USA / Wisconsin / Verona 2020-03-22 University of Wisconsin-Madison AIDS Vaccine Research Laboratories University of Wisconsin-Madison AIDS Vaccine Research Laboratories Gage Moreno, Katarina Braun, et al. AIDS Vaccine Research Laboratories
EPI_ISL_421296 hCoV-19/USA/WI-UW-38/2020 North America / USA / Wisconsin / Stoughton 2020-03-25 University of Wisconsin-Madison AIDS Vaccine Research Laboratories University of Wisconsin-Madison AIDS Vaccine Research Laboratories Gage Moreno, Katarina Braun, et al. AIDS Vaccine Research Laboratories
EPI_ISL_421297 hCoV-19/USA/WI-UW-39/2020 North America / USA / Wisconsin / Marshall 2020-03-22 University of Wisconsin-Madison AIDS Vaccine Research Laboratories University of Wisconsin-Madison AIDS Vaccine Research Laboratories Gage Moreno, Katarina Braun, et al. AIDS Vaccine Research Laboratories
EPI_ISL_421298 hCoV-19/USA/WI-UW-40/2020 North America / USA / Wisconsin / Waunakee 2020-03-24 University of Wisconsin-Madison AIDS Vaccine Research Laboratories University of Wisconsin-Madison AIDS Vaccine Research Laboratories Gage Moreno, Katarina Braun, et al. AIDS Vaccine Research Laboratories
EPI_ISL_421299 hCoV-19/USA/WI-UW-41/2020 North America / USA / Wisconsin / DeForest 2020-03-25 University of Wisconsin-Madison AIDS Vaccine Research Laboratories University of Wisconsin-Madison AIDS Vaccine Research Laboratories Gage Moreno, Katarina Braun, et al. AIDS Vaccine Research Laboratories
EPI_ISL_421303 hCoV-19/USA/WI-UW-45/2020 North America / USA / Wisconsin / Oregon 2020-03-22 University of Wisconsin-Madison AIDS Vaccine Research Laboratories University of Wisconsin-Madison AIDS

Vaccine Research Laboratories   Gage Moreno, Katarina Braun, et al. AIDS Vaccine Research Laboratories
EPI_ISL_421324  hCoV-19/USA/WI-UW-66/2020       North America / USA / Wisconsin / Madison       2020-03-24
University of Wisconsin-Madison AIDS Vaccine Research Laboratories      University of Wisconsin-Madison AIDS Vaccine Research Laboratories   Gage Moreno, Katarina Braun, et al. AIDS Vaccine Research Laboratories
EPI_ISL_421325  hCoV-19/USA/WI-UW-67/2020       North America / USA / Wisconsin / Sun Prarie    2020-03-25
University of Wisconsin-Madison AIDS Vaccine Research Laboratories      University of Wisconsin-Madison AIDS Vaccine Research Laboratories   Gage Moreno, Katarina Braun, et al. AIDS Vaccine Research Laboratories
EPI_ISL_421326  hCoV-19/USA/WI-UW-68/2020       North America / USA / Wisconsin / Madison       2020-03-24
University of Wisconsin-Madison AIDS Vaccine Research Laboratories      University of Wisconsin-Madison AIDS Vaccine Research Laboratories   Gage Moreno, Katarina Braun, et al. AIDS Vaccine Research Laboratories
EPI_ISL_421327  hCoV-19/USA/WI-UW-69/2020       North America / USA / Wisconsin / Stoughton     2020-03-19
University of Wisconsin-Madison AIDS Vaccine Research Laboratories      University of Wisconsin-Madison AIDS Vaccine Research Laboratories   Gage Moreno, Katarina Braun, et al. AIDS Vaccine Research Laboratories
EPI_ISL_421328  hCoV-19/USA/WI-UW-70/2020       North America / USA / Wisconsin / Sun Prarie    2020-03-19
University of Wisconsin-Madison AIDS Vaccine Research Laboratories      University of Wisconsin-Madison AIDS Vaccine Research Laboratories   Gage Moreno, Katarina Braun, et al. AIDS Vaccine Research Laboratories
EPI_ISL_421329  hCoV-19/USA/WI-UW-71/2020       North America / USA / Wisconsin / Madison       2020-03-24
University of Wisconsin-Madison AIDS Vaccine Research Laboratories      University of Wisconsin-Madison AIDS Vaccine Research Laboratories   Gage Moreno, Katarina Braun, et al. AIDS Vaccine Research Laboratories
EPI_ISL_421330  hCoV-19/USA/WI-UW-72/2020       North America / USA / Wisconsin / Madison       2020-03-25
University of Wisconsin-Madison AIDS Vaccine Research Laboratories      University of Wisconsin-Madison AIDS Vaccine Research Laboratories   Gage Moreno, Katarina Braun, et al. AIDS Vaccine Research Laboratories
EPI_ISL_421331  hCoV-19/USA/WI-UW-73/2020       North America / USA / Wisconsin / Madison       2020-03-24
University of Wisconsin-Madison AIDS Vaccine Research Laboratories      University of Wisconsin-Madison AIDS Vaccine Research Laboratories   Gage Moreno, Katarina Braun, et al. AIDS Vaccine Research Laboratories
EPI_ISL_421332  hCoV-19/USA/WI-UW-74/2020       North America / USA / Wisconsin / Fitchburg     2020-03-20
University of Wisconsin-Madison AIDS Vaccine Research Laboratories      University of Wisconsin-Madison AIDS Vaccine Research Laboratories   Gage Moreno, Katarina Braun, et al. AIDS Vaccine Research Laboratories
EPI_ISL_421333  hCoV-19/USA/WI-UW-75/2020       North America / USA / Wisconsin / Madison       2020-03-26
University of Wisconsin-Madison AIDS Vaccine Research Laboratories      University of Wisconsin-Madison AIDS Vaccine Research Laboratories   Gage Moreno, Katarina Braun, et al. AIDS Vaccine Research Laboratories
EPI_ISL_421334  hCoV-19/USA/WI-UW-76/2020       North America / USA / Wisconsin / Marshall      2020-03-22
University of Wisconsin-Madison AIDS Vaccine Research Laboratories      University of Wisconsin-Madison AIDS Vaccine Research Laboratories   Gage Moreno, Katarina Braun, et al. AIDS Vaccine Research Laboratories
EPI_ISL_421335  hCoV-19/USA/WI-UW-77/2020       North America / USA / Wisconsin / Stoughton     2020-03-19
University of Wisconsin-Madison AIDS Vaccine Research Laboratories      University of Wisconsin-Madison AIDS Vaccine Research Laboratories   Gage Moreno, Katarina Braun, et al. AIDS Vaccine Research Laboratories
EPI_ISL_421336  hCoV-19/USA/WI-UW-78/2020       North America / USA / Wisconsin / Madison       2020-03-24
University of Wisconsin-Madison AIDS Vaccine Research Laboratories      University of Wisconsin-Madison AIDS Vaccine Research Laboratories   Gage Moreno, Katarina Braun, et al. AIDS Vaccine Research Laboratories
EPI_ISL_421338  hCoV-19/USA/WI-UW-79/2020       North America / USA / Wisconsin / Madison       2020-03-23
University of Wisconsin-Madison AIDS Vaccine Research Laboratories      University of Wisconsin-Madison AIDS Vaccine Research Laboratories   Gage Moreno, Katarina Braun, et al. AIDS Vaccine Research Laboratories
EPI_ISL_421339  hCoV-19/USA/WI-UW-80/2020       North America / USA / Wisconsin / Mount Horeb   2020-03-24
University of Wisconsin-Madison AIDS Vaccine Research Laboratories      University of Wisconsin-Madison AIDS Vaccine Research Laboratories   Gage Moreno, Katarina Braun, et al. AIDS Vaccine Research Laboratories
EPI_ISL_421340  hCoV-19/USA/WI-UW-81/2020       North America / USA / Wisconsin / Verona        2020-03-25
University of Wisconsin-Madison AIDS Vaccine Research Laboratories      University of Wisconsin-Madison AIDS Vaccine Research Laboratories   Gage Moreno, Katarina Braun, et al. AIDS Vaccine Research Laboratories
EPI_ISL_421341  hCoV-19/USA/WI-UW-82/2020       North America / USA / Wisconsin / Monona        2020-03-25
University of Wisconsin-Madison AIDS Vaccine Research Laboratories      University of Wisconsin-Madison AIDS Vaccine Research Laboratories   Gage Moreno, Katarina Braun, et al. AIDS Vaccine Research Laboratories
EPI_ISL_421342  hCoV-19/USA/WI-UW-83/2020       North America / USA / Wisconsin / Columbus      2020-03-25
University of Wisconsin-Madison AIDS Vaccine Research Laboratories      University of Wisconsin-Madison AIDS Vaccine Research Laboratories   Gage Moreno, Katarina Braun, et al. AIDS Vaccine Research Laboratories
EPI_ISL_421343  hCoV-19/USA/WI-UW-84/2020       North America / USA / Wisconsin / Middleton     2020-03-24
University of Wisconsin-Madison AIDS Vaccine Research Laboratories      University of Wisconsin-Madison AIDS Vaccine Research Laboratories   Gage Moreno, Katarina Braun, et al. AIDS Vaccine Research Laboratories
EPI_ISL_422556  hCoV-19/USA/NY-PV09301/2020     North America / USA / New York / Brooklyn       2020-03-21
MSHS Clinical Microbiology Laboratories MSHS Pathogen Surveillance Program      Ana S. Gonzalez-Reiche, Mitchell Sullivan, Ajay Obla, Gopi Patel, Emilia Sordillo, Melissa Gitman, Alberto Paniz-mondolfi, Matthew Hernandez, Shelcie Fabre, Jose Polanco, Zenab Khan, Bremy Albuquerque, Jayeeta Dutta, Juan Soto, Shwetha Sridhar Hara, Ying-Chih Wang, Melissa Smith, Robert Sebra, Lisa Miorin, Wen-chun Liu, Randy Albrecht, Judith Aberg, Florian Krammer, Adolfo Garcia-Sarstre, Viviana Simon, Harm van Bakel
EPI_ISL_422557  hCoV-19/USA/NY-PV09303/2020     North America / USA / New York / Manhattan      2020-03-21
MSHS Clinical Microbiology Laboratories MSHS Pathogen Surveillance Program      Ana S. Gonzalez-Reiche, Mitchell Sullivan, Ajay Obla, Gopi Patel, Emilia Sordillo, Melissa Gitman, Alberto Paniz-mondolfi, Matthew Hernandez, Shelcie Fabre, Jose Polanco, Zenab Khan, Bremy Albuquerque, Jayeeta Dutta, Juan Soto, Shwetha Sridhar Hara, Ying-Chih Wang, Melissa Smith, Robert Sebra, Lisa Miorin, Wen-chun Liu, Randy Albrecht, Judith Aberg, Florian Krammer, Adolfo Garcia-Sarstre, Viviana Simon, Harm van Bakel

EPI_ISL_422558 hCoV-19/USA/NY-PV09304/2020 North America / USA / New York / Brooklyn 2020-03-21 MSHS Clinical Microbiology Laboratories MSHS Pathogen Surveillance Program Ana S. Gonzalez-Reiche, Mitchell Sullivan, Ajay Obla, Gopi Patel, Emilia Sordillo, Melissa Gitman, Alberto Paniz-mondolfi, Matthew Hernandez, Shelcie Fabre, Jose Polanco, Zenab Khan, Bremy Albuquerque, Jayeeta Dutta, Juan Soto, Shwetha Sridhar Hara, Ying-Chih Wang, Melissa Smith, Robert Sebra, Lisa Miorin, Wen-chun Liu, Randy Albrecht, Judith Aberg, Florian Krammer, Adolfo Garcia-Sarstre, Viviana Simon, Harm van Bakel
EPI_ISL_422559 hCoV-19/USA/NY-PV09305/2020 North America / USA / New York / Brooklyn 2020-03-22 MSHS Clinical Microbiology Laboratories MSHS Pathogen Surveillance Program Ana S. Gonzalez-Reiche, Mitchell Sullivan, Ajay Obla, Gopi Patel, Emilia Sordillo, Melissa Gitman, Alberto Paniz-mondolfi, Matthew Hernandez, Shelcie Fabre, Jose Polanco, Zenab Khan, Bremy Albuquerque, Jayeeta Dutta, Juan Soto, Shwetha Sridhar Hara, Ying-Chih Wang, Melissa Smith, Robert Sebra, Lisa Miorin, Wen-chun Liu, Randy Albrecht, Judith Aberg, Florian Krammer, Adolfo Garcia-Sarstre, Viviana Simon, Harm van Bakel
EPI_ISL_422560 hCoV-19/USA/NY-PV09307/2020 North America / USA / New York / 2020-03-22 MSHS Clinical Microbiology Laboratories MSHS Pathogen Surveillance Program Ana S. Gonzalez-Reiche, Mitchell Sullivan, Ajay Obla, Gopi Patel, Emilia Sordillo, Melissa Gitman, Alberto Paniz-mondolfi, Matthew Hernandez, Shelcie Fabre, Jose Polanco, Zenab Khan, Bremy Albuquerque, Jayeeta Dutta, Juan Soto, Shwetha Sridhar Hara, Ying-Chih Wang, Melissa Smith, Robert Sebra, Lisa Miorin, Wen-chun Liu, Randy Albrecht, Judith Aberg, Florian Krammer, Adolfo Garcia-Sarstre, Viviana Simon, Harm van Bakel
EPI_ISL_422561 hCoV-19/USA/NY-PV09308/2020 North America / USA / New York / Manhattan 2020-03-22 MSHS Clinical Microbiology Laboratories MSHS Pathogen Surveillance Program Ana S. Gonzalez-Reiche, Mitchell Sullivan, Ajay Obla, Gopi Patel, Emilia Sordillo, Melissa Gitman, Alberto Paniz-mondolfi, Matthew Hernandez, Shelcie Fabre, Jose Polanco, Zenab Khan, Bremy Albuquerque, Jayeeta Dutta, Juan Soto, Shwetha Sridhar Hara, Ying-Chih Wang, Melissa Smith, Robert Sebra, Lisa Miorin, Wen-chun Liu, Randy Albrecht, Judith Aberg, Florian Krammer, Adolfo Garcia-Sarstre, Viviana Simon, Harm van Bakel
EPI_ISL_422562 hCoV-19/USA/NY-PV09309/2020 North America / USA / New York / Bronx 2020-03-22 MSHS Clinical Microbiology Laboratories MSHS Pathogen Surveillance Program Ana S. Gonzalez-Reiche, Mitchell Sullivan, Ajay Obla, Gopi Patel, Emilia Sordillo, Melissa Gitman, Alberto Paniz-mondolfi, Matthew Hernandez, Shelcie Fabre, Jose Polanco, Zenab Khan, Bremy Albuquerque, Jayeeta Dutta, Juan Soto, Shwetha Sridhar Hara, Ying-Chih Wang, Melissa Smith, Robert Sebra, Lisa Miorin, Wen-chun Liu, Randy Albrecht, Judith Aberg, Florian Krammer, Adolfo Garcia-Sarstre, Viviana Simon, Harm van Bakel
EPI_ISL_422566 hCoV-19/Netherlands/Friesland_10/2020 Europe / Netherlands / Friesland 2020-03-30 Dutch COVID-19 response team Erasmus Medical Center Bas Oude Munnink, David Nieuwenhuijse, Reina Sikkema, Claudia Schapendonk, Irina Chestakova, Anne van der Linden, Theo Bestebroer, Stefan van Nieuwkoop, Mark Pronk, Pascal Lexmond, Corien Swaan, Manon Haverkate, Madelief Mollers, Mart Stein, Sandra Kengne Kamga Mobou, Jeroen van Kampen, Jolanda Voermans, Aura Timen, Corine GeurtsvanKessel, Annemiek van der Eijk, Richard Molenkamp, Marion Koopmans, on behalf of the Dutch national COVID-19 response team.
EPI_ISL_422568 hCoV-19/Netherlands/Friesland_3/2020 Europe / Netherlands / Friesland 2020-03-28 Dutch COVID-19 response team Erasmus Medical Center Bas Oude Munnink, David Nieuwenhuijse, Reina Sikkema, Claudia Schapendonk, Irina Chestakova, Anne van der Linden, Theo Bestebroer, Stefan van Nieuwkoop, Mark Pronk, Pascal Lexmond, Corien Swaan, Manon Haverkate, Madelief Mollers, Mart Stein, Sandra Kengne Kamga Mobou, Jeroen van Kampen, Jolanda Voermans, Aura Timen, Corine GeurtsvanKessel, Annemiek van der Eijk, Richard Molenkamp, Marion Koopmans, on behalf of the Dutch national COVID-19 response team.
EPI_ISL_422569 hCoV-19/Netherlands/Friesland_4/2020 Europe / Netherlands / Friesland 2020-03-27 Dutch COVID-19 response team Erasmus Medical Center Bas Oude Munnink, David Nieuwenhuijse, Reina Sikkema, Claudia Schapendonk, Irina Chestakova, Anne van der Linden, Theo Bestebroer, Stefan van Nieuwkoop, Mark Pronk, Pascal Lexmond, Corien Swaan, Manon Haverkate, Madelief Mollers, Mart Stein, Sandra Kengne Kamga Mobou, Jeroen van Kampen, Jolanda Voermans, Aura Timen, Corine GeurtsvanKessel, Annemiek van der Eijk, Richard Molenkamp, Marion Koopmans, on behalf of the Dutch national COVID-19 response team.
EPI_ISL_422571 hCoV-19/Netherlands/Friesland_6/2020 Europe / Netherlands / Friesland 2020-03-28 Dutch COVID-19 response team Erasmus Medical Center Bas Oude Munnink, David Nieuwenhuijse, Reina Sikkema, Claudia Schapendonk, Irina Chestakova, Anne van der Linden, Theo Bestebroer, Stefan van Nieuwkoop, Mark Pronk, Pascal Lexmond, Corien Swaan, Manon Haverkate, Madelief Mollers, Mart Stein, Sandra Kengne Kamga Mobou, Jeroen van Kampen, Jolanda Voermans, Aura Timen, Corine GeurtsvanKessel, Annemiek van der Eijk, Richard Molenkamp, Marion Koopmans, on behalf of the Dutch national COVID-19 response team.
EPI_ISL_422572 hCoV-19/Netherlands/Friesland_7/2020 Europe / Netherlands / Friesland 2020-03-28 Dutch COVID-19 response team Erasmus Medical Center Bas Oude Munnink, David Nieuwenhuijse, Reina Sikkema, Claudia Schapendonk, Irina Chestakova, Anne van der Linden, Theo Bestebroer, Stefan van Nieuwkoop, Mark Pronk, Pascal Lexmond, Corien Swaan, Manon Haverkate, Madelief Mollers, Mart Stein, Sandra Kengne Kamga Mobou, Jeroen van Kampen, Jolanda Voermans, Aura Timen, Corine GeurtsvanKessel, Annemiek van der Eijk, Richard Molenkamp, Marion Koopmans, on behalf of the Dutch national COVID-19 response team.
EPI_ISL_422576 hCoV-19/Netherlands/NA_265/2020 Europe / Netherlands 2020-03-30 Dutch COVID-19 response team Erasmus Medical Center Bas Oude Munnink, David Nieuwenhuijse, Reina Sikkema, Claudia Schapendonk, Irina Chestakova, Anne van der Linden, Theo Bestebroer, Stefan van Nieuwkoop, Mark Pronk, Pascal Lexmond, Corien Swaan, Manon Haverkate, Madelief Mollers, Mart Stein, Sandra Kengne Kamga Mobou, Jeroen van Kampen, Jolanda Voermans, Aura Timen, Corine GeurtsvanKessel, Annemiek van der Eijk, Richard Molenkamp, Marion Koopmans, on behalf of the Dutch national COVID-19 response team.
EPI_ISL_422577 hCoV-19/Netherlands/NA_266/2020 Europe / Netherlands 2020-03-30 Dutch COVID-19 response team Erasmus Medical Center Bas Oude Munnink, David Nieuwenhuijse, Reina Sikkema, Claudia Schapendonk, Irina Chestakova, Anne van der Linden, Theo Bestebroer, Stefan van Nieuwkoop, Mark Pronk, Pascal Lexmond, Corien Swaan, Manon Haverkate, Madelief Mollers, Mart Stein, Sandra Kengne Kamga Mobou, Jeroen van Kampen,

Jolanda Voermans, Aura Timen, Corine GeurtsvanKessel, Annemiek van der Eijk, Richard Molenkamp, Marion Koopmans, on behalf of the Dutch national COVID-19 response team.
EPI_ISL_422578 hCoV-19/Netherlands/NA_267/2020 Europe / Netherlands 2020-03-30 Dutch COVID-19 response team Erasmus Medical Center Bas Oude Munnink, David Nieuwenhuijse, Reina Sikkema, Claudia Schapendonk, Irina Chestakova, Anne van der Linden, Theo Bestebroer, Stefan van Nieuwkoop, Mark Pronk, Pascal Lexmond, Corien Swaan, Manon Haverkate, Madelief Mollers, Mart Stein, Sandra Kengne Kamga Mobou, Jeroen van Kampen, Jolanda Voermans, Aura Timen, Corine GeurtsvanKessel, Annemiek van der Eijk, Richard Molenkamp, Marion Koopmans, on behalf of the Dutch national COVID-19 response team.
EPI_ISL_422579 hCoV-19/Netherlands/NA_268/2020 Europe / Netherlands 2020-03-30 Dutch COVID-19 response team Erasmus Medical Center Bas Oude Munnink, David Nieuwenhuijse, Reina Sikkema, Claudia Schapendonk, Irina Chestakova, Anne van der Linden, Theo Bestebroer, Stefan van Nieuwkoop, Mark Pronk, Pascal Lexmond, Corien Swaan, Manon Haverkate, Madelief Mollers, Mart Stein, Sandra Kengne Kamga Mobou, Jeroen van Kampen, Jolanda Voermans, Aura Timen, Corine GeurtsvanKessel, Annemiek van der Eijk, Richard Molenkamp, Marion Koopmans, on behalf of the Dutch national COVID-19 response team.
EPI_ISL_422580 hCoV-19/Netherlands/NA_269/2020 Europe / Netherlands 2020-03-30 Dutch COVID-19 response team Erasmus Medical Center Bas Oude Munnink, David Nieuwenhuijse, Reina Sikkema, Claudia Schapendonk, Irina Chestakova, Anne van der Linden, Theo Bestebroer, Stefan van Nieuwkoop, Mark Pronk, Pascal Lexmond, Corien Swaan, Manon Haverkate, Madelief Mollers, Mart Stein, Sandra Kengne Kamga Mobou, Jeroen van Kampen, Jolanda Voermans, Aura Timen, Corine GeurtsvanKessel, Annemiek van der Eijk, Richard Molenkamp, Marion Koopmans, on behalf of the Dutch national COVID-19 response team.
EPI_ISL_422581 hCoV-19/Netherlands/NA_270/2020 Europe / Netherlands 2020-03-30 Dutch COVID-19 response team Erasmus Medical Center Bas Oude Munnink, David Nieuwenhuijse, Reina Sikkema, Claudia Schapendonk, Irina Chestakova, Anne van der Linden, Theo Bestebroer, Stefan van Nieuwkoop, Mark Pronk, Pascal Lexmond, Corien Swaan, Manon Haverkate, Madelief Mollers, Mart Stein, Sandra Kengne Kamga Mobou, Jeroen van Kampen, Jolanda Voermans, Aura Timen, Corine GeurtsvanKessel, Annemiek van der Eijk, Richard Molenkamp, Marion Koopmans, on behalf of the Dutch national COVID-19 response team.
EPI_ISL_422582 hCoV-19/Netherlands/NA_271/2020 Europe / Netherlands 2020-03-30 Dutch COVID-19 response team Erasmus Medical Center Bas Oude Munnink, David Nieuwenhuijse, Reina Sikkema, Claudia Schapendonk, Irina Chestakova, Anne van der Linden, Theo Bestebroer, Stefan van Nieuwkoop, Mark Pronk, Pascal Lexmond, Corien Swaan, Manon Haverkate, Madelief Mollers, Mart Stein, Sandra Kengne Kamga Mobou, Jeroen van Kampen, Jolanda Voermans, Aura Timen, Corine GeurtsvanKessel, Annemiek van der Eijk, Richard Molenkamp, Marion Koopmans, on behalf of the Dutch national COVID-19 response team.
EPI_ISL_422583 hCoV-19/Netherlands/NA_272/2020 Europe / Netherlands 2020-03-30 Dutch COVID-19 response team Erasmus Medical Center Bas Oude Munnink, David Nieuwenhuijse, Reina Sikkema, Claudia Schapendonk, Irina Chestakova, Anne van der Linden, Theo Bestebroer, Stefan van Nieuwkoop, Mark Pronk, Pascal Lexmond, Corien Swaan, Manon Haverkate, Madelief Mollers, Mart Stein, Sandra Kengne Kamga Mobou, Jeroen van Kampen, Jolanda Voermans, Aura Timen, Corine GeurtsvanKessel, Annemiek van der Eijk, Richard Molenkamp, Marion Koopmans, on behalf of the Dutch national COVID-19 response team.
EPI_ISL_422585 hCoV-19/Netherlands/NA_274/2020 Europe / Netherlands 2020-03-30 Dutch COVID-19 response team Erasmus Medical Center Bas Oude Munnink, David Nieuwenhuijse, Reina Sikkema, Claudia Schapendonk, Irina Chestakova, Anne van der Linden, Theo Bestebroer, Stefan van Nieuwkoop, Mark Pronk, Pascal Lexmond, Corien Swaan, Manon Haverkate, Madelief Mollers, Mart Stein, Sandra Kengne Kamga Mobou, Jeroen van Kampen, Jolanda Voermans, Aura Timen, Corine GeurtsvanKessel, Annemiek van der Eijk, Richard Molenkamp, Marion Koopmans, on behalf of the Dutch national COVID-19 response team.
EPI_ISL_422586 hCoV-19/Netherlands/NA_275/2020 Europe / Netherlands 2020-03-30 Dutch COVID-19 response team Erasmus Medical Center Bas Oude Munnink, David Nieuwenhuijse, Reina Sikkema, Claudia Schapendonk, Irina Chestakova, Anne van der Linden, Theo Bestebroer, Stefan van Nieuwkoop, Mark Pronk, Pascal Lexmond, Corien Swaan, Manon Haverkate, Madelief Mollers, Mart Stein, Sandra Kengne Kamga Mobou, Jeroen van Kampen, Jolanda Voermans, Aura Timen, Corine GeurtsvanKessel, Annemiek van der Eijk, Richard Molenkamp, Marion Koopmans, on behalf of the Dutch national COVID-19 response team.
EPI_ISL_422587 hCoV-19/Netherlands/NA_276/2020 Europe / Netherlands 2020-03-31 Dutch COVID-19 response team Erasmus Medical Center Bas Oude Munnink, David Nieuwenhuijse, Reina Sikkema, Claudia Schapendonk, Irina Chestakova, Anne van der Linden, Theo Bestebroer, Stefan van Nieuwkoop, Mark Pronk, Pascal Lexmond, Corien Swaan, Manon Haverkate, Madelief Mollers, Mart Stein, Sandra Kengne Kamga Mobou, Jeroen van Kampen, Jolanda Voermans, Aura Timen, Corine GeurtsvanKessel, Annemiek van der Eijk, Richard Molenkamp, Marion Koopmans, on behalf of the Dutch national COVID-19 response team.
EPI_ISL_422588 hCoV-19/Netherlands/NA_277/2020 Europe / Netherlands 2020-03-31 Dutch COVID-19 response team Erasmus Medical Center Bas Oude Munnink, David Nieuwenhuijse, Reina Sikkema, Claudia Schapendonk, Irina Chestakova, Anne van der Linden, Theo Bestebroer, Stefan van Nieuwkoop, Mark Pronk, Pascal Lexmond, Corien Swaan, Manon Haverkate, Madelief Mollers, Mart Stein, Sandra Kengne Kamga Mobou, Jeroen van Kampen, Jolanda Voermans, Aura Timen, Corine GeurtsvanKessel, Annemiek van der Eijk, Richard Molenkamp, Marion Koopmans, on behalf of the Dutch national COVID-19 response team.
EPI_ISL_422591 hCoV-19/Netherlands/NA_282/2020 Europe / Netherlands 2020-03-31 Dutch COVID-19 response team Erasmus Medical Center Bas Oude Munnink, David Nieuwenhuijse, Reina Sikkema, Claudia Schapendonk, Irina Chestakova, Anne van der Linden, Theo Bestebroer, Stefan van Nieuwkoop, Mark Pronk, Pascal Lexmond, Corien Swaan, Manon Haverkate, Madelief Mollers, Mart Stein, Sandra Kengne Kamga Mobou, Jeroen van Kampen, Jolanda Voermans, Aura Timen, Corine GeurtsvanKessel, Annemiek van der Eijk, Richard Molenkamp, Marion Koopmans, on behalf of the Dutch national COVID-19 response team.
EPI_ISL_422592 hCoV-19/Netherlands/NA_283/2020 Europe / Netherlands 2020-03-31 Dutch COVID-19 response team Erasmus Medical Center Bas Oude Munnink, David Nieuwenhuijse, Reina Sikkema, Claudia Schapendonk,

Irina Chestakova, Anne van der Linden, Theo Bestebroer, Stefan van Nieuwkoop, Mark Pronk, Pascal Lexmond, Corien Swaan, Manon Haverkate, Madelief Mollers, Mart Stein, Sandra Kengne Kamga Mobou, Jeroen van Kampen, Jolanda Voermans, Aura Timen, Corine GeurtsvanKessel, Annemiek van der Eijk, Richard Molenkamp, Marion Koopmans, on behalf of the Dutch national COVID-19 response team.
EPI_ISL_422593 hCoV-19/Netherlands/NA_284/2020 Europe / Netherlands 2020-03-31 Dutch COVID-19 response team Erasmus Medical Center Bas Oude Munnink, David Nieuwenhuijse, Reina Sikkema, Claudia Schapendonk, Irina Chestakova, Anne van der Linden, Theo Bestebroer, Stefan van Nieuwkoop, Mark Pronk, Pascal Lexmond, Corien Swaan, Manon Haverkate, Madelief Mollers, Mart Stein, Sandra Kengne Kamga Mobou, Jeroen van Kampen, Jolanda Voermans, Aura Timen, Corine GeurtsvanKessel, Annemiek van der Eijk, Richard Molenkamp, Marion Koopmans, on behalf of the Dutch national COVID-19 response team.
EPI_ISL_422594 hCoV-19/Netherlands/NA_285/2020 Europe / Netherlands 2020-03-31 Dutch COVID-19 response team Erasmus Medical Center Bas Oude Munnink, David Nieuwenhuijse, Reina Sikkema, Claudia Schapendonk, Irina Chestakova, Anne van der Linden, Theo Bestebroer, Stefan van Nieuwkoop, Mark Pronk, Pascal Lexmond, Corien Swaan, Manon Haverkate, Madelief Mollers, Mart Stein, Sandra Kengne Kamga Mobou, Jeroen van Kampen, Jolanda Voermans, Aura Timen, Corine GeurtsvanKessel, Annemiek van der Eijk, Richard Molenkamp, Marion Koopmans, on behalf of the Dutch national COVID-19 response team.
EPI_ISL_422595 hCoV-19/Netherlands/NA_286/2020 Europe / Netherlands 2020-03-31 Dutch COVID-19 response team Erasmus Medical Center Bas Oude Munnink, David Nieuwenhuijse, Reina Sikkema, Claudia Schapendonk, Irina Chestakova, Anne van der Linden, Theo Bestebroer, Stefan van Nieuwkoop, Mark Pronk, Pascal Lexmond, Corien Swaan, Manon Haverkate, Madelief Mollers, Mart Stein, Sandra Kengne Kamga Mobou, Jeroen van Kampen, Jolanda Voermans, Aura Timen, Corine GeurtsvanKessel, Annemiek van der Eijk, Richard Molenkamp, Marion Koopmans, on behalf of the Dutch national COVID-19 response team.
EPI_ISL_422596 hCoV-19/Netherlands/NA_287/2020 Europe / Netherlands 2020-03-31 Dutch COVID-19 response team Erasmus Medical Center Bas Oude Munnink, David Nieuwenhuijse, Reina Sikkema, Claudia Schapendonk, Irina Chestakova, Anne van der Linden, Theo Bestebroer, Stefan van Nieuwkoop, Mark Pronk, Pascal Lexmond, Corien Swaan, Manon Haverkate, Madelief Mollers, Mart Stein, Sandra Kengne Kamga Mobou, Jeroen van Kampen, Jolanda Voermans, Aura Timen, Corine GeurtsvanKessel, Annemiek van der Eijk, Richard Molenkamp, Marion Koopmans, on behalf of the Dutch national COVID-19 response team.
EPI_ISL_422597 hCoV-19/Netherlands/NA_288/2020 Europe / Netherlands 2020-03-31 Dutch COVID-19 response team Erasmus Medical Center Bas Oude Munnink, David Nieuwenhuijse, Reina Sikkema, Claudia Schapendonk, Irina Chestakova, Anne van der Linden, Theo Bestebroer, Stefan van Nieuwkoop, Mark Pronk, Pascal Lexmond, Corien Swaan, Manon Haverkate, Madelief Mollers, Mart Stein, Sandra Kengne Kamga Mobou, Jeroen van Kampen, Jolanda Voermans, Aura Timen, Corine GeurtsvanKessel, Annemiek van der Eijk, Richard Molenkamp, Marion Koopmans, on behalf of the Dutch national COVID-19 response team.
EPI_ISL_422598 hCoV-19/Netherlands/NA_289/2020 Europe / Netherlands 2020-03-31 Dutch COVID-19 response team Erasmus Medical Center Bas Oude Munnink, David Nieuwenhuijse, Reina Sikkema, Claudia Schapendonk, Irina Chestakova, Anne van der Linden, Theo Bestebroer, Stefan van Nieuwkoop, Mark Pronk, Pascal Lexmond, Corien Swaan, Manon Haverkate, Madelief Mollers, Mart Stein, Sandra Kengne Kamga Mobou, Jeroen van Kampen, Jolanda Voermans, Aura Timen, Corine GeurtsvanKessel, Annemiek van der Eijk, Richard Molenkamp, Marion Koopmans, on behalf of the Dutch national COVID-19 response team.
EPI_ISL_422599 hCoV-19/Netherlands/NA_290/2020 Europe / Netherlands 2020-03-31 Dutch COVID-19 response team Erasmus Medical Center Bas Oude Munnink, David Nieuwenhuijse, Reina Sikkema, Claudia Schapendonk, Irina Chestakova, Anne van der Linden, Theo Bestebroer, Stefan van Nieuwkoop, Mark Pronk, Pascal Lexmond, Corien Swaan, Manon Haverkate, Madelief Mollers, Mart Stein, Sandra Kengne Kamga Mobou, Jeroen van Kampen, Jolanda Voermans, Aura Timen, Corine GeurtsvanKessel, Annemiek van der Eijk, Richard Molenkamp, Marion Koopmans, on behalf of the Dutch national COVID-19 response team.
EPI_ISL_422634 hCoV-19/Netherlands/ZuidHolland_98/2020 Europe / Netherlands / Zuid Holland 2020-03-31 Dutch COVID-19 response team Erasmus Medical Center Bas Oude Munnink, David Nieuwenhuijse, Reina Sikkema, Claudia Schapendonk, Irina Chestakova, Anne van der Linden, Theo Bestebroer, Stefan van Nieuwkoop, Mark Pronk, Pascal Lexmond, Corien Swaan, Manon Haverkate, Madelief Mollers, Mart Stein, Sandra Kengne Kamga Mobou, Jeroen van Kampen, Jolanda Voermans, Aura Timen, Corine GeurtsvanKessel, Annemiek van der Eijk, Richard Molenkamp, Marion Koopmans, on behalf of the Dutch national COVID-19 response team.
EPI_ISL_422655 hCoV-19/Netherlands/NA_106/2020 Europe / Netherlands 2020-03-18 Dutch COVID-19 response team Erasmus Medical Center Bas Oude Munnink, David Nieuwenhuijse, Reina Sikkema, Claudia Schapendonk, Irina Chestakova, Anne van der Linden, Theo Bestebroer, Stefan van Nieuwkoop, Mark Pronk, Pascal Lexmond, Corien Swaan, Manon Haverkate, Madelief Mollers, Mart Stein, Sandra Kengne Kamga Mobou, Jeroen van Kampen, Jolanda Voermans, Aura Timen, Corine GeurtsvanKessel, Annemiek van der Eijk, Richard Molenkamp, Marion Koopmans, on behalf of the Dutch national COVID-19 response team.
EPI_ISL_422656 hCoV-19/Netherlands/NA_107/2020 Europe / Netherlands 2020-03-18 Dutch COVID-19 response team Erasmus Medical Center Bas Oude Munnink, David Nieuwenhuijse, Reina Sikkema, Claudia Schapendonk, Irina Chestakova, Anne van der Linden, Theo Bestebroer, Stefan van Nieuwkoop, Mark Pronk, Pascal Lexmond, Corien Swaan, Manon Haverkate, Madelief Mollers, Mart Stein, Sandra Kengne Kamga Mobou, Jeroen van Kampen, Jolanda Voermans, Aura Timen, Corine GeurtsvanKessel, Annemiek van der Eijk, Richard Molenkamp, Marion Koopmans, on behalf of the Dutch national COVID-19 response team.
EPI_ISL_422657 hCoV-19/Netherlands/NA_109/2020 Europe / Netherlands 2020-03-18 Dutch COVID-19 response team Erasmus Medical Center Bas Oude Munnink, David Nieuwenhuijse, Reina Sikkema, Claudia Schapendonk, Irina Chestakova, Anne van der Linden, Theo Bestebroer, Stefan van Nieuwkoop, Mark Pronk, Pascal Lexmond, Corien Swaan, Manon Haverkate, Madelief Mollers, Mart Stein, Sandra Kengne Kamga Mobou, Jeroen van Kampen, Jolanda Voermans, Aura Timen, Corine GeurtsvanKessel, Annemiek van der Eijk, Richard Molenkamp, Marion Koopmans, on behalf of the Dutch national COVID-19 response team.

EPI_ISL_422659 hCoV-19/Netherlands/NA_111/2020 Europe / Netherlands 2020-03-15 Dutch COVID-19 response team Erasmus Medical Center Bas Oude Munnink, David Nieuwenhuijse, Reina Sikkema, Claudia Schapendonk, Irina Chestakova, Anne van der Linden, Theo Bestebroer, Stefan van Nieuwkoop, Mark Pronk, Pascal Lexmond, Corien Swaan, Manon Haverkate, Madelief Mollers, Mart Stein, Sandra Kengne Kamga Mobou, Jeroen van Kampen, Jolanda Voermans, Aura Timen, Corine GeurtsvanKessel, Annemiek van der Eijk, Richard Molenkamp, Marion Koopmans, on behalf of the Dutch national COVID-19 response team.
EPI_ISL_422660 hCoV-19/Netherlands/NA_112/2020 Europe / Netherlands 2020-03-10 Dutch COVID-19 response team Erasmus Medical Center Bas Oude Munnink, David Nieuwenhuijse, Reina Sikkema, Claudia Schapendonk, Irina Chestakova, Anne van der Linden, Theo Bestebroer, Stefan van Nieuwkoop, Mark Pronk, Pascal Lexmond, Corien Swaan, Manon Haverkate, Madelief Mollers, Mart Stein, Sandra Kengne Kamga Mobou, Jeroen van Kampen, Jolanda Voermans, Aura Timen, Corine GeurtsvanKessel, Annemiek van der Eijk, Richard Molenkamp, Marion Koopmans, on behalf of the Dutch national COVID-19 response team.
EPI_ISL_422661 hCoV-19/Netherlands/NA_113/2020 Europe / Netherlands 2020-03-10 Dutch COVID-19 response team Erasmus Medical Center Bas Oude Munnink, David Nieuwenhuijse, Reina Sikkema, Claudia Schapendonk, Irina Chestakova, Anne van der Linden, Theo Bestebroer, Stefan van Nieuwkoop, Mark Pronk, Pascal Lexmond, Corien Swaan, Manon Haverkate, Madelief Mollers, Mart Stein, Sandra Kengne Kamga Mobou, Jeroen van Kampen, Jolanda Voermans, Aura Timen, Corine GeurtsvanKessel, Annemiek van der Eijk, Richard Molenkamp, Marion Koopmans, on behalf of the Dutch national COVID-19 response team.
EPI_ISL_422662 hCoV-19/Netherlands/NA_114/2020 Europe / Netherlands 2020-03-10 Dutch COVID-19 response team Erasmus Medical Center Bas Oude Munnink, David Nieuwenhuijse, Reina Sikkema, Claudia Schapendonk, Irina Chestakova, Anne van der Linden, Theo Bestebroer, Stefan van Nieuwkoop, Mark Pronk, Pascal Lexmond, Corien Swaan, Manon Haverkate, Madelief Mollers, Mart Stein, Sandra Kengne Kamga Mobou, Jeroen van Kampen, Jolanda Voermans, Aura Timen, Corine GeurtsvanKessel, Annemiek van der Eijk, Richard Molenkamp, Marion Koopmans, on behalf of the Dutch national COVID-19 response team.
EPI_ISL_422664 hCoV-19/Netherlands/NA_117/2020 Europe / Netherlands 2020-03-16 Dutch COVID-19 response team Erasmus Medical Center Bas Oude Munnink, David Nieuwenhuijse, Reina Sikkema, Claudia Schapendonk, Irina Chestakova, Anne van der Linden, Theo Bestebroer, Stefan van Nieuwkoop, Mark Pronk, Pascal Lexmond, Corien Swaan, Manon Haverkate, Madelief Mollers, Mart Stein, Sandra Kengne Kamga Mobou, Jeroen van Kampen, Jolanda Voermans, Aura Timen, Corine GeurtsvanKessel, Annemiek van der Eijk, Richard Molenkamp, Marion Koopmans, on behalf of the Dutch national COVID-19 response team.
EPI_ISL_422665 hCoV-19/Netherlands/NA_118/2020 Europe / Netherlands 2020-03-16 Dutch COVID-19 response team Erasmus Medical Center Bas Oude Munnink, David Nieuwenhuijse, Reina Sikkema, Claudia Schapendonk, Irina Chestakova, Anne van der Linden, Theo Bestebroer, Stefan van Nieuwkoop, Mark Pronk, Pascal Lexmond, Corien Swaan, Manon Haverkate, Madelief Mollers, Mart Stein, Sandra Kengne Kamga Mobou, Jeroen van Kampen, Jolanda Voermans, Aura Timen, Corine GeurtsvanKessel, Annemiek van der Eijk, Richard Molenkamp, Marion Koopmans, on behalf of the Dutch national COVID-19 response team.
EPI_ISL_422666 hCoV-19/Netherlands/NA_119/2020 Europe / Netherlands 2020-03-16 Dutch COVID-19 response team Erasmus Medical Center Bas Oude Munnink, David Nieuwenhuijse, Reina Sikkema, Claudia Schapendonk, Irina Chestakova, Anne van der Linden, Theo Bestebroer, Stefan van Nieuwkoop, Mark Pronk, Pascal Lexmond, Corien Swaan, Manon Haverkate, Madelief Mollers, Mart Stein, Sandra Kengne Kamga Mobou, Jeroen van Kampen, Jolanda Voermans, Aura Timen, Corine GeurtsvanKessel, Annemiek van der Eijk, Richard Molenkamp, Marion Koopmans, on behalf of the Dutch national COVID-19 response team.
EPI_ISL_422667 hCoV-19/Netherlands/NA_120/2020 Europe / Netherlands 2020-03-16 Dutch COVID-19 response team Erasmus Medical Center Bas Oude Munnink, David Nieuwenhuijse, Reina Sikkema, Claudia Schapendonk, Irina Chestakova, Anne van der Linden, Theo Bestebroer, Stefan van Nieuwkoop, Mark Pronk, Pascal Lexmond, Corien Swaan, Manon Haverkate, Madelief Mollers, Mart Stein, Sandra Kengne Kamga Mobou, Jeroen van Kampen, Jolanda Voermans, Aura Timen, Corine GeurtsvanKessel, Annemiek van der Eijk, Richard Molenkamp, Marion Koopmans, on behalf of the Dutch national COVID-19 response team.
EPI_ISL_422668 hCoV-19/Netherlands/NA_121/2020 Europe / Netherlands 2020-03-17 Dutch COVID-19 response team Erasmus Medical Center Bas Oude Munnink, David Nieuwenhuijse, Reina Sikkema, Claudia Schapendonk, Irina Chestakova, Anne van der Linden, Theo Bestebroer, Stefan van Nieuwkoop, Mark Pronk, Pascal Lexmond, Corien Swaan, Manon Haverkate, Madelief Mollers, Mart Stein, Sandra Kengne Kamga Mobou, Jeroen van Kampen, Jolanda Voermans, Aura Timen, Corine GeurtsvanKessel, Annemiek van der Eijk,
