## Supplementary material for "Emergence of Novel SARS-CoV-2 Variants in the Netherlands": supplementary-file-3.pdf

Aysun Urhan1, Thomas Abeel1,2,*
1 Delft Bioinformatics Lab, Delft University of Technology Van Mourik Broekmanweg 6, 2628 XE Delft, The Netherlands
2 Infectious Disease and Microbiome Program, Broad Institute of MIT and Harvard, 415 Main Street, Cambridge, MA 02142, USA

MT020781
MT049951
MT066156
MT077125
MT123292
MT123293
MT126808
MT192765
MT226610
MT233526
MT246667
MT256924
MT263459
MT276324
MT276325
MT276328
MT276597
MT291826
MT291827
MT291828
MT291829
MT291830
MT292569
MT292570
MT292571
MT292572
MT292573
MT292574
MT292575
MT293156
MT293158
MT293159
MT293160
MT293161
MT293162
MT293163
MT293164
MT293165
MT293166
MT293167
MT293168
MT293169
MT293170
MT293171
MT293172
MT293173
MT293174
MT293175
MT293176
MT293177
MT293178
MT293179
MT293181
MT293182
MT293183
MT293184
MT293185
MT293186
MT293187
MT293188
MT293189

MT293190
MT293191
MT293192
MT293194
MT293195
MT293196
MT293197
MT293198
MT293199
MT293200
MT293201
MT293202
MT293204
MT293205
MT293206
MT293207
MT293208
MT293209
MT293210
MT293211
MT293212
MT293213
MT293215
MT293216
MT293218
MT293219
MT293220
MT293222
MT293224
MT293225
MT300186
MT308702
MT308703
MT308704
MT318827
MT320538
MT320891
MT322396
MT322397
MT325561
MT325562
MT325563
MT325564
MT325565
MT325566
MT325567
MT325568
MT325569
MT325570
MT325571
MT325572
MT325573
MT325574
MT325575
MT325576
MT325577
MT325578
MT325579
MT325580
MT325581
MT325582
MT325583
MT325584
MT325585
MT325586
MT325587
MT325588
MT325589
MT325590
MT325591

MT325592
MT325593
MT325594
MT325595
MT325596
MT325597
MT325598
MT325599
MT325600
MT325601
MT325602
MT325603
MT325604
MT325605
MT325606
MT325607
MT325608
MT325609
MT325610
MT325611
MT325612
MT325613
MT325614
MT325615
MT325616
MT325617
MT325618
MT325619
MT325620
MT325621
MT325622
MT325623
MT325624
MT325625
MT325626
MT325627
MT325628
MT325629
MT325630
MT325631
MT325632
MT325633
MT325634
MT325635
MT325636
MT325637
MT325638
MT325639
MT325640
MT327745
MT328032
MT328033
MT328034
MT328035
MT334523
MT334526
MT345802
MT345816
MT345868
MT345869
MT345870
MT345873
MT345874
MT350263
MT350264
MT350265
MT350266
MT350267
MT350268
MT350269

MT350270
MT350271
MT350272
MT350273
MT350274
MT350275
MT350276
MT350277
MT350278
MT350279
MT350280
MT358401
MT358402
MT371002
MT374109
MT374110
MT374111
MT374112
MT374113
MT374114
MT374115
MT374116
MT380728
MT380729
MT380730
MT380731
MT380732
MT380733
MT380734
MT412310
MT415320
MT415321
MT415322
MT415323
MT444148
MT446339
MT452574
MT452575
MT452576
MT457403
MT459837
MT459838
MT459839
MT459840
MT459841
MT459842
MT459843
MT459844
MT459845
MT459846
MT459847
MT459849
MT459850
MT459851
MT459852
MT459853
MT459854
MT459855
MT459856
MT459857
MT459858
MT459859
MT459860
MT459862
MT459863
MT459865
MT459866
MT459867
MT459868
MT459869

MT459898
MT459906
MT459985
MT459986
MT459987
MT459988
MT459989
MT459990
MT460089
MT460090
MT460091
MT460092
MT460113
MT460114
MT460115
MT460116
MT460117
MT460119
MT460120
MT460121
MT460124
MT460125
MT460127
MT460128
MT460129
MT460130
MT460132
MT460133
MT460135
MT460136
MT460137
MT460138
MT460139
MT460140
MT466583
MT477835
MT477836
MT477837
MT477838
MT477839
MT477840
MT477841
MT477842
MT477843
MT477844
MT477845
MT477846
MT477847
MT477848
MT477849
MT477850
MT477851
MT477852
MT477853
MT477854
MT477855
MT477856
MT477857
MT477858
MT477859
MT477860
MT477861
MT477862
MT477885
MT477903
MT477904
MT483553
MT483554
MT483555
MT483556

MT483557
MT483558
MT483559
MT483560
MT483563
MT483702
MT506707
MT506888
MT506889
MT506891
MT506892
MT506894
MT506896
MT506897
MT506899
MT506900
MT506902
MT506903
MT506904
MT506905
MT506907
MT509452
MT509453
MT509454
MT509455
MT509456
MT509457
MT509458
MT509459
MT509460
MT509461
MT509462
MT509463
MT509464
MT509465
MT509466
MT509467
MT509468
MT509471
MT509472
MT509474
MT509475
MT509477
MT509480
MT509486
MT509488
MT509489
MT509490
MT509491
MT509492
MT509493
MT510718
MT510719
MT510721
MT510722
MT510725
MT510726
MT528598
MT528599
MT528600
MT528601
MT528602
MT528603
MT528604
MT528605
MT528606
MT528607
MT528608
MT528609
MT528610

MT528611
MT528612
MT528613
MT528614
MT528615
MT528616
MT528617
MT528618
MT528619
MT528622
MT528623
MT528624
MT528625
MT528626
MT528627
MT528628
MT528629
MT528630
MT533201
MT535472
MT535481
MT539726
MT551604
MT551605
MT551606
MT551607
MT551608
MT551609
MT554052
MT557568
MT557569
MT557570
MT559037
MT559038
MT559800
MT559801
MT559802
MT559803
MT565495
MT565496
MT565497
MT565498
MT565499
MT566435
MT566436
MT566437
MT576529
MT576530
MT576531
MT576532
MT576639
MT576640
MT576641
MT576642
MT576643
MT576644
MT576689
MT578015
MT578016
MT578017
MT598171
MT598172
MT598173
MT598174
MT598175
MT598176
MT598177
MT598178
MT598633
MT598634

MT598636
MT598639
MT598640
MT598641
MT598642
MT601275
MT601276
MT601277
MT601278
MT601279
MT601280
MT601281
MT601282
MT601283
MT601284
MT601285
MT601286
MT601287
MT601288
MT601289
MT601290
MT601291
MT601292
MT601293
MT601294
MT601295
NC_045512
NMDC60013002-06
NMDC60013002-07
NMDC60013002-08
NMDC60013002-09
NMDC60013002-10
CNA0013697
CNA0013698
CNA0013699
CNA0013700
CNA0013701
CNA0013702
CNA0013703
CNA0013704
CNA0013705
CNA0013708
